# Emergence of N antigen SARS-CoV-2 genetic variants escaping detection of antigenic tests

**DOI:** 10.1101/2021.03.25.21253802

**Authors:** Claudia Del Vecchio, Giuseppina Brancaccio, Alessandra Rosalba Brazzale, Enrico Lavezzo, Francesco Onelia, Elisa Franchin, Laura Manuto, Federico Bianca, Vito Cianci, Annamaria Cattelan, Stefano Toppo, Andrea Crisanti

## Abstract

SARS-CoV-2 genetic variants are emerging as a major threat to vaccination efforts worldwide as they may increase virus transmission rate and/or confer the ability to escape vaccine induced immunity with knock on effects on the level of herd immunity and vaccine efficacy respectively. These variants concern the Spike protein, which is encoded by the S gene, involved in virus entry into host cells and the major target of vaccine development. We report here that genetic variants of the N gene can impair our ability to utilize antigenic tests for both diagnosis and mass testing efforts aimed at controlling virus transmission. While conducting a large validation study on the Abbott Panbio™ COVID-19 Ag test, we noticed that some swab samples failed to generate a positive result in spite of a high viral load in Rt-PCR assays. Sequencing analysis of viruses showing discordant results in the RT-PCR and antigen assays revealed the presence of multiple disruptive amino-acid substitutions in the N antigen (the viral protein detected in the antigen test) clustered from position 229 to 374 a region known to contain an immunodominant epitope. A relevant fraction of the variants, undetected by the antigen test, contained the mutations A376T coupled to M241I. Intriguingly we found that virus sequences with this mutation were over-represented in the antigen-test-negative and PCR-positive samples and progressively increased in frequency over time in Veneto, a region of Italy that has aggressively scaled up the utilization of antigen tests, which reached nearly 68% of all the SARS-CoV-2 swab assays performed there. We speculate that mass utilization of antigen assays could create a selection pressure on the target that may favor the spread of undetectable virus variants.

## INTRODUCTION

SARS-CoV-2 rapid antigen tests are immunoassays designed to identify in the nasopharyngeal or nasal swab the presence of a viral antigens thereby corroborating the diagnosis of the infection. Typically, these assays utilize immuno-chromatographic lateral flow design and high affinity monoclonal antibodies against an abundant protein that in the case of SARS-CoV-2 is the N antigen^1^. On August 26, 2020, the U.S. Food and Drug Administration (FDA) has granted emergency use authorization (EUA) for rapid antigen tests that can identify SARS-CoV-2, thereafter most countries followed suit ^2^. The major advantage of antigen tests is their flexibility and ease of use. They can be performed in local settings with no convenient access to a molecular biology laboratory. They generate a result in about 15 minutes but require complex logistics when thousands of individuals are tested on the same occasion as they lack both automation and parallelism. The main limitation of SARS-CoV-2 antigen tests is sensitivity. These tests are less sensitive than real-time reverse transcription polymerase chain reaction (RT-PCR) and other nucleic amplification technology ^3^. Preliminary results on the Liverpool pilot study with Innova lateral flow test (LFT) indicated that this assay had a sensitivity of 66% ^4^ and an even lower 49% ^5^ sensitivity was reported in other community testing in UK. A screening with the same test performed on the students at Birmingham University reported a worryingly low sensitivity of 3.2% amongst asymptomatic young individuals ^4,6^. It has been argued that this discrepancy was possibly due to the lower viral loads of Birmingham students compared with those examined at Liverpool.

Rapid antigenic tests have been proposed as a flexible and powerful tool to implement effective surveillance regimens aimed at reducing virus prevalence in large target communities and to reduce transmission from asymptomatic individuals. It has been argued that if the test is repeated at least a couple of times on the same person at two-three days interval the sensitivity will increases because of the higher probability to intercept the peak of viral load in an infected individual ^7^. Accordingly, a negative result on multiple occasions would indicate that the individual is a true negative or is in the declining phase of viral multiplication and therefore unlikely to be infectious. During October 2020 almost the entire adult population of Slovakia was tested using SARS-CoV-2 antigen assays ^8^. In an unprecedented logistic effort 3.3 million individuals were tested in few days with a positivity rate of about 1% ^8^. To minimize sensitivity problems the test was repeated a week later in regions with high prevalence. Unfortunately, no data is available on the overall sensitivity of the test utilized there under those epidemiological conditions. However, a few weeks after the testing was completed hospitalization plateaued suggesting that this effort may have helped to temporarily reduce transmission ^8^. More recently massive population screenings with antigen tests have been carried out in different regions of the world including Italy ^9,10^. Since the beginning of October, the Regione Veneto, one of the twenty-one administrative districts of Italy, has progressively scaled up the use of antigen tests in the effort to expand its testing and tracing capability. Out of 226,904 tests performed from the 15^th^ to the 22^th^ of February 2021, 153,020 were antigen swabs accounting for 67.4% of the total, the great majority of the them being manufactured by Abbott (Panbio™ COVID-19 Ag Rapid Test Device, ABBOTT Lake Country, IL, U.S.A.). Shortly after the introduction of this test as a screening tool for surveillance monitoring purposes country wide, we conducted a validation study to evaluate its overall performance and to assess its suitability to rapidly identify SARS-CoV-2 infections in patients admitted to hospital. We compared the performance of this test with RT-PCR on 1,441 patients examined at the A&E and the Infectious Diseases wards over a period of one month. We also systematically analysed the sequence of several viral samples that generated non concordant results in the antigen test and RT-PCR to discriminate between lack of sensitivity and the presence of virus variants undetectable to the antigen tests.

## METHODS

### Study design

The samples were collected from the 15th of September to the 16th of October 2020 from patients admitted to the A&E and Infectious disease wards of the University Hospital of Padua and from subjects who required SARS-CoV-2 testing for the following reasons: a) presence of symptoms indicating a possible SARS-CoV-2 infection (fever and/or cough and/or headache, diarrhea, asthenia, muscle pain, joint pain, loss of taste or smell, or shortness of breath, with or without pneumonia); and b) patients who were asymptomatic but had a contact with a confirmed case of SARS-CoV-2 infection during the previous ten days. All subjects were tested within one hour with both antigen (ANCOV, Panbio™ COVID-19 Ag Rapid Test Device, ABBOTT Lake Country, IL, U.S.A.) and molecular (DNCOV, Simplexa™ COVID-19 Direct Kit, Diasorin Cypress, CA, U.S.A.) swab assay. Information was collected from each individual about age, sex, date of sampling, symptoms, time interval from the onset of symptoms and the test, Ct value of the molecular test. The DNCOV and ANCOV test results were grouped as concordant (DNCOV+/ANCOV+) or discordant (DNCOV+/ANCOV-).

### Testing for SARS-CoV-2 with antigenic and molecular assays

Two swabs were collected from each patient. One sample was processed with Panbio™ COVID-19 Ag Rapid Test right after sampling according to manufacturer’s instructions. The second swab was processed with real time RT-PCR, DiaSorin Molecular Simplexa™ COVID-19 Direct assay system (Diasorin Cypress, CA, U.S.A.) which amplifies two targets of the SARS-CoV-2 genome (the S gene and the ORF1ab gene). The assay also reveals the presence of host mRNA in the same reaction to confirm the correct execution of the test. Samples showing a positive result for both viral targets were considered positive. Samples with either a single positive target or with Ct value ≥30 were confirmed with an in-house real-time RT-PCR targeting the N2 gene ^11^.

### Viral Culture

Viral isolation was carried out within a biosafety-level 3 (BSL3) containment facility. Typically, 100 µl of not inactivated swab medium were inoculated in monolayers of Vero cells (ATCC CCL-81) with Dulbecco’s Modified Eagle Medium (DMEM) (Gibco) supplemented with 2% (v/v) of fetal bovine serum (FBS) (Gibco) and 1% (v/v) of penicillin/streptomycin (Gibco) and incubated at 37°C with 5% CO2. 24 hours after inoculation culture supernatant was replaced with fresh culture medium. The supernatant was collected, centrifuged and the presence of SARS-CoV-2 confirmed by in-house real-time RT-PCR targeting the N2 gene. Some of the supernatants were evaluated with. Supernatant were also tested with Panbio™ COVID-19 Ag Rapid Test, COVID-19 Ag Respi-Strip (Coris BioConcept, Belgium) as well as with LumiraDX SARS-CoV-2.

### Synthesis of cDNA and library preparation protocol

RNA samples were treated with DNase using the DNA-freeTM Kit (Ambion, Life Technologies) following the manufacturer’s instructions. The cDNA was synthetised using the ProtoScript® II First Strand cDNA Synthesis kit (New England Biolabs Inc.). The first strand synthesis reaction was performed in a total volume of 20µl at 95°C for 5 minutes, then the temperature ramped down to 20°C. The cDNA synthesis was completed at 25°C for 5 minutes followed by 1 hour at 42°C and 5 minutes at 80°C. The cDNA libraries were prepared following the Twist Library Preparation Kit for ssRNA Virus Detection protocol (Twist Bioscience) and fragmented to generate 300 bp dA-tailed DNA fragments. The Twist Universal Adapters were ligated to the dA-tailed DNA fragments to prepare the cDNA libraries ready for indexing. The samples were amplified using the Twist Unique Dual Index Primers for 10 cycles, set up as follows: denaturation step at 98°C for 25 seconds; annealing step at 60°C for 30 seconds and extension step at 72°C for 30 seconds. The PCR products were then purified using the DNA Purification Beads (Twist Bioscience).

### Viral cDNA enrichment protocol and sequencing

The libraries enrichment was performed according to the Twist Target Enrichment protocol (Twist Bioscience). The samples were pooled in groups of four to a final concentration of 1,500 ng. The hybridisation reaction to SARS-CoV-2 specific probes was performed by incubating the library pools and the probes at 70°C for 16 hours. The hybridised targets were then captured by streptavidin beads following the manufacturer’s instructions. The library pools were then enriched through a post-capture PCR. The amplification products were purified using the DNA Purification beads (Twist Bioscience). After quantification and validation check, libraries were normalised and sequenced 2×150 paired-end on both the Illumina MiSeq and NextSeq 550 platforms at the Polo GGB facility (Siena, Italy). The sequencing was performed using the Standard V2 and the Mid Output V2.5 flowcells, respectively.

### Quality check and mapping of the reads

Raw sequences were filtered for length and quality with Trimmomatic v0.40 ^12^ according to the following parameters: ILLUMINACLIP:TruSeq3-PE-2:2:30:10 LEADING:30 TRAILING:30 SLIDINGWINDOW:4:20 MINLEN:90. High quality reads were aligned on the SARS-CoV-2 reference genome (genbank ACC: NC_045512) with BWA-MEM v0.7.17 ^13^. Duplicated reads were then removed with Picard tool v2.25.0 (http://broadinstitute.github.io/picard/). Consensus sequence were generated using a combination of SAMtools v1.11 ^14^ and VarScan v2.4.1 ^15^ variant caller. Consensus sequences were reconstructed from VarScan output with an in-house script that automatically introduce ‘N’ in low quality or uncertain/uncovered regions of the reference sequence. Sequence details are available on **Suppl. Mat**.

### Analysis of the amino acid variants in N protein

Sequence data and corresponding metadata, released from the 17th of February 2021, were downloaded from GISAID databank ^16^ and further analyzed to map the variants in N protein. Sequences with missing information and incomplete N protein were discarded. The analysis was conducted on the variants found in concordant (DNCOV+/ANCOV+) and discordant cases (DNCOV+/ANCOV-) (see **Suppl. Mat**. and **GISAID acknowledgments** where the list of accession numbers of considered sequences is available). The monthly prevalence trend of the variants found in concordant and discordant cases was plotted from January to December 2020 and reported for Europe, Italy, and the Veneto region (**Figure 7**).

### Statistical analysis

P-values are evaluated at the 5% significance level; confidence intervals were computed with a confidence level of 95%. Continuous outcomes were compared using the two-sided Wilcoxon-Mann-Whitney test (two groups) and the Kruskall-Wallis test (> two groups) ^17^. Pearson’s chi-squared test statistic was used to compare proportions. The distribution of Ct values was estimated using a two-component Gaussian mixture. Logistic regression is used to model ANCOV test sensitivity against Ct level.

## RESULTS

### Analysis of antigen assay performance

A total of 1,441 subjects representing 44% of all patients examined (3,290) from the 15^th^ of September to the 16^th^ of October in the A&E and the Infectious Diseases wards were tested within an hour with both antigen (ANCOV, Abbott) and molecular (DNCOV, Simplexa™ COVID-19 Direct Kit, Diasorin Cypress, CA, U.S.A.) tests (**Figure 1**). The antigen test failed to correctly identify the presence of SARS-CoV-2 in 19 out 61 samples that showed a clear positive signal in RT-PCR against both the S and ORF1 viral sequences. Compared to RT PCR the Panbio™ COVID-19 Ag test showed an overall specificity of 99.9% (95% CI: 99.6 – 100) a sensitivity of 68.9% (95% CI: 55.7 – 80.1) and a positive predictive value of 82.7% (95% CI: 40.1 – 97.2) at P= 0.005 and of 48.7% (95% CI: 11.7 – 87.2) at P=0.001. At Ct values below 33 for both the S and ORF1 genes the sensitivity of the antigen test increased to 77.8 (95% CI: 64.4 – 88.0) and 79.2 (95% CI: 65.9 – 89.2) respectively (**Table I**). We performed a logistic regression analysis to estimate the sensitivity of the antigen test at different levels of Ct values for both the S and ORF1 gene showing that the sensitivity of the test increases at progressively lowering Ct (**Table II** and **Figure 2**). The analysis of the distribution values of the discordant DNCOV+/ANCOV- samples identified for both the S and ORF1 antigens two well separate groups with only slightly overlapping normal distributions (**Figure 3**). One group of 6 DNCOV+/ANCOV- samples showed Ct values for the S gene ranging from 33 to 38, above the sensitivity threshold declared by the manufacturer. The other group of 12 DNCOV+/ANCOV- samples showed Ct values ranging from 17 to 32, within the declared sensitivity performance of the test. A similar distribution was observed when using the ORF1 gene as positive reference with 2 DNCOV+/ANCOV- samples with Ct of 37-38 above the threshold and 11 samples with Ct ranging from 20.2 to 32.5 within the sensitivity of the antigen assay. Five samples were only analysed for the S gene and are not included in this analysis. Welch Two Sample t-tests revealed that the distributions of Ct values of concordant and discordant assays are not equal at the 5% significance level, with all test results having p-value < 0.001 (**Table III**). The occurrence of a number of DNCOV+/ANCOV- discordant samples with values well below the threshold of the Abbott test sensitivity raises questions about the occasional occurrence of genetic variants of the viral N gene that are not detected by the capture antibody reagent.

**Figure 1:**
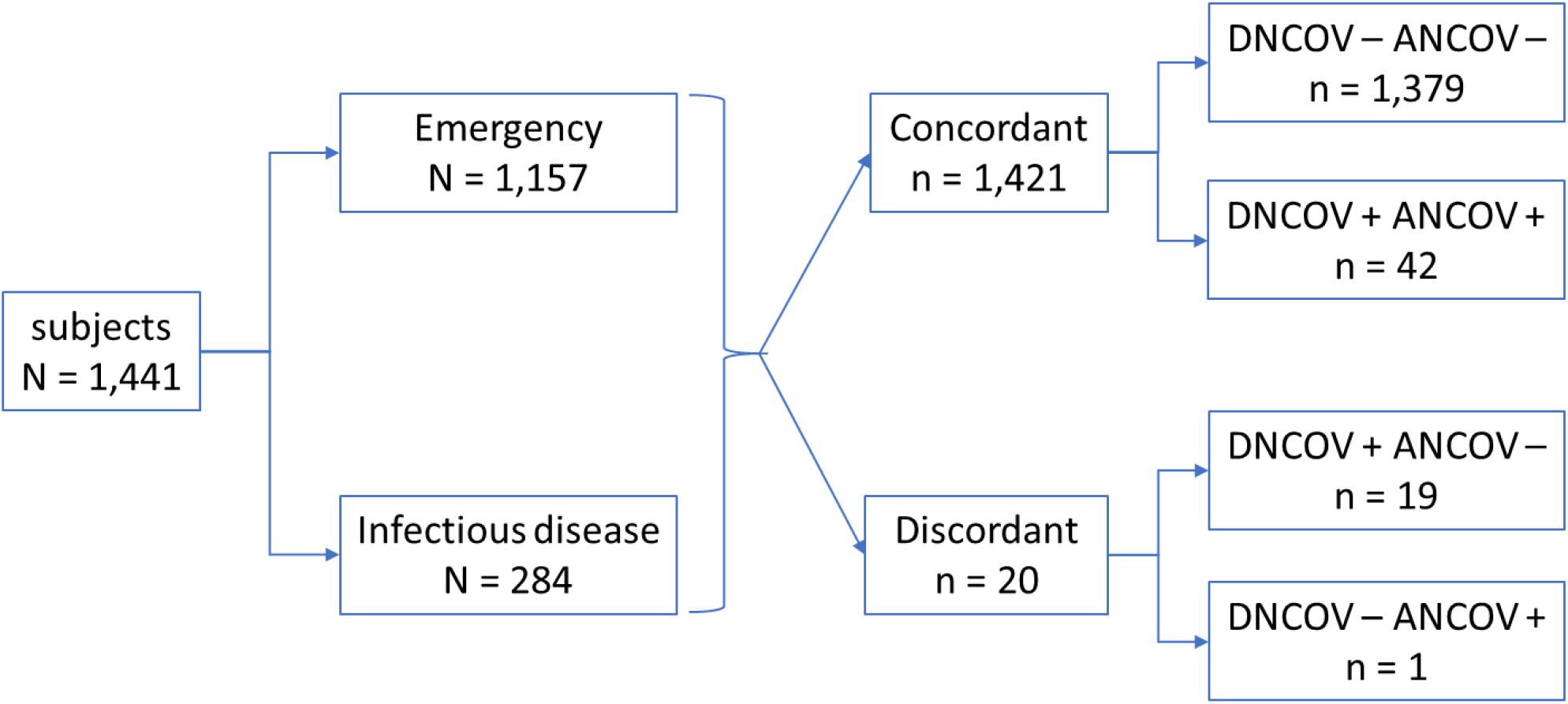
Diagram showing the total number of patients tested with both molecular (DNCOV) and antigen (ANCOV) swabs from A&E and the Infectious Diseases wards in the period from the 15^th^ of September to the 16^th^ of October 2020 and further arranged in four groups according to test result concordant (DNCOV-/+ANCOV- /+).

**Figure 2:**
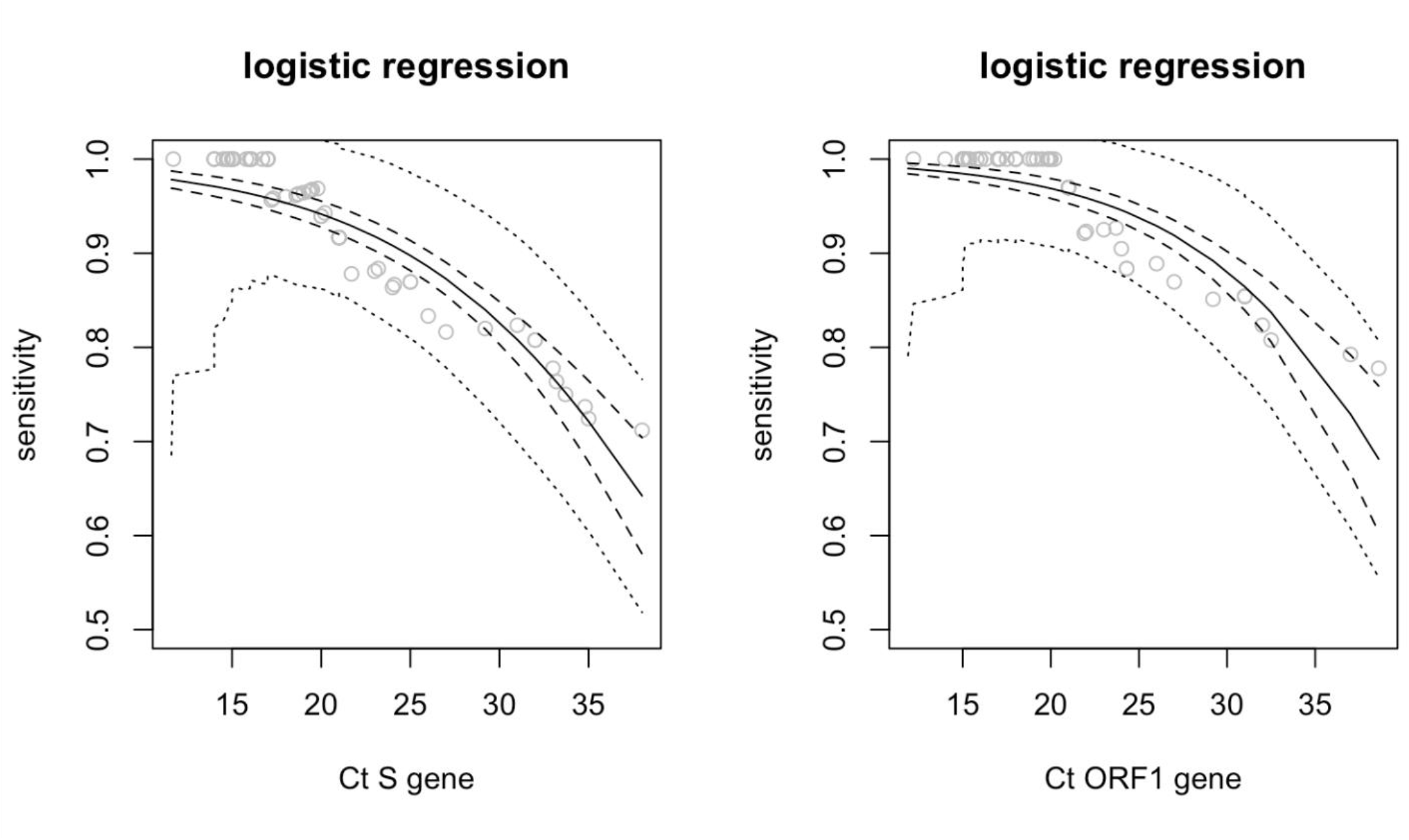
Sensitivity estimates (solid line) of ANCOV test for increasing Ct values of the S gene (left panel) and Ct ORF1 (right panel) gene as obtained from a logistic regression model. The dashed lines give the 95% confidence band for the fitted regression line. The dotted lines represent the 95% band for the fitted sensitivities.

**Figure 3:**
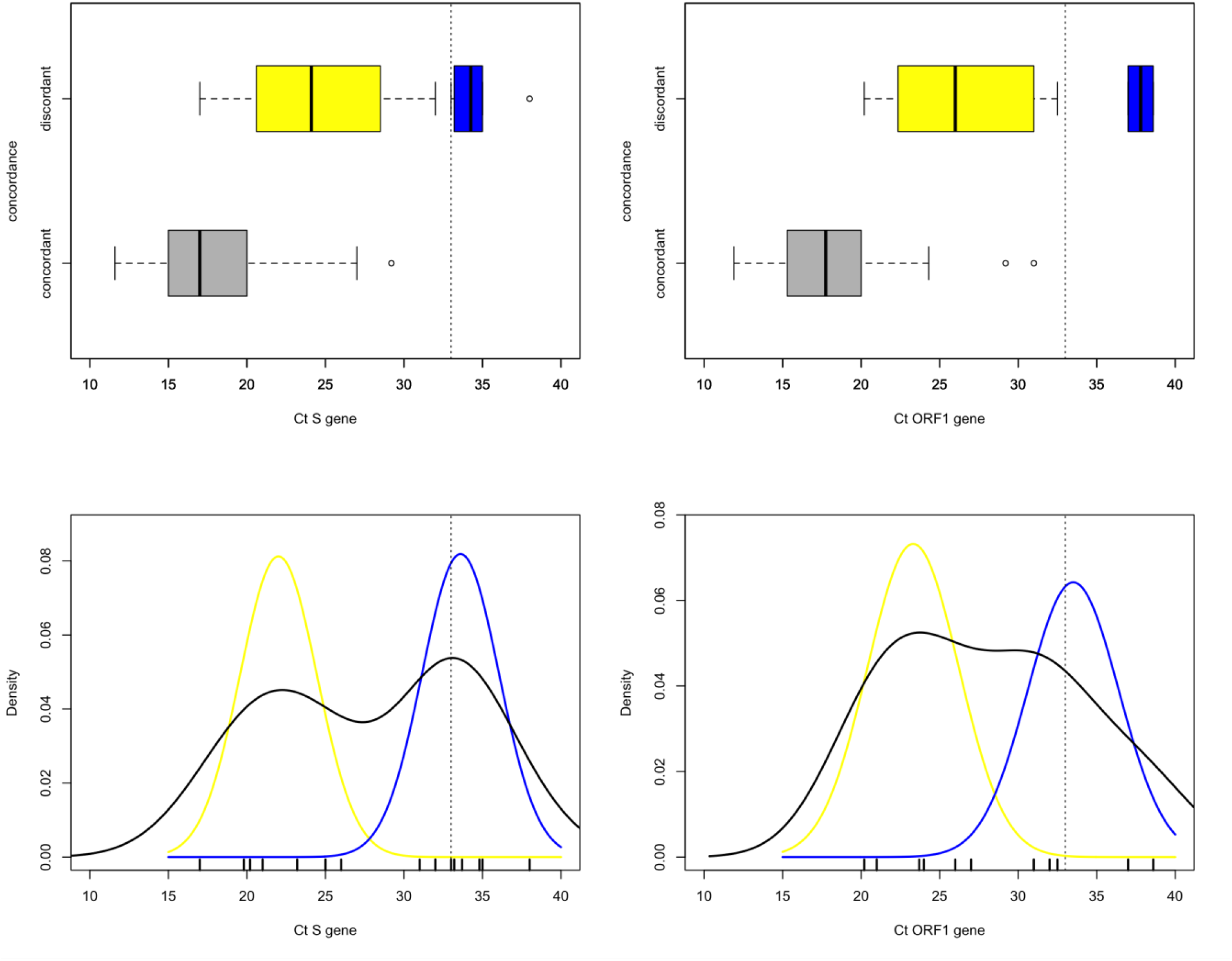
Distribution of Ct values for concordant DNCOV+/ANCOV+ and discordant DNCOV+ / ANCOV- test results. Top row: Boxplots of Ct values for the S (left) and ORF1 (right) gene. Dotted line at Ct = 33 shows the critical value of sensitivity (95%) declared for the antigen test above which there is an expected drop and an unreliable result. DNCOV+/ANCOV+ (grey box), DNCOV+/ANCOV- below sensitive threshold (yellow box) DNCOV+/ANCOV- above sensitive threshold (blue box). Bottom row: Nonparametric density estimation for Ct values of the S (left) and ORF1(right) gene, with superimposed the two densities fitted using a two-component Gaussian mixture model. DNCOV+/ANCOV- below sensitive threshold (yellow line) DNCOV+/ANCOV- above sensitive threshold (blue line).

**Table I:**
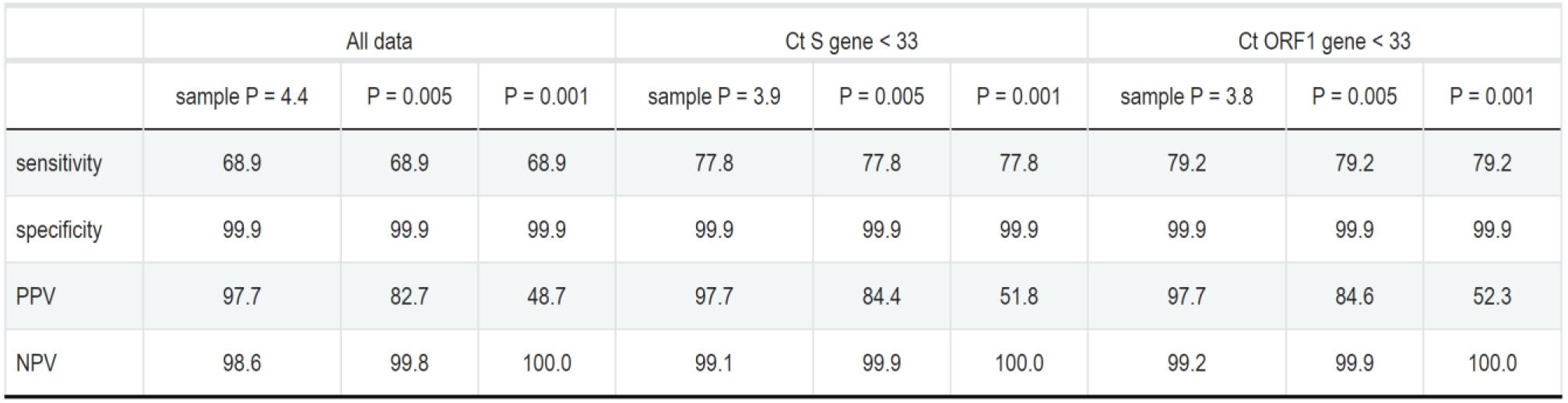
Overall sensitivity/specificity and PPV/NPV for ANCOV test.^*1*^

**Table II:**
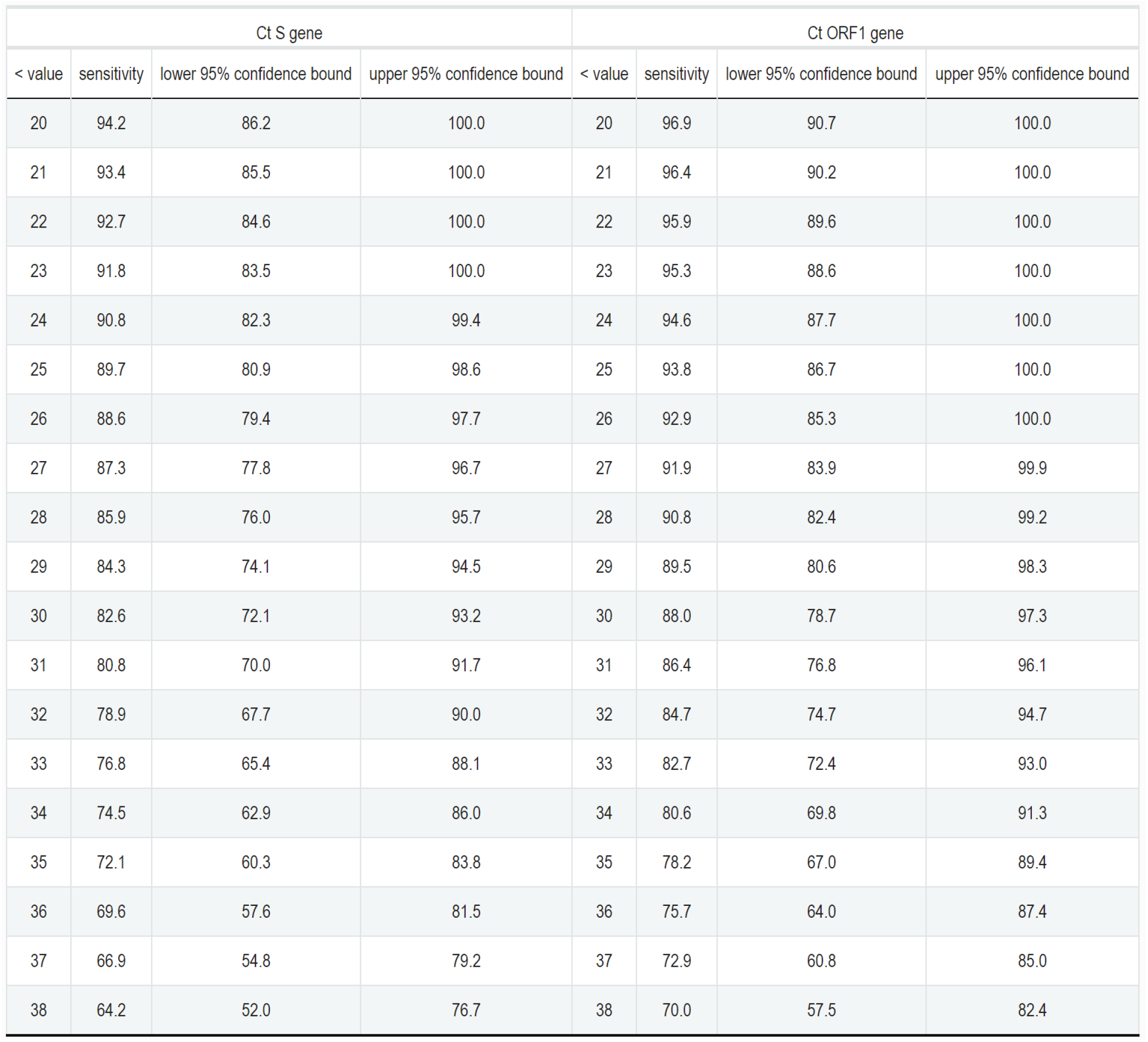
Sensitivity estimates of ANCOV test at increasing Ct value for the S and ORF1 gene generated by a logistic regression model, with 95% confidence intervals.

| Ct S gene |  |  |  | Ct ORF1 gene |  |  |  |
| --- | --- | --- | --- | --- | --- | --- | --- |
| < value | sensitivity | lower 95% confidence bound | upper 95% confidence bound | < value | sensitivity | lower 95% confidence bound | upper 95% confidence bound |
| 20 | 94.2 | 86.2 | 100.0 | 20 | 96.9 | 90.7 | 100.0 |
| 21 | 93.4 | 85.5 | 100.0 | 21 | 96.4 | 90.2 | 100.0 |
| 22 | 92.7 | 84.6 | 100.0 | 22 | 95.9 | 89.6 | 100.0 |
| 23 | 91.8 | 83.5 | 100.0 | 23 | 95.3 | 88.6 | 100.0 |
| 24 | 90.8 | 82.3 | 99.4 | 24 | 94.6 | 87.7 | 100.0 |
| 25 | 89.7 | 80.9 | 98.6 | 25 | 93.8 | 86.7 | 100.0 |
| 26 | 88.6 | 79.4 | 97.7 | 26 | 92.9 | 85.3 | 100.0 |
| 27 | 87.3 | 77.8 | 96.7 | 27 | 91.9 | 83.9 | 99.9 |
| 28 | 85.9 | 76.0 | 95.7 | 28 | 90.8 | 82.4 | 99.2 |
| 29 | 84.3 | 74.1 | 94.5 | 29 | 89.5 | 80.6 | 98.3 |
| 30 | 82.6 | 72.1 | 93.2 | 30 | 88.0 | 78.7 | 97.3 |
| 31 | 80.8 | 70.0 | 91.7 | 31 | 86.4 | 76.8 | 96.1 |
| 32 | 78.9 | 67.7 | 90.0 | 32 | 84.7 | 74.7 | 94.7 |
| 33 | 76.8 | 65.4 | 88.1 | 33 | 82.7 | 72.4 | 93.0 |
| 34 | 74.5 | 62.9 | 86.0 | 34 | 80.6 | 69.8 | 91.3 |
| 35 | 72.1 | 60.3 | 83.8 | 35 | 78.2 | 67.0 | 89.4 |
| 36 | 69.6 | 57.6 | 81.5 | 36 | 75.7 | 64.0 | 87.4 |
| 37 | 66.9 | 54.8 | 79.2 | 37 | 72.9 | 60.8 | 85.0 |
| 38 | 64.2 | 52.0 | 76.7 | 38 | 70.0 | 57.5 | 82.4 |

**Table III:** Wilcoxon-Mann-Whitney two-sample test. The hypothesis that the distributions of Ct values from DNCOV+ / ANCOV+ and discordant DNCOV+ / ANCOV- test results are equal is rejected at the 5% significance level.

|  | *Mean of Concordant* | *Mean of Discordant* | *p-value* |
| --- | --- | --- | --- |
| *Ct S gene all* | 17.9 | 27.8 | < 0.001 |
| *Ct S gene < 33* | 17.9 | 24.4 | <0.001 |
| *Ct ORF1 gene all* | 18.3 | 28.1 | < 0.001 |
| *Ct ORF1 gene < 33* | 18.3 | 26.3 | < 0.001 |

### Stratification of test performance

In spite of a lower sensitivity, antigen tests could find a useful application to rapidly confirm the diagnosis of Covid-19 in symptomatic patients with respiratory symptoms showing at A&E thus avoiding stationing of patients in a key hospital ward. The stratification of DNCOV+/ANCOV- and DNCOV+/ANCOV+ samples by age, sex and the presence of severe disease (pneumonia) at the time of testing did not show any statistical difference between concordant and discordant results at the 5% significance level. In addition, there is no difference in the distribution at the 5% significance level of the Ct values for the S and ORF1 gene when comparing samples stratified by age, sex and presence of disease severity. The date of symptom onset is known for all the 51 symptomatic patients. The time interval between symptom onset and test ranges from 0 (patients declare symptoms the same date of admission to the hospital ward) to 20 days, with a median and mean of 2 and 4 days respectively. The sensitivity of the assay decreased from 75% (95% CI: 55 – 89) to 60% (95% CI: 17 – 93) and 50% (95% CI: 29 – 71) when the test was performed within 3 days, on days 4 or 5 and after 5 days after the development of symptoms (**Table IV**). However, this gradient is not statistically significant at the 5% significance level possibly due the small sample size of each group (28, 5 and 18 patients). The viral load is known to decrease with time ^18^. Accordingly, the Ct values of both genes increases with the time interval (**Figure 4**). Ct values of 33 and above tend to be associated with a larger time interval (**Figure 5**), with Pearson’s correlation coefficient being marginally significant at the 5% significance level (S gene: 0.34 with p-value = 0.016; ORF1 gene: 0.48 with p-value < 0.001). The sensitivity of the 10 asymptomatic patients is 90 % (95% CI: 54 – 99) which have a Ct value for both S gene and ORF 1 gene below 30. Notably, these patients performed the test shortly after the diagnosis of infected relatives possible on after exposure as part of surveillance measures.

**Table IV:**
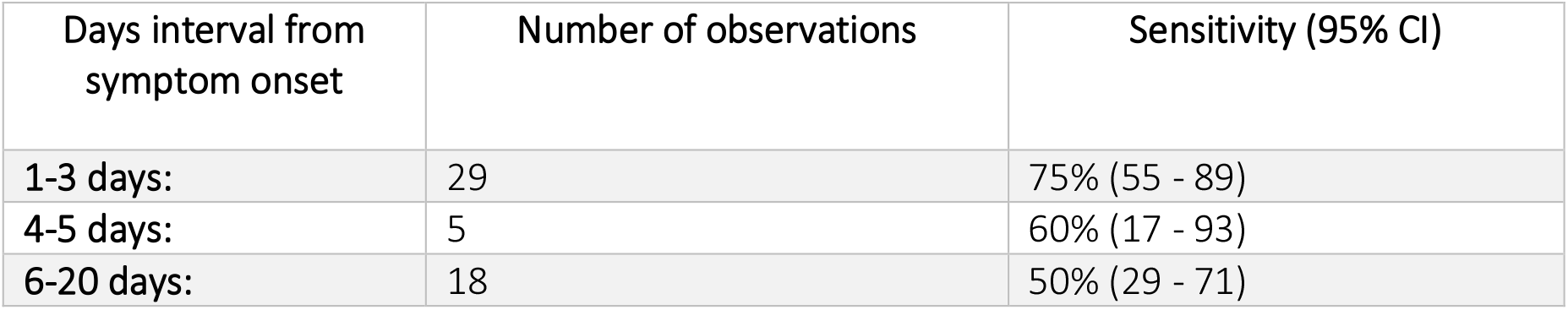
Test sensitivity at increasing time interval from symptom onset with 95% confidence intervals. P-value = 0.22 for equality of proportions.

**Figure 4:**
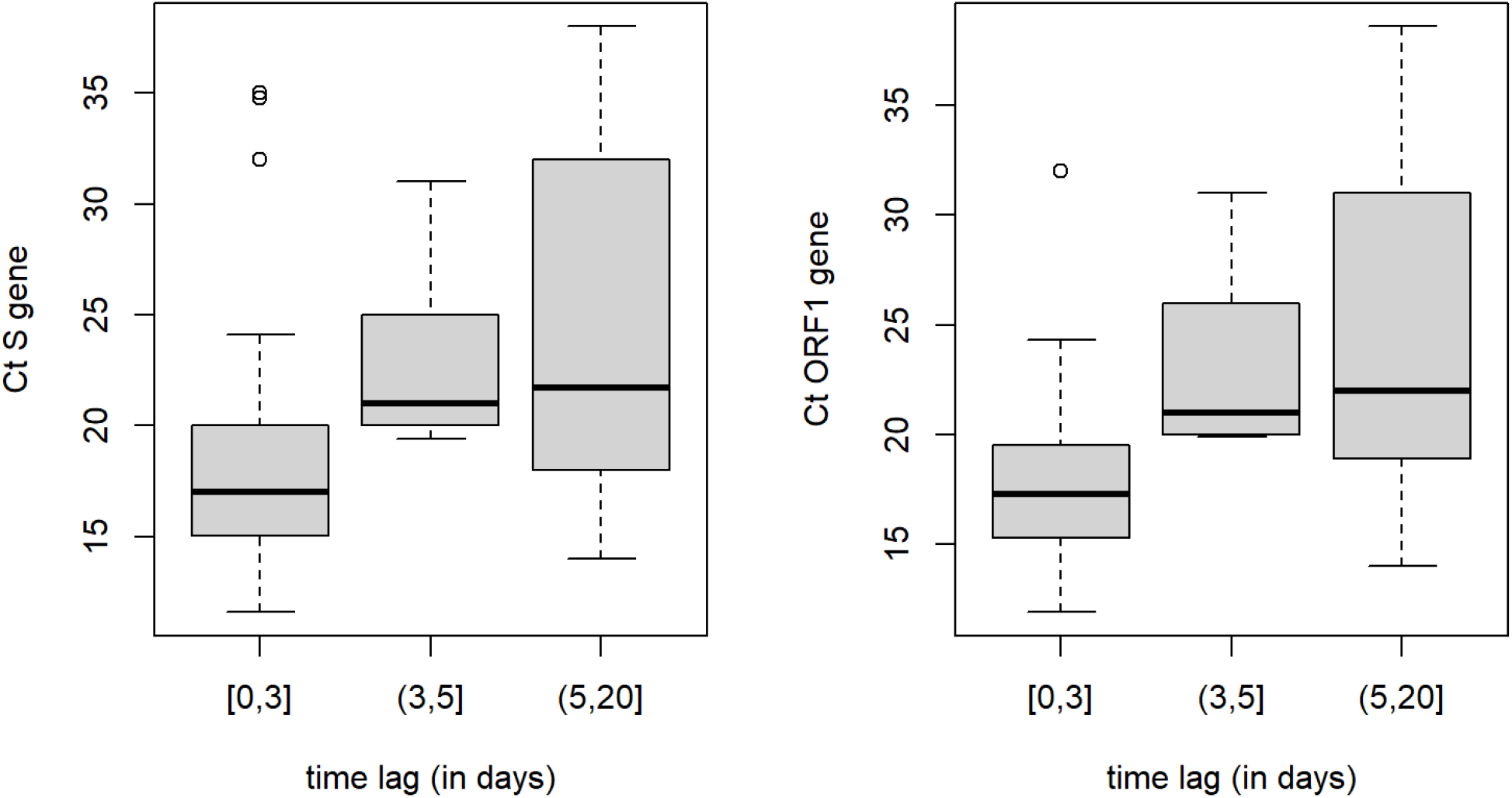
Boxplots of Ct values of S (left panel) and ORF1 (right panel) gene recorded at increasing time intervals (in days) from symptoms onset to day of testing 0-3 days, 4-5 days and above 5 days. P-value for Kruskall-Wallis test is 0.004 for Ct of ORF1 gene and 0.018 for Ct of S gene.

**Figure 5:**
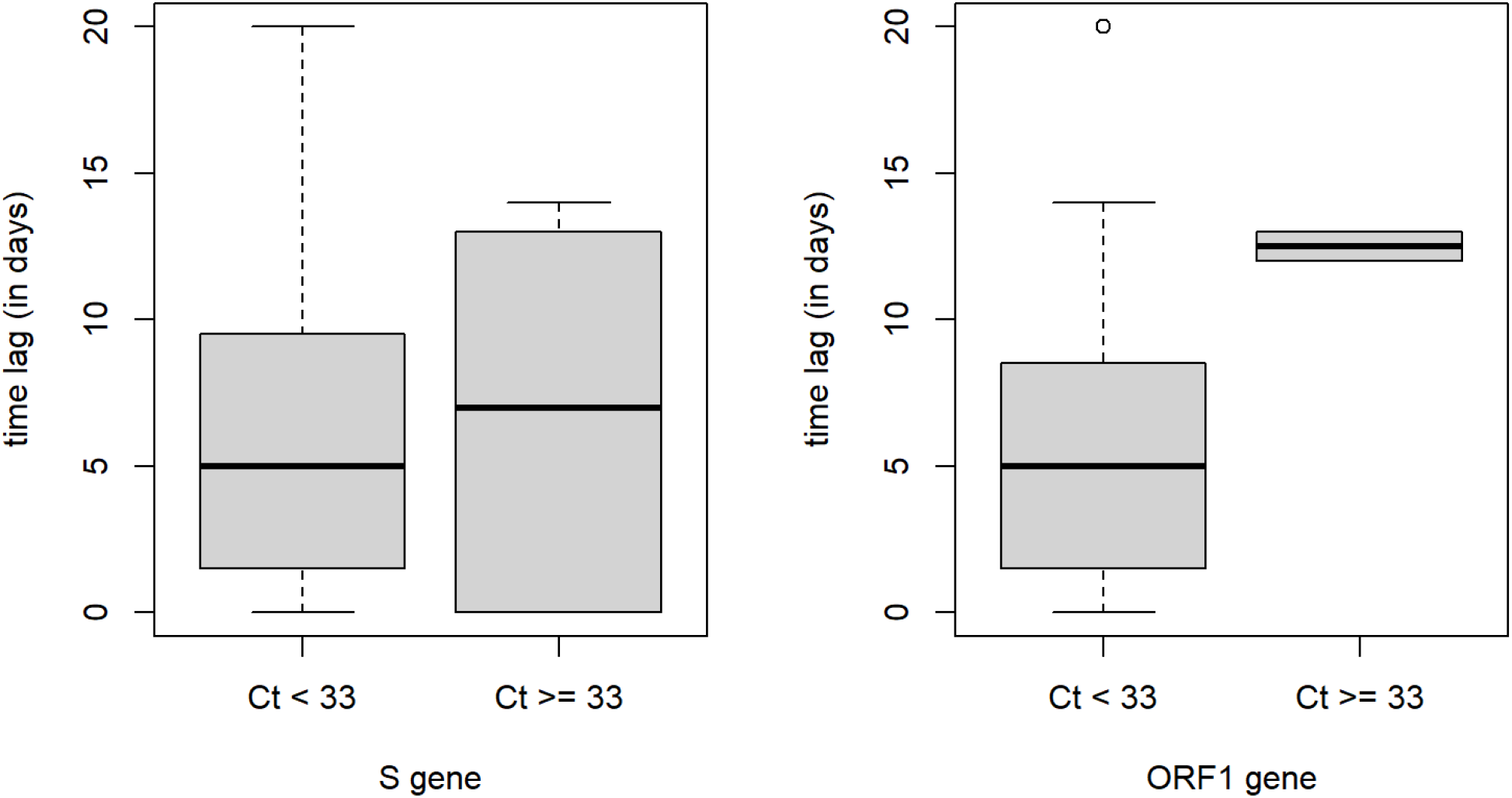
Boxplots of time interval (in days) between time of testing and day of symptom onset against different levels of Ct values Ct < 33 and Ct >= 33 for S (left panel) and Ct ORF1 (right panel) gene. P-value for two-sided Wilcoxon-Mann-Whitney test is 1.0 for Ct S gene and 0.2 for ORF1 gene.

### Genotyping of DNCOV+/ANCOV- and DNCOV+/ANCOV+ viruses

We carried full length sequencing on the viral RNA present on a collection of DNCOV+/ANCOV- and DNCOV+/ANCOV+ swab extracts to investigate whether the presence of genetic variants could account for the inability of the Abbott antigen test to identify positive samples in spite of the presence of a high viral load as documented by their Ct values for both the ORF1 and S gene. Initially, we selected two DNCOV+/ANCOV- (D1_7_B.1.1.119 and D1_8_B.1.177.7) samples that on RT-PCR showed Ct values for both the S and the ORF1 ranging from 22 to 25 and the sample C1_6_B.1.177 DNCOV+/ANCOV+ as control (well below the assay sensitivity threshold ^19^). The analysis of assembled viral sequences revealed the presence in both DNCOV+/ANCOV- samples of two distinct rare virus variants belonging to lineage B.1.1.119 and B.1.177.7. Notably both virus sequences were characterized by disruptive mutations (P365S for sample D1_8_B.1.177.7 and R209I for sample D1_7_B.1.1.119) in a region of the N protein, known to contain immunodominant epitopes that function as the target of capture antibodies in antigen tests ^20^, (**Figure 6**). To confirm this observation, we sequenced additional 6 DNCOV+/ANCOV- and 8 DNCOV+/ANCOV+ samples. Analysis of assembled viral sequences confirmed that the great majority of DNCOV+/ANCOV- were characterized by disruptive amino-acid substitutions mapping within or in proximity of B cell epitopes of the N antigen (**Figure 6**). One virus contained the mutation P365S (D2_6_B.1.177), already found in the virus variant D1_8_B.1.177.7, three viruses (D2_1_B.1.160, D2_4_B.1.160 and D2_7_B.1.160) showed the same amino-acid substitutions A376T and M234I, two additional samples revealed different mutations; D348Y (D2_3_B.1.177.4) and Q229H (D2_2_B1.1.177). None of the viruses present in the DNCOV+/ANCOV+ samples showed amino-acid substitution downstream to position 220. We also tested the ability of Abbott Panbio™ COVID-19 Ag and two additional antigen tests (COVID-19 Ag Respi-Strip from Coris BioConcept, Belgium and LumiraDX SARS-CoV-2) to correctly identify 5 cultured viruses isolated at the beginning of the pandemic in the municipality of Vo’ when lineage B (original lineage first to be discovered in Wuhan province) was circulating. Culture supernatants of Vo’ virus isolates (samples C_Vo_110_B, C_Vo_15_B, C_Vo_40_B, C_Vo_42_B, C_Vo_46_B) produced a positive signal in all antigen tests. Accordingly, their sequence did not show any mutation in the N antigen. On the contrary all antigen tests failed to detect the presence of the viruses D2_3_B.1.177.4, D2_1_B.1.160 and D1_7_B.1.1.119. RT-PCR analysis confirmed that the supernatant contained the viruses. We analysed the frequency of both DNCOV+/ANCOV+ and DNCOV+/ANCOV- virus variants in Europe (230,756 analysed sequences), in Italy (3,014 analysed sequences) and the in Regione Veneto (339 analysed sequences) deposited in SARS-CoV-2 sequence database from 01-03-2021 to 31-01-2021 (GISAID databank ^16^) to investigate their temporal dynamics and spatial distribution (**Figure 7** and **Suppl. Mat**.). The amino-acid mutations D348Y, Q229H, R209I, and T141I appeared sporadically in Europe over the period examined but never increased in frequency. One conservative amino-acid substitution A220V found in different clades increased in frequency to 60% in both Europe and Italy. The mutations G204R and R203K appeared very early from the start of the epidemics reaching 80% frequency by the end of June and declined thereafter. The amino-acid substitution A376T coupled with M234I found in 38% of all DNCOV+/ANCOV- virus sequences analysed appeared in both Europe and Italy at the beginning of July and after a modest rise this variant decreased to less than 5% by the end of December 2020. On the contrary in the Veneto region, where antigen tests have been massively used starting from the beginning of June, the frequency of this variant progressively increased to reach 20% of all sequences analysed during the same period.

**Figure 6:**
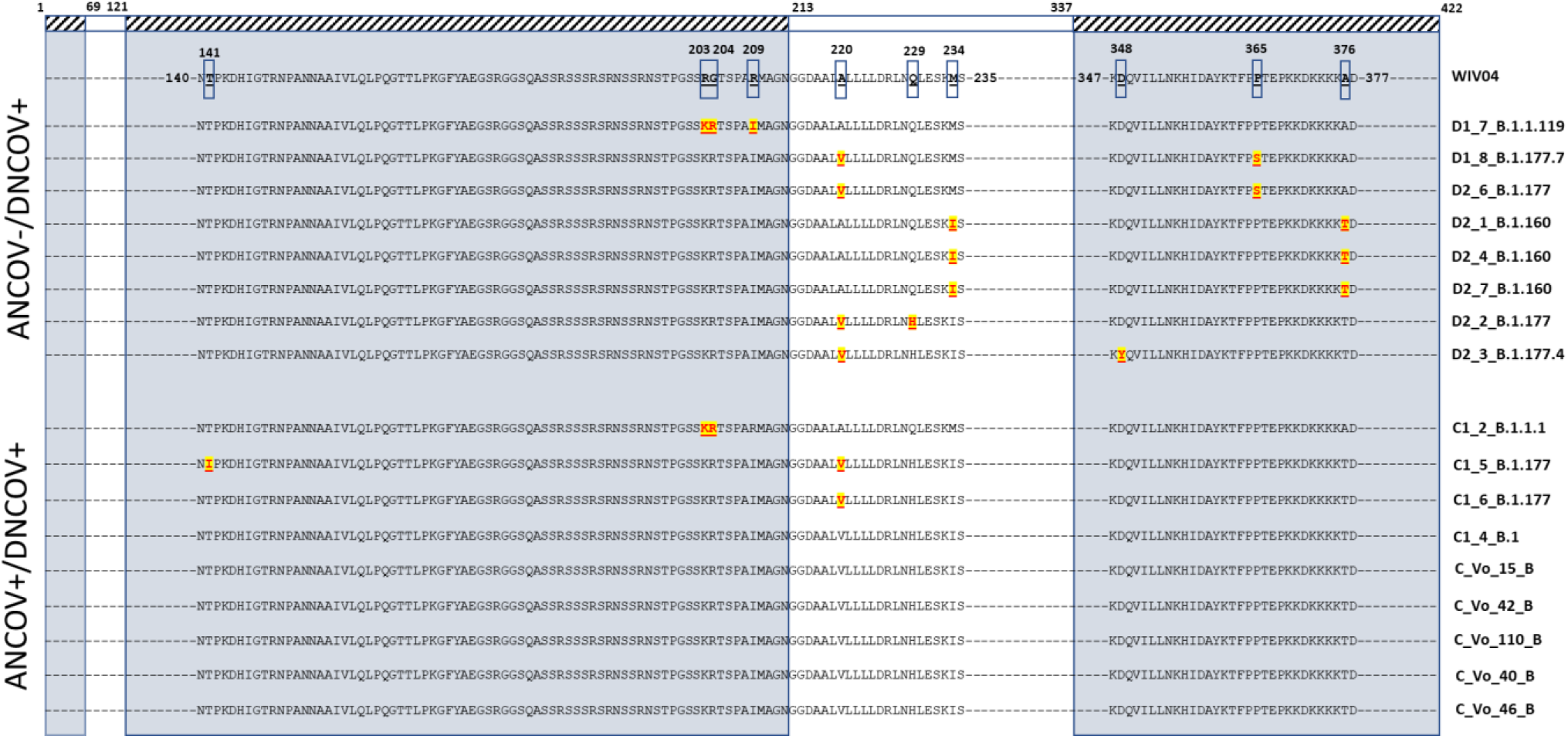
The relative positions of amino-acid substitutions (with respect to the Wuhan reference sequence - WIV04) found in the N gene of 17 full-length sequences of SARS-CoV-2 are shown. The sequences are from individuals showing either discordant, ANCOV- /DNCOV+ or concordant ANCOV+/DNCOV+, at molecular and antigenic swab tests. Regions of mapped B cell epitopes are highlighted on the Wuhan reference (hatched boxes) and the sequences of the N gene (gray shaded boxes). Only sequences around amino acid substitutions (boxed on the Wuhan reference WIV04) are shown. Mutations found in DNCOV+/ANCOV+, and DNCOV+/ANCOV- sequences are shaded in yellow and highlighted in red. The last six sequences do not contain any mutation according to the Wuhan reference sequence.

**Figure 7:**
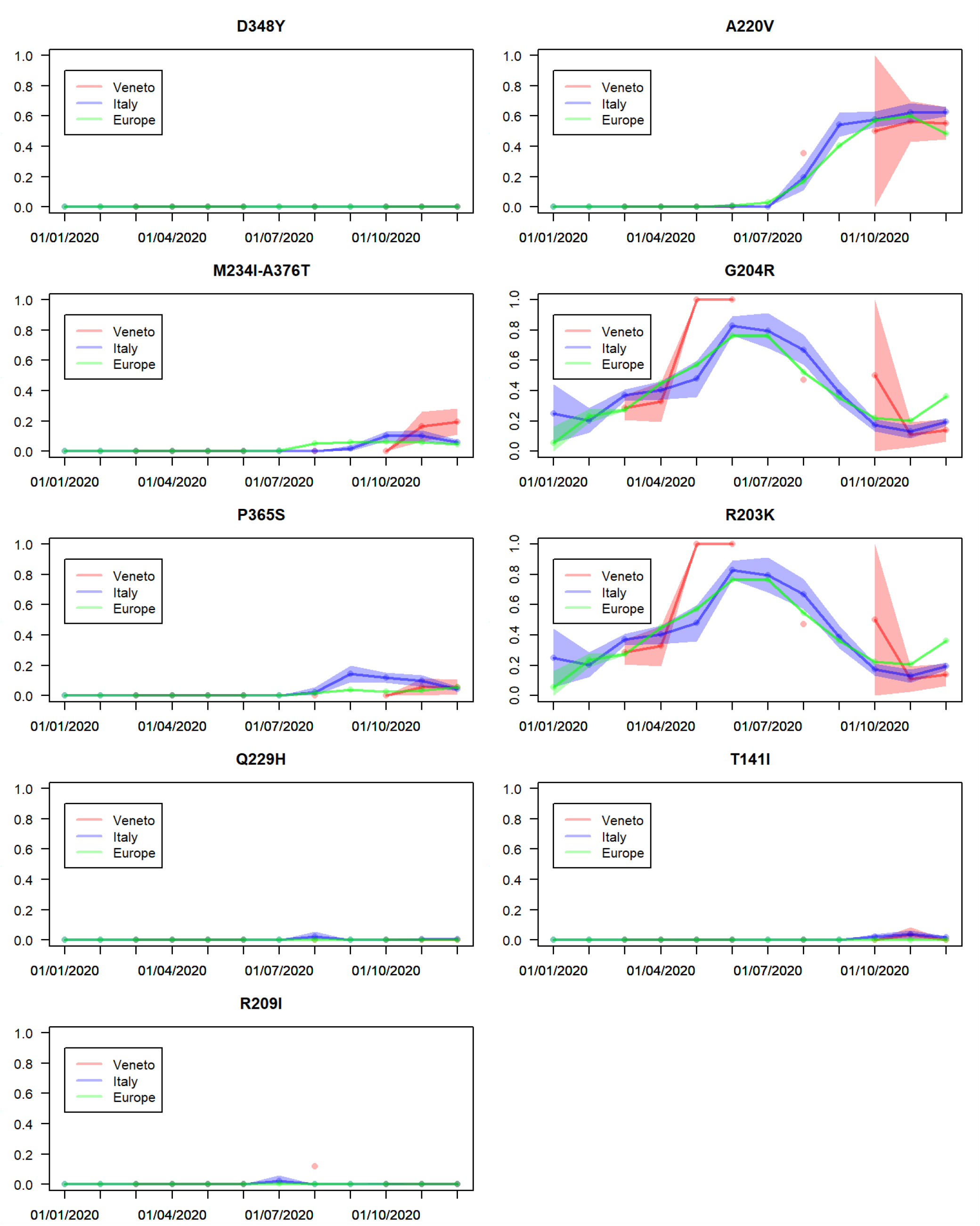
Monthly prevalence (percentage over total sequences) of N variants generating discordant DNCOV+ / ANCOV- (left panels) and concordant DNCOV+ / ANCOV+ (right panels) result in molecular and antigen swab. Shaded areas show the 95% confidence bands for each different region. No sequence data are available for Veneto region in the months of January, February, July, and September.

## DISCUSSION

After the introduction of the Abbott Panbio™ COVID-19 Ag Rapid Test as a screening tool in hospital settings across Veneto ^21^ we systematically tested 1,441 subjects admitted to the A&E and the Infectious Disease wards with both the antigen and RT-PCR tests. The sample represented 44% of all patients examined (3,290) at the University Hospital. Statistical analysis indicated that no bias was introduced in age, gender and underlying pathological conditions. The Abbott Panbio™ COVID-19 test showed an overall specificity of 99.9% (95% CI: 99.5 – 100) and a sensitivity of 68.9% (95% CI: 55.6 – 79.8) the latter value being much lower than that declared by the manufacturer though in agreement with previous reports ^22,23^. The analysis indicated that the sensitivity was clearly affected by the Ct value. Accordingly, samples with Ct values above 33 invariably gave a negative result whereas the sensitivity progressively increased with lowering Ct values. The analysis also confirmed that the performance of the test was not affected by the severity of the disease. We also investigated whether the time of testing from symptom onset impacted on the sensitivity as consequence of changing the Ct value (hence viral load) during the course of the diseases with the aim to provide insight on the best timing to recommend the use of the antigen test. This analysis showed that the sensitivity of the Abbott Panbio™ COVID-19 decreased with the number of days that passed between symptoms onset and time of testing in line with the observed decrease in vital load.

Unexpectedly, we came across a number of samples with low Ct values (well below the antigen test threshold) that failed to generate a positive result. An observation that could not be explained in term of test performance. We challenged the hypothesis that the negative results were due to the presence of variants of the N antigen that escaped detection by the capture antibody. Sequencing analysis of viruses found in RNA extract of nasopharyngeal swabs from concordant DNCOV+/ANCOV+ and discordant DNCOV+/ANCOV- samples revealed a pattern of amino-acid substitutions mapping around major epitopes of the N antigen ^20^. Compared to viral sequences of isolates collected in February 2020 the N antigen presented several mutations at its carboxyl-end starting from amino-acid 141. The amino-acid substitution A220V was the most common but unlikely to affect test sensitivity as it was equally present in both DNCOV+/ANCOV+ and DNCOV+/ANCOV- viruses. Most of the mutations found in DNCOV+/ANCOV- viruses mapped within a major B cell epitope of the N protein previously identified from position 337 and 422 and involved disruptive amino-acid substitutions ^20^. Unfortunately, the nature of the B cell epitope recognized by the antibody utilised in the Abbott test is not known but apparently other SARS-CoV-2 antigen tests, COVID-19 Ag Respi-Strip and LumiraDX SARS-CoV-2, seem to suffer of the same limitation as supernatant of cultured cells infected with virus isolates D1_7_B.1.1.119, D1_8_B.1.177.7, D2_1_B.1.160, and D2_3_B.1.177.4 carrying respectively mutations R209I, P365S, M234I-A376T, and D348Y failed to generate a positive signal thus suggesting that these assays use antibodies with similar specificities.

The fact that some virus variants are “undetectable” irrespectively of the Ct value combined with a lower sensitivity of antigen tests may have accounted for the partial and disappointing results obtained when antigen tests were utilised in large population screens. It should also be considered that the massive use of antigen tests could exert a positive selection for undetectable virus variants and thereby causing their relative increase in frequency over time. Our results would support this notion: The DNCOV+/ANCOV- A376T-M241I virus variant accounted for 42% of all mutation found in the N-antigen B cell epitope 337-422. Analysis of virus sequences isolated from January 2020 to December 2020 indicated that in Veneto the A376T-M241I variant progressively increased in frequency from the beginning of October 2020 to 20% by the end of December 2020 while in Europe and the rest of Italy, after a small peak in October 2020, it followed an opposite temporal pattern. This increase in frequency of A376T-M241I variant paralleled the scaling up of rapid antigen tests in this region of Italy that reached nearly 67.4% of all swab tests performed there in February 2021. During the progressive and massive introduction of antigen tests from October 2020, Veneto suffered one of the highest daily infection rates with more than 7,000 deaths in the period from October 2020 to February 2021 one third of which amongst the elderly people hosted in nursing home ^24,25^. All together the findings presented here question the use of antigenic tests for surveillance approaches and highlight urgent need to revisit their use indication ^26^ as well as for the producers to disclose the targets of the capture antibodies.

## Supporting information

Suppl. Mat.

GISAID acknowledgments

## Data Availability

Data are available as supplementary material

## ETHICAL APPROVAL STATEMENT

This study is a retrospective diagnostic analysis. Patient consent for diagnostic investigation was taken at the time of admission. The purpose and design of the study has been approved by the Local Ethics Committee of the Province of Padova (Italy) which has acknowledged to be in line with ethical practices and ICH-CGP guidelines.

## ACKNOWLEDGEMENTS

COVID-19 emergency fund from University of Padova (grant number: TOPP_PRIV20_01) to ST. We gratefully acknowledge the Authors from the Originating Laboratories and the Submitting Laboratories who generated and shared via GISAID the data on which this research is based.

## Footnotes

1 The estimates of sensitivity slightly differ from the ones reported in **Table II**, as the latter obtains the estimates of sensitivity from the fitted logistic regression model.

