## Supplementary material for "Emergence of N antigen SARS-CoV-2 genetic variants escaping detection of antigenic tests": GISAID acknowledgments

We gratefully acknowledge the following Authors from the Originating laboratories responsible for obtaining the specimens, as well as the Submitting laboratories where the genome data were generated and shared via GISAID, on which this research is based.

All Submitters of data may be contacted directly via www.gisaid.org

Authors are sorted alphabetically.

| Accession ID | Originating Laboratory | Submitting Laboratory | Authors |
|---|---|---|---|
| EPI_ISL_875674 | "1.AO Universitaria 'S. Giovanni di Dio e Ruggi D'Aragona, Scuola Medica Salernitana' Hospital / 2.UOC di Virologia e Microbiologia, Università della Campania 'L. Vanvitelli' / 3.AO Universitaria 'Federico II' Napoli Hospital / 4.AORN 'San Giuseppe Moscati' Avellino Hospital / 5.AO 'San Pio - presidio G. Rummo' Benevento Hospital / 6.AO 'Sant'Anna e San Sebastiano' Caserta Hospital / 7.PO 'Maria Santissima Addolorata' Eboli Hospital / 8.Biogem Istituto di Ricerche Genetiche" | "1. Genome Research Center for Health (CRGS) / 2. Laboratory of Molecular Medicine and Genomics(LMMGe) / 3. Center for Research in Pure and Applied Mathematics (CRMPA)" | "Giorgio Giurato (Corresponding Author); Alessandro Weisz (Corresponding Author); Alessia Cossu; Aniello Gentile; Annamaria Salvati; Antonello Saccomanno; Arnolfo Petruzziello; Assunta Sellitto; Carlo Ferravante; Domenico Memoli; Domenico Palumbo; Elena Alexandrova; Emilia Vaccaro; Francesca Marciano; Francesca Rizzo (Corresponding Author); Gianluigi Franci; Giovanni Nassa; Giovanni Pecoraro; Giuseppe Fenza; Giuseppe Portella; Gregorio Goffredi; Ilaria Terenzi; Jessica Lamberti; Maddalena Schioppa; Maria Grazia Foti; Maria Landi; Marianna Scrima"; Mariarosaria Ingino; Massimiliano Galdiero; Maurizio Fumi; Michele Caraglia; Michele Cennamo; Oriana Strianese; Pasquale Pagliano; Rita Greco; Roberta Tarallo; Sonia Amabile; Teresa Rocco; Valeria Mirici Cappa; Vincenzo Rocco; Viola Melone; Vittoria Letizia; Ylenia D'Agostino |
| EPI_ISL_833052 to 833054 | "Presidio Ospedaliero "San Liberatore" Atri | Istituto Zooprofilattico Sperimentale dell'Abruzzo e Molise "G. Caporale" | Ancora M; Calistri P; Cammà C; Curini V; Delli Compagni E; Di Domenico M; Di Pasquale A; Lorusso A; Mangone I; Marcacci M; Puglia I; Rinaldi A; Savini G |
| EPI_ISL_833055 to 833056 | "SIESP DIPARTIMENTO DI PREVENZIONE CHIETi | Istituto Zooprofilattico Sperimentale dell'Abruzzo e Molise "G. Caporale" | Ancora M; Calistri P; Cammà C; Curini V; Delli Compagni E; Di Domenico M; Di Pasquale A; Lorusso A; Mangone I; Marcacci M; Puglia I; Rinaldi A; Savini G |
| EPI_ISL_451971 to 451977, EPI_ISL_451979 to 451982, EPI_ISL_451984 to 451987 | 1. ViroGenetics - BSL3 Laboratory of Virology, Maopolska Centre of Biotechnology, Jagiellonian University; 2. II Department of Internal Medicine, Faculty of Medicine, Jagiellonian University Medical College; 3. DIAGNOSTYKA Ltd. | 1. ViroGenetics - BSL3 Laboratory of Virology, Maopolska Centre of Biotechnology, Jagiellonian University; 2. II Department of Internal Medicine, Faculty of Medicine, Jagiellonian University Medical College. | Jakub Swadba; Krzysztof Pyr; Marcin Surmiak; Marek Sanak; Marta Rogalska-Kupiec; Monika Gsecka-Czapla; Pawe P abaj; Wojciech Branicki |
| EPI_ISL_455440 to 455453, EPI_ISL_492066 to 492073 | 1. ViroGenetics - BSL3 Laboratory of Virology, Maopolska Centre of Biotechnology, Jagiellonian University; 2. II Department of Internal Medicine, Faculty of Medicine, Jagiellonian University Medical College; 3. Narodowy Instytut Zdrowia Publicznego - Pastwowy Zakad Higieny (NIZP-PZH) | 1. ViroGenetics - BSL3 Laboratory of Virology, Maopolska Centre of Biotechnology, Jagiellonian University; 2. II Department of Internal Medicine, Faculty of Medicine, Jagiellonian University Medical College; 3. Narodowy Instytut Zdrowia Publicznego - Pastwowy Zakad Higieny (NIZP-PZH). | Agnieszka Koakowska-Kulesza; Aleksandra A. Zasada; Aleksandra Milewska; Ewelina Hallman-Szeliska; Katarzyna Owczarek; Katarzyna Pancer; Katarzyna Zacharczuk; Krzysztof Pyr; Magdalena Rzeczkowska; Marek Sanak; Natalia Wolaniuk; Pawe P abaj; Tomasz Wokowicz; Wojciech Branicki |
| EPI_ISL_876042 to 876043, EPI_ISL_876969 to 877034, EPI_ISL_877036, EPI_ISL_882674 to 882688, EPI_ISL_882690 to 882768, EPI_ISL_883500, EPI_ISL_887506 to 887507, EPI_ISL_888679 to 888682, EPI_ISL_894218, EPI_ISL_918128, EPI_ISL_918412 to 918422, EPI_ISL_918457 to 918460, EPI_ISL_918462 to 918482, EPI_ISL_925093 to 925136, EPI_ISL_925497 to 925501, EPI_ISL_934419 to 934423, EPI_ISL_935104 to 935127, EPI_ISL_935129 to 935154, EPI_ISL_954183 to 954212, EPI_ISL_954214 to 954217 | | | |
| see above | 1.AO Universitaria 'S. Giovanni di Dio e Ruggi D'Aragona, Scuola Medica Salernitana' Hospital / 2.UOC di Virologia e Microbiologia, Università della Campania 'L. Vanvitelli' / 3.AO Universitaria 'Federico II' Napoli Hospital / 4.AORN 'San Giuseppe Moscati' Avellino Hospital / 5.AO 'San Pio - presidio G. Rummo' Benevento Hospital / 6.AO 'Sant'Anna e San Sebastiano' Caserta Hospital / 7.PO 'Maria Santissima Addolorata' Eboli Hospital / 8.Biogem Istituto di Ricerche Genetiche | 1. Genome Research Center for Health (CRGS) / 2. Laboratory of Molecular Medicine and Genomics(LMMGe) / 3. Center for Research in Pure and Applied Mathematics (CRMPA) | Alessandro Weisz; Alessandro Weisz (Corresponding Author); Alessia Cossu; Andreina Baj; Aniello Gentile; Annamaria Salvati; Antonello Saccomanno; Arnolfo Petruzziello; Assunta Sellitto; Carlo Ferravante; Domenico Memoli; Domenico Palumbo; Edmondo Adorisio; Elena Alexandrova; Emilia Vaccaro; Fausto Sessa.; Francesca Marciano; Francesca Rizzo; Francesca Rizzo (Corresponding Author); Francesco Curcio; Gianluigi Franci; Giorgio Dirani; Giorgio Giurato; Giorgio Giurato (Corresponding Author); Giovanni Nassa; Giovanni Pecoraro; Giuseppe Fenza; Giuseppe Portella; Gregorio Goffredi; Ilaria Terenzi; Jessica Lamberti; Maddalena Schioppa; Maria Grazia Foti; Maria Landi; Marianna Scrima; Mariarosaria Ingino; Massimiliano Galdiero; Maurizio Fumi; Michela Iacobellis; Michele Caraglia; Michele Cennamo; Morena D'Avenia; Oriana Strianese; Pasquale Pagliano; Rita Greco; Roberta Tarallo; Rosanna Piluscio; Silvia Zanoli; Simona Semprini; Sonia Amabile; Stefania Marzinotto; Teresa Rocco; Valeria Mirici Cappa; Vincenzo Rocco; Viola Melone; Vittoria Letizia; Vittorio Sambri; Ylenia D'Agostino |
| EPI_ISL_729338, EPI_ISL_729340, EPI_ISL_729342 to 729345, EPI_ISL_729347, EPI_ISL_729351 to 729352, EPI_ISL_729354, EPI_ISL_729358 to 729359, EPI_ISL_729361 to 729364, EPI_ISL_729367 to 729369, EPI_ISL_729371 to 729376, EPI_ISL_729379 to 729380, EPI_ISL_729383 to 729385, EPI_ISL_729389 to 729392, EPI_ISL_729394 to 729396, EPI_ISL_729399, EPI_ISL_729401, EPI_ISL_729403, EPI_ISL_729407, EPI_ISL_729411, EPI_ISL_729414, EPI_ISL_729416 to 729467, EPI_ISL_729469 to 729472, EPI_ISL_729498, EPI_ISL_729500 to 729506, EPI_ISL_729508 to 729513, EPI_ISL_729515 to 729528, EPI_ISL_729535 to 729547, EPI_ISL_729549 to 729552, EPI_ISL_729562 to 729595, EPI_ISL_729608 to 729687, EPI_ISL_729689 to 729717, EPI_ISL_729719 to 729734, EPI_ISL_824742 to 824786, EPI_ISL_849794 to 849866 | | | |
| see above | A. Krumbholz, Labor Dr. Krause und Kollegen MVZ GmbH, Kiel | Charité Universitätsmedizin Berlin, Institut für Virologie | Barbara Mühlemann; Christian Drosten; Julia Schneider; Julia Tesch; Jörn Beheim-Schwarzbach; Talitha Veith; Terry Jones; Tobias Bleicker; Victor M Corman |
| EPI_ISL_766695, EPI_ISL_766698 to 766699, EPI_ISL_766708 to 766709, EPI_ISL_766715 to 766723 | A05 Biomedicum | The Public Health Agency of Sweden | Department of Microbiology; The Public Health Agency of Sweden |
| EPI_ISL_775533 to 775541, EPI_ISL_775550 to 775571, EPI_ISL_775575 to 775582, EPI_ISL_788991 to 788998, EPI_ISL_789000 to 789011, EPI_ISL_789014 to 789016, EPI_ISL_789018 to 789028, EPI_ISL_831944, EPI_ISL_831947 to 831952, EPI_ISL_831959, EPI_ISL_831982 to 831985 | | | |
| see above | ABC | The Public Health Agency of Sweden | Department of Microbiology; The Public Health Agency of Sweden |
| EPI_ISL_763362 to 763363, EPI_ISL_768752 to 768753, EPI_ISL_768755, EPI_ISL_768757, EPI_ISL_768759 to 768760, EPI_ISL_768762 to 768797, EPI_ISL_768833 | | | |
| see above | AIID | Irish Coronavirus Sequencing Consortium - National Virus Reference Laboratory | Alejandro Abner Garcia Leon; Gabriel Gonzalez; Michael Carr; Patrick Mallon |
| EPI_ISL_778819 to 778841, EPI_ISL_871793 to 871814 | AIID | Irish Coronavirus Sequencing Consortium-Teagasc Grange | Aljandro Abner Garcia Leon; Calum Walsh; Fiona Crispie; Gabriel Gonzalez; John Kenny; Matthew McCabe; Michael Carr; Patrick Mallon; Paul Cotter |
| EPI_ISL_418000 to 418001, EPI_ISL_418004 | ARS Algarve - Laboratório Laura Ayres | Instituto Nacional de Saude (INSA) | Guiomar et al |
| EPI_ISL_542098 to 542277, EPI_ISL_542400 to 542443 | ASST GOM Niguarda | Dep. Of Oncology and Hemato-Oncology University of Milan | Antonio Piralla; Carlo Federico Perno; Chiara Vismara; Claudia Alteri; Elisa Matarazzo; Fausto Baldanti; Federica Giardina; Federica Novazzi; Luna Colagrossi; Maria Antonello; Massimo Puoti; Monica Tallarita; Oscar Massimiliano Epis; Roberto Fumagalli; Silvia Renica; Stefano Gaiarsa; Valentino Costabile; Valeria Cento |
| EPI_ISL_476135 | Achima Care Fristadens VC | The Public Health Agency of Sweden | Anna Risberg; Anna-Malin Linde; Karin Tegmark-Wisell; Maria Lind Karlberg; Mattias Haukland; Mia Brytting; Olov Svartstrom; Oskar Karlsson Lindsjo; Petra Edquist; Reza Advani; Sandra Broddesson |
| EPI_ISL_509412 to 509413, EPI_ISL_509415 to 509417 | Acibadem Labcell Cellular Therapy Laboratory | Acibadem Mehmet Ali Aydinlar University School of Medicine, Medical Genetics Department | Bulut Yurtsever; Cihan Tastan; Derya Dilek Kancagi; Ercument Ovali; Gozde Sir Karakus; Gunseli Bayram Akcapinar; Ilayda Sahin; Ozden Hatirnaz Ng; Ozkan Ozdemir; Sezer Akyoney; Ugur Ozbek |
| EPI_ISL_528934 to 528949 | Agenzia di Tutela della Salute di Bergamo | Istituto Zooprofilattico Sperimentale dell'Abruzzo e Molise "G.Caporale" | Ancora M; Cammà C; Curini V; Di Domenico M; Di Pasquale A; Lorusso A; Mangone I; Marcacci M; Puglia I; Rinaldi A; Savini G. |
| EPI_ISL_420134, EPI_ISL_420136, EPI_ISL_420139, EPI_ISL_420311 to 420312, EPI_ISL_500779 to 500783, EPI_ISL_500793 to 500796, EPI_ISL_549083 to 549084, EPI_ISL_549089 to 549091, EPI_ISL_549169 to 549171, EPI_ISL_590889 to 590892, EPI_ISL_590951 to 590952, EPI_ISL_635102, EPI_ISL_635194, EPI_ISL_668391, EPI_ISL_668393, EPI_ISL_775269 to 775270, EPI_ISL_775275, EPI_ISL_775344, EPI_ISL_775387 to 775390, EPI_ISL_775479, EPI_ISL_775528 to 775530, EPI_ISL_796649, EPI_ISL_796665 to 796666, EPI_ISL_796699, EPI_ISL_796712 to 796713, EPI_ISL_813975, EPI_ISL_860223 | | | |
| see above | Akershus University Hospital, Department for Microbiology and Infectious Disease Control | Norwegian Institute of Public Health, Department of Virology | Atiya R Ali; Hilde Elshaug; Hilde Synnøve Vollan; Hilde Vollan; Kamilla Heddeland Instefjord; Karoline Bragstad; Kathrine Stene-Johansen; Marie Paulsen Madsen; Olav Hungnes; Rasmus Riis Kopperud |
| EPI_ISL_569865 to 569886 | Amedeo di savoia | Crosetto lab, Karolinska Institutet, SciLifeLab | Anna Sapino; Luuk Harbers; Maria Grazia Milia; Michele Simonetti; Nicola Crosetto; Ning Zhang; Valeria Ghisetti |

| | | | |
|---|---|---|---|
| EPI_ISL_766587 | Analytica Medizinische Laboratorien AG | Institute of Medical Virology, University of Zurich | Alexandra Trkola; Annette Audigé; Cyril Shah; Jon Huder; Jürg Böni; Kevin Steiner; Maria Grünberg; Maryam Zaheri; Michael Huber; Riccarda Capaul; Stefan Schmutz; Verena Kufner |
| EPI_ISL_450818 | Aneby VC | The Public Health Agency of Sweden | Anna Risberg; Anna-Malin Linde; Karin Tegmark-Wisell; Ken Granath; Maria Lind Karlberg; Mia Brytting; Olov Svartstrom; Oskar Karlsson Lindsjo; Theresa Enkirch |
| EPI_ISL_582127 to 582132 | Antwerp University Hospital | Institute of Tropical Medicine | Colin Anthony; Philippe Selhorst |
| EPI_ISL_512656 | Area De Salud Pavas (Coopesalud) [Pavas/San Jose] | Inciensa, Instituto Costarricense de Investigación y Enseñanza en Nutrición y Salud | Adriana Godinez & Melany Calderon; Claudio Soto-Garita; Estela Cordero; Francisco Duarte; Hebleen Porras |
| EPI_ISL_457826 | Army Medical Center - Scientific Department | Army Medical and Veterinary Research Center | Anna Anselmo; Antonella Fortunato; Florigio Lista; Francesco Giordani; Giovanni Faggioni; Nino D'Amore; Riccardo De Sanctis; Silvia Fillo; Vanessa Vera Fain |
| EPI_ISL_717978 | Army Medical Center, Scientific Department, Virology Laboratory | Army Medical Center, Scientific Department, Virology Laboratory | Alessandra Amoroso; Andrea Ciammaruconi; Anella Monte; Anna Anselmo; Annalisa Pelo; Antonella Fortunato; Federica Galeano; Florigio Lista.; Francesco Giordani; Giovanni Faggioni; Giulia Campoli; Margherita De Santis; Marzia Cavalli; Nino D'Amore; Riccardo De Santis; Roberta Sorrentino; Rossella Tirelli; Silvia Fillo; Stella Lia; Vanessa Vera Fain |
| EPI_ISL_457825 | Army Medical Research Center - Scientific Department | Army Medical and Veterinary Research Center | Anna Anselmo; Antonella Fortunato; Florigio Lista; Francesco Giordani; Giovanni Faggioni; Nino D'Amore; Riccardo De Sanctis; Silvia Fillo; Vanessa Vera Fain |
| EPI_ISL_475830 to 475872, EPI_ISL_475874 to 475882, EPI_ISL_475884 to 475886, EPI_ISL_583573 to 583578, EPI_ISL_583630 to 583631, EPI_ISL_583633 to 583637, EPI_ISL_583639 to 583640, EPI_ISL_583642 to 583655, EPI_ISL_583657 to 583676, EPI_ISL_583678 to 583690, EPI_ISL_583883 to 583892, EPI_ISL_853733, EPI_ISL_853738, EPI_ISL_853749, EPI_ISL_853758, EPI_ISL_853779 to 853782, EPI_ISL_853790 to 853791, EPI_ISL_853793 to 853794, EPI_ISL_853797, EPI_ISL_853800 to 853801, EPI_ISL_853803 to 853804, EPI_ISL_853815, EPI_ISL_853818, EPI_ISL_853965 to 853972, EPI_ISL_853975 to 854040, EPI_ISL_854113 to 854175, EPI_ISL_854209, EPI_ISL_854213, EPI_ISL_854215 to 854216, EPI_ISL_854228, EPI_ISL_854243 to 854252, EPI_ISL_854254, EPI_ISL_854262 to 854281, EPI_ISL_854285 to 854294, EPI_ISL_854297, EPI_ISL_854301, EPI_ISL_854304 | | | |
| see above | Austrian Agency for Health and Food Safety (AGES) | Bergthaler laboratory, CeMM Research Center for Molecular Medicine of the Austrian Academy of Sciences | Adi Steinrigl; Alexander Lercher; Alexandra Popa; Andreas Bergthaler; Anna Schedl; Benedikt Agerer; Christian Paar; Christoph Bock; Christoph Bock; Daniela Schmid; Dorothee von Laer; Elisabeth Puchhammer-Stoeckl; Franz Allerberger; Gernot Walder; Gregor Hörmann; Guenter Weiss; Gunther Vogl; Henrique Colaco; Jakob-Wendelin Genger; Jan Laine; Judith Aberle; Kinga Rigler-Hohenwarter; Lukas Endler; Manfred Nairz; Mark Smyth; Martin Senekowitsch; Martin Senekowitsch; Michael Schuster; Michael Schuster; Peter Hufnagl; Peter Obrist; Rainer Gattringer; Sabine Sussitz-Rack; Stephan Aberle; Thomas Penz; Wegene Borena |
| EPI_ISL_415590 | BCCDC Public Health Laboratory | BCCDC Public Health Laboratory | Choi; Gilmour; Harrigan; Hoang; Kamelian; Krajden; Lapointe; Lee; Levett; Li; Loman; Prystajecky; Quick; Sekirov; Snutch; Tyson |
| EPI_ISL_500716 | BSL3 Lab Pendik Veterinary Control Institute | Department of Medicinal Genetics, Bursa Uluda University, Faculty of medicine By Sehime Gülsün Temel, Adem Alemdar, Kadir Yeilba | Ahmet SAIT; Cumhur ADIAY; Fahriye SARAC; Hakan ENUL; Kadir YESILBAG; Mustafa HASOKSUZ; Oguz KARABEY; Orbay SAYI; Osman ERGANIS; Serdar UZAR |
| EPI_ISL_491476 | BSL3 Lab, Pendik Veterinary Control Enstitue | Genomic Laboratory (GLAB), Istanbul Technical University | Ahmet SAIT; Cumhur ADIAY; Fahriye SARAC; Hakan ENUL; Kadir YESILBAG; Mustafa HASOKSUZ; Oguz KARABEY; Orbay SAYI; Osman ERGANIS; Serdar UZAR |
| EPI_ISL_539886, EPI_ISL_710613, EPI_ISL_732657, EPI_ISL_766645 | Barnakuten | The Public Health Agency of Sweden | Anna Risberg; Anna-Malin Linde; Department of Microbiology; Karin Tegmark-Wisell; Maria Lind Karlberg; Mattias Haukland; Mia Brytting; Olov Svartstrom; Oskar Karlsson Lindsjo; Reza Advani; Sandra Broddesson; The Public Health Agency of Sweden; Theresa Enkirch |
| EPI_ISL_855549 | Barts Health NHS Trust | Barts Health NHS Trust | BROAD; Beatrix; CUTINO-MOGUEL; Claire; David; Dola; HARRINGTON; KELE; KULASEGARAN-SHYLINI; Maria-Teresa; OWOYEMI; Raghavendran |
| EPI_ISL_842793 to 842802, EPI_ISL_842804 to 842809, EPI_ISL_842811 to 842825, EPI_ISL_842827 to 842832, EPI_ISL_842834 to 842835, EPI_ISL_842837 to 842840, EPI_ISL_842844, EPI_ISL_842847 to 842854, EPI_ISL_842856 to 842857, EPI_ISL_842859, EPI_ISL_842862 to 842865, EPI_ISL_842867 to 842868, EPI_ISL_842870 to 842871, EPI_ISL_842873 to 842886, EPI_ISL_842890 to 842895, EPI_ISL_842897 to 842901, EPI_ISL_842903 to 842905, EPI_ISL_842907 to 842929, EPI_ISL_842931, EPI_ISL_842933 to 842944, EPI_ISL_842947 to 842986, EPI_ISL_842988 to 842993, EPI_ISL_842995 to 843005, EPI_ISL_843007 to 843008, EPI_ISL_843010 to 843043, EPI_ISL_843045 to 843047, EPI_ISL_843049 to 843062, EPI_ISL_843064 to 843118, EPI_ISL_843121 to 843133, EPI_ISL_843135, EPI_ISL_843137 to 843145, EPI_ISL_843147 to 843150, EPI_ISL_865488 to 865489, EPI_ISL_865492 to 865498, EPI_ISL_865501 to 865522, EPI_ISL_865524 to 865527, EPI_ISL_865529 to 865560, EPI_ISL_865562 to 865564, EPI_ISL_865566 to 865570, EPI_ISL_865576 to 865580, EPI_ISL_865582 to 865583 | | | |
| see above | Barts Health NHS Trust | COVID-19 Genomics UK (COG-UK) Consortium | BROAD; Beatrix; CUTINO-MOGUEL; Claire; David; Dola; HARRINGTON; KELE; KULASEGARAN-SHYLINI; Maria-Teresa; OWOYEMI; Raghavendran; SHYLINI |
| EPI_ISL_541752, EPI_ISL_541754 to 541755, EPI_ISL_541757 to 541759, EPI_ISL_541761 to 541766, EPI_ISL_541768 to 541772, EPI_ISL_541775 to 541781, EPI_ISL_541783 | | | |
| see above | Barts Health NHS Trust | Wellcome Sanger Institute for the COVID-19 Genomics UK (COG-UK) consortium | Beatrix Kele; Cordelia Langford; David Harrington and Alex Alderton; David K. Jackson; Dominic Kwiatkowski; Ewan Harrison; Ian Johnston; John Sillitoe on behalf of the Wellcome Sanger Institute COVID-19 Surveillance Team; Mark Hopkins; Roberto Amato; Sonia Goncalves; Teresa Cutino-Moguel |
| EPI_ISL_457824 | Bezmialem Vakif University, Dept Microbiology, Medical School, Fatih, Istanbul, Turkey | Bezmialem Vakif University, Medical School & Beykoz Institute of Life Sciences & Biotechnology | Bilge Sumbul; Elif Karaaslan; Filiz Guney; Mehmet Z. Doymaz; Merve Kalkan; Nesibe Cetin |
| EPI_ISL_861865 to 861866 | Bioanalytika AG | University Hospital Basel, Clinical Bacteriology | Adrian Egli; Adrian Härri; Alfredo Mari; Hans Hirsch; Helena MB Seth-Smith; Julia Bielicki; Karoline Leuzinger; Madlen Stange; Manuel Battegay; Tim Roloff |
| EPI_ISL_814287 to 814290, EPI_ISL_814292, EPI_ISL_814351, EPI_ISL_814357 to 814361, EPI_ISL_814417 to 814421, EPI_ISL_814424 to 814438, EPI_ISL_814517 to 814522, EPI_ISL_814524 to 814525, EPI_ISL_814530, EPI_ISL_814532, EPI_ISL_814538 to 814539, EPI_ISL_814543 to 814544, EPI_ISL_814548, EPI_ISL_814610 to 814619, EPI_ISL_816686, EPI_ISL_816688 to 816693, EPI_ISL_816695 to 816728, EPI_ISL_816730 to 816769, EPI_ISL_816771 to 816934, EPI_ISL_816936 to 817001, EPI_ISL_817003 to 817064, EPI_ISL_817066 to 817068, EPI_ISL_817070 to 817122, EPI_ISL_842355 to 842398, EPI_ISL_842400 to 842445, EPI_ISL_842447 to 842470, EPI_ISL_842472 to 842493, EPI_ISL_842496 to 842502, EPI_ISL_842504 to 842511, EPI_ISL_842513 to 842524, EPI_ISL_842526 to 842544, EPI_ISL_842546 to 842582, EPI_ISL_842584 to 842591, EPI_ISL_842593 to 842596, EPI_ISL_842598 to 842608, EPI_ISL_868716 to 868769, EPI_ISL_868771 to 868785, EPI_ISL_868787 to 868791, EPI_ISL_868793 to 868808, EPI_ISL_868810 to 868817, EPI_ISL_868819 to 868826, EPI_ISL_868828 to 868830, EPI_ISL_868832 to 868888, EPI_ISL_868890 to 868900, EPI_ISL_868902 to 868903, EPI_ISL_868905, EPI_ISL_868907, EPI_ISL_868909 to 868929, EPI_ISL_868931 to 868937, EPI_ISL_868940, EPI_ISL_868942 to 868948, EPI_ISL_868950 to 869025, EPI_ISL_869027 to 869044, EPI_ISL_869047, EPI_ISL_869051 to 869054, EPI_ISL_869056, EPI_ISL_869058, EPI_ISL_869061 to 869062, EPI_ISL_869067, EPI_ISL_869069, EPI_ISL_869071 to 869075, EPI_ISL_869077 to 869078 | | | |
| see above | Bioinformatics and Biostatistics Lab, Advanced Sequencing Facility | COVID-19 Genomics UK (COG-UK) Consortium | Aengus Stewart; Chelsea Sawyer; Harshil Patel; Jerome Nicod; Laura Cubitt; Margaret Crawford |
| EPI_ISL_505003 | Biology Dpt | Microbiology and Infections Diseases | Annabelle Garnier; Audrey Ferrier-Rembert; Clarisse Vigne; Emilie Tessier; Emmanuelle Billon-Denis; Flora Nolent; Isabelle Drouet; Jean-Nicolas Tournier; Jessica Denis; Laurence Cheutin; Noémie Verguet; Olivier Ferraris; Olivier Gorgé |
| EPI_ISL_506041 | Biology Dpt, HIA Percy | Microbiology and Infectious Diseases Dpt | Annabelle Garnier; Audrey Ferrier-Rembert; Clarisse Vigne; Emilie Tessier; Emmanuelle Billon-Denis; Flora Nolent; Isabelle Drouet; Jean-Nicolas Tournier; Jessica Denis; Laurence Cheutin; Noémie Verguet; Olivier Ferraris; Olivier Gorgé |
| EPI_ISL_516079 to 516088 | Biomedical Sciences and Public Health, Polytechnic University of Marche | Biomedical Sciences and Public Health, Polytechnic University of Marche | Alessandrini, F.; Bagnarelli, P.; Caucci, S.; Di Sante, L.; Melchionda, F.; Menzo, S.; Onofri, V.; Tagliabracci, A.; Turchi, C. |
| EPI_ISL_450813, EPI_ISL_475540 | Bla Kustens halsocentral | The Public Health Agency of Sweden | Anna Risberg; Anna-Malin Linde; Karin Tegmark-Wisell; Maria Lind Karlberg; Mattias Haukland; Mia Brytting; Olof Norrby; Olov Svartstrom; Oskar Karlsson Lindsjo; Reza Advani; Sandra Broddesson; Theresa Enkirch |
| EPI_ISL_414521, EPI_ISL_732525, EPI_ISL_732528 to 732541, EPI_ISL_732543 to 732564, EPI_ISL_732658, EPI_ISL_755639 | Bundeswehr Institute of Microbiology | Bundeswehr Institute of Microbiology | Alexandra Rehn; Christina Bugert; Elham Khatamzas; Enrico Georgi; Joachim Bugert; Malena Bestehorn-Willmann; Markus Antwerpen; Markus H Antwerpen and Roman Wölfel; Mathias C Walter; Mathias Walter; Michael von Bergwelt-Baildon; Roman Wölfel; Sabine Zange |
| EPI_ISL_450832 | Byjorden vardcentral | The Public Health Agency of Sweden | Anna Risberg; Anna-Malin Linde; Karin Tegmark-Wisell; Maria Lind Karlberg; Mia Brytting; Olov Svartstrom; Oskar Karlsson Lindsjo; Pernilla Brunman; Theresa Enkirch |
| EPI_ISL_524476 | Bülach Hospital | Institute of Medical Virology, University of Zurich | Alexandra Trkola; Andrea Zbinden; Fiona Steiner; Gabriela Ziltener; Jon Huder; Jürg Böni; Maryam Zaheri; Michael Huber; Patrick Redli; Riccarda Capaul; Stefan Schmutz; Verena Kufner |
| EPI_ISL_539531 | C.H.U Nuestra Señora de Candelaria | Instituto de Salud Carlos III | A. Monzón; F. Casas; I; I. Jiménez; Iglesias-Caballero; M. Camarero; M. Cuesta; M. González-Esguevillas; M. Molinero Calamita; M. Zaballos; O. Díez; P. Jiménez; S. Juliá; S. Pozo; S. Varona |
| EPI_ISL_644229 to 644230, EPI_ISL_644250 to 644254, EPI_ISL_644256, EPI_ISL_644258 | CEPHR / Mater Hospital | Irish Coronavirus Sequencing Consortium - National Virus Reference Laboratory | Alejandro Abner Garcia Leon; Gabriel Gonzalez; Michael Carr; Patrick Mallon |
| EPI_ISL_644209 to 644215, EPI_ISL_644219 to 644227, EPI_ISL_644231 to 644236, EPI_ISL_644238 to 644240, EPI_ISL_644242 to 644245, EPI_ISL_644247 to 644248, EPI_ISL_644255, EPI_ISL_644257, EPI_ISL_644259 to 644265, EPI_ISL_644267 to 644319, EPI_ISL_644321, EPI_ISL_644323 to 644324, EPI_ISL_644326 | | | |

| | | | |
|---|---|---|---|
| to 644345 | | | |
| see above | CEPHR / Vincent's Hospital | Irish Coronavirus Sequencing Consortium - National Virus Reference Laboratory | Alejandro Abner Garcia Leon; Gabriel Gonzalez; Michael Carr; Patrick Mallon |
| EPI_ISL_780418 | CERBA LAB | CERBA LAB | Costa JM; Haïm-Boukobza S; Hedbaut E; Lecorche E; Malek Ramdane; Olivi M; Roquebert B; Trombert S; Verdurme L |
| EPI_ISL_421455, EPI_ISL_421479 to 421480, EPI_ISL_421485 to 421486 | CH Barreiro Montijo | Instituto Nacional de Saude (INSA) | Guiomar et al |
| EPI_ISL_420041, EPI_ISL_420049 to 420050, EPI_ISL_420056 to 420057, EPI_ISL_421500, EPI_ISL_421509 to 421511, EPI_ISL_428353, EPI_ISL_428359 to 428360, EPI_ISL_443309, EPI_ISL_443316 | | | |
| see above | CH Compiègne Laboratoire de Biologie | National Reference Center for Viruses of Respiratory Infections, Institut Pasteur, Paris | Angela Brisebarre; Etienne Simon-Lorière; Flora Donati; Marion Barbet; Maud Vanpeene; Mélanie Albert; Méline Bizard; Olivia Raulin; Raulin Olivia; Sylvie Behillil; Sylvie van der Werf; Vincent Enouf |
| EPI_ISL_416493, EPI_ISL_420044, EPI_ISL_420053, EPI_ISL_428350 | CH Jean de Navarre Laboratoire de Biologie | National Reference Center for Viruses of Respiratory Infections, Institut Pasteur, Paris | Angela Brisebarre; Etienne Simon-Lorière; Flora Donati; Marion Barbet; Maud Vanpeene; Mélanie Albert; Méline Bizard; Mélnie Albert; Sylvie Behillil; Sylvie van der Werf; Vincent Enouf |
| EPI_ISL_428358, EPI_ISL_428366 | CH Jeanne de Navarre Laboratoire de Biologie | National Reference Center for Viruses of Respiratory Infections, Institut Pasteur, Paris | Angela Brisebarre; Etienne Simon-Lorière; Flora Donati; Marion Barbet; Maud Vanpeene; Mélanie Albert; Méline Bizard; Sylvie Behillil; Sylvie van der Werf; Vincent Enouf |
| EPI_ISL_421482 to 421484 | CH VN Gaia - Espinho | Instituto Nacional de Saude (INSA) | Guiomar et al |
| EPI_ISL_792008 to 792015 | CH de Bethune - Laboratoire de Biologie Médicale | National Reference Center for Viruses of Respiratory Infections, Institut Pasteur, Paris | Angela Brisebarre; Camille Capel; Etienne Simon-Lorière; Léa Pilorgé; Marion Barbet; Maud Vanpeene; Méline Bizard; Sylvie Behillil; Sylvie van der Werf; Vincent Enouf |
| EPI_ISL_418006 | CHBarreiro Montijo | Instituto Nacional de Saude (INSA) | Guiomar et al |
| EPI_ISL_418017 | CHMT | Instituto Nacional de Saude (INSA) | Guiomar et al |
| EPI_ISL_418222 | CHRU Bretonneau - Serv. Bacterio-Virol. | National Reference Center for Viruses of Respiratory Infections, Institut Pasteur, Paris | Angela Brisebarre; Etienne Simon-Lorière; Fabiana Gambaro; Flora Donati; Julien Marlet; Marion Barbet; Maud Vanpeene; Mélanie Albert; Méline Bizard; Sylvie Behillil; Sylvie van der Werf; Vincent Enouf |
| EPI_ISL_416502 to 416513, EPI_ISL_443289 to 443294, EPI_ISL_734168 to 734169 | CHRU Pontchaillou - Laboratoire de Virologie | National Reference Center for Viruses of Respiratory Infections, Institut Pasteur, Paris | Angela Brisebarre; Camille Capel; Etienne Simon-Lorière; Flora Donati; Gisèle Lagathu; Marion Barbet; Maud Vanpeene; Mélanie Albert; Méline Bizard; Mélnie Albert; Sylvie Behillil; Sylvie van der Werf; Vincent Enouf |
| EPI_ISL_613545 to 613559, EPI_ISL_614281 | CHRU Pontchaillou - Laboratoire de Virologie 2, rue Henri Le Guilloux | National Reference Center for Viruses of Respiratory Infections, Institut Pasteur, Paris | Angela Brisebarre; Camille Capel; Etienne Simon-Lorière; Gisèle Lagathu; Marion Barbet; Maud Vanpeene; Méline Bizard; Sylvie Behillil; Sylvie van der Werf; Vincent Enouf |
| EPI_ISL_418027, EPI_ISL_421453, EPI_ISL_421464 to 421465 | CHTMAD | Instituto Nacional de Saude (INSA) | Guiomar et al |
| EPI_ISL_791991 to 791999 | CHU - Hôpital Cavale Blanche | National Reference Center for Viruses of Respiratory Infections, Institut Pasteur, Paris | Angela Brisebarre; Camille Capel; Etienne Simon-Lorière; Léa Pilorgé; Marion Barbet; Maud Vanpeene; Méline Bizard; Sylvie Behillil; Sylvie van der Werf; Vincent Enouf |
| EPI_ISL_418219, EPI_ISL_443265 to 443283, EPI_ISL_754853 to 754856 | CHU - Hôpital Cavale Blanche - Labo. de Virologie | National Reference Center for Viruses of Respiratory Infections, Institut Pasteur, Paris | Angela Brisebarre; Camille Capel; Etienne Simon-Lorière; Fabiana Gambaro; Flora Donati; Léa Pilorge; Marion Barbet; Maud Vanpeene; Mélanie Albert; Méline Bizard; Pilorge léa; Sylvie Behillil; Sylvie van der Werf; Vincent Enouf |
| EPI_ISL_754842 | CHU Amiens - Labo virologie | National Reference Center for Viruses of Respiratory Infections, Institut Pasteur, Paris | Angela Brisebarre; Camille Capel; Castelain Sandrine; Etienne Simon-Lorière; François Catherine; Marion Barbet; Maud Vanpeene; Méline Bizard; Sylvie Behillil; Sylvie van der Werf; Vincent Enouf |
| EPI_ISL_645169 to 645172, EPI_ISL_649954 to 649971, EPI_ISL_660372, EPI_ISL_700326, EPI_ISL_768823, EPI_ISL_779786, EPI_ISL_779837 to 779844 | | | |
| see above | CHU Bordeaux | CNR Virus des Infections Respiratoires - France SUD | Antonin Bal; Bruno Lina; Camille Ciccone; Gregory Destras; Gwendolyne Burfin; Hadrien Règue; Isabelle Garrigue; Laurence Josset; Marie-Edith Lafon; Martine Valette; Pantxika Bellecave; Pascale Trimoulet; Quentin Semanas |
| EPI_ISL_641546 to 641550, EPI_ISL_645188 to 645196, EPI_ISL_660353 to 660371, EPI_ISL_745312 to 745315, EPI_ISL_745335 to 745336 | CHU Clermont-Ferrand | CNR Virus des Infections Respiratoires - France SUD | Amélie Brebion; Antonin Bal; Audrey Mirand; Bruno Lina; Christel Regagnon; Christine Archimbaud; Cécile Henquell; Gregory Destras; Gwendolyne Burfin; Hadrien Règue; Hélène Chabrolles; Laurence Josset; Martine Chambon; Martine Valette; Maxime Bisseux; Patricia Combes; Quentin Semanas |
| EPI_ISL_418002 | CHU Coimbra | Instituto Nacional de Saude (INSA) | Guiomar et al |
| EPI_ISL_418005 | CHU Coimbra - Pediátrico | Instituto Nacional de Saude (INSA) | Guiomar et al |
| EPI_ISL_416751 to 416752 | CHU Gabriel Montpied | CNR Virus des Infections Respiratoires - France SUD | Alexandre; Antonin; Bal; Bouscambert-Duchamp; Brengel-Pesce; Bruno.; Cheynet; Destras; Florence; Gaymard; Gregory; Josset; Karen; Laurence; Lina; Martine; Maude; Morfin-Sherpa; Valette; Valérie |
| EPI_ISL_641556, EPI_ISL_644681 to 644696, EPI_ISL_660672 to 660703, EPI_ISL_660705 to 660709 | CHU Montpellier | CNR Virus des Infections Respiratoires - France SUD | Antonin Bal; Bruno Lina; Gregory Destras; Gwendolyne Burfin; Hadrien Règue; Laurence Josset; Martine Valette; Michel Segondy; Quentin Semanas; Vincent Foulongne |
| EPI_ISL_645217 to 645218, EPI_ISL_649173 to 649187, EPI_ISL_663249 to 663284, EPI_ISL_666721 to 666724, EPI_ISL_700371 | CHU Nantes | CNR Virus des Infections Respiratoires - France SUD | Antonin Bal; Bruno Lina; Celine Bressollette; Gregory Destras; Gwendolyne Burfin; Hadrien Règue; Laurence Josset; Louise Castain; Martine Valette; Quentin Semanas; Virginie Ferré |
| EPI_ISL_644697 to 644702, EPI_ISL_645197 to 645209, EPI_ISL_645215, EPI_ISL_660710 to 660731, EPI_ISL_663243 to 663248 | CHU Nîmes | CNR Virus des Infections Respiratoires - France SUD | Antonin Bal; Bruno Lina; Gregory Destras; Gwendolyne Burfin; Hadrien Règue; Jean-Philippe Lavigne; Laurence Josset; Marie-Josée Carles; Martine Valette; Maxence Lotellier; Quentin Semanas; Stephan Robin |
| EPI_ISL_645216, EPI_ISL_663227 to 663242, EPI_ISL_707781 to 707782, EPI_ISL_768822, EPI_ISL_779785 | CHU Poitiers | CNR Virus des Infections Respiratoires - France SUD | Agnès Beby-Defaux; Antonin Bal; Bruno Lina; Clément Jousselin; Gregory Destras; Gwendolyne Burfin; Hadrien Règue; Laurence Josset; Magali Garcia; Martine Valette; Nicolas Lévêque; Quentin Semanas |
| EPI_ISL_591099, EPI_ISL_591541 to 591547, EPI_ISL_593855 to 593901, EPI_ISL_603216 to 603220, EPI_ISL_671941 to 671972, EPI_ISL_671975 to 671977, EPI_ISL_751448 to 751499, EPI_ISL_754136 to 754139, EPI_ISL_804370 to 804378, EPI_ISL_833457 to 833480, EPI_ISL_848063 to 848064, EPI_ISL_852814 to 852818 | | | |
| see above | CHU Purpan - Laboratoire de Virologie - Institut Fédératif de Biologie | CHU Purpan - Laboratoire de Virologie - Institut Fédératif de Biologie | Boyer P.; Carcenac R.; Dubois M.; Harter A.; Izopet J.; Latour J.; Ranger N.; Tremeaux P. |
| EPI_ISL_424993, EPI_ISL_434616 to 434635, EPI_ISL_482879 to 482889 | CHU Purpan - Laboratoire de Virologie - Institut Fédératif de Biologie | Laboratoire de virologie - École Nationale Vétérinaire de Toulouse | Croville, G.; Guerin; Guillaume Croville; J.-L. and Izopet, J.; Jacques Izopet; Jean-Luc Guérin |
| EPI_ISL_641551 to 641555, EPI_ISL_660373 to 660377, EPI_ISL_660665 to 660671 | CHU Toulouse | CNR Virus des Infections Respiratoires - France SUD | Antonin Bal; Bruno Lina; Gregory Destras; Gwendolyne Burfin; Hadrien Règue; Jean Michel Mansuy; Laurence Josset; Martine Valette; Quentin Semanas |
| EPI_ISL_735391 | CHU Tours | CNR Virus des Infections Respiratoires - France SUD | Antonin Bal; Bruno Lina; Catherine Gaudy-Graffin; Claudia Gonzalez; Florence Morfin; Gregory Destras; Gwendolyne Burfin; Hadrien Regue; Julien Marlet; Karl Stefic; Laurence Josset; Martine Valette; Maude Bouscambert; Quentin Semanas; Thibault Guinoiseau; Yahia Mekki |
| EPI_ISL_443261 to 443264 | CHU de Dijon - Laboratoire de Virologie | National Reference Center for Viruses of Respiratory Infections, Institut Pasteur, Paris | Angela Brisebarre; Etienne Simon-Lorière; Flora Donati; Jean-Baptiste Bour; Marion Barbet; Maud Vanpeene; Mélanie Albert; Méline Bizard; Sylvie Behillil; Sylvie van der Werf; Vincent Enouf |
| EPI_ISL_644673 to 644680, EPI_ISL_645174 to 645183, EPI_ISL_663206 to 663226, EPI_ISL_666711 to 666720 | CHU de Limoges | CNR Virus des Infections Respiratoires - France SUD | Antonin Bal; Bruno Lina; Gregory Destras; Gwendolyne Burfin; Hadrien Règue; Laurence Josset; Martine Valette; Quentin Semanas; Sylvie Rogez |

| | | | |
|---|---|---|---|
| EPI_ISL_645173 | CHU de Nice | CNR Virus des Infections Respiratoires - France SUD | Antonin Bal; Bruno Lina; Gregory Destras; Gwendolyne Burfin; Géraldine Gonfrier; Hadrien Règue; Laurence Josset; Martine Valette; Quentin Semanas; Valérie Giordanengo |
| EPI_ISL_641528 | CHU de Nice - Hôpital Archet 10 | CNR Virus des Infections Respiratoires - France SUD | Antonin Bal; Bruno Lina; Gregory Destras; Gwendolyne Burfin; Géraldine Gonfrier; Hadrien Règue; Laurence Josset; Martine Valette; Quentin Semanas; Valérie Giordanengo |
| EPI_ISL_641529 | CHU de Nice - Hôpital Archet 11 | CNR Virus des Infections Respiratoires - France SUD | Antonin Bal; Bruno Lina; Gregory Destras; Gwendolyne Burfin; Géraldine Gonfrier; Hadrien Règue; Laurence Josset; Martine Valette; Quentin Semanas; Valérie Giordanengo |
| EPI_ISL_641530 | CHU de Nice - Hôpital Archet 12 | CNR Virus des Infections Respiratoires - France SUD | Antonin Bal; Bruno Lina; Gregory Destras; Gwendolyne Burfin; Géraldine Gonfrier; Hadrien Règue; Laurence Josset; Martine Valette; Quentin Semanas; Valérie Giordanengo |
| EPI_ISL_641531 | CHU de Nice - Hôpital Archet 13 | CNR Virus des Infections Respiratoires - France SUD | Antonin Bal; Bruno Lina; Gregory Destras; Gwendolyne Burfin; Géraldine Gonfrier; Hadrien Règue; Laurence Josset; Martine Valette; Quentin Semanas; Valérie Giordanengo |
| EPI_ISL_641532 | CHU de Nice - Hôpital Archet 14 | CNR Virus des Infections Respiratoires - France SUD | Antonin Bal; Bruno Lina; Gregory Destras; Gwendolyne Burfin; Géraldine Gonfrier; Hadrien Règue; Laurence Josset; Martine Valette; Quentin Semanas; Valérie Giordanengo |
| EPI_ISL_641533 | CHU de Nice - Hôpital Archet 15 | CNR Virus des Infections Respiratoires - France SUD | Antonin Bal; Bruno Lina; Gregory Destras; Gwendolyne Burfin; Géraldine Gonfrier; Hadrien Règue; Laurence Josset; Martine Valette; Quentin Semanas; Valérie Giordanengo |
| EPI_ISL_641534 | CHU de Nice - Hôpital Archet 16 | CNR Virus des Infections Respiratoires - France SUD | Antonin Bal; Bruno Lina; Gregory Destras; Gwendolyne Burfin; Géraldine Gonfrier; Hadrien Règue; Laurence Josset; Martine Valette; Quentin Semanas; Valérie Giordanengo |
| EPI_ISL_641520, EPI_ISL_693387 to 693390, EPI_ISL_779846 | CHU de Nice - Hôpital Archet 2 | CNR Virus des Infections Respiratoires - France SUD | Antonin Bal; Bruno Lina; Gregory Destras; Gwendolyne Burfin; Géraldine Gonfrier; Hadrien Règue; Laurence Josset; Martine Valette; Quentin Semanas; Valérie Giordanengo |
| EPI_ISL_641522 | CHU de Nice - Hôpital Archet 3 | CNR Virus des Infections Respiratoires - France SUD | Antonin Bal; Bruno Lina; Gregory Destras; Gwendolyne Burfin; Géraldine Gonfrier; Hadrien Règue; Laurence Josset; Martine Valette; Quentin Semanas; Valérie Giordanengo |
| EPI_ISL_641523 | CHU de Nice - Hôpital Archet 4 | CNR Virus des Infections Respiratoires - France SUD | Antonin Bal; Bruno Lina; Gregory Destras; Gwendolyne Burfin; Géraldine Gonfrier; Hadrien Règue; Laurence Josset; Martine Valette; Quentin Semanas; Valérie Giordanengo |
| EPI_ISL_641524 | CHU de Nice - Hôpital Archet 5 | CNR Virus des Infections Respiratoires - France SUD | Antonin Bal; Bruno Lina; Gregory Destras; Gwendolyne Burfin; Géraldine Gonfrier; Hadrien Règue; Laurence Josset; Martine Valette; Quentin Semanas; Valérie Giordanengo |
| EPI_ISL_641525 | CHU de Nice - Hôpital Archet 6 | CNR Virus des Infections Respiratoires - France SUD | Antonin Bal; Bruno Lina; Gregory Destras; Gwendolyne Burfin; Géraldine Gonfrier; Hadrien Règue; Laurence Josset; Martine Valette; Quentin Semanas; Valérie Giordanengo |
| EPI_ISL_641526 | CHU de Nice - Hôpital Archet 7 | CNR Virus des Infections Respiratoires - France SUD | Antonin Bal; Bruno Lina; Gregory Destras; Gwendolyne Burfin; Géraldine Gonfrier; Hadrien Règue; Laurence Josset; Martine Valette; Quentin Semanas; Valérie Giordanengo |
| EPI_ISL_641521 | CHU de Nice - Hôpital Archet 8 | CNR Virus des Infections Respiratoires - France SUD | Antonin Bal; Bruno Lina; Gregory Destras; Gwendolyne Burfin; Géraldine Gonfrier; Hadrien Règue; Laurence Josset; Martine Valette; Quentin Semanas; Valérie Giordanengo |
| EPI_ISL_641527 | CHU de Nice - Hôpital Archet 9 | CNR Virus des Infections Respiratoires - France SUD | Antonin Bal; Bruno Lina; Gregory Destras; Gwendolyne Burfin; Géraldine Gonfrier; Hadrien Règue; Laurence Josset; Martine Valette; Quentin Semanas; Valérie Giordanengo |
| EPI_ISL_641519, EPI_ISL_641535 to 641545, EPI_ISL_645184 to 645187, EPI_ISL_649942 to 649953, EPI_ISL_660326 to 660352, EPI_ISL_660432 | CHU de Saint-Étienne Hôpital Nord | CNR Virus des Infections Respiratoires - France SUD | Antonin Bal; Bruno Lina; Bruno Pozzetto; Gregory Destras; Gwendolyne Burfin; Hadrien Règue; Issam Bechri; Laurence Josset; Manon Vogrig; Marine Delorme; Martine Valette; Quentin Semanas; Sylvie Gonzalo; Sylvie Pillet; Thomas Bourlet |
| EPI_ISL_418024 | CHUA - Faro | Instituto Nacional de Saude (INSA) | Guiomar et al |
| EPI_ISL_417988, EPI_ISL_417990 to 417991, EPI_ISL_417994 to 417996, EPI_ISL_418010 to 418016 | CHULC - H Curry Cabral | Instituto Nacional de Saude (INSA) | Guiomar et al |
| EPI_ISL_417992 to 417993 | CHULC - H D Estefania | Instituto Nacional de Saude (INSA) | Guiomar et al |
| EPI_ISL_420043, EPI_ISL_420061 | CMIP | National Reference Center for Viruses of Respiratory Infections, Institut Pasteur, Paris | Angela Brisebarre; Etienne Simon-Lorière; Flora Donati; Marion Barbet; Maud Vanpeene; Mélanie Albert; Méline Bizard; Sylvie Behillil; Sylvie van der Werf; Vincent Enouf |
| EPI_ISL_410486, EPI_ISL_416745 to 416746, EPI_ISL_508912 to 508930, EPI_ISL_508933, EPI_ISL_508937, EPI_ISL_508939 to 508940, EPI_ISL_508942, EPI_ISL_508945, EPI_ISL_508948, EPI_ISL_508951, EPI_ISL_508953 to 508957, EPI_ISL_508961 to 508965, EPI_ISL_508967, EPI_ISL_508969 to 508974, EPI_ISL_508976 to 508977, EPI_ISL_508979 to 508997, EPI_ISL_508999 to 509002, EPI_ISL_525536 to 525538, EPI_ISL_525540 to 525543, EPI_ISL_578176 to 578177, EPI_ISL_582110 to 582120, EPI_ISL_582122, EPI_ISL_582508, EPI_ISL_623098 to 623102, EPI_ISL_636476 to 636477, EPI_ISL_636479, EPI_ISL_636484 to 636485, EPI_ISL_636487, EPI_ISL_636489 to 636490, EPI_ISL_639974 to 639977, EPI_ISL_639979 to 639983, EPI_ISL_639985 to 640014, EPI_ISL_676542 to 676573, EPI_ISL_678494 to 678508, EPI_ISL_678534 to 678547, EPI_ISL_681261 to 681263, EPI_ISL_683334 to 683384, EPI_ISL_683386 to 683402, EPI_ISL_684107, EPI_ISL_692733 to 692768, EPI_ISL_693488 to 693514, EPI_ISL_700327, EPI_ISL_728304 to 728320, EPI_ISL_728516 to 728551, EPI_ISL_730640 to 730651, EPI_ISL_732677 to 732703, EPI_ISL_745307 to 745311, EPI_ISL_745316 to 745334, EPI_ISL_745337 to 745397, EPI_ISL_768827 to 768828, EPI_ISL_779780, EPI_ISL_779783 to 779784, EPI_ISL_779787 to 779813, EPI_ISL_779845 | | | |
| see above | CNR Virus des Infections Respiratoires - France SUD | CNR Virus des Infections Respiratoires - France SUD | Alexandre; Alexandre Gaymard; Antonin; Antonin Bal; Bal; Bouscambert-Duchamp; Brengel-Pesce; Bruno Lina; Bruno.; Carine Moustaud; Cheynet; Claudia Gonzalez; Destras; Emilie Frobert; Florence; Florence Morfin-Sherpa; Gaymard; Gregory; Gregory Destras; Gregory Queromes; Gwendolyne Burfin; Hadrien Règue; Josset; Karen; Laurence; Laurence Josset; Lina; Martine; Martine Valette; Maude; Maude Bouscambert-Duchamp; Morfin-Sherpa; Quentin Semanas; Raphaelle Lamy; Solenne Brun; Valette; Valérie |
| EPI_ISL_416994, EPI_ISL_418248 to 418249 | COMPLEJO ASISTENCIAL UNIVERSITARIO DE BURGOS | Instituto de Salud Carlos III | A. Monzón; F. Casas; G. Hospital: ------; I. Jiménez; I. Megias-Lobon G.; Iglesias-Caballero; M. Camarero; M. Camarero S. Pozo F. Casas I. Jiménez P. Jiménez M. Zaballos A. Monzón; M. Cuesta; M. González-Esguevillas; M. Molinero Calamita; M. Zaballos; P. Jiménez; S. Juliá; S. Juliá M. Cuesta I. Megias Lobón; S. Pozo; S. Varona |
| EPI_ISL_451935, EPI_ISL_452140, EPI_ISL_452142, EPI_ISL_452148 to 452152 | CUB Hopital Erasme Laboratoire d'Anatomie Pathologique | CUB Hopital Erasme Laboratoire d'Anatomie Pathologique | Dr Nicky D'Haene; Dr. Nicky D'Haene; Dr.Nikcy D'Haene; Isabelle Salmon; Nicky D'Haene; Nikcy D'Haene; Prof. Isabelle Salmon |
| EPI_ISL_428364, EPI_ISL_428367, EPI_ISL_443300 to 443302, EPI_ISL_443306, EPI_ISL_443311 to 443313, EPI_ISL_443317 | Cabinet Médical | National Reference Center for Viruses of Respiratory Infections, Institut Pasteur, Paris | Angela Brisebarre; Etienne Simon-Lorière; Flora Donati; Marion Barbet; Maud Vanpeene; Mélanie Albert; Méline Bizard; Sylvie Behillil; Sylvie van der Werf; Vincent Enouf |
| EPI_ISL_418235 | Cabinet médical | National Reference Center for Viruses of Respiratory Infections, Institut Pasteur, Paris | Angela Brisebarre; Etienne Simon-Lorière; Flora Donati; Marion Barbet; Maud Vanpeene; Mélanie Albert; Méline Bizard; Sylvie Behillil; Sylvie van der Werf; Vincent Enouf |
| EPI_ISL_447054 | Cantacuzino National Military-Medical Institute for Research and Development | Cantacuzino Institute | A.Cretu; L.Ustea; M.Lazar |
| EPI_ISL_730623 | Cantonal Hospital Frauenfeld | Institute of Medical Virology, University of Zurich | Alexandra Trkola; Gabriela Ziltener; Jürg Böni; Maryam Zaheri; Michael Huber; Stefan Schmutz; Verena Kufner |
| EPI_ISL_524478 | Cantonal Hospital Winterthur | Institute of Medical Virology, University of Zurich | Alexandra Trkola; Andrea Zbinden; Fiona Steiner; Gabriela Ziltener; Jon Huder; Jürg Böni; Maryam Zaheri; Michael Huber; Patrick Redli; Riccarda Capaul; Stefan Schmutz; Verena Kufner |
| EPI_ISL_710612 | Capio Bro VC | The Public Health Agency of Sweden | Department of Microbiology; The Public Health Agency of Sweden |
| EPI_ISL_534230 to 534233, EPI_ISL_560981 | Capio S:t Gorans sjukhus | The Public Health Agency of Sweden | Anna Risberg; Anna-Malin Linde; Karin Tegmark-Wisell; Maria Lind Karlberg; Mattias Haukland; Mia Brytting; Olov Svartstrom; Oskar Karlsson Lindsjo; Petra Edquist; Reza Advani; Sandra Broddesson |
| EPI_ISL_806783 | Casa di cura Di Lorenzo- Avezzano | Istituto Zooprofilattico Sperimentale dell'Abruzzo e Molise "G. Caporale" | Ancora M; Calistri P; Cammà C; Curini V; Di Domenico M; Di Pasquale A; Lorusso A; Mangone I; Marcacci M; Puglia I; Rinaldi A; Savini G |

| | | | |
|---|---|---|---|
| EPI_ISL_450413 | Center for Diagnostics, Institute of Medical Microbiology, Virology and Hygiene | University Medical Center Hamburg-Eppendorf | Huang, J.; Pfefferle; S. and Fischer, N. |
| EPI_ISL_419654 to 419656, EPI_ISL_419658 to 419674, EPI_ISL_437993 to 438009, EPI_ISL_438011 to 438020, EPI_ISL_438022 to 438026, EPI_ISL_438028 to 438043, EPI_ISL_438045 to 438103, EPI_ISL_438105 to 438115, EPI_ISL_438117 to 438123, EPI_ISL_438125 to 438128, EPI_ISL_475770, EPI_ISL_475772 to 475796, EPI_ISL_475798 to 475804, EPI_ISL_475806 to 475812, EPI_ISL_583563 to 583564, EPI_ISL_583566, EPI_ISL_583568 to 583570, EPI_ISL_583692 to 583694, EPI_ISL_583696 to 583698, EPI_ISL_583700 to 583702, EPI_ISL_583704 to 583715, EPI_ISL_583718 to 583721, EPI_ISL_583723 to 583726, EPI_ISL_583869 to 583881, EPI_ISL_853751, EPI_ISL_853759, EPI_ISL_853822 to 853853, EPI_ISL_853877 to 853913, EPI_ISL_853957 to 853961, EPI_ISL_854218 to 854219, EPI_ISL_854241, EPI_ISL_854253, EPI_ISL_854258, EPI_ISL_854295, EPI_ISL_854299 | | | |
| see above | Center for Virology, Medical University of Vienna | Bergthaler laboratory, CeMM Research Center for Molecular Medicine of the Austrian Academy of Sciences | Adi Steinrigl; Alexander Lercher; Alexandra Popa; Andreas Bergthaler; Anna Schedl; Benedikt Agerer; Christian Paar; Christoph Bock; Christoph Bock; Daniela Schmid; Dorothee von Laer; Elisabeth Puchhammer-Stoeckl; Elisabeth Puchhammer-Stöckl; Franz Allerberger; Gernot Walder; Gregor Hörmann; Guenter Weiss; Gunther Vogl; Henrique Colaco; Jakob-Wendelin Genger; Jan Laine; Judith Aberle; Kinga Rigler-Hohenwarter; Lukas Endler; Manfred Nairz; Mark Smyth; Martin Senekowitsch; Martin Senekowitsch; Michael Schuster; Michael Schuster; Peter Hufnagl; Peter Obrist; Rainer Gattringer; Sabine Sussitz-Rack; Stephan Aberle; Thomas Penz; Wegene Borena |
| EPI_ISL_516414 to 516425, EPI_ISL_677700 to 677702, EPI_ISL_677706, EPI_ISL_677708, EPI_ISL_678248 to 678249 | Center for public health - Skopje | Research Center for Genetic Engineering and Biotechnology "Georgi D. Efremov" , Macedonian Academy of Sciences and Arts | RCGEB - MASA |
| EPI_ISL_755572 to 755574 | Center of Advanced Studies and Technology, CAST | Center of Advanced Studies and Technology, CAST | De Fabritiis, S.; Ferrante, R.; Mandatori, D. |
| EPI_ISL_888674 to 888676 | Center of Advanced Studies and Technology, Molecular Genetics Laboratory | Center of Advanced Studies and Technology, Molecular Genetics Laboratory | De Fabritiis Simone; Ferrante Rossella; Mandatori Domitilla |
| EPI_ISL_523950 | Center of Medical Microbiology, Virology, and Hospital Hygiene, University of Duesseldorf | Center of Medical Microbiology, Virology, and Hospital Hygiene, Heinrich Heine University Düsseldorf | Alexander Dilthey; Andreas Walker; Daniel Strelow; Hendrik Streeck; Jessica Nicolai; Jörg Timm; Klaus Pfeffer; Malte Kohns Vasconcelos; Marek Korencak; Maximilian Damagnez; Tobias Wienemann; Torsten Houwaart |
| EPI_ISL_413488, EPI_ISL_414497 to 414499, EPI_ISL_414504 to 414509, EPI_ISL_414574, EPI_ISL_417457 to 417459, EPI_ISL_417461 to 417468, EPI_ISL_419541 to 419543, EPI_ISL_419545 to 419547, EPI_ISL_419549 to 419552, EPI_ISL_425120 to 425140, EPI_ISL_452180, EPI_ISL_523927 to 523929, EPI_ISL_523931 to 523949, EPI_ISL_602510, EPI_ISL_602512, EPI_ISL_602514 to 602517, EPI_ISL_602533 to 602540, EPI_ISL_602542 to 602545, EPI_ISL_602547 to 602552, EPI_ISL_717976, EPI_ISL_779922 to 779953, EPI_ISL_832364 to 832374 | | | |
| see above | Center of Medical Microbiology, Virology, and Hospital Hygiene, University of Duesseldorf | Center of Medical Microbiology, Virology, and Hospital Hygiene, University of Duesseldorf | Alexander Dilthey; Andreas Walker; Ashley-Jane Duplessis; Bjorn-Erik Jensen; Björn-Erik Jensen; Björn-ErikJensen; Daniel Strelow; Detlef Kindgen-Milles; Hendrik Streeck; Jessica Nicolai; Jorg Timm; Jörg Timm; Klaus Pfeffer; Lisanna Hülse; Malte Kohns Vasconcelos; Marcel Andree; Marek Korencak; Maximilian Damagnez; Nadine Lübke; Ortwin Adams; Sandra Hauka; Teresa Tamayo; Tina Senff; Tobias Wienemann; Torsten Feldt; Torsten Houwaart |
| EPI_ISL_523930 | Center of Medical Microbiology, Virology, and Hospital Hygiene, University of Duesseldorf | Universitätstr.1 40225 Düsseldorf Germany | Alexander Dilthey; Andreas Walker; Daniel Strelow; Hendrik Streeck; Jessica Nicolai; Jörg Timm; Klaus Pfeffer; Malte Kohns Vasconcelos; Marek Korencak; Maximilian Damagnez; Tobias Wienemann; Torsten Houwaart |
| EPI_ISL_815257 to 815382, EPI_ISL_815384 to 815397 | Centogene | Centogene | Krishna Kumar Kandaswamy; Peter Bauer; Vivi Hue-Trang Lieu |
| EPI_ISL_457750, EPI_ISL_459962 to 459964 | Centogene AG | Centogene AG | Dr. Krishna Kumar Kandaswamy; Prof. Dr. Peter Bauer |
| EPI_ISL_486410, EPI_ISL_486417 | Centrala laboratorija | Latvian Biomedical Research and Study Centre | Ivars Silamielis; Jana Oste; Jnis Kloviš; Kaspars Megnis; Marta Priedte; Monta Ustinova; Stella Lapia; Uga Dumpis; Vita Rovte; ikita Zrelovs |
| EPI_ISL_548256 | Centralsjukhuset | The Public Health Agency of Sweden | Anna Risberg; Anna-Malin Linde; Karin Tegmark-Wisell; Maria Lind Karlberg; Mattias Haukland; Mia Brytting; Olov Svartstrom; Oskar Karlsson Lindsjo; Petra Edquist; Reza Advani; Sandra Broddesson |
| EPI_ISL_414624, EPI_ISL_416494 | Centre Hositalier Universitaire de Rouen Laboratoire de Virologie | National Reference Center for Viruses of Respiratory Infections, Institut Pasteur, Paris | Angela Brisebarre; Etienne Simon-Lorière; Flora Donati; Flora Donati Vincent Enouf; Jean-Christophe Plantier; Marion Barbet; Maud Vanpeene; Méline Bizard; Mélnie Albert; Sylvie Behillil; Sylvie van der Werf; Vincent Enouf |
| EPI_ISL_508958, EPI_ISL_509012 | Centre Hospitalier Alpes Leman | CNR Virus des Infections Respiratoires - France SUD | Alexandre Gaymard; Antonin Bal; Bruno Lina; Carine Moustaud; Florence Morfin-Sherpa; Gregory Destras; Gwendolyne Burfin; Laurence Josset; Martine Valette; Maude Bouscambert-Duchamp; Raphaelle Lamy; Solenne Brun |
| EPI_ISL_414627 to 414630, EPI_ISL_414634 to 414638, EPI_ISL_415653 to 415654, EPI_ISL_416495 to 416497, EPI_ISL_418218, EPI_ISL_418220 to 418221, EPI_ISL_418223 to 418225, EPI_ISL_418227 to 418228, EPI_ISL_418231, EPI_ISL_418236 to 418239, EPI_ISL_429968 | | | |
| see above | Centre Hospitalier Compiègne Laboratoire de Biologie | National Reference Center for Viruses of Respiratory Infections, Institut Pasteur, Paris | Angela Brisebarre; Etienne Simon-Lorière; Fabiana Gambaro; Flora Donati; Flora Donati Vincent Enouf; Marion Barbet; Maud Vanpeene; Mélanie Albert; Méline Bizard; Mélnie Albert; Raulin Olivia; Sylvie Behillil; Sylvie van der Werf; Vincent Enouf |
| EPI_ISL_860898 | Centre Hospitalier Compiègne-Noyon - Laboratoire | National Reference Center for Viruses of Respiratory Infections, Institut Pasteur, Paris | Angela Brisebarre; Camille Capel; Emond Jean-Philippe; Etienne Simon-Lorière; Marion Barbet; Maud Vanpeene; Méline Bizard; Sylvie Behillil; Sylvie van der Werf; Vincent Enouf |
| EPI_ISL_418428, EPI_ISL_508947 | Centre Hospitalier Lucien Hussel | CNR Virus des Infections Respiratoires - France SUD | Alexandre Gaymard; Antonin Bal; Bruno Lina; Carine Moustaud; Florence Morfin-Sherpa; Gregory Destras; Gwendolyne Burfin; Laurence Josset; Martine Valette; Maude Bouscambert-Duchamp; Raphaelle Lamy; Solenne Brun |
| EPI_ISL_508966, EPI_ISL_508975, EPI_ISL_508998, EPI_ISL_636478, EPI_ISL_636480, EPI_ISL_636482 to 636483, EPI_ISL_636486 | | | |
| see above | Centre Hospitalier Pierre Oudot | CNR Virus des Infections Respiratoires - France SUD | Alexandre Gaymard; Antonin Bal; Bruno Lina; Carine Moustaud; Florence Morfin-Sherpa; Gregory Destras; Gwendolyne Burfin; Hadrien Règue; Laurence Josset; Martine Valette; Maude Bouscambert-Duchamp; Raphaelle Lamy; Solenne Brun |
| EPI_ISL_414633 | Centre Hospitalier René Dubois Laboratoire de Microbiologie - Bât A | National Reference Center for Viruses of Respiratory Infections, Institut Pasteur, Paris | Angela Brisebarre; Flora Donati Vincent Enouf; Marion Barbet; Maud Vanpeene; Méline Bizard; Mélnie Albert; Pascale Martres; Sylvie Behillil; Sylvie van der Werf |
| EPI_ISL_414625 | Centre Hospitalier Régional Universitaire de Nantes Laboratoire de Virologie | National Reference Center for Viruses of Respiratory Infections, Institut Pasteur, Paris | Angela Brisebarre; Flora Donati Vincent Enouf; Marianne Coste-Burel; Marion Barbet; Maud Vanpeene; Méline Bizard; Mélnie Albert; Sylvie Behillil; Sylvie van der Werf |
| EPI_ISL_418418 to 418419, EPI_ISL_420617, EPI_ISL_508879 to 508880, EPI_ISL_508959 to 508960, EPI_ISL_639978 | Centre Hospitalier Saint Joseph Saint Luc | CNR Virus des Infections Respiratoires - France SUD | Alexandre Gaymard; Antonin Bal; Bruno Lina; Carine Moustaud; Florence Morfin-Sherpa; Gregory Destras; Gwendolyne Burfin; Hadrien Règue; Laurence Josset; Martine Valette; Maude Bouscambert-Duchamp; Raphaelle Lamy; Solenne Brun |
| EPI_ISL_416757, EPI_ISL_417340, EPI_ISL_418426, EPI_ISL_419183, EPI_ISL_419185 to 419186, EPI_ISL_420620, EPI_ISL_508938, EPI_ISL_508944, EPI_ISL_508968, EPI_ISL_525539, EPI_ISL_582121, EPI_ISL_636481, EPI_ISL_639984 | | | |
| see above | Centre Hospitalier de Bourg en Bresse | CNR Virus des Infections Respiratoires - France SUD | Alexandre; Alexandre Gaymard; Antonin; Antonin Bal; Bal; Bouscambert-Duchamp; Brengel-Pesce; Bruno Lina; Bruno.; Carine Moustaud; Cheynet; Destras; Florence; Florence Morfin-Sherpa; Gaymard; Gregory; Gregory Destras; Gwendolyne Burfin; Hadrien Règue; Josset; Karen; Laurence; Laurence Josset; Lina; Martine; Martine Valette; Maude; Maude Bouscambert-Duchamp; Morfin-Sherpa; Raphaelle Lamy; Solenne Brun; Valette; Valérie |
| EPI_ISL_417338, EPI_ISL_418413, EPI_ISL_419174 to 419176, EPI_ISL_419187 to 419188, EPI_ISL_420612 to 420614, EPI_ISL_508875 to 508876, EPI_ISL_508941, EPI_ISL_508943, EPI_ISL_508946, EPI_ISL_508949 to 508950, EPI_ISL_508952, EPI_ISL_509005 | | | |
| see above | Centre Hospitalier de Macon | CNR Virus des Infections Respiratoires - France SUD | Alexandre Gaymard; Antonin Bal; Bruno Lina; Carine Moustaud; Florence Morfin-Sherpa; Gregory Destras; Gwendolyne Burfin; Laurence Josset; Martine Valette; Maude Bouscambert-Duchamp; Raphaelle Lamy; Solenne Brun |
| EPI_ISL_416749, EPI_ISL_418414 to 418415, EPI_ISL_418417, EPI_ISL_419168, EPI_ISL_508881 | Centre Hospitalier de Valence | CNR Virus des Infections Respiratoires - France SUD | Alexandre; Alexandre Gaymard; Antonin; Antonin Bal; Bal; Bouscambert-Duchamp; Brengel-Pesce; Bruno Lina; Bruno.; Carine Moustaud; Cheynet; Destras; Florence; Florence Morfin-Sherpa; Gaymard; Gregory; Gregory Destras; Gwendolyne Burfin; Josset; Karen; Laurence; Laurence Josset; Lina; Martine; Martine Valette; Maude; Maude Bouscambert-Duchamp; Morfin-Sherpa; Raphaelle Lamy; Solenne Brun; Valette; Valérie |
| EPI_ISL_508932, EPI_ISL_508978, EPI_ISL_509006, EPI_ISL_509015 | Centre Hospitalier de Villefranche | CNR Virus des Infections Respiratoires - France SUD | Alexandre Gaymard; Antonin Bal; Bruno Lina; Carine Moustaud; Florence Morfin-Sherpa; Gregory Destras; Gwendolyne Burfin; Laurence Josset; Martine Valette; Maude Bouscambert-Duchamp; Raphaelle Lamy; Solenne Brun |
| EPI_ISL_418412 | Centre Hospitalier des Vals d'Ardeche | CNR Virus des Infections Respiratoires - France SUD | Alexandre Gaymard; Antonin Bal; Bruno Lina; Carine Moustaud; Florence Morfin-Sherpa; Gregory Destras; Gwendolyne Burfin; Laurence Josset; Martine Valette; Maude Bouscambert-Duchamp; Raphaelle Lamy; Solenne Brun |
| EPI_ISL_509016 | Centre Hospitalier du Haut-Bugey | CNR Virus des Infections Respiratoires - France SUD | Alexandre Gaymard; Antonin Bal; Bruno Lina; Carine Moustaud; Florence Morfin-Sherpa; Gregory Destras; Gwendolyne Burfin; Laurence Josset; Martine Valette; Maude Bouscambert-Duchamp; Raphaelle Lamy; Solenne Brun |
| EPI_ISL_779847 | Centre Hospotalier Cannes | CNR Virus des Infections Respiratoires - France SUD | Antonin Bal; Bruno Lina; Gregory Destras; Gwendolyne Burfin; Hadrien Règue; Laurence Josset; Martine Valette; Quentin Semanas |
| EPI_ISL_443310 | Centre de santé Filieris | National Reference Center for Viruses of Respiratory | Angela Brisebarre; Etienne Simon-Lorière; Flora Donati; Marion Barbet; Maud Vanpeene; Mélanie Albert; Méline Bizard; Sylvie Behillil; Sylvie van der |

| | | | |
|---|---|---|---|
| | | Infections, Institut Pasteur, Paris | Werf; Vincent Enouf |
| EPI_ISL_466873 | Centre for Clinical Infection and Diagnostics Research and Genomics Innovation Unit | Respiratory Virus Unit, Microbiology Services Colindale, Public Health England | Ali Awan; Chloe Fisher; Gaia Nebbia; Luke Snell; PHE Covid Sequencing Team |
| EPI_ISL_483922 to 483924, EPI_ISL_483928 to 483929, EPI_ISL_483931, EPI_ISL_483937, EPI_ISL_483945, EPI_ISL_483947 to 483948, EPI_ISL_483952 to 483953, EPI_ISL_483956, EPI_ISL_483959, EPI_ISL_483961, EPI_ISL_483964, EPI_ISL_483966 to 483968, EPI_ISL_483971, EPI_ISL_483976, EPI_ISL_483982, EPI_ISL_483986, EPI_ISL_483992, EPI_ISL_484005, EPI_ISL_484008, EPI_ISL_484012, EPI_ISL_484019 to 484020, EPI_ISL_484028, EPI_ISL_484034, EPI_ISL_484036, EPI_ISL_484053, EPI_ISL_484056, EPI_ISL_484059, EPI_ISL_484064, EPI_ISL_484070, EPI_ISL_484072, EPI_ISL_484079, EPI_ISL_484082, EPI_ISL_484085, EPI_ISL_484087, EPI_ISL_484091, EPI_ISL_484093, EPI_ISL_484095 to 484096, EPI_ISL_484104, EPI_ISL_484110, EPI_ISL_484115, EPI_ISL_484117, EPI_ISL_484125 to 484126, EPI_ISL_484129, EPI_ISL_484131, EPI_ISL_484134, EPI_ISL_484136, EPI_ISL_484145, EPI_ISL_484150, EPI_ISL_484152, EPI_ISL_484168 to 484170, EPI_ISL_484174, EPI_ISL_484189, EPI_ISL_484191 | | | |
| see above | Centre for Clinical Infection and Diagnostics Research and Genomics Innovation Unit, Guy's and St. Thomas' NHS Trust | COVID-19 Genomics UK (COG-UK) Consortium | Ali Raza Awan; Chloe Fisher; Jonathan Edgeworth; Luke Snell; Penny Cliff; Rahul Batra |
| EPI_ISL_560650, EPI_ISL_560652, EPI_ISL_560656, EPI_ISL_560658, EPI_ISL_560660 to 560661, EPI_ISL_560668, EPI_ISL_560671, EPI_ISL_560673, EPI_ISL_560678 to 560679, EPI_ISL_560684, EPI_ISL_560686, EPI_ISL_560697, EPI_ISL_560700 to 560701, EPI_ISL_560705, EPI_ISL_560719, EPI_ISL_560723, EPI_ISL_560727, EPI_ISL_560734, EPI_ISL_560736, EPI_ISL_561363, EPI_ISL_561365 | | | |
| see above | Centre for Clinical Infection and Diagnostics Research and Genomics Innovation Unit, Guy's and St. Thomas' NHS Trust | Centre for Clinical Infection and Diagnostics Research and Genomics Innovation Unit, Guy's and St. Thomas' NHS Trust | Ali Raza Awan; Chloe Fisher; Jonathan Edgeworth; Luke Snell; Rahul Batra |
| EPI_ISL_453666 to 453668, EPI_ISL_453670 to 453674, EPI_ISL_453676 to 453693, EPI_ISL_461970 to 461971, EPI_ISL_461973 to 461981, EPI_ISL_461983 to 461997, EPI_ISL_475239 to 475249, EPI_ISL_475251 to 475265, EPI_ISL_475267 to 475293, EPI_ISL_475295 to 475303, EPI_ISL_475306 to 475341, EPI_ISL_479175 to 479181, EPI_ISL_479183 to 479194, EPI_ISL_484407 to 484408, EPI_ISL_484410 to 484413, EPI_ISL_484415 to 484432, EPI_ISL_493615 to 493623, EPI_ISL_493625 to 493634, EPI_ISL_493636 to 493638, EPI_ISL_493640 to 493641, EPI_ISL_493643 to 493665, EPI_ISL_499270 to 499273, EPI_ISL_499275 to 499329, EPI_ISL_507131 to 507157, EPI_ISL_512385 to 512388, EPI_ISL_512392 to 512422, EPI_ISL_512424 to 512435, EPI_ISL_514455 to 514457, EPI_ISL_514459 to 514466, EPI_ISL_514468 to 514513, EPI_ISL_517510 to 517530, EPI_ISL_517532 to 517541, EPI_ISL_526433 to 526436, EPI_ISL_529272, EPI_ISL_529303, EPI_ISL_529341 to 529352, EPI_ISL_529373 to 529377, EPI_ISL_529431 to 529432, EPI_ISL_529481, EPI_ISL_529491, EPI_ISL_529511, EPI_ISL_529525, EPI_ISL_529668 to 529674, EPI_ISL_535024 to 535025, EPI_ISL_535027 to 535042, EPI_ISL_549341, EPI_ISL_549343, EPI_ISL_549360, EPI_ISL_549363 to 549364, EPI_ISL_549366, EPI_ISL_549460 to 549476, EPI_ISL_559983, EPI_ISL_572715 to 572716, EPI_ISL_572721, EPI_ISL_573253 to 573265, EPI_ISL_573870, EPI_ISL_577040 to 577063, EPI_ISL_577065 to 577067, EPI_ISL_577193 to 577205, EPI_ISL_577207 to 577219, EPI_ISL_577299 to 577307, EPI_ISL_577309 to 577326, EPI_ISL_577328 to 577339, EPI_ISL_577341 to 577345, EPI_ISL_577381 to 577382, EPI_ISL_577385 to 577391, EPI_ISL_577393 to 577400, EPI_ISL_577545 to 577546, EPI_ISL_584169 to 584184, EPI_ISL_584187, EPI_ISL_584193, EPI_ISL_584195, EPI_ISL_584200 to 584206, EPI_ISL_584212 to 584215, EPI_ISL_584809 to 584810, EPI_ISL_595100 to 595114, EPI_ISL_611526, EPI_ISL_611532, EPI_ISL_611535, EPI_ISL_611537 to 611538, EPI_ISL_611555, EPI_ISL_611557, EPI_ISL_611585, EPI_ISL_611589 to 611590, EPI_ISL_611592, EPI_ISL_611638 to 611639, EPI_ISL_611689, EPI_ISL_611754 to 611755, EPI_ISL_611761, EPI_ISL_611771 to 611773, EPI_ISL_611841, EPI_ISL_611916, EPI_ISL_611927 to 611928, EPI_ISL_611933 to 611934, EPI_ISL_611957 to 611960, EPI_ISL_611962, EPI_ISL_611971, EPI_ISL_611983, EPI_ISL_611998, EPI_ISL_612068 to 612069, EPI_ISL_612089, EPI_ISL_612092, EPI_ISL_612094, EPI_ISL_613282 to 613283, EPI_ISL_613286 to 613298, EPI_ISL_613302 to 613306, EPI_ISL_626677 to 626678, EPI_ISL_626684, EPI_ISL_626689 to 626690, EPI_ISL_626738, EPI_ISL_626778, EPI_ISL_626792, EPI_ISL_626878 to 626880, EPI_ISL_626895, EPI_ISL_626936, EPI_ISL_627056 to 627057, EPI_ISL_627116, EPI_ISL_627128, EPI_ISL_627144, EPI_ISL_627150, EPI_ISL_628312 to 628333, EPI_ISL_637278 to 637279, EPI_ISL_637305, EPI_ISL_637369 to 637372, EPI_ISL_637387, EPI_ISL_637402, EPI_ISL_637411, EPI_ISL_637432, EPI_ISL_637453, EPI_ISL_637459, EPI_ISL_637466 to 637467, EPI_ISL_637491 to 637499, EPI_ISL_637502, EPI_ISL_637528, EPI_ISL_637610, EPI_ISL_637659, EPI_ISL_637674, EPI_ISL_637714 to 637715, EPI_ISL_637717 to 637720, EPI_ISL_637780 to 637783, EPI_ISL_637853, EPI_ISL_637889, EPI_ISL_637892, EPI_ISL_637920 to 637921, EPI_ISL_637931 to 637932, EPI_ISL_637949, EPI_ISL_637951, EPI_ISL_637986, EPI_ISL_638002, EPI_ISL_638006, EPI_ISL_638026, EPI_ISL_638050, EPI_ISL_638065, EPI_ISL_638134 to 638137, EPI_ISL_638249 to 638257, EPI_ISL_638265 to 638273, EPI_ISL_638275 to 638292, EPI_ISL_638454 to 638458, EPI_ISL_638911 to 638913, EPI_ISL_639006 to 639007, EPI_ISL_650168 to 650169, EPI_ISL_650221, EPI_ISL_650248 to 650249, EPI_ISL_650379, EPI_ISL_650388, EPI_ISL_650401, EPI_ISL_650566 to 650571, EPI_ISL_650575 to 650578, EPI_ISL_650581, EPI_ISL_650584 to 650585, EPI_ISL_650600 to 650605, EPI_ISL_650608, EPI_ISL_650711, EPI_ISL_650941, EPI_ISL_650947, EPI_ISL_650949, EPI_ISL_650956 to 650957, EPI_ISL_650964 to 650966, EPI_ISL_650980 to 650982, EPI_ISL_651066 to 651069, EPI_ISL_651099, EPI_ISL_651187, EPI_ISL_651277, EPI_ISL_652103 to 652107, EPI_ISL_652109 to 652110, EPI_ISL_652620 to 652626, EPI_ISL_664137, EPI_ISL_664191, EPI_ISL_664302, EPI_ISL_664374 to 664377, EPI_ISL_664492, EPI_ISL_664526, EPI_ISL_664556 to 664559, EPI_ISL_664679, EPI_ISL_664690, EPI_ISL_664746, EPI_ISL_664763, EPI_ISL_664767, EPI_ISL_664840 to 664855, EPI_ISL_665338, EPI_ISL_665347 to 665348, EPI_ISL_665391, EPI_ISL_665487 to 665488, EPI_ISL_665616, EPI_ISL_665625, EPI_ISL_665652, EPI_ISL_665723, EPI_ISL_665726, EPI_ISL_665849, EPI_ISL_666107 to 666109, EPI_ISL_678831, EPI_ISL_679971, EPI_ISL_680016 to 680036, EPI_ISL_680562 to 680570, EPI_ISL_702870, EPI_ISL_723521 to 723557, EPI_ISL_727697 to 727723, EPI_ISL_727734 to 727738, EPI_ISL_727740 to 727750, EPI_ISL_727774 to 727806, EPI_ISL_741348, EPI_ISL_741357 to 741358, EPI_ISL_741658, EPI_ISL_741675, EPI_ISL_741693, EPI_ISL_741697, EPI_ISL_741716 to 741723, EPI_ISL_741725 to 741759, EPI_ISL_742260, EPI_ISL_763396, EPI_ISL_763407, EPI_ISL_763425, EPI_ISL_763451, EPI_ISL_763465, EPI_ISL_763471, EPI_ISL_763476, EPI_ISL_763495, EPI_ISL_763505, EPI_ISL_763512, EPI_ISL_763516, EPI_ISL_763521, EPI_ISL_763541, EPI_ISL_763550, EPI_ISL_763595, EPI_ISL_763606 to 763612, EPI_ISL_763617, EPI_ISL_763624, EPI_ISL_763659 to 763661, EPI_ISL_763672, EPI_ISL_763676, EPI_ISL_763690, EPI_ISL_763693, EPI_ISL_763734, EPI_ISL_763755, EPI_ISL_763791, EPI_ISL_763798 to 763800, EPI_ISL_763804 to 763806, EPI_ISL_763823 to 763825, EPI_ISL_763827 to 763836, EPI_ISL_763865, EPI_ISL_764007 to 764027, EPI_ISL_764029 to 764030, EPI_ISL_764036, EPI_ISL_764081, EPI_ISL_764094 to 764096, EPI_ISL_764101 to 764102, EPI_ISL_764118 to 764125, EPI_ISL_764136, EPI_ISL_764169, EPI_ISL_764172, EPI_ISL_764193, EPI_ISL_765112 to 765116, EPI_ISL_765119 to 765142, EPI_ISL_765144 to 765148, EPI_ISL_816153, EPI_ISL_816172, EPI_ISL_816174, EPI_ISL_816194 to 816205, EPI_ISL_816207 to 816209, EPI_ISL_816213 to 816215, EPI_ISL_816220 to 816225, EPI_ISL_841755 to 841759, EPI_ISL_841767, EPI_ISL_841769 to 841775, EPI_ISL_841778, EPI_ISL_841780 to 841784, EPI_ISL_841786 to 841806, EPI_ISL_841808 to 841819, EPI_ISL_841821 to 841848, EPI_ISL_841850, EPI_ISL_841852 to 841861, EPI_ISL_841863 to 841864, EPI_ISL_841866 to 841870, EPI_ISL_841978, EPI_ISL_842004, EPI_ISL_842010 to 842014, EPI_ISL_867930 to 867945, EPI_ISL_867948 to 867967, EPI_ISL_867969 to 867972, EPI_ISL_867974 to 867978, EPI_ISL_867980 to 867981, EPI_ISL_868356 | | | |
| see above | Centre for Enzyme Innovation, University of Portsmouth / Translational Research Laboratory, Portsmouth Hospitals NHS Trust | COVID-19 Genomics UK (COG-UK) Consortium | ; Allyson Lloyd; Angela Beckett; Anoop Chauhan; Ethan Butcher; Garry Scarlett; Kelly Bicknell; Robert Impey; Samuel Robson; Sarah Wyllie; Scott Elliott; Sharon Glaysher; Yann Bourgeois |
| EPI_ISL_453665 | Centre for Enzyme Innovation, University of Portsmouth / Translational Research Laboratory, Portsmouth Hospitals NHS Trust | University of Portsmouth | ; Allyson Lloyd; Angela Beckett; Anoop Chauhan; Ethan Butcher; Garry Scarlett; Kelly Bicknell; Robert Impey; Samuel Robson; Sarah Wyllie; Scott Elliott; Sharon Glaysher; Yann Bourgeois |
| EPI_ISL_413550 | Centre for Human and Zoonotic Virology (CHAZVY), College of Medicine University of Lagos/Lagos University Teaching Hospital (LUTH), part of the Laboratory Network of the Nigeria Centre for Disease Control (NCDC) | African Centre of Excellence for Genomics of Infectious Diseases (ACEGID), Redeemer's University, Ede, Osun State, Nigeria | Ajogbasile F.V.; Folarin O.A.; Ihekweazu C. Happi C.T.; Kayode A.; Oguzie J.; Oluniyi P.E. |
| EPI_ISL_508934 to 508936, EPI_ISL_666682 to 666686, EPI_ISL_693391, EPI_ISL_700372 to 700410 | Centre hospitalier Métropole Savoie | CNR Virus des Infections Respiratoires - France SUD | Alexandre Gaymard; Antonin Bal; Bruno Lina; Carine Dumollard; Carine Moustaud; Florence Morfin-Sherpa; Gregory Destras; Gwendolyne Burfin; Hadrien Règue; Jérôme Grosjean; Laurence Josset; Martine Valette; Maude Bouscambert-Duchamp; Quentin Semanas; Raphaelle Lamy; Solenne Brun |
| EPI_ISL_639739 | Centre of Nanotechnologies, INCD IMT-Bucuresti (National Institute for Research and Development in Microtechnologies - Bucharest) | Centre of Nanotechnologies, INCD IMT-Bucuresti (National Institute for Research and Development in Microtechnologies - Bucharest) | Gogianu; L. and Baisan, M.; Salceanu, A. |
| EPI_ISL_413647, EPI_ISL_417997 to 417999 | Centro Hospital do Porto, E.P.E. - H. Geral de Santo Antonio | Instituto Nacional de Saude (INSA) | Guiomar et al; Helena Ramos; Inês Costa; Joana Isidro; Joana Mendonça; João Paulo Gomes; Luís Vieira; Pedro Pechirra; Raquel Guiomar; Vítor Borges |
| EPI_ISL_417986 to 417987, EPI_ISL_417989 | Centro Hospitalar e Universitario de Sao Joao, Porto | Instituto Nacional de Saude (INSA) | Guiomar et al |
| EPI_ISL_413648 | Centro Hospitalar e Universitário de Sao Joao, Porto | Instituto Nacional de Saude (INSA) | Inês Costa; Joana Isidro; Joana Mendonça; João Paulo Gomes; João Tiago Guimarães; Luís Vieira; Pedro Pechirra; Raquel Guiomar; Vítor Borges |
| EPI_ISL_468761 to 468764, EPI_ISL_500369 to 500372, EPI_ISL_500374 to 500381, EPI_ISL_500383, EPI_ISL_500386 to 500394, EPI_ISL_500396 to 500401, EPI_ISL_500403 to 500411, EPI_ISL_500413, EPI_ISL_500415 to 500417, EPI_ISL_500419, EPI_ISL_500421 to 500435, EPI_ISL_500438 to 500442, EPI_ISL_500444 to 500458, EPI_ISL_537382 to 537400, EPI_ISL_537402 to 537420, EPI_ISL_537423 to 537437, EPI_ISL_537439 to 537444, EPI_ISL_537447 to 537449, EPI_ISL_537452 to 537455, EPI_ISL_537457 to 537464, EPI_ISL_537466, EPI_ISL_537811 to 537833, EPI_ISL_537835 to 537847, EPI_ISL_537849 to 537852, EPI_ISL_537854 to 537866, EPI_ISL_537868, EPI_ISL_537871 to 537873, EPI_ISL_732748 to 732753, EPI_ISL_732755 to 732759, EPI_ISL_732761 to 732764, EPI_ISL_732766 to 732777, EPI_ISL_732780, EPI_ISL_732783, EPI_ISL_732785 to 732789, EPI_ISL_732793 to 732801, EPI_ISL_732804 to 732808, EPI_ISL_732810, EPI_ISL_732813 to 732814, EPI_ISL_732816 to 732818 | | | |
| see above | Centro de Investigación Biomédica de La Rioja - Hospital San Pedro Logroño | SeqCOVID-SPAIN consortium/IBV(CSIC) | José Manuel Azcona Gutiérrez; María Pilar Bea Escudero; María de Toro; Miriam Blasco Alberdi and SeqCOVID-SPAIN consortium |
| EPI_ISL_450520 to 450524, EPI_ISL_639632 to 639633, EPI_ISL_639636, EPI_ISL_639640, EPI_ISL_639642, EPI_ISL_639648, EPI_ISL_639650 to 639651, EPI_ISL_639654 to 639657, EPI_ISL_639663, EPI_ISL_639672, EPI_ISL_639676 to 639680, EPI_ISL_639683, EPI_ISL_770038 to 770039, EPI_ISL_770042, EPI_ISL_770045, EPI_ISL_770048 to 770049, EPI_ISL_770052, EPI_ISL_770054, EPI_ISL_770056, EPI_ISL_770058 to 770059, EPI_ISL_770061 | | | |
| see above | Centrl Laboratorija | Latvian Biomedical Research and Study Centre | Ivars Silamielis; Jana Oste; Jnis Kloviš; Jnis Pjalkovskis; Kaspars Megnis; Marta Priedte; Monta Ustinova; Stella Lapia; Uga Dumpis; Vita Rovte; ikita Zrelovs |
| EPI_ISL_486390 to 486391, EPI_ISL_486411 to 486416, EPI_ISL_486418 to 486421, EPI_ISL_486437, EPI_ISL_492988 to 492992, EPI_ISL_501286 to 501289, EPI_ISL_501808, EPI_ISL_501817, EPI_ISL_501895 to 501896, EPI_ISL_501915, EPI_ISL_501922, EPI_ISL_515196, EPI_ISL_534200 to 534202, EPI_ISL_534206 to 534208, EPI_ISL_534210 to 534211, EPI_ISL_534220, EPI_ISL_534222 to 534223 | | | |
| see above | Centrl laboratorija | Latvian Biomedical Research and Study Centre | Ivars Silamielis; Jana Oste; Jnis Kloviš; Jnis Pjalkovskis; Kaspars Megnis; Marta Priedte; Monta Ustinova; Stella Lapia; Uga Dumpis; Vita Rovte; ikita Zrelovs |
| EPI_ISL_802994 to 802995 | Charite Universitatsmedizin Berlin, Institut fur Virologie, Charitéplatz 1, 10117 Berlin, Germany | Charite Universitatsmedizin Berlin, Institut fur Virologie, Charitéplatz 1, 10117 Berlin, Germany | Barbara Mühlemann; Christian Drosten; Julia Schneider; Julia Tesch; Jörn Beheim-Schwarzbach; Talitha Veith; Terry Jones; Tobias Bleicker; Victor M Corman |
| EPI_ISL_516629 to 516638, EPI_ISL_516640 to 516645 | Charite Universitatsmedizin Berlin, Institut fur Virologie/Labor Berlin | Charite Universitatsmedizin Berlin, Institut fur Virologie/Labor Berlin | Barbara Muhlemann; Christian Drosten; Julia Schneider; Jörn Beheim-Schwarzbach; Talitha Veith; Terry Jones; Victor M Corman |
| EPI_ISL_862125, EPI_ISL_862127 to 862128, EPI_ISL_862132, EPI_ISL_862137 to 862138, EPI_ISL_862145, EPI_ISL_862159 | Charité Universitätsmedizin Berlin, Institut fur Virologie/Labor Berlin | Charité Universitätsmedizin Berlin, Institut für Virologie | Barbara Mühlemann; Christian Drosten; Julia Schneider; Julia Tesch; Jörn Beheim-Schwarzbach; Talitha Veith; Terry Jones; Tobias Bleicker; Victor M Corman |

| | | | |
|---|---|---|---|
| EPI_ISL_754174 to 754175 | Charité Universitätsmedizin Berlin, Institut für Virologie | Charité Universitätsmedizin Berlin, Institut für Virologie | Barbara Mühlemann; Christian Drosten; Julia Schneider; Jörn Beheim-Schwarzbach; Talitha Veith; Terry Jones; Victor M Corman |
| EPI_ISL_729346, EPI_ISL_729348, EPI_ISL_729355 to 729357, EPI_ISL_729360, EPI_ISL_729365 to 729366, EPI_ISL_729370, EPI_ISL_729377, EPI_ISL_729382, EPI_ISL_729387 to 729388, EPI_ISL_729398, EPI_ISL_729400, EPI_ISL_729405 to 729406, EPI_ISL_729408 to 729410, EPI_ISL_729413, EPI_ISL_729473 to 729497, EPI_ISL_729499, EPI_ISL_729514, EPI_ISL_729529 to 729534, EPI_ISL_729555 to 729556, EPI_ISL_729558, EPI_ISL_729598 to 729607, EPI_ISL_753702 to 753712, EPI_ISL_753714 to 753715, EPI_ISL_753717 to 753721, EPI_ISL_753723 to 753727, EPI_ISL_753729 to 753739, EPI_ISL_753741 to 753747, EPI_ISL_753749 to 753762, EPI_ISL_753764 to 753765, EPI_ISL_753767 to 753769, EPI_ISL_753771 to 753800, EPI_ISL_753802 to 753813, EPI_ISL_753815 to 753921, EPI_ISL_753923 to 754058, EPI_ISL_754185 to 754194, EPI_ISL_806526 to 806542, EPI_ISL_853401 to 853404, EPI_ISL_869118 | | | |
| see above | Charité Universitätsmedizin Berlin, Institut für Virologie/Labor Berlin | Charité Universitätsmedizin Berlin, Institut für Virologie | Barbara Mühlemann; Christian Drosten; Cornelia Schlee; Julia Schneider; Julia Tesch; Jörn Beheim-Schwarzbach; Talitha Veith; Terry Jones; Tobias Bleicker; Tomasz Zemojtel; Victor M Corman |
| EPI_ISL_818342 | Charité Universitätsmedizin Berlin, Institute of Virology | Charité Universitätsmedizin Berlin, Institute of Virology | Barbara Mühlemann; Christian Drosten; Julia Schneider; Jörn Beheim-Schwarzbach; Talitha Veith; Terry Jones; Victor M Corman |
| EPI_ISL_824920 to 824921, EPI_ISL_824942, EPI_ISL_824944, EPI_ISL_856680 to 856681, EPI_ISL_856683, EPI_ISL_868949 | Charité Universitätsmedizin Berlin, Institute of Virology, Charitéplatz 1, 10117 Berlin, Germany | Charité Universitätsmedizin Berlin, Institute of Virology, Charitéplatz 1, 10117 Berlin, Germany | Barbara Mühlemann; Christian Drosten; Julia Schneider; Julia Tesch; Jörn Beheim-Schwarzbach; Talitha Veith; Terry Jones; Tobias Bleicker; Victor M Corman |
| EPI_ISL_406862 | Charité Universitätsmedizin Berlin, Institute of Virology; Institut für Mikrobiologie der Bundeswehr, Munich | Charité Universitätsmedizin Berlin, Institute of Virology | Barbara Mühlemann; Christian Drosten; Julia Schneider; Markus Antwerpen; Roman Wölfel; Talitha Veith; Victor M Corman |
| EPI_ISL_872100 to 872103 | Chu Tivoli | GIGA Medical Genomics | Bouchra Boujemla; Cécile Meex; Keith Durkin; Maria Artesi; Marie-Pierre Hayette; Pierrette Melin; Raphaël Boreux; Sébastien Bontems; Vincent Bours |
| EPI_ISL_443315 | Château de la Source | National Reference Center for Viruses of Respiratory Infections, Institut Pasteur, Paris | Angela Brisebarre; Etienne Simon-Lorière; Flora Donati; Marion Barbet; Maud Vanpeene; Mélanie Albert; Méline Bizard; Sylvie Behillil; Sylvie van der Werf; Vincent Enouf |
| EPI_ISL_491115 | Cicin-Sain Lab | Cicin-Sain Lab | Kathrin Eschke; Luka Cicin-Sain; M. Zeeshan Chaudhry; Yeonsu Kim |
| EPI_ISL_462450 to 462476 | Clinical Center, University of Sarajevo | Charite Universitatsmedizin Berlin, Institute of Virology | Almedina Hadzihasanovic-Moro; Amela Dedeic-Ljubovic; Barbara Muehlemann; Christian Drosten; Irma Salimovic-Besic; Jorn Beheim-Schwarzbach; Julia Schneider; Selma Mutevelic; Suzana Arapcic; Talitha Veith; Terry Jones; Victor M Corman |
| EPI_ISL_677715, EPI_ISL_677718 | Clinical Hospital - Bitola | Research Center for Genetic Engineering and Biotechnology "Georgi D. Efremov" , Macedonian Academy of Sciences and Arts | RCGEB - MASA |
| EPI_ISL_516426 to 516431, EPI_ISL_677676 to 677677, EPI_ISL_677716 to 677717, EPI_ISL_677720 to 677721, EPI_ISL_677724, EPI_ISL_678250, EPI_ISL_678259 | | | |
| see above | Clinical Hospital - Shtip | Research Center for Genetic Engineering and Biotechnology "Georgi D. Efremov" , Macedonian Academy of Sciences and Arts | RCGEB - MASA |
| EPI_ISL_483570 | Clinical Microbiology Laboratory- Basurto University Hospita | Biocruces-Bizkaia | Ana Belén Belén de la Hoz; Estibaliz Ugalde-Zarraga; José Luis Díaz de Tuesta del Arco; Matxalen Vidal-García; Mikel J. Urrutikoetxea-Gutierrez; Mº Carmen Nieto Toboso |
| EPI_ISL_483566, EPI_ISL_483571 to 483573, EPI_ISL_486876, EPI_ISL_489833 to 489835, EPI_ISL_490202 to 490204, EPI_ISL_490977 | Clinical Microbiology Laboratory- Basurto University Hospital | Biocruces-Bizkaia | Ana Belén Belén de la Hoz; Estibaliz Ugalde-Zarraga; José Luis Díaz de Tuesta del Arco; Matxalen Vidal-García; Mikel J. Urrutikoetxea-Gutierrez; Mº Carmen Nieto Toboso |
| EPI_ISL_735511 | Clinical Pathology and Microbiology, San Gallicano Dermatologic Institute IRCCS | National Institute for Infectious Diseases, INMI, "L. Spallanzani" IRCCS | A. Di Caro; B. Bartolini; C.E.M Gruber; E. Giombini; F. Carletti; F. Ensoli; F. Messina; F. Pimpinelli; M. Rueca; M.R. Capobianchi |
| EPI_ISL_418230 | Clinique AVERAY LA BROUSTE, Med. Polyvalente | National Reference Center for Viruses of Respiratory Infections, Institut Pasteur, Paris | Angela Brisebarre; Elsa Ngwem; Etienne Simon-Lorière; Flora Donati; Marion Barbet; Maud Vanpeene; Mélanie Albert; Méline Bizard; Sylvie Behillil; Sylvie van der Werf; Vincent Enouf |
| EPI_ISL_862652 to 862653 | Clínica Universidad de Navarra | Instituto de Salud Carlos III | A. Monzón; F. Casas; I. Fernández, M.; I. Jiménez; Iglesias-Caballero; M. Pozo; M. Cuesta; M. González-Esguevillas; M. Zaballos; M.Camarero; P. Jiménez; S. Juliá; S. Molinero Calamita; S. Varona |
| EPI_ISL_452473 to 452474, EPI_ISL_452476 to 452515, EPI_ISL_452517 to 452543, EPI_ISL_538082, EPI_ISL_538084 to 538098, EPI_ISL_538100 to 538104, EPI_ISL_538107 to 538117, EPI_ISL_538608 to 538612 | | | |
| see above | Clínica Universidad de Navarra. Servicio de Enfermedades Infecciosas y Microbiología clínica | SeqCOVID-SPAIN consortium/IBV(CSIC) | Jose Luis del Pozo and SeqCOVID-SPAIN consortium; Mirian Fernández-Alonso |
| EPI_ISL_862544, EPI_ISL_862553 to 862555 | Compejo Hospitalario Xeral-Calde | Instituto de Salud Carlos III | A. Monzón; F. Casas; I. Alonso, P.; I. Jiménez; Iglesias-Caballero; M. Pozo; M. Cuesta; M. González-Esguevillas; M. Zaballos; M.Camarero; P. Jiménez; S. Juliá; S. Molinero Calamita; S. Varona |
| EPI_ISL_455328 to 455331, EPI_ISL_539563 to 539564, EPI_ISL_578194 | Complejo Hospitalario Universitario La Coruna | Instituto de Salud Carlos III | A. Monzón; F. Casas; I; I. Jiménez; Iglesias-Caballero; J. López; M. Camarero; M. Cuesta; M. González-Esguevillas; M. Molinero Calamita; M. Zaballos; M.A Canizares; M.I Paz; P. Jiménez; S. Juliá; S. Pozo; S. Varona |
| EPI_ISL_691684 | Complejo Hospitalario Universitario de Albacete | Instituto de Salud Carlos III | A. Monzón; F. Casas; I. Jiménez; I. Martínez, E.; Iglesias-Caballero; M. Camarero; M. Cuesta; M. González-Esguevillas; M. Pozo; M. Zaballos; P. Jiménez; S. Juliá; S. Molinero Calamita; S. Varona |
| EPI_ISL_474798, EPI_ISL_474800 to 474813, EPI_ISL_474815 to 474820, EPI_ISL_474822 to 474831, EPI_ISL_474837 to 474838, EPI_ISL_474840, EPI_ISL_474847 to 474849, EPI_ISL_474853, EPI_ISL_474901 to 474904, EPI_ISL_474919, EPI_ISL_474921, EPI_ISL_474933, EPI_ISL_474940 to 474941, EPI_ISL_474946, EPI_ISL_474951 to 474955, EPI_ISL_500157 to 500159, EPI_ISL_500166, EPI_ISL_500207, EPI_ISL_500218 | | | |
| see above | Complejo Hospitalario Universitario de Albacete | SeqCOVID-SPAIN consortium/IBV(CSIC) | Caridad Sainz de Baranda Camino and SeqCOVID-SPAIN consortium; Encarnacion Simarro Córdoba; Julia Lozano Serra; Lorena Robles Fonseca; Monica Parra Grandes |
| EPI_ISL_455333 | Complejo Hospitalario Universitario de Santiago | Instituto de Salud Carlos III | A. Monzón; F. Casas; I; I. Jiménez; Iglesias-Caballero; J. Llovo; M. Camarero; M. Cuesta; M. González-Esguevillas; M. Molinero Calamita; M. Zaballos; P. Jiménez; S. Juliá; S. Pozo; S. Varona |
| EPI_ISL_537380 to 537381 | Complejo Hospitalario Universitario de Vigo | SeqCOVID-SPAIN consortium/IBV(CSIC) | Benito Regueiro and SeqCOVID-SPAIN consortium |
| EPI_ISL_539569 to 539572, EPI_ISL_862550, EPI_ISL_862552, EPI_ISL_862570 to 862572, EPI_ISL_862645 to 862651 | Complejo Hospitalario de Navarra | Instituto de Salud Carlos III | A. Monzón; F. Casas; I; I. Ezpeleta, C.; I. Jiménez; Iglesias-Caballero; J. López; M. Pozo; M. Camarero; M. Cuesta; M. González-Esguevillas; M. Molinero Calamita; M. Zaballos; M.Camarero; P. Jiménez; S. Juliá; S. Molinero Calamita; S. Pozo; S. Varona |
| EPI_ISL_455334 to 455335, EPI_ISL_578195 | Complejo Hospitalario de Orense | Instituto de Salud Carlos III | A. Monzón; F. Casas; I; I. Jiménez; Iglesias-Caballero; M. Camarero; M. Cuesta; M. García; M. González-Esguevillas; M. Molinero Calamita; M. Paz; M. Zaballos; P. Jiménez; S. Juliá; S. Pozo; S. Varona |
| EPI_ISL_455327, EPI_ISL_539526 to 539529, EPI_ISL_691681 | Consejeria de Sanidad y Asuntos Sociales | Instituto de Salud Carlos III | A. Monzón; F. Casas; G. Gutiérrez; I; I. Gutiérrez, G.; I. Jiménez; Iglesias-Caballero; M. Camarero; M. Cuesta; M. González-Esguevillas; M. Molinero Calamita; M. Pozo; M. Zaballos; P. Jiménez; S. Juliá; S. Molinero Calamita; S. Pozo; S. Varona |
| EPI_ISL_862546 to 862547, EPI_ISL_862557 to 862563, EPI_ISL_862566, EPI_ISL_862641 to 862644 | Consejería de Sanidad y Asuntos Sociales | Instituto de Salud Carlos III | A. Monzón; F. Casas; I. Gutiérrez, G.; I. Jiménez; Iglesias-Caballero; M. Camarero; M. Cuesta; M. González-Esguevillas; M. Pozo; M. Zaballos; P. Jiménez; S. Juliá; S. Molinero Calamita; S. Varona |
| EPI_ISL_514751 | CoronaNet Lab- TaskForce Regione Campania, CEINGE Biotecnologie Avanzate, Via G. Salvatore | CoronaNet Lab- TaskForce Regione Campania, CEINGE Biotecnologie Avanzate, Via G. Salvatore | Asadzadeh, F.; Atripaldi, L.; Bianchi, M.; Boccia, A.; Borriello, G.; Brandi, S.; Castaldo, G.; Cerino, R.; Chiariotti, L.; Comegna, M.; Ferrucci, V.; Fusco, G.; H.Y.; J.H.; J.M.; Jung; K.S. and Kim; Kong, Dy.; Lee; Marrone, L.; Paolella, G.; Pascarella, S.; Siciliano, R.; Tiberio, C.; Viscardi, M.; Yun; Zollo, M. |
| EPI_ISL_454602 | Croatian Institute of Public Health | University of Zagreb, Centre for research and knowledge transfer in biotechnology | Anamarija Slovic; Irena Tabain; Jelena Ivancic Jelecki; Tatjana Vilibic-Cavlek |
| EPI_ISL_833050 to 833051, EPI_ISL_833057 to 833067 | DIP. PREV. AVEZZANO SERVIZIO DI IGIENE EPIDEMIOLOGIAE SANITA' PUBBLICA | Istituto Zooprofilattico Sperimentale dell'Abruzzo e Molise "G. Caporale" | Ancora M; Calistri P; Cammà C; Curini V; Delli Compagni E; Di Domenico M; Di Pasquale A; Lorusso A; Mangone I; Marcacci M; Puglia I; Rinaldi A; Savini G |
| EPI_ISL_849635 to 849636 | DIPARTIMENTO PREVENZIONE AVEZZANO-SERVIZIO DI IGIENE EPIDEMIOLOGIA E SANITA' PUBBLICA | Istitituto Zooprofilattico Sperimentale dell'Abruzzo e Molise "G.Caporale" | Ancora M; Cammà C; Curini V; Di Domenico M; Di Pasquale A; Lorusso A; Mangone I; Marcacci M; Puglia I; Rinaldi A; Savini G. |

| | | | |
|---|---|---|---|
| EPI_ISL_806758 to 806761, EPI_ISL_806763 to 806765, EPI_ISL_806781 to 806782, EPI_ISL_833283, EPI_ISL_833301 | DIPARTIMENTO PREVENZIONE AVEZZANO-SERVIZIO DI IGIENE EPIDEMIOLOGIA E SANITA' PUBBLICA | Istituto Zooprofilattico Sperimentale dell'Abruzzo e Molise "G. Caporale" | Ancora M; Calistri P; Cammà C; Curini V; Di Domenico M; Di Pasquale A; Lorusso A; Mangone I; Marcacci M; Puglia I; Rinaldi A; Savini G |
| EPI_ISL_529147 to 529148 | Democritus University of Thrace, Department of Medicine | Democritus University of Thrace, Department of Medicine | Bampali, M.; Dovrolis, N.; Froukala, E.; Gatzidou, E.; Karakasiliotis, I.; Kassela, K.; Spanakis, N.; Stavropoulou, A.; Tsakris, A.; Veletza, S. |
| EPI_ISL_508615 to 508631, EPI_ISL_508635 to 508643, EPI_ISL_508645 to 508650, EPI_ISL_508652 to 508657, EPI_ISL_508660 to 508661, EPI_ISL_508663 to 508665, EPI_ISL_508667 to 508670, EPI_ISL_508672 to 508673, EPI_ISL_508675 to 508676, EPI_ISL_508678 to 508683, EPI_ISL_508685 | | | |
| see above | Departamento de Microbiología, CDB, Hospital Clínic, Barcelona | SeqCOVID-SPAIN consortium/IBV(CSIC) | Aida Peiró and SeqCOVID-SPAIN consortium; Andrea Vergara; Elisa Rubio; Jéssica Navero; Mikel Martínez |
| EPI_ISL_516922 to 516933 | Department for Molecular Diagnostics, Centre for Medical Microbiology, Institute of Public Health of Montenegro | Charite Universitatsmedizin Berlin, Institut fur Virologie | Barbara Muehlemann; Christian Drosten; Julia Schneider; Jörn Beheim-Schwarzbach; Marija Govedarica and Danijela Vujoševi; Talitha Veith; Terry Jones; Victor M Corman |
| EPI_ISL_416997, EPI_ISL_417004 to 417006, EPI_ISL_417008 to 417009, EPI_ISL_417011 to 417023, EPI_ISL_417025, EPI_ISL_418623 to 418646, EPI_ISL_418648 to 418666, EPI_ISL_421182 to 421196, EPI_ISL_421198 to 421214, EPI_ISL_424628 to 424634, EPI_ISL_424636 to 424665, EPI_ISL_427340 to 427390, EPI_ISL_447120 to 447162, EPI_ISL_455958 to 455979, EPI_ISL_471427 to 471437, EPI_ISL_484693 to 484704, EPI_ISL_484706 to 484708, EPI_ISL_498127 to 498151, EPI_ISL_498628 to 498629, EPI_ISL_515055 to 515081, EPI_ISL_540469 to 540518, EPI_ISL_540520 to 540556, EPI_ISL_540558 to 540578, EPI_ISL_581575 to 581582, EPI_ISL_581585 to 581667, EPI_ISL_582027, EPI_ISL_626231 to 626240, EPI_ISL_626242 to 626339, EPI_ISL_636605 to 636685, EPI_ISL_641557 to 641606, EPI_ISL_644611 to 644626, EPI_ISL_661201 to 661251, EPI_ISL_666871 to 666889, EPI_ISL_678399 to 678409, EPI_ISL_678411 to 678427, EPI_ISL_678429 to 678485, EPI_ISL_681264 to 681270, EPI_ISL_707715 to 707721, EPI_ISL_707723 to 707770, EPI_ISL_708454 to 708459, EPI_ISL_722926 to 722966, EPI_ISL_737310 to 737338, EPI_ISL_737340, EPI_ISL_737342 to 737375, EPI_ISL_737377 to 737387, EPI_ISL_737390 to 737394, EPI_ISL_824790 to 824831, EPI_ISL_824833 to 824834, EPI_ISL_832207, EPI_ISL_833186 to 833187, EPI_ISL_872040 to 872041, EPI_ISL_872058 to 872060, EPI_ISL_872062 to 872064, EPI_ISL_872109, EPI_ISL_872153 to 872155 | | | |
| see above | Department of Clinical Microbiology | GIGA Medical Genomics | Artesi Maria; Axelle Chaslain; Bontems Sébastien; Boreux Raphaël; Bouchra Boujemla; Bours Vincent.; Cecile Meex; Celine Fombellida-Lopez; Cécile Meex; Céline Fombellida-Lopez; Durkin Keith; Gilles Darcis; Hayette Marie-Pierre; Justine Defêche; Keith Durkin; Maria Artesi; Marie-Pierre Hayette; Meex Cécile; Melin Pierrette; Michel Moutschen; Pierrette Melin; Raphael Boreux; Raphaël Boreux; Sebastien Bontems; Sébastien Bontems; Vincent Bours; Vincent Bours. |
| EPI_ISL_429262 to 429289, EPI_ISL_429291, EPI_ISL_429293 to 429318, EPI_ISL_429320 to 429331, EPI_ISL_451988 to 452001, EPI_ISL_452003 to 452004, EPI_ISL_452006 to 452007, EPI_ISL_452009 to 452010, EPI_ISL_452012 to 452016, EPI_ISL_452018 to 452060, EPI_ISL_452063 to 452071, EPI_ISL_452073, EPI_ISL_452075 to 452100 | | | |
| see above | Department of Clinical Microbiology, Copenhagen University Hospital, Hvidovre, Kettegaard Alle 30, 2650 Hvidovre. | Albertsen lab, Department of Chemistry and Bioscience, Aalborg University, Denmark | Rasmus Kirkegaard |
| EPI_ISL_507206 to 507215, EPI_ISL_605793 to 605796 | Department of Experimental Modeling and Pathogenesis of Infectious Diseases | WHO National Influenza Centre Russian Federation | Alekseev A.Yu.; Andrey Komissarov; Anna Ivanova; Artem Fadeev; Chepurnov A.A.; Daria Danilenko; Kononova Yu.V.; Kseniya Komissarova; Mariia Sergeeva; Shestopalov A.M.; Sobolev I.A. |
| EPI_ISL_487276 | Department of Food Safety, Nutrition and Veterinary public health, Istituto Superiore di Sanita' | Department of Biomedical, Surgical and Dental Sciences and Department of Biomedical Sciences for Health | Anselmi, G.; Basilico, N.; Binda, S.; D'Alessandro, S.; Delbue, S.; Ferrante, P.; Galli, C.; Parapini, S.; Pariani, E.; Primache, V.; Signorini, L. |
| EPI_ISL_522407 to 522408 | Department of Infection Prevention and Infectious Diseases, University Hospital Regensburg | Department of Infection Prevention and Infectious Diseases, University Hospital Regensburg | Fritsch, J.; Holzmann, T.; Schneider-Brachert, W. |
| EPI_ISL_513298 to 513307 | Department of Infection Prevention and Infectious Diseases, University Hospital Regensburg | University Hospital Regensburg | Fritsch, J.; Holzmann, T.; Schneider-Brachert, W. |
| EPI_ISL_524474, EPI_ISL_524480 to 524481 | Department of Infectious Diseases, Cantonal Hospital Baden | Institute of Medical Virology, University of Zurich | Alexandra Trkola; Andrea Zbinden; Fiona Steiner; Gabriela Ziltener; Jon Huder; Jürg Böni; Maryam Zaheri; Michael Huber; Patrick Redli; Riccarda Capaul; Stefan Schmutz; Verena Kufner |
| EPI_ISL_457699 to 457700, EPI_ISL_457721, EPI_ISL_457724, EPI_ISL_457728, EPI_ISL_457732, EPI_ISL_457736, EPI_ISL_457749 | | | |
| see above | Department of Infectious Diseases, Istituto Superiore di Sanità, Roma , Italy | Army Medical and Veterinary Research Center | Alessandra Lo Presti; Anna Anselmo; Antonella Fortunato; Antonella Marchi; Concetta Fabiani Silvia Fillo; Concetta Fabiani Silvia Fillo; Eleonora Benedetti; Florigio Lista; Francesco Giordani; Giovanni Faggioni; Nino D'Amore; Paola Stefanelli; Riccardo De Sanctis; Stefano Fiore; Vanessa Vera Fain |
| EPI_ISL_412973 | Department of Infectious Diseases, Istituto Superiore di Sanità, Roma , Italy | Virology Laboratory, Scientific Department, Army Medical Center | Andrea Ciammaruconi; Anna Anselmo; Antonella Fortunato; Antonella Marchi; Concetta Fabiani; Eleonora Benedetti; Florigio Lista; Giovanni Faggioni; Paola Stefanelli; Riccardo De Santis; Silvia Fillo; Stefano Fiore; Stefano Palomba |
| EPI_ISL_856889, EPI_ISL_856904 | Department of Infectious Diseases, Istituto Superiore di Sanità, Roma, Italy | Virology Laboratory, Scientific Department, Army Medical Center | Alessandra Lo Presti; Angela Di Martino; Anna Anselmo; Antonella Fortunato; Florigio Lista; Francesco Giordani; Giovanni Faggioni; Nino D'Amore; Paola Stefanelli; Riccardo De Sanctis; Silvia Fillo; Stefano Fiore; Vanessa Vera Fain |
| EPI_ISL_856888 | Department of Infectious Diseases, Istituto Superiore di Sanità, Roma, Italy; A.O.R. San Carlo di Potenza, Laboratorio Microbiologia e Virologia, Potenza, Italy | Virology Laboratory, Scientific Department, Army Medical Center | Alessandra Lo Presti; Angela Di Martino; Anna Anselmo; Antonella Fortunato; Antonio Picerno; Florigio Lista; Francesco Giordani; Giovanni Faggioni; Nino D'Amore; Paola Stefanelli; Riccardo De Sanctis; Silvia Fillo; Stefano Fiore; Vanessa Vera Fain |
| EPI_ISL_856890 | Department of Infectious Diseases, Istituto Superiore di Sanità, Roma, Italy; AO Annunziata, UOC Microbiologia e Virologia, Cosenza, Italy | Virology Laboratory, Scientific Department, Army Medical Center | Alessandra Lo Presti; Angela Di Martino; Anna Anselmo; Antonella Fortunato; Cristina Giraldi; Florigio Lista; Francesco Giordani; Giovanni Faggioni; Nino D'Amore; Paola Stefanelli; Riccardo De Sanctis; Silvia Fillo; Stefano Fiore; Vanessa Vera Fain |
| EPI_ISL_856905 to 856906 | Department of Infectious Diseases, Istituto Superiore di Sanità, Roma, Italy; AO S.M. della Misericordia, S.C. Microbiologia, Perugia, Italy | Virology Laboratory, Scientific Department, Army Medical Center | Alessandra Lo Presti; Angela Di Martino; Anna Anselmo; Antonella Fortunato; Barbara Camilloni; Florigio Lista; Francesco Giordani; Giovanni Faggioni; Nino D'Amore; Paola Stefanelli; Riccardo De Sanctis; Silvia Fillo; Stefano Fiore; Vanessa Vera Fain |
| EPI_ISL_856871, EPI_ISL_856891 to 856892 | Department of Infectious Diseases, Istituto Superiore di Sanità, Roma, Italy; AORN dei Colli, UOC Microbiologia Virologia, Napoli, Italy | Virology Laboratory, Scientific Department, Army Medical Center | Alessandra Lo Presti; Angela Di Martino; Anna Anselmo; Antonella Fortunato; Florigio Lista; Francesco Giordani; Giovanni Faggioni; Luigi Atripaldi; Nino D'Amore; Paola Stefanelli; Riccardo De Sanctis; Silvia Fillo; Stefano Fiore; Vanessa Vera Fain |
| EPI_ISL_856900 to 856901 | Department of Infectious Diseases, Istituto Superiore di Sanità, Roma, Italy; AOUP V. Emanuele di Catania, P.O. Gaspare Rodolico, Catania, Italy | Virology Laboratory, Scientific Department, Army Medical Center | Alessandra Lo Presti; Angela Di Martino; Anna Anselmo; Antonella Fortunato; Florigio Lista; Francesco Giordani; Giovanni Faggioni; Guido Scalia; Nino D'Amore; Paola Stefanelli; Riccardo De Sanctis; Silvia Fillo; Stefano Fiore; Vanessa Vera Fain |
| EPI_ISL_856902 | Department of Infectious Diseases, Istituto Superiore di Sanità, Roma, Italy; AS Alto Adige, Microbiologia e Virologia, Bolzano, Italy | Virology Laboratory, Scientific Department, Army Medical Center | Alessandra Lo Presti; Angela Di Martino; Anna Anselmo; Antonella Fortunato; Elisabetta Pagani; Florigio Lista; Francesco Giordani; Giovanni Faggioni; Nino D'Amore; Paola Stefanelli; Riccardo De Sanctis; Silvia Fillo; Stefano Fiore; Vanessa Vera Fain |
| EPI_ISL_856898 to 856899 | Department of Infectious Diseases, Istituto Superiore di Sanità, Roma, Italy; ASL Città di Torino, S.C. Microbiologia e Virologia, Torino, Italy | Virology Laboratory, Scientific Department, Army Medical Center | Alessandra Lo Presti; Angela Di Martino; Anna Anselmo; Antonella Fortunato; Florigio Lista; Francesco Giordani; Giovanni Faggioni; Nino D'Amore; Paola Stefanelli; Riccardo De Sanctis; Silvia Fillo; Stefano Fiore; Valeria Ghisetti; Vanessa Vera Fain |
| EPI_ISL_856893 to 856894 | Department of Infectious Diseases, Istituto Superiore di Sanità, Roma, Italy; ASUITS - Ospedale Maggiore, S.C. UOC Igiene e Sanità Pubblica, Trieste, Italy | Virology Laboratory, Scientific Department, Army Medical Center | Alessandra Lo Presti; Angela Di Martino; Anna Anselmo; Antonella Fortunato; Florigio Lista; Francesco Giordani; Giovanni Faggioni; Nino D'Amore; Paola Stefanelli; Pierlanfranco D'Agaro; Riccardo De Sanctis; Silvia Fillo; Stefano Fiore; Vanessa Vera Fain |
| EPI_ISL_856887 | Department of Infectious Diseases, Istituto Superiore di Sanità, Roma, Italy; AUOC Policlinico Bari, UOC Igiene, Bari, Italy | Virology Laboratory, Scientific Department, Army Medical Center | Alessandra Lo Presti; Angela Di Martino; Anna Anselmo; Antonella Fortunato; Florigio Lista; Francesco Giordani; Giovanni Faggioni; Maria Chironna; Nino D'Amore; Paola Stefanelli; Riccardo De Sanctis; Silvia Fillo; Stefano Fiore; Vanessa Vera Fain |
| EPI_ISL_856907 | Department of Infectious Diseases, Istituto Superiore di Sanità, Roma, Italy; AUSL Valle d'Aosta, Aosta, Italy | Virology Laboratory, Scientific Department, Army Medical Center | Alessandra Lo Presti; Angela Di Martino; Anna Anselmo; Antonella Fortunato; Florigio Lista; Francesco Giordani; Giovanni Faggioni; Massimo Di Benedetto; Nino D'Amore; Paola Stefanelli; Riccardo De Sanctis; Silvia Fillo; Stefano Fiore; Vanessa Vera Fain |
| EPI_ISL_856880, EPI_ISL_856895 to 856896 | Department of Infectious Diseases, Istituto Superiore di Sanità, Roma, Italy; Ospedali Riuniti, Laboratorio Virologia, Ancona, Italy | Virology Laboratory, Scientific Department, Army Medical Center | Alessandra Lo Presti; Angela Di Martino; Anna Anselmo; Antonella Fortunato; Florigio Lista; Francesco Giordani; Giovanni Faggioni; Nino D'Amore; Paola Stefanelli; Patrizia Bagnarelli; Riccardo De Sanctis; Silvia Fillo; Stefano Fiore; Vanessa Vera Fain |
| EPI_ISL_856881, EPI_ISL_856897 | Department of Infectious Diseases, Istituto Superiore di Sanità, Roma, Italy; PO Cardarelli, Laboratorio Analisi Microbiologia e Virologia, Campobasso, Italy | Virology Laboratory, Scientific Department, Army Medical Center | Alessandra Lo Presti; Angela Di Martino; Anna Anselmo; Antonella Fortunato; Florigio Lista; Francesco Giordani; Giovanni Faggioni; Massimiliano Scutellà; Nino D'Amore; Paola Stefanelli; Riccardo De Sanctis; Silvia Fillo; Stefano Fiore; Vanessa Vera Fain |

| | | | |
|---|---|---|---|
| EPI_ISL_856908 | Department of Infectious Diseases, Istituto Superiore di Sanità, Roma, Italy; PO Madre Teresa di Calcutta, UOC. Laboratorio Analisi ULSS 6 Euganea, Padova, Italy | Virology Laboratory, Scientific Department, Army Medical Center | Alessandra Lo Presti; Angela Di Martino; Anna Anselmo; Antonella Fortunato; Florigio Lista; Francesco Giordani; Giacomo Mezzapelle; Giovanni Faggioni; Nino D'Amore; Paola Stefanelli; Riccardo De Sanctis; Silvia Fillo; Stefano Fiore; Vanessa Vera Fain |
| EPI_ISL_856884, EPI_ISL_856903 | Department of Infectious Diseases, Istituto Superiore di Sanità, Roma, Italy; PO Santa Chiara, Microbiologia e Virologia, Trento, Italy | Virology Laboratory, Scientific Department, Army Medical Center | Alessandra Lo Presti; Angela Di Martino; Anna Anselmo; Antonella Fortunato; Florigio Lista; Francesco Giordani; Giovanni Faggioni; Nino D'Amore; Paola Stefanelli; Paolo Lanzafame; Riccardo De Sanctis; Silvia Fillo; Stefano Fiore; Vanessa Vera Fain |
| EPI_ISL_856886 | Department of Infectious Diseases, Istituto Superiore di Sanità, Roma, Italy; PO Spirito Santo, UOC Microbiologia e Virologia Clinica, Pescara, Italy | Virology Laboratory, Scientific Department, Army Medical Center | Alessandra Lo Presti; Angela Di Martino; Anna Anselmo; Antonella Fortunato; Florigio Lista; Francesco Giordani; Giovanni Faggioni; Nino D'Amore; Paola Stefanelli; Paolo Fazii; Riccardo De Sanctis; Silvia Fillo; Stefano Fiore; Vanessa Vera Fain |
| EPI_ISL_856909 | Department of Infectious Diseases, Istituto Superiore di Sanità, Roma, Italy; ULSS 8 Berica Vicenza, UOC Microbiologia, Vicenza, Italy | Virology Laboratory, Scientific Department, Army Medical Center | Alessandra Lo Presti; Angela Di Martino; Anna Anselmo; Antonella Fortunato; Florigio Lista; Francesco Giordani; Giovanni Faggioni; Mario Rassu; Nino D'Amore; Paola Stefanelli; Riccardo De Sanctis; Silvia Fillo; Stefano Fiore; Vanessa Vera Fain |
| EPI_ISL_855550, EPI_ISL_855553 | Department of Infectious Diseases, Istituto Superiore di Sanità, Rome, Italy | Istituto Superiore di Sanità (ISS) | Alessandra Lo Presti; Angela Di Martino; Gabriele Vaccari; Giovanni Ianiro; Ilaria Di Bartolo; Luca De Sabato; Manuela Marra; Marco Crescenzi; Maria Carollo; Paola Stefanelli; Stefano Fiore |
| EPI_ISL_412974 | Department of Infectious Diseases, Istituto Superiore di Sanità, Rome, Italy | Virology Laboratory, Scientific Department, Army Medical Center | Andrea Ciammaruconi; Antonella Fortunato; Antonella Marchi; Concetta Fabiani; Eleonora Benedetti; Filippo Molinari; Florigio Lista; Giancarlo Petralito; Giovanni Faggioni; Paola Stefanelli; Riccardo De Santis; Silvia Fillo; Stefano Fiore |
| EPI_ISL_910331 to 910332 | Department of Infectious Diseases, Istituto Superiore di Sanità, Rome, Italy; AOUP Paolo Giaccone, Palermo, Italy | Istituto Superiore di Sanità (ISS) | Alessandra Lo Presti; Angela Di Martino; Carmelo Maida; Daniela Di Naro; Fabio Tramuto; Francesco Vitale; Giulia Randazzo; Manuela Marra; Marco Crescenzi; Maria Carollo; Paola Stefanelli; Stefano Fiore |
| EPI_ISL_855554 to 855555 | Department of Infectious Diseases, Istituto Superiore di Sanità, Rome, Italy; ASST G.O.M. Niguarda, Sezione Microbiologia e Virologia, Milano, Italy | Istituto Superiore di Sanità (ISS) | Alessandra Lo Presti; Angela Di Martino; Carlo Federico Perno; Gabriele Vaccari; Giovanni Ianiro; Ilaria Di Bartolo; Luca De Sabato; Manuela Marra; Marco Crescenzi; Maria Carollo; Paola Stefanelli; Stefano Fiore |
| EPI_ISL_855552 | Department of Infectious Diseases, Istituto Superiore di Sanità, Rome, Italy; Ospedali Riuniti, Laboratorio Virologia, Ancona, Italy | Istituto Superiore di Sanità (ISS) | Alessandra Lo Presti; Angela Di Martino; Gabriele Vaccari; Giovanni Ianiro; Ilaria Di Bartolo; Luca De Sabato; Manuela Marra; Marco Crescenzi; Maria Carollo; Paola Stefanelli; Patrizia Bagnarelli; Stefano Fiore |
| EPI_ISL_875687 | Department of Infectious Diseases, Istituto Superiore di Sanità, Rome, Italy; UCO Igiene e Sanità Pubblica, ASUGI, Trieste, Italy; Laboratorio di Genomica ed Epigenomica sistema Argo, Area SciencePark, Trieste, Italy; Laboratorio di Virologia Molecolare, ICGEB, Trieste, Italy | Istituto Superiore di Sanità (ISS) | Alessandra Lo Presti; Alessandro Marcello; Angela Di Martino; Danilo Licastro; Ludovica Segat; Manuela Marra; Marco Crescenzi; Maria Carollo; Paola Stefanelli; Pierlanfranco D'Agaro; Stefano Fiore |
| EPI_ISL_411218 to 411220 | Department of Infectious and Tropical Diseases, Bichat Claude Bernard Hospital, Paris | Laboratoire Virpath, CIRI U111, UCBL1, INSERM, CNRS, ENS Lyon | Alexandre Gaymard; Aurélien Traversier; Bruno Lina; Catherine Legras-Lachuer; Julien Fouret; Manuel Rosa-Calatrava; Olivier Terrier; Xavier Lescure; Yazdan Yazdanpanah |
| EPI_ISL_410720, EPI_ISL_410984 | Department of Infectious and Tropical Diseases, Bichat Claude Bernard Hospital, Paris | National Reference Center for Viruses of Respiratory Infections, Institut Pasteur, Paris | Angela Brisebarre; Flora Donati; Marion Barbet; Maud Vanpeene; Mélanie Albert; Méline Bizard; Sylvie Behillil; Sylvie van der Werf; Vincent Enouf; Xavier Lescure; Xavier Lescure.; Yazdan Yazdanpanah |
| EPI_ISL_413019 to 413020 | Department of Internal Medicine, Triemli Hospital | Institute of Medical Virology, University of Zurich | Alexandra Trkola; Andrea Zbinden; Fiona Steiner; Gerhard Eich; Jon Huder; Jürg Böni; Maryam Zaheri; Michael Huber; Patrick Redli; Riccarda Capaul; Stefan Schmutz; Verena Kufner |
| EPI_ISL_413592, EPI_ISL_422408 to 422409, EPI_ISL_422412 to 422414, EPI_ISL_422416, EPI_ISL_422418, EPI_ISL_422420 to 422422, EPI_ISL_447619, EPI_ISL_693305, EPI_ISL_872597 | | | |
| see above | Department of Laboratory Medicine, National Taiwan University Hospital | Microbial Genomics Core Lab, National Taiwan University Centers of Genomic and Precision Medicine | Chiao-Ling Li; Pei-Jer Chen; Shan-Chwen Chang; Shiou-Hwei Yeh; Sui-Yuan Chang; Ya-Yun Lai; You-Yu Lin |
| EPI_ISL_803880 to 803888, EPI_ISL_803891 to 803893, EPI_ISL_803895 | Department of Medical Biotechnologies, University of Siena | Laboratory of Infectious Diseases, Department of Biomedical and Clinical Sciences L. Sacco, University of Milan | Alessia Lai; Annalisa Bergna; Carla Della Ventura; Claudia Balotta; Filippo Dragoni; Gianguglielmo Zehender on behalf of SARS-CoV-2 ITALIAN RESEARCH ENTERPRISE-(SCIRE) Collaborative Group; Ilaria Vicenti; Maria Grazia Cusi; Massimo Galli; Maurizio Zazzi |
| EPI_ISL_591012, EPI_ISL_591018 to 591019, EPI_ISL_775428 to 775437, EPI_ISL_775467, EPI_ISL_775523, EPI_ISL_796653, EPI_ISL_796681 to 796683 | | | |
| see above | Department of Medical Microbiology - section Molde, Molde Hospital | Norwegian Institute of Public Health, Department of Virology | Atiya R Ali; Hilde Elshaug; Hilde Vollan; Kamilla Heddeland Instefjord; Karoline Bragstad; Kathrine Stene-Johansen; Marie Paulsen Madsen; Olav Hungnes; Rasmus Riis Kopperud |
| EPI_ISL_454416 | Department of Medical Microbiology, Leiden University Medical Center | Department of Medical Microbiology, Leiden University Medical Center | Dalebout; E.J.; J.C.; J.J. and Sidorov, I.; N.S.; Ogando; Snijder; T.J.; Zevenhoven; de Vries |
| EPI_ISL_591007, EPI_ISL_591014, EPI_ISL_635096, EPI_ISL_635100, EPI_ISL_635110 to 635111, EPI_ISL_635126, EPI_ISL_635135 to 635136, EPI_ISL_635165, EPI_ISL_668404, EPI_ISL_668417 to 668418, EPI_ISL_668420 to 668421, EPI_ISL_668441, EPI_ISL_708042 to 708043, EPI_ISL_708106 to 708109, EPI_ISL_708142 to 708146, EPI_ISL_759969 to 759972, EPI_ISL_775306, EPI_ISL_775464 to 775466, EPI_ISL_775475, EPI_ISL_775481, EPI_ISL_775491 to 775494, EPI_ISL_775505 to 775508, EPI_ISL_796687, EPI_ISL_796706, EPI_ISL_796711, EPI_ISL_796717 to 796721, EPI_ISL_796725, EPI_ISL_796727 to 796728, EPI_ISL_842643 | | | |
| see above | Department of Medical Microbiology, St. Olavs hospital | Norwegian Institute of Public Health, Department of Virology | Atiya R Ali; Hilde Elshaug; Hilde Vollan; Kamilla Heddeland Instefjord; Karoline Bragstad; Kathrine Stene-Johansen; Marie Paulsen Madsen; Olav Hungnes; Rasmus Riis Kopperud |
| EPI_ISL_488840, EPI_ISL_488845, EPI_ISL_488847, EPI_ISL_488853 to 488856, EPI_ISL_488861, EPI_ISL_488863 to 488864, EPI_ISL_488872, EPI_ISL_492836, EPI_ISL_492838 to 492839, EPI_ISL_492841 to 492842, EPI_ISL_492848 to 492849, EPI_ISL_492853 to 492855, EPI_ISL_492857 to 492861, EPI_ISL_492867, EPI_ISL_492870 to 492871, EPI_ISL_492873, EPI_ISL_492875, EPI_ISL_492878, EPI_ISL_492881, EPI_ISL_492884, EPI_ISL_492886, EPI_ISL_492889, EPI_ISL_492891 to 492893, EPI_ISL_492895 to 492900, EPI_ISL_492903 to 492904, EPI_ISL_492907, EPI_ISL_492909 to 492910, EPI_ISL_492912 to 492913, EPI_ISL_492915, EPI_ISL_495081 to 495096, EPI_ISL_501557, EPI_ISL_501562, EPI_ISL_501602 | | | |
| see above | Department of Medical Microbiology, Western Sussex Hospitals NHS Foundation Trust, St Richard's Hospital | Wellcome Sanger Institute for the COVID-19 Genomics UK (COG-UK) consortium | Cordelia Langford; David K. Jackson; Dominic Kwiatkowski; Ewan Harrison; Ian Johnston; John Sillitoe on behalf of the Wellcome Sanger Institute COVID-19 Surveillance Team (http://www.sanger.ac.uk/covid-team); Jonathan Lewis; Manasa Mutingwende; Michelle Erkiert; Olga Podplomyk; Paul Randell and Alex Alderton; Roberto Amato; Sarah Lowdon; Sonia Goncalves |
| EPI_ISL_454733 | Department of Medical, Biotechnologies University of Siena | Department of Medical, Biotechnologies University of Siena | Anichini, G.; Cusi; G. and Santoro, F.; Gandolfo, C.; M.G.; Pinzauti, D.; Pozzi |
| EPI_ISL_431102 | Department of MicroBiology,Gandhi Medical College and Hospital,Secendrabad,Hyderabad,India | Department of Microbiology, Gandhi Medical College and Hospital, Secendrabad, Hyderabad | Amit A. Upadhyay; Anand Kumar K; Kalyani Putty; Muttineni Radhakrishna; Nagamani K; Pankaj Singh D; Raja Rao M; Rama Amara; Ravikumar P; Steven E. Bosinger; Sunitha P; Thrilok Chander B |
| EPI_ISL_431117 | Department of Microbiology, Gandhi Medical College and Hospital, Secendrabad, Hyderabad, India | Department of Microbiology, Gandhi Medical College and Hospital, Secendrabad, Hyderabad, India | Amit A. Upadhyay; Anand Kumar K; Kalyani Putty; Muttineni Radhakrishna; Nagamani K; Pankaj Singh D; Raja Rao M; Rama Amara; Ravikumar P; Steven E.Bosinger; Sunitha P; Thrilok Chander B |
| EPI_ISL_516799 | Department of Microbiology, The University of Hong Kong | Department of Microbiology, The University of Hong Kong | Kelvin K.W. To; Kwok-Yung Yuen |
| EPI_ISL_812968 | Department of Molecular Medicine, University of Padova | Department of Molecular Medicine, University of Padova | Abate, D.; Barzon, L.; Besutti, V.; De Canale, E.; Del Vecchio, C.; Franchin, E.; Lavezzo, E.; Loregian, A.; M.C.; Manganelli, R.; Manuto, L.; Masi, G.; Onelia, F.; Pacenti, M.; Parisi; Rossi, L.; S. and Crisanti, A.; S.G.; Saluzzo, F.; Sciro, M.; Toppo; Trevisan, M.; Vanuzzo |
| EPI_ISL_463741 to 463748 | Department of Molecular Virology, Cyprus Institute of Neurology and Genetics | Department of Molecular Virology, Cyprus Institute of Neurology and Genetics | Christina Christodoulou; Christina Tryfonos; Dana Koptides; George Krashias; Jan Richter; Stavros Bashiardes |
| EPI_ISL_853819 to 853821, EPI_ISL_854239 | Department of Pathology, Landeskrankenhaus Graz II, Medical University Graz | Bergthaler laboratory, CeMM Research Center for Molecular Medicine of the Austrian Academy of Sciences | Alexander Lercher; Alexandra Popa; Andreas Bergthaler; Anna Schedl; Benedikt Agerer; Christoph Bock; Jakob-Wendelin Genger; Jan Laine; Lukas Endler; Martin Senekowitsch; Michael Schuster; Thomas Penz |
| EPI_ISL_425231 to 425257, EPI_ISL_425260, EPI_ISL_425262 to 425273, EPI_ISL_425275 to 425279, EPI_ISL_425281 to 425301, EPI_ISL_425303 to 425305, EPI_ISL_425307 to 425333, EPI_ISL_425335 to 425345, EPI_ISL_425347 to 425348, EPI_ISL_425351 to 425352, EPI_ISL_425354, EPI_ISL_425358 to 425371, EPI_ISL_425373 to 425385, EPI_ISL_425387 to 425404, EPI_ISL_425406 to 425409, EPI_ISL_425411 to 425418, EPI_ISL_425420 to 425421, EPI_ISL_425423 to 425438, EPI_ISL_425441 to 425460, EPI_ISL_433466 to 433487, EPI_ISL_433490 to 433497, EPI_ISL_433666 to 433686, EPI_ISL_433688 to 433707, EPI_ISL_433709 to 433715, EPI_ISL_433717 to 433739, EPI_ISL_433741 to 433746, EPI_ISL_433748 to 433765, EPI_ISL_433767 to 433776, EPI_ISL_433778 to 433806, EPI_ISL_433808 to 433837, EPI_ISL_433839 to 433971, EPI_ISL_433974 to 433983, EPI_ISL_433986 to 433997, EPI_ISL_433999 to 434006, EPI_ISL_434008 to 434021, EPI_ISL_434023 to 434041, EPI_ISL_434043 to 434058, EPI_ISL_434060 to 434062, EPI_ISL_438550 to 438571, EPI_ISL_438573 to 438610, EPI_ISL_438612 to 438654, EPI_ISL_438656 to 438683, EPI_ISL_438685 to 438687, EPI_ISL_438689 to 438693, EPI_ISL_438695 to 438730, EPI_ISL_438732 to 438748, EPI_ISL_444313 to 444342, EPI_ISL_444344, EPI_ISL_444346 to 444348, EPI_ISL_444350 to 444354, EPI_ISL_444356 to 444374, EPI_ISL_444377 to 444378, EPI_ISL_444380 to 444382, EPI_ISL_444384 to 444408, EPI_ISL_444410 to 444433, EPI_ISL_444435 to 444443, EPI_ISL_444446 to | | | |

444453, EPI_ISL_447945, EPI_ISL_447947 to 447981, EPI_ISL_447983, EPI_ISL_447987 to 447990, EPI_ISL_447993 to 448008, EPI_ISL_448010 to 448013, EPI_ISL_448015 to 448024, EPI_ISL_448026 to 448029, EPI_ISL_448032 to 448037, EPI_ISL_448039 to 448071, EPI_ISL_448074 to 448084, EPI_ISL_448086 to 448090, EPI_ISL_448092, EPI_ISL_448096 to 448103, EPI_ISL_448106 to 448115, EPI_ISL_452857 to 452874, EPI_ISL_452876 to 452901, EPI_ISL_452903 to 452906, EPI_ISL_452910 to 452930, EPI_ISL_452932, EPI_ISL_452934 to 452940, EPI_ISL_452942 to 452949, EPI_ISL_452951 to 452954, EPI_ISL_452957 to 452986, EPI_ISL_452988 to 453002, EPI_ISL_453004 to 453005, EPI_ISL_456677 to 456682, EPI_ISL_456686, EPI_ISL_456688, EPI_ISL_456692, EPI_ISL_456695 to 456706, EPI_ISL_456708 to 456725, EPI_ISL_456727 to 456733, EPI_ISL_456735 to 456737, EPI_ISL_456739 to 456743, EPI_ISL_456745, EPI_ISL_456747 to 456755, EPI_ISL_461546 to 461563, EPI_ISL_461565, EPI_ISL_461567 to 461568, EPI_ISL_461570 to 461585, EPI_ISL_461587 to 461588, EPI_ISL_473451 to 473493, EPI_ISL_473495 to 473503, EPI_ISL_473505 to 473507, EPI_ISL_477783 to 477785, EPI_ISL_477787 to 477816, EPI_ISL_477818 to 477819, EPI_ISL_483913 to 483915, EPI_ISL_490317 to 490321, EPI_ISL_490323 to 490328, EPI_ISL_499853, EPI_ISL_499855, EPI_ISL_499905, EPI_ISL_499908, EPI_ISL_499966, EPI_ISL_499968 to 499973, EPI_ISL_512330 to 512335, EPI_ISL_514443 to 514451, EPI_ISL_516990 to 517000, EPI_ISL_529224 to 529225, EPI_ISL_529236, EPI_ISL_529257, EPI_ISL_529265, EPI_ISL_529269, EPI_ISL_529283, EPI_ISL_529299, EPI_ISL_529445, EPI_ISL_529656 to 529660, EPI_ISL_549374, EPI_ISL_549386, EPI_ISL_549388 to 549394, EPI_ISL_572471 to 572472, EPI_ISL_572484, EPI_ISL_572519, EPI_ISL_572524, EPI_ISL_572541, EPI_ISL_572592, EPI_ISL_572616, EPI_ISL_572642, EPI_ISL_572779, EPI_ISL_572792, EPI_ISL_572851 to 572852, EPI_ISL_572862 to 572868, EPI_ISL_573138 to 573151, EPI_ISL_573153, EPI_ISL_576897 to 576902, EPI_ISL_576906 to 576919, EPI_ISL_584283, EPI_ISL_584286 to 584291, EPI_ISL_584294 to 584302, EPI_ISL_584304 to 584307, EPI_ISL_584309 to 584314, EPI_ISL_584316 to 584318, EPI_ISL_584321, EPI_ISL_611593, EPI_ISL_611751, EPI_ISL_611938, EPI_ISL_611964 to 611967, EPI_ISL_612007, EPI_ISL_612034, EPI_ISL_612099, EPI_ISL_612106, EPI_ISL_612123, EPI_ISL_612186, EPI_ISL_612262 to 612283, EPI_ISL_637261, EPI_ISL_637267 to 637272, EPI_ISL_637275 to 637277, EPI_ISL_637281 to 637282, EPI_ISL_637287, EPI_ISL_637292, EPI_ISL_637294, EPI_ISL_637299, EPI_ISL_637308, EPI_ISL_637310, EPI_ISL_637314 to 637318, EPI_ISL_637324, EPI_ISL_637328, EPI_ISL_637331, EPI_ISL_637334 to 637335, EPI_ISL_637340 to 637341, EPI_ISL_637345, EPI_ISL_637348, EPI_ISL_637353 to 637355, EPI_ISL_637357, EPI_ISL_637360, EPI_ISL_637362 to 637368, EPI_ISL_637384, EPI_ISL_637386, EPI_ISL_637388 to 637389, EPI_ISL_637392, EPI_ISL_637394, EPI_ISL_637397 to 637398, EPI_ISL_637403 to 637408, EPI_ISL_637410, EPI_ISL_637413 to 637418, EPI_ISL_637420 to 637421, EPI_ISL_637424 to 637426, EPI_ISL_637428 to 637431, EPI_ISL_637433, EPI_ISL_637435, EPI_ISL_637440, EPI_ISL_637451, EPI_ISL_637456, EPI_ISL_637460, EPI_ISL_637462, EPI_ISL_637464 to 637465, EPI_ISL_637468 to 637490, EPI_ISL_637500 to 637501, EPI_ISL_637503 to 637519, EPI_ISL_637526 to 637527, EPI_ISL_637529 to 637530, EPI_ISL_637545, EPI_ISL_637547, EPI_ISL_637549, EPI_ISL_637552, EPI_ISL_637554, EPI_ISL_637560, EPI_ISL_637562 to 637579, EPI_ISL_637583, EPI_ISL_637590 to 637592, EPI_ISL_637595, EPI_ISL_637598, EPI_ISL_637613, EPI_ISL_637620, EPI_ISL_637623 to 637624, EPI_ISL_637628 to 637638, EPI_ISL_637640 to 637642, EPI_ISL_637644 to 637648, EPI_ISL_637650 to 637654, EPI_ISL_637656 to 637658, EPI_ISL_637662 to 637666, EPI_ISL_637668 to 637673, EPI_ISL_637676 to 637678, EPI_ISL_637680 to 637713, EPI_ISL_637716, EPI_ISL_637721 to 637725, EPI_ISL_637727 to 637764, EPI_ISL_637766 to 637779, EPI_ISL_637801 to 637852, EPI_ISL_637856, EPI_ISL_637859 to 637860, EPI_ISL_637873, EPI_ISL_637877, EPI_ISL_637880, EPI_ISL_637888, EPI_ISL_637890, EPI_ISL_637903, EPI_ISL_637906, EPI_ISL_637913, EPI_ISL_637916, EPI_ISL_637936, EPI_ISL_637938 to 637940, EPI_ISL_637942, EPI_ISL_637947, EPI_ISL_637953, EPI_ISL_637955, EPI_ISL_637966 to 637967, EPI_ISL_637974, EPI_ISL_637978, EPI_ISL_637980, EPI_ISL_637982 to 637983, EPI_ISL_638001, EPI_ISL_638008, EPI_ISL_638013 to 638014, EPI_ISL_638016, EPI_ISL_638018 to 638020, EPI_ISL_638027, EPI_ISL_638029 to 638030, EPI_ISL_638038 to 638040, EPI_ISL_638066 to 638069, EPI_ISL_638071 to 638072, EPI_ISL_638079 to 638083, EPI_ISL_638131 to 638133, EPI_ISL_638138, EPI_ISL_638140 to 638143, EPI_ISL_638146, EPI_ISL_638165 to 638167, EPI_ISL_638169, EPI_ISL_638174, EPI_ISL_638177 to 638205, EPI_ISL_638208 to 638248, EPI_ISL_638258 to 638264, EPI_ISL_638293 to 638299, EPI_ISL_638302 to 638314, EPI_ISL_638316 to 638325, EPI_ISL_638327 to 638330, EPI_ISL_638332, EPI_ISL_638334 to 638335, EPI_ISL_638337 to 638425, EPI_ISL_638427 to 638448, EPI_ISL_638614 to 638681, EPI_ISL_638731 to 638735, EPI_ISL_638895 to 638910, EPI_ISL_638914, EPI_ISL_638916 to 638927, EPI_ISL_638936 to 638943, EPI_ISL_639003 to 639005, EPI_ISL_639624 to 639625, EPI_ISL_650122, EPI_ISL_650129, EPI_ISL_650133, EPI_ISL_650143, EPI_ISL_650149, EPI_ISL_650200, EPI_ISL_650217, EPI_ISL_650228, EPI_ISL_650236, EPI_ISL_650239 to 650240, EPI_ISL_650273, EPI_ISL_650364 to 650365, EPI_ISL_650376, EPI_ISL_650383, EPI_ISL_650390 to 650391, EPI_ISL_650396 to 650398, EPI_ISL_650400, EPI_ISL_650481, EPI_ISL_650553, EPI_ISL_650562, EPI_ISL_650564, EPI_ISL_650573 to 650574, EPI_ISL_650580, EPI_ISL_650583, EPI_ISL_650589 to 650596, EPI_ISL_650609, EPI_ISL_650612, EPI_ISL_650614 to 650617, EPI_ISL_650626 to 650633, EPI_ISL_650661, EPI_ISL_650664, EPI_ISL_650698, EPI_ISL_650707 to 650708, EPI_ISL_650768, EPI_ISL_650774, EPI_ISL_650836, EPI_ISL_650899, EPI_ISL_650910, EPI_ISL_650916, EPI_ISL_650922, EPI_ISL_650951 to 650952, EPI_ISL_650959, EPI_ISL_650962, EPI_ISL_650969 to 650972, EPI_ISL_650978, EPI_ISL_650983, EPI_ISL_650985 to 650986, EPI_ISL_650988 to 650994, EPI_ISL_651001 to 651010, EPI_ISL_651014 to 651047, EPI_ISL_651049 to 651052, EPI_ISL_651080 to 651081, EPI_ISL_651122, EPI_ISL_651136, EPI_ISL_651186, EPI_ISL_651276, EPI_ISL_651278, EPI_ISL_651281, EPI_ISL_651292, EPI_ISL_651302, EPI_ISL_651346, EPI_ISL_651348, EPI_ISL_651360, EPI_ISL_651363, EPI_ISL_651365, EPI_ISL_651370, EPI_ISL_651383, EPI_ISL_651423, EPI_ISL_651435 to 651436, EPI_ISL_651488 to 651490, EPI_ISL_651492, EPI_ISL_651542, EPI_ISL_651550 to 651582, EPI_ISL_651653 to 651657, EPI_ISL_651659, EPI_ISL_651662, EPI_ISL_651664, EPI_ISL_651666 to 651670, EPI_ISL_651672 to 651677, EPI_ISL_651679 to 651681, EPI_ISL_651683 to 651698, EPI_ISL_651702 to 651714, EPI_ISL_652157 to 652170, EPI_ISL_652172 to 652179, EPI_ISL_652286 to 652288, EPI_ISL_664120, EPI_ISL_664139 to 664141, EPI_ISL_664143, EPI_ISL_664149 to 664150, EPI_ISL_664167, EPI_ISL_664198 to 664200, EPI_ISL_664217, EPI_ISL_664221, EPI_ISL_664234, EPI_ISL_664237, EPI_ISL_664249, EPI_ISL_664282, EPI_ISL_664286, EPI_ISL_664291, EPI_ISL_664304, EPI_ISL_664316 to 664317, EPI_ISL_664326, EPI_ISL_664346, EPI_ISL_664378 to 664384, EPI_ISL_664397, EPI_ISL_664421 to 664422, EPI_ISL_664428, EPI_ISL_664432 to 664434, EPI_ISL_664446, EPI_ISL_664451, EPI_ISL_664453 to 664455, EPI_ISL_664470, EPI_ISL_664514, EPI_ISL_664516, EPI_ISL_664518, EPI_ISL_664527 to 664538, EPI_ISL_664542 to 664543, EPI_ISL_664546 to 664555, EPI_ISL_664567 to 664582, EPI_ISL_664584 to 664591, EPI_ISL_664598 to 664599, EPI_ISL_664601, EPI_ISL_664610, EPI_ISL_664616, EPI_ISL_664622, EPI_ISL_664625, EPI_ISL_664627, EPI_ISL_664631, EPI_ISL_664635, EPI_ISL_664655 to 664656, EPI_ISL_664659 to 664660, EPI_ISL_664662, EPI_ISL_664666, EPI_ISL_664684, EPI_ISL_664697 to 664698, EPI_ISL_664719, EPI_ISL_664721, EPI_ISL_664731, EPI_ISL_664748 to 664749, EPI_ISL_664765 to 664766, EPI_ISL_664768 to 664836, EPI_ISL_664941 to 664986, EPI_ISL_664988 to 664990, EPI_ISL_664992 to 665022, EPI_ISL_678832 to 678851, EPI_ISL_678853 to 678854, EPI_ISL_678856 to 678865, EPI_ISL_678867 to 678887, EPI_ISL_678889 to 678915, EPI_ISL_679134 to 679147, EPI_ISL_679149 to 679221, EPI_ISL_679223 to 679224, EPI_ISL_679226, EPI_ISL_679228, EPI_ISL_679230, EPI_ISL_679232, EPI_ISL_679234, EPI_ISL_679236, EPI_ISL_679238, EPI_ISL_679240, EPI_ISL_679242, EPI_ISL_679244, EPI_ISL_679246, EPI_ISL_679248, EPI_ISL_680037 to 680039, EPI_ISL_680041 to 680084, EPI_ISL_680086 to 680100, EPI_ISL_680571 to 680577, EPI_ISL_681260, EPI_ISL_702474, EPI_ISL_702500, EPI_ISL_702561, EPI_ISL_702703, EPI_ISL_702723, EPI_ISL_702734, EPI_ISL_702782, EPI_ISL_702787, EPI_ISL_702834, EPI_ISL_702847 to 702848, EPI_ISL_702860, EPI_ISL_702876, EPI_ISL_702883, EPI_ISL_702886, EPI_ISL_702891, EPI_ISL_702971, EPI_ISL_702977, EPI_ISL_703032, EPI_ISL_703076, EPI_ISL_703093, EPI_ISL_703115, EPI_ISL_703118, EPI_ISL_703127, EPI_ISL_703213, EPI_ISL_703238, EPI_ISL_703406, EPI_ISL_703432, EPI_ISL_703435, EPI_ISL_703438, EPI_ISL_703440, EPI_ISL_703451, EPI_ISL_703457, EPI_ISL_703459, EPI_ISL_703462, EPI_ISL_703465, EPI_ISL_703468, EPI_ISL_703471, EPI_ISL_703473, EPI_ISL_703476, EPI_ISL_703479, EPI_ISL_703508, EPI_ISL_703513, EPI_ISL_703516, EPI_ISL_703519, EPI_ISL_703521, EPI_ISL_703524, EPI_ISL_703619, EPI_ISL_703629, EPI_ISL_703632, EPI_ISL_703635, EPI_ISL_703679, EPI_ISL_703693, EPI_ISL_703708 to 703710, EPI_ISL_703713, EPI_ISL_703715 to 703716, EPI_ISL_703724, EPI_ISL_703741 to 703742, EPI_ISL_703747, EPI_ISL_704060, EPI_ISL_704076, EPI_ISL_704115, EPI_ISL_704203, EPI_ISL_704219, EPI_ISL_704222, EPI_ISL_704225, EPI_ISL_704247, EPI_ISL_704250, EPI_ISL_704252, EPI_ISL_704255, EPI_ISL_704257, EPI_ISL_704269, EPI_ISL_704273, EPI_ISL_704276, EPI_ISL_704280, EPI_ISL_704283, EPI_ISL_704286, EPI_ISL_704289, EPI_ISL_704291, EPI_ISL_704294, EPI_ISL_704297, EPI_ISL_704299, EPI_ISL_704302, EPI_ISL_704305, EPI_ISL_704307, EPI_ISL_704309, EPI_ISL_704328, EPI_ISL_704331, EPI_ISL_704333, EPI_ISL_704336, EPI_ISL_704339, EPI_ISL_704341, EPI_ISL_704344, EPI_ISL_704347, EPI_ISL_704350, EPI_ISL_704353, EPI_ISL_704356, EPI_ISL_704359, EPI_ISL_704361, EPI_ISL_704364, EPI_ISL_704366, EPI_ISL_704369, EPI_ISL_704374, EPI_ISL_704377, EPI_ISL_704379, EPI_ISL_704382, EPI_ISL_704384, EPI_ISL_704389, EPI_ISL_704392, EPI_ISL_704395, EPI_ISL_704427, EPI_ISL_704430, EPI_ISL_704433, EPI_ISL_704435, EPI_ISL_704438, EPI_ISL_704441, EPI_ISL_704443, EPI_ISL_704446, EPI_ISL_704449, EPI_ISL_704452, EPI_ISL_704454, EPI_ISL_704457, EPI_ISL_704460, EPI_ISL_704462, EPI_ISL_704465, EPI_ISL_704467, EPI_ISL_704470, EPI_ISL_704473, EPI_ISL_704478, EPI_ISL_704481, EPI_ISL_704484, EPI_ISL_704486, EPI_ISL_704489, EPI_ISL_704492, EPI_ISL_704494, EPI_ISL_704497, EPI_ISL_704499, EPI_ISL_704502, EPI_ISL_704505, EPI_ISL_704507, EPI_ISL_704510, EPI_ISL_704513, EPI_ISL_704516, EPI_ISL_704518, EPI_ISL_704521, EPI_ISL_704524, EPI_ISL_704526, EPI_ISL_704529, EPI_ISL_704532, EPI_ISL_704535, EPI_ISL_704537, EPI_ISL_704540, EPI_ISL_704543, EPI_ISL_704545, EPI_ISL_704548, EPI_ISL_704551, EPI_ISL_704554, EPI_ISL_704557, EPI_ISL_704606, EPI_ISL_704610 to 704613, EPI_ISL_704616 to 704618, EPI_ISL_704640 to 704641, EPI_ISL_704643, EPI_ISL_704646, EPI_ISL_704648, EPI_ISL_704651, EPI_ISL_704654, EPI_ISL_704656, EPI_ISL_704659, EPI_ISL_704662, EPI_ISL_704664, EPI_ISL_704834, EPI_ISL_704957, EPI_ISL_705110, EPI_ISL_705140, EPI_ISL_705217, EPI_ISL_705257, EPI_ISL_705289 to 705290, EPI_ISL_705333, EPI_ISL_705339, EPI_ISL_705346, EPI_ISL_705365, EPI_ISL_705376, EPI_ISL_705382, EPI_ISL_705410, EPI_ISL_705502, EPI_ISL_705529 to 705538, EPI_ISL_705540 to 705541, EPI_ISL_705544, EPI_ISL_705546 to 705548, EPI_ISL_705550 to 705551, EPI_ISL_705555 to 705557, EPI_ISL_705559 to 705566, EPI_ISL_705629, EPI_ISL_705631 to 705635, EPI_ISL_723852 to 723864, EPI_ISL_723866 to 723867, EPI_ISL_723869 to 723871, EPI_ISL_723873 to 723876, EPI_ISL_723878 to 723882, EPI_ISL_723885 to 723899, EPI_ISL_723901 to 723913, EPI_ISL_723915 to 723916, EPI_ISL_723919 to 723948, EPI_ISL_740953 to 740970, EPI_ISL_740972, EPI_ISL_740975 to 740985, EPI_ISL_740990 to 740994, EPI_ISL_740996 to 740998, EPI_ISL_741000 to 741002, EPI_ISL_741005, EPI_ISL_741007 to 741010, EPI_ISL_741012 to 741017, EPI_ISL_763370, EPI_ISL_763375, EPI_ISL_763379 to 763380, EPI_ISL_763448, EPI_ISL_763464, EPI_ISL_763469, EPI_ISL_763526, EPI_ISL_763542, EPI_ISL_763569, EPI_ISL_763592 to 763593, EPI_ISL_763637 to 763638, EPI_ISL_763666, EPI_ISL_763702, EPI_ISL_763790, EPI_ISL_763792 to 763794, EPI_ISL_763796, EPI_ISL_763801, EPI_ISL_763815 to 763816, EPI_ISL_763839, EPI_ISL_763843, EPI_ISL_763848 to 763850, EPI_ISL_763872 to 763881, EPI_ISL_763883 to 763891, EPI_ISL_764141, EPI_ISL_764159, EPI_ISL_764162, EPI_ISL_764164, EPI_ISL_764171, EPI_ISL_764176 to 764177, EPI_ISL_764191, EPI_ISL_764202, EPI_ISL_764209, EPI_ISL_764236, EPI_ISL_764276, EPI_ISL_764281, EPI_ISL_764284 to 764295, EPI_ISL_764297 to 764305, EPI_ISL_764307, EPI_ISL_764309, EPI_ISL_764312 to 764314, EPI_ISL_764316 to 764317, EPI_ISL_764319, EPI_ISL_764322 to 764327, EPI_ISL_764329 to 764355, EPI_ISL_813154 to 813159, EPI_ISL_813162 to 813173, EPI_ISL_813175 to 813187, EPI_ISL_813190 to 813194, EPI_ISL_813196, EPI_ISL_813198, EPI_ISL_813201 to 813207, EPI_ISL_813209 to 813217, EPI_ISL_813219 to 813227, EPI_ISL_813230 to 813235, EPI_ISL_813237 to 813243, EPI_ISL_813245 to 813246, EPI_ISL_813249 to 813262, EPI_ISL_813264 to 813268, EPI_ISL_813272 to 813279, EPI_ISL_813281 to 813290, EPI_ISL_813292 to 813294, EPI_ISL_813297 to 813324, EPI_ISL_813327, EPI_ISL_813329 to 813330, EPI_ISL_813333 to 813338, EPI_ISL_813340 to 813343, EPI_ISL_813345, EPI_ISL_813347 to 813355, EPI_ISL_837837 to 837842, EPI_ISL_837851, EPI_ISL_837853, EPI_ISL_837858, EPI_ISL_837866 to 837868, EPI_ISL_837872 to 837874, EPI_ISL_837880 to 837883, EPI_ISL_837885, EPI_ISL_837888 to 837889, EPI_ISL_837891, EPI_ISL_837893, EPI_ISL_837895, EPI_ISL_837897 to 837902, EPI_ISL_842110, EPI_ISL_864738 to 864745, EPI_ISL_864752 to 864753, EPI_ISL_864782 to 864783, EPI_ISL_864786 to 864788, EPI_ISL_864793, EPI_ISL_864796, EPI_ISL_864802, EPI_ISL_864804, EPI_ISL_864806, EPI_ISL_864808, EPI_ISL_864833, EPI_ISL_864837, EPI_ISL_864839, EPI_ISL_864847, EPI_ISL_864849, EPI_ISL_864851 to 864852, EPI_ISL_864854, EPI_ISL_864868 to 864871, EPI_ISL_864873 to 864874, EPI_ISL_864882, EPI_ISL_864888, EPI_ISL_864890, EPI_ISL_864892, EPI_ISL_864894

| | | | |
|---|---|---|---|
| see above | Department of Pathology, University of Cambridge | COVID-19 Genomics UK (COG-UK) Consortium | Aminu S. Jahun; Anna Yakovleva; Charlotte J. Houldcroft; Fahad A Khokhar; Grant Hall; Ian Goodfellow; Iliana Georgana; Laura G Caller; Luke W Meredith; M. Estee Torok; M. Estée Török; M. Est©e Trk; Malte Pinckert; Martin D. Curran; Myra Hosmillo; Sarah L. Caddy; Surendra Parmar; Theresa Feltwell; William L. Hamilton; Yasmin Chaudhry |
| EPI_ISL_458571, EPI_ISL_459173, EPI_ISL_459243, EPI_ISL_459258, EPI_ISL_524590, EPI_ISL_524595 | Department of Pathology, University of Cambridge | Wellcome Sanger Institute for the COVID-19 Genomics UK (COG-UK) Consortium | Aminu S. Jahun; Anna Yakovleva; Charlotte J. Houldcroft; Cordelia Langford; David K. Jackson; Dominic Kwiatkowski; Ewan Harrison; Fahad A Khokhar; Grant Hall; Ian Goodfellow; Ian Johnston; John Sillitoe on behalf of the Wellcome Sanger Institute COVID-19 Surveillance Team; Laura G Caller; Luke W Meredith; M. Estée Török; Martin D. Curran; Myra Hosmillo; Roberto Amato; Sarah L. Caddy; Sonia Goncalves; Theresa Feltwell; William L. Hamilton; and Alex Alderton |

EPI_ISL_438249 to 438253, EPI_ISL_438255 to 438268, EPI_ISL_438270 to 438285, EPI_ISL_438287 to 438324, EPI_ISL_438326 to 438370, EPI_ISL_438372 to 438380, EPI_ISL_438382 to 438404, EPI_ISL_438406 to 438434, EPI_ISL_438436 to 438443, EPI_ISL_438445 to 438464, EPI_ISL_438466 to 438472, EPI_ISL_438474 to 438493, EPI_ISL_438495 to 438525, EPI_ISL_438527 to 438545, EPI_ISL_439368 to 439391, EPI_ISL_439393 to 439399, EPI_ISL_439401 to 439446, EPI_ISL_439448 to 439470, EPI_ISL_439472 to 439477, EPI_ISL_439479 to 439527, EPI_ISL_439529 to 439568, EPI_ISL_439570 to 439573, EPI_ISL_439575 to 439664, EPI_ISL_439863 to 439885, EPI_ISL_439887 to 439912, EPI_ISL_439914 to 439938, EPI_ISL_439940 to 439947, EPI_ISL_439949 to 439952, EPI_ISL_439954, EPI_ISL_439956, EPI_ISL_439958, EPI_ISL_439961, EPI_ISL_439963, EPI_ISL_439970, EPI_ISL_439975, EPI_ISL_439977, EPI_ISL_439980, EPI_ISL_439987, EPI_ISL_439989 to 439991, EPI_ISL_439993 to 439994, EPI_ISL_439996 to 439997, EPI_ISL_439999, EPI_ISL_440003 to 440004, EPI_ISL_440007, EPI_ISL_440009 to 440012, EPI_ISL_440014 to 440015, EPI_ISL_440017, EPI_ISL_440019, EPI_ISL_440024 to 440025, EPI_ISL_440030, EPI_ISL_440034, EPI_ISL_440041, EPI_ISL_440044 to 440049, EPI_ISL_440053, EPI_ISL_440055, EPI_ISL_440057, EPI_ISL_440059 to 440062, EPI_ISL_440064, EPI_ISL_440066 to 440067, EPI_ISL_440069, EPI_ISL_440072 to 440080, EPI_ISL_440082 to 440083, EPI_ISL_440085 to 440086, EPI_ISL_440088 to 440090, EPI_ISL_440096 to 440097, EPI_ISL_440099, EPI_ISL_440104, EPI_ISL_440107 to 440112, EPI_ISL_440117, EPI_ISL_440120 to 440122, EPI_ISL_440125, EPI_ISL_440129, EPI_ISL_440132 to 440133, EPI_ISL_440138, EPI_ISL_440143 to 440144, EPI_ISL_440147, EPI_ISL_440150, EPI_ISL_440152, EPI_ISL_440155, EPI_ISL_440159, EPI_ISL_440161, EPI_ISL_440163 to 440164, EPI_ISL_440166, EPI_ISL_440169 to 440171, EPI_ISL_440173, EPI_ISL_440176 to 440177, EPI_ISL_440179 to 440186, EPI_ISL_440188, EPI_ISL_440193, EPI_ISL_440195, EPI_ISL_440197, EPI_ISL_440202 to 440203, EPI_ISL_440207 to 440209, EPI_ISL_440211, EPI_ISL_440213, EPI_ISL_440217, EPI_ISL_440223, EPI_ISL_440226, EPI_ISL_440230 to 440231, EPI_ISL_440234, EPI_ISL_440237, EPI_ISL_440239 to 440240, EPI_ISL_440244, EPI_ISL_440246 to 440247, EPI_ISL_440254, EPI_ISL_440257 to 440260, EPI_ISL_440263 to 440265, EPI_ISL_440267 to 440268, EPI_ISL_440270 to 440271, EPI_ISL_440279 to 440285, EPI_ISL_440287 to 440290, EPI_ISL_440293, EPI_ISL_440295 to 440297, EPI_ISL_440300 to 440328, EPI_ISL_440330 to 440341, EPI_ISL_440343 to 440365, EPI_ISL_440367 to 440393, EPI_ISL_440395 to 440440, EPI_ISL_440442 to 440444, EPI_ISL_440448, EPI_ISL_440450 to 440452, EPI_ISL_440455 to 440456, EPI_ISL_440459 to 440462, EPI_ISL_440464 to 440467, EPI_ISL_440469 to 440523, EPI_ISL_440525 to 440539, EPI_ISL_440541 to 440550, EPI_ISL_440552 to 440593, EPI_ISL_440595, EPI_ISL_440597 to 440622, EPI_ISL_440810 to 440829, EPI_ISL_440831 to 440850, EPI_ISL_441052 to 441063, EPI_ISL_441065 to 441121, EPI_ISL_441123 to 441162, EPI_ISL_441164 to 441195, EPI_ISL_441197 to 441225, EPI_ISL_441227 to 441310, EPI_ISL_441312 to 441317, EPI_ISL_441319 to 441327, EPI_ISL_441329 to 441335, EPI_ISL_441337 to 441349, EPI_ISL_441547 to 441566, EPI_ISL_441568 to 441583, EPI_ISL_441585 to 441624, EPI_ISL_441626 to 441652, EPI_ISL_441654 to 441658, EPI_ISL_441660 to 441661, EPI_ISL_441664 to 441669, EPI_ISL_441671 to 441674, EPI_ISL_441677 to 441684, EPI_ISL_441686 to 441689, EPI_ISL_441692, EPI_ISL_441695, EPI_ISL_441700 to 441703, EPI_ISL_441705 to 441707, EPI_ISL_441709 to 441711, EPI_ISL_441713, EPI_ISL_441715, EPI_ISL_441718, EPI_ISL_441720 to 441729, EPI_ISL_441731 to 441732, EPI_ISL_441734 to 441736, EPI_ISL_441738 to 441746, EPI_ISL_441748 to 441750, EPI_ISL_441752, EPI_ISL_441754 to 441755, EPI_ISL_441757 to 441760, EPI_ISL_441762 to 441764, EPI_ISL_441767 to 441769, EPI_ISL_441772 to 441774, EPI_ISL_441777 to 441780, EPI_ISL_441782 to 441804, EPI_ISL_441806 to 441809, EPI_ISL_441811 to 441833, EPI_ISL_441835 to 441842, EPI_ISL_441844 to 441846, EPI_ISL_442045 to 442072, EPI_ISL_442074 to 442226, EPI_ISL_442228 to 442245, EPI_ISL_442247 to 442270, EPI_ISL_442272 to 442297, EPI_ISL_442299 to 442343, EPI_ISL_442524 to 442532, EPI_ISL_442534 to 442565, EPI_ISL_442567 to 442573, EPI_ISL_442575 to 442608, EPI_ISL_442610 to 442620, EPI_ISL_442622 to 442623, EPI_ISL_442811 to 442847, EPI_ISL_442849 to 442876, EPI_ISL_442878 to 442890, EPI_ISL_442892 to 442910, EPI_ISL_442912 to 442944, EPI_ISL_442946 to 442977, EPI_ISL_442979 to 442986, EPI_ISL_442988 to 443037, EPI_ISL_443039 to 443051, EPI_ISL_443053 to 443056, EPI_ISL_443058 to 443068, EPI_ISL_443070 to 443107, EPI_ISL_443109 to 443118, EPI_ISL_443318 to 443331, EPI_ISL_443333 to 443353, EPI_ISL_443355 to 443403, EPI_ISL_443405 to 443423, EPI_ISL_443425 to 443432, EPI_ISL_443434 to 443437, EPI_ISL_443439 to 443451, EPI_ISL_443453 to 443466, EPI_ISL_443468 to 443474, EPI_ISL_443476 to 443520, EPI_ISL_443522 to 443623, EPI_ISL_443625 to 443654, EPI_ISL_443656 to 443674, EPI_ISL_443676 to 443685, EPI_ISL_458516 to 458531, EPI_ISL_458533 to 458563, EPI_ISL_458565 to 458566, EPI_ISL_458568 to 458570, EPI_ISL_458572 to 458577, EPI_ISL_459166 to 459172, EPI_ISL_459174 to 459187, EPI_ISL_459189 to 459200, EPI_ISL_459202 to 459224, EPI_ISL_459226 to 459240, EPI_ISL_459242, EPI_ISL_459244 to 459257, EPI_ISL_459259 to 459295, EPI_ISL_459297, EPI_ISL_459304, EPI_ISL_459306 to 459317, EPI_ISL_459319 to 459322, EPI_ISL_459410 to 459421, EPI_ISL_459423 to 459425, EPI_ISL_459427 to 459429, EPI_ISL_459431 to 459438, EPI_ISL_459440 to 459445, EPI_ISL_459447 to 459464, EPI_ISL_459466 to 459477, EPI_ISL_459479 to 459485, EPI_ISL_459488 to 459489, EPI_ISL_459491 to 459493, EPI_ISL_459495 to 459504, EPI_ISL_469891 to 469913, EPI_ISL_470090 to 470105, EPI_ISL_470107 to 470119, EPI_ISL_470121 to 470160, EPI_ISL_470162 to 470171, EPI_ISL_470173, EPI_ISL_470175 to 470178, EPI_ISL_470180 to 470196, EPI_ISL_470198 to 470211, EPI_ISL_470213 to 470232, EPI_ISL_470234 to 470246, EPI_ISL_470248 to 470256, EPI_ISL_470258 to 470344, EPI_ISL_470346 to 470353, EPI_ISL_470355, EPI_ISL_470358 to 470413, EPI_ISL_470415 to 470462, EPI_ISL_470464 to 470472, EPI_ISL_470474 to 470516, EPI_ISL_470518 to 470528, EPI_ISL_470531 to 470533, EPI_ISL_470535 to 470537, EPI_ISL_489381 to 489390, EPI_ISL_489394 to 489395, EPI_ISL_489397 to 489398, EPI_ISL_489400, EPI_ISL_489402 to 489406, EPI_ISL_489409, EPI_ISL_489411 to 489412, EPI_ISL_489414 to 489417, EPI_ISL_489419 to 489428, EPI_ISL_489430 to 489440, EPI_ISL_489443 to 489448, EPI_ISL_489450 to 489454, EPI_ISL_489456 to 489469, EPI_ISL_489471 to 489479, EPI_ISL_489481 to 489488, EPI_ISL_489490 to 489512, EPI_ISL_489514 to 489522, EPI_ISL_489524 to 489529, EPI_ISL_489531, EPI_ISL_489533 to 489535, EPI_ISL_489538 to 489542, EPI_ISL_489544 to 489546, EPI_ISL_489548 to 489565, EPI_ISL_489568 to 489577, EPI_ISL_492189 to 492194, EPI_ISL_492196, EPI_ISL_492198 to 492201, EPI_ISL_492204 to 492206, EPI_ISL_511959, EPI_ISL_511970, EPI_ISL_511982, EPI_ISL_524493, EPI_ISL_524495, EPI_ISL_524497 to 524498, EPI_ISL_524501 to 524503, EPI_ISL_524505 to 524506, EPI_ISL_524508 to 524509, EPI_ISL_524515, EPI_ISL_524521 to 524522, EPI_ISL_524528 to 524531, EPI_ISL_524537 to 524538, EPI_ISL_524541, EPI_ISL_524544, EPI_ISL_524546, EPI_ISL_524548, EPI_ISL_524551, EPI_ISL_524553, EPI_ISL_524558, EPI_ISL_524560, EPI_ISL_524564, EPI_ISL_524568, EPI_ISL_524570, EPI_ISL_524573 to 524584, EPI_ISL_524586 to 524589, EPI_ISL_524591 to 524594, EPI_ISL_524596 to 524610, EPI_ISL_524612 to 524618, EPI_ISL_591076 to 591079, EPI_ISL_591081, EPI_ISL_591091 to 591093

| | | | |
|---|---|---|---|
| see above | Department of Pathology, University of Cambridge | Wellcome Sanger Institute for the COVID-19 Genomics UK | Alex Alderton; Aminu S. Jahun; Anna Yakovleva; Charlotte J. Houldcroft; Cordelia Langford; David K. Jackson; Dominic Kwiatkowski; Ewan Harrison; |

| | | | |
|---|---|---|---|
| | | (COG-UK) consortium | Fahad A Khokhar; Grant Hall; Ian Goodfellow; Ian Johnston; John Sillitoe on behalf of the Wellcome Sanger Institute COVID-19 Surveillance Team; John Sillitoe on behalf of the Wellcome Sanger Institute COVID-19 Surveillance Team (http://www.sanger.ac.uk/covid-team); Laura G Caller; Luke W Meredith; M. Estée Török; Martin D. Curran; Myra Hosmillo; Roberto Amato; Sarah L. Caddy; Sonia Goncalves; Theresa Feltwell; William L. Hamilton; and Alex Alderton |
| EPI_ISL_576146 to 576149 | Department of Respiratory & Other Viral Infections of L.V. Gromashevsky Institute of Epidemiology & Infectious Diseases NAMS of Ukraine | Department of Respiratory & Other Viral Infections of L.V. Gromashevsky Institute of Epidemiology & Infectious Diseases NAMS of Ukraine, JSC "Farmak" | Alla Mironenko; Andriy Goy; Ihor Kravchuk; Larysa Radchenko; Liudmyla Bolotova; Nataliia Teteriuk |
| EPI_ISL_582509 | Department of Respiratory and other Viral Infections of L.V.Gromashevsky Institute of Epidemiology & Infectious Diseases NAMS of Ukraine | Department of Respiratory and other Viral Infections of L.V.Gromashevsky Institute of Epidemiology & Infectious Diseases NAMS of Ukraine, JSC "Farmak" | Alla Mironenko; Andriy Goy; Ihor Kravchuk; Larysa Radchenko; Ludmyla Bolotova; Nataliia Teteriuk |
| EPI_ISL_582510 | Department of Respiratory and other Viral Infections of L.V.Gromashevsky Institute of Epidemiology & Infectious Diseases NAMS of Ukraine | Department of Respiratory and other Viral Infections of L.V.Gromashevsky Institute of Epidemiology & Infectious Diseases NAMS of Ukrain, JSC "Farmak" | Alla Mironenko; Andriy Goy; Ihor Kravchuk; Larysa Radchenko; Ludmyla Bolotova; Nataliia Teteriuk |
| EPI_ISL_582511 to 582513, EPI_ISL_654819 to 654820 | Department of Respiratory and other Viral Infections of L.V.Gromashevsky Institute of Epidemiology & Infectious Diseases NAMS of Ukrain | Department of Respiratory and other Viral Infections of L.V.Gromashevsky Institute of Epidemiology & Infectious Diseases NAMS of Ukrain, JSC "Farmak" | Alla Mironenko; Andriy Goy; Ihor Kravchuk; Larysa Radchenko; Ludmyla Bolotova; Nataliia Teteriuk |
| EPI_ISL_654818 | Department of Respiratory and other Viral Infections of L.V.Gromashevsky Institute of Epidemiology & Infectious Diseases NAMS of Ukrain | Department of Respiratory and other Viral Infections of L.V.Gromashevsky Institute of Epidemiology & Infectious Diseases NAMS of Ukrain, JSC "Farmak" | Alla Mironenko; Andriy Goy; Ihor Kravchuk; Larysa Radchenko; Ludmyla Bolotova; Nataliia Teteriuk |
| EPI_ISL_450198 to 450210 | Department of Virology | Department of Virology | Ackermann, N.; Antwerpen, M.; Bengs, K.; Berger, A.; Boehm, S.; Boehmer; Boender; Buchholz, U.; Cai, W.; Corman; D.V.; Dangel, A.; Drosten, C.; Eberle, U.; Fingerle, V.; Grahl, A.; Haas, W.; Hamouda, O.; Hoch, M.; Hoermansdorfer, S.; Ippisch, S.; Jones; Katz, K.; Konrad, R.; Liebl, B.; M.M.; Marosevic; Muehlemann, B.; Muller, N.; Poertner, K.; Protzer, U.; Reich, A.; Rexroth, U.; Schneider, J.; Sing, A.; T.C.; T.S.; Treis, B.; V.M.; Veith, T.; Walter, M.; Wicklein, B.; Woelfel, R.; Woudenberg, T.; Zapf, A.; Zeitlmann, N.; an der Heiden, M. |
| EPI_ISL_768526 to 768527, EPI_ISL_804007 to 804008 | Department of Virology I, National Institute of Infectious Diseases | Pathogen Genomics Center, National Institute of Infectious Diseases | Kentaro Itokawa; Makoto Kuroda; Masanori Hashino; Rina Tanaka; Shuetsu Fukushi; Souichi Yamada; Tsuyoshi Sekizuka |
| EPI_ISL_529135 | Department of Virology III, National Institute of Infectious Diseases | Pathogen Genomics Center, National Institute of Infectious Diseases | Kentaro Itokawa; Makoto Kuroda; Makoto Takeda; Masanori Hashino; Rina Tanaka; Shutoku Matsuyama; Takaji Wakita; Tsuyoshi Sekizuka |
| EPI_ISL_413602 to 413605, EPI_ISL_414640 to 414643, EPI_ISL_414645, EPI_ISL_418385 to 418406, EPI_ISL_418408 to 418411, EPI_ISL_481513 to 481715, EPI_ISL_481717 to 481740, EPI_ISL_732565 to 732598, EPI_ISL_732600 to 732631, EPI_ISL_732633 to 732640, EPI_ISL_732642 to 732644, EPI_ISL_732646 to 732653, EPI_ISL_737034 to 737080, EPI_ISL_737209 to 737235, EPI_ISL_738134 to 738135, EPI_ISL_755943 to 756125, EPI_ISL_756165 to 756183, EPI_ISL_756247 to 756252, EPI_ISL_756282 to 756292, EPI_ISL_757288 to 757394, EPI_ISL_759763 to 759841, EPI_ISL_759899 to 759954, EPI_ISL_766016 to 766027, EPI_ISL_833201 to 833205, EPI_ISL_833207, EPI_ISL_833210 to 833212, EPI_ISL_833214 to 833219, EPI_ISL_833227, EPI_ISL_862052 to 862074, EPI_ISL_862082 to 862120 | | | |
| see above | Department of Virology and Immunology, University of Helsinki and Helsinki University Hospital, Huslab Finland | Department of Virology, Faculty of Medicine, University of Helsinki, Helsinki, Finland | Essi Korhonen; Fathiah Zakham; Hanna Liimatainen; Hannimari Kallio-Kokko; Harri Kangas; Hussein Alburkat; Jenni Virtanen; Maija Lappalainen; Maija Suvanto; Olli Vapalahti; Pekka Ellonen; Phuoc Truong; Ravi Kant; Sari Hannula; Satu Kurkela; Teemu Smura |
| EPI_ISL_515974 | Department of Virus and Microbiological Special Diagnostics, Statens Serum Institut | Albertsen lab, Department of Chemistry and Bioscience, Aalborg University, Denmark | Danish Corona Danica Consortia |
| EPI_ISL_856612 to 856614, EPI_ISL_856617, EPI_ISL_856620 to 856644, EPI_ISL_856646 to 856651, EPI_ISL_856653 to 856672, EPI_ISL_856675 to 856676 | Department of Virus and Microbiological Special Diagnostics, Statens Serum Institut, Copenhagen, Denmark | Aalborg University | Danish Covid-19 Genome Consortium |
| EPI_ISL_668458 to 668464, EPI_ISL_668467 to 668502, EPI_ISL_668504 to 668509, EPI_ISL_668511 to 668513, EPI_ISL_668516 to 668517, EPI_ISL_668519 to 668522, EPI_ISL_668524 to 668543, EPI_ISL_668546 to 668556, EPI_ISL_668558 to 668596, EPI_ISL_668598 to 668608, EPI_ISL_668613, EPI_ISL_668615 to 668616, EPI_ISL_668618, EPI_ISL_668620 to 668621, EPI_ISL_668623, EPI_ISL_668627 to 668631, EPI_ISL_668633 to 668658, EPI_ISL_668660 to 668680, EPI_ISL_668682 to 668686, EPI_ISL_668688 to 668700, EPI_ISL_668702 to 668721, EPI_ISL_668723 to 668750, EPI_ISL_668752 to 668758, EPI_ISL_668760, EPI_ISL_668762 to 668789, EPI_ISL_668791 to 668830, EPI_ISL_668832 to 668837, EPI_ISL_668839 to 668853, EPI_ISL_668855, EPI_ISL_668858 to 668861, EPI_ISL_668863 to 668864, EPI_ISL_668867 to 668875, EPI_ISL_668877 to 668878, EPI_ISL_668880 to 668885, EPI_ISL_668887, EPI_ISL_668889, EPI_ISL_668893 to 668894, EPI_ISL_668896, EPI_ISL_668900, EPI_ISL_668903 to 668905, EPI_ISL_668907 to 668916, EPI_ISL_668918, EPI_ISL_668923 to 668925, EPI_ISL_668927 to 668928, EPI_ISL_668930 to 668933, EPI_ISL_668935 to 668937, EPI_ISL_668940, EPI_ISL_668943 to 668944, EPI_ISL_668946 to 668952, EPI_ISL_668954, EPI_ISL_668956 to 668959, EPI_ISL_668961 to 668966, EPI_ISL_668968 to 668970, EPI_ISL_668972 to 668977, EPI_ISL_668979, EPI_ISL_668981 to 668995, EPI_ISL_668997 to 668999, EPI_ISL_669001 to 669005, EPI_ISL_669008, EPI_ISL_669011, EPI_ISL_669013, EPI_ISL_669016 to 669018, EPI_ISL_669020 to 669023, EPI_ISL_669025 to 669033, EPI_ISL_669035, EPI_ISL_669037 to 669039, EPI_ISL_669041 to 669049, EPI_ISL_669051 to 669054, EPI_ISL_669057 to 669063, EPI_ISL_669066 to 669074, EPI_ISL_669076 to 669081, EPI_ISL_669083 to 669093, EPI_ISL_669095 to 669100, EPI_ISL_669102 to 669104, EPI_ISL_669106, EPI_ISL_669109, EPI_ISL_669111 to 669115, EPI_ISL_669117 to 669120, EPI_ISL_669122 to 669130, EPI_ISL_669132 to 669141, EPI_ISL_669143 to 669145, EPI_ISL_669147 to 669153, EPI_ISL_669155 to 669158, EPI_ISL_669160 to 669165, EPI_ISL_669167 to 669169, EPI_ISL_669171 to 669175, EPI_ISL_669178 to 669184, EPI_ISL_669186 to 669188, EPI_ISL_669190 to 669191, EPI_ISL_669193 to 669213, EPI_ISL_669215, EPI_ISL_669217, EPI_ISL_669219 to 669220, EPI_ISL_669222 to 669226, EPI_ISL_669228 to 669234, EPI_ISL_669237 to 669255, EPI_ISL_669257 to 669259, EPI_ISL_669261 to 669265, EPI_ISL_669267, EPI_ISL_669269 to 669272, EPI_ISL_669275, EPI_ISL_669277 to 669283, EPI_ISL_669285 to 669294, EPI_ISL_669296 to 669297, EPI_ISL_669299 to 669323, EPI_ISL_669325 to 669326, EPI_ISL_669328 to 669329, EPI_ISL_669331, EPI_ISL_669333 to 669337, EPI_ISL_669339, EPI_ISL_669342 to 669357, EPI_ISL_669359 to 669360, EPI_ISL_669363 to 669379, EPI_ISL_669381 to 669401, EPI_ISL_669403 to 669404, EPI_ISL_669406 to 669407, EPI_ISL_669409 to 669420, EPI_ISL_669423 to 669428, EPI_ISL_669430 to 669433, EPI_ISL_669435 to 669439, EPI_ISL_669441 to 669449, EPI_ISL_669452 to 669453, EPI_ISL_669455, EPI_ISL_669458, EPI_ISL_669460 to 669478, EPI_ISL_669480 to 669498, EPI_ISL_669500, EPI_ISL_669502, EPI_ISL_669504 to 669507, EPI_ISL_669510 to 669512, EPI_ISL_669515 to 669539, EPI_ISL_669541 to 669558, EPI_ISL_669560 to 669566, EPI_ISL_669570 to 669571, EPI_ISL_669573 to 669581, EPI_ISL_669584 to 669590, EPI_ISL_669593, EPI_ISL_669596, EPI_ISL_669599 to 669638, EPI_ISL_669641 to 669648, EPI_ISL_669650 to 669652, EPI_ISL_669654 to 669688, EPI_ISL_669690 to 669698, EPI_ISL_669700 to 669727, EPI_ISL_669729 to 669731, EPI_ISL_669733 to 669750, EPI_ISL_669752 to 669755, EPI_ISL_669757 to 669817, EPI_ISL_669819 to 669827, EPI_ISL_669829 to 669835, EPI_ISL_669837 to 669841, EPI_ISL_669844 to 669845, EPI_ISL_669847, EPI_ISL_669849 to 669856, EPI_ISL_669859 to 669867, EPI_ISL_669869, EPI_ISL_669872 to 669874, EPI_ISL_669876 to 669878, EPI_ISL_669880 to 669888, EPI_ISL_669890 to 669891, EPI_ISL_669894 to 669896, EPI_ISL_669899 to 669901, EPI_ISL_669906 to 669907, EPI_ISL_669909 to 669910, EPI_ISL_669913 to 669916, EPI_ISL_669918 to 669919, EPI_ISL_669921, EPI_ISL_669923, EPI_ISL_669926, EPI_ISL_669929 to 669935, EPI_ISL_669938, EPI_ISL_669940 to 669941, EPI_ISL_669943 to 669945, EPI_ISL_669948 to 669949, EPI_ISL_669951 to 669975, EPI_ISL_669977 to 669983, EPI_ISL_669985 to 669990, EPI_ISL_669993 to 669995, EPI_ISL_669997 to 670000, EPI_ISL_670002, EPI_ISL_670005 to 670010, EPI_ISL_670012, EPI_ISL_670014 to 670016, EPI_ISL_670018 to 670038, EPI_ISL_670040 to 670042, EPI_ISL_670044 to 670045, EPI_ISL_670048 to 670051, EPI_ISL_670053 to 670054, EPI_ISL_670056 to 670061, EPI_ISL_670063 to 670069, EPI_ISL_670072 to 670076, EPI_ISL_670079 to 670087, EPI_ISL_670089 to 670102, EPI_ISL_670104 to 670105, EPI_ISL_670109, EPI_ISL_670112 to 670123, EPI_ISL_670126 to 670127, EPI_ISL_670129 to 670133, EPI_ISL_670135 to 670136, EPI_ISL_670138, EPI_ISL_670140 to 670141, EPI_ISL_670143 to 670145, EPI_ISL_670148 to 670153, EPI_ISL_670155 to 670171, EPI_ISL_670174, EPI_ISL_670176 to 670184, EPI_ISL_670186 to 670190, EPI_ISL_670192 to 670194, EPI_ISL_670196 to 670199, EPI_ISL_670201 to 670205, EPI_ISL_670207 to 670217, EPI_ISL_670219 to 670224, EPI_ISL_670226, EPI_ISL_670228 to 670233, EPI_ISL_670235, EPI_ISL_670237, EPI_ISL_670239 to 670242, EPI_ISL_670244 to 670247, EPI_ISL_670249 to 670259, EPI_ISL_670261 to 670271, EPI_ISL_670273 to 670282, EPI_ISL_670284 to 670300, EPI_ISL_670302 to 670304, EPI_ISL_670306 to 670307, EPI_ISL_670309, EPI_ISL_670311 to 670313, EPI_ISL_670319, EPI_ISL_670321 to 670329, EPI_ISL_670331 to 670335, EPI_ISL_670337 to 670341, EPI_ISL_670343 to 670351, EPI_ISL_670353 to 670357, EPI_ISL_670359, EPI_ISL_670362, EPI_ISL_670364 to 670370, EPI_ISL_670373 to 670382, EPI_ISL_670384 to 670389, EPI_ISL_670393 to 670401, EPI_ISL_670403 to 670406, EPI_ISL_670408 to 670414, EPI_ISL_670416 to 670423, EPI_ISL_670427, EPI_ISL_670429 to 670430, EPI_ISL_670432, EPI_ISL_670436 to 670443, EPI_ISL_670445 to 670446, EPI_ISL_670448 to 670454, EPI_ISL_670457 to 670461, EPI_ISL_670463 to 670469, EPI_ISL_670471 to 670516, EPI_ISL_670518 to 670519, EPI_ISL_670521 to 670522, EPI_ISL_670524 to 670525, EPI_ISL_670527 to 670530, EPI_ISL_670533, EPI_ISL_670535 to 670536, EPI_ISL_670538 to 670541, EPI_ISL_670543 to 670545, EPI_ISL_670547 to 670548, EPI_ISL_670550 to 670580, EPI_ISL_670582 to 670588, EPI_ISL_670590 to 670591, EPI_ISL_670593, EPI_ISL_670595 to 670600, EPI_ISL_670602 to 670608, EPI_ISL_670610 to 670625, EPI_ISL_670627 to 670628, EPI_ISL_670630, EPI_ISL_670632 to 670639, EPI_ISL_670641 to 670652, EPI_ISL_670654 to 670656, EPI_ISL_670658 to 670675, EPI_ISL_670678 to 670698, EPI_ISL_670700, EPI_ISL_670702 to 670709, EPI_ISL_670711 to 670712, EPI_ISL_670714 to 670721, EPI_ISL_670723 to 670729, EPI_ISL_670731 to 670745, EPI_ISL_670747 to 670765, EPI_ISL_670767 to 670771, EPI_ISL_670774, EPI_ISL_670776 to 670783, EPI_ISL_670785 to 670789, EPI_ISL_670791 to 670795, EPI_ISL_670798 to 670799, EPI_ISL_670801 to 670804, EPI_ISL_670806, EPI_ISL_670808 to 670816, EPI_ISL_670818 to 670826, EPI_ISL_670829 to 670839, EPI_ISL_670843 to 670845, EPI_ISL_670847, EPI_ISL_670850 to 670853, EPI_ISL_670855 to 670857, EPI_ISL_670859 to 670860, EPI_ISL_670863 to 670872, EPI_ISL_670874 to 670884, EPI_ISL_670886 to 670887, EPI_ISL_670891 to 670895, EPI_ISL_670899 to 670902, EPI_ISL_670904 to 670905, EPI_ISL_670907 to 670912, EPI_ISL_670914, EPI_ISL_670916 to 670918, EPI_ISL_670920 to 670922, EPI_ISL_670924 to 670925, EPI_ISL_670927 to 670930, EPI_ISL_670932, EPI_ISL_670936 to 670945, EPI_ISL_670947, EPI_ISL_670949 to 670953, EPI_ISL_670955 to 670959, EPI_ISL_670961 to 670962, EPI_ISL_670964 to 670969, EPI_ISL_670971 to 670973, EPI_ISL_670975 to 670982, EPI_ISL_670984 to 670985, EPI_ISL_670988 to 670989, EPI_ISL_670991 to 670996, EPI_ISL_670998 to 671001, EPI_ISL_671003 to 671007, EPI_ISL_671009 to 671017, EPI_ISL_671019 to 671023, EPI_ISL_671028 to 671030, EPI_ISL_671032 to 671034, EPI_ISL_671036 to 671039, EPI_ISL_671041 to 671042, EPI_ISL_671044 to 671046, EPI_ISL_671048 to 671049, EPI_ISL_671051 to 671054, EPI_ISL_671056 to 671061, EPI_ISL_671063 to 671064, EPI_ISL_671066 to 671072, EPI_ISL_671074 to 671084, EPI_ISL_671086 to 671091, EPI_ISL_671093 to 671098, EPI_ISL_671100 to 671101, EPI_ISL_671105 to 671112, EPI_ISL_671116, EPI_ISL_671118 to 671150, EPI_ISL_671152 to 671156, EPI_ISL_671158, EPI_ISL_671161 to 671162, EPI_ISL_671164, EPI_ISL_671166 to 671175, EPI_ISL_671178, EPI_ISL_671180 to 671184, EPI_ISL_671186 to 671187, EPI_ISL_671190 to 671204, EPI_ISL_671206 to 671220, EPI_ISL_671223 to 671231, EPI_ISL_671233 to 671242, EPI_ISL_671244 to 671245, EPI_ISL_671249 to 671252, EPI_ISL_671254 to 671261, EPI_ISL_671263 to 671266, EPI_ISL_671268 to 671269, EPI_ISL_682353 to 682404, EPI_ISL_682406 to 682420, EPI_ISL_682422 to 682425, EPI_ISL_682427 to 682454, EPI_ISL_682456 to 682517, EPI_ISL_682519 to 682529, EPI_ISL_682531 to 682581, EPI_ISL_682583 to 682589, EPI_ISL_682591 to 682651, EPI_ISL_682653 to 682700, EPI_ISL_682702 to 682715, EPI_ISL_682717 to 682789, EPI_ISL_682791 to 682859, EPI_ISL_682862 to 682876, EPI_ISL_682878 to 682890, EPI_ISL_682892 to 682908, EPI_ISL_682910 to 682959, EPI_ISL_682961 to 682963, EPI_ISL_682965 to 682966, EPI_ISL_682970 to 682974, EPI_ISL_682977 to 682978, EPI_ISL_682980 to 682982, EPI_ISL_682984 to 682985, EPI_ISL_682987 to 682988, EPI_ISL_682990 to 682999, EPI_ISL_683001 to 683002, EPI_ISL_683004, EPI_ISL_711060 to 711070, EPI_ISL_711075 to 711076, EPI_ISL_711078 to 711175, EPI_ISL_711177 to 711187, EPI_ISL_711190 to 711192, EPI_ISL_711195 to 711213, EPI_ISL_711217 to 711272, EPI_ISL_711275, EPI_ISL_711277 to 711280, EPI_ISL_711283 to 711302, EPI_ISL_711304 to 711428, EPI_ISL_711430 to 711545, EPI_ISL_711547 to 711701, EPI_ISL_711703 to 711743, EPI_ISL_711745 to 711840, EPI_ISL_711842 to 711865, EPI_ISL_711867 to 711941, EPI_ISL_711943 to 712004, EPI_ISL_712006 to 712034, EPI_ISL_712036 to 712045, EPI_ISL_712047 to 712059, EPI_ISL_712097 to 712112, EPI_ISL_712114 to 712225, EPI_ISL_712227 to 712308, EPI_ISL_712310 to 712318, EPI_ISL_712320 to 712336, EPI_ISL_712338 to 712567, EPI_ISL_712569 to 712799, EPI_ISL_712801 to 713122, EPI_ISL_713125 to 713208, EPI_ISL_713210 to 713241, EPI_ISL_713247 to 713267, EPI_ISL_713269 to 713294, EPI_ISL_713296 to 713303, EPI_ISL_713305 to 713306, EPI_ISL_713309 to 713373, EPI_ISL_713375 to 713511, EPI_ISL_713513 to 713559, EPI_ISL_713561 to 713563, EPI_ISL_713565 to 713571, EPI_ISL_713573 to 713586, EPI_ISL_713588 to 713614, EPI_ISL_713616 to 713727, EPI_ISL_713729 to 713769, EPI_ISL_713771 to 713890, EPI_ISL_713892 to 713922, EPI_ISL_713924 to 713980, EPI_ISL_713982 to 713992, EPI_ISL_713994 to 714029, EPI_ISL_714031 to 714104, EPI_ISL_714106 to 714107, EPI_ISL_714109 to 714190, EPI_ISL_714192 to 714220, EPI_ISL_714224 to 714292, EPI_ISL_714294 to 714335, EPI_ISL_714337 to 714365, EPI_ISL_714368 to 714398, EPI_ISL_714400 to 714479, EPI_ISL_714481 to 714564, EPI_ISL_714566 to 714864, EPI_ISL_714866 to 714927, EPI_ISL_714929 to 714937, EPI_ISL_747527 to 747530, EPI_ISL_747533 to 747536, EPI_ISL_747538 to 747592, EPI_ISL_747594 to 747595, EPI_ISL_747599 to 747614, EPI_ISL_747616 to 747638, EPI_ISL_747640 to 747641, EPI_ISL_747643 to 747644, EPI_ISL_747646, EPI_ISL_747648 to 747650, EPI_ISL_747652, EPI_ISL_747654, EPI_ISL_747656 to 747732, EPI_ISL_747734 to 747771, EPI_ISL_747773 to 747831, EPI_ISL_747833 to 747851, EPI_ISL_747853 to 747905, EPI_ISL_747907 to 747943, EPI_ISL_747945, EPI_ISL_747947 to 747956, EPI_ISL_747958 to 747983, EPI_ISL_747985 to 747999, EPI_ISL_748001 to 748092, EPI_ISL_748094 to 748097, EPI_ISL_748099 to 748105, EPI_ISL_748107 to 748137, EPI_ISL_748146 to 748157, EPI_ISL_748161, EPI_ISL_748164 to 748219, EPI_ISL_748221 to 748242, EPI_ISL_748244 to 748257, EPI_ISL_748260 to 748327, EPI_ISL_748329 to 748340, EPI_ISL_748342 to 748350, EPI_ISL_748352 to 748476, EPI_ISL_748478 to 748507, EPI_ISL_748509 to 748513, EPI_ISL_748515 to 748546, EPI_ISL_748548 to 748550, EPI_ISL_748552 to 748592, EPI_ISL_748594 to 748636, EPI_ISL_748638 to 748666, EPI_ISL_748668 to 748712, EPI_ISL_748714 to 748895, EPI_ISL_748897 to 748966, EPI_ISL_748968 to 748989, EPI_ISL_748991 to 748996, EPI_ISL_748998 to 749035, EPI_ISL_749037 to 749084, EPI_ISL_749086, EPI_ISL_749088 to 749128, EPI_ISL_749130 to 749147, EPI_ISL_749156 to 749158, EPI_ISL_749160 to 749167, EPI_ISL_749169 to 749171, EPI_ISL_749174 to 749184, EPI_ISL_749186 to 749237, EPI_ISL_749239 to 749265, EPI_ISL_749267 to 749274, EPI_ISL_749277 to 749361, EPI_ISL_749363, EPI_ISL_749365 to 749366, EPI_ISL_749368, EPI_ISL_749370 to 749375, EPI_ISL_749377 to 749435, EPI_ISL_749437 to | | | |

749473, EPI_ISL_749475 to 749621, EPI_ISL_749623 to 749705, EPI_ISL_749707 to 749723, EPI_ISL_749725 to 749737, EPI_ISL_749739 to 749812, EPI_ISL_749814 to 749816, EPI_ISL_749818 to 749905, EPI_ISL_749907 to 750098, EPI_ISL_750100 to 750107, EPI_ISL_750109 to 750160, EPI_ISL_750180 to 750187, EPI_ISL_750189 to 750254, EPI_ISL_750258 to 750259, EPI_ISL_750261 to 750283, EPI_ISL_750285 to 750363, EPI_ISL_750365 to 750372, EPI_ISL_750374 to 750376, EPI_ISL_750378 to 750391, EPI_ISL_750393 to 750411, EPI_ISL_750413 to 750429, EPI_ISL_750431 to 750515, EPI_ISL_750517 to 750617, EPI_ISL_750619 to 750625, EPI_ISL_750627 to 750731, EPI_ISL_750733 to 750819, EPI_ISL_750821 to 750835, EPI_ISL_750837 to 751010, EPI_ISL_751012 to 751039, EPI_ISL_751041 to 751123, EPI_ISL_751125 to 751152, EPI_ISL_751155 to 751183, EPI_ISL_757395 to 757443, EPI_ISL_757445, EPI_ISL_757447 to 757706, EPI_ISL_757708, EPI_ISL_757714 to 757852, EPI_ISL_757854, EPI_ISL_757857 to 757859, EPI_ISL_757862 to 757864, EPI_ISL_757868, EPI_ISL_757871 to 757989, EPI_ISL_757991 to 758082, EPI_ISL_758084 to 758158, EPI_ISL_758160 to 758250, EPI_ISL_758252 to 758618, EPI_ISL_758620 to 758692, EPI_ISL_758694 to 758708, EPI_ISL_758711, EPI_ISL_758714 to 758850, EPI_ISL_758852 to 758856, EPI_ISL_758858 to 758868, EPI_ISL_758870 to 758942, EPI_ISL_758944 to 758953, EPI_ISL_758955 to 758958, EPI_ISL_758960 to 758963, EPI_ISL_758966 to 759082, EPI_ISL_759084 to 759227, EPI_ISL_759229 to 759263, EPI_ISL_759265 to 759438, EPI_ISL_759440 to 759459, EPI_ISL_759461 to 759469, EPI_ISL_759471 to 759700, EPI_ISL_792708 to 792710, EPI_ISL_792712 to 792713, EPI_ISL_792716 to 792788, EPI_ISL_792790 to 792797, EPI_ISL_792799 to 792885, EPI_ISL_792887 to 792919, EPI_ISL_792921 to 793012, EPI_ISL_793015 to 793018, EPI_ISL_793020 to 793021, EPI_ISL_793024, EPI_ISL_793027 to 793066, EPI_ISL_793068 to 793114, EPI_ISL_793116 to 793123, EPI_ISL_793125 to 793128, EPI_ISL_793131 to 793135, EPI_ISL_793137 to 793139, EPI_ISL_793141, EPI_ISL_793144 to 793178, EPI_ISL_793181 to 793196, EPI_ISL_793198 to 793254, EPI_ISL_793256, EPI_ISL_793258 to 793269, EPI_ISL_793272 to 793292, EPI_ISL_793294 to 793389, EPI_ISL_793391, EPI_ISL_793393 to 793412, EPI_ISL_793415 to 793660, EPI_ISL_793662 to 793664, EPI_ISL_793666 to 793700, EPI_ISL_793702 to 793704, EPI_ISL_793706 to 793733, EPI_ISL_793736 to 793807, EPI_ISL_793809 to 793822, EPI_ISL_793824 to 793890, EPI_ISL_793892 to 793919, EPI_ISL_793921 to 793923, EPI_ISL_793925 to 794013, EPI_ISL_794324 to 794363, EPI_ISL_794365 to 794372, EPI_ISL_794374 to 794519, EPI_ISL_794521 to 794591, EPI_ISL_794830 to 794859, EPI_ISL_794861 to 794867, EPI_ISL_794869 to 794879, EPI_ISL_794881 to 795021, EPI_ISL_795023 to 795024, EPI_ISL_795028 to 795161, EPI_ISL_795166 to 795171, EPI_ISL_795175 to 795184, EPI_ISL_795186 to 795216, EPI_ISL_795218 to 795252, EPI_ISL_795254 to 795256, EPI_ISL_795258 to 795290, EPI_ISL_795292 to 795335, EPI_ISL_795337 to 795424, EPI_ISL_795426 to 795450, EPI_ISL_795452, EPI_ISL_795454 to 795463, EPI_ISL_795465 to 795552, EPI_ISL_795554 to 795778, EPI_ISL_795780 to 795855, EPI_ISL_795857 to 795999, EPI_ISL_796001 to 796010, EPI_ISL_815401 to 815402, EPI_ISL_815406 to 815432, EPI_ISL_815434 to 815448, EPI_ISL_815450 to 815469, EPI_ISL_815471, EPI_ISL_815473 to 815474, EPI_ISL_815476 to 815489, EPI_ISL_815491, EPI_ISL_815493 to 815533, EPI_ISL_815535 to 815536, EPI_ISL_815539 to 815620, EPI_ISL_815622 to 815634, EPI_ISL_815636 to 815646, EPI_ISL_815648 to 815662, EPI_ISL_815664, EPI_ISL_815666 to 815670, EPI_ISL_815672 to 815701, EPI_ISL_815703 to 815711, EPI_ISL_815713 to 815714, EPI_ISL_815716 to 815766, EPI_ISL_815768 to 815820, EPI_ISL_815822 to 815870, EPI_ISL_815872 to 816037, EPI_ISL_816039 to 816112, EPI_ISL_816115 to 816134, EPI_ISL_817123 to 817134, EPI_ISL_817140 to 817141, EPI_ISL_817144, EPI_ISL_817150 to 817158, EPI_ISL_817177 to 817183, EPI_ISL_817185 to 817198, EPI_ISL_817200 to 817262, EPI_ISL_817266 to 817268, EPI_ISL_817274 to 817277, EPI_ISL_817280, EPI_ISL_817287, EPI_ISL_817289, EPI_ISL_817293 to 817295, EPI_ISL_817298, EPI_ISL_817300, EPI_ISL_817302, EPI_ISL_817304 to 817305, EPI_ISL_817307, EPI_ISL_817309 to 817311, EPI_ISL_817313 to 817314, EPI_ISL_817318 to 817327, EPI_ISL_817329 to 817332, EPI_ISL_817336 to 817338, EPI_ISL_817340 to 817365, EPI_ISL_817367 to 817368, EPI_ISL_817370 to 817374, EPI_ISL_817379 to 817382, EPI_ISL_817387 to 817388, EPI_ISL_817390 to 817391, EPI_ISL_817394, EPI_ISL_817396 to 817397, EPI_ISL_817400, EPI_ISL_817402, EPI_ISL_817406 to 817412, EPI_ISL_817420 to 817486, EPI_ISL_817488 to 817495, EPI_ISL_817498, EPI_ISL_817500, EPI_ISL_817502 to 817507, EPI_ISL_817512 to 817529, EPI_ISL_817531, EPI_ISL_817533 to 817536, EPI_ISL_817539 to 817554, EPI_ISL_817653 to 817668, EPI_ISL_817670 to 817785, EPI_ISL_817787 to 818011, EPI_ISL_818013 to 818063, EPI_ISL_818065, EPI_ISL_818067 to 818076, EPI_ISL_818078 to 818122, EPI_ISL_843341, EPI_ISL_843360, EPI_ISL_843370, EPI_ISL_843384, EPI_ISL_843547, EPI_ISL_843574, EPI_ISL_843591, EPI_ISL_843595, EPI_ISL_843598, EPI_ISL_843608, EPI_ISL_843616 to 843617, EPI_ISL_843619 to 843622, EPI_ISL_843624 to 843646, EPI_ISL_843649 to 843671, EPI_ISL_843673 to 843688, EPI_ISL_843692 to 843698, EPI_ISL_843701 to 843702, EPI_ISL_843704 to 843763, EPI_ISL_843765 to 843776, EPI_ISL_843778 to 843788, EPI_ISL_843790 to 843795, EPI_ISL_843797 to 843802, EPI_ISL_843804 to 843815, EPI_ISL_843817 to 843900, EPI_ISL_843902 to 843911, EPI_ISL_843913 to 843924, EPI_ISL_843926 to 843943, EPI_ISL_843945 to 843966, EPI_ISL_843968 to 843972, EPI_ISL_843974 to 843993, EPI_ISL_843995 to 843999, EPI_ISL_844001 to 844007, EPI_ISL_844009 to 844018, EPI_ISL_844020 to 844025, EPI_ISL_844027 to 844064, EPI_ISL_844066, EPI_ISL_844068 to 844107, EPI_ISL_844109, EPI_ISL_844111 to 844112, EPI_ISL_844114, EPI_ISL_844116 to 844119, EPI_ISL_844121 to 844128, EPI_ISL_844130 to 844184, EPI_ISL_844186 to 844200, EPI_ISL_844202 to 844243, EPI_ISL_844245 to 844256, EPI_ISL_844258 to 844263, EPI_ISL_844265 to 844295, EPI_ISL_844297 to 844498, EPI_ISL_844501 to 844527, EPI_ISL_844529 to 844542, EPI_ISL_846818 to 846820, EPI_ISL_846851 to 846853, EPI_ISL_846861, EPI_ISL_846866, EPI_ISL_846894 to 846895, EPI_ISL_846913, EPI_ISL_847168 to 847179, EPI_ISL_847205, EPI_ISL_847256, EPI_ISL_847274, EPI_ISL_847320, EPI_ISL_847348 to 847349, EPI_ISL_847368 to 847370, EPI_ISL_847510 to 847514

| | | | |
|---|---|---|---|
| see above | Department of Virus and Microbiological Special Diagnostics, Statens Serum Institut, Copenhagen, Denmark | Albertsen Lab, Department of Chemistry and Bioscience, Aalborg University, Denmark | Danish Covid-19 Genome Consortium |

EPI_ISL_429333 to 429337, EPI_ISL_429339 to 429346, EPI_ISL_429348, EPI_ISL_429350 to 429358, EPI_ISL_429360 to 429362, EPI_ISL_429364 to 429379, EPI_ISL_429381 to 429399, EPI_ISL_429401 to 429429, EPI_ISL_429431 to 429454, EPI_ISL_429456, EPI_ISL_429458, EPI_ISL_429460 to 429466, EPI_ISL_429468 to 429469, EPI_ISL_429471 to 429474, EPI_ISL_429476 to 429486, EPI_ISL_429488 to 429499, EPI_ISL_429501 to 429503, EPI_ISL_429505 to 429521, EPI_ISL_429524 to 429530, EPI_ISL_429533 to 429575, EPI_ISL_429577 to 429590, EPI_ISL_436962 to 436978, EPI_ISL_436980 to 436985, EPI_ISL_436987 to 436991, EPI_ISL_436993, EPI_ISL_436995 to 437004, EPI_ISL_437006 to 437009, EPI_ISL_437011 to 437042, EPI_ISL_437627 to 437640, EPI_ISL_437642, EPI_ISL_437644 to 437647, EPI_ISL_437650, EPI_ISL_437652 to 437663, EPI_ISL_437665 to 437672, EPI_ISL_437674, EPI_ISL_437676 to 437679, EPI_ISL_437681 to 437683, EPI_ISL_444817 to 444829, EPI_ISL_444831 to 444841, EPI_ISL_444843 to 444850, EPI_ISL_444852 to 444864, EPI_ISL_444866 to 444867, EPI_ISL_444869 to 444870, EPI_ISL_444872 to 444880, EPI_ISL_444883 to 444888, EPI_ISL_444890 to 444906, EPI_ISL_444908 to 444913, EPI_ISL_444915 to 444935, EPI_ISL_444937 to 444942, EPI_ISL_444944 to 444950, EPI_ISL_444952, EPI_ISL_444954 to 444955, EPI_ISL_444957 to 444961, EPI_ISL_444963, EPI_ISL_444965 to 444968, EPI_ISL_452101 to 452102

| | | | |
|---|---|---|---|
| see above | Department of Virus and Microbiological Special Diagnostics, Statens Serum Institut, Copenhagen, Denmark, Artillerivej 5, 2300 Copenahgen S | Albertsen lab, Department of Chemistry and Bioscience, Aalborg University, Denmark | Rasmus Kirkegaard |

EPI_ISL_613541 to 613543, EPI_ISL_614403, EPI_ISL_614406 to 614408, EPI_ISL_614410 to 614416, EPI_ISL_614418 to 614420, EPI_ISL_614422 to 614453, EPI_ISL_614455 to 614479, EPI_ISL_614481 to 614487, EPI_ISL_614489 to 614517, EPI_ISL_614519 to 614520, EPI_ISL_614522 to 614526, EPI_ISL_614528 to 614534, EPI_ISL_614536 to 614549, EPI_ISL_614551 to 614567, EPI_ISL_614569 to 614573, EPI_ISL_614575 to 614636, EPI_ISL_614638 to 614645, EPI_ISL_614647 to 614654, EPI_ISL_614656 to 614661, EPI_ISL_614663 to 614725, EPI_ISL_614727 to 614751, EPI_ISL_614753 to 614759, EPI_ISL_614761 to 614762, EPI_ISL_614764, EPI_ISL_614766 to 614775, EPI_ISL_614777 to 614780, EPI_ISL_614783 to 614801, EPI_ISL_614803 to 614864, EPI_ISL_614866 to 614885, EPI_ISL_614887 to 614888, EPI_ISL_615166 to 615167, EPI_ISL_615169 to 615229, EPI_ISL_615231 to 615233, EPI_ISL_615235 to 615236, EPI_ISL_615239 to 615264, EPI_ISL_615266, EPI_ISL_615268 to 615270, EPI_ISL_615272 to 615275, EPI_ISL_615277 to 615280, EPI_ISL_615282 to 615286, EPI_ISL_615288 to 615303, EPI_ISL_615305 to 615325, EPI_ISL_615327 to 615331, EPI_ISL_615333, EPI_ISL_615336, EPI_ISL_615338 to 615343, EPI_ISL_615345 to 615354, EPI_ISL_615356 to 615364, EPI_ISL_615366 to 615397, EPI_ISL_615399 to 615412, EPI_ISL_615414 to 615416, EPI_ISL_615418 to 615422, EPI_ISL_615424 to 615461, EPI_ISL_615463 to 615475, EPI_ISL_615477 to 615478, EPI_ISL_615480 to 615499, EPI_ISL_615501 to 615505, EPI_ISL_615507 to 615510, EPI_ISL_615512 to 615514, EPI_ISL_615516 to 615520, EPI_ISL_615522 to 615534, EPI_ISL_615536 to 615547, EPI_ISL_615549, EPI_ISL_615551 to 615647, EPI_ISL_615649 to 615661, EPI_ISL_615663 to 615664, EPI_ISL_615666 to 615677, EPI_ISL_615679 to 615687, EPI_ISL_615689 to 615712, EPI_ISL_615714 to 615738, EPI_ISL_615740 to 615747, EPI_ISL_615749 to 615753, EPI_ISL_615755 to 615777, EPI_ISL_615779 to 615802, EPI_ISL_615804 to 615810, EPI_ISL_615812 to 615822, EPI_ISL_615824 to 615838, EPI_ISL_615840 to 615871, EPI_ISL_615873 to 615876, EPI_ISL_615879 to 615886, EPI_ISL_615888 to 615902, EPI_ISL_615904 to 615914, EPI_ISL_615916 to 615922, EPI_ISL_615924 to 615929, EPI_ISL_615931 to 615935, EPI_ISL_615937 to 615942, EPI_ISL_615944 to 615989, EPI_ISL_615991 to 616051, EPI_ISL_616054 to 616057, EPI_ISL_616059, EPI_ISL_616061 to 616065, EPI_ISL_616067 to 616070, EPI_ISL_616072 to 616078, EPI_ISL_616080 to 616100, EPI_ISL_616102 to 616117, EPI_ISL_616119 to 616128, EPI_ISL_616131 to 616141, EPI_ISL_616143 to 616175, EPI_ISL_616177 to 616187, EPI_ISL_616189 to 616199, EPI_ISL_616201 to 616210, EPI_ISL_616212 to 616283, EPI_ISL_616285 to 616289, EPI_ISL_616291 to 616303, EPI_ISL_616305 to 616349, EPI_ISL_616351 to 616371, EPI_ISL_616373 to 616403, EPI_ISL_616405 to 616410, EPI_ISL_616412 to 616435, EPI_ISL_616437 to 616479, EPI_ISL_616481 to 616521, EPI_ISL_616523 to 616536, EPI_ISL_616538 to 616593, EPI_ISL_616595 to 616604, EPI_ISL_616606 to 616608, EPI_ISL_616610 to 616641, EPI_ISL_616643 to 616646, EPI_ISL_616648 to 616657, EPI_ISL_616659 to 616666, EPI_ISL_616668 to 616687, EPI_ISL_616689 to 616705, EPI_ISL_616707 to 616711, EPI_ISL_616713 to 616729, EPI_ISL_616731, EPI_ISL_616733 to 616734, EPI_ISL_616736 to 616737, EPI_ISL_616739 to 616775, EPI_ISL_616777 to 616787, EPI_ISL_616789 to 616852, EPI_ISL_616854 to 616884, EPI_ISL_616886 to 616903, EPI_ISL_616905 to 616909, EPI_ISL_616911 to 616917, EPI_ISL_616919, EPI_ISL_616921, EPI_ISL_616923, EPI_ISL_616925 to 616928, EPI_ISL_616930 to 616932, EPI_ISL_616934 to 616948, EPI_ISL_616950 to 616956, EPI_ISL_616958 to 616965, EPI_ISL_616968 to 616973, EPI_ISL_616976 to 617002, EPI_ISL_617004 to 617029, EPI_ISL_617031 to 617057, EPI_ISL_617059 to 617097, EPI_ISL_617099 to 617100, EPI_ISL_617102 to 617103, EPI_ISL_617105 to 617106, EPI_ISL_617108 to 617112, EPI_ISL_617114 to 617116, EPI_ISL_617118 to 617133, EPI_ISL_617135 to 617142, EPI_ISL_617144 to 617147, EPI_ISL_617151 to 617158, EPI_ISL_617160 to 617164, EPI_ISL_617166 to 617173, EPI_ISL_617175 to 617187, EPI_ISL_617189 to 617209, EPI_ISL_617211 to 617227, EPI_ISL_617229 to 617235, EPI_ISL_617237 to 617241, EPI_ISL_617243 to 617246, EPI_ISL_617249 to 617266, EPI_ISL_617268 to 617269, EPI_ISL_617271 to 617275, EPI_ISL_617277 to 617290, EPI_ISL_617292 to 617294, EPI_ISL_617296 to 617297, EPI_ISL_617300, EPI_ISL_617302, EPI_ISL_617304 to 617312, EPI_ISL_617315 to 617317, EPI_ISL_617319 to 617322, EPI_ISL_617325 to 617326, EPI_ISL_617328 to 617330, EPI_ISL_617332 to 617334, EPI_ISL_617336 to 617348, EPI_ISL_617350 to 617352, EPI_ISL_617354 to 617359, EPI_ISL_617361 to 617366, EPI_ISL_617368 to 617391, EPI_ISL_617393 to 617403, EPI_ISL_617405 to 617435, EPI_ISL_617437 to 617443, EPI_ISL_617446, EPI_ISL_617448 to 617456, EPI_ISL_617458 to 617473, EPI_ISL_617475 to 617501, EPI_ISL_617503 to 617516, EPI_ISL_617518 to 617563, EPI_ISL_617565 to 617577, EPI_ISL_617579 to 617596, EPI_ISL_617598, EPI_ISL_617600 to 617610, EPI_ISL_617612 to 617629, EPI_ISL_617631 to 617636, EPI_ISL_617638 to 617646, EPI_ISL_617648 to 617655, EPI_ISL_617657 to 617683, EPI_ISL_617685 to 617691, EPI_ISL_617693 to 617696, EPI_ISL_617698 to 617714, EPI_ISL_617716 to 617736, EPI_ISL_617738 to 617740, EPI_ISL_617742 to 617877, EPI_ISL_617881 to 617906, EPI_ISL_617908 to 617937, EPI_ISL_617939 to 617941, EPI_ISL_617944 to 617958, EPI_ISL_617960 to 617964, EPI_ISL_617966 to 617974, EPI_ISL_617976 to 617977, EPI_ISL_617979 to 618006, EPI_ISL_618009 to 618049, EPI_ISL_618051 to 618060, EPI_ISL_618062 to 618072, EPI_ISL_618074 to 618125, EPI_ISL_618128 to 618136, EPI_ISL_618139 to 618140, EPI_ISL_618142 to 618167, EPI_ISL_618169 to 618172, EPI_ISL_618174 to 618265, EPI_ISL_618267 to 618279, EPI_ISL_618281 to 618291, EPI_ISL_618294 to 618374, EPI_ISL_618376 to 618387, EPI_ISL_618389 to 618418, EPI_ISL_618420 to 618429, EPI_ISL_618431, EPI_ISL_618433 to 618448, EPI_ISL_618450 to 618451, EPI_ISL_618453 to 618465, EPI_ISL_618467 to 618507, EPI_ISL_618509 to 618519, EPI_ISL_618521 to 618561, EPI_ISL_618563 to 618576, EPI_ISL_618578 to 618585, EPI_ISL_618588 to 618591, EPI_ISL_618593 to 618596, EPI_ISL_618598 to 618600, EPI_ISL_618602 to 618603, EPI_ISL_618605 to 618620, EPI_ISL_618622, EPI_ISL_618624 to 618627, EPI_ISL_618629, EPI_ISL_618631 to 618635, EPI_ISL_618637 to 618645, EPI_ISL_618647 to 618657, EPI_ISL_618659, EPI_ISL_618661 to 618664, EPI_ISL_618666 to 618693, EPI_ISL_618695 to 618698, EPI_ISL_618700 to 618714, EPI_ISL_618716 to 618740, EPI_ISL_618742, EPI_ISL_618744 to 618761, EPI_ISL_618763, EPI_ISL_618766 to 618771, EPI_ISL_618773, EPI_ISL_618775 to 618778, EPI_ISL_618780 to 618781, EPI_ISL_618783, EPI_ISL_618785 to 618796, EPI_ISL_618798 to 618799, EPI_ISL_618801 to 618820, EPI_ISL_618822 to 618825, EPI_ISL_618827 to 618832, EPI_ISL_618834 to 618842, EPI_ISL_618845, EPI_ISL_618847 to 618858, EPI_ISL_618860 to 618871, EPI_ISL_618873 to 618875, EPI_ISL_618877 to 618880, EPI_ISL_618882 to 618887, EPI_ISL_618889, EPI_ISL_618891 to 618910, EPI_ISL_618912 to 618936, EPI_ISL_618938 to 618965, EPI_ISL_618968, EPI_ISL_618970 to 618971, EPI_ISL_618973 to 619015, EPI_ISL_619017 to 619025, EPI_ISL_619027 to 619040, EPI_ISL_619042 to 619056, EPI_ISL_619058 to 619059, EPI_ISL_619063 to 619117, EPI_ISL_619121 to 619134, EPI_ISL_619137 to 619157, EPI_ISL_619159 to 619160, EPI_ISL_619162 to 619164, EPI_ISL_619166 to 619171, EPI_ISL_619173 to 619186, EPI_ISL_619188 to 619190, EPI_ISL_619192 to 619194, EPI_ISL_619196 to 619199, EPI_ISL_619201 to 619212, EPI_ISL_619214 to 619222, EPI_ISL_619224 to 619231, EPI_ISL_619233 to 619252, EPI_ISL_619254 to 619282, EPI_ISL_619284 to 619293, EPI_ISL_619295, EPI_ISL_619297, EPI_ISL_619299 to 619326, EPI_ISL_619328 to 619337, EPI_ISL_619339 to 619349, EPI_ISL_619351 to 619355, EPI_ISL_619357 to 619371, EPI_ISL_619374 to 619394, EPI_ISL_619396 to 619428, EPI_ISL_619430, EPI_ISL_619432 to 619438, EPI_ISL_619440 to 619458, EPI_ISL_619460 to 619463, EPI_ISL_619465, EPI_ISL_619467 to 619506, EPI_ISL_619510 to 619511, EPI_ISL_619513 to 619514, EPI_ISL_619516 to 619518, EPI_ISL_619520 to 619554, EPI_ISL_619556 to 619557, EPI_ISL_619559 to 619560, EPI_ISL_619564 to 619573, EPI_ISL_619577 to 619582, EPI_ISL_619584 to 619588, EPI_ISL_619591 to 619599, EPI_ISL_619601, EPI_ISL_619603 to 619626, EPI_ISL_619630 to 619632, EPI_ISL_619634 to 619661, EPI_ISL_619663 to 619702, EPI_ISL_619704 to 619705, EPI_ISL_619707 to 619708, EPI_ISL_619710 to 619718, EPI_ISL_619720 to 619735, EPI_ISL_619737 to 619750, EPI_ISL_619752 to 619763, EPI_ISL_619765 to 619786, EPI_ISL_619788 to 619797, EPI_ISL_619799 to 619825, EPI_ISL_619827 to 619849, EPI_ISL_619851 to 619865, EPI_ISL_619867 to 619889, EPI_ISL_619891, EPI_ISL_619893 to 619903, EPI_ISL_619905 to 619922, EPI_ISL_619924 to 619928, EPI_ISL_619930 to 619942, EPI_ISL_619944 to 619946, EPI_ISL_619948 to 619953, EPI_ISL_619955, EPI_ISL_619957 to 619963, EPI_ISL_619965 to 619973, EPI_ISL_619976 to 619983, EPI_ISL_619985 to 619987, EPI_ISL_619989 to 619997, EPI_ISL_619999 to 620032, EPI_ISL_620035 to 620055, EPI_ISL_620058, EPI_ISL_620060 to 620088, EPI_ISL_620090 to 620123, EPI_ISL_620125 to 620136, EPI_ISL_620138 to 620142, EPI_ISL_620144 to 620161, EPI_ISL_620163 to 620177, EPI_ISL_620179, EPI_ISL_620181 to 620218, EPI_ISL_620221 to 620268, EPI_ISL_620270 to 620303, EPI_ISL_620305 to 620336, EPI_ISL_620338 to 620351, EPI_ISL_620353 to 620365, EPI_ISL_620367 to 620374, EPI_ISL_620376 to 620377, EPI_ISL_620379 to 620397, EPI_ISL_620401 to 620405, EPI_ISL_620762 to 620781, EPI_ISL_620783 to 620803, EPI_ISL_620805 to 620812, EPI_ISL_620814 to 620827, EPI_ISL_620829 to 620857, EPI_ISL_620859 to 620864, EPI_ISL_620866 to 620879, EPI_ISL_620881 to 620887, EPI_ISL_620889, EPI_ISL_620891 to 620894, EPI_ISL_620896 to 620912, EPI_ISL_620914 to 620932, EPI_ISL_620934 to 620935, EPI_ISL_620937 to 620960, EPI_ISL_620962 to 620969, EPI_ISL_620971 to 621003, EPI_ISL_621005 to 621008, EPI_ISL_621011 to 621019, EPI_ISL_621021 to 621042, EPI_ISL_621044 to 621052, EPI_ISL_621056, EPI_ISL_621058, EPI_ISL_621060 to 621063, EPI_ISL_621066 to 621068, EPI_ISL_621072 to 621080, EPI_ISL_621082 to 621085, EPI_ISL_621087, EPI_ISL_621090 to 621096, EPI_ISL_621098, EPI_ISL_621100 to 621104, EPI_ISL_621106, EPI_ISL_621108, EPI_ISL_621110 to 621111, EPI_ISL_621116, EPI_ISL_621119, EPI_ISL_621124 to 621130, EPI_ISL_621132, EPI_ISL_621134, EPI_ISL_621137 to 621151, EPI_ISL_621153 to 621159, EPI_ISL_621161 to 621164, EPI_ISL_621167, EPI_ISL_621169 to 621171, EPI_ISL_621173, EPI_ISL_621175, EPI_ISL_621179 to 621180, EPI_ISL_621182 to 621188, EPI_ISL_621190 to 621232, EPI_ISL_621234 to 621251, EPI_ISL_621254 to 621273, EPI_ISL_621275 to 621282, EPI_ISL_621285 to 621289, EPI_ISL_621291 to 621310, EPI_ISL_621312 to 621318, EPI_ISL_621321 to 621324, EPI_ISL_621326 to 621347, EPI_ISL_621349 to 621355, EPI_ISL_621357 to 621361, EPI_ISL_621363 to 621378, EPI_ISL_621380 to 621411, EPI_ISL_621413 to 621437, EPI_ISL_621439 to 621449, EPI_ISL_621451 to 621453, EPI_ISL_621456 to 621479, EPI_ISL_621481 to 621494, EPI_ISL_621496 to 621498, EPI_ISL_621500 to 621513, EPI_ISL_621515 to 621526, EPI_ISL_621528 to 621542, EPI_ISL_621544 to 621560, EPI_ISL_621562 to 621564, EPI_ISL_621566, EPI_ISL_621568, EPI_ISL_621570 to 621583, EPI_ISL_621585 to 621589, EPI_ISL_621591 to 621596, EPI_ISL_621598, EPI_ISL_621600 to 621602, EPI_ISL_621604 to 621610, EPI_ISL_621613 to 621621, EPI_ISL_621623 to 621632, EPI_ISL_621634 to 621635, EPI_ISL_621637 to 621642, EPI_ISL_621644 to 621685, EPI_ISL_621687, EPI_ISL_621689 to 621693, EPI_ISL_621695 to 621700, EPI_ISL_621703 to 621708, EPI_ISL_621710 to 621746, EPI_ISL_621748 to 621761, EPI_ISL_621763 to 621771, EPI_ISL_621773 to 621798, EPI_ISL_621800 to 621802, EPI_ISL_621804 to 621823, EPI_ISL_621825 to 621848, EPI_ISL_621851 to 621853, EPI_ISL_621855 to 621860, EPI_ISL_621862 to 621897, EPI_ISL_621899 to 621937, EPI_ISL_621939 to 621946, EPI_ISL_621948 to 621951, EPI_ISL_621953 to 621957, EPI_ISL_621959 to 621966, EPI_ISL_621968 to 621973, EPI_ISL_621975 to 622008, EPI_ISL_622010 to 622031, EPI_ISL_622033 to 622038, EPI_ISL_622040 to 622066, EPI_ISL_622068 to 622080, EPI_ISL_622082 to 622097, EPI_ISL_622099 to 622100, EPI_ISL_622102 to 622123, EPI_ISL_622125 to 622127, EPI_ISL_622129 to 622130, EPI_ISL_622132 to 622140, EPI_ISL_622142, EPI_ISL_622144 to 622197, EPI_ISL_622199 to 622202, EPI_ISL_622204 to 622205, EPI_ISL_622207 to 622213, EPI_ISL_622215 to 622227, EPI_ISL_622229 to 622248, EPI_ISL_622250 to 622265, EPI_ISL_622270 to 622276, EPI_ISL_622278, EPI_ISL_622280 to 622308, EPI_ISL_622310 to 622333, EPI_ISL_622335 to 622402, EPI_ISL_622404, EPI_ISL_622406 to 622411, EPI_ISL_622413 to 622442, EPI_ISL_622444 to 622448, EPI_ISL_622450 to 622457, EPI_ISL_622459 to 622465, EPI_ISL_622467 to 622481, EPI_ISL_622483 to 622489, EPI_ISL_622491 to 622495, EPI_ISL_622497, EPI_ISL_622499 to 622511, EPI_ISL_622513 to 622517, EPI_ISL_622519 to 622524, EPI_ISL_622526 to 622534, EPI_ISL_622536 to 622556, EPI_ISL_622558 to 622631, EPI_ISL_622633 to 622641, EPI_ISL_622643 to 622667, EPI_ISL_622669 to 622730, EPI_ISL_622732 to 622737, EPI_ISL_622739 to 622760, EPI_ISL_625684, EPI_ISL_625686 to 625696, EPI_ISL_625698 to 625700, EPI_ISL_625702 to 625722, EPI_ISL_625724 to 625740, EPI_ISL_625743, EPI_ISL_625746, EPI_ISL_625748 to 625781, EPI_ISL_625784 to 625835, EPI_ISL_625837 to 625857, EPI_ISL_625859 to 625872, EPI_ISL_625874, EPI_ISL_625876 to 625884, EPI_ISL_625887 to 625888, EPI_ISL_625890 to 625902, EPI_ISL_625904 to 625910, EPI_ISL_625912 to 625917, EPI_ISL_625919 to 625922, EPI_ISL_625924 to 625932, EPI_ISL_625934 to 625941, EPI_ISL_625944 to 625959, EPI_ISL_625961 to 625965, EPI_ISL_625967 to 625971, EPI_ISL_625974 to 625975, EPI_ISL_625977 to 625978, EPI_ISL_625980 to 625986, EPI_ISL_625988 to 625995, EPI_ISL_625997, EPI_ISL_625999 to 626006, EPI_ISL_626008 to 626009, EPI_ISL_626011 to 626014, EPI_ISL_626016 to 626017, EPI_ISL_626020 to 626023, EPI_ISL_626025 to 626026, EPI_ISL_626028 to 626031, EPI_ISL_626033 to 626035, EPI_ISL_626037 to 626042, EPI_ISL_626045, EPI_ISL_626047 to 626051, EPI_ISL_626053, EPI_ISL_626055 to 626057, EPI_ISL_626060, EPI_ISL_626062 to 626064, EPI_ISL_626066 to 626068, EPI_ISL_626070 to 626077, EPI_ISL_626080 to 626082, EPI_ISL_626084 to 626085, EPI_ISL_626087 to 626091, EPI_ISL_626093, EPI_ISL_626095 to 626101, EPI_ISL_626103 to 626105, EPI_ISL_626108 to 626118, EPI_ISL_626120 to 626124, EPI_ISL_626126 to 626130, EPI_ISL_626132 to 626136, EPI_ISL_626138 to 626153, EPI_ISL_626157 to 626161, EPI_ISL_626164 to 626175, EPI_ISL_626177 to 626181, EPI_ISL_626183 to 626184, EPI_ISL_626187 to 626191, EPI_ISL_626193 to 626195, EPI_ISL_626197, EPI_ISL_626199, EPI_ISL_626201 to 626206, EPI_ISL_626208 to 626210

| | | | |
|---|---|---|---|
| see above | Department of Virus and Microbiological Special Diagnostics, Statens Serum Institut, Denmark | Albertsen lab, Department of Chemistry and Bioscience, Aalborg University, Denmark | Danish Corona Genome Consortia; Danish Covid-19 Genome Consortia |

| | | | |
|---|---|---|---|
| EPI_ISL_415647, EPI_ISL_416140 to 416142 | Department of Virus and Microbiological Special diagnostics, Statens Serum Institut, Copenhagen, Denmark. | Statens Serum Institute | Anders Fomsgaard; Maiken Worsoe Rosenstierne; Morten Rasmussen |
| EPI_ISL_415646, EPI_ISL_415648, EPI_ISL_416143 to 416144, EPI_ISL_416153 | Department of Virus and Microbiological Special diagnostics, Statens Serum Institut, Copenhagen, Denmark. | ViFU | Anders Fomsgaard; Maiken Worsoe Rosenstierne; Morten Rasmussen |
| EPI_ISL_775346 to 775347 | Department of medical microbiology, section Aalesund, Aalesund Hospital | Norwegian Institute of Public Health, Department of Virology | Atiya R Ali; Hilde Elshaug; Hilde Vollan; Kamilla Heddeland Instefjord; Karoline Bragstad; Kathrine Stene-Johansen; Marie Paulsen Madsen; Olav Hungnes; Rasmus Riis Kopperud |
| EPI_ISL_496482 | Dept. Infectious, Tropical Diseases & Microbiology, IRCCS Sacro Cuore Don Calabria Hospital | 1) Dept. Infectious, Tropical Diseases & Microbiology, IRCCS Sacro Cuore Don Calabria Hospital; 2) Centro Piattaforme Tecnologiche, University of Verona; 3) Dept. Neurosciences, Biomedicine and Movement Sciences, University of Verona. | 1) Antonio Mori; 2) Monica Castellucci and Francesca Griggio; 3) Giovanni Malerba; Chiara Piubelli; Elena Pomari; Michela Deiana |
| EPI_ISL_449789 to 449794, EPI_ISL_590877, EPI_ISL_590953 to 590977, EPI_ISL_591021, EPI_ISL_635120, EPI_ISL_635144, EPI_ISL_635146, EPI_ISL_635166, EPI_ISL_635196, EPI_ISL_668407 to 668408, EPI_ISL_708044 to 708045, EPI_ISL_708081, EPI_ISL_708131 to 708135, EPI_ISL_759988, EPI_ISL_775272, EPI_ISL_775280, EPI_ISL_775314, EPI_ISL_775327 to 775329, EPI_ISL_775353 to 775355, EPI_ISL_775386, EPI_ISL_796658, EPI_ISL_796663, EPI_ISL_796677, EPI_ISL_796698, EPI_ISL_860289 | | | |
| see above | Dept. of Medical Microbiology, Stavanger University Hospital, Helse Stavanger HF | Norwegian Institute of Public Health, Department of Virology | Atiya R Ali; Hilde Elshaug; Hilde Vollan; Iren Löhr; Kamilla Heddeland Instefjord; Karoline Bragstad; Kathrine Stene-Johansen; Marie Paulsen Madsen; Olav Hungnes; Rasmus Riis Kopperud |
| EPI_ISL_447837 | Dept. of Medical Microbiology, Stavanger University Hospital, Helse Stavanger HF, | Norwegian Institute of Public Health, Department of Virology | Hilde Elshaug; Kamilla Heddeland Instefjord; Karoline Bragstad; Kathrine Stene-Johansen; Olav Hungnes; Rasmus Riis Kopperud |
| EPI_ISL_664101, EPI_ISL_664103, EPI_ISL_664105 to 664106, EPI_ISL_665254 to 665257, EPI_ISL_665259 to 665260, EPI_ISL_666568, EPI_ISL_666593 to 666608, EPI_ISL_666610 to 666612, EPI_ISL_666614 to 666615 | | | |
| see above | Dept. of Microbiology and Infection Control, Akershus University Hospital HF | Dept. of Microbiology and Infection Control, Akershus University Hospital HF | Alexander Hesselberg Løvestad; Hege Vangstein Aamot; Nina Handal; Ole Herman Ambur; Silje Bakken Jørgensen |
| EPI_ISL_807155 | Deva County Emergency Hospital | National Institute of Infectious Diseases-Prof. Dr. Matei Bals Molecular Diagnostics Laboratory | Andreea Tudor; Corina Casangiu; Dan Otelea; Leontina Banica; Marius Surleac; Petre Milu; Simona Paraschiv |
| EPI_ISL_437197 to 437203, EPI_ISL_437298 to 437303, EPI_ISL_653916 | Diagnostic- and Research Institute of Pathology, Medical University of Graz | Diagnostic- and Research Institute of Pathology, Medical University of Graz | Gregor Gorkiewicz; Karl Kashofer; Martin Zacharias; Peter Regitnig |
| EPI_ISL_413573 | Dienst Gezondheid & Jeugd Zuid-Holland Zuid | Erasmus Medical Center | Anne van der Linden; Annemiek van der Eijk; Aura Timen; Bas Oude Munnink; Claudia Schapendonk; Corien Swaan; Corine GeurtsvanKessel; David Nieuwenhuijse; Irina Chestakova; Jeroen van Kampen; Jolanda Voermans; Madelief Mollers; Manon Haverkate; Marion Koopmans; Mark Pronk; Mart Stein; Pascal Lexmond; Reina Sikkema; Richard Molenkamp; Sandra Kengne Kamga Mobou; on behalf of the Dutch national COVID-19 response team. |
| EPI_ISL_751196 to 751200 | Dienststelle Gesundheit und Sport Kanton Luzern | Institute of Medical Virology, University of Zurich | Alexandra Trkola; Claudia Schmutz; Eva Spieler; Gabriela Ziltener; Jürg Böni; Kevin Steiner; Maryam Zaheri; Michael Huber; Stefan Schmutz; Verena Kufner |
| EPI_ISL_452234, EPI_ISL_475512, EPI_ISL_475562 to 475563 | Din Klinik | The Public Health Agency of Sweden | Anna Risberg; Anna-Malin Linde; Helene Warnborg; Karin Tegmark-Wisell; Maria Lind Karlberg; Mattias Haukland; Mia Brytting; Olov Svartstrom; Oskar Karlsson Lindsjo; Reza Advani; Sandra Broddesson; Theresa Enkirch |
| EPI_ISL_591326 to 591338, EPI_ISL_591340 | Dipartimento di Biotecnologie Mediche, University of Siena | Dipartimento di Biotecnologie Mediche, University of Siena | Anichini, G.; COVID; Cusi; Gandolfo, C.; M.G.; Pinzauti, D.; Pozzi, G.; Santoro, F. |
| EPI_ISL_722855 to 722858, EPI_ISL_722872, EPI_ISL_722896 to 722897, EPI_ISL_722899, EPI_ISL_722912 | Dipartimento di Scienze Biomediche e Oncologia Umana - Azienda Ospedaliero Universitaria Consorziale Policlinico | Istituto Zooprofilattico Sperimentale della Puglia e della Basilicata | Bianco A.; Capozzi L.; Chironna M.; Del Sambro L.; Loconsole D.; Parisi A. |
| EPI_ISL_738139, EPI_ISL_738141 to 738142, EPI_ISL_747370, EPI_ISL_747421, EPI_ISL_760096 to 760097, EPI_ISL_760129, EPI_ISL_760132, EPI_ISL_760135, EPI_ISL_760137 to 760153, EPI_ISL_760156 to 760157, EPI_ISL_760189 to 760190, EPI_ISL_760202 to 760205, EPI_ISL_760214, EPI_ISL_760225, EPI_ISL_850235, EPI_ISL_850237, EPI_ISL_850253, EPI_ISL_850261, EPI_ISL_850277, EPI_ISL_850283 to 850287, EPI_ISL_850291 to 850292, EPI_ISL_850294, EPI_ISL_850305, EPI_ISL_850314 to 850315, EPI_ISL_850319, EPI_ISL_850347, EPI_ISL_850356, EPI_ISL_850653 to 850662 | | | |
| see above | Division of Emerging Infectious Diseases, Bureau of Infectious Diseases Diagnosis Control, Korea Disease Control and Prevention Agency | Division of Emerging Infectious Diseases, Bureau of Infectious Diseases Diagnosis Control, Korea Disease Control and Prevention Agency | Ae Kyung Park; Chaeyoung Lee; Eun-Jin Kim; Heui Man Kim; Il-Hwan Kim; Jeong-Min Kim; Namjoo Lee; Sang Hee Woo |
| EPI_ISL_485603 | Division of Infectious Disease | Steininger Lab | Christoph Steininger; Ingeborg Klymiuk; Jakob Thannesberger; Lorenz Schubert; Nicolas Rascovan; Oliver Robak |
| EPI_ISL_508864 to 508870, EPI_ISL_509222 | Division of Infectious Diseases and Hospital Epidemiology, University Hospital Zürich | Institute of Medical Virology, University of Zurich | Aline Wolfensberger; Dana Weissberg; Hugo Sax; Irene A. Abela; Jürg Böni; Maryam Zaheri; Michael Huber; Peter W. Schreiber; Silvana K. Rampini; Verena Kufner |
| EPI_ISL_413022 to 413024 | Division of Infectious Diseases, University Hospital Zurich | Institute of Medical Virology, University of Zurich | Alexandra Trkola; Andrea Zbinden; Fiona Steiner; Gabriela Ziltener; Jon Huder; Jürg Böni; Maryam Zaheri; Michael Huber; Patrick Redli; Riccarda Capaul; Roberto Speck; Stefan Schmutz; Verena Kufner |
| EPI_ISL_524475, EPI_ISL_524477, EPI_ISL_524483, EPI_ISL_524485 | Division of Infectious Diseases, University Hospital Zürich | Institute of Medical Virology, University of Zurich | Alexandra Trkola; Andrea Zbinden; Fiona Steiner; Gabriela Ziltener; Jon Huder; Jürg Böni; Maryam Zaheri; Michael Huber; Patrick Redli; Riccarda Capaul; Stefan Schmutz; Verena Kufner |
| EPI_ISL_722209 | Dom Zdravlja Sarajevo | Alea Genetic Center | Konjhodzic R.; Pecar D.; Salihefendic L. |
| EPI_ISL_766589 to 766590 | Dr. Boubaker Karim laboratory | Institute of Medical Virology, University of Zurich | Alexandra Trkola; Annette Audigé; Cyril Shah; Jon Huder; Jürg Böni; Kevin Steiner; Maria Grünberg; Maryam Zaheri; Michael Huber; Riccarda Capaul; Stefan Schmutz; Verena Kufner |
| EPI_ISL_583727 to 583733, EPI_ISL_583735 to 583763, EPI_ISL_583765 to 583771, EPI_ISL_583773 to 583814, EPI_ISL_583816 to 583831, EPI_ISL_583834 to 583852 | Dr. Gernot Walder GmbH | Bergthaler laboratory, CeMM Research Center for Molecular Medicine of the Austrian Academy of Sciences | Adi Steinrigl; Alexander Lercher; Alexandra Popa; Andreas Bergthaler; Benedikt Agerer; Christian Paar; Christoph Bock; Daniela Schmid; Dorothee von Laer; Elisabeth Puchhammer-Stoeckl; Franz Allerberger; Gernot Walder; Gregor Hörmann; Guenter Weiss; Gunther Vogl; Henrique Colaco; Jakob-Wendelin Genger; Jan Laine; Judith Aberle; Kinga Rigler-Hohenwarter; Lukas Endler; Manfred Nairz; Mark Smyth; Martin Senekowitsch; Michael Schuster; Peter Hufnagl; Peter Obrist; Rainer Gattringer; Sabine Sussitz-Rack; Stephan Aberle; Thomas Penz; Wegene Borena |
| EPI_ISL_414423 to 414446, EPI_ISL_414448 to 414471, EPI_ISL_414529 to 414545, EPI_ISL_414547 to 414566, EPI_ISL_415460 to 415463, EPI_ISL_415465 to 415492, EPI_ISL_415495 to 415498, EPI_ISL_415500 to 415506, EPI_ISL_415508, EPI_ISL_415510 to 415516, EPI_ISL_415524 to 415535, EPI_ISL_422566 to 422574, EPI_ISL_422576 to 422589, EPI_ISL_422591 to 422612, EPI_ISL_422615 to 422635, EPI_ISL_422637 to 422881, EPI_ISL_422885 to 422899, EPI_ISL_422901 to 422960, EPI_ISL_423034, EPI_ISL_455112 to 455149, EPI_ISL_455151 to 455180, EPI_ISL_455195 to 455200, EPI_ISL_455202 to 455222, EPI_ISL_455226 to 455239, EPI_ISL_455241 to 455242, EPI_ISL_455244 to 455258, EPI_ISL_455261, EPI_ISL_455263 to 455307, EPI_ISL_460636 to 460678, EPI_ISL_460680 to 460705, EPI_ISL_460714 to 460721, EPI_ISL_460723 to 460738, EPI_ISL_460753 to 460764, EPI_ISL_460766, EPI_ISL_460768, EPI_ISL_460770, EPI_ISL_460773 to 460821, EPI_ISL_460823 to 460854, EPI_ISL_460856, EPI_ISL_460858 to 460890, EPI_ISL_460895, EPI_ISL_460898, EPI_ISL_460901 to 460915, EPI_ISL_460917 to 460956, EPI_ISL_460958 to 460990, EPI_ISL_460992 to 461001, EPI_ISL_461003 to 461048, EPI_ISL_461050, EPI_ISL_461061 to 461080, EPI_ISL_461082 to 461087, EPI_ISL_461101 to 461106, EPI_ISL_461108 to 461111, EPI_ISL_461113 to 461118, EPI_ISL_461120 to 461123, EPI_ISL_461125 to 461159, EPI_ISL_461161, EPI_ISL_461163 to 461167, EPI_ISL_461172, EPI_ISL_461177, EPI_ISL_461179, EPI_ISL_461181 to 461189, EPI_ISL_461193, EPI_ISL_461195 to 461199, EPI_ISL_461201 to 461202, EPI_ISL_461204 to 461224, EPI_ISL_461226 to 461232, EPI_ISL_461234 to 461247, EPI_ISL_461249 to 461261, EPI_ISL_461263 to 461278, EPI_ISL_461280 to 461289, EPI_ISL_461291 to 461335, EPI_ISL_461337 to 461371, EPI_ISL_461373, EPI_ISL_461375 to 461381, EPI_ISL_461383 to 461391, EPI_ISL_461394, EPI_ISL_461396 to 461398, EPI_ISL_523121 to 523135, EPI_ISL_523137 to 523164, EPI_ISL_523166 to 523200, EPI_ISL_523202 to 523211, EPI_ISL_523235 to 523241, EPI_ISL_523243 to 523252, EPI_ISL_523254 to 523265, EPI_ISL_523267 to 523273, EPI_ISL_523275 to 523291, EPI_ISL_523301 to 523309, EPI_ISL_523332, EPI_ISL_523334 to 523385, EPI_ISL_523401 to 523403, EPI_ISL_523405 to 523420, EPI_ISL_523422 to 523480, EPI_ISL_523483 to 523491, EPI_ISL_523493 to 523495, EPI_ISL_523497 to 523499, EPI_ISL_523501 to 523509, EPI_ISL_523511 to 523517, EPI_ISL_523523 to 523526, EPI_ISL_523528 to 523558, EPI_ISL_523568 to 523578, EPI_ISL_523595 to 523611, EPI_ISL_523613 to 523614, EPI_ISL_523616 to 523641, EPI_ISL_523643 to 523657, EPI_ISL_523659 to 523809, EPI_ISL_574779 to 574781, EPI_ISL_574786 to 574789, EPI_ISL_577743 to 577744, EPI_ISL_577746 to 577747, EPI_ISL_577829 to 577835, EPI_ISL_577837 to 577844, EPI_ISL_577847 to 577850, EPI_ISL_577853 to 577866, EPI_ISL_577873 to 577877, EPI_ISL_577879 to 577882, EPI_ISL_577886 to 577905, EPI_ISL_577909 to 577916, EPI_ISL_577926 to 577927, EPI_ISL_577929 to 577931, EPI_ISL_577933 to 577935, EPI_ISL_577937 to 577944, EPI_ISL_577948 to 577954, EPI_ISL_577964 to 577967, EPI_ISL_577969 to 577985, EPI_ISL_577989 to 578000, EPI_ISL_578004 to 578009, EPI_ISL_578011, EPI_ISL_578013 to 578030, EPI_ISL_578033 to 578048, EPI_ISL_578050 to 578051, EPI_ISL_578053 to 578054, EPI_ISL_578057 to 578071, EPI_ISL_578073 to 578079, EPI_ISL_632323, EPI_ISL_632325 to 632327, EPI_ISL_632330 to 632332, EPI_ISL_632335, EPI_ISL_632337, EPI_ISL_632339 to 632340, EPI_ISL_632342 to 632347, EPI_ISL_632349 to 632350, EPI_ISL_632352, EPI_ISL_632354 to 632356, EPI_ISL_632359 to 632364, EPI_ISL_632367, EPI_ISL_632369 to 632377, EPI_ISL_632379, EPI_ISL_632381, EPI_ISL_632383, EPI_ISL_632386 to 632391, EPI_ISL_632394, EPI_ISL_632400 to 632401, EPI_ISL_632406 to 632407, EPI_ISL_632410, EPI_ISL_632413, EPI_ISL_632415, EPI_ISL_632418 to 632419, EPI_ISL_632421 to 632422, EPI_ISL_632426, EPI_ISL_632440 to 632449, EPI_ISL_632460 to 632461, EPI_ISL_632480 to 632482, EPI_ISL_632493 to 632649, EPI_ISL_632651 to 632774, EPI_ISL_632776 to 632795, EPI_ISL_722273, EPI_ISL_722277, EPI_ISL_722279 to 722289, EPI_ISL_722291, EPI_ISL_722293 to 722299, EPI_ISL_722302 to 722304, EPI_ISL_722306 to 722312, EPI_ISL_722314, EPI_ISL_722316 to 722319, EPI_ISL_722321 to 722324, EPI_ISL_722326 to 722329, EPI_ISL_722331 to 722335, EPI_ISL_722337 to 722339, EPI_ISL_722341 to 722354, EPI_ISL_722356 to 722357, EPI_ISL_722359 to 722372, EPI_ISL_722374 to 722376, EPI_ISL_722378 to 722379, EPI_ISL_722381, EPI_ISL_722383 to 722387, EPI_ISL_722389 to 722402, EPI_ISL_722404 to 722405, EPI_ISL_722407 to 722411, EPI_ISL_722413, EPI_ISL_722419 to 722447, EPI_ISL_722452 to 722453, EPI_ISL_722455 to 722465, EPI_ISL_722493 to 722560, EPI_ISL_722562 to 722605, EPI_ISL_722607 to 722850, EPI_ISL_734166, EPI_ISL_763099 to 763103, EPI_ISL_763109 to 763116, EPI_ISL_763118 to 763122, EPI_ISL_763124 to 763127, EPI_ISL_763130 to 763137, EPI_ISL_763139 to 763153, EPI_ISL_763155 to 763172, EPI_ISL_763174 to 763181, EPI_ISL_763188 to 763189, EPI_ISL_763191 to 763221, EPI_ISL_763223 to 763231, EPI_ISL_763233, EPI_ISL_763235 to 763239, EPI_ISL_763241, EPI_ISL_763243 to 763244, EPI_ISL_763246 to 763259, EPI_ISL_763261 to 763272, EPI_ISL_763274 to 763280, EPI_ISL_763284 to 763289, EPI_ISL_763291 to 763292, EPI_ISL_763294 to 763307, EPI_ISL_763309 to 763314, EPI_ISL_763348 to 763349, EPI_ISL_763353 to 763359, EPI_ISL_801404 to 801409, EPI_ISL_801411 to 801420, EPI_ISL_801422, EPI_ISL_801425 to 801513, EPI_ISL_801515, EPI_ISL_802506 to 802509, EPI_ISL_802512 to 802514, EPI_ISL_802516 to 802533, EPI_ISL_802536 to 802544 | | | |
| see above | Dutch COVID-19 response team | Erasmus Medical Center | Anne van der Linden; Annemiek van der Eijk; Aura Timen; Bas Oude Munnink; Claudia Schapendonk; Corien Swaan; Corine GeurtsvanKessel; David Nieuwenhuijse; Emmanuelle Munger; Irina Chestakova; Jeroen van Kampen; Jolanda Voermans; Madelief Mollers; Manon Haverkate; Marion Koopmans; |

| | | | |
|---|---|---|---|
| | | | Marjan Boter; Mark Pronk; Mart Stein; Pascal Lexmond; Reina Sikkema; Richard Molenkamp; Sandra Kengne Kamga Mobou; Stefan van Nieuwkoop; Theo Bestebroer; on behalf of the Dutch national COVID-19 response team. |
| EPI_ISL_454750 to 454767, EPI_ISL_454769 to 454794, EPI_ISL_547455 to 547458, EPI_ISL_547460 to 547465, EPI_ISL_547469 to 547470, EPI_ISL_547473 to 547486, EPI_ISL_547491 to 547499, EPI_ISL_547501 to 547509, EPI_ISL_547511, EPI_ISL_547513 to 547522, EPI_ISL_547524 to 547529, EPI_ISL_547531 to 547534, EPI_ISL_547536 to 547539, EPI_ISL_547541, EPI_ISL_547557 to 547560, EPI_ISL_636491, EPI_ISL_636494 to 636504, EPI_ISL_636506 to 636507, EPI_ISL_636511, EPI_ISL_636525 to 636526, EPI_ISL_636528 to 636544, EPI_ISL_636546 to 636561, EPI_ISL_636564 to 636585, EPI_ISL_636587 to 636591, EPI_ISL_636593, EPI_ISL_636598, EPI_ISL_636600 to 636603, EPI_ISL_723155 to 723467, EPI_ISL_728564 to 728763, EPI_ISL_747521 to 747525, EPI_ISL_790570 to 790577, EPI_ISL_790579 to 790596, EPI_ISL_790598 to 790623, EPI_ISL_790627 to 791083, EPI_ISL_804379 to 804462, EPI_ISL_823976 to 823988, EPI_ISL_823990 to 824095, EPI_ISL_824101 to 824140, EPI_ISL_824142 to 824144, EPI_ISL_824151 to 824152, EPI_ISL_824155 to 824168, EPI_ISL_824170 to 824208, EPI_ISL_824210 to 824252, EPI_ISL_824255 to 824260, EPI_ISL_824265 to 824266, EPI_ISL_824269, EPI_ISL_824271 to 824278, EPI_ISL_824281 to 824283, EPI_ISL_826523 to 826536 | | | |
| see above | Dutch COVID-19 response team | National Institute for Public Health and the Environment (RIVM) | Adam Meijer; AnneMarie van den Brandt; Bas van der Veer; Chantal Reusken; Dennis Schmitz; Dirk Eggink; Florian Zwagemaker; Harry Vennema; Jeroen Cremer; Matthijs Welkers; Pieter Overduin; Sharon van den Brink; on behalf of the national COVID-19 response team |
| EPI_ISL_419691 | E. Gulbja Laboratorija | Charite Universitatsmedizin Berlin, Institute of Virology | Barbara Mühlemann; Christian Drosten; Dmitrijs Perminovs; Dr. Didzis Gavars; Jorn Beheim-Schwarzbach; Julia Schneider; Mikus Gavars; Talitha Veith; Terry Jones; Victor M Corman |
| EPI_ISL_421653 to 421656, EPI_ISL_426285 to 426289, EPI_ISL_450518 to 450519, EPI_ISL_492993 to 493000, EPI_ISL_501275, EPI_ISL_501284 to 501285, EPI_ISL_501823, EPI_ISL_501829, EPI_ISL_501833, EPI_ISL_501839, EPI_ISL_501849, EPI_ISL_501894, EPI_ISL_501929, EPI_ISL_501936, EPI_ISL_512313 to 512314, EPI_ISL_512645 to 512646, EPI_ISL_515187 to 515195, EPI_ISL_534199, EPI_ISL_534203, EPI_ISL_534212 to 534216, EPI_ISL_534218 to 534219, EPI_ISL_534221, EPI_ISL_639631, EPI_ISL_639635, EPI_ISL_639637 to 639638, EPI_ISL_639641, EPI_ISL_639643 to 639647, EPI_ISL_639649, EPI_ISL_639652 to 639653, EPI_ISL_639658 to 639662, EPI_ISL_639664 to 639671, EPI_ISL_639673 to 639675, EPI_ISL_639681 to 639682, EPI_ISL_639684 to 639694, EPI_ISL_770033 to 770037, EPI_ISL_770040 to 770041, EPI_ISL_770043 to 770044, EPI_ISL_770046, EPI_ISL_770050 to 770051, EPI_ISL_770053, EPI_ISL_770055, EPI_ISL_770057, EPI_ISL_770062 | | | |
| see above | E. Gulbja Laboratorija | Latvian Biomedical Research and Study Centre | Dmitrijs Perminovs; Ivars Silamielis; Jnis Kloviš; Jnis Pjalkovskis; Kaspars Megnis; Mikus Gavars; Monta Ustinova; Uga Dumpis; Vita Rovte; ikita Zrelovs |
| EPI_ISL_486438 | E. Gulbja laboratorija | Latvian Biomedical Research and Study Centre | Dmitrijs Perminovs; Ivars Silamielis; Jnis Kloviš; Kaspars Megnis; Mikus Gavars; Monta Ustinova; Uga Dumpis; Vita Rovte; ikita Zrelovs |
| EPI_ISL_418226 | EHPAD - Résidences les Cèdres | National Reference Center for Viruses of Respiratory Infections, Institut Pasteur, Paris | Angela Brisebarre; Etienne Simon-Lorière; Flora Donati; Marion Barbet; Maud Vanpeene; Mélanie Albert; Méline Bizard; Sylvie Behillil; Sylvie van der Werf; Vincent Enouf |
| EPI_ISL_434652, EPI_ISL_710610 to 710611 | Ektorps Vardcentral | The Public Health Agency of Sweden | Anna Risberg; Anna-Malin Linde; Department of Microbiology; Eva Espmark; Karin Tegmark-Wisell; Maria Lind Karlberg; Mia Brytting; Olov Svartstrom; Oskar Karlsson Lindsjo; The Public Health Agency of Sweden; Theresa Enkirch |
| EPI_ISL_486854 | Emergency County Hospital Suceava | Stefan cel Mare, University Metagenomics lab | Lobiuc Andrei et al. |
| EPI_ISL_486855 | Emergency county Hospital Suceava | "Stefan cel Mare" University Metagenomics Lab | Lobiuc Andrei et al. |
| EPI_ISL_413582 | ErasmusMC | Erasmus Medical Center | Anne van der Linden; Annemiek van der Eijk; Aura Timen; Bas Oude Munnink; Claudia Schapendonk; Corien Swaan; Corine GeurtsvanKessel; David Nieuwenhuijse; Irina Chestakova; Jeroen van Kampen; Jolanda Voermans; Madelief Mollers; Manon Haverkate; Marion Koopmans; Mark Pronk; Mart Stein; Pascal Lexmond; Reina Sikkema; Richard Molenkamp; Sandra Kengne Kamga Mobou; on behalf of the Dutch national COVID-19 response team. |
| EPI_ISL_648166 to 648168 | Eskilstuna | The Public Health Agency of Sweden | Anna Risberg; Anna-Malin Linde; Karin Tegmark-Wisell; Maria Lind Karlberg; Mattias Haukland; Mia Brytting; Olov Svartstrom; Oskar Karlsson Lindsjo; Petra Edquist; Reza Advani; Sandra Broddesson |
| EPI_ISL_636970 | Etlik Veterinary Control Central Research Institute | Etlik Veterinary Control Central Research Institute | Ahu Pakdemirli; Cevdet Yarali; Dilek Dulger; Erdem Danyer; Ozcan Yildirim; Sabri Hacioglu; Ummu Sena Sari |
| EPI_ISL_614265 to 614280 | Eurofins | National Reference Center for Viruses of Respiratory Infections, Institut Pasteur, Paris | Angela Brisebarre; Camille Capel; Etienne Simon-Lorière; Marion Barbet; Maud Vanpeene; Méline Bizard; Sylvie Behillil; Sylvie van der Werf; Vincent Enouf |
| EPI_ISL_602282 to 602303 | Evangelisches Klinikum Bethel, Institut für Laboratoriumsmedizin, Mikrobiologie und Hygiene | Bielefeld University | Alexander Sczyrba; Christiane Scherer; David Brandt; Jörn Kalinowski; Levin-Joe Klages; Marina Simunovic; Markus Haak; Svenja Vinke; Tobias Busche |
| EPI_ISL_833046, EPI_ISL_833068 to 833075 | FPAM | Istituto Zooprofilattico Sperimentale dell'Abruzzo e Molise "G. Caporale" | Ancora M; Calistri P; Cammà C; Curini V; Delli Compagni E; Di Domenico M; Di Pasquale A; Lorusso A; Mangone I; Marcacci M; Puglia I; Rinaldi A; Savini G |
| EPI_ISL_428851 to 428852 | FSBSI "Chumakov Federal Scientific Center for Research and Development of Immune-and-Biological Products of Russian Academy of Sciences" | FSBSI "Chumakov Federal Scientific Center for Research and Development of Immune-and-Biological Products of Russian Academy of Sciences" & NRC "Kurchatov institute" | Anastasia Berestovskaya; Anastasia Piniaeva; Anna Shishova; Aydar Ishmukhametov; Denis Protsenko; Egor Prokhorchuk; Georgy Ignatyev; Liubov Kozlovskaya; Mikhail Rychev |
| EPI_ISL_417010, EPI_ISL_418252 | FUNDACION JIMENEZ DIAZ | Instituto de Salud Carlos III | A. Monzón; F. Casas; I. Fernández Roblas, R.; I. Jiménez; Iglesias-Caballero; M. Camarero; M. Cuesta; M. González-Esguevillas; M. Molinero Calamita; M. Zaballos; P. Jiménez; S. Juliá; S. Pozo; S. Varona |
| EPI_ISL_510533 to 510534 | Faculty of Medicine, Bursa Uludag University | Bursa Uludag University | Alemdar, A.; S.G.; Saglik, I.; Temel; Özemri Sa, . |
| EPI_ISL_526937 to 526941, EPI_ISL_526943 to 526944, EPI_ISL_526946 to 526948, EPI_ISL_614298, EPI_ISL_614301 to 614302, EPI_ISL_614304, EPI_ISL_614307 to 614308, EPI_ISL_614310 to 614313, EPI_ISL_614316 | | | |
| see above | Faroese National Reference Laboratory for Fish and Animal Diseases | Faroese National Reference Laboratory for Fish and Animal Diseases | Debes Hammershaimb Christiansen; Maria Marjunardóttir Dahl; Petra Elisabeth Petersen |
| EPI_ISL_451963 to 451970 | Federal Budget Institution of Science, State Research Center for Applied Microbiology & Biotechnology | Federal Budget Institution of Science, State Research Center for Applied Microbiology & Biotechnology | Abaimova A; Bakhteeva I; Blagodatskikh S; Bogun A; Borzilov A; Chekan L; Chernysh S; Denisenko E; Dentovskaya S; Detushev K; Detusheva E; Dyatlov I; Firstova V; Frolov V; Fursov M; Fursova N; Galkina E; Gapelchenkova T; Goncharova J; Gorbatov A; Hlyntseva A; Ivanov S; Kalmantayev T; Kalmantayeva O; Kanashenko M; Kartsev N; Kartseva A; Khomyakov A; Khramov M; Kislichkina A; Kolchanova A; Koroleva-Ushakova A; Kosilova I; Krasilnikova E; Kuzin V; Kuzina E; Makarova M; Marin M; Novikova T; Platonov M; Podkopaev Y; Ryabko A; Shaikhutdinova R; Shemyakin I; Shishkina L; Silkina M; Sizova A; Skryabin Y; Slukin P; Slukina N; Solomentsev V; Solovieva A; Teymurazov M; Timofeev V; Titareva G; Trunyakova A; Tyurin E; Vagayskaya A; Zeninskaya N; Zhumakaev R |
| EPI_ISL_470882, EPI_ISL_500776 to 500778, EPI_ISL_590883 to 590884, EPI_ISL_590899, EPI_ISL_590981, EPI_ISL_590985, EPI_ISL_591008, EPI_ISL_635063 to 635065, EPI_ISL_635098, EPI_ISL_635127, EPI_ISL_635167, EPI_ISL_635170, EPI_ISL_635173, EPI_ISL_635176 to 635177, EPI_ISL_708073 to 708078, EPI_ISL_708176 to 708179, EPI_ISL_738313, EPI_ISL_775323 to 775326, EPI_ISL_775333, EPI_ISL_775362 to 775370 | | | |
| see above | Foerde Hospital, Department of Microbiology | Norwegian Institute of Public Health, Department of Virology | Atiya R Ali; Hilde Elshaug; Hilde Vollan; Kamilla Heddeland Instefjord; Karoline Bragstad; Kathrine Stene-Johansen; Marie Paulsen Madsen; Olav Hungnes; Rasmus Riis Kopperud |
| EPI_ISL_455903, EPI_ISL_475152, EPI_ISL_476139 | Folkhalsomyndigheten | The Public Health Agency of Sweden | Anna Risberg; Anna-Malin Linde; Karin Tegmark-Wisell; Maria Lind Karlberg; Mattias Haukland; Olov Svartstrom; Oskar Karlsson Lindsjo; Petra Edquist; Reza Advani; Sandra Broddesson; Shamam Muradrasoli |
| EPI_ISL_434650, EPI_ISL_475536, EPI_ISL_475541 | Follinge Halsocentral | The Public Health Agency of Sweden | Anna Risberg; Anna-Malin Linde; Karin Tegmark-Wisell; Kerstin Persson Moberg; Maria Lind Karlberg; Mattias Haukland; Mia Brytting; Olov Svartstrom; Oskar Karlsson Lindsjo; Reza Advani; Sandra Broddesson; Theresa Enkirch |
| EPI_ISL_420145 | Forde Hospital Department of Microbiology | Norwegian Institute of Public Health, Department of Virology | Hilde Elshaug; Kamilla Heddeland Instefjord; Karoline Bragstad; Kathrine Stene-Johansen; Olav Hungnes |
| EPI_ISL_413586 to 413587 | Foundation Elisabeth-Tweesteden Ziekenhuis | Erasmus Medical Center | Anne van der Linden; Annemiek van der Eijk; Aura Timen; Bas Oude Munnink; Claudia Schapendonk; Corien Swaan; Corine GeurtsvanKessel; David Nieuwenhuijse; Irina Chestakova; Jeroen van Kampen; Jolanda Voermans; Madelief Mollers; Manon Haverkate; Marion Koopmans; Mark Pronk; Mart Stein; Pascal Lexmond; Reina Sikkema; Richard Molenkamp; Sandra Kengne Kamga Mobou; on behalf of the Dutch national COVID-19 response team. |
| EPI_ISL_413565 | Foundation Pamm | Erasmus Medical Center | Anne van der Linden; Annemiek van der Eijk; Aura Timen; Bas Oude Munnink; Claudia Schapendonk; Corien Swaan; Corine GeurtsvanKessel; David Nieuwenhuijse; Irina Chestakova; Jeroen van Kampen; Jolanda Voermans; Madelief Mollers; Manon Haverkate; Marion Koopmans; Mark Pronk; Mart Stein; Pascal Lexmond; Reina Sikkema; Richard Molenkamp; Sandra Kengne Kamga Mobou; on behalf of the Dutch national COVID-19 response team. |
| EPI_ISL_419235 to 419237 | Fundacion Jimenez Diaz | Instituto de Salud Carlos III | Camarero, S.; Casas, I.; Cuesta, I.; Fernández, R.; González-Esguevillas, M.; Iglesias-Caballero, M.; Jiménez, M.; Jiménez, P.; Juliá, M.; Molinero Calamita, M.; Monzón, S.; Pozo, F.; Varona, S.; Zaballos, A. |
| EPI_ISL_462447 to 462449 | Fundació Lluita contra la SIDA (FLSida)/Hospital Universitari Germans Trias i Pujol | IrsiCaixa AIDS Research Lab | Bonaventura Clotet; Joaquim Segalés; Jorge Carrillo; Julia Blanco; Lidia Ruiz; Marc Corbacho; Marc Noguera-Julian; Maria Pilar Armengol; Maria Ubals; Mariona Parera; Nuria Izquierdo; Oriol Mitjà; Roger Paredes |
| EPI_ISL_462478 | Fundación Jiménez Díaz | Instituto de Salud Carlos III | A. Monzón; F. Casas; I; I. Jiménez; Iglesias-Caballero; M. Camarero; M. Cuesta; M. González-Esguevillas; M. Molinero Calamita; M. Zaballos; P. Jiménez; R. Fernández; S. Juliá; S. Pozo; S. Varona |

| | | | |
|---|---|---|---|
| EPI_ISL_420141, EPI_ISL_420146, EPI_ISL_420313, EPI_ISL_449787, EPI_ISL_493356 to 493380, EPI_ISL_500768 to 500774, EPI_ISL_500784 to 500790, EPI_ISL_549027 to 549028, EPI_ISL_549038, EPI_ISL_549048 to 549049, EPI_ISL_549056 to 549058, EPI_ISL_549060 to 549069, EPI_ISL_549071 to 549080, EPI_ISL_549082, EPI_ISL_549088, EPI_ISL_549112, EPI_ISL_549119 to 549122, EPI_ISL_549124, EPI_ISL_549132, EPI_ISL_549143, EPI_ISL_549154, EPI_ISL_549165, EPI_ISL_549168, EPI_ISL_590881, EPI_ISL_590898, EPI_ISL_590901 to 590903, EPI_ISL_590912, EPI_ISL_635086 to 635087, EPI_ISL_668396 to 668397, EPI_ISL_668403, EPI_ISL_708035, EPI_ISL_708057 to 708059, EPI_ISL_708126 to 708130, EPI_ISL_759973 to 759974, EPI_ISL_759984 to 759985, EPI_ISL_775271, EPI_ISL_775273 to 775274, EPI_ISL_775279, EPI_ISL_775294, EPI_ISL_775299 to 775300, EPI_ISL_775303 to 775304, EPI_ISL_775427, EPI_ISL_775484, EPI_ISL_775488 to 775490, EPI_ISL_775496 to 775497, EPI_ISL_775524 to 775527, EPI_ISL_796722 to 796724, EPI_ISL_796726, EPI_ISL_796730 to 796732, EPI_ISL_860213 | | | |
| see above | Furst Medical Laboratory | Norwegian Institute of Public Health, Department of Virology | Atiya R Ali; Hilde Elshaug; Hilde Synnøve Vollan; Hilde Vollan; Kamilla Heddeland Instefjord; Karoline Bragstad; Kathrine Stene-Johansen; Marie Paulsen Madsen; Olav Hungnes; Rasmus Riis Kopperud |
| EPI_ISL_522549 to 522550 | Félix Guyon Hospital | UMR PIMIT Université de La Réunion | Camille Lebarbenchon; David Wilkinson; Patrick Mavingui |
| EPI_ISL_418416, EPI_ISL_508878, EPI_ISL_509003 to 509004 | GH Les Portes du Sud | CNR Virus des Infections Respiratoires - France SUD | Alexandre Gaymard; Antonin Bal; Bruno Lina; Carine Moustaud; Florence Morfin-Sherpa; Gregory Destras; Gwendolyne Burfin; Laurence Josset; Martine Valette; Maude Bouscambert-Duchamp; Raphaelle Lamy; Solenne Brun |
| EPI_ISL_428351 to 428352, EPI_ISL_428363 | GH Nord Essonne Service de Biologie clinique | National Reference Center for Viruses of Respiratory Infections, Institut Pasteur, Paris | Angela Brisebarre; Etienne Simon-Lorière; Flora Donati; Marion Barbet; Maud Vanpeene; Mélanie Albert; Méline Bizard; Sylvie Behillil; Sylvie van der Werf; Vincent Enouf |
| EPI_ISL_825054 | GMERS Medical College and Hospital, Gotri, Vadodara | Gujarat Biotechnology Research Centre | Afzal Ansari; Apurvasinh Puvar; Bithika Duttaroy; Chaitanya Joshi; Dinesh Kumar; Janvi Raval; Kalpesh Mistry; Madhvi Joshi; Mitesh Kamothi; Nikha Trivedi; Nitin Savaliya; Ramesh Pandit; Zarna Patel; Zuber Saiyed |
| EPI_ISL_534249 to 534251 | Gavle Sjukhus | The Public Health Agency of Sweden | Anna Risberg; Anna-Malin Linde; Karin Tegmark-Wisell; Maria Lind Karlberg; Mattias Haukland; Mia Brytting; Olov Svartstrom; Oskar Karlsson Lindsjo; Petra Edquist; Reza Advani; Sandra Broddesson |
| EPI_ISL_475110, EPI_ISL_475115, EPI_ISL_582801, EPI_ISL_582809, EPI_ISL_582834, EPI_ISL_615101 to 615103, EPI_ISL_623095, EPI_ISL_648140 to 648142, EPI_ISL_648144, EPI_ISL_648163, EPI_ISL_661283, EPI_ISL_661286 to 661287, EPI_ISL_710608 to 710609, EPI_ISL_831977 to 831978 | | | |
| see above | Gavle klinisk mikrobiologi | The Public Health Agency of Sweden | Anna Risberg; Anna-Malin Linde; Department of Microbiology; Karin Tegmark-Wisell; Maria Lind Karlberg; Mattias Haukland; Mia Brytting; Olov Svartstrom; Oskar Karlsson Lindsjo; Petra Edquist; Reza Advani; Sandra Broddesson; Shamam Muradrasoli; The Public Health Agency of Sweden |
| EPI_ISL_676514 | Gavleborg | The Public Health Agency of Sweden | Department of Microbiology; The Public Health Agency of Sweden |
| EPI_ISL_730567 to 730568, EPI_ISL_730570 to 730577, EPI_ISL_730620 | Gazi University Faculty of Medicine, Medical Virology Laboratory | Gazi University Faculty of Medicine, Medical Virology Laboratory | Erdem ahin; Gülendam Bozday; Hager Muftah; Il Fidan; Kayhan Çalar; Murat Dizbay; Selin Yiit; Shaknoza Sarzhanova; Özlem Güzel Tunçcan |
| EPI_ISL_467300 | General Hospital "Abdulah Nakas" | Alea Genetic Center | Damir Marjanovic; Dino Pecar; Enis Kandic; Lana Salihefendic; Nihad Fejzic; Rijad Konjhodzic; Sead Jazic; Teufik Goletic |
| EPI_ISL_516413, EPI_ISL_677705, EPI_ISL_677707, EPI_ISL_678252 | General Hospital - Kumanovo | Research Center for Genetic Engineering and Biotechnology "Georgi D. Efremov" , Macedonian Academy of Sciences and Arts | RCGEB - MASA |
| EPI_ISL_677704, EPI_ISL_677709, EPI_ISL_677711, EPI_ISL_677722, EPI_ISL_677725 to 677726, EPI_ISL_678255 to 678256 | General Hospital - Ohrid | Research Center for Genetic Engineering and Biotechnology "Georgi D. Efremov" , Macedonian Academy of Sciences and Arts | RCGEB - MASA |
| EPI_ISL_514354, EPI_ISL_677674, EPI_ISL_677703, EPI_ISL_677712 to 677714, EPI_ISL_678258 | General Hospital - Prilep | Research Center for Genetic Engineering and Biotechnology "Georgi D. Efremov" , Macedonian Academy of Sciences and Arts | RCGEB - MASA |
| EPI_ISL_677719, EPI_ISL_678260 | General Hospital - Struga | Research Center for Genetic Engineering and Biotechnology "Georgi D. Efremov" , Macedonian Academy of Sciences and Arts | RCGEB - MASA |
| EPI_ISL_678254 | General Hospital - Strumica | Research Center for Genetic Engineering and Biotechnology "Georgi D. Efremov" , Macedonian Academy of Sciences and Arts | RCGEB - MASA |
| EPI_ISL_677675, EPI_ISL_677710 | General Hospital - Veles | Research Center for Genetic Engineering and Biotechnology "Georgi D. Efremov" , Macedonian Academy of Sciences and Arts | RCGEB - MASA |
| EPI_ISL_430846 | General Intensive Care Unit, Raymond Poincaré Hospital (AP-HP), Lab Inflammation & Infection, U1173 University Paris Saclay-UVSQ/INSERM, Garches, France. | Institut Pasteur, Laboratory for Urgent Response to biological Threats | Annane Djillali; Caro Valérie; Manuguerra Jean-Claude; Vanhomwegen Jessica |
| EPI_ISL_614282 to 614293, EPI_ISL_623096 to 623097, EPI_ISL_623103, EPI_ISL_660324 to 660325, EPI_ISL_693766 to 693774, EPI_ISL_792042 to 792053 | General practitioner | National Reference Center for Viruses of Respiratory Infections, Institut Pasteur, Paris | ; Angela Brisebarre; Brieuc Lefaure; Camille Capel; Etienne Simon-Lorière; Marion Barbet; Maud Vanpeene; Méline Bizard; Sylvie Behillil; Sylvie van der Werf; Vincent Enouf |
| EPI_ISL_427391, EPI_ISL_428346, EPI_ISL_428368, EPI_ISL_480228 to 480292, EPI_ISL_735259 to 735279, EPI_ISL_735281 to 735373, EPI_ISL_735434 to 735435, EPI_ISL_833141 to 833146 | | | |
| see above | Genomic Laboratory (GLAB) (Conjoint lab of Health Directorate of Istanbul and Istanbul Technical University) | Genomic Laboratory (GLAB), Istanbul Technical University | Arzu Irvem; Ayse Serra Ozel; Betsi Kose; Betsi Köse; Bugra Agaoglu; Elifnaz Çelik; Gizem Alkurt; Gizem Dinler Doganay; Ilker Karacan; Jale Yildiz; Levent Doganay; Mehtap Aydin; Mehtap Aydn; Nihat Bugra Agaoglu; Nilsun Altunal; Nisan Denizce Can; Ozlem Akgun Dogan; Pari Sharifli; Payam Zolfagharian; Tugba Kizilboga Akgun; Yasemin Kendir Demirkol |
| EPI_ISL_882921 | Genomic Medicine Laboratory, IRCCS Santa Lucia Foundation | INMI Lazzaro Spallanzani IRCCS | A Di Caro; B Bartolini; C.E.M Gruber; E Giardina; E Giombini; F Messina; M Rueca; MR Capobianchi; O Butera |
| EPI_ISL_803899 | Genomic Medicine Laboratory, IRCCS Santa Lucia Foundation | National Institute for Infectious Diseases, INMI, "L. Spallanzani" IRCCS | A Di Caro; B Bartolini; C.E.M Gruber; E Giardina; E Giombini; F Messina; M. Rueca; MR Capobianchi; O Butera |
| EPI_ISL_436715 to 436717 | Genomics and Computational Biology Lab, Scientific Research Institute of Physical-Chemical Medicine, FMBA of Russia | Genomics and Computational Biology Lab, Scientific Research Institute of Physical-Chemical Medicine, FMBA of Russia | A. Manolov; A. Pavlenko; D. Fedorov; K. Klimina; O. Guskova; V. Govorun and E. Ilina; V. Veselovsky |
| EPI_ISL_539524, EPI_ISL_862556, EPI_ISL_862564 to 862565, EPI_ISL_862567, EPI_ISL_862640 | Gerencia de Asistencia Sanitaria de Soria | Instituto de Salud Carlos III | A. Monzón; C. Aldea; F. Casas; I; I. Aldea, C.; I. Jiménez; Iglesias-Caballero; M. Pozo; M. Camarero; M. Cuesta; M. González-Esguevillas; M. Molinero Calamita; M. Zaballos; M.Camarero; P. Jiménez; S. Juliá; S. Molinero Calamita; S. Pozo; S. Varona |
| EPI_ISL_539557 | Gerencia del área de salud de Badajoz, Llerena y Zafra | Instituto de Salud Carlos III | A. Monzón; C. Pazos; F. Casas; I; I. Jiménez; Iglesias-Caballero; M. Camarero; M. Cuesta; M. González-Esguevillas; M. Molinero Calamita; M. Zaballos; P. Jiménez; S. Juliá; S. Pozo; S. Varona |
| EPI_ISL_833260 to 833261 | Giulianova | Istituto Zooprofilattico Sperimentale dell'Abruzzo e Molise "G. Caporale" | Ancora M; Calistri P; Cammà C; Curini V; Di Domenico M; Di Pasquale A; Lorusso A; Mangone I; Marcacci M; Puglia I; Rinaldi A; Savini G |
| EPI_ISL_447608 to 447613, EPI_ISL_452219 to 452222 | Goethe University Hospital Frankfurt | Institute for Medical Virology, Goethe University Hospital Frankfurt | Annemarie Berger; Björn Rotter; Denisa Bojkova; Jindrich Cinatl; Klaus Hoffmeier; Sandra Westhaus; Sandra Ciesek; Sebastian Hoehl; Tuna Toptan; and Marek Widera |
| EPI_ISL_529146 | Goethe University Hospital Frankfurt, Institute for Medical Virology | Goethe University Hospital Frankfurt, Institute for Medical Virology | Berger, A.; Bojkova, D.; Ciesek, S.; Cinatl, J.; Grabmair; Hoehl, S.; Hoffmeier, K.; Rotter, B.; T.T.; Westhaus, S.; Widera, M. |
| EPI_ISL_415152 | Gorgas Memorial Institute for Health Studies | Gorgas Memorial Institute for Health Studies | Alexander A. Martinez.; Ambar Moreno; Claudia Gonzalez; Danilo Franco; Elimelec Valdespino; Juan M. Pascale; Leyda Abrego; Oris Chavarria; Sandra Lopez-Verges; Yamilka Diaz |
| EPI_ISL_794818 to 794820 | Greek Genome Center, Biomedical Research Foundation of | Greek Genome Center, Biomedical Research Foundation of | Christina Maria Kravvari; Dimitrios Thanos; Emmanouil Athanasiadis; Ioannis Vatsellas; Katerina Zoi; Thodoris Loupis |

| | | | |
|---|---|---|---|
| | the Academy of Athens (BRFAA) | the Academy of Athens (BRFAA) | |
| EPI_ISL_421454, EPI_ISL_421458, EPI_ISL_421481, EPI_ISL_421487, EPI_ISL_421491 | H Beatriz Angelo | Instituto Nacional de Saude (INSA) | Guiomar et al |
| EPI_ISL_418003, EPI_ISL_418007 to 418008, EPI_ISL_418019 to 418022 | H Braga | Instituto Nacional de Saude (INSA) | Guiomar et al |
| EPI_ISL_421457 | H Dr Nelio Mendonca - Funchal | Instituto Nacional de Saude (INSA) | Guiomar et al |
| EPI_ISL_418026, EPI_ISL_421449, EPI_ISL_421459 to 421461 | H Dr. Nelio Mendonca - Funchal | Instituto Nacional de Saude (INSA) | Guiomar et al |
| EPI_ISL_418023, EPI_ISL_421466 to 421467 | H Evora | Instituto Nacional de Saude (INSA) | Guiomar et al |
| EPI_ISL_418018 | H Garcia de Orta | Instituto Nacional de Saude (INSA) | Guiomar et al |
| EPI_ISL_421446 to 421448 | H Guimaraes | Instituto Nacional de Saude (INSA) | Guiomar et al |
| EPI_ISL_418025, EPI_ISL_421462, EPI_ISL_421468 to 421471, EPI_ISL_421488, EPI_ISL_421490, EPI_ISL_421492, EPI_ISL_421495 | | | |
| see above | H Santarem | Instituto Nacional de Saude (INSA) | Guiomar et al |
| EPI_ISL_596250 to 596263, EPI_ISL_596268 to 596317, EPI_ISL_596357 to 596377 | HELIX LCC | WHO National Influenza Centre Russian Federation | Andrey Komissarov; Anna Ivanova; Artem Fadeev; Daria Danilenko; Dmitry Bazhenov; Kseniya Komissarova |
| EPI_ISL_602329 to 602411, EPI_ISL_602417 to 602461, EPI_ISL_639948 to 639973, EPI_ISL_733023 to 733051, EPI_ISL_733053 to 733153, EPI_ISL_733301 to 733391, EPI_ISL_733393, EPI_ISL_733396 to 733401, EPI_ISL_733417 to 733458, EPI_ISL_872985 to 873026, EPI_ISL_873034 | | | |
| see above | HELIX LLC | WHO National Influenza Centre Russian Federation | Andrey Komissarov; Anna Ivanova; Artem Fadeev; Daria Danilenko; Dmitry Bazhenov; Dmitry Lioznov; Elena Nabieva; Georgii Bazykin; Ksenia Safina; Kseniya Komissarova; Mikhail Bakaev |
| EPI_ISL_860899 | HOPITAL PAUL BROUSSE - Service Microbiologie | National Reference Center for Viruses of Respiratory Infections, Institut Pasteur, Paris | Angela Brisebarre; Camille Capel; Etienne Simon-Lorière; Marion Barbet; Maud Vanpeene; Méline Bizard; Roque-Afonso Anne-Marie; Sylvie Behillil; Sylvie van der Werf; Vincent Enouf |
| EPI_ISL_419238 | HOSPITAL DE CRUCES. | Instituto de Salud Carlos III | A. Monzón; F. Casas; I. Aranzamendi, M.; I. Jiménez; Iglesias-Caballero; M. Camarero; M. Cuesta; M. González-Esguevillas; M. Molinero Calamita; M. Zaballos; P. Jiménez; S. Juliá; S. Pozo; S. Varona |
| EPI_ISL_418247 | HOSPITAL GENERAL DE SEGOVIA | Instituto de Salud Carlos III | A. Monzón; F. Casas; I. Hernando-Real S.; I. Jiménez; Iglesias-Caballero; M. Camarero; M. Cuesta; M. González-Esguevillas; M. Molinero Calamita; M. Zaballos; P. Jiménez; S. Juliá; S. Pozo; S. Varona |
| EPI_ISL_417007 | HOSPITAL SANTA MARIA NAI | Instituto de Salud Carlos III | A. Monzón; I. García Costa, J.; Iglesias-Caballero; M. Camarero S. Pozo F. Casas I. Jiménez; M. Cuesta; M. González-Esguevillas; M. Molinero Calamita; M. Zaballos; P. Jiménez; S. Juliá; S. Varona |
| EPI_ISL_418253, EPI_ISL_419240, EPI_ISL_419709 | HOSPITAL TXAGORRITXU | Instituto de Salud Carlos III | A. Monzón; C; F. Casas; I. Gomez-Gonzalez C.; I. Gómez; I. Gómez, C.; I. Jiménez; Iglesias-Caballero; M. Camarero; M. Camarero S. Pozo F. Casas I. Jiménez; M. Cuesta; M. González-Esguevillas; M. Molinero Calamita; M. Zaballos; P. Jiménez; S. Juliá; S. Pozo; S. Varona |
| EPI_ISL_418251 | HOSPITAL UNIVERSITARIO LA PAZ | Instituto de Salud Carlos III | A. Monzón; F. Casas; I. Jiménez; I. Romero P.; Iglesias-Caballero; M. Camarero; M. Cuesta; M. González-Esguevillas; M. Molinero Calamita; M. Zaballos; P. Jiménez; S. Juliá; S. Pozo; S. Varona |
| EPI_ISL_418243 to 418244 | HOSPITAL UNIVERSITARIO VIRGEN DE LAS NIEVES | Instituto de Salud Carlos III | A. Monzón; F. Casas; I. Jiménez; I. Sanbonmatsu S.; Iglesias-Caballero; M. Camarero; M. Cuesta; M. González-Esguevillas; M. Molinero Calamita; M. Zaballos; P. Jiménez; S. Juliá; S. Pozo; S. Varona |
| EPI_ISL_430847 | HS mikrobiologi virus | The Public Health Agency of Sweden | Anna Risberg; Anna-Malin Linde; Karin Tegmark-Wisell; Maria Lind Karlberg; Olov Svartstrom; Oskar Karlsson Lindsjo; Shaman Muradrasoli; Zhibing Yun |
| EPI_ISL_418009, EPI_ISL_421463, EPI_ISL_421489, EPI_ISL_421493 to 421494 | HSE Ilha Terceira - Angra do Heroismo | Instituto Nacional de Saude (INSA) | Guiomar et al |
| EPI_ISL_412971 | HUS Diagnostiikkakeskus, Hallinto | Department of Virology Faculty of Medicine, Medicum University of Helsinki | Hannimari Kallio-Kokko; Olli Vapalahti; Suvi Kuivanen; Teemu Smura |
| EPI_ISL_648151 | Halmstad | The Public Health Agency of Sweden | Anna Risberg; Anna-Malin Linde; Karin Tegmark-Wisell; Maria Lind Karlberg; Mattias Haukland; Mia Brytting; Olov Svartstrom; Oskar Karlsson Lindsjo; Petra Edquist; Reza Advani; Sandra Broddesson |
| EPI_ISL_454436 to 454446, EPI_ISL_455902, EPI_ISL_469073 to 469074, EPI_ISL_475093 to 475094, EPI_ISL_475096 to 475098, EPI_ISL_475100, EPI_ISL_475114, EPI_ISL_475116 to 475117, EPI_ISL_475120 to 475121, EPI_ISL_475123, EPI_ISL_475125, EPI_ISL_475156 to 475159, EPI_ISL_475164, EPI_ISL_475548, EPI_ISL_475553, EPI_ISL_475556 to 475557, EPI_ISL_510831, EPI_ISL_510866, EPI_ISL_548247, EPI_ISL_548249, EPI_ISL_582803 to 582805, EPI_ISL_594148, EPI_ISL_615104 to 615107, EPI_ISL_623086, EPI_ISL_648148 | | | |
| see above | Halmstad klinisk mikrobiologi | The Public Health Agency of Sweden | Anna Risberg; Anna-Malin Linde; Karin Tegmark-Wisell; Maria Lind Karlberg; Mattias Haukland; Mia Brytting; Olov Svartstrom; Oskar Karlsson Lindsjo; Petra Edquist; Reza Advani; Sandra Broddesson; Shamam Muradrasoli; Shaman Muradrasoli |
| EPI_ISL_710606 to 710607, EPI_ISL_766591 | Halsomedicinskt Center | The Public Health Agency of Sweden | Department of Microbiology; The Public Health Agency of Sweden |
| EPI_ISL_766028 | Hannover Medical School, Institute of Virology | Hannover Medical School, Institute of Virology | Lars Steinbrück |
| EPI_ISL_635080 to 635084, EPI_ISL_635089 to 635091, EPI_ISL_635133, EPI_ISL_635184 | Haukeland University Hospital, Department of Medical Microbiology | Norwegian Institute of Public Health, Department of Virology | Hilde Elshaug; Hilde Vollan; Kamilla Heddeland Instefjord; Karoline Bragstad; Kathrine Stene-Johansen; Marie Paulsen Madsen; Olav Hungnes; Rasmus Riis Kopperud |
| EPI_ISL_635128, EPI_ISL_668392, EPI_ISL_668405, EPI_ISL_759975, EPI_ISL_775276, EPI_ISL_775335 to 775336, EPI_ISL_775357, EPI_ISL_775407 to 775409, EPI_ISL_796652, EPI_ISL_860208, EPI_ISL_860211 | | | |
| see above | Haukeland University Hospital, Dept. of Microbiology | Norwegian Institute of Public Health, Department of Virology | Atiya R Ali; Hilde Elshaug; Hilde Vollan; Kamilla Heddeland Instefjord; Karoline Bragstad; Kathrine Stene-Johansen; Marie Paulsen Madsen; Olav Hungnes; Rasmus Riis Kopperud |
| EPI_ISL_450498 | Health Board Laboratory of Communicable Diseases | Charite Universitatsmedizin Berlin, Institute of Virology | Barbara Muhlemann; Christian Drosten; Julia Schneider; Jörn Beheim-Schwarzbach; Liidia Dotsenko; Natalja Kuznetsova; Talitha Veith; Terry Jones; Victor M Corman |
| EPI_ISL_420065 to 420066 | Health Board Laboratory of Communicable Diseases | Charite Universitatsmedizin Berlin, Institute of Virology | Barbara Mühlemann; Christian Drosten; Jorn Beheim-Schwarzbach; Julia Schneider; Liidia Dotsenko; Natalja Kuznetsova; Talitha Veith; Terry Jones; Victor M Corman |
| EPI_ISL_429256 | Health Sciences Technology Park, Avicena, 8, 18016 Granada. Spain | Sequencing and Bioinformatics Service FISABIO-Public Health | Almudena Rojas; Joaquín Mendoza; Pablo Mendoza |
| EPI_ISL_451310 to 451311, EPI_ISL_487370, EPI_ISL_487377, EPI_ISL_487379, EPI_ISL_487381, EPI_ISL_501231 to 501238, EPI_ISL_501241 to 501242, EPI_ISL_501248 to 501251 | | | |
| see above | Hellenic Pasteur Institute, National Influenza Reference laboratory of Southern Greece & Unit of Bioinformatics and Applied Genomics | Hellenic Pasteur Institute, National Influenza Reference laboratory of Southern Greece & Unit of Bioinformatics and Applied Genomics | Andreas Mentis; Androniki Voulgari-Kokota; Antonios Kalliaropoulos; Aspasia Kontou; Athanasios Kossyvakis; Evangelidou Maria; Horefti Elina; Timokratis Karamitros; Vasiliki Pogka |
| EPI_ISL_501230 | Hellenic Pasteur Institute, Public Health Laboratories | Hellenic Pasteur Institute, National Influenza Reference laboratory of Southern Greece & Unit of Bioinformatics and Applied Genomics | Andreas Mentis; Androniki Voulgari-Kokota; Antonios Kalliaropoulos; Aspasia Kontou; Athanasios Kossyvakis; Evangelidou Maria; Horefti Elina; Timokratis Karamitros; Vasiliki Pogka |
| EPI_ISL_430469 | Hellenic Pasteur Institute, Public Health Laboratories | Hellenic Pasteur Institute, Public Health Laboratories, Unit of Bioinformatics and Applied Genomics | Andreas Mentis; Androniki Voulgari-Kokota; Antonios Kalliaropoulos; Aspasia Kontou; Athanasios Kossyvakis; Evangelidou Maria; Horefti Elina; Timokratis Karamitros; Vasiliki Pogka |
| EPI_ISL_428232 to 428236 | Hematology Laboratory, Section of Molecular Diagnostics, University Clinical Centre, Medical University of Gdansk | Department of Virology, Faculty of Medicine, University of Helsinki, Helsinki, Finland | Aneta Szulc; Maciej Grzybek; Marlena Robakowska; Olli Vapalahti; Teemu Smura |

| | | | |
|---|---|---|---|
| EPI_ISL_450525 to 450530, EPI_ISL_512296 to 512303 | Hematology Laboratory, Section of Molecular Diagnostics, University Clinical Centre, Medical University of Gdansk | Department of Virology, Faculty of Medicine, University of Helsinki, Helsinki, Finland | Aneta Szulc; Ewa Miosz; Maciej Grzybek; Marlena Robakowska; Olli Vapalahti; Teemu Smura |
| EPI_ISL_451648 to 451649, EPI_ISL_451652 to 451654 | Hematology Laboratory, Section of Molecular Diagnostics, University Clinical Centre, Medical University of Gdansk | Laboratory of Recombinant Vaccines | Adam Sodol; Aneta Szulc; Boguslaw Szewczyk; Ewa Milosz; Krystyna Bienkowska-Szewczyk; Krzysztof Lewandowski; Lukasz Rabalski; Marlena Robakowska |
| EPI_ISL_560581 to 560584, EPI_ISL_560586 to 560590, EPI_ISL_792056 to 792058 | Hopital | National Reference Center for Viruses of Respiratory Infections, Institut Pasteur, Paris | Angela Brisebarre; Camille Capel; Etienne Simon-Lorière; Fabiana Gambaro; Marion Barbet; Maud Vanpeene; Méline Bizard; Patricia Stoessel; Sylvie Behillil; Sylvie van der Werf; Vincent Enouf |
| EPI_ISL_418427 | Hopital Privé de l'Est Lyonnais | CNR Virus des Infections Respiratoires - France SUD | Alexandre Gaymard; Antonin Bal; Bruno Lina; Carine Moustaud; Florence Morfin-Sherpa; Gregory Destras; Gwendolyne Burfin; Laurence Josset; Martine Valette; Maude Bouscambert-Duchamp; Raphaelle Lamy; Solenne Brun |
| EPI_ISL_418229 | Hopital franco britannique - Laboratoire | National Reference Center for Viruses of Respiratory Infections, Institut Pasteur, Paris | Angela Brisebarre; Etienne Simon-Lorière; Flora Donati; Marianne Asso Bonnet; Marion Barbet; Maud Vanpeene; Mélanie Albert; Méline Bizard; Sylvie Behillil; Sylvie van der Werf; Vincent Enouf |
| EPI_ISL_416501 | Hopital franco britannique - Service des Urgences | National Reference Center for Viruses of Respiratory Infections, Institut Pasteur, Paris | Angela Brisebarre; Etienne Simon-Lorière; Flora Donati; Marion Barbet; Maud Vanpeene; Méline Bizard; Mélnie Albert; Sylvie Behillil; Sylvie van der Werf; Vincent Enouf |
| EPI_ISL_434666 | Hornefors Halsocentral | The Public Health Agency of Sweden | Anna Risberg; Anna-Malin Linde; Camilla Eiback; Karin Tegmark-Wisell; Maria Lind Karlberg; Mia Brytting; Olov Svartstrom; Oskar Karlsson Lindsjo; Theresa Enkirch |
| EPI_ISL_560598 to 560636, EPI_ISL_560643 to 560646, EPI_ISL_660323, EPI_ISL_693758 to 693764, EPI_ISL_718226 to 718249, EPI_ISL_832130 to 832166, EPI_ISL_832169 to 832177, EPI_ISL_855354 to 855356, EPI_ISL_855382, EPI_ISL_855388 | | | |
| see above | Hospital | National Reference Center for Viruses of Respiratory Infections, Institut Pasteur, Paris | Alexandra Ducancelle; Angela Brisebarre; Aurélie Guigon; Camille Capel; Clémence Guillaume; Ducancelle Alexandra; Etienne Simon-Lorière; Fabiana Gambaro; Gisèle Lagathu; Laurent Andreoletti; Léa Pilorge; Marion Barbet; Maud Vanpeene; Méline Bizard; Patricia Stoessel-Thouvenin; Sylvie Behillil; Sylvie van der Werf; Vincent Enouf |
| EPI_ISL_434366 to 434384, EPI_ISL_434386, EPI_ISL_450724 to 450732, EPI_ISL_450734 to 450737 | Hospital AZ Rivierenland | Institute of Tropical Medicine | ; Colin Anthony; Philippe Selhorst |
| EPI_ISL_491439 | Hospital Calderon Guardia | Inciensa, Instituto Costarricense de Investigación y Enseñanza en Nutrición y Salud | Adriana Godinez & Melany Calderon; Claudio Soto-Garita; Estela Cordero; Francisco Duarte; Hebleen Brenes |
| EPI_ISL_539559 to 539560 | Hospital Campo Arañuelo | Instituto de Salud Carlos III | A. Monzón; F. Casas; G. Rodríguez; I; I. Jiménez; Iglesias-Caballero; J. López; M. Camarero; M. Cuesta; M. González-Esguevillas; M. Molinero Calamita; M. Zaballos; P. Jiménez; S. Juliá; S. Pozo; S. Varona |
| EPI_ISL_462479 | Hospital Clinic | Instituto de Salud Carlos III | A. Monzón; F. Casas; I; I. Jiménez; Iglesias-Caballero; M. Camarero; M. Cuesta; M. González-Esguevillas; M. Molinero Calamita; M. Zaballos; M.A Marcos; P. Jiménez; S. Juliá; S. Pozo; S. Varona |
| EPI_ISL_539533 to 539556, EPI_ISL_862573 to 862588, EPI_ISL_862627 to 862636 | Hospital Clínic | Instituto de Salud Carlos III | A. Monzón; F. Casas; I; I. Jiménez; I. Marcos; Iglesias-Caballero; M. Pozo; M. Camarero; M. Cuesta; M. González-Esguevillas; M. Molinero Calamita; M. Zaballos; M.A Marcos; M.A.; M.Camarero; P. Jiménez; S. Juliá; S. Molinero Calamita; S. Pozo; S. Varona |
| EPI_ISL_802726 to 802749 | Hospital Clínic de Barcelona | Instituto de Salud Carlos III | A. Monzón; F. Casas; I; I. Jiménez; Iglesias-Caballero; M. Camarero; M. Cuesta; M. González-Esguevillas; M. Molinero Calamita; M. Zaballos; M.A Marcos.; P. Jiménez; S. Juliá; S. Pozo; S. Varona |
| EPI_ISL_691700 to 691728, EPI_ISL_862548, EPI_ISL_862569, EPI_ISL_871985 to 871990, EPI_ISL_871992 to 871993 | Hospital Clínico San Carlos | Instituto de Salud Carlos III | A. Monzón; F. Casas; I. Jiménez; I. Rodríguez, I.; Iglesias-Caballero; M. Camarero; M. Cuesta; M. González-Esguevillas; M. Pozo; M. Zaballos; P. Jiménez; S. Juliá; S. Molinero Calamita; S. Varona |
| EPI_ISL_539500 to 539502, EPI_ISL_539509 to 539512, EPI_ISL_578190 to 578191 | Hospital Clínico Universitario Lozano Blesa | Instituto de Salud Carlos III | A. Monzón; F. Casas; I; I. Jiménez; Iglesias-Caballero; M. Camarero; M. Cuesta; M. González-Esguevillas; M. Molinero Calamita; M. Zaballos; P. Jiménez; R. Benito; S. Juliá; S. Pozo; S. Varona |
| EPI_ISL_671794 to 671804, EPI_ISL_683595 to 683597 | Hospital Clínico Universitario Lozano Blesa de Zaragoza (España) | SeqCOVID-SPAIN consortium/IBV(CSIC) | Jessica Bueno and SeqCOVID-SPAIN consortium; Rafael Benito; Sonia Algarate |
| EPI_ISL_467262 to 467265, EPI_ISL_467267 to 467285, EPI_ISL_467287, EPI_ISL_467289 to 467297, EPI_ISL_500162, EPI_ISL_500167, EPI_ISL_500209, EPI_ISL_500219, EPI_ISL_500221, EPI_ISL_510268 to 510281, EPI_ISL_510283 to 510302 | | | |
| see above | Hospital Clínico Universitario de Santiago de Compostela | SeqCOVID-SPAIN consortium/IBV(CSIC) | Amparo Coira Nieto; Antonio Aguilera Guirao; Gema Barbeito Castiñeiras; José Javier Costa Alcalde; Mª Luisa Pérez del Molino Bernal; Rocio Trastoy Pena and SeqCOVID-SPAIN consortium |
| EPI_ISL_541037 to 541066 | Hospital Clínico Universitario de Santiago de Compostela | SeqCOVID-SPAIN consortium/Institute of Biomedicine of Valencia, IBV-CSIC | Amparo Coira Nieto; Antonio Aguilera Guirao; Gema Barbeito Castiñeiras; José Javier Costa Alcalde; Mª Luisa Pérez del Molino Bernal; Rocio Trastoy Pena and SeqCOVID-SPAIN consortium |
| EPI_ISL_455344 to 455349, EPI_ISL_539567 to 539568, EPI_ISL_804949, EPI_ISL_804952 to 804979, EPI_ISL_862589 to 862626 | Hospital Comarcal de Melilla | Instituto de Salud Carlos III | A. Monzón; C. Ezpeleta; F. Casas; I; I. Jiménez; I. López, J.; I. Pérez; Iglesias-Caballero; J. López; M. Camarero; M. Cuesta; M. González-Esguevillas; M. Molinero Calamita; M. Pozo; M. Zaballos; P. Jiménez; S. Juliá; S. Molinero Calamita; S. Pozo; S. Varona |
| EPI_ISL_462477, EPI_ISL_539499 | Hospital Costa del Sol | Instituto de Salud Carlos III | A. Monzón; F. Casas; F. Fernández; I; I. Jiménez; Iglesias-Caballero; M. Camarero; M. Cuesta; M. González-Esguevillas; M. Molinero Calamita; M. Zaballos; P. Jiménez; S. Juliá; S. Pozo; S. Varona |
| EPI_ISL_512654 | Hospital Dr. Rafael A. Calderon Guardia [San Jose/San Jose] | Inciensa, Instituto Costarricense de Investigación y Enseñanza en Nutrición y Salud | Adriana Godinez & Melany Calderon; Claudio Soto-Garita; Estela Cordero; Francisco Duarte; Hebleen Porras |
| EPI_ISL_578186 | Hospital General Juan Ramón Jiménez | Instituto de Salud Carlos III | A. Monzón; F. Casas; I; I. Jiménez; Iglesias-Caballero; J. Saavedra; M. Camarero; M. Cuesta; M. González-Esguevillas; M. Molinero Calamita; M. Zaballos; P. Jiménez; S. Juliá; S. Pozo; S. Varona |
| EPI_ISL_756271, EPI_ISL_756273 to 756274, EPI_ISL_776770, EPI_ISL_776875, EPI_ISL_776877, EPI_ISL_777490, EPI_ISL_777902, EPI_ISL_778445, EPI_ISL_778644 to 778645, EPI_ISL_803854 to 803864 | | | |
| see above | Hospital General Universitario Gregorio Marañón | Hospital General Universitario Gregorio Marañón | Darío García de Viedma; Darío García de Viedma.; Laura Perez-Lago; Laura Pérez-Lago; Patricia Muñoz; Pedro Sola Campoy; Pilar Catalán; Sergio Buenestado Serrano |
| EPI_ISL_467184 to 467199, EPI_ISL_467201 to 467261, EPI_ISL_481041 to 481045, EPI_ISL_481047 to 481060, EPI_ISL_481062 to 481084, EPI_ISL_481086 to 481109, EPI_ISL_510104 to 510106, EPI_ISL_510109 to 510128, EPI_ISL_510130 to 510151, EPI_ISL_510165 to 510167, EPI_ISL_510169 to 510186, EPI_ISL_510188 to 510213, EPI_ISL_510215 to 510243, EPI_ISL_510245, EPI_ISL_541878, EPI_ISL_541880 to 541895, EPI_ISL_541897 to 541941, EPI_ISL_582812 to 582832, EPI_ISL_582843 to 582844, EPI_ISL_654021 to 654050, EPI_ISL_654052 to 654063, EPI_ISL_654065 to 654124, EPI_ISL_654126 to 654134, EPI_ISL_654137 to 654138, EPI_ISL_654140 to 654162, EPI_ISL_654164 to 654169, EPI_ISL_654171, EPI_ISL_654174 to 654193, EPI_ISL_654195 to 654205, EPI_ISL_654207 to 654226, EPI_ISL_654228 to 654236, EPI_ISL_654238 to 654239, EPI_ISL_654241 to 654251, EPI_ISL_654253 to 654255, EPI_ISL_654257 to 654268, EPI_ISL_654270 to 654275, EPI_ISL_654277 to 654297, EPI_ISL_654299 to 654305, EPI_ISL_654307 to 654316, EPI_ISL_654318 to 654329, EPI_ISL_654331 to 654333, EPI_ISL_654335 to 654336, EPI_ISL_654341, EPI_ISL_654343 to 654353, EPI_ISL_654355 to 654357, EPI_ISL_654360 to 654384, EPI_ISL_654386 to 654393, EPI_ISL_654395 to 654400, EPI_ISL_779955 to 780021, EPI_ISL_780023 to 780028, EPI_ISL_780030 to 780032, EPI_ISL_780034 to 780037, EPI_ISL_780039 to 780043, EPI_ISL_780046 to 780049, EPI_ISL_780051, EPI_ISL_780053, EPI_ISL_780055, EPI_ISL_780057 to 780068, EPI_ISL_780070 to 780073, EPI_ISL_780075 to 780081, EPI_ISL_780083 to 780095, EPI_ISL_780414 to 780415, EPI_ISL_813980 to 813982, EPI_ISL_813984 to 813986, EPI_ISL_813989 to 813994, EPI_ISL_813997 to 813999, EPI_ISL_814001 to 814006, EPI_ISL_814008, EPI_ISL_814015 to 814016, EPI_ISL_814018 to 814020, EPI_ISL_814022, EPI_ISL_814026 to 814033, EPI_ISL_814037 to 814038, EPI_ISL_814040, EPI_ISL_814051 to 814061, EPI_ISL_852953, EPI_ISL_852958, EPI_ISL_852961, EPI_ISL_852974, EPI_ISL_852983, EPI_ISL_852988 to 852989, EPI_ISL_852991, EPI_ISL_852998, EPI_ISL_853000, EPI_ISL_853002, EPI_ISL_853016, EPI_ISL_861774 to 861777, EPI_ISL_861779 to 861784, EPI_ISL_861786 to 861798, EPI_ISL_861800 to 861807, EPI_ISL_861809 to 861820, EPI_ISL_861822, EPI_ISL_861825 to 861845, EPI_ISL_861847, EPI_ISL_861849 to 861855 | | | |
| see above | Hospital General Universitario Gregorio Marañón | SeqCOVID-SPAIN consortium/IBV(CSIC) | Darío García de Viedma; Darío García de Viedma and SeqCOVID-SPAIN consortium; Jon Sicilia; Julia Suárez; Laura Pérez-Lago; Marta Herranz; Patricia Muñoz; Patricia Muñoz and SeqCOVID-SPAIN consortium; Pedro J Sola-Campoy; Pilar Catalán; Sergio Buenestado-Serrano; Victor Manuel de la Cueva |
| EPI_ISL_756272 | Hospital General Universitario La Princesa | Hospital General Universitario Gregorio Marañón | Andrés Von Wernitz; Arturo fraile Torres; Carmen del Arco; Darío García de Viedma.; Laura Cardeñoso; Patricia Muñoz; Pedro Sola Campoy; Pilar Catalán; Sergio Buenestado Serrano |
| EPI_ISL_691682 to 691683 | Hospital General Universitario de Ciudad Real | Instituto de Salud Carlos III | A. Monzón; F. Casas; I. Jarilla, M.; I. Jiménez; I. Madrigal, M.; Iglesias-Caballero; M. Camarero; M. Cuesta; M. González-Esguevillas; M. Pozo; M. Zaballos; P. Jiménez; S. Juliá; S. Molinero Calamita; S. Varona |
| EPI_ISL_539523 | Hospital General de Segovia | Instituto de Salud Carlos III | A. Monzón; F. Casas; I; I. Jiménez; Iglesias-Caballero; M. Camarero; M. Cuesta; M. González-Esguevillas; M. Molinero Calamita; M. Zaballos; P. Jiménez; S. Hernando; S. Juliá; S. Pozo; S. Varona |
| EPI_ISL_418245 to 418246 | Hospital General y Universitario de Guadalajara | Instituto de Salud Carlos III | A. Monzón; F. Casas; I. Gonzalez-Praetorius A.; I. Jiménez; Iglesias-Caballero; M. Camarero; M. Cuesta; M. González-Esguevillas; M. Molinero Calamita; |

| | | | |
|---|---|---|---|
| | | | M. Zaballos; P. Jiménez; S. Juliá; S. Pozo; S. Varona |
| EPI_ISL_871974 to 871975, EPI_ISL_871979 | Hospital Infanta Cristina | Instituto de Salud Carlos III | A. Monzón; F. Casas; I. García, P.; I. Jiménez; Iglesias-Caballero; M. Camarero; M. Cuesta; M. González-Esguevillas; M. Pozo; M. Zaballos; P. Jiménez; S. Juliá; S. Molinero Calamita; S. Varona |
| EPI_ISL_412964 | Hospital Israelita Albert Einstein | Instituto Adolfo Lutz Interdisciplinary Procedures Center Strategic Laboratory | Andrew Rambaut; Claudia Regina Gonçalves; Claudio Tavares Sacchi; Daniela Bernardes Borges da Silva; Ester Cerdeira Sabino; Flávia Cristina da Silva Sales; Ingra Morales Claro; Jaqueline Goes de Jesus; Joshua Quick; Maria do Carmo; Nicholas James Loman; Nuno Rodrigues Faria; Sampaio Tavares Timenetsky |
| EPI_ISL_414014, EPI_ISL_416033 | Hospital Israelita Albert Einstein | Instituto Adolfo Lutz, Interdiciplinary Procedures Center, Strategic Laboratory | Adriana Bugno; Adriano Abbud; Carlos Henrique Camargo; Claudia Regina Gonçalves; Claudio Tavares Sacchi; Daniela Bernardes Borges da Silva; Erica Valessa Ramos Gomes; Ester Cerdeira Sabino; Fabiana Cristina Pereira dos Santos; Katia Correia dos Santos; Maria do Carmo Sampaio Tavares Timenetsky; Maria do Carmo Sampaito Tavares Timenetsky; Simone Guadagnucci Morillo; Terezinha Maria de Paiva |
| EPI_ISL_413016 | Hospital Israelita Albert Einstein | Instituto Adolfo Lutz, Interdisciplinary Procedures Center, Strategic Laboratory | Andrew Rambaut; Claudia Regina Gonçalves; Claudio Tavares Sacchi; Ester Cerdeira Sabino; Fabiana Cristina Pereira dos Santos; Flávia Cristina da Silva Sales; Ingra Morales Claro; Jaqueline Goes de Jesus; Joshua Quick; Maria do Carmo Sampaio Tavares Timenetsky; Nicholas James Loman; Nuno Rodrigues Faria |
| EPI_ISL_862551 | Hospital J.M. Morales Meseguer | Instituto de Salud Carlos III | A. Monzón; F. Casas; I. Guerrero, C.; I. Jiménez; Iglesias-Caballero; M. . Pozo; M. Camarero; M. Cuesta; M. González-Esguevillas; M. Zaballos; P. Jiménez; S. Juliá; S. Molinero Calamita; S. Varona |
| EPI_ISL_539496 | Hospital Nostra Senyora de Meritxell | Instituto de Salud Carlos III | A. Monzón; F. Casas; F. Fernández; I; I. Jiménez; Iglesias-Caballero; M. Camarero; M. Cuesta; M. González-Esguevillas; M. Molinero Calamita; M. Zaballos; P. Jiménez; S. Juliá; S. Pozo; S. Varona |
| EPI_ISL_539525, EPI_ISL_862637 to 862639 | Hospital Nuestra Señora de Sonsoles | Instituto de Salud Carlos III | A. Monzón; A. San Pedro; F. Casas; I; I. Jiménez; Iglesias-Caballero; M. Camarero; M. Cuesta; M. González-Esguevillas; M. Molinero Calamita; M. Zaballos; P. Jiménez; S. Juliá; S. Pozo; S. Varona |
| EPI_ISL_419386 to 419387 | Hospital Prof. Doutor Fernando Fonseca, EPE | Instituto Gulbenkian de Ciência | Cathy Paulino; Joao Sobral; João Costa; Ricardo Leite; Susana Ladeiro |
| EPI_ISL_491454 | Hospital San Juan de Dios | Inciensa, Instituto Costarricense de Investigación y Enseñanza en Nutrición y Salud | Adriana Godinez & Melany Calderon; Claudio Soto-Garita; Estela Cordero; Francisco Duarte; Hebleen Brenes |
| EPI_ISL_419234, EPI_ISL_455336 to 455343, EPI_ISL_578196 to 578201 | Hospital San Pedro | Instituto de Salud Carlos III | A. Monzón; Alonso, C.; C. Alonso; Camarero, S.; Casas, I.; Cuesta, I.; F. Casas; González-Esguevillas, M.; I; I. Jiménez; Iglesias-Caballero; Iglesias-Caballero, M.; J.M Azcona; Jiménez, M.; Jiménez, P.; Juliá, M.; M. Blasco; M. Camarero; M. Cuesta; M. González-Esguevillas; M. Molinero Calamita; M. Zaballos; Molinero Calamita, M.; Monzón, S.; P. Jiménez; Pozo, F.; S. Juliá; S. Pozo; S. Varona; Varona, S.; Zaballos, A. |
| EPI_ISL_539558, EPI_ISL_539561 to 539562, EPI_ISL_862654 | Hospital San Pedro de Alcántara | Instituto de Salud Carlos III | A. Monzón; E. Cerro; F. Casas; I; I. Jiménez; I. Rodríguez, G.; Iglesias-Caballero; J. López; M. Pozo; M. Camarero; M. Cuesta; M. González-Esguevillas; M. Molinero Calamita; M. Zaballos; M.A Cañizares; M.Camarero; P. Jiménez; S. Juliá; S. Molinero Calamita; S. Pozo; S. Varona |
| EPI_ISL_510305, EPI_ISL_510307 to 510332 | Hospital San Pedro de Alcántara (Cáceres) | SeqCOVID-SPAIN consortium/IBV(CSIC) | Cristina Muñoz Cuevas; Guadalupe Rodriguez Rodriguez and SeqCOVID-SPAIN consortium |
| EPI_ISL_802750 to 802776 | Hospital Santa María Nai | Instituto de Salud Carlos III | A. Monzón; F. Casas; I; I. Jiménez; Iglesias-Caballero; J. García.; M. Camarero; M. Cuesta; M. González-Esguevillas; M. Molinero Calamita; M. Zaballos; P. Jiménez; S. Juliá; S. Pozo; S. Varona |
| EPI_ISL_524484 | Hospital Schwyz | Institute of Medical Virology, University of Zurich | Alexandra Trkola; Andrea Zbinden; Fiona Steiner; Gabriela Ziltener; Jon Huder; Jürg Böni; Maryam Zaheri; Michael Huber; Patrick Redli; Riccarda Capaul; Stefan Schmutz; Verena Kufner |
| EPI_ISL_414017 | Hospital São Joaquim Beneficencia Portuguesa | Instituto Adolfo Lutz, Interdiciplinary Procedures Center, Strategic Laboratory | Carlos Henrique Camargo; Claudia Regina Gonçalves; Claudio Tavares Sacchi; Daniela Bernardes Borges da Silva; Ester Cerdeira Sabino; Fabiana Cristina Pereira dos Santos; Maria do Carmo Sampaito Tavares Timenetsky; Terezinha Maria de Paiva |
| EPI_ISL_455350 to 455351 | Hospital Txagorritxu | Instituto de Salud Carlos III | A. Monzón; C. Gómez; F. Casas; I; I. Jiménez; Iglesias-Caballero; M. Camarero; M. Cuesta; M. González-Esguevillas; M. Molinero Calamita; M. Zaballos; P. Jiménez; S. Juliá; S. Pozo; S. Varona |
| EPI_ISL_483059 | Hospital Universitari Germans Trias i Pujol | IrsiCaixa AIDS Research Lab | A. Valencia; B. Clotet; C. Avila-Nieto; E. Vidal; G. Cantero; I. Blanco; J. Blanco; J. Carrillo; J. Rodon; J. Segalés; J. Vergara-Alert; M. Noguera-Julián; M. Parera; M. Puig; M.T. Terrón; N. Izquierdo-Useros; S. Cruz; V. Guallar |
| EPI_ISL_535575, EPI_ISL_538552 | Hospital Universitari Germans Trias i Pujol(HUGTiP)/Fundació Lluita contra la SIDA (FLSida) | IrsiCaixa AIDS Research Lab | Bonaventura Clotet; Ester Ballana; Joaquim Segalés; Jorge Carrillo; Julia Blanco; Lidia Ruiz; Marc Noguera-Julian; Maria Pilar Armengol; Mariona Parera; Marta Massanella; Nuria Izquierdo; Roger Paredes |
| EPI_ISL_471472, EPI_ISL_510689 | Hospital Universitari Germans Trias i Pujol(HUGTiP)/Fundació Lluita contra la SIDA (FLSida)/IRTA-CReSA | IrsiCaixa AIDS Research Lab | Albert Bensaid; Bonaventura Clotet; Joaquim Segalés; Jordi Rodón; Jorge Carrillo; Julia Blanco; Julia Vergara; Lidia Ruiz; Marc Noguera-Julian; Nuria Izquierdo; Pilar Armengol; Roger Paredes |
| EPI_ISL_516193 | Hospital Universitari Germans Trias i Pujol. | IrsiCaixa AIDS Research Lab | Bonaventura Clotet; Ester Ballana; Joaquim Segalés; Jorge Carrillo; Julia Blanco; Lidia Ruiz; Marc Noguera-Julian; Maria Pilar Armengol; Mariona Parera; Marta Massanella; Nuria Izquierdo; Roger Paredes |
| EPI_ISL_418860 to 418861, EPI_ISL_728364 to 728365 | Hospital Universitari Vall d'Hebron (HUVH) - Vall d'Hebron Research Institute (VHIR) | Hospital Universitari Vall d'Hebron (HUVH) - Vall d'Hebron Research Institute (VHIR) | Andrés Antón; Ariadna Rando; Aroa Silgado; Carla Castillo; Cristina Andrés; Damir Garcia-Cehic; Dàmir Garcia-Cehic; Elena Sulleiro; Josep F Abril; Josep Quer; Juliana Esperalba; Maria Carmen Martín; Maria Gema Codina; Maria Piñana; Mercedes Guerrero-Murillo; Tomàs Pumarola |
| EPI_ISL_728362 | Hospital Universitari Vall d'Hebron (HUVH) - Vall d'Hebron Research Institute (VHIR) | Hospital Universitari Vall d'Hebron - Vall d'Hebron Research Institute (VHIR) | Andrés Antón; Ariadna Rando; Aroa Silgado; Carla Castillo; Cristina Andrés; Damir Garcia-Cehic; Elena Sulleiro; Josep F Abril; Josep Quer; Juliana Esperalba; Maria Carmen Martín; Maria Gema Codina; Maria Piñana; Tomàs Pumarola |
| EPI_ISL_728361 | Hospital Universitari Vall d'Hebron (HUVH) - Vall d'Hebron Research Institute (VHIR) | Hospital Universitari Vall d'Hebron - Vall d'hebron Research Institut (VHIR) | Andrés Antón; Ariadna Rando; Aroa Silgado; Carla Castillo; Cristina Andrés; Damir Garcia-Cehic; Elena Sulleiro; Josep F Abril; Josep Quer; Juliana Esperalba; Maria Carmen Martín; Maria Gema Codina; Maria Piñana; Tomàs Pumarola |
| EPI_ISL_444980 to 444983 | Hospital Universitari Vall d'Hebron - Vall d''hebron Institut de Recerca | Hospital Universitari Vall d'Hebron | Andrés Antón; Ariadna Rando; Cristina Andrés; Damir Garcia-Cehic; Josep Quer; Juliana Esperalba; Maria Gema Codina; Maria Piñana; Mercedes Guerrero-Murillo; Tomàs Pumarola |
| EPI_ISL_444971 to 444972, EPI_ISL_444974 to 444979, EPI_ISL_444984 to 444990, EPI_ISL_447532 to 447533, EPI_ISL_450337, EPI_ISL_458130 to 458131 | Hospital Universitari Vall d'Hebron - Vall d'Hebron Institut de Recerca | Hospital Universitari Vall d'Hebron | Andrés Antón; Ariadna Rando; Cristina Andrés; DAmir Garcia-Cehic; Damir Garcia-Cehic; Josep Gregori; Josep Quer; Juliana Esperalba; Maria Carmen Martin; Maria Carmen Martín; Maria Gema Codina; Maria Piñana; Mercedes Guerrero-Murillo; Tomàs Pumarola |
| EPI_ISL_458132, EPI_ISL_819293 to 819327, EPI_ISL_819329 to 819363, EPI_ISL_824411 to 824512, EPI_ISL_825326 to 825327, EPI_ISL_825335 to 825384, EPI_ISL_825620 to 825625, EPI_ISL_825627 to 825628, EPI_ISL_825630 to 825631 | | | |
| see above | Hospital Universitari Vall d'Hebron - Vall d'Hebron Institut de Rercerca | Hospital Universitari Vall d'Hebron | Andrés Antón; Ariadna Rando; Carla Castillo; Cristina Andrés; Damir Garcia-Cehic; Josep F Abril; Josep Gregori; Josep Quer; Juliana Esperalba; Maria Carmen Martin; Maria Gema Codina; Maria Piñana; Mercedes Guerrero-Murillo; Tomàs Pumarola |
| EPI_ISL_444973 | Hospital Universitari Vall d`Hebron - Vall d'Hebron Institut de Recerca | Hospital Universitari Vall d'Hebron | Andrés Antón; Ariadna Rando; Cristina Andrés; DAmir Garcia-Cehic; Josep Quer; Juliana Esperalba; Maria Gema Codina; Maria Piñana; Mercedes Guerrero-Murillo; Tomàs Pumarola |
| EPI_ISL_538003 to 538009, EPI_ISL_538011 to 538054, EPI_ISL_539245 | Hospital Universitari i Politècnic La Fe de València | SeqCOVID-SPAIN consortium/IBV(CSIC) | Ana Gil Brusola; Eva González Barbera; José Luis López Hontangas and SeqCOVID-SPAIN consortium; María Dolores Gómez Ruiz; Salvador Giner Almaraz |
| EPI_ISL_421171 to 421180, EPI_ISL_428683 to 428711, EPI_ISL_430718, EPI_ISL_529972 to 530024, EPI_ISL_534327 to 534329, EPI_ISL_737930 | Hospital Universitario 12 de Octubre | Hospital Universitario 12 de Octubre | Elias Dahdouh; Esther Viedma; Fernando Lázaro; Irene Múñoz-Gallego; Jesús Mingorance; Juan Carlos Galán; Julio García; Ma Dolores Folgueira; Mª Dolores Folgueira; Natalia Stella; Rafael Cantón; Rafael Delgado; Raul Recio; Raúl Recio; Sara González |
| EPI_ISL_417952, EPI_ISL_417954, EPI_ISL_417956 to 417957, EPI_ISL_417961, EPI_ISL_417963, EPI_ISL_417967, EPI_ISL_418182 | | | |
| see above | Hospital Universitario 12 de Octubre | Hospital Universitario La Paz | Elias Dahdouh; Esther Viedma; Fernando Lázaro; Jesús Mingorance; Juan Carlos Galán; Julio García; Mª Dolores Folgueira; Natalia Stella; Rafael Cantón; Rafael Delgado; Sara González |
| EPI_ISL_467092 to 467117, EPI_ISL_467119 to 467183, EPI_ISL_500228 to 500235, EPI_ISL_500237 to 500286, EPI_ISL_509616 to 509618, EPI_ISL_537874 to 537880, EPI_ISL_537882, EPI_ISL_537884 to 537905, EPI_ISL_537907 to 537924, EPI_ISL_537926 to 537945, EPI_ISL_538118, EPI_ISL_538231 to 538232 | | | |
| see above | Hospital Universitario Araba. Vitoria-Gasteiz | SeqCOVID-SPAIN consortium/IBV(CSIC) | Amaia Aguirre Quiñonero; Andrés Canut Blasco and SeqCOVID-SPAIN consortium; Andrés Canut Blasco. and SeqCOVID-SPAIN consortium; Carmen |

| | | | |
|---|---|---|---|
| | | | Gómez González; Ma Concepción Lecaroz Agara; Ma Rosario Almela Ferrer; Marina Fernández Torres; Mª Concepción Lecaroz Agara; Mª Rosario Almela Ferrer; Silvia Hernáez Crespo |
| EPI_ISL_452692 to 452756, EPI_ISL_452758 to 452786 | Hospital Universitario Araba. Vitoria-Gasteiz, | SeqCOVID-SPAIN consortium/IBV(CSIC) | Amaia Aguirre Quiñonero; Andrés Canut Blasco and SeqCOVID-SPAIN consortium; Carmen Gómez González; Maria Concepción Lecaroz Agara; Maria Rosario Almela Ferrer; Marina Fernández Torres; Silvia Hernáez Crespo |
| EPI_ISL_455326 | Hospital Universitario Insular de Gran Canaria | Instituto de Salud Carlos III | A. Hernández; A. Monzón; F. Casas; I; I. Jiménez; Iglesias-Caballero; M. Camarero; M. Cuesta; M. González-Esguevillas; M. Molinero Calamita; M. Zaballos; P. Jiménez; S. Juliá; S. Pozo; S. Varona |
| EPI_ISL_428674 to 428682, EPI_ISL_430719 to 430721, EPI_ISL_435144 | Hospital Universitario La Paz | Hospital Universitario 12 de Octubre | Elias Dahdouh; Esther Viedma; Fernando Lázaro; Jesús Mingorance; Juan Carlos Galán; Julio García; Mª Dolores Folgueira; Natalia Stella; Rafael Cantón; Rafael Delgado; Raúl Recio; Sara González |
| EPI_ISL_417969, EPI_ISL_417972, EPI_ISL_417975, EPI_ISL_417978, EPI_ISL_530025 to 530093, EPI_ISL_534330 to 534333 | Hospital Universitario La Paz | Hospital Universitario La Paz | Elias Dahdouh; Esther Viedma; Fernando Lázaro; Jesús Mingorance; Juan Carlos Galán; Julio García; Ma Dolores Folgueira; María Rodríguez; Mª Dolores Folgueira; Natalia Stella; Rafael Cantón; Rafael Delgado; Raúl Recio; Sara González |
| EPI_ISL_831021 to 831116, EPI_ISL_831118 to 831288 | Hospital Universitario La Paz (Madrid) | SeqCOVID-SPAIN consortium/IBV(CSIC) | Elias Dahdouh; Fernando Lázaro-Perona; Jesús Mingorance and SeqCOVID-SPAIN consortium; María Rodríguez-Tejedor |
| EPI_ISL_871976 to 871977 | Hospital Universitario Lozano Bielsa | Instituto de Salud Carlos III | A. Monzón; F. Casas; I. Benito, R.; I. Jiménez; Iglesias-Caballero; M. Camarero; M. Cuesta; M. González-Esguevillas; M. Pozo; M. Zaballos; P. Jiménez; S. Juliá; S. Molinero Calamita; S. Varona |
| EPI_ISL_537809 to 537810 | Hospital Universitario Marqués de Valdecilla (Santander), Servicio de Microbiología | SeqCOVID-SPAIN consortium/IBV(CSIC) | Daniel Pablo Marcos; Jesús Rodríguez Lozano; Jose Manuel Méndez Legaza; María Eliecer Cano García; María Siller Ruiz and SeqCOVID-SPAIN consortium; Mónica Gozalo Margüello |
| EPI_ISL_538690 to 538722, EPI_ISL_538724 to 538736, EPI_ISL_538738 to 538747, EPI_ISL_582091, EPI_ISL_582093 to 582094, EPI_ISL_582097 to 582105, EPI_ISL_582108 to 582109, EPI_ISL_871893, EPI_ISL_871896, EPI_ISL_871911 | | | |
| see above | Hospital Universitario Marqués de Valdecilla - IDIVAL (Santander, Cantabria) | SeqCOVID-SPAIN consortium/IBV(CSIC) | Daniel Pablo Marcos; Jesús Rodríguez Rodríguez; Jose Manuel Méndez Legaza; María Eliecer Cano García; María Siller Ruiz and SeqCOVID-SPAIN consortium; Mónica Gozalo Margüello |
| EPI_ISL_539503 to 539508, EPI_ISL_539513 to 539519, EPI_ISL_578189, EPI_ISL_578193, EPI_ISL_871978, EPI_ISL_871980 to 871983 | Hospital Universitario Miguel Servet | Instituto de Salud Carlos III | A. Monzón; A. Rezusta; F. Casas; I; I. Jiménez; I. Rezusta, A.; Iglesias-Caballero; M. Camarero; M. Cuesta; M. González-Esguevillas; M. Molinero Calamita; M. Pozo; M. Zaballos; P. Jiménez; R. Benito; S. Juliá; S. Molinero Calamita; S. Pozo; S. Varona |
| EPI_ISL_691731 | Hospital Universitario Puerta de Hierro | Instituto de Salud Carlos III | A. Monzón; F. Casas; I. Jiménez; I. Velasco, A.; Iglesias-Caballero; M. Camarero; M. Cuesta; M. González-Esguevillas; M. Pozo; M. Zaballos; P. Jiménez; S. Juliá; S. Molinero Calamita; S. Varona |
| EPI_ISL_452453 to 452457, EPI_ISL_452459 to 452471, EPI_ISL_467064 to 467066, EPI_ISL_467068 to 467085 | Hospital Universitario Puerta del Mar de Cádiz - INIBICA | SeqCOVID-SPAIN consortium/IBV(CSIC) | Fátima-Galán-Sánchez; Manuel Rodríguez-Iglesias and SeqCOVID-SPAIN consortium; Manuel Rodrí-guez-Iglesias and SeqCOVID-SPAIN consortium; Salud Rodríguez-Pallares |
| EPI_ISL_530094 to 530099 | Hospital Universitario Ramón y Cajal | Hospital Universitario 12 de Octubre | Elias Dahdouh; Esther Viedma; Fernando Lázaro; Jesús Mingorance; Juan Carlos Galán; Julio García; Mª Dolores Folgueira; Natalia Stella; Rafael Cantón; Rafael Delgado; Raúl Recio; Sara González |
| EPI_ISL_417979 to 417981, EPI_ISL_530100 to 530119, EPI_ISL_534334 | Hospital Universitario Ramón y Cajal | Hospital Universitario La Paz | Elias Dahdouh; Esther Viedma; Fernando Lázaro; Jesús Mingorance; Juan Carlos Galán; Julio García; Ma Dolores Folgueira; María Rodríguez; Mª Dolores Folgueira; Natalia Stella; Rafael Cantón; Rafael Delgado; Raúl Recio; Sara González |
| EPI_ISL_691729 to 691730, EPI_ISL_871973, EPI_ISL_871984, EPI_ISL_871991 | Hospital Universitario Severo Ochoa | Instituto de Salud Carlos III | A. Monzón; F. Casas; I. García; I. García, M.; I. Jiménez; Iglesias-Caballero; M. Camarero; M. Cuesta; M. González-Esguevillas; M. Pozo; M. Zaballos; M.L.; P. Jiménez; S. Juliá; S. Molinero Calamita; S. Varona |
| EPI_ISL_862545, EPI_ISL_862549, EPI_ISL_862568 | Hospital Universitario Virgen de la Arrixaca | Instituto de Salud Carlos III | A. Monzón; F. Casas; I; I. Jiménez; Iglesias-Caballero; M. Camarero; M. Cuesta; M. González-Esguevillas; M. Pozo; M. Zaballos; Moreno, A.; P. Jiménez; S. Juliá; S. Molinero Calamita; S. Varona |
| EPI_ISL_419230 | Hospital Universitario Virgen de las Nieves | Instituto de Salud Carlos III | Camarero, S.; Casas, I.; Cuesta, I.; González-Esguevillas, M.; Iglesias-Caballero, M.; Jiménez, M.; Jiménez, P.; Juliá, M.; Molinero Calamita, M.; Monzón, S.; Pozo, F.; Sanbonmatsu, S.; Varona, S.; Zaballos, A. |
| EPI_ISL_467059 to 467063, EPI_ISL_474797, EPI_ISL_474832 to 474836, EPI_ISL_474841 to 474846, EPI_ISL_474850 to 474852, EPI_ISL_474854 to 474900, EPI_ISL_474907, EPI_ISL_474909, EPI_ISL_474920, EPI_ISL_474922 to 474932, EPI_ISL_474934 to 474939, EPI_ISL_474942 to 474943, EPI_ISL_474948 to 474950, EPI_ISL_474957, EPI_ISL_500160 to 500161, EPI_ISL_500163 to 500165, EPI_ISL_500169 to 500170, EPI_ISL_500172 to 500176, EPI_ISL_500178 to 500199, EPI_ISL_500201 to 500206, EPI_ISL_500208, EPI_ISL_500210 to 500216, EPI_ISL_500220, EPI_ISL_500222 to 500227, EPI_ISL_510424 to 510425, EPI_ISL_510427 to 510460, EPI_ISL_538672 to 538689, EPI_ISL_654626 | | | |
| see above | Hospital Universitario Virgen de las Nieves de Granada-SAS | SeqCOVID-SPAIN consortium/IBV(CSIC) | Irene Pedrosa Corral; José M. Navarro-Marí and SeqCOVID-SPAIN consortium; Mercedes Pérez Ruiz; Sara Sanbonmatsu Gámez |
| EPI_ISL_419233, EPI_ISL_455325, EPI_ISL_539532 | Hospital Universitario de Canarias | Instituto de Salud Carlos III | A. Monzón; B. Castro; Camarero, S.; Casas, I.; Castro, B.; Cuesta, I.; F. Casas; González-Esguevillas, M.; I; I. Jiménez; Iglesias-Caballero; Iglesias-Caballero, M.; Jiménez, M.; Jiménez, P.; Juliá, M.; M. Camarero; M. Cuesta; M. González-Esguevillas; M. Molinero Calamita; M. Zaballos; Molinero Calamita, M.; Monzón, S.; P. Jiménez; Pozo, F.; S. Juliá; S. Pozo; S. Varona; Varona, S.; Zaballos, A. |
| EPI_ISL_539520 to 539522, EPI_ISL_691686 to 691699, EPI_ISL_862655 to 862658, EPI_ISL_871995 | Hospital Universitario de Ceuta | Instituto de Salud Carlos III | A. Monzón; F. Casas; G. Sánchez; I; I. Hijano, S.; I. Jiménez; Iglesias-Caballero; J. López; M. Camarero; M. Cuesta; M. González-Esguevillas; M. Molinero Calamita; M. Pozo; M. Zaballos; P. Jiménez; S. Juliá; S. Molinero Calamita; S. Pozo; S. Varona |
| EPI_ISL_467086 to 467091, EPI_ISL_474910 to 474918, EPI_ISL_537691 to 537728, EPI_ISL_537788 to 537796, EPI_ISL_537798 to 537799, EPI_ISL_537801 to 537808, EPI_ISL_561370 to 561372, EPI_ISL_660600 | | | |
| see above | Hospital Universitario de Gran Canaria Dr. Negrín | SeqCOVID-SPAIN consortium/IBV(CSIC) | Ana Bordes Benítez and SeqCOVID-SPAIN consortium; Francisco J. Chamizo López; M. Carmen Pérez González |
| EPI_ISL_539246 to 539247, EPI_ISL_539249, EPI_ISL_539251 to 539258, EPI_ISL_539262, EPI_ISL_539264, EPI_ISL_539266 to 539267, EPI_ISL_539270, EPI_ISL_539272 to 539283 | | | |
| see above | Hospital Universitario de La Ribera (Alzira, València) | SeqCOVID-SPAIN consortium/IBV(CSIC) | Julia González and SeqCOVID-SPAIN consortium; Olalla Martínez Macias |
| EPI_ISL_693516, EPI_ISL_693518 to 693520, EPI_ISL_693523 to 693524, EPI_ISL_693534 to 693536, EPI_ISL_693630 to 693657 | Hospital Vila Franca de Xira | Instituto Nacional de Saude (INSA) | Borges et al |
| EPI_ISL_455315 to 455322, EPI_ISL_455324, EPI_ISL_539497 to 539498, EPI_ISL_578187 to 578188 | Hospital Virgen de las Nieves | Instituto de Salud Carlos III | A. Monzón; F. Casas; I; I. Jiménez; Iglesias-Caballero; J. Lepe; J.M Navarro; M. Camarero; M. Cuesta; M. González-Esguevillas; M. Molinero Calamita; M. Zaballos; P. Jiménez; S. Juliá; S. Pozo; S. Sanbonmatsu; S. Varona |
| EPI_ISL_455314, EPI_ISL_455323 | Hospital Virgen del Rocio | Instituto de Salud Carlos III | A. Monzón; F. Casas; I; I. Jiménez; Iglesias-Caballero; J. Lepe; M. Camarero; M. Cuesta; M. González-Esguevillas; M. Molinero Calamita; M. Zaballos; P. Jiménez; S. Juliá; S. Pozo; S. Varona |
| EPI_ISL_511577 | Hospital de Braga | Instituto Nacional de Saude (INSA) | Borges et al |
| EPI_ISL_455352 to 455354 | Hospital de Cruces | Instituto de Salud Carlos III | A. Monzón; F. Casas; I; I. Jiménez; Iglesias-Caballero; M. Aranzamendi; M. Camarero; M. Cuesta; M. González-Esguevillas; M. Molinero Calamita; M. Zaballos; P. Jiménez; S. Juliá; S. Pozo; S. Varona |
| EPI_ISL_691685 | Hospital de Leon | Instituto de Salud Carlos III | A. Monzón; F. Casas; I. Jiménez; I. Vidan, J.; Iglesias-Caballero; M. Camarero; M. Cuesta; M. González-Esguevillas; M. Pozo; M. Zaballos; P. Jiménez; S. Juliá; S. Molinero Calamita; S. Varona |
| EPI_ISL_691732, EPI_ISL_871994 | Hospital de Madrid | Instituto de Salud Carlos III | A. Monzón; F. Casas; I. Alhambra, A.; I. Jiménez; Iglesias-Caballero; M. Camarero; M. Cuesta; M. González-Esguevillas; M. Pozo; M. Zaballos; P. Jiménez; S. Juliá; S. Molinero Calamita; S. Varona |
| EPI_ISL_850508 | Hospital de Tortosa Verge de la Cinta | Hospital Universitari Vall d'Hebron - Vall d'Hebron Institut de Rercerca | Andrés Antón; Ariadna Rando; Carla Castillo; Cristina Andrés; Damir Garcia-Cehic; Josep F Abril; Josep Quer; Juliana Esperalba; Maria Carmen Martin; Maria Gema Codina; Maria Piñana; Tomàs Pumarola |
| EPI_ISL_510247 to 510267, EPI_ISL_671807, EPI_ISL_671809 to 671810, EPI_ISL_671815, EPI_ISL_671826, EPI_ISL_671829 to 671830, EPI_ISL_671833 | | | |

| | | | |
|---|---|---|---|
| see above | Hospital de la Santa Creu i Sant Pau. Servicio de Microbiología | SeqCOVID-SPAIN consortium/IBV(CSIC) | Elisenda Miró and SeqCOVID-SPAIN consortium; Ferran Navarro; Núria Rabella |
| EPI_ISL_541067 to 541076, EPI_ISL_541078 to 541081 | Hospital de la Santa Creu i Sant Pau. Servicio de Microbiología | SeqCOVID-SPAIN consortium/Institute of Biomedicine of Valencia, IBV-CSIC | Elisenda Miró and SeqCOVID-SPAIN consortium; Ferran Navarro; Núria Rabella |
| EPI_ISL_417486 to 417487, EPI_ISL_420147, EPI_ISL_471173, EPI_ISL_471176, EPI_ISL_493384, EPI_ISL_500775, EPI_ISL_500791 to 500792, EPI_ISL_500797 to 500799, EPI_ISL_549036 to 549037, EPI_ISL_549052, EPI_ISL_549167, EPI_ISL_590904 to 590907, EPI_ISL_590915, EPI_ISL_590936 to 590937, EPI_ISL_590984, EPI_ISL_635097, EPI_ISL_635121 to 635123, EPI_ISL_635138, EPI_ISL_635143, EPI_ISL_635161, EPI_ISL_635189, EPI_ISL_668394 to 668395, EPI_ISL_668427 to 668428, EPI_ISL_708055 to 708056, EPI_ISL_708122 to 708125, EPI_ISL_759976, EPI_ISL_775285, EPI_ISL_775371, EPI_ISL_775518 to 775522, EPI_ISL_796664, EPI_ISL_796688, EPI_ISL_796714 to 796716, EPI_ISL_796729, EPI_ISL_813977, EPI_ISL_860214, EPI_ISL_860222, EPI_ISL_860236 | | | |
| see above | Hospital of Southern Norway - Kristiansand, Department of Medical Microbiology | Norwegian Institute of Public Health, Department of Virology | Atiya R Ali; Hilde Elshaug; Hilde Synnøve Vollan; Hilde Vollan; Kamilla Heddeland Instefjord; Karoline Bragstad; Kathrine Stene-Johansen; Marie Paulsen Madsen; Olav Hungnes; Rasmus Riis Kopperud |
| EPI_ISL_593915 | Hospital, Amneville | National Reference Center for Viruses of Respiratory Infections, Institut Pasteur, Paris | Etienne Simon-Lorière; Fabiana Gambaro; Maud Vanpeene; Sylvie Behillil; Sylvie van der Werf; Vincent Enouf |
| EPI_ISL_593903 | Hospital, Argenteuil | National Reference Center for Viruses of Respiratory Infections, Institut Pasteur, Paris | Etienne Simon-Lorière; Fabiana Gambaro; Maud Vanpeene; Sylvie Behillil; Sylvie van der Werf; Vincent Enouf |
| EPI_ISL_593910 | Hospital, Boulogne-Billancourt | National Reference Center for Viruses of Respiratory Infections, Institut Pasteur, Paris | Etienne Simon-Lorière; Fabiana Gambaro; Maud Vanpeene; Sylvie Behillil; Sylvie van der Werf; Vincent Enouf |
| EPI_ISL_593921 | Hospital, Fameck | National Reference Center for Viruses of Respiratory Infections, Institut Pasteur, Paris | Etienne Simon-Lorière; Fabiana Gambaro; Maud Vanpeene; Sylvie Behillil; Sylvie van der Werf; Vincent Enouf |
| EPI_ISL_593918 | Hospital, Joeuf | National Reference Center for Viruses of Respiratory Infections, Institut Pasteur, Paris | Etienne Simon-Lorière; Fabiana Gambaro; Maud Vanpeene; Sylvie Behillil; Sylvie van der Werf; Vincent Enouf |
| EPI_ISL_593911 | Hospital, Le Chesnay Cédex | National Reference Center for Viruses of Respiratory Infections, Institut Pasteur, Paris | Etienne Simon-Lorière; Fabiana Gambaro; Maud Vanpeene; Sylvie Behillil; Sylvie van der Werf; Vincent Enouf |
| EPI_ISL_593922 to 593923 | Hospital, Maizières les Metz | National Reference Center for Viruses of Respiratory Infections, Institut Pasteur, Paris | Etienne Simon-Lorière; Fabiana Gambaro; Maud Vanpeene; Sylvie Behillil; Sylvie van der Werf; Vincent Enouf |
| EPI_ISL_593912 | Hospital, Marange Silvange | National Reference Center for Viruses of Respiratory Infections, Institut Pasteur, Paris | Etienne Simon-Lorière; Fabiana Gambaro; Maud Vanpeene; Sylvie Behillil; Sylvie van der Werf; Vincent Enouf |
| EPI_ISL_593914, EPI_ISL_593920 | Hospital, Metz | National Reference Center for Viruses of Respiratory Infections, Institut Pasteur, Paris | Etienne Simon-Lorière; Fabiana Gambaro; Maud Vanpeene; Sylvie Behillil; Sylvie van der Werf; Vincent Enouf |
| EPI_ISL_593917 | Hospital, Montigny les Metz | National Reference Center for Viruses of Respiratory Infections, Institut Pasteur, Paris | Etienne Simon-Lorière; Fabiana Gambaro; Maud Vanpeene; Sylvie Behillil; Sylvie van der Werf; Vincent Enouf |
| EPI_ISL_593919 | Hospital, Ottonville | National Reference Center for Viruses of Respiratory Infections, Institut Pasteur, Paris | Etienne Simon-Lorière; Fabiana Gambaro; Maud Vanpeene; Sylvie Behillil; Sylvie van der Werf; Vincent Enouf |
| EPI_ISL_593916 | Hospital, Sarreguemines | National Reference Center for Viruses of Respiratory Infections, Institut Pasteur, Paris | Etienne Simon-Lorière; Fabiana Gambaro; Maud Vanpeene; Sylvie Behillil; Sylvie van der Werf; Vincent Enouf |
| EPI_ISL_593913 | Hospital, Talange | National Reference Center for Viruses of Respiratory Infections, Institut Pasteur, Paris | Etienne Simon-Lorière; Fabiana Gambaro; Maud Vanpeene; Sylvie Behillil; Sylvie van der Werf; Vincent Enouf |
| EPI_ISL_450819, EPI_ISL_469059, EPI_ISL_469068 | Hovas Askim Familjelakare och BVC | The Public Health Agency of Sweden | Anna Risberg; Anna Wendel; Anna-Malin Linde; Karin Tegmark-Wisell; Maria Lind Karlberg; Mattias Haukland; Mia Brytting; Olov Svartstrom; Oskar Karlsson Lindsjo; Petra Edquist; Reza Advani; Sandra Broddesson; Shamam Muradrasoli; Theresa Enkirch |
| EPI_ISL_452237, EPI_ISL_452240, EPI_ISL_469062, EPI_ISL_469064 to 469065, EPI_ISL_475513, EPI_ISL_475522 to 475523, EPI_ISL_475525, EPI_ISL_475567 | | | |
| see above | Huddinge VC | The Public Health Agency of Sweden | Anders Johansson; Anna Risberg; Anna-Malin Linde; Karin Tegmark-Wisell; Maria Lind Karlberg; Mattias Haukland; Mia Brytting; Olov Svartstrom; Oskar Karlsson Lindsjo; Petra Edquist; Reza Advani; Sandra Broddesson; Shamam Muradrasoli; Theresa Enkirch |
| EPI_ISL_645000 to 645014, EPI_ISL_645016 to 645028, EPI_ISL_645030 to 645104, EPI_ISL_645106 to 645109 | Human Genome Variation Research Group, Malopolska Centre of Biotechnology | Human Genome Variation Research Group, Malopolska Centre of Biotechnology | Botwina, P.; Branicki, W.; Dabrowska, A.; Foremny, J.; Gromowski, T.; Klajmon, A.; Kopera, K.; Kowalski, M.; Labaj; Marszalek, K.; Owczarek, K.; P.P.; Pisarek, A.; Pospiech, E.; Pyrc, K.; Sanak, M.; Swadzba, J.; Szczepanski, A. |
| EPI_ISL_526215 to 526238 | Hungarian Defence Forces Military Medical Centre | National Laboratory of Virology, Szentágothai Research Centre | Balázs Somogyi; Bálint Eszenyi; Endre Gábor Tóth; Ferenc Jakab; Gábor Kemenesi |
| EPI_ISL_450814, EPI_ISL_452241, EPI_ISL_455103 | Huslakarna Varmbadhuset Varberg | The Public Health Agency of Sweden | Anna Risberg; Anna-Malin Linde; Johanna Hilmersson; Karin Tegmark-Wisell; Maria Lind Karlberg; Mia Brytting; Olov Svartstrom; Oskar Karlsson Lindsjo; Theresa Enkirch |
| EPI_ISL_833191 to 833200 | Hôpital Bichat Claude Bernard, Laboratoire de Virologie | IAME UMR1137 Inserm, Université de Paris, Hôpital Bichat | Alexandre Storto; Amélie Recoing; Antoine Bridier; Benoit Visseaux; Charlotte Charpentier; Diane Descamps; Gilles Collin; Lena Daniel; Mélanie Bertine; Nadhira Houhou-Fidouh; Quentin Le Hingrat; Siham Hamri |
| EPI_ISL_447654 to 447655, EPI_ISL_447657 to 447733 | Hôpital Henri-Mondor Ap-Hp | Hôpital Henri-Mondor Ap-Hp | Canoui-Poitrine, F.; Clinical Study Group, C.; De Prost, N.; Demontant, V.; Deveaux, I.; Fourati, S.; Gricourt, G.; J.-M.; Lamoureux, C.; Lepeule, R.; Mekontso-Dessap, A.; N'debi, M.; P.-L.; Pawlotsky; Picard, O.; Rodriguez, C.; Schmitz, D.; Surgers, L.; Trawinski, E.; Woerther |
| EPI_ISL_415650 | Hôpital Instruction des Armées - BEGIN | National Reference Center for Viruses of Respiratory Infections, Institut Pasteur, Paris | Angela Brisebarre; Christine Bigaillon; Flora Donati Vincent Enouf; Marion Barbet; Maud Vanpeene; Méline Bizard; Mélnie Albert; Sylvie Behillil; Sylvie van der Werf |
| EPI_ISL_443295 to 443299 | Hôpital Necker - Enfants - Malades Laboratoire de Virologie | National Reference Center for Viruses of Respiratory Infections, Institut Pasteur, Paris | Angela Brisebarre; Etienne Simon-Lorière; Flora Donati; Marianne Leruez-Ville; Marion Barbet; Maud Vanpeene; Mélanie Albert; Méline Bizard; Sylvie Behillil; Sylvie van der Werf; Vincent Enouf |
| EPI_ISL_414631 to 414632 | Hôpital Robert Debré Laboratoire de Virologie | National Reference Center for Viruses of Respiratory Infections, Institut Pasteur, Paris | Angela Brisebarre; Flora Donati Vincent Enouf; Laurent Andreoletti; Marion Barbet; Maud Vanpeene; Méline Bizard; Mélnie Albert; Sylvie Behillil; Sylvie van der Werf |
| EPI_ISL_768829 | Hôpital d'Instruction des Armées Sainte-Anne | CNR Virus des Infections Respiratoires - France SUD | Antonin Bal; Bruno Lina; Gregory Destras; Gwendolyne Burfin; Hadrien Règue; Laurence Josset; Martine Valette; Quentin Semanas |
| EPI_ISL_649785, EPI_ISL_649938 to 649939, EPI_ISL_653763 to 653783, EPI_ISL_653787 to 653812, EPI_ISL_653814 to 653823, EPI_ISL_722851 to 722854, EPI_ISL_722859 to 722871, EPI_ISL_722895, EPI_ISL_722898, EPI_ISL_722900 | | | |
| see above | I.R.C.C.S. "S. De Bellis" - Ente Ospedaliero | Istituto Zooprofilattico Sperimentale della Puglia e della Basilicata | Bianco A.; Capozzi L.; Cipolletta D.; Del Sambro L.; Galante D.; Lippolis A.; Manzulli V; Notarnicola M.; Pace L.; Parisi A.; Rondinone V. |
| EPI_ISL_853854 to 853855, EPI_ISL_853914, EPI_ISL_853924 to 853928 | ILV Kärnten | Bergthaler laboratory, CeMM Research Center for Molecular Medicine of the Austrian Academy of Sciences | Alexander Lercher; Alexandra Popa; Andreas Bergthaler; Anna Schedl; Benedikt Agerer; Christoph Bock; Jakob-Wendelin Genger; Jan Laine; Lukas Endler; Martin Senekowitsch; Michael Schuster; Thomas Penz |
| EPI_ISL_419255, EPI_ISL_493328 to 493331, EPI_ISL_590693 to 590698, EPI_ISL_603137 to 603187, EPI_ISL_609989 to 609999, EPI_ISL_882924 to 882925, EPI_ISL_913448, EPI_ISL_983323 | | | |
| see above | INMI Lazzaro Spallanzani IRCCS | INMI Lazzaro Spallanzani IRCCS | A Di Caro; Annunziata Tamburro; Antonino Di Caro; B Bartolini; Barbara Bartolini; Beatrice Valli; C.E.M Gruber; CEM Gruber; Cesare E. M. Gruber; Cesare E.M. Gruber; Cesare EM Gruber; Concetta Castilletti; Daniele Lapa; E Giombini; Eleonora Lalle; Emanuela Giombini; F Messina; Fabrizio Carletti; Francesca Colavita; Francesco Messina; Francesco Vairo; Fulvia Pimpinelli; Giuseppe Ippolito.; Giuseppina Cappiello; M Rueca; MR Capobianchi; Marcello Meledandri; Maria Letizia Schiavone; Maria R Capobianchi; Maria R. Capobianchi; Mariarosa Gaudio; Martina Rueca; O Butera; Ornella Butera; Patrizia Massarelli; Silvia Meschi; Simone Lanini |
| EPI_ISL_410545 to 410546, EPI_ISL_417921 to | INMI Lazzaro Spallanzani IRCCS | Laboratory of Virology, INMI Lazzaro Spallanzani IRCCS | Antonino Di Caro; Barbara Bartolini; Cesare E. M. Gruber; Cesare Ernesto Maria Gruber; Concetta Castilletti; Daniele Lapa; Eleonora Lalle; Emanuela |

| | | | |
|---|---|---|---|
| 417923, EPI_ISL_419254, EPI_ISL_424342 to 424344 | | | Giombini; Emanuele Nicastri; Fabrizio Carletti; Francesca Colavita; Francesco Messina; Francesco Vairo; Giulia Matusali; Giuseppe Ippolito; Giuseppe Ippolito.; Licia Bordi; Maria R. Capobianchi; Maria Rosaria Capobianchi; Martina Rueca |
| EPI_ISL_492184, EPI_ISL_493197, EPI_ISL_493213, EPI_ISL_494756 to 494757, EPI_ISL_494759, EPI_ISL_494761 to 494763, EPI_ISL_494771, EPI_ISL_494774 to 494776, EPI_ISL_775239 to 775240, EPI_ISL_775243 to 775244, EPI_ISL_775246, EPI_ISL_775257, EPI_ISL_775259, EPI_ISL_775261 | | | |
| see above | INT Fondazione Pascale | INT Fondazione Pascale | INT Fondazione Pascale; Pascale |
| EPI_ISL_492980 to 492987, EPI_ISL_751318 to 751447 | IRCCS Sacro Cuore Don Calabria Hospital, Department of Infectious, Tropical Diseases & Microbiology | University of Verona, Department of Biotechnology | Antonio Mori; Chiara Degli Esposti; Chiara Piubelli; Cristina Beltrami; Elena Pomari; Emanuela Cosentino; Giulia Lopatriello; Luca Marcolungo; Massimo Delledonne; Michela Deiana |
| EPI_ISL_480961, EPI_ISL_480975, EPI_ISL_480981, EPI_ISL_480989, EPI_ISL_480995, EPI_ISL_481003, EPI_ISL_481017, EPI_ISL_481025, EPI_ISL_481029, EPI_ISL_481034 to 481035 | | | |
| see above | ISGlobal, Institut de Salut Global de Barcelona | SeqCOVID-SPAIN consortium/IBV(CSIC) | Alberto L Garcia-Basteiro; Alfredo Mayor; Carlota Dobaño; Gemma Moncunill; Pau Cisteró and SeqCOVID-SPAIN consortium |
| EPI_ISL_833076 | IZSAM | Istituto Zooprofilattico Sperimentale dell'Abruzzo e Molise "G. Caporale" | Ancora M; Calistri P; Cammà C; Curini V; Delli Compagni E; Di Domenico M; Di Pasquale A; Lorusso A; Mangone I; Marcacci M; Puglia I; Rinaldi A; Savini G |
| EPI_ISL_572320 to 572324, EPI_ISL_584069, EPI_ISL_584071 to 584072 | IZSM | IZSM | Giovanna Fusco; Lorena Cardillo; Maurizio Viscardi |
| EPI_ISL_900574 to 900690 | IZSM | TIGEM | Andrea Ballabio; Anna Manfredi; Antonio Grimaldi; Antonio Limone; Biancamaria Pierri; Chiara Colantuono; Davide Cacchiarelli.; Denise Di Concilio; Francesco Panariello; Lucio Di Filippo; Marcello Salvi; Maria Concetta Cuomo; Patrizia Annunziata; Pellegrino Cerino; Valentina Bouche |
| EPI_ISL_738121 to 738132 | IZSM-U.O.C. Virologia | Istituto Zooprofilattico Sperimentale del Mezzogiorno | Giovanna Fusco; Lorena Cardillo; Maurizio Viscardi |
| EPI_ISL_683657 to 683670, EPI_ISL_683672 to 683677, EPI_ISL_779715 to 779729, EPI_ISL_779738 to 779739, EPI_ISL_779741 to 779752, EPI_ISL_779754 to 779773, EPI_ISL_855402 to 855403, EPI_ISL_855405, EPI_ISL_855407 | | | |
| see above | IdISSC/Hospital Clínico San Carlos de Madrid | SeqCOVID-SPAIN consortium/IBV(CSIC) | Alberto Delgado-Iribarren; Esther Culebras Lopez; Esther Culebras López; Jorge Matias-Guiu; Luis Ortega Medina; Silvia Sánchez Ramón; Ulises Gómez-Pinedo and SeqCOVID-SPAIN consortium; Vicente Estrada Pérez |
| EPI_ISL_475572 | Imperial College London | Imperial College London | Jie Zhou; Wendy Barclay |
| EPI_ISL_856712 to 856721 | Infectious Diseases Unit, Department of Internal Medicine, Azienda Ospedaliera-Universitaria di Padova | Laboratory of Infectious Diseases, Department of Biomedical and Clinical Sciences L. Sacco, University of Milan | Alessia Lai; Anna Maria Cattelan; Annalisa Bergna; Carla Della Ventura; Claudia Balotta; Davide Leoni; Gianguglielmo Zehender on behalf of SARS-CoV-2 ITALIAN RESEARCH ENTERPRISE-(SCIRE) Collaborative Group; Lolita Sasset; Massimo Galli |
| EPI_ISL_541970 | Influenza Centre, University of Bergen | Norwegian Institute of Public Health, Department of Virology | Bjørn Blomberg; Fan Zhou; Hilde Elshaug; Hilde Synnøve Vollan; Kamilla Heddeland Instefjord; Karl A Brokstad; Karoline Bragstad; Kathrine Stene-Johansen; Olav Hungnes; Rasmus Riis Kopperud; Rebecca J Cox |
| EPI_ISL_470900 to 470904, EPI_ISL_872641 to 872642 | Influenza etiology and epidemiology laboratory | Pathogenic Microorganisms Variability Laboratory | Alexander Gintsburg; Alexey Shchetinin; Andrei Siniavin; Andrey Botikov; Andrey Pochtovyy; Anna Ignatieva; Anna Ignatjeva; Daria Grousova; Denis Logunov; Elena Burtseva; Elena Shidlovskaya; Elizaveta Divisenko; Evgeniya Mukasheva; Evgeny Usachev; Inna Dolzhikova; Kirill Krasnoslobotsev; Ludmila Kolobukhina; Maria Nikiforova; Nadezhda Kuznetsova; Olesya Venchakova; Olga Burgasova; Svetlana Smetanina; Svetlana Trushakova; Valeria Bacalin; Vladimir Gushchin |
| EPI_ISL_469057 | Inger Landgren | The Public Health Agency of Sweden | Anna Risberg; Anna-Malin Linde; Karin Tegmark-Wisell; Maria Lind Karlberg; Mattias Haukland; Olov Svartstrom; Oskar Karlsson Lindsjo; Petra Edquist; Reza Advani; Sandra Broddesson; Shamam Muradrasoli |
| EPI_ISL_635124 to 635125, EPI_ISL_635130 to 635131, EPI_ISL_635183, EPI_ISL_668412 to 668416, EPI_ISL_708030 to 708031, EPI_ISL_708036 to 708039, EPI_ISL_708151 to 708153, EPI_ISL_775440 to 775441, EPI_ISL_860226 to 860228, EPI_ISL_860291 to 860292 | | | |
| see above | Innlandet Hospital Trust, Division Lillehammer, Department for Medical Microbiology | Norwegian Institute of Public Health, Department of Virology | Atiya R Ali; Hilde Elshaug; Hilde Vollan; Kamilla Heddeland Instefjord; Karoline Bragstad; Kathrine Stene-Johansen; Marie Paulsen Madsen; Olav Hungnes; Rasmus Riis Kopperud |
| EPI_ISL_416498, EPI_ISL_428355 to 428357 | Institut Médico légal- Hop R. Poincaré | National Reference Center for Viruses of Respiratory Infections, Institut Pasteur, Paris | Angela Brisebarre; Etienne Simon-Lorière; Flora Donati; Marion Barbet; Maud Vanpeene; Mélanie Albert; Méline Bizard; Mélnie Albert; Sylvie Behillil; Sylvie van der Werf; Vincent Enouf |
| EPI_ISL_416747 to 416748, EPI_ISL_416750, EPI_ISL_416753 to 416754, EPI_ISL_416756, EPI_ISL_416758 | Institut des Agents Infectieux (IAI) Hospices Civils de Lyon | CNR Virus des Infections Respiratoires - France SUD | Alexandre; Antonin; Bal; Bouscambert-Duchamp; Brengel-Pesce; Bruno.; Cheynet; Destras; Florence; Gaymard; Gregory; Josset; Karen; Laurence; Lina; Martine; Maude; Morfin-Sherpa; Valette; Valérie |
| EPI_ISL_417333 to 417337, EPI_ISL_417339, EPI_ISL_418420 to 418425, EPI_ISL_418429 to 418432, EPI_ISL_419169 to 419173, EPI_ISL_419177 to 419182, EPI_ISL_419184, EPI_ISL_420604 to 420611, EPI_ISL_420615 to 420616, EPI_ISL_420618 to 420619, EPI_ISL_420621 to 420625, EPI_ISL_508871 to 508874, EPI_ISL_508877, EPI_ISL_508882 to 508911, EPI_ISL_509007 to 509011, EPI_ISL_509013 to 509014 | | | |
| see above | Institut des Agents Infectieux (IAI), Hospices Civils de Lyon | CNR Virus des Infections Respiratoires - France SUD | Alexandre Gaymard; Antonin Bal; Bruno Lina; Carine Moustaud; Florence Morfin-Sherpa; Gregory Destras; Gwendolyne Burfin; Laurence Josset; Martine Valette; Maude Bouscambert-Duchamp; Raphaelle Lamy; Solenne Brun |
| EPI_ISL_437913 to 437932, EPI_ISL_437974 to 437978, EPI_ISL_437980 to 437981, EPI_ISL_437984 to 437991, EPI_ISL_475763, EPI_ISL_475769, EPI_ISL_475813 to 475814, EPI_ISL_475816, EPI_ISL_475818 to 475820, EPI_ISL_475822 to 475823, EPI_ISL_475825 to 475829, EPI_ISL_475916 to 475927, EPI_ISL_583601 to 583613, EPI_ISL_583615 to 583616, EPI_ISL_583691 | | | |
| see above | Institut für Virologie am Department für Hygiene, Mikrobiologie und Public Health | Bergthaler laboratory, CeMM Research Center for Molecular Medicine of the Austrian Academy of Sciences | Adi Steinrigl; Alexander Lercher; Alexandra Popa; Andreas Bergthaler; Benedikt Agerer; Christian Paar; Christoph Bock; Daniela Schmid; Dorothee von Laer; Elisabeth Puchhammer-Stoeckl; Franz Allerberger; Gernot Walder; Gregor Hörmann; Guenter Weiss; Gunther Vogl; Henrique Colaco; Jakob-Wendelin Genger; Jan Laine; Judith Aberle; Kinga Rigler-Hohenwarter; Lukas Endler; Manfred Nairz; Mark Smyth; Martin Senekowitsch; Michael Schuster; Peter Hufnagl; Peter Obrist; Rainer Gattringer; Sabine Sussitz-Rack; Stephan Aberle; Thomas Penz; Wegene Borena |
| EPI_ISL_485809 to 485813, EPI_ISL_508687 to 508704 | Institut für Virologie und Epidemiologie der Viruskrankheiten, Universitätsklinikum Tübingen | NGS Competence Center Tübingen, Institut für Medizinische Mikrobiologie und Hygiene, Universitätsklinikum Tübingen | Angel Angelov; Angelov at al.; Angelov et al. |
| EPI_ISL_574790 to 574794, EPI_ISL_574796 to 574858, EPI_ISL_574860 to 574862 | Institute for Infectious Diseases, University of Bern | Institute for Infectious Diseases, University of Bern | Alban Ramette; Christian Baumann; Cora Sägesser; Franziska Suter-Riniker; Michel C Koch; Miguel A Terrazos Miani; Peter Keller; Stephen L Leib |
| EPI_ISL_831649 to 831650, EPI_ISL_831652, EPI_ISL_831654, EPI_ISL_831656 to 831659 | Institute for Infectious Diseases, University of Bern, Switzerland | Institute for Infectious Diseases, University of Bern, Switzerland | Alban Ramette; Christian Baumann; Cora Sägesser; Franziska Suter-Riniker; Michel C Koch; Miguel A Terrazos Miani; Pascal Bittel; Peter Keller; Stephen L Leib |
| EPI_ISL_583853, EPI_ISL_583855, EPI_ISL_583857 to 583868, EPI_ISL_853856 to 853873, EPI_ISL_853915 to 853916 | Institute for Laboratory Diagnostics and Microbiology, Klinikum Klagenfurt am Worthersee | Bergthaler laboratory, CeMM Research Center for Molecular Medicine of the Austrian Academy of Sciences | Adi Steinrigl; Alexander Lercher; Alexandra Popa; Andreas Bergthaler; Anna Schedl; Benedikt Agerer; Christian Paar; Christoph Bock; Christoph Bock; Daniela Schmid; Dorothee von Laer; Elisabeth Puchhammer-Stoeckl; Franz Allerberger; Gernot Walder; Gregor Hörmann; Guenter Weiss; Gunther Vogl; Henrique Colaco; Jakob-Wendelin Genger; Jan Laine; Judith Aberle; Kinga Rigler-Hohenwarter; Lukas Endler; Manfred Nairz; Mark Smyth; Martin Senekowitsch; Martin Senekowitsch; Michael Schuster; Michael Schuster; Peter Hufnagl; Peter Obrist; Rainer Gattringer; Sabine Sussitz-Rack; Stephan Aberle; Thomas Penz; Wegene Borena |
| EPI_ISL_677723, EPI_ISL_678251, EPI_ISL_678253 | Institute for Lung Diseases in Children - Skopje | Research Center for Genetic Engineering and Biotechnology "Georgi D. Efremov" , Macedonian Academy of Sciences and Arts | RCGEB - MASA |
| EPI_ISL_583580 to 583598 | Institute for Medical and Chemical Laboratory Diagnostics, Kepler Universitätsklinikum | Bergthaler laboratory, CeMM Research Center for Molecular Medicine of the Austrian Academy of Sciences | Adi Steinrigl; Alexander Lercher; Alexandra Popa; Andreas Bergthaler; Benedikt Agerer; Christian Paar; Christoph Bock; Daniela Schmid; Dorothee von Laer; Elisabeth Puchhammer-Stoeckl; Franz Allerberger; Gernot Walder; Gregor Hörmann; Guenter Weiss; Gunther Vogl; Henrique Colaco; Jakob-Wendelin Genger; Jan Laine; Judith Aberle; Kinga Rigler-Hohenwarter; Lukas Endler; Manfred Nairz; Mark Smyth; Martin Senekowitsch; Michael Schuster; Peter Hufnagl; Peter Obrist; Rainer Gattringer; Sabine Sussitz-Rack; Stephan Aberle; Thomas Penz; Wegene Borena |
| EPI_ISL_429659, EPI_ISL_429663, EPI_ISL_429705, EPI_ISL_429792, EPI_ISL_429802 to 429803, EPI_ISL_429805, EPI_ISL_454605 to 454606, EPI_ISL_455566 to 455567, EPI_ISL_468591, EPI_ISL_468656 | | | |
| see above | Institute for Public Health | Laboratory for advanced genomics | Filip Roki; Igor Jurak; Lovro Trgovec-Greif; Neven Sui; Oliver Vugrek; Tomislav Rukavina |
| EPI_ISL_492075 to 492086 | Institute for Public Health of the Republic of North Macedonia | Charite Universitatsmedizin Berlin, Institute of Virology | Barbara Muhlemann; Christian Drosten; Elizabeta Jancheska; Golubinka Bosevska; Joern Beheim-Schwarzbach; Julia Schneider; Maja Kuzmanovska; Talitha Veith; Terry Jones; Victor M Corman |
| EPI_ISL_728560 to 728561 | Institute for Urban Disease Control and Prevention | COVID-19 Network Investigations (CONI) Alliance | Amornmas Kongklieng; Anek Mungaomklang; Angkana Huang; Anthony R. Jones; Arporn Wangwiwatsin; Bhakbhoom Panthan; Chonticha Klungtong; Ekawat Pasomsub; Elizabeth Batty; Insee Sensorn; Janjira Thaipadungpanit; Kamolthip Atsawawaranunt; Khajohn Joonsalak; Kingkan Rakmanee; |

| | | | |
|---|---|---|---|
| | | | Krittikorn Kumpornsin; Namfon Kotanan; Prayuth Kaewmalang; Pukkaporn Parnwijitkul; Stefan Fernandez; Thanat Chookajorn; Theerarat Kochakarn; Treewat Watthanachockchai; Wasun Chantratita; Wudtichai Manasatienkij |
| EPI_ISL_572330 to 572331 | Institute for Virology, University Hospital Duesseldorf, Medical Faculty, Heinrich-Heine-University Duesseldorf | Institute for Virology, University Hospital Duesseldorf, Medical Faculty, Heinrich-Heine-University Duesseldorf | ; Alexander Killer; Andreas Walker; Annemarie Mohring; Anselm Kunstein; Ansgar Schulz; Björn Jensen; Caroline Klindt; Edwin Bölke; Gerald Antoch; Heiner Schaal; Jennifer Neubert; Johannes Bode; Johannes C. Fischer; Jörg Timm; Lisa Müller; Maximilian Damagnez; Nadine Lübke; Ortwin Adams; Philipp Albrecht; Philipp Ostermann; Saskia Elben; Tina Senff; Tom Lüdde; Torsten Feldt; Verena Keitel |
| EPI_ISL_602462 to 602474, EPI_ISL_602476 to 602498, EPI_ISL_602500 to 602509, EPI_ISL_602518 to 602521, EPI_ISL_602523 to 602532, EPI_ISL_626211 to 626230, EPI_ISL_717977 | | | |
| see above | Institute for Virology, University Hospital Essen | Center of Medical Microbiology, Virology, and Hospital Hygiene, University of Duesseldorf | Alexander Dilthey; Andreas Walker; Daniel Strelow; Jessica Nicolai; Jörg Timm; Klaus Pfeffer; Lisanna Hülse; Malte Kohns Vasconcelos; Maximilian Damagnez; Nadine Lübke; Olympia E. Anastasiou; Tobias Wienemann; Torsten Houwaart; Ulf Dittmer |
| EPI_ISL_525472 to 525473 | Institute of Clinical Microbiology and Hygiene, University Hospital Regensburg | Institute of Clinical Microbiology and Hygiene, University Hospital Regensburg | Hiergeist, A. |
| EPI_ISL_462480, EPI_ISL_485399, EPI_ISL_582031 to 582032 | Institute of Human Genetics, Polish Academy of Sciences | Institute of Human Genetics, Polish Academy of Sciences | Adam Ustaszewski; Andrzej Pawski; Emilia Lis; Ewa Zitkiewicz; Marta Kaczmarek-Ry; Micha Witt; Szymon Hryhorowicz |
| EPI_ISL_485400 | Institute of Human Genetics, Polish Academy of Sciences | Institute of Human Genetics, Polish Academy of Sciences, | Adam Ustaszewski; Andrzej Pawski; Emilia Lis; Ewa Zitkiewicz; Marta Kaczmarek-Ry; Micha Witt; Szymon Hryhorowicz |
| EPI_ISL_450294 | Institute of Human Genetics, Polish Academy of Sciences Sanitary and Epidemiological Station in Pozna | Institute of Human Genetics, Polish Academy of Sciences | Adam Ustaszewski; Andrzej Pawski; Emilia Lis; Marta Kaczmarek-Ry; Micha Witt; Szymon Hryhorowicz |
| EPI_ISL_450295 | Institute of Human Genetics, Polish Academy of Sciences; Sanitary and Epidemiological Station in Pozna | Institute of Human Genetics, Polish Academy of Sciences | Adam Ustaszewski; Andrzej Pawski; Emilia Lis; Marta Kaczmarek-Ry; Micha Witt; Szymon Hryhorowicz |
| EPI_ISL_450338 | Institute of Human Genetics, Polish Academy of Sciences; Sanitary and Epidemiological Station in Pozna | Institute of Human Genetics, Polish Academy of Sciences | Adam Ustaszewski; Andrzej Pawski; Emilia Lis; Marta Kaczmarek-Ry; Micha Witt; Szymon Hryhorowicz |
| EPI_ISL_723077 to 723117 | Institute of Medical Genetics and Applied Genomics | Institute of Medical Genetics and Applied Genomics | Angel Angelov; Caspar Gross; Daniela Bezdan; Michael Bitzer; Michael Sonnabend; Michaela Pogoda; Nicolas Casadei; Siri Göpel; Stephan Ossowski; Thomas Iftner; Tina Ganzenmüller |
| EPI_ISL_803871, EPI_ISL_803874 to 803875, EPI_ISL_864563, EPI_ISL_864566, EPI_ISL_864568 to 864572, EPI_ISL_864574, EPI_ISL_864576, EPI_ISL_864578 | | | |
| see above | Institute of Medical Microbiology and Hospital Hygiene | Institute of Medical Microbiology and Hospital Hygiene | Aljoscha Tersteegen; Prof. Dr. Achim Kaasch |
| EPI_ISL_590823 to 590836 | Institute of Medical Virology, University of Zurich | Institute of Medical Virology, University of Zurich | Alexandra Trkola; Benjamin G. Hale; Idoia Busnadiego; Irene Abela; Marie O. Pohl; Maryam Zaheri; Michael Huber; Silke Stertz; Stefan Schmutz; Verena Kufner |
| EPI_ISL_420294 to 420295, EPI_ISL_420541, EPI_ISL_635200 to 635203, EPI_ISL_635205 to 635219, EPI_ISL_635221 to 635270, EPI_ISL_635272 to 635279, EPI_ISL_635281 to 635294, EPI_ISL_635296 to 635297, EPI_ISL_635299, EPI_ISL_636459 to 636461 | | | |
| see above | Institute of Microbiology and Immunology, Faculty of Medicine, University of Ljubljana | Institute of Microbiology and Immunology, Faculty of Medicine, University of Ljubljana | Lucijan Skubic; Mario Poljak; Miša Korva; Samo Zakotnik; Tatjana Avši - Županc; Tatjana Avši - Županc; Tomaž Mark Zorec |
| EPI_ISL_635204 | Institute of Microbiology and Immunology, Faculty of Medicine, University of Ljubljana | The National Laboratory of Health, Environment and Food - Centre for Medical Microbiology Maribor | Mario Poljak; Miša Korva; Mojca Cimerman; Samo Zakotnik; Tatjana Avši - Županc; Tomaž Mark Zorec |
| EPI_ISL_463008 | Institute of Molecular Virology, University Münster | Institute of Molecular Virology, University Münster | Alexander Mellmann; Angeles Mecate Zambrano; Joachim Kühn; Linda Brunotte; Stephan Ludwig |
| EPI_ISL_577734 to 577735, EPI_ISL_577737 to 577742, EPI_ISL_583481 to 583488, EPI_ISL_717975, EPI_ISL_718250 to 718260, EPI_ISL_779403, EPI_ISL_779651 to 779660, EPI_ISL_788979 to 788988, EPI_ISL_791990 | | | |
| see above | Institute of Virology, Biomedical Research Center of the Slovak Academy of Sciences, Bratislava | Faculty of Natural Sciences, Comenius University, Bratislava | Alena Košálová; Andrej Belák; Boris Klempa; Broa Brejová; Dominika Friová; Edita Staroová; Elena Tichá; Jozef Nosek; Juraj Kopáek; Kristína Boršová; Martina Liková; Martina Neboháová; Monika Sláviková; Peter Sabaka; Sabina Fumaová Havlíková; Tomáš Vina; Viktória Hodorová; Viktória abanová; ubomíra Lukáiková |
| EPI_ISL_417877 to 417880 | Institute of Virology, Biomedical Research Center of the Slovak Academy of Sciences, Bratislava; Public Health Authority of the Slovak Republic, Bratislava | Institute of Virology, Biomedical Research Center of the Slovak Academy of Sciences, Bratislava; Comenius University Science Park, Bratislava | Boris Klempa; Diana Rusáková; Edita Staroová; Elena Tichá; Jaroslav Budiš; Juraj Kopáek; Juraj Koi; Martina Liková; Miroslav Böhmer; Monika Slávikova; Sabina Fumaová Havlíková; Tomáš Szemeš; Werner Krampl |
| EPI_ISL_852633 to 852812 | Institute of Virology, Medical Center, University of Freiburg, Freiburg, Germany | Institute of Virology, Clinial Virus Genomics, Medical Center, University of Freiburg, Freiburg, Germany | Hajo Grundmann; Jonas Fuchs; Lisa Kern; Marcus Panning; Sandra Reuter |
| EPI_ISL_796027 to 796038, EPI_ISL_796041, EPI_ISL_796043, EPI_ISL_796045 to 796048, EPI_ISL_796050 to 796063, EPI_ISL_803866, EPI_ISL_827042 | | | |
| see above | Institute of Virology, University of Cologne | Institute of Virology, University of Cologne | Alex Thielen; Alexander Dilthey; Andreas Walker; Dominik Aschenmeier; Elena Knops; Eva Heger; Gibran Rubio; Jörg Timm; Martin Däumer; Maximilian Damagnez; Rolf Kaiser; Saleta Sierra; Zevanya Tessalonica |
| EPI_ISL_728346, EPI_ISL_728348, EPI_ISL_728351, EPI_ISL_728354, EPI_ISL_728357, EPI_ISL_776711, EPI_ISL_776771 | | | |
| see above | Institute of medical Microbiology and hospital Hygiene | Institute of medical Microbiology and hospital Hygiene | Aljoscha Tersteegen; Prof. Dr. Achim Kaasch |
| EPI_ISL_491174 to 491175, EPI_ISL_491178, EPI_ISL_491180, EPI_ISL_491182 to 491184, EPI_ISL_491186 to 491189, EPI_ISL_491191 to 491192, EPI_ISL_491194, EPI_ISL_491199, EPI_ISL_491202, EPI_ISL_491206 to 491211, EPI_ISL_491216 to 491219, EPI_ISL_491221 to 491222, EPI_ISL_491224 to 491264, EPI_ISL_491266 to 491271, EPI_ISL_491273 to 491280, EPI_ISL_491282 to 491287, EPI_ISL_491289, EPI_ISL_491291 to 491293, EPI_ISL_491297 | | | |
| see above | Instituto Gulbenkian de Ciência | Instituto Gulbenkian de Ciência | Cathy Paulino; Joao Sobral; João Costa; Ricardo Leite; Susana Ladeiro |
| EPI_ISL_424345, EPI_ISL_424348, EPI_ISL_424626 | Instituto Nacional de Enfermedades Respiratorias | Instituto Nacional de Enfermedades Respiratorias | Adnan Araiza Rodríguez; Alejandro Sánchez; Alfredo Ponce de León Garduño; Blanca Taboada; Carlos F. Arias.; Carolina González Torres; Celia Boukadida; Cesar Raúl González Bonilla; Concepción Grajales Muñiz; Edgar Mendieta Condado; Eduardo Becerril Vargas; Fabiola Garcés Ayala; Fernando Ledesma Barrientos; Francisco Javier Gaytán Cervantes; Francisco Pulido; Gisela Barrera Badillo; Gloria Vázquez; Guillermo M. Ruiz-Palacios; Irma López Martínez; Joel Armando Vázquez Pérez; Jorge Salas Hernández; José Arturo Martínez Orozco; José Ernesto Ramírez González; José Esteban Muñoz Medina; Lucia Hernández Rivas; Luis Alberto García Andrade; Mario Mújica Sánchez; Pavel Isa; Pilar Ramos Cervantes; Ricardo Grande; Santiago Avila Ríos; Victor Hugo Borja Aburto; Violeta Ibarra Gonzalez |
| EPI_ISL_412972 | Instituto Nacional de Enfermedades Respiratorias | Instituto de Diagnostico y Referencia Epidemiologicos (INDRE) | Araiza-Rodriguez Adnan; Arias Carlos; Barrera-Badillo Gisela; Boukadida Celia; Garces-Ayala Fabiola; Hernandez-Rivas Lucia; Isa Pavel; Lopez Susana; Lopez-Martinez Irma; Martinez Arturo; Mendieta-Condado Edgar; Munoz-Medina Esteban; Ramirez-Gonzalez Ernesto; Rodriguez-Maldonado Abril; Sanchez Alejandro; Taboada Blanca; Vazquez-Perez Joel; Wong-Arambula Claudia |
| EPI_ISL_421450 to 421452, EPI_ISL_421456, EPI_ISL_421472 to 421478, EPI_ISL_421496 to 421499, EPI_ISL_510889 to 510909, EPI_ISL_510911 to 510983, EPI_ISL_510985 to 511019, EPI_ISL_511021 to 511025, EPI_ISL_511027 to 511104, EPI_ISL_511106 to 511141, EPI_ISL_511144 to 511152, EPI_ISL_511154 to 511170, EPI_ISL_511172 to 511181, EPI_ISL_511189, EPI_ISL_511316, EPI_ISL_511319, EPI_ISL_511322, EPI_ISL_511324, EPI_ISL_511326 to 511343, EPI_ISL_511345 to 511348, EPI_ISL_511350 to 511474, EPI_ISL_511477 to 511483, EPI_ISL_511509 to 511511, EPI_ISL_511513 to 511546, EPI_ISL_511548 to 511549, EPI_ISL_511551, EPI_ISL_511553, EPI_ISL_511556 to 511576, EPI_ISL_511579, EPI_ISL_511581 to 511582, EPI_ISL_511584, EPI_ISL_511591 to 511594, EPI_ISL_511596 to 511598, EPI_ISL_511600, EPI_ISL_511603, EPI_ISL_511605, EPI_ISL_511607 to 511643, EPI_ISL_511645 to 511663, EPI_ISL_511665 to 511680, EPI_ISL_511683 to 511693, EPI_ISL_511695 to 511699, EPI_ISL_511702, EPI_ISL_511704, EPI_ISL_511706, EPI_ISL_511708 to 511711, EPI_ISL_511713 to 511730, EPI_ISL_511732 to 511738, EPI_ISL_511740 to 511744, EPI_ISL_511746 to 511750, EPI_ISL_693517, EPI_ISL_693521 to 693522, EPI_ISL_693525 to 693533, EPI_ISL_693538, EPI_ISL_693540 to 693629, EPI_ISL_731895 to 731896, EPI_ISL_731898 to 731899, EPI_ISL_731901 to 731902, EPI_ISL_731904 to 731905, EPI_ISL_731908 to 731909, EPI_ISL_731911, EPI_ISL_731916, EPI_ISL_731918 to 731923, EPI_ISL_731931 to 731939, EPI_ISL_731941 to 732117, EPI_ISL_738088 to 738110, EPI_ISL_738112 to 738120, EPI_ISL_738146, EPI_ISL_759707 to 759709, EPI_ISL_759711, EPI_ISL_759713 to 759716, EPI_ISL_759718 to 759721, EPI_ISL_759723, EPI_ISL_759761, EPI_ISL_765210 to 765215, EPI_ISL_765219 to 765221, EPI_ISL_796734 to 796735, EPI_ISL_796737 to 796754, EPI_ISL_796759 to 796761, EPI_ISL_796763 to 796764, EPI_ISL_796772 to 796773, EPI_ISL_796776 to 796781, EPI_ISL_801517 to 801518, EPI_ISL_801521, EPI_ISL_861530 to 861534, EPI_ISL_861537 to 861538, EPI_ISL_861552, EPI_ISL_861556, EPI_ISL_861567 to 861571, EPI_ISL_861581 to 861591 | | | |
| see above | Instituto Nacional de Saude (INSA) | Instituto Nacional de Saude (INSA) | Borges et al; Guiomar et al |
| EPI_ISL_511484 to 511490, EPI_ISL_511492 to 511508, EPI_ISL_511751 to 511757, EPI_ISL_511759 to 511771 | Instituto Nacional de Saude (INSA) | Instituto Nacional de Saude (INSA) and Instituto Gulbenkian de Ciencia (IGC) | Borges et al |
| EPI_ISL_511182 to 511188, EPI_ISL_511190 to 511226, EPI_ISL_511228 to 511257, EPI_ISL_511259, EPI_ISL_511261 to 511262, EPI_ISL_511264 to 511265, EPI_ISL_511267 to 511277, EPI_ISL_511280 to 511281, EPI_ISL_511283 to 511302, EPI_ISL_511304, EPI_ISL_511307 to 511308, EPI_ISL_511310 to 511315, EPI_ISL_731897, EPI_ISL_731900, EPI_ISL_731903, EPI_ISL_731906 to 731907, EPI_ISL_731912, EPI_ISL_731917, EPI_ISL_731924 to 731930, EPI_ISL_731940, EPI_ISL_732118 to 732293, EPI_ISL_765216 to 765218 | | | |
| see above | Instituto Nacional de Saude (INSA) and Instituto Gulbenkian de | Instituto Nacional de Saude (INSA) and Instituto Gulbenkian | Borges et al |

| | | | |
|---|---|---|---|
| | Ciencia (IGC) | de Ciencia (IGC) | |
| EPI_ISL_452139, EPI_ISL_452141 | Instituto de Diagnostico y Referencia Epidemiologicos (INDRE) | Instituto de diagnóstico y Referencia Epidemiologicos (INDRE) | Araiza-Rodriguez Adnan; Barrera-Badillo Gisela; Garces-Ayala Fabiola; Hernandez-Rivas Lucia; Lopez-Martinez Irma; Lopez-Martinez Irma.; Mendieta-Condado Edgar; Ramirez-Gonzalez Ernesto; Rodriguez-Maldonado Abril; Wong-Arambula Claudia |
| EPI_ISL_510064, EPI_ISL_510066 to 510069, EPI_ISL_510071 to 510075, EPI_ISL_510077, EPI_ISL_510462 to 510466 | Instituto de Investigaciones Biomédicas de Barcelona (CSIC), Hospital Clinic i Provincial de Barcelona, Instituto de Biomedicina de Valencia (CSIC), Hospital de Sant Pau | SeqCOVID-SPAIN consortium/IBV(CSIC) | Alex Soriano; Andrea Vergara; Anna M. Planas; Israel Fernández Cadenas and SeqCOVID-SPAIN consortium; Jordi Pérez Tur; Miguel J. Martínez; Mª Angeles Marcos |
| EPI_ISL_871972, EPI_ISL_871996 to 871998 | Instituto de Salud Carlos III | Instituto de Salud Carlos III | A. Monzón; F. Casas; I. Jiménez; Iglesias-Caballero; M. Camarero; M. Cuesta, I.; M. González-Esguevillas; M. Pozo; M. Zaballos; P. Jiménez; S. Juliá; S. Molinero Calamita; S. Varona |
| EPI_ISL_812250 | Invenimus AG | Institute of Medical Virology, University of Zurich | Alexandra Trkola; Annette Audigé; Cyril Shah; Guido Bloemberg; Jon Huder; Jürg Böni; Kevin Steiner; Maria Grünberg; Maryam Zaheri; Michael Huber; Riccarda Capaul; Stefan Schmutz; Verena Kufner |
| EPI_ISL_824407 to 824408 | Istituto Nazionale Malattie Infettive Lazzaro Spallanzani IRCCS | Istituto Nazionale Malattie Infettive Lazzaro Spallanzani IRCCS | Antonino Di Caro; Barbara Bartolini; Cesare E.M. Gruber; Concetta Castilletti; Emanuela Giombini; Francesca Colavita; Francesco Messina; Maria R. Capobianchi; Martina Rueca; Ornella Butera; Silvia Meschi |
| EPI_ISL_477193 to 477194, EPI_ISL_477202 to 477203 | Istituto Zooprofilattico Sperimentale Puglia e Basilicata; | Beaconlab (Bioinformatics, Evolution and Comparative Genomics lab), Dept of Biosciences, University on Mila | Chiara M.; Manzari C.; Parisi A.; Pesole G. |
| EPI_ISL_477195 to 477201 | Istituto Zooprofilattico Sperimentale Puglia e Basilicata; | Beaconlab (Bioinformatics, Evolution and Comparative Genomics lab), Dept of Biosciences, University on Milan | Chiara M.; Manzari C.; Parisi A.; Pesole G. |
| EPI_ISL_451961 | Istituto Zooprofilattico Sperimentale Puglia e Basilicata; Dipartimento di Bioscienze, Biotecnologie e Biofarmaceutica dell'Università degli Studi di Bari "A.Moro"; Istituto di Biomembrane. Bioenergetica e Biotecnologie Molecolari del Consiglio Nazionale delle Ricerche di Bari | Beaconlab (Bioinformatics Evolution and Comparative Genomics lab), Dept of Biosciences, University of Milan | Chiara M.; Manzari C.; Parisi A.; Pesole G. |
| EPI_ISL_451962 | Istituto Zooprofilattico Sperimentale Puglia e Basilicata; Dipartimento di Bioscienze, Biotecnologie e Biofarmaceutica dell'Università degli Studi di Bari "A.Moro"; Istituto di Biomembrane. Bioenergetica e Biotecnologie Molecolari del Consiglio Nazionale delle Ricerche di Bari | Beaconlab (Bioinformatics, Evolution and Comparative Genomics lab), Dept of Biosciences, University on Milan | Chiara M.; Manzari C.; Parisi A.; Pesole G. |
| EPI_ISL_468914, EPI_ISL_469016, EPI_ISL_469018 to 469022, EPI_ISL_469050, EPI_ISL_525553 to 525571, EPI_ISL_525573, EPI_ISL_527380 | | | |
| see above | Istituto Zooprofilattico Sperimentale Puglia e Basilicata; Dipartimento di Bioscienze, Biotecnologie e Biofarmaceutica dell'Università degli Studi di Bari "A.Moro"; Istituto di Biomembrane. Bioenergetica e Biotecnologie Molecolari del Consiglio Nazionale delle Ricerche di Bari | Beaconlab (Bioinformatics, Evolution and Comparative Genomics lab), Dept of Biosciences, University on Milan | Chiara M; Chiara M.; Manzari C.; Parisi A.; Pesole G. |
| EPI_ISL_493332 to 493333, EPI_ISL_560407, EPI_ISL_882923, EPI_ISL_913449 | Istituto Zooprofilattico Sperimentale del Mezzogiorno | INMI Lazzaro Spallanzani IRCCS | A Di Caro; Antonino Di Caro; B Bartolini; B Pierri; Barbara Bartolini; C Buonerba; C.E.M Gruber; Cesare E.M. Gruber; Cesare EM Gruber; Concetta Castilletti; D Di Concilio; E Giombini; Emanuela Giombini; F Messina; Francesca Colavita; Francesco Messina; Giorgia Borriello; Giovanna Fusco; M. Rueca; MC Cuomo; MR Capobianchi; Maria R Capobianchi; Maria R. Capobianchi; Martina Rueca; Maurizio Viscardi; O Butera; Ornella Butera; P Cerino; Sergio Brandi; Silvia Meschi |
| EPI_ISL_736996 to 736997 | Istituto Zooprofilattico Sperimentale del Mezzogiorno | National Institute for Infectious Diseases, INMI, "L. Spallanzani" IRCCS | A. Di Caro; B. Bartolini; B. Pierri; C. Buonerba; C.E.M Gruber; D. Di Concilio; E. Giombini; F. Carletti; F. Messina; M. Rueca; M.C. Cuomo; M.R Capobianchi; M.R. Capobianchi; P. Cerino |
| EPI_ISL_736777 to 736890, EPI_ISL_776879 to 776993, EPI_ISL_776995 to 777009, EPI_ISL_778646 to 778817, EPI_ISL_833522 to 833570, EPI_ISL_837247 to 837330, EPI_ISL_837433 to 837504, EPI_ISL_960442 to 960447, EPI_ISL_960449 to 960458, EPI_ISL_960460, EPI_ISL_960463 to 960464, EPI_ISL_960466 to 960469, EPI_ISL_960471 to 960571, EPI_ISL_960574 to 960661 | | | |
| see above | Istituto Zooprofilattico Sperimentale del Mezzogiorno | TIGEM | Andrea Ballabio; Anna Manfredi; Antonio Grimaldi; Antonio Limone; Biancamaria Pierri; Chiara Colantuono; Davide Cacchiarelli.; Denise Di Concilio; Francesco Panariello; Lucio Di Filippo; Marcello Salvi; Maria Concetta Cuomo; Patrizia Annunziata; Pellegrino Cerino; Valentina Bouche |
| EPI_ISL_602304 | Istituto Zooprofilattico Sperimentale del Mezzogiorno | U.O. Diagnostica Virologica Dip. Sanità Animale IZSM | Lorena Cardillo; Maurizio Viscardi; e Giovanna Fusco |
| EPI_ISL_763069, EPI_ISL_763071 to 763073, EPI_ISL_763094 to 763098, EPI_ISL_763138, EPI_ISL_763318, EPI_ISL_763320, EPI_ISL_763322 to 763330 | | | |
| see above | Istituto Zooprofilattico Sperimentale dell' Umbria e delle Marche -Togo Rosati | Istituto Superiore di Sanità | Gabriele Vaccari; Giovanni Ianiro; Ilaria Di Bartolo; Luca De Sabato; Massimo Biagetti; Monica Giammarioli |
| EPI_ISL_649189 to 649191, EPI_ISL_649940, EPI_ISL_653784 to 653786, EPI_ISL_653813, EPI_ISL_722873 to 722894, EPI_ISL_722901 to 722911, EPI_ISL_722913 to 722925, EPI_ISL_794745, EPI_ISL_794749, EPI_ISL_794752, EPI_ISL_794754 to 794755, EPI_ISL_794759 to 794817, EPI_ISL_876738 to 876740, EPI_ISL_876744 to 876813 | | | |
| see above | Istituto Zooprofilattico Sperimentale della Puglia e della Basilicata | Istituto Zooprofilattico Sperimentale della Puglia e della Basilicata | Bianco A.; Capozzi L.; Cipolletta D.; Del Sambro L.; Galante D.; Manzulli V; Pace L.; Parisi A.; Rondinone V. |
| EPI_ISL_422427 | JABER AL AHMAD AL SABAH HOSPITAL - KUWAIT CITY | Dasman Diabetes Institute | Ebaa Al-Ozairi; Fahd Al-Mulla; Qais Al-Duwairi; Rasheeba Iqbal; Sumi John |
| EPI_ISL_450821 to 450822 | Jarpens HC | The Public Health Agency of Sweden | Anna Risberg; Anna-Malin Linde; Gunilla Johansson; Karin Tegmark-Wisell; Maria Lind Karlberg; Mia Brytting; Olov Svartstrom; Oskar Karlsson Lindsjo; Theresa Enkirch |
| EPI_ISL_728288 | Jena University Hospital, Institute for Infectious Diseases and Infection Control | Institute for infectious medicine & hospital hygiene, CaSe-Group | Brandt; Christian; Marquet; Matthias W.; Mike; Pletz; Riccardo; Spott |
| EPI_ISL_728292 to 728293, EPI_ISL_728296, EPI_ISL_728302 to 728303, EPI_ISL_728321, EPI_ISL_728323, EPI_ISL_728331, EPI_ISL_728333, EPI_ISL_728335, EPI_ISL_728338, EPI_ISL_728341, EPI_ISL_763077, EPI_ISL_763086 to 763087, EPI_ISL_763089 to 763093, EPI_ISL_768633, EPI_ISL_768725, EPI_ISL_768727 | | | |
| see above | Jena University Hospital, Institute for Infectious Diseases and Infection Control | Institute of infectious medicine & hospital hygiene, CaSe-Group | Brandt; Christian; Marquet; Matthias W.; Mike; Pletz; Riccardo; Spott |
| EPI_ISL_632938 | Jena University Hospital; Institute for Infectious Diseases and Infection Control | Institute of infectious medicine & hospital hygiene, CaSe-Group | Brandt; Christian; Marquet; Matthias W.; Mike; Pletz; Riccardo; Spott |
| EPI_ISL_445242 | Jokkmokks Halsocentral | The Public Health Agency of Sweden | Anna Risberg; Anna-Malin Linde; Karin Tegmark-Wisell; Maria Lind Karlberg; Markus Beland; Mia Brytting; Olov Svartstrom; Oskar Karlsson Lindsjo; Theresa Enkirch |
| EPI_ISL_455101, EPI_ISL_469055 to 469056 | Jourcentralen | The Public Health Agency of Sweden | Anna Risberg; Anna-Malin Linde; Karin Tegmark-Wisell; Maria Lind Karlberg; Mattias Haukland; Mia Brytting; Olov Svartstrom; Oskar Karlsson Lindsjo; Petra Edquist; Reza Advani; Salvatore Ascione; Sandra Broddesson; Shamam Muradrasoli; Theresa Enkirch |
| EPI_ISL_415154 | KU Leuven, Clincal and Epidemiological Virology | KU Leuven, Clincal and Epidemiological Virology | Bert Vanmechelen; Joan Marti-Careras; Marc Van Ranst; Piet Maes.; Tony Wawina |
| EPI_ISL_415153, EPI_ISL_415155 to 415159, EPI_ISL_416467 to 416472, EPI_ISL_416475 to 416476, EPI_ISL_417422, EPI_ISL_417424 to 417430, EPI_ISL_418270, EPI_ISL_418792 to 418798, EPI_ISL_418800, EPI_ISL_418805 to 418806, EPI_ISL_418863, EPI_ISL_418981 to 418987, EPI_ISL_418989, EPI_ISL_420314 to 420345, EPI_ISL_420347, EPI_ISL_420349 to 420368, EPI_ISL_420371, EPI_ISL_420373 to 420454 | | | |
| see above | KU Leuven, Clinical and Epidemiological Virology | KU Leuven, Clinical and Epidemiological Virology | Bert Vanmechelen; Joan Marti-Carerras; Joan Marti-Carreras; Marc Van Ranst; Piet Maes; Tony Wawina |
| EPI_ISL_458156 to 458234, EPI_ISL_462151 to 462275, EPI_ISL_464065 to 464090, EPI_ISL_476941 to 477007, EPI_ISL_522349 to 522350, EPI_ISL_845568 to 845570, EPI_ISL_845572, EPI_ISL_845578, EPI_ISL_845580, EPI_ISL_845583, EPI_ISL_845585, EPI_ISL_845587 to 845589, EPI_ISL_845592, EPI_ISL_845594, EPI_ISL_845596, EPI_ISL_845602, EPI_ISL_845604, EPI_ISL_845606, EPI_ISL_845608, EPI_ISL_845610 to 845611, EPI_ISL_845614 to 845615 | | | |

| | | | |
|---|---|---|---|
| see above | KU Leuven, Rega Institute, Clinical and Epidemiological Virology | KU Leuven, Rega Institute, Clinical and Epidemiological Virology | Bert Vanmechelen; Joan Marti-Carerras; Piet Maes; Tony Wawina-Bokalanga |
| EPI_ISL_495411 to 495450, EPI_ISL_495452 to 495457 | Kafkas University, Faculty of Medicine, Department of Medical Microbiology | Kafkas University, Faculty of Medicine, Department of Medical Microbiology | Didem Ozgur; E. Ediz Tutuncu; Murat Karamese |
| EPI_ISL_475137 to 475138, EPI_ISL_510825 to 510826, EPI_ISL_510832 to 510833, EPI_ISL_510836 to 510837, EPI_ISL_510859, EPI_ISL_510864 to 510865, EPI_ISL_623091, EPI_ISL_648164, EPI_ISL_676534, EPI_ISL_710605 | | | |
| see above | Kalmar klinisk mikrobiologi | The Public Health Agency of Sweden | Anna Risberg; Anna-Malin Linde; Department of Microbiology; Karin Tegmark-Wisell; Maria Lind Karlberg; Mattias Haukland; Mia Brytting; Olov Svartstrom; Oskar Karlsson Lindsjo; Petra Edquist; Reza Advani; Sandra Broddesson; Shamam Muradrasoli; The Public Health Agency of Sweden |
| EPI_ISL_872571 | Kamchatka Regional Children's Infectious Diseases Hospital | WHO National Influenza Centre Russian Federation | Andrey Komissarov; Anna Ivanova; Artem Fadeev; Daria Danilenko; Dmitry Bazhenov; Dmitry Lioznov; Elena Nabieva; Georgii Bazykin; Ksenia Safina; Kseniya Komissarova; Mikhail Bakaev |
| EPI_ISL_444487 to 444488, EPI_ISL_444490 to 444492 | Karolinska Universitetslaboratoriet | CTMR, Karolinska Institutet, Stockholm, Sweden | Caroline Bjurnemark; Fredrik Boulund; Jing Wang; Jingkai Ji; Lars Engstrand; Linnéa Påvénius; Marica Hamsten; Shuiqin Li; Stefanie Prast-Nielsen; Vivien Lan Yang Swartz; Yinghua Zha; Yue Hu |
| EPI_ISL_454447 to 454493, EPI_ISL_454868 to 454902, EPI_ISL_455846 to 455901, EPI_ISL_469075, EPI_ISL_469077 to 469079, EPI_ISL_475086 to 475091, EPI_ISL_475095, EPI_ISL_475136, EPI_ISL_475141 to 475142, EPI_ISL_475149 to 475150, EPI_ISL_475544 to 475547, EPI_ISL_475551 to 475552, EPI_ISL_475558 to 475561, EPI_ISL_510834 to 510835, EPI_ISL_510838 to 510840, EPI_ISL_510852, EPI_ISL_510860 to 510863, EPI_ISL_560974, EPI_ISL_615086 to 615089, EPI_ISL_676533 | | | |
| see above | Karolinska Universitetslaboratoriet | The Public Health Agency of Sweden | Anna Risberg; Anna-Malin Linde; Department of Microbiology; Karin Tegmark-Wisell; Maria Lind Karlberg; Mattias Haukland; Mia Brytting; Olov Svartstrom; Oskar Karlsson Lindsjo; Petra Edquist; Reza Advani; Sandra Broddesson; Shamam Muradrasoli; Shaman Muradrasoli; The Public Health Agency of Sweden |
| EPI_ISL_534259, EPI_ISL_548257 to 548258, EPI_ISL_560989 to 560990 | Karolinska universitetslaboratoriet | The Public Health Agency of Sweden | Anna Risberg; Anna-Malin Linde; Karin Tegmark-Wisell; Maria Lind Karlberg; Mattias Haukland; Mia Brytting; Olov Svartstrom; Oskar Karlsson Lindsjo; Petra Edquist; Reza Advani; Sandra Broddesson |
| EPI_ISL_534234 to 534236, EPI_ISL_560982 to 560985 | Karolinska universitetslaboratoriet SOLNA | The Public Health Agency of Sweden | Anna Risberg; Anna-Malin Linde; Karin Tegmark-Wisell; Maria Lind Karlberg; Mattias Haukland; Mia Brytting; Olov Svartstrom; Oskar Karlsson Lindsjo; Petra Edquist; Reza Advani; Sandra Broddesson |
| EPI_ISL_813655 to 813656 | Kettering General Hospital | COVID-19 Genomics UK (COG-UK) Consortium | Christopher Moore; Fei Sang; Johnny Debebe; Jonathan Ball; Joseph Chappell; Matthew Carlisle; Matthew Loose; Nadine Holmes; Patrick McClure; Theocharis Tsoleridis; Victoria Wright |
| EPI_ISL_413021 | Klinik Hirslanden Zurich | Institute of Medical Virology, University of Zurich | Alexandra Trkola; Andrea Zbinden; Christian Ruef; Fiona Steiner; Gabriela Ziltener; Jon Huder; Jürg Böni; Maryam Zaheri; Michael Huber; Patrick Redli; Riccarda Capaul; Stefan Schmutz; Verena Kufner |
| EPI_ISL_524479 | Klinik Hirslanden Zürich | Institute of Medical Virology, University of Zurich | Alexandra Trkola; Andrea Zbinden; Fiona Steiner; Gabriela Ziltener; Jon Huder; Jürg Böni; Maryam Zaheri; Michael Huber; Patrick Redli; Riccarda Capaul; Stefan Schmutz; Verena Kufner |
| EPI_ISL_475910 to 475915 | Klinikum Wels-Grieskirchen | Bergthaler laboratory, CeMM Research Center for Molecular Medicine of the Austrian Academy of Sciences | Alexander Lercher; Alexandra Popa; Andreas Bergthaler; Benedikt Agerer; Christoph Bock; Daniela Schmid; Dorothee von Laer; Elisabeth Puchhammer-Stoeckl; Franz Allerberger; Gregor Hörmann; Guenter Weiss; Henrique Colaco; Jakob-Wendelin Genger; Jan Laine; Judith Aberle; Kinga Rigler-Hohenwarter; Lukas Endler; Manfred Nairz; Mark Smyth; Martin Senekowitsch; Michael Schuster; Peter Hufnagl; Rainer Gattringer; Stephan Aberle; Thomas Penz; Wegene Borena |
| EPI_ISL_475101 to 475104, EPI_ISL_476140 to 476142, EPI_ISL_582808, EPI_ISL_615108, EPI_ISL_654954, EPI_ISL_660427 to 660431, EPI_ISL_661301, EPI_ISL_676492, EPI_ISL_676497 to 676501, EPI_ISL_676518, EPI_ISL_766696, EPI_ISL_766713, EPI_ISL_775546 to 775548, EPI_ISL_789017 | | | |
| see above | Klinisk Mikrobiologi | The Public Health Agency of Sweden | Anna Risberg; Anna-Malin Linde; Department of Microbiology; Karin Tegmark-Wisell; Maria Lind Karlberg; Mattias Haukland; Mia Brytting; Olov Svartstrom; Oskar Karlsson Lindsjo; Petra Edquist; Reza Advani; Sandra Broddesson; Shamam Muradrasoli; The Public Health Agency of Sweden |
| EPI_ISL_654892 to 654899, EPI_ISL_654942, EPI_ISL_654944 to 654951, EPI_ISL_660416 to 660421, EPI_ISL_661274 to 661278, EPI_ISL_661280 to 661281, EPI_ISL_661288 to 661289, EPI_ISL_661293, EPI_ISL_661302 to 661303, EPI_ISL_676489 to 676491, EPI_ISL_676494 to 676496, EPI_ISL_676502 to 676508, EPI_ISL_676520, EPI_ISL_676531 to 676532, EPI_ISL_710595 to 710604, EPI_ISL_732654, EPI_ISL_766592 to 766593, EPI_ISL_766608 to 766611, EPI_ISL_766613, EPI_ISL_766615 to 766629, EPI_ISL_766633 to 766639, EPI_ISL_766694, EPI_ISL_766707, EPI_ISL_766710 to 766712, EPI_ISL_766714, EPI_ISL_766725 to 766730, EPI_ISL_775549, EPI_ISL_775572 to 775574, EPI_ISL_775584, EPI_ISL_775588, EPI_ISL_775590 to 775592, EPI_ISL_788989 to 788990, EPI_ISL_789036 to 789045, EPI_ISL_789047, EPI_ISL_831958, EPI_ISL_831973, EPI_ISL_831980 to 831981, EPI_ISL_831990 to 831992, EPI_ISL_831996 to 831997, EPI_ISL_832001 to 832002 | | | |
| see above | Klinisk mikrobiologi | The Public Health Agency of Sweden | Anna Risberg; Anna-Malin Linde; Department of Microbiology; Karin Tegmark-Wisell; Maria Lind Karlberg; Mattias Haukland; Mia Brytting; Olov Svartstrom; Oskar Karlsson Lindsjo; Petra Edquist; Reza Advani; Sandra Broddesson; The Public Health Agency of Sweden |
| EPI_ISL_615097 to 615100, EPI_ISL_623092 to 623094 | Klinisk mikrobiologi Lanssjukhuset Ryhov, Jonkoping | The Public Health Agency of Sweden | Anna Risberg; Anna-Malin Linde; Karin Tegmark-Wisell; Maria Lind Karlberg; Mattias Haukland; Mia Brytting; Olov Svartstrom; Oskar Karlsson Lindsjo; Petra Edquist; Reza Advani; Sandra Broddesson |
| EPI_ISL_623089 | Klinisk mikrobiologi Linkoping | The Public Health Agency of Sweden | Anna Risberg; Anna-Malin Linde; Karin Tegmark-Wisell; Maria Lind Karlberg; Mattias Haukland; Mia Brytting; Olov Svartstrom; Oskar Karlsson Lindsjo; Petra Edquist; Reza Advani; Sandra Broddesson |
| EPI_ISL_510841 to 510844, EPI_ISL_510868 to 510870, EPI_ISL_548250 to 548251, EPI_ISL_548253 to 548254, EPI_ISL_560973, EPI_ISL_615090 to 615091, EPI_ISL_615109, EPI_ISL_623087 to 623088, EPI_ISL_648169, EPI_ISL_648173, EPI_ISL_831974 | | | |
| see above | Klinisk mikrobiologi NAL Trollhattan | The Public Health Agency of Sweden | Anna Risberg; Anna-Malin Linde; Department of Microbiology; Karin Tegmark-Wisell; Maria Lind Karlberg; Mattias Haukland; Mia Brytting; Olov Svartstrom; Oskar Karlsson Lindsjo; Petra Edquist; Reza Advani; Sandra Broddesson; The Public Health Agency of Sweden |
| EPI_ISL_429136 to 429148, EPI_ISL_429150 to 429158, EPI_ISL_429160, EPI_ISL_430860 to 430861, EPI_ISL_430863 | Klinisk mikrobiologi Orebro | The Public Health Agency of Sweden | Anna Risberg; Anna-Malin Linde; Karin Tegmark-Wisell; Maria Lind Karlberg; Martin Sundqvist; Olov Svartstrom; Oskar Karlsson Lindsjo; Shaman Muradrasoli |
| EPI_ISL_582806 to 582807, EPI_ISL_648193 | Klinisk mikrobiologi SAS Boras | The Public Health Agency of Sweden | Anna Risberg; Anna-Malin Linde; Karin Tegmark-Wisell; Maria Lind Karlberg; Mattias Haukland; Mia Brytting; Olov Svartstrom; Oskar Karlsson Lindsjo; Petra Edquist; Reza Advani; Sandra Broddesson |
| EPI_ISL_475144 to 475148, EPI_ISL_475153 to 475155, EPI_ISL_475160 to 475163, EPI_ISL_510818 to 510819, EPI_ISL_510853 to 510858, EPI_ISL_582797 to 582798, EPI_ISL_582800, EPI_ISL_582835 to 582838, EPI_ISL_623090, EPI_ISL_648162, EPI_ISL_654504 to 654505, EPI_ISL_654957, EPI_ISL_710594 | | | |
| see above | Klinisk mikrobiologi Vasternorrland | The Public Health Agency of Sweden | Anna Risberg; Anna-Malin Linde; Department of Microbiology; Karin Tegmark-Wisell; Maria Lind Karlberg; Mattias Haukland; Mia Brytting; Olov Svartstrom; Oskar Karlsson Lindsjo; Petra Edquist; Reza Advani; Sandra Broddesson; Shamam Muradrasoli; The Public Health Agency of Sweden |
| EPI_ISL_510820 to 510824, EPI_ISL_510867, EPI_ISL_548252, EPI_ISL_560975, EPI_ISL_648133 | Klinisk mikrobiologi centralsjukhuset Karlstad | The Public Health Agency of Sweden | Anna Risberg; Anna-Malin Linde; Karin Tegmark-Wisell; Maria Lind Karlberg; Mattias Haukland; Mia Brytting; Olov Svartstrom; Oskar Karlsson Lindsjo; Petra Edquist; Reza Advani; Sandra Broddesson |
| EPI_ISL_429115 to 429125, EPI_ISL_430848 to 430855, EPI_ISL_450834 | Klinisk mikrobiologi och vardhygien Halmstad | The Public Health Agency of Sweden | Anna Risberg; Anna-Malin Linde; Arne Kotz; Karin Tegmark-Wisell; Maria Lind Karlberg; Mia Brytting; Olov Svartstrom; Oskar Karlsson Lindsjo; Shaman Muradrasoli; Theresa Enkirch |
| EPI_ISL_766692 to 766693, EPI_ISL_789012 to 789013, EPI_ISL_789029 to 789035 | Klinisk mikrobiologi, Laboratoriemedicin | The Public Health Agency of Sweden | Department of Microbiology; The Public Health Agency of Sweden |
| EPI_ISL_534256, EPI_ISL_560988 | Klinisk mikrobiologi, Laboratoriemedicin Gavleborg | The Public Health Agency of Sweden | Anna Risberg; Anna-Malin Linde; Karin Tegmark-Wisell; Maria Lind Karlberg; Mattias Haukland; Mia Brytting; Olov Svartstrom; Oskar Karlsson Lindsjo; Petra Edquist; Reza Advani; Sandra Broddesson |
| EPI_ISL_424703, EPI_ISL_428148, EPI_ISL_428201, EPI_ISL_548962 to 548963, EPI_ISL_549024, EPI_ISL_549026, EPI_ISL_549176 | | | |
| see above | Klinisk mikrobiologi, Region Västerbotten | Unit for Biological Agents, Department for CBRN Defence and Security, Swedish Defence Research Agency | FOI Bioinformatics team; FOI bioinformatics team |
| EPI_ISL_615080 to 615085, EPI_ISL_623080 to 623085 | Klinisk mikrobiologi, Skanes universitetssjukhus, Lund | The Public Health Agency of Sweden | Anna Risberg; Anna-Malin Linde; Karin Tegmark-Wisell; Maria Lind Karlberg; Mattias Haukland; Mia Brytting; Olov Svartstrom; Oskar Karlsson Lindsjo; Petra Edquist; Reza Advani; Sandra Broddesson |
| EPI_ISL_454903, EPI_ISL_455904 to 455907 | Klinisk mikrobiologi, UAS | The Public Health Agency of Sweden | Anna Risberg; Anna-Malin Linde; Karin Tegmark-Wisell; Maria Lind Karlberg; Mattias Haukland; Olov Svartstrom; Oskar Karlsson Lindsjo; Petra Edquist; Reza Advani; Shamam Muradrasoli |
| EPI_ISL_661290, EPI_ISL_676515, EPI_ISL_766630 to 766632, EPI_ISL_775531 to 775532, EPI_ISL_775542 to 775545, EPI_ISL_789048 to 789053, EPI_ISL_789055 to 789059, EPI_ISL_831979 | | | |

| | | | |
|---|---|---|---|
| see above | Klinisk mikrobiologi, Viruslab | The Public Health Agency of Sweden | Department of Microbiology; The Public Health Agency of Sweden |
| EPI_ISL_766697 | Klinisk mikrobiologi, bakteriologi | The Public Health Agency of Sweden | Department of Microbiology; The Public Health Agency of Sweden |
| EPI_ISL_788999, EPI_ISL_831945 to 831946 | Klinisk mikrobiologi, virus F68 | The Public Health Agency of Sweden | Department of Microbiology; The Public Health Agency of Sweden |
| EPI_ISL_534245, EPI_ISL_560987 | Kliniskt mikrobiologiska laboratoriet | The Public Health Agency of Sweden | Anna Risberg; Anna-Malin Linde; Karin Tegmark-Wisell; Maria Lind Karlberg; Mattias Haukland; Mia Brytting; Olov Svartstrom; Oskar Karlsson Lindsjo; Petra Edquist; Reza Advani; Sandra Broddesson |
| EPI_ISL_510871 to 510877, EPI_ISL_560976, EPI_ISL_582771 to 582775, EPI_ISL_594149 to 594153, EPI_ISL_615110, EPI_ISL_615117 to 615119, EPI_ISL_654952, EPI_ISL_660384 to 660414, EPI_ISL_661279, EPI_ISL_661284 to 661285, EPI_ISL_676512, EPI_ISL_676519 | | | |
| see above | Klinsisk mikrobiologi Linkoping | The Public Health Agency of Sweden | Anna Risberg; Anna-Malin Linde; Department of Microbiology; Karin Tegmark-Wisell; Maria Lind Karlberg; Mattias Haukland; Mia Brytting; Olov Svartstrom; Oskar Karlsson Lindsjo; Petra Edquist; Reza Advani; Sandra Broddesson; The Public Health Agency of Sweden |
| EPI_ISL_864547 to 864562 | Knappschaftskrankenhaus Bochum | Bundeswehr Institute of Microbiology | Alexandra Rehn; Antonios Katsounas; Enrico Georgi; Malena Bestehorn-Willmann; Markus Antwerpen; Mathias Walter; Mustafa Özçürümez; Roman Wölfel; Sabine Zange |
| EPI_ISL_434659, EPI_ISL_450811 | Knivsta VC | The Public Health Agency of Sweden | Anna Risberg; Anna-Malin Linde; Johanna Carlson; Karin Tegmark-Wisell; Maria Lind Karlberg; Mia Brytting; Olov Svartstrom; Oskar Karlsson Lindsjo; Theresa Enkirch |
| EPI_ISL_754847 to 754848 | Korian Villa d'Azon | National Reference Center for Viruses of Respiratory Infections, Institut Pasteur, Paris | Angela Brisebarre; Camille Capel; Etienne Simon-Lorière; Marion Barbet; Maud Vanpeene; Méline Bizard; Sylvie Behillil; Sylvie van der Werf; Vincent Enouf |
| EPI_ISL_869247, EPI_ISL_869250, EPI_ISL_869262 to 869264, EPI_ISL_872570 | Krasnoyarsk Regional Clinical Hospital | WHO National Influenza Centre Russian Federation | Andrey Komissarov; Anna Ivanova; Artem Fadeev; Daria Danilenko; Dmitry Bazhenov; Dmitry Lioznov; Elena Nabieva; Georgii Bazykin; Ksenia Safina; Kseniya Komissarova; Mikhail Bakaev |
| EPI_ISL_434657 | Kristianstadkliniken | The Public Health Agency of Sweden | Anna Risberg; Anna-Malin Linde; Karin Tegmark-Wisell; Maria Lind Karlberg; Mia Brytting; Mia Settergren Hammer; Olov Svartstrom; Oskar Karlsson Lindsjo; Theresa Enkirch |
| EPI_ISL_434651 | Krokoms Halsocentral | The Public Health Agency of Sweden | Anna Risberg; Anna-Malin Linde; Karin Tegmark-Wisell; Maria Lind Karlberg; Martin Ersson; Mia Brytting; Olov Svartstrom; Oskar Karlsson Lindsjo; Theresa Enkirch |
| EPI_ISL_539881 | Kungsbacka Narakut | The Public Health Agency of Sweden | Anna Risberg; Anna-Malin Linde; Karin Tegmark-Wisell; Maria Lind Karlberg; Mattias Haukland; Mia Brytting; Olov Svartstrom; Oskar Karlsson Lindsjo; Reza Advani; Sandra Broddesson; Theresa Enkirch |
| EPI_ISL_434644, EPI_ISL_450809, EPI_ISL_469067, EPI_ISL_475542, EPI_ISL_475569 | Kungsholmsdoktorn | The Public Health Agency of Sweden | Anna Risberg; Anna-Malin Linde; Karin Tegmark-Wisell; Linus Hammar; Maria Lind Karlberg; Mattias Haukland; Mia Brytting; Olov Svartstrom; Oskar Karlsson Lindsjo; Petra Edquist; Reza Advani; Sandra Broddesson; Shamam Muradrasoli; Theresa Enkirch |
| EPI_ISL_434668, EPI_ISL_445232, EPI_ISL_455102, EPI_ISL_475529 to 475532, EPI_ISL_475568 | Kungsors VC | The Public Health Agency of Sweden | Anna Risberg; Anna-Malin Linde; Jessica Karlsson; Karin Tegmark-Wisell; Maria Lind Karlberg; Mattias Haukland; Mia Brytting; Olov Svartstrom; Oskar Karlsson Lindsjo; Reza Advani; Sandra Broddesson; Theresa Enkirch |
| EPI_ISL_420039 to 420040 | L'Air du Temps | National Reference Center for Viruses of Respiratory Infections, Institut Pasteur, Paris | Angela Brisebarre; Etienne Simon-Lorière; Flora Donati; Marion Barbet; Maud Vanpeene; Mélanie Albert; Méline Bizard; Sylvie Behillil; Sylvie van der Werf; Vincent Enouf |
| EPI_ISL_508931 | L'Hôpital Nord-Ouest Tarare-Grandris | CNR Virus des Infections Respiratoires - France SUD | Alexandre Gaymard; Antonin Bal; Bruno Lina; Carine Moustaud; Florence Morfin-Sherpa; Gregory Destras; Gwendolyne Burfin; Laurence Josset; Martine Valette; Maude Bouscambert-Duchamp; Raphaelle Lamy; Solenne Brun |
| EPI_ISL_416499 to 416500, EPI_ISL_418234, EPI_ISL_418240 | LABM GH nord Essonne | National Reference Center for Viruses of Respiratory Infections, Institut Pasteur, Paris | Angela Brisebarre; Christine Lambert; Etienne Simon-Lorière; Flora Donati; Marion Barbet; Maud Vanpeene; Mélanie Albert; Méline Bizard; Mélnie Albert; Sylvie Behillil; Sylvie van der Werf; Vincent Enouf |
| EPI_ISL_428354, EPI_ISL_428361 to 428362, EPI_ISL_428365, EPI_ISL_443260, EPI_ISL_443305, EPI_ISL_443314 | LABM GH nord Essonne de Longjumeau - BP 125 | National Reference Center for Viruses of Respiratory Infections, Institut Pasteur, Paris | Angela Brisebarre; Etienne Simon-Lorière; Flora Donati; Marion Barbet; Maud Vanpeene; Mélanie Albert; Méline Bizard; Sylvie Behillil; Sylvie van der Werf; Vincent Enouf |
| EPI_ISL_498226 | LIC | LIC | LIC |
| EPI_ISL_443307 | La Villa Papyri | National Reference Center for Viruses of Respiratory Infections, Institut Pasteur, Paris | Angela Brisebarre; Etienne Simon-Lorière; Flora Donati; Marion Barbet; Maud Vanpeene; Mélanie Albert; Méline Bizard; Sylvie Behillil; Sylvie van der Werf; Vincent Enouf |
| EPI_ISL_501614, EPI_ISL_501616 to 501631 | Lab Microbiology, Pathology Department, William Harvey Hospital | Wellcome Sanger Institute for the COVID-19 Genomics UK (COG-UK) consortium | Cordelia Langford; David K. Jackson; Dominic Kwiatkowski; Ewan Harrison; Felicity Ryan and Alex Alderton; Hannah Lowe; Ian Johnston; John Sillitoe on behalf of the Wellcome Sanger Institute COVID-19 Surveillance Team (http://www.sanger.ac.uk/covid-team); Roberto Amato; Samuel Moses; Sonia Goncalves |
| EPI_ISL_419259, EPI_ISL_419264 to 419266, EPI_ISL_425048, EPI_ISL_425050 to 425064, EPI_ISL_434347 to 434365, EPI_ISL_451164 to 451182, EPI_ISL_468727 to 468746, EPI_ISL_475060 to 475082, EPI_ISL_481221 to 481233, EPI_ISL_634876 to 634895, EPI_ISL_717573 to 717575, EPI_ISL_717577 to 717593, EPI_ISL_717596 to 717633, EPI_ISL_717980 to 717983, EPI_ISL_717985, EPI_ISL_717987, EPI_ISL_717990 to 718026, EPI_ISL_745202 to 745212, EPI_ISL_745214 to 745221, EPI_ISL_794825, EPI_ISL_855902, EPI_ISL_855906, EPI_ISL_855908, EPI_ISL_855910, EPI_ISL_855912, EPI_ISL_855914, EPI_ISL_855916 to 855917, EPI_ISL_855919, EPI_ISL_855921, EPI_ISL_855923, EPI_ISL_855925, EPI_ISL_855927 to 855928, EPI_ISL_855932, EPI_ISL_855934, EPI_ISL_855942 | | | |
| see above | Lab voor klinische biologie | Onderzoeksgroep Virologie | Basiel Cole; Bruno Verhasselt; Hans Nauwynck; Jozefien De Clercq; Laurens Lambrechts; Linos Vandekerckhove; Marthe Pauwels; Nick Vereecke; Sebastiaan Theuns |
| EPI_ISL_852619, EPI_ISL_852622, EPI_ISL_852628, EPI_ISL_855556, EPI_ISL_855573, EPI_ISL_855610 to 855614, EPI_ISL_856679, EPI_ISL_856682, EPI_ISL_856694, EPI_ISL_856698, EPI_ISL_856700, EPI_ISL_860095, EPI_ISL_860203 to 860205, EPI_ISL_860257, EPI_ISL_860290, EPI_ISL_860297, EPI_ISL_860794 | | | |
| see above | Lab. Microbiologia e Virologia, Cotugno, A.O. dei Colli | Lab. Microbiologia e Virologia, Cotugno, A.O. dei Colli | Anna Perfetti; Claudia Tiberio; Luigi Atripaldi |
| EPI_ISL_560637 to 560642, EPI_ISL_860864, EPI_ISL_860869 to 860871, EPI_ISL_860882 to 860884, EPI_ISL_872203, EPI_ISL_872209, EPI_ISL_872258 to 872259, EPI_ISL_872290, EPI_ISL_872323 | | | |
| see above | Labo Analyses Med | National Reference Center for Viruses of Respiratory Infections, Institut Pasteur, Paris | Angela Brisebarre; Camille Capel; Etienne Simon-Lorière; Fabiana Gambaro; Girard Sophie; Lefaure Brieuc; Marion Barbet; Maud Vanpeene; Méline Bizard; Rouah Raquel; Sylvie Behillil; Sylvie van der Werf; Vincent Enouf |
| EPI_ISL_593924 to 593933 | Labo Analyses Med, Puteaux | National Reference Center for Viruses of Respiratory Infections, Institut Pasteur, Paris | Etienne Simon-Lorière; Fabiana Gambaro; Maud Vanpeene; Sylvie Behillil; Sylvie van der Werf; Vincent Enouf |
| EPI_ISL_593904 to 593909 | Labo Analyses Med, Sarcelles | National Reference Center for Viruses of Respiratory Infections, Institut Pasteur, Paris | Etienne Simon-Lorière; Fabiana Gambaro; Maud Vanpeene; Sylvie Behillil; Sylvie van der Werf; Vincent Enouf |
| EPI_ISL_420063 | Labo BM - Site de Juvisy - Hopital Général | National Reference Center for Viruses of Respiratory Infections, Institut Pasteur, Paris | Angela Brisebarre; Etienne Simon-Lorière; Flora Donati; Marion Barbet; Maud Vanpeene; Mélanie Albert; Méline Bizard; Sylvie Behillil; Sylvie van der Werf; Vincent Enouf |
| EPI_ISL_872313 | Labo analyses med | National Reference Center for Viruses of Respiratory Infections, Institut Pasteur, Paris | Amzalag Jonas; Angela Brisebarre; Camille Capel; Etienne Simon-Lorière; Marion Barbet; Maud Vanpeene; Méline Bizard; Sylvie Behillil; Sylvie van der Werf; Vincent Enouf |
| EPI_ISL_831404, EPI_ISL_831412, EPI_ISL_831417 to 831419, EPI_ISL_831423, EPI_ISL_831426 | Labor Dr. Krause & Kollegen MVZ GmbH | Robert Koch Institute, Influenza and respiratory viruses FG17 & Bioinformatics MF1, Berlin, Germany | Aleksandar Radonic; Dr. Lorentz; Marianne Wedde; Oliver Drechsel; Ralf Duerrwald; Rene Kmiecinski; Stefan Kroeger; Stephan Fuchs; Thorsten Wolff |
| EPI_ISL_803955 to 803957, EPI_ISL_831409, EPI_ISL_831411, EPI_ISL_831420 to 831421, EPI_ISL_831424 to 831425 | Labor Dr. Wisplinghoff - Köln | Robert Koch Institute, Influenza and respiratory viruses FG17 & Bioinformatics MF1, Berlin, Germany | Aleksandar Radonic; Dr. R. Grosser; Marianne Wedde; Oliver Drechsel; Ralf Duerrwald; Rene Kmiecinski; Stefan Kroeger; Stephan Fuchs; Thorsten Wolff |
| EPI_ISL_476705, EPI_ISL_487398 to 487431 | Labor Kneißler GmbH & Co. KG | Heinrich Pette Institute, Leibniz Institute for Experimental Virology | Adam Grundhoff; Manja Czech-Sioli; Matthias Ottinger; Melanie M. Brinkmann; Nicole Fischer; Thomas Günther |
| EPI_ISL_831370 to 831372, EPI_ISL_831374, EPI_ISL_831379, EPI_ISL_831382 to 831385, EPI_ISL_831392, EPI_ISL_831394 to 831395, EPI_ISL_831398 to 831399, EPI_ISL_831401 | | | |

| | | | |
|---|---|---|---|
| see above | Labor Krone | Robert Koch Institute, Influenza and respiratory viruses FG17 & Bioinformatics MF1, Berlin, Germany | Aleksandar Radonic; Dr. Münstermann. Prof. Tiemann; Marianne Wedde; Oliver Drechsel; Ralf Duerrwald; Rene Kmiecinski; Stefan Kroeger; Stephan Fuchs; Thorsten Wolff |
| EPI_ISL_831414 to 831416 | Labor Prof. Dr. G. Enders MVZ GbR | Robert Koch Institute, Influenza and respiratory viruses FG17 & Bioinformatics MF1, Berlin, Germany | Aleksandar Radonic; Dr. Friedemann Tewald; Marianne Wedde; Oliver Drechsel; Ralf Duerrwald; Rene Kmiecinski; Stefan Kroeger; Stephan Fuchs; Thorsten Wolff |
| EPI_ISL_445220 | Laborarory for Respiratory Viruses, "Cantacuzino" National Military-Medical Institute for Resararch and Development | Cantacuzino Institute | A.Cretu; L.Ustea; M.Lazar |
| EPI_ISL_768818 | Laboratoire 2A2B | CNR Virus des Infections Respiratoires - France SUD | Antonin Bal; Bruno Lina; Gregory Destras; Gwendolyne Burfin; Hadrien Règue; Laurence Josset; Martine Valette; Quentin Semanas |
| EPI_ISL_768825 to 768826 | Laboratoire AX Bio Ocean | CNR Virus des Infections Respiratoires - France SUD | Antonin Bal; Bruno Lina; Gregory Destras; Gwendolyne Burfin; Hadrien Règue; Laurence Josset; Martine Valette; Quentin Semanas |
| EPI_ISL_768824 | Laboratoire BIOLAB33 | CNR Virus des Infections Respiratoires - France SUD | Antonin Bal; Bruno Lina; Gregory Destras; Gwendolyne Burfin; Hadrien Règue; Laurence Josset; Martine Valette; Quentin Semanas |
| EPI_ISL_768830, EPI_ISL_779848 to 779849 | Laboratoire BIOMED 05 | CNR Virus des Infections Respiratoires - France SUD | Antonin Bal; Bruno Lina; Gregory Destras; Gwendolyne Burfin; Hadrien Règue; Laurence Josset; Martine Valette; Quentin Semanas |
| EPI_ISL_768819 to 768820 | Laboratoire BPO-BIOEPINE | CNR Virus des Infections Respiratoires - France SUD | Antonin Bal; Bruno Lina; Gregory Destras; Gwendolyne Burfin; Hadrien Règue; Laurence Josset; Martine Valette; Quentin Semanas |
| EPI_ISL_768817, EPI_ISL_779836 | Laboratoire Mirialis | CNR Virus des Infections Respiratoires - France SUD | Antonin Bal; Bruno Lina; Gregory Destras; Gwendolyne Burfin; Hadrien Règue; Laurence Josset; Martine Valette; Quentin Semanas |
| EPI_ISL_421734 to 421763, EPI_ISL_428939 to 428962, EPI_ISL_429706 to 429750, EPI_ISL_429752 to 429791, EPI_ISL_429793 to 429800, EPI_ISL_434487 to 434515, EPI_ISL_445053 to 445076, EPI_ISL_459893 to 459906 | | | |
| see above | Laboratoire National de Sante, Microbiology, Virology | Laboratoire National de Sante, Microbiology, Epidemiology and Microbial Genomics | Anke Wienecke-Baldacchino; Ardashel Latsuzbaia; Catherine Ragimbeau; Guillaume Fournier; Jessica Tapp; Joel Mossong; Tamir Abdelrahman; Trung Nguyen Nguyen |
| EPI_ISL_413593 | Laboratoire National de Santé | Erasmus Medical Center | Anne van der Linden; Annemiek van der Eijk; Bas Oude Munnink; Claudia Schapendonk; Corine GeurtsvanKessel; David Nieuwenhuijse; G. Fournier; Irina Chestakova; J. Mossong; Jeroen van Kampen; Jolanda Voermans; Marion Koopmans; Mark Pronk; Pascal Lexmond; Reina Sikkema; Richard Molenkamp; T. Abdelrahman; T. Nguyen; on behalf of the Dutch national COVID-19 response team. |
| EPI_ISL_419562 to 419607 | Laboratoire National de Santé, Microbiology, Virology | Laboratoire National de Santé, Microbiology, Epidemiology and Microbial Genomics | Anke Wienecke-Baldacchino; Ardashel Latsuzbaia; Catherine Ragimbeau; Guillaume Fournier; Jessica Tapp; Joel Mossong; Tamir Abdelrahman; Trung Nguyen Nguyen |
| EPI_ISL_417526, EPI_ISL_417528 to 417534 | Laboratoire Nationale de Santé, Microbiology, Virology | Laboratoire Nationale de Santé, Microbiology, Epidemiology and Microbial Genomics | Anke Wienecke-Baldacchino; Ardashel Latsuzbaia; Catherine Ragimbeau; Guillaume Fournier; Jessica Tapp; Joel Mossong; Tamir Abdelrahman; Trung Nguyen Nguyen |
| EPI_ISL_768821 | Laboratoire Novabio | CNR Virus des Infections Respiratoires - France SUD | Antonin Bal; Bruno Lina; Gregory Destras; Gwendolyne Burfin; Hadrien Règue; Laurence Josset; Martine Valette; Quentin Semanas |
| EPI_ISL_754141 to 754172 | Laboratoire de Microbiologie | National Reference Center for Viruses of Respiratory Infections, Institut Pasteur, Paris | Angela Brisebarre; Camille Capel; Etienne Simon-Lorière; Marie-Sarah Fangous; Marion Barbet; Maud Vanpeene; Méline Bizard; Sylvie Behillil; Sylvie van der Werf; Vincent Enouf |
| EPI_ISL_443284 to 443288 | Laboratoire de Microbiologie - Bât A - CH René Dubois | National Reference Center for Viruses of Respiratory Infections, Institut Pasteur, Paris | Angela Brisebarre; Etienne Simon-Lorière; Flora Donati; Marion Barbet; Maud Vanpeene; Mélanie Albert; Méline Bizard; Pascale Martres; Sylvie Behillil; Sylvie van der Werf; Vincent Enouf |
| EPI_ISL_754843 to 754846, EPI_ISL_754849 to 754852 | Laboratoire de Virologie Hôpital Robert Debré | National Reference Center for Viruses of Respiratory Infections, Institut Pasteur, Paris | Andreoletti Laurent; Angela Brisebarre; Camille Capel; Etienne Simon-Lorière; Marion Barbet; Maud Vanpeene; Méline Bizard; Sylvie Behillil; Sylvie van der Werf; Vincent Enouf |
| EPI_ISL_792000 to 792002, EPI_ISL_792004 to 792007 | Laboratoire de Virologie Hôpital Robert Debré | National Reference Center for Viruses of Respiratory Infections, Institut Pasteur, Paris | Andreoletti Laurent; Angela Brisebarre; Camille Capel; Etienne Simon-Lorière; Marion Barbet; Maud Vanpeene; Méline Bizard; Sylvie Behillil; Sylvie van der Werf; Vincent Enouf |
| EPI_ISL_414600, EPI_ISL_414623 | Laboratoire de Virologie Institut de Virologie - INSERM U 1109 Hôpitaux Universitaires de Strasbourg | National Reference Center for Viruses of Respiratory Infections, Institut Pasteur, Paris | Angela Brisebarre; Flora Donati Vincent Enouf; Marion Barbet; Maud Vanpeene; Méline Bizard; Mélnie Albert; Samira Fafi-Kremer; Sylvie Behillil; Sylvie van der Werf |
| EPI_ISL_666870 | Laboratoire de Virologie, CHU de Caen, Normandie, France. | GRAM2.0, Université de Caen Normandie | Astrid VABRET; Estelle LEPERCHOIS; Januel VOUTEAU; Juliette ECHE; Meriadeg LE GOUIL |
| EPI_ISL_413996 to 413997, EPI_ISL_413999, EPI_ISL_414019 to 414023, EPI_ISL_548147 to 548175, EPI_ISL_548177 to 548199, EPI_ISL_548201 to 548202, EPI_ISL_548204 to 548235, EPI_ISL_548237 to 548242 | | | |
| see above | Laboratoire de Virologie, HUG | Swiss National Reference Centre for Influenza | LAUBSCHER F.; LAUBSCHER Florian et al. |
| EPI_ISL_765479 | Laboratoire de Virologie, Hôpital Robert Debré | National Reference Center for Viruses of Respiratory Infections, Institut Pasteur, Paris | Andreoletti Laurent; Angela Brisebarre; Camille Capel; Etienne Simon-Lorière; Marion Barbet; Maud Vanpeene; Méline Bizard; Sylvie Behillil; Sylvie van der Werf; Vincent Enouf |
| EPI_ISL_779781 to 779782, EPI_ISL_779814 to 779835 | Laboratoire de virologie, CHU de Grenoble | CNR Virus des Infections Respiratoires - France SUD | Antonin Bal; Bruno Lina; Gregory Destras; Gwendolyne Burfin; Hadrien Règue; Laurence Josset; Martine Valette; Quentin Semanas; Sylvie Larrat |
| EPI_ISL_634818, EPI_ISL_700349 | Laboratoire de virologie, CHU de Grenoble - CS 10217 - 38043 Grenoble cedex 10 | CNR Virus des Infections Respiratoires - France SUD | Antonin Bal; Bruno Lina; Gregory Destras; Gwendolyne Burfin; Hadrien Règue; Laurence Josset; Martine Valette; Quentin Semanas; Sylvie Larrat |
| EPI_ISL_634819, EPI_ISL_700350 | Laboratoire de virologie, CHU de Grenoble - CS 10217 - 38043 Grenoble cedex 11 | CNR Virus des Infections Respiratoires - France SUD | Antonin Bal; Bruno Lina; Gregory Destras; Gwendolyne Burfin; Hadrien Règue; Laurence Josset; Martine Valette; Quentin Semanas; Sylvie Larrat |
| EPI_ISL_634820, EPI_ISL_700351 | Laboratoire de virologie, CHU de Grenoble - CS 10217 - 38043 Grenoble cedex 12 | CNR Virus des Infections Respiratoires - France SUD | Antonin Bal; Bruno Lina; Gregory Destras; Gwendolyne Burfin; Hadrien Règue; Laurence Josset; Martine Valette; Quentin Semanas; Sylvie Larrat |
| EPI_ISL_634821, EPI_ISL_700352 | Laboratoire de virologie, CHU de Grenoble - CS 10217 - 38043 Grenoble cedex 13 | CNR Virus des Infections Respiratoires - France SUD | Antonin Bal; Bruno Lina; Gregory Destras; Gwendolyne Burfin; Hadrien Règue; Laurence Josset; Martine Valette; Quentin Semanas; Sylvie Larrat |
| EPI_ISL_634822, EPI_ISL_700353 | Laboratoire de virologie, CHU de Grenoble - CS 10217 - 38043 Grenoble cedex 14 | CNR Virus des Infections Respiratoires - France SUD | Antonin Bal; Bruno Lina; Gregory Destras; Gwendolyne Burfin; Hadrien Règue; Laurence Josset; Martine Valette; Quentin Semanas; Sylvie Larrat |
| EPI_ISL_634823, EPI_ISL_700354 | Laboratoire de virologie, CHU de Grenoble - CS 10217 - 38043 Grenoble cedex 15 | CNR Virus des Infections Respiratoires - France SUD | Antonin Bal; Bruno Lina; Gregory Destras; Gwendolyne Burfin; Hadrien Règue; Laurence Josset; Martine Valette; Quentin Semanas; Sylvie Larrat |
| EPI_ISL_634824, EPI_ISL_700355 | Laboratoire de virologie, CHU de Grenoble - CS 10217 - 38043 Grenoble cedex 16 | CNR Virus des Infections Respiratoires - France SUD | Antonin Bal; Bruno Lina; Gregory Destras; Gwendolyne Burfin; Hadrien Règue; Laurence Josset; Martine Valette; Quentin Semanas; Sylvie Larrat |
| EPI_ISL_634825, EPI_ISL_700356 | Laboratoire de virologie, CHU de Grenoble - CS 10217 - 38043 Grenoble cedex 17 | CNR Virus des Infections Respiratoires - France SUD | Antonin Bal; Bruno Lina; Gregory Destras; Gwendolyne Burfin; Hadrien Règue; Laurence Josset; Martine Valette; Quentin Semanas; Sylvie Larrat |
| EPI_ISL_634826, EPI_ISL_700357 | Laboratoire de virologie, CHU de Grenoble - CS 10217 - 38043 Grenoble cedex 18 | CNR Virus des Infections Respiratoires - France SUD | Antonin Bal; Bruno Lina; Gregory Destras; Gwendolyne Burfin; Hadrien Règue; Laurence Josset; Martine Valette; Quentin Semanas; Sylvie Larrat |
| EPI_ISL_634827, EPI_ISL_700358 | Laboratoire de virologie, CHU de Grenoble - CS 10217 - 38043 Grenoble cedex 19 | CNR Virus des Infections Respiratoires - France SUD | Antonin Bal; Bruno Lina; Gregory Destras; Gwendolyne Burfin; Hadrien Règue; Laurence Josset; Martine Valette; Quentin Semanas; Sylvie Larrat |
| EPI_ISL_634828, EPI_ISL_700359 | Laboratoire de virologie, CHU de Grenoble - CS 10217 - 38043 Grenoble cedex 20 | CNR Virus des Infections Respiratoires - France SUD | Antonin Bal; Bruno Lina; Gregory Destras; Gwendolyne Burfin; Hadrien Règue; Laurence Josset; Martine Valette; Quentin Semanas; Sylvie Larrat |
| EPI_ISL_634829, EPI_ISL_700360 | Laboratoire de virologie, CHU de Grenoble - CS 10217 - 38043 Grenoble cedex 21 | CNR Virus des Infections Respiratoires - France SUD | Antonin Bal; Bruno Lina; Gregory Destras; Gwendolyne Burfin; Hadrien Règue; Laurence Josset; Martine Valette; Quentin Semanas; Sylvie Larrat |
| EPI_ISL_634830, EPI_ISL_700361 | Laboratoire de virologie, CHU de Grenoble - CS 10217 - 38043 Grenoble cedex 22 | CNR Virus des Infections Respiratoires - France SUD | Antonin Bal; Bruno Lina; Gregory Destras; Gwendolyne Burfin; Hadrien Règue; Laurence Josset; Martine Valette; Quentin Semanas; Sylvie Larrat |
| EPI_ISL_634831, EPI_ISL_700362 | Laboratoire de virologie, CHU de Grenoble - CS 10217 - 38043 Grenoble cedex 23 | CNR Virus des Infections Respiratoires - France SUD | Antonin Bal; Bruno Lina; Gregory Destras; Gwendolyne Burfin; Hadrien Règue; Laurence Josset; Martine Valette; Quentin Semanas; Sylvie Larrat |

| | | | |
|---|---|---|---|
| EPI_ISL_634832, EPI_ISL_700363 | Laboratoire de virologie, CHU de Grenoble - CS 10217 - 38043 Grenoble cedex 24 | CNR Virus des Infections Respiratoires - France SUD | Antonin Bal; Bruno Lina; Gregory Destras; Gwendolyne Burfin; Hadrien Règue; Laurence Josset; Martine Valette; Quentin Semanas; Sylvie Larrat |
| EPI_ISL_634833, EPI_ISL_700364 | Laboratoire de virologie, CHU de Grenoble - CS 10217 - 38043 Grenoble cedex 25 | CNR Virus des Infections Respiratoires - France SUD | Antonin Bal; Bruno Lina; Gregory Destras; Gwendolyne Burfin; Hadrien Règue; Laurence Josset; Martine Valette; Quentin Semanas; Sylvie Larrat |
| EPI_ISL_634834, EPI_ISL_700365 | Laboratoire de virologie, CHU de Grenoble - CS 10217 - 38043 Grenoble cedex 26 | CNR Virus des Infections Respiratoires - France SUD | Antonin Bal; Bruno Lina; Gregory Destras; Gwendolyne Burfin; Hadrien Règue; Laurence Josset; Martine Valette; Quentin Semanas; Sylvie Larrat |
| EPI_ISL_634835, EPI_ISL_700366 | Laboratoire de virologie, CHU de Grenoble - CS 10217 - 38043 Grenoble cedex 27 | CNR Virus des Infections Respiratoires - France SUD | Antonin Bal; Bruno Lina; Gregory Destras; Gwendolyne Burfin; Hadrien Règue; Laurence Josset; Martine Valette; Quentin Semanas; Sylvie Larrat |
| EPI_ISL_634836, EPI_ISL_700367 | Laboratoire de virologie, CHU de Grenoble - CS 10217 - 38043 Grenoble cedex 28 | CNR Virus des Infections Respiratoires - France SUD | Antonin Bal; Bruno Lina; Gregory Destras; Gwendolyne Burfin; Hadrien Règue; Laurence Josset; Martine Valette; Quentin Semanas; Sylvie Larrat |
| EPI_ISL_634837, EPI_ISL_700368 | Laboratoire de virologie, CHU de Grenoble - CS 10217 - 38043 Grenoble cedex 29 | CNR Virus des Infections Respiratoires - France SUD | Antonin Bal; Bruno Lina; Gregory Destras; Gwendolyne Burfin; Hadrien Règue; Laurence Josset; Martine Valette; Quentin Semanas; Sylvie Larrat |
| EPI_ISL_700369 | Laboratoire de virologie, CHU de Grenoble - CS 10217 - 38043 Grenoble cedex 30 | CNR Virus des Infections Respiratoires - France SUD | Antonin Bal; Bruno Lina; Gregory Destras; Gwendolyne Burfin; Hadrien Règue; Laurence Josset; Martine Valette; Quentin Semanas; Sylvie Larrat |
| EPI_ISL_700370 | Laboratoire de virologie, CHU de Grenoble - CS 10217 - 38043 Grenoble cedex 31 | CNR Virus des Infections Respiratoires - France SUD | Antonin Bal; Bruno Lina; Gregory Destras; Gwendolyne Burfin; Hadrien Règue; Laurence Josset; Martine Valette; Quentin Semanas; Sylvie Larrat |
| EPI_ISL_634817, EPI_ISL_700348 | Laboratoire de virologie, CHU de Grenoble - CS 10217 - 38043 Grenoble cedex 9 | CNR Virus des Infections Respiratoires - France SUD | Antonin Bal; Bruno Lina; Gregory Destras; Gwendolyne Burfin; Hadrien Règue; Laurence Josset; Martine Valette; Quentin Semanas; Sylvie Larrat |
| EPI_ISL_476823, EPI_ISL_476830 to 476831, EPI_ISL_476833 to 476834 | Laboratoire des Fièvres Hémorragiques Virales du Benin | Charité-Universitätsmedizin Berlin | Anges; Drexler; Jan Felix; Moreira-Soto Andres; Sander Anna-Lena; Yadouleton |
| EPI_ISL_629081 to 629108, EPI_ISL_666653 to 666665, EPI_ISL_666668 to 666681, EPI_ISL_683331 to 683333 | Laboratoire du Centre Hospitalier Annecy Genevois | CNR Virus des Infections Respiratoires - France SUD | Antonin Bal; Bruno Chanzy; Bruno Lina; Gregory Destras; Gwendolyne Burfin; Hadrien Règue; Hélène Petitprez; Laurence Josset; Martine Valette; Quentin Semanas |
| EPI_ISL_739700, EPI_ISL_739880, EPI_ISL_740129, EPI_ISL_744295, EPI_ISL_744819, EPI_ISL_744830 | Laboratoire national de santé, Microbiology, Virology | Laboratoire national de santé, Microbiology, Epidemiology and Microbial Genomics | Anke Wienecke-Baldacchino; Catherine Ragimbeau; Fatu Djabi; Jessica Tapp; Tamir Abdelrahman; Trung Nguyen Nguyen |
| EPI_ISL_739693 to 739699, EPI_ISL_739701 to 739713, EPI_ISL_739715 to 739746, EPI_ISL_739748 to 739757, EPI_ISL_739759 to 739788, EPI_ISL_739790 to 739807, EPI_ISL_739809 to 739855, EPI_ISL_739857 to 739876, EPI_ISL_739878 to 739879, EPI_ISL_739881 to 739894, EPI_ISL_739896 to 739943, EPI_ISL_739945 to 739954, EPI_ISL_739956 to 739961, EPI_ISL_739963 to 739967, EPI_ISL_739969 to 739975, EPI_ISL_739977 to 739987, EPI_ISL_739989 to 739993, EPI_ISL_739995 to 740008, EPI_ISL_740010 to 740015, EPI_ISL_740017 to 740024, EPI_ISL_740026 to 740032, EPI_ISL_740034 to 740037, EPI_ISL_740039, EPI_ISL_740041 to 740042, EPI_ISL_740044 to 740049, EPI_ISL_740051 to 740095, EPI_ISL_740097 to 740104, EPI_ISL_740106 to 740124, EPI_ISL_740126 to 740128, EPI_ISL_740130 to 740156, EPI_ISL_740158 to 740167, EPI_ISL_740169 to 740181, EPI_ISL_740183, EPI_ISL_740185 to 740200, EPI_ISL_740202, EPI_ISL_740204, EPI_ISL_740206 to 740245, EPI_ISL_740247 to 740251, EPI_ISL_740253 to 740257, EPI_ISL_740259 to 740293, EPI_ISL_740295 to 740306, EPI_ISL_740309 to 740318, EPI_ISL_740320, EPI_ISL_740322 to 740336, EPI_ISL_740338 to 740347, EPI_ISL_740349 to 740352, EPI_ISL_740354 to 740361, EPI_ISL_740363 to 740368, EPI_ISL_740370 to 740376, EPI_ISL_740378 to 740420, EPI_ISL_740422 to 740428, EPI_ISL_740430, EPI_ISL_740432 to 740464, EPI_ISL_740466 to 740467, EPI_ISL_740470 to 740472, EPI_ISL_740474 to 740546, EPI_ISL_744135 to 744150, EPI_ISL_744152 to 744188, EPI_ISL_744190 to 744196, EPI_ISL_744198 to 744210, EPI_ISL_744212 to 744286, EPI_ISL_744288 to 744290, EPI_ISL_744292 to 744294, EPI_ISL_744296 to 744314, EPI_ISL_744316 to 744328, EPI_ISL_744331 to 744339, EPI_ISL_744341 to 744364, EPI_ISL_744367 to 744376, EPI_ISL_744378 to 744383, EPI_ISL_744385 to 744412, EPI_ISL_744414 to 744420, EPI_ISL_744423 to 744427, EPI_ISL_744429 to 744444, EPI_ISL_744446 to 744464, EPI_ISL_744466 to 744476, EPI_ISL_744478 to 744483, EPI_ISL_744486 to 744487, EPI_ISL_744489 to 744530, EPI_ISL_744533 to 744550, EPI_ISL_744552 to 744560, EPI_ISL_744562 to 744596, EPI_ISL_744598 to 744609, EPI_ISL_744611 to 744637, EPI_ISL_744639 to 744643, EPI_ISL_744645 to 744650, EPI_ISL_744653 to 744655, EPI_ISL_744657 to 744709, EPI_ISL_744711 to 744730, EPI_ISL_744732 to 744737, EPI_ISL_744739 to 744743, EPI_ISL_744745 to 744746, EPI_ISL_744748, EPI_ISL_744750 to 744780, EPI_ISL_744782 to 744789, EPI_ISL_744792 to 744818, EPI_ISL_744820 to 744829, EPI_ISL_744831 to 744866, EPI_ISL_744868 to 744869, EPI_ISL_744871 to 744881, EPI_ISL_744883 to 744893, EPI_ISL_744895 to 744902, EPI_ISL_744904 to 744909, EPI_ISL_744911 to 744948, EPI_ISL_744950 to 745014, EPI_ISL_745016 to 745023, EPI_ISL_745025 to 745027, EPI_ISL_755589, EPI_ISL_770845, EPI_ISL_770851, EPI_ISL_770853, EPI_ISL_770855, EPI_ISL_770857 to 770858, EPI_ISL_770860, EPI_ISL_770862, EPI_ISL_770867 to 770870, EPI_ISL_770872 to 770874, EPI_ISL_770877 to 770882, EPI_ISL_770888 to 770946, EPI_ISL_770948 to 770953, EPI_ISL_770955 to 770961, EPI_ISL_770963 to 770970, EPI_ISL_770972 to 770973, EPI_ISL_770976 to 770984, EPI_ISL_770986 to 771053, EPI_ISL_771055 to 771078, EPI_ISL_771080 to 771100, EPI_ISL_771102 to 771150 | | | |
| see above | Laboratoire national de santé, Microbiology, Virology | Laboratoire national de santé, Microbiology, Microbial Genomics Platform | Anke Wienecke-Baldacchino; Catherine Ragimbeau; Fatu Djabi; Jessica Tapp; Lise Pignon; Raoul Salmon; Tamir Abdelrahman |
| EPI_ISL_429128 to 429135, EPI_ISL_430856 to 430859, EPI_ISL_434641 to 434642, EPI_ISL_450836 to 450837 | Laboratoriemedicin | The Public Health Agency of Sweden | Anna Risberg; Anna-Malin Linde; Karin Tegmark-Wisell; Maria Lind Karlberg; Mia Brytting; Olov Svartstrom; Oskar Karlsson Lindsjo; Shaman Muradrasoli; Theresa Enkirch |
| EPI_ISL_534248, EPI_ISL_534252 to 534255, EPI_ISL_534257 to 534258 | Laboratoriemedicin Vasternorrland | The Public Health Agency of Sweden | Anna Risberg; Anna-Malin Linde; Karin Tegmark-Wisell; Maria Lind Karlberg; Mattias Haukland; Mia Brytting; Olov Svartstrom; Oskar Karlsson Lindsjo; Petra Edquist; Reza Advani; Sandra Broddesson |
| EPI_ISL_661299 to 661300, EPI_ISL_676516 to 676517, EPI_ISL_676535 to 676540, EPI_ISL_766640 to 766641, EPI_ISL_831953 to 831957, EPI_ISL_831998 to 831999 | Laboratoriemedicin, Klinisk mikrobiologi | The Public Health Agency of Sweden | Department of Microbiology; The Public Health Agency of Sweden |
| EPI_ISL_965179 | Laboratorio Biologia Molecolare SARS-COV2- UOC Laboratorio Analisi- Servizio Medicina di Laboratorio, Ospedale San Francesco - ATS-ASSLNUoro | LABORATORIO SPECIALISTICO UOC EMATOLOGIA-Ospedale San Francesco - ATS-ASSLNUoro | ASPRONI ROSANNA; FANCELLO TATIANA; FIAMMA MAURA; FLORE ANNA RITA. PALMAS ANGELO DOMENICO; IANA LO MAGLIO; MAMELI GIUSEPPE; MONNE MARIA ITRIA; PIRAS GIOVANNA; SANNA FILOMENA; SULIS VINCENZO; TOJA ALESSANDRO |
| EPI_ISL_458085 | Laboratorio Biologia Molecolare Sars Cov2 - UOC Laboratorio Analisi - Servizio Medicina di Laboratorio , Ospedale "San Francesco" - ATS- ASSL Nuoro | Laboratorio specialistico UOC Ematologia - Ospedale "San Francesco" - ATS-ASSL Nuoro | Asproni Rosanna; Casu Gavino; Fancello Tatiana; Fiamma Maura; Floris Anna Rita; Lo Maglio Iana; Mameli Giuseppe.; Monne Maria Itria; Palmas Angelo Domenico; Piras Giovanna; Sanna Filomena; Sulis Vincenzo; Toja Alessandro |
| EPI_ISL_458084 | Laboratorio Biologia Molecolare Sars Cov2 - UOC Laboratorio Analisi - Servizio Medicina di Laboratorio, Ospedale "San Francesco" - ATS-ASSL Nuoro | Laboratorio specialistico UOC Ematologia - Ospedale "San Francesco" - ATS-ASSL Nuoro | Asproni Rosanna; Casu Gavino; Fancello Tatiana; Fiamma Maura; Floris Anna Rita; Lo Maglio Iana; Mameli Giuseppe.; Monne Maria Itria; Palmas Angelo Domenico; Piras Giovanna; Sanna Filomena; Sulis Vincenzo; Toja Alessandro |
| EPI_ISL_613706, EPI_ISL_613710, EPI_ISL_613953, EPI_ISL_613955, EPI_ISL_614396 to 614398, EPI_ISL_614889, EPI_ISL_637107 to 637109 | | | |
| see above | Laboratorio Biologia Molecolare Sars Cov2 - UOC Laboratorio Analisi - Servizio Medicina di Laboratorio, Ospedale "San Francesco" - ATS-ASSL Nuoro | Laboratorio specialistico UOC Ematologia - Ospedale "San Francesco" - ATS-ASSL Nuoro | Asproni Rosanna; Casu Gavino; Fancello Tatiana; Fiamma Maura; Floris Anna Rita; Lo Maglio Iana; Mameli Giuseppe; Mameli Giuseppe.; Monne Maria Itria; Palmas Angelo Domenico; Piras Giovanna; Sanna Filomena; Sulis Vincenzo; Toja Alessandro |
| EPI_ISL_613560 | Laboratorio Biologia Molecolare Sars Cov2 - UOC Laboratorio Analisi - Servizio Medicina di Laboratorio, Ospedale "San Francesco" - ATS-ASSL Nuoro Via Mannironi 1, 08100 Nuoro | Laboratorio specialistico UOC Ematologia - Ospedale "San Francesco" - ATS-ASSL Nuoro Nuoro | Asproni Rosanna; Casu Gavino; Fancello Tatiana; Fiamma Maura; Floris Anna Rita; Lo Maglio Iana; Mameli Giuseppe; Monne Maria Itria; Palmas Angelo Domenico; Piras Giovanna; Sanna Filomena; Sulis Vincenzo; Toja Alessandro |
| EPI_ISL_547965 | Laboratorio Biologia Molecolare SarsCov2 UOC LAboratorio Analisi SErvizio Medicina di Laboratorio Ospedale San Francesco ATS-ASSL Nuoro | Laboratorio Specialistico UOC Ematologia Ospedale San Francesco - ATS ASSL NUORO | Asproni Rosanna; Casu Gavino; Fancello Tatiana; Fiamma Maura; Floris Anna Rita; Lo Maglio Iana; Mameli Giuseppe.; Monne Maria Itria; Palmas Angelo Domenico; Piras Giovanna; Sanna Filomena; Sulis Vincenzo; Toja Alessandro |
| EPI_ISL_842652 | Laboratorio de Biología Molecular Hospital Pedro de Elizalde | Grupo de Genómica y Bioinformática del Instituto de Investigación de la Cadena Láctea CONICET-INTA on behalf of 'Proyecto Argentino Interinstitucional de genomica de SARS-CoV-2' (PAIS Consortium) | A; AF; Alegre; Alexay; Amadio; Aulicino; B; Bressan; C; Chamorro; Claps; D; Diaz; E; Eberhardt; F; FJ; G; Gondolessi; Goya; Gómez; Indart; Irazoqui; J; König; L; Lorenzo; Lusso; M; ME; MF; ML; MS; Marchetti; Martín; Montoto Piazza; Morandi; N; Nabaes Jodar; Natale; Osaba; P; Paez; Rocovich; Rosales; S; Sanchez; Sueiro; Torres; Valinotto; Viegas, M.; Wenk; Zamora |
| EPI_ISL_796127 to 796133 | Laboratorio de Microbiología. Hospital General Universitario de Elda, Alicante | SeqCOVID-SPAIN consortium/IBV(CSIC) | Cristina Torregrosa Hetland; Eva Pastor Boix; Mª Isabel Gascón Ros; Paloma Cascales Ramos and SeqCOVID-SPAIN consortium |
| EPI_ISL_430804, EPI_ISL_430807, | Laboratorio de Virología del Hospital de Niños Dr. Ricardo | Área de Secuenciación del Laboratorio de Virología del | AS; E; Goya; Gravis; LE; Lusso; MI; MS; Mistchenko; Nabaes Jodar; Natale; S; Valinotto; Viegas, M. |

| | | | |
|---|---|---|---|
| EPI_ISL_430811 | Gutierrez | Hospital de Niños Dr. Ricardo Gutierrez on behalf of 'Proyecto Argentino Interinstitucional de genomica de SARS-CoV-2' (PAIS Consortium) | |
| EPI_ISL_413489 | Laboratorio di Microbiologia e Virologia, Università Vita-Salute San Raffaele, Milano | Laboratorio di Microbiologia e Virologia, Università Vita-Salute San Raffaele, Milano | C. Di Resta; E. Boeri; E. Criscuolo; G. Lo Raso; I. Negri; M. Castelli; M. Clementi; M. Sampaolo; N. Mancini & N. Clementi; R. Burioni; R. Ferrarese; R.A Diotti; V. Amato; V. Caputo |
| EPI_ISL_582692 to 582694, EPI_ISL_582767 to 582770, EPI_ISL_582802, EPI_ISL_582810 to 582811, EPI_ISL_582833, EPI_ISL_582842, EPI_ISL_582845 to 582850 | | | |
| see above | Laboratorio di Riferimento Regionale della Sicilia Occidentale per l'Emergenza COVID-19 | Istituto Zooprofilattico Sperimentale della Sicilia | Lo Presti Alessandra; Maida Carmelo Massimo; Mazzucco Walter; Pulvirenti Claudio; Purpari Giuseppa; Reale Stefano; Rezza Giovanni; Scibetta Silvia; Stefanelli Paola; Tramuto Fabio; Vitale Fabrizio; Vitale Francesco; Zichichi Salvatore |
| EPI_ISL_710503 | Laboratorio specialistico UOC Ematologia - Ospedale "San Francesco" - ATS-ASSL Nuoro | Laboratorio specialistico UOC Ematologia - Ospedale "San Francesco" - ATS-ASSL Nuoro | Giovanna Piras |
| EPI_ISL_775589, EPI_ISL_831989 | Laboratorium for klinisk mikrobiologi | The Public Health Agency of Sweden | Department of Microbiology; The Public Health Agency of Sweden |
| EPI_ISL_832192 to 832195 | Laboratory Analyses Med | National Reference Center for Viruses of Respiratory Infections, Institut Pasteur, Paris | Angela Brisebarre; Brieuc Gestin; Brieuc Lefaure; Camille Capel; Etienne Simon-Lorière; Marion Barbet; Maud Vanpeene; Méline Bizard; Sylvie Behillil; Sylvie van der Werf; Vincent Enouf |
| EPI_ISL_514425 to 514431, EPI_ISL_541649 to 541655, EPI_ISL_541657 to 541662, EPI_ISL_582530, EPI_ISL_644565 to 644567 | Laboratory Diagnostic, Veterinary Specialized Institute Kraljevo | Laboratory Diagnostic, Veterinary Specialized Institute Kraljevo | Afonso, C.; Banovic Djeri, B.; Jankovic, M.; Jovanovic, T.; Knezevic, A.; Petrovic, T.; Sekler, M.; Tesovic, B.; Vidanovic, D.; Volkening, J. |
| EPI_ISL_445086 to 445087, EPI_ISL_482744, EPI_ISL_541656 | Laboratory Diagnostic, Veterinary Specialized Institute Kraljevo | Laboratory Diagnostic, Veterinary Specialized Institute Kraljevo | Afonso, C.; Banovic Djeri, B.; C.L.; Debeljak, Z.; Dmitric, M.; J. and Alfonso; J. and Alfonso, C.; Jankovic, M.; Jovanovic, T.; Knezevic, A.; Matovic, K.; Petrovic, T.; Sekler, M.; Tesovic, B.; Vaskovic, N.; Vidanovic, D.; Volkening; Volkening, J. |
| EPI_ISL_754902, EPI_ISL_754904 to 754913 | Laboratory Diagnostics and Clinical Immunology of Developmental Age, Medical University of Warsaw | genXone SA, Research & Development Laboratory; The Faculty of Mathematics, Informatics and Mechanics of the University of Warsaw | Anna Gambin; Grzegorz Nowicki; Jakub Grabowski; Maciej Sykulski; Micha Kaszuba; Monika Makowska-Woniak; Natalia Drwska-Matelska; Urszula Demkow; ukasz Krych |
| EPI_ISL_445243, EPI_ISL_455468 to 455477, EPI_ISL_455479 | Laboratory for Respiratory Viruses, Cantacuzino National Military-Medical Institute for Research and Development | Cantacuzino Institute | A.Cretu; L.Ustea; M.Lazar; T.Durfee; Tim Durfee |
| EPI_ISL_754238 | Laboratory for Respiratory Viruses, Cantacuzino National Military-Medical Institute for Research and Development | Cantacuzino Institute Virology | Luiza Ustea; Mihaela Lazar; Mühlemann Barbara |
| EPI_ISL_471416 to 471424 | Laboratory for Respiratory Viruses, National Influenza Centre, Cantacuzino National Military-Medical Institute for Research and Development | Cantacuzino Institute | Luiza Ustea; Mihaela Lazar; Nicoleta Paraschiv; Tim Durfee |
| EPI_ISL_437689 | Laboratory for Urgent Response to Biological Threats | Institut Pasteur CIBU / ERI | A. Kwasiborski; C. Balière; C. Batéjat; J. Vanhomwegen; JC. Manuguerra; V. Caro; V. Hourdel |
| EPI_ISL_437690 | Laboratory for Urgent Response to Biological Threats | Institut Pasteur CIBU /ERI | A. Kwasiborski; C. Balière; C. Batéjat; H. Hourdel; J. Vanhomwegen; JC. Manuguerra; V. Caro |
| EPI_ISL_572398 | Laboratory of Biology and Identification of Arboviruses | Pathogenic Microorganisms Variability Laboratory | Alexander Gintsburg; Alexey Prilipov; Alexey Shchetinin; Alina Kozlova; Andrei Siniavin; Maria Nikiforova; Muhammad Saifullin; Victor Larichev; Vladimir Gushchin |
| EPI_ISL_735392 to 735395, EPI_ISL_760059, EPI_ISL_760246 | Laboratory of Genetics and Personalized Medicine, Zan Mitrev Clinic | Laboratory of Genetics and Personalized Medicine, Zan Mitrev Clinic | 2020; Kungulovski et al.; Kungulovski et al. 2020 |
| EPI_ISL_435723 | Laboratory of Genomics & Bioinformatics, Institute of Immunology and Experimental Therapy, Polish Academy of Sciences Oddzia Mikrobiologii Wojewódzkiej Stacji Sanitarno Epidemiologiczna | Laboratory of Genomics & Bioinformatics, Institute of Immunology and Experimental Therapy, Polish Academy of Sciences | Aleksandra Herud; Dariusz Martynowski; Dorota Kujawa; Grayna Zalewska; Joanna Sikorska; Krzysztof Jakub Pawlik; Oskar Karpiski and ukasz aczmaski; Paulina ebrowska |
| EPI_ISL_437625 | Laboratory of Genomics & Bioinformatics, Institute of Immunology and Experimental Therapy, Polish Academy of Sciences Oddzial Mikrobiologii Wojewodzkiej Stacji Sanitarno-Epidemiologicznej. | Laboratory of Genomics & Bioinformatics, Institute of Immunology and Experimental Therapy, Polish Academy of Sciences | Aleksandra Herud; Dariusz Martynowski; Dorota Kujawa; Grazyna Zalewska; Joanna Sikorska; Krzysztof Jakub Pawlik; Oskar Karpinski and Lukasz Laczmanski; Paulina Zebrowska |
| EPI_ISL_452327, EPI_ISL_452329, EPI_ISL_452333 to 452337, EPI_ISL_452342 to 452356 | Laboratory of Infectious Diseases Center of Beijing Ditan Hospital | Laboratory of Infectious Diseases Center of Beijing Ditan Hospital | Chengjie Jie; Fengting Yu; Linghang Wang; Liting Yan; Siyuan Yang; Yunxia Tang |
| EPI_ISL_417445 to 417447, EPI_ISL_778860, EPI_ISL_779704, EPI_ISL_779709, EPI_ISL_779712 to 779714, EPI_ISL_780375 to 780376, EPI_ISL_780378, EPI_ISL_780381, EPI_ISL_780411 to 780413 | | | |
| see above | Laboratory of Infectious Diseases, Department of Biomedical and Clinical Sciences L. Sacco, University of Milan | Laboratory of Infectious Diseases, Department of Biomedical and Clinical Sciences L. Sacco, University of Milan | Agostino Riva; Alessia Lai; Annalisa Bergna; Arianna Gabrieli; Carla Della Ventura; Claudia Balotta; Dario Bernacchia; Gianguglielmo Zehender; Gianguglielmo Zehender on behalf of SARS-CoV-2 ITALIAN RESEARCH ENTERPRISE-(SCIRE) Collaborative Group; Gianguglielmo Zehender on behalf of SARS-CoV-2 ITALIAN RESEARCH ENTERPRISE-(SCIRE) Collaborative Group; Giuliano Rizzardini; Luca Meroni; Maciej Tarkowski; Massimo Galli; Spinello Antinori; Stefano Rusconi |
| EPI_ISL_418263 to 418265 | Laboratory of Microbiology, Department of Medicine, National and Kapodistrian University of Athens, Greece | Laboratory of Biology, Department of Medicine, Democritus University of Thrace, Greece | Elisavet Gatzidou; Ioannis Karakasiliotis; Maria Bampali; Nikolaos Dovrolis; Nikolaos Spanakis; Stavroula Veletza |
| EPI_ISL_427043, EPI_ISL_434455 to 434486, EPI_ISL_437874 to 437911 | Laboratory of Microbiology, Medical School, National and Kapodistrian University of Athens | Laboratory of Biology, Department of Medicine, Democritus University of Thrace | Bampali, M.; Dovrolis, N.; Froukala, E.; Gatzidou, E.; Kassela K.; N. and Karakasiliotis, I.; Spanakis; Stavropoulou, A.; Tsakris, A.; Veletza, S. |
| EPI_ISL_428209, EPI_ISL_450746, EPI_ISL_451644 to 451647 | Laboratory of Molecular Biology, Diagnostyka sp. z o.o. | Laboratory of Recombinant Vaccines | Anna Piotrowska-Mietelska; Boguslaw Szewczyk; Krystyna Bienkowska-Szewczyk; Lukasz Rabalski; Maciej Kosinski |
| EPI_ISL_852842 | Laboratory of Molecular Biology, Diagnostyka sp. z o.o. | genXone SA, Research & Development Laboratory | Grzegorz Nowicki; Jakub Grabowski; Maciej Sykulski; Micha Kaszuba; Natalia Drwska-Matelska; ukasz Krych |
| EPI_ISL_425228 to 425229, EPI_ISL_426051, EPI_ISL_426356 to 426360 | Laboratory of Molecular Genetics, 2nd Faculty of Medicine, Charles University in Prague, Prague, Czech Republic | Laboratory of Molecular Genetics, 2nd Faculty of Medicine, Charles University in Prague, Prague, Czech Republic | Katerina Polackova; Kateina Poláková; Lenka Kramna; Lenka Kramná; Ondrej Cinek; Ondej Cinek |
| EPI_ISL_428853, EPI_ISL_498558 to 498563 | Laboratory of Molecular Virology International Center for Genetic Engineering and Biotechnology (ICGEB) | ARGO Open Lab Platform for Genome Sequencing | D'Agaro P; Dal Monego S; D'Agaro P; Licastro D; Marcello A; Rajasekharan S; Segat L |
| EPI_ISL_417419, EPI_ISL_417421, EPI_ISL_417423, EPI_ISL_428854 | Laboratory of Molecular Virology International Center for Genetic Engineering and Biotechnology (ICGEB) | ARGO Open Lab Platform for Genome sequencing | D'Agaro P; Dal Monego S; Licastro D; Marcello A; Rajasekharan; Rajasekharan S; Segat L |
| EPI_ISL_417418 | Laboratory of Molecular Virology International Center fro Genetic Engineering and Biotechnology (ICGEB) | ARGO Open Lab Platform for Genome sequencing | D'Agaro P; Dal Monego S; Licastro D; Marcello A; Rajasekharan S; Segat L |
| EPI_ISL_479616 to 479619, EPI_ISL_479790 to 479791, EPI_ISL_525495 to 525496 | Laboratory of Molecular Virology of the International Centre for Genetic Engineering and Biotechnology (ICGEB) | ARGO Open Lab Platform for Genome Sequencing | Confalonieri M; Confalonieri M Marcello A; Confalonieri P; D; D'Agaro P; Dal Monego S; Licastro; Licastro D; Marcello A; Rajasekharan S; Salton F; Segat L |
| EPI_ISL_584048 to 584049, EPI_ISL_584051 to 584052 | Laboratory of Molecular Virology, Department of Biomedical, Surgical and Dental Sciences University of Milano | Laboratory of Molecular Virology, Department of Biomedical, Surgical and Dental Sciences University of Milano | Anselmi; B. and Ferrante, P.; Basilico, N.; Bianchi, M.; Binda, S.; C. and Ferrante, P.; D'Alessandro, S.; Delbue, S.; Dolci, M.; E; Fattori, M.; Ferrante, P.; G. and Pariani; Galli; Galli, C.; Incorvaia; Modenese, A.; Parapini, S.; Pariani, E.; Primache, V.; Signorini, L.; Taramelli, D. |
| EPI_ISL_451298 to 451305 | Laboratory of Virology, INMI Lazzaro Spallanzani IRCCS | Laboratory of Virology, INMI Lazzaro Spallanzani IRCCS | Antonino Di Caro; Barbara Bartolini; Cesare E.M. Gruber; Francesco Messina; Giuseppe Ippolito; Maria R. Capobianchi; Martina Rueca |
| EPI_ISL_576117 to 576118, EPI_ISL_576120 to | Laboratory, The Bio Arte Limited | Laboratory, The Bio Arte Limited | Biazzo, M.; Madeddu, S.; Pinzauti, D.; Santoro, F. |

| | | | |
|---|---|---|---|
| 576126 | | | |
| EPI_ISL_541370 to 541371 | Laboratório Central de Saúde Pública do Estado de Santa Catarina (LACEN-SC) | Laboratory of Respiratory Viruses and Measles, Oswaldo Cruz Institute, FIOCRUZ | Ana Carolina Mendonça; Anna Carolina Paixão; Fernando Motta; Jonathan Lopes; Luciana Appolinario; Marilda Siqueira on behalf of the Fiocruz COVID-19 Genomic Surveillance Network; Paola Resende; Sandra Bianchini |
| EPI_ISL_541375 | Laboratório Central de Saúde Pública do Estado de Sergipe (LACEN-SE) | Laboratory of Respiratory Viruses and Measles, Oswaldo Cruz Institute, FIOCRUZ | Ana Carolina Mendonça; Anna Carolina Paixão; Clioma Santos; Fernando Motta; Jonathan Lopes; Luciana Appolinario; Marilda Siqueira on behalf of the Fiocruz COVID-19 Genomic Surveillance Network; Paola Resende |
| EPI_ISL_541341, EPI_ISL_541345 | Laboratório Central de Saúde Pública do Estado do Paraná (LACEN-PR) | Laboratory of Respiratory Viruses and Measles, Oswaldo Cruz Institute, FIOCRUZ | Ana Carolina Mendonça; Anna Carolina Paixão; Fernando Motta; Irina Riediger; Jonathan Lopes; Luciana Appolinario; Maria do Carmo Debur; Marilda Siqueira on behalf of the Fiocruz COVID-19 Genomic Surveillance Network; Paola Resende |
| EPI_ISL_861856 to 861863 | Labormedizinisches Zentrum Dr Risch | University Hospital Basel, Clinical Bacteriology | Adrian Egli; Alfredo Mari; Hans Hirsch; Helena MB Seth-Smith; Julia Bielicki; Karoline Leuzinger; Lorenz Risch; Madlen Stange; Manuel Battegay; Martin Risch; Nadia Wohlwend; Tim Roloff |
| EPI_ISL_434669, EPI_ISL_475515 | Lakargruppen | The Public Health Agency of Sweden | Anna Risberg; Anna-Malin Linde; Boris Klanger; Karin Tegmark-Wisell; Maria Lind Karlberg; Mattias Haukland; Mia Brytting; Olov Svartstrom; Oskar Karlsson Lindsjo; Reza Advani; Sandra Broddesson; Theresa Enkirch |
| EPI_ISL_738317 to 738322, EPI_ISL_738324 to 738349 | Landstuhl Regional Medical Center | United States Air Force School of Aerospace Medicine | Amanda Javorina; Anthony Fries; Clarise Starr; Cole Anderson; Elizabeth Macias; Fritz Castillo; Jennifer Meyer; Sarah Purves; William Gruner |
| EPI_ISL_534237 to 534238, EPI_ISL_548255 | Lanssjukhuset Kalmar | The Public Health Agency of Sweden | Anna Risberg; Anna-Malin Linde; Karin Tegmark-Wisell; Maria Lind Karlberg; Mattias Haukland; Mia Brytting; Olov Svartstrom; Oskar Karlsson Lindsjo; Petra Edquist; Reza Advani; Sandra Broddesson |
| EPI_ISL_437089 to 437096, EPI_ISL_486422 to 486426, EPI_ISL_486428, EPI_ISL_486430 to 486436, EPI_ISL_512647 to 512652, EPI_ISL_515185 to 515186, EPI_ISL_534204 to 534205, EPI_ISL_534209, EPI_ISL_639634, EPI_ISL_639639, EPI_ISL_770047, EPI_ISL_770060 | | | |
| see above | Latvijas Infektoloijas centrs | Latvian Biomedical Research and Study Centre | Ivars Silamielis; Jeena Storoženko; Jnis Kloviš; Jnis Pjalkovskis; Kaspars Megnis; Monta Ustinova; Oksana Savicka; Tatjana Kolupajeva; Uga Dumpis; Vita Rovte; ikita Zrelovs |
| EPI_ISL_421507 to 421508 | Le Château de Seine-Port | National Reference Center for Viruses of Respiratory Infections, Institut Pasteur, Paris | Angela Brisebarre; Etienne Simon-Lorière; Flora Donati; Marion Barbet; Maud Vanpeene; Mélanie Albert; Méline Bizard; Sylvie Behillil; Sylvie van der Werf; Vincent Enouf |
| EPI_ISL_538811, EPI_ISL_538818, EPI_ISL_538841, EPI_ISL_538851, EPI_ISL_538863, EPI_ISL_538866, EPI_ISL_538917, EPI_ISL_538945, EPI_ISL_538987, EPI_ISL_538994, EPI_ISL_539006, EPI_ISL_539030, EPI_ISL_539070, EPI_ISL_539116 to 539117, EPI_ISL_539120, EPI_ISL_539143, EPI_ISL_539150, EPI_ISL_539152, EPI_ISL_539157, EPI_ISL_539185, EPI_ISL_539197, EPI_ISL_539213 | | | |
| see above | Leeds Teaching Hospitals NHS Trust and Public Health England, National Infection Service (Leeds laboratory) | Wellcome Sanger Institute for the COVID-19 Genomics UK (COG-UK) Consortium | Antony Hale and Alex Alderton; Cordelia Langford; David K. Jackson; Dominic Kwiatkowski; Ewan Harrison; Holli Carden; Ian Johnston; John Sillitoe on behalf of the Wellcome Sanger Institute COVID-19 Surveillance Team; Katherine L. Harper; Louissa Macfarlane-Smith; Roberto Amato; Sonia Goncalves |
| EPI_ISL_538748 to 538771, EPI_ISL_538773 to 538786, EPI_ISL_538788 to 538800, EPI_ISL_538802 to 538810, EPI_ISL_538812 to 538816, EPI_ISL_538819 to 538832, EPI_ISL_538834 to 538840, EPI_ISL_538843 to 538850, EPI_ISL_538852 to 538854, EPI_ISL_538856 to 538862, EPI_ISL_538864 to 538865, EPI_ISL_538867 to 538872, EPI_ISL_538874 to 538898, EPI_ISL_538900 to 538915, EPI_ISL_538918 to 538933, EPI_ISL_538935 to 538944, EPI_ISL_538946, EPI_ISL_538948, EPI_ISL_538950 to 538963, EPI_ISL_538965 to 538986, EPI_ISL_538988 to 538992, EPI_ISL_538995 to 539005, EPI_ISL_539007 to 539029, EPI_ISL_539031 to 539064, EPI_ISL_539066 to 539069, EPI_ISL_539071 to 539093, EPI_ISL_539096 to 539098, EPI_ISL_539100 to 539115, EPI_ISL_539118 to 539119, EPI_ISL_539121 to 539124, EPI_ISL_539126 to 539142, EPI_ISL_539144 to 539146, EPI_ISL_539148 to 539149, EPI_ISL_539151, EPI_ISL_539153 to 539156, EPI_ISL_539158 to 539164, EPI_ISL_539167 to 539169, EPI_ISL_539171 to 539184, EPI_ISL_539186 to 539196, EPI_ISL_539198 to 539212, EPI_ISL_539214 to 539215, EPI_ISL_539217 to 539220 | | | |
| see above | Leeds Teaching Hospitals NHS Trust and Public Health England, National Infection Service (Leeds laboratory) | Wellcome Sanger Institute for the COVID-19 Genomics UK (COG-UK) consortium | Antony Hale and Alex Alderton; Cordelia Langford; David K. Jackson; Dominic Kwiatkowski; Ewan Harrison; Holli Carden; Ian Johnston; John Sillitoe on behalf of the Wellcome Sanger Institute COVID-19 Surveillance Team; Katherine L. Harper; Louissa Macfarlane-Smith; Roberto Amato; Sonia Goncalves |
| EPI_ISL_551294, EPI_ISL_551327, EPI_ISL_551412, EPI_ISL_551453, EPI_ISL_551484, EPI_ISL_551496, EPI_ISL_551530, EPI_ISL_552492, EPI_ISL_552972 to 552973, EPI_ISL_553421, EPI_ISL_553568, EPI_ISL_553778, EPI_ISL_553784, EPI_ISL_553812, EPI_ISL_553846, EPI_ISL_553866, EPI_ISL_553946, EPI_ISL_554478, EPI_ISL_554484, EPI_ISL_554497, EPI_ISL_554537, EPI_ISL_554667, EPI_ISL_554709, EPI_ISL_554763, EPI_ISL_554845, EPI_ISL_554868, EPI_ISL_554929, EPI_ISL_555041, EPI_ISL_555129, EPI_ISL_555148, EPI_ISL_555259, EPI_ISL_555303 to 555304, EPI_ISL_555391, EPI_ISL_555398, EPI_ISL_555432 to 555433, EPI_ISL_555502, EPI_ISL_555510 to 555511, EPI_ISL_555650 to 555651, EPI_ISL_555660, EPI_ISL_555794, EPI_ISL_555807, EPI_ISL_555815, EPI_ISL_555887, EPI_ISL_555947, EPI_ISL_555986, EPI_ISL_555990, EPI_ISL_556000, EPI_ISL_556141, EPI_ISL_556348, EPI_ISL_556350, EPI_ISL_556411, EPI_ISL_556433, EPI_ISL_556435, EPI_ISL_556505, EPI_ISL_556510, EPI_ISL_557002, EPI_ISL_557043, EPI_ISL_557110, EPI_ISL_557142, EPI_ISL_557625, EPI_ISL_557711, EPI_ISL_557728, EPI_ISL_557730, EPI_ISL_557739, EPI_ISL_557760, EPI_ISL_557852, EPI_ISL_557896, EPI_ISL_557999, EPI_ISL_558024, EPI_ISL_558126, EPI_ISL_558185, EPI_ISL_558222, EPI_ISL_558362, EPI_ISL_558387, EPI_ISL_558422, EPI_ISL_558432, EPI_ISL_558543, EPI_ISL_558605, EPI_ISL_558617, EPI_ISL_558796, EPI_ISL_558846, EPI_ISL_558872, EPI_ISL_558877, EPI_ISL_558888, EPI_ISL_558950, EPI_ISL_558994, EPI_ISL_559042, EPI_ISL_559046, EPI_ISL_559120, EPI_ISL_559129, EPI_ISL_559213, EPI_ISL_559228, EPI_ISL_559261, EPI_ISL_559275, EPI_ISL_559353, EPI_ISL_559447, EPI_ISL_559576, EPI_ISL_559591, EPI_ISL_559616, EPI_ISL_559644, EPI_ISL_567811, EPI_ISL_575536, EPI_ISL_579951, EPI_ISL_579971, EPI_ISL_580062, EPI_ISL_580138, EPI_ISL_580187, EPI_ISL_580210, EPI_ISL_580285, EPI_ISL_580563, EPI_ISL_580686, EPI_ISL_580708, EPI_ISL_580780, EPI_ISL_580824, EPI_ISL_581423, EPI_ISL_586617, EPI_ISL_586621, EPI_ISL_586709, EPI_ISL_586907, EPI_ISL_587085, EPI_ISL_587265, EPI_ISL_587977, EPI_ISL_588007, EPI_ISL_588026, EPI_ISL_589289, EPI_ISL_589302, EPI_ISL_589329, EPI_ISL_589342, EPI_ISL_589344, EPI_ISL_589376, EPI_ISL_597633, EPI_ISL_597984, EPI_ISL_598163, EPI_ISL_606948, EPI_ISL_606960, EPI_ISL_607189, EPI_ISL_607200, EPI_ISL_607894, EPI_ISL_608147, EPI_ISL_608216, EPI_ISL_608310, EPI_ISL_609209, EPI_ISL_610332, EPI_ISL_610502 to 610503, EPI_ISL_611093, EPI_ISL_623864, EPI_ISL_623904, EPI_ISL_623974, EPI_ISL_623983, EPI_ISL_624094, EPI_ISL_624244, EPI_ISL_624291, EPI_ISL_624350, EPI_ISL_624354, EPI_ISL_624909, EPI_ISL_625213, EPI_ISL_625240, EPI_ISL_630474, EPI_ISL_630533, EPI_ISL_630547, EPI_ISL_630780, EPI_ISL_630891, EPI_ISL_644095 to 644098, EPI_ISL_644100, EPI_ISL_644103 to 644106, EPI_ISL_644108, EPI_ISL_644110 to 644111, EPI_ISL_644113, EPI_ISL_644118, EPI_ISL_645240, EPI_ISL_645266, EPI_ISL_645297, EPI_ISL_645303, EPI_ISL_645362, EPI_ISL_645368, EPI_ISL_645418, EPI_ISL_645452, EPI_ISL_645459, EPI_ISL_645464, EPI_ISL_645473, EPI_ISL_645499, EPI_ISL_645502, EPI_ISL_645508, EPI_ISL_645530 to 645540, EPI_ISL_645542 to 645713, EPI_ISL_645715 to 645786, EPI_ISL_645788 to 645809, EPI_ISL_645811 to 645821, EPI_ISL_645823 to 645825, EPI_ISL_645827, EPI_ISL_645830 to 645831, EPI_ISL_645833 to 645835, EPI_ISL_645837, EPI_ISL_645839, EPI_ISL_645841 to 645849, EPI_ISL_645853 to 645865, EPI_ISL_645867, EPI_ISL_645870 to 645876, EPI_ISL_645878 to 645879, EPI_ISL_645881, EPI_ISL_645883 to 645884, EPI_ISL_645887 to 645895, EPI_ISL_645898, EPI_ISL_645900 to 645901, EPI_ISL_645903 to 645904, EPI_ISL_645906 to 645907, EPI_ISL_645909 to 645915, EPI_ISL_645919, EPI_ISL_645921 to 645922, EPI_ISL_645924, EPI_ISL_645928 to 645929, EPI_ISL_645931 to 645944, EPI_ISL_645946 to 645948, EPI_ISL_645950 to 645957, EPI_ISL_645960, EPI_ISL_645963 to 645967, EPI_ISL_645969, EPI_ISL_645971 to 645978, EPI_ISL_645982, EPI_ISL_645984 to 645986, EPI_ISL_645988, EPI_ISL_645990 to 645991, EPI_ISL_645996 to 645997, EPI_ISL_645999 to 646008, EPI_ISL_646010 to 646013, EPI_ISL_646015 to 646016, EPI_ISL_646019 to 646021, EPI_ISL_646023 to 646026, EPI_ISL_646029, EPI_ISL_646033, EPI_ISL_646036 to 646038, EPI_ISL_646040, EPI_ISL_646042 to 646045, EPI_ISL_646048 to 646051, EPI_ISL_646053 to 646055, EPI_ISL_646057, EPI_ISL_646059 to 646066, EPI_ISL_646068 to 646069, EPI_ISL_646071 to 646075, EPI_ISL_646077 to 646082, EPI_ISL_646087 to 646088, EPI_ISL_646090, EPI_ISL_646092 to 646100, EPI_ISL_646103 to 646104, EPI_ISL_646106 to 646114, EPI_ISL_646116 to 646121, EPI_ISL_646123 to 646127, EPI_ISL_646130, EPI_ISL_646132 to 646134, EPI_ISL_646137 to 646138, EPI_ISL_646148, EPI_ISL_646150, EPI_ISL_646155 to 646157, EPI_ISL_646162, EPI_ISL_646166, EPI_ISL_646169 to 646170, EPI_ISL_646173, EPI_ISL_646180, EPI_ISL_646182, EPI_ISL_646185, EPI_ISL_646189, EPI_ISL_646191, EPI_ISL_646193, EPI_ISL_646198 to 646199, EPI_ISL_646204, EPI_ISL_646207 to 646208, EPI_ISL_646212, EPI_ISL_646215, EPI_ISL_646224, EPI_ISL_646231, EPI_ISL_646235, EPI_ISL_646242, EPI_ISL_646245, EPI_ISL_646248 to 646249, EPI_ISL_646252, EPI_ISL_646261, EPI_ISL_646268 to 646269, EPI_ISL_646271, EPI_ISL_646276 to 646279, EPI_ISL_646283, EPI_ISL_646291, EPI_ISL_646297, EPI_ISL_646299, EPI_ISL_646303, EPI_ISL_646305, EPI_ISL_646307, EPI_ISL_646309, EPI_ISL_646313, EPI_ISL_646318, EPI_ISL_646320, EPI_ISL_646323, EPI_ISL_646327, EPI_ISL_646334, EPI_ISL_646342 to 646343, EPI_ISL_646345 to 646347, EPI_ISL_646349 to 646350, EPI_ISL_646354, EPI_ISL_646356, EPI_ISL_646362 to 646363, EPI_ISL_646366, EPI_ISL_646369 to 646370, EPI_ISL_646373, EPI_ISL_646375 to 646377, EPI_ISL_646382, EPI_ISL_646385, EPI_ISL_646388, EPI_ISL_646397, EPI_ISL_646402, EPI_ISL_646412, EPI_ISL_646418 to 646420, EPI_ISL_646426, EPI_ISL_646428 to 646432, EPI_ISL_646434, EPI_ISL_646441 to 646443, EPI_ISL_646448, EPI_ISL_646454, EPI_ISL_646463, EPI_ISL_646466 to 646468, EPI_ISL_646476 to 646479, EPI_ISL_646485 to 646494, EPI_ISL_646497, EPI_ISL_646500, EPI_ISL_646502, EPI_ISL_646505 to 646506, EPI_ISL_646508 to 646516, EPI_ISL_646518, EPI_ISL_646520 to 646522, EPI_ISL_646525 to 646526, EPI_ISL_646528 to 646529, EPI_ISL_646532 to 646533, EPI_ISL_646535, EPI_ISL_646537 to 646538, EPI_ISL_646540 to 646545, EPI_ISL_646548 to 646549, EPI_ISL_646551, EPI_ISL_646555, EPI_ISL_646557 to 646561, EPI_ISL_646563 to 646564, EPI_ISL_646566 to 646569, EPI_ISL_646571 to 646573, EPI_ISL_646576 to 646577, EPI_ISL_646580 to 646581, EPI_ISL_646583, EPI_ISL_646585 to 646586, EPI_ISL_646591 to 646592, EPI_ISL_646594 to 646595, EPI_ISL_646597, EPI_ISL_646599 to 646601, EPI_ISL_646603, EPI_ISL_646605 to 646607, EPI_ISL_646610, EPI_ISL_646612 to 646614, EPI_ISL_646616, EPI_ISL_646618, EPI_ISL_646621, EPI_ISL_646623, EPI_ISL_646625 to 646626, EPI_ISL_646628, EPI_ISL_646631, EPI_ISL_646635, EPI_ISL_646638 to 646640, EPI_ISL_646642, EPI_ISL_646648, EPI_ISL_646651 to 646653, EPI_ISL_646655, EPI_ISL_646658, EPI_ISL_646660, EPI_ISL_646662, EPI_ISL_646664 to 646666, EPI_ISL_646668, EPI_ISL_646671, EPI_ISL_646673, EPI_ISL_646675 to 646676, EPI_ISL_646678, EPI_ISL_646683 to 646684, EPI_ISL_646686 to 646687, EPI_ISL_646689, EPI_ISL_646691 to 646693, EPI_ISL_646695 to 646698, EPI_ISL_646702, EPI_ISL_646706 to 646708, EPI_ISL_646710 to 646713, EPI_ISL_646715 to 646716, EPI_ISL_646718 to 646719, EPI_ISL_646721 to 646722, EPI_ISL_646724, EPI_ISL_646727, EPI_ISL_646731, EPI_ISL_646734, EPI_ISL_646736 to 646741, EPI_ISL_646743 to 646744, EPI_ISL_646746 to 646747, EPI_ISL_646749, EPI_ISL_646751 to 646753, EPI_ISL_646755 to 646758, EPI_ISL_646760 to 646763, EPI_ISL_646767 to 646771, EPI_ISL_646773, EPI_ISL_646776, EPI_ISL_646782, EPI_ISL_646784, EPI_ISL_646787, EPI_ISL_646789 to 646791, EPI_ISL_646793, EPI_ISL_646795 to 646797, EPI_ISL_646799 to 646809, EPI_ISL_646811 to 646817, EPI_ISL_646819 to 646824, EPI_ISL_646827 to 646836, EPI_ISL_646838 to 646839, EPI_ISL_646841 to 646849, EPI_ISL_646851 to 646852, EPI_ISL_646855 to 646867, EPI_ISL_646871, EPI_ISL_646873, EPI_ISL_646875 to 646876, EPI_ISL_646878 to 646879, EPI_ISL_646881 to 646884, EPI_ISL_646886 to 646890, EPI_ISL_646893 to 646897, EPI_ISL_646899 to 646901, EPI_ISL_646903 to 646904, EPI_ISL_646906 to 646911, EPI_ISL_646913 to 646914, EPI_ISL_646916, EPI_ISL_646918 to 646919, EPI_ISL_646922 to 646928, EPI_ISL_646930 to 646935, EPI_ISL_646938, EPI_ISL_646940 to 646947, EPI_ISL_646949 to 646954, EPI_ISL_646956 to 646959, EPI_ISL_646961 to 646963, EPI_ISL_646965 to 646966, EPI_ISL_646968 to 646973, EPI_ISL_646978 to 646982, EPI_ISL_646984 to 646987, EPI_ISL_646989 to 646991, EPI_ISL_646993 to 646995, EPI_ISL_646997 to 647000, EPI_ISL_647002, EPI_ISL_647004 to 647005, EPI_ISL_647007 to 647013, EPI_ISL_647015 to 647017, EPI_ISL_647019 to 647022, EPI_ISL_647024 to 647030, EPI_ISL_647033 to 647034, EPI_ISL_647036 to 647037, EPI_ISL_647039 to 647044, EPI_ISL_647046 to 647049, EPI_ISL_647051 to 647052, EPI_ISL_647054 to 647058, EPI_ISL_647060, EPI_ISL_647062 to 647066, EPI_ISL_647068 to 647072, EPI_ISL_647074 to 647075, EPI_ISL_647077 to 647080, EPI_ISL_647082 to 647086, EPI_ISL_647088 to 647089, EPI_ISL_647091 to 647096, EPI_ISL_647098 to 647099, EPI_ISL_647101, EPI_ISL_647103 to 647104, EPI_ISL_647106 to 647108, EPI_ISL_647111 to 647113, EPI_ISL_647116, EPI_ISL_647118 to 647120, EPI_ISL_647122, EPI_ISL_647468 to 647469, EPI_ISL_647473 to 647474, EPI_ISL_647477, EPI_ISL_647497, EPI_ISL_647499, EPI_ISL_647503, EPI_ISL_647506, EPI_ISL_647511, EPI_ISL_647513, EPI_ISL_647526, EPI_ISL_647528, EPI_ISL_647535 to 647536, EPI_ISL_647541, EPI_ISL_647546, EPI_ISL_647550, EPI_ISL_647553, EPI_ISL_647558, EPI_ISL_647560, EPI_ISL_647572 to 647573, EPI_ISL_647576, EPI_ISL_647579 to 647580, EPI_ISL_647584, EPI_ISL_647591, EPI_ISL_647594, EPI_ISL_647603, EPI_ISL_647606, EPI_ISL_647608, EPI_ISL_647612, EPI_ISL_647615, EPI_ISL_647618, EPI_ISL_647624, EPI_ISL_647630, EPI_ISL_647632, EPI_ISL_647634 to 647635, EPI_ISL_647638, EPI_ISL_647643, EPI_ISL_647650, EPI_ISL_647655, EPI_ISL_647659, EPI_ISL_647667 to 647668, EPI_ISL_647672, EPI_ISL_647677, EPI_ISL_647682 to 647683, EPI_ISL_647693, EPI_ISL_647695, EPI_ISL_647698, EPI_ISL_647700, EPI_ISL_647704 to 647705, EPI_ISL_647708, EPI_ISL_647711, EPI_ISL_647726, EPI_ISL_647733, EPI_ISL_647741, EPI_ISL_647750, EPI_ISL_647752, EPI_ISL_647762, EPI_ISL_647771, EPI_ISL_647776, EPI_ISL_647780, EPI_ISL_647786 to 647787, EPI_ISL_647792, EPI_ISL_649193 to 649204, EPI_ISL_649206 to 649254, EPI_ISL_649256 to 649266, EPI_ISL_649268 to 649282, EPI_ISL_649284 to 649319, EPI_ISL_649321, EPI_ISL_649323 to 649361, EPI_ISL_649363 to 649385, EPI_ISL_649387 to 649404, EPI_ISL_649406 to 649423, EPI_ISL_649425 to 649460, EPI_ISL_649462 to 649469, EPI_ISL_649471 to 649478, EPI_ISL_649480 to 649481, EPI_ISL_649483 to 649491, EPI_ISL_655273 to 655288, EPI_ISL_655290 to 655321, EPI_ISL_655323 to 655424, EPI_ISL_655426 to 655444, EPI_ISL_655446 to 655517, EPI_ISL_655519 to 655538, EPI_ISL_655540 to 655574, EPI_ISL_655576 to 655650, EPI_ISL_655652 to 655659, EPI_ISL_655661, EPI_ISL_655663 to 655671, EPI_ISL_655673 to 655676, EPI_ISL_655679 to 655693, EPI_ISL_655696, EPI_ISL_655698, EPI_ISL_655700 to 655701, EPI_ISL_655703 to 655710, EPI_ISL_655712 to 655718, EPI_ISL_655720 to 655722, EPI_ISL_655724 to 655726, EPI_ISL_655728 to 655734, EPI_ISL_655736, EPI_ISL_655738, EPI_ISL_655740 to 655743, EPI_ISL_655746 to 655755, EPI_ISL_655757 to 655761, EPI_ISL_655763 to 655767, EPI_ISL_655771 to 655772, EPI_ISL_655774 to 655778, EPI_ISL_655781 to 655782, EPI_ISL_655784 to 655787, EPI_ISL_655789 to 655797, EPI_ISL_655799 to 655803, EPI_ISL_655805, EPI_ISL_655808, EPI_ISL_655810 to 655816, EPI_ISL_655818, EPI_ISL_655820 to 655821, EPI_ISL_655823 to 655827, EPI_ISL_655829 to 655831, EPI_ISL_655833 to 655840, EPI_ISL_655842 to 655846, EPI_ISL_655849, EPI_ISL_655851 to 655853, EPI_ISL_655855 to 655860, EPI_ISL_655862 to 655863, EPI_ISL_655866 to 655867, EPI_ISL_655869 to 655872, EPI_ISL_655875 to 655878, EPI_ISL_655880 to 655882, EPI_ISL_655884 to 655890, EPI_ISL_655892 to 655906, EPI_ISL_655908 to 655910, EPI_ISL_655913 to 655917, EPI_ISL_655919, EPI_ISL_655921 to 655923, EPI_ISL_655925 to 655928, EPI_ISL_655930 to 655932, EPI_ISL_655934 to 655946, EPI_ISL_655949, EPI_ISL_655951, EPI_ISL_655953 to 655956, EPI_ISL_655958, EPI_ISL_655960 to 655966, EPI_ISL_655968 to 655970, EPI_ISL_655972, EPI_ISL_655974, EPI_ISL_655976 to 656108, EPI_ISL_656110 to 656148, EPI_ISL_656150 to 656173, EPI_ISL_656175 to 656209, EPI_ISL_656211 to 656221, EPI_ISL_656223 to 656263, EPI_ISL_656265 to 656359, EPI_ISL_656362 to 656372, EPI_ISL_656374 to 656380, EPI_ISL_656382 to 656384, EPI_ISL_656386, EPI_ISL_656388 to 656389, EPI_ISL_656391 to 656397, EPI_ISL_656399 to 656412, EPI_ISL_656414, EPI_ISL_656416 to 656420, EPI_ISL_656422, EPI_ISL_656424 to 656427, EPI_ISL_656429 to 656452, EPI_ISL_656455 to 656467, EPI_ISL_656469, EPI_ISL_656472 to 656479, EPI_ISL_656481 to 656497, EPI_ISL_656499 to 656503, EPI_ISL_656505 to 656508, EPI_ISL_656510 to 656519, EPI_ISL_656521 to 656534, EPI_ISL_656536, EPI_ISL_656538 to 656544, EPI_ISL_656546 to 656547, EPI_ISL_656549 to 656571, EPI_ISL_656573 to 656579, EPI_ISL_656581 to 656587, EPI_ISL_656590 to 656610, EPI_ISL_656613, EPI_ISL_656615 to 656620, EPI_ISL_656622 to 656625, EPI_ISL_656628 to 656631, EPI_ISL_656633 to 656642, EPI_ISL_656644 to 656645, EPI_ISL_656647 to 656671, EPI_ISL_656673 to 656676, EPI_ISL_656678 to 656682, EPI_ISL_656684, EPI_ISL_661956, EPI_ISL_661958, EPI_ISL_661961 to 661962, EPI_ISL_661964 to 661966, EPI_ISL_661968, EPI_ISL_661970, EPI_ISL_661974, EPI_ISL_661977 to 661978, EPI_ISL_661980, EPI_ISL_661982 to 661983, EPI_ISL_661985 to 661986, EPI_ISL_661988 to 661989, EPI_ISL_661991 to 661992, EPI_ISL_661995, EPI_ISL_661998 to 662002, EPI_ISL_662004, EPI_ISL_662006, EPI_ISL_662008, EPI_ISL_662013 to 662015, EPI_ISL_662017 to 662020, EPI_ISL_662022, EPI_ISL_662025, EPI_ISL_662029, EPI_ISL_662032, EPI_ISL_662034, EPI_ISL_662036, EPI_ISL_662038, EPI_ISL_662043, EPI_ISL_662045 to 662051, EPI_ISL_662053, EPI_ISL_662055 to 662058, EPI_ISL_662060 to 662063, EPI_ISL_662065, EPI_ISL_662067 to 662068, EPI_ISL_662070 to 662071, EPI_ISL_662074, EPI_ISL_662078 to 662081, EPI_ISL_662086, EPI_ISL_662093, EPI_ISL_662097 to 662101, EPI_ISL_662105 to 662108, EPI_ISL_662114 to 662116, EPI_ISL_662118, EPI_ISL_662120, EPI_ISL_662122 to 662124, EPI_ISL_662129, EPI_ISL_662131 to 662133, EPI_ISL_662135, EPI_ISL_662137 to 662138, EPI_ISL_662140 to 662141, EPI_ISL_662143, EPI_ISL_662145 to 662146, EPI_ISL_662150 to 662151, EPI_ISL_662156, EPI_ISL_662159 to 662160, EPI_ISL_662162 to 662167, EPI_ISL_662169 to 662170, | | | |

EPI_ISL_662173 to 662179, EPI_ISL_662181 to 662184, EPI_ISL_662186 to 662187, EPI_ISL_662191 to 662192, EPI_ISL_662194, EPI_ISL_662196, EPI_ISL_662198, EPI_ISL_662200, EPI_ISL_662202 to 662203, EPI_ISL_662206 to 662209, EPI_ISL_662212, EPI_ISL_662214 to 662215, EPI_ISL_662217 to 662219,
EPI_ISL_662223, EPI_ISL_662227 to 662229, EPI_ISL_662231, EPI_ISL_662233 to 662240, EPI_ISL_662244 to 662245, EPI_ISL_662247 to 662249, EPI_ISL_662252, EPI_ISL_662254 to 662255, EPI_ISL_662258, EPI_ISL_662260, EPI_ISL_662263, EPI_ISL_662265 to 662267, EPI_ISL_662269 to 662270, EPI_ISL_662272 to
662276, EPI_ISL_662279, EPI_ISL_662281, EPI_ISL_662285 to 662286, EPI_ISL_662290, EPI_ISL_662292, EPI_ISL_662296, EPI_ISL_662305, EPI_ISL_662308 to 662309, EPI_ISL_662312, EPI_ISL_662320, EPI_ISL_662327, EPI_ISL_662334, EPI_ISL_662342 to 662344, EPI_ISL_662352, EPI_ISL_662354, EPI_ISL_662356,
EPI_ISL_662358 to 662359, EPI_ISL_662365, EPI_ISL_662369, EPI_ISL_662373 to 662375, EPI_ISL_662381 to 662382, EPI_ISL_662385, EPI_ISL_662389 to 662391, EPI_ISL_662393 to 662394, EPI_ISL_662403, EPI_ISL_662405, EPI_ISL_662410 to 662411, EPI_ISL_662415, EPI_ISL_662418 to 662419, EPI_ISL_662427,
EPI_ISL_662429, EPI_ISL_662431, EPI_ISL_662433, EPI_ISL_662444, EPI_ISL_662446, EPI_ISL_662452, EPI_ISL_662457, EPI_ISL_662462 to 662463, EPI_ISL_662468, EPI_ISL_662472, EPI_ISL_662474, EPI_ISL_662477 to 662478, EPI_ISL_662480, EPI_ISL_662486 to 662489, EPI_ISL_662492, EPI_ISL_662496,
EPI_ISL_662499, EPI_ISL_662502, EPI_ISL_662507 to 662508, EPI_ISL_662514, EPI_ISL_662518, EPI_ISL_662520, EPI_ISL_662526, EPI_ISL_662535, EPI_ISL_662549, EPI_ISL_662553, EPI_ISL_662558, EPI_ISL_662564, EPI_ISL_662576, EPI_ISL_662578 to 662581, EPI_ISL_662583, EPI_ISL_662588, EPI_ISL_662593,
EPI_ISL_662597, EPI_ISL_662604 to 662605, EPI_ISL_662609 to 662610, EPI_ISL_662614, EPI_ISL_662616 to 662617, EPI_ISL_662621 to 662626, EPI_ISL_662628 to 662630, EPI_ISL_662647 to 662650, EPI_ISL_674159 to 674162, EPI_ISL_674164 to 674165, EPI_ISL_674168 to 674172, EPI_ISL_674174 to 674177,
EPI_ISL_674179, EPI_ISL_674181 to 674182, EPI_ISL_674186, EPI_ISL_674188, EPI_ISL_674190, EPI_ISL_674192 to 674195, EPI_ISL_674197, EPI_ISL_674199, EPI_ISL_674203 to 674204, EPI_ISL_674206, EPI_ISL_674208, EPI_ISL_674211, EPI_ISL_674217, EPI_ISL_674222 to 674224, EPI_ISL_674226,
EPI_ISL_674228 to 674229, EPI_ISL_674231, EPI_ISL_674233 to 674234, EPI_ISL_674240 to 674242, EPI_ISL_674245, EPI_ISL_674249, EPI_ISL_674253 to 674255, EPI_ISL_674260 to 674261, EPI_ISL_674264, EPI_ISL_674267, EPI_ISL_674272, EPI_ISL_674276 to 674277, EPI_ISL_674281, EPI_ISL_674283 to 674287,
EPI_ISL_674292, EPI_ISL_674294 to 674297, EPI_ISL_674300 to 674301, EPI_ISL_674308, EPI_ISL_674310 to 674311, EPI_ISL_674313, EPI_ISL_674317 to 674319, EPI_ISL_674322 to 674323, EPI_ISL_674328 to 674329, EPI_ISL_674332, EPI_ISL_674337, EPI_ISL_674339 to 674341, EPI_ISL_674343, EPI_ISL_674345 to
674347, EPI_ISL_674352, EPI_ISL_674354, EPI_ISL_674360 to 674362, EPI_ISL_674364, EPI_ISL_674366, EPI_ISL_674369 to 674370, EPI_ISL_674372, EPI_ISL_674374 to 674376, EPI_ISL_674380, EPI_ISL_674383, EPI_ISL_674385, EPI_ISL_674389 to 674392, EPI_ISL_674396 to 674397, EPI_ISL_674400,
EPI_ISL_674402, EPI_ISL_674404, EPI_ISL_674406, EPI_ISL_674408 to 674409, EPI_ISL_674411 to 674413, EPI_ISL_674416, EPI_ISL_674418, EPI_ISL_674420 to 674421, EPI_ISL_674428 to 674429, EPI_ISL_674431 to 674432, EPI_ISL_674434 to 674436, EPI_ISL_674438 to 674439, EPI_ISL_674445 to 674447,
EPI_ISL_674451 to 674452, EPI_ISL_674454, EPI_ISL_674456, EPI_ISL_674459, EPI_ISL_674462, EPI_ISL_674465, EPI_ISL_674467, EPI_ISL_674469 to 674472, EPI_ISL_674474, EPI_ISL_674476 to 674478, EPI_ISL_674484 to 674488, EPI_ISL_674490, EPI_ISL_674495, EPI_ISL_675130, EPI_ISL_675133,
EPI_ISL_675135 to 675137, EPI_ISL_675139 to 675144, EPI_ISL_675146, EPI_ISL_675148 to 675152, EPI_ISL_675154 to 675155, EPI_ISL_675158 to 675159, EPI_ISL_675161, EPI_ISL_675163 to 675164, EPI_ISL_675166 to 675169, EPI_ISL_675171 to 675177, EPI_ISL_675181 to 675182, EPI_ISL_675184, EPI_ISL_675186
to 675187, EPI_ISL_675189, EPI_ISL_675192, EPI_ISL_675194 to 675198, EPI_ISL_675200 to 675206, EPI_ISL_675209, EPI_ISL_675211, EPI_ISL_675214 to 675216, EPI_ISL_675218, EPI_ISL_675220 to 675223, EPI_ISL_675225, EPI_ISL_675230 to 675235, EPI_ISL_675237, EPI_ISL_675239 to 675240, EPI_ISL_675246 to
675256, EPI_ISL_675258 to 675259, EPI_ISL_675261 to 675263, EPI_ISL_675265 to 675266, EPI_ISL_675268 to 675274, EPI_ISL_675278 to 675281, EPI_ISL_675283, EPI_ISL_675287 to 675288, EPI_ISL_675291, EPI_ISL_675294 to 675300, EPI_ISL_675302, EPI_ISL_675304, EPI_ISL_675310, EPI_ISL_675312,
EPI_ISL_675315 to 675316, EPI_ISL_675321 to 675325, EPI_ISL_675327 to 675328, EPI_ISL_675330, EPI_ISL_675333 to 675339, EPI_ISL_675341 to 675352, EPI_ISL_675354 to 675356, EPI_ISL_675358 to 675363, EPI_ISL_675365 to 675367, EPI_ISL_675373 to 675385, EPI_ISL_675387 to 675388, EPI_ISL_675390,
EPI_ISL_675392, EPI_ISL_675394 to 675396, EPI_ISL_675398, EPI_ISL_675400 to 675403, EPI_ISL_675405, EPI_ISL_675407 to 675408, EPI_ISL_675410 to 675415, EPI_ISL_675421 to 675422, EPI_ISL_675425 to 675430, EPI_ISL_675433 to 675435, EPI_ISL_675437 to 675438, EPI_ISL_675441 to 675442, EPI_ISL_675445,
EPI_ISL_675448 to 675455, EPI_ISL_675457 to 675458, EPI_ISL_675460 to 675461, EPI_ISL_675463 to 675466, EPI_ISL_675468 to 675471, EPI_ISL_675473 to 675474, EPI_ISL_675476 to 675479, EPI_ISL_675481 to 675484, EPI_ISL_675487 to 675489, EPI_ISL_675491 to 675494, EPI_ISL_675497 to 675498,
EPI_ISL_675501 to 675502, EPI_ISL_675508 to 675512, EPI_ISL_675514 to 675515, EPI_ISL_675517, EPI_ISL_675519 to 675527, EPI_ISL_675529 to 675532, EPI_ISL_675534, EPI_ISL_675536, EPI_ISL_675539, EPI_ISL_675541, EPI_ISL_675543 to 675546, EPI_ISL_675548 to 675549, EPI_ISL_675551 to 675554,
EPI_ISL_675557, EPI_ISL_675559, EPI_ISL_675561 to 675563, EPI_ISL_675567 to 675568, EPI_ISL_675570 to 675571, EPI_ISL_675573 to 675575, EPI_ISL_675577, EPI_ISL_675579 to 675595, EPI_ISL_675597 to 675600, EPI_ISL_675603 to 675605, EPI_ISL_675607 to 675620, EPI_ISL_675622 to 675626, EPI_ISL_675628
to 675630, EPI_ISL_675632 to 675633, EPI_ISL_675635 to 675643, EPI_ISL_675645 to 675651, EPI_ISL_675653 to 675659, EPI_ISL_675662, EPI_ISL_675664 to 675666, EPI_ISL_675668 to 675675, EPI_ISL_675677 to 675681, EPI_ISL_675683, EPI_ISL_675685 to 675689, EPI_ISL_675691 to 675692, EPI_ISL_675696 to
675697, EPI_ISL_675699 to 675703, EPI_ISL_675705 to 675708, EPI_ISL_675710, EPI_ISL_675715 to 675716, EPI_ISL_675719 to 675721, EPI_ISL_675735, EPI_ISL_675740, EPI_ISL_675746, EPI_ISL_675750 to 675751, EPI_ISL_675756 to 675757, EPI_ISL_675759 to 675760, EPI_ISL_675766, EPI_ISL_675771,
EPI_ISL_675774 to 675775, EPI_ISL_675777, EPI_ISL_675779, EPI_ISL_675785, EPI_ISL_675788, EPI_ISL_675794, EPI_ISL_675798, EPI_ISL_675800, EPI_ISL_675802 to 675803, EPI_ISL_675806, EPI_ISL_675810, EPI_ISL_675812 to 675813, EPI_ISL_675817 to 675818, EPI_ISL_675820, EPI_ISL_675822 to 675823,
EPI_ISL_675825 to 675826, EPI_ISL_675829, EPI_ISL_675833, EPI_ISL_675839, EPI_ISL_675847, EPI_ISL_675855, EPI_ISL_675868, EPI_ISL_675882, EPI_ISL_675892, EPI_ISL_675894, EPI_ISL_675897, EPI_ISL_675901 to 675903, EPI_ISL_675908, EPI_ISL_675913, EPI_ISL_675916, EPI_ISL_675920, EPI_ISL_675923 to
675925, EPI_ISL_675929 to 675930, EPI_ISL_675934, EPI_ISL_675937, EPI_ISL_675939, EPI_ISL_675942, EPI_ISL_675944, EPI_ISL_675952, EPI_ISL_675954, EPI_ISL_675962, EPI_ISL_675965, EPI_ISL_675968, EPI_ISL_675974, EPI_ISL_675976, EPI_ISL_675978, EPI_ISL_675980, EPI_ISL_675982, EPI_ISL_675986 to
675988, EPI_ISL_675993, EPI_ISL_676000, EPI_ISL_676002, EPI_ISL_676006, EPI_ISL_676010, EPI_ISL_681661 to 681662, EPI_ISL_681665, EPI_ISL_693332 to 693333, EPI_ISL_701673 to 701685, EPI_ISL_701687, EPI_ISL_701689 to 701696, EPI_ISL_701698 to 701699, EPI_ISL_701701, EPI_ISL_701704 to 701709,
EPI_ISL_701711, EPI_ISL_701713 to 701716, EPI_ISL_701718 to 701719, EPI_ISL_701721 to 701723, EPI_ISL_701725 to 701730, EPI_ISL_701732 to 701750, EPI_ISL_701752 to 701754, EPI_ISL_701756, EPI_ISL_701758, EPI_ISL_701760 to 701762, EPI_ISL_701764, EPI_ISL_701766 to 701770, EPI_ISL_701772 to 701774,
EPI_ISL_701776 to 701783, EPI_ISL_701785, EPI_ISL_701787 to 701791, EPI_ISL_701794 to 701802, EPI_ISL_701804 to 701811, EPI_ISL_701814 to 701815, EPI_ISL_701817 to 701827, EPI_ISL_701829, EPI_ISL_701831 to 701841, EPI_ISL_701843, EPI_ISL_701845 to 701851, EPI_ISL_701853 to 701856, EPI_ISL_701859
to 701862, EPI_ISL_701864 to 701867, EPI_ISL_701869, EPI_ISL_701871 to 701873, EPI_ISL_701875, EPI_ISL_701877 to 701883, EPI_ISL_701885, EPI_ISL_701888 to 701889, EPI_ISL_701891, EPI_ISL_701893 to 701920, EPI_ISL_701922 to 701925, EPI_ISL_701927 to 701934, EPI_ISL_701936 to 701950, EPI_ISL_701952,
EPI_ISL_701954 to 701957, EPI_ISL_701960, EPI_ISL_701963, EPI_ISL_701965 to 701968, EPI_ISL_701970 to 701977, EPI_ISL_701979, EPI_ISL_701981 to 701983, EPI_ISL_701985 to 701986, EPI_ISL_701988, EPI_ISL_701990 to 701991, EPI_ISL_701993 to 702004, EPI_ISL_702006 to 702012, EPI_ISL_702015 to 702018,
EPI_ISL_702020 to 702021, EPI_ISL_702370 to 702380, EPI_ISL_702382 to 702384, EPI_ISL_702387 to 702391, EPI_ISL_702393 to 702402, EPI_ISL_702404 to 702408, EPI_ISL_702410, EPI_ISL_702412 to 702420, EPI_ISL_702422 to 702437, EPI_ISL_702439 to 702445, EPI_ISL_702447 to 702454, EPI_ISL_702456 to
702463, EPI_ISL_702465, EPI_ISL_702468, EPI_ISL_702472, EPI_ISL_702475, EPI_ISL_702479 to 702480, EPI_ISL_702482 to 702483, EPI_ISL_702485 to 702486, EPI_ISL_702488, EPI_ISL_702490 to 702491, EPI_ISL_702494, EPI_ISL_702497, EPI_ISL_702499, EPI_ISL_702501 to 702503, EPI_ISL_702506,
EPI_ISL_702509, EPI_ISL_702511, EPI_ISL_702514, EPI_ISL_702516 to 702517, EPI_ISL_702519, EPI_ISL_702521 to 702522, EPI_ISL_702524, EPI_ISL_702526 to 702527, EPI_ISL_702529 to 702530, EPI_ISL_702532, EPI_ISL_702534 to 702535, EPI_ISL_702537 to 702538, EPI_ISL_702542 to 702543, EPI_ISL_702546,
EPI_ISL_702548, EPI_ISL_702551 to 702552, EPI_ISL_702554 to 702555, EPI_ISL_702557, EPI_ISL_702559, EPI_ISL_702563, EPI_ISL_702565, EPI_ISL_702567 to 702568, EPI_ISL_702570, EPI_ISL_702572 to 702573, EPI_ISL_702575 to 702576, EPI_ISL_702578, EPI_ISL_702580 to 702581, EPI_ISL_702583,
EPI_ISL_702586, EPI_ISL_702589, EPI_ISL_702592 to 702593, EPI_ISL_702595 to 702596, EPI_ISL_702598 to 702599, EPI_ISL_702601, EPI_ISL_702603 to 702604, EPI_ISL_702606, EPI_ISL_702608 to 702609, EPI_ISL_702611, EPI_ISL_702613 to 702614, EPI_ISL_702616 to 702617, EPI_ISL_702619, EPI_ISL_702621 to
702622, EPI_ISL_702624, EPI_ISL_702627, EPI_ISL_702629 to 702630, EPI_ISL_702632, EPI_ISL_702634 to 702635, EPI_ISL_702637, EPI_ISL_702640 to 702641, EPI_ISL_702643, EPI_ISL_702645 to 702646, EPI_ISL_702648, EPI_ISL_702650 to 702651, EPI_ISL_702653 to 702654, EPI_ISL_702658, EPI_ISL_702660,
EPI_ISL_702662, EPI_ISL_702668, EPI_ISL_702670 to 702671, EPI_ISL_702673, EPI_ISL_702675 to 702676, EPI_ISL_702678, EPI_ISL_702680, EPI_ISL_702683, EPI_ISL_702686, EPI_ISL_702688 to 702689, EPI_ISL_702691, EPI_ISL_702693, EPI_ISL_702695 to 702696, EPI_ISL_702701 to 702702, EPI_ISL_702704,
EPI_ISL_702706 to 702707, EPI_ISL_702709, EPI_ISL_702711 to 702712, EPI_ISL_702714, EPI_ISL_702717, EPI_ISL_702719 to 702720, EPI_ISL_702722, EPI_ISL_702724 to 702725, EPI_ISL_702727 to 702728, EPI_ISL_702730, EPI_ISL_702732 to 702733, EPI_ISL_702735, EPI_ISL_702737 to 702738, EPI_ISL_702740 to
702741, EPI_ISL_702743, EPI_ISL_702745, EPI_ISL_702748, EPI_ISL_702750, EPI_ISL_702752, EPI_ISL_702755, EPI_ISL_702757, EPI_ISL_702759 to 702760, EPI_ISL_702762, EPI_ISL_702765, EPI_ISL_702767 to 702768, EPI_ISL_702772 to 702773, EPI_ISL_702775, EPI_ISL_702777 to 702778, EPI_ISL_702781,
EPI_ISL_702783, EPI_ISL_702785 to 702786, EPI_ISL_702788, EPI_ISL_702791, EPI_ISL_702793, EPI_ISL_702796 to 702797, EPI_ISL_702799 to 702800, EPI_ISL_702802, EPI_ISL_702804, EPI_ISL_702807 to 702808, EPI_ISL_702810 to 702811, EPI_ISL_702813, EPI_ISL_702816, EPI_ISL_702818, EPI_ISL_702821 to
702822, EPI_ISL_702898 to 702899, EPI_ISL_702901, EPI_ISL_702906, EPI_ISL_702909, EPI_ISL_702912, EPI_ISL_702914, EPI_ISL_702916, EPI_ISL_702918, EPI_ISL_702921, EPI_ISL_702923 to 702924, EPI_ISL_702927, EPI_ISL_702929, EPI_ISL_702931, EPI_ISL_702933 to 702934, EPI_ISL_702936, EPI_ISL_702938,
EPI_ISL_702940, EPI_ISL_702942 to 702943, EPI_ISL_702945 to 702946, EPI_ISL_702948, EPI_ISL_702951 to 702952, EPI_ISL_702954 to 702955, EPI_ISL_702957 to 702958, EPI_ISL_702960, EPI_ISL_702962 to 702963, EPI_ISL_702965, EPI_ISL_702967 to 702968, EPI_ISL_702970, EPI_ISL_702972 to 702973,
EPI_ISL_702975 to 702976, EPI_ISL_702978 to 702979, EPI_ISL_702981, EPI_ISL_702983 to 702984, EPI_ISL_702986, EPI_ISL_702988 to 702989, EPI_ISL_702991, EPI_ISL_702993, EPI_ISL_702995, EPI_ISL_702997 to 702998, EPI_ISL_703000 to 703001, EPI_ISL_703003 to 703004, EPI_ISL_703006, EPI_ISL_703009,
EPI_ISL_703011 to 703012, EPI_ISL_703014, EPI_ISL_703016 to 703017, EPI_ISL_703019, EPI_ISL_703022, EPI_ISL_703027 to 703028, EPI_ISL_703030 to 703031, EPI_ISL_703033, EPI_ISL_703035, EPI_ISL_703037, EPI_ISL_703039, EPI_ISL_703041 to 703042, EPI_ISL_703044, EPI_ISL_703047, EPI_ISL_703049 to
703050, EPI_ISL_703052, EPI_ISL_703054 to 703055, EPI_ISL_703058, EPI_ISL_703063, EPI_ISL_703065, EPI_ISL_703068, EPI_ISL_703070, EPI_ISL_703075, EPI_ISL_703078 to 703079, EPI_ISL_703081, EPI_ISL_703083 to 703084, EPI_ISL_703086 to 703087, EPI_ISL_703089, EPI_ISL_703091 to 703092,
EPI_ISL_703094 to 703095, EPI_ISL_703097 to 703098, EPI_ISL_703100 to 703101, EPI_ISL_703103, EPI_ISL_703106, EPI_ISL_703108 to 703109, EPI_ISL_703111 to 703112, EPI_ISL_703116 to 703117, EPI_ISL_703119 to 703120, EPI_ISL_703122 to 703123, EPI_ISL_703125 to 703126, EPI_ISL_703128, EPI_ISL_703130
to 703131, EPI_ISL_703134 to 703135, EPI_ISL_703137, EPI_ISL_703140, EPI_ISL_703142 to 703143, EPI_ISL_703145 to 703146, EPI_ISL_703149, EPI_ISL_703151, EPI_ISL_703153 to 703154, EPI_ISL_703156 to 703157, EPI_ISL_703159 to 703161, EPI_ISL_703163 to 703164, EPI_ISL_703166, EPI_ISL_703168 to 703169,
EPI_ISL_703171, EPI_ISL_703173, EPI_ISL_703175, EPI_ISL_703178, EPI_ISL_703180 to 703181, EPI_ISL_703183, EPI_ISL_703185, EPI_ISL_703188 to 703189, EPI_ISL_703191, EPI_ISL_703193 to 703194, EPI_ISL_703196 to 703197, EPI_ISL_703199, EPI_ISL_703201 to 703202, EPI_ISL_703204 to 703205,
EPI_ISL_703207, EPI_ISL_703209 to 703210, EPI_ISL_703212, EPI_ISL_703214 to 703215, EPI_ISL_703217 to 703218, EPI_ISL_703220 to 703221, EPI_ISL_703223 to 703224, EPI_ISL_703226, EPI_ISL_703228 to 703229, EPI_ISL_703231 to 703232, EPI_ISL_703234, EPI_ISL_703236 to 703237, EPI_ISL_703239,
EPI_ISL_703241 to 703242, EPI_ISL_703244 to 703245, EPI_ISL_703247 to 703248, EPI_ISL_703250, EPI_ISL_703252 to 703253, EPI_ISL_703255, EPI_ISL_703257 to 703258, EPI_ISL_703260, EPI_ISL_703265 to 703266, EPI_ISL_703268, EPI_ISL_703271, EPI_ISL_703274, EPI_ISL_703277, EPI_ISL_703282,
EPI_ISL_703293 to 703294, EPI_ISL_703298, EPI_ISL_703301, EPI_ISL_703306 to 703307, EPI_ISL_703311 to 703312, EPI_ISL_703314, EPI_ISL_703317 to 703318, EPI_ISL_703322, EPI_ISL_703326, EPI_ISL_703328 to 703329, EPI_ISL_703333, EPI_ISL_703336, EPI_ISL_703339, EPI_ISL_703341 to 703342,
EPI_ISL_703344, EPI_ISL_703347, EPI_ISL_703365, EPI_ISL_703368, EPI_ISL_703373 to 703374, EPI_ISL_703376, EPI_ISL_703386, EPI_ISL_703388, EPI_ISL_703394, EPI_ISL_703396 to 703397, EPI_ISL_703402, EPI_ISL_703404 to 703405, EPI_ISL_703413, EPI_ISL_703416, EPI_ISL_703418, EPI_ISL_703420,
EPI_ISL_703425, EPI_ISL_703433 to 703434, EPI_ISL_703436 to 703437, EPI_ISL_703441, EPI_ISL_703448, EPI_ISL_703450, EPI_ISL_703452, EPI_ISL_703455, EPI_ISL_703460 to 703461, EPI_ISL_703464, EPI_ISL_703467, EPI_ISL_703470, EPI_ISL_703475, EPI_ISL_703477 to 703478, EPI_ISL_703483,
EPI_ISL_703488, EPI_ISL_703490, EPI_ISL_703493 to 703494, EPI_ISL_703498, EPI_ISL_703503 to 703504, EPI_ISL_703507, EPI_ISL_703509, EPI_ISL_703518, EPI_ISL_703520, EPI_ISL_703523, EPI_ISL_703526, EPI_ISL_703528, EPI_ISL_703531 to 703532, EPI_ISL_703536, EPI_ISL_703539, EPI_ISL_703547,
EPI_ISL_703549, EPI_ISL_703552, EPI_ISL_703558, EPI_ISL_703563, EPI_ISL_703565 to 703566, EPI_ISL_703568, EPI_ISL_703570 to 703571, EPI_ISL_703573 to 703574, EPI_ISL_703576, EPI_ISL_703580 to 703581, EPI_ISL_703586, EPI_ISL_703588 to 703589, EPI_ISL_703591 to 703592, EPI_ISL_703594 to 703595,
EPI_ISL_703602 to 703603, EPI_ISL_703605, EPI_ISL_703613, EPI_ISL_703615, EPI_ISL_703618, EPI_ISL_703620, EPI_ISL_703623, EPI_ISL_703625, EPI_ISL_703628, EPI_ISL_703630, EPI_ISL_703637, EPI_ISL_703646, EPI_ISL_703648, EPI_ISL_703662, EPI_ISL_703664 to 703665, EPI_ISL_703667, EPI_ISL_703670,
EPI_ISL_703675, EPI_ISL_703678, EPI_ISL_703680 to 703681, EPI_ISL_703683, EPI_ISL_703686, EPI_ISL_703688 to 703689, EPI_ISL_703691 to 703692, EPI_ISL_703696, EPI_ISL_703699, EPI_ISL_703702, EPI_ISL_703705, EPI_ISL_703707, EPI_ISL_703777 to 703778, EPI_ISL_703780 to 703781, EPI_ISL_703783,
EPI_ISL_703785, EPI_ISL_703788, EPI_ISL_703791, EPI_ISL_703793, EPI_ISL_703795 to 703796, EPI_ISL_703798, EPI_ISL_703803 to 703804, EPI_ISL_703809, EPI_ISL_703812, EPI_ISL_703814, EPI_ISL_703817, EPI_ISL_708841 to 708842, EPI_ISL_708845 to 708846, EPI_ISL_708851 to 708854, EPI_ISL_708859,
EPI_ISL_708861, EPI_ISL_708863 to 708868, EPI_ISL_708870, EPI_ISL_708872 to 708873, EPI_ISL_708875 to 708878, EPI_ISL_708880, EPI_ISL_708886 to 708888, EPI_ISL_708891 to 708892, EPI_ISL_708894 to 708895, EPI_ISL_708902 to 708903, EPI_ISL_708907, EPI_ISL_708911, EPI_ISL_708914 to 708915,
EPI_ISL_708919, EPI_ISL_708923 to 708924, EPI_ISL_708927, EPI_ISL_708929, EPI_ISL_708931, EPI_ISL_708934 to 708937, EPI_ISL_708939 to 708940, EPI_ISL_708942 to 708943, EPI_ISL_708947, EPI_ISL_708949 to 708950, EPI_ISL_708954 to 708956, EPI_ISL_708959 to 708960, EPI_ISL_708962, EPI_ISL_708965,
EPI_ISL_708967 to 708968, EPI_ISL_708970 to 708974, EPI_ISL_708977 to 708978, EPI_ISL_708980 to 708982, EPI_ISL_708987 to 708988, EPI_ISL_709295 to 709296, EPI_ISL_709298 to 709300, EPI_ISL_709302 to 709306, EPI_ISL_709308 to 709310, EPI_ISL_709313, EPI_ISL_709315 to 709319, EPI_ISL_709321 to
709322, EPI_ISL_709325 to 709326, EPI_ISL_709328 to 709331, EPI_ISL_709333, EPI_ISL_709335, EPI_ISL_709337 to 709340, EPI_ISL_709342, EPI_ISL_709344, EPI_ISL_709346 to 709352, EPI_ISL_709355 to 709357, EPI_ISL_709361 to 709363, EPI_ISL_709368 to 709372, EPI_ISL_709374, EPI_ISL_709377,
EPI_ISL_709379, EPI_ISL_709381 to 709382, EPI_ISL_709386 to 709387, EPI_ISL_709389 to 709391, EPI_ISL_709393, EPI_ISL_709397, EPI_ISL_709400 to 709402, EPI_ISL_709405, EPI_ISL_709408, EPI_ISL_709410, EPI_ISL_709413 to 709416, EPI_ISL_709418, EPI_ISL_709420, EPI_ISL_709425, EPI_ISL_709428,
EPI_ISL_709430 to 709435, EPI_ISL_709437, EPI_ISL_709442, EPI_ISL_709444 to 709445, EPI_ISL_709447 to 709449, EPI_ISL_709451 to 709452, EPI_ISL_709455, EPI_ISL_709460 to 709461, EPI_ISL_709463, EPI_ISL_709467 to 709468, EPI_ISL_709470 to 709471, EPI_ISL_709473, EPI_ISL_709475 to 709478,
EPI_ISL_709480, EPI_ISL_709484 to 709485, EPI_ISL_709489, EPI_ISL_709491 to 709492, EPI_ISL_709499, EPI_ISL_709504 to 709508, EPI_ISL_709512 to 709514, EPI_ISL_709516, EPI_ISL_709519 to 709521, EPI_ISL_709523, EPI_ISL_709526, EPI_ISL_709529, EPI_ISL_709531 to 709532, EPI_ISL_709534 to 709536,
EPI_ISL_709538, EPI_ISL_709543 to 709546, EPI_ISL_709550 to 709552, EPI_ISL_709554, EPI_ISL_709559 to 709560, EPI_ISL_709563 to 709564, EPI_ISL_709566, EPI_ISL_710614, EPI_ISL_710616 to 710617, EPI_ISL_710620, EPI_ISL_710625, EPI_ISL_710628, EPI_ISL_710631, EPI_ISL_710638, EPI_ISL_710645,
EPI_ISL_710656, EPI_ISL_710661 to 710663, EPI_ISL_710665, EPI_ISL_710671, EPI_ISL_710676, EPI_ISL_710689, EPI_ISL_710693 to 710694, EPI_ISL_710698 to 710709, EPI_ISL_710711, EPI_ISL_710714 to 710747, EPI_ISL_710749 to 710752, EPI_ISL_710837, EPI_ISL_710865 to 710866, EPI_ISL_710869 to 710871,
EPI_ISL_710873 to 710876, EPI_ISL_710887 to 710902, EPI_ISL_710908 to 710940, EPI_ISL_710942 to 710980, EPI_ISL_710982 to 710994, EPI_ISL_711008 to 711013, EPI_ISL_711015 to 711022, EPI_ISL_711027, EPI_ISL_711029 to 711037, EPI_ISL_711039 to 711047, EPI_ISL_718629, EPI_ISL_718631, EPI_ISL_718633,
EPI_ISL_718635, EPI_ISL_718637, EPI_ISL_718639 to 718640, EPI_ISL_718643 to 718644, EPI_ISL_718647, EPI_ISL_718653, EPI_ISL_718657 to 718658, EPI_ISL_718662 to 718666, EPI_ISL_718670, EPI_ISL_718672, EPI_ISL_718674 to 718676, EPI_ISL_718678, EPI_ISL_718682, EPI_ISL_718686 to 718687,
EPI_ISL_718691, EPI_ISL_718694 to 718695, EPI_ISL_718700, EPI_ISL_718705, EPI_ISL_718707, EPI_ISL_718710 to 718716, EPI_ISL_718718 to 718719, EPI_ISL_718721, EPI_ISL_718723 to 718725, EPI_ISL_718728 to 718729, EPI_ISL_718735, EPI_ISL_718738 to 718740, EPI_ISL_718742, EPI_ISL_718744 to 718747,
EPI_ISL_718750, EPI_ISL_718754 to 718755, EPI_ISL_718759, EPI_ISL_718761 to 718762, EPI_ISL_718764 to 718767, EPI_ISL_718769, EPI_ISL_718772 to 718774, EPI_ISL_718777, EPI_ISL_718779 to 718783, EPI_ISL_718787, EPI_ISL_718789, EPI_ISL_718791, EPI_ISL_718795 to 718796, EPI_ISL_718798 to 718799,
EPI_ISL_718801 to 718803, EPI_ISL_718806 to 718808, EPI_ISL_718811 to 718813, EPI_ISL_718819, EPI_ISL_718821, EPI_ISL_718823, EPI_ISL_718827 to 718833, EPI_ISL_718837, EPI_ISL_718839, EPI_ISL_718841, EPI_ISL_718844, EPI_ISL_718847, EPI_ISL_718849 to 718850, EPI_ISL_718853 to 718854,
EPI_ISL_718858 to 718859, EPI_ISL_718864 to 718866, EPI_ISL_718870 to 718873, EPI_ISL_718877 to 718878, EPI_ISL_718880 to 718881, EPI_ISL_718885 to 718891, EPI_ISL_718894 to 718895, EPI_ISL_718901 to 718903, EPI_ISL_718907 to 718908, EPI_ISL_718911, EPI_ISL_718914 to 718915, EPI_ISL_718917,
EPI_ISL_718919, EPI_ISL_718922 to 718923, EPI_ISL_718926, EPI_ISL_718929 to 718930, EPI_ISL_718932, EPI_ISL_718935 to 718936, EPI_ISL_718939, EPI_ISL_718943, EPI_ISL_718945, EPI_ISL_718950, EPI_ISL_718952 to 718953, EPI_ISL_718955, EPI_ISL_718957, EPI_ISL_718959, EPI_ISL_718962 to 718964,
EPI_ISL_718966 to 718967, EPI_ISL_718970, EPI_ISL_718974 to 718979, EPI_ISL_720683, EPI_ISL_720685 to 720686, EPI_ISL_720688, EPI_ISL_720694, EPI_ISL_720697, EPI_ISL_720709, EPI_ISL_720711, EPI_ISL_720715, EPI_ISL_720722, EPI_ISL_720741, EPI_ISL_720756, EPI_ISL_720764 to 720765,
EPI_ISL_720770, EPI_ISL_720773, EPI_ISL_720779 to 720780, EPI_ISL_720796, EPI_ISL_720810, EPI_ISL_720814, EPI_ISL_720823, EPI_ISL_720825, EPI_ISL_720827, EPI_ISL_720836, EPI_ISL_720842, EPI_ISL_720845 to 720846, EPI_ISL_720848 to 720849, EPI_ISL_720853, EPI_ISL_720858, EPI_ISL_720864,
EPI_ISL_720869, EPI_ISL_720872, EPI_ISL_720875, EPI_ISL_720880, EPI_ISL_720885 to 720886, EPI_ISL_720893 to 720894, EPI_ISL_720898, EPI_ISL_720907, EPI_ISL_721185, EPI_ISL_721187, EPI_ISL_721189 to 721192, EPI_ISL_721194 to 721207, EPI_ISL_721209 to 721227, EPI_ISL_721229 to 721231,
EPI_ISL_721233, EPI_ISL_721235 to 721262, EPI_ISL_721264 to 721303, EPI_ISL_721305 to 721329, EPI_ISL_721331 to 721334, EPI_ISL_721336 to 721345, EPI_ISL_721348 to 721352, EPI_ISL_721355 to 721361, EPI_ISL_721364 to 721371, EPI_ISL_721373 to 721388, EPI_ISL_721390 to 721394, EPI_ISL_721396 to
721421, EPI_ISL_721423, EPI_ISL_721425 to 721431, EPI_ISL_721433 to 721441, EPI_ISL_721443 to 721452, EPI_ISL_721454, EPI_ISL_721458 to 721465, EPI_ISL_721467 to 721472, EPI_ISL_721475, EPI_ISL_721477 to 721483, EPI_ISL_721485 to 721486, EPI_ISL_721488 to 721510, EPI_ISL_721512 to 721517,
EPI_ISL_721520, EPI_ISL_721523, EPI_ISL_721525, EPI_ISL_730654 to 730655, EPI_ISL_730658, EPI_ISL_730660 to 730663, EPI_ISL_730665 to 730672, EPI_ISL_730674, EPI_ISL_730677, EPI_ISL_730679 to 730684, EPI_ISL_730686 to 730696, EPI_ISL_730698 to 730704, EPI_ISL_730706 to 730708, EPI_ISL_730711 to
730717, EPI_ISL_730719 to 730722, EPI_ISL_730724 to 730731, EPI_ISL_730733 to 730736, EPI_ISL_730738 to 730751, EPI_ISL_730753 to 730758, EPI_ISL_730760 to 730761, EPI_ISL_730763 to 730764, EPI_ISL_730766 to 730774, EPI_ISL_730776, EPI_ISL_730779, EPI_ISL_730781 to 730784, EPI_ISL_730786 to
730788, EPI_ISL_730790 to 730792, EPI_ISL_730794, EPI_ISL_730797 to 730800, EPI_ISL_730807, EPI_ISL_730810 to 730812, EPI_ISL_730814, EPI_ISL_730816 to 730823, EPI_ISL_730825 to 730827, EPI_ISL_730829 to 730830, EPI_ISL_730834 to 730839, EPI_ISL_730841 to 730845, EPI_ISL_730848, EPI_ISL_730851 to
730857, EPI_ISL_730859, EPI_ISL_730861, EPI_ISL_730863, EPI_ISL_730865 to 730866, EPI_ISL_730868, EPI_ISL_730870 to 730874, EPI_ISL_730878 to 730886, EPI_ISL_730888, EPI_ISL_730890 to 730893, EPI_ISL_730895 to 730897, EPI_ISL_730899 to 730907, EPI_ISL_730909, EPI_ISL_730912, EPI_ISL_730914 to
730917, EPI_ISL_730919 to 730920, EPI_ISL_730922 to 730925, EPI_ISL_730927 to 730931, EPI_ISL_730933 to 730938, EPI_ISL_730940 to 730941, EPI_ISL_730943 to 730947, EPI_ISL_730950 to 730954, EPI_ISL_730957 to 730964, EPI_ISL_730966, EPI_ISL_730968 to 730977, EPI_ISL_730980 to 730992, EPI_ISL_730994
to 731006, EPI_ISL_731008 to 731020, EPI_ISL_731022 to 731036, EPI_ISL_731038 to 731046, EPI_ISL_731048 to 731051, EPI_ISL_731053 to 731090, EPI_ISL_731092 to 731106, EPI_ISL_731108 to 731113, EPI_ISL_731115 to 731133, EPI_ISL_731135, EPI_ISL_731137 to 731150, EPI_ISL_731152 to 731157,
EPI_ISL_731159 to 731161, EPI_ISL_731163 to 731170, EPI_ISL_731172 to 731178, EPI_ISL_731181 to 731186, EPI_ISL_731188 to 731192, EPI_ISL_731196 to 731198, EPI_ISL_731200 to 731209, EPI_ISL_731211 to 731224, EPI_ISL_731228 to 731243, EPI_ISL_731245, EPI_ISL_731247 to 731250, EPI_ISL_731254,

EPI_ISL_731260, EPI_ISL_731263, EPI_ISL_731265, EPI_ISL_731268 to 731270, EPI_ISL_731272, EPI_ISL_731274 to 731279, EPI_ISL_731285, EPI_ISL_731288 to 731289, EPI_ISL_731292, EPI_ISL_731294, EPI_ISL_731298, EPI_ISL_731300 to 731304, EPI_ISL_731307, EPI_ISL_731311 to 731313, EPI_ISL_731315 to
731318, EPI_ISL_731323 to 731325, EPI_ISL_731329, EPI_ISL_731331, EPI_ISL_731336, EPI_ISL_731339, EPI_ISL_731342 to 731344, EPI_ISL_731350 to 731352, EPI_ISL_731356, EPI_ISL_731358 to 731360, EPI_ISL_731362 to 731366, EPI_ISL_731369 to 731371, EPI_ISL_731374 to 731375, EPI_ISL_731377,
EPI_ISL_731381, EPI_ISL_731385, EPI_ISL_731387 to 731389, EPI_ISL_731391, EPI_ISL_731395, EPI_ISL_731399, EPI_ISL_731409 to 731410, EPI_ISL_731412 to 731413, EPI_ISL_731415 to 731416, EPI_ISL_731418 to 731419, EPI_ISL_731422, EPI_ISL_731424, EPI_ISL_731427 to 731428, EPI_ISL_731430,
EPI_ISL_731434, EPI_ISL_731436, EPI_ISL_731439 to 731443, EPI_ISL_731445 to 731446, EPI_ISL_731448, EPI_ISL_731451, EPI_ISL_731455, EPI_ISL_731457, EPI_ISL_731460 to 731465, EPI_ISL_731468, EPI_ISL_731470, EPI_ISL_731473 to 731475, EPI_ISL_731480 to 731481, EPI_ISL_731483, EPI_ISL_731487,
EPI_ISL_731489, EPI_ISL_731492, EPI_ISL_731495, EPI_ISL_731498, EPI_ISL_731500, EPI_ISL_731502 to 731504, EPI_ISL_731506, EPI_ISL_731509 to 731512, EPI_ISL_731514 to 731516, EPI_ISL_731518, EPI_ISL_731520 to 731521, EPI_ISL_731524 to 731528, EPI_ISL_731530 to 731532, EPI_ISL_731534,
EPI_ISL_731538 to 731540, EPI_ISL_731542 to 731543, EPI_ISL_731545 to 731547, EPI_ISL_731549, EPI_ISL_731552 to 731554, EPI_ISL_731556, EPI_ISL_731558 to 731562, EPI_ISL_731564 to 731565, EPI_ISL_731567 to 731568, EPI_ISL_731570, EPI_ISL_731572 to 731589, EPI_ISL_731592 to 731593, EPI_ISL_731595,
EPI_ISL_731597 to 731601, EPI_ISL_731603 to 731611, EPI_ISL_731613 to 731620, EPI_ISL_731622 to 731625, EPI_ISL_731627 to 731649, EPI_ISL_731651 to 731656, EPI_ISL_731658 to 731665, EPI_ISL_731667 to 731673, EPI_ISL_731675, EPI_ISL_731677 to 731689, EPI_ISL_731691, EPI_ISL_731693 to 731709,
EPI_ISL_731711 to 731713, EPI_ISL_731716 to 731722, EPI_ISL_731724, EPI_ISL_731726 to 731745, EPI_ISL_731747, EPI_ISL_731749 to 731772, EPI_ISL_731774 to 731784, EPI_ISL_731786 to 731790, EPI_ISL_731792 to 731806, EPI_ISL_731808 to 731810, EPI_ISL_731812 to 731814, EPI_ISL_731816 to 731821,
EPI_ISL_731823 to 731825, EPI_ISL_731827 to 731846, EPI_ISL_731849 to 731878, EPI_ISL_731880 to 731884, EPI_ISL_731886 to 731890, EPI_ISL_733598, EPI_ISL_733603, EPI_ISL_733609, EPI_ISL_733611, EPI_ISL_733613, EPI_ISL_733618, EPI_ISL_733621, EPI_ISL_733623, EPI_ISL_733635, EPI_ISL_733638,
EPI_ISL_733641 to 733643, EPI_ISL_733647, EPI_ISL_733659, EPI_ISL_733682 to 733683, EPI_ISL_733693, EPI_ISL_733697, EPI_ISL_733699, EPI_ISL_733701, EPI_ISL_733703, EPI_ISL_733710, EPI_ISL_733712, EPI_ISL_733725, EPI_ISL_733731, EPI_ISL_733734, EPI_ISL_733737, EPI_ISL_733742, EPI_ISL_733754 to
733755, EPI_ISL_733759, EPI_ISL_733766, EPI_ISL_733769, EPI_ISL_733777, EPI_ISL_733783, EPI_ISL_733785, EPI_ISL_733794 to 733795, EPI_ISL_733797, EPI_ISL_733803, EPI_ISL_733806 to 733808, EPI_ISL_733815, EPI_ISL_733819, EPI_ISL_733830, EPI_ISL_733833, EPI_ISL_733837, EPI_ISL_733844,
EPI_ISL_733848, EPI_ISL_733877, EPI_ISL_733879 to 733880, EPI_ISL_733882, EPI_ISL_733885, EPI_ISL_733897, EPI_ISL_733901, EPI_ISL_733912 to 733913, EPI_ISL_735860, EPI_ISL_735862 to 735863, EPI_ISL_735865 to 735877, EPI_ISL_735881 to 735885, EPI_ISL_735887 to 735893, EPI_ISL_735895 to 735908,
EPI_ISL_735910 to 735919, EPI_ISL_735921 to 735926, EPI_ISL_735928 to 735936, EPI_ISL_735938, EPI_ISL_735940 to 735944, EPI_ISL_735946 to 735948, EPI_ISL_735950 to 735951, EPI_ISL_735953 to 735955, EPI_ISL_735957 to 735960, EPI_ISL_735962 to 735964, EPI_ISL_735966 to 735967, EPI_ISL_735969 to
735971, EPI_ISL_735974 to 735977, EPI_ISL_735979 to 735981, EPI_ISL_735983 to 735986, EPI_ISL_735988 to 735992, EPI_ISL_735994, EPI_ISL_735997 to 736000, EPI_ISL_736002, EPI_ISL_736004 to 736006, EPI_ISL_736008 to 736014, EPI_ISL_736016, EPI_ISL_736020 to 736023, EPI_ISL_736026, EPI_ISL_736028 to
736032, EPI_ISL_736034 to 736037, EPI_ISL_736039 to 736040, EPI_ISL_736042 to 736051, EPI_ISL_736053 to 736056, EPI_ISL_736058, EPI_ISL_736060 to 736063, EPI_ISL_736065, EPI_ISL_736067 to 736071, EPI_ISL_736073 to 736089, EPI_ISL_736093 to 736095, EPI_ISL_736097 to 736098, EPI_ISL_736103,
EPI_ISL_736105 to 736106, EPI_ISL_736108 to 736114, EPI_ISL_736116 to 736119, EPI_ISL_736121 to 736123, EPI_ISL_736125 to 736129, EPI_ISL_736133 to 736134, EPI_ISL_736136 to 736142, EPI_ISL_736144 to 736145, EPI_ISL_736147 to 736148, EPI_ISL_736150, EPI_ISL_736152 to 736158, EPI_ISL_736160 to
736161, EPI_ISL_736163 to 736166, EPI_ISL_736170 to 736174, EPI_ISL_736176 to 736181, EPI_ISL_736185 to 736189, EPI_ISL_736192 to 736200, EPI_ISL_736481 to 736482, EPI_ISL_736484 to 736486, EPI_ISL_736493 to 736496, EPI_ISL_736498 to 736500, EPI_ISL_736505 to 736507, EPI_ISL_736509 to 736511,
EPI_ISL_736513, EPI_ISL_736516 to 736518, EPI_ISL_736520 to 736522, EPI_ISL_736526, EPI_ISL_736530 to 736531, EPI_ISL_736534 to 736535, EPI_ISL_736537, EPI_ISL_736539 to 736541, EPI_ISL_736543 to 736546, EPI_ISL_736548, EPI_ISL_736553 to 736555, EPI_ISL_736557, EPI_ISL_736559 to 736561,
EPI_ISL_736564, EPI_ISL_736568 to 736570, EPI_ISL_736572, EPI_ISL_736574 to 736577, EPI_ISL_736579, EPI_ISL_736581, EPI_ISL_736583 to 736585, EPI_ISL_736587, EPI_ISL_736589, EPI_ISL_736591, EPI_ISL_736595 to 736596, EPI_ISL_736599 to 736600, EPI_ISL_736603 to 736604, EPI_ISL_736606 to 736607,
EPI_ISL_736611 to 736616, EPI_ISL_736619, EPI_ISL_736621, EPI_ISL_736623, EPI_ISL_736625 to 736626, EPI_ISL_736629, EPI_ISL_736632, EPI_ISL_736634 to 736636, EPI_ISL_736638 to 736642, EPI_ISL_736644 to 736645, EPI_ISL_736647, EPI_ISL_736649, EPI_ISL_736652, EPI_ISL_736654, EPI_ISL_736656,
EPI_ISL_736660 to 736667, EPI_ISL_736669 to 736671, EPI_ISL_736673 to 736674, EPI_ISL_736676, EPI_ISL_736679 to 736680, EPI_ISL_736684, EPI_ISL_736687 to 736688, EPI_ISL_736690, EPI_ISL_736692, EPI_ISL_736698, EPI_ISL_736701, EPI_ISL_736704 to 736705, EPI_ISL_736707 to 736711, EPI_ISL_736714 to
736715, EPI_ISL_736717 to 736718, EPI_ISL_736720, EPI_ISL_736722, EPI_ISL_736726, EPI_ISL_736729 to 736730, EPI_ISL_736732, EPI_ISL_736739, EPI_ISL_736741 to 736742, EPI_ISL_736748, EPI_ISL_736750 to 736751, EPI_ISL_736753, EPI_ISL_736755 to 736756, EPI_ISL_736758, EPI_ISL_736760 to 736768,
EPI_ISL_736770 to 736772, EPI_ISL_756378, EPI_ISL_756381 to 756383, EPI_ISL_756387 to 756388, EPI_ISL_756390 to 756391, EPI_ISL_756394, EPI_ISL_756396 to 756400, EPI_ISL_756402, EPI_ISL_756404 to 756408, EPI_ISL_756411 to 756412, EPI_ISL_756414, EPI_ISL_756421, EPI_ISL_756423, EPI_ISL_756425,
EPI_ISL_756428, EPI_ISL_756433, EPI_ISL_756435 to 756437, EPI_ISL_756439, EPI_ISL_756441, EPI_ISL_756443, EPI_ISL_756445, EPI_ISL_756447, EPI_ISL_756449 to 756452, EPI_ISL_756455, EPI_ISL_756457 to 756459, EPI_ISL_756461, EPI_ISL_756463 to 756464, EPI_ISL_756466, EPI_ISL_756468,
EPI_ISL_756470, EPI_ISL_756472 to 756473, EPI_ISL_756476 to 756477, EPI_ISL_756479, EPI_ISL_756482 to 756483, EPI_ISL_756488 to 756489, EPI_ISL_756493 to 756495, EPI_ISL_756503 to 756504, EPI_ISL_756510, EPI_ISL_756514 to 756516, EPI_ISL_756520, EPI_ISL_756523 to 756524, EPI_ISL_756527,
EPI_ISL_756530 to 756533, EPI_ISL_756536 to 756539, EPI_ISL_756541, EPI_ISL_756544, EPI_ISL_756546 to 756548, EPI_ISL_756550 to 756552, EPI_ISL_756555 to 756556, EPI_ISL_756560 to 756563, EPI_ISL_756565 to 756566, EPI_ISL_756569, EPI_ISL_756571, EPI_ISL_756574, EPI_ISL_756578, EPI_ISL_756580,
EPI_ISL_756583, EPI_ISL_756587, EPI_ISL_756592, EPI_ISL_756596, EPI_ISL_756598 to 756600, EPI_ISL_756602 to 756603, EPI_ISL_756609, EPI_ISL_756612, EPI_ISL_756616, EPI_ISL_756618 to 756620, EPI_ISL_756624, EPI_ISL_756626, EPI_ISL_756634 to 756636, EPI_ISL_756638 to 756641, EPI_ISL_756644 to
756645, EPI_ISL_756647 to 756648, EPI_ISL_756650 to 756651, EPI_ISL_756654, EPI_ISL_756656 to 756658, EPI_ISL_756661 to 756664, EPI_ISL_756669 to 756670, EPI_ISL_756673, EPI_ISL_756681 to 756683, EPI_ISL_756685 to 756686, EPI_ISL_756688, EPI_ISL_760249, EPI_ISL_760259, EPI_ISL_760261,
EPI_ISL_760263, EPI_ISL_760266 to 760268, EPI_ISL_760273, EPI_ISL_760276, EPI_ISL_760278, EPI_ISL_760281, EPI_ISL_760284, EPI_ISL_760286, EPI_ISL_760288, EPI_ISL_760293, EPI_ISL_760296, EPI_ISL_760301, EPI_ISL_760305 to 760306, EPI_ISL_760311, EPI_ISL_760326, EPI_ISL_760333, EPI_ISL_760341,
EPI_ISL_760344, EPI_ISL_760346, EPI_ISL_760358 to 760360, EPI_ISL_760364, EPI_ISL_760369 to 760372, EPI_ISL_760381, EPI_ISL_760386 to 760387, EPI_ISL_760390, EPI_ISL_760392, EPI_ISL_760397, EPI_ISL_760405, EPI_ISL_760408, EPI_ISL_760415, EPI_ISL_760418 to 760419, EPI_ISL_760421,
EPI_ISL_760424, EPI_ISL_760430, EPI_ISL_760432, EPI_ISL_760436, EPI_ISL_760441 to 760442, EPI_ISL_760445, EPI_ISL_760448, EPI_ISL_760450, EPI_ISL_760460 to 760461, EPI_ISL_760471, EPI_ISL_760474 to 760475, EPI_ISL_760485 to 760486, EPI_ISL_760491, EPI_ISL_760495, EPI_ISL_760498,
EPI_ISL_760500 to 760501, EPI_ISL_760508 to 760509, EPI_ISL_760512, EPI_ISL_760516, EPI_ISL_760521, EPI_ISL_760524, EPI_ISL_760530 to 760531, EPI_ISL_760534 to 760535, EPI_ISL_760538, EPI_ISL_760543 to 760544, EPI_ISL_760546, EPI_ISL_760549, EPI_ISL_760551 to 760552, EPI_ISL_760555,
EPI_ISL_760558 to 760559, EPI_ISL_760561 to 760562, EPI_ISL_760564 to 760565, EPI_ISL_760567, EPI_ISL_760572, EPI_ISL_760574, EPI_ISL_760582, EPI_ISL_760586 to 760589, EPI_ISL_760591, EPI_ISL_760593, EPI_ISL_760596 to 760597, EPI_ISL_760599 to 760601, EPI_ISL_760608 to 760609, EPI_ISL_760612 to
760613, EPI_ISL_760615, EPI_ISL_760617, EPI_ISL_760623 to 760625, EPI_ISL_760627 to 760628, EPI_ISL_760633, EPI_ISL_760637, EPI_ISL_760640, EPI_ISL_760644, EPI_ISL_760647, EPI_ISL_760649 to 760650, EPI_ISL_760652, EPI_ISL_760654, EPI_ISL_760656, EPI_ISL_760660 to 760661, EPI_ISL_760663 to
760667, EPI_ISL_760670, EPI_ISL_760677, EPI_ISL_760679 to 760680, EPI_ISL_760684 to 760685, EPI_ISL_760688, EPI_ISL_760691, EPI_ISL_760693, EPI_ISL_760699, EPI_ISL_760701 to 760702, EPI_ISL_760705, EPI_ISL_760708, EPI_ISL_760710 to 760711, EPI_ISL_760715, EPI_ISL_760719, EPI_ISL_760721 to
760723, EPI_ISL_760726 to 760728, EPI_ISL_760733 to 760734, EPI_ISL_760736 to 760738, EPI_ISL_760745, EPI_ISL_760748, EPI_ISL_760752 to 760753, EPI_ISL_760755 to 760756, EPI_ISL_760759, EPI_ISL_760761 to 760762, EPI_ISL_760769, EPI_ISL_760776, EPI_ISL_760780, EPI_ISL_760783 to 760786,
EPI_ISL_760788, EPI_ISL_760790, EPI_ISL_760792, EPI_ISL_760794 to 760795, EPI_ISL_760799, EPI_ISL_760802, EPI_ISL_760804, EPI_ISL_760806, EPI_ISL_760808, EPI_ISL_760812, EPI_ISL_760817, EPI_ISL_760825 to 760826, EPI_ISL_760832, EPI_ISL_760838 to 760839, EPI_ISL_760852 to 760853,
EPI_ISL_760857, EPI_ISL_760860 to 760862, EPI_ISL_760864 to 760865, EPI_ISL_760867 to 760868, EPI_ISL_760874, EPI_ISL_760878, EPI_ISL_760884, EPI_ISL_760887, EPI_ISL_760889, EPI_ISL_760892, EPI_ISL_760897 to 760898, EPI_ISL_760900, EPI_ISL_760906, EPI_ISL_760908, EPI_ISL_760912,
EPI_ISL_760915 to 760916, EPI_ISL_760918, EPI_ISL_760921, EPI_ISL_760925, EPI_ISL_760928 to 760929, EPI_ISL_760931, EPI_ISL_760933, EPI_ISL_760937 to 760938, EPI_ISL_760941 to 760942, EPI_ISL_760945, EPI_ISL_760948 to 760950, EPI_ISL_760952, EPI_ISL_760960, EPI_ISL_760962, EPI_ISL_760967 to
760968, EPI_ISL_760971, EPI_ISL_760978 to 760979, EPI_ISL_760982, EPI_ISL_760989, EPI_ISL_760994, EPI_ISL_761000 to 761001, EPI_ISL_761011, EPI_ISL_761013, EPI_ISL_761022, EPI_ISL_761025, EPI_ISL_761027, EPI_ISL_761029, EPI_ISL_761040, EPI_ISL_761046, EPI_ISL_761048 to 761049, EPI_ISL_761052,
EPI_ISL_761054, EPI_ISL_761056, EPI_ISL_761060 to 761061, EPI_ISL_761064 to 761066, EPI_ISL_761069, EPI_ISL_761080 to 761090, EPI_ISL_761092 to 761093, EPI_ISL_761096 to 761104, EPI_ISL_761106 to 761112, EPI_ISL_761115, EPI_ISL_761117, EPI_ISL_761119, EPI_ISL_761124, EPI_ISL_761126 to 761127,
EPI_ISL_761129 to 761130, EPI_ISL_761132, EPI_ISL_761136 to 761138, EPI_ISL_761140 to 761142, EPI_ISL_761144, EPI_ISL_761147 to 761153, EPI_ISL_761155 to 761156, EPI_ISL_761159 to 761161, EPI_ISL_761163 to 761167, EPI_ISL_761169 to 761171, EPI_ISL_761173 to 761174, EPI_ISL_761177 to 761186,
EPI_ISL_761188 to 761189, EPI_ISL_761192, EPI_ISL_761195 to 761196, EPI_ISL_761198, EPI_ISL_761200 to 761202, EPI_ISL_761205 to 761206, EPI_ISL_761208, EPI_ISL_761210 to 761211, EPI_ISL_761213, EPI_ISL_761215 to 761217, EPI_ISL_761219 to 761220, EPI_ISL_761222 to 761224, EPI_ISL_761230 to 761238,
EPI_ISL_761240, EPI_ISL_761243 to 761251, EPI_ISL_761253 to 761254, EPI_ISL_761256, EPI_ISL_761260 to 761265, EPI_ISL_761269 to 761275, EPI_ISL_761277, EPI_ISL_761279, EPI_ISL_761281 to 761282, EPI_ISL_761284 to 761286, EPI_ISL_761288, EPI_ISL_761290 to 761291, EPI_ISL_761294, EPI_ISL_761297,
EPI_ISL_761299 to 761301, EPI_ISL_761304 to 761306, EPI_ISL_761308 to 761310, EPI_ISL_761312, EPI_ISL_761314 to 761325, EPI_ISL_761327 to 761330, EPI_ISL_761332 to 761337, EPI_ISL_761341 to 761347, EPI_ISL_761349 to 761350, EPI_ISL_761352, EPI_ISL_761354, EPI_ISL_761356, EPI_ISL_761665 to 761678,
EPI_ISL_761680 to 761684, EPI_ISL_761686 to 761693, EPI_ISL_761695, EPI_ISL_761697 to 761703, EPI_ISL_761705, EPI_ISL_761708 to 761713, EPI_ISL_761715 to 761719, EPI_ISL_761722 to 761732, EPI_ISL_761734, EPI_ISL_761736 to 761740, EPI_ISL_761742 to 761747, EPI_ISL_761750 to 761763, EPI_ISL_761765
to 761769, EPI_ISL_761772, EPI_ISL_761774 to 761783, EPI_ISL_761785 to 761792, EPI_ISL_761794 to 761800, EPI_ISL_761802 to 761805, EPI_ISL_761807 to 761818, EPI_ISL_761820 to 761822, EPI_ISL_761824 to 761827, EPI_ISL_761829 to 761832, EPI_ISL_761835 to 761846, EPI_ISL_761848 to 761858,
EPI_ISL_761860 to 761863, EPI_ISL_761866 to 761875, EPI_ISL_761877 to 761901, EPI_ISL_761903 to 761904, EPI_ISL_761906 to 761910, EPI_ISL_761912 to 761913, EPI_ISL_761915 to 761917, EPI_ISL_761919 to 761920, EPI_ISL_762547, EPI_ISL_762554, EPI_ISL_762557 to 762558, EPI_ISL_762562, EPI_ISL_762564,
EPI_ISL_762581, EPI_ISL_762588, EPI_ISL_762599, EPI_ISL_762605 to 762606, EPI_ISL_762610, EPI_ISL_762617, EPI_ISL_762620, EPI_ISL_762625, EPI_ISL_762629, EPI_ISL_762638, EPI_ISL_762642 to 762643, EPI_ISL_762647, EPI_ISL_762655, EPI_ISL_762659, EPI_ISL_762666, EPI_ISL_762671 to 762672,
EPI_ISL_762674, EPI_ISL_762683, EPI_ISL_762686, EPI_ISL_762691 to 762693, EPI_ISL_762702 to 762703, EPI_ISL_762706 to 762707, EPI_ISL_762709 to 762710, EPI_ISL_762712, EPI_ISL_762714, EPI_ISL_762720, EPI_ISL_762722, EPI_ISL_762724, EPI_ISL_762730, EPI_ISL_762735, EPI_ISL_762750,
EPI_ISL_762754, EPI_ISL_762756, EPI_ISL_762763 to 762764, EPI_ISL_762777, EPI_ISL_762786, EPI_ISL_762789, EPI_ISL_762791, EPI_ISL_762793, EPI_ISL_762796, EPI_ISL_762798 to 762799, EPI_ISL_762815, EPI_ISL_762819, EPI_ISL_762821, EPI_ISL_762823, EPI_ISL_762830, EPI_ISL_762832, EPI_ISL_762835,
EPI_ISL_762839, EPI_ISL_762841, EPI_ISL_762843 to 762845, EPI_ISL_762847, EPI_ISL_762849, EPI_ISL_762869, EPI_ISL_762878 to 762879, EPI_ISL_762883, EPI_ISL_762886, EPI_ISL_762890, EPI_ISL_762894, EPI_ISL_762900 to 762902, EPI_ISL_762906, EPI_ISL_762910 to 762911, EPI_ISL_762913,
EPI_ISL_762919, EPI_ISL_762921 to 762922, EPI_ISL_762926, EPI_ISL_762931, EPI_ISL_762935, EPI_ISL_762939, EPI_ISL_762947, EPI_ISL_762949, EPI_ISL_762951 to 762952, EPI_ISL_762955, EPI_ISL_762957, EPI_ISL_762964, EPI_ISL_762973 to 762975, EPI_ISL_762977, EPI_ISL_762988 to 762989,
EPI_ISL_767091, EPI_ISL_767093 to 767095, EPI_ISL_767097, EPI_ISL_767103 to 767106, EPI_ISL_767116, EPI_ISL_767120 to 767133, EPI_ISL_767135 to 767137, EPI_ISL_767139 to 767153, EPI_ISL_767155 to 767218, EPI_ISL_767220 to 767224, EPI_ISL_767226 to 767229, EPI_ISL_767231 to 767236, EPI_ISL_767238,
EPI_ISL_767240 to 767250, EPI_ISL_767252 to 767259, EPI_ISL_767261 to 767265, EPI_ISL_767268 to 767270, EPI_ISL_767272 to 767279, EPI_ISL_767281 to 767284, EPI_ISL_767286 to 767293, EPI_ISL_767295 to 767296, EPI_ISL_767298, EPI_ISL_767301 to 767303, EPI_ISL_767305 to 767330, EPI_ISL_767332 to
767333, EPI_ISL_768841 to 768844, EPI_ISL_768898 to 768900, EPI_ISL_777011 to 777012, EPI_ISL_777014 to 777015, EPI_ISL_777017 to 777018, EPI_ISL_777020, EPI_ISL_777024 to 777028, EPI_ISL_777030, EPI_ISL_777032 to 777033, EPI_ISL_777035 to 777036, EPI_ISL_777043 to 777046, EPI_ISL_777048 to
777053, EPI_ISL_777057, EPI_ISL_777060, EPI_ISL_777063, EPI_ISL_777067, EPI_ISL_777069 to 777072, EPI_ISL_777074 to 777079, EPI_ISL_777083, EPI_ISL_777087 to 777088, EPI_ISL_777090 to 777094, EPI_ISL_777096, EPI_ISL_777103, EPI_ISL_777107, EPI_ISL_777109 to 777111, EPI_ISL_777113 to 777114,
EPI_ISL_777116, EPI_ISL_777119, EPI_ISL_777123, EPI_ISL_777125 to 777126, EPI_ISL_777128 to 777129, EPI_ISL_777134, EPI_ISL_777136 to 777139, EPI_ISL_777143 to 777144, EPI_ISL_777147, EPI_ISL_777150, EPI_ISL_777153, EPI_ISL_777155, EPI_ISL_777157, EPI_ISL_777159, EPI_ISL_777163,
EPI_ISL_777165 to 777168, EPI_ISL_777170 to 777171, EPI_ISL_777174 to 777175, EPI_ISL_777177 to 777179, EPI_ISL_777183, EPI_ISL_777185 to 777186, EPI_ISL_777188, EPI_ISL_777190, EPI_ISL_777193, EPI_ISL_777195, EPI_ISL_777197 to 777198, EPI_ISL_777202 to 777204, EPI_ISL_777210, EPI_ISL_777212 to
777214, EPI_ISL_777216 to 777217, EPI_ISL_777220 to 777221, EPI_ISL_777226, EPI_ISL_777228, EPI_ISL_777231, EPI_ISL_777234, EPI_ISL_777237, EPI_ISL_777239 to 777240, EPI_ISL_777243 to 777244, EPI_ISL_777247 to 777249, EPI_ISL_777251, EPI_ISL_777254, EPI_ISL_777256 to 777260, EPI_ISL_777262 to
777266, EPI_ISL_777270 to 777272, EPI_ISL_777274, EPI_ISL_777276 to 777281, EPI_ISL_777285 to 777288, EPI_ISL_777291 to 777292, EPI_ISL_777294, EPI_ISL_777296 to 777297, EPI_ISL_777299, EPI_ISL_777307 to 777309, EPI_ISL_777311 to 777312, EPI_ISL_777314 to 777315, EPI_ISL_777318 to 777319,
EPI_ISL_777326 to 777327, EPI_ISL_777333, EPI_ISL_777336, EPI_ISL_777338, EPI_ISL_777375, EPI_ISL_777380, EPI_ISL_777386, EPI_ISL_777389, EPI_ISL_777411 to 777412, EPI_ISL_777416, EPI_ISL_777424, EPI_ISL_777427, EPI_ISL_777430, EPI_ISL_777435, EPI_ISL_777437, EPI_ISL_777440, EPI_ISL_777448,
EPI_ISL_777451, EPI_ISL_777458, EPI_ISL_777465, EPI_ISL_777467, EPI_ISL_777470, EPI_ISL_777477, EPI_ISL_777479, EPI_ISL_777481, EPI_ISL_777489, EPI_ISL_777491, EPI_ISL_777495, EPI_ISL_777502, EPI_ISL_777509, EPI_ISL_777522, EPI_ISL_777524, EPI_ISL_777529 to 777530, EPI_ISL_777532,
EPI_ISL_777534 to 777535, EPI_ISL_777538 to 777539, EPI_ISL_777548, EPI_ISL_777550, EPI_ISL_777552, EPI_ISL_777558, EPI_ISL_777561, EPI_ISL_777563, EPI_ISL_777567 to 777568, EPI_ISL_777573, EPI_ISL_777576, EPI_ISL_777580, EPI_ISL_777582, EPI_ISL_777584, EPI_ISL_777587, EPI_ISL_777589,
EPI_ISL_777591 to 777593, EPI_ISL_777595, EPI_ISL_777597, EPI_ISL_777605, EPI_ISL_777621, EPI_ISL_777627, EPI_ISL_777629, EPI_ISL_777633 to 777634, EPI_ISL_777638 to 777639, EPI_ISL_777644, EPI_ISL_777649 to 777651, EPI_ISL_777655, EPI_ISL_777659, EPI_ISL_777669, EPI_ISL_777672,
EPI_ISL_778016, EPI_ISL_778018, EPI_ISL_778021, EPI_ISL_778029, EPI_ISL_778033, EPI_ISL_778035, EPI_ISL_778040, EPI_ISL_778043 to 778044, EPI_ISL_778048, EPI_ISL_778050 to 778052, EPI_ISL_778055, EPI_ISL_778059, EPI_ISL_778064 to 778065, EPI_ISL_778069 to 778070, EPI_ISL_778074,
EPI_ISL_778083, EPI_ISL_778089, EPI_ISL_778101, EPI_ISL_778104, EPI_ISL_778108, EPI_ISL_778113, EPI_ISL_778116, EPI_ISL_778121, EPI_ISL_778128, EPI_ISL_778132, EPI_ISL_778144, EPI_ISL_778163 to 778166, EPI_ISL_778183, EPI_ISL_778187, EPI_ISL_778190 to 778191, EPI_ISL_778196, EPI_ISL_778201,
EPI_ISL_778207, EPI_ISL_778212, EPI_ISL_778225, EPI_ISL_778228, EPI_ISL_778230, EPI_ISL_778233 to 778234, EPI_ISL_778238 to 778240, EPI_ISL_778246, EPI_ISL_778249, EPI_ISL_778256, EPI_ISL_778259, EPI_ISL_778261, EPI_ISL_778264, EPI_ISL_778266, EPI_ISL_778272, EPI_ISL_778282, EPI_ISL_778287 to
778288, EPI_ISL_778290, EPI_ISL_778292, EPI_ISL_778295, EPI_ISL_778305, EPI_ISL_778309, EPI_ISL_778311, EPI_ISL_778315, EPI_ISL_778327 to 778328, EPI_ISL_778331, EPI_ISL_778333, EPI_ISL_778335, EPI_ISL_778346 to 778348, EPI_ISL_778353, EPI_ISL_778355, EPI_ISL_778357 to 778358, EPI_ISL_778360,
EPI_ISL_778365, EPI_ISL_778368, EPI_ISL_778374, EPI_ISL_778380, EPI_ISL_778382, EPI_ISL_778385, EPI_ISL_778390, EPI_ISL_778392, EPI_ISL_778396, EPI_ISL_778399, EPI_ISL_778401, EPI_ISL_778404, EPI_ISL_778408, EPI_ISL_778410, EPI_ISL_778412, EPI_ISL_778415 to 778417, EPI_ISL_778426,
EPI_ISL_778430, EPI_ISL_778432, EPI_ISL_778444, EPI_ISL_778448, EPI_ISL_778454 to 778455, EPI_ISL_778471 to 778473, EPI_ISL_778478, EPI_ISL_778484, EPI_ISL_778487 to 778488, EPI_ISL_778490 to 778491, EPI_ISL_778494, EPI_ISL_778500, EPI_ISL_778510, EPI_ISL_778512, EPI_ISL_778517,
EPI_ISL_778524, EPI_ISL_778528 to 778529, EPI_ISL_778531, EPI_ISL_778539, EPI_ISL_778543, EPI_ISL_778547, EPI_ISL_778556, EPI_ISL_778558, EPI_ISL_778562, EPI_ISL_778564, EPI_ISL_778569, EPI_ISL_778571, EPI_ISL_778578, EPI_ISL_778584, EPI_ISL_778593, EPI_ISL_778599, EPI_ISL_778601,
EPI_ISL_778605, EPI_ISL_778607, EPI_ISL_778614 to 778616, EPI_ISL_778618, EPI_ISL_778621, EPI_ISL_778623, EPI_ISL_778627 to 778628, EPI_ISL_778630, EPI_ISL_778635, EPI_ISL_781186, EPI_ISL_781195, EPI_ISL_781201 to 781202, EPI_ISL_781205 to 781207, EPI_ISL_781210, EPI_ISL_781214,
EPI_ISL_781219, EPI_ISL_781230, EPI_ISL_781234 to 781235, EPI_ISL_781250, EPI_ISL_781256, EPI_ISL_781273, EPI_ISL_781280, EPI_ISL_781282, EPI_ISL_781288, EPI_ISL_781291 to 781292, EPI_ISL_781294, EPI_ISL_781297, EPI_ISL_781301, EPI_ISL_781307, EPI_ISL_781313, EPI_ISL_781318, EPI_ISL_781323 to
781324, EPI_ISL_781330, EPI_ISL_781336 to 781337, EPI_ISL_781348 to 781349, EPI_ISL_781352, EPI_ISL_781356, EPI_ISL_781366, EPI_ISL_781374 to 781375, EPI_ISL_781378, EPI_ISL_781389, EPI_ISL_781400, EPI_ISL_781405 to 781406, EPI_ISL_781408, EPI_ISL_781412, EPI_ISL_781433, EPI_ISL_781435,
EPI_ISL_781437, EPI_ISL_781443, EPI_ISL_781446 to 781447, EPI_ISL_781451, EPI_ISL_781457 to 781458, EPI_ISL_781463 to 781465, EPI_ISL_781469, EPI_ISL_781473, EPI_ISL_781475, EPI_ISL_781478, EPI_ISL_781499, EPI_ISL_781502, EPI_ISL_781505, EPI_ISL_781508, EPI_ISL_781510, EPI_ISL_781516,
EPI_ISL_781872, EPI_ISL_781874, EPI_ISL_781877, EPI_ISL_781879, EPI_ISL_781881, EPI_ISL_781883 to 781884, EPI_ISL_781887, EPI_ISL_781892 to 781894, EPI_ISL_781896, EPI_ISL_781899 to 781902, EPI_ISL_781904 to 781905, EPI_ISL_781911 to 781912, EPI_ISL_781917, EPI_ISL_781923, EPI_ISL_781926 to
781927, EPI_ISL_781929, EPI_ISL_781931 to 781932, EPI_ISL_781934 to 781937, EPI_ISL_781939, EPI_ISL_781943, EPI_ISL_781945, EPI_ISL_781949, EPI_ISL_781951, EPI_ISL_781954, EPI_ISL_781957 to 781959, EPI_ISL_781961 to 781966, EPI_ISL_781969 to 781970, EPI_ISL_781972, EPI_ISL_781974 to 781976,
EPI_ISL_781981, EPI_ISL_781983, EPI_ISL_781985 to 781986, EPI_ISL_781989, EPI_ISL_781991, EPI_ISL_781993, EPI_ISL_781995 to 781996, EPI_ISL_782002, EPI_ISL_782005 to 782007, EPI_ISL_782010 to 782013, EPI_ISL_782016, EPI_ISL_782019 to 782020, EPI_ISL_782022 to 782025, EPI_ISL_782029,
EPI_ISL_782033 to 782034, EPI_ISL_782041, EPI_ISL_782043, EPI_ISL_782045, EPI_ISL_782047 to 782048, EPI_ISL_782050, EPI_ISL_782053, EPI_ISL_782057, EPI_ISL_782060, EPI_ISL_782063, EPI_ISL_782068, EPI_ISL_782070 to 782072, EPI_ISL_782076, EPI_ISL_782078, EPI_ISL_782082 to 782083,
EPI_ISL_782085 to 782086, EPI_ISL_782089 to 782091, EPI_ISL_782095 to 782097, EPI_ISL_782099 to 782100, EPI_ISL_782102, EPI_ISL_782104, EPI_ISL_782108 to 782111, EPI_ISL_782117, EPI_ISL_782119 to 782120, EPI_ISL_782122 to 782124, EPI_ISL_782126, EPI_ISL_782132, EPI_ISL_782135, EPI_ISL_782139 to
782142, EPI_ISL_782148 to 782150, EPI_ISL_782161, EPI_ISL_782165, EPI_ISL_782201 to 782202, EPI_ISL_782207, EPI_ISL_782211 to 782212, EPI_ISL_782220, EPI_ISL_782224, EPI_ISL_782226, EPI_ISL_782235, EPI_ISL_782242 to 782243, EPI_ISL_782245, EPI_ISL_782249, EPI_ISL_782252, EPI_ISL_782259,
EPI_ISL_782264, EPI_ISL_782267, EPI_ISL_782278, EPI_ISL_782282, EPI_ISL_782287, EPI_ISL_782292, EPI_ISL_782307, EPI_ISL_782309, EPI_ISL_782313 to 782315, EPI_ISL_782326, EPI_ISL_782335, EPI_ISL_782345, EPI_ISL_782352, EPI_ISL_782355, EPI_ISL_782358, EPI_ISL_782361 to 782362, EPI_ISL_782369,
EPI_ISL_782372, EPI_ISL_782377, EPI_ISL_782380, EPI_ISL_782388, EPI_ISL_782393, EPI_ISL_782402, EPI_ISL_782409, EPI_ISL_782412, EPI_ISL_782420, EPI_ISL_782423, EPI_ISL_782425 to 782426, EPI_ISL_782435, EPI_ISL_782439, EPI_ISL_782451, EPI_ISL_782454, EPI_ISL_782471, EPI_ISL_782474,
EPI_ISL_782478, EPI_ISL_782737, EPI_ISL_782745, EPI_ISL_782747 to 782748, EPI_ISL_782758, EPI_ISL_782768, EPI_ISL_782771, EPI_ISL_782774, EPI_ISL_782783 to 782784, EPI_ISL_782787, EPI_ISL_782791 to 782792, EPI_ISL_782795, EPI_ISL_782801, EPI_ISL_782805 to 782806, EPI_ISL_782808,
EPI_ISL_782813, EPI_ISL_782820, EPI_ISL_782825, EPI_ISL_782827, EPI_ISL_782838, EPI_ISL_782841, EPI_ISL_782843, EPI_ISL_782849, EPI_ISL_782855 to 782856, EPI_ISL_782869, EPI_ISL_782872 to 782873, EPI_ISL_782877 to 782878, EPI_ISL_782892 to 782894, EPI_ISL_782897, EPI_ISL_782899,
EPI_ISL_782902, EPI_ISL_782904 to 782905, EPI_ISL_782918 to 782919, EPI_ISL_782924, EPI_ISL_782926, EPI_ISL_782931, EPI_ISL_782936, EPI_ISL_782945 to 782946, EPI_ISL_782956, EPI_ISL_782963 to 782964, EPI_ISL_782969 to 782970, EPI_ISL_782976, EPI_ISL_782988, EPI_ISL_782993 to 782994,

EPI_ISL_782997, EPI_ISL_783000, EPI_ISL_783002, EPI_ISL_783008, EPI_ISL_783014 to 783015, EPI_ISL_783026, EPI_ISL_783030, EPI_ISL_783033, EPI_ISL_783045 to 783046, EPI_ISL_783052, EPI_ISL_783056, EPI_ISL_783068, EPI_ISL_796784 to 796858, EPI_ISL_796860 to 796889, EPI_ISL_796891,
EPI_ISL_796893 to 796960, EPI_ISL_796962 to 797013, EPI_ISL_797015 to 797076, EPI_ISL_797078 to 797117, EPI_ISL_797399, EPI_ISL_797402, EPI_ISL_797409, EPI_ISL_797413, EPI_ISL_797416, EPI_ISL_797420 to 797421, EPI_ISL_797424, EPI_ISL_797430, EPI_ISL_797435, EPI_ISL_797456, EPI_ISL_797468,
EPI_ISL_797480, EPI_ISL_797489, EPI_ISL_797491, EPI_ISL_797499, EPI_ISL_797512, EPI_ISL_797533, EPI_ISL_797535, EPI_ISL_797540 to 797541, EPI_ISL_797543, EPI_ISL_797547, EPI_ISL_797549, EPI_ISL_797568, EPI_ISL_797587, EPI_ISL_797599, EPI_ISL_797606, EPI_ISL_797610, EPI_ISL_797613,
EPI_ISL_797616, EPI_ISL_797637, EPI_ISL_797641 to 797642, EPI_ISL_797644, EPI_ISL_797654, EPI_ISL_797660, EPI_ISL_797663, EPI_ISL_797666, EPI_ISL_797699, EPI_ISL_797709, EPI_ISL_797714 to 797715, EPI_ISL_797717, EPI_ISL_797999, EPI_ISL_798002, EPI_ISL_798007, EPI_ISL_798009, EPI_ISL_798011,
EPI_ISL_798013, EPI_ISL_798015, EPI_ISL_798025, EPI_ISL_798027, EPI_ISL_798029, EPI_ISL_798035, EPI_ISL_798054, EPI_ISL_798061 to 798062, EPI_ISL_798064, EPI_ISL_798066, EPI_ISL_798068, EPI_ISL_798073, EPI_ISL_798083 to 798084, EPI_ISL_798096, EPI_ISL_798101, EPI_ISL_798103, EPI_ISL_798106,
EPI_ISL_798108, EPI_ISL_798119, EPI_ISL_798122 to 798125, EPI_ISL_798127, EPI_ISL_798131, EPI_ISL_798133 to 798134, EPI_ISL_798139, EPI_ISL_798152, EPI_ISL_798154, EPI_ISL_798169 to 798170, EPI_ISL_798173 to 798176, EPI_ISL_798181 to 798182, EPI_ISL_798202, EPI_ISL_798204, EPI_ISL_798208,
EPI_ISL_798220 to 798222, EPI_ISL_798225, EPI_ISL_798229, EPI_ISL_798234 to 798236, EPI_ISL_798239, EPI_ISL_798241 to 798242, EPI_ISL_798245 to 798246, EPI_ISL_798250, EPI_ISL_798255, EPI_ISL_798268, EPI_ISL_798274 to 798275, EPI_ISL_798277, EPI_ISL_798280, EPI_ISL_798282 to 798285,
EPI_ISL_798292, EPI_ISL_798295 to 798296, EPI_ISL_798299, EPI_ISL_798305, EPI_ISL_798310, EPI_ISL_798314, EPI_ISL_798321, EPI_ISL_798323 to 798324, EPI_ISL_798328 to 798329, EPI_ISL_798334, EPI_ISL_798336, EPI_ISL_798341, EPI_ISL_798345 to 798346, EPI_ISL_798349, EPI_ISL_798369 to 798370,
EPI_ISL_798379, EPI_ISL_798384, EPI_ISL_798386, EPI_ISL_798388 to 798389, EPI_ISL_798399, EPI_ISL_798404, EPI_ISL_798413, EPI_ISL_798416 to 798417, EPI_ISL_798431, EPI_ISL_798436, EPI_ISL_798444, EPI_ISL_798455, EPI_ISL_798465, EPI_ISL_798468, EPI_ISL_798475, EPI_ISL_798479, EPI_ISL_798492 to
798493, EPI_ISL_798500, EPI_ISL_798502, EPI_ISL_798508, EPI_ISL_798510, EPI_ISL_798513, EPI_ISL_798516 to 798517, EPI_ISL_798519, EPI_ISL_798524, EPI_ISL_798534, EPI_ISL_798539, EPI_ISL_798541 to 798542, EPI_ISL_798544, EPI_ISL_798548, EPI_ISL_798550, EPI_ISL_798555, EPI_ISL_798561,
EPI_ISL_798564, EPI_ISL_798591, EPI_ISL_798593, EPI_ISL_798598, EPI_ISL_798601, EPI_ISL_798609, EPI_ISL_798612, EPI_ISL_798615, EPI_ISL_798622, EPI_ISL_798631 to 798633, EPI_ISL_798635, EPI_ISL_798637, EPI_ISL_798639 to 798641, EPI_ISL_798643, EPI_ISL_798652, EPI_ISL_798659, EPI_ISL_798665,
EPI_ISL_798667 to 798669, EPI_ISL_798671, EPI_ISL_798676, EPI_ISL_798684, EPI_ISL_798688 to 798690, EPI_ISL_798692, EPI_ISL_798695, EPI_ISL_798704 to 798705, EPI_ISL_798709, EPI_ISL_798719, EPI_ISL_798722, EPI_ISL_798727, EPI_ISL_798729, EPI_ISL_798735, EPI_ISL_798742, EPI_ISL_798744,
EPI_ISL_798749, EPI_ISL_798768, EPI_ISL_798771, EPI_ISL_798784, EPI_ISL_798791, EPI_ISL_798793, EPI_ISL_798798, EPI_ISL_798804, EPI_ISL_798811, EPI_ISL_798814, EPI_ISL_798819, EPI_ISL_798824, EPI_ISL_798826, EPI_ISL_798833, EPI_ISL_798845, EPI_ISL_798850, EPI_ISL_798852 to 798853,
EPI_ISL_798869, EPI_ISL_798872, EPI_ISL_798878, EPI_ISL_798880, EPI_ISL_798884 to 798885, EPI_ISL_798888, EPI_ISL_798891, EPI_ISL_798898, EPI_ISL_798902, EPI_ISL_798905 to 798906, EPI_ISL_798914, EPI_ISL_798921, EPI_ISL_798924, EPI_ISL_798926, EPI_ISL_798928, EPI_ISL_798934 to 798935,
EPI_ISL_798939, EPI_ISL_798941, EPI_ISL_798951 to 798952, EPI_ISL_798957, EPI_ISL_798962 to 798964, EPI_ISL_798968 to 798969, EPI_ISL_798984, EPI_ISL_798986, EPI_ISL_798988, EPI_ISL_798991, EPI_ISL_799000, EPI_ISL_799003 to 799005, EPI_ISL_799010, EPI_ISL_799021 to 799022, EPI_ISL_799025 to
799026, EPI_ISL_799029, EPI_ISL_799040, EPI_ISL_799045, EPI_ISL_799053, EPI_ISL_799055, EPI_ISL_799057, EPI_ISL_799059, EPI_ISL_799069 to 799070, EPI_ISL_799079, EPI_ISL_799085, EPI_ISL_799099, EPI_ISL_799103, EPI_ISL_799105 to 799106, EPI_ISL_799111, EPI_ISL_799113 to 799114, EPI_ISL_799122,
EPI_ISL_799124, EPI_ISL_799137 to 799139, EPI_ISL_799143, EPI_ISL_799145, EPI_ISL_799149, EPI_ISL_799154, EPI_ISL_799160 to 799161, EPI_ISL_799165 to 799166, EPI_ISL_799170 to 799171, EPI_ISL_799177 to 799178, EPI_ISL_799182, EPI_ISL_799187, EPI_ISL_799191, EPI_ISL_799198, EPI_ISL_799200,
EPI_ISL_799202, EPI_ISL_799212 to 799214, EPI_ISL_799219, EPI_ISL_799227 to 799229, EPI_ISL_799231, EPI_ISL_799233 to 799235, EPI_ISL_799237, EPI_ISL_799244, EPI_ISL_799248, EPI_ISL_799252, EPI_ISL_799257, EPI_ISL_799265, EPI_ISL_799269, EPI_ISL_799271, EPI_ISL_799276, EPI_ISL_799278,
EPI_ISL_799281, EPI_ISL_799637 to 799638, EPI_ISL_799644, EPI_ISL_799652, EPI_ISL_799655, EPI_ISL_799662, EPI_ISL_799665, EPI_ISL_799678, EPI_ISL_799683, EPI_ISL_799687, EPI_ISL_799693, EPI_ISL_799702, EPI_ISL_799706, EPI_ISL_799711, EPI_ISL_799732, EPI_ISL_799734, EPI_ISL_799744,
EPI_ISL_799747, EPI_ISL_799749 to 799751, EPI_ISL_799754, EPI_ISL_799760, EPI_ISL_799767, EPI_ISL_799784, EPI_ISL_799787, EPI_ISL_799796, EPI_ISL_799799, EPI_ISL_799809, EPI_ISL_799814, EPI_ISL_799818, EPI_ISL_799821 to 799823, EPI_ISL_799840 to 799841, EPI_ISL_799844, EPI_ISL_799846,
EPI_ISL_799859, EPI_ISL_799868, EPI_ISL_799877, EPI_ISL_799881, EPI_ISL_799891, EPI_ISL_799897 to 799899, EPI_ISL_799902, EPI_ISL_799908, EPI_ISL_799914, EPI_ISL_799920, EPI_ISL_799922, EPI_ISL_799924 to 799925, EPI_ISL_799930, EPI_ISL_799933, EPI_ISL_799938, EPI_ISL_799940 to 799943,
EPI_ISL_799948, EPI_ISL_799956, EPI_ISL_799961, EPI_ISL_799964, EPI_ISL_799966 to 799967, EPI_ISL_799970, EPI_ISL_799975, EPI_ISL_799977 to 799979, EPI_ISL_799982 to 799984, EPI_ISL_799989, EPI_ISL_799991, EPI_ISL_799995, EPI_ISL_799998 to 799999, EPI_ISL_800002 to 800003, EPI_ISL_800009,
EPI_ISL_800011, EPI_ISL_800013 to 800014, EPI_ISL_800016 to 800017, EPI_ISL_800019 to 800023, EPI_ISL_800025, EPI_ISL_800031, EPI_ISL_800033, EPI_ISL_800035 to 800036, EPI_ISL_800039 to 800044, EPI_ISL_800046, EPI_ISL_800049 to 800051, EPI_ISL_800053, EPI_ISL_800058 to 800063, EPI_ISL_800066 to
800067, EPI_ISL_800069, EPI_ISL_800073 to 800076, EPI_ISL_800080 to 800081, EPI_ISL_800087, EPI_ISL_800090 to 800091, EPI_ISL_800094, EPI_ISL_800097, EPI_ISL_800099, EPI_ISL_800101, EPI_ISL_800104 to 800105, EPI_ISL_800107 to 800110, EPI_ISL_800112, EPI_ISL_800118, EPI_ISL_800126 to 800130,
EPI_ISL_800132, EPI_ISL_800134 to 800136, EPI_ISL_800140, EPI_ISL_800143 to 800146, EPI_ISL_800148, EPI_ISL_800150, EPI_ISL_800152, EPI_ISL_800154 to 800155, EPI_ISL_800159, EPI_ISL_800162 to 800163, EPI_ISL_800165, EPI_ISL_800169 to 800171, EPI_ISL_800179, EPI_ISL_800181, EPI_ISL_800184 to
800185, EPI_ISL_800187, EPI_ISL_800193 to 800194, EPI_ISL_800197 to 800199, EPI_ISL_800202, EPI_ISL_800205 to 800209, EPI_ISL_800212 to 800213, EPI_ISL_800215 to 800216, EPI_ISL_800219, EPI_ISL_800222 to 800224, EPI_ISL_800226 to 800227, EPI_ISL_800229 to 800232, EPI_ISL_800239 to 800240,
EPI_ISL_800242 to 800243, EPI_ISL_800246 to 800249, EPI_ISL_800262, EPI_ISL_800267, EPI_ISL_800271, EPI_ISL_800274, EPI_ISL_800280 to 800282, EPI_ISL_800286, EPI_ISL_800299, EPI_ISL_800301, EPI_ISL_800303, EPI_ISL_800308, EPI_ISL_800317, EPI_ISL_800338, EPI_ISL_800340, EPI_ISL_800342,
EPI_ISL_800347 to 800348, EPI_ISL_800350 to 800351, EPI_ISL_800355, EPI_ISL_800359 to 800360, EPI_ISL_800362, EPI_ISL_800365, EPI_ISL_800367, EPI_ISL_800370, EPI_ISL_800376, EPI_ISL_800384, EPI_ISL_800395, EPI_ISL_800407, EPI_ISL_800420, EPI_ISL_800428, EPI_ISL_800441, EPI_ISL_800444,
EPI_ISL_800446, EPI_ISL_800452 to 800454, EPI_ISL_800456 to 800457, EPI_ISL_800460, EPI_ISL_800468, EPI_ISL_800474, EPI_ISL_800481, EPI_ISL_800485, EPI_ISL_800494 to 800495, EPI_ISL_800501 to 800502, EPI_ISL_800506, EPI_ISL_800509, EPI_ISL_800516, EPI_ISL_800518, EPI_ISL_800524 to 800526,
EPI_ISL_800528, EPI_ISL_800531, EPI_ISL_800542, EPI_ISL_800555, EPI_ISL_800558, EPI_ISL_800566, EPI_ISL_800575, EPI_ISL_800577 to 800580, EPI_ISL_820129 to 820130, EPI_ISL_820132 to 820133, EPI_ISL_820135, EPI_ISL_820137, EPI_ISL_820139 to 820140, EPI_ISL_820142 to 820143, EPI_ISL_820145,
EPI_ISL_820148, EPI_ISL_820153, EPI_ISL_820155, EPI_ISL_820157 to 820158, EPI_ISL_820160, EPI_ISL_820162 to 820163, EPI_ISL_820165 to 820166, EPI_ISL_820168, EPI_ISL_820170, EPI_ISL_820173 to 820174, EPI_ISL_820179, EPI_ISL_820181, EPI_ISL_820185, EPI_ISL_820187, EPI_ISL_820189,
EPI_ISL_820192, EPI_ISL_820195, EPI_ISL_820198, EPI_ISL_820201, EPI_ISL_820203, EPI_ISL_820205, EPI_ISL_820207 to 820208, EPI_ISL_820210, EPI_ISL_820214, EPI_ISL_820217, EPI_ISL_820219, EPI_ISL_820222, EPI_ISL_820228, EPI_ISL_820230, EPI_ISL_820232, EPI_ISL_820235, EPI_ISL_820237,
EPI_ISL_820239 to 820240, EPI_ISL_820245, EPI_ISL_820247 to 820248, EPI_ISL_820250, EPI_ISL_820252, EPI_ISL_820254 to 820255, EPI_ISL_820257 to 820258, EPI_ISL_820263, EPI_ISL_820265, EPI_ISL_820274 to 820275, EPI_ISL_820277 to 820278, EPI_ISL_820281, EPI_ISL_820285 to 820286, EPI_ISL_820288,
EPI_ISL_820290 to 820291, EPI_ISL_820293, EPI_ISL_820296, EPI_ISL_820298 to 820299, EPI_ISL_820301, EPI_ISL_820305, EPI_ISL_820307 to 820308, EPI_ISL_820310, EPI_ISL_820313, EPI_ISL_820315, EPI_ISL_820317, EPI_ISL_820322, EPI_ISL_820324 to 820325, EPI_ISL_820329 to 820330, EPI_ISL_820332,
EPI_ISL_820334, EPI_ISL_820336, EPI_ISL_820338 to 820339, EPI_ISL_820341 to 820342, EPI_ISL_820344, EPI_ISL_820346, EPI_ISL_820351 to 820352, EPI_ISL_820354, EPI_ISL_820357, EPI_ISL_820359, EPI_ISL_820361 to 820362, EPI_ISL_820364, EPI_ISL_820366 to 820367, EPI_ISL_820369 to 820370,
EPI_ISL_820374 to 820375, EPI_ISL_820377, EPI_ISL_820379 to 820380, EPI_ISL_820382, EPI_ISL_820389 to 820390, EPI_ISL_820392 to 820393, EPI_ISL_820398, EPI_ISL_820400 to 820401, EPI_ISL_820403, EPI_ISL_820405 to 820406, EPI_ISL_820408, EPI_ISL_820411, EPI_ISL_820413 to 820414, EPI_ISL_820416,
EPI_ISL_820419, EPI_ISL_820422, EPI_ISL_820424 to 820425, EPI_ISL_820427, EPI_ISL_820429 to 820430, EPI_ISL_820432, EPI_ISL_820434 to 820435, EPI_ISL_820438, EPI_ISL_820440, EPI_ISL_820442 to 820443, EPI_ISL_820447, EPI_ISL_820450, EPI_ISL_820452 to 820453, EPI_ISL_820455, EPI_ISL_820457 to
820458, EPI_ISL_820460, EPI_ISL_820463, EPI_ISL_820466, EPI_ISL_820469, EPI_ISL_820471, EPI_ISL_820473 to 820474, EPI_ISL_820476, EPI_ISL_820482, EPI_ISL_820486 to 820487, EPI_ISL_820491 to 820492, EPI_ISL_820494 to 820495, EPI_ISL_820497, EPI_ISL_820499 to 820500, EPI_ISL_820502 to 820503,
EPI_ISL_820508, EPI_ISL_820515, EPI_ISL_820517, EPI_ISL_820520, EPI_ISL_820523, EPI_ISL_820525, EPI_ISL_820527, EPI_ISL_820529 to 820530, EPI_ISL_820532, EPI_ISL_820534 to 820535, EPI_ISL_820537 to 820538, EPI_ISL_820540, EPI_ISL_820542, EPI_ISL_820544 to 820545, EPI_ISL_820550 to 820551,
EPI_ISL_820553, EPI_ISL_820555 to 820556, EPI_ISL_820559, EPI_ISL_820561, EPI_ISL_820564, EPI_ISL_820566 to 820567, EPI_ISL_820569, EPI_ISL_820571 to 820572, EPI_ISL_820574, EPI_ISL_820576, EPI_ISL_820579, EPI_ISL_820581 to 820582, EPI_ISL_820584 to 820585, EPI_ISL_820587, EPI_ISL_820592,
EPI_ISL_820594 to 820595, EPI_ISL_820599 to 820600, EPI_ISL_820602, EPI_ISL_820604, EPI_ISL_820606, EPI_ISL_820608, EPI_ISL_820611 to 820612, EPI_ISL_820614 to 820616, EPI_ISL_820619, EPI_ISL_820622 to 820627, EPI_ISL_820629 to 820631, EPI_ISL_820635 to 820636, EPI_ISL_820638, EPI_ISL_820640,
EPI_ISL_820642 to 820644, EPI_ISL_820646, EPI_ISL_821039, EPI_ISL_821128, EPI_ISL_821140, EPI_ISL_821163, EPI_ISL_821165, EPI_ISL_821172, EPI_ISL_821197, EPI_ISL_821263, EPI_ISL_821270 to 821271, EPI_ISL_821318, EPI_ISL_821456, EPI_ISL_821525, EPI_ISL_821617, EPI_ISL_821626 to 821628,
EPI_ISL_821971, EPI_ISL_821977 to 821982, EPI_ISL_821984, EPI_ISL_821986, EPI_ISL_821990, EPI_ISL_821998 to 822000, EPI_ISL_822002, EPI_ISL_822007 to 822010, EPI_ISL_822012, EPI_ISL_822014 to 822018, EPI_ISL_822020 to 822021, EPI_ISL_822024 to 822025, EPI_ISL_822027 to 822028, EPI_ISL_822031 to
822032, EPI_ISL_822037 to 822039, EPI_ISL_822045 to 822046, EPI_ISL_822050, EPI_ISL_822054 to 822058, EPI_ISL_822061 to 822062, EPI_ISL_822068, EPI_ISL_822070 to 822071, EPI_ISL_822073 to 822074, EPI_ISL_822076 to 822077, EPI_ISL_822082, EPI_ISL_822084 to 822085, EPI_ISL_822088 to 822090,
EPI_ISL_822092 to 822093, EPI_ISL_822095 to 822096, EPI_ISL_822102 to 822103, EPI_ISL_822106, EPI_ISL_822108 to 822112, EPI_ISL_822114 to 822116, EPI_ISL_822118, EPI_ISL_822120, EPI_ISL_822123 to 822126, EPI_ISL_822128 to 822129, EPI_ISL_822131, EPI_ISL_822136 to 822138, EPI_ISL_822143 to 822147,
EPI_ISL_822149, EPI_ISL_822151 to 822152, EPI_ISL_822154 to 822155, EPI_ISL_822157, EPI_ISL_822161 to 822164, EPI_ISL_822166 to 822168, EPI_ISL_822171 to 822172, EPI_ISL_822174 to 822175, EPI_ISL_822177, EPI_ISL_822180 to 822181, EPI_ISL_822185 to 822190, EPI_ISL_822192, EPI_ISL_822194,
EPI_ISL_822196, EPI_ISL_822198, EPI_ISL_822200, EPI_ISL_822203 to 822208, EPI_ISL_822210 to 822213, EPI_ISL_822215 to 822216, EPI_ISL_822218 to 822220, EPI_ISL_822222, EPI_ISL_822226 to 822229, EPI_ISL_822231, EPI_ISL_822233 to 822236, EPI_ISL_822238 to 822239, EPI_ISL_822241, EPI_ISL_822246,
EPI_ISL_822251, EPI_ISL_822254, EPI_ISL_822257 to 822259, EPI_ISL_822261 to 822263, EPI_ISL_822266, EPI_ISL_822269 to 822271, EPI_ISL_822277 to 822280, EPI_ISL_822284 to 822287, EPI_ISL_822289 to 822291, EPI_ISL_822294, EPI_ISL_822297 to 822298, EPI_ISL_822300, EPI_ISL_822303, EPI_ISL_822308,
EPI_ISL_822317, EPI_ISL_826464 to 826465, EPI_ISL_826472, EPI_ISL_826476, EPI_ISL_826480 to 826507, EPI_ISL_826509 to 826515, EPI_ISL_826517, EPI_ISL_826519, EPI_ISL_834146, EPI_ISL_834211 to 834212, EPI_ISL_834223, EPI_ISL_834226, EPI_ISL_834228, EPI_ISL_834231, EPI_ISL_834233,
EPI_ISL_834236 to 834237, EPI_ISL_834248, EPI_ISL_834256, EPI_ISL_834264, EPI_ISL_834271, EPI_ISL_834280, EPI_ISL_834283, EPI_ISL_834288, EPI_ISL_834290, EPI_ISL_834293, EPI_ISL_834298 to 834299, EPI_ISL_834311 to 834313, EPI_ISL_834343 to 834344, EPI_ISL_834352 to 834353, EPI_ISL_834360,
EPI_ISL_834373, EPI_ISL_834376 to 834378, EPI_ISL_834380 to 834382, EPI_ISL_834388, EPI_ISL_834404, EPI_ISL_834407, EPI_ISL_834409, EPI_ISL_834417, EPI_ISL_834423, EPI_ISL_834427, EPI_ISL_834440, EPI_ISL_834443, EPI_ISL_834445, EPI_ISL_834448 to 834449, EPI_ISL_834459, EPI_ISL_834466 to
834468, EPI_ISL_834485 to 834486, EPI_ISL_834488, EPI_ISL_834494, EPI_ISL_834497, EPI_ISL_834499 to 834500, EPI_ISL_834503, EPI_ISL_834511, EPI_ISL_834517, EPI_ISL_834529, EPI_ISL_834541, EPI_ISL_834545, EPI_ISL_834804, EPI_ISL_834814, EPI_ISL_834823, EPI_ISL_834826, EPI_ISL_834833 to 834834,
EPI_ISL_834837, EPI_ISL_834848, EPI_ISL_834865, EPI_ISL_834870, EPI_ISL_834874, EPI_ISL_834903, EPI_ISL_834909 to 834910, EPI_ISL_834920, EPI_ISL_834925, EPI_ISL_834931, EPI_ISL_834933, EPI_ISL_834949, EPI_ISL_834959, EPI_ISL_834972, EPI_ISL_834979, EPI_ISL_834981, EPI_ISL_834983,
EPI_ISL_834989, EPI_ISL_834991, EPI_ISL_835001, EPI_ISL_835004, EPI_ISL_835008, EPI_ISL_835017, EPI_ISL_835038, EPI_ISL_835047, EPI_ISL_835055, EPI_ISL_835058, EPI_ISL_835060, EPI_ISL_835067, EPI_ISL_835114 to 835115, EPI_ISL_835127, EPI_ISL_835130, EPI_ISL_835132, EPI_ISL_835140,
EPI_ISL_835474, EPI_ISL_835483, EPI_ISL_835492, EPI_ISL_835495, EPI_ISL_835532, EPI_ISL_835546, EPI_ISL_835550, EPI_ISL_835562, EPI_ISL_835615, EPI_ISL_835639, EPI_ISL_835655, EPI_ISL_835680, EPI_ISL_835698, EPI_ISL_846526, EPI_ISL_846536, EPI_ISL_851989, EPI_ISL_852074, EPI_ISL_852083,
EPI_ISL_852191, EPI_ISL_857891, EPI_ISL_858408 to 858409, EPI_ISL_858411, EPI_ISL_858413 to 858414, EPI_ISL_858418, EPI_ISL_858422, EPI_ISL_858573, EPI_ISL_858575, EPI_ISL_858578 to 858579, EPI_ISL_858582, EPI_ISL_858644, EPI_ISL_858648 to 858649, EPI_ISL_858653, EPI_ISL_858655,
EPI_ISL_858663, EPI_ISL_858666 to 858667, EPI_ISL_858669 to 858710, EPI_ISL_858713 to 858714, EPI_ISL_858719, EPI_ISL_858725, EPI_ISL_858732, EPI_ISL_858737, EPI_ISL_858739, EPI_ISL_858748, EPI_ISL_858752 to 858753, EPI_ISL_858779 to 858780, EPI_ISL_858785 to 858790, EPI_ISL_858809,
EPI_ISL_858811, EPI_ISL_858813 to 858814, EPI_ISL_858816 to 858827, EPI_ISL_858829, EPI_ISL_858831 to 858841, EPI_ISL_858845 to 858858, EPI_ISL_858861 to 858862, EPI_ISL_858864, EPI_ISL_858866, EPI_ISL_858868 to 858873, EPI_ISL_858875 to 858899, EPI_ISL_858902, EPI_ISL_858905, EPI_ISL_858907,
EPI_ISL_858909, EPI_ISL_858913 to 858915, EPI_ISL_858917 to 858918, EPI_ISL_858923 to 858926, EPI_ISL_858929, EPI_ISL_858931 to 858934, EPI_ISL_858938 to 858942, EPI_ISL_858944, EPI_ISL_858946 to 858952, EPI_ISL_858954 to 858957, EPI_ISL_858959, EPI_ISL_858961 to 858968, EPI_ISL_858971,
EPI_ISL_858973, EPI_ISL_858976 to 858980, EPI_ISL_858982 to 858984, EPI_ISL_858986 to 858989, EPI_ISL_858991 to 858993, EPI_ISL_858995 to 858996, EPI_ISL_858998 to 859002, EPI_ISL_859004 to 859011, EPI_ISL_859013 to 859029, EPI_ISL_859043 to 859044, EPI_ISL_859049, EPI_ISL_859051, EPI_ISL_859056,
EPI_ISL_859061, EPI_ISL_859063, EPI_ISL_859066 to 859067, EPI_ISL_859075 to 859076, EPI_ISL_859082, EPI_ISL_859085 to 859086, EPI_ISL_859089 to 859091, EPI_ISL_859095 to 859097, EPI_ISL_859099, EPI_ISL_859101, EPI_ISL_859103, EPI_ISL_859107, EPI_ISL_859110 to 859111, EPI_ISL_859113,
EPI_ISL_859115, EPI_ISL_859117, EPI_ISL_859121 to 859122, EPI_ISL_859124, EPI_ISL_859126, EPI_ISL_859128 to 859129, EPI_ISL_859132, EPI_ISL_859135, EPI_ISL_859138, EPI_ISL_859141 to 859148, EPI_ISL_859154, EPI_ISL_859161, EPI_ISL_859165, EPI_ISL_859181, EPI_ISL_859186, EPI_ISL_859194,
EPI_ISL_859198, EPI_ISL_859202 to 859204, EPI_ISL_859206, EPI_ISL_859208 to 859210, EPI_ISL_859212 to 859213, EPI_ISL_859215 to 859217, EPI_ISL_859220 to 859222, EPI_ISL_859224 to 859228, EPI_ISL_859230 to 859231, EPI_ISL_859233, EPI_ISL_859235, EPI_ISL_859237 to 859244, EPI_ISL_859246 to 859252,
EPI_ISL_859259 to 859261, EPI_ISL_859264, EPI_ISL_859266 to 859267, EPI_ISL_859269, EPI_ISL_859273 to 859275, EPI_ISL_859277 to 859279, EPI_ISL_859281, EPI_ISL_859283 to 859284, EPI_ISL_859286 to 859287, EPI_ISL_859289 to 859290, EPI_ISL_859292 to 859293, EPI_ISL_859296, EPI_ISL_859298 to 859299,
EPI_ISL_859302, EPI_ISL_859305, EPI_ISL_859307 to 859308, EPI_ISL_859310, EPI_ISL_859314 to 859315, EPI_ISL_859328 to 859344, EPI_ISL_859346 to 859355, EPI_ISL_859357 to 859391, EPI_ISL_859393 to 859394, EPI_ISL_859396 to 859397, EPI_ISL_859399 to 859406, EPI_ISL_859408 to 859411, EPI_ISL_859414
to 859415, EPI_ISL_859417 to 859419, EPI_ISL_859421, EPI_ISL_859423 to 859425, EPI_ISL_859427 to 859430, EPI_ISL_859432 to 859434, EPI_ISL_859437 to 859438, EPI_ISL_859440 to 859451, EPI_ISL_859453 to 859455, EPI_ISL_859534 to 859539

| | | | |
|---|---|---|---|
| see above | Lighthouse Lab in Alderley Park | Wellcome Sanger Institute for the COVID-19 Genomics UK (COG-UK) Consortium | Cordelia Langford; David K. Jackson; Dominic Kwiatkowski; Ewan Harrison; Ian Johnston; Jacquelyn Wynn; John Sillitoe on behalf of the Wellcome Sanger Institute COVID-19 Surveillance Team; John Sillitoe on behalf of the Wellcome Sanger Institute COVID-19 Surveillance Team (http://www.sanger.ac.uk/covid-team); Mairead Hyland; Roberto Amato; Sonia Goncalves; The Lighthouse Lab in Alderley Park and Alex Alderton |

EPI_ISL_551285 to 551293, EPI_ISL_551295 to 551297, EPI_ISL_551299 to 551326, EPI_ISL_551328 to 551390, EPI_ISL_551392 to 551401, EPI_ISL_551403 to 551411, EPI_ISL_551413 to 551414, EPI_ISL_551416 to 551433, EPI_ISL_551435 to 551452, EPI_ISL_551454 to 551482, EPI_ISL_551485 to 551486,
EPI_ISL_551488 to 551495, EPI_ISL_551497 to 551502, EPI_ISL_551504 to 551518, EPI_ISL_551521, EPI_ISL_551523, EPI_ISL_551525 to 551529, EPI_ISL_551531 to 551576, EPI_ISL_551735 to 551736, EPI_ISL_551740, EPI_ISL_551742 to 551743, EPI_ISL_551749, EPI_ISL_551751, EPI_ISL_551755 to 551756,
EPI_ISL_551759 to 551764, EPI_ISL_551769, EPI_ISL_551772, EPI_ISL_551776, EPI_ISL_551779 to 551780, EPI_ISL_551784 to 551785, EPI_ISL_551789, EPI_ISL_551795 to 551796, EPI_ISL_551798 to 551799, EPI_ISL_551801, EPI_ISL_551804, EPI_ISL_551806 to 551808, EPI_ISL_551810 to 551811, EPI_ISL_551815,
EPI_ISL_551818 to 551819, EPI_ISL_551823 to 551827, EPI_ISL_551832, EPI_ISL_551834, EPI_ISL_552446, EPI_ISL_552449, EPI_ISL_552463 to 552464, EPI_ISL_552466, EPI_ISL_552468, EPI_ISL_552473, EPI_ISL_552476, EPI_ISL_552479, EPI_ISL_552481, EPI_ISL_552486, EPI_ISL_552488, EPI_ISL_552506 to
552508, EPI_ISL_552512, EPI_ISL_552515 to 552516, EPI_ISL_552523, EPI_ISL_552526 to 552527, EPI_ISL_552530, EPI_ISL_552533 to 552534, EPI_ISL_552539 to 552540, EPI_ISL_552547, EPI_ISL_552556, EPI_ISL_552562, EPI_ISL_552565 to 552566, EPI_ISL_552574, EPI_ISL_552576 to 552577, EPI_ISL_552579 to
552580, EPI_ISL_552587, EPI_ISL_552590 to 552591, EPI_ISL_552595 to 552596, EPI_ISL_552598, EPI_ISL_552607 to 552608, EPI_ISL_552614, EPI_ISL_552617 to 552618, EPI_ISL_552620, EPI_ISL_552622, EPI_ISL_552629, EPI_ISL_552639, EPI_ISL_552642, EPI_ISL_552644, EPI_ISL_552646, EPI_ISL_552655,
EPI_ISL_552659, EPI_ISL_552665 to 552666, EPI_ISL_552669, EPI_ISL_552671, EPI_ISL_552679, EPI_ISL_552681, EPI_ISL_552686, EPI_ISL_552689, EPI_ISL_552699, EPI_ISL_552703, EPI_ISL_552706 to 552707, EPI_ISL_552712 to 552713, EPI_ISL_552717, EPI_ISL_552719 to 552724, EPI_ISL_552726,
EPI_ISL_552728 to 552732, EPI_ISL_552735, EPI_ISL_552741 to 552742, EPI_ISL_552745 to 552746, EPI_ISL_552748 to 552750, EPI_ISL_552760 to 552761, EPI_ISL_552768, EPI_ISL_552770 to 552771, EPI_ISL_552773 to 552777, EPI_ISL_552779 to 552780, EPI_ISL_552782 to 552783, EPI_ISL_552785, EPI_ISL_552792
to 552794, EPI_ISL_552796 to 552798, EPI_ISL_552800 to 552801, EPI_ISL_552803 to 552808, EPI_ISL_552812 to 552813, EPI_ISL_552815 to 552817, EPI_ISL_552819, EPI_ISL_552821 to 552822, EPI_ISL_552824 to 552840, EPI_ISL_552843 to 552851, EPI_ISL_552853 to 552859, EPI_ISL_552861 to 552866,
EPI_ISL_552868 to 552873, EPI_ISL_552876 to 552878, EPI_ISL_552881 to 552904, EPI_ISL_552906 to 552909, EPI_ISL_552911 to 552915, EPI_ISL_552917 to 552923, EPI_ISL_552925 to 552935, EPI_ISL_552937, EPI_ISL_552941 to 552942, EPI_ISL_552944 to 552946, EPI_ISL_552951 to 552952, EPI_ISL_552954 to
552960, EPI_ISL_552962, EPI_ISL_552964 to 552966, EPI_ISL_552968 to 552971, EPI_ISL_552974, EPI_ISL_552976 to 552978, EPI_ISL_552980 to 552986, EPI_ISL_552988 to 552989, EPI_ISL_552991 to 552994, EPI_ISL_552996, EPI_ISL_552998 to 553001, EPI_ISL_553003 to 553006, EPI_ISL_553269, EPI_ISL_553271,
EPI_ISL_553273 to 553274, EPI_ISL_553277 to 553284, EPI_ISL_553286 to 553292, EPI_ISL_553294, EPI_ISL_553296 to 553303, EPI_ISL_553305 to 553322, EPI_ISL_553324 to 553326, EPI_ISL_553328 to 553332, EPI_ISL_553334 to 553370, EPI_ISL_553372 to 553374, EPI_ISL_553376 to 553417, EPI_ISL_553419 to
553420, EPI_ISL_553422 to 553453, EPI_ISL_553455 to 553464, EPI_ISL_553466 to 553467, EPI_ISL_553469 to 553480, EPI_ISL_553482 to 553566, EPI_ISL_553569 to 553596, EPI_ISL_553598 to 553644, EPI_ISL_553646 to 553663, EPI_ISL_553665 to 553666, EPI_ISL_553668, EPI_ISL_553670 to 553707, EPI_ISL_553709
to 553731, EPI_ISL_553733 to 553742, EPI_ISL_553744 to 553777, EPI_ISL_553779 to 553783, EPI_ISL_553785 to 553811, EPI_ISL_553813 to 553838, EPI_ISL_553840 to 553845, EPI_ISL_553847 to 553865, EPI_ISL_553867 to 553870, EPI_ISL_553872 to 553878, EPI_ISL_553881, EPI_ISL_553883 to 553884,
EPI_ISL_553886 to 553888, EPI_ISL_553890 to 553892, EPI_ISL_553895, EPI_ISL_553899 to 553904, EPI_ISL_553906, EPI_ISL_553908 to 553928, EPI_ISL_553930 to 553933, EPI_ISL_553935 to 553938, EPI_ISL_553940 to 553945, EPI_ISL_553947 to 553951, EPI_ISL_553953, EPI_ISL_553958 to 553964, EPI_ISL_553966
to 553970, EPI_ISL_553973 to 553982, EPI_ISL_553984, EPI_ISL_553986, EPI_ISL_553988, EPI_ISL_553990 to 553992, EPI_ISL_553995 to 554001, EPI_ISL_554004, EPI_ISL_554006 to 554008, EPI_ISL_554010 to 554014, EPI_ISL_554016 to 554023, EPI_ISL_554027, EPI_ISL_554029 to 554041, EPI_ISL_554044 to 554047,
EPI_ISL_554049 to 554053, EPI_ISL_554055 to 554056, EPI_ISL_554059 to 554064, EPI_ISL_554066 to 554079, EPI_ISL_554081 to 554087, EPI_ISL_554089 to 554096, EPI_ISL_554098, EPI_ISL_554102, EPI_ISL_554104 to 554106, EPI_ISL_554108 to 554117, EPI_ISL_554120, EPI_ISL_554122, EPI_ISL_554126,

EPI_ISL_554129 to 554131, EPI_ISL_554136, EPI_ISL_554140, EPI_ISL_554151, EPI_ISL_554155 to 554158, EPI_ISL_554164 to 554165, EPI_ISL_554170 to 554172, EPI_ISL_554174 to 554176, EPI_ISL_554183, EPI_ISL_554186 to 554190, EPI_ISL_554192 to 554194, EPI_ISL_554196 to 554199, EPI_ISL_554202 to 554203,
EPI_ISL_554206 to 554207, EPI_ISL_554209, EPI_ISL_554212, EPI_ISL_554215, EPI_ISL_554219, EPI_ISL_554225, EPI_ISL_554227, EPI_ISL_554229 to 554230, EPI_ISL_554233, EPI_ISL_554235, EPI_ISL_554238, EPI_ISL_554241, EPI_ISL_554252, EPI_ISL_554257, EPI_ISL_554260, EPI_ISL_554263, EPI_ISL_554265 to
554266, EPI_ISL_554268, EPI_ISL_554270, EPI_ISL_554275, EPI_ISL_554278 to 554280, EPI_ISL_554282 to 554283, EPI_ISL_554288, EPI_ISL_554292, EPI_ISL_554296, EPI_ISL_554298 to 554299, EPI_ISL_554301, EPI_ISL_554305, EPI_ISL_554309, EPI_ISL_554318, EPI_ISL_554320 to 554324, EPI_ISL_554326 to
554339, EPI_ISL_554341 to 554381, EPI_ISL_554383 to 554385, EPI_ISL_554387 to 554477, EPI_ISL_554479 to 554483, EPI_ISL_554485 to 554496, EPI_ISL_554498 to 554536, EPI_ISL_554538 to 554600, EPI_ISL_554602 to 554603, EPI_ISL_554605 to 554618, EPI_ISL_554623 to 554626, EPI_ISL_554630 to 554631,
EPI_ISL_554640, EPI_ISL_554642, EPI_ISL_554646, EPI_ISL_554651, EPI_ISL_554653 to 554654, EPI_ISL_554656 to 554657, EPI_ISL_554661, EPI_ISL_554664, EPI_ISL_554666, EPI_ISL_554677, EPI_ISL_554680 to 554681, EPI_ISL_554686, EPI_ISL_554688 to 554689, EPI_ISL_554692, EPI_ISL_554694,
EPI_ISL_554696 to 554697, EPI_ISL_554703, EPI_ISL_554708, EPI_ISL_554710, EPI_ISL_554716, EPI_ISL_554725 to 554727, EPI_ISL_554730, EPI_ISL_554732, EPI_ISL_554735, EPI_ISL_554741, EPI_ISL_554754 to 554757, EPI_ISL_554759, EPI_ISL_554761, EPI_ISL_554767, EPI_ISL_554771 to 554772,
EPI_ISL_554776 to 554777, EPI_ISL_554780, EPI_ISL_554782, EPI_ISL_554792, EPI_ISL_554794 to 554795, EPI_ISL_554797, EPI_ISL_554799, EPI_ISL_554802, EPI_ISL_554804, EPI_ISL_554807 to 554813, EPI_ISL_554815 to 554816, EPI_ISL_554820, EPI_ISL_554827, EPI_ISL_554830, EPI_ISL_554832 to 554834,
EPI_ISL_554836 to 554837, EPI_ISL_554841, EPI_ISL_554846 to 554848, EPI_ISL_554850, EPI_ISL_554853, EPI_ISL_554861 to 554867, EPI_ISL_554869 to 554905, EPI_ISL_554907 to 554915, EPI_ISL_554917 to 554928, EPI_ISL_554930 to 554957, EPI_ISL_554959 to 554983, EPI_ISL_554985 to 555018, EPI_ISL_555020
to 555021, EPI_ISL_555023 to 555040, EPI_ISL_555042 to 555083, EPI_ISL_555085 to 555086, EPI_ISL_555088 to 555090, EPI_ISL_555092 to 555109, EPI_ISL_555111 to 555117, EPI_ISL_555119 to 555126, EPI_ISL_555131 to 555147, EPI_ISL_555149 to 555188, EPI_ISL_555191 to 555219, EPI_ISL_555221 to 555246,
EPI_ISL_555248 to 555258, EPI_ISL_555260 to 555275, EPI_ISL_555277 to 555302, EPI_ISL_555305 to 555313, EPI_ISL_555315 to 555359, EPI_ISL_555361 to 555376, EPI_ISL_555378 to 555390, EPI_ISL_555392 to 555397, EPI_ISL_555399 to 555431, EPI_ISL_555434 to 555447, EPI_ISL_555449 to 555458,
EPI_ISL_555460 to 555473, EPI_ISL_555475 to 555501, EPI_ISL_555503 to 555508, EPI_ISL_555512 to 555520, EPI_ISL_555522 to 555531, EPI_ISL_555533 to 555557, EPI_ISL_555559 to 555562, EPI_ISL_555564 to 555567, EPI_ISL_555569 to 555570, EPI_ISL_555572 to 555577, EPI_ISL_555579 to 555597,
EPI_ISL_555599 to 555607, EPI_ISL_555609 to 555615, EPI_ISL_555617 to 555630, EPI_ISL_555632 to 555639, EPI_ISL_555643 to 555649, EPI_ISL_555653 to 555659, EPI_ISL_555661 to 555679, EPI_ISL_555681 to 555718, EPI_ISL_555720 to 555745, EPI_ISL_555747 to 555793, EPI_ISL_555795 to 555805,
EPI_ISL_555809 to 555812, EPI_ISL_555814, EPI_ISL_555816 to 555827, EPI_ISL_555829 to 555842, EPI_ISL_555845 to 555863, EPI_ISL_555865 to 555867, EPI_ISL_555869 to 555874, EPI_ISL_555876, EPI_ISL_555878 to 555886, EPI_ISL_555888 to 555905, EPI_ISL_555907 to 555909, EPI_ISL_555911 to 555938,
EPI_ISL_555940 to 555944, EPI_ISL_555946, EPI_ISL_555949, EPI_ISL_555951 to 555954, EPI_ISL_555956 to 555972, EPI_ISL_555974 to 555975, EPI_ISL_555977 to 555984, EPI_ISL_555987 to 555988, EPI_ISL_555991 to 555992, EPI_ISL_555994 to 555996, EPI_ISL_555998 to 555999, EPI_ISL_556001 to 556006,
EPI_ISL_556008 to 556011, EPI_ISL_556013 to 556020, EPI_ISL_556023 to 556025, EPI_ISL_556027 to 556028, EPI_ISL_556031 to 556038, EPI_ISL_556040, EPI_ISL_556042 to 556046, EPI_ISL_556048 to 556057, EPI_ISL_556059 to 556062, EPI_ISL_556064 to 556065, EPI_ISL_556067 to 556068, EPI_ISL_556070 to
556074, EPI_ISL_556078, EPI_ISL_556080 to 556082, EPI_ISL_556084 to 556086, EPI_ISL_556088 to 556089, EPI_ISL_556096, EPI_ISL_556099 to 556101, EPI_ISL_556103, EPI_ISL_556105 to 556108, EPI_ISL_556110 to 556111, EPI_ISL_556113 to 556114, EPI_ISL_556117 to 556122, EPI_ISL_556124 to 556125,
EPI_ISL_556127 to 556132, EPI_ISL_556135 to 556137, EPI_ISL_556142 to 556145, EPI_ISL_556147, EPI_ISL_556149, EPI_ISL_556151 to 556157, EPI_ISL_556159 to 556162, EPI_ISL_556167, EPI_ISL_556169, EPI_ISL_556171, EPI_ISL_556174 to 556175, EPI_ISL_556177 to 556181, EPI_ISL_556184 to 556185,
EPI_ISL_556188 to 556189, EPI_ISL_556191 to 556194, EPI_ISL_556196, EPI_ISL_556200, EPI_ISL_556203, EPI_ISL_556205 to 556206, EPI_ISL_556209 to 556210, EPI_ISL_556212, EPI_ISL_556215 to 556217, EPI_ISL_556219 to 556222, EPI_ISL_556224 to 556231, EPI_ISL_556234, EPI_ISL_556236, EPI_ISL_556238 to
556241, EPI_ISL_556243, EPI_ISL_556245 to 556247, EPI_ISL_556249 to 556250, EPI_ISL_556252, EPI_ISL_556254 to 556258, EPI_ISL_556260, EPI_ISL_556262, EPI_ISL_556264 to 556265, EPI_ISL_556268, EPI_ISL_556270, EPI_ISL_556272 to 556274, EPI_ISL_556277 to 556281, EPI_ISL_556283 to 556284,
EPI_ISL_556286, EPI_ISL_556288, EPI_ISL_556290, EPI_ISL_556292, EPI_ISL_556295 to 556297, EPI_ISL_556299, EPI_ISL_556301 to 556303, EPI_ISL_556306 to 556308, EPI_ISL_556310 to 556315, EPI_ISL_556320, EPI_ISL_556322 to 556323, EPI_ISL_556325 to 556337, EPI_ISL_556339 to 556343, EPI_ISL_556345 to
556347, EPI_ISL_556352 to 556356, EPI_ISL_556358 to 556410, EPI_ISL_556412, EPI_ISL_556414 to 556420, EPI_ISL_556422 to 556424, EPI_ISL_556426 to 556427, EPI_ISL_556429 to 556431, EPI_ISL_556436 to 556445, EPI_ISL_556447 to 556458, EPI_ISL_556460 to 556470, EPI_ISL_556472 to 556481, EPI_ISL_556483
to 556487, EPI_ISL_556489 to 556493, EPI_ISL_556495 to 556499, EPI_ISL_556501 to 556504, EPI_ISL_556506 to 556507, EPI_ISL_556512 to 556515, EPI_ISL_556576 to 556638, EPI_ISL_556985 to 557001, EPI_ISL_557003 to 557006, EPI_ISL_557008 to 557031, EPI_ISL_557033 to 557042, EPI_ISL_557044 to 557073,
EPI_ISL_557075 to 557109, EPI_ISL_557111 to 557118, EPI_ISL_557120 to 557128, EPI_ISL_557130 to 557141, EPI_ISL_557143 to 557155, EPI_ISL_557157 to 557165, EPI_ISL_557351, EPI_ISL_557354, EPI_ISL_557356, EPI_ISL_557359, EPI_ISL_557361, EPI_ISL_557366, EPI_ISL_557371, EPI_ISL_557373 to 557374,
EPI_ISL_557378, EPI_ISL_557380 to 557381, EPI_ISL_557383, EPI_ISL_557388, EPI_ISL_557390, EPI_ISL_557398 to 557399, EPI_ISL_557404, EPI_ISL_557406, EPI_ISL_557408, EPI_ISL_557411 to 557412, EPI_ISL_557414 to 557415, EPI_ISL_557417, EPI_ISL_557419, EPI_ISL_557423, EPI_ISL_557425,
EPI_ISL_557428, EPI_ISL_557430, EPI_ISL_557433, EPI_ISL_557436, EPI_ISL_557439 to 557440, EPI_ISL_557446 to 557447, EPI_ISL_557449, EPI_ISL_557452, EPI_ISL_557454, EPI_ISL_557466, EPI_ISL_557468 to 557469, EPI_ISL_557471, EPI_ISL_557474, EPI_ISL_557476, EPI_ISL_557482, EPI_ISL_557484 to
557485, EPI_ISL_557487 to 557488, EPI_ISL_557491, EPI_ISL_557493, EPI_ISL_557497, EPI_ISL_557499 to 557500, EPI_ISL_557507, EPI_ISL_557511, EPI_ISL_557518, EPI_ISL_557522, EPI_ISL_557526 to 557528, EPI_ISL_557530 to 557531, EPI_ISL_557533, EPI_ISL_557537, EPI_ISL_557539, EPI_ISL_557541,
EPI_ISL_557543 to 557544, EPI_ISL_557551, EPI_ISL_557553 to 557557, EPI_ISL_557559 to 557624, EPI_ISL_557626 to 557629, EPI_ISL_557631 to 557646, EPI_ISL_557648 to 557659, EPI_ISL_557661 to 557663, EPI_ISL_557665 to 557700, EPI_ISL_557702 to 557706, EPI_ISL_557708 to 557710, EPI_ISL_557712 to
557715, EPI_ISL_557717 to 557727, EPI_ISL_557729, EPI_ISL_557731 to 557738, EPI_ISL_557743 to 557744, EPI_ISL_557746 to 557747, EPI_ISL_557749, EPI_ISL_557751 to 557752, EPI_ISL_557754 to 557756, EPI_ISL_557759, EPI_ISL_557761 to 557763, EPI_ISL_557767, EPI_ISL_557769 to 557770, EPI_ISL_557772 to
557774, EPI_ISL_557776 to 557777, EPI_ISL_557779 to 557780, EPI_ISL_557783, EPI_ISL_557785, EPI_ISL_557787, EPI_ISL_557790, EPI_ISL_557792, EPI_ISL_557794, EPI_ISL_557797 to 557799, EPI_ISL_557801, EPI_ISL_557803 to 557806, EPI_ISL_557809 to 557810, EPI_ISL_557813 to 557815, EPI_ISL_557818 to
557820, EPI_ISL_557825 to 557826, EPI_ISL_557831, EPI_ISL_557833 to 557837, EPI_ISL_557839, EPI_ISL_557841 to 557842, EPI_ISL_557844, EPI_ISL_557846, EPI_ISL_557850 to 557851, EPI_ISL_557855 to 557857, EPI_ISL_557859 to 557864, EPI_ISL_557872 to 557875, EPI_ISL_557881 to 557883, EPI_ISL_557886 to
557888, EPI_ISL_557891 to 557892, EPI_ISL_557897, EPI_ISL_557899 to 557904, EPI_ISL_557906, EPI_ISL_557910, EPI_ISL_557912 to 557914, EPI_ISL_557916, EPI_ISL_557918, EPI_ISL_557920, EPI_ISL_557922 to 557924, EPI_ISL_557928 to 557932, EPI_ISL_557934 to 557936, EPI_ISL_557939, EPI_ISL_557943,
EPI_ISL_557945 to 557946, EPI_ISL_557950 to 557951, EPI_ISL_557955, EPI_ISL_557960 to 557961, EPI_ISL_557966, EPI_ISL_557968 to 557969, EPI_ISL_557971, EPI_ISL_557973, EPI_ISL_557975, EPI_ISL_557978, EPI_ISL_557980 to 557983, EPI_ISL_557985 to 557988, EPI_ISL_557990 to 557998, EPI_ISL_558000,
EPI_ISL_558002 to 558020, EPI_ISL_558022 to 558023, EPI_ISL_558025, EPI_ISL_558027, EPI_ISL_558029, EPI_ISL_558032 to 558035, EPI_ISL_558038 to 558040, EPI_ISL_558042 to 558057, EPI_ISL_558059 to 558067, EPI_ISL_558069 to 558079, EPI_ISL_558081 to 558088, EPI_ISL_558090 to 558094, EPI_ISL_558097
to 558102, EPI_ISL_558104 to 558105, EPI_ISL_558107 to 558111, EPI_ISL_558114 to 558115, EPI_ISL_558117 to 558125, EPI_ISL_558128, EPI_ISL_558130 to 558140, EPI_ISL_558142, EPI_ISL_558144 to 558154, EPI_ISL_558156 to 558184, EPI_ISL_558186 to 558194, EPI_ISL_558196 to 558221, EPI_ISL_558223 to
558233, EPI_ISL_558235 to 558236, EPI_ISL_558238, EPI_ISL_558240, EPI_ISL_558244, EPI_ISL_558256, EPI_ISL_558259 to 558260, EPI_ISL_558263, EPI_ISL_558268 to 558270, EPI_ISL_558274, EPI_ISL_558276, EPI_ISL_558288 to 558289, EPI_ISL_558291, EPI_ISL_558307, EPI_ISL_558312, EPI_ISL_558316,
EPI_ISL_558321, EPI_ISL_558324, EPI_ISL_558329, EPI_ISL_558332, EPI_ISL_558335, EPI_ISL_558338, EPI_ISL_558343, EPI_ISL_558345, EPI_ISL_558348, EPI_ISL_558350, EPI_ISL_558354 to 558355, EPI_ISL_558364, EPI_ISL_558379, EPI_ISL_558389, EPI_ISL_558394 to 558396, EPI_ISL_558404 to 558421,
EPI_ISL_558423 to 558431, EPI_ISL_558433 to 558482, EPI_ISL_558484 to 558511, EPI_ISL_558513 to 558542, EPI_ISL_558544 to 558553, EPI_ISL_558555 to 558585, EPI_ISL_558588 to 558601, EPI_ISL_558603 to 558604, EPI_ISL_558606 to 558616, EPI_ISL_558618 to 558637, EPI_ISL_558639 to 558666,
EPI_ISL_558668 to 558730, EPI_ISL_558732 to 558791, EPI_ISL_558793 to 558795, EPI_ISL_558797 to 558811, EPI_ISL_558813 to 558845, EPI_ISL_558847 to 558871, EPI_ISL_558873 to 558876, EPI_ISL_558878 to 558886, EPI_ISL_558889 to 558902, EPI_ISL_558904 to 558913, EPI_ISL_558915 to 558919,
EPI_ISL_558922, EPI_ISL_558924, EPI_ISL_558927, EPI_ISL_558929 to 558931, EPI_ISL_558933, EPI_ISL_558936, EPI_ISL_558938, EPI_ISL_558940 to 558948, EPI_ISL_558952 to 558958, EPI_ISL_558960 to 558964, EPI_ISL_558966 to 558969, EPI_ISL_558971 to 558972, EPI_ISL_558974, EPI_ISL_558976 to 558977,
EPI_ISL_558979, EPI_ISL_558982 to 558986, EPI_ISL_558988 to 558989, EPI_ISL_558992, EPI_ISL_558995 to 558999, EPI_ISL_559001 to 559004, EPI_ISL_559006 to 559008, EPI_ISL_559010, EPI_ISL_559012 to 559016, EPI_ISL_559020 to 559021, EPI_ISL_559023, EPI_ISL_559025, EPI_ISL_559027, EPI_ISL_559029 to
559033, EPI_ISL_559035 to 559041, EPI_ISL_559044 to 559045, EPI_ISL_559048, EPI_ISL_559050 to 559056, EPI_ISL_559058 to 559060, EPI_ISL_559064 to 559070, EPI_ISL_559072 to 559079, EPI_ISL_559085 to 559087, EPI_ISL_559089 to 559099, EPI_ISL_559102, EPI_ISL_559105, EPI_ISL_559107 to 559116,
EPI_ISL_559118, EPI_ISL_559121 to 559124, EPI_ISL_559127 to 559128, EPI_ISL_559130, EPI_ISL_559134 to 559135, EPI_ISL_559137 to 559146, EPI_ISL_559148, EPI_ISL_559150 to 559153, EPI_ISL_559155 to 559156, EPI_ISL_559159 to 559162, EPI_ISL_559165, EPI_ISL_559169 to 559170, EPI_ISL_559172 to 559173,
EPI_ISL_559175 to 559180, EPI_ISL_559182 to 559186, EPI_ISL_559189 to 559191, EPI_ISL_559193, EPI_ISL_559196 to 559200, EPI_ISL_559202 to 559203, EPI_ISL_559205 to 559206, EPI_ISL_559208 to 559212, EPI_ISL_559214, EPI_ISL_559216 to 559217, EPI_ISL_559219 to 559222, EPI_ISL_559224 to 559227,
EPI_ISL_559229 to 559234, EPI_ISL_559237 to 559240, EPI_ISL_559242 to 559243, EPI_ISL_559245, EPI_ISL_559248 to 559252, EPI_ISL_559255, EPI_ISL_559257 to 559258, EPI_ISL_559260, EPI_ISL_559262, EPI_ISL_559265, EPI_ISL_559267 to 559274, EPI_ISL_559276, EPI_ISL_559279 to 559282,
EPI_ISL_559285 to 559286, EPI_ISL_559288 to 559289, EPI_ISL_559291 to 559295, EPI_ISL_559297 to 559299, EPI_ISL_559301 to 559311, EPI_ISL_559313, EPI_ISL_559315 to 559318, EPI_ISL_559321 to 559325, EPI_ISL_559327 to 559330, EPI_ISL_559332 to 559334, EPI_ISL_559336, EPI_ISL_559338 to 559349,
EPI_ISL_559351, EPI_ISL_559356 to 559367, EPI_ISL_559370 to 559390, EPI_ISL_559392 to 559405, EPI_ISL_559407 to 559434, EPI_ISL_559436 to 559446, EPI_ISL_559448 to 559469, EPI_ISL_559471 to 559480, EPI_ISL_559482 to 559489, EPI_ISL_559491 to 559493, EPI_ISL_559495 to 559503, EPI_ISL_559505 to
559521, EPI_ISL_559523, EPI_ISL_559525 to 559554, EPI_ISL_559556 to 559574, EPI_ISL_559577 to 559590, EPI_ISL_559592 to 559609, EPI_ISL_559611 to 559615, EPI_ISL_559617, EPI_ISL_559619 to 559620, EPI_ISL_559622 to 559627, EPI_ISL_559629 to 559643, EPI_ISL_559645 to 559651, EPI_ISL_559653 to
559661, EPI_ISL_566110 to 566164, EPI_ISL_566166, EPI_ISL_566168 to 566292, EPI_ISL_566294 to 566312, EPI_ISL_566314 to 566323, EPI_ISL_566325 to 566337, EPI_ISL_566339 to 566343, EPI_ISL_566345 to 566420, EPI_ISL_567533 to 567557, EPI_ISL_567559 to 567587, EPI_ISL_567589 to 567621, EPI_ISL_567623
to 567669, EPI_ISL_567671 to 567721, EPI_ISL_567723 to 567786, EPI_ISL_567788 to 567802, EPI_ISL_567804 to 567810, EPI_ISL_567813 to 567825, EPI_ISL_567827 to 567830, EPI_ISL_567832 to 567844, EPI_ISL_567846 to 567886, EPI_ISL_567888 to 567902, EPI_ISL_567904 to 568061, EPI_ISL_568063 to 568067,
EPI_ISL_568069 to 568095, EPI_ISL_568097 to 568131, EPI_ISL_568133 to 568188, EPI_ISL_568190 to 568222, EPI_ISL_568467, EPI_ISL_568474 to 568475, EPI_ISL_568477, EPI_ISL_575399 to 575452, EPI_ISL_575454 to 575514, EPI_ISL_575516 to 575535, EPI_ISL_575537 to 575597, EPI_ISL_575599 to 575658,
EPI_ISL_575660 to 575760, EPI_ISL_575982 to 575983, EPI_ISL_579919 to 579950, EPI_ISL_579952 to 579970, EPI_ISL_579972 to 580019, EPI_ISL_580021 to 580061, EPI_ISL_580063 to 580072, EPI_ISL_580074 to 580096, EPI_ISL_580098 to 580109, EPI_ISL_580111 to 580137, EPI_ISL_580139 to 580169,
EPI_ISL_580171 to 580186, EPI_ISL_580188 to 580197, EPI_ISL_580199 to 580209, EPI_ISL_580211 to 580212, EPI_ISL_580214 to 580281, EPI_ISL_580284, EPI_ISL_580288 to 580298, EPI_ISL_580302 to 580304, EPI_ISL_580306 to 580312, EPI_ISL_580314 to 580321, EPI_ISL_580323 to 580324, EPI_ISL_580326,
EPI_ISL_580328 to 580331, EPI_ISL_580334 to 580337, EPI_ISL_580339, EPI_ISL_580342 to 580352, EPI_ISL_580355 to 580361, EPI_ISL_580363, EPI_ISL_580365 to 580366, EPI_ISL_580370, EPI_ISL_580372, EPI_ISL_580374 to 580384, EPI_ISL_580386 to 580390, EPI_ISL_580392, EPI_ISL_580395, EPI_ISL_580397 to
580399, EPI_ISL_580401 to 580413, EPI_ISL_580415 to 580417, EPI_ISL_580419 to 580420, EPI_ISL_580422 to 580434, EPI_ISL_580436 to 580443, EPI_ISL_580445 to 580448, EPI_ISL_580450 to 580452, EPI_ISL_580454 to 580455, EPI_ISL_580457 to 580458, EPI_ISL_580460 to 580464, EPI_ISL_580466, EPI_ISL_580468
to 580470, EPI_ISL_580472 to 580477, EPI_ISL_580483 to 580484, EPI_ISL_580487 to 580492, EPI_ISL_580494, EPI_ISL_580496 to 580498, EPI_ISL_580500 to 580501, EPI_ISL_580503 to 580507, EPI_ISL_580510, EPI_ISL_580512 to 580514, EPI_ISL_580517 to 580531, EPI_ISL_580534 to 580535, EPI_ISL_580537 to
580549, EPI_ISL_580551 to 580556, EPI_ISL_580558, EPI_ISL_580560 to 580562, EPI_ISL_580566, EPI_ISL_580568, EPI_ISL_580570 to 580572, EPI_ISL_580574 to 580575, EPI_ISL_580578, EPI_ISL_580580 to 580581, EPI_ISL_580583 to 580584, EPI_ISL_580588 to 580592, EPI_ISL_580595 to 580602, EPI_ISL_580605 to
580606, EPI_ISL_580608 to 580609, EPI_ISL_580611, EPI_ISL_580613 to 580617, EPI_ISL_580619 to 580620, EPI_ISL_580623 to 580624, EPI_ISL_580627 to 580630, EPI_ISL_580633, EPI_ISL_580635 to 580636, EPI_ISL_580640, EPI_ISL_580645, EPI_ISL_580648, EPI_ISL_580650 to 580651, EPI_ISL_580655,
EPI_ISL_580660, EPI_ISL_580663, EPI_ISL_580674 to 580675, EPI_ISL_580677 to 580682, EPI_ISL_580689 to 580691, EPI_ISL_580695, EPI_ISL_580697, EPI_ISL_580699, EPI_ISL_580706, EPI_ISL_580710, EPI_ISL_580720, EPI_ISL_580722 to 580723, EPI_ISL_580730, EPI_ISL_580734, EPI_ISL_580737 to 580738,
EPI_ISL_580740, EPI_ISL_580754, EPI_ISL_580763 to 580764, EPI_ISL_580769, EPI_ISL_580771, EPI_ISL_580784, EPI_ISL_580787 to 580789, EPI_ISL_580791, EPI_ISL_580821 to 580823, EPI_ISL_580825, EPI_ISL_580828, EPI_ISL_580833, EPI_ISL_580835, EPI_ISL_580849, EPI_ISL_580851, EPI_ISL_580853,
EPI_ISL_581367, EPI_ISL_581378 to 581380, EPI_ISL_581391, EPI_ISL_581404, EPI_ISL_581409, EPI_ISL_581411 to 581414, EPI_ISL_581417, EPI_ISL_581424 to 581427, EPI_ISL_581429 to 581433, EPI_ISL_581435 to 581436, EPI_ISL_581442, EPI_ISL_581444 to 581446, EPI_ISL_581448, EPI_ISL_586572 to 586611,
EPI_ISL_586613 to 586616, EPI_ISL_586618 to 586620, EPI_ISL_586622, EPI_ISL_586624 to 586656, EPI_ISL_586658 to 586661, EPI_ISL_586663 to 586708, EPI_ISL_586710 to 586906, EPI_ISL_586908 to 586971, EPI_ISL_586973 to 586988, EPI_ISL_586990 to 587026, EPI_ISL_587028 to 587032, EPI_ISL_587034 to
587077, EPI_ISL_587079 to 587084, EPI_ISL_587086 to 587221, EPI_ISL_587223 to 587240, EPI_ISL_587243 to 587249, EPI_ISL_587251 to 587258, EPI_ISL_587260 to 587264, EPI_ISL_587266 to 587302, EPI_ISL_587711 to 587722, EPI_ISL_587724 to 587730, EPI_ISL_587732 to 587749, EPI_ISL_587751 to 587776,
EPI_ISL_587778 to 587807, EPI_ISL_587809 to 587811, EPI_ISL_587813 to 587853, EPI_ISL_587855 to 587872, EPI_ISL_587874 to 587917, EPI_ISL_587919 to 587934, EPI_ISL_587936 to 587976, EPI_ISL_587978 to 587980, EPI_ISL_587982 to 587987, EPI_ISL_587989 to 587991, EPI_ISL_587993 to 587998,
EPI_ISL_588000 to 588006, EPI_ISL_588008 to 588012, EPI_ISL_588014 to 588025, EPI_ISL_588027 to 588049, EPI_ISL_588051 to 588058, EPI_ISL_589267 to 589279, EPI_ISL_589282 to 589288, EPI_ISL_589290 to 589296, EPI_ISL_589298 to 589301, EPI_ISL_589303, EPI_ISL_589305 to 589315, EPI_ISL_589317 to
589320, EPI_ISL_589322 to 589328, EPI_ISL_589331 to 589333, EPI_ISL_589337 to 589341, EPI_ISL_589343, EPI_ISL_589345 to 589346, EPI_ISL_589348 to 589354, EPI_ISL_589356 to 589375, EPI_ISL_589378 to 589389, EPI_ISL_589391 to 589406, EPI_ISL_589408 to 589416, EPI_ISL_589418 to 589422, EPI_ISL_589424
to 589429, EPI_ISL_589431 to 589439, EPI_ISL_589441 to 589476, EPI_ISL_589478 to 589492, EPI_ISL_589494 to 589518, EPI_ISL_589522 to 589525, EPI_ISL_589527 to 589531, EPI_ISL_589533 to 589566, EPI_ISL_589568 to 589571, EPI_ISL_589762, EPI_ISL_589765, EPI_ISL_589778 to 589786, EPI_ISL_589802,
EPI_ISL_590181, EPI_ISL_590186, EPI_ISL_590193 to 590194, EPI_ISL_590198, EPI_ISL_590200, EPI_ISL_590206, EPI_ISL_590210, EPI_ISL_590214 to 590215, EPI_ISL_590220, EPI_ISL_590224 to 590226, EPI_ISL_590228, EPI_ISL_590234 to 590235, EPI_ISL_590239, EPI_ISL_590246, EPI_ISL_590250,
EPI_ISL_590255, EPI_ISL_590257, EPI_ISL_590259 to 590261, EPI_ISL_590265, EPI_ISL_590274 to 590277, EPI_ISL_590282, EPI_ISL_590284, EPI_ISL_590286, EPI_ISL_590288, EPI_ISL_590295 to 590296, EPI_ISL_590300, EPI_ISL_590306 to 590307, EPI_ISL_590309, EPI_ISL_590314, EPI_ISL_590319,
EPI_ISL_590321, EPI_ISL_590327 to 590329, EPI_ISL_590335, EPI_ISL_590337, EPI_ISL_590346 to 590347, EPI_ISL_590351 to 590352, EPI_ISL_590355, EPI_ISL_590357, EPI_ISL_590359, EPI_ISL_590361, EPI_ISL_590364, EPI_ISL_590367, EPI_ISL_590369, EPI_ISL_590373, EPI_ISL_590379 to 590380,
EPI_ISL_590383, EPI_ISL_590386, EPI_ISL_590388, EPI_ISL_590390, EPI_ISL_590392, EPI_ISL_590397 to 590399, EPI_ISL_590403, EPI_ISL_590405, EPI_ISL_590419, EPI_ISL_590422 to 590423, EPI_ISL_590438, EPI_ISL_590441 to 590442, EPI_ISL_590446, EPI_ISL_590448, EPI_ISL_590452, EPI_ISL_590462,
EPI_ISL_597523 to 597548, EPI_ISL_597550 to 597620, EPI_ISL_597622 to 597630, EPI_ISL_597632, EPI_ISL_597634 to 597672, EPI_ISL_597674 to 597718, EPI_ISL_597720 to 597803, EPI_ISL_597805 to 597810, EPI_ISL_597812 to 597843, EPI_ISL_597845 to 597866, EPI_ISL_597868 to 597870, EPI_ISL_597872 to
597890, EPI_ISL_597892 to 597922, EPI_ISL_597924 to 597975, EPI_ISL_597977 to 597980, EPI_ISL_597982 to 597983, EPI_ISL_597985 to 597990, EPI_ISL_597992 to 598000, EPI_ISL_598002 to 598023, EPI_ISL_598025 to 598043, EPI_ISL_598045 to 598080, EPI_ISL_598082 to 598146, EPI_ISL_598148 to 598152,
EPI_ISL_598154 to 598162, EPI_ISL_598164 to 598193, EPI_ISL_598195 to 598222, EPI_ISL_598224 to 598233, EPI_ISL_598235 to 598248, EPI_ISL_598824 to 598830, EPI_ISL_600521, EPI_ISL_600523, EPI_ISL_600525 to 600526, EPI_ISL_600528 to 600529, EPI_ISL_600531 to 600532, EPI_ISL_600534, EPI_ISL_600536
to 600537, EPI_ISL_602155, EPI_ISL_602158 to 602159, EPI_ISL_602161, EPI_ISL_605932 to 605936, EPI_ISL_605938 to 605971, EPI_ISL_605973 to 605982, EPI_ISL_605984 to 605991, EPI_ISL_605993 to 606030, EPI_ISL_606032 to 606070, EPI_ISL_606072 to 606104, EPI_ISL_606106 to 606109, EPI_ISL_606111 to
606138, EPI_ISL_606140 to 606154, EPI_ISL_606156 to 606175, EPI_ISL_606177 to 606178, EPI_ISL_606180 to 606205, EPI_ISL_606207 to 606225, EPI_ISL_606227 to 606266, EPI_ISL_606268 to 606295, EPI_ISL_606924 to 606947, EPI_ISL_606949 to 606959, EPI_ISL_606961 to 606986, EPI_ISL_606988 to 607047,
EPI_ISL_607049 to 607154, EPI_ISL_607156 to 607188, EPI_ISL_607190 to 607199, EPI_ISL_607201 to 607248, EPI_ISL_607250 to 607290, EPI_ISL_607623 to 607659, EPI_ISL_607661 to 607677, EPI_ISL_607679 to 607691, EPI_ISL_607693 to 607695, EPI_ISL_607697 to 607746, EPI_ISL_607748 to 607757,
EPI_ISL_607759 to 607775, EPI_ISL_607777 to 607828, EPI_ISL_607830 to 607844, EPI_ISL_607846 to 607893, EPI_ISL_607895 to 607977, EPI_ISL_607979 to 608014, EPI_ISL_608016 to 608058, EPI_ISL_608060 to 608068, EPI_ISL_608070 to 608119, EPI_ISL_608121 to 608146, EPI_ISL_608148 to 608173,
EPI_ISL_608175 to 608189, EPI_ISL_608191 to 608214, EPI_ISL_608217 to 608238, EPI_ISL_608240 to 608251, EPI_ISL_608253 to 608308, EPI_ISL_608311 to 608346, EPI_ISL_608350 to 608352, EPI_ISL_608355 to 608356, EPI_ISL_608358, EPI_ISL_608361 to 608362, EPI_ISL_608364 to 608367, EPI_ISL_608369,
EPI_ISL_608371 to 608372, EPI_ISL_608374 to 608380, EPI_ISL_608383 to 608387, EPI_ISL_608390, EPI_ISL_608399, EPI_ISL_608401, EPI_ISL_608403, EPI_ISL_608405 to 608406, EPI_ISL_608409, EPI_ISL_608413 to 608415, EPI_ISL_608418 to 608422, EPI_ISL_608425, EPI_ISL_608431, EPI_ISL_608434 to 608435,
EPI_ISL_608438, EPI_ISL_608440 to 608441, EPI_ISL_608443 to 608445, EPI_ISL_608448, EPI_ISL_608452 to 608454, EPI_ISL_608457, EPI_ISL_608460 to 608462, EPI_ISL_608465, EPI_ISL_608468 to 608470, EPI_ISL_608473 to 608474, EPI_ISL_608479, EPI_ISL_608481, EPI_ISL_608483, EPI_ISL_608487 to 608489,
EPI_ISL_608491, EPI_ISL_608496, EPI_ISL_608499 to 608500, EPI_ISL_608502 to 608503, EPI_ISL_608505, EPI_ISL_608511 to 608513, EPI_ISL_608516 to 608520, EPI_ISL_608526, EPI_ISL_608528, EPI_ISL_608533 to 608534, EPI_ISL_608539, EPI_ISL_608541 to 608543, EPI_ISL_608548 to 608549, EPI_ISL_608551,
EPI_ISL_608553 to 608556, EPI_ISL_608562 to 608563, EPI_ISL_608567, EPI_ISL_608569, EPI_ISL_608574 to 608575, EPI_ISL_608579 to 608584, EPI_ISL_608586 to 608587, EPI_ISL_608589 to 608595, EPI_ISL_608599 to 608601, EPI_ISL_608603, EPI_ISL_608605 to 608606, EPI_ISL_608609 to 608610, EPI_ISL_608614,
EPI_ISL_608617, EPI_ISL_608621 to 608622, EPI_ISL_608625, EPI_ISL_608630, EPI_ISL_608634 to 608636, EPI_ISL_608639 to 608640, EPI_ISL_608646 to 608649, EPI_ISL_608652, EPI_ISL_608655, EPI_ISL_608657 to 608659, EPI_ISL_608661 to 608663, EPI_ISL_608665, EPI_ISL_608667 to 608670, EPI_ISL_608672,
EPI_ISL_608674 to 608677, EPI_ISL_608680, EPI_ISL_608684 to 608687, EPI_ISL_608690 to 608692, EPI_ISL_608694, EPI_ISL_608847, EPI_ISL_608849, EPI_ISL_608851, EPI_ISL_608853, EPI_ISL_608855 to 608856, EPI_ISL_608858, EPI_ISL_608860, EPI_ISL_608862 to 608866, EPI_ISL_608869, EPI_ISL_608872,
EPI_ISL_608874, EPI_ISL_608877 to 608878, EPI_ISL_608880, EPI_ISL_608883 to 608891, EPI_ISL_608895 to 608900, EPI_ISL_608903, EPI_ISL_608909, EPI_ISL_608911, EPI_ISL_608913 to 608917, EPI_ISL_608919 to 608922, EPI_ISL_608925 to 608926, EPI_ISL_608928 to 608934, EPI_ISL_608938, EPI_ISL_608941 to
608942, EPI_ISL_608944 to 608946, EPI_ISL_608948, EPI_ISL_608950, EPI_ISL_608952, EPI_ISL_608955 to 608957, EPI_ISL_608961 to 608964, EPI_ISL_608966, EPI_ISL_608970 to 608973, EPI_ISL_608976, EPI_ISL_608979 to 608981, EPI_ISL_608983 to 608985, EPI_ISL_608988 to 608989, EPI_ISL_608991 to 608993,
EPI_ISL_608995, EPI_ISL_608998 to 609000, EPI_ISL_609002, EPI_ISL_609004, EPI_ISL_609007, EPI_ISL_609009 to 609013, EPI_ISL_609016, EPI_ISL_609019 to 609020, EPI_ISL_609022, EPI_ISL_609028, EPI_ISL_609031 to 609032, EPI_ISL_609034 to 609035, EPI_ISL_609039 to 609041, EPI_ISL_609043,

EPI_ISL_609047, EPI_ISL_609051 to 609052, EPI_ISL_609055 to 609057, EPI_ISL_609060 to 609061, EPI_ISL_609064 to 609065, EPI_ISL_609068 to 609070, EPI_ISL_609073 to 609075, EPI_ISL_609078 to 609080, EPI_ISL_609083, EPI_ISL_609085, EPI_ISL_609087, EPI_ISL_609089 to 609090, EPI_ISL_609092 to 609096,
EPI_ISL_609098, EPI_ISL_609102 to 609107, EPI_ISL_609109, EPI_ISL_609120, EPI_ISL_609124 to 609125, EPI_ISL_609127, EPI_ISL_609130, EPI_ISL_609132 to 609135, EPI_ISL_609141, EPI_ISL_609144, EPI_ISL_609146 to 609148, EPI_ISL_609150, EPI_ISL_609152 to 609154, EPI_ISL_609157, EPI_ISL_609207 to
609208, EPI_ISL_609210 to 609218, EPI_ISL_610286 to 610331, EPI_ISL_610333 to 610361, EPI_ISL_610363, EPI_ISL_610365 to 610397, EPI_ISL_610399 to 610406, EPI_ISL_610408 to 610410, EPI_ISL_610412, EPI_ISL_610414 to 610447, EPI_ISL_610449 to 610455, EPI_ISL_610457 to 610493, EPI_ISL_610495 to
610501, EPI_ISL_610504 to 610529, EPI_ISL_610531 to 610555, EPI_ISL_610558 to 610573, EPI_ISL_610575 to 610591, EPI_ISL_610593 to 610602, EPI_ISL_610604 to 610628, EPI_ISL_610922 to 610956, EPI_ISL_610958 to 611013, EPI_ISL_611015 to 611071, EPI_ISL_611073 to 611092, EPI_ISL_611094 to 611155,
EPI_ISL_611157 to 611261, EPI_ISL_611263 to 611286, EPI_ISL_623858 to 623862, EPI_ISL_623866 to 623867, EPI_ISL_623869 to 623871, EPI_ISL_623875, EPI_ISL_623877 to 623882, EPI_ISL_623885, EPI_ISL_623887 to 623892, EPI_ISL_623894, EPI_ISL_623896 to 623902, EPI_ISL_623905 to 623906, EPI_ISL_623909
to 623910, EPI_ISL_623912 to 623915, EPI_ISL_623918 to 623920, EPI_ISL_623922 to 623923, EPI_ISL_623925 to 623929, EPI_ISL_623931 to 623932, EPI_ISL_623935 to 623941, EPI_ISL_623943, EPI_ISL_623945 to 623948, EPI_ISL_623950 to 623953, EPI_ISL_623955, EPI_ISL_623957 to 623958, EPI_ISL_623960 to
623962, EPI_ISL_623964 to 623973, EPI_ISL_623978 to 623982, EPI_ISL_623984 to 623985, EPI_ISL_623987 to 623989, EPI_ISL_623992 to 623993, EPI_ISL_623996 to 624009, EPI_ISL_624011, EPI_ISL_624013 to 624017, EPI_ISL_624020 to 624021, EPI_ISL_624023 to 624026, EPI_ISL_624030 to 624031,
EPI_ISL_624033, EPI_ISL_624035 to 624042, EPI_ISL_624045 to 624048, EPI_ISL_624050 to 624066, EPI_ISL_624068 to 624072, EPI_ISL_624074 to 624076, EPI_ISL_624078, EPI_ISL_624080 to 624091, EPI_ISL_624093, EPI_ISL_624095 to 624104, EPI_ISL_624106 to 624111, EPI_ISL_624113, EPI_ISL_624115 to 624119,
EPI_ISL_624121, EPI_ISL_624123 to 624127, EPI_ISL_624129 to 624131, EPI_ISL_624133 to 624139, EPI_ISL_624141 to 624144, EPI_ISL_624146, EPI_ISL_624149, EPI_ISL_624151 to 624157, EPI_ISL_624159 to 624162, EPI_ISL_624166 to 624175, EPI_ISL_624177 to 624180, EPI_ISL_624182 to 624185, EPI_ISL_624187,
EPI_ISL_624190 to 624196, EPI_ISL_624198 to 624215, EPI_ISL_624217 to 624223, EPI_ISL_624225 to 624243, EPI_ISL_624245 to 624290, EPI_ISL_624292 to 624305, EPI_ISL_624307 to 624334, EPI_ISL_624336 to 624348, EPI_ISL_624352, EPI_ISL_624355 to 624370, EPI_ISL_624372 to 624523, EPI_ISL_624525 to
624577, EPI_ISL_624579 to 624592, EPI_ISL_624594 to 624612, EPI_ISL_624614 to 624686, EPI_ISL_624688 to 624715, EPI_ISL_624717 to 624723, EPI_ISL_624725 to 624760, EPI_ISL_624762 to 624845, EPI_ISL_624847 to 624870, EPI_ISL_624872 to 624902, EPI_ISL_624904 to 624908, EPI_ISL_624910 to 625167,
EPI_ISL_625170 to 625179, EPI_ISL_625181 to 625212, EPI_ISL_625214 to 625239, EPI_ISL_625241 to 625246, EPI_ISL_625248 to 625280, EPI_ISL_625396 to 625404, EPI_ISL_625422, EPI_ISL_625425, EPI_ISL_625427, EPI_ISL_625429, EPI_ISL_625431, EPI_ISL_625433 to 625436, EPI_ISL_630348 to 630435,
EPI_ISL_630437 to 630473, EPI_ISL_630475 to 630499, EPI_ISL_630501 to 630532, EPI_ISL_630534 to 630546, EPI_ISL_630548 to 630562, EPI_ISL_630564 to 630616, EPI_ISL_630618 to 630620, EPI_ISL_630622 to 630654, EPI_ISL_630656 to 630676, EPI_ISL_630678 to 630701, EPI_ISL_630706 to 630709,
EPI_ISL_630711, EPI_ISL_630713 to 630717, EPI_ISL_630720 to 630724, EPI_ISL_630726 to 630730, EPI_ISL_630732, EPI_ISL_630734 to 630740, EPI_ISL_630742 to 630744, EPI_ISL_630746 to 630748, EPI_ISL_630750 to 630760, EPI_ISL_630762 to 630763, EPI_ISL_630765 to 630774, EPI_ISL_630776 to 630777,
EPI_ISL_630779, EPI_ISL_630781, EPI_ISL_630783 to 630786, EPI_ISL_630788 to 630803, EPI_ISL_630805 to 630812, EPI_ISL_630814, EPI_ISL_630816 to 630823, EPI_ISL_630825 to 630826, EPI_ISL_630828 to 630829, EPI_ISL_630831 to 630832, EPI_ISL_630834 to 630837, EPI_ISL_630839 to 630849, EPI_ISL_630852,
EPI_ISL_630854 to 630858, EPI_ISL_630860 to 630861, EPI_ISL_630863, EPI_ISL_630865 to 630869, EPI_ISL_630874, EPI_ISL_630877 to 630882, EPI_ISL_630884 to 630887, EPI_ISL_630889 to 630890, EPI_ISL_630892 to 630899, EPI_ISL_630901 to 630914, EPI_ISL_630916 to 630918, EPI_ISL_630920 to 630922,
EPI_ISL_630924 to 630925, EPI_ISL_630927 to 630930, EPI_ISL_630932, EPI_ISL_630934 to 630940, EPI_ISL_630942 to 630947, EPI_ISL_630950, EPI_ISL_630952 to 630954, EPI_ISL_630956, EPI_ISL_630958, EPI_ISL_630960 to 630964, EPI_ISL_630966 to 630971, EPI_ISL_630975 to 630976, EPI_ISL_630982 to 630983,
EPI_ISL_630985, EPI_ISL_630992, EPI_ISL_630994, EPI_ISL_630999, EPI_ISL_631011 to 631012, EPI_ISL_631014, EPI_ISL_631018, EPI_ISL_631023, EPI_ISL_631027, EPI_ISL_631031, EPI_ISL_631033, EPI_ISL_631035, EPI_ISL_631041, EPI_ISL_631044, EPI_ISL_631046, EPI_ISL_631050, EPI_ISL_631056 to 631057,
EPI_ISL_631059 to 631060, EPI_ISL_631074, EPI_ISL_631081, EPI_ISL_631084, EPI_ISL_631089, EPI_ISL_631093, EPI_ISL_631096, EPI_ISL_631099, EPI_ISL_631107, EPI_ISL_631111, EPI_ISL_631113, EPI_ISL_631116, EPI_ISL_631127 to 631128, EPI_ISL_631131, EPI_ISL_631133, EPI_ISL_631150, EPI_ISL_631163,
EPI_ISL_631165, EPI_ISL_631173, EPI_ISL_631175, EPI_ISL_631184, EPI_ISL_631189 to 631190, EPI_ISL_631202, EPI_ISL_631212, EPI_ISL_631215, EPI_ISL_631217, EPI_ISL_631222, EPI_ISL_631228, EPI_ISL_631231, EPI_ISL_631241, EPI_ISL_631250, EPI_ISL_631265, EPI_ISL_631269, EPI_ISL_634754 to 634788

| | | | |
|---|---|---|---|
| see above | Lighthouse Lab in Alderley Park | Wellcome Sanger Institute for the COVID-19 Genomics UK (COG-UK) consortium | Cordelia Langford; David K. Jackson; Dominic Kwiatkowski; Ewan Harrison; Ian Johnston; Jacquelyn Wynn; John Sillitoe on behalf of the Wellcome Sanger Institute COVID-19 Surveillance Team; John Sillitoe on behalf of the Wellcome Sanger Institute COVID-19 Surveillance Team (http://www.sanger.ac.uk/covid-team); Mairead Hyland; Roberto Amato; Sonia Goncalves; The Lighthouse Lab in Alderley Park and Alex Alderton |

EPI_ISL_551850, EPI_ISL_556698, EPI_ISL_556770, EPI_ISL_556940, EPI_ISL_566455, EPI_ISL_566474, EPI_ISL_580332, EPI_ISL_580594, EPI_ISL_596969, EPI_ISL_597074, EPI_ISL_597099, EPI_ISL_597189, EPI_ISL_608347, EPI_ISL_608455, EPI_ISL_608681, EPI_ISL_608907, EPI_ISL_609121, EPI_ISL_609378,
EPI_ISL_609393, EPI_ISL_609450, EPI_ISL_609527, EPI_ISL_609534, EPI_ISL_609600, EPI_ISL_609746, EPI_ISL_610702, EPI_ISL_610779, EPI_ISL_623477, EPI_ISL_623781, EPI_ISL_625289, EPI_ISL_631159, EPI_ISL_633165, EPI_ISL_633221, EPI_ISL_643315 to 643330, EPI_ISL_643332 to 643385, EPI_ISL_643387 to
643393, EPI_ISL_643395 to 643447, EPI_ISL_643449 to 643457, EPI_ISL_643459 to 643497, EPI_ISL_643499 to 643523, EPI_ISL_643525 to 643530, EPI_ISL_643532 to 643571, EPI_ISL_643573 to 643645, EPI_ISL_643647 to 643662, EPI_ISL_643664 to 643675, EPI_ISL_643677, EPI_ISL_643679 to 643696, EPI_ISL_643698
to 643699, EPI_ISL_643701 to 643709, EPI_ISL_643711 to 643733, EPI_ISL_643735 to 643839, EPI_ISL_643841 to 643859, EPI_ISL_643861 to 643914, EPI_ISL_643916 to 643927, EPI_ISL_643929 to 643950, EPI_ISL_643952 to 643982, EPI_ISL_644115, EPI_ISL_644117, EPI_ISL_645219 to 645220, EPI_ISL_645222 to
645225, EPI_ISL_645227 to 645239, EPI_ISL_645242, EPI_ISL_645245 to 645262, EPI_ISL_645265, EPI_ISL_645267 to 645284, EPI_ISL_645286 to 645295, EPI_ISL_645298 to 645299, EPI_ISL_645301, EPI_ISL_645304 to 645307, EPI_ISL_645309 to 645318, EPI_ISL_645320 to 645323, EPI_ISL_645325 to 645326,
EPI_ISL_645328, EPI_ISL_645330 to 645344, EPI_ISL_645346 to 645353, EPI_ISL_645355 to 645359, EPI_ISL_645361, EPI_ISL_645363 to 645366, EPI_ISL_645370 to 645373, EPI_ISL_645376, EPI_ISL_645378 to 645382, EPI_ISL_645384 to 645390, EPI_ISL_645392, EPI_ISL_645394, EPI_ISL_645396 to 645398,
EPI_ISL_645400 to 645404, EPI_ISL_645407 to 645417, EPI_ISL_645419 to 645420, EPI_ISL_645422 to 645423, EPI_ISL_645425 to 645437, EPI_ISL_645439 to 645440, EPI_ISL_645442 to 645451, EPI_ISL_645454 to 645458, EPI_ISL_645460 to 645463, EPI_ISL_645465 to 645472, EPI_ISL_645474 to 645476,
EPI_ISL_645478 to 645480, EPI_ISL_645482 to 645488, EPI_ISL_645490 to 645498, EPI_ISL_645500 to 645501, EPI_ISL_645504 to 645507, EPI_ISL_645509, EPI_ISL_645511 to 645512, EPI_ISL_645514 to 645516, EPI_ISL_645519, EPI_ISL_645521 to 645529, EPI_ISL_645826, EPI_ISL_645828 to 645829, EPI_ISL_645832,
EPI_ISL_645836, EPI_ISL_645838, EPI_ISL_645840, EPI_ISL_645850 to 645852, EPI_ISL_645866, EPI_ISL_645868 to 645869, EPI_ISL_645877, EPI_ISL_645880, EPI_ISL_645882, EPI_ISL_645885 to 645886, EPI_ISL_645896 to 645897, EPI_ISL_645899, EPI_ISL_645902, EPI_ISL_645905, EPI_ISL_645908,
EPI_ISL_645916 to 645918, EPI_ISL_645920, EPI_ISL_645923, EPI_ISL_645925 to 645927, EPI_ISL_645930, EPI_ISL_645945, EPI_ISL_645949, EPI_ISL_645959, EPI_ISL_645962, EPI_ISL_645968, EPI_ISL_645970, EPI_ISL_645979 to 645981, EPI_ISL_645983, EPI_ISL_645987, EPI_ISL_645989, EPI_ISL_645992 to
645995, EPI_ISL_645998, EPI_ISL_646009, EPI_ISL_646014, EPI_ISL_646017 to 646018, EPI_ISL_646022, EPI_ISL_646027 to 646028, EPI_ISL_646030 to 646032, EPI_ISL_646034 to 646035, EPI_ISL_646039, EPI_ISL_646041, EPI_ISL_646046 to 646047, EPI_ISL_646052, EPI_ISL_646056, EPI_ISL_646058,
EPI_ISL_646067, EPI_ISL_646070, EPI_ISL_646076, EPI_ISL_646083 to 646086, EPI_ISL_646089, EPI_ISL_646091, EPI_ISL_646101 to 646102, EPI_ISL_646105, EPI_ISL_646115, EPI_ISL_646122, EPI_ISL_646128 to 646129, EPI_ISL_646131, EPI_ISL_646135 to 646136, EPI_ISL_646139, EPI_ISL_647123 to 647132,
EPI_ISL_647134 to 647162, EPI_ISL_647164 to 647206, EPI_ISL_647208 to 647254, EPI_ISL_647256 to 647287, EPI_ISL_647290 to 647365, EPI_ISL_647367 to 647385, EPI_ISL_647387 to 647443, EPI_ISL_647445 to 647467, EPI_ISL_647470 to 647472, EPI_ISL_647475 to 647476, EPI_ISL_647478 to 647496,
EPI_ISL_647498, EPI_ISL_647500 to 647502, EPI_ISL_647504 to 647505, EPI_ISL_647507 to 647510, EPI_ISL_647512, EPI_ISL_647514 to 647517, EPI_ISL_647519 to 647521, EPI_ISL_647523 to 647525, EPI_ISL_647527, EPI_ISL_647529 to 647534, EPI_ISL_647537 to 647540, EPI_ISL_647542 to 647543, EPI_ISL_647545,
EPI_ISL_647547 to 647549, EPI_ISL_647551 to 647552, EPI_ISL_647554 to 647555, EPI_ISL_647559, EPI_ISL_647561 to 647571, EPI_ISL_647574 to 647575, EPI_ISL_647577 to 647578, EPI_ISL_647581 to 647583, EPI_ISL_647585 to 647590, EPI_ISL_647592 to 647593, EPI_ISL_647595 to 647602, EPI_ISL_647604 to
647605, EPI_ISL_647607, EPI_ISL_647609 to 647611, EPI_ISL_647613 to 647614, EPI_ISL_647616 to 647617, EPI_ISL_647619 to 647623, EPI_ISL_647625 to 647629, EPI_ISL_647631, EPI_ISL_647633, EPI_ISL_647636 to 647637, EPI_ISL_647639 to 647642, EPI_ISL_647644 to 647649, EPI_ISL_647651 to 647654,
EPI_ISL_647656 to 647658, EPI_ISL_647660 to 647666, EPI_ISL_647669 to 647671, EPI_ISL_647673 to 647676, EPI_ISL_647678 to 647681, EPI_ISL_647684 to 647688, EPI_ISL_647690 to 647691, EPI_ISL_647694, EPI_ISL_647696 to 647697, EPI_ISL_647699, EPI_ISL_647701 to 647703, EPI_ISL_647706 to 647707,
EPI_ISL_647709 to 647710, EPI_ISL_647712 to 647725, EPI_ISL_647727 to 647732, EPI_ISL_647734 to 647740, EPI_ISL_647742 to 647749, EPI_ISL_647751, EPI_ISL_647753 to 647761, EPI_ISL_647763 to 647770, EPI_ISL_647772 to 647775, EPI_ISL_647777 to 647779, EPI_ISL_647781 to 647785, EPI_ISL_647788 to
647791, EPI_ISL_647793 to 647796, EPI_ISL_647900 to 647901, EPI_ISL_647906, EPI_ISL_647908 to 647909, EPI_ISL_647913, EPI_ISL_647916, EPI_ISL_649482, EPI_ISL_649492 to 649493, EPI_ISL_649625 to 649632, EPI_ISL_656686 to 656696, EPI_ISL_656698 to 656703, EPI_ISL_656706 to 656727, EPI_ISL_656729 to
656730, EPI_ISL_656734 to 656742, EPI_ISL_656747 to 656753, EPI_ISL_656755 to 656756, EPI_ISL_656758 to 656773, EPI_ISL_656775 to 656776, EPI_ISL_656781 to 656787, EPI_ISL_656790 to 656794, EPI_ISL_656796 to 656797, EPI_ISL_656800 to 656806, EPI_ISL_656809, EPI_ISL_656812 to 656817, EPI_ISL_656819
to 656823, EPI_ISL_656825 to 656826, EPI_ISL_656828, EPI_ISL_656830 to 656833, EPI_ISL_656836 to 656837, EPI_ISL_656839, EPI_ISL_656842 to 656845, EPI_ISL_656847, EPI_ISL_656849 to 656859, EPI_ISL_656863 to 656864, EPI_ISL_656866, EPI_ISL_656868, EPI_ISL_656872 to 656876, EPI_ISL_656878,
EPI_ISL_656880 to 656886, EPI_ISL_656888 to 656891, EPI_ISL_656894 to 656905, EPI_ISL_656907 to 656909, EPI_ISL_656911 to 656912, EPI_ISL_656914 to 656919, EPI_ISL_656921, EPI_ISL_656923, EPI_ISL_656925 to 656929, EPI_ISL_656931, EPI_ISL_656933 to 656934, EPI_ISL_656937, EPI_ISL_656939 to 656941,
EPI_ISL_656943 to 656944, EPI_ISL_656946, EPI_ISL_656948 to 656949, EPI_ISL_656951 to 656954, EPI_ISL_656956, EPI_ISL_656959 to 656960, EPI_ISL_656963 to 656964, EPI_ISL_656967 to 656968, EPI_ISL_656970 to 656972, EPI_ISL_656975, EPI_ISL_656977 to 656978, EPI_ISL_656981, EPI_ISL_656983 to 656986,
EPI_ISL_656988 to 656990, EPI_ISL_656992 to 656996, EPI_ISL_656998 to 657002, EPI_ISL_657005 to 657007, EPI_ISL_657009 to 657010, EPI_ISL_657012 to 657013, EPI_ISL_657015, EPI_ISL_657018 to 657020, EPI_ISL_657022, EPI_ISL_657024 to 657025, EPI_ISL_657356 to 657358, EPI_ISL_657362 to 657363,
EPI_ISL_657366, EPI_ISL_657368, EPI_ISL_657374, EPI_ISL_657376, EPI_ISL_657378, EPI_ISL_657380, EPI_ISL_657387 to 657389, EPI_ISL_657394 to 657395, EPI_ISL_657397, EPI_ISL_657399, EPI_ISL_657402 to 657404, EPI_ISL_657407, EPI_ISL_657409, EPI_ISL_657412, EPI_ISL_657415, EPI_ISL_657421 to
657423, EPI_ISL_657426 to 657427, EPI_ISL_657431 to 657437, EPI_ISL_657442, EPI_ISL_657444 to 657445, EPI_ISL_657447 to 657449, EPI_ISL_657454, EPI_ISL_657456, EPI_ISL_657461, EPI_ISL_657468 to 657471, EPI_ISL_657474 to 657478, EPI_ISL_657481 to 657482, EPI_ISL_657489, EPI_ISL_657496,
EPI_ISL_657498 to 657499, EPI_ISL_657501 to 657502, EPI_ISL_657504, EPI_ISL_657506 to 657508, EPI_ISL_657511, EPI_ISL_657514, EPI_ISL_657516, EPI_ISL_657520 to 657525, EPI_ISL_657528 to 657529, EPI_ISL_657534, EPI_ISL_657537 to 657541, EPI_ISL_657545 to 657546, EPI_ISL_657548 to 657549,
EPI_ISL_657551 to 657552, EPI_ISL_657559, EPI_ISL_657561 to 657562, EPI_ISL_657565 to 657566, EPI_ISL_657569, EPI_ISL_657572 to 657573, EPI_ISL_657576, EPI_ISL_657578, EPI_ISL_657580, EPI_ISL_657582, EPI_ISL_657586, EPI_ISL_657588, EPI_ISL_657590, EPI_ISL_657592, EPI_ISL_657595,
EPI_ISL_657597 to 657598, EPI_ISL_657600 to 657601, EPI_ISL_657603 to 657604, EPI_ISL_657612 to 657613, EPI_ISL_657617 to 657623, EPI_ISL_657626, EPI_ISL_657628, EPI_ISL_657630, EPI_ISL_657633 to 657634, EPI_ISL_657637 to 657638, EPI_ISL_657641, EPI_ISL_657643, EPI_ISL_657645 to 657646,
EPI_ISL_657649, EPI_ISL_657652 to 657656, EPI_ISL_657658 to 657660, EPI_ISL_657664, EPI_ISL_657668 to 657672, EPI_ISL_657674 to 657675, EPI_ISL_657677 to 657679, EPI_ISL_657684 to 657687, EPI_ISL_657689 to 657691, EPI_ISL_657693, EPI_ISL_657697 to 657699, EPI_ISL_657701, EPI_ISL_657704,
EPI_ISL_657707 to 657708, EPI_ISL_657713, EPI_ISL_657720 to 657721, EPI_ISL_657723 to 657726, EPI_ISL_657728, EPI_ISL_657731, EPI_ISL_657736 to 657737, EPI_ISL_657739, EPI_ISL_657744, EPI_ISL_657746 to 657747, EPI_ISL_657749 to 657750, EPI_ISL_657754, EPI_ISL_657756, EPI_ISL_657759 to 657760,
EPI_ISL_657762 to 657767, EPI_ISL_657769, EPI_ISL_657771, EPI_ISL_657780, EPI_ISL_657782, EPI_ISL_657786 to 657787, EPI_ISL_657790, EPI_ISL_657793, EPI_ISL_657795, EPI_ISL_657798 to 657799, EPI_ISL_657801 to 657807, EPI_ISL_657809 to 657810, EPI_ISL_657813, EPI_ISL_657815 to 657818,
EPI_ISL_657824 to 657826, EPI_ISL_657828, EPI_ISL_657831, EPI_ISL_657833 to 657834, EPI_ISL_657837 to 657839, EPI_ISL_657841, EPI_ISL_657845 to 657847, EPI_ISL_657849, EPI_ISL_657851 to 657853, EPI_ISL_657855, EPI_ISL_657857, EPI_ISL_657860, EPI_ISL_657863 to 657864, EPI_ISL_657866 to 657868,
EPI_ISL_657871, EPI_ISL_657873, EPI_ISL_657875 to 657877, EPI_ISL_657880 to 657885, EPI_ISL_657890 to 657891, EPI_ISL_657893, EPI_ISL_657895 to 657897, EPI_ISL_657899 to 657900, EPI_ISL_657902 to 657904, EPI_ISL_657907 to 657909, EPI_ISL_657911 to 657912, EPI_ISL_657914, EPI_ISL_657917,
EPI_ISL_657920, EPI_ISL_657924, EPI_ISL_657926, EPI_ISL_657931, EPI_ISL_657933 to 657937, EPI_ISL_657939, EPI_ISL_657942 to 657943, EPI_ISL_657945 to 657947, EPI_ISL_657949, EPI_ISL_657951 to 657953, EPI_ISL_657957, EPI_ISL_657960, EPI_ISL_657964, EPI_ISL_657966 to 657968, EPI_ISL_657974,
EPI_ISL_657976, EPI_ISL_657978, EPI_ISL_657983, EPI_ISL_657986 to 657987, EPI_ISL_657989 to 657990, EPI_ISL_657992, EPI_ISL_657994, EPI_ISL_657998, EPI_ISL_658001, EPI_ISL_658007 to 658008, EPI_ISL_658010 to 658014, EPI_ISL_658019 to 658020, EPI_ISL_658024, EPI_ISL_658028 to 658034,
EPI_ISL_658036 to 658037, EPI_ISL_658041, EPI_ISL_658043, EPI_ISL_658045 to 658046, EPI_ISL_658048, EPI_ISL_658052, EPI_ISL_658054 to 658056, EPI_ISL_658063 to 658064, EPI_ISL_658069, EPI_ISL_658071 to 658078, EPI_ISL_658080, EPI_ISL_658083 to 658086, EPI_ISL_658089, EPI_ISL_658092 to 658093,
EPI_ISL_658095 to 658101, EPI_ISL_658104, EPI_ISL_658110, EPI_ISL_658116, EPI_ISL_658119 to 658120, EPI_ISL_658122, EPI_ISL_658124 to 658125, EPI_ISL_658128, EPI_ISL_658131, EPI_ISL_658133 to 658134, EPI_ISL_658138, EPI_ISL_658142, EPI_ISL_658144, EPI_ISL_658148, EPI_ISL_658153,
EPI_ISL_658156 to 658157, EPI_ISL_658159 to 658164, EPI_ISL_658166 to 658167, EPI_ISL_658171, EPI_ISL_658177 to 658179, EPI_ISL_658181, EPI_ISL_658186 to 658192, EPI_ISL_658194 to 658203, EPI_ISL_658206 to 658209, EPI_ISL_658214 to 658215, EPI_ISL_658218, EPI_ISL_658220 to 658221, EPI_ISL_658224,
EPI_ISL_658226 to 658228, EPI_ISL_658231, EPI_ISL_658234, EPI_ISL_658236, EPI_ISL_658242, EPI_ISL_658246, EPI_ISL_658252, EPI_ISL_658256, EPI_ISL_658258, EPI_ISL_658260, EPI_ISL_658263, EPI_ISL_658270 to 658272, EPI_ISL_658274 to 658276, EPI_ISL_658278 to 658279, EPI_ISL_658285 to 658287,
EPI_ISL_658292, EPI_ISL_658299, EPI_ISL_658304 to 658305, EPI_ISL_658307, EPI_ISL_658310 to 658314, EPI_ISL_658316 to 658317, EPI_ISL_658320 to 658321, EPI_ISL_658323, EPI_ISL_658326 to 658327, EPI_ISL_658330 to 658332, EPI_ISL_658334 to 658335, EPI_ISL_658337, EPI_ISL_658340 to 658341,
EPI_ISL_658343, EPI_ISL_658345, EPI_ISL_658348, EPI_ISL_658351, EPI_ISL_658355 to 658356, EPI_ISL_658360 to 658364, EPI_ISL_658366 to 658367, EPI_ISL_658370 to 658371, EPI_ISL_658376, EPI_ISL_658381 to 658383, EPI_ISL_658387 to 658388, EPI_ISL_658390, EPI_ISL_658394, EPI_ISL_658396 to 658398,
EPI_ISL_658402, EPI_ISL_658744 to 658745, EPI_ISL_658750, EPI_ISL_658752 to 658753, EPI_ISL_658756, EPI_ISL_658759 to 658760, EPI_ISL_658763 to 658764, EPI_ISL_658767 to 658769, EPI_ISL_658775, EPI_ISL_658781, EPI_ISL_658787 to 658788, EPI_ISL_658795, EPI_ISL_658807, EPI_ISL_658809,
EPI_ISL_658816, EPI_ISL_658829, EPI_ISL_658833, EPI_ISL_658836 to 658838, EPI_ISL_658846, EPI_ISL_658852, EPI_ISL_658864, EPI_ISL_658866, EPI_ISL_658877, EPI_ISL_658881, EPI_ISL_658885, EPI_ISL_658900, EPI_ISL_658905, EPI_ISL_658909, EPI_ISL_658912 to 658914, EPI_ISL_658916, EPI_ISL_658919,
EPI_ISL_658921, EPI_ISL_658923, EPI_ISL_658925, EPI_ISL_658930 to 658931, EPI_ISL_658933, EPI_ISL_658936, EPI_ISL_658948, EPI_ISL_658954, EPI_ISL_658959, EPI_ISL_658963, EPI_ISL_658970 to 658971, EPI_ISL_658976, EPI_ISL_658979, EPI_ISL_658995 to 658996, EPI_ISL_659000, EPI_ISL_659005,
EPI_ISL_659007, EPI_ISL_659011, EPI_ISL_659017 to 659019, EPI_ISL_659025, EPI_ISL_659028, EPI_ISL_659040, EPI_ISL_659045, EPI_ISL_659050, EPI_ISL_659053 to 659054, EPI_ISL_659070 to 659073, EPI_ISL_659076 to 659077, EPI_ISL_659079 to 659080, EPI_ISL_659082 to 659084, EPI_ISL_659090 to 659091,
EPI_ISL_659093 to 659094, EPI_ISL_659097 to 659099, EPI_ISL_659101 to 659122, EPI_ISL_659124 to 659138, EPI_ISL_659140 to 659196, EPI_ISL_659198 to 659204, EPI_ISL_659206 to 659232, EPI_ISL_659234 to 659450, EPI_ISL_659452 to 659606, EPI_ISL_659608 to 659635, EPI_ISL_659637 to 659669,
EPI_ISL_659671 to 659703, EPI_ISL_659705 to 659730, EPI_ISL_659732 to 659736, EPI_ISL_659738 to 659743, EPI_ISL_661638, EPI_ISL_661640 to 661641, EPI_ISL_661647, EPI_ISL_661651 to 661654, EPI_ISL_661656, EPI_ISL_661664, EPI_ISL_661675, EPI_ISL_661679 to 661680, EPI_ISL_661684, EPI_ISL_661687,
EPI_ISL_661691, EPI_ISL_661693, EPI_ISL_661695 to 661697, EPI_ISL_661702, EPI_ISL_661710 to 661712, EPI_ISL_661717, EPI_ISL_661726, EPI_ISL_661729, EPI_ISL_661732, EPI_ISL_661734, EPI_ISL_661741 to 661743, EPI_ISL_661747, EPI_ISL_661749, EPI_ISL_661751 to 661752, EPI_ISL_661759,
EPI_ISL_661761, EPI_ISL_661775, EPI_ISL_661778 to 661779, EPI_ISL_661781, EPI_ISL_661783, EPI_ISL_661789, EPI_ISL_661791, EPI_ISL_661797, EPI_ISL_661799, EPI_ISL_661801, EPI_ISL_661805, EPI_ISL_661811, EPI_ISL_661815, EPI_ISL_661817, EPI_ISL_661830, EPI_ISL_661834, EPI_ISL_661836,
EPI_ISL_661844 to 661845, EPI_ISL_661853 to 661854, EPI_ISL_661856, EPI_ISL_661863, EPI_ISL_661869, EPI_ISL_661873 to 661874, EPI_ISL_661876, EPI_ISL_661881, EPI_ISL_661883, EPI_ISL_661886 to 661887, EPI_ISL_661895, EPI_ISL_661897, EPI_ISL_661900, EPI_ISL_661902 to 661903, EPI_ISL_661917 to
661918, EPI_ISL_661925 to 661926, EPI_ISL_661928, EPI_ISL_661930, EPI_ISL_661941, EPI_ISL_661944, EPI_ISL_661947, EPI_ISL_661951, EPI_ISL_661953 to 661954, EPI_ISL_662631 to 662632, EPI_ISL_662635, EPI_ISL_662651 to 662653, EPI_ISL_667811 to 667813, EPI_ISL_667815, EPI_ISL_667817 to 667821,
EPI_ISL_667823, EPI_ISL_667826 to 667827, EPI_ISL_667830, EPI_ISL_667834 to 667835, EPI_ISL_667837, EPI_ISL_667839 to 667842, EPI_ISL_667845 to 667846, EPI_ISL_667852 to 667854, EPI_ISL_667856, EPI_ISL_667858, EPI_ISL_667861 to 667862, EPI_ISL_667865, EPI_ISL_667869 to 667871, EPI_ISL_667874 to
667875, EPI_ISL_667877, EPI_ISL_667880, EPI_ISL_667882, EPI_ISL_667884, EPI_ISL_667886, EPI_ISL_667890, EPI_ISL_667892, EPI_ISL_667894, EPI_ISL_667896 to 667897, EPI_ISL_667899, EPI_ISL_667901 to 667903, EPI_ISL_667905 to 667906, EPI_ISL_667908 to 667911, EPI_ISL_667913 to 667914,
EPI_ISL_667916, EPI_ISL_667921, EPI_ISL_667923, EPI_ISL_667926, EPI_ISL_667933 to 667936, EPI_ISL_667942, EPI_ISL_667944 to 667946, EPI_ISL_667949, EPI_ISL_667951, EPI_ISL_667953 to 667956, EPI_ISL_667961, EPI_ISL_667964 to 667965, EPI_ISL_667968, EPI_ISL_667971, EPI_ISL_667974,
EPI_ISL_667979, EPI_ISL_667981, EPI_ISL_667984 to 667985, EPI_ISL_667988, EPI_ISL_667991, EPI_ISL_667993 to 667995, EPI_ISL_667999, EPI_ISL_668002 to 668003, EPI_ISL_668007 to 668009, EPI_ISL_668011 to 668012, EPI_ISL_668014 to 668019, EPI_ISL_668023, EPI_ISL_668025 to 668027, EPI_ISL_668037,
EPI_ISL_668039 to 668043, EPI_ISL_668046, EPI_ISL_668048, EPI_ISL_668051, EPI_ISL_668058 to 668059, EPI_ISL_668061, EPI_ISL_668065, EPI_ISL_668068 to 668072, EPI_ISL_668076 to 668077, EPI_ISL_668079 to 668081, EPI_ISL_668085 to 668092, EPI_ISL_668097, EPI_ISL_668100 to 668101, EPI_ISL_668103 to
668104, EPI_ISL_668108 to 668110, EPI_ISL_668112, EPI_ISL_668115, EPI_ISL_668122 to 668123, EPI_ISL_668127 to 668128, EPI_ISL_668136 to 668137, EPI_ISL_668148, EPI_ISL_668155, EPI_ISL_668171, EPI_ISL_668180, EPI_ISL_668188, EPI_ISL_668190, EPI_ISL_668200 to 668201, EPI_ISL_668211 to 668212,
EPI_ISL_668219, EPI_ISL_668222, EPI_ISL_668227, EPI_ISL_668237, EPI_ISL_668252, EPI_ISL_668254 to 668255, EPI_ISL_668265, EPI_ISL_668267, EPI_ISL_668283, EPI_ISL_668289, EPI_ISL_668299, EPI_ISL_668313, EPI_ISL_668316, EPI_ISL_668319, EPI_ISL_668336, EPI_ISL_668353, EPI_ISL_668369,
EPI_ISL_668374, EPI_ISL_668376, EPI_ISL_668385, EPI_ISL_668388 to 668389, EPI_ISL_672912 to 672914, EPI_ISL_672916 to 672923, EPI_ISL_672925 to 672931, EPI_ISL_672933 to 672938, EPI_ISL_672940 to 672948, EPI_ISL_672950, EPI_ISL_672952 to 672971, EPI_ISL_672973 to 672978, EPI_ISL_672982 to 672986,
EPI_ISL_672988, EPI_ISL_672990 to 673003, EPI_ISL_673005 to 673008, EPI_ISL_673010, EPI_ISL_673012 to 673033, EPI_ISL_673035 to 673048, EPI_ISL_673050 to 673060, EPI_ISL_673062 to 673074, EPI_ISL_673076 to 673077, EPI_ISL_673079 to 673082, EPI_ISL_673084 to 673085, EPI_ISL_673087 to 673094,
EPI_ISL_673096, EPI_ISL_673098 to 673101, EPI_ISL_673103 to 673113, EPI_ISL_673115 to 673120, EPI_ISL_673122, EPI_ISL_673124 to 673133, EPI_ISL_673135 to 673144, EPI_ISL_673146, EPI_ISL_673150, EPI_ISL_673152 to 673173, EPI_ISL_673175 to 673180, EPI_ISL_673182 to 673188, EPI_ISL_673190,
EPI_ISL_673192 to 673204, EPI_ISL_673206 to 673225, EPI_ISL_673227 to 673230, EPI_ISL_673233 to 673234, EPI_ISL_673237 to 673244, EPI_ISL_673247 to 673253, EPI_ISL_673255 to 673262, EPI_ISL_673264 to 673266, EPI_ISL_673268 to 673269, EPI_ISL_673271 to 673272, EPI_ISL_673274, EPI_ISL_673276 to

673283, EPI_ISL_673285 to 673306, EPI_ISL_673308 to 673316, EPI_ISL_673318 to 673321, EPI_ISL_673324 to 673330, EPI_ISL_673332 to 673337, EPI_ISL_673339 to 673341, EPI_ISL_673345 to 673359, EPI_ISL_673361 to 673362, EPI_ISL_673364 to 673366, EPI_ISL_673368 to 673370, EPI_ISL_673372 to 673381, EPI_ISL_673383 to 673389, EPI_ISL_673391 to 673395, EPI_ISL_673397 to 673409, EPI_ISL_673411 to 673413, EPI_ISL_673415 to 673420, EPI_ISL_673422 to 673424, EPI_ISL_673426 to 673459, EPI_ISL_673461, EPI_ISL_673463 to 673464, EPI_ISL_673467 to 673476, EPI_ISL_673478 to 673480, EPI_ISL_673482 to 673483, EPI_ISL_673485, EPI_ISL_673488 to 673490, EPI_ISL_673492, EPI_ISL_673494 to 673499, EPI_ISL_673501, EPI_ISL_673503 to 673512, EPI_ISL_673514 to 673522, EPI_ISL_673524 to 673532, EPI_ISL_673534 to 673544, EPI_ISL_673546 to 673548, EPI_ISL_673550 to 673565, EPI_ISL_673567, EPI_ISL_673569 to 673570, EPI_ISL_673572 to 673576, EPI_ISL_673578 to 673591, EPI_ISL_673593, EPI_ISL_673595, EPI_ISL_673597 to 673598, EPI_ISL_673600, EPI_ISL_673602, EPI_ISL_673604 to 673611, EPI_ISL_673613 to 673615, EPI_ISL_673617 to 673618, EPI_ISL_673620, EPI_ISL_673622 to 673623, EPI_ISL_673626, EPI_ISL_673628 to 673632, EPI_ISL_673636, EPI_ISL_673639 to 673640, EPI_ISL_673643 to 673644, EPI_ISL_673647 to 673650, EPI_ISL_673653 to 673654, EPI_ISL_673656, EPI_ISL_673658, EPI_ISL_673660, EPI_ISL_673662, EPI_ISL_673664 to 673669, EPI_ISL_673671 to 673672, EPI_ISL_673674 to 673675, EPI_ISL_673678 to 673681, EPI_ISL_673683 to 673685, EPI_ISL_673687, EPI_ISL_673689 to 673690, EPI_ISL_673692 to 673694, EPI_ISL_673697 to 673701, EPI_ISL_673703 to 673707, EPI_ISL_673710 to 673713, EPI_ISL_673716 to 673721, EPI_ISL_673723 to 673725, EPI_ISL_673727 to 673728, EPI_ISL_673731 to 673732, EPI_ISL_673734, EPI_ISL_673736 to 673737, EPI_ISL_673739, EPI_ISL_673743 to 673744, EPI_ISL_673747 to 673748, EPI_ISL_673751 to 673756, EPI_ISL_673758 to 673759, EPI_ISL_673761 to 673763, EPI_ISL_673766, EPI_ISL_673768 to 673769, EPI_ISL_673771 to 673776, EPI_ISL_673779 to 673784, EPI_ISL_673786, EPI_ISL_673788 to 673789, EPI_ISL_673791 to 673795, EPI_ISL_673797, EPI_ISL_673800, EPI_ISL_673802, EPI_ISL_673805 to 673806, EPI_ISL_673808 to 673809, EPI_ISL_673811, EPI_ISL_673815 to 673820, EPI_ISL_673823 to 673826, EPI_ISL_673830 to 673831, EPI_ISL_673836 to 673839, EPI_ISL_673841, EPI_ISL_673843 to 673845, EPI_ISL_673847, EPI_ISL_673849 to 673850, EPI_ISL_673852, EPI_ISL_673854 to 673855, EPI_ISL_673858, EPI_ISL_673860 to 673861, EPI_ISL_673864, EPI_ISL_673868 to 673870, EPI_ISL_674826, EPI_ISL_674828, EPI_ISL_674834, EPI_ISL_674837 to 674838, EPI_ISL_674840, EPI_ISL_674845 to 674847, EPI_ISL_674849 to 674850, EPI_ISL_674859, EPI_ISL_674862, EPI_ISL_674867 to 674868, EPI_ISL_674873, EPI_ISL_674876, EPI_ISL_674883, EPI_ISL_674906, EPI_ISL_674908 to 674910, EPI_ISL_674917, EPI_ISL_674919 to 674920, EPI_ISL_674922 to 674924, EPI_ISL_674933, EPI_ISL_674936, EPI_ISL_674939 to 674940, EPI_ISL_674944, EPI_ISL_674946, EPI_ISL_674950, EPI_ISL_674952 to 674953, EPI_ISL_674957, EPI_ISL_674959, EPI_ISL_674962, EPI_ISL_674972 to 674973, EPI_ISL_674977, EPI_ISL_674983, EPI_ISL_674992, EPI_ISL_675006, EPI_ISL_675012, EPI_ISL_675018, EPI_ISL_675024, EPI_ISL_675026, EPI_ISL_675033, EPI_ISL_675038 to 675040, EPI_ISL_675042, EPI_ISL_675046, EPI_ISL_675053, EPI_ISL_675056, EPI_ISL_675064, EPI_ISL_675071, EPI_ISL_675075, EPI_ISL_675080, EPI_ISL_675082, EPI_ISL_675087, EPI_ISL_675104, EPI_ISL_675110, EPI_ISL_675117, EPI_ISL_675120, EPI_ISL_675126, EPI_ISL_676383, EPI_ISL_681657, EPI_ISL_681659, EPI_ISL_681671 to 681672, EPI_ISL_702025 to 702026, EPI_ISL_702028 to 702040, EPI_ISL_702042 to 702044, EPI_ISL_702046 to 702049, EPI_ISL_702051 to 702065, EPI_ISL_702067 to 702081, EPI_ISL_702084 to 702093, EPI_ISL_702096, EPI_ISL_702098 to 702102, EPI_ISL_702104 to 702132, EPI_ISL_702135 to 702138, EPI_ISL_702140, EPI_ISL_702142 to 702153, EPI_ISL_702155 to 702172, EPI_ISL_702174, EPI_ISL_702177 to 702179, EPI_ISL_702181 to 702195, EPI_ISL_702197 to 702207, EPI_ISL_702209 to 702221, EPI_ISL_702223 to 702235, EPI_ISL_702237, EPI_ISL_702239 to 702243, EPI_ISL_702245 to 702259, EPI_ISL_702261 to 702274, EPI_ISL_702276 to 702289, EPI_ISL_702291 to 702309, EPI_ISL_702311 to 702341, EPI_ISL_702343 to 702348, EPI_ISL_702350 to 702361, EPI_ISL_702363 to 702369, EPI_ISL_704688 to 704689, EPI_ISL_704691, EPI_ISL_704693 to 704694, EPI_ISL_704696 to 704697, EPI_ISL_704699 to 704700, EPI_ISL_704702, EPI_ISL_704704 to 704705, EPI_ISL_704707 to 704708, EPI_ISL_704710, EPI_ISL_704712 to 704713, EPI_ISL_704715, EPI_ISL_704717 to 704718, EPI_ISL_704721, EPI_ISL_704723 to 704724, EPI_ISL_704726 to 704727, EPI_ISL_704729 to 704730, EPI_ISL_704732, EPI_ISL_704735, EPI_ISL_704738, EPI_ISL_704740, EPI_ISL_704742 to 704743, EPI_ISL_704745, EPI_ISL_704747 to 704748, EPI_ISL_704750 to 704751, EPI_ISL_704753, EPI_ISL_704755 to 704756, EPI_ISL_704758, EPI_ISL_704760 to 704761, EPI_ISL_704763 to 704764, EPI_ISL_704766 to 704767, EPI_ISL_704769, EPI_ISL_704771, EPI_ISL_704773 to 704774, EPI_ISL_704776, EPI_ISL_704778 to 704779, EPI_ISL_704781 to 704782, EPI_ISL_704784 to 704785, EPI_ISL_704787, EPI_ISL_704790, EPI_ISL_704792 to 704793, EPI_ISL_704795 to 704796, EPI_ISL_704798, EPI_ISL_704800 to 704801, EPI_ISL_704803, EPI_ISL_704805 to 704806, EPI_ISL_704808, EPI_ISL_704810 to 704811, EPI_ISL_704813 to 704814, EPI_ISL_704816, EPI_ISL_704818 to 704819, EPI_ISL_704821 to 704822, EPI_ISL_704824 to 704825, EPI_ISL_704827, EPI_ISL_704829 to 704830, EPI_ISL_704832 to 704833, EPI_ISL_704835, EPI_ISL_704837, EPI_ISL_704840 to 704841, EPI_ISL_704843 to 704844, EPI_ISL_704846, EPI_ISL_704848 to 704849, EPI_ISL_704851 to 704852, EPI_ISL_704856 to 704857, EPI_ISL_704859 to 704860, EPI_ISL_704862 to 704863, EPI_ISL_704865, EPI_ISL_704867 to 704868, EPI_ISL_704873, EPI_ISL_704875 to 704876, EPI_ISL_704880 to 704881, EPI_ISL_704883 to 704884, EPI_ISL_704886, EPI_ISL_704888 to 704889, EPI_ISL_704891, EPI_ISL_704893 to 704894, EPI_ISL_704896, EPI_ISL_704898 to 704899, EPI_ISL_704901 to 704902, EPI_ISL_704904, EPI_ISL_704906 to 704907, EPI_ISL_704909 to 704910, EPI_ISL_704912 to 704913, EPI_ISL_704915, EPI_ISL_704917, EPI_ISL_704919 to 704920, EPI_ISL_704922, EPI_ISL_704924 to 704925, EPI_ISL_704927 to 704928, EPI_ISL_704930, EPI_ISL_704932 to 704933, EPI_ISL_704935 to 704936, EPI_ISL_704938, EPI_ISL_704940 to 704941, EPI_ISL_704943 to 704944, EPI_ISL_704946, EPI_ISL_704949, EPI_ISL_704951, EPI_ISL_704953, EPI_ISL_704956, EPI_ISL_704958 to 704959, EPI_ISL_704961 to 704962, EPI_ISL_704964, EPI_ISL_704966 to 704967, EPI_ISL_704969, EPI_ISL_704971 to 704972, EPI_ISL_704974 to 704975, EPI_ISL_704977, EPI_ISL_704979 to 704980, EPI_ISL_704982, EPI_ISL_704984 to 704985, EPI_ISL_704987 to 704988, EPI_ISL_704990, EPI_ISL_704992 to 704993, EPI_ISL_704995 to 704996, EPI_ISL_704998, EPI_ISL_705000 to 705001, EPI_ISL_705003, EPI_ISL_705006, EPI_ISL_705008 to 705009, EPI_ISL_705011 to 705012, EPI_ISL_705014, EPI_ISL_705016 to 705017, EPI_ISL_705019 to 705020, EPI_ISL_705022, EPI_ISL_705025, EPI_ISL_705027 to 705029, EPI_ISL_705031, EPI_ISL_705033 to 705034, EPI_ISL_705036 to 705037, EPI_ISL_705039, EPI_ISL_705041 to 705042, EPI_ISL_705044 to 705045, EPI_ISL_705047, EPI_ISL_705049 to 705050, EPI_ISL_705052 to 705053, EPI_ISL_705055, EPI_ISL_705057 to 705058, EPI_ISL_705060, EPI_ISL_705063, EPI_ISL_705065 to 705066, EPI_ISL_705068 to 705069, EPI_ISL_705071, EPI_ISL_705073 to 705074, EPI_ISL_705076, EPI_ISL_705079 to 705080, EPI_ISL_705082, EPI_ISL_705084 to 705085, EPI_ISL_705087, EPI_ISL_705089 to 705090, EPI_ISL_705092 to 705093, EPI_ISL_705095, EPI_ISL_705097, EPI_ISL_705100, EPI_ISL_705103, EPI_ISL_705106 to 705107, EPI_ISL_705109, EPI_ISL_705111 to 705112, EPI_ISL_705114, EPI_ISL_705117, EPI_ISL_705120, EPI_ISL_705122 to 705123, EPI_ISL_705125, EPI_ISL_705127 to 705128, EPI_ISL_705130 to 705131, EPI_ISL_705135 to 705136, EPI_ISL_705138 to 705139, EPI_ISL_705141 to 705142, EPI_ISL_705144, EPI_ISL_705146 to 705147, EPI_ISL_705149, EPI_ISL_705151 to 705152, EPI_ISL_705155, EPI_ISL_705157, EPI_ISL_705159 to 705160, EPI_ISL_705162, EPI_ISL_705164 to 705165, EPI_ISL_705167, EPI_ISL_705169 to 705170, EPI_ISL_705173, EPI_ISL_705175, EPI_ISL_705177 to 705178, EPI_ISL_705180 to 705181, EPI_ISL_705183, EPI_ISL_705185 to 705186, EPI_ISL_705188 to 705189, EPI_ISL_708992 to 708997, EPI_ISL_708999 to 709003, EPI_ISL_709006 to 709008, EPI_ISL_709011, EPI_ISL_709014 to 709015, EPI_ISL_709019 to 709021, EPI_ISL_709023 to 709030, EPI_ISL_709032, EPI_ISL_709035, EPI_ISL_709037 to 709039, EPI_ISL_709042 to 709043, EPI_ISL_709046, EPI_ISL_709049, EPI_ISL_709051 to 709053, EPI_ISL_709055, EPI_ISL_709057, EPI_ISL_709059, EPI_ISL_709061 to 709062, EPI_ISL_709064, EPI_ISL_709067 to 709076, EPI_ISL_709080 to 709081, EPI_ISL_709086 to 709088, EPI_ISL_709090, EPI_ISL_709093 to 709096, EPI_ISL_709099 to 709100, EPI_ISL_709102, EPI_ISL_709105 to 709106, EPI_ISL_709113 to 709118, EPI_ISL_709121 to 709125, EPI_ISL_709129 to 709130, EPI_ISL_709134, EPI_ISL_709136, EPI_ISL_709138, EPI_ISL_709143, EPI_ISL_709145 to 709147, EPI_ISL_709152 to 709158, EPI_ISL_709162 to 709165, EPI_ISL_709167, EPI_ISL_709169, EPI_ISL_709173, EPI_ISL_709175 to 709180, EPI_ISL_709185 to 709186, EPI_ISL_709188, EPI_ISL_709193 to 709197, EPI_ISL_709200, EPI_ISL_709204 to 709210, EPI_ISL_709212 to 709214, EPI_ISL_709217 to 709219, EPI_ISL_709223 to 709224, EPI_ISL_709226 to 709227, EPI_ISL_709229, EPI_ISL_709232 to 709233, EPI_ISL_709238, EPI_ISL_709241, EPI_ISL_709244, EPI_ISL_709246, EPI_ISL_709249 to 709251, EPI_ISL_709253 to 709257, EPI_ISL_709259 to 709262, EPI_ISL_709265, EPI_ISL_709267 to 709269, EPI_ISL_709272 to 709273, EPI_ISL_709275, EPI_ISL_709278 to 709281, EPI_ISL_709284, EPI_ISL_709286 to 709287, EPI_ISL_709293 to 709294, EPI_ISL_709804 to 709805, EPI_ISL_709807 to 709808, EPI_ISL_709810, EPI_ISL_709813 to 709814, EPI_ISL_709817, EPI_ISL_709821 to 709822, EPI_ISL_709825, EPI_ISL_709827, EPI_ISL_709829, EPI_ISL_709836 to 709837, EPI_ISL_709842 to 709844, EPI_ISL_709846, EPI_ISL_709854, EPI_ISL_709864 to 709865, EPI_ISL_709867, EPI_ISL_709869 to 709870, EPI_ISL_709873, EPI_ISL_709876 to 709878, EPI_ISL_709880, EPI_ISL_710821, EPI_ISL_710823 to 710829, EPI_ISL_710835 to 710836, EPI_ISL_710878, EPI_ISL_711023 to 711026, EPI_ISL_711038, EPI_ISL_718318 to 718325, EPI_ISL_718327 to 718335, EPI_ISL_718338 to 718361, EPI_ISL_718364, EPI_ISL_718366 to 718372, EPI_ISL_718374 to 718388, EPI_ISL_718390 to 718395, EPI_ISL_718397 to 718399, EPI_ISL_718401 to 718403, EPI_ISL_718405 to 718410, EPI_ISL_718413 to 718420, EPI_ISL_718422 to 718431, EPI_ISL_718433 to 718463, EPI_ISL_718465 to 718467, EPI_ISL_718469 to 718485, EPI_ISL_718487 to 718499, EPI_ISL_718501 to 718510, EPI_ISL_718512 to 718531, EPI_ISL_718533 to 718534, EPI_ISL_718536 to 718563, EPI_ISL_718565 to 718568, EPI_ISL_718570 to 718577, EPI_ISL_718580 to 718581, EPI_ISL_718583 to 718597, EPI_ISL_718601 to 718604, EPI_ISL_718607 to 718608, EPI_ISL_718612 to 718617, EPI_ISL_718619 to 718626, EPI_ISL_718628, EPI_ISL_718630, EPI_ISL_718632, EPI_ISL_718634, EPI_ISL_718636, EPI_ISL_718638, EPI_ISL_718642, EPI_ISL_718645 to 718646, EPI_ISL_718648 to 718649, EPI_ISL_718651, EPI_ISL_718654 to 718656, EPI_ISL_718659 to 718661, EPI_ISL_718667 to 718669, EPI_ISL_718671, EPI_ISL_718673, EPI_ISL_718677, EPI_ISL_718679 to 718681, EPI_ISL_718683 to 718685, EPI_ISL_718688 to 718690, EPI_ISL_718692 to 718693, EPI_ISL_718696 to 718699, EPI_ISL_718701 to 718704, EPI_ISL_718706, EPI_ISL_718708 to 718709, EPI_ISL_718717, EPI_ISL_718720, EPI_ISL_718722, EPI_ISL_718726 to 718727, EPI_ISL_718730 to 718734, EPI_ISL_718736 to 718737, EPI_ISL_718741, EPI_ISL_718743, EPI_ISL_718748 to 718749, EPI_ISL_718751 to 718753, EPI_ISL_718756, EPI_ISL_718758, EPI_ISL_718760, EPI_ISL_718763, EPI_ISL_718768, EPI_ISL_718770 to 718771, EPI_ISL_718775 to 718776, EPI_ISL_718778, EPI_ISL_718784 to 718785, EPI_ISL_718788, EPI_ISL_718790, EPI_ISL_718792 to 718794, EPI_ISL_718797, EPI_ISL_718800, EPI_ISL_718804 to 718805, EPI_ISL_718809 to 718810, EPI_ISL_718814 to 718818, EPI_ISL_718820, EPI_ISL_718822, EPI_ISL_718824 to 718826, EPI_ISL_718834 to 718836, EPI_ISL_718840, EPI_ISL_718842 to 718843, EPI_ISL_718845, EPI_ISL_718848, EPI_ISL_718851 to 718852, EPI_ISL_718855 to 718857, EPI_ISL_718860 to 718863, EPI_ISL_718867 to 718869, EPI_ISL_718874 to 718876, EPI_ISL_718879, EPI_ISL_718882 to 718884, EPI_ISL_718892 to 718893, EPI_ISL_718896 to 718900, EPI_ISL_718904 to 718906, EPI_ISL_718909 to 718910, EPI_ISL_718912 to 718913, EPI_ISL_718916, EPI_ISL_718918, EPI_ISL_718921, EPI_ISL_718924 to 718925, EPI_ISL_718927 to 718928, EPI_ISL_718931, EPI_ISL_718933 to 718934, EPI_ISL_718937 to 718938, EPI_ISL_718940 to 718942, EPI_ISL_718944, EPI_ISL_718946 to 718949, EPI_ISL_718951, EPI_ISL_718954, EPI_ISL_718956, EPI_ISL_718958, EPI_ISL_718960 to 718961, EPI_ISL_718965, EPI_ISL_718968 to 718969, EPI_ISL_718971 to 718973, EPI_ISL_718981 to 719015, EPI_ISL_719017 to 719030, EPI_ISL_719033 to 719036, EPI_ISL_719038 to 719040, EPI_ISL_719043 to 719045, EPI_ISL_719047 to 719070, EPI_ISL_719072, EPI_ISL_719074 to 719090, EPI_ISL_719093 to 719097, EPI_ISL_719099 to 719124, EPI_ISL_719126 to 719128, EPI_ISL_719130 to 719139, EPI_ISL_719141 to 719143, EPI_ISL_719145, EPI_ISL_719147 to 719155, EPI_ISL_719157, EPI_ISL_719159 to 719168, EPI_ISL_719170 to 719183, EPI_ISL_719185 to 719189, EPI_ISL_719191 to 719195, EPI_ISL_719197 to 719211, EPI_ISL_719213, EPI_ISL_719215 to 719216, EPI_ISL_719218 to 719224, EPI_ISL_719226 to 719242, EPI_ISL_719244 to 719252, EPI_ISL_719254 to 719256, EPI_ISL_719258 to 719260, EPI_ISL_719262 to 719276, EPI_ISL_719280 to 719306, EPI_ISL_719308 to 719310, EPI_ISL_719312 to 719320, EPI_ISL_719322 to 719323, EPI_ISL_719325 to 719333, EPI_ISL_719335 to 719338, EPI_ISL_719340 to 719341, EPI_ISL_719343 to 719344, EPI_ISL_719346, EPI_ISL_719348 to 719353, EPI_ISL_719355 to 719362, EPI_ISL_719365 to 719381, EPI_ISL_719383 to 719388, EPI_ISL_719390 to 719404, EPI_ISL_719406 to 719408, EPI_ISL_719410 to 719420, EPI_ISL_719424, EPI_ISL_719427 to 719432, EPI_ISL_719434 to 719442, EPI_ISL_719444 to 719464, EPI_ISL_719466 to 719491, EPI_ISL_719493, EPI_ISL_719495 to 719498, EPI_ISL_719500 to 719501, EPI_ISL_719503 to 719514, EPI_ISL_719516 to 719519, EPI_ISL_719522 to 719528, EPI_ISL_719530 to 719535, EPI_ISL_719537 to 719550, EPI_ISL_719552 to 719553, EPI_ISL_719555 to 719560, EPI_ISL_719562 to 719566, EPI_ISL_719568 to 719592, EPI_ISL_719594 to 719604, EPI_ISL_728299, EPI_ISL_733596, EPI_ISL_733599, EPI_ISL_733601 to 733602, EPI_ISL_733604, EPI_ISL_733608, EPI_ISL_733610, EPI_ISL_733614 to 733615, EPI_ISL_733619, EPI_ISL_733622, EPI_ISL_733624, EPI_ISL_733626 to 733627, EPI_ISL_733629, EPI_ISL_733631 to 733632, EPI_ISL_733634, EPI_ISL_733637, EPI_ISL_733640, EPI_ISL_733649 to 733651, EPI_ISL_733655 to 733656, EPI_ISL_733660 to 733663, EPI_ISL_733665, EPI_ISL_733668, EPI_ISL_733670 to 733672, EPI_ISL_733674 to 733677, EPI_ISL_733680 to 733681, EPI_ISL_733684, EPI_ISL_733689, EPI_ISL_733691, EPI_ISL_733695 to 733696, EPI_ISL_733698, EPI_ISL_733704 to 733705, EPI_ISL_733709, EPI_ISL_733714 to 733717, EPI_ISL_733719 to 733723, EPI_ISL_733726 to 733727, EPI_ISL_733729, EPI_ISL_733732 to 733733, EPI_ISL_733738, EPI_ISL_733740 to 733741, EPI_ISL_733744 to 733745, EPI_ISL_733751 to 733753, EPI_ISL_733758, EPI_ISL_733763, EPI_ISL_733765, EPI_ISL_733767, EPI_ISL_733770, EPI_ISL_733772 to 733775, EPI_ISL_733778 to 733779, EPI_ISL_733782, EPI_ISL_733784, EPI_ISL_733787 to 733788, EPI_ISL_733790 to 733793, EPI_ISL_733799, EPI_ISL_733802, EPI_ISL_733804 to 733805, EPI_ISL_733809 to 733810, EPI_ISL_733812, EPI_ISL_733814, EPI_ISL_733816 to 733817, EPI_ISL_733821 to 733824, EPI_ISL_733834 to 733835, EPI_ISL_733839, EPI_ISL_733841, EPI_ISL_733846 to 733847, EPI_ISL_733849, EPI_ISL_733852 to 733854, EPI_ISL_733857, EPI_ISL_733860, EPI_ISL_733862, EPI_ISL_733868 to 733871, EPI_ISL_733876, EPI_ISL_733878, EPI_ISL_733883, EPI_ISL_733886, EPI_ISL_733888, EPI_ISL_733892, EPI_ISL_733895 to 733896, EPI_ISL_733898 to 733899, EPI_ISL_733906 to 733910, EPI_ISL_733915, EPI_ISL_756384, EPI_ISL_756386, EPI_ISL_756389, EPI_ISL_756395, EPI_ISL_756409, EPI_ISL_756413, EPI_ISL_756415 to 756418, EPI_ISL_756420, EPI_ISL_756422, EPI_ISL_756424, EPI_ISL_756426, EPI_ISL_756429, EPI_ISL_756431 to 756432, EPI_ISL_756434, EPI_ISL_756438, EPI_ISL_756442, EPI_ISL_756444, EPI_ISL_756446, EPI_ISL_756448, EPI_ISL_756453 to 756454, EPI_ISL_756456, EPI_ISL_756460, EPI_ISL_756467, EPI_ISL_756469, EPI_ISL_756474 to 756475, EPI_ISL_756478, EPI_ISL_756480 to 756481, EPI_ISL_756484 to 756485, EPI_ISL_756490 to 756491, EPI_ISL_756496 to 756497, EPI_ISL_756499, EPI_ISL_756501 to 756502, EPI_ISL_756505 to 756508, EPI_ISL_756511 to 756513, EPI_ISL_756517 to 756519, EPI_ISL_756521 to 756522, EPI_ISL_756525, EPI_ISL_756528, EPI_ISL_756534 to 756535, EPI_ISL_756540, EPI_ISL_756542, EPI_ISL_756545, EPI_ISL_756549, EPI_ISL_756554, EPI_ISL_756558 to 756559, EPI_ISL_756567 to 756568, EPI_ISL_756570, EPI_ISL_756572, EPI_ISL_756576 to 756577, EPI_ISL_756579, EPI_ISL_756582, EPI_ISL_756585 to 756586, EPI_ISL_756588 to 756591, EPI_ISL_756594, EPI_ISL_756597, EPI_ISL_756604 to 756608, EPI_ISL_756610 to 756611, EPI_ISL_756613, EPI_ISL_756615, EPI_ISL_756617, EPI_ISL_756621 to 756623, EPI_ISL_756625, EPI_ISL_756628, EPI_ISL_756630 to 756631, EPI_ISL_756637, EPI_ISL_756642, EPI_ISL_756649, EPI_ISL_756653, EPI_ISL_756655, EPI_ISL_756659 to 756660, EPI_ISL_756665 to 756668, EPI_ISL_756671 to 756672, EPI_ISL_756674 to 756675, EPI_ISL_756677 to 756680, EPI_ISL_757119, EPI_ISL_760813, EPI_ISL_760822, EPI_ISL_760833, EPI_ISL_760858 to 760859, EPI_ISL_760894, EPI_ISL_760920, EPI_ISL_760922, EPI_ISL_760932, EPI_ISL_760936, EPI_ISL_760939, EPI_ISL_760969, EPI_ISL_760976, EPI_ISL_761017, EPI_ISL_761021, EPI_ISL_761057, EPI_ISL_761074 to 761075, EPI_ISL_761922, EPI_ISL_761927, EPI_ISL_761929, EPI_ISL_761935, EPI_ISL_761937, EPI_ISL_761939, EPI_ISL_761946, EPI_ISL_761952 to 761953, EPI_ISL_761956, EPI_ISL_761961, EPI_ISL_761967 to 761968, EPI_ISL_761970, EPI_ISL_761979, EPI_ISL_761981, EPI_ISL_761994 to 761995, EPI_ISL_761997, EPI_ISL_761999, EPI_ISL_762007 to 762009, EPI_ISL_762014 to 762016, EPI_ISL_762018 to 762019, EPI_ISL_762023 to 762030, EPI_ISL_762036 to 762038, EPI_ISL_762041 to 762043, EPI_ISL_762045, EPI_ISL_762050, EPI_ISL_762057, EPI_ISL_762059 to 762061, EPI_ISL_762063 to 762067, EPI_ISL_762069, EPI_ISL_762072 to 762075, EPI_ISL_762078, EPI_ISL_762085, EPI_ISL_762087, EPI_ISL_762089 to 762091, EPI_ISL_762096 to 762098, EPI_ISL_762103, EPI_ISL_762109, EPI_ISL_762112 to 762114, EPI_ISL_762117 to 762118, EPI_ISL_762122, EPI_ISL_762124 to 762126, EPI_ISL_762131, EPI_ISL_762133, EPI_ISL_762135 to 762140, EPI_ISL_762142, EPI_ISL_762146 to 762148, EPI_ISL_762152 to 762153, EPI_ISL_762155, EPI_ISL_762163, EPI_ISL_762166, EPI_ISL_762173, EPI_ISL_762175, EPI_ISL_762178, EPI_ISL_762181, EPI_ISL_762183, EPI_ISL_762186 to 762187, EPI_ISL_762189, EPI_ISL_762192 to 762193, EPI_ISL_762195, EPI_ISL_762546, EPI_ISL_762549 to 762553, EPI_ISL_762559, EPI_ISL_762563, EPI_ISL_762565 to 762566, EPI_ISL_762568 to 762572, EPI_ISL_762575, EPI_ISL_762577 to 762580, EPI_ISL_762584 to 762585, EPI_ISL_762587, EPI_ISL_762590 to 762596, EPI_ISL_762598, EPI_ISL_762600 to 762601, EPI_ISL_762603, EPI_ISL_762607 to 762608, EPI_ISL_762611 to 762613, EPI_ISL_762615 to 762616, EPI_ISL_762619, EPI_ISL_762623 to 762624, EPI_ISL_762626 to 762627, EPI_ISL_762632, EPI_ISL_762635 to 762637, EPI_ISL_762640, EPI_ISL_762645, EPI_ISL_762649 to 762651, EPI_ISL_762658, EPI_ISL_762660, EPI_ISL_762664 to 762665, EPI_ISL_762667 to 762668, EPI_ISL_762670, EPI_ISL_762673, EPI_ISL_762675, EPI_ISL_762677, EPI_ISL_762679 to 762682, EPI_ISL_762684 to 762685, EPI_ISL_762687 to 762689, EPI_ISL_762695 to 762697, EPI_ISL_762699 to 762700, EPI_ISL_762705, EPI_ISL_762713, EPI_ISL_762715 to 762719, EPI_ISL_762721, EPI_ISL_762723, EPI_ISL_762725, EPI_ISL_762727, EPI_ISL_762729, EPI_ISL_762736 to 762738, EPI_ISL_762740 to 762742, EPI_ISL_762745, EPI_ISL_762747 to 762748, EPI_ISL_762751 to 762753, EPI_ISL_762755, EPI_ISL_762757, EPI_ISL_762762, EPI_ISL_762767, EPI_ISL_762769, EPI_ISL_762772, EPI_ISL_762774, EPI_ISL_762778, EPI_ISL_762780 to 762784, EPI_ISL_762790, EPI_ISL_762792, EPI_ISL_762795, EPI_ISL_762797, EPI_ISL_762800, EPI_ISL_762803 to 762808, EPI_ISL_762811, EPI_ISL_762818, EPI_ISL_762820, EPI_ISL_762822, EPI_ISL_762825, EPI_ISL_762827, EPI_ISL_762829, EPI_ISL_762831, EPI_ISL_762833 to 762834, EPI_ISL_762836, EPI_ISL_762838, EPI_ISL_762840, EPI_ISL_762842, EPI_ISL_762848, EPI_ISL_762852, EPI_ISL_762855, EPI_ISL_762858, EPI_ISL_767115, EPI_ISL_767138, EPI_ISL_768845, EPI_ISL_768901 to 768903, EPI_ISL_768905 to 768913, EPI_ISL_768915 to 768930, EPI_ISL_768932 to 768933, EPI_ISL_768935 to 768950, EPI_ISL_768952 to 768971, EPI_ISL_768973 to 768987, EPI_ISL_768989 to 769003, EPI_ISL_769005 to 769027, EPI_ISL_769030 to 769033, EPI_ISL_769035 to 769037, EPI_ISL_769039 to 769048, EPI_ISL_769050 to 769053, EPI_ISL_769056 to 769060, EPI_ISL_769062 to 769064, EPI_ISL_769066 to 769067, EPI_ISL_769069 to 769070, EPI_ISL_769072 to 769076, EPI_ISL_769078 to 769084, EPI_ISL_769086 to 769106, EPI_ISL_769108 to 769113, EPI_ISL_769115 to 769136, EPI_ISL_769138 to 769141, EPI_ISL_769145 to 769160, EPI_ISL_769162 to 769177, EPI_ISL_769179 to 769190, EPI_ISL_769192 to 769201, EPI_ISL_769203 to 769204, EPI_ISL_769206 to 769222, EPI_ISL_777133, EPI_ISL_777173, EPI_ISL_777201, EPI_ISL_777218, EPI_ISL_777275, EPI_ISL_777289, EPI_ISL_777293, EPI_ISL_777323, EPI_ISL_777328, EPI_ISL_777330 to 777332, EPI_ISL_777334, EPI_ISL_777339, EPI_ISL_777342 to 777345, EPI_ISL_777347 to 777348, EPI_ISL_777350 to 777352, EPI_ISL_777354 to 777355, EPI_ISL_777357, EPI_ISL_777359 to 777360, EPI_ISL_777362, EPI_ISL_777365 to 777368, EPI_ISL_777370 to 777371, EPI_ISL_777374, EPI_ISL_777378 to 777379, EPI_ISL_777381, EPI_ISL_777384, EPI_ISL_777387 to 777388, EPI_ISL_777390 to 777391, EPI_ISL_777396, EPI_ISL_777398, EPI_ISL_777400 to 777401, EPI_ISL_777406, EPI_ISL_777408 to 777409, EPI_ISL_777413 to 777415, EPI_ISL_777418 to 777419, EPI_ISL_777422, EPI_ISL_777426, EPI_ISL_777428 to 777429, EPI_ISL_777431, EPI_ISL_777433 to 777434, EPI_ISL_777439, EPI_ISL_777442, EPI_ISL_777445 to 777446, EPI_ISL_777452 to 777454, EPI_ISL_777456 to 777457, EPI_ISL_777460, EPI_ISL_777464, EPI_ISL_777469, EPI_ISL_777473, EPI_ISL_777476, EPI_ISL_777478, EPI_ISL_777480, EPI_ISL_777482 to 777483, EPI_ISL_777486, EPI_ISL_777492, EPI_ISL_777494, EPI_ISL_777499 to 777501, EPI_ISL_777503, EPI_ISL_777507 to 777508, EPI_ISL_777511 to 777513, EPI_ISL_777516 to 777517, EPI_ISL_777519, EPI_ISL_777521, EPI_ISL_777528, EPI_ISL_777536, EPI_ISL_777541 to 777542, EPI_ISL_777544 to 777547, EPI_ISL_777549, EPI_ISL_777551, EPI_ISL_777553, EPI_ISL_777555, EPI_ISL_777559, EPI_ISL_777565 to 777566, EPI_ISL_777570 to 777571, EPI_ISL_777575, EPI_ISL_777577 to 777578, EPI_ISL_777581, EPI_ISL_777585 to 777586, EPI_ISL_777588, EPI_ISL_777590, EPI_ISL_777594, EPI_ISL_777596, EPI_ISL_777598 to 777599, EPI_ISL_777603 to 777604, EPI_ISL_777606 to 777609, EPI_ISL_777611, EPI_ISL_777613 to 777620, EPI_ISL_777624, EPI_ISL_777626, EPI_ISL_777632, EPI_ISL_777636 to 777637, EPI_ISL_777640, EPI_ISL_777642 to 777643, EPI_ISL_777645, EPI_ISL_777647, EPI_ISL_777652, EPI_ISL_777656, EPI_ISL_777661, EPI_ISL_777664 to 777666, EPI_ISL_777668, EPI_ISL_777673 to 777675, EPI_ISL_777677 to 777689, EPI_ISL_777691 to 777700, EPI_ISL_777702 to 777705, EPI_ISL_777707 to 777718, EPI_ISL_777720 to 777736, EPI_ISL_777738 to 777740, EPI_ISL_777743 to 777745, EPI_ISL_777747 to 777749, EPI_ISL_777751 to 777763, EPI_ISL_777765, EPI_ISL_777767, EPI_ISL_777769 to 777771, EPI_ISL_777775 to 777777, EPI_ISL_777779, EPI_ISL_777781 to 777789, EPI_ISL_777791 to 777792, EPI_ISL_777794 to 777803, EPI_ISL_777805 to 777815, EPI_ISL_777817, EPI_ISL_777820 to 777822, EPI_ISL_777824 to 777838, EPI_ISL_777840 to 777847, EPI_ISL_777849, EPI_ISL_777851 to 777854, EPI_ISL_777856 to 777863, EPI_ISL_777865 to 777874, EPI_ISL_777876 to 777886, EPI_ISL_777888 to 777891, EPI_ISL_777894 to 777901, EPI_ISL_777903 to 777911, EPI_ISL_777913 to 777916, EPI_ISL_777918 to 777920, EPI_ISL_777922 to 777923, EPI_ISL_777927 to 777937, EPI_ISL_777939 to 777990, EPI_ISL_777992 to 777999, EPI_ISL_778002, EPI_ISL_778005 to 778006, EPI_ISL_778010 to 778011, EPI_ISL_778013, EPI_ISL_778015, EPI_ISL_778017, EPI_ISL_778019 to 778020, EPI_ISL_778022 to 778023, EPI_ISL_778025, EPI_ISL_778027 to 778028, EPI_ISL_778030, EPI_ISL_778032, EPI_ISL_778038 to 778039, EPI_ISL_778041, EPI_ISL_778045, EPI_ISL_778049, EPI_ISL_778054, EPI_ISL_778056 to 778057, EPI_ISL_778063, EPI_ISL_778066 to 778067, EPI_ISL_778072, EPI_ISL_778075, EPI_ISL_778078 to 778079, EPI_ISL_778081, EPI_ISL_778084 to 778087, EPI_ISL_778091, EPI_ISL_778094 to 778097, EPI_ISL_778099 to 778100, EPI_ISL_778103, EPI_ISL_778110, EPI_ISL_778112, EPI_ISL_778114 to 778115, EPI_ISL_778120, EPI_ISL_778123, EPI_ISL_778125 to 778126, EPI_ISL_778130, EPI_ISL_778134 to 778135, EPI_ISL_778137 to 778139, EPI_ISL_778141 to 778143, EPI_ISL_778147 to 778148, EPI_ISL_778152, EPI_ISL_778157,

EPI_ISL_778160 to 778162, EPI_ISL_778169 to 778170, EPI_ISL_778172 to 778175, EPI_ISL_778177 to 778182, EPI_ISL_778185 to 778186, EPI_ISL_778188 to 778189, EPI_ISL_778194, EPI_ISL_778197 to 778200, EPI_ISL_778204 to 778206, EPI_ISL_778208, EPI_ISL_778210 to 778211, EPI_ISL_778213 to 778214, EPI_ISL_778217 to 778218, EPI_ISL_778220 to 778221, EPI_ISL_778223 to 778224, EPI_ISL_778226, EPI_ISL_778231 to 778232, EPI_ISL_778235, EPI_ISL_778237, EPI_ISL_778241 to 778243, EPI_ISL_778251, EPI_ISL_778253 to 778255, EPI_ISL_778257, EPI_ISL_778262, EPI_ISL_778265, EPI_ISL_778267, EPI_ISL_778271, EPI_ISL_778273, EPI_ISL_778275, EPI_ISL_778277 to 778278, EPI_ISL_778280, EPI_ISL_778284 to 778285, EPI_ISL_778291, EPI_ISL_778293 to 778294, EPI_ISL_778296, EPI_ISL_781183, EPI_ISL_781185, EPI_ISL_781187 to 781190, EPI_ISL_781199, EPI_ISL_781220 to 781221, EPI_ISL_781223, EPI_ISL_781248 to 781249, EPI_ISL_781251, EPI_ISL_781258, EPI_ISL_781260, EPI_ISL_781269, EPI_ISL_781281, EPI_ISL_781287, EPI_ISL_781299, EPI_ISL_781304, EPI_ISL_781306, EPI_ISL_781308, EPI_ISL_781325, EPI_ISL_781327, EPI_ISL_781333, EPI_ISL_781340 to 781342, EPI_ISL_781345, EPI_ISL_781354 to 781355, EPI_ISL_781358 to 781360, EPI_ISL_781364, EPI_ISL_781367 to 781368, EPI_ISL_781371, EPI_ISL_781376 to 781377, EPI_ISL_781379, EPI_ISL_781385, EPI_ISL_781396, EPI_ISL_781401, EPI_ISL_781407, EPI_ISL_781413, EPI_ISL_781426, EPI_ISL_781431, EPI_ISL_781434, EPI_ISL_781436, EPI_ISL_781438, EPI_ISL_781440 to 781442, EPI_ISL_781448, EPI_ISL_781466, EPI_ISL_781471 to 781472, EPI_ISL_781480 to 781481, EPI_ISL_781483, EPI_ISL_781488 to 781490, EPI_ISL_781497 to 781498, EPI_ISL_781506, EPI_ISL_781512 to 781514, EPI_ISL_781517 to 781521, EPI_ISL_781523 to 781526, EPI_ISL_781528, EPI_ISL_781530 to 781535, EPI_ISL_781538, EPI_ISL_781540, EPI_ISL_781542 to 781543, EPI_ISL_781545 to 781546, EPI_ISL_781549 to 781553, EPI_ISL_781557, EPI_ISL_781559 to 781566, EPI_ISL_781568 to 781575, EPI_ISL_781577, EPI_ISL_781580 to 781581, EPI_ISL_781583 to 781596, EPI_ISL_781600 to 781604, EPI_ISL_781606 to 781608, EPI_ISL_781610, EPI_ISL_781612 to 781614, EPI_ISL_781616, EPI_ISL_781620, EPI_ISL_781622, EPI_ISL_781625 to 781626, EPI_ISL_781628 to 781630, EPI_ISL_781633 to 781635, EPI_ISL_781637 to 781641, EPI_ISL_781643 to 781648, EPI_ISL_781650, EPI_ISL_781653 to 781654, EPI_ISL_781656 to 781660, EPI_ISL_781662, EPI_ISL_781664, EPI_ISL_781666 to 781668, EPI_ISL_781670 to 781673, EPI_ISL_781675 to 781676, EPI_ISL_781678 to 781679, EPI_ISL_781682 to 781683, EPI_ISL_781687 to 781694, EPI_ISL_781696 to 781698, EPI_ISL_781700, EPI_ISL_781702 to 781719, EPI_ISL_781722 to 781723, EPI_ISL_781726 to 781729, EPI_ISL_781731, EPI_ISL_781733 to 781734, EPI_ISL_781736 to 781741, EPI_ISL_781744 to 781745, EPI_ISL_781748, EPI_ISL_781750 to 781752, EPI_ISL_781755 to 781760, EPI_ISL_781762, EPI_ISL_781764 to 781766, EPI_ISL_781768 to 781769, EPI_ISL_781771 to 781772, EPI_ISL_781774 to 781776, EPI_ISL_781778 to 781782, EPI_ISL_781786 to 781787, EPI_ISL_781790 to 781794, EPI_ISL_781796 to 781797, EPI_ISL_781799 to 781800, EPI_ISL_781802 to 781806, EPI_ISL_781808 to 781831, EPI_ISL_781833, EPI_ISL_781835 to 781836, EPI_ISL_781838, EPI_ISL_781842 to 781850, EPI_ISL_781852 to 781853, EPI_ISL_781855 to 781857, EPI_ISL_781859 to 781861, EPI_ISL_781865 to 781868, EPI_ISL_782739, EPI_ISL_782746, EPI_ISL_782757, EPI_ISL_782759, EPI_ISL_782761, EPI_ISL_782763, EPI_ISL_782769, EPI_ISL_782776 to 782777, EPI_ISL_782780, EPI_ISL_782788, EPI_ISL_782796, EPI_ISL_782800, EPI_ISL_782803, EPI_ISL_782809 to 782810, EPI_ISL_782814, EPI_ISL_782816 to 782818, EPI_ISL_782822, EPI_ISL_782830, EPI_ISL_782834, EPI_ISL_782840, EPI_ISL_782842, EPI_ISL_782846, EPI_ISL_782850, EPI_ISL_782853, EPI_ISL_782857, EPI_ISL_782860, EPI_ISL_782862, EPI_ISL_782866 to 782868, EPI_ISL_782880, EPI_ISL_782882 to 782883, EPI_ISL_782885 to 782887, EPI_ISL_782889 to 782890, EPI_ISL_782907 to 782910, EPI_ISL_782912 to 782913, EPI_ISL_782920, EPI_ISL_782923, EPI_ISL_782928 to 782930, EPI_ISL_782932, EPI_ISL_782941 to 782942, EPI_ISL_782947, EPI_ISL_782954, EPI_ISL_782959 to 782960, EPI_ISL_782966, EPI_ISL_782974, EPI_ISL_782979, EPI_ISL_782984 to 782985, EPI_ISL_782987, EPI_ISL_782991, EPI_ISL_782996, EPI_ISL_782998, EPI_ISL_783012, EPI_ISL_783019, EPI_ISL_783023, EPI_ISL_783028, EPI_ISL_783031, EPI_ISL_783036 to 783038, EPI_ISL_783042, EPI_ISL_783051, EPI_ISL_783055, EPI_ISL_783057, EPI_ISL_783066, EPI_ISL_783367, EPI_ISL_783369, EPI_ISL_783371 to 783373, EPI_ISL_783375 to 783381, EPI_ISL_783383 to 783390, EPI_ISL_783392 to 783399, EPI_ISL_783401 to 783402, EPI_ISL_783404 to 783416, EPI_ISL_783418 to 783423, EPI_ISL_783425 to 783442, EPI_ISL_783444 to 783457, EPI_ISL_783459 to 783484, EPI_ISL_783486, EPI_ISL_783489 to 783496, EPI_ISL_783499 to 783501, EPI_ISL_783503 to 783529, EPI_ISL_783531 to 783535, EPI_ISL_797397 to 797398, EPI_ISL_797401, EPI_ISL_797403 to 797404, EPI_ISL_797406 to 797407, EPI_ISL_797410, EPI_ISL_797412, EPI_ISL_797414 to 797415, EPI_ISL_797417, EPI_ISL_797419, EPI_ISL_797423, EPI_ISL_797425 to 797427, EPI_ISL_797429, EPI_ISL_797431, EPI_ISL_797433 to 797434, EPI_ISL_797436 to 797437, EPI_ISL_797439 to 797440, EPI_ISL_797442 to 797444, EPI_ISL_797446 to 797448, EPI_ISL_797450, EPI_ISL_797452, EPI_ISL_797454, EPI_ISL_797457 to 797458, EPI_ISL_797460, EPI_ISL_797463 to 797465, EPI_ISL_797467, EPI_ISL_797469 to 797474, EPI_ISL_797476 to 797479, EPI_ISL_797481 to 797483, EPI_ISL_797485 to 797488, EPI_ISL_797490, EPI_ISL_797492 to 797498, EPI_ISL_797501, EPI_ISL_797503, EPI_ISL_797505 to 797508, EPI_ISL_797511, EPI_ISL_797513, EPI_ISL_797515 to 797530, EPI_ISL_797532, EPI_ISL_797534, EPI_ISL_797536 to 797539, EPI_ISL_797542, EPI_ISL_797544 to 797546, EPI_ISL_797548, EPI_ISL_797550 to 797552, EPI_ISL_797555, EPI_ISL_797557 to 797558, EPI_ISL_797561 to 797562, EPI_ISL_797565 to 797567, EPI_ISL_797569 to 797572, EPI_ISL_797574, EPI_ISL_797577 to 797585, EPI_ISL_797589 to 797590, EPI_ISL_797592 to 797598, EPI_ISL_797600 to 797601, EPI_ISL_797603 to 797604, EPI_ISL_797607 to 797609, EPI_ISL_797611 to 797612, EPI_ISL_797614, EPI_ISL_797617 to 797618, EPI_ISL_797622 to 797623, EPI_ISL_797625 to 797626, EPI_ISL_797628 to 797630, EPI_ISL_797633 to 797634, EPI_ISL_797636, EPI_ISL_797638, EPI_ISL_797643, EPI_ISL_797645, EPI_ISL_797647 to 797651, EPI_ISL_797653, EPI_ISL_797656 to 797657, EPI_ISL_797659, EPI_ISL_797661 to 797662, EPI_ISL_797664 to 797665, EPI_ISL_797667 to 797672, EPI_ISL_797675 to 797676, EPI_ISL_797679 to 797684, EPI_ISL_797687 to 797697, EPI_ISL_797700, EPI_ISL_797702 to 797703, EPI_ISL_797705 to 797708, EPI_ISL_797710 to 797711, EPI_ISL_798000, EPI_ISL_798003, EPI_ISL_798018, EPI_ISL_798032, EPI_ISL_798040, EPI_ISL_798046, EPI_ISL_798048, EPI_ISL_798053, EPI_ISL_798055, EPI_ISL_798058, EPI_ISL_798060, EPI_ISL_798069, EPI_ISL_798072, EPI_ISL_798076 to 798079, EPI_ISL_798086, EPI_ISL_798088, EPI_ISL_798093 to 798095, EPI_ISL_798097, EPI_ISL_798105, EPI_ISL_798110 to 798113, EPI_ISL_798115, EPI_ISL_798117, EPI_ISL_798121, EPI_ISL_798126, EPI_ISL_798128, EPI_ISL_798130, EPI_ISL_798132, EPI_ISL_798137, EPI_ISL_798140, EPI_ISL_798143, EPI_ISL_798145, EPI_ISL_798148 to 798149, EPI_ISL_798151, EPI_ISL_798153, EPI_ISL_798160 to 798161, EPI_ISL_798166 to 798168, EPI_ISL_798183, EPI_ISL_798185 to 798186, EPI_ISL_798193, EPI_ISL_798197, EPI_ISL_798199, EPI_ISL_798201, EPI_ISL_798203, EPI_ISL_798211, EPI_ISL_798213 to 798214, EPI_ISL_798216 to 798218, EPI_ISL_798224, EPI_ISL_798230, EPI_ISL_798243 to 798244, EPI_ISL_798251 to 798254, EPI_ISL_798256, EPI_ISL_798258, EPI_ISL_798266, EPI_ISL_798270, EPI_ISL_798272 to 798273, EPI_ISL_798281, EPI_ISL_798288 to 798289, EPI_ISL_798294, EPI_ISL_798298, EPI_ISL_798300, EPI_ISL_798307, EPI_ISL_798311 to 798312, EPI_ISL_798315 to 798316, EPI_ISL_798319, EPI_ISL_798322, EPI_ISL_798326 to 798327, EPI_ISL_798664, EPI_ISL_798673 to 798674, EPI_ISL_798677 to 798679, EPI_ISL_798681 to 798683, EPI_ISL_798685 to 798687, EPI_ISL_798694, EPI_ISL_798697 to 798700, EPI_ISL_798703, EPI_ISL_798706, EPI_ISL_798708, EPI_ISL_798711, EPI_ISL_798713 to 798715, EPI_ISL_798717 to 798718, EPI_ISL_798723, EPI_ISL_798726, EPI_ISL_798730 to 798734, EPI_ISL_798736 to 798737, EPI_ISL_798743, EPI_ISL_798745 to 798748, EPI_ISL_798751, EPI_ISL_798753, EPI_ISL_798756 to 798763, EPI_ISL_798765 to 798766, EPI_ISL_798772 to 798773, EPI_ISL_798775, EPI_ISL_798778, EPI_ISL_798780, EPI_ISL_798782 to 798783, EPI_ISL_798785 to 798790, EPI_ISL_798792, EPI_ISL_798794 to 798797, EPI_ISL_798799 to 798800, EPI_ISL_798802, EPI_ISL_798805 to 798807, EPI_ISL_798810, EPI_ISL_798813, EPI_ISL_798815 to 798816, EPI_ISL_798818, EPI_ISL_798820, EPI_ISL_798825, EPI_ISL_798827 to 798829, EPI_ISL_798831, EPI_ISL_798834 to 798836, EPI_ISL_798839 to 798843, EPI_ISL_798846 to 798849, EPI_ISL_798854, EPI_ISL_798857 to 798862, EPI_ISL_798865 to 798868, EPI_ISL_798871, EPI_ISL_798873, EPI_ISL_798877, EPI_ISL_798881, EPI_ISL_798883, EPI_ISL_798886 to 798887, EPI_ISL_798890, EPI_ISL_798893 to 798897, EPI_ISL_798899 to 798901, EPI_ISL_798903 to 798904, EPI_ISL_798907 to 798909, EPI_ISL_798912 to 798913, EPI_ISL_798916 to 798918, EPI_ISL_798920, EPI_ISL_798923, EPI_ISL_798927, EPI_ISL_798929, EPI_ISL_798931 to 798932, EPI_ISL_798937 to 798938, EPI_ISL_798940, EPI_ISL_798943, EPI_ISL_798945 to 798947, EPI_ISL_798949, EPI_ISL_798953 to 798954, EPI_ISL_798956, EPI_ISL_798958 to 798960, EPI_ISL_798965, EPI_ISL_798971, EPI_ISL_798974, EPI_ISL_798976, EPI_ISL_798978, EPI_ISL_798980, EPI_ISL_798982, EPI_ISL_798985, EPI_ISL_798989 to 798990, EPI_ISL_798992 to 798993, EPI_ISL_798997 to 798999, EPI_ISL_799001 to 799002, EPI_ISL_799007 to 799008, EPI_ISL_799017 to 799018, EPI_ISL_799020, EPI_ISL_799031 to 799032, EPI_ISL_799035 to 799037, EPI_ISL_799039, EPI_ISL_799048, EPI_ISL_799052, EPI_ISL_799058, EPI_ISL_799061, EPI_ISL_799074, EPI_ISL_799076, EPI_ISL_799078, EPI_ISL_799083, EPI_ISL_799087 to 799088, EPI_ISL_799091 to 799095, EPI_ISL_799097, EPI_ISL_799109 to 799110, EPI_ISL_799116, EPI_ISL_799119, EPI_ISL_799121, EPI_ISL_799128, EPI_ISL_799133, EPI_ISL_799135 to 799136, EPI_ISL_799141 to 799142, EPI_ISL_799146, EPI_ISL_799148, EPI_ISL_799152 to 799153, EPI_ISL_799159, EPI_ISL_799173 to 799174, EPI_ISL_799181, EPI_ISL_799184 to 799186, EPI_ISL_799195, EPI_ISL_799206, EPI_ISL_799208, EPI_ISL_799210 to 799211, EPI_ISL_799217, EPI_ISL_799220, EPI_ISL_799223, EPI_ISL_799225 to 799226, EPI_ISL_799238, EPI_ISL_799241 to 799242, EPI_ISL_799247, EPI_ISL_799251, EPI_ISL_799253 to 799254, EPI_ISL_799259, EPI_ISL_799284, EPI_ISL_799291 to 799292, EPI_ISL_799295, EPI_ISL_799935 to 799936, EPI_ISL_799939, EPI_ISL_799944, EPI_ISL_799947, EPI_ISL_799950, EPI_ISL_799952 to 799953, EPI_ISL_799955, EPI_ISL_799957 to 799960, EPI_ISL_799962 to 799963, EPI_ISL_799972, EPI_ISL_799974, EPI_ISL_799988, EPI_ISL_799993 to 799994, EPI_ISL_799996, EPI_ISL_800004, EPI_ISL_800006 to 800008, EPI_ISL_800012, EPI_ISL_800018, EPI_ISL_800024, EPI_ISL_800026, EPI_ISL_800028, EPI_ISL_800037, EPI_ISL_800045, EPI_ISL_800047, EPI_ISL_800054 to 800055, EPI_ISL_800065, EPI_ISL_800072, EPI_ISL_800077, EPI_ISL_800082 to 800083, EPI_ISL_800085, EPI_ISL_800088 to 800089, EPI_ISL_800092 to 800093, EPI_ISL_800095 to 800096, EPI_ISL_800098, EPI_ISL_800100, EPI_ISL_800102 to 800103, EPI_ISL_800114 to 800116, EPI_ISL_800119, EPI_ISL_800121, EPI_ISL_800123, EPI_ISL_800125, EPI_ISL_800133, EPI_ISL_800137, EPI_ISL_800139, EPI_ISL_800149, EPI_ISL_800153, EPI_ISL_800156 to 800157, EPI_ISL_800166 to 800168, EPI_ISL_800172 to 800173, EPI_ISL_800177 to 800178, EPI_ISL_800182, EPI_ISL_800192, EPI_ISL_800195 to 800196, EPI_ISL_800203, EPI_ISL_800211, EPI_ISL_800214, EPI_ISL_800218, EPI_ISL_800220, EPI_ISL_800225, EPI_ISL_800228, EPI_ISL_800233, EPI_ISL_800238, EPI_ISL_800244, EPI_ISL_800250, EPI_ISL_800252, EPI_ISL_800255 to 800261, EPI_ISL_800263 to 800266, EPI_ISL_800268 to 800270, EPI_ISL_800272 to 800273, EPI_ISL_800275 to 800279, EPI_ISL_800283 to 800285, EPI_ISL_800287 to 800292, EPI_ISL_800294 to 800298, EPI_ISL_800300, EPI_ISL_800302, EPI_ISL_800304 to 800307, EPI_ISL_800309 to 800316, EPI_ISL_800318 to 800337, EPI_ISL_800339, EPI_ISL_800341, EPI_ISL_800343 to 800346, EPI_ISL_800349, EPI_ISL_800352 to 800353, EPI_ISL_800356 to 800358, EPI_ISL_800361, EPI_ISL_800363 to 800364, EPI_ISL_800366, EPI_ISL_800368 to 800369, EPI_ISL_800371 to 800375, EPI_ISL_800377 to 800383, EPI_ISL_800385 to 800391, EPI_ISL_800393 to 800394, EPI_ISL_800396 to 800406, EPI_ISL_800408 to 800419, EPI_ISL_800422 to 800427, EPI_ISL_800429 to 800440, EPI_ISL_800442 to 800443, EPI_ISL_800445, EPI_ISL_800447 to 800450, EPI_ISL_800455, EPI_ISL_800458 to 800459, EPI_ISL_800461 to 800467, EPI_ISL_800469 to 800473, EPI_ISL_800475 to 800480, EPI_ISL_800482 to 800484, EPI_ISL_800486 to 800493, EPI_ISL_800496 to 800500, EPI_ISL_800503 to 800505, EPI_ISL_800507 to 800508, EPI_ISL_800510 to 800515, EPI_ISL_800517, EPI_ISL_800519 to 800523, EPI_ISL_800527, EPI_ISL_800529 to 800530, EPI_ISL_800532 to 800541, EPI_ISL_800543 to 800554, EPI_ISL_800556 to 800557, EPI_ISL_800559 to 800565, EPI_ISL_800567 to 800574, EPI_ISL_800586 to 800587, EPI_ISL_800589 to 800590, EPI_ISL_800592, EPI_ISL_800597, EPI_ISL_800600, EPI_ISL_800606, EPI_ISL_800609, EPI_ISL_800615, EPI_ISL_800626, EPI_ISL_800637, EPI_ISL_800641, EPI_ISL_800647 to 800649, EPI_ISL_800658, EPI_ISL_800663, EPI_ISL_800665, EPI_ISL_800669, EPI_ISL_800672, EPI_ISL_800679 to 800680, EPI_ISL_800682 to 800683, EPI_ISL_800686 to 800687, EPI_ISL_800702, EPI_ISL_800707 to 800709, EPI_ISL_800712, EPI_ISL_800716, EPI_ISL_800720, EPI_ISL_800724, EPI_ISL_800730, EPI_ISL_800732 to 800734, EPI_ISL_800737, EPI_ISL_800744, EPI_ISL_800746, EPI_ISL_800755, EPI_ISL_800761, EPI_ISL_800763, EPI_ISL_800768, EPI_ISL_800770 to 800771, EPI_ISL_800792 to 800793, EPI_ISL_800795, EPI_ISL_800798, EPI_ISL_800802 to 800803, EPI_ISL_800806, EPI_ISL_800808, EPI_ISL_800810, EPI_ISL_800816 to 800817, EPI_ISL_800819, EPI_ISL_800828, EPI_ISL_800832, EPI_ISL_800839, EPI_ISL_800842, EPI_ISL_800853 to 800855, EPI_ISL_800859, EPI_ISL_800865, EPI_ISL_800870, EPI_ISL_800872 to 800873, EPI_ISL_800883, EPI_ISL_800885 to 800886, EPI_ISL_800888 to 800891, EPI_ISL_800894 to 800898, EPI_ISL_800901, EPI_ISL_800903, EPI_ISL_800905, EPI_ISL_800907 to 800908, EPI_ISL_800910 to 800924, EPI_ISL_800926 to 800927, EPI_ISL_800929 to 800936, EPI_ISL_800938 to 800947, EPI_ISL_800949 to 800953, EPI_ISL_800955, EPI_ISL_800957 to 800958, EPI_ISL_800960 to 800967, EPI_ISL_800969, EPI_ISL_800971 to 800977, EPI_ISL_800979 to 800981, EPI_ISL_800985, EPI_ISL_800988 to 800991, EPI_ISL_800993 to 800995, EPI_ISL_800998, EPI_ISL_801000 to 801008, EPI_ISL_801010 to 801011, EPI_ISL_801013 to 801014, EPI_ISL_801016 to 801018, EPI_ISL_801023 to 801028, EPI_ISL_801031 to 801033, EPI_ISL_801035 to 801054, EPI_ISL_801056 to 801061, EPI_ISL_801064 to 801070, EPI_ISL_801072, EPI_ISL_801074 to 801086, EPI_ISL_801089 to 801095, EPI_ISL_801099 to 801103, EPI_ISL_801105 to 801113, EPI_ISL_801116, EPI_ISL_801118 to 801140, EPI_ISL_801142 to 801145, EPI_ISL_801147 to 801148, EPI_ISL_801150 to 801154, EPI_ISL_801157 to 801178, EPI_ISL_811713, EPI_ISL_821278, EPI_ISL_821311, EPI_ISL_821316, EPI_ISL_821320, EPI_ISL_821368, EPI_ISL_821376, EPI_ISL_821393, EPI_ISL_821399, EPI_ISL_821421, EPI_ISL_821427, EPI_ISL_821442, EPI_ISL_821444, EPI_ISL_821454, EPI_ISL_821463, EPI_ISL_821473, EPI_ISL_821485, EPI_ISL_821513, EPI_ISL_821546, EPI_ISL_821602, EPI_ISL_821613, EPI_ISL_821662 to 821665, EPI_ISL_821667, EPI_ISL_821670 to 821673, EPI_ISL_821676, EPI_ISL_821679 to 821686, EPI_ISL_821689 to 821697, EPI_ISL_821699, EPI_ISL_821701 to 821707, EPI_ISL_821709 to 821720, EPI_ISL_821722 to 821723, EPI_ISL_821725 to 821728, EPI_ISL_821730 to 821734, EPI_ISL_821736, EPI_ISL_821738 to 821751, EPI_ISL_821753 to 821757, EPI_ISL_821759 to 821766, EPI_ISL_821769 to 821772, EPI_ISL_821777 to 821780, EPI_ISL_821783, EPI_ISL_821785, EPI_ISL_821787 to 821788, EPI_ISL_821790 to 821791, EPI_ISL_821795 to 821796, EPI_ISL_821798 to 821800, EPI_ISL_821802, EPI_ISL_821804 to 821809, EPI_ISL_821811 to 821816, EPI_ISL_821818 to 821827, EPI_ISL_821830, EPI_ISL_821832 to 821833, EPI_ISL_821835, EPI_ISL_821837 to 821838, EPI_ISL_821840 to 821841, EPI_ISL_821843 to 821844, EPI_ISL_821846, EPI_ISL_821848 to 821853, EPI_ISL_821855 to 821857, EPI_ISL_821859 to 821860, EPI_ISL_821862 to 821864, EPI_ISL_821866, EPI_ISL_821868 to 821872, EPI_ISL_821876 to 821881, EPI_ISL_821883, EPI_ISL_821885, EPI_ISL_821888, EPI_ISL_821891 to 821894, EPI_ISL_821896, EPI_ISL_821898, EPI_ISL_821900 to 821902, EPI_ISL_821904, EPI_ISL_821906 to 821907, EPI_ISL_821909, EPI_ISL_821911 to 821914, EPI_ISL_821916 to 821918, EPI_ISL_821920 to 821922, EPI_ISL_821924 to 821933, EPI_ISL_821935 to 821943, EPI_ISL_821945 to 821949, EPI_ISL_821951 to 821955, EPI_ISL_821957 to 821959, EPI_ISL_821962, EPI_ISL_821965, EPI_ISL_821968, EPI_ISL_821970, EPI_ISL_822299, EPI_ISL_822301, EPI_ISL_822310 to 822316, EPI_ISL_822322, EPI_ISL_826518, EPI_ISL_859554, EPI_ISL_859556 to 859560

| | | | |
|---|---|---|---|
| see above | Lighthouse Lab in Cambridge | Wellcome Sanger Institute for the COVID-19 Genomics UK (COG-UK) Consortium | Cordelia Langford; David K. Jackson; Dominic Kwiatkowski; Ewan Harrison; Ian Johnston; John Sillitoe on behalf of the Wellcome Sanger Institute COVID-19 Surveillance Team; John Sillitoe on behalf of the Wellcome Sanger Institute COVID-19 Surveillance Team (http://www.sanger.ac.uk/covid-team); Rob Howes; Roberto Amato; Sonia Goncalves; The Lighthouse Lab in Cambridge and Alex Alderton |

EPI_ISL_551835 to 551849, EPI_ISL_551851 to 551863, EPI_ISL_551865 to 551881, EPI_ISL_551883 to 551884, EPI_ISL_551886 to 551887, EPI_ISL_551889 to 551920, EPI_ISL_551925, EPI_ISL_551932, EPI_ISL_551951, EPI_ISL_551953, EPI_ISL_551967, EPI_ISL_551980, EPI_ISL_551983, EPI_ISL_551990, EPI_ISL_551994, EPI_ISL_552001, EPI_ISL_552005, EPI_ISL_552008 to 552010, EPI_ISL_552017, EPI_ISL_552051, EPI_ISL_552065, EPI_ISL_552073, EPI_ISL_552091, EPI_ISL_552094, EPI_ISL_552098, EPI_ISL_552105, EPI_ISL_552124, EPI_ISL_552151, EPI_ISL_552166, EPI_ISL_552170, EPI_ISL_552173, EPI_ISL_552180, EPI_ISL_552185, EPI_ISL_555864, EPI_ISL_555868, EPI_ISL_555875, EPI_ISL_555910, EPI_ISL_555939, EPI_ISL_555945, EPI_ISL_555948, EPI_ISL_555955, EPI_ISL_555973, EPI_ISL_555976, EPI_ISL_555993, EPI_ISL_556012, EPI_ISL_556021 to 556022, EPI_ISL_556026, EPI_ISL_556039, EPI_ISL_556047, EPI_ISL_556058, EPI_ISL_556063, EPI_ISL_556066, EPI_ISL_556069, EPI_ISL_556309, EPI_ISL_556317 to 556318, EPI_ISL_556324, EPI_ISL_556338, EPI_ISL_556344, EPI_ISL_556349, EPI_ISL_556351, EPI_ISL_556357, EPI_ISL_556425, EPI_ISL_556434, EPI_ISL_556446, EPI_ISL_556459, EPI_ISL_556471, EPI_ISL_556482, EPI_ISL_556488, EPI_ISL_556494, EPI_ISL_556500, EPI_ISL_556508 to 556509, EPI_ISL_556516 to 556529, EPI_ISL_556532 to 556539, EPI_ISL_556541 to 556549, EPI_ISL_556551 to 556555, EPI_ISL_556557 to 556575, EPI_ISL_556639 to 556679, EPI_ISL_556681 to 556696, EPI_ISL_556699 to 556714, EPI_ISL_556716 to 556720, EPI_ISL_556722 to 556745, EPI_ISL_556747 to 556750, EPI_ISL_556752 to 556758, EPI_ISL_556760 to 556769, EPI_ISL_556771 to 556789, EPI_ISL_556791 to 556799, EPI_ISL_556801 to 556821, EPI_ISL_556823 to 556862, EPI_ISL_556864 to 556886, EPI_ISL_556888 to 556895, EPI_ISL_556897 to 556900, EPI_ISL_556902 to 556926, EPI_ISL_556928 to 556939, EPI_ISL_556941 to 556962, EPI_ISL_556964 to 556965, EPI_ISL_556967 to 556977, EPI_ISL_556979 to 556984, EPI_ISL_566423, EPI_ISL_566429, EPI_ISL_566444 to 566445, EPI_ISL_566453, EPI_ISL_566457, EPI_ISL_566467, EPI_ISL_566493, EPI_ISL_566500, EPI_ISL_566508, EPI_ISL_566518 to 566519, EPI_ISL_566524, EPI_ISL_566528, EPI_ISL_566534, EPI_ISL_566543 to 566544, EPI_ISL_566567, EPI_ISL_566576, EPI_ISL_566586, EPI_ISL_566589, EPI_ISL_566610 to 566611, EPI_ISL_566625, EPI_ISL_566638, EPI_ISL_566659, EPI_ISL_566661 to 566662, EPI_ISL_566666, EPI_ISL_566672, EPI_ISL_566674, EPI_ISL_568385, EPI_ISL_568436, EPI_ISL_568453 to 568454, EPI_ISL_568458, EPI_ISL_568461, EPI_ISL_568472 to 568473, EPI_ISL_580282 to 580283, EPI_ISL_580286 to 580287, EPI_ISL_580299 to 580301, EPI_ISL_580305, EPI_ISL_580313, EPI_ISL_580322, EPI_ISL_580325, EPI_ISL_580327, EPI_ISL_580333, EPI_ISL_580338, EPI_ISL_580340 to 580341, EPI_ISL_580353 to 580354, EPI_ISL_580362, EPI_ISL_580364, EPI_ISL_580367 to 580369, EPI_ISL_580371, EPI_ISL_580373, EPI_ISL_580385, EPI_ISL_580391, EPI_ISL_580393 to 580394, EPI_ISL_580396, EPI_ISL_580400, EPI_ISL_580414, EPI_ISL_580418, EPI_ISL_580421, EPI_ISL_580435, EPI_ISL_580444, EPI_ISL_580449, EPI_ISL_580453, EPI_ISL_580459, EPI_ISL_580465, EPI_ISL_580467, EPI_ISL_580471, EPI_ISL_580478 to 580481, EPI_ISL_580485 to 580486, EPI_ISL_580495, EPI_ISL_580499, EPI_ISL_580502, EPI_ISL_580509, EPI_ISL_580511, EPI_ISL_580515, EPI_ISL_580532 to 580533, EPI_ISL_580536, EPI_ISL_580550, EPI_ISL_580557, EPI_ISL_580559, EPI_ISL_580564 to 580565, EPI_ISL_580567, EPI_ISL_580569, EPI_ISL_580573, EPI_ISL_580576 to 580577, EPI_ISL_580579, EPI_ISL_580582, EPI_ISL_580586 to 580587, EPI_ISL_580593, EPI_ISL_580603, EPI_ISL_580607, EPI_ISL_580610, EPI_ISL_580618, EPI_ISL_580621, EPI_ISL_580625 to 580626, EPI_ISL_581394, EPI_ISL_581396, EPI_ISL_590173, EPI_ISL_596928 to 596931, EPI_ISL_596933 to 596968, EPI_ISL_596970 to 596978, EPI_ISL_596980 to 597037, EPI_ISL_597039 to 597051, EPI_ISL_597053 to 597064, EPI_ISL_597066 to 597073, EPI_ISL_597075 to 597078, EPI_ISL_597080 to 597098, EPI_ISL_597100 to 597133, EPI_ISL_597135 to 597141, EPI_ISL_597143 to 597149, EPI_ISL_597151 to 597188, EPI_ISL_597190 to 597193, EPI_ISL_597195 to 597232, EPI_ISL_597237 to 597239, EPI_ISL_597241, EPI_ISL_597243, EPI_ISL_597246 to 597248, EPI_ISL_597250 to 597251, EPI_ISL_597253 to 597259, EPI_ISL_597262, EPI_ISL_597264 to 597266, EPI_ISL_597269 to 597271, EPI_ISL_597273 to 597274, EPI_ISL_597279, EPI_ISL_597282 to 597287, EPI_ISL_597289, EPI_ISL_597293 to 597295, EPI_ISL_597297, EPI_ISL_597300 to 597301, EPI_ISL_597304 to 597310, EPI_ISL_597316 to 597317, EPI_ISL_597319 to 597321, EPI_ISL_597323 to 597325, EPI_ISL_597327, EPI_ISL_597331, EPI_ISL_597335, EPI_ISL_597337, EPI_ISL_597340 to 597346, EPI_ISL_597349 to 597350, EPI_ISL_597354, EPI_ISL_597361, EPI_ISL_597365 to 597368, EPI_ISL_597371 to 597375, EPI_ISL_597377 to 597378, EPI_ISL_597380 to 597381, EPI_ISL_597388, EPI_ISL_597390, EPI_ISL_597392, EPI_ISL_597396 to 597399, EPI_ISL_597401 to 597403, EPI_ISL_597405, EPI_ISL_597412, EPI_ISL_597414, EPI_ISL_597418, EPI_ISL_597425 to 597428, EPI_ISL_597432, EPI_ISL_597435, EPI_ISL_597437 to 597438, EPI_ISL_597441, EPI_ISL_597445 to 597446, EPI_ISL_597450 to 597451, EPI_ISL_597453, EPI_ISL_597458 to 597462, EPI_ISL_597464, EPI_ISL_597466 to 597470, EPI_ISL_597472 to 597473, EPI_ISL_597476 to 597480, EPI_ISL_597483, EPI_ISL_597487 to 597490, EPI_ISL_597494 to 597495, EPI_ISL_597497, EPI_ISL_597502, EPI_ISL_597504 to 597505, EPI_ISL_597507 to 597508, EPI_ISL_597510, EPI_ISL_597512, EPI_ISL_597519 to 597520, EPI_ISL_601493, EPI_ISL_601495, EPI_ISL_601499, EPI_ISL_601501, EPI_ISL_601506, EPI_ISL_601515, EPI_ISL_601522, EPI_ISL_601530, EPI_ISL_601541, EPI_ISL_601549 to 601550, EPI_ISL_601555, EPI_ISL_601567, EPI_ISL_601569, EPI_ISL_601571, EPI_ISL_601578 to 601580, EPI_ISL_601585, EPI_ISL_601588 to 601591, EPI_ISL_601594, EPI_ISL_601604, EPI_ISL_601606, EPI_ISL_601611, EPI_ISL_601615, EPI_ISL_601617 to 601619, EPI_ISL_601622, EPI_ISL_601624, EPI_ISL_601627 to 601628, EPI_ISL_601634, EPI_ISL_601639, EPI_ISL_601648, EPI_ISL_601657, EPI_ISL_601663 to 601664, EPI_ISL_601666, EPI_ISL_601668, EPI_ISL_601673, EPI_ISL_601682, EPI_ISL_601685, EPI_ISL_601690 to 601691, EPI_ISL_601700, EPI_ISL_601715, EPI_ISL_601728, EPI_ISL_601730, EPI_ISL_601733, EPI_ISL_601740, EPI_ISL_601747 to 601748, EPI_ISL_601764 to 601765, EPI_ISL_601768, EPI_ISL_601771, EPI_ISL_601778, EPI_ISL_601783, EPI_ISL_601785, EPI_ISL_602154, EPI_ISL_608348 to 608349, EPI_ISL_608353 to 608354, EPI_ISL_608357, EPI_ISL_608359 to 608360, EPI_ISL_608363, EPI_ISL_608368, EPI_ISL_608370, EPI_ISL_608373, EPI_ISL_608381 to 608382, EPI_ISL_608388 to 608389, EPI_ISL_608391 to 608398, EPI_ISL_608400, EPI_ISL_608402, EPI_ISL_608404, EPI_ISL_608407 to 608408, EPI_ISL_608410 to 608412, EPI_ISL_608416 to 608417, EPI_ISL_608423 to 608424, EPI_ISL_608426 to 608430, EPI_ISL_608432 to 608433, EPI_ISL_608436 to 608437, EPI_ISL_608439, EPI_ISL_608442, EPI_ISL_608446 to 608447, EPI_ISL_608449 to 608451, EPI_ISL_608456,

EPI_ISL_608458 to 608459, EPI_ISL_608463 to 608464, EPI_ISL_608466 to 608467, EPI_ISL_608471 to 608472, EPI_ISL_608475 to 608478, EPI_ISL_608480, EPI_ISL_608482, EPI_ISL_608484 to 608486, EPI_ISL_608490, EPI_ISL_608492 to 608495, EPI_ISL_608497 to 608498, EPI_ISL_608501, EPI_ISL_608504, EPI_ISL_608506 to 608510, EPI_ISL_608514 to 608515, EPI_ISL_608521 to 608522, EPI_ISL_608524, EPI_ISL_608527, EPI_ISL_608529, EPI_ISL_608531 to 608532, EPI_ISL_608535 to 608538, EPI_ISL_608540, EPI_ISL_608544 to 608547, EPI_ISL_608550, EPI_ISL_608552, EPI_ISL_608557 to 608561, EPI_ISL_608565 to 608566, EPI_ISL_608568, EPI_ISL_608570 to 608573, EPI_ISL_608576 to 608578, EPI_ISL_608585, EPI_ISL_608588, EPI_ISL_608596 to 608598, EPI_ISL_608602, EPI_ISL_608604, EPI_ISL_608607 to 608608, EPI_ISL_608611 to 608613, EPI_ISL_608615 to 608616, EPI_ISL_608618 to 608620, EPI_ISL_608624, EPI_ISL_608626 to 608629, EPI_ISL_608631 to 608632, EPI_ISL_608636 to 608638, EPI_ISL_608641 to 608645, EPI_ISL_608650 to 608651, EPI_ISL_608653 to 608654, EPI_ISL_608656, EPI_ISL_608660, EPI_ISL_608664, EPI_ISL_608666, EPI_ISL_608671, EPI_ISL_608673, EPI_ISL_608678 to 608679, EPI_ISL_608682 to 608683, EPI_ISL_608688 to 608689, EPI_ISL_608693, EPI_ISL_608695 to 608696, EPI_ISL_608845, EPI_ISL_608852, EPI_ISL_608867 to 608868, EPI_ISL_608873, EPI_ISL_608875 to 608876, EPI_ISL_608879, EPI_ISL_608881, EPI_ISL_608892, EPI_ISL_608902, EPI_ISL_608906, EPI_ISL_608908, EPI_ISL_608910, EPI_ISL_608912, EPI_ISL_608918, EPI_ISL_608923 to 608924, EPI_ISL_608927, EPI_ISL_608935, EPI_ISL_608939, EPI_ISL_608943, EPI_ISL_608947, EPI_ISL_608949, EPI_ISL_608953, EPI_ISL_608958 to 608960, EPI_ISL_608965, EPI_ISL_608967 to 608969, EPI_ISL_608974, EPI_ISL_608982, EPI_ISL_608987, EPI_ISL_608990, EPI_ISL_608996 to 608997, EPI_ISL_609003, EPI_ISL_609005, EPI_ISL_609014, EPI_ISL_609018, EPI_ISL_609023, EPI_ISL_609025, EPI_ISL_609033, EPI_ISL_609037, EPI_ISL_609045, EPI_ISL_609048, EPI_ISL_609050, EPI_ISL_609054, EPI_ISL_609067, EPI_ISL_609072, EPI_ISL_609077, EPI_ISL_609081 to 609082, EPI_ISL_609086, EPI_ISL_609088, EPI_ISL_609091, EPI_ISL_609097, EPI_ISL_609099 to 609101, EPI_ISL_609111, EPI_ISL_609113 to 609114, EPI_ISL_609118 to 609119, EPI_ISL_609122 to 609123, EPI_ISL_609129, EPI_ISL_609131, EPI_ISL_609142 to 609143, EPI_ISL_609145, EPI_ISL_609156, EPI_ISL_609158 to 609177, EPI_ISL_609180 to 609181, EPI_ISL_609183, EPI_ISL_609188, EPI_ISL_609193, EPI_ISL_609195 to 609196, EPI_ISL_609200 to 609201, EPI_ISL_609203 to 609205, EPI_ISL_609308, EPI_ISL_609310 to 609330, EPI_ISL_609332 to 609377, EPI_ISL_609379 to 609392, EPI_ISL_609394 to 609418, EPI_ISL_609420, EPI_ISL_609422 to 609437, EPI_ISL_609439 to 609449, EPI_ISL_609451 to 609471, EPI_ISL_609473 to 609476, EPI_ISL_609478 to 609483, EPI_ISL_609485 to 609487, EPI_ISL_609489 to 609506, EPI_ISL_609509 to 609511, EPI_ISL_609513 to 609526, EPI_ISL_609528 to 609533, EPI_ISL_609535 to 609562, EPI_ISL_609564 to 609599, EPI_ISL_609601 to 609655, EPI_ISL_609657, EPI_ISL_609659 to 609662, EPI_ISL_609664 to 609665, EPI_ISL_609667 to 609671, EPI_ISL_609673 to 609705, EPI_ISL_609707 to 609735, EPI_ISL_609737 to 609745, EPI_ISL_609747 to 609753, EPI_ISL_609755 to 609760, EPI_ISL_609763 to 609767, EPI_ISL_609769 to 609792, EPI_ISL_609794 to 609800, EPI_ISL_610629 to 610653, EPI_ISL_610655, EPI_ISL_610657 to 610674, EPI_ISL_610676 to 610684, EPI_ISL_610686 to 610693, EPI_ISL_610695 to 610701, EPI_ISL_610703 to 610712, EPI_ISL_610714 to 610743, EPI_ISL_610745 to 610763, EPI_ISL_610765 to 610778, EPI_ISL_610781 to 610842, EPI_ISL_610844 to 610860, EPI_ISL_610862 to 610867, EPI_ISL_610869 to 610884, EPI_ISL_610886 to 610907, EPI_ISL_610909 to 610913, EPI_ISL_610915 to 610921, EPI_ISL_623220, EPI_ISL_623222, EPI_ISL_623234, EPI_ISL_623245, EPI_ISL_623247 to 623248, EPI_ISL_623256, EPI_ISL_623268, EPI_ISL_623273, EPI_ISL_623276, EPI_ISL_623281, EPI_ISL_623286, EPI_ISL_623288, EPI_ISL_623301, EPI_ISL_623308, EPI_ISL_623312, EPI_ISL_623321, EPI_ISL_623323, EPI_ISL_623330, EPI_ISL_623333 to 623334, EPI_ISL_623337, EPI_ISL_623343 to 623347, EPI_ISL_623359 to 623360, EPI_ISL_623363 to 623364, EPI_ISL_623366, EPI_ISL_623370, EPI_ISL_623385 to 623386, EPI_ISL_623390, EPI_ISL_623392 to 623393, EPI_ISL_623397 to 623398, EPI_ISL_623404, EPI_ISL_623409, EPI_ISL_623411, EPI_ISL_623424 to 623427, EPI_ISL_623433, EPI_ISL_623437, EPI_ISL_623440, EPI_ISL_623444 to 623445, EPI_ISL_623448 to 623449, EPI_ISL_623451, EPI_ISL_623474 to 623475, EPI_ISL_623482, EPI_ISL_623485, EPI_ISL_623488 to 623490, EPI_ISL_623494 to 623495, EPI_ISL_623520, EPI_ISL_623527 to 623528, EPI_ISL_623531 to 623532, EPI_ISL_623537, EPI_ISL_623539, EPI_ISL_623544 to 623545, EPI_ISL_623547 to 623548, EPI_ISL_623550 to 623551, EPI_ISL_623555 to 623560, EPI_ISL_623562, EPI_ISL_623568 to 623570, EPI_ISL_623572, EPI_ISL_623574 to 623576, EPI_ISL_623578, EPI_ISL_623581, EPI_ISL_623584 to 623585, EPI_ISL_623587, EPI_ISL_623592 to 623593, EPI_ISL_623595, EPI_ISL_623600, EPI_ISL_623605, EPI_ISL_623608 to 623609, EPI_ISL_623611 to 623612, EPI_ISL_623616 to 623617, EPI_ISL_623619, EPI_ISL_623626, EPI_ISL_623633, EPI_ISL_623639 to 623641, EPI_ISL_623643 to 623645, EPI_ISL_623647, EPI_ISL_623650 to 623652, EPI_ISL_623654 to 623655, EPI_ISL_623657 to 623658, EPI_ISL_623661 to 623663, EPI_ISL_623665 to 623667, EPI_ISL_623674 to 623675, EPI_ISL_623679 to 623680, EPI_ISL_623683, EPI_ISL_623688, EPI_ISL_623690, EPI_ISL_623694, EPI_ISL_623700, EPI_ISL_623702 to 623703, EPI_ISL_623710, EPI_ISL_623712, EPI_ISL_623716 to 623717, EPI_ISL_623721, EPI_ISL_623724 to 623726, EPI_ISL_623731, EPI_ISL_623733, EPI_ISL_623735 to 623736, EPI_ISL_623740 to 623742, EPI_ISL_623746, EPI_ISL_623749 to 623751, EPI_ISL_623754, EPI_ISL_623758 to 623759, EPI_ISL_623762 to 623764, EPI_ISL_623772, EPI_ISL_623774 to 623776, EPI_ISL_623785, EPI_ISL_623788, EPI_ISL_623790 to 623791, EPI_ISL_623793 to 623794, EPI_ISL_623796 to 623800, EPI_ISL_623802, EPI_ISL_623806, EPI_ISL_623808, EPI_ISL_623810 to 623812, EPI_ISL_623815 to 623816, EPI_ISL_623822 to 623823, EPI_ISL_623828 to 623829, EPI_ISL_623831 to 623834, EPI_ISL_623838, EPI_ISL_623840 to 623841, EPI_ISL_623843 to 623844, EPI_ISL_623846 to 623847, EPI_ISL_623849, EPI_ISL_623854 to 623857, EPI_ISL_625281 to 625288, EPI_ISL_625290 to 625291, EPI_ISL_625293 to 625296, EPI_ISL_625298 to 625303, EPI_ISL_625305, EPI_ISL_625307 to 625311, EPI_ISL_625313 to 625314, EPI_ISL_625432, EPI_ISL_625437 to 625438, EPI_ISL_625440 to 625453, EPI_ISL_630972, EPI_ISL_630977, EPI_ISL_630979, EPI_ISL_630984, EPI_ISL_630988, EPI_ISL_630996 to 630997, EPI_ISL_631001, EPI_ISL_631003, EPI_ISL_631005, EPI_ISL_631008, EPI_ISL_631016 to 631017, EPI_ISL_631025, EPI_ISL_631028, EPI_ISL_631038 to 631039, EPI_ISL_631045, EPI_ISL_631053, EPI_ISL_631055, EPI_ISL_631062 to 631063, EPI_ISL_631069, EPI_ISL_631078, EPI_ISL_631080, EPI_ISL_631094, EPI_ISL_631101, EPI_ISL_631105, EPI_ISL_631117 to 631118, EPI_ISL_631123, EPI_ISL_631129, EPI_ISL_631132, EPI_ISL_631135, EPI_ISL_631137, EPI_ISL_631142, EPI_ISL_631144, EPI_ISL_631148 to 631149, EPI_ISL_631154 to 631155, EPI_ISL_631160, EPI_ISL_631164, EPI_ISL_631171, EPI_ISL_631174, EPI_ISL_631177, EPI_ISL_631180 to 631181, EPI_ISL_631185, EPI_ISL_631191 to 631192, EPI_ISL_631194, EPI_ISL_631199, EPI_ISL_631204, EPI_ISL_631208, EPI_ISL_631219 to 631221, EPI_ISL_631224 to 631225, EPI_ISL_631235, EPI_ISL_631237, EPI_ISL_631245, EPI_ISL_631254, EPI_ISL_631257, EPI_ISL_631262, EPI_ISL_631266, EPI_ISL_631268, EPI_ISL_631270 to 631274, EPI_ISL_633100 to 633102, EPI_ISL_633104 to 633105, EPI_ISL_633107 to 633108, EPI_ISL_633114, EPI_ISL_633116 to 633117, EPI_ISL_633120 to 633124, EPI_ISL_633126 to 633131, EPI_ISL_633135 to 633136, EPI_ISL_633140 to 633142, EPI_ISL_633147 to 633150, EPI_ISL_633153, EPI_ISL_633155 to 633157, EPI_ISL_633159, EPI_ISL_633161, EPI_ISL_633163 to 633164, EPI_ISL_633170, EPI_ISL_633172 to 633173, EPI_ISL_633176 to 633177, EPI_ISL_633183, EPI_ISL_633186 to 633189, EPI_ISL_633192, EPI_ISL_633194, EPI_ISL_633197 to 633200, EPI_ISL_633205, EPI_ISL_633208, EPI_ISL_633211 to 633212, EPI_ISL_633214 to 633220, EPI_ISL_633223 to 633225, EPI_ISL_633227, EPI_ISL_633229, EPI_ISL_633231, EPI_ISL_633234 to 633236, EPI_ISL_633238 to 633239, EPI_ISL_633242, EPI_ISL_633245 to 633250, EPI_ISL_633252 to 633255, EPI_ISL_633257 to 633258, EPI_ISL_633260, EPI_ISL_633263 to 633267, EPI_ISL_633270, EPI_ISL_633273, EPI_ISL_633275, EPI_ISL_633277 to 633278, EPI_ISL_633281, EPI_ISL_633284, EPI_ISL_633286, EPI_ISL_633288, EPI_ISL_633291 to 633292, EPI_ISL_633294 to 633295, EPI_ISL_633298 to 633300, EPI_ISL_633302, EPI_ISL_633308, EPI_ISL_633311, EPI_ISL_633313 to 633315, EPI_ISL_633319, EPI_ISL_633321 to 633322, EPI_ISL_633324 to 633325, EPI_ISL_633329 to 633330, EPI_ISL_633333 to 633334, EPI_ISL_633336 to 633337, EPI_ISL_633340 to 633341, EPI_ISL_633344 to 633347, EPI_ISL_633349, EPI_ISL_633356 to 633357, EPI_ISL_633359 to 633360, EPI_ISL_633363, EPI_ISL_633365 to 633366, EPI_ISL_633369 to 633370, EPI_ISL_633372 to 633375, EPI_ISL_633378, EPI_ISL_633380, EPI_ISL_633383, EPI_ISL_633385, EPI_ISL_633388 to 633390, EPI_ISL_633393, EPI_ISL_633395, EPI_ISL_633397 to 633399, EPI_ISL_633403, EPI_ISL_633405, EPI_ISL_633407 to 633408, EPI_ISL_634789 to 634816

| | | | |
|---|---|---|---|
| see above | Lighthouse Lab in Cambridge | Wellcome Sanger Institute for the COVID-19 Genomics UK (COG-UK) consortium | Cordelia Langford; David K. Jackson; Dominic Kwiatkowski; Ewan Harrison; Ian Johnston; John Sillitoe on behalf of the Wellcome Sanger Institute COVID-19 Surveillance Team; John Sillitoe on behalf of the Wellcome Sanger Institute COVID-19 Surveillance Team (http://www.sanger.ac.uk/covid-team); Rob Howes; Roberto Amato; Sonia Goncalves; The Lighthouse Lab in Cambridge and Alex Alderton |

EPI_ISL_530482, EPI_ISL_530899, EPI_ISL_530963, EPI_ISL_531211, EPI_ISL_531688, EPI_ISL_531872, EPI_ISL_532012, EPI_ISL_532511, EPI_ISL_532583, EPI_ISL_533086, EPI_ISL_533403, EPI_ISL_536889, EPI_ISL_537017, EPI_ISL_539906, EPI_ISL_540030, EPI_ISL_540278, EPI_ISL_540281, EPI_ISL_540351, EPI_ISL_549772, EPI_ISL_567024, EPI_ISL_567059, EPI_ISL_567184, EPI_ISL_567231, EPI_ISL_568301, EPI_ISL_568315, EPI_ISL_568344, EPI_ISL_568392, EPI_ISL_568398, EPI_ISL_576050, EPI_ISL_576064, EPI_ISL_576088, EPI_ISL_579794, EPI_ISL_579835, EPI_ISL_579843, EPI_ISL_580909, EPI_ISL_581045, EPI_ISL_581359, EPI_ISL_587386, EPI_ISL_587477, EPI_ISL_588125, EPI_ISL_588128, EPI_ISL_588175, EPI_ISL_588496, EPI_ISL_588541, EPI_ISL_588572, EPI_ISL_588674, EPI_ISL_588717, EPI_ISL_588844 to 588845, EPI_ISL_588927, EPI_ISL_588940, EPI_ISL_588962, EPI_ISL_589248, EPI_ISL_589936, EPI_ISL_590046, EPI_ISL_590497, EPI_ISL_598970, EPI_ISL_599010, EPI_ISL_599077, EPI_ISL_599166, EPI_ISL_599183, EPI_ISL_599194, EPI_ISL_599201, EPI_ISL_599270, EPI_ISL_599590, EPI_ISL_599627, EPI_ISL_599816, EPI_ISL_600139, EPI_ISL_600221, EPI_ISL_600643, EPI_ISL_600824, EPI_ISL_600904, EPI_ISL_601022, EPI_ISL_601150, EPI_ISL_601192, EPI_ISL_601214, EPI_ISL_601262, EPI_ISL_601797, EPI_ISL_601834, EPI_ISL_601950, EPI_ISL_602006, EPI_ISL_602015, EPI_ISL_602077, EPI_ISL_602083, EPI_ISL_602090 to 602091, EPI_ISL_602102, EPI_ISL_602106, EPI_ISL_602109, EPI_ISL_602122, EPI_ISL_609187, EPI_ISL_609192, EPI_ISL_611304, EPI_ISL_611378, EPI_ISL_624197, EPI_ISL_629940, EPI_ISL_633118, EPI_ISL_633282, EPI_ISL_633361, EPI_ISL_633492, EPI_ISL_633577, EPI_ISL_633653, EPI_ISL_633700, EPI_ISL_633760, EPI_ISL_633782, EPI_ISL_633799, EPI_ISL_634156, EPI_ISL_634678, EPI_ISL_641607 to 641609, EPI_ISL_641611 to 641675, EPI_ISL_641677 to 641747, EPI_ISL_641749 to 641773, EPI_ISL_641775 to 641796, EPI_ISL_641798 to 641936, EPI_ISL_641938 to 641965, EPI_ISL_641967 to 642005, EPI_ISL_642007 to 642028, EPI_ISL_642030 to 642036, EPI_ISL_642038 to 642055, EPI_ISL_642057 to 642082, EPI_ISL_642084 to 642117, EPI_ISL_642119 to 642129, EPI_ISL_642131 to 642141, EPI_ISL_642143 to 642208, EPI_ISL_642210 to 642275, EPI_ISL_642277 to 642291, EPI_ISL_642293 to 642296, EPI_ISL_642298 to 642319, EPI_ISL_642321 to 642343, EPI_ISL_642345 to 642358, EPI_ISL_642360 to 642399, EPI_ISL_642401 to 642457, EPI_ISL_642459 to 642482, EPI_ISL_642484 to 642504, EPI_ISL_642506 to 642561, EPI_ISL_642563 to 642565, EPI_ISL_642568 to 642607, EPI_ISL_642609, EPI_ISL_642611 to 642624, EPI_ISL_642626 to 642642, EPI_ISL_642644 to 642726, EPI_ISL_642728 to 642760, EPI_ISL_643044 to 643052, EPI_ISL_643054 to 643063, EPI_ISL_643065 to 643176, EPI_ISL_643178 to 643215, EPI_ISL_643217 to 643235, EPI_ISL_643237 to 643270, EPI_ISL_643272 to 643314, EPI_ISL_643992 to 644051, EPI_ISL_644093 to 644094, EPI_ISL_644099, EPI_ISL_644101 to 644102, EPI_ISL_644107, EPI_ISL_644112, EPI_ISL_646140 to 646147, EPI_ISL_646149, EPI_ISL_646151 to 646154, EPI_ISL_646158 to 646161, EPI_ISL_646163 to 646165, EPI_ISL_646167 to 646168, EPI_ISL_646171 to 646172, EPI_ISL_646174 to 646179, EPI_ISL_646181, EPI_ISL_646183 to 646184, EPI_ISL_646186 to 646188, EPI_ISL_646190, EPI_ISL_646192, EPI_ISL_646194 to 646197, EPI_ISL_646200 to 646203, EPI_ISL_646205 to 646206, EPI_ISL_646209 to 646211, EPI_ISL_646213 to 646214, EPI_ISL_646216 to 646222, EPI_ISL_646225 to 646227, EPI_ISL_646229 to 646230, EPI_ISL_646232 to 646234, EPI_ISL_646236 to 646241, EPI_ISL_646243 to 646244, EPI_ISL_646246 to 646247, EPI_ISL_646250 to 646251, EPI_ISL_646253 to 646260, EPI_ISL_646262 to 646267, EPI_ISL_646270, EPI_ISL_646272 to 646275, EPI_ISL_646280 to 646282, EPI_ISL_646284 to 646286, EPI_ISL_646288 to 646290, EPI_ISL_646292 to 646296, EPI_ISL_646298, EPI_ISL_646300 to 646301, EPI_ISL_646304, EPI_ISL_646306, EPI_ISL_646308, EPI_ISL_646310 to 646312, EPI_ISL_646315 to 646317, EPI_ISL_646319, EPI_ISL_646321 to 646322, EPI_ISL_646324 to 646326, EPI_ISL_646328 to 646333, EPI_ISL_646335 to 646341, EPI_ISL_646344, EPI_ISL_646348, EPI_ISL_646351 to 646353, EPI_ISL_646355, EPI_ISL_646357 to 646361, EPI_ISL_646364 to 646365, EPI_ISL_646367 to 646368, EPI_ISL_646371 to 646372, EPI_ISL_646374, EPI_ISL_646378 to 646381, EPI_ISL_646383 to 646384, EPI_ISL_646386 to 646387, EPI_ISL_646389 to 646396, EPI_ISL_646398 to 646401, EPI_ISL_646403 to 646411, EPI_ISL_646413 to 646417, EPI_ISL_646421 to 646425, EPI_ISL_646427, EPI_ISL_646433, EPI_ISL_646435 to 646440, EPI_ISL_646444 to 646447, EPI_ISL_646449, EPI_ISL_646451 to 646453, EPI_ISL_646455 to 646462, EPI_ISL_646464 to 646465, EPI_ISL_646469 to 646475, EPI_ISL_646480 to 646484, EPI_ISL_646495 to 646496, EPI_ISL_646498 to 646499, EPI_ISL_646501, EPI_ISL_646503 to 646504, EPI_ISL_646507, EPI_ISL_646517, EPI_ISL_646519, EPI_ISL_646523 to 646524, EPI_ISL_646527, EPI_ISL_646530 to 646531, EPI_ISL_646534, EPI_ISL_646536, EPI_ISL_646539, EPI_ISL_646547, EPI_ISL_646550, EPI_ISL_646552, EPI_ISL_646554, EPI_ISL_646556, EPI_ISL_646562, EPI_ISL_646565, EPI_ISL_646570, EPI_ISL_646574 to 646575, EPI_ISL_646578 to 646579, EPI_ISL_646582, EPI_ISL_646584, EPI_ISL_646587 to 646590, EPI_ISL_646593, EPI_ISL_646596, EPI_ISL_646598, EPI_ISL_646602, EPI_ISL_646604, EPI_ISL_646608 to 646609, EPI_ISL_646611, EPI_ISL_646615, EPI_ISL_646617, EPI_ISL_646619 to 646620, EPI_ISL_646622, EPI_ISL_646624, EPI_ISL_646627, EPI_ISL_646629 to 646630, EPI_ISL_646632 to 646634, EPI_ISL_646636 to 646637, EPI_ISL_646641, EPI_ISL_646643 to 646647, EPI_ISL_646649 to 646650, EPI_ISL_646656 to 646657, EPI_ISL_646659, EPI_ISL_646661, EPI_ISL_646663, EPI_ISL_646667, EPI_ISL_646669 to 646670, EPI_ISL_646672, EPI_ISL_646674, EPI_ISL_646677, EPI_ISL_646679 to 646682, EPI_ISL_646685, EPI_ISL_646688, EPI_ISL_646690, EPI_ISL_646694, EPI_ISL_646699 to 646701, EPI_ISL_646703 to 646705, EPI_ISL_646714, EPI_ISL_646717, EPI_ISL_646720, EPI_ISL_646723, EPI_ISL_646725 to 646726, EPI_ISL_646728 to 646730, EPI_ISL_646732 to 646733, EPI_ISL_646735, EPI_ISL_646742, EPI_ISL_646745, EPI_ISL_646748, EPI_ISL_646754, EPI_ISL_646759, EPI_ISL_646764 to 646766, EPI_ISL_646772, EPI_ISL_646774 to 646775, EPI_ISL_646777 to 646781, EPI_ISL_646783, EPI_ISL_646785 to 646786, EPI_ISL_646788, EPI_ISL_646792, EPI_ISL_646794, EPI_ISL_646798, EPI_ISL_646810, EPI_ISL_646818, EPI_ISL_646825 to 646826, EPI_ISL_646837, EPI_ISL_646850, EPI_ISL_646853 to 646854, EPI_ISL_646868 to 646870, EPI_ISL_646872, EPI_ISL_646874, EPI_ISL_646877, EPI_ISL_646880, EPI_ISL_646885, EPI_ISL_646891 to 646892, EPI_ISL_646902, EPI_ISL_646905, EPI_ISL_646912, EPI_ISL_646915, EPI_ISL_646921, EPI_ISL_646929, EPI_ISL_646936 to 646937, EPI_ISL_646939, EPI_ISL_646948, EPI_ISL_646955, EPI_ISL_646960, EPI_ISL_646964, EPI_ISL_646967, EPI_ISL_646974 to 646977, EPI_ISL_646983, EPI_ISL_646988, EPI_ISL_646992, EPI_ISL_646996, EPI_ISL_647001, EPI_ISL_647003, EPI_ISL_647006, EPI_ISL_647014, EPI_ISL_647023, EPI_ISL_647031 to 647032, EPI_ISL_647035, EPI_ISL_647038, EPI_ISL_647045, EPI_ISL_647050, EPI_ISL_647059, EPI_ISL_647067, EPI_ISL_647073, EPI_ISL_647076, EPI_ISL_647081, EPI_ISL_647087, EPI_ISL_647090, EPI_ISL_647097, EPI_ISL_647102, EPI_ISL_647105, EPI_ISL_647109 to 647110, EPI_ISL_647114 to 647115, EPI_ISL_647117, EPI_ISL_647121, EPI_ISL_647797 to 647806, EPI_ISL_647854 to 647875, EPI_ISL_647877 to 647892, EPI_ISL_647894 to 647899, EPI_ISL_647902 to 647905, EPI_ISL_647907, EPI_ISL_647910 to 647912, EPI_ISL_647914 to 647915, EPI_ISL_647917 to 647956, EPI_ISL_647958 to 647968, EPI_ISL_649494 to 649540, EPI_ISL_649542 to 649547, EPI_ISL_649556 to 649557, EPI_ISL_649559 to 649563, EPI_ISL_649565 to 649602, EPI_ISL_649605 to 649624, EPI_ISL_649634 to 649640, EPI_ISL_649642 to 649663, EPI_ISL_649665 to 649698, EPI_ISL_649700 to 649739, EPI_ISL_649741 to 649780, EPI_ISL_649782 to 649783, EPI_ISL_654959 to 654999, EPI_ISL_655001 to 655056, EPI_ISL_655058 to 655068, EPI_ISL_655070 to 655116, EPI_ISL_655118 to 655198, EPI_ISL_655200 to 655272, EPI_ISL_655651, EPI_ISL_655660, EPI_ISL_655662, EPI_ISL_655672, EPI_ISL_655677, EPI_ISL_655694 to 655695, EPI_ISL_655697, EPI_ISL_655699, EPI_ISL_655711, EPI_ISL_655719, EPI_ISL_655723, EPI_ISL_655727, EPI_ISL_655735, EPI_ISL_655737, EPI_ISL_655739, EPI_ISL_655744 to 655745, EPI_ISL_655756, EPI_ISL_655762, EPI_ISL_655768 to 655770, EPI_ISL_655773, EPI_ISL_655779 to 655780, EPI_ISL_655783, EPI_ISL_655788, EPI_ISL_655798, EPI_ISL_655804, EPI_ISL_655806 to 655807, EPI_ISL_655809, EPI_ISL_655817, EPI_ISL_655819, EPI_ISL_655822, EPI_ISL_655828, EPI_ISL_655832, EPI_ISL_655841, EPI_ISL_655847, EPI_ISL_655850, EPI_ISL_655854, EPI_ISL_655861, EPI_ISL_655864 to 655865, EPI_ISL_655868, EPI_ISL_655873 to 655874, EPI_ISL_655879, EPI_ISL_655883, EPI_ISL_655891, EPI_ISL_655907, EPI_ISL_655911 to 655912, EPI_ISL_655918, EPI_ISL_655920, EPI_ISL_655924, EPI_ISL_655929, EPI_ISL_655933, EPI_ISL_655947 to 655948, EPI_ISL_655957, EPI_ISL_655959, EPI_ISL_655967, EPI_ISL_655971, EPI_ISL_655973, EPI_ISL_655975, EPI_ISL_657027 to 657126, EPI_ISL_657128 to 657155, EPI_ISL_657157 to 657158, EPI_ISL_657160 to 657231, EPI_ISL_657233 to 657354, EPI_ISL_658404 to 658466, EPI_ISL_658468 to 658523, EPI_ISL_658525 to 658573, EPI_ISL_658575 to 658598, EPI_ISL_658600 to 658740, EPI_ISL_659744 to 660037, EPI_ISL_660039 to 660067, EPI_ISL_661314 to 661349, EPI_ISL_661351 to 661395, EPI_ISL_661397 to 661525, EPI_ISL_661527 to 661542, EPI_ISL_661544 to 661567, EPI_ISL_661569 to 661620, EPI_ISL_661622 to 661635, EPI_ISL_661637, EPI_ISL_661639, EPI_ISL_661642 to 661646, EPI_ISL_661648 to 661650, EPI_ISL_661655, EPI_ISL_661657 to 661663, EPI_ISL_661665 to 661666, EPI_ISL_661668 to 661674, EPI_ISL_661676 to 661678, EPI_ISL_661681, EPI_ISL_661683, EPI_ISL_661685 to 661686, EPI_ISL_661688 to 661690, EPI_ISL_661692, EPI_ISL_661694, EPI_ISL_661698 to 661699, EPI_ISL_661701, EPI_ISL_661703 to 661709, EPI_ISL_661715 to 661716, EPI_ISL_661718 to 661720, EPI_ISL_661722, EPI_ISL_661724 to 661725, EPI_ISL_661727 to 661728, EPI_ISL_661730 to 661731, EPI_ISL_661733, EPI_ISL_661735 to 661740, EPI_ISL_661744 to 661746, EPI_ISL_661748, EPI_ISL_661750, EPI_ISL_661753, EPI_ISL_661755 to 661758, EPI_ISL_661760, EPI_ISL_661762, EPI_ISL_661764 to 661774, EPI_ISL_661776 to 661777, EPI_ISL_661780, EPI_ISL_661782, EPI_ISL_661784, EPI_ISL_661786 to 661788, EPI_ISL_661790, EPI_ISL_661792 to 661796, EPI_ISL_661798, EPI_ISL_661800, EPI_ISL_661802, EPI_ISL_661804, EPI_ISL_661806 to 661810, EPI_ISL_661812 to 661814, EPI_ISL_661816, EPI_ISL_661818 to 661819, EPI_ISL_661821 to 661829, EPI_ISL_661831 to 661832, EPI_ISL_661835, EPI_ISL_661837 to 661843, EPI_ISL_661846 to 661852, EPI_ISL_661855, EPI_ISL_661857 to 661862, EPI_ISL_661864 to 661868, EPI_ISL_661870 to 661872, EPI_ISL_661875, EPI_ISL_661877 to 661880, EPI_ISL_661882, EPI_ISL_661884 to 661885, EPI_ISL_661888 to 661894, EPI_ISL_661896, EPI_ISL_661898 to 661899, EPI_ISL_661901, EPI_ISL_661904 to 661916, EPI_ISL_661919 to 661924, EPI_ISL_661927, EPI_ISL_661929, EPI_ISL_661931 to 661940, EPI_ISL_661942 to 661943, EPI_ISL_661945 to 661946, EPI_ISL_661948 to 661950, EPI_ISL_661952, EPI_ISL_661955, EPI_ISL_661957, EPI_ISL_661959 to 661960, EPI_ISL_661963, EPI_ISL_661967, EPI_ISL_661969, EPI_ISL_661971 to 661973, EPI_ISL_661975 to 661976, EPI_ISL_661979, EPI_ISL_661981, EPI_ISL_661984, EPI_ISL_661987, EPI_ISL_661993 to 661994, EPI_ISL_661996 to 661997, EPI_ISL_662003, EPI_ISL_662005, EPI_ISL_662007, EPI_ISL_662009, EPI_ISL_662011 to 662012, EPI_ISL_662016, EPI_ISL_662021, EPI_ISL_662023 to 662024, EPI_ISL_662026 to 662028, EPI_ISL_662030, EPI_ISL_662033, EPI_ISL_662035, EPI_ISL_662037, EPI_ISL_662039 to 662042, EPI_ISL_662044, EPI_ISL_662052, EPI_ISL_662054, EPI_ISL_662059, EPI_ISL_662064, EPI_ISL_662066, EPI_ISL_662069, EPI_ISL_662072 to 662073, EPI_ISL_662075 to 662077, EPI_ISL_662082 to 662085, EPI_ISL_662087 to 662092, EPI_ISL_662094 to 662096, EPI_ISL_662102 to 662104, EPI_ISL_662109 to 662113, EPI_ISL_662117, EPI_ISL_662119, EPI_ISL_662121, EPI_ISL_662125 to 662128, EPI_ISL_662130, EPI_ISL_662134, EPI_ISL_662136, EPI_ISL_662139, EPI_ISL_662142, EPI_ISL_662144, EPI_ISL_662147, EPI_ISL_662149, EPI_ISL_662152 to 662155, EPI_ISL_662157 to 662158, EPI_ISL_662161, EPI_ISL_662168, EPI_ISL_662171 to 662172, EPI_ISL_662180, EPI_ISL_662188 to 662190, EPI_ISL_662193, EPI_ISL_662195, EPI_ISL_662197, EPI_ISL_662199, EPI_ISL_662201, EPI_ISL_662204 to 662205, EPI_ISL_662210 to 662211, EPI_ISL_662213, EPI_ISL_662216, EPI_ISL_662220 to 662222, EPI_ISL_662224 to 662226, EPI_ISL_662230, EPI_ISL_662232, EPI_ISL_662241 to 662243, EPI_ISL_662246, EPI_ISL_662250 to 662251, EPI_ISL_662253, EPI_ISL_662256 to 662257, EPI_ISL_662259, EPI_ISL_662261 to 662262, EPI_ISL_662264, EPI_ISL_662268, EPI_ISL_662271, EPI_ISL_662277 to 662278, EPI_ISL_662280, EPI_ISL_662282 to 662284, EPI_ISL_662287 to 662289, EPI_ISL_662291, EPI_ISL_662293 to 662295, EPI_ISL_662297 to 662299, EPI_ISL_662301 to 662304, EPI_ISL_662306 to 662307, EPI_ISL_662310 to 662311, EPI_ISL_662313 to 662319, EPI_ISL_662321 to 662326, EPI_ISL_662328 to 662333, EPI_ISL_662335 to 662341, EPI_ISL_662345 to 662351, EPI_ISL_662353, EPI_ISL_662355, EPI_ISL_662357, EPI_ISL_662360 to 662364, EPI_ISL_662366 to 662368, EPI_ISL_662370 to 662372, EPI_ISL_662376 to 662380, EPI_ISL_662383 to 662384, EPI_ISL_662386 to 662388, EPI_ISL_662392, EPI_ISL_662395 to 662397, EPI_ISL_662399 to 662402, EPI_ISL_662404, EPI_ISL_662406 to 662409, EPI_ISL_662413 to 662414, EPI_ISL_662416 to 662417, EPI_ISL_662420 to 662426, EPI_ISL_662428, EPI_ISL_662430, EPI_ISL_662432, EPI_ISL_662434 to 662443, EPI_ISL_662445, EPI_ISL_662447 to 662451, EPI_ISL_662453 to 662456, EPI_ISL_662458 to 662459, EPI_ISL_662461, EPI_ISL_662464 to 662467, EPI_ISL_662469, EPI_ISL_662471, EPI_ISL_662473, EPI_ISL_662475 to 662476, EPI_ISL_662479, EPI_ISL_662481 to 662485, EPI_ISL_662490 to 662491, EPI_ISL_662493 to 662495, EPI_ISL_662497 to 662498, EPI_ISL_662500 to 662501, EPI_ISL_662503 to 662506, EPI_ISL_662509 to 662513, EPI_ISL_662515 to 662517, EPI_ISL_662519, EPI_ISL_662521 to 662522, EPI_ISL_662524 to 662525, EPI_ISL_662527 to 662534, EPI_ISL_662536 to 662548, EPI_ISL_662550 to 662552, EPI_ISL_662554 to 662557, EPI_ISL_662559 to 662563, EPI_ISL_662565 to 662574, EPI_ISL_662577, EPI_ISL_662582, EPI_ISL_662584 to 662587, EPI_ISL_662589 to 662592, EPI_ISL_662594 to 662596, EPI_ISL_662598 to 662601, EPI_ISL_662603, EPI_ISL_662606 to 662608, EPI_ISL_662611 to 662613, EPI_ISL_662615, EPI_ISL_662618 to 662620, EPI_ISL_662627, EPI_ISL_662654 to 662657, EPI_ISL_662659 to 662665, EPI_ISL_662667 to 662668, EPI_ISL_662670 to 662671, EPI_ISL_662675 to 662677, EPI_ISL_662679 to 662680, EPI_ISL_662682 to 662685, EPI_ISL_662688 to 662692, EPI_ISL_662694 to 662696, EPI_ISL_662698 to 662699, EPI_ISL_662701 to 662707, EPI_ISL_662710 to 662711, EPI_ISL_662713, EPI_ISL_662715 to 662716, EPI_ISL_662719 to 662720, EPI_ISL_662723 to

662728, EPI_ISL_662730 to 662735, EPI_ISL_662737, EPI_ISL_662739 to 662740, EPI_ISL_662743 to 662744, EPI_ISL_662746 to 662750, EPI_ISL_662752 to 662758, EPI_ISL_662760 to 662780, EPI_ISL_662782 to 662785, EPI_ISL_662787 to 662790, EPI_ISL_662792 to 662800, EPI_ISL_662802 to 662805, EPI_ISL_662807 to 662809, EPI_ISL_662811, EPI_ISL_662813 to 662815, EPI_ISL_662817 to 662819, EPI_ISL_662822 to 662827, EPI_ISL_662829 to 662831, EPI_ISL_662833 to 662838, EPI_ISL_662840 to 662844, EPI_ISL_662846, EPI_ISL_662848 to 662851, EPI_ISL_662853, EPI_ISL_662855 to 662858, EPI_ISL_662860 to 662868, EPI_ISL_662870 to 662876, EPI_ISL_662878, EPI_ISL_662881 to 662884, EPI_ISL_662886 to 662893, EPI_ISL_662895 to 662951, EPI_ISL_662954 to 662958, EPI_ISL_662960 to 662978, EPI_ISL_662980 to 662983, EPI_ISL_662985 to 663000, EPI_ISL_663002, EPI_ISL_663004 to 663005, EPI_ISL_663007 to 663016, EPI_ISL_663018 to 663027, EPI_ISL_663029 to 663039, EPI_ISL_663041 to 663057, EPI_ISL_663059 to 663104, EPI_ISL_663106 to 663126, EPI_ISL_663128 to 663156, EPI_ISL_663158 to 663160, EPI_ISL_663162 to 663196, EPI_ISL_663198 to 663205, EPI_ISL_667814, EPI_ISL_667816, EPI_ISL_667822, EPI_ISL_667824 to 667825, EPI_ISL_667828, EPI_ISL_667831 to 667832, EPI_ISL_667836, EPI_ISL_667838, EPI_ISL_667843, EPI_ISL_667847, EPI_ISL_667849 to 667851, EPI_ISL_667855, EPI_ISL_667857, EPI_ISL_667859 to 667860, EPI_ISL_667863 to 667864, EPI_ISL_667866 to 667868, EPI_ISL_667872 to 667873, EPI_ISL_667881, EPI_ISL_667883, EPI_ISL_667885, EPI_ISL_667887 to 667888, EPI_ISL_667891, EPI_ISL_667893, EPI_ISL_667895, EPI_ISL_667898, EPI_ISL_667900, EPI_ISL_667904, EPI_ISL_667907, EPI_ISL_667912, EPI_ISL_667915, EPI_ISL_667917, EPI_ISL_667919 to 667920, EPI_ISL_667922, EPI_ISL_667924 to 667925, EPI_ISL_667927 to 667931, EPI_ISL_667938 to 667941, EPI_ISL_667943, EPI_ISL_667947 to 667948, EPI_ISL_667950, EPI_ISL_667952, EPI_ISL_667957 to 667960, EPI_ISL_667962 to 667963, EPI_ISL_667966 to 667967, EPI_ISL_667969 to 667970, EPI_ISL_667972 to 667973, EPI_ISL_667975 to 667978, EPI_ISL_667980, EPI_ISL_667982 to 667983, EPI_ISL_667986 to 667987, EPI_ISL_667989 to 667990, EPI_ISL_667992, EPI_ISL_667996 to 667998, EPI_ISL_668000 to 668001, EPI_ISL_668004 to 668006, EPI_ISL_668010, EPI_ISL_668013, EPI_ISL_668020 to 668022, EPI_ISL_668024, EPI_ISL_668028 to 668031, EPI_ISL_668033 to 668036, EPI_ISL_668038, EPI_ISL_668044 to 668045, EPI_ISL_668049 to 668050, EPI_ISL_668052 to 668057, EPI_ISL_668060, EPI_ISL_668062 to 668064, EPI_ISL_668066 to 668067, EPI_ISL_668073 to 668075, EPI_ISL_668078, EPI_ISL_668082 to 668084, EPI_ISL_668093 to 668096, EPI_ISL_668102, EPI_ISL_668105 to 668107, EPI_ISL_668111, EPI_ISL_668113 to 668114, EPI_ISL_668116 to 668121, EPI_ISL_668124 to 668126, EPI_ISL_668129 to 668130, EPI_ISL_668132, EPI_ISL_668135, EPI_ISL_668144, EPI_ISL_668150, EPI_ISL_668152, EPI_ISL_668157, EPI_ISL_668161 to 668162, EPI_ISL_668167 to 668168, EPI_ISL_668170, EPI_ISL_668173 to 668177, EPI_ISL_668181 to 668183, EPI_ISL_668186, EPI_ISL_668193, EPI_ISL_668202 to 668203, EPI_ISL_668205, EPI_ISL_668208 to 668210, EPI_ISL_668213, EPI_ISL_668217 to 668218, EPI_ISL_668220 to 668221, EPI_ISL_668223 to 668224, EPI_ISL_668226, EPI_ISL_668230 to 668232, EPI_ISL_668235 to 668236, EPI_ISL_668238, EPI_ISL_668240 to 668243, EPI_ISL_668248 to 668249, EPI_ISL_668253, EPI_ISL_668256 to 668260, EPI_ISL_668264, EPI_ISL_668266, EPI_ISL_668269, EPI_ISL_668271, EPI_ISL_668273, EPI_ISL_668276 to 668278, EPI_ISL_668282, EPI_ISL_668284, EPI_ISL_668286 to 668288, EPI_ISL_668290 to 668291, EPI_ISL_668303, EPI_ISL_668306 to 668308, EPI_ISL_668312, EPI_ISL_668318, EPI_ISL_668320 to 668321, EPI_ISL_668325, EPI_ISL_668328, EPI_ISL_668330 to 668334, EPI_ISL_668338 to 668340, EPI_ISL_668344, EPI_ISL_668346 to 668347, EPI_ISL_668349, EPI_ISL_668352, EPI_ISL_668357, EPI_ISL_668359 to 668360, EPI_ISL_668362, EPI_ISL_668364, EPI_ISL_668366, EPI_ISL_668372, EPI_ISL_668380 to 668381, EPI_ISL_668383, EPI_ISL_668387, EPI_ISL_672753 to 672754, EPI_ISL_672757 to 672758, EPI_ISL_672760 to 672761, EPI_ISL_672764 to 672765, EPI_ISL_672767 to 672770, EPI_ISL_672774, EPI_ISL_672777 to 672782, EPI_ISL_672786 to 672791, EPI_ISL_672796 to 672799, EPI_ISL_672804 to 672805, EPI_ISL_672807, EPI_ISL_672810 to 672811, EPI_ISL_672813, EPI_ISL_672819, EPI_ISL_672824 to 672826, EPI_ISL_672829, EPI_ISL_672834 to 672838, EPI_ISL_672840 to 672841, EPI_ISL_672843 to 672846, EPI_ISL_672848, EPI_ISL_672850, EPI_ISL_672853 to 672854, EPI_ISL_672857 to 672864, EPI_ISL_672866 to 672870, EPI_ISL_672872, EPI_ISL_672874, EPI_ISL_672876, EPI_ISL_672878 to 672879, EPI_ISL_672881, EPI_ISL_672884, EPI_ISL_672886, EPI_ISL_672889, EPI_ISL_672893 to 672895, EPI_ISL_672897 to 672900, EPI_ISL_672902 to 672904, EPI_ISL_672907 to 672908, EPI_ISL_672911, EPI_ISL_674498 to 674499, EPI_ISL_674501, EPI_ISL_674514, EPI_ISL_674516, EPI_ISL_674522, EPI_ISL_674527 to 674528, EPI_ISL_674543 to 674545, EPI_ISL_674550, EPI_ISL_674558, EPI_ISL_674563, EPI_ISL_674565, EPI_ISL_674569, EPI_ISL_674585, EPI_ISL_674589 to 674590, EPI_ISL_674592, EPI_ISL_674595, EPI_ISL_674603, EPI_ISL_674605, EPI_ISL_674609, EPI_ISL_674613, EPI_ISL_674635, EPI_ISL_674652, EPI_ISL_674655, EPI_ISL_674657 to 674658, EPI_ISL_674661 to 674662, EPI_ISL_674667, EPI_ISL_674678, EPI_ISL_674685, EPI_ISL_674690 to 674692, EPI_ISL_674698, EPI_ISL_674707, EPI_ISL_674714, EPI_ISL_674716 to 674717, EPI_ISL_674723 to 674725, EPI_ISL_674736, EPI_ISL_674740, EPI_ISL_674743 to 674747, EPI_ISL_674751 to 674752, EPI_ISL_674762 to 674763, EPI_ISL_674772, EPI_ISL_674779, EPI_ISL_674781, EPI_ISL_674786, EPI_ISL_674791, EPI_ISL_674794, EPI_ISL_674808 to 674809, EPI_ISL_676313 to 676382, EPI_ISL_676384 to 676388, EPI_ISL_676390 to 676440, EPI_ISL_676442 to 676444, EPI_ISL_676446 to 676449, EPI_ISL_676451 to 676483, EPI_ISL_676486 to 676488, EPI_ISL_703262 to 703263, EPI_ISL_703270, EPI_ISL_703273, EPI_ISL_703280, EPI_ISL_703285 to 703286, EPI_ISL_703290 to 703291, EPI_ISL_703296, EPI_ISL_703300, EPI_ISL_703304, EPI_ISL_703309, EPI_ISL_703315, EPI_ISL_703320, EPI_ISL_703323, EPI_ISL_703325, EPI_ISL_703331, EPI_ISL_703334, EPI_ISL_703337, EPI_ISL_703345, EPI_ISL_703350, EPI_ISL_703352, EPI_ISL_703354 to 703355, EPI_ISL_703358, EPI_ISL_703362, EPI_ISL_703370 to 703371, EPI_ISL_703378 to 703379, EPI_ISL_703381 to 703382, EPI_ISL_703384, EPI_ISL_703389, EPI_ISL_703391, EPI_ISL_703393, EPI_ISL_703399, EPI_ISL_703401, EPI_ISL_703407 to 703408, EPI_ISL_703410 to 703411, EPI_ISL_703422, EPI_ISL_703426, EPI_ISL_703428 to 703429, EPI_ISL_703431, EPI_ISL_703439, EPI_ISL_703442, EPI_ISL_703444 to 703445, EPI_ISL_703447, EPI_ISL_703456, EPI_ISL_703458, EPI_ISL_703463, EPI_ISL_703469, EPI_ISL_703472, EPI_ISL_703474, EPI_ISL_703482, EPI_ISL_703486, EPI_ISL_703491, EPI_ISL_703496, EPI_ISL_703499, EPI_ISL_703501, EPI_ISL_703506, EPI_ISL_703510, EPI_ISL_703514 to 703515, EPI_ISL_703517, EPI_ISL_703522, EPI_ISL_703525, EPI_ISL_703529, EPI_ISL_703534, EPI_ISL_703538, EPI_ISL_703541, EPI_ISL_703546, EPI_ISL_703550, EPI_ISL_703553, EPI_ISL_703557, EPI_ISL_703560 to 703561, EPI_ISL_703578, EPI_ISL_703583 to 703584, EPI_ISL_703598, EPI_ISL_703600, EPI_ISL_703607 to 703608, EPI_ISL_703612, EPI_ISL_703617, EPI_ISL_703626, EPI_ISL_703631, EPI_ISL_703633 to 703634, EPI_ISL_703640 to 703641, EPI_ISL_703643, EPI_ISL_703645, EPI_ISL_703650 to 703651, EPI_ISL_703653 to 703654, EPI_ISL_703657, EPI_ISL_703659, EPI_ISL_703661, EPI_ISL_703669, EPI_ISL_703672 to 703673, EPI_ISL_703677, EPI_ISL_703685, EPI_ISL_703694, EPI_ISL_703697, EPI_ISL_703700, EPI_ISL_703703, EPI_ISL_703786, EPI_ISL_703790, EPI_ISL_703799, EPI_ISL_703801, EPI_ISL_703806, EPI_ISL_703808, EPI_ISL_703816, EPI_ISL_703819 to 703820, EPI_ISL_703822, EPI_ISL_703824 to 703825, EPI_ISL_703827, EPI_ISL_703830, EPI_ISL_703833, EPI_ISL_703836, EPI_ISL_703838, EPI_ISL_703840 to 703841, EPI_ISL_703843, EPI_ISL_703851 to 703852, EPI_ISL_703854, EPI_ISL_703857, EPI_ISL_703859, EPI_ISL_703861 to 703862, EPI_ISL_703864, EPI_ISL_703866 to 703867, EPI_ISL_703869, EPI_ISL_703874 to 703875, EPI_ISL_703877 to 703878, EPI_ISL_703880, EPI_ISL_703882 to 703883, EPI_ISL_703887, EPI_ISL_703889 to 703890, EPI_ISL_703895, EPI_ISL_703900 to 703901, EPI_ISL_703904, EPI_ISL_703906 to 703907, EPI_ISL_703909 to 703910, EPI_ISL_703912, EPI_ISL_703914, EPI_ISL_703917, EPI_ISL_703919, EPI_ISL_703925, EPI_ISL_703928, EPI_ISL_703930, EPI_ISL_703933 to 703934, EPI_ISL_703936, EPI_ISL_703938, EPI_ISL_703941, EPI_ISL_703943 to 703944, EPI_ISL_703949, EPI_ISL_703951, EPI_ISL_703956 to 703957, EPI_ISL_703959, EPI_ISL_703962, EPI_ISL_703964 to 703965, EPI_ISL_703967, EPI_ISL_703972 to 703973, EPI_ISL_703976, EPI_ISL_703978, EPI_ISL_703980 to 703981, EPI_ISL_703983 to 703984, EPI_ISL_703986, EPI_ISL_703988 to 703989, EPI_ISL_703991, EPI_ISL_703993 to 703994, EPI_ISL_703997, EPI_ISL_703999 to 704000, EPI_ISL_704002, EPI_ISL_704004 to 704005, EPI_ISL_704007 to 704008, EPI_ISL_704010, EPI_ISL_704013, EPI_ISL_704015 to 704016, EPI_ISL_704018 to 704019, EPI_ISL_704023 to 704024, EPI_ISL_704026 to 704027, EPI_ISL_704029, EPI_ISL_704031 to 704032, EPI_ISL_704034 to 704035, EPI_ISL_704037, EPI_ISL_704040, EPI_ISL_704042 to 704043, EPI_ISL_704045, EPI_ISL_704047 to 704048, EPI_ISL_704051, EPI_ISL_704053, EPI_ISL_704055 to 704056, EPI_ISL_704058 to 704059, EPI_ISL_704061, EPI_ISL_704063 to 704064, EPI_ISL_704066 to 704067, EPI_ISL_704071 to 704072, EPI_ISL_704074, EPI_ISL_704077 to 704078, EPI_ISL_704080, EPI_ISL_704082 to 704083, EPI_ISL_704085, EPI_ISL_704088, EPI_ISL_704090 to 704091, EPI_ISL_704093 to 704094, EPI_ISL_704096, EPI_ISL_704098 to 704099, EPI_ISL_704101, EPI_ISL_704104, EPI_ISL_704106 to 704107, EPI_ISL_704109 to 704110, EPI_ISL_704114, EPI_ISL_704116 to 704117, EPI_ISL_704119, EPI_ISL_704121 to 704122, EPI_ISL_704124 to 704125, EPI_ISL_704127 to 704128, EPI_ISL_704130, EPI_ISL_704133, EPI_ISL_704135 to 704136, EPI_ISL_704140 to 704141, EPI_ISL_704143 to 704144, EPI_ISL_704146, EPI_ISL_704151 to 704152, EPI_ISL_704154, EPI_ISL_704157, EPI_ISL_704159, EPI_ISL_704162, EPI_ISL_704164 to 704165, EPI_ISL_704167, EPI_ISL_704169 to 704170, EPI_ISL_704172 to 704173, EPI_ISL_704175, EPI_ISL_704177 to 704178, EPI_ISL_704180, EPI_ISL_704182 to 704183, EPI_ISL_704185 to 704186, EPI_ISL_704188 to 704189, EPI_ISL_704191 to 704192, EPI_ISL_704197, EPI_ISL_704199 to 704200, EPI_ISL_704202, EPI_ISL_704204 to 704205, EPI_ISL_704207 to 704208, EPI_ISL_704211 to 704216, EPI_ISL_704218, EPI_ISL_704220 to 704221, EPI_ISL_704223 to 704224, EPI_ISL_704226 to 704227, EPI_ISL_704229 to 704230, EPI_ISL_704232, EPI_ISL_704234 to 704235, EPI_ISL_704237, EPI_ISL_704239 to 704240, EPI_ISL_704242, EPI_ISL_704244 to 704246, EPI_ISL_704248 to 704249, EPI_ISL_704251, EPI_ISL_704253 to 704254, EPI_ISL_704256, EPI_ISL_704258, EPI_ISL_704261, EPI_ISL_704263 to 704265, EPI_ISL_704267 to 704268, EPI_ISL_704270, EPI_ISL_704272, EPI_ISL_704274 to 704275, EPI_ISL_704277 to 704279, EPI_ISL_704281 to 704282, EPI_ISL_704284 to 704285, EPI_ISL_704287 to 704288, EPI_ISL_704290, EPI_ISL_704292 to 704293, EPI_ISL_704295 to 704296, EPI_ISL_704298, EPI_ISL_704300 to 704301, EPI_ISL_704303 to 704304, EPI_ISL_704306, EPI_ISL_704310 to 704311, EPI_ISL_704313 to 704314, EPI_ISL_704316, EPI_ISL_704318 to 704319, EPI_ISL_704321, EPI_ISL_704323 to 704324, EPI_ISL_704326 to 704327, EPI_ISL_704329 to 704330, EPI_ISL_704332, EPI_ISL_704334 to 704335, EPI_ISL_704337 to 704338, EPI_ISL_704340, EPI_ISL_704342 to 704343, EPI_ISL_704345 to 704346, EPI_ISL_704348 to 704349, EPI_ISL_704351 to 704352, EPI_ISL_704354 to 704355, EPI_ISL_704357 to 704358, EPI_ISL_704360, EPI_ISL_704362 to 704363, EPI_ISL_704365, EPI_ISL_704367 to 704368, EPI_ISL_704370 to 704371, EPI_ISL_704373, EPI_ISL_704375 to 704376, EPI_ISL_704378, EPI_ISL_704380 to 704381, EPI_ISL_704383, EPI_ISL_704385 to 704386, EPI_ISL_704388, EPI_ISL_704390 to 704391, EPI_ISL_704394, EPI_ISL_704396 to 704397, EPI_ISL_704399 to 704400, EPI_ISL_704402, EPI_ISL_704404 to 704405, EPI_ISL_704407, EPI_ISL_704410, EPI_ISL_704412, EPI_ISL_704414 to 704415, EPI_ISL_704417 to 704418, EPI_ISL_704420 to 704421, EPI_ISL_704423 to 704424, EPI_ISL_704426, EPI_ISL_704428 to 704429, EPI_ISL_704431 to 704432, EPI_ISL_704434, EPI_ISL_704436 to 704437, EPI_ISL_704439 to 704440, EPI_ISL_704442, EPI_ISL_704444 to 704445, EPI_ISL_704447 to 704448, EPI_ISL_704450 to 704451, EPI_ISL_704453, EPI_ISL_704455 to 704456, EPI_ISL_704458, EPI_ISL_704461, EPI_ISL_704463 to 704464, EPI_ISL_704466, EPI_ISL_704468 to 704469, EPI_ISL_704471 to 704472, EPI_ISL_704474, EPI_ISL_704476 to 704477, EPI_ISL_704479 to 704480, EPI_ISL_704482 to 704483, EPI_ISL_704485, EPI_ISL_704487 to 704488, EPI_ISL_704490 to 704491, EPI_ISL_704493, EPI_ISL_704496, EPI_ISL_704498, EPI_ISL_704500 to 704501, EPI_ISL_704503 to 704504, EPI_ISL_704506, EPI_ISL_704509, EPI_ISL_704511 to 704512, EPI_ISL_704514 to 704515, EPI_ISL_704519 to 704520, EPI_ISL_704522 to 704523, EPI_ISL_704525, EPI_ISL_704527 to 704528, EPI_ISL_704530 to 704531, EPI_ISL_704533 to 704534, EPI_ISL_704536, EPI_ISL_704538 to 704539, EPI_ISL_704541 to 704542, EPI_ISL_704544, EPI_ISL_704546 to 704547, EPI_ISL_704549 to 704550, EPI_ISL_704552 to 704553, EPI_ISL_704556, EPI_ISL_704558, EPI_ISL_704560 to 704561, EPI_ISL_704563, EPI_ISL_704565 to 704566, EPI_ISL_704568 to 704569, EPI_ISL_704571, EPI_ISL_704642, EPI_ISL_704644 to 704645, EPI_ISL_704647, EPI_ISL_704649 to 704650, EPI_ISL_704652 to 704653, EPI_ISL_704655, EPI_ISL_704657 to 704658, EPI_ISL_704660 to 704661, EPI_ISL_704663, EPI_ISL_704665, EPI_ISL_704667 to 704668, EPI_ISL_704670, EPI_ISL_704672 to 704673, EPI_ISL_704675, EPI_ISL_704678, EPI_ISL_704680 to 704681, EPI_ISL_704683 to 704684, EPI_ISL_704686, EPI_ISL_708843, EPI_ISL_708855, EPI_ISL_708884, EPI_ISL_708890, EPI_ISL_708905 to 708906, EPI_ISL_708910, EPI_ISL_708921 to 708922, EPI_ISL_708925, EPI_ISL_708933, EPI_ISL_708953, EPI_ISL_708957, EPI_ISL_708985, EPI_ISL_708991, EPI_ISL_708998, EPI_ISL_709004 to 709005, EPI_ISL_709009, EPI_ISL_709013, EPI_ISL_709022, EPI_ISL_709031, EPI_ISL_709036, EPI_ISL_709040, EPI_ISL_709048, EPI_ISL_709056, EPI_ISL_709058, EPI_ISL_709060, EPI_ISL_709065, EPI_ISL_709077 to 709079, EPI_ISL_709082, EPI_ISL_709091, EPI_ISL_709098, EPI_ISL_709108, EPI_ISL_709119, EPI_ISL_709126, EPI_ISL_709131 to 709133, EPI_ISL_709135, EPI_ISL_709139 to 709142, EPI_ISL_709144, EPI_ISL_709150 to 709151, EPI_ISL_709160 to 709161, EPI_ISL_709170 to 709171, EPI_ISL_709174, EPI_ISL_709182, EPI_ISL_709189 to 709191, EPI_ISL_709199, EPI_ISL_709201 to 709202, EPI_ISL_709215, EPI_ISL_709231, EPI_ISL_709234, EPI_ISL_709239, EPI_ISL_709242 to 709243, EPI_ISL_709245, EPI_ISL_709248, EPI_ISL_709264, EPI_ISL_709274, EPI_ISL_709277, EPI_ISL_709282 to 709283, EPI_ISL_709285, EPI_ISL_709290 to 709292, EPI_ISL_709301, EPI_ISL_709307, EPI_ISL_709311 to 709312, EPI_ISL_709314, EPI_ISL_709320, EPI_ISL_709324, EPI_ISL_709327, EPI_ISL_709332, EPI_ISL_709334, EPI_ISL_709336, EPI_ISL_709343, EPI_ISL_709345, EPI_ISL_709353 to 709354, EPI_ISL_709359 to 709360, EPI_ISL_709364 to 709367, EPI_ISL_709373, EPI_ISL_709376, EPI_ISL_709378, EPI_ISL_709380, EPI_ISL_709383 to 709384, EPI_ISL_709392, EPI_ISL_709395 to 709396, EPI_ISL_709399, EPI_ISL_709403 to 709404, EPI_ISL_709406 to 709407, EPI_ISL_709409, EPI_ISL_709412, EPI_ISL_709417, EPI_ISL_709419, EPI_ISL_709422 to 709424, EPI_ISL_709426, EPI_ISL_709429, EPI_ISL_709436, EPI_ISL_709438 to 709441, EPI_ISL_709443, EPI_ISL_709446, EPI_ISL_709450, EPI_ISL_709453 to 709454, EPI_ISL_709456 to 709459, EPI_ISL_709464 to 709466, EPI_ISL_709469, EPI_ISL_709472, EPI_ISL_709474, EPI_ISL_709481 to 709482, EPI_ISL_709486 to 709487, EPI_ISL_709490, EPI_ISL_709493 to 709494, EPI_ISL_709496 to 709498, EPI_ISL_709500, EPI_ISL_709502 to 709503, EPI_ISL_709509, EPI_ISL_709511, EPI_ISL_709515, EPI_ISL_709517 to 709518, EPI_ISL_709522, EPI_ISL_709524 to 709525, EPI_ISL_709527 to 709528, EPI_ISL_709530, EPI_ISL_709533, EPI_ISL_709537, EPI_ISL_709539 to 709541, EPI_ISL_709549, EPI_ISL_709555 to 709558, EPI_ISL_709561, EPI_ISL_709565, EPI_ISL_709800, EPI_ISL_709806, EPI_ISL_709809, EPI_ISL_709811 to 709812, EPI_ISL_709816, EPI_ISL_709818, EPI_ISL_709820, EPI_ISL_709826, EPI_ISL_709831 to 709832, EPI_ISL_709834 to 709835, EPI_ISL_709840, EPI_ISL_709845, EPI_ISL_709848 to 709849, EPI_ISL_709851, EPI_ISL_709857 to 709859, EPI_ISL_709861 to 709862, EPI_ISL_709875, EPI_ISL_710615, EPI_ISL_710619, EPI_ISL_710621 to 710623, EPI_ISL_710626 to 710627, EPI_ISL_710629, EPI_ISL_710633 to 710635, EPI_ISL_710637, EPI_ISL_710640 to 710644, EPI_ISL_710646, EPI_ISL_710651 to 710652, EPI_ISL_710654 to 710655, EPI_ISL_710657 to 710660, EPI_ISL_710664, EPI_ISL_710666 to 710670, EPI_ISL_710672 to 710675, EPI_ISL_710677 to 710680, EPI_ISL_710684 to 710686, EPI_ISL_710688, EPI_ISL_710690 to 710692, EPI_ISL_710696 to 710697, EPI_ISL_710748, EPI_ISL_710753 to 710755, EPI_ISL_710758 to 710766, EPI_ISL_710768, EPI_ISL_710770 to 710772, EPI_ISL_710774 to 710777, EPI_ISL_710779 to 710790, EPI_ISL_710793 to 710820, EPI_ISL_710831 to 710832, EPI_ISL_710838 to 710841, EPI_ISL_710843 to 710846, EPI_ISL_710848 to 710855, EPI_ISL_710857 to 710859, EPI_ISL_710861 to 710864, EPI_ISL_710867 to 710868, EPI_ISL_710872, EPI_ISL_710877, EPI_ISL_710879 to 710886, EPI_ISL_710903 to 710906, EPI_ISL_710995 to 711007, EPI_ISL_711014, EPI_ISL_711048 to 711049, EPI_ISL_711053 to 711054, EPI_ISL_719605 to 719613, EPI_ISL_719615 to 719626, EPI_ISL_719629, EPI_ISL_719631, EPI_ISL_719633 to 719637, EPI_ISL_719640 to 719641, EPI_ISL_719643 to 719644, EPI_ISL_719646, EPI_ISL_719648, EPI_ISL_719650 to 719653, EPI_ISL_719655 to 719658, EPI_ISL_719660 to 719666, EPI_ISL_719668 to 719673, EPI_ISL_719678 to 719681, EPI_ISL_719683 to 719688, EPI_ISL_719690 to 719695, EPI_ISL_719698 to 719705, EPI_ISL_719707 to 719710, EPI_ISL_719712 to 719717, EPI_ISL_719719 to 719726, EPI_ISL_719728 to 719733, EPI_ISL_719735 to 719736, EPI_ISL_719738 to 719740, EPI_ISL_719742, EPI_ISL_719744, EPI_ISL_719746 to 719753, EPI_ISL_719755 to 719756, EPI_ISL_719758 to 719761, EPI_ISL_719763 to 719764, EPI_ISL_719766, EPI_ISL_719768 to 719775, EPI_ISL_719777, EPI_ISL_719779 to 719783, EPI_ISL_719786 to 719793, EPI_ISL_719796, EPI_ISL_719798 to 719800, EPI_ISL_719802 to 719819, EPI_ISL_719821 to 719824, EPI_ISL_719826 to 719853, EPI_ISL_719855 to 719859, EPI_ISL_719861 to 719880, EPI_ISL_719882 to 719892, EPI_ISL_719894 to 719895, EPI_ISL_719897 to 719904, EPI_ISL_719906 to 719914, EPI_ISL_719916 to 719930, EPI_ISL_719932 to 719954, EPI_ISL_719956 to 719970, EPI_ISL_719972 to 719975, EPI_ISL_719977 to 719980, EPI_ISL_719982 to 719986, EPI_ISL_720668, EPI_ISL_720672, EPI_ISL_720684, EPI_ISL_720704, EPI_ISL_720728, EPI_ISL_720732 to 720733, EPI_ISL_720738, EPI_ISL_720749, EPI_ISL_720752, EPI_ISL_720755, EPI_ISL_720761, EPI_ISL_720767, EPI_ISL_720776, EPI_ISL_720786 to 720787, EPI_ISL_720792, EPI_ISL_720802, EPI_ISL_720811, EPI_ISL_720819, EPI_ISL_720831, EPI_ISL_720834, EPI_ISL_720841, EPI_ISL_720850, EPI_ISL_720856, EPI_ISL_720870, EPI_ISL_720877, EPI_ISL_720889, EPI_ISL_720897, EPI_ISL_720910 to 720915, EPI_ISL_720917 to 720950, EPI_ISL_720952 to 720955, EPI_ISL_720957 to 721016, EPI_ISL_721018 to 721031, EPI_ISL_721033 to 721036, EPI_ISL_721038 to 721043, EPI_ISL_721045 to 721064, EPI_ISL_721066 to 721068, EPI_ISL_721070 to 721081, EPI_ISL_721083 to 721096, EPI_ISL_721098 to 721115, EPI_ISL_721117 to 721121, EPI_ISL_721123 to 721134, EPI_ISL_721136 to 721142, EPI_ISL_721144 to 721157, EPI_ISL_721159, EPI_ISL_721161 to 721166, EPI_ISL_721168 to 721180, EPI_ISL_721182 to 721184, EPI_ISL_721519, EPI_ISL_721521, EPI_ISL_721533 to 721543, EPI_ISL_721545 to 721567, EPI_ISL_728300, EPI_ISL_730656 to 730657, EPI_ISL_730659, EPI_ISL_730664, EPI_ISL_730673, EPI_ISL_730675 to 730676, EPI_ISL_730678, EPI_ISL_730685, EPI_ISL_730697, EPI_ISL_730705, EPI_ISL_730709 to 730710, EPI_ISL_730718, EPI_ISL_730723, EPI_ISL_730737, EPI_ISL_730752, EPI_ISL_730759, EPI_ISL_730765, EPI_ISL_730777 to 730778, EPI_ISL_730780, EPI_ISL_730785, EPI_ISL_730789, EPI_ISL_730793, EPI_ISL_730795 to 730796, EPI_ISL_730801 to 730806, EPI_ISL_730808 to 730809, EPI_ISL_730813, EPI_ISL_730815, EPI_ISL_730824, EPI_ISL_730828, EPI_ISL_730831 to 730833, EPI_ISL_730840, EPI_ISL_730846 to 730847, EPI_ISL_730849 to 730850, EPI_ISL_730858, EPI_ISL_730860, EPI_ISL_730862, EPI_ISL_730864, EPI_ISL_730867, EPI_ISL_730869, EPI_ISL_730875 to 730877, EPI_ISL_730887, EPI_ISL_730889, EPI_ISL_730894, EPI_ISL_730898, EPI_ISL_730908, EPI_ISL_730910 to 730911, EPI_ISL_730913, EPI_ISL_731255 to 731259, EPI_ISL_731261 to 731262, EPI_ISL_731264, EPI_ISL_731267, EPI_ISL_731271, EPI_ISL_731273, EPI_ISL_731280 to 731284, EPI_ISL_731286 to 731287, EPI_ISL_731291, EPI_ISL_731293, EPI_ISL_731295, EPI_ISL_731299, EPI_ISL_731305, EPI_ISL_731309 to 731310, EPI_ISL_731314, EPI_ISL_731319 to 731321, EPI_ISL_731326 to 731327, EPI_ISL_731330, EPI_ISL_731332 to 731334, EPI_ISL_731337 to 731338, EPI_ISL_731341, EPI_ISL_731345 to 731347, EPI_ISL_731349, EPI_ISL_731353 to 731354, EPI_ISL_731357, EPI_ISL_731361, EPI_ISL_731367 to 731368, EPI_ISL_731372 to 731373, EPI_ISL_731376, EPI_ISL_731378, EPI_ISL_731380, EPI_ISL_731382, EPI_ISL_731386, EPI_ISL_731390, EPI_ISL_731392 to 731393, EPI_ISL_731396, EPI_ISL_731398, EPI_ISL_731400 to 731407, EPI_ISL_731411, EPI_ISL_731414, EPI_ISL_731417, EPI_ISL_731421, EPI_ISL_731425 to 731426, EPI_ISL_731431 to 731432, EPI_ISL_731435, EPI_ISL_731438, EPI_ISL_731444, EPI_ISL_731447, EPI_ISL_731449 to 731450, EPI_ISL_731452 to 731454, EPI_ISL_731456, EPI_ISL_731458 to 731459, EPI_ISL_731466, EPI_ISL_731472, EPI_ISL_731476 to 731477, EPI_ISL_731479, EPI_ISL_731482, EPI_ISL_731484 to 731486, EPI_ISL_731488, EPI_ISL_731491, EPI_ISL_731493 to 731494, EPI_ISL_731496 to 731497, EPI_ISL_731499, EPI_ISL_731501, EPI_ISL_731507 to 731508, EPI_ISL_731513, EPI_ISL_731519, EPI_ISL_731535 to 731537, EPI_ISL_731541, EPI_ISL_731548, EPI_ISL_731550 to 731551, EPI_ISL_731894, EPI_ISL_733916 to 733931, EPI_ISL_733934 to 733941, EPI_ISL_733943 to 733944, EPI_ISL_733946 to 733962, EPI_ISL_733964 to 733970, EPI_ISL_733972 to 733984, EPI_ISL_733986 to 734018, EPI_ISL_734020 to 734027, EPI_ISL_734029 to 734042, EPI_ISL_734044 to 734047, EPI_ISL_734050, EPI_ISL_734053 to 734063, EPI_ISL_734065, EPI_ISL_734068 to 734090, EPI_ISL_734093 to 734106, EPI_ISL_734108 to 734118, EPI_ISL_734120 to 734127, EPI_ISL_734129 to 734131, EPI_ISL_734133 to 734139, EPI_ISL_734141 to 734152, EPI_ISL_734154 to 734165, EPI_ISL_735861, EPI_ISL_735864, EPI_ISL_735878, EPI_ISL_735880, EPI_ISL_735886, EPI_ISL_735894, EPI_ISL_735909, EPI_ISL_735920, EPI_ISL_735927, EPI_ISL_735937, EPI_ISL_735939, EPI_ISL_735945, EPI_ISL_735949, EPI_ISL_735952, EPI_ISL_735956, EPI_ISL_735961, EPI_ISL_735965, EPI_ISL_735968, EPI_ISL_735972 to 735973, EPI_ISL_735978, EPI_ISL_735982, EPI_ISL_735987, EPI_ISL_735993, EPI_ISL_735995 to 735996, EPI_ISL_736001, EPI_ISL_736003, EPI_ISL_736007, EPI_ISL_736015, EPI_ISL_736017 to 736019, EPI_ISL_736024 to 736025, EPI_ISL_736027, EPI_ISL_736033, EPI_ISL_736038, EPI_ISL_736041, EPI_ISL_736052, EPI_ISL_736057, EPI_ISL_736059, EPI_ISL_736064, EPI_ISL_736066, EPI_ISL_736072, EPI_ISL_736090 to 736092, EPI_ISL_736096, EPI_ISL_736099 to 736102, EPI_ISL_736104, EPI_ISL_736107, EPI_ISL_736115, EPI_ISL_736124, EPI_ISL_736130 to 736132, EPI_ISL_736135, EPI_ISL_736143, EPI_ISL_736146, EPI_ISL_736149, EPI_ISL_736151, EPI_ISL_736159, EPI_ISL_736162, EPI_ISL_736167 to 736169, EPI_ISL_736175, EPI_ISL_736182 to 736184, EPI_ISL_736190 to 736191, EPI_ISL_736204 to 736205, EPI_ISL_736209, EPI_ISL_736212, EPI_ISL_736220 to 736223, EPI_ISL_736231, EPI_ISL_736234 to 736238, EPI_ISL_736241 to 736243, EPI_ISL_736245, EPI_ISL_736248, EPI_ISL_736250, EPI_ISL_736254 to 736255, EPI_ISL_736257 to 736258, EPI_ISL_736261, EPI_ISL_736266, EPI_ISL_736269, EPI_ISL_736275, EPI_ISL_736277 to 736278, EPI_ISL_736284, EPI_ISL_736287, EPI_ISL_736290, EPI_ISL_736301, EPI_ISL_736304, EPI_ISL_736306, EPI_ISL_736309, EPI_ISL_736311 to 736313, EPI_ISL_736315 to 736316, EPI_ISL_736318, EPI_ISL_736320 to 736321, EPI_ISL_736323, EPI_ISL_736325, EPI_ISL_736327, EPI_ISL_736330 to 736332, EPI_ISL_736334, EPI_ISL_736343, EPI_ISL_736346 to 736347, EPI_ISL_736350, EPI_ISL_736352, EPI_ISL_736355 to 736356, EPI_ISL_736358, EPI_ISL_736362, EPI_ISL_736367, EPI_ISL_736370 to 736371, EPI_ISL_736373, EPI_ISL_736376, EPI_ISL_736378, EPI_ISL_736382 to 736383, EPI_ISL_736386, EPI_ISL_736388 to 736391, EPI_ISL_736394, EPI_ISL_736398 to 736400, EPI_ISL_736404, EPI_ISL_736411 to 736412, EPI_ISL_736414, EPI_ISL_736416 to 736417, EPI_ISL_736419, EPI_ISL_736423, EPI_ISL_736425 to 736426, EPI_ISL_736428 to 736431, EPI_ISL_736437 to 736438, EPI_ISL_736442, EPI_ISL_736444, EPI_ISL_736451, EPI_ISL_736455 to 736456, EPI_ISL_736458 to 736461, EPI_ISL_736464, EPI_ISL_736466, EPI_ISL_736468, EPI_ISL_736470 to 736471, EPI_ISL_736476, EPI_ISL_736478 to 736479, EPI_ISL_736483, EPI_ISL_736488 to 736490, EPI_ISL_736492, EPI_ISL_736503, EPI_ISL_736512, EPI_ISL_736519, EPI_ISL_736529, EPI_ISL_736532 to 736533, EPI_ISL_736556, EPI_ISL_736567, EPI_ISL_736573, EPI_ISL_736580, EPI_ISL_736582, EPI_ISL_736605, EPI_ISL_736620, EPI_ISL_736624, EPI_ISL_736627 to 736628, EPI_ISL_736631, EPI_ISL_736633, EPI_ISL_736646, EPI_ISL_736650, EPI_ISL_736657,

EPI_ISL_736659, EPI_ISL_736668, EPI_ISL_736672, EPI_ISL_736678, EPI_ISL_736685, EPI_ISL_736689, EPI_ISL_736695, EPI_ISL_736700, EPI_ISL_736721, EPI_ISL_736734, EPI_ISL_736736 to 736737, EPI_ISL_736743, EPI_ISL_736745, EPI_ISL_736752, EPI_ISL_736754, EPI_ISL_736774, EPI_ISL_756690, EPI_ISL_756695, EPI_ISL_756702, EPI_ISL_756705 to 756706, EPI_ISL_756709, EPI_ISL_756711, EPI_ISL_756716, EPI_ISL_756723, EPI_ISL_756729, EPI_ISL_756737, EPI_ISL_756744 to 756745, EPI_ISL_756748, EPI_ISL_756750, EPI_ISL_756761, EPI_ISL_756773, EPI_ISL_756775, EPI_ISL_756780, EPI_ISL_756784, EPI_ISL_756789, EPI_ISL_756795, EPI_ISL_756800, EPI_ISL_756803, EPI_ISL_756805, EPI_ISL_756824, EPI_ISL_756835, EPI_ISL_756840, EPI_ISL_756849 to 756850, EPI_ISL_756852, EPI_ISL_756861, EPI_ISL_756867, EPI_ISL_756877 to 756879, EPI_ISL_756881, EPI_ISL_756885 to 756886, EPI_ISL_756888 to 756889, EPI_ISL_756893 to 756895, EPI_ISL_756897, EPI_ISL_756900, EPI_ISL_756909 to 756910, EPI_ISL_756912, EPI_ISL_756918, EPI_ISL_756920, EPI_ISL_756923, EPI_ISL_756928, EPI_ISL_756931, EPI_ISL_756935, EPI_ISL_756940, EPI_ISL_756947, EPI_ISL_756951 to 756953, EPI_ISL_756955, EPI_ISL_756959, EPI_ISL_756970 to 756971, EPI_ISL_756973, EPI_ISL_756977, EPI_ISL_756998, EPI_ISL_757005, EPI_ISL_757007 to 757009, EPI_ISL_757011 to 757012, EPI_ISL_757018, EPI_ISL_757021, EPI_ISL_757027 to 757028, EPI_ISL_757032, EPI_ISL_757034 to 757060, EPI_ISL_757062 to 757067, EPI_ISL_757069 to 757070, EPI_ISL_757072 to 757073, EPI_ISL_757075 to 757077, EPI_ISL_757079 to 757083, EPI_ISL_757085 to 757087, EPI_ISL_757089, EPI_ISL_757091 to 757113, EPI_ISL_757115 to 757118, EPI_ISL_757120 to 757145, EPI_ISL_757147 to 757167, EPI_ISL_757169 to 757182, EPI_ISL_757184, EPI_ISL_757186 to 757194, EPI_ISL_757196 to 757204, EPI_ISL_757207 to 757209, EPI_ISL_757211 to 757228, EPI_ISL_757230 to 757256, EPI_ISL_757258 to 757284, EPI_ISL_760250 to 760258, EPI_ISL_760260, EPI_ISL_760262, EPI_ISL_760264, EPI_ISL_760269 to 760272, EPI_ISL_760274 to 760275, EPI_ISL_760277, EPI_ISL_760279 to 760280, EPI_ISL_760283, EPI_ISL_760287, EPI_ISL_760289 to 760291, EPI_ISL_760294, EPI_ISL_760297, EPI_ISL_760299 to 760300, EPI_ISL_760302 to 760304, EPI_ISL_760307 to 760310, EPI_ISL_760312, EPI_ISL_760314 to 760322, EPI_ISL_760324 to 760325, EPI_ISL_760327 to 760332, EPI_ISL_760334 to 760340, EPI_ISL_760342 to 760343, EPI_ISL_760345, EPI_ISL_760347, EPI_ISL_760349, EPI_ISL_760351 to 760354, EPI_ISL_760356 to 760357, EPI_ISL_760361 to 760363, EPI_ISL_760365 to 760368, EPI_ISL_760373 to 760380, EPI_ISL_760383, EPI_ISL_760385, EPI_ISL_760391, EPI_ISL_760393 to 760396, EPI_ISL_760398 to 760399, EPI_ISL_760401 to 760404, EPI_ISL_760406 to 760407, EPI_ISL_760409 to 760414, EPI_ISL_760417, EPI_ISL_760422 to 760423, EPI_ISL_760425 to 760429, EPI_ISL_760431, EPI_ISL_760433, EPI_ISL_760435, EPI_ISL_760437 to 760440, EPI_ISL_760443 to 760444, EPI_ISL_760446 to 760447, EPI_ISL_760449, EPI_ISL_760451 to 760459, EPI_ISL_760462 to 760470, EPI_ISL_760472 to 760473, EPI_ISL_760476 to 760484, EPI_ISL_760487 to 760488, EPI_ISL_760490, EPI_ISL_760492 to 760494, EPI_ISL_760496 to 760497, EPI_ISL_760499, EPI_ISL_760502, EPI_ISL_760504 to 760507, EPI_ISL_760510 to 760511, EPI_ISL_760513 to 760515, EPI_ISL_760517 to 760520, EPI_ISL_760522 to 760523, EPI_ISL_760525 to 760529, EPI_ISL_760532 to 760533, EPI_ISL_760536 to 760537, EPI_ISL_760539 to 760542, EPI_ISL_760547 to 760548, EPI_ISL_760553, EPI_ISL_760556, EPI_ISL_760563, EPI_ISL_760566, EPI_ISL_760571, EPI_ISL_760573, EPI_ISL_760575 to 760581, EPI_ISL_760583 to 760585, EPI_ISL_760590, EPI_ISL_760592, EPI_ISL_760594, EPI_ISL_760598, EPI_ISL_760602, EPI_ISL_760604 to 760607, EPI_ISL_760610 to 760611, EPI_ISL_760614, EPI_ISL_760616, EPI_ISL_760618 to 760619, EPI_ISL_760621 to 760622, EPI_ISL_760626, EPI_ISL_760630 to 760632, EPI_ISL_760634 to 760636, EPI_ISL_760638, EPI_ISL_760641, EPI_ISL_760643, EPI_ISL_760645 to 760646, EPI_ISL_760648, EPI_ISL_760651, EPI_ISL_760655, EPI_ISL_760657 to 760659, EPI_ISL_760662, EPI_ISL_760668 to 760669, EPI_ISL_760671, EPI_ISL_760673 to 760676, EPI_ISL_760678, EPI_ISL_760681 to 760683, EPI_ISL_760686 to 760687, EPI_ISL_760689 to 760690, EPI_ISL_760692, EPI_ISL_760694 to 760696, EPI_ISL_760700, EPI_ISL_760703 to 760704, EPI_ISL_760706 to 760707, EPI_ISL_760712 to 760713, EPI_ISL_760716 to 760717, EPI_ISL_760720, EPI_ISL_760729 to 760730, EPI_ISL_760732, EPI_ISL_760735, EPI_ISL_760740 to 760744, EPI_ISL_760746, EPI_ISL_760749 to 760751, EPI_ISL_760754, EPI_ISL_760757, EPI_ISL_760760, EPI_ISL_760764 to 760768, EPI_ISL_760770, EPI_ISL_760772 to 760775, EPI_ISL_760777, EPI_ISL_760779, EPI_ISL_760781 to 760782, EPI_ISL_760791, EPI_ISL_760793, EPI_ISL_760797 to 760798, EPI_ISL_760800, EPI_ISL_760803, EPI_ISL_760805, EPI_ISL_760807, EPI_ISL_760811, EPI_ISL_760814, EPI_ISL_760816, EPI_ISL_760818 to 760821, EPI_ISL_760823 to 760824, EPI_ISL_760827 to 760831, EPI_ISL_760834 to 760837, EPI_ISL_760840 to 760841, EPI_ISL_760843, EPI_ISL_760845, EPI_ISL_760847 to 760851, EPI_ISL_760854 to 760856, EPI_ISL_760866, EPI_ISL_760869 to 760870, EPI_ISL_760872, EPI_ISL_760877, EPI_ISL_760879 to 760880, EPI_ISL_760882, EPI_ISL_760885 to 760886, EPI_ISL_760895, EPI_ISL_760899, EPI_ISL_760901 to 760902, EPI_ISL_760907, EPI_ISL_760909 to 760911, EPI_ISL_760913 to 760914, EPI_ISL_760917, EPI_ISL_760919, EPI_ISL_760923, EPI_ISL_760926 to 760927, EPI_ISL_760930, EPI_ISL_760934 to 760935, EPI_ISL_760944, EPI_ISL_760946 to 760947, EPI_ISL_760955 to 760958, EPI_ISL_760964 to 760966, EPI_ISL_760970, EPI_ISL_760972 to 760974, EPI_ISL_760980 to 760981, EPI_ISL_760984, EPI_ISL_760986, EPI_ISL_760990 to 760993, EPI_ISL_760995 to 760999, EPI_ISL_761003, EPI_ISL_761005 to 761007, EPI_ISL_761010, EPI_ISL_761012, EPI_ISL_761014, EPI_ISL_761016, EPI_ISL_761018 to 761019, EPI_ISL_761024, EPI_ISL_761026, EPI_ISL_761028, EPI_ISL_761030 to 761031, EPI_ISL_761033 to 761035, EPI_ISL_761037 to 761038, EPI_ISL_761041, EPI_ISL_761043 to 761045, EPI_ISL_761047, EPI_ISL_761051, EPI_ISL_761053, EPI_ISL_761055, EPI_ISL_761058, EPI_ISL_761062 to 761063, EPI_ISL_761067, EPI_ISL_761070 to 761073, EPI_ISL_761076 to 761078, EPI_ISL_761094 to 761095, EPI_ISL_761114, EPI_ISL_761116, EPI_ISL_761118, EPI_ISL_761120, EPI_ISL_761125, EPI_ISL_761131, EPI_ISL_761133, EPI_ISL_761143, EPI_ISL_761145, EPI_ISL_761154, EPI_ISL_761157 to 761158, EPI_ISL_761162, EPI_ISL_761168, EPI_ISL_761172, EPI_ISL_761176, EPI_ISL_761187, EPI_ISL_761191, EPI_ISL_761193 to 761194, EPI_ISL_761204, EPI_ISL_761207, EPI_ISL_761209, EPI_ISL_761221, EPI_ISL_761225, EPI_ISL_761227 to 761229, EPI_ISL_761241, EPI_ISL_761252, EPI_ISL_761255, EPI_ISL_761259, EPI_ISL_761266, EPI_ISL_761276, EPI_ISL_761278, EPI_ISL_761280, EPI_ISL_761283, EPI_ISL_761287, EPI_ISL_761292 to 761293, EPI_ISL_761296, EPI_ISL_761298, EPI_ISL_761302 to 761303, EPI_ISL_761307, EPI_ISL_761326, EPI_ISL_761331, EPI_ISL_761338, EPI_ISL_761348, EPI_ISL_761351, EPI_ISL_761353, EPI_ISL_761355, EPI_ISL_761921, EPI_ISL_761923 to 761926, EPI_ISL_761928, EPI_ISL_761930 to 761933, EPI_ISL_761936, EPI_ISL_761938, EPI_ISL_761940 to 761945, EPI_ISL_761947 to 761951, EPI_ISL_761954, EPI_ISL_761957 to 761960, EPI_ISL_761962 to 761966, EPI_ISL_761969, EPI_ISL_761971 to 761978, EPI_ISL_761980, EPI_ISL_761982 to 761984, EPI_ISL_761986 to 761993, EPI_ISL_761996, EPI_ISL_761998, EPI_ISL_762000 to 762006, EPI_ISL_762010 to 762013, EPI_ISL_762017, EPI_ISL_762020 to 762022, EPI_ISL_762031 to 762035, EPI_ISL_762039 to 762040, EPI_ISL_762044, EPI_ISL_762046 to 762049, EPI_ISL_762051 to 762056, EPI_ISL_762058, EPI_ISL_762062, EPI_ISL_762068, EPI_ISL_762070 to 762071, EPI_ISL_762076 to 762077, EPI_ISL_762079 to 762084, EPI_ISL_762086, EPI_ISL_762088, EPI_ISL_762092 to 762095, EPI_ISL_762099 to 762102, EPI_ISL_762104 to 762108, EPI_ISL_762110 to 762111, EPI_ISL_762115 to 762116, EPI_ISL_762119, EPI_ISL_762121, EPI_ISL_762123, EPI_ISL_762127 to 762130, EPI_ISL_762132, EPI_ISL_762134, EPI_ISL_762141, EPI_ISL_762143 to 762145, EPI_ISL_762149 to 762151, EPI_ISL_762154, EPI_ISL_762156 to 762158, EPI_ISL_762161 to 762162, EPI_ISL_762164 to 762165, EPI_ISL_762167 to 762172, EPI_ISL_762176 to 762177, EPI_ISL_762179, EPI_ISL_762182, EPI_ISL_762184 to 762185, EPI_ISL_762188, EPI_ISL_762190 to 762191, EPI_ISL_762194, EPI_ISL_762196 to 762197, EPI_ISL_762548, EPI_ISL_762555 to 762556, EPI_ISL_762560 to 762561, EPI_ISL_762567, EPI_ISL_762574, EPI_ISL_762576, EPI_ISL_762583, EPI_ISL_762586, EPI_ISL_762589, EPI_ISL_762597, EPI_ISL_762602, EPI_ISL_762604, EPI_ISL_762609, EPI_ISL_762614, EPI_ISL_762618, EPI_ISL_762622, EPI_ISL_762630 to 762631, EPI_ISL_762633, EPI_ISL_762639, EPI_ISL_762644, EPI_ISL_762646, EPI_ISL_762648, EPI_ISL_762652 to 762654, EPI_ISL_762656 to 762657, EPI_ISL_762661 to 762663, EPI_ISL_762669, EPI_ISL_762676, EPI_ISL_762678, EPI_ISL_762690, EPI_ISL_762694, EPI_ISL_762701, EPI_ISL_762704, EPI_ISL_762708, EPI_ISL_762726, EPI_ISL_762728, EPI_ISL_762731 to 762732, EPI_ISL_762734, EPI_ISL_762739, EPI_ISL_762743 to 762744, EPI_ISL_762746, EPI_ISL_762749, EPI_ISL_762758 to 762761, EPI_ISL_762766, EPI_ISL_762770, EPI_ISL_762773, EPI_ISL_762775, EPI_ISL_762779, EPI_ISL_762785, EPI_ISL_762794, EPI_ISL_762801, EPI_ISL_762809, EPI_ISL_762813 to 762814, EPI_ISL_762816, EPI_ISL_762824, EPI_ISL_762826, EPI_ISL_762837, EPI_ISL_762846, EPI_ISL_762850 to 762851, EPI_ISL_762853 to 762854, EPI_ISL_762856, EPI_ISL_762859 to 762861, EPI_ISL_762865 to 762866, EPI_ISL_762870 to 762872, EPI_ISL_762874, EPI_ISL_762876 to 762877, EPI_ISL_762881, EPI_ISL_762885, EPI_ISL_762888, EPI_ISL_762893, EPI_ISL_762897 to 762898, EPI_ISL_762905, EPI_ISL_762907, EPI_ISL_762909, EPI_ISL_762918, EPI_ISL_762923 to 762925, EPI_ISL_762927 to 762930, EPI_ISL_762934, EPI_ISL_762936, EPI_ISL_762940 to 762941, EPI_ISL_762950, EPI_ISL_762954, EPI_ISL_762956, EPI_ISL_762959 to 762961, EPI_ISL_762966, EPI_ISL_762980, EPI_ISL_762982 to 762987, EPI_ISL_767092, EPI_ISL_767096, EPI_ISL_767113 to 767114, EPI_ISL_767117 to 767118, EPI_ISL_769223 to 769225, EPI_ISL_769229, EPI_ISL_769233, EPI_ISL_769236, EPI_ISL_769243 to 769244, EPI_ISL_769246, EPI_ISL_769249 to 769250, EPI_ISL_769252, EPI_ISL_769254 to 769255, EPI_ISL_769257, EPI_ISL_769259 to 769261, EPI_ISL_769267, EPI_ISL_769275 to 769276, EPI_ISL_769280 to 769282, EPI_ISL_769284, EPI_ISL_769286, EPI_ISL_769288 to 769291, EPI_ISL_769294, EPI_ISL_769297, EPI_ISL_769306, EPI_ISL_769308, EPI_ISL_769310, EPI_ISL_769312, EPI_ISL_769314, EPI_ISL_769316, EPI_ISL_769319 to 769323, EPI_ISL_769325, EPI_ISL_769327, EPI_ISL_769330, EPI_ISL_769334 to 769335, EPI_ISL_769341, EPI_ISL_769343, EPI_ISL_769345, EPI_ISL_769348 to 769349, EPI_ISL_769352 to 769353, EPI_ISL_769355, EPI_ISL_769360 to 769361, EPI_ISL_769365, EPI_ISL_769367, EPI_ISL_769370, EPI_ISL_769374, EPI_ISL_769380 to 769381, EPI_ISL_769384 to 769385, EPI_ISL_769387 to 769392, EPI_ISL_769394, EPI_ISL_769399, EPI_ISL_769401 to 769402, EPI_ISL_769404 to 769406, EPI_ISL_769412 to 769413, EPI_ISL_769416, EPI_ISL_769418 to 769422, EPI_ISL_769427 to 769428, EPI_ISL_769430 to 769431, EPI_ISL_769434, EPI_ISL_769440 to 769441, EPI_ISL_769449 to 769450, EPI_ISL_769452, EPI_ISL_769454, EPI_ISL_769457 to 769460, EPI_ISL_769462, EPI_ISL_769466, EPI_ISL_769468, EPI_ISL_769470 to 769471, EPI_ISL_769473 to 769476, EPI_ISL_769481 to 769482, EPI_ISL_769485, EPI_ISL_769489, EPI_ISL_769497, EPI_ISL_769499 to 769504, EPI_ISL_769506, EPI_ISL_769511 to 769513, EPI_ISL_769517 to 769519, EPI_ISL_769522, EPI_ISL_769524 to 769525, EPI_ISL_769528, EPI_ISL_769530, EPI_ISL_769533, EPI_ISL_769535, EPI_ISL_769540, EPI_ISL_769542 to 769543, EPI_ISL_769551, EPI_ISL_769556, EPI_ISL_769567 to 769568, EPI_ISL_769577, EPI_ISL_769583, EPI_ISL_769589, EPI_ISL_769594, EPI_ISL_769606 to 769607, EPI_ISL_769609, EPI_ISL_769611 to 769614, EPI_ISL_769617, EPI_ISL_769619, EPI_ISL_769625, EPI_ISL_769641, EPI_ISL_769644, EPI_ISL_769648, EPI_ISL_769652, EPI_ISL_769655, EPI_ISL_769661 to 769662, EPI_ISL_769666 to 769667, EPI_ISL_769677, EPI_ISL_769687, EPI_ISL_769689, EPI_ISL_769691, EPI_ISL_769703, EPI_ISL_769705, EPI_ISL_769711, EPI_ISL_769713 to 769714, EPI_ISL_769716, EPI_ISL_769720, EPI_ISL_769724, EPI_ISL_769735, EPI_ISL_769741, EPI_ISL_769743, EPI_ISL_769751 to 769753, EPI_ISL_769758 to 769759, EPI_ISL_769761, EPI_ISL_769768, EPI_ISL_769773, EPI_ISL_769787 to 769788, EPI_ISL_769794, EPI_ISL_769802, EPI_ISL_769806, EPI_ISL_769813, EPI_ISL_769815, EPI_ISL_769818, EPI_ISL_769826, EPI_ISL_769834, EPI_ISL_769836 to 769837, EPI_ISL_769842, EPI_ISL_769850, EPI_ISL_769861 to 769862, EPI_ISL_777013, EPI_ISL_777016, EPI_ISL_777019, EPI_ISL_777021 to 777023, EPI_ISL_777029, EPI_ISL_777031, EPI_ISL_777034, EPI_ISL_777040, EPI_ISL_777047, EPI_ISL_777055, EPI_ISL_777058 to 777059, EPI_ISL_777061 to 777062, EPI_ISL_777065 to 777066, EPI_ISL_777068, EPI_ISL_777073, EPI_ISL_777081, EPI_ISL_777089, EPI_ISL_777095, EPI_ISL_777098, EPI_ISL_777101, EPI_ISL_777105, EPI_ISL_777112, EPI_ISL_777115, EPI_ISL_777117 to 777118, EPI_ISL_777120 to 777122, EPI_ISL_777127, EPI_ISL_777130 to 777131, EPI_ISL_777140, EPI_ISL_777142, EPI_ISL_777146, EPI_ISL_777148 to 777149, EPI_ISL_777151 to 777152, EPI_ISL_777154, EPI_ISL_777160 to 777162, EPI_ISL_777164, EPI_ISL_777176, EPI_ISL_777189, EPI_ISL_777191 to 777192, EPI_ISL_777196, EPI_ISL_777205, EPI_ISL_777208 to 777209, EPI_ISL_777215, EPI_ISL_777219, EPI_ISL_777222 to 777223, EPI_ISL_777225, EPI_ISL_777227, EPI_ISL_777230, EPI_ISL_777232 to 777233, EPI_ISL_777235 to 777236, EPI_ISL_777238, EPI_ISL_777241 to 777242, EPI_ISL_777246, EPI_ISL_777250, EPI_ISL_777252 to 777253, EPI_ISL_777255, EPI_ISL_777261, EPI_ISL_777268, EPI_ISL_777283 to 777284, EPI_ISL_777290, EPI_ISL_777298, EPI_ISL_777300 to 777302, EPI_ISL_777305 to 777306, EPI_ISL_777316, EPI_ISL_777321 to 777322, EPI_ISL_777324, EPI_ISL_777335, EPI_ISL_777337, EPI_ISL_777340 to 777341, EPI_ISL_777346, EPI_ISL_777353, EPI_ISL_777356, EPI_ISL_777358, EPI_ISL_777361, EPI_ISL_777363 to 777364, EPI_ISL_777372, EPI_ISL_777376 to 777377, EPI_ISL_777382, EPI_ISL_777385, EPI_ISL_777392 to 777393, EPI_ISL_777395, EPI_ISL_777397, EPI_ISL_777399, EPI_ISL_777402, EPI_ISL_777407, EPI_ISL_777410, EPI_ISL_777417, EPI_ISL_777420, EPI_ISL_777423, EPI_ISL_777432, EPI_ISL_777436, EPI_ISL_777438, EPI_ISL_777443 to 777444, EPI_ISL_777447, EPI_ISL_777449, EPI_ISL_777459, EPI_ISL_777461 to 777462, EPI_ISL_777466, EPI_ISL_777471, EPI_ISL_777474, EPI_ISL_777484, EPI_ISL_777487, EPI_ISL_777493, EPI_ISL_777496 to 777498, EPI_ISL_777504 to 777506, EPI_ISL_777510, EPI_ISL_777514, EPI_ISL_777523, EPI_ISL_777526, EPI_ISL_777531, EPI_ISL_777533, EPI_ISL_777554, EPI_ISL_777557, EPI_ISL_777560, EPI_ISL_777562, EPI_ISL_777569, EPI_ISL_777574, EPI_ISL_777579, EPI_ISL_777583, EPI_ISL_777600 to 777601, EPI_ISL_777610, EPI_ISL_777612, EPI_ISL_777622, EPI_ISL_777628, EPI_ISL_777630, EPI_ISL_777641, EPI_ISL_777646, EPI_ISL_777653, EPI_ISL_777657 to 777658, EPI_ISL_777660, EPI_ISL_777662 to 777663, EPI_ISL_777667, EPI_ISL_777670, EPI_ISL_778003 to 778004, EPI_ISL_778007 to 778009, EPI_ISL_778014, EPI_ISL_778024, EPI_ISL_778026, EPI_ISL_778034, EPI_ISL_778042, EPI_ISL_778053, EPI_ISL_778058, EPI_ISL_778060, EPI_ISL_778068, EPI_ISL_778073, EPI_ISL_778076, EPI_ISL_778082, EPI_ISL_778088, EPI_ISL_778090, EPI_ISL_778092 to 778093, EPI_ISL_778098, EPI_ISL_778102, EPI_ISL_778106 to 778107, EPI_ISL_778111, EPI_ISL_778117, EPI_ISL_778119, EPI_ISL_778122, EPI_ISL_778127, EPI_ISL_778129, EPI_ISL_778131, EPI_ISL_778133, EPI_ISL_778136, EPI_ISL_778149, EPI_ISL_778153, EPI_ISL_778155 to 778156, EPI_ISL_778167 to 778168, EPI_ISL_778176, EPI_ISL_778192, EPI_ISL_778202 to 778203, EPI_ISL_778215 to 778216, EPI_ISL_778229, EPI_ISL_778236, EPI_ISL_778244, EPI_ISL_778247 to 778248, EPI_ISL_778250, EPI_ISL_778258, EPI_ISL_778260, EPI_ISL_778263, EPI_ISL_778269, EPI_ISL_778276, EPI_ISL_778279, EPI_ISL_778281, EPI_ISL_778289, EPI_ISL_778297, EPI_ISL_778299, EPI_ISL_778301 to 778303, EPI_ISL_778322, EPI_ISL_778325, EPI_ISL_778330, EPI_ISL_778332, EPI_ISL_778334, EPI_ISL_778336 to 778337, EPI_ISL_778345, EPI_ISL_778352, EPI_ISL_778356, EPI_ISL_778359, EPI_ISL_778361 to 778362, EPI_ISL_778364, EPI_ISL_778372 to 778373, EPI_ISL_778375 to 778378, EPI_ISL_778397, EPI_ISL_778400, EPI_ISL_778407, EPI_ISL_778411, EPI_ISL_778419 to 778424, EPI_ISL_778428, EPI_ISL_778433, EPI_ISL_778447, EPI_ISL_778450 to 778451, EPI_ISL_778457, EPI_ISL_778459, EPI_ISL_778461 to 778463, EPI_ISL_778465, EPI_ISL_778482, EPI_ISL_778496 to 778497, EPI_ISL_778499, EPI_ISL_778504, EPI_ISL_778506, EPI_ISL_778522, EPI_ISL_778525, EPI_ISL_778527, EPI_ISL_778530, EPI_ISL_778534, EPI_ISL_778540 to 778541, EPI_ISL_778548, EPI_ISL_778551, EPI_ISL_778553, EPI_ISL_778560, EPI_ISL_778568, EPI_ISL_778570, EPI_ISL_778577, EPI_ISL_778579, EPI_ISL_778586, EPI_ISL_778594, EPI_ISL_778617, EPI_ISL_778619 to 778620, EPI_ISL_778629, EPI_ISL_778631, EPI_ISL_781870 to 781871, EPI_ISL_781875, EPI_ISL_781882, EPI_ISL_781886, EPI_ISL_781890, EPI_ISL_781895, EPI_ISL_781910, EPI_ISL_781913, EPI_ISL_781918 to 781919, EPI_ISL_781922, EPI_ISL_781928, EPI_ISL_781930, EPI_ISL_781933, EPI_ISL_781953, EPI_ISL_781956, EPI_ISL_781960, EPI_ISL_781967, EPI_ISL_781971, EPI_ISL_781978, EPI_ISL_781980, EPI_ISL_781982, EPI_ISL_781990, EPI_ISL_781992, EPI_ISL_781997, EPI_ISL_782001, EPI_ISL_782003 to 782004, EPI_ISL_782008, EPI_ISL_782014, EPI_ISL_782018, EPI_ISL_782027, EPI_ISL_782036, EPI_ISL_782038 to 782039, EPI_ISL_782056, EPI_ISL_782059, EPI_ISL_782062, EPI_ISL_782064 to 782067, EPI_ISL_782074 to 782075, EPI_ISL_782079, EPI_ISL_782084, EPI_ISL_782101, EPI_ISL_782107, EPI_ISL_782112, EPI_ISL_782114 to 782116, EPI_ISL_782121, EPI_ISL_782127 to 782128, EPI_ISL_782133, EPI_ISL_782138, EPI_ISL_782145 to 782146, EPI_ISL_782151 to 782160, EPI_ISL_782162, EPI_ISL_782166 to 782170, EPI_ISL_782172 to 782183, EPI_ISL_782185 to 782190, EPI_ISL_782192 to 782194, EPI_ISL_782196, EPI_ISL_782198, EPI_ISL_782200, EPI_ISL_782203 to 782206, EPI_ISL_782208 to 782210, EPI_ISL_782213 to 782216, EPI_ISL_782218, EPI_ISL_782221 to 782223, EPI_ISL_782225, EPI_ISL_782227, EPI_ISL_782229, EPI_ISL_782231, EPI_ISL_782233 to 782234, EPI_ISL_782236 to 782241, EPI_ISL_782244, EPI_ISL_782246 to 782248, EPI_ISL_782251, EPI_ISL_782253 to 782255, EPI_ISL_782257, EPI_ISL_782260 to 782263, EPI_ISL_782265, EPI_ISL_782269, EPI_ISL_782272, EPI_ISL_782275 to 782277, EPI_ISL_782279 to 782281, EPI_ISL_782283 to 782286, EPI_ISL_782288 to 782290, EPI_ISL_782293 to 782302, EPI_ISL_782305 to 782306, EPI_ISL_782308, EPI_ISL_782310 to 782312, EPI_ISL_782316, EPI_ISL_782318 to 782324, EPI_ISL_782327, EPI_ISL_782329 to 782334, EPI_ISL_782336 to 782338, EPI_ISL_782340, EPI_ISL_782342 to 782344, EPI_ISL_782346, EPI_ISL_782348 to 782349, EPI_ISL_782353 to 782354, EPI_ISL_782356 to 782357, EPI_ISL_782359 to 782360, EPI_ISL_782363 to 782368, EPI_ISL_782370, EPI_ISL_782373 to 782376, EPI_ISL_782378 to 782379, EPI_ISL_782383, EPI_ISL_782385 to 782387, EPI_ISL_782391 to 782392, EPI_ISL_782395, EPI_ISL_782397 to 782400, EPI_ISL_782403 to 782408, EPI_ISL_782410 to 782411, EPI_ISL_782413 to 782418, EPI_ISL_782421 to 782422, EPI_ISL_782424, EPI_ISL_782427 to 782434, EPI_ISL_782436 to 782438, EPI_ISL_782440 to 782447, EPI_ISL_782449 to 782450, EPI_ISL_782452 to 782453, EPI_ISL_782455, EPI_ISL_782457 to 782460, EPI_ISL_782463, EPI_ISL_782465, EPI_ISL_782468 to 782470, EPI_ISL_782472 to 782473, EPI_ISL_782475 to 782476, EPI_ISL_782479, EPI_ISL_782481 to 782486, EPI_ISL_782488 to 782506, EPI_ISL_782508 to 782511, EPI_ISL_782514 to 782524, EPI_ISL_782526 to 782551, EPI_ISL_782553 to 782560, EPI_ISL_782562 to 782597, EPI_ISL_782599 to 782600, EPI_ISL_782602, EPI_ISL_782604, EPI_ISL_782606 to 782624, EPI_ISL_782626 to 782636, EPI_ISL_782638 to 782646, EPI_ISL_782648 to 782656, EPI_ISL_782658 to 782660, EPI_ISL_782662 to 782680, EPI_ISL_782682 to 782683, EPI_ISL_782686 to 782688, EPI_ISL_782690 to 782695, EPI_ISL_782697 to 782699, EPI_ISL_782701 to 782717, EPI_ISL_782719 to 782725, EPI_ISL_782727, EPI_ISL_782729 to 782733, EPI_ISL_797719 to 797744, EPI_ISL_797746 to 797771, EPI_ISL_797773 to 797790, EPI_ISL_797792 to 797815, EPI_ISL_797817 to 797839, EPI_ISL_797841 to 797846, EPI_ISL_797848 to 797849, EPI_ISL_797851 to 797856, EPI_ISL_797858 to 797866, EPI_ISL_797868 to 797869, EPI_ISL_797871 to 797876, EPI_ISL_797878 to 797913, EPI_ISL_797915 to 797928, EPI_ISL_797930 to 797944, EPI_ISL_797946 to 797960, EPI_ISL_797962 to 797969, EPI_ISL_797971 to 797979, EPI_ISL_797981 to 797995, EPI_ISL_797997, EPI_ISL_798001, EPI_ISL_798004 to 798006, EPI_ISL_798008, EPI_ISL_798010, EPI_ISL_798012, EPI_ISL_798014, EPI_ISL_798016 to 798017, EPI_ISL_798019 to 798024, EPI_ISL_798026, EPI_ISL_798028, EPI_ISL_798030 to 798031, EPI_ISL_798033 to 798034, EPI_ISL_798036 to 798037, EPI_ISL_798039, EPI_ISL_798041 to 798045, EPI_ISL_798047, EPI_ISL_798049 to 798052, EPI_ISL_798056 to 798057, EPI_ISL_798059, EPI_ISL_798063, EPI_ISL_798065, EPI_ISL_798067, EPI_ISL_798070 to 798071, EPI_ISL_798074 to 798075, EPI_ISL_798080 to 798082, EPI_ISL_798085, EPI_ISL_798087, EPI_ISL_798089 to 798092, EPI_ISL_798098 to 798100, EPI_ISL_798102, EPI_ISL_798104, EPI_ISL_798107, EPI_ISL_798109, EPI_ISL_798114, EPI_ISL_798116, EPI_ISL_798118, EPI_ISL_798120, EPI_ISL_798129, EPI_ISL_798135 to 798136, EPI_ISL_798138, EPI_ISL_798141 to 798142, EPI_ISL_798144, EPI_ISL_798146 to 798147, EPI_ISL_798150, EPI_ISL_798155 to 798159, EPI_ISL_798162 to 798165, EPI_ISL_798171 to 798172, EPI_ISL_798177 to 798180, EPI_ISL_798184, EPI_ISL_798187 to 798192, EPI_ISL_798194 to 798196, EPI_ISL_798198, EPI_ISL_798200, EPI_ISL_798205 to 798207, EPI_ISL_798209 to 798210, EPI_ISL_798212, EPI_ISL_798215, EPI_ISL_798219, EPI_ISL_798223, EPI_ISL_798226 to 798228, EPI_ISL_798231 to 798233, EPI_ISL_798237 to 798238, EPI_ISL_798240, EPI_ISL_798247, EPI_ISL_798249, EPI_ISL_798257, EPI_ISL_798259 to 798265, EPI_ISL_798267, EPI_ISL_798269, EPI_ISL_798271, EPI_ISL_798276, EPI_ISL_798278 to 798279, EPI_ISL_798286 to 798287, EPI_ISL_798290 to 798291, EPI_ISL_798293, EPI_ISL_798297, EPI_ISL_798301 to 798304, EPI_ISL_798306, EPI_ISL_798308 to 798309, EPI_ISL_798313, EPI_ISL_798317 to 798318, EPI_ISL_798320, EPI_ISL_798325, EPI_ISL_798330 to 798332, EPI_ISL_799006, EPI_ISL_799011 to 799012, EPI_ISL_799014 to 799016, EPI_ISL_799019, EPI_ISL_799024, EPI_ISL_799027, EPI_ISL_799030, EPI_ISL_799033 to 799034, EPI_ISL_799038, EPI_ISL_799041 to 799044, EPI_ISL_799046, EPI_ISL_799049, EPI_ISL_799051, EPI_ISL_799054, EPI_ISL_799056, EPI_ISL_799060, EPI_ISL_799063 to 799068, EPI_ISL_799071 to 799073, EPI_ISL_799075, EPI_ISL_799077, EPI_ISL_799080 to 799082, EPI_ISL_799084, EPI_ISL_799089 to 799090, EPI_ISL_799096, EPI_ISL_799098, EPI_ISL_799100 to 799102, EPI_ISL_799107 to 799108, EPI_ISL_799112, EPI_ISL_799115, EPI_ISL_799120, EPI_ISL_799123, EPI_ISL_799125 to 799127, EPI_ISL_799129 to 799132, EPI_ISL_799134, EPI_ISL_799140, EPI_ISL_799144, EPI_ISL_799147, EPI_ISL_799150, EPI_ISL_799155 to 799158, EPI_ISL_799162 to 799164, EPI_ISL_799167, EPI_ISL_799169, EPI_ISL_799172, EPI_ISL_799175 to 799176, EPI_ISL_799183, EPI_ISL_799189 to 799190, EPI_ISL_799192 to 799194, EPI_ISL_799196 to 799197, EPI_ISL_799201, EPI_ISL_799203 to 799205, EPI_ISL_799207, EPI_ISL_799209, EPI_ISL_799215 to 799216, EPI_ISL_799218, EPI_ISL_799221 to 799222, EPI_ISL_799224, EPI_ISL_799230, EPI_ISL_799232, EPI_ISL_799240, EPI_ISL_799243, EPI_ISL_799245 to 799246, EPI_ISL_799249 to 799250, EPI_ISL_799255 to 799256, EPI_ISL_799258, EPI_ISL_799260, EPI_ISL_799262 to 799264, EPI_ISL_799267 to 799268, EPI_ISL_799270, EPI_ISL_799272 to 799273, EPI_ISL_799275, EPI_ISL_799277, EPI_ISL_799282, EPI_ISL_799285 to 799290, EPI_ISL_799297 to 799298, EPI_ISL_799300 to 799302, EPI_ISL_799304, EPI_ISL_799307, EPI_ISL_799309 to 799310, EPI_ISL_799312, EPI_ISL_799315 to 799316, EPI_ISL_799321 to 799322, EPI_ISL_799324, EPI_ISL_799326, EPI_ISL_799328 to 799330, EPI_ISL_799332, EPI_ISL_799336 to 799340, EPI_ISL_799342 to 799345, EPI_ISL_799349, EPI_ISL_799355, EPI_ISL_799358 to 799359, EPI_ISL_799361, EPI_ISL_799365, EPI_ISL_799367, EPI_ISL_799369, EPI_ISL_799371, EPI_ISL_799374, EPI_ISL_799377, EPI_ISL_799379, EPI_ISL_799381, EPI_ISL_799383, EPI_ISL_799385, EPI_ISL_799389, EPI_ISL_799391, EPI_ISL_799393, EPI_ISL_799396, EPI_ISL_799398 to 799399, EPI_ISL_799401, EPI_ISL_799405, EPI_ISL_799407 to 799409, EPI_ISL_799414, EPI_ISL_799417 to 799419, EPI_ISL_799421 to 799425, EPI_ISL_799428 to 799429, EPI_ISL_799431, EPI_ISL_799433, EPI_ISL_799435, EPI_ISL_799438, EPI_ISL_799442, EPI_ISL_799444, EPI_ISL_799449 to 799453, EPI_ISL_799455 to 799464, EPI_ISL_799471 to 799472, EPI_ISL_799474 to 799475, EPI_ISL_799478, EPI_ISL_799480 to 799481, EPI_ISL_799483 to 799484, EPI_ISL_799486, EPI_ISL_799489, EPI_ISL_799491, EPI_ISL_799495, EPI_ISL_799499, EPI_ISL_799503 to 799504, EPI_ISL_799507, EPI_ISL_799510 to 799511, EPI_ISL_799516 to 799518, EPI_ISL_799524, EPI_ISL_799526 to 799527, EPI_ISL_799530, EPI_ISL_799537, EPI_ISL_799539, EPI_ISL_799541 to 799544, EPI_ISL_799548 to 799549, EPI_ISL_799552 to 799554, EPI_ISL_799556 to 799561, EPI_ISL_799564, EPI_ISL_799566, EPI_ISL_799568 to 799569, EPI_ISL_799571, EPI_ISL_799574,

EPI_ISL_799576, EPI_ISL_799581, EPI_ISL_799584 to 799586, EPI_ISL_799589, EPI_ISL_799591 to 799594, EPI_ISL_799596, EPI_ISL_799599, EPI_ISL_799601, EPI_ISL_799605, EPI_ISL_799609, EPI_ISL_799615, EPI_ISL_799622, EPI_ISL_799634, EPI_ISL_799641, EPI_ISL_799649 to 799650, EPI_ISL_799659, EPI_ISL_799661, EPI_ISL_799664, EPI_ISL_799669, EPI_ISL_799676, EPI_ISL_799679, EPI_ISL_799681 to 799682, EPI_ISL_799686, EPI_ISL_799707 to 799708, EPI_ISL_799713, EPI_ISL_799718, EPI_ISL_799722, EPI_ISL_799726, EPI_ISL_799729 to 799731, EPI_ISL_799735, EPI_ISL_799737, EPI_ISL_799739, EPI_ISL_799748, EPI_ISL_799758, EPI_ISL_799761, EPI_ISL_799763, EPI_ISL_799765, EPI_ISL_799776, EPI_ISL_799780, EPI_ISL_799783, EPI_ISL_799786, EPI_ISL_799791 to 799792, EPI_ISL_799801, EPI_ISL_799805, EPI_ISL_799811, EPI_ISL_799819, EPI_ISL_799825, EPI_ISL_799830, EPI_ISL_799838, EPI_ISL_799843, EPI_ISL_799855, EPI_ISL_799861, EPI_ISL_799864 to 799866, EPI_ISL_799869, EPI_ISL_799871, EPI_ISL_799875, EPI_ISL_799883, EPI_ISL_799888 to 799889, EPI_ISL_799892 to 799894, EPI_ISL_799901, EPI_ISL_799906, EPI_ISL_799910, EPI_ISL_799912, EPI_ISL_799916 to 799919, EPI_ISL_799932, EPI_ISL_799937, EPI_ISL_799945 to 799946, EPI_ISL_799949, EPI_ISL_799951, EPI_ISL_799954, EPI_ISL_799965, EPI_ISL_799968 to 799969, EPI_ISL_799973, EPI_ISL_799976, EPI_ISL_799980 to 799981, EPI_ISL_799985 to 799987, EPI_ISL_799990, EPI_ISL_799992, EPI_ISL_799997, EPI_ISL_800000, EPI_ISL_800005, EPI_ISL_800010, EPI_ISL_800015, EPI_ISL_800027, EPI_ISL_800029 to 800030, EPI_ISL_800032, EPI_ISL_800034, EPI_ISL_800038, EPI_ISL_800048, EPI_ISL_800052, EPI_ISL_800056 to 800057, EPI_ISL_800064, EPI_ISL_800068, EPI_ISL_800070 to 800071, EPI_ISL_800078 to 800079, EPI_ISL_800084, EPI_ISL_800086, EPI_ISL_800106, EPI_ISL_800111, EPI_ISL_800113, EPI_ISL_800117, EPI_ISL_800120, EPI_ISL_800122, EPI_ISL_800124, EPI_ISL_800131, EPI_ISL_800138, EPI_ISL_800141 to 800142, EPI_ISL_800147, EPI_ISL_800151, EPI_ISL_800158, EPI_ISL_800161, EPI_ISL_800164, EPI_ISL_800174 to 800176, EPI_ISL_800180, EPI_ISL_800183, EPI_ISL_800186, EPI_ISL_800188 to 800189, EPI_ISL_800191, EPI_ISL_800200 to 800201, EPI_ISL_800204, EPI_ISL_800210, EPI_ISL_800217, EPI_ISL_800221, EPI_ISL_800234 to 800237, EPI_ISL_800241, EPI_ISL_800245, EPI_ISL_800251, EPI_ISL_800253, EPI_ISL_800576, EPI_ISL_800594, EPI_ISL_800596, EPI_ISL_800601 to 800602, EPI_ISL_800604 to 800605, EPI_ISL_800607 to 800608, EPI_ISL_800611 to 800612, EPI_ISL_800614, EPI_ISL_800616, EPI_ISL_800618, EPI_ISL_800620 to 800621, EPI_ISL_800625, EPI_ISL_800627 to 800631, EPI_ISL_800633 to 800635, EPI_ISL_800638 to 800640, EPI_ISL_800643, EPI_ISL_800646, EPI_ISL_800651, EPI_ISL_800653 to 800654, EPI_ISL_800656, EPI_ISL_800659, EPI_ISL_800662, EPI_ISL_800666 to 800668, EPI_ISL_800670 to 800671, EPI_ISL_800675, EPI_ISL_800688 to 800689, EPI_ISL_800691, EPI_ISL_800693 to 800697, EPI_ISL_800700, EPI_ISL_800703 to 800706, EPI_ISL_800715, EPI_ISL_800722 to 800723, EPI_ISL_800725 to 800728, EPI_ISL_800731, EPI_ISL_800735 to 800736, EPI_ISL_800739, EPI_ISL_800742, EPI_ISL_800745, EPI_ISL_800748 to 800749, EPI_ISL_800753 to 800754, EPI_ISL_800759 to 800760, EPI_ISL_800766 to 800767, EPI_ISL_800772, EPI_ISL_800775, EPI_ISL_800779 to 800781, EPI_ISL_800785 to 800787, EPI_ISL_800794, EPI_ISL_800796 to 800797, EPI_ISL_800799 to 800801, EPI_ISL_800805, EPI_ISL_800812 to 800813, EPI_ISL_800815, EPI_ISL_800821, EPI_ISL_800825, EPI_ISL_800827, EPI_ISL_800829, EPI_ISL_800833, EPI_ISL_800835, EPI_ISL_800838, EPI_ISL_800841, EPI_ISL_800843, EPI_ISL_800847 to 800848, EPI_ISL_800850 to 800851, EPI_ISL_800856, EPI_ISL_800858, EPI_ISL_800860 to 800862, EPI_ISL_800869, EPI_ISL_800874 to 800875, EPI_ISL_800882, EPI_ISL_800884, EPI_ISL_801179, EPI_ISL_801231 to 801238, EPI_ISL_801240 to 801244, EPI_ISL_801246 to 801249, EPI_ISL_801251 to 801282, EPI_ISL_801284 to 801293, EPI_ISL_801295 to 801305, EPI_ISL_801307 to 801312, EPI_ISL_801314 to 801323, EPI_ISL_801325 to 801327, EPI_ISL_801329 to 801340, EPI_ISL_801343 to 801349, EPI_ISL_801354 to 801363, EPI_ISL_801365, EPI_ISL_801367 to 801384, EPI_ISL_820147, EPI_ISL_820150, EPI_ISL_820152, EPI_ISL_820171, EPI_ISL_820176 to 820177, EPI_ISL_820182, EPI_ISL_820184, EPI_ISL_820190, EPI_ISL_820197, EPI_ISL_820212, EPI_ISL_820215, EPI_ISL_820220, EPI_ISL_820227, EPI_ISL_820233, EPI_ISL_820260, EPI_ISL_820262, EPI_ISL_820270, EPI_ISL_820272, EPI_ISL_820283, EPI_ISL_820295, EPI_ISL_820312, EPI_ISL_820319 to 820320, EPI_ISL_820349, EPI_ISL_820356, EPI_ISL_820384, EPI_ISL_820386 to 820387, EPI_ISL_820397, EPI_ISL_820409, EPI_ISL_820417, EPI_ISL_820421, EPI_ISL_820437, EPI_ISL_820448, EPI_ISL_820461, EPI_ISL_820465, EPI_ISL_820468, EPI_ISL_820477, EPI_ISL_820479, EPI_ISL_820481, EPI_ISL_820489, EPI_ISL_820505, EPI_ISL_820507, EPI_ISL_820512 to 820513, EPI_ISL_820518, EPI_ISL_820547, EPI_ISL_820558, EPI_ISL_820577, EPI_ISL_820590, EPI_ISL_820597, EPI_ISL_820613, EPI_ISL_820617 to 820618, EPI_ISL_820620, EPI_ISL_820628, EPI_ISL_820633 to 820634, EPI_ISL_820639, EPI_ISL_820641, EPI_ISL_820645, EPI_ISL_820647, EPI_ISL_821659, EPI_ISL_822292 to 822293, EPI_ISL_822295 to 822296, EPI_ISL_822302, EPI_ISL_822304, EPI_ISL_822307, EPI_ISL_822318 to 822321, EPI_ISL_822323 to 822328, EPI_ISL_822330 to 822341, EPI_ISL_822343, EPI_ISL_826516, EPI_ISL_834578, EPI_ISL_858313, EPI_ISL_858410, EPI_ISL_858412, EPI_ISL_858415, EPI_ISL_858417, EPI_ISL_858419 to 858420, EPI_ISL_858423 to 858424, EPI_ISL_858589 to 858591, EPI_ISL_858593, EPI_ISL_858595 to 858597, EPI_ISL_858599, EPI_ISL_858602 to 858603, EPI_ISL_858605 to 858606, EPI_ISL_858611, EPI_ISL_858619 to 858620, EPI_ISL_858622, EPI_ISL_858625, EPI_ISL_858629, EPI_ISL_858631 to 858632, EPI_ISL_858639, EPI_ISL_858711 to 858712, EPI_ISL_858715 to 858718, EPI_ISL_858721 to 858722, EPI_ISL_858726 to 858731, EPI_ISL_858734 to 858736, EPI_ISL_858738, EPI_ISL_858740 to 858747, EPI_ISL_858749 to 858751, EPI_ISL_858754 to 858756, EPI_ISL_858758 to 858778, EPI_ISL_858781 to 858783, EPI_ISL_858793 to 858794, EPI_ISL_858796 to 858797, EPI_ISL_858801, EPI_ISL_858806, EPI_ISL_858810, EPI_ISL_858815, EPI_ISL_858830, EPI_ISL_858842 to 858844, EPI_ISL_858859 to 858860, EPI_ISL_858863, EPI_ISL_858865, EPI_ISL_858867, EPI_ISL_858874, EPI_ISL_858900 to 858901, EPI_ISL_858903 to 858904, EPI_ISL_858906, EPI_ISL_858908, EPI_ISL_858910, EPI_ISL_858912, EPI_ISL_858916, EPI_ISL_858919 to 858922, EPI_ISL_858927 to 858928, EPI_ISL_858930, EPI_ISL_858935 to 858937, EPI_ISL_858943, EPI_ISL_858945, EPI_ISL_858953, EPI_ISL_858958, EPI_ISL_858960, EPI_ISL_858969 to 858970, EPI_ISL_858974 to 858975, EPI_ISL_859003, EPI_ISL_859012, EPI_ISL_859030, EPI_ISL_859032 to 859034, EPI_ISL_859036 to 859038, EPI_ISL_859041 to 859042, EPI_ISL_859045 to 859048, EPI_ISL_859050, EPI_ISL_859052 to 859055, EPI_ISL_859057, EPI_ISL_859059, EPI_ISL_859062, EPI_ISL_859064, EPI_ISL_859068, EPI_ISL_859074, EPI_ISL_859077 to 859079, EPI_ISL_859081, EPI_ISL_859083 to 859084, EPI_ISL_859087, EPI_ISL_859093 to 859094, EPI_ISL_859098, EPI_ISL_859100, EPI_ISL_859106, EPI_ISL_859108, EPI_ISL_859112, EPI_ISL_859114, EPI_ISL_859116, EPI_ISL_859118 to 859119, EPI_ISL_859123, EPI_ISL_859125, EPI_ISL_859127, EPI_ISL_859130 to 859131, EPI_ISL_859133, EPI_ISL_859137, EPI_ISL_859139, EPI_ISL_859149 to 859153, EPI_ISL_859155 to 859160, EPI_ISL_859162 to 859164, EPI_ISL_859166 to 859168, EPI_ISL_859170 to 859178, EPI_ISL_859180, EPI_ISL_859182 to 859185, EPI_ISL_859187 to 859193, EPI_ISL_859195 to 859197, EPI_ISL_859199 to 859200, EPI_ISL_859205, EPI_ISL_859211, EPI_ISL_859214, EPI_ISL_859218 to 859219, EPI_ISL_859232, EPI_ISL_859234, EPI_ISL_859236, EPI_ISL_859253, EPI_ISL_859256, EPI_ISL_859265, EPI_ISL_859268, EPI_ISL_859270, EPI_ISL_859282, EPI_ISL_859288, EPI_ISL_859294 to 859295, EPI_ISL_859297, EPI_ISL_859301, EPI_ISL_859303 to 859304, EPI_ISL_859306, EPI_ISL_859312 to 859313, EPI_ISL_859316 to 859326, EPI_ISL_859422, EPI_ISL_859435, EPI_ISL_859456 to 859459, EPI_ISL_859461 to 859463, EPI_ISL_859465 to 859468, EPI_ISL_859470 to 859474, EPI_ISL_859476 to 859496, EPI_ISL_859498 to 859506, EPI_ISL_859508 to 859511, EPI_ISL_859513 to 859518, EPI_ISL_859520 to 859525, EPI_ISL_859527 to 859532, EPI_ISL_859541 to 859552, EPI_ISL_863532, EPI_ISL_863590, EPI_ISL_863600, EPI_ISL_863631, EPI_ISL_863635, EPI_ISL_863639, EPI_ISL_863645, EPI_ISL_863651, EPI_ISL_863707, EPI_ISL_863721, EPI_ISL_863729, EPI_ISL_863746, EPI_ISL_863803, EPI_ISL_863830, EPI_ISL_863841

| | | | |
|---|---|---|---|
| see above | Lighthouse Lab in Glasgow | Wellcome Sanger Institute for the COVID-19 Genomics UK (COG-UK) Consortium | Anna Dominiczak and Alex Alderton; Carol Clugston; Cordelia Langford; David Gray; David K. Jackson; Dominic Kwiatkowski; Ewan Harrison; Harper VanSteenhouse; Ian Johnston; John Sillitoe on behalf of the Wellcome Sanger Institute COVID-19 Surveillance Team; John Sillitoe on behalf of the Wellcome Sanger Institute COVID-19 Surveillance Team (http://www.sanger.ac.uk/covid-team); Roberto Amato; Sonia Goncalves; Yumi Kasai |

EPI_ISL_530351 to 530360, EPI_ISL_530362 to 530365, EPI_ISL_530367 to 530369, EPI_ISL_530371 to 530376, EPI_ISL_530378 to 530385, EPI_ISL_530387, EPI_ISL_530389 to 530390, EPI_ISL_530392 to 530397, EPI_ISL_530399 to 530400, EPI_ISL_530402 to 530409, EPI_ISL_530411 to 530419, EPI_ISL_530421 to 530423, EPI_ISL_530425 to 530428, EPI_ISL_530430 to 530434, EPI_ISL_530436 to 530439, EPI_ISL_530441 to 530467, EPI_ISL_530469 to 530478, EPI_ISL_530481, EPI_ISL_530483 to 530487, EPI_ISL_530490 to 530508, EPI_ISL_530510 to 530522, EPI_ISL_530524 to 530527, EPI_ISL_530529 to 530532, EPI_ISL_530534, EPI_ISL_530536 to 530537, EPI_ISL_530539 to 530541, EPI_ISL_530543 to 530552, EPI_ISL_530554 to 530561, EPI_ISL_530563 to 530595, EPI_ISL_530597 to 530599, EPI_ISL_530601 to 530641, EPI_ISL_530643 to 530658, EPI_ISL_530660 to 530666, EPI_ISL_530668 to 530672, EPI_ISL_530674 to 530681, EPI_ISL_530683 to 530687, EPI_ISL_530690 to 530696, EPI_ISL_530698 to 530703, EPI_ISL_530705 to 530707, EPI_ISL_530709 to 530721, EPI_ISL_530723 to 530739, EPI_ISL_530741 to 530742, EPI_ISL_530744 to 530745, EPI_ISL_530747 to 530753, EPI_ISL_530755, EPI_ISL_530757 to 530760, EPI_ISL_530762 to 530763, EPI_ISL_530765, EPI_ISL_530770 to 530798, EPI_ISL_530800 to 530825, EPI_ISL_530827 to 530835, EPI_ISL_530837 to 530844, EPI_ISL_530847 to 530861, EPI_ISL_530863 to 530870, EPI_ISL_530872, EPI_ISL_530874, EPI_ISL_530876 to 530898, EPI_ISL_530900 to 530903, EPI_ISL_530905, EPI_ISL_530907 to 530908, EPI_ISL_530910 to 530917, EPI_ISL_530919 to 530920, EPI_ISL_530922, EPI_ISL_530924 to 530928, EPI_ISL_530930 to 530952, EPI_ISL_530954 to 530958, EPI_ISL_530960, EPI_ISL_530964 to 530972, EPI_ISL_530974 to 530975, EPI_ISL_530977 to 530978, EPI_ISL_530980 to 530982, EPI_ISL_530984, EPI_ISL_530988 to 530989, EPI_ISL_530991 to 530995, EPI_ISL_530998 to 531001, EPI_ISL_531003, EPI_ISL_531005, EPI_ISL_531007 to 531011, EPI_ISL_531013, EPI_ISL_531015 to 531016, EPI_ISL_531018, EPI_ISL_531021 to 531031, EPI_ISL_531034 to 531037, EPI_ISL_531039, EPI_ISL_531041 to 531044, EPI_ISL_531047, EPI_ISL_531050 to 531051, EPI_ISL_531053, EPI_ISL_531055, EPI_ISL_531058 to 531064, EPI_ISL_531066 to 531078, EPI_ISL_531080 to 531082, EPI_ISL_531084 to 531086, EPI_ISL_531088 to 531090, EPI_ISL_531092, EPI_ISL_531094 to 531095, EPI_ISL_531097 to 531098, EPI_ISL_531101 to 531102, EPI_ISL_531105, EPI_ISL_531107 to 531113, EPI_ISL_531115, EPI_ISL_531117, EPI_ISL_531120, EPI_ISL_531122, EPI_ISL_531124 to 531125, EPI_ISL_531127 to 531129, EPI_ISL_531132 to 531141, EPI_ISL_531143 to 531146, EPI_ISL_531148 to 531153, EPI_ISL_531158, EPI_ISL_531160 to 531165, EPI_ISL_531167 to 531173, EPI_ISL_531176 to 531177, EPI_ISL_531179 to 531187, EPI_ISL_531190 to 531191, EPI_ISL_531193 to 531197, EPI_ISL_531199 to 531206, EPI_ISL_531209 to 531210, EPI_ISL_531213, EPI_ISL_531215 to 531222, EPI_ISL_531224 to 531227, EPI_ISL_531229 to 531242, EPI_ISL_531244 to 531248, EPI_ISL_531250 to 531265, EPI_ISL_531267 to 531290, EPI_ISL_531292 to 531293, EPI_ISL_531295 to 531320, EPI_ISL_531322 to 531329, EPI_ISL_531331 to 531334, EPI_ISL_531336 to 531360, EPI_ISL_531362 to 531364, EPI_ISL_531366 to 531370, EPI_ISL_531372 to 531373, EPI_ISL_531375 to 531381, EPI_ISL_531383 to 531394, EPI_ISL_531396 to 531411, EPI_ISL_531413 to 531415, EPI_ISL_531417 to 531419, EPI_ISL_531421, EPI_ISL_531423 to 531431, EPI_ISL_531433 to 531441, EPI_ISL_531443 to 531447, EPI_ISL_531449 to 531463, EPI_ISL_531465 to 531469, EPI_ISL_531471 to 531472, EPI_ISL_531474 to 531475, EPI_ISL_531481 to 531482, EPI_ISL_531486 to 531489, EPI_ISL_531491, EPI_ISL_531493 to 531494, EPI_ISL_531496 to 531499, EPI_ISL_531501 to 531503, EPI_ISL_531505 to 531523, EPI_ISL_531525 to 531545, EPI_ISL_531549 to 531556, EPI_ISL_531558 to 531562, EPI_ISL_531566 to 531568, EPI_ISL_531571 to 531574, EPI_ISL_531576, EPI_ISL_531578 to 531582, EPI_ISL_531584 to 531589, EPI_ISL_531591 to 531594, EPI_ISL_531596 to 531597, EPI_ISL_531600, EPI_ISL_531602 to 531606, EPI_ISL_531608, EPI_ISL_531610 to 531616, EPI_ISL_531619, EPI_ISL_531621 to 531624, EPI_ISL_531626 to 531629, EPI_ISL_531632 to 531633, EPI_ISL_531635 to 531637, EPI_ISL_531640 to 531648, EPI_ISL_531650 to 531654, EPI_ISL_531656 to 531659, EPI_ISL_531661 to 531667, EPI_ISL_531669 to 531671, EPI_ISL_531673, EPI_ISL_531675 to 531676, EPI_ISL_531679 to 531680, EPI_ISL_531682 to 531686, EPI_ISL_531692 to 531701, EPI_ISL_531704 to 531713, EPI_ISL_531715 to 531725, EPI_ISL_531728 to 531729, EPI_ISL_531731, EPI_ISL_531733 to 531742, EPI_ISL_531744 to 531752, EPI_ISL_531756 to 531757, EPI_ISL_531759 to 531767, EPI_ISL_531769 to 531778, EPI_ISL_531780 to 531783, EPI_ISL_531785 to 531787, EPI_ISL_531789 to 531791, EPI_ISL_531793 to 531800, EPI_ISL_531803 to 531804, EPI_ISL_531806 to 531812, EPI_ISL_531816 to 531820, EPI_ISL_531822 to 531823, EPI_ISL_531825 to 531831, EPI_ISL_531833 to 531841, EPI_ISL_531843 to 531850, EPI_ISL_531852 to 531855, EPI_ISL_531859 to 531861, EPI_ISL_531864, EPI_ISL_531866 to 531871, EPI_ISL_531874 to 531876, EPI_ISL_531878 to 531879, EPI_ISL_531882 to 531883, EPI_ISL_531885 to 531886, EPI_ISL_531888, EPI_ISL_531891, EPI_ISL_531893, EPI_ISL_531895 to 531901, EPI_ISL_531903 to 531904, EPI_ISL_531906 to 531911, EPI_ISL_531914, EPI_ISL_531916 to 531917, EPI_ISL_531920 to 531924, EPI_ISL_531926 to 531928, EPI_ISL_531930 to 531931, EPI_ISL_531934, EPI_ISL_531937 to 531942, EPI_ISL_531944, EPI_ISL_531946 to 531950, EPI_ISL_531952 to 531953, EPI_ISL_531955 to 531962, EPI_ISL_531964 to 531972, EPI_ISL_531974 to 531977, EPI_ISL_531980, EPI_ISL_531983, EPI_ISL_531985 to 531986, EPI_ISL_531988 to 531989, EPI_ISL_531991 to 531994, EPI_ISL_531996 to 532002, EPI_ISL_532006, EPI_ISL_532008 to 532011, EPI_ISL_532013 to 532017, EPI_ISL_532019 to 532023, EPI_ISL_532025 to 532031, EPI_ISL_532033 to 532037, EPI_ISL_532039 to 532048, EPI_ISL_532050, EPI_ISL_532052, EPI_ISL_532054, EPI_ISL_532056, EPI_ISL_532058 to 532061, EPI_ISL_532063 to 532069, EPI_ISL_532072 to 532075, EPI_ISL_532079, EPI_ISL_532082 to 532084, EPI_ISL_532086 to 532092, EPI_ISL_532095 to 532097, EPI_ISL_532099 to 532101, EPI_ISL_532103 to 532104, EPI_ISL_532106 to 532117, EPI_ISL_532119 to 532122, EPI_ISL_532124, EPI_ISL_532126 to 532128, EPI_ISL_532132 to 532137, EPI_ISL_532139 to 532140, EPI_ISL_532143, EPI_ISL_532146, EPI_ISL_532150 to 532152, EPI_ISL_532157, EPI_ISL_532159 to 532160, EPI_ISL_532162, EPI_ISL_532164 to 532166, EPI_ISL_532169, EPI_ISL_532171 to 532174, EPI_ISL_532176, EPI_ISL_532180 to 532181, EPI_ISL_532183 to 532185, EPI_ISL_532188 to 532191, EPI_ISL_532193, EPI_ISL_532199, EPI_ISL_532201, EPI_ISL_532204 to 532206, EPI_ISL_532209, EPI_ISL_532211 to 532213, EPI_ISL_532216 to 532217, EPI_ISL_532220, EPI_ISL_532222 to 532224, EPI_ISL_532226 to 532228, EPI_ISL_532230 to 532231, EPI_ISL_532233 to 532234, EPI_ISL_532236 to 532237, EPI_ISL_532239, EPI_ISL_532243, EPI_ISL_532247 to 532248, EPI_ISL_532251 to 532252, EPI_ISL_532254 to 532257, EPI_ISL_532261 to 532262, EPI_ISL_532265, EPI_ISL_532267, EPI_ISL_532270, EPI_ISL_532273, EPI_ISL_532277, EPI_ISL_532279 to 532280, EPI_ISL_532282, EPI_ISL_532284 to 532287, EPI_ISL_532289 to 532291, EPI_ISL_532295 to 532300, EPI_ISL_532302 to 532310, EPI_ISL_532312 to 532315, EPI_ISL_532318 to 532320, EPI_ISL_532322 to 532326, EPI_ISL_532329 to 532333, EPI_ISL_532336 to 532337, EPI_ISL_532339, EPI_ISL_532343 to 532346, EPI_ISL_532348, EPI_ISL_532350, EPI_ISL_532354, EPI_ISL_532357 to 532362, EPI_ISL_532365 to 532378, EPI_ISL_532380 to 532383, EPI_ISL_532385 to 532387, EPI_ISL_532389, EPI_ISL_532391 to 532393, EPI_ISL_532395 to 532398, EPI_ISL_532400 to 532401, EPI_ISL_532405 to 532406, EPI_ISL_532408 to 532413, EPI_ISL_532415 to 532429, EPI_ISL_532433 to 532437, EPI_ISL_532439, EPI_ISL_532441 to 532442, EPI_ISL_532445, EPI_ISL_532447, EPI_ISL_532449 to 532451, EPI_ISL_532453, EPI_ISL_532455, EPI_ISL_532457 to 532462, EPI_ISL_532465 to 532468, EPI_ISL_532470 to 532471, EPI_ISL_532474, EPI_ISL_532476 to 532479, EPI_ISL_532481 to 532483, EPI_ISL_532485, EPI_ISL_532487 to 532489, EPI_ISL_532491, EPI_ISL_532494 to 532505, EPI_ISL_532507 to 532509, EPI_ISL_532513, EPI_ISL_532516 to 532517, EPI_ISL_532519 to 532522, EPI_ISL_532524 to 532530, EPI_ISL_532532 to 532537, EPI_ISL_532539, EPI_ISL_532541 to 532545, EPI_ISL_532547 to 532551, EPI_ISL_532553 to 532562, EPI_ISL_532565, EPI_ISL_532567 to 532571, EPI_ISL_532573 to 532581, EPI_ISL_532584 to 532586, EPI_ISL_532588 to 532593, EPI_ISL_532595 to 532597, EPI_ISL_532599 to 532614, EPI_ISL_532616 to 532619, EPI_ISL_532622 to 532624, EPI_ISL_532627 to 532632, EPI_ISL_532634 to 532638, EPI_ISL_532641 to 532647, EPI_ISL_532649 to 532658, EPI_ISL_532660 to 532669, EPI_ISL_532671 to 532672, EPI_ISL_532674 to 532682, EPI_ISL_532684 to 532688, EPI_ISL_532690 to 532696, EPI_ISL_532698 to 532704, EPI_ISL_532706 to 532714, EPI_ISL_532716 to 532732, EPI_ISL_532734 to 532737, EPI_ISL_532739 to 532743, EPI_ISL_532745, EPI_ISL_532747 to 532749, EPI_ISL_532751 to 532752, EPI_ISL_532754, EPI_ISL_532756 to 532764, EPI_ISL_532766, EPI_ISL_532768 to 532778, EPI_ISL_532780 to 532783, EPI_ISL_532785, EPI_ISL_532787 to 532790, EPI_ISL_532792 to 532810, EPI_ISL_532812 to 532834, EPI_ISL_532836 to 532837, EPI_ISL_532840, EPI_ISL_532842, EPI_ISL_532845, EPI_ISL_532847 to 532848, EPI_ISL_532853 to 532856, EPI_ISL_532860, EPI_ISL_532862, EPI_ISL_532867, EPI_ISL_532869, EPI_ISL_532877, EPI_ISL_532879, EPI_ISL_532881 to 532882, EPI_ISL_532887, EPI_ISL_532890, EPI_ISL_532892 to 532893, EPI_ISL_532897, EPI_ISL_532899 to 532901, EPI_ISL_532904, EPI_ISL_532911 to 532912, EPI_ISL_532914, EPI_ISL_532917 to 532918, EPI_ISL_532923 to 532924, EPI_ISL_532929 to 532930, EPI_ISL_532935, EPI_ISL_532938, EPI_ISL_532940 to 532941, EPI_ISL_532944 to 532945, EPI_ISL_532947, EPI_ISL_532949 to 532950, EPI_ISL_532953 to 532956, EPI_ISL_532959, EPI_ISL_532961, EPI_ISL_532963 to 532975, EPI_ISL_532977, EPI_ISL_532979 to 532982, EPI_ISL_532984 to 532987, EPI_ISL_532989, EPI_ISL_532991, EPI_ISL_532993, EPI_ISL_532995 to 532999, EPI_ISL_533001 to 533013, EPI_ISL_533015, EPI_ISL_533017 to 533020, EPI_ISL_533022 to 533026, EPI_ISL_533028 to 533031, EPI_ISL_533033 to 533053, EPI_ISL_533055 to 533071, EPI_ISL_533073 to 533085, EPI_ISL_533087 to 533099, EPI_ISL_533102 to 533112, EPI_ISL_533114 to 533116, EPI_ISL_533118, EPI_ISL_533121 to 533153, EPI_ISL_533155 to 533224, EPI_ISL_533226 to 533227, EPI_ISL_533229, EPI_ISL_533233 to 533235, EPI_ISL_533238 to 533240, EPI_ISL_533242 to 533250, EPI_ISL_533253 to 533255, EPI_ISL_533257 to 533261, EPI_ISL_533263 to 533269, EPI_ISL_533272 to 533282, EPI_ISL_533284 to 533288, EPI_ISL_533290 to 533297, EPI_ISL_533299 to 533310, EPI_ISL_533313 to 533342, EPI_ISL_533344 to 533345, EPI_ISL_533347 to 533349, EPI_ISL_533352 to 533354, EPI_ISL_533356 to 533357, EPI_ISL_533359 to 533364, EPI_ISL_533366 to 533375, EPI_ISL_533377 to 533386, EPI_ISL_533388 to 533393, EPI_ISL_533395, EPI_ISL_533397 to 533402, EPI_ISL_533404 to 533405, EPI_ISL_533407 to 533410, EPI_ISL_533412, EPI_ISL_533414 to 533418, EPI_ISL_533420, EPI_ISL_533422 to 533434, EPI_ISL_536819 to 536843, EPI_ISL_536845 to 536888, EPI_ISL_536890, EPI_ISL_536892 to 536900, EPI_ISL_536902 to 536977, EPI_ISL_536980, EPI_ISL_536982 to 537016, EPI_ISL_537018 to 537019, EPI_ISL_537021 to 537034, EPI_ISL_537036 to 537044, EPI_ISL_537047 to 537124, EPI_ISL_537126 to 537151, EPI_ISL_537154 to 537185, EPI_ISL_537187 to 537205, EPI_ISL_539897 to 539900, EPI_ISL_539902 to 539905, EPI_ISL_539907 to 539960, EPI_ISL_539963 to 539981, EPI_ISL_539983 to 540000, EPI_ISL_540002 to 540019, EPI_ISL_540021 to 540024, EPI_ISL_540026 to 540029, EPI_ISL_540031 to 540048, EPI_ISL_540050 to 540058, EPI_ISL_540060 to 540099, EPI_ISL_540101 to 540152, EPI_ISL_540154 to 540155, EPI_ISL_540157 to 540246, EPI_ISL_540248 to 540252, EPI_ISL_540254 to 540277, EPI_ISL_540279 to 540280, EPI_ISL_540282 to 540286, EPI_ISL_540288 to 540316, EPI_ISL_540319 to 540320, EPI_ISL_540323 to 540333, EPI_ISL_540335 to 540350, EPI_ISL_540352 to 540358, EPI_ISL_540360 to 540411, EPI_ISL_540413 to 540420, EPI_ISL_541784 to 541838, EPI_ISL_541840, EPI_ISL_541842 to 541846, EPI_ISL_549547 to 549627, EPI_ISL_549629 to 549649, EPI_ISL_549651, EPI_ISL_549653, EPI_ISL_549655 to 549682, EPI_ISL_549684 to 549695, EPI_ISL_549697 to 549699, EPI_ISL_549701 to 549705, EPI_ISL_549707 to 549744, EPI_ISL_549746 to 549750, EPI_ISL_549752 to 549771, EPI_ISL_549773 to 549783, EPI_ISL_549785 to 549788, EPI_ISL_549790 to 549806, EPI_ISL_553732, EPI_ISL_566998 to 567023, EPI_ISL_567025 to 567033, EPI_ISL_567035 to 567058, EPI_ISL_567060 to 567109, EPI_ISL_567111 to 567183, EPI_ISL_567185, EPI_ISL_567187 to 567201, EPI_ISL_567203 to 567230, EPI_ISL_567232 to 567275, EPI_ISL_567278 to 567445, EPI_ISL_567447 to 567452, EPI_ISL_567454 to 567455, EPI_ISL_567457 to 567475, EPI_ISL_567477 to 567532, EPI_ISL_568223 to 568288, EPI_ISL_568291 to 568300, EPI_ISL_568302 to 568309, EPI_ISL_568311 to 568314, EPI_ISL_568316, EPI_ISL_568318 to 568340, EPI_ISL_568342 to 568343, EPI_ISL_568345 to 568363, EPI_ISL_568366 to 568384, EPI_ISL_568388 to 568391, EPI_ISL_568393 to 568394, EPI_ISL_568396 to 568397, EPI_ISL_568399 to 568435, EPI_ISL_568437 to 568452, EPI_ISL_568455 to 568457, EPI_ISL_568459 to 568460, EPI_ISL_568462 to 568465, EPI_ISL_575338 to 575355, EPI_ISL_575357 to 575366, EPI_ISL_575368 to 575379, EPI_ISL_575381 to 575398, EPI_ISL_575995 to 576049, EPI_ISL_576051 to 576063, EPI_ISL_576065 to 576069, EPI_ISL_576071 to 576079, EPI_ISL_576081 to 576087, EPI_ISL_576089 to 576093, EPI_ISL_579759 to 579793, EPI_ISL_579795 to 579796, EPI_ISL_579798 to 579820, EPI_ISL_579822 to 579834, EPI_ISL_579836 to 579842, EPI_ISL_579844 to 579917, EPI_ISL_580858 to 580860, EPI_ISL_580862 to 580863, EPI_ISL_580865 to 580872, EPI_ISL_580874, EPI_ISL_580876, EPI_ISL_580878 to 580881, EPI_ISL_580883 to 580884, EPI_ISL_580886 to 580891, EPI_ISL_580893 to 580895, EPI_ISL_580899, EPI_ISL_580901, EPI_ISL_580905 to 580908, EPI_ISL_580911 to 580912, EPI_ISL_580914, EPI_ISL_580916, EPI_ISL_580919 to 580920, EPI_ISL_580922 to 580928, EPI_ISL_580930 to 580934, EPI_ISL_580936 to 580939, EPI_ISL_580941 to 580947, EPI_ISL_580949, EPI_ISL_580952 to 580958, EPI_ISL_580960, EPI_ISL_580962 to 580975, EPI_ISL_580977 to 580978, EPI_ISL_580981 to 580989, EPI_ISL_580992, EPI_ISL_580994, EPI_ISL_580997, EPI_ISL_580999, EPI_ISL_581001 to 581003, EPI_ISL_581005 to 581007, EPI_ISL_581010 to 581017, EPI_ISL_581019 to 581022, EPI_ISL_581024, EPI_ISL_581027, EPI_ISL_581030 to 581031, EPI_ISL_581033 to 581034, EPI_ISL_581036 to 581037, EPI_ISL_581039, EPI_ISL_581042 to 581044, EPI_ISL_581047 to 581053, EPI_ISL_581055, EPI_ISL_581057, EPI_ISL_581059 to 581063, EPI_ISL_581066, EPI_ISL_581068, EPI_ISL_581070 to 581073, EPI_ISL_581075, EPI_ISL_581077, EPI_ISL_581079 to 581083, EPI_ISL_581085, EPI_ISL_581087, EPI_ISL_581089 to 581092, EPI_ISL_581094 to 581096, EPI_ISL_581098, EPI_ISL_581100 to 581102, EPI_ISL_581104 to 581105, EPI_ISL_581108 to 581110, EPI_ISL_581112, EPI_ISL_581127, EPI_ISL_581132 to 581133, EPI_ISL_581141, EPI_ISL_581146, EPI_ISL_581154, EPI_ISL_581166, EPI_ISL_581174, EPI_ISL_581182 to 581183, EPI_ISL_581189, EPI_ISL_581193 to 581194, EPI_ISL_581196, EPI_ISL_581214, EPI_ISL_581217, EPI_ISL_581223, EPI_ISL_581226, EPI_ISL_581229 to 581230, EPI_ISL_581245, EPI_ISL_581247, EPI_ISL_581267, EPI_ISL_581285, EPI_ISL_581291, EPI_ISL_581310, EPI_ISL_581323 to 581324, EPI_ISL_581329, EPI_ISL_581334, EPI_ISL_581337, EPI_ISL_581341, EPI_ISL_581344, EPI_ISL_581346, EPI_ISL_581350, EPI_ISL_581354, EPI_ISL_581358, EPI_ISL_581377,

EPI_ISL_581381, EPI_ISL_581384, EPI_ISL_581387, EPI_ISL_581392, EPI_ISL_581395, EPI_ISL_581402 to 581403, EPI_ISL_581405 to 581408, EPI_ISL_581410, EPI_ISL_581418 to 581420, EPI_ISL_581437, EPI_ISL_581439, EPI_ISL_581441, EPI_ISL_581443, EPI_ISL_587304 to 587322, EPI_ISL_587324 to 587349, EPI_ISL_587351 to 587378, EPI_ISL_587380 to 587385, EPI_ISL_587387 to 587436, EPI_ISL_587438 to 587474, EPI_ISL_587476, EPI_ISL_587478 to 587528, EPI_ISL_587530 to 587534, EPI_ISL_587536, EPI_ISL_587538 to 587596, EPI_ISL_587598 to 587601, EPI_ISL_587603 to 587633, EPI_ISL_587635 to 587641, EPI_ISL_587643 to 587667, EPI_ISL_587669 to 587710, EPI_ISL_588060 to 588074, EPI_ISL_588076 to 588113, EPI_ISL_588115 to 588117, EPI_ISL_588119 to 588124, EPI_ISL_588126 to 588127, EPI_ISL_588129 to 588174, EPI_ISL_588176 to 588182, EPI_ISL_588184 to 588217, EPI_ISL_588219 to 588248, EPI_ISL_588250 to 588251, EPI_ISL_588253 to 588279, EPI_ISL_588282 to 588284, EPI_ISL_588286 to 588297, EPI_ISL_588299 to 588304, EPI_ISL_588306 to 588308, EPI_ISL_588310 to 588334, EPI_ISL_588336 to 588346, EPI_ISL_588348, EPI_ISL_588350 to 588387, EPI_ISL_588389 to 588400, EPI_ISL_588402 to 588426, EPI_ISL_588428 to 588437, EPI_ISL_588439 to 588443, EPI_ISL_588445 to 588454, EPI_ISL_588456 to 588495, EPI_ISL_588497 to 588522, EPI_ISL_588524 to 588535, EPI_ISL_588537 to 588540, EPI_ISL_588542 to 588571, EPI_ISL_588573 to 588663, EPI_ISL_588665 to 588673, EPI_ISL_588675 to 588694, EPI_ISL_588696 to 588697, EPI_ISL_588699 to 588716, EPI_ISL_588718 to 588843, EPI_ISL_588846 to 588866, EPI_ISL_588868 to 588918, EPI_ISL_588920 to 588926, EPI_ISL_588928 to 588939, EPI_ISL_588941 to 588961, EPI_ISL_588963 to 589011, EPI_ISL_589013 to 589014, EPI_ISL_589016, EPI_ISL_589020, EPI_ISL_589022 to 589023, EPI_ISL_589025, EPI_ISL_589027, EPI_ISL_589031 to 589033, EPI_ISL_589035, EPI_ISL_589038, EPI_ISL_589042, EPI_ISL_589044 to 589049, EPI_ISL_589052 to 589056, EPI_ISL_589058 to 589062, EPI_ISL_589064 to 589066, EPI_ISL_589068 to 589069, EPI_ISL_589071 to 589073, EPI_ISL_589076, EPI_ISL_589078, EPI_ISL_589082, EPI_ISL_589084, EPI_ISL_589087 to 589089, EPI_ISL_589092 to 589094, EPI_ISL_589096, EPI_ISL_589100, EPI_ISL_589102, EPI_ISL_589105 to 589107, EPI_ISL_589112 to 589113, EPI_ISL_589116, EPI_ISL_589119 to 589120, EPI_ISL_589124 to 589126, EPI_ISL_589128 to 589129, EPI_ISL_589131 to 589132, EPI_ISL_589134, EPI_ISL_589136, EPI_ISL_589138 to 589146, EPI_ISL_589148 to 589152, EPI_ISL_589154 to 589156, EPI_ISL_589158 to 589159, EPI_ISL_589162 to 589165, EPI_ISL_589175, EPI_ISL_589177 to 589178, EPI_ISL_589180 to 589181, EPI_ISL_589186, EPI_ISL_589188 to 589194, EPI_ISL_589197, EPI_ISL_589201 to 589204, EPI_ISL_589206 to 589210, EPI_ISL_589213 to 589220, EPI_ISL_589222 to 589223, EPI_ISL_589228, EPI_ISL_589230, EPI_ISL_589232, EPI_ISL_589234, EPI_ISL_589236, EPI_ISL_589238 to 589241, EPI_ISL_589244 to 589246, EPI_ISL_589249 to 589250, EPI_ISL_589252, EPI_ISL_589254 to 589255, EPI_ISL_589257, EPI_ISL_589263, EPI_ISL_589774 to 589777, EPI_ISL_589788, EPI_ISL_589790, EPI_ISL_589793, EPI_ISL_589803 to 589834, EPI_ISL_589836 to 589844, EPI_ISL_589846 to 589890, EPI_ISL_589892 to 589903, EPI_ISL_589905 to 589912, EPI_ISL_589914 to 589934, EPI_ISL_589937 to 589962, EPI_ISL_589964 to 589972, EPI_ISL_589975 to 590007, EPI_ISL_590009 to 590034, EPI_ISL_590036 to 590045, EPI_ISL_590047 to 590069, EPI_ISL_590071 to 590092, EPI_ISL_590094 to 590105, EPI_ISL_590107 to 590126, EPI_ISL_590128 to 590147, EPI_ISL_590149 to 590171, EPI_ISL_590176 to 590180, EPI_ISL_590182 to 590185, EPI_ISL_590187 to 590192, EPI_ISL_590195 to 590197, EPI_ISL_590199, EPI_ISL_590201 to 590205, EPI_ISL_590207 to 590209, EPI_ISL_590211 to 590212, EPI_ISL_590216 to 590219, EPI_ISL_590221 to 590223, EPI_ISL_590227, EPI_ISL_590229 to 590233, EPI_ISL_590236 to 590238, EPI_ISL_590240 to 590245, EPI_ISL_590247 to 590249, EPI_ISL_590251 to 590254, EPI_ISL_590256, EPI_ISL_590258, EPI_ISL_590262, EPI_ISL_590264, EPI_ISL_590266 to 590271, EPI_ISL_590273, EPI_ISL_590278 to 590281, EPI_ISL_590285, EPI_ISL_590287, EPI_ISL_590289 to 590291, EPI_ISL_590293 to 590294, EPI_ISL_590297 to 590299, EPI_ISL_590301 to 590305, EPI_ISL_590308, EPI_ISL_590310 to 590311, EPI_ISL_590313, EPI_ISL_590315 to 590318, EPI_ISL_590320, EPI_ISL_590322 to 590326, EPI_ISL_590330 to 590334, EPI_ISL_590336, EPI_ISL_590338 to 590345, EPI_ISL_590348 to 590350, EPI_ISL_590353 to 590354, EPI_ISL_590356, EPI_ISL_590358, EPI_ISL_590360, EPI_ISL_590362 to 590363, EPI_ISL_590365, EPI_ISL_590368, EPI_ISL_590370 to 590372, EPI_ISL_590374 to 590378, EPI_ISL_590381 to 590382, EPI_ISL_590384 to 590385, EPI_ISL_590387, EPI_ISL_590389, EPI_ISL_590391, EPI_ISL_590393 to 590395, EPI_ISL_590400 to 590402, EPI_ISL_590404, EPI_ISL_590406 to 590418, EPI_ISL_590420 to 590421, EPI_ISL_590424 to 590437, EPI_ISL_590439 to 590440, EPI_ISL_590443 to 590445, EPI_ISL_590447, EPI_ISL_590449 to 590451, EPI_ISL_590454 to 590461, EPI_ISL_590463 to 590468, EPI_ISL_590470 to 590496, EPI_ISL_590498 to 590562, EPI_ISL_590564 to 590607, EPI_ISL_590609 to 590621, EPI_ISL_590623 to 590681, EPI_ISL_590683 to 590688, EPI_ISL_597234 to 597236, EPI_ISL_597240, EPI_ISL_597242, EPI_ISL_597244 to 597245, EPI_ISL_597249, EPI_ISL_597252, EPI_ISL_597260 to 597261, EPI_ISL_597263, EPI_ISL_597267 to 597268, EPI_ISL_597272, EPI_ISL_597275 to 597278, EPI_ISL_597280 to 597281, EPI_ISL_597288, EPI_ISL_597290 to 597292, EPI_ISL_597296, EPI_ISL_597298 to 597299, EPI_ISL_597302 to 597303, EPI_ISL_597311 to 597315, EPI_ISL_597318, EPI_ISL_597322, EPI_ISL_597326, EPI_ISL_597328 to 597330, EPI_ISL_597332 to 597334, EPI_ISL_597336, EPI_ISL_597338 to 597339, EPI_ISL_597347 to 597348, EPI_ISL_597351 to 597353, EPI_ISL_597355 to 597357, EPI_ISL_597359 to 597360, EPI_ISL_597362 to 597364, EPI_ISL_597369 to 597370, EPI_ISL_597376, EPI_ISL_597379, EPI_ISL_597382 to 597387, EPI_ISL_597389, EPI_ISL_597391, EPI_ISL_597393 to 597395, EPI_ISL_597400, EPI_ISL_597404, EPI_ISL_597406 to 597410, EPI_ISL_597413, EPI_ISL_597415 to 597417, EPI_ISL_597419 to 597423, EPI_ISL_597429 to 597431, EPI_ISL_597433 to 597434, EPI_ISL_597436, EPI_ISL_597439 to 597440, EPI_ISL_597442 to 597444, EPI_ISL_597447 to 597448, EPI_ISL_597452, EPI_ISL_597454 to 597457, EPI_ISL_597463, EPI_ISL_597465, EPI_ISL_597471, EPI_ISL_597474 to 597475, EPI_ISL_597481 to 597482, EPI_ISL_597484 to 597485, EPI_ISL_597491 to 597493, EPI_ISL_597496, EPI_ISL_597498 to 597501, EPI_ISL_597503, EPI_ISL_597506, EPI_ISL_597509, EPI_ISL_597511, EPI_ISL_597513 to 597516, EPI_ISL_597518, EPI_ISL_597521 to 597522, EPI_ISL_598831 to 598901, EPI_ISL_598903 to 598912, EPI_ISL_598914 to 598941, EPI_ISL_598943 to 598953, EPI_ISL_598955 to 598969, EPI_ISL_598971 to 598991, EPI_ISL_598993 to 599009, EPI_ISL_599011 to 599022, EPI_ISL_599024 to 599042, EPI_ISL_599044 to 599050, EPI_ISL_599052 to 599057, EPI_ISL_599059 to 599064, EPI_ISL_599066 to 599076, EPI_ISL_599078 to 599093, EPI_ISL_599095 to 599119, EPI_ISL_599121 to 599164, EPI_ISL_599167, EPI_ISL_599169 to 599170, EPI_ISL_599172 to 599182, EPI_ISL_599184 to 599193, EPI_ISL_599195 to 599200, EPI_ISL_599202 to 599204, EPI_ISL_599206 to 599227, EPI_ISL_599229 to 599232, EPI_ISL_599234 to 599269, EPI_ISL_599271, EPI_ISL_599273 to 599313, EPI_ISL_599315 to 599335, EPI_ISL_599337 to 599342, EPI_ISL_599344 to 599359, EPI_ISL_599361 to 599373, EPI_ISL_599375 to 599393, EPI_ISL_599395 to 599448, EPI_ISL_599450 to 599477, EPI_ISL_599479 to 599489, EPI_ISL_599491 to 599506, EPI_ISL_599508 to 599527, EPI_ISL_599529 to 599553, EPI_ISL_599555, EPI_ISL_599557, EPI_ISL_599559 to 599589, EPI_ISL_599591 to 599626, EPI_ISL_599629, EPI_ISL_599631 to 599647, EPI_ISL_599649 to 599662, EPI_ISL_599664 to 599698, EPI_ISL_599700 to 599725, EPI_ISL_599727 to 599730, EPI_ISL_599732 to 599782, EPI_ISL_599784 to 599786, EPI_ISL_599788 to 599789, EPI_ISL_599791 to 599810, EPI_ISL_599812 to 599815, EPI_ISL_599817, EPI_ISL_600128 to 600138, EPI_ISL_600141 to 600197, EPI_ISL_600199 to 600205, EPI_ISL_600208 to 600211, EPI_ISL_600213 to 600220, EPI_ISL_600222 to 600236, EPI_ISL_600238 to 600271, EPI_ISL_600273 to 600286, EPI_ISL_600288 to 600301, EPI_ISL_600303 to 600319, EPI_ISL_600321 to 600323, EPI_ISL_600325 to 600326, EPI_ISL_600328 to 600337, EPI_ISL_600339 to 600385, EPI_ISL_600387 to 600407, EPI_ISL_600409 to 600427, EPI_ISL_600447, EPI_ISL_600450, EPI_ISL_600452 to 600453, EPI_ISL_600455, EPI_ISL_600457 to 600458, EPI_ISL_600460, EPI_ISL_600462, EPI_ISL_600464 to 600465, EPI_ISL_600467, EPI_ISL_600469 to 600470, EPI_ISL_600472, EPI_ISL_600474 to 600475, EPI_ISL_600477, EPI_ISL_600479, EPI_ISL_600481 to 600483, EPI_ISL_600485 to 600486, EPI_ISL_600488, EPI_ISL_600490 to 600491, EPI_ISL_600493 to 600494, EPI_ISL_600496 to 600497, EPI_ISL_600499, EPI_ISL_600501, EPI_ISL_600503 to 600504, EPI_ISL_600506, EPI_ISL_600508 to 600509, EPI_ISL_600511, EPI_ISL_600513 to 600514, EPI_ISL_600516, EPI_ISL_600518, EPI_ISL_600520, EPI_ISL_600539 to 600540, EPI_ISL_600542, EPI_ISL_600544 to 600545, EPI_ISL_600547 to 600548, EPI_ISL_600550 to 600551, EPI_ISL_600553, EPI_ISL_600555 to 600556, EPI_ISL_600558, EPI_ISL_600560 to 600561, EPI_ISL_600563, EPI_ISL_600565 to 600566, EPI_ISL_600568 to 600569, EPI_ISL_600571 to 600592, EPI_ISL_600594 to 600602, EPI_ISL_600604, EPI_ISL_600606 to 600634, EPI_ISL_600637 to 600642, EPI_ISL_600644 to 600696, EPI_ISL_600698 to 600710, EPI_ISL_600712, EPI_ISL_600714 to 600783, EPI_ISL_600785 to 600814, EPI_ISL_600816 to 600823, EPI_ISL_600825 to 600850, EPI_ISL_600852 to 600881, EPI_ISL_600883 to 600891, EPI_ISL_600893 to 600900, EPI_ISL_600902 to 600903, EPI_ISL_600905 to 600911, EPI_ISL_600914, EPI_ISL_600916 to 600922, EPI_ISL_600924, EPI_ISL_600932 to 600955, EPI_ISL_600957 to 601021, EPI_ISL_601023 to 601087, EPI_ISL_601089 to 601109, EPI_ISL_601111 to 601142, EPI_ISL_601144 to 601149, EPI_ISL_601151 to 601161, EPI_ISL_601163 to 601191, EPI_ISL_601194 to 601203, EPI_ISL_601205 to 601209, EPI_ISL_601211 to 601213, EPI_ISL_601215 to 601223, EPI_ISL_601225 to 601233, EPI_ISL_601236 to 601238, EPI_ISL_601241, EPI_ISL_601244, EPI_ISL_601246, EPI_ISL_601250 to 601255, EPI_ISL_601257, EPI_ISL_601259 to 601261, EPI_ISL_601265 to 601266, EPI_ISL_601269, EPI_ISL_601271, EPI_ISL_601276, EPI_ISL_601279, EPI_ISL_601284 to 601286, EPI_ISL_601288, EPI_ISL_601295 to 601296, EPI_ISL_601300, EPI_ISL_601308 to 601309, EPI_ISL_601315, EPI_ISL_601323, EPI_ISL_601328, EPI_ISL_601331 to 601336, EPI_ISL_601341 to 601342, EPI_ISL_601344, EPI_ISL_601347 to 601348, EPI_ISL_601351 to 601353, EPI_ISL_601355 to 601358, EPI_ISL_601361 to 601362, EPI_ISL_601364, EPI_ISL_601372, EPI_ISL_601374, EPI_ISL_601377, EPI_ISL_601380, EPI_ISL_601384, EPI_ISL_601386 to 601387, EPI_ISL_601389, EPI_ISL_601391, EPI_ISL_601393, EPI_ISL_601400 to 601403, EPI_ISL_601405, EPI_ISL_601407 to 601408, EPI_ISL_601411, EPI_ISL_601413 to 601414, EPI_ISL_601416, EPI_ISL_601421, EPI_ISL_601423 to 601424, EPI_ISL_601426, EPI_ISL_601429 to 601430, EPI_ISL_601432 to 601433, EPI_ISL_601435 to 601436, EPI_ISL_601445 to 601446, EPI_ISL_601453 to 601454, EPI_ISL_601459 to 601462, EPI_ISL_601466 to 601467, EPI_ISL_601469, EPI_ISL_601471, EPI_ISL_601477, EPI_ISL_601479, EPI_ISL_601481 to 601483, EPI_ISL_601486, EPI_ISL_601488 to 601489, EPI_ISL_601787 to 601796, EPI_ISL_601798 to 601833, EPI_ISL_601835 to 601928, EPI_ISL_601930 to 601946, EPI_ISL_601948 to 601949, EPI_ISL_601951 to 601992, EPI_ISL_601994 to 601995, EPI_ISL_601997 to 601999, EPI_ISL_602001 to 602005, EPI_ISL_602007, EPI_ISL_602009 to 602014, EPI_ISL_602016 to 602026, EPI_ISL_602030 to 602066, EPI_ISL_602068 to 602076, EPI_ISL_602078 to 602080, EPI_ISL_602082, EPI_ISL_602084 to 602088, EPI_ISL_602092 to 602101, EPI_ISL_602103 to 602105, EPI_ISL_602107 to 602108, EPI_ISL_602110 to 602119, EPI_ISL_602121, EPI_ISL_602123 to 602140, EPI_ISL_602142 to 602144, EPI_ISL_602156 to 602157, EPI_ISL_609178 to 609179, EPI_ISL_609182, EPI_ISL_609184 to 609186, EPI_ISL_609189 to 609191, EPI_ISL_609197 to 609199, EPI_ISL_609202, EPI_ISL_609206, EPI_ISL_611287 to 611303, EPI_ISL_611305 to 611376, EPI_ISL_611379 to 611467, EPI_ISL_611469 to 611484, EPI_ISL_623863, EPI_ISL_623865, EPI_ISL_623868, EPI_ISL_623872 to 623874, EPI_ISL_623876, EPI_ISL_623883 to 623884, EPI_ISL_623886, EPI_ISL_623893, EPI_ISL_623895, EPI_ISL_623903, EPI_ISL_623908, EPI_ISL_623911, EPI_ISL_623916 to 623917, EPI_ISL_623921, EPI_ISL_623924, EPI_ISL_623930, EPI_ISL_623933 to 623934, EPI_ISL_623942, EPI_ISL_623944, EPI_ISL_623949, EPI_ISL_623954, EPI_ISL_623956, EPI_ISL_623959, EPI_ISL_623963, EPI_ISL_623975 to 623977, EPI_ISL_623986, EPI_ISL_623990 to 623991, EPI_ISL_623994 to 623995, EPI_ISL_624010, EPI_ISL_624012, EPI_ISL_624018 to 624019, EPI_ISL_624022, EPI_ISL_624027 to 624029, EPI_ISL_624032, EPI_ISL_624034, EPI_ISL_624043 to 624044, EPI_ISL_624049, EPI_ISL_624067, EPI_ISL_624073, EPI_ISL_624077, EPI_ISL_624079, EPI_ISL_624092, EPI_ISL_624105, EPI_ISL_624112, EPI_ISL_624114, EPI_ISL_624120, EPI_ISL_624122, EPI_ISL_624128, EPI_ISL_624132, EPI_ISL_624140, EPI_ISL_624145, EPI_ISL_624147 to 624148, EPI_ISL_624158, EPI_ISL_624163 to 624165, EPI_ISL_624176, EPI_ISL_624181, EPI_ISL_624188 to 624189, EPI_ISL_625315 to 625389, EPI_ISL_625391 to 625395, EPI_ISL_629754 to 629844, EPI_ISL_629846 to 629939, EPI_ISL_629941 to 629949, EPI_ISL_629951 to 630001, EPI_ISL_630003 to 630006, EPI_ISL_630702 to 630705, EPI_ISL_630718 to 630719, EPI_ISL_630725, EPI_ISL_630731, EPI_ISL_630733, EPI_ISL_630741, EPI_ISL_630745, EPI_ISL_630749, EPI_ISL_630761, EPI_ISL_630764, EPI_ISL_630775, EPI_ISL_630778, EPI_ISL_630782, EPI_ISL_630787, EPI_ISL_630804, EPI_ISL_630813, EPI_ISL_630815, EPI_ISL_630824, EPI_ISL_630827, EPI_ISL_630833, EPI_ISL_630838, EPI_ISL_630850 to 630851, EPI_ISL_630853, EPI_ISL_630859, EPI_ISL_630862, EPI_ISL_630864, EPI_ISL_630870, EPI_ISL_630872 to 630873, EPI_ISL_630875 to 630876, EPI_ISL_630883, EPI_ISL_630888, EPI_ISL_630900, EPI_ISL_630915, EPI_ISL_630919, EPI_ISL_630923, EPI_ISL_630926, EPI_ISL_630931, EPI_ISL_630933, EPI_ISL_630941, EPI_ISL_630948 to 630949, EPI_ISL_630951, EPI_ISL_630955, EPI_ISL_630957, EPI_ISL_630959, EPI_ISL_630965, EPI_ISL_633103, EPI_ISL_633111 to 633113, EPI_ISL_633115, EPI_ISL_633119, EPI_ISL_633125, EPI_ISL_633132 to 633134, EPI_ISL_633137 to 633139, EPI_ISL_633143 to 633146, EPI_ISL_633151 to 633152, EPI_ISL_633154, EPI_ISL_633158, EPI_ISL_633160, EPI_ISL_633162, EPI_ISL_633166 to 633169, EPI_ISL_633171, EPI_ISL_633174 to 633175, EPI_ISL_633178 to 633182, EPI_ISL_633184 to 633185, EPI_ISL_633190 to 633191, EPI_ISL_633193, EPI_ISL_633195 to 633196, EPI_ISL_633201 to 633204, EPI_ISL_633206 to 633207, EPI_ISL_633209 to 633210, EPI_ISL_633213, EPI_ISL_633222, EPI_ISL_633226, EPI_ISL_633228, EPI_ISL_633230, EPI_ISL_633232 to 633233, EPI_ISL_633237, EPI_ISL_633240 to 633241, EPI_ISL_633243 to 633244, EPI_ISL_633251, EPI_ISL_633256, EPI_ISL_633259, EPI_ISL_633261 to 633262, EPI_ISL_633268 to 633269, EPI_ISL_633271 to 633272, EPI_ISL_633274, EPI_ISL_633276, EPI_ISL_633279 to 633280, EPI_ISL_633283, EPI_ISL_633285, EPI_ISL_633287, EPI_ISL_633289 to 633290, EPI_ISL_633293, EPI_ISL_633296, EPI_ISL_633301, EPI_ISL_633303 to 633307, EPI_ISL_633309 to 633310, EPI_ISL_633312, EPI_ISL_633316 to 633318, EPI_ISL_633320, EPI_ISL_633323, EPI_ISL_633326 to 633328, EPI_ISL_633331 to 633332, EPI_ISL_633335, EPI_ISL_633338 to 633339, EPI_ISL_633342, EPI_ISL_633348, EPI_ISL_633351 to 633353, EPI_ISL_633355, EPI_ISL_633358, EPI_ISL_633362, EPI_ISL_633364, EPI_ISL_633367 to 633368, EPI_ISL_633371, EPI_ISL_633376 to 633377, EPI_ISL_633379, EPI_ISL_633381 to 633382, EPI_ISL_633384, EPI_ISL_633386 to 633387, EPI_ISL_633391 to 633392, EPI_ISL_633394, EPI_ISL_633396, EPI_ISL_633400 to 633401, EPI_ISL_633404, EPI_ISL_633406, EPI_ISL_633409 to 633491, EPI_ISL_633493 to 633498, EPI_ISL_633500 to 633539, EPI_ISL_633541 to 633576, EPI_ISL_633578 to 633582, EPI_ISL_633584 to 633614, EPI_ISL_633616, EPI_ISL_633618 to 633652, EPI_ISL_633654 to 633699, EPI_ISL_633701 to 633759, EPI_ISL_633761 to 633781, EPI_ISL_633783 to 633798, EPI_ISL_633802 to 633820, EPI_ISL_633822 to 633916, EPI_ISL_633919 to 633930, EPI_ISL_633932 to 633961, EPI_ISL_633964 to 634035, EPI_ISL_634037 to 634119, EPI_ISL_634121 to 634155, EPI_ISL_634157 to 634162, EPI_ISL_634164 to 634166, EPI_ISL_634168 to 634194, EPI_ISL_634196 to 634255, EPI_ISL_634521 to 634586, EPI_ISL_634588 to 634597, EPI_ISL_634599 to 634612, EPI_ISL_634614 to 634664, EPI_ISL_634666 to 634667, EPI_ISL_634670 to 634677, EPI_ISL_634679 to 634736, EPI_ISL_634738 to 634753

| | | | |
|---|---|---|---|
| see above | Lighthouse Lab in Glasgow | Wellcome Sanger Institute for the COVID-19 Genomics UK (COG-UK) consortium | Anna Dominiczak and Alex Alderton; Carol Clugston; Cordelia Langford; David Gray; David K. Jackson; Dominic Kwiatkowski; Ewan Harrison; Harper VanSteenhouse; Ian Johnston; John Sillitoe; John Sillitoe on behalf of the Wellcome Sanger Institute COVID-19 Surveillance Team; John Sillitoe on behalf of the Wellcome Sanger Institute COVID-19 Surveillance Team (http://www.sanger.ac.uk/covid-team); Roberto Amato; Sonia Goncalves; Yumi Kasai |

EPI_ISL_540823 to 540870, EPI_ISL_559947 to 559970, EPI_ISL_573769, EPI_ISL_585297 to 585341, EPI_ISL_585343 to 585344, EPI_ISL_585346 to 585421, EPI_ISL_650844, EPI_ISL_651424, EPI_ISL_651790 to 651810, EPI_ISL_651886, EPI_ISL_865000 to 865003, EPI_ISL_865008

| | | | |
|---|---|---|---|
| see above | Lighthouse Lab in Glasgow / MRC-University of Glasgow Centre for Virus Research | COVID-19 Genomics UK (COG-UK) Consortium | Alasdair MacLean; Alice Broos; Ana da Silva Filipe; Anna Dominiczak; Antonia Ho; Carol Clugston; Daniel Mair; David Gray; David L Robertson; Elihu Aranday-Cortes; Emma Thomson; Harper VanSteenhouse; James Shepherd; Jenna Nichols; Joseph Hughes; Kathy Li; Kathy Smollett; Kyriaki Nomikou; Lily Tong; Marc Niebel; Matthew Holden; Natasha Jesudason; Natasha Johnson; Patawee Asamaphan; Rajiv Shah; Richard Orton; Rory Gunson; Sarah McDonald; Sharif Shaaban; Sreenu Vattipally; Stephen Carmichael; Yumi Kasai |

EPI_ISL_549861, EPI_ISL_550069, EPI_ISL_550849, EPI_ISL_550983, EPI_ISL_551007, EPI_ISL_551092, EPI_ISL_551218, EPI_ISL_552106, EPI_ISL_552157, EPI_ISL_552240, EPI_ISL_552458, EPI_ISL_553084, EPI_ISL_554195, EPI_ISL_554213, EPI_ISL_554273, EPI_ISL_554281, EPI_ISL_554662, EPI_ISL_554798, EPI_ISL_556207, EPI_ISL_557186, EPI_ISL_557189, EPI_ISL_557238, EPI_ISL_557355, EPI_ISL_557379, EPI_ISL_557843, EPI_ISL_557885, EPI_ISL_557952, EPI_ISL_558237, EPI_ISL_558261, EPI_ISL_558328, EPI_ISL_558921, EPI_ISL_559017, EPI_ISL_559264, EPI_ISL_559287, EPI_ISL_559670, EPI_ISL_559706, EPI_ISL_559724, EPI_ISL_559733 to 559734, EPI_ISL_559740, EPI_ISL_566951, EPI_ISL_566986, EPI_ISL_580840, EPI_ISL_581149, EPI_ISL_581366, EPI_ISL_589050, EPI_ISL_589130, EPI_ISL_589184, EPI_ISL_589262, EPI_ISL_589334, EPI_ISL_589567, EPI_ISL_598249, EPI_ISL_598305, EPI_ISL_598330 to 598331, EPI_ISL_598361, EPI_ISL_598408, EPI_ISL_598433, EPI_ISL_598436, EPI_ISL_598606, EPI_ISL_598638, EPI_ISL_598667, EPI_ISL_599845, EPI_ISL_599888, EPI_ISL_599948, EPI_ISL_599960, EPI_ISL_600056, EPI_ISL_601290, EPI_ISL_601652, EPI_ISL_601667, EPI_ISL_606311, EPI_ISL_606314, EPI_ISL_606353, EPI_ISL_606462, EPI_ISL_606516, EPI_ISL_606524, EPI_ISL_606608, EPI_ISL_606654, EPI_ISL_606797, EPI_ISL_607461, EPI_ISL_607516, EPI_ISL_607550, EPI_ISL_607572, EPI_ISL_607579, EPI_ISL_608748, EPI_ISL_608834, EPI_ISL_609230, EPI_ISL_609267, EPI_ISL_609302, EPI_ISL_623214, EPI_ISL_623259, EPI_ISL_623298, EPI_ISL_623311, EPI_ISL_623313, EPI_ISL_623428, EPI_ISL_623456, EPI_ISL_623484, EPI_ISL_623515, EPI_ISL_623522, EPI_ISL_623602, EPI_ISL_629138, EPI_ISL_629199, EPI_ISL_629219, EPI_ISL_629245, EPI_ISL_629409, EPI_ISL_629490, EPI_ISL_629512, EPI_ISL_629526, EPI_ISL_629580, EPI_ISL_629590, EPI_ISL_629592, EPI_ISL_629727, EPI_ISL_630191, EPI_ISL_630219, EPI_ISL_630249, EPI_ISL_631090, EPI_ISL_631102, EPI_ISL_634266, EPI_ISL_634341, EPI_ISL_634376, EPI_ISL_634384 to 634385, EPI_ISL_634406, EPI_ISL_634428, EPI_ISL_634465, EPI_ISL_634483, EPI_ISL_634499, EPI_ISL_642761 to 642835, EPI_ISL_642837 to 642838, EPI_ISL_642840 to 642918, EPI_ISL_642920 to 643043, EPI_ISL_643983 to 643991, EPI_ISL_644052 to 644064, EPI_ISL_644066 to 644092, EPI_ISL_644114, EPI_ISL_644116, EPI_ISL_644119 to 644120, EPI_ISL_645221, EPI_ISL_645226, EPI_ISL_645241, EPI_ISL_645243 to 645244, EPI_ISL_645263 to 645264, EPI_ISL_645285, EPI_ISL_645296, EPI_ISL_645300, EPI_ISL_645302, EPI_ISL_645308, EPI_ISL_645319, EPI_ISL_645324, EPI_ISL_645327, EPI_ISL_645329, EPI_ISL_645345, EPI_ISL_645354, EPI_ISL_645360, EPI_ISL_645367, EPI_ISL_645369, EPI_ISL_645374, EPI_ISL_645383, EPI_ISL_645391, EPI_ISL_645395, EPI_ISL_645399, EPI_ISL_645405 to 645406, EPI_ISL_645421, EPI_ISL_645424, EPI_ISL_645438, EPI_ISL_645441, EPI_ISL_645453, EPI_ISL_645477, EPI_ISL_645489, EPI_ISL_645513, EPI_ISL_645517 to 645518, EPI_ISL_645520, EPI_ISL_647807 to 647835, EPI_ISL_647837 to 647853, EPI_ISL_649192, EPI_ISL_649205, EPI_ISL_649255, EPI_ISL_649283, EPI_ISL_649320, EPI_ISL_649362, EPI_ISL_649386, EPI_ISL_649405, EPI_ISL_649424, EPI_ISL_649479, EPI_ISL_649548 to 649555, EPI_ISL_656705, EPI_ISL_656728, EPI_ISL_656731 to 656733, EPI_ISL_656744 to 656746, EPI_ISL_656757, EPI_ISL_656774, EPI_ISL_656777 to 656779, EPI_ISL_656788 to 656789, EPI_ISL_656798 to 656799, EPI_ISL_656807 to 656808, EPI_ISL_656810 to 656811, EPI_ISL_656824, EPI_ISL_656827, EPI_ISL_656829, EPI_ISL_656834 to 656835, EPI_ISL_656838, EPI_ISL_656840 to 656841, EPI_ISL_656846, EPI_ISL_656848, EPI_ISL_656860 to 656862, EPI_ISL_656865, EPI_ISL_656867, EPI_ISL_656869, EPI_ISL_656871, EPI_ISL_656877, EPI_ISL_656879, EPI_ISL_656887, EPI_ISL_656892 to 656893, EPI_ISL_656906, EPI_ISL_656910, EPI_ISL_656913, EPI_ISL_656920, EPI_ISL_656922, EPI_ISL_656924, EPI_ISL_656930, EPI_ISL_656935 to 656936, EPI_ISL_656938, EPI_ISL_656942, EPI_ISL_656945, EPI_ISL_656947, EPI_ISL_656955, EPI_ISL_656957 to 656958, EPI_ISL_656961 to 656962, EPI_ISL_656965 to 656966, EPI_ISL_656969, EPI_ISL_656973 to 656974, EPI_ISL_656976, EPI_ISL_656979 to 656980, EPI_ISL_656982, EPI_ISL_656987, EPI_ISL_656991, EPI_ISL_656997, EPI_ISL_657003 to 657004, EPI_ISL_657008, EPI_ISL_657011, EPI_ISL_657014, EPI_ISL_657016 to 657017, EPI_ISL_657021, EPI_ISL_657023, EPI_ISL_657026, EPI_ISL_657355, EPI_ISL_657359 to 657361, EPI_ISL_657364 to 657365, EPI_ISL_657367, EPI_ISL_657369 to 657373, EPI_ISL_657375, EPI_ISL_657377, EPI_ISL_657379, EPI_ISL_657381 to 657382, EPI_ISL_657384 to 657386, EPI_ISL_657390 to 657393, EPI_ISL_657396, EPI_ISL_657398, EPI_ISL_657400 to 657401, EPI_ISL_657405 to 657406, EPI_ISL_657408, EPI_ISL_657410 to 657411, EPI_ISL_657413 to 657414, EPI_ISL_657416 to 657420, EPI_ISL_657424 to 657425, EPI_ISL_657428 to 657430, EPI_ISL_657438 to 657441, EPI_ISL_657443, EPI_ISL_657446, EPI_ISL_657451, EPI_ISL_657453, EPI_ISL_657457, EPI_ISL_657459, EPI_ISL_657462, EPI_ISL_657464, EPI_ISL_657472 to 657473, EPI_ISL_657479 to 657480, EPI_ISL_657483 to 657488, EPI_ISL_657490 to 657491, EPI_ISL_657493 to 657495, EPI_ISL_657497, EPI_ISL_657500, EPI_ISL_657503, EPI_ISL_657505, EPI_ISL_657509 to 657510, EPI_ISL_657512 to 657513, EPI_ISL_657515, EPI_ISL_657518 to 657519, EPI_ISL_657526 to 657527, EPI_ISL_657531 to 657533, EPI_ISL_657535 to 657536, EPI_ISL_657542 to 657544, EPI_ISL_657547, EPI_ISL_657553 to 657558, EPI_ISL_657560, EPI_ISL_657563 to 657564, EPI_ISL_657567 to 657568, EPI_ISL_657570 to 657571, EPI_ISL_657574 to 657575, EPI_ISL_657577, EPI_ISL_657579, EPI_ISL_657581, EPI_ISL_657583 to 657584, EPI_ISL_657587, EPI_ISL_657589, EPI_ISL_657591, EPI_ISL_657593 to 657594, EPI_ISL_657596, EPI_ISL_657599, EPI_ISL_657602, EPI_ISL_657605 to 657611, EPI_ISL_657614 to 657616, EPI_ISL_657625, EPI_ISL_657627, EPI_ISL_657629, EPI_ISL_657631 to 657632, EPI_ISL_657635 to

657636, EPI_ISL_657639 to 657640, EPI_ISL_657642, EPI_ISL_657644, EPI_ISL_657647 to 657648, EPI_ISL_657650 to 657651, EPI_ISL_657657, EPI_ISL_657661 to 657663, EPI_ISL_657665, EPI_ISL_657667, EPI_ISL_657676, EPI_ISL_657680 to 657683, EPI_ISL_657688, EPI_ISL_657692, EPI_ISL_657694 to 657696, EPI_ISL_657700, EPI_ISL_657702 to 657703, EPI_ISL_657705 to 657706, EPI_ISL_657709 to 657712, EPI_ISL_657714 to 657719, EPI_ISL_657722, EPI_ISL_657727, EPI_ISL_657729 to 657730, EPI_ISL_657732 to 657735, EPI_ISL_657738, EPI_ISL_657740 to 657743, EPI_ISL_657745, EPI_ISL_657748, EPI_ISL_657751 to 657753, EPI_ISL_657755, EPI_ISL_657757 to 657758, EPI_ISL_657761, EPI_ISL_657768, EPI_ISL_657770, EPI_ISL_657772 to 657779, EPI_ISL_657781, EPI_ISL_657783 to 657785, EPI_ISL_657788 to 657789, EPI_ISL_657791 to 657792, EPI_ISL_657794, EPI_ISL_657796 to 657797, EPI_ISL_657800, EPI_ISL_657808, EPI_ISL_657811 to 657812, EPI_ISL_657814, EPI_ISL_657819 to 657823, EPI_ISL_657827, EPI_ISL_657829 to 657830, EPI_ISL_657832, EPI_ISL_657835 to 657836, EPI_ISL_657840, EPI_ISL_657842 to 657844, EPI_ISL_657848, EPI_ISL_657850, EPI_ISL_657854, EPI_ISL_657856, EPI_ISL_657858 to 657859, EPI_ISL_657861 to 657862, EPI_ISL_657865, EPI_ISL_657869 to 657870, EPI_ISL_657872, EPI_ISL_657874, EPI_ISL_657878 to 657879, EPI_ISL_657886 to 657887, EPI_ISL_657889, EPI_ISL_657892, EPI_ISL_657894, EPI_ISL_657898, EPI_ISL_657901, EPI_ISL_657905 to 657906, EPI_ISL_657910, EPI_ISL_657913, EPI_ISL_657915 to 657916, EPI_ISL_657918 to 657919, EPI_ISL_657921 to 657923, EPI_ISL_657925, EPI_ISL_657927 to 657930, EPI_ISL_657932, EPI_ISL_657938, EPI_ISL_657940 to 657941, EPI_ISL_657944, EPI_ISL_657948, EPI_ISL_657950, EPI_ISL_657954 to 657956, EPI_ISL_657958 to 657959, EPI_ISL_657961 to 657963, EPI_ISL_657965, EPI_ISL_657969 to 657973, EPI_ISL_657975, EPI_ISL_657977, EPI_ISL_657979 to 657982, EPI_ISL_657984 to 657985, EPI_ISL_657988, EPI_ISL_657991, EPI_ISL_657993, EPI_ISL_657995 to 657997, EPI_ISL_657999 to 658000, EPI_ISL_658002 to 658006, EPI_ISL_658009, EPI_ISL_658015 to 658018, EPI_ISL_658021 to 658023, EPI_ISL_658025 to 658027, EPI_ISL_658035, EPI_ISL_658038 to 658040, EPI_ISL_658042, EPI_ISL_658044, EPI_ISL_658047, EPI_ISL_658049 to 658051, EPI_ISL_658053, EPI_ISL_658057 to 658062, EPI_ISL_658065 to 658068, EPI_ISL_658070, EPI_ISL_658079, EPI_ISL_658081 to 658082, EPI_ISL_658087 to 658088, EPI_ISL_658090 to 658091, EPI_ISL_658094, EPI_ISL_658102 to 658103, EPI_ISL_658105 to 658109, EPI_ISL_658111 to 658115, EPI_ISL_658117 to 658118, EPI_ISL_658121, EPI_ISL_658123, EPI_ISL_658126 to 658127, EPI_ISL_658129 to 658130, EPI_ISL_658132, EPI_ISL_658135 to 658137, EPI_ISL_658139 to 658141, EPI_ISL_658143, EPI_ISL_658145 to 658147, EPI_ISL_658149 to 658152, EPI_ISL_658154 to 658155, EPI_ISL_658158, EPI_ISL_658165, EPI_ISL_658168 to 658170, EPI_ISL_658172 to 658174, EPI_ISL_658176, EPI_ISL_658180, EPI_ISL_658182 to 658185, EPI_ISL_658193, EPI_ISL_658204 to 658205, EPI_ISL_658210 to 658211, EPI_ISL_658213, EPI_ISL_658216 to 658217, EPI_ISL_658219, EPI_ISL_658223, EPI_ISL_658225, EPI_ISL_658229 to 658230, EPI_ISL_658232 to 658233, EPI_ISL_658235, EPI_ISL_658237 to 658238, EPI_ISL_658240 to 658241, EPI_ISL_658243 to 658245, EPI_ISL_658247 to 658249, EPI_ISL_658251, EPI_ISL_658253 to 658255, EPI_ISL_658257, EPI_ISL_658259, EPI_ISL_658261 to 658262, EPI_ISL_658264 to 658265, EPI_ISL_658267 to 658269, EPI_ISL_658273, EPI_ISL_658277, EPI_ISL_658280 to 658284, EPI_ISL_658289 to 658291, EPI_ISL_658293 to 658298, EPI_ISL_658300 to 658303, EPI_ISL_658306, EPI_ISL_658308 to 658309, EPI_ISL_658315, EPI_ISL_658318 to 658319, EPI_ISL_658322, EPI_ISL_658324 to 658325, EPI_ISL_658328 to 658329, EPI_ISL_658333, EPI_ISL_658336, EPI_ISL_658338 to 658339, EPI_ISL_658342, EPI_ISL_658344, EPI_ISL_658346 to 658347, EPI_ISL_658349 to 658350, EPI_ISL_658353 to 658354, EPI_ISL_658357 to 658359, EPI_ISL_658365, EPI_ISL_658368 to 658369, EPI_ISL_658372 to 658375, EPI_ISL_658378, EPI_ISL_658380, EPI_ISL_658384 to 658386, EPI_ISL_658389, EPI_ISL_658391 to 658393, EPI_ISL_658395, EPI_ISL_658399 to 658401, EPI_ISL_658403, EPI_ISL_658741 to 658743, EPI_ISL_658746 to 658749, EPI_ISL_658751, EPI_ISL_658754 to 658755, EPI_ISL_658757 to 658758, EPI_ISL_658761 to 658762, EPI_ISL_658765, EPI_ISL_658770, EPI_ISL_658772 to 658774, EPI_ISL_658776 to 658777, EPI_ISL_658779 to 658780, EPI_ISL_658782 to 658786, EPI_ISL_658789, EPI_ISL_658791 to 658794, EPI_ISL_658796 to 658799, EPI_ISL_658801 to 658804, EPI_ISL_658806, EPI_ISL_658808, EPI_ISL_658810 to 658812, EPI_ISL_658814 to 658815, EPI_ISL_658817 to 658828, EPI_ISL_658830 to 658832, EPI_ISL_658834 to 658835, EPI_ISL_658839 to 658845, EPI_ISL_658847 to 658851, EPI_ISL_658853 to 658862, EPI_ISL_658867, EPI_ISL_658869, EPI_ISL_658871 to 658872, EPI_ISL_658874, EPI_ISL_658876, EPI_ISL_658879, EPI_ISL_658882, EPI_ISL_658884, EPI_ISL_658887, EPI_ISL_658889 to 658890, EPI_ISL_658892, EPI_ISL_658894 to 658895, EPI_ISL_658897 to 658898, EPI_ISL_658903, EPI_ISL_658907 to 658908, EPI_ISL_658910 to 658911, EPI_ISL_658915, EPI_ISL_658917 to 658918, EPI_ISL_658920, EPI_ISL_658922, EPI_ISL_658924, EPI_ISL_658926 to 658929, EPI_ISL_658932, EPI_ISL_658934 to 658935, EPI_ISL_658937 to 658947, EPI_ISL_658949 to 658953, EPI_ISL_658955 to 658958, EPI_ISL_658960 to 658962, EPI_ISL_658965 to 658969, EPI_ISL_658972 to 658975, EPI_ISL_658977 to 658978, EPI_ISL_658981 to 658987, EPI_ISL_658989 to 658994, EPI_ISL_658997 to 658998, EPI_ISL_659001 to 659004, EPI_ISL_659006, EPI_ISL_659008 to 659010, EPI_ISL_659012 to 659016, EPI_ISL_659020 to 659024, EPI_ISL_659026 to 659027, EPI_ISL_659029 to 659034, EPI_ISL_659036 to 659039, EPI_ISL_659042 to 659044, EPI_ISL_659046 to 659049, EPI_ISL_659051 to 659052, EPI_ISL_659055 to 659069, EPI_ISL_659075, EPI_ISL_659078, EPI_ISL_659081, EPI_ISL_659085 to 659089, EPI_ISL_659092, EPI_ISL_659095 to 659096, EPI_ISL_662633 to 662634, EPI_ISL_662636 to 662641, EPI_ISL_662643 to 662646, EPI_ISL_673577, EPI_ISL_673592, EPI_ISL_673594, EPI_ISL_673599, EPI_ISL_673601, EPI_ISL_673603, EPI_ISL_673616, EPI_ISL_673621, EPI_ISL_673633 to 673634, EPI_ISL_673642, EPI_ISL_673646, EPI_ISL_673657, EPI_ISL_673659, EPI_ISL_673663, EPI_ISL_673673, EPI_ISL_673676, EPI_ISL_673686, EPI_ISL_673688, EPI_ISL_673691, EPI_ISL_673696, EPI_ISL_673702, EPI_ISL_673708 to 673709, EPI_ISL_673715, EPI_ISL_673722, EPI_ISL_673729 to 673730, EPI_ISL_673733, EPI_ISL_673738, EPI_ISL_673745, EPI_ISL_673750, EPI_ISL_673760, EPI_ISL_673765, EPI_ISL_673770, EPI_ISL_673778, EPI_ISL_673785, EPI_ISL_673787, EPI_ISL_673790, EPI_ISL_673798, EPI_ISL_673801, EPI_ISL_673803, EPI_ISL_673812 to 673814, EPI_ISL_673822, EPI_ISL_673827 to 673829, EPI_ISL_673832 to 673835, EPI_ISL_673840, EPI_ISL_673848, EPI_ISL_673856 to 673857, EPI_ISL_673859, EPI_ISL_673865, EPI_ISL_673867, EPI_ISL_673871 to 673878, EPI_ISL_673880 to 673885, EPI_ISL_673887, EPI_ISL_673891 to 673897, EPI_ISL_673900, EPI_ISL_673902 to 673903, EPI_ISL_673905 to 673908, EPI_ISL_673910 to 673917, EPI_ISL_673919 to 673924, EPI_ISL_673926 to 673930, EPI_ISL_673932, EPI_ISL_673934 to 673937, EPI_ISL_673939 to 673946, EPI_ISL_673948 to 673949, EPI_ISL_673952 to 673957, EPI_ISL_673959, EPI_ISL_673961, EPI_ISL_673963 to 673978, EPI_ISL_673980 to 673981, EPI_ISL_673983 to 673993, EPI_ISL_673995 to 673997, EPI_ISL_674000, EPI_ISL_674002 to 674004, EPI_ISL_674006 to 674009, EPI_ISL_674012 to 674018, EPI_ISL_674020 to 674031, EPI_ISL_674033 to 674037, EPI_ISL_674041 to 674048, EPI_ISL_674051 to 674057, EPI_ISL_674059 to 674068, EPI_ISL_674070 to 674071, EPI_ISL_674073 to 674081, EPI_ISL_674085 to 674086, EPI_ISL_674088 to 674091, EPI_ISL_674093 to 674106, EPI_ISL_674108 to 674109, EPI_ISL_674111 to 674115, EPI_ISL_674117 to 674132, EPI_ISL_674134 to 674135, EPI_ISL_674137 to 674143, EPI_ISL_674145 to 674151, EPI_ISL_674153 to 674155, EPI_ISL_674158, EPI_ISL_674163, EPI_ISL_674166 to 674167, EPI_ISL_674173, EPI_ISL_674180, EPI_ISL_674183 to 674185, EPI_ISL_674187, EPI_ISL_674189, EPI_ISL_674196, EPI_ISL_674198, EPI_ISL_674200, EPI_ISL_674202, EPI_ISL_674205, EPI_ISL_674209 to 674210, EPI_ISL_674212 to 674216, EPI_ISL_674219 to 674221, EPI_ISL_674225, EPI_ISL_674230, EPI_ISL_674232, EPI_ISL_674235 to 674239, EPI_ISL_674244, EPI_ISL_674246 to 674248, EPI_ISL_674250, EPI_ISL_674252, EPI_ISL_674256 to 674259, EPI_ISL_674262 to 674263, EPI_ISL_674265, EPI_ISL_674269 to 674270, EPI_ISL_674273 to 674275, EPI_ISL_674278 to 674280, EPI_ISL_674282, EPI_ISL_674288, EPI_ISL_674290 to 674291, EPI_ISL_674298, EPI_ISL_674302, EPI_ISL_674304, EPI_ISL_674306, EPI_ISL_674309, EPI_ISL_674312, EPI_ISL_674314 to 674316, EPI_ISL_674320 to 674321, EPI_ISL_674325 to 674326, EPI_ISL_674330 to 674331, EPI_ISL_674333, EPI_ISL_674335 to 674336, EPI_ISL_674338, EPI_ISL_674342, EPI_ISL_674344, EPI_ISL_674348 to 674350, EPI_ISL_674353, EPI_ISL_674355 to 674356, EPI_ISL_674358 to 674359, EPI_ISL_674363, EPI_ISL_674365, EPI_ISL_674371, EPI_ISL_674373, EPI_ISL_674378 to 674379, EPI_ISL_674382, EPI_ISL_674384, EPI_ISL_674386 to 674388, EPI_ISL_674393, EPI_ISL_674395, EPI_ISL_674398, EPI_ISL_674401, EPI_ISL_674403, EPI_ISL_674405, EPI_ISL_674407, EPI_ISL_674410, EPI_ISL_674414 to 674415, EPI_ISL_674417, EPI_ISL_674422 to 674423, EPI_ISL_674425 to 674427, EPI_ISL_674430, EPI_ISL_674433, EPI_ISL_674437, EPI_ISL_674440, EPI_ISL_674443 to 674444, EPI_ISL_674448 to 674450, EPI_ISL_674453, EPI_ISL_674457 to 674458, EPI_ISL_674460 to 674461, EPI_ISL_674464, EPI_ISL_674468, EPI_ISL_674473, EPI_ISL_674475, EPI_ISL_674480 to 674483, EPI_ISL_674489, EPI_ISL_674491 to 674492, EPI_ISL_674494, EPI_ISL_674496 to 674497, EPI_ISL_674502 to 674503, EPI_ISL_674505 to 674513, EPI_ISL_674515, EPI_ISL_674517 to 674521, EPI_ISL_674523, EPI_ISL_674525 to 674526, EPI_ISL_674529, EPI_ISL_674531 to 674540, EPI_ISL_674542, EPI_ISL_674546 to 674549, EPI_ISL_674551 to 674554, EPI_ISL_674556 to 674557, EPI_ISL_674560 to 674562, EPI_ISL_674564, EPI_ISL_674567 to 674568, EPI_ISL_674570 to 674583, EPI_ISL_674586 to 674588, EPI_ISL_674591, EPI_ISL_674593 to 674594, EPI_ISL_674596, EPI_ISL_674598 to 674602, EPI_ISL_674604, EPI_ISL_674606, EPI_ISL_674608, EPI_ISL_674610 to 674612, EPI_ISL_674614 to 674615, EPI_ISL_674617 to 674624, EPI_ISL_674626 to 674628, EPI_ISL_674631 to 674634, EPI_ISL_674636, EPI_ISL_674638 to 674644, EPI_ISL_674646 to 674651, EPI_ISL_674654, EPI_ISL_674656, EPI_ISL_674659 to 674660, EPI_ISL_674663 to 674666, EPI_ISL_674668 to 674669, EPI_ISL_674671 to 674673, EPI_ISL_674675 to 674677, EPI_ISL_674679 to 674684, EPI_ISL_674686 to 674689, EPI_ISL_674693 to 674697, EPI_ISL_674699 to 674706, EPI_ISL_674708 to 674710, EPI_ISL_674712, EPI_ISL_674715, EPI_ISL_674719 to 674721, EPI_ISL_674726, EPI_ISL_674728 to 674735, EPI_ISL_674737 to 674739, EPI_ISL_674742, EPI_ISL_674748, EPI_ISL_674750, EPI_ISL_674753 to 674755, EPI_ISL_674757, EPI_ISL_674759, EPI_ISL_674761, EPI_ISL_674764 to 674766, EPI_ISL_674768 to 674769, EPI_ISL_674771, EPI_ISL_674774, EPI_ISL_674776 to 674778, EPI_ISL_674780, EPI_ISL_674784 to 674785, EPI_ISL_674787 to 674790, EPI_ISL_674793, EPI_ISL_674796 to 674802, EPI_ISL_674805 to 674806, EPI_ISL_674810 to 674825, EPI_ISL_674827, EPI_ISL_674829 to 674833, EPI_ISL_674835 to 674836, EPI_ISL_674839, EPI_ISL_674841 to 674844, EPI_ISL_674848, EPI_ISL_674853 to 674858, EPI_ISL_674860 to 674861, EPI_ISL_674863 to 674865, EPI_ISL_674869 to 674870, EPI_ISL_674872, EPI_ISL_674874 to 674875, EPI_ISL_674877 to 674881, EPI_ISL_674884 to 674888, EPI_ISL_674890, EPI_ISL_674892, EPI_ISL_674894 to 674895, EPI_ISL_674897 to 674898, EPI_ISL_674900, EPI_ISL_674903 to 674905, EPI_ISL_674907, EPI_ISL_674911 to 674912, EPI_ISL_674914 to 674916, EPI_ISL_674918, EPI_ISL_674921, EPI_ISL_674927 to 674932, EPI_ISL_674934 to 674935, EPI_ISL_674938, EPI_ISL_674941, EPI_ISL_674943, EPI_ISL_674945, EPI_ISL_674947 to 674948, EPI_ISL_674954 to 674956, EPI_ISL_674958, EPI_ISL_674960 to 674961, EPI_ISL_674963 to 674971, EPI_ISL_674974 to 674976, EPI_ISL_674978 to 674979, EPI_ISL_674981 to 674982, EPI_ISL_674985 to 674987, EPI_ISL_674989 to 674991, EPI_ISL_674993 to 674994, EPI_ISL_674997, EPI_ISL_674999 to 675002, EPI_ISL_675004 to 675005, EPI_ISL_675007, EPI_ISL_675010 to 675011, EPI_ISL_675013 to 675015, EPI_ISL_675017, EPI_ISL_675020 to 675023, EPI_ISL_675025, EPI_ISL_675028, EPI_ISL_675031 to 675032, EPI_ISL_675035 to 675037, EPI_ISL_675041, EPI_ISL_675043 to 675045, EPI_ISL_675047 to 675052, EPI_ISL_675054 to 675055, EPI_ISL_675057 to 675058, EPI_ISL_675060 to 675063, EPI_ISL_675065 to 675070, EPI_ISL_675072, EPI_ISL_675074, EPI_ISL_675076 to 675079, EPI_ISL_675081, EPI_ISL_675083 to 675086, EPI_ISL_675088 to 675093, EPI_ISL_675095 to 675103, EPI_ISL_675105 to 675106, EPI_ISL_675109, EPI_ISL_675111, EPI_ISL_675113, EPI_ISL_675115 to 675116, EPI_ISL_675119, EPI_ISL_675121 to 675123, EPI_ISL_675125, EPI_ISL_675127 to 675129, EPI_ISL_675131 to 675132, EPI_ISL_675134, EPI_ISL_675145, EPI_ISL_675160, EPI_ISL_675170, EPI_ISL_675178 to 675179, EPI_ISL_675183, EPI_ISL_675185, EPI_ISL_675210, EPI_ISL_675212, EPI_ISL_675217, EPI_ISL_675219, EPI_ISL_675224, EPI_ISL_675227 to 675228, EPI_ISL_675238, EPI_ISL_675243, EPI_ISL_675260, EPI_ISL_675264, EPI_ISL_675275 to 675277, EPI_ISL_675282, EPI_ISL_675284 to 675286, EPI_ISL_675292, EPI_ISL_675301, EPI_ISL_675305 to 675309, EPI_ISL_675311, EPI_ISL_675313, EPI_ISL_675317 to 675318, EPI_ISL_675320, EPI_ISL_675326, EPI_ISL_675331 to 675332, EPI_ISL_675340, EPI_ISL_675353, EPI_ISL_675357, EPI_ISL_675364, EPI_ISL_675368, EPI_ISL_675371, EPI_ISL_675386, EPI_ISL_675393, EPI_ISL_675397, EPI_ISL_675399, EPI_ISL_675404, EPI_ISL_675417, EPI_ISL_675420, EPI_ISL_675423 to 675424, EPI_ISL_675432, EPI_ISL_675436, EPI_ISL_675439, EPI_ISL_675443 to 675444, EPI_ISL_675446, EPI_ISL_675467, EPI_ISL_675472, EPI_ISL_675475, EPI_ISL_675480, EPI_ISL_675486, EPI_ISL_675495 to 675496, EPI_ISL_675499 to 675500, EPI_ISL_675503 to 675506, EPI_ISL_675513, EPI_ISL_675518, EPI_ISL_675528, EPI_ISL_675538, EPI_ISL_675542, EPI_ISL_675550, EPI_ISL_675555 to 675556, EPI_ISL_675558, EPI_ISL_675560, EPI_ISL_675564 to 675566, EPI_ISL_675572, EPI_ISL_675576, EPI_ISL_675578, EPI_ISL_675601 to 675602, EPI_ISL_675606, EPI_ISL_675621, EPI_ISL_675631, EPI_ISL_675634, EPI_ISL_675644, EPI_ISL_675652, EPI_ISL_675660 to 675661, EPI_ISL_675663, EPI_ISL_675667, EPI_ISL_675676, EPI_ISL_675682, EPI_ISL_675693 to 675694, EPI_ISL_675698, EPI_ISL_675704, EPI_ISL_675709, EPI_ISL_675711 to 675714, EPI_ISL_675717 to 675718, EPI_ISL_675722 to 675723, EPI_ISL_675725 to 675726, EPI_ISL_675729 to 675732, EPI_ISL_675734, EPI_ISL_675736 to 675738, EPI_ISL_675741 to 675745, EPI_ISL_675747 to 675749, EPI_ISL_675752 to 675755, EPI_ISL_675758, EPI_ISL_675761 to 675765, EPI_ISL_675768 to 675769, EPI_ISL_675772 to 675773, EPI_ISL_675778, EPI_ISL_675781 to 675784, EPI_ISL_675786 to 675787, EPI_ISL_675789 to 675790, EPI_ISL_675792 to 675793, EPI_ISL_675795 to 675797, EPI_ISL_675801, EPI_ISL_675804 to 675805, EPI_ISL_675807 to 675809, EPI_ISL_675811, EPI_ISL_675814 to 675816, EPI_ISL_675819, EPI_ISL_675821, EPI_ISL_675824, EPI_ISL_675827 to 675828, EPI_ISL_675830, EPI_ISL_675834 to 675838, EPI_ISL_675840 to 675841, EPI_ISL_675843 to 675846, EPI_ISL_675849 to 675851, EPI_ISL_675853 to 675854, EPI_ISL_675856 to 675867, EPI_ISL_675869 to 675877, EPI_ISL_675879, EPI_ISL_675881, EPI_ISL_675884 to 675889, EPI_ISL_675891, EPI_ISL_675893, EPI_ISL_675896, EPI_ISL_675898 to 675899, EPI_ISL_675904 to 675905, EPI_ISL_675910 to 675912, EPI_ISL_675914 to 675915, EPI_ISL_675917 to 675919, EPI_ISL_675921 to 675922, EPI_ISL_675926 to 675928, EPI_ISL_675932 to 675933, EPI_ISL_675935 to 675936, EPI_ISL_675938, EPI_ISL_675940 to 675941, EPI_ISL_675943, EPI_ISL_675946 to 675950, EPI_ISL_675953, EPI_ISL_675955, EPI_ISL_675958 to 675961, EPI_ISL_675963 to 675964, EPI_ISL_675966 to 675967, EPI_ISL_675969 to 675972, EPI_ISL_675975, EPI_ISL_675977, EPI_ISL_675981, EPI_ISL_675983 to 675985, EPI_ISL_675989, EPI_ISL_675991 to 675992, EPI_ISL_675994 to 675999, EPI_ISL_676001, EPI_ISL_676003 to 676005, EPI_ISL_676007 to 676009, EPI_ISL_676011 to 676025, EPI_ISL_676027 to 676045, EPI_ISL_676047 to 676054, EPI_ISL_676056 to 676063, EPI_ISL_676066 to 676070, EPI_ISL_676072 to 676079, EPI_ISL_676081 to 676090, EPI_ISL_676092, EPI_ISL_676094 to 676095, EPI_ISL_676097, EPI_ISL_676099 to 676100, EPI_ISL_676102 to 676103, EPI_ISL_676105 to 676111, EPI_ISL_676113 to 676142, EPI_ISL_676144, EPI_ISL_676146 to 676150, EPI_ISL_676152 to 676154, EPI_ISL_676156 to 676157, EPI_ISL_676159 to 676163, EPI_ISL_676165 to 676168, EPI_ISL_676170 to 676178, EPI_ISL_676180 to 676182, EPI_ISL_676185 to 676193, EPI_ISL_676195 to 676201, EPI_ISL_676203 to 676204, EPI_ISL_676206 to 676210, EPI_ISL_676212 to 676219, EPI_ISL_676222 to 676224, EPI_ISL_676226 to 676250, EPI_ISL_676252, EPI_ISL_676254 to 676262, EPI_ISL_676264 to 676266, EPI_ISL_676268 to 676271, EPI_ISL_676273 to 676276, EPI_ISL_676278 to 676281, EPI_ISL_676283 to 676285, EPI_ISL_676287 to 676306, EPI_ISL_676308 to 676312, EPI_ISL_681325 to 681344, EPI_ISL_681346 to 681409, EPI_ISL_681411 to 681412, EPI_ISL_681414 to 681420, EPI_ISL_681422 to 681424, EPI_ISL_681426 to 681434, EPI_ISL_681436 to 681438, EPI_ISL_681440 to 681457, EPI_ISL_681459 to 681465, EPI_ISL_681467 to 681505, EPI_ISL_681507 to 681512, EPI_ISL_681514 to 681532, EPI_ISL_681534 to 681546, EPI_ISL_681548 to 681566, EPI_ISL_681569 to 681584, EPI_ISL_681586 to 681595, EPI_ISL_681597 to 681598, EPI_ISL_681600 to 681619, EPI_ISL_681621 to 681631, EPI_ISL_681633 to 681642, EPI_ISL_681644 to 681655, EPI_ISL_681660, EPI_ISL_681666 to 681668, EPI_ISL_705191, EPI_ISL_705196 to 705197, EPI_ISL_705199 to 705200, EPI_ISL_705202, EPI_ISL_708844, EPI_ISL_708848 to 708850, EPI_ISL_708856 to 708858, EPI_ISL_708862, EPI_ISL_708869, EPI_ISL_708871, EPI_ISL_708879, EPI_ISL_708881, EPI_ISL_708889, EPI_ISL_708896 to 708898, EPI_ISL_708900 to 708901, EPI_ISL_708908, EPI_ISL_708913, EPI_ISL_708916, EPI_ISL_708920, EPI_ISL_708928, EPI_ISL_708945, EPI_ISL_708958, EPI_ISL_708963, EPI_ISL_708969, EPI_ISL_708975, EPI_ISL_708979, EPI_ISL_708983 to 708984, EPI_ISL_708986, EPI_ISL_709567 to 709580, EPI_ISL_709582 to 709586, EPI_ISL_709588 to 709600, EPI_ISL_709602 to 709603, EPI_ISL_709605 to 709610, EPI_ISL_709613 to 709621, EPI_ISL_709623 to 709626, EPI_ISL_709629 to 709634, EPI_ISL_709636 to 709666, EPI_ISL_709668 to 709669, EPI_ISL_709671 to 709681, EPI_ISL_709683 to 709684, EPI_ISL_709686 to 709691, EPI_ISL_709693 to 709698, EPI_ISL_709700 to 709706, EPI_ISL_709709 to 709711, EPI_ISL_709713 to 709714, EPI_ISL_709716 to 709721, EPI_ISL_709723 to 709731, EPI_ISL_709733 to 709735, EPI_ISL_709737 to 709739, EPI_ISL_709741 to 709745, EPI_ISL_709747 to 709771, EPI_ISL_709774 to 709775, EPI_ISL_709777, EPI_ISL_709779 to 709799, EPI_ISL_709801 to 709803, EPI_ISL_709815, EPI_ISL_709819, EPI_ISL_709823 to 709824, EPI_ISL_709828, EPI_ISL_709830, EPI_ISL_709833, EPI_ISL_709838 to 709839, EPI_ISL_709841, EPI_ISL_709847, EPI_ISL_709852, EPI_ISL_709856, EPI_ISL_709860, EPI_ISL_709863, EPI_ISL_709866, EPI_ISL_709868, EPI_ISL_709871 to 709872, EPI_ISL_709874, EPI_ISL_709879, EPI_ISL_709881 to 709889, EPI_ISL_709891 to 709893, EPI_ISL_709895 to 709896, EPI_ISL_709898 to 709928, EPI_ISL_709930 to 709936, EPI_ISL_709938 to 709947, EPI_ISL_710907, EPI_ISL_719988, EPI_ISL_719991 to 719999, EPI_ISL_720001 to 720014, EPI_ISL_720016 to 720024, EPI_ISL_720026 to 720027, EPI_ISL_720029 to 720030, EPI_ISL_720033 to 720039, EPI_ISL_720041 to 720044, EPI_ISL_720046 to 720056, EPI_ISL_720058 to 720074, EPI_ISL_720077 to 720090, EPI_ISL_720092 to 720094, EPI_ISL_720097 to 720099, EPI_ISL_720101 to 720112, EPI_ISL_720114 to 720137, EPI_ISL_720140 to 720145, EPI_ISL_720147 to 720150, EPI_ISL_720152 to 720162, EPI_ISL_720164 to 720171, EPI_ISL_720173 to 720174, EPI_ISL_720176 to 720180, EPI_ISL_720183 to 720189, EPI_ISL_720191 to 720198, EPI_ISL_720200 to 720218, EPI_ISL_720220 to 720223, EPI_ISL_720225 to 720227, EPI_ISL_720229 to 720244, EPI_ISL_720246 to 720266, EPI_ISL_720268 to 720272, EPI_ISL_720274 to 720283, EPI_ISL_720285 to 720287, EPI_ISL_720289 to 720290, EPI_ISL_720292, EPI_ISL_720294 to 720297, EPI_ISL_720299 to 720318, EPI_ISL_720320 to 720328, EPI_ISL_720330 to 720331, EPI_ISL_720334 to 720344, EPI_ISL_720346 to 720357, EPI_ISL_720359 to 720369, EPI_ISL_720371 to 720372, EPI_ISL_720374 to 720401, EPI_ISL_720403 to 720409, EPI_ISL_720411 to 720413, EPI_ISL_720415 to 720418, EPI_ISL_720420 to 720448, EPI_ISL_720450 to 720493, EPI_ISL_720496 to 720501, EPI_ISL_720503 to 720504, EPI_ISL_720506 to 720507, EPI_ISL_720510 to 720528, EPI_ISL_720530 to 720531, EPI_ISL_720533 to 720535, EPI_ISL_720537 to 720542, EPI_ISL_720544 to 720565, EPI_ISL_720567 to 720618, EPI_ISL_720620 to 720628, EPI_ISL_720630, EPI_ISL_720632 to 720661, EPI_ISL_720663, EPI_ISL_720665 to 720667, EPI_ISL_720669, EPI_ISL_720671, EPI_ISL_720673 to 720676, EPI_ISL_720678 to 720682, EPI_ISL_720687, EPI_ISL_720689, EPI_ISL_720691 to 720693, EPI_ISL_720696, EPI_ISL_720698 to 720699, EPI_ISL_720701, EPI_ISL_720705 to 720708, EPI_ISL_720710, EPI_ISL_720712, EPI_ISL_720714, EPI_ISL_720716 to 720721, EPI_ISL_720723 to 720725, EPI_ISL_720727, EPI_ISL_720729 to 720731, EPI_ISL_720736 to 720737, EPI_ISL_720739, EPI_ISL_720742 to 720743, EPI_ISL_720746 to 720748, EPI_ISL_720751, EPI_ISL_720753, EPI_ISL_720757 to 720760, EPI_ISL_720763, EPI_ISL_720766, EPI_ISL_720768 to 720769, EPI_ISL_720771 to 720772, EPI_ISL_720774 to 720775, EPI_ISL_720777 to 720778, EPI_ISL_720781, EPI_ISL_720783 to 720785, EPI_ISL_720789, EPI_ISL_720791, EPI_ISL_720793 to 720795, EPI_ISL_720797, EPI_ISL_720799, EPI_ISL_720801, EPI_ISL_720803 to 720804, EPI_ISL_720806 to 720809, EPI_ISL_720812, EPI_ISL_720817 to 720818, EPI_ISL_720820, EPI_ISL_720822, EPI_ISL_720824, EPI_ISL_720826, EPI_ISL_720828 to 720830, EPI_ISL_720832 to 720833, EPI_ISL_720835, EPI_ISL_720838 to 720840, EPI_ISL_720843 to 720844, EPI_ISL_720847, EPI_ISL_720851 to 720852, EPI_ISL_720854 to 720855, EPI_ISL_720859 to 720863, EPI_ISL_720866, EPI_ISL_720868, EPI_ISL_720871, EPI_ISL_720873 to 720874, EPI_ISL_720876, EPI_ISL_720878 to 720879, EPI_ISL_720882 to 720884, EPI_ISL_720887 to 720888, EPI_ISL_720890, EPI_ISL_720892, EPI_ISL_720895 to 720896, EPI_ISL_720899, EPI_ISL_720901 to 720903, EPI_ISL_720906, EPI_ISL_720908, EPI_ISL_721526, EPI_ISL_721529 to 721532, EPI_ISL_731251 to 731253, EPI_ISL_731892 to 731893, EPI_ISL_733607, EPI_ISL_733617, EPI_ISL_733628, EPI_ISL_733636, EPI_ISL_733644, EPI_ISL_733648, EPI_ISL_733652, EPI_ISL_733657 to 733658, EPI_ISL_733664, EPI_ISL_733666, EPI_ISL_733669, EPI_ISL_733673, EPI_ISL_733678, EPI_ISL_733687, EPI_ISL_733707, EPI_ISL_733711, EPI_ISL_733713, EPI_ISL_733735, EPI_ISL_733739, EPI_ISL_733746 to 733747, EPI_ISL_733749, EPI_ISL_733757, EPI_ISL_733760 to 733761, EPI_ISL_733764, EPI_ISL_733768, EPI_ISL_733771, EPI_ISL_733776, EPI_ISL_733781, EPI_ISL_733796, EPI_ISL_733801, EPI_ISL_733811, EPI_ISL_733813, EPI_ISL_733820, EPI_ISL_733825 to 733826, EPI_ISL_733828 to 733829, EPI_ISL_733832, EPI_ISL_733838, EPI_ISL_733842, EPI_ISL_733845, EPI_ISL_733851, EPI_ISL_733855, EPI_ISL_733858, EPI_ISL_733861, EPI_ISL_733863 to 733865, EPI_ISL_733872 to 733873, EPI_ISL_733884, EPI_ISL_733889, EPI_ISL_733891, EPI_ISL_733894, EPI_ISL_733900, EPI_ISL_733902 to 733905, EPI_ISL_733914, EPI_ISL_735512 to 735514, EPI_ISL_735516 to 735522, EPI_ISL_735524 to 735529, EPI_ISL_735531 to 735533, EPI_ISL_735535 to 735540, EPI_ISL_735542 to 735543, EPI_ISL_735545, EPI_ISL_735547 to 735556, EPI_ISL_735558 to 735563, EPI_ISL_735565 to 735577, EPI_ISL_735579 to 735587, EPI_ISL_735589 to 735598, EPI_ISL_735601 to 735604, EPI_ISL_735606 to 735616, EPI_ISL_735618 to 735631, EPI_ISL_735633 to 735645, EPI_ISL_735647 to 735652, EPI_ISL_735654 to 735656, EPI_ISL_735658 to 735674, EPI_ISL_735677 to 735711, EPI_ISL_735713 to 735722, EPI_ISL_735724 to 735732, EPI_ISL_735734 to 735742, EPI_ISL_735744 to 735745, EPI_ISL_735747 to 735750, EPI_ISL_735752 to 735755, EPI_ISL_735757 to 735758, EPI_ISL_735760 to 735772, EPI_ISL_735774 to 735790, EPI_ISL_735792 to 735793, EPI_ISL_735795 to 735822, EPI_ISL_735824 to 735829, EPI_ISL_735831 to 735835, EPI_ISL_735837 to 735859, EPI_ISL_736201 to 736203, EPI_ISL_736207 to 736208, EPI_ISL_736210 to 736211, EPI_ISL_736213 to 736219, EPI_ISL_736224 to 736228, EPI_ISL_736230, EPI_ISL_736232 to 736233, EPI_ISL_736240, EPI_ISL_736244, EPI_ISL_736249, EPI_ISL_736251 to 736252, EPI_ISL_736256, EPI_ISL_736260, EPI_ISL_736262 to 736265, EPI_ISL_736267 to 736268, EPI_ISL_736270, EPI_ISL_736272, EPI_ISL_736274, EPI_ISL_736276, EPI_ISL_736279, EPI_ISL_736281 to 736283, EPI_ISL_736285 to 736286, EPI_ISL_736288 to 736289, EPI_ISL_736291 to 736300, EPI_ISL_736302, EPI_ISL_736305, EPI_ISL_736307 to 736308, EPI_ISL_736310, EPI_ISL_736314, EPI_ISL_736317, EPI_ISL_736319, EPI_ISL_736322, EPI_ISL_736324, EPI_ISL_736326, EPI_ISL_736328 to 736329, EPI_ISL_736333, EPI_ISL_736335 to 736342, EPI_ISL_736344 to 736345, EPI_ISL_736348 to 736349, EPI_ISL_736353 to 736354, EPI_ISL_736357, EPI_ISL_736359 to 736361, EPI_ISL_736363 to 736366, EPI_ISL_736368 to 736369, EPI_ISL_736372, EPI_ISL_736374 to 736375, EPI_ISL_736377, EPI_ISL_736379 to 736381, EPI_ISL_736384 to 736385, EPI_ISL_736387, EPI_ISL_736392 to 736393, EPI_ISL_736395 to 736397, EPI_ISL_736401 to 736403,

EPI_ISL_736405 to 736410, EPI_ISL_736413, EPI_ISL_736415, EPI_ISL_736418, EPI_ISL_736420 to 736422, EPI_ISL_736424, EPI_ISL_736427, EPI_ISL_736432 to 736436, EPI_ISL_736439 to 736441, EPI_ISL_736443, EPI_ISL_736446 to 736450, EPI_ISL_736452 to 736454, EPI_ISL_736457, EPI_ISL_736462 to 736463, EPI_ISL_736465, EPI_ISL_736467, EPI_ISL_736469, EPI_ISL_736472 to 736473, EPI_ISL_736475, EPI_ISL_736480, EPI_ISL_736487, EPI_ISL_736497, EPI_ISL_736501 to 736502, EPI_ISL_736504, EPI_ISL_736508, EPI_ISL_736514 to 736515, EPI_ISL_736523, EPI_ISL_736525, EPI_ISL_736527 to 736528, EPI_ISL_736536, EPI_ISL_736538, EPI_ISL_736542, EPI_ISL_736547, EPI_ISL_736549 to 736552, EPI_ISL_736558, EPI_ISL_736563, EPI_ISL_736565, EPI_ISL_736571, EPI_ISL_736578, EPI_ISL_736586, EPI_ISL_736588, EPI_ISL_736590, EPI_ISL_736593 to 736594, EPI_ISL_736597 to 736598, EPI_ISL_736601 to 736602, EPI_ISL_736608, EPI_ISL_736610, EPI_ISL_736617 to 736618, EPI_ISL_736622, EPI_ISL_736630, EPI_ISL_736637, EPI_ISL_736643, EPI_ISL_736648, EPI_ISL_736675, EPI_ISL_736677, EPI_ISL_736681 to 736683, EPI_ISL_736686, EPI_ISL_736693 to 736694, EPI_ISL_736696 to 736697, EPI_ISL_736703, EPI_ISL_736712 to 736713, EPI_ISL_736716, EPI_ISL_736719, EPI_ISL_736723 to 736725, EPI_ISL_736727 to 736728, EPI_ISL_736731, EPI_ISL_736733, EPI_ISL_736738, EPI_ISL_736740, EPI_ISL_736744, EPI_ISL_736746, EPI_ISL_736749, EPI_ISL_736757, EPI_ISL_756377, EPI_ISL_756689, EPI_ISL_756691 to 756694, EPI_ISL_756698 to 756701, EPI_ISL_756703 to 756704, EPI_ISL_756707 to 756708, EPI_ISL_756710, EPI_ISL_756712 to 756715, EPI_ISL_756718 to 756719, EPI_ISL_756721 to 756722, EPI_ISL_756724 to 756728, EPI_ISL_756730 to 756736, EPI_ISL_756738 to 756743, EPI_ISL_756746 to 756747, EPI_ISL_756749, EPI_ISL_756751 to 756760, EPI_ISL_756762 to 756771, EPI_ISL_756774, EPI_ISL_756776 to 756779, EPI_ISL_756781 to 756783, EPI_ISL_756785 to 756788, EPI_ISL_756790 to 756792, EPI_ISL_756794, EPI_ISL_756796 to 756799, EPI_ISL_756801 to 756802, EPI_ISL_756804, EPI_ISL_756806 to 756819, EPI_ISL_756821 to 756823, EPI_ISL_756825 to 756834, EPI_ISL_756836 to 756839, EPI_ISL_756841 to 756848, EPI_ISL_756851, EPI_ISL_756853 to 756855, EPI_ISL_756857 to 756860, EPI_ISL_756862 to 756866, EPI_ISL_756868 to 756876, EPI_ISL_756880, EPI_ISL_756882 to 756884, EPI_ISL_756887, EPI_ISL_756890 to 756892, EPI_ISL_756896, EPI_ISL_756898 to 756899, EPI_ISL_756901 to 756905, EPI_ISL_756908, EPI_ISL_756911, EPI_ISL_756913 to 756917, EPI_ISL_756919, EPI_ISL_756921 to 756922, EPI_ISL_756924 to 756927, EPI_ISL_756929 to 756930, EPI_ISL_756932, EPI_ISL_756934, EPI_ISL_756936 to 756939, EPI_ISL_756941 to 756946, EPI_ISL_756948 to 756950, EPI_ISL_756954, EPI_ISL_756956 to 756958, EPI_ISL_756961, EPI_ISL_756963 to 756968, EPI_ISL_756972, EPI_ISL_756975 to 756976, EPI_ISL_756979 to 756981, EPI_ISL_756983 to 756988, EPI_ISL_756990 to 756994, EPI_ISL_756996 to 756997, EPI_ISL_756999 to 757004, EPI_ISL_757006, EPI_ISL_757010, EPI_ISL_757013 to 757016, EPI_ISL_757019, EPI_ISL_757022 to 757026, EPI_ISL_757029 to 757030, EPI_ISL_760698, EPI_ISL_760801, EPI_ISL_760809 to 760810, EPI_ISL_760842, EPI_ISL_760844, EPI_ISL_760863, EPI_ISL_760871, EPI_ISL_760873, EPI_ISL_760881, EPI_ISL_760896, EPI_ISL_760904 to 760905, EPI_ISL_760943, EPI_ISL_760954, EPI_ISL_760959, EPI_ISL_760961, EPI_ISL_760963, EPI_ISL_760975, EPI_ISL_760983, EPI_ISL_760987, EPI_ISL_761002, EPI_ISL_761023, EPI_ISL_761039, EPI_ISL_761050, EPI_ISL_761059, EPI_ISL_761079, EPI_ISL_761357 to 761371, EPI_ISL_761373 to 761379, EPI_ISL_761382 to 761397, EPI_ISL_761399 to 761425, EPI_ISL_761427 to 761437, EPI_ISL_761439 to 761447, EPI_ISL_761449 to 761457, EPI_ISL_761459 to 761480, EPI_ISL_761482 to 761484, EPI_ISL_761486 to 761504, EPI_ISL_761506 to 761535, EPI_ISL_761537 to 761538, EPI_ISL_761540 to 761551, EPI_ISL_761553 to 761556, EPI_ISL_761558 to 761561, EPI_ISL_761563 to 761600, EPI_ISL_761602 to 761605, EPI_ISL_761607 to 761624, EPI_ISL_761626 to 761632, EPI_ISL_761634 to 761647, EPI_ISL_761650 to 761664, EPI_ISL_762199 to 762284, EPI_ISL_762286 to 762287, EPI_ISL_762289 to 762307, EPI_ISL_762309 to 762321, EPI_ISL_762323 to 762378, EPI_ISL_762380, EPI_ISL_762382 to 762386, EPI_ISL_762388 to 762419, EPI_ISL_762421 to 762424, EPI_ISL_762426 to 762427, EPI_ISL_762429 to 762448, EPI_ISL_762450 to 762457, EPI_ISL_762459 to 762465, EPI_ISL_762467, EPI_ISL_762469 to 762487, EPI_ISL_762489 to 762496, EPI_ISL_762498 to 762499, EPI_ISL_762501 to 762505, EPI_ISL_762507 to 762518, EPI_ISL_762520 to 762545, EPI_ISL_762864, EPI_ISL_762867, EPI_ISL_762880, EPI_ISL_762884, EPI_ISL_762891, EPI_ISL_762896, EPI_ISL_762903, EPI_ISL_762908, EPI_ISL_762912, EPI_ISL_762914, EPI_ISL_762920, EPI_ISL_762937, EPI_ISL_762943, EPI_ISL_762945, EPI_ISL_762963, EPI_ISL_762967 to 762968, EPI_ISL_762976, EPI_ISL_762978, EPI_ISL_762981, EPI_ISL_767090, EPI_ISL_767098 to 767101, EPI_ISL_767107 to 767109, EPI_ISL_767111 to 767112, EPI_ISL_767134, EPI_ISL_767239, EPI_ISL_768847 to 768850, EPI_ISL_768852 to 768858, EPI_ISL_768860 to 768889, EPI_ISL_768891 to 768897, EPI_ISL_769226 to 769228, EPI_ISL_769230 to 769232, EPI_ISL_769234 to 769235, EPI_ISL_769237 to 769242, EPI_ISL_769247 to 769248, EPI_ISL_769251, EPI_ISL_769253, EPI_ISL_769258, EPI_ISL_769262 to 769265, EPI_ISL_769268, EPI_ISL_769270, EPI_ISL_769272, EPI_ISL_769274, EPI_ISL_769277, EPI_ISL_769279, EPI_ISL_769283, EPI_ISL_769285, EPI_ISL_769287, EPI_ISL_769293, EPI_ISL_769295 to 769296, EPI_ISL_769298 to 769305, EPI_ISL_769307, EPI_ISL_769309, EPI_ISL_769311, EPI_ISL_769313, EPI_ISL_769315, EPI_ISL_769317 to 769318, EPI_ISL_769324, EPI_ISL_769326, EPI_ISL_769331 to 769333, EPI_ISL_769337 to 769340, EPI_ISL_769342, EPI_ISL_769344, EPI_ISL_769346 to 769347, EPI_ISL_769350 to 769351, EPI_ISL_769354, EPI_ISL_769356 to 769357, EPI_ISL_769359, EPI_ISL_769362 to 769364, EPI_ISL_769366, EPI_ISL_769368 to 769369, EPI_ISL_769371 to 769373, EPI_ISL_769375 to 769379, EPI_ISL_769382, EPI_ISL_769386, EPI_ISL_769393, EPI_ISL_769395, EPI_ISL_769397 to 769398, EPI_ISL_769400, EPI_ISL_769403, EPI_ISL_769407 to 769411, EPI_ISL_769414 to 769415, EPI_ISL_769423 to 769426, EPI_ISL_769429, EPI_ISL_769432 to 769433, EPI_ISL_769435 to 769439, EPI_ISL_769443 to 769444, EPI_ISL_769447 to 769448, EPI_ISL_769451, EPI_ISL_769453, EPI_ISL_769455 to 769456, EPI_ISL_769461, EPI_ISL_769463 to 769464, EPI_ISL_769467, EPI_ISL_769472, EPI_ISL_769477 to 769480, EPI_ISL_769483 to 769484, EPI_ISL_769486 to 769488, EPI_ISL_769490 to 769493, EPI_ISL_769495, EPI_ISL_769505, EPI_ISL_769507 to 769510, EPI_ISL_769514, EPI_ISL_769516, EPI_ISL_769520 to 769521, EPI_ISL_769523, EPI_ISL_769526 to 769527, EPI_ISL_769529, EPI_ISL_769531, EPI_ISL_769534, EPI_ISL_769536 to 769538, EPI_ISL_769541, EPI_ISL_769544 to 769550, EPI_ISL_769552 to 769555, EPI_ISL_769557 to 769566, EPI_ISL_769569 to 769576, EPI_ISL_769578 to 769582, EPI_ISL_769584 to 769588, EPI_ISL_769590 to 769593, EPI_ISL_769595 to 769605, EPI_ISL_769608, EPI_ISL_769610, EPI_ISL_769615 to 769616, EPI_ISL_769618, EPI_ISL_769620 to 769624, EPI_ISL_769626 to 769640, EPI_ISL_769642 to 769643, EPI_ISL_769645, EPI_ISL_769647, EPI_ISL_769649 to 769651, EPI_ISL_769653 to 769654, EPI_ISL_769656 to 769660, EPI_ISL_769663 to 769665, EPI_ISL_769668 to 769676, EPI_ISL_769678 to 769686, EPI_ISL_769688, EPI_ISL_769690, EPI_ISL_769692 to 769696, EPI_ISL_769698 to 769702, EPI_ISL_769704, EPI_ISL_769706 to 769710, EPI_ISL_769712, EPI_ISL_769715, EPI_ISL_769717 to 769719, EPI_ISL_769721 to 769723, EPI_ISL_769725 to 769734, EPI_ISL_769736 to 769740, EPI_ISL_769742, EPI_ISL_769744 to 769750, EPI_ISL_769754 to 769757, EPI_ISL_769760, EPI_ISL_769762 to 769767, EPI_ISL_769769 to 769772, EPI_ISL_769774 to 769786, EPI_ISL_769789 to 769793, EPI_ISL_769795 to 769801, EPI_ISL_769803 to 769805, EPI_ISL_769807 to 769812, EPI_ISL_769814, EPI_ISL_769817, EPI_ISL_769819 to 769825, EPI_ISL_769827 to 769833, EPI_ISL_769835, EPI_ISL_769838 to 769841, EPI_ISL_769843 to 769849, EPI_ISL_769851 to 769860, EPI_ISL_777039, EPI_ISL_777056, EPI_ISL_777064, EPI_ISL_777080, EPI_ISL_777084 to 777085, EPI_ISL_777102, EPI_ISL_777104, EPI_ISL_777108, EPI_ISL_777141, EPI_ISL_777158, EPI_ISL_777184, EPI_ISL_777187, EPI_ISL_777200, EPI_ISL_777207, EPI_ISL_777224, EPI_ISL_777229, EPI_ISL_777245, EPI_ISL_777269, EPI_ISL_777295, EPI_ISL_777304, EPI_ISL_777310, EPI_ISL_777313, EPI_ISL_778298, EPI_ISL_778304, EPI_ISL_778307 to 778308, EPI_ISL_778310, EPI_ISL_778312 to 778314, EPI_ISL_778316 to 778321, EPI_ISL_778326, EPI_ISL_778329, EPI_ISL_778339 to 778341, EPI_ISL_778343 to 778344, EPI_ISL_778350 to 778351, EPI_ISL_778354, EPI_ISL_778363, EPI_ISL_778366 to 778367, EPI_ISL_778369 to 778371, EPI_ISL_778379, EPI_ISL_778381, EPI_ISL_778383 to 778384, EPI_ISL_778386 to 778389, EPI_ISL_778391, EPI_ISL_778393 to 778395, EPI_ISL_778398, EPI_ISL_778402, EPI_ISL_778405 to 778406, EPI_ISL_778409, EPI_ISL_778413 to 778414, EPI_ISL_778418, EPI_ISL_778425, EPI_ISL_778427, EPI_ISL_778429, EPI_ISL_778431, EPI_ISL_778434 to 778436, EPI_ISL_778438 to 778439, EPI_ISL_778441 to 778443, EPI_ISL_778446, EPI_ISL_778449, EPI_ISL_778452 to 778453, EPI_ISL_778456, EPI_ISL_778458, EPI_ISL_778460, EPI_ISL_778464, EPI_ISL_778466 to 778470, EPI_ISL_778474 to 778477, EPI_ISL_778480 to 778481, EPI_ISL_778483, EPI_ISL_778485 to 778486, EPI_ISL_778489, EPI_ISL_778492 to 778493, EPI_ISL_778495, EPI_ISL_778501 to 778502, EPI_ISL_778505, EPI_ISL_778507 to 778509, EPI_ISL_778511, EPI_ISL_778513 to 778515, EPI_ISL_778518, EPI_ISL_778520 to 778521, EPI_ISL_778523, EPI_ISL_778526, EPI_ISL_778532, EPI_ISL_778535 to 778538, EPI_ISL_778542, EPI_ISL_778544 to 778546, EPI_ISL_778549 to 778550, EPI_ISL_778552, EPI_ISL_778554 to 778555, EPI_ISL_778557, EPI_ISL_778559, EPI_ISL_778561, EPI_ISL_778563, EPI_ISL_778565 to 778567, EPI_ISL_778572 to 778576, EPI_ISL_778580 to 778583, EPI_ISL_778585, EPI_ISL_778587 to 778589, EPI_ISL_778591, EPI_ISL_778595 to 778597, EPI_ISL_778600, EPI_ISL_778602 to 778604, EPI_ISL_778606, EPI_ISL_778608 to 778611, EPI_ISL_778613, EPI_ISL_778622, EPI_ISL_778624 to 778626, EPI_ISL_778632 to 778634, EPI_ISL_778636 to 778637, EPI_ISL_778639 to 778643, EPI_ISL_781180, EPI_ISL_781182, EPI_ISL_781184, EPI_ISL_781191, EPI_ISL_781193, EPI_ISL_781196 to 781197, EPI_ISL_781208, EPI_ISL_781211 to 781213, EPI_ISL_781215, EPI_ISL_781217 to 781218, EPI_ISL_781222, EPI_ISL_781227 to 781229, EPI_ISL_781231, EPI_ISL_781233, EPI_ISL_781236, EPI_ISL_781238 to 781239, EPI_ISL_781241, EPI_ISL_781243 to 781244, EPI_ISL_781246 to 781247, EPI_ISL_781252, EPI_ISL_781255, EPI_ISL_781257, EPI_ISL_781264 to 781268, EPI_ISL_781270 to 781272, EPI_ISL_781274 to 781275, EPI_ISL_781277, EPI_ISL_781283 to 781284, EPI_ISL_781286, EPI_ISL_781289 to 781290, EPI_ISL_781293, EPI_ISL_781296, EPI_ISL_781298, EPI_ISL_781300, EPI_ISL_781302 to 781303, EPI_ISL_781305, EPI_ISL_781309 to 781312, EPI_ISL_781314, EPI_ISL_781316 to 781317, EPI_ISL_781320 to 781322, EPI_ISL_781326, EPI_ISL_781329, EPI_ISL_781334 to 781335, EPI_ISL_781338 to 781339, EPI_ISL_781343, EPI_ISL_781346 to 781347, EPI_ISL_781350 to 781351, EPI_ISL_781353, EPI_ISL_781357, EPI_ISL_781361 to 781363, EPI_ISL_781365, EPI_ISL_781373, EPI_ISL_781380 to 781384, EPI_ISL_781386, EPI_ISL_781390 to 781391, EPI_ISL_781393 to 781395, EPI_ISL_781398, EPI_ISL_781404, EPI_ISL_781410, EPI_ISL_781414 to 781422, EPI_ISL_781425, EPI_ISL_781427 to 781430, EPI_ISL_781449 to 781450, EPI_ISL_781452 to 781454, EPI_ISL_781460 to 781462, EPI_ISL_781467 to 781468, EPI_ISL_781470, EPI_ISL_781476 to 781477, EPI_ISL_781479, EPI_ISL_781482, EPI_ISL_781484 to 781486, EPI_ISL_781491, EPI_ISL_781496, EPI_ISL_781501, EPI_ISL_781504, EPI_ISL_781507, EPI_ISL_781509, EPI_ISL_781511, EPI_ISL_781522, EPI_ISL_781527, EPI_ISL_781529, EPI_ISL_781536 to 781537, EPI_ISL_781539, EPI_ISL_781541, EPI_ISL_781544, EPI_ISL_781547 to 781548, EPI_ISL_781554 to 781556, EPI_ISL_781558, EPI_ISL_781567, EPI_ISL_781576, EPI_ISL_781578 to 781579, EPI_ISL_781582, EPI_ISL_781597 to 781599, EPI_ISL_781605, EPI_ISL_781609, EPI_ISL_781611, EPI_ISL_781615, EPI_ISL_781617 to 781619, EPI_ISL_781621, EPI_ISL_781623, EPI_ISL_781627, EPI_ISL_781632, EPI_ISL_781636, EPI_ISL_781649, EPI_ISL_781651 to 781652, EPI_ISL_781655, EPI_ISL_781663, EPI_ISL_781665, EPI_ISL_781669, EPI_ISL_781674, EPI_ISL_781677, EPI_ISL_781680 to 781681, EPI_ISL_781684, EPI_ISL_781686, EPI_ISL_781695, EPI_ISL_781699, EPI_ISL_781701, EPI_ISL_781720 to 781721, EPI_ISL_781724 to 781725, EPI_ISL_781730, EPI_ISL_781732, EPI_ISL_781735, EPI_ISL_781742 to 781743, EPI_ISL_781746 to 781747, EPI_ISL_781749, EPI_ISL_781753 to 781754, EPI_ISL_781763, EPI_ISL_781767, EPI_ISL_781770, EPI_ISL_781773, EPI_ISL_781777, EPI_ISL_781783 to 781785, EPI_ISL_781788 to 781789, EPI_ISL_781795, EPI_ISL_781798, EPI_ISL_781801, EPI_ISL_781807, EPI_ISL_781832, EPI_ISL_781834, EPI_ISL_781837, EPI_ISL_781839 to 781841, EPI_ISL_781851, EPI_ISL_781854, EPI_ISL_781858, EPI_ISL_781862, EPI_ISL_781864, EPI_ISL_781869, EPI_ISL_781873, EPI_ISL_781876, EPI_ISL_781878, EPI_ISL_781880, EPI_ISL_781885, EPI_ISL_781891, EPI_ISL_781897 to 781898, EPI_ISL_781903, EPI_ISL_781907 to 781909, EPI_ISL_781914 to 781916, EPI_ISL_781920 to 781921, EPI_ISL_781924 to 781925, EPI_ISL_781938, EPI_ISL_781940 to 781942, EPI_ISL_781944, EPI_ISL_781946 to 781948, EPI_ISL_781950, EPI_ISL_781952, EPI_ISL_781955, EPI_ISL_781968, EPI_ISL_781973, EPI_ISL_781977, EPI_ISL_781979, EPI_ISL_781984, EPI_ISL_781987 to 781988, EPI_ISL_781994, EPI_ISL_781998 to 782000, EPI_ISL_782009, EPI_ISL_782015, EPI_ISL_782017, EPI_ISL_782021, EPI_ISL_782026, EPI_ISL_782028, EPI_ISL_782030 to 782032, EPI_ISL_782035, EPI_ISL_782037, EPI_ISL_782040, EPI_ISL_782042, EPI_ISL_782044, EPI_ISL_782046, EPI_ISL_782049, EPI_ISL_782051 to 782052, EPI_ISL_782054 to 782055, EPI_ISL_782058, EPI_ISL_782061, EPI_ISL_782069, EPI_ISL_782073, EPI_ISL_782080 to 782081, EPI_ISL_782087 to 782088, EPI_ISL_782092 to 782094, EPI_ISL_782098, EPI_ISL_782103, EPI_ISL_782105 to 782106, EPI_ISL_782118, EPI_ISL_782125, EPI_ISL_782129 to 782131, EPI_ISL_782134, EPI_ISL_782136 to 782137, EPI_ISL_782143 to 782144, EPI_ISL_782147, EPI_ISL_782734, EPI_ISL_782736, EPI_ISL_782738, EPI_ISL_782740 to 782744, EPI_ISL_782749 to 782756, EPI_ISL_782760, EPI_ISL_782762, EPI_ISL_782765 to 782767, EPI_ISL_782770, EPI_ISL_782772 to 782773, EPI_ISL_782775, EPI_ISL_782778 to 782779, EPI_ISL_782781 to 782782, EPI_ISL_782789 to 782790, EPI_ISL_782793 to 782794, EPI_ISL_782798 to 782799, EPI_ISL_782802, EPI_ISL_782804, EPI_ISL_782807, EPI_ISL_782811 to 782812, EPI_ISL_782815, EPI_ISL_782819, EPI_ISL_782821, EPI_ISL_782823 to 782824, EPI_ISL_782826, EPI_ISL_782828 to 782829, EPI_ISL_782831, EPI_ISL_782833, EPI_ISL_782835 to 782837, EPI_ISL_782839, EPI_ISL_782845, EPI_ISL_782847 to 782848, EPI_ISL_782852, EPI_ISL_782858 to 782859, EPI_ISL_782861, EPI_ISL_782865, EPI_ISL_782870 to 782871, EPI_ISL_782874 to 782876, EPI_ISL_782879, EPI_ISL_782881, EPI_ISL_782884, EPI_ISL_782888, EPI_ISL_782891, EPI_ISL_782895 to 782896, EPI_ISL_782898, EPI_ISL_782900 to 782901, EPI_ISL_782914 to 782916, EPI_ISL_782921 to 782922, EPI_ISL_782927, EPI_ISL_782933 to 782935, EPI_ISL_782938 to 782940, EPI_ISL_782943 to 782944, EPI_ISL_782948 to 782951, EPI_ISL_782953, EPI_ISL_782955, EPI_ISL_782957 to 782958, EPI_ISL_782961 to 782962, EPI_ISL_782965, EPI_ISL_782968, EPI_ISL_782972 to 782973, EPI_ISL_782975, EPI_ISL_782978, EPI_ISL_782980 to 782982, EPI_ISL_782986, EPI_ISL_782989, EPI_ISL_782995, EPI_ISL_782999, EPI_ISL_783003 to 783006, EPI_ISL_783010 to 783011, EPI_ISL_783013, EPI_ISL_783016 to 783018, EPI_ISL_783020 to 783022, EPI_ISL_783024 to 783025, EPI_ISL_783027, EPI_ISL_783029, EPI_ISL_783035, EPI_ISL_783040 to 783041, EPI_ISL_783048 to 783050, EPI_ISL_783053 to 783054, EPI_ISL_783058 to 783060, EPI_ISL_783062 to 783063, EPI_ISL_783065, EPI_ISL_783067, EPI_ISL_783069, EPI_ISL_783071 to 783076, EPI_ISL_783078 to 783081, EPI_ISL_783083 to 783086, EPI_ISL_783088 to 783090, EPI_ISL_783092 to 783100, EPI_ISL_783103 to 783110, EPI_ISL_783114 to 783125, EPI_ISL_783127 to 783129, EPI_ISL_783131 to 783135, EPI_ISL_783137 to 783150, EPI_ISL_783152 to 783160, EPI_ISL_783162 to 783164, EPI_ISL_783166 to 783184, EPI_ISL_783186 to 783193, EPI_ISL_783195 to 783204, EPI_ISL_783206 to 783207, EPI_ISL_783210 to 783215, EPI_ISL_783217 to 783218, EPI_ISL_783220 to 783233, EPI_ISL_783235 to 783241, EPI_ISL_783243 to 783251, EPI_ISL_783253 to 783259, EPI_ISL_783262 to 783263, EPI_ISL_783265 to 783272, EPI_ISL_783275 to 783280, EPI_ISL_783282, EPI_ISL_783285, EPI_ISL_783287 to 783289, EPI_ISL_783291 to 783297, EPI_ISL_783299 to 783308, EPI_ISL_783310 to 783311, EPI_ISL_783314, EPI_ISL_783316 to 783317, EPI_ISL_783320 to 783322, EPI_ISL_783324, EPI_ISL_783326 to 783333, EPI_ISL_783335 to 783337, EPI_ISL_783339 to 783366, EPI_ISL_797118 to 797135, EPI_ISL_797138 to 797139, EPI_ISL_797141 to 797145, EPI_ISL_797147 to 797150, EPI_ISL_797154 to 797155, EPI_ISL_797158 to 797163, EPI_ISL_797166 to 797177, EPI_ISL_797180 to 797187, EPI_ISL_797189 to 797194, EPI_ISL_797197, EPI_ISL_797199 to 797203, EPI_ISL_797205 to 797214, EPI_ISL_797217 to 797223, EPI_ISL_797225 to 797240, EPI_ISL_797242 to 797258, EPI_ISL_797260 to 797270, EPI_ISL_797273 to 797279, EPI_ISL_797283 to 797285, EPI_ISL_797287, EPI_ISL_797289 to 797298, EPI_ISL_797300 to 797304, EPI_ISL_797306 to 797308, EPI_ISL_797310 to 797311, EPI_ISL_797313 to 797318, EPI_ISL_797321 to 797324, EPI_ISL_797326, EPI_ISL_797328 to 797331, EPI_ISL_797333 to 797337, EPI_ISL_797339 to 797346, EPI_ISL_797348 to 797349, EPI_ISL_797352 to 797353, EPI_ISL_797355 to 797359, EPI_ISL_797361, EPI_ISL_797363 to 797365, EPI_ISL_797367 to 797376, EPI_ISL_797378 to 797388, EPI_ISL_797390 to 797396, EPI_ISL_798333, EPI_ISL_798335, EPI_ISL_798338 to 798340, EPI_ISL_798342 to 798344, EPI_ISL_798348, EPI_ISL_798351 to 798355, EPI_ISL_798357 to 798359, EPI_ISL_798361, EPI_ISL_798363 to 798367, EPI_ISL_798373 to 798377, EPI_ISL_798380 to 798383, EPI_ISL_798385, EPI_ISL_798387, EPI_ISL_798390 to 798395, EPI_ISL_798397 to 798398, EPI_ISL_798400, EPI_ISL_798402 to 798403, EPI_ISL_798406, EPI_ISL_798408 to 798412, EPI_ISL_798414 to 798415, EPI_ISL_798418 to 798423, EPI_ISL_798425 to 798429, EPI_ISL_798432 to 798435, EPI_ISL_798437, EPI_ISL_798439 to 798441, EPI_ISL_798443, EPI_ISL_798447 to 798454, EPI_ISL_798456, EPI_ISL_798458 to 798464, EPI_ISL_798466, EPI_ISL_798470, EPI_ISL_798472 to 798474, EPI_ISL_798476 to 798478, EPI_ISL_798480 to 798482, EPI_ISL_798484 to 798486, EPI_ISL_798488 to 798491, EPI_ISL_798494 to 798495, EPI_ISL_798497, EPI_ISL_798499, EPI_ISL_798501, EPI_ISL_798503 to 798507, EPI_ISL_798509, EPI_ISL_798512, EPI_ISL_798514 to 798515, EPI_ISL_798518, EPI_ISL_798521 to 798523, EPI_ISL_798525 to 798530, EPI_ISL_798532 to 798533, EPI_ISL_798536 to 798538, EPI_ISL_798540, EPI_ISL_798543, EPI_ISL_798545 to 798547, EPI_ISL_798552 to 798554, EPI_ISL_798556 to 798559, EPI_ISL_798562 to 798563, EPI_ISL_798565 to 798573, EPI_ISL_798576 to 798578, EPI_ISL_798580 to 798581, EPI_ISL_798583 to 798588, EPI_ISL_798592, EPI_ISL_798594 to 798597, EPI_ISL_798599, EPI_ISL_798602 to 798606, EPI_ISL_798608, EPI_ISL_798610 to 798611, EPI_ISL_798613, EPI_ISL_798616 to 798621, EPI_ISL_798624 to 798625, EPI_ISL_798627 to 798630, EPI_ISL_798634, EPI_ISL_798636, EPI_ISL_798638, EPI_ISL_798644 to 798648, EPI_ISL_798650 to 798651, EPI_ISL_798653 to 798658, EPI_ISL_798660, EPI_ISL_798663, EPI_ISL_798666, EPI_ISL_798670, EPI_ISL_798672, EPI_ISL_798675, EPI_ISL_798680, EPI_ISL_798691, EPI_ISL_798693, EPI_ISL_798696, EPI_ISL_798701 to 798702, EPI_ISL_798707, EPI_ISL_798710, EPI_ISL_798712, EPI_ISL_798716, EPI_ISL_798720 to 798721, EPI_ISL_798724 to 798725, EPI_ISL_798728, EPI_ISL_798738 to 798741, EPI_ISL_798750, EPI_ISL_798752, EPI_ISL_798755, EPI_ISL_798764, EPI_ISL_798767, EPI_ISL_798769 to 798770, EPI_ISL_798774, EPI_ISL_798776 to 798777, EPI_ISL_798779, EPI_ISL_798781, EPI_ISL_798801, EPI_ISL_798803, EPI_ISL_798808 to 798809, EPI_ISL_798812, EPI_ISL_798817, EPI_ISL_798821 to 798823, EPI_ISL_798830, EPI_ISL_798832, EPI_ISL_798837 to 798838, EPI_ISL_798844, EPI_ISL_798851, EPI_ISL_798855 to 798856, EPI_ISL_798863 to 798864, EPI_ISL_798870, EPI_ISL_798874 to 798876, EPI_ISL_798879, EPI_ISL_798882, EPI_ISL_798889, EPI_ISL_798892, EPI_ISL_798910 to 798911, EPI_ISL_798915, EPI_ISL_798919, EPI_ISL_798922, EPI_ISL_798925, EPI_ISL_798930, EPI_ISL_798933, EPI_ISL_798936, EPI_ISL_798942, EPI_ISL_798948, EPI_ISL_798950, EPI_ISL_798955, EPI_ISL_798961, EPI_ISL_798966 to 798967, EPI_ISL_798970, EPI_ISL_798972 to 798973, EPI_ISL_798975, EPI_ISL_798977, EPI_ISL_798979, EPI_ISL_798981, EPI_ISL_798983, EPI_ISL_798987, EPI_ISL_798994 to 798996, EPI_ISL_799296, EPI_ISL_799299, EPI_ISL_799303, EPI_ISL_799305 to 799306, EPI_ISL_799308, EPI_ISL_799311, EPI_ISL_799314, EPI_ISL_799317, EPI_ISL_799319 to 799320, EPI_ISL_799323, EPI_ISL_799325, EPI_ISL_799327, EPI_ISL_799334, EPI_ISL_799346 to 799347, EPI_ISL_799350 to 799354, EPI_ISL_799356, EPI_ISL_799360, EPI_ISL_799362 to 799364, EPI_ISL_799366, EPI_ISL_799368, EPI_ISL_799372 to 799373, EPI_ISL_799375 to 799376, EPI_ISL_799380, EPI_ISL_799384, EPI_ISL_799386 to 799388, EPI_ISL_799392, EPI_ISL_799394 to 799395, EPI_ISL_799397, EPI_ISL_799400, EPI_ISL_799403 to 799404, EPI_ISL_799406, EPI_ISL_799410 to 799413, EPI_ISL_799415, EPI_ISL_799420, EPI_ISL_799430, EPI_ISL_799432, EPI_ISL_799434, EPI_ISL_799436 to 799437, EPI_ISL_799439 to 799441, EPI_ISL_799443, EPI_ISL_799445, EPI_ISL_799447 to 799448, EPI_ISL_799454, EPI_ISL_799465 to 799470, EPI_ISL_799473, EPI_ISL_799476 to 799477, EPI_ISL_799479, EPI_ISL_799482, EPI_ISL_799485, EPI_ISL_799488, EPI_ISL_799490, EPI_ISL_799494, EPI_ISL_799500 to 799502, EPI_ISL_799505 to 799506, EPI_ISL_799508 to 799509, EPI_ISL_799514 to 799515, EPI_ISL_799519 to 799520, EPI_ISL_799522 to 799523, EPI_ISL_799528, EPI_ISL_799531, EPI_ISL_799533 to 799534, EPI_ISL_799536, EPI_ISL_799538, EPI_ISL_799540, EPI_ISL_799545 to 799547, EPI_ISL_799550 to 799551, EPI_ISL_799555, EPI_ISL_799562, EPI_ISL_799567, EPI_ISL_799570, EPI_ISL_799572 to 799573, EPI_ISL_799575, EPI_ISL_799577 to 799580, EPI_ISL_799582 to 799583, EPI_ISL_799587 to 799588, EPI_ISL_799590, EPI_ISL_799595, EPI_ISL_799598, EPI_ISL_799600, EPI_ISL_799604, EPI_ISL_799607 to 799608, EPI_ISL_799610, EPI_ISL_799614, EPI_ISL_799616 to 799618, EPI_ISL_799620 to 799621, EPI_ISL_799623 to 799628, EPI_ISL_799632 to 799633, EPI_ISL_799635 to 799636, EPI_ISL_799639 to 799640, EPI_ISL_799642, EPI_ISL_799645 to 799647, EPI_ISL_799651, EPI_ISL_799653 to 799654, EPI_ISL_799657 to 799658, EPI_ISL_799660, EPI_ISL_799666 to 799668, EPI_ISL_799670, EPI_ISL_799672 to 799673, EPI_ISL_799675, EPI_ISL_799677, EPI_ISL_799680, EPI_ISL_799685, EPI_ISL_799688, EPI_ISL_799690, EPI_ISL_799692, EPI_ISL_799694 to 799698, EPI_ISL_799700 to 799701, EPI_ISL_799703 to 799705, EPI_ISL_799709, EPI_ISL_799712, EPI_ISL_799714 to 799717, EPI_ISL_799719 to 799721, EPI_ISL_799723 to 799725, EPI_ISL_799728, EPI_ISL_799733, EPI_ISL_799736, EPI_ISL_799740 to 799743, EPI_ISL_799745, EPI_ISL_799752 to 799753, EPI_ISL_799755 to 799757, EPI_ISL_799762, EPI_ISL_799764, EPI_ISL_799766, EPI_ISL_799769 to 799774, EPI_ISL_799777 to 799779, EPI_ISL_799782, EPI_ISL_799788 to 799790, EPI_ISL_799793 to 799795, EPI_ISL_799798, EPI_ISL_799800, EPI_ISL_799802, EPI_ISL_799804, EPI_ISL_799806, EPI_ISL_799808, EPI_ISL_799810, EPI_ISL_799813, EPI_ISL_799815 to 799817, EPI_ISL_799820, EPI_ISL_799824, EPI_ISL_799826, EPI_ISL_799828 to 799829, EPI_ISL_799833 to 799837, EPI_ISL_799842, EPI_ISL_799845, EPI_ISL_799847 to 799852, EPI_ISL_799854, EPI_ISL_799857 to 799858, EPI_ISL_799860, EPI_ISL_799862 to 799863, EPI_ISL_799867, EPI_ISL_799870, EPI_ISL_799872 to 799874, EPI_ISL_799876, EPI_ISL_799880, EPI_ISL_799885 to 799887, EPI_ISL_799890, EPI_ISL_799895 to 799896, EPI_ISL_799900, EPI_ISL_799904 to 799905, EPI_ISL_799909, EPI_ISL_799911, EPI_ISL_799913, EPI_ISL_799915, EPI_ISL_799921, EPI_ISL_799923, EPI_ISL_799926 to 799928, EPI_ISL_799931, EPI_ISL_800581 to 800585, EPI_ISL_800593, EPI_ISL_800595, EPI_ISL_800599, EPI_ISL_800613, EPI_ISL_800617, EPI_ISL_800623 to 800624, EPI_ISL_800632, EPI_ISL_800636, EPI_ISL_800642, EPI_ISL_800644, EPI_ISL_800650, EPI_ISL_800652, EPI_ISL_800657, EPI_ISL_800660 to 800661, EPI_ISL_800664, EPI_ISL_800673 to 800674, EPI_ISL_800676 to 800677, EPI_ISL_800681, EPI_ISL_800684, EPI_ISL_800698, EPI_ISL_800701, EPI_ISL_800710 to 800711, EPI_ISL_800718, EPI_ISL_800729, EPI_ISL_800738, EPI_ISL_800740, EPI_ISL_800747, EPI_ISL_800751 to 800752, EPI_ISL_800757 to 800758, EPI_ISL_800762, EPI_ISL_800764, EPI_ISL_800769, EPI_ISL_800773, EPI_ISL_800777 to 800778, EPI_ISL_800783, EPI_ISL_800788, EPI_ISL_800807, EPI_ISL_800811, EPI_ISL_800814, EPI_ISL_800820, EPI_ISL_800822, EPI_ISL_800824, EPI_ISL_800830 to 800831, EPI_ISL_800834, EPI_ISL_800836 to 800837, EPI_ISL_800840, EPI_ISL_800844, EPI_ISL_800846, EPI_ISL_800849, EPI_ISL_800857, EPI_ISL_800863 to 800864, EPI_ISL_800866 to 800868, EPI_ISL_800871, EPI_ISL_800877 to 800879, EPI_ISL_800881, EPI_ISL_801180 to 801185, EPI_ISL_801187 to 801198, EPI_ISL_801200 to 801201, EPI_ISL_801203 to 801207, EPI_ISL_801209 to 801220, EPI_ISL_801222 to 801230, EPI_ISL_811214,

EPI_ISL_811234, EPI_ISL_811236, EPI_ISL_811241, EPI_ISL_811247, EPI_ISL_811275, EPI_ISL_811293, EPI_ISL_811310, EPI_ISL_811312, EPI_ISL_811321, EPI_ISL_811345, EPI_ISL_811350, EPI_ISL_811353, EPI_ISL_811377, EPI_ISL_811414, EPI_ISL_811422, EPI_ISL_811472, EPI_ISL_811490, EPI_ISL_811500, EPI_ISL_811516, EPI_ISL_811519, EPI_ISL_811530, EPI_ISL_811548, EPI_ISL_811563, EPI_ISL_811573, EPI_ISL_811580, EPI_ISL_811584, EPI_ISL_811594, EPI_ISL_811617 to 811618, EPI_ISL_811620 to 811621, EPI_ISL_811627, EPI_ISL_811645, EPI_ISL_811650 to 811651, EPI_ISL_811661, EPI_ISL_811669 to 811671, EPI_ISL_811677 to 811678, EPI_ISL_811682, EPI_ISL_811688, EPI_ISL_811720, EPI_ISL_811727, EPI_ISL_811750, EPI_ISL_811753, EPI_ISL_811756, EPI_ISL_811759, EPI_ISL_811767, EPI_ISL_811785, EPI_ISL_811808 to 811844, EPI_ISL_811846 to 811957, EPI_ISL_811959 to 812102, EPI_ISL_812104 to 812113, EPI_ISL_821275, EPI_ISL_821277, EPI_ISL_821291, EPI_ISL_821299, EPI_ISL_821302, EPI_ISL_821305, EPI_ISL_821312 to 821313, EPI_ISL_821315, EPI_ISL_821321 to 821322, EPI_ISL_821325 to 821326, EPI_ISL_821330, EPI_ISL_821332, EPI_ISL_821337, EPI_ISL_821345, EPI_ISL_821349, EPI_ISL_821355, EPI_ISL_821357, EPI_ISL_821361, EPI_ISL_821364, EPI_ISL_821369, EPI_ISL_821396, EPI_ISL_821413, EPI_ISL_821420, EPI_ISL_821429 to 821430, EPI_ISL_821450, EPI_ISL_821455, EPI_ISL_821464, EPI_ISL_821471, EPI_ISL_821481, EPI_ISL_821489, EPI_ISL_821495, EPI_ISL_821499, EPI_ISL_821505, EPI_ISL_821508, EPI_ISL_821512, EPI_ISL_821531, EPI_ISL_821557, EPI_ISL_821578, EPI_ISL_821581, EPI_ISL_821591, EPI_ISL_821611, EPI_ISL_821619, EPI_ISL_821629, EPI_ISL_821631 to 821658, EPI_ISL_822305, EPI_ISL_852017, EPI_ISL_858792, EPI_ISL_858795, EPI_ISL_858798 to 858799, EPI_ISL_858803 to 858805, EPI_ISL_858807 to 858808, EPI_ISL_859327, EPI_ISL_859469, EPI_ISL_859540, EPI_ISL_859553

| | | | |
|---|---|---|---|
| see above | Lighthouse Lab in Milton Keynes | Wellcome Sanger Institute for the COVID-19 Genomics UK (COG-UK) Consortium | Cordelia Langford; David K. Jackson; Dominic Kwiatkowski; Ewan Harrison; Ian Johnston; John Sillitoe on behalf of the Wellcome Sanger Institute COVID-19 Surveillance Team; John Sillitoe on behalf of the Wellcome Sanger Institute COVID-19 Surveillance Team (http://www.sanger.ac.uk/covid-team); Roberto Amato; Sonia Goncalves; The Lighthouse Lab in Alderley Park and Alex Alderton; The Lighthouse Lab in Milton Keynes and Alex Alderton |

EPI_ISL_549808 to 549828, EPI_ISL_549830 to 549846, EPI_ISL_549848 to 549860, EPI_ISL_549863 to 549890, EPI_ISL_549892 to 549901, EPI_ISL_549904 to 549921, EPI_ISL_549923 to 549926, EPI_ISL_549928 to 549951, EPI_ISL_549953 to 549980, EPI_ISL_549982 to 549988, EPI_ISL_549990 to 550024, EPI_ISL_550026 to 550032, EPI_ISL_550034 to 550065, EPI_ISL_550067 to 550068, EPI_ISL_550070 to 550075, EPI_ISL_550077 to 550153, EPI_ISL_550155 to 550165, EPI_ISL_550167 to 550178, EPI_ISL_550180 to 550219, EPI_ISL_550221 to 550227, EPI_ISL_550229, EPI_ISL_550231 to 550327, EPI_ISL_550329 to 550333, EPI_ISL_550335 to 550339, EPI_ISL_550341 to 550415, EPI_ISL_550419, EPI_ISL_550422 to 550424, EPI_ISL_550428 to 550432, EPI_ISL_550434 to 550435, EPI_ISL_550437 to 550440, EPI_ISL_550442 to 550443, EPI_ISL_550445 to 550451, EPI_ISL_550453 to 550454, EPI_ISL_550456 to 550465, EPI_ISL_550467, EPI_ISL_550470 to 550473, EPI_ISL_550475, EPI_ISL_550479 to 550481, EPI_ISL_550487 to 550488, EPI_ISL_550490 to 550491, EPI_ISL_550496, EPI_ISL_550498, EPI_ISL_550500, EPI_ISL_550503 to 550510, EPI_ISL_550514 to 550516, EPI_ISL_550518 to 550528, EPI_ISL_550530 to 550541, EPI_ISL_550543 to 550561, EPI_ISL_550563 to 550616, EPI_ISL_550618 to 550699, EPI_ISL_550701 to 550717, EPI_ISL_550719 to 550848, EPI_ISL_550851 to 550869, EPI_ISL_550871 to 550876, EPI_ISL_550878 to 550908, EPI_ISL_550910 to 550929, EPI_ISL_550931 to 550982, EPI_ISL_550984, EPI_ISL_550986 to 551000, EPI_ISL_551002 to 551006, EPI_ISL_551008 to 551012, EPI_ISL_551014 to 551019, EPI_ISL_551021 to 551041, EPI_ISL_551043 to 551046, EPI_ISL_551048 to 551067, EPI_ISL_551069 to 551074, EPI_ISL_551076 to 551091, EPI_ISL_551093 to 551105, EPI_ISL_551107, EPI_ISL_551109 to 551113, EPI_ISL_551115 to 551132, EPI_ISL_551134 to 551140, EPI_ISL_551142 to 551156, EPI_ISL_551158, EPI_ISL_551160 to 551179, EPI_ISL_551181 to 551200, EPI_ISL_551202 to 551214, EPI_ISL_551216 to 551217, EPI_ISL_551219 to 551225, EPI_ISL_551227 to 551253, EPI_ISL_551255 to 551277, EPI_ISL_551280 to 551284, EPI_ISL_551577 to 551645, EPI_ISL_551647 to 551659, EPI_ISL_551661 to 551662, EPI_ISL_551664 to 551697, EPI_ISL_551699 to 551708, EPI_ISL_551710 to 551718, EPI_ISL_551720 to 551734, EPI_ISL_551737 to 551739, EPI_ISL_551741, EPI_ISL_551744 to 551748, EPI_ISL_551750, EPI_ISL_551752 to 551754, EPI_ISL_551757 to 551758, EPI_ISL_551765 to 551768, EPI_ISL_551770 to 551771, EPI_ISL_551773 to 551775, EPI_ISL_551777 to 551778, EPI_ISL_551781 to 551783, EPI_ISL_551786 to 551787, EPI_ISL_551790 to 551794, EPI_ISL_551797, EPI_ISL_551800, EPI_ISL_551802, EPI_ISL_551805, EPI_ISL_551809, EPI_ISL_551813 to 551814, EPI_ISL_551816 to 551817, EPI_ISL_551820 to 551822, EPI_ISL_551828 to 551831, EPI_ISL_551833, EPI_ISL_551922 to 551924, EPI_ISL_551926 to 551931, EPI_ISL_551934 to 551936, EPI_ISL_551938 to 551950, EPI_ISL_551952, EPI_ISL_551954 to 551956, EPI_ISL_551958 to 551966, EPI_ISL_551968 to 551979, EPI_ISL_551981 to 551982, EPI_ISL_551984 to 551987, EPI_ISL_551989, EPI_ISL_551991 to 551993, EPI_ISL_551995 to 552000, EPI_ISL_552002 to 552004, EPI_ISL_552006 to 552007, EPI_ISL_552011 to 552016, EPI_ISL_552018 to 552036, EPI_ISL_552038 to 552050, EPI_ISL_552052 to 552064, EPI_ISL_552066 to 552072, EPI_ISL_552074 to 552090, EPI_ISL_552092 to 552093, EPI_ISL_552095 to 552097, EPI_ISL_552099 to 552104, EPI_ISL_552107 to 552123, EPI_ISL_552125 to 552150, EPI_ISL_552152 to 552156, EPI_ISL_552158 to 552159, EPI_ISL_552161 to 552165, EPI_ISL_552167 to 552169, EPI_ISL_552171 to 552172, EPI_ISL_552174 to 552179, EPI_ISL_552181 to 552184, EPI_ISL_552186 to 552211, EPI_ISL_552213 to 552226, EPI_ISL_552228 to 552239, EPI_ISL_552241 to 552275, EPI_ISL_552277 to 552282, EPI_ISL_552284 to 552289, EPI_ISL_552291 to 552293, EPI_ISL_552295 to 552305, EPI_ISL_552307, EPI_ISL_552309 to 552341, EPI_ISL_552343 to 552373, EPI_ISL_552375 to 552377, EPI_ISL_552379 to 552409, EPI_ISL_552411, EPI_ISL_552413 to 552445, EPI_ISL_552447 to 552448, EPI_ISL_552450 to 552457, EPI_ISL_552459 to 552462, EPI_ISL_552465, EPI_ISL_552467, EPI_ISL_552469 to 552472, EPI_ISL_552474 to 552475, EPI_ISL_552477 to 552478, EPI_ISL_552480, EPI_ISL_552482 to 552485, EPI_ISL_552487, EPI_ISL_552489 to 552491, EPI_ISL_552493 to 552505, EPI_ISL_552509 to 552511, EPI_ISL_552513, EPI_ISL_552517 to 552522, EPI_ISL_552524 to 552525, EPI_ISL_552528 to 552529, EPI_ISL_552531 to 552532, EPI_ISL_552535 to 552538, EPI_ISL_552541 to 552546, EPI_ISL_552548 to 552554, EPI_ISL_552557 to 552561, EPI_ISL_552563 to 552564, EPI_ISL_552568 to 552571, EPI_ISL_552573, EPI_ISL_552575, EPI_ISL_552578, EPI_ISL_552581 to 552582, EPI_ISL_552584 to 552586, EPI_ISL_552588 to 552589, EPI_ISL_552592 to 552594, EPI_ISL_552597, EPI_ISL_552599 to 552606, EPI_ISL_552609 to 552613, EPI_ISL_552615 to 552616, EPI_ISL_552619, EPI_ISL_552621, EPI_ISL_552623 to 552628, EPI_ISL_552630, EPI_ISL_552632 to 552638, EPI_ISL_552640 to 552641, EPI_ISL_552643, EPI_ISL_552645, EPI_ISL_552647 to 552654, EPI_ISL_552656 to 552658, EPI_ISL_552660 to 552662, EPI_ISL_552664, EPI_ISL_552667 to 552668, EPI_ISL_552670, EPI_ISL_552672 to 552678, EPI_ISL_552680, EPI_ISL_552682 to 552685, EPI_ISL_552688, EPI_ISL_552690 to 552698, EPI_ISL_552700 to 552702, EPI_ISL_552704 to 552705, EPI_ISL_552708 to 552711, EPI_ISL_552714 to 552716, EPI_ISL_552718, EPI_ISL_552725, EPI_ISL_552727, EPI_ISL_552733 to 552734, EPI_ISL_552736 to 552740, EPI_ISL_552744, EPI_ISL_552747, EPI_ISL_552751 to 552759, EPI_ISL_552762 to 552767, EPI_ISL_552769, EPI_ISL_552772, EPI_ISL_552778, EPI_ISL_552781, EPI_ISL_552784, EPI_ISL_552786 to 552791, EPI_ISL_552795, EPI_ISL_552799, EPI_ISL_552802, EPI_ISL_552809 to 552811, EPI_ISL_552814, EPI_ISL_552820, EPI_ISL_552823, EPI_ISL_552842, EPI_ISL_552852, EPI_ISL_552867, EPI_ISL_552874 to 552875, EPI_ISL_552879, EPI_ISL_552905, EPI_ISL_552910, EPI_ISL_552916, EPI_ISL_552936, EPI_ISL_552938 to 552939, EPI_ISL_552947 to 552949, EPI_ISL_552953, EPI_ISL_552963, EPI_ISL_552967, EPI_ISL_552975, EPI_ISL_552979, EPI_ISL_552987, EPI_ISL_552990, EPI_ISL_552995, EPI_ISL_552997, EPI_ISL_553007 to 553037, EPI_ISL_553039 to 553043, EPI_ISL_553045 to 553083, EPI_ISL_553086 to 553092, EPI_ISL_553094 to 553097, EPI_ISL_553099 to 553107, EPI_ISL_553109 to 553127, EPI_ISL_553129 to 553141, EPI_ISL_553143 to 553147, EPI_ISL_553149 to 553181, EPI_ISL_553183 to 553185, EPI_ISL_553187 to 553230, EPI_ISL_553232 to 553241, EPI_ISL_553243 to 553268, EPI_ISL_553270, EPI_ISL_553275 to 553276, EPI_ISL_553295, EPI_ISL_553323, EPI_ISL_553879 to 553880, EPI_ISL_553885, EPI_ISL_553889, EPI_ISL_553893 to 553894, EPI_ISL_553896 to 553898, EPI_ISL_553905, EPI_ISL_553907, EPI_ISL_553929, EPI_ISL_553934, EPI_ISL_553939, EPI_ISL_553952, EPI_ISL_553954 to 553957, EPI_ISL_553965, EPI_ISL_553972, EPI_ISL_553983, EPI_ISL_553985, EPI_ISL_553987, EPI_ISL_553989, EPI_ISL_553993 to 553994, EPI_ISL_554003, EPI_ISL_554005, EPI_ISL_554009, EPI_ISL_554015, EPI_ISL_554024 to 554026, EPI_ISL_554028, EPI_ISL_554042, EPI_ISL_554048, EPI_ISL_554054, EPI_ISL_554057 to 554058, EPI_ISL_554065, EPI_ISL_554080, EPI_ISL_554097, EPI_ISL_554099 to 554101, EPI_ISL_554118 to 554119, EPI_ISL_554121, EPI_ISL_554123 to 554125, EPI_ISL_554127 to 554128, EPI_ISL_554132 to 554135, EPI_ISL_554137 to 554139, EPI_ISL_554141 to 554144, EPI_ISL_554146 to 554148, EPI_ISL_554150, EPI_ISL_554152 to 554154, EPI_ISL_554159 to 554163, EPI_ISL_554166 to 554169, EPI_ISL_554173, EPI_ISL_554177 to 554182, EPI_ISL_554184 to 554185, EPI_ISL_554191, EPI_ISL_554200 to 554201, EPI_ISL_554204 to 554205, EPI_ISL_554208, EPI_ISL_554210 to 554211, EPI_ISL_554214, EPI_ISL_554216 to 554218, EPI_ISL_554221, EPI_ISL_554223 to 554224, EPI_ISL_554226, EPI_ISL_554228, EPI_ISL_554231 to 554232, EPI_ISL_554234, EPI_ISL_554236 to 554237, EPI_ISL_554239 to 554240, EPI_ISL_554242 to 554249, EPI_ISL_554251, EPI_ISL_554253 to 554256, EPI_ISL_554258 to 554259, EPI_ISL_554261 to 554262, EPI_ISL_554264, EPI_ISL_554267, EPI_ISL_554269, EPI_ISL_554271 to 554272, EPI_ISL_554274, EPI_ISL_554276 to 554277, EPI_ISL_554284 to 554287, EPI_ISL_554289 to 554291, EPI_ISL_554293 to 554295, EPI_ISL_554297, EPI_ISL_554300, EPI_ISL_554302 to 554304, EPI_ISL_554306 to 554308, EPI_ISL_554310 to 554317, EPI_ISL_554319, EPI_ISL_554619, EPI_ISL_554621 to 554622, EPI_ISL_554627 to 554629, EPI_ISL_554633 to 554636, EPI_ISL_554638 to 554639, EPI_ISL_554641, EPI_ISL_554643 to 554645, EPI_ISL_554647 to 554650, EPI_ISL_554652, EPI_ISL_554655, EPI_ISL_554658 to 554660, EPI_ISL_554665, EPI_ISL_554668 to 554674, EPI_ISL_554676, EPI_ISL_554678 to 554679, EPI_ISL_554682 to 554685, EPI_ISL_554687, EPI_ISL_554690, EPI_ISL_554695, EPI_ISL_554698 to 554702, EPI_ISL_554704 to 554707, EPI_ISL_554711 to 554715, EPI_ISL_554717 to 554724, EPI_ISL_554728 to 554729, EPI_ISL_554731, EPI_ISL_554733 to 554734, EPI_ISL_554736 to 554740, EPI_ISL_554742, EPI_ISL_554745 to 554753, EPI_ISL_554758, EPI_ISL_554760, EPI_ISL_554762, EPI_ISL_554764 to 554766, EPI_ISL_554768 to 554770, EPI_ISL_554773 to 554775, EPI_ISL_554783 to 554791, EPI_ISL_554793, EPI_ISL_554796, EPI_ISL_554800 to 554801, EPI_ISL_554803, EPI_ISL_554805 to 554806, EPI_ISL_554814, EPI_ISL_554817 to 554819, EPI_ISL_554821 to 554826, EPI_ISL_554828 to 554829, EPI_ISL_554831, EPI_ISL_554835, EPI_ISL_554838 to 554839, EPI_ISL_554842 to 554844, EPI_ISL_554851 to 554852, EPI_ISL_554854 to 554855, EPI_ISL_554857 to 554860, EPI_ISL_555558, EPI_ISL_555616, EPI_ISL_556075 to 556076, EPI_ISL_556079, EPI_ISL_556083, EPI_ISL_556087, EPI_ISL_556090 to 556095, EPI_ISL_556097 to 556098, EPI_ISL_556102, EPI_ISL_556104, EPI_ISL_556109, EPI_ISL_556112, EPI_ISL_556115 to 556116, EPI_ISL_556123, EPI_ISL_556126, EPI_ISL_556133 to 556134, EPI_ISL_556138 to 556139, EPI_ISL_556146, EPI_ISL_556148, EPI_ISL_556150, EPI_ISL_556158, EPI_ISL_556163, EPI_ISL_556165 to 556166, EPI_ISL_556170, EPI_ISL_556172, EPI_ISL_556176, EPI_ISL_556183, EPI_ISL_556186 to 556187, EPI_ISL_556195, EPI_ISL_556197 to 556199, EPI_ISL_556201, EPI_ISL_556204, EPI_ISL_556208, EPI_ISL_556211, EPI_ISL_556213 to 556214, EPI_ISL_556223, EPI_ISL_556233, EPI_ISL_556235, EPI_ISL_556237, EPI_ISL_556242, EPI_ISL_556244, EPI_ISL_556248, EPI_ISL_556251, EPI_ISL_556253, EPI_ISL_556259, EPI_ISL_556261, EPI_ISL_556263, EPI_ISL_556266 to 556267, EPI_ISL_556269, EPI_ISL_556275 to 556276, EPI_ISL_556282, EPI_ISL_556285, EPI_ISL_556287, EPI_ISL_556289, EPI_ISL_556291, EPI_ISL_556293 to 556294, EPI_ISL_556298, EPI_ISL_556300, EPI_ISL_556304, EPI_ISL_557166 to 557185, EPI_ISL_557187 to 557188, EPI_ISL_557190 to 557212, EPI_ISL_557214 to 557232, EPI_ISL_557234 to 557237, EPI_ISL_557239 to 557248, EPI_ISL_557250 to 557279, EPI_ISL_557281 to 557282, EPI_ISL_557284 to 557307, EPI_ISL_557309 to 557323, EPI_ISL_557325 to 557344, EPI_ISL_557346 to 557350, EPI_ISL_557352, EPI_ISL_557357 to 557358, EPI_ISL_557360, EPI_ISL_557362, EPI_ISL_557364 to 557365, EPI_ISL_557367 to 557370, EPI_ISL_557372, EPI_ISL_557375 to 557377, EPI_ISL_557384 to 557387, EPI_ISL_557389, EPI_ISL_557391 to 557397, EPI_ISL_557400 to 557403, EPI_ISL_557407, EPI_ISL_557409 to 557410, EPI_ISL_557413, EPI_ISL_557416, EPI_ISL_557418, EPI_ISL_557420 to 557422, EPI_ISL_557424, EPI_ISL_557426 to 557427, EPI_ISL_557429, EPI_ISL_557431 to 557432, EPI_ISL_557435, EPI_ISL_557437 to 557438, EPI_ISL_557441 to 557445, EPI_ISL_557450 to 557451, EPI_ISL_557455 to 557465, EPI_ISL_557467, EPI_ISL_557470, EPI_ISL_557472 to 557473, EPI_ISL_557475, EPI_ISL_557477 to 557480, EPI_ISL_557483, EPI_ISL_557486, EPI_ISL_557489 to 557490, EPI_ISL_557492, EPI_ISL_557494, EPI_ISL_557496, EPI_ISL_557498, EPI_ISL_557501 to 557503, EPI_ISL_557505 to 557506, EPI_ISL_557508 to 557510, EPI_ISL_557512 to 557517, EPI_ISL_557519 to 557521, EPI_ISL_557524 to 557525, EPI_ISL_557529, EPI_ISL_557532, EPI_ISL_557534 to 557536, EPI_ISL_557538, EPI_ISL_557540, EPI_ISL_557542, EPI_ISL_557545 to 557550, EPI_ISL_557552, EPI_ISL_557701, EPI_ISL_557740 to 557742, EPI_ISL_557745, EPI_ISL_557748, EPI_ISL_557750, EPI_ISL_557753, EPI_ISL_557757 to 557758, EPI_ISL_557764 to 557766, EPI_ISL_557768, EPI_ISL_557771, EPI_ISL_557775, EPI_ISL_557778, EPI_ISL_557781 to 557782, EPI_ISL_557784, EPI_ISL_557786, EPI_ISL_557789, EPI_ISL_557791, EPI_ISL_557793, EPI_ISL_557795 to 557796, EPI_ISL_557800, EPI_ISL_557802, EPI_ISL_557807 to 557808, EPI_ISL_557811 to 557812, EPI_ISL_557816 to 557817, EPI_ISL_557821 to 557824, EPI_ISL_557827 to 557830, EPI_ISL_557832, EPI_ISL_557838, EPI_ISL_557845, EPI_ISL_557847 to 557849, EPI_ISL_557853 to 557854, EPI_ISL_557858, EPI_ISL_557865 to 557870, EPI_ISL_557876 to 557880, EPI_ISL_557884, EPI_ISL_557889 to 557890, EPI_ISL_557893 to 557895, EPI_ISL_557898, EPI_ISL_557905, EPI_ISL_557907 to 557909, EPI_ISL_557911, EPI_ISL_557915, EPI_ISL_557917, EPI_ISL_557919, EPI_ISL_557921, EPI_ISL_557925 to 557927, EPI_ISL_557933, EPI_ISL_557937 to 557938, EPI_ISL_557940 to 557942, EPI_ISL_557944, EPI_ISL_557947, EPI_ISL_557949, EPI_ISL_557953 to 557954, EPI_ISL_557956, EPI_ISL_557958 to 557959, EPI_ISL_557963 to 557965, EPI_ISL_557967, EPI_ISL_557970, EPI_ISL_557972, EPI_ISL_557974, EPI_ISL_557976 to 557977, EPI_ISL_557979, EPI_ISL_557984, EPI_ISL_557989, EPI_ISL_558001, EPI_ISL_558021, EPI_ISL_558026, EPI_ISL_558028, EPI_ISL_558030, EPI_ISL_558037, EPI_ISL_558041, EPI_ISL_558058, EPI_ISL_558068, EPI_ISL_558089, EPI_ISL_558096, EPI_ISL_558103, EPI_ISL_558106, EPI_ISL_558127, EPI_ISL_558155, EPI_ISL_558195, EPI_ISL_558234, EPI_ISL_558239, EPI_ISL_558241 to 558243, EPI_ISL_558245 to 558255, EPI_ISL_558257 to 558258, EPI_ISL_558262, EPI_ISL_558264 to 558267, EPI_ISL_558271 to 558273, EPI_ISL_558275, EPI_ISL_558278 to 558282, EPI_ISL_558284 to 558287, EPI_ISL_558292 to 558306, EPI_ISL_558308 to 558310, EPI_ISL_558313 to 558315, EPI_ISL_558317 to 558320, EPI_ISL_558322 to 558323, EPI_ISL_558325 to 558327, EPI_ISL_558330 to 558331, EPI_ISL_558333 to 558334, EPI_ISL_558336 to 558337, EPI_ISL_558339 to 558342, EPI_ISL_558346 to 558347, EPI_ISL_558349, EPI_ISL_558351 to 558353, EPI_ISL_558356 to 558361, EPI_ISL_558363, EPI_ISL_558365 to 558367, EPI_ISL_558369 to 558378, EPI_ISL_558380 to 558386, EPI_ISL_558388, EPI_ISL_558390 to 558391, EPI_ISL_558393, EPI_ISL_558397 to 558401, EPI_ISL_558403, EPI_ISL_558638, EPI_ISL_558731, EPI_ISL_558903, EPI_ISL_558914, EPI_ISL_558920, EPI_ISL_558923, EPI_ISL_558925, EPI_ISL_558928, EPI_ISL_558932, EPI_ISL_558934, EPI_ISL_558937, EPI_ISL_558951, EPI_ISL_558959, EPI_ISL_558965, EPI_ISL_558970, EPI_ISL_558973, EPI_ISL_558975, EPI_ISL_558980 to 558981, EPI_ISL_558987, EPI_ISL_558993, EPI_ISL_559000, EPI_ISL_559005, EPI_ISL_559009, EPI_ISL_559011, EPI_ISL_559022, EPI_ISL_559026, EPI_ISL_559028, EPI_ISL_559043, EPI_ISL_559047, EPI_ISL_559049, EPI_ISL_559057, EPI_ISL_559061, EPI_ISL_559063, EPI_ISL_559071, EPI_ISL_559080 to 559081, EPI_ISL_559083 to 559084, EPI_ISL_559088, EPI_ISL_559100, EPI_ISL_559104, EPI_ISL_559106, EPI_ISL_559117, EPI_ISL_559119, EPI_ISL_559126, EPI_ISL_559131 to 559133, EPI_ISL_559136, EPI_ISL_559147, EPI_ISL_559149, EPI_ISL_559154, EPI_ISL_559157 to 559158, EPI_ISL_559163 to 559164, EPI_ISL_559166 to 559167, EPI_ISL_559171, EPI_ISL_559174, EPI_ISL_559187, EPI_ISL_559194 to 559195, EPI_ISL_559201, EPI_ISL_559204, EPI_ISL_559207, EPI_ISL_559218, EPI_ISL_559223, EPI_ISL_559232, EPI_ISL_559235 to 559236, EPI_ISL_559241, EPI_ISL_559246 to 559247, EPI_ISL_559253 to 559254, EPI_ISL_559256, EPI_ISL_559259, EPI_ISL_559263, EPI_ISL_559277 to 559278, EPI_ISL_559283 to 559284, EPI_ISL_559290, EPI_ISL_559296, EPI_ISL_559300, EPI_ISL_559312, EPI_ISL_559314, EPI_ISL_559319 to 559320, EPI_ISL_559326, EPI_ISL_559331, EPI_ISL_559335, EPI_ISL_559352, EPI_ISL_559354 to 559355, EPI_ISL_559368, EPI_ISL_559662 to 559669, EPI_ISL_559671 to 559698, EPI_ISL_559700 to 559705, EPI_ISL_559707 to 559715, EPI_ISL_559717 to 559723, EPI_ISL_559725 to 559729, EPI_ISL_559731, EPI_ISL_559735 to 559739, EPI_ISL_559741 to 559751, EPI_ISL_566421 to 566422, EPI_ISL_566424 to 566425, EPI_ISL_566430 to 566434, EPI_ISL_566436 to 566443, EPI_ISL_566446 to 566451, EPI_ISL_566454, EPI_ISL_566458 to 566466, EPI_ISL_566468 to 566472, EPI_ISL_566475 to 566478, EPI_ISL_566480 to 566491, EPI_ISL_566494, EPI_ISL_566496 to 566499, EPI_ISL_566501, EPI_ISL_566503 to 566506, EPI_ISL_566509 to 566510, EPI_ISL_566512, EPI_ISL_566514 to 566517, EPI_ISL_566520 to 566523, EPI_ISL_566525 to 566527, EPI_ISL_566529 to 566533, EPI_ISL_566535, EPI_ISL_566538 to 566542, EPI_ISL_566545 to 566560, EPI_ISL_566562 to 566566, EPI_ISL_566568 to 566575, EPI_ISL_566577 to 566585, EPI_ISL_566587 to 566588, EPI_ISL_566590 to 566609, EPI_ISL_566612 to 566624, EPI_ISL_566626 to 566630, EPI_ISL_566632 to 566634, EPI_ISL_566636 to 566637, EPI_ISL_566639, EPI_ISL_566641 to 566658, EPI_ISL_566660, EPI_ISL_566663 to 566665, EPI_ISL_566667 to 566671, EPI_ISL_566673, EPI_ISL_566675 to 566693, EPI_ISL_566695 to 566723, EPI_ISL_566725 to 566746, EPI_ISL_566748 to 566760, EPI_ISL_566762 to 566790, EPI_ISL_566792 to 566794, EPI_ISL_566796 to 566810, EPI_ISL_566812 to 566822, EPI_ISL_566824 to 566840, EPI_ISL_566842 to 566852, EPI_ISL_566854 to 566855, EPI_ISL_566857 to 566865, EPI_ISL_566867 to 566869, EPI_ISL_566871 to 566921, EPI_ISL_566923 to 566931, EPI_ISL_566933 to 566940, EPI_ISL_566942, EPI_ISL_566944 to 566950, EPI_ISL_566952 to 566963, EPI_ISL_566965 to 566985, EPI_ISL_566987 to 566997, EPI_ISL_568289 to 568290, EPI_ISL_568364 to 568365, EPI_ISL_568386 to 568387, EPI_ISL_568395, EPI_ISL_568466, EPI_ISL_568469 to 568471, EPI_ISL_568476, EPI_ISL_575367, EPI_ISL_575762 to 575766, EPI_ISL_575768 to 575787, EPI_ISL_575789 to 575795, EPI_ISL_575797 to 575800, EPI_ISL_575802 to 575804, EPI_ISL_575806 to 575807, EPI_ISL_575809 to 575811, EPI_ISL_575813 to 575821, EPI_ISL_575823 to 575830, EPI_ISL_575832 to 575838, EPI_ISL_575840 to 575843, EPI_ISL_575846 to 575849, EPI_ISL_575851 to 575869, EPI_ISL_575872 to 575873, EPI_ISL_575875 to 575876, EPI_ISL_575878 to 575881, EPI_ISL_575884 to 575892, EPI_ISL_575894 to 575911, EPI_ISL_575913 to 575927, EPI_ISL_575929 to 575930, EPI_ISL_575932 to 575951, EPI_ISL_575953, EPI_ISL_575955 to 575956, EPI_ISL_575958 to 575965, EPI_ISL_575967 to 575977, EPI_ISL_575979 to 575981, EPI_ISL_575984 to 575994, EPI_ISL_576094, EPI_ISL_580631 to 580632, EPI_ISL_580634, EPI_ISL_580637 to 580639, EPI_ISL_580641 to 580644, EPI_ISL_580646 to 580647, EPI_ISL_580649, EPI_ISL_580652 to 580654, EPI_ISL_580656 to 580659, EPI_ISL_580661 to 580662, EPI_ISL_580664 to 580673, EPI_ISL_580676, EPI_ISL_580683 to 580685, EPI_ISL_580687 to 580688, EPI_ISL_580693 to 580694, EPI_ISL_580698, EPI_ISL_580700 to 580705, EPI_ISL_580707, EPI_ISL_580709, EPI_ISL_580711 to 580719, EPI_ISL_580721, EPI_ISL_580724 to 580729, EPI_ISL_580731 to 580733, EPI_ISL_580735 to 580736, EPI_ISL_580739, EPI_ISL_580741 to 580753, EPI_ISL_580755, EPI_ISL_580757 to 580762, EPI_ISL_580766 to 580768, EPI_ISL_580772 to 580779, EPI_ISL_580781 to 580783, EPI_ISL_580785 to 580786, EPI_ISL_580790, EPI_ISL_580792 to 580793, EPI_ISL_580795 to 580820, EPI_ISL_580826 to 580827, EPI_ISL_580829 to 580832, EPI_ISL_580834, EPI_ISL_580836 to 580839, EPI_ISL_580841 to 580848, EPI_ISL_580850, EPI_ISL_580852, EPI_ISL_580854, EPI_ISL_580857, EPI_ISL_580861, EPI_ISL_580864, EPI_ISL_580873, EPI_ISL_580875, EPI_ISL_580877, EPI_ISL_580882, EPI_ISL_580885, EPI_ISL_580892, EPI_ISL_580896 to 580898, EPI_ISL_580900, EPI_ISL_580902 to 580904, EPI_ISL_580910, EPI_ISL_580913, EPI_ISL_580915, EPI_ISL_580917 to 580918, EPI_ISL_580921, EPI_ISL_580929, EPI_ISL_580935, EPI_ISL_580940, EPI_ISL_580948, EPI_ISL_580950 to 580951, EPI_ISL_580959, EPI_ISL_580961, EPI_ISL_580976, EPI_ISL_580979 to 580980, EPI_ISL_580990 to 580991, EPI_ISL_580993, EPI_ISL_580995 to 580996, EPI_ISL_580998, EPI_ISL_581000, EPI_ISL_581004, EPI_ISL_581008 to 581009, EPI_ISL_581018, EPI_ISL_581023, EPI_ISL_581025 to 581026, EPI_ISL_581028 to 581029, EPI_ISL_581032, EPI_ISL_581035, EPI_ISL_581038, EPI_ISL_581040 to 581041, EPI_ISL_581046, EPI_ISL_581054, EPI_ISL_581056, EPI_ISL_581058, EPI_ISL_581064, EPI_ISL_581069, EPI_ISL_581074, EPI_ISL_581076, EPI_ISL_581078, EPI_ISL_581084, EPI_ISL_581086, EPI_ISL_581093, EPI_ISL_581097, EPI_ISL_581099, EPI_ISL_581103, EPI_ISL_581106 to 581107, EPI_ISL_581111, EPI_ISL_581113 to 581117, EPI_ISL_581119 to 581126, EPI_ISL_581128 to 581129, EPI_ISL_581131, EPI_ISL_581134 to 581140, EPI_ISL_581142 to 581145, EPI_ISL_581147 to 581148, EPI_ISL_581150 to 581153, EPI_ISL_581155 to 581165, EPI_ISL_581167 to 581171, EPI_ISL_581173, EPI_ISL_581175, EPI_ISL_581177, EPI_ISL_581179 to 581181, EPI_ISL_581184 to 581188, EPI_ISL_581190 to 581192, EPI_ISL_581195, EPI_ISL_581197 to 581213, EPI_ISL_581215 to 581216, EPI_ISL_581218 to 581222, EPI_ISL_581224 to 581225, EPI_ISL_581227 to 581228, EPI_ISL_581231 to 581236, EPI_ISL_581238 to 581244, EPI_ISL_581246, EPI_ISL_581248, EPI_ISL_581250 to 581260, EPI_ISL_581263 to 581266, EPI_ISL_581268 to 581284, EPI_ISL_581287 to 581290, EPI_ISL_581292 to 581306, EPI_ISL_581308 to 581309, EPI_ISL_581311 to 581322, EPI_ISL_581325 to 581328, EPI_ISL_581330 to 581333, EPI_ISL_581335 to 581336, EPI_ISL_581339 to 581340, EPI_ISL_581342 to 581343, EPI_ISL_581345, EPI_ISL_581347 to 581349, EPI_ISL_581351 to 581353, EPI_ISL_581355 to 581357, EPI_ISL_581360 to 581365, EPI_ISL_581368 to 581372, EPI_ISL_581374 to 581376, EPI_ISL_581382 to 581383, EPI_ISL_581385 to 581386, EPI_ISL_581389 to 581390, EPI_ISL_581393, EPI_ISL_581397 to 581401, EPI_ISL_581415, EPI_ISL_581421 to 581422, EPI_ISL_581428, EPI_ISL_581434, EPI_ISL_581438, EPI_ISL_581440, EPI_ISL_581447, EPI_ISL_589017 to 589019, EPI_ISL_589021, EPI_ISL_589024, EPI_ISL_589026, EPI_ISL_589028 to 589030, EPI_ISL_589034, EPI_ISL_589039 to 589041, EPI_ISL_589043, EPI_ISL_589051, EPI_ISL_589057, EPI_ISL_589063,

| | | | |
|---|---|---|---|
| EPI_ISL_589067, EPI_ISL_589070, EPI_ISL_589074 to 589075, EPI_ISL_589077, EPI_ISL_589079 to 589081, EPI_ISL_589086, EPI_ISL_589090 to 589091, EPI_ISL_589095, EPI_ISL_589097 to 589099, EPI_ISL_589101, EPI_ISL_589103, EPI_ISL_589108 to 589111, EPI_ISL_589114 to 589115, EPI_ISL_589117 to 589118, EPI_ISL_589121 to 589123, EPI_ISL_589127, EPI_ISL_589133, EPI_ISL_589137, EPI_ISL_589147, EPI_ISL_589153, EPI_ISL_589157, EPI_ISL_589160 to 589161, EPI_ISL_589166 to 589174, EPI_ISL_589176, EPI_ISL_589179, EPI_ISL_589182 to 589183, EPI_ISL_589185, EPI_ISL_589187, EPI_ISL_589195 to 589196, EPI_ISL_589198 to 589200, EPI_ISL_589205, EPI_ISL_589211 to 589212, EPI_ISL_589221, EPI_ISL_589224 to 589227, EPI_ISL_589229, EPI_ISL_589231, EPI_ISL_589233, EPI_ISL_589235, EPI_ISL_589237, EPI_ISL_589242 to 589243, EPI_ISL_589247, EPI_ISL_589251, EPI_ISL_589253, EPI_ISL_589256, EPI_ISL_589259 to 589261, EPI_ISL_589264 to 589266, EPI_ISL_589297, EPI_ISL_589304, EPI_ISL_589335, EPI_ISL_589355, EPI_ISL_589377, EPI_ISL_589407, EPI_ISL_589417, EPI_ISL_589423, EPI_ISL_589430, EPI_ISL_589440, EPI_ISL_589493, EPI_ISL_589519 to 589521, EPI_ISL_589526, EPI_ISL_589572 to 589575, EPI_ISL_589577 to 589605, EPI_ISL_589607 to 589634, EPI_ISL_589636 to 589758, EPI_ISL_589760 to 589761, EPI_ISL_589763 to 589764, EPI_ISL_589766 to 589773, EPI_ISL_589787, EPI_ISL_589789, EPI_ISL_589791 to 589792, EPI_ISL_589794 to 589801, EPI_ISL_590172, EPI_ISL_590174 to 590175, EPI_ISL_598250 to 598280, EPI_ISL_598282 to 598284, EPI_ISL_598286 to 598303, EPI_ISL_598306 to 598329, EPI_ISL_598332 to 598360, EPI_ISL_598362 to 598405, EPI_ISL_598407, EPI_ISL_598409 to 598432, EPI_ISL_598434 to 598435, EPI_ISL_598437 to 598484, EPI_ISL_598486 to 598488, EPI_ISL_598490 to 598513, EPI_ISL_598515 to 598524, EPI_ISL_598526 to 598529, EPI_ISL_598531 to 598532, EPI_ISL_598534 to 598587, EPI_ISL_598589, EPI_ISL_598591 to 598602, EPI_ISL_598604, EPI_ISL_598607 to 598614, EPI_ISL_598616 to 598637, EPI_ISL_598639 to 598666, EPI_ISL_598668 to 598686, EPI_ISL_598688 to 598706, EPI_ISL_598708 to 598795, EPI_ISL_598797 to 598820, EPI_ISL_598822 to 598823, EPI_ISL_599818 to 599820, EPI_ISL_599822 to 599831, EPI_ISL_599833, EPI_ISL_599835 to 599844, EPI_ISL_599846 to 599866, EPI_ISL_599868 to 599885, EPI_ISL_599887, EPI_ISL_599889 to 599894, EPI_ISL_599896 to 599903, EPI_ISL_599905 to 599909, EPI_ISL_599911 to 599912, EPI_ISL_599914 to 599930, EPI_ISL_599932 to 599947, EPI_ISL_599949, EPI_ISL_599951 to 599959, EPI_ISL_599961 to 599962, EPI_ISL_599964 to 599972, EPI_ISL_599974 to 599994, EPI_ISL_599996 to 600006, EPI_ISL_600008 to 600032, EPI_ISL_600034 to 600046, EPI_ISL_600048 to 600055, EPI_ISL_600057 to 600065, EPI_ISL_600067 to 600075, EPI_ISL_600077 to 600082, EPI_ISL_600084 to 600090, EPI_ISL_600092 to 600106, EPI_ISL_600108 to 600122, EPI_ISL_600124 to 600127, EPI_ISL_600892, EPI_ISL_600901, EPI_ISL_600912 to 600913, EPI_ISL_600915, EPI_ISL_600923, EPI_ISL_600925 to 600931, EPI_ISL_601224, EPI_ISL_601234 to 601235, EPI_ISL_601239 to 601240, EPI_ISL_601242 to 601243, EPI_ISL_601247 to 601249, EPI_ISL_601256, EPI_ISL_601263 to 601264, EPI_ISL_601267 to 601268, EPI_ISL_601270, EPI_ISL_601272 to 601273, EPI_ISL_601275, EPI_ISL_601277, EPI_ISL_601280 to 601283, EPI_ISL_601287, EPI_ISL_601289, EPI_ISL_601291 to 601294, EPI_ISL_601297 to 601299, EPI_ISL_601301 to 601307, EPI_ISL_601310 to 601313, EPI_ISL_601316 to 601317, EPI_ISL_601320 to 601322, EPI_ISL_601324 to 601327, EPI_ISL_601329 to 601330, EPI_ISL_601337, EPI_ISL_601340, EPI_ISL_601343, EPI_ISL_601345 to 601346, EPI_ISL_601349 to 601350, EPI_ISL_601354, EPI_ISL_601359 to 601360, EPI_ISL_601363, EPI_ISL_601365, EPI_ISL_601367 to 601371, EPI_ISL_601373, EPI_ISL_601375 to 601376, EPI_ISL_601378 to 601379, EPI_ISL_601381 to 601383, EPI_ISL_601385, EPI_ISL_601388, EPI_ISL_601390, EPI_ISL_601392, EPI_ISL_601394, EPI_ISL_601396 to 601398, EPI_ISL_601404, EPI_ISL_601406, EPI_ISL_601409 to 601410, EPI_ISL_601412, EPI_ISL_601415, EPI_ISL_601417 to 601420, EPI_ISL_601422, EPI_ISL_601427 to 601428, EPI_ISL_601431, EPI_ISL_601434, EPI_ISL_601437 to 601444, EPI_ISL_601447 to 601452, EPI_ISL_601455 to 601458, EPI_ISL_601463 to 601465, EPI_ISL_601468, EPI_ISL_601470, EPI_ISL_601472 to 601476, EPI_ISL_601478, EPI_ISL_601484 to 601485, EPI_ISL_601487, EPI_ISL_601490 to 601492, EPI_ISL_601494, EPI_ISL_601496 to 601498, EPI_ISL_601500, EPI_ISL_601502 to 601505, EPI_ISL_601507 to 601514, EPI_ISL_601516 to 601521, EPI_ISL_601523 to 601527, EPI_ISL_601529, EPI_ISL_601531 to 601540, EPI_ISL_601542 to 601546, EPI_ISL_601551 to 601554, EPI_ISL_601557 to 601566, EPI_ISL_601568, EPI_ISL_601570, EPI_ISL_601572 to 601577, EPI_ISL_601581 to 601584, EPI_ISL_601586 to 601587, EPI_ISL_601592 to 601593, EPI_ISL_601595 to 601599, EPI_ISL_601601 to 601603, EPI_ISL_601605, EPI_ISL_601607 to 601610, EPI_ISL_601612 to 601614, EPI_ISL_601616, EPI_ISL_601620 to 601621, EPI_ISL_601623, EPI_ISL_601625 to 601626, EPI_ISL_601629 to 601633, EPI_ISL_601635 to 601638, EPI_ISL_601640, EPI_ISL_601642 to 601647, EPI_ISL_601649, EPI_ISL_601651, EPI_ISL_601653 to 601656, EPI_ISL_601658 to 601662, EPI_ISL_601665, EPI_ISL_601669 to 601672, EPI_ISL_601674 to 601679, EPI_ISL_601681, EPI_ISL_601684, EPI_ISL_601686 to 601688, EPI_ISL_601692 to 601699, EPI_ISL_601701 to 601703, EPI_ISL_601705 to 601708, EPI_ISL_601710 to 601714, EPI_ISL_601716 to 601720, EPI_ISL_601722 to 601727, EPI_ISL_601729, EPI_ISL_601731 to 601732, EPI_ISL_601734 to 601739, EPI_ISL_601741 to 601746, EPI_ISL_601749 to 601756, EPI_ISL_601758 to 601763, EPI_ISL_601767, EPI_ISL_601769 to 601770, EPI_ISL_601772 to 601777, EPI_ISL_601779 to 601782, EPI_ISL_601784, EPI_ISL_601786, EPI_ISL_602027 to 602029, EPI_ISL_602145 to 602153, EPI_ISL_606296 to 606310, EPI_ISL_606312 to 606313, EPI_ISL_606315 to 606318, EPI_ISL_606320, EPI_ISL_606322 to 606352, EPI_ISL_606354 to 606357, EPI_ISL_606359 to 606369, EPI_ISL_606371 to 606378, EPI_ISL_606380 to 606401, EPI_ISL_606403 to 606414, EPI_ISL_606416 to 606436, EPI_ISL_606438 to 606461, EPI_ISL_606463 to 606515, EPI_ISL_606517 to 606523, EPI_ISL_606525 to 606587, EPI_ISL_606589 to 606593, EPI_ISL_606595 to 606605, EPI_ISL_606607, EPI_ISL_606609 to 606623, EPI_ISL_606625 to 606627, EPI_ISL_606629 to 606653, EPI_ISL_606656 to 606674, EPI_ISL_606676 to 606701, EPI_ISL_606703 to 606727, EPI_ISL_606729 to 606732, EPI_ISL_606734 to 606748, EPI_ISL_606750 to 606773, EPI_ISL_606775, EPI_ISL_606777 to 606779, EPI_ISL_606781 to 606796, EPI_ISL_606798 to 606808, EPI_ISL_606810 to 606828, EPI_ISL_606830 to 606843, EPI_ISL_606845 to 606891, EPI_ISL_606893 to 606903, EPI_ISL_606905 to 606909, EPI_ISL_606911 to 606915, EPI_ISL_606917 to 606921, EPI_ISL_606923, EPI_ISL_607292 to 607302, EPI_ISL_607304 to 607321, EPI_ISL_607323 to 607342, EPI_ISL_607344 to 607352, EPI_ISL_607355 to 607362, EPI_ISL_607364 to 607372, EPI_ISL_607374 to 607404, EPI_ISL_607406 to 607415, EPI_ISL_607417 to 607426, EPI_ISL_607428 to 607429, EPI_ISL_607431 to 607447, EPI_ISL_607449 to 607454, EPI_ISL_607456 to 607460, EPI_ISL_607462 to 607474, EPI_ISL_607476 to 607505, EPI_ISL_607507 to 607515, EPI_ISL_607517 to 607518, EPI_ISL_607520 to 607521, EPI_ISL_607523 to 607542, EPI_ISL_607544 to 607549, EPI_ISL_607551 to 607565, EPI_ISL_607567 to 607571, EPI_ISL_607573 to 607578, EPI_ISL_607580 to 607586, EPI_ISL_607588 to 607622, EPI_ISL_608697, EPI_ISL_608700 to 608747, EPI_ISL_608749 to 608765, EPI_ISL_608767 to 608833, EPI_ISL_608835 to 608844, EPI_ISL_608846, EPI_ISL_608848, EPI_ISL_608850, EPI_ISL_608854, EPI_ISL_608857, EPI_ISL_608859, EPI_ISL_608861, EPI_ISL_608870 to 608871, EPI_ISL_608882, EPI_ISL_608893 to 608894, EPI_ISL_608901, EPI_ISL_608905, EPI_ISL_608936, EPI_ISL_608940, EPI_ISL_608954, EPI_ISL_608975, EPI_ISL_608977 to 608978, EPI_ISL_608986, EPI_ISL_609001, EPI_ISL_609006, EPI_ISL_609008, EPI_ISL_609015, EPI_ISL_609017, EPI_ISL_609021, EPI_ISL_609027, EPI_ISL_609029 to 609030, EPI_ISL_609036, EPI_ISL_609038, EPI_ISL_609042, EPI_ISL_609044, EPI_ISL_609046, EPI_ISL_609049, EPI_ISL_609058 to 609059, EPI_ISL_609062 to 609063, EPI_ISL_609066, EPI_ISL_609071, EPI_ISL_609076, EPI_ISL_609084, EPI_ISL_609108, EPI_ISL_609110, EPI_ISL_609115 to 609117, EPI_ISL_609126, EPI_ISL_609128, EPI_ISL_609136 to 609140, EPI_ISL_609149, EPI_ISL_609151, EPI_ISL_609155, EPI_ISL_609219 to 609227, EPI_ISL_609229, EPI_ISL_609231 to 609253, EPI_ISL_609256 to 609266, EPI_ISL_609268 to 609290, EPI_ISL_609292 to 609293, EPI_ISL_609295 to 609301, EPI_ISL_609303 to 609307, EPI_ISL_623211 to 623213, EPI_ISL_623215 to 623219, EPI_ISL_623221, EPI_ISL_623223 to 623233, EPI_ISL_623235 to 623244, EPI_ISL_623246, EPI_ISL_623249 to 623255, EPI_ISL_623257 to 623258, EPI_ISL_623260 to 623263, EPI_ISL_623265 to 623267, EPI_ISL_623269 to 623272, EPI_ISL_623274 to 623275, EPI_ISL_623277 to 623280, EPI_ISL_623282 to 623285, EPI_ISL_623287, EPI_ISL_623289 to 623297, EPI_ISL_623299 to 623300, EPI_ISL_623302 to 623307, EPI_ISL_623309 to 623310, EPI_ISL_623314 to 623320, EPI_ISL_623322, EPI_ISL_623325 to 623329, EPI_ISL_623331 to 623332, EPI_ISL_623335 to 623336, EPI_ISL_623338 to 623342, EPI_ISL_623348 to 623358, EPI_ISL_623361 to 623362, EPI_ISL_623365, EPI_ISL_623367 to 623369, EPI_ISL_623371 to 623384, EPI_ISL_623387 to 623389, EPI_ISL_623394 to 623396, EPI_ISL_623399 to 623403, EPI_ISL_623405 to 623408, EPI_ISL_623410, EPI_ISL_623412 to 623421, EPI_ISL_623423, EPI_ISL_623429 to 623432, EPI_ISL_623434 to 623436, EPI_ISL_623438 to 623439, EPI_ISL_623441 to 623443, EPI_ISL_623446 to 623447, EPI_ISL_623450, EPI_ISL_623452 to 623455, EPI_ISL_623457 to 623473, EPI_ISL_623476, EPI_ISL_623478 to 623481, EPI_ISL_623483, EPI_ISL_623486 to 623487, EPI_ISL_623491 to 623493, EPI_ISL_623496 to 623514, EPI_ISL_623516 to 623519, EPI_ISL_623521, EPI_ISL_623523 to 623526, EPI_ISL_623529 to 623530, EPI_ISL_623533 to 623536, EPI_ISL_623538, EPI_ISL_623540 to 623543, EPI_ISL_623546, EPI_ISL_623549, EPI_ISL_623552 to 623554, EPI_ISL_623561, EPI_ISL_623563 to 623566, EPI_ISL_623571, EPI_ISL_623573, EPI_ISL_623577, EPI_ISL_623579 to 623580, EPI_ISL_623582 to 623583, EPI_ISL_623586, EPI_ISL_623588 to 623591, EPI_ISL_623594, EPI_ISL_623596 to 623599, EPI_ISL_623601, EPI_ISL_623603 to 623604, EPI_ISL_623606 to 623607, EPI_ISL_623610, EPI_ISL_623613 to 623615, EPI_ISL_623618, EPI_ISL_623620 to 623625, EPI_ISL_623627 to 623629, EPI_ISL_623631 to 623632, EPI_ISL_623634 to 623638, EPI_ISL_623642, EPI_ISL_623646, EPI_ISL_623648 to 623649, EPI_ISL_623656, EPI_ISL_623659 to 623660, EPI_ISL_623668 to 623673, EPI_ISL_623676 to 623678, EPI_ISL_623681 to 623682, EPI_ISL_623684 to 623687, EPI_ISL_623689, EPI_ISL_623691 to 623693, EPI_ISL_623695 to 623699, EPI_ISL_623701, EPI_ISL_623704 to 623706, EPI_ISL_623708 to 623709, EPI_ISL_623711, EPI_ISL_623713 to 623715, EPI_ISL_623718 to 623720, EPI_ISL_623722 to 623723, EPI_ISL_623727 to 623730, EPI_ISL_623732, EPI_ISL_623734, EPI_ISL_623737 to 623739, EPI_ISL_623743 to 623745, EPI_ISL_623747 to 623748, EPI_ISL_623752 to 623753, EPI_ISL_623755 to 623757, EPI_ISL_623760 to 623761, EPI_ISL_623765 to 623766, EPI_ISL_623768 to 623771, EPI_ISL_623773, EPI_ISL_623777 to 623780, EPI_ISL_623782 to 623784, EPI_ISL_623786 to 623787, EPI_ISL_623789, EPI_ISL_623792, EPI_ISL_623795, EPI_ISL_623801, EPI_ISL_623803 to 623805, EPI_ISL_623807, EPI_ISL_623813 to 623814, EPI_ISL_623817 to 623821, EPI_ISL_623824 to 623827, EPI_ISL_623830, EPI_ISL_623835 to 623837, EPI_ISL_623839, EPI_ISL_623842, EPI_ISL_623845, EPI_ISL_623850 to 623853, EPI_ISL_625405 to 625421, EPI_ISL_625424, EPI_ISL_625426, EPI_ISL_625428, EPI_ISL_625430, EPI_ISL_629109 to 629137, EPI_ISL_629139 to 629179, EPI_ISL_629181 to 629198, EPI_ISL_629200 to 629218, EPI_ISL_629220 to 629244, EPI_ISL_629246 to 629352, EPI_ISL_629354 to 629397, EPI_ISL_629399 to 629408, EPI_ISL_629410 to 629483, EPI_ISL_629485 to 629489, EPI_ISL_629491 to 629511, EPI_ISL_629513 to 629525, EPI_ISL_629527 to 629552, EPI_ISL_629554 to 629566, EPI_ISL_629568 to 629579, EPI_ISL_629581 to 629589, EPI_ISL_629591, EPI_ISL_629593 to 629631, EPI_ISL_629633 to 629659, EPI_ISL_629661 to 629667, EPI_ISL_629669 to 629704, EPI_ISL_629706 to 629709, EPI_ISL_629711 to 629726, EPI_ISL_629728 to 629753, EPI_ISL_630007 to 630062, EPI_ISL_630064 to 630126, EPI_ISL_630128 to 630150, EPI_ISL_630152 to 630178, EPI_ISL_630180 to 630190, EPI_ISL_630192 to 630218, EPI_ISL_630220 to 630244, EPI_ISL_630246 to 630248, EPI_ISL_630251 to 630347, EPI_ISL_630973 to 630974, EPI_ISL_630978, EPI_ISL_630980 to 630981, EPI_ISL_630986 to 630987, EPI_ISL_630989 to 630991, EPI_ISL_630993, EPI_ISL_630995, EPI_ISL_630998, EPI_ISL_631000, EPI_ISL_631002, EPI_ISL_631004, EPI_ISL_631006 to 631007, EPI_ISL_631009 to 631010, EPI_ISL_631013, EPI_ISL_631015, EPI_ISL_631019 to 631022, EPI_ISL_631024, EPI_ISL_631026, EPI_ISL_631029 to 631030, EPI_ISL_631032, EPI_ISL_631034, EPI_ISL_631036 to 631037, EPI_ISL_631040, EPI_ISL_631042 to 631043, EPI_ISL_631047 to 631049, EPI_ISL_631051 to 631052, EPI_ISL_631054, EPI_ISL_631058, EPI_ISL_631061, EPI_ISL_631064 to 631068, EPI_ISL_631070 to 631073, EPI_ISL_631075 to 631077, EPI_ISL_631079, EPI_ISL_631082 to 631083, EPI_ISL_631085 to 631088, EPI_ISL_631091 to 631092, EPI_ISL_631095, EPI_ISL_631097 to 631098, EPI_ISL_631100, EPI_ISL_631103 to 631104, EPI_ISL_631106, EPI_ISL_631108, EPI_ISL_631110, EPI_ISL_631114 to 631115, EPI_ISL_631119 to 631122, EPI_ISL_631124 to 631126, EPI_ISL_631130, EPI_ISL_631134, EPI_ISL_631136, EPI_ISL_631138 to 631141, EPI_ISL_631143, EPI_ISL_631145 to 631146, EPI_ISL_631151 to 631153, EPI_ISL_631156 to 631158, EPI_ISL_631161 to 631162, EPI_ISL_631166 to 631170, EPI_ISL_631172, EPI_ISL_631176, EPI_ISL_631178 to 631179, EPI_ISL_631182 to 631183, EPI_ISL_631186 to 631188, EPI_ISL_631193, EPI_ISL_631195 to 631198, EPI_ISL_631200 to 631201, EPI_ISL_631203, EPI_ISL_631205 to 631207, EPI_ISL_631209 to 631211, EPI_ISL_631213 to 631214, EPI_ISL_631216, EPI_ISL_631218, EPI_ISL_631223, EPI_ISL_631226 to 631227, EPI_ISL_631229 to 631230, EPI_ISL_631232 to 631234, EPI_ISL_631236, EPI_ISL_631238 to 631240, EPI_ISL_631242 to 631244, EPI_ISL_631246 to 631249, EPI_ISL_631251 to 631253, EPI_ISL_631255 to 631256, EPI_ISL_631258 to 631261, EPI_ISL_631263 to 631264, EPI_ISL_631267, EPI_ISL_631275, EPI_ISL_634256 to 634265, EPI_ISL_634267 to 634340, EPI_ISL_634342 to 634375, EPI_ISL_634377 to 634381, EPI_ISL_634383, EPI_ISL_634386 to 634405, EPI_ISL_634407 to 634427, EPI_ISL_634429 to 634464, EPI_ISL_634466 to 634482, EPI_ISL_634484 to 634498, EPI_ISL_634500 to 634519 | | | |
| see above | Lighthouse Lab in Milton Keynes | Wellcome Sanger Institute for the COVID-19 Genomics UK (COG-UK) consortium | Cordelia Langford; David K. Jackson; Dominic Kwiatkowski; Ewan Harrison; Ian Johnston; John Sillitoe on behalf of the Wellcome Sanger Institute COVID-19 Surveillance Team; John Sillitoe on behalf of the Wellcome Sanger Institute COVID-19 Surveillance Team (http://www.sanger.ac.uk/covid-team); Roberto Amato; Sonia Goncalves; The Lighthouse Lab in Alderley Park and Alex Alderton; The Lighthouse Lab in Milton Keynes and Alex Alderton |
| EPI_ISL_831373, EPI_ISL_831375 to 831378, EPI_ISL_831380 to 831381, EPI_ISL_831386 to 831389, EPI_ISL_831391, EPI_ISL_831393, EPI_ISL_831396 | | | |
| see above | Limbach - MVZ Labor Dr. Limbach & Kollegen | Robert Koch Institute, Influenza and respiratory viruses FG17 & Bioinformatics MF1, Berlin, Germany | Aleksandar Radonic; Dr. Konrad Bode; Marianne Wedde; Oliver Drechsel; Ralf Duerrwald; Rene Kmiecinski; Stefan Kroeger; Stephan Fuchs; Thorsten Wolff |
| EPI_ISL_730621, EPI_ISL_730624 to 730625, EPI_ISL_730630, EPI_ISL_730635, EPI_ISL_751191 to 751192, EPI_ISL_766586 | Limmattal Hospital | Institute of Medical Virology, University of Zurich | Alexandra Trkola; Annette Audigé; Cyril Shah; Gabriela Ziltener; Jon Huder; Jürg Böni; Kevin Steiner; Maria Grünberg; Maryam Zaheri; Michael Huber; Riccarda Capaul; Roberto Buonomano; Stefan Schmutz; Verena Kufner |
| EPI_ISL_484354 to 484366, EPI_ISL_484368 to 484373, EPI_ISL_484392 to 484406, EPI_ISL_490563 to 490580, EPI_ISL_493549 to 493558, EPI_ISL_493560 to 493600, EPI_ISL_493603 to 493605, EPI_ISL_493610 to 493613, EPI_ISL_526431 to 526432, EPI_ISL_549352, EPI_ISL_549356, EPI_ISL_549362, EPI_ISL_549365, EPI_ISL_549371 to 549372, EPI_ISL_549375, EPI_ISL_549381, EPI_ISL_549387, EPI_ISL_549517 to 549522, EPI_ISL_572507 to 572508, EPI_ISL_572510, EPI_ISL_572527, EPI_ISL_572705, EPI_ISL_572938, EPI_ISL_572943 to 572944, EPI_ISL_573437 to 573449, EPI_ISL_679946 to 679968, EPI_ISL_725540, EPI_ISL_725542 to 725545, EPI_ISL_725547 to 725604, EPI_ISL_819476, EPI_ISL_819478 to 819482, EPI_ISL_819487 to 819489, EPI_ISL_819493, EPI_ISL_819499, EPI_ISL_819503 to 819504, EPI_ISL_819506 to 819507, EPI_ISL_819520 to 819523, EPI_ISL_819529, EPI_ISL_819532, EPI_ISL_840017 to 840018, EPI_ISL_840020 to 840028, EPI_ISL_840030 to 840050, EPI_ISL_840062 to 840074, EPI_ISL_840076 to 840086, EPI_ISL_840088 to 840091, EPI_ISL_840093 to 840120, EPI_ISL_840146 to 840169, EPI_ISL_840172 to 840185, EPI_ISL_866992 to 867007, EPI_ISL_867010 | | | |
| see above | Lincolnshire Hospitals and DeepSeq Nottingham | COVID-19 Genomics UK (COG-UK) Consortium | Christopher Moore; Fei Sang; Joeseph Chappell; Johnny Debebe; Jonathan Ball; Matthew Carlisle; Matthew Loose; Nadine Holmes; Nichola Duckworth; Patrick McClure; Sarah Walsh; Tim Sloan; Victoria Wright |
| EPI_ISL_648200 to 648203 | Linkoping | The Public Health Agency of Sweden | Anna Risberg; Anna-Malin Linde; Karin Tegmark-Wisell; Maria Lind Karlberg; Mattias Haukland; Mia Brytting; Olov Svartstrom; Oskar Karlsson Lindsjo; Petra Edquist; Reza Advani; Sandra Broddesson |
| EPI_ISL_541847 to 541877 | Lithuanian University of Health Sciences Hospital, Department of Laboratory Medicine | Lithuanian University of Health Sciences, Laboratory of Molecular Cardiology | Arnoldas Pautienius; Astra Vitkauskiene; Dovydas Gecys; Kamile Tamusauskaite; Laura Pareckaite; Lukas Zemaitis; Vaiva Lesauskaite |
| EPI_ISL_536398, EPI_ISL_603082 to 603120, EPI_ISL_603122, EPI_ISL_636604, EPI_ISL_636839 to 636906, EPI_ISL_770498 to 770532, EPI_ISL_770550 | | | |
| see above | Lithuanian University of Health Sciences Hospital, Department of Laboratory Medicine | Lithuanian University of Health Sciences, Molecular cardiology lab. | Arnoldas Pautienius; Astra Vitkauskiene; Dovydas Gecys; Ingrida Olendrait; Kamile Tamusauskaite; Laura Pareckaite; Lukas Zemaitis; Vaiva Lesauskaite |
| EPI_ISL_439672 to 439697, EPI_ISL_439699 to 439724, EPI_ISL_439726 to 439731, EPI_ISL_439733 to 439738, EPI_ISL_439740 to 439741, EPI_ISL_439743 to 439745, EPI_ISL_439747 to 439772, EPI_ISL_439774 to 439832, EPI_ISL_439834 to 439845, EPI_ISL_439847 to 439862, EPI_ISL_440852 to 440906, EPI_ISL_440908 to 440937, EPI_ISL_440939 to 440950, EPI_ISL_449330 to 449335, EPI_ISL_449337 to 449344, EPI_ISL_449346 to 449351, EPI_ISL_449353 to 449358, EPI_ISL_449360 to 449361, EPI_ISL_449363 to 449374, EPI_ISL_449377 to 449384, EPI_ISL_449386 to 449389, EPI_ISL_449391 to 449400, EPI_ISL_449403, EPI_ISL_449407 to 449409, EPI_ISL_449411 to 449417, EPI_ISL_449420 to 449423, EPI_ISL_449425 to 449426, EPI_ISL_449429 to 449430, EPI_ISL_449432, EPI_ISL_449435, EPI_ISL_449438 to 449443, EPI_ISL_449447 to 449449, EPI_ISL_449451 to 449457, EPI_ISL_449459 to 449460, EPI_ISL_449462 to 449464, EPI_ISL_449467 to 449469, EPI_ISL_449471 to 449475, EPI_ISL_449488 to 449492, EPI_ISL_449494 to 449519, EPI_ISL_449521 to 449534, EPI_ISL_449536 to 449538, EPI_ISL_449540, EPI_ISL_449542 to 449559, EPI_ISL_449561 to 449571, EPI_ISL_449573, EPI_ISL_449576 to 449578, EPI_ISL_449580 to 449588, EPI_ISL_449590, EPI_ISL_449593 to 449596, EPI_ISL_449598 to 449625, EPI_ISL_453195 to 453199, EPI_ISL_453201 to 453211, EPI_ISL_453213, EPI_ISL_453215 to 453223, EPI_ISL_453225 to 453256, EPI_ISL_453258 to 453269, EPI_ISL_453271 to 453272, EPI_ISL_453276 to 453280, EPI_ISL_453282, EPI_ISL_453284, EPI_ISL_453288 to 453302, EPI_ISL_453304 to 453306, EPI_ISL_453308 to 453325, EPI_ISL_453327 to 453346, EPI_ISL_453348 to 453354, EPI_ISL_453357 to 453370, EPI_ISL_453372 to 453388, EPI_ISL_453390 to 453417, EPI_ISL_453419 to 453428, EPI_ISL_453430 to 453459, EPI_ISL_457260 to 457269, EPI_ISL_471993 to 472028, EPI_ISL_472030 to 472034, EPI_ISL_472037 to 472067, EPI_ISL_472069, EPI_ISL_472071 to 472084, EPI_ISL_472087 to 472088, EPI_ISL_472090 to 472135, EPI_ISL_478372 to 478375, EPI_ISL_478377 to 478403, EPI_ISL_484254 to 484263, EPI_ISL_490331 to 490351, EPI_ISL_490353 to 490357, EPI_ISL_490359 to 490379, EPI_ISL_490382 to 490390, EPI_ISL_490393 to 490420, EPI_ISL_499460 to 499471, EPI_ISL_499473, EPI_ISL_499477, EPI_ISL_499480 to 499481, EPI_ISL_499488, EPI_ISL_499491 to 499492, EPI_ISL_499496 to 499501, EPI_ISL_499509, EPI_ISL_499511 to 499514, EPI_ISL_499516 to 499545, EPI_ISL_499557, EPI_ISL_499564 to 499567, EPI_ISL_499569 to 499571, EPI_ISL_499573 to 499586, EPI_ISL_499589 to 499590, EPI_ISL_499593 to 499594, EPI_ISL_499600, EPI_ISL_499604 to 499605, EPI_ISL_499607 to 499608, EPI_ISL_499611 to 499612, EPI_ISL_499616 to 499622, EPI_ISL_499624, | | | |

EPI_ISL_499627 to 499630, EPI_ISL_499632 to 499635, EPI_ISL_499637 to 499704, EPI_ISL_499706 to 499754, EPI_ISL_499756 to 499776, EPI_ISL_499811 to 499827, EPI_ISL_499832 to 499838, EPI_ISL_499840 to 499850, EPI_ISL_499854, EPI_ISL_499856 to 499857, EPI_ISL_499859 to 499860, EPI_ISL_499862 to 499868, EPI_ISL_499870 to 499880, EPI_ISL_499882 to 499893, EPI_ISL_499896, EPI_ISL_499898, EPI_ISL_499900 to 499904, EPI_ISL_499906 to 499907, EPI_ISL_499909, EPI_ISL_499911, EPI_ISL_499914 to 499922, EPI_ISL_499924 to 499928, EPI_ISL_499930 to 499932, EPI_ISL_499934 to 499946, EPI_ISL_499948 to 499949, EPI_ISL_499951 to 499958, EPI_ISL_499960 to 499965, EPI_ISL_499967, EPI_ISL_499984 to 500101, EPI_ISL_500103 to 500144, EPI_ISL_500146 to 500156, EPI_ISL_512342 to 512345, EPI_ISL_512347, EPI_ISL_517001 to 517008, EPI_ISL_517010 to 517014, EPI_ISL_517016 to 517025, EPI_ISL_517027 to 517053, EPI_ISL_517055 to 517061, EPI_ISL_517063 to 517148, EPI_ISL_517150 to 517229, EPI_ISL_517231 to 517280, EPI_ISL_517282 to 517308, EPI_ISL_517310 to 517354, EPI_ISL_517356 to 517371, EPI_ISL_517373 to 517394, EPI_ISL_517396 to 517435, EPI_ISL_517437 to 517462, EPI_ISL_517464 to 517487, EPI_ISL_517489 to 517490, EPI_ISL_517492 to 517498, EPI_ISL_517500 to 517509, EPI_ISL_526368 to 526371, EPI_ISL_526373 to 526379, EPI_ISL_526381 to 526384, EPI_ISL_526386 to 526394, EPI_ISL_529254, EPI_ISL_529258 to 529260, EPI_ISL_529274, EPI_ISL_529280, EPI_ISL_529284, EPI_ISL_529296, EPI_ISL_529305, EPI_ISL_529466, EPI_ISL_529484, EPI_ISL_529498, EPI_ISL_529510, EPI_ISL_529516, EPI_ISL_534732, EPI_ISL_534734 to 534743, EPI_ISL_534746, EPI_ISL_534748 to 534755, EPI_ISL_540584 to 540625, EPI_ISL_540627 to 540639, EPI_ISL_549354, EPI_ISL_572402, EPI_ISL_572404, EPI_ISL_572426, EPI_ISL_572429, EPI_ISL_572431, EPI_ISL_572436, EPI_ISL_572443, EPI_ISL_572445, EPI_ISL_572451, EPI_ISL_572466, EPI_ISL_572473, EPI_ISL_572479, EPI_ISL_572500, EPI_ISL_572502, EPI_ISL_572506, EPI_ISL_572511, EPI_ISL_572552, EPI_ISL_572554, EPI_ISL_572557, EPI_ISL_572562, EPI_ISL_572578, EPI_ISL_572584, EPI_ISL_572588, EPI_ISL_572601, EPI_ISL_572604, EPI_ISL_572650, EPI_ISL_572669, EPI_ISL_572677, EPI_ISL_572679, EPI_ISL_572682, EPI_ISL_572687 to 572688, EPI_ISL_572691, EPI_ISL_572696, EPI_ISL_572710, EPI_ISL_572733 to 572734, EPI_ISL_572736 to 572739, EPI_ISL_572748, EPI_ISL_572750, EPI_ISL_572769, EPI_ISL_572771, EPI_ISL_572773, EPI_ISL_572813, EPI_ISL_572815, EPI_ISL_572817, EPI_ISL_572822, EPI_ISL_572824, EPI_ISL_572827, EPI_ISL_572843, EPI_ISL_572904 to 572908, EPI_ISL_573236 to 573246, EPI_ISL_576962 to 576965, EPI_ISL_576967 to 576968, EPI_ISL_576970 to 576973, EPI_ISL_576976 to 576977, EPI_ISL_576980 to 576981, EPI_ISL_584592, EPI_ISL_584595 to 584603, EPI_ISL_584605 to 584621, EPI_ISL_594849 to 594851, EPI_ISL_594856 to 594860, EPI_ISL_611514 to 611517, EPI_ISL_611521 to 611522, EPI_ISL_611542 to 611543, EPI_ISL_611565, EPI_ISL_611567 to 611571, EPI_ISL_611575, EPI_ISL_611579 to 611580, EPI_ISL_611600, EPI_ISL_611603, EPI_ISL_611613, EPI_ISL_611621, EPI_ISL_611627, EPI_ISL_611629 to 611630, EPI_ISL_611632, EPI_ISL_611634 to 611635, EPI_ISL_611642, EPI_ISL_611653 to 611654, EPI_ISL_611658, EPI_ISL_611664, EPI_ISL_611667, EPI_ISL_611672, EPI_ISL_611675, EPI_ISL_611694 to 611695, EPI_ISL_611721, EPI_ISL_611723, EPI_ISL_611727 to 611731, EPI_ISL_611734, EPI_ISL_611736 to 611737, EPI_ISL_611739, EPI_ISL_611741, EPI_ISL_611743, EPI_ISL_611764 to 611765, EPI_ISL_611784 to 611786, EPI_ISL_611790, EPI_ISL_611796, EPI_ISL_611811, EPI_ISL_611815, EPI_ISL_611821, EPI_ISL_611827, EPI_ISL_611845, EPI_ISL_611851, EPI_ISL_611853, EPI_ISL_611857, EPI_ISL_611885, EPI_ISL_611889, EPI_ISL_611891 to 611893, EPI_ISL_611895, EPI_ISL_611897 to 611899, EPI_ISL_611908, EPI_ISL_611910 to 611911, EPI_ISL_611914 to 611915, EPI_ISL_611918, EPI_ISL_611921, EPI_ISL_611929, EPI_ISL_611974, EPI_ISL_611978, EPI_ISL_611985, EPI_ISL_611995, EPI_ISL_611999, EPI_ISL_612001, EPI_ISL_612003, EPI_ISL_612016 to 612017, EPI_ISL_612021, EPI_ISL_612024, EPI_ISL_612035 to 612036, EPI_ISL_612039, EPI_ISL_612041, EPI_ISL_612043 to 612044, EPI_ISL_612046, EPI_ISL_612051, EPI_ISL_612059 to 612061, EPI_ISL_612065 to 612066, EPI_ISL_612070 to 612072, EPI_ISL_612075, EPI_ISL_612080, EPI_ISL_612101, EPI_ISL_612108, EPI_ISL_612111 to 612112, EPI_ISL_612114 to 612115, EPI_ISL_612190 to 612191, EPI_ISL_612401 to 612449, EPI_ISL_626712, EPI_ISL_626736, EPI_ISL_626742, EPI_ISL_626760, EPI_ISL_626766, EPI_ISL_626789, EPI_ISL_626807, EPI_ISL_626815, EPI_ISL_626832, EPI_ISL_626866, EPI_ISL_626994, EPI_ISL_627004, EPI_ISL_627017, EPI_ISL_627106, EPI_ISL_627141, EPI_ISL_627153, EPI_ISL_627156, EPI_ISL_627162, EPI_ISL_627178, EPI_ISL_627445 to 627448, EPI_ISL_637257, EPI_ISL_637259, EPI_ISL_637262 to 637263, EPI_ISL_637274, EPI_ISL_637295, EPI_ISL_637306, EPI_ISL_637313, EPI_ISL_637323, EPI_ISL_637325, EPI_ISL_637336 to 637339, EPI_ISL_637390, EPI_ISL_637401, EPI_ISL_637434, EPI_ISL_637438 to 637439, EPI_ISL_637443 to 637444, EPI_ISL_637617, EPI_ISL_637908, EPI_ISL_637926, EPI_ISL_637929 to 637930, EPI_ISL_637934, EPI_ISL_637948, EPI_ISL_637959, EPI_ISL_637961, EPI_ISL_637965, EPI_ISL_637972, EPI_ISL_637976, EPI_ISL_637981, EPI_ISL_637989 to 637990, EPI_ISL_637993, EPI_ISL_638015, EPI_ISL_638021, EPI_ISL_638023, EPI_ISL_638025, EPI_ISL_638449 to 638453, EPI_ISL_639002, EPI_ISL_650112, EPI_ISL_650114, EPI_ISL_650132, EPI_ISL_650140, EPI_ISL_650160 to 650162, EPI_ISL_650166, EPI_ISL_650183 to 650184, EPI_ISL_650189, EPI_ISL_650194, EPI_ISL_650196, EPI_ISL_650198 to 650199, EPI_ISL_650202 to 650203, EPI_ISL_650205, EPI_ISL_650207, EPI_ISL_650242, EPI_ISL_650257, EPI_ISL_650268 to 650269, EPI_ISL_650272, EPI_ISL_650278, EPI_ISL_650284, EPI_ISL_650290, EPI_ISL_650299, EPI_ISL_650315, EPI_ISL_650318, EPI_ISL_650321, EPI_ISL_650324, EPI_ISL_650326, EPI_ISL_650331, EPI_ISL_650333, EPI_ISL_650336 to 650337, EPI_ISL_650341 to 650342, EPI_ISL_650344, EPI_ISL_650346, EPI_ISL_650366, EPI_ISL_650381 to 650382, EPI_ISL_650419 to 650420, EPI_ISL_650440, EPI_ISL_650448, EPI_ISL_650453, EPI_ISL_650473, EPI_ISL_650479, EPI_ISL_650486, EPI_ISL_650496, EPI_ISL_650515, EPI_ISL_650527, EPI_ISL_650529, EPI_ISL_650532 to 650533, EPI_ISL_650537, EPI_ISL_650539 to 650540, EPI_ISL_650544, EPI_ISL_650547, EPI_ISL_650560 to 650561, EPI_ISL_650563, EPI_ISL_650619 to 650623, EPI_ISL_650654, EPI_ISL_650656 to 650658, EPI_ISL_650671 to 650672, EPI_ISL_650688, EPI_ISL_650694, EPI_ISL_650710, EPI_ISL_650714, EPI_ISL_650755, EPI_ISL_650782, EPI_ISL_650801, EPI_ISL_650831, EPI_ISL_650847, EPI_ISL_650877, EPI_ISL_650881 to 650882, EPI_ISL_650886 to 650887, EPI_ISL_650889, EPI_ISL_650891, EPI_ISL_650893, EPI_ISL_650897, EPI_ISL_650904, EPI_ISL_650907 to 650909, EPI_ISL_650920 to 650921, EPI_ISL_650924, EPI_ISL_650926, EPI_ISL_650928, EPI_ISL_650932 to 650936, EPI_ISL_650939, EPI_ISL_650945 to 650946, EPI_ISL_651095, EPI_ISL_651114 to 651115, EPI_ISL_651129, EPI_ISL_651131, EPI_ISL_651135, EPI_ISL_651138, EPI_ISL_651145, EPI_ISL_651149, EPI_ISL_651153, EPI_ISL_651163, EPI_ISL_651165, EPI_ISL_651175, EPI_ISL_651199, EPI_ISL_651205, EPI_ISL_651208, EPI_ISL_651212, EPI_ISL_651219, EPI_ISL_651225, EPI_ISL_651236, EPI_ISL_651239, EPI_ISL_651245, EPI_ISL_651248, EPI_ISL_651251, EPI_ISL_651259, EPI_ISL_651270 to 651272, EPI_ISL_651295 to 651297, EPI_ISL_651307, EPI_ISL_651311, EPI_ISL_651319, EPI_ISL_651325, EPI_ISL_651334, EPI_ISL_651336, EPI_ISL_651344, EPI_ISL_651358, EPI_ISL_651364, EPI_ISL_651368, EPI_ISL_651375, EPI_ISL_651391, EPI_ISL_651396, EPI_ISL_651403, EPI_ISL_651407, EPI_ISL_651429 to 651430, EPI_ISL_651499 to 651500, EPI_ISL_652073 to 652081, EPI_ISL_652083 to 652102, EPI_ISL_664111, EPI_ISL_664113, EPI_ISL_664134, EPI_ISL_664138, EPI_ISL_664142, EPI_ISL_664146, EPI_ISL_664151, EPI_ISL_664164, EPI_ISL_664203 to 664204, EPI_ISL_664219 to 664220, EPI_ISL_664223, EPI_ISL_664229 to 664230, EPI_ISL_664278, EPI_ISL_664299, EPI_ISL_664332 to 664333, EPI_ISL_664336, EPI_ISL_664338 to 664339, EPI_ISL_664341, EPI_ISL_664343, EPI_ISL_664345, EPI_ISL_664348, EPI_ISL_664350, EPI_ISL_664354 to 664355, EPI_ISL_664362 to 664364, EPI_ISL_664368, EPI_ISL_664424, EPI_ISL_664430, EPI_ISL_664435 to 664436, EPI_ISL_664440, EPI_ISL_664443 to 664445, EPI_ISL_664448, EPI_ISL_664468 to 664469, EPI_ISL_664475, EPI_ISL_664478, EPI_ISL_664480 to 664481, EPI_ISL_664487, EPI_ISL_664489, EPI_ISL_664496, EPI_ISL_664501, EPI_ISL_664511, EPI_ISL_664513, EPI_ISL_664519, EPI_ISL_664523 to 664524, EPI_ISL_664539, EPI_ISL_664606, EPI_ISL_664613, EPI_ISL_664619, EPI_ISL_664633, EPI_ISL_664643, EPI_ISL_664648 to 664649, EPI_ISL_664652 to 664654, EPI_ISL_664663 to 664665, EPI_ISL_664667, EPI_ISL_664671, EPI_ISL_664675, EPI_ISL_664678, EPI_ISL_664681, EPI_ISL_664692, EPI_ISL_664695, EPI_ISL_664705, EPI_ISL_664710, EPI_ISL_664726, EPI_ISL_664730, EPI_ISL_664733, EPI_ISL_664738, EPI_ISL_665113 to 665119, EPI_ISL_665121 to 665135, EPI_ISL_665137 to 665142, EPI_ISL_665696, EPI_ISL_665704, EPI_ISL_666022, EPI_ISL_679384 to 679388, EPI_ISL_679390 to 679395, EPI_ISL_702471, EPI_ISL_702498, EPI_ISL_702510, EPI_ISL_702512, EPI_ISL_702515, EPI_ISL_702523, EPI_ISL_702536, EPI_ISL_702541, EPI_ISL_702556, EPI_ISL_702566, EPI_ISL_702569, EPI_ISL_702571, EPI_ISL_702574, EPI_ISL_702577, EPI_ISL_702587, EPI_ISL_702591, EPI_ISL_702597, EPI_ISL_702600, EPI_ISL_702605, EPI_ISL_702612, EPI_ISL_702615, EPI_ISL_702623, EPI_ISL_702631, EPI_ISL_702636, EPI_ISL_702639, EPI_ISL_702642, EPI_ISL_702644, EPI_ISL_702647, EPI_ISL_702649, EPI_ISL_702656, EPI_ISL_702664, EPI_ISL_702667, EPI_ISL_702672, EPI_ISL_702674, EPI_ISL_702677, EPI_ISL_702684, EPI_ISL_702697, EPI_ISL_702708, EPI_ISL_702710, EPI_ISL_702718, EPI_ISL_702721, EPI_ISL_702726, EPI_ISL_702729, EPI_ISL_702739, EPI_ISL_702753, EPI_ISL_702756, EPI_ISL_702758, EPI_ISL_702766, EPI_ISL_702769, EPI_ISL_702774, EPI_ISL_702779, EPI_ISL_702792, EPI_ISL_702801, EPI_ISL_702806, EPI_ISL_702838, EPI_ISL_702845, EPI_ISL_702849, EPI_ISL_702854, EPI_ISL_702856, EPI_ISL_702866, EPI_ISL_702869, EPI_ISL_702872, EPI_ISL_702874, EPI_ISL_702877, EPI_ISL_702884, EPI_ISL_702887 to 702888, EPI_ISL_702894, EPI_ISL_702897, EPI_ISL_702907, EPI_ISL_702913, EPI_ISL_702917, EPI_ISL_702920, EPI_ISL_702922, EPI_ISL_702928, EPI_ISL_702932, EPI_ISL_702935, EPI_ISL_702939, EPI_ISL_702947, EPI_ISL_702953, EPI_ISL_702956, EPI_ISL_702966, EPI_ISL_702974, EPI_ISL_702980, EPI_ISL_703023, EPI_ISL_703043, EPI_ISL_703046, EPI_ISL_703051, EPI_ISL_703059, EPI_ISL_703061, EPI_ISL_703080, EPI_ISL_703085, EPI_ISL_703090, EPI_ISL_703102, EPI_ISL_703110, EPI_ISL_703121, EPI_ISL_703147, EPI_ISL_703150, EPI_ISL_703158, EPI_ISL_703162, EPI_ISL_703167, EPI_ISL_703170, EPI_ISL_703174, EPI_ISL_703176, EPI_ISL_703182, EPI_ISL_703184, EPI_ISL_703195, EPI_ISL_703206, EPI_ISL_703208, EPI_ISL_703216, EPI_ISL_703219, EPI_ISL_703230, EPI_ISL_703235, EPI_ISL_703254, EPI_ISL_703256, EPI_ISL_703272, EPI_ISL_703275, EPI_ISL_703281, EPI_ISL_703284, EPI_ISL_703289, EPI_ISL_703305, EPI_ISL_703308, EPI_ISL_703310, EPI_ISL_703313, EPI_ISL_703316, EPI_ISL_703324, EPI_ISL_703348, EPI_ISL_703361, EPI_ISL_703364, EPI_ISL_703367, EPI_ISL_703372, EPI_ISL_703375, EPI_ISL_703387, EPI_ISL_703403, EPI_ISL_703409, EPI_ISL_703412, EPI_ISL_703417, EPI_ISL_703421, EPI_ISL_703427, EPI_ISL_703481, EPI_ISL_703527, EPI_ISL_703530, EPI_ISL_703577, EPI_ISL_703596, EPI_ISL_703611, EPI_ISL_703624, EPI_ISL_703652, EPI_ISL_703663, EPI_ISL_703690, EPI_ISL_703695, EPI_ISL_703698, EPI_ISL_703711 to 703712, EPI_ISL_703725 to 703726, EPI_ISL_703729 to 703732, EPI_ISL_703734 to 703735, EPI_ISL_703738, EPI_ISL_703744, EPI_ISL_703759, EPI_ISL_703762, EPI_ISL_703770, EPI_ISL_703774, EPI_ISL_703807, EPI_ISL_703815, EPI_ISL_703818, EPI_ISL_703821, EPI_ISL_703823, EPI_ISL_703826, EPI_ISL_703847, EPI_ISL_703850, EPI_ISL_703876, EPI_ISL_703881, EPI_ISL_703884, EPI_ISL_703888, EPI_ISL_703891, EPI_ISL_703897, EPI_ISL_703913, EPI_ISL_703916, EPI_ISL_703935, EPI_ISL_703937, EPI_ISL_703952, EPI_ISL_703958, EPI_ISL_703961, EPI_ISL_703971, EPI_ISL_703977, EPI_ISL_703990, EPI_ISL_703992, EPI_ISL_703995, EPI_ISL_704003, EPI_ISL_704006, EPI_ISL_704009, EPI_ISL_704012, EPI_ISL_704020, EPI_ISL_704022, EPI_ISL_704028, EPI_ISL_704033, EPI_ISL_704049, EPI_ISL_704057, EPI_ISL_704062, EPI_ISL_704065, EPI_ISL_704068, EPI_ISL_704079, EPI_ISL_704097, EPI_ISL_704108, EPI_ISL_704129, EPI_ISL_704131, EPI_ISL_704134, EPI_ISL_704137, EPI_ISL_704153, EPI_ISL_704156, EPI_ISL_704201, EPI_ISL_704209, EPI_ISL_704231, EPI_ISL_704260, EPI_ISL_704562, EPI_ISL_704895, EPI_ISL_704897, EPI_ISL_704900, EPI_ISL_704903, EPI_ISL_704905, EPI_ISL_704934, EPI_ISL_704947, EPI_ISL_704970, EPI_ISL_704973, EPI_ISL_704983, EPI_ISL_704986, EPI_ISL_704989, EPI_ISL_705013, EPI_ISL_705021, EPI_ISL_705024, EPI_ISL_705038, EPI_ISL_705040, EPI_ISL_705056, EPI_ISL_705059, EPI_ISL_705062, EPI_ISL_705064, EPI_ISL_705075, EPI_ISL_705150, EPI_ISL_705168, EPI_ISL_705176, EPI_ISL_705182, EPI_ISL_705190, EPI_ISL_705193, EPI_ISL_705198, EPI_ISL_705206, EPI_ISL_705208, EPI_ISL_705227, EPI_ISL_705231, EPI_ISL_705240, EPI_ISL_705242, EPI_ISL_705246, EPI_ISL_705248, EPI_ISL_705252, EPI_ISL_705258 to 705260, EPI_ISL_705263 to 705264, EPI_ISL_705267, EPI_ISL_705269, EPI_ISL_705271, EPI_ISL_705274 to 705275, EPI_ISL_705284, EPI_ISL_705286 to 705287, EPI_ISL_705292, EPI_ISL_705298 to 705299, EPI_ISL_705301, EPI_ISL_705304 to 705305, EPI_ISL_705312 to 705315, EPI_ISL_705318, EPI_ISL_705321 to 705324, EPI_ISL_705330, EPI_ISL_705334, EPI_ISL_705340, EPI_ISL_705344, EPI_ISL_705347 to 705348, EPI_ISL_705354, EPI_ISL_705360, EPI_ISL_705362 to 705363, EPI_ISL_705372, EPI_ISL_705377, EPI_ISL_705386 to 705388, EPI_ISL_705393, EPI_ISL_705398, EPI_ISL_705402, EPI_ISL_705404, EPI_ISL_705406 to 705407, EPI_ISL_705411, EPI_ISL_705423 to 705424, EPI_ISL_705446 to 705449, EPI_ISL_705471, EPI_ISL_705525, EPI_ISL_705806 to 705847, EPI_ISL_705851 to 705855, EPI_ISL_705858 to 705892, EPI_ISL_705894 to 705936, EPI_ISL_705938 to 705941, EPI_ISL_724347, EPI_ISL_724349, EPI_ISL_724351, EPI_ISL_724353 to 724372, EPI_ISL_724374 to 724375, EPI_ISL_724377 to 724380, EPI_ISL_724382 to 724384, EPI_ISL_724386 to 724392, EPI_ISL_724394, EPI_ISL_724396 to 724399, EPI_ISL_724401, EPI_ISL_724403 to 724439, EPI_ISL_724441 to 724444, EPI_ISL_724446 to 724455, EPI_ISL_724457 to 724458, EPI_ISL_724460 to 724507, EPI_ISL_724509 to 724526, EPI_ISL_724528 to 724531, EPI_ISL_724533 to 724536, EPI_ISL_724538 to 724539, EPI_ISL_813657, EPI_ISL_813659 to 813662, EPI_ISL_813666, EPI_ISL_813668, EPI_ISL_813671 to 813677, EPI_ISL_813679 to 813690, EPI_ISL_813692 to 813704, EPI_ISL_813706, EPI_ISL_813712, EPI_ISL_813715, EPI_ISL_813720 to 813721, EPI_ISL_813723, EPI_ISL_813725 to 813726, EPI_ISL_813729 to 813732, EPI_ISL_813734, EPI_ISL_813736 to 813738, EPI_ISL_813740, EPI_ISL_813742 to 813746, EPI_ISL_813748 to 813756, EPI_ISL_813758 to 813762, EPI_ISL_813764 to 813767, EPI_ISL_813769, EPI_ISL_813772, EPI_ISL_813776 to 813777, EPI_ISL_813781, EPI_ISL_813783, EPI_ISL_813785, EPI_ISL_813790, EPI_ISL_813794, EPI_ISL_813798, EPI_ISL_813801, EPI_ISL_813803 to 813808, EPI_ISL_813811, EPI_ISL_813813, EPI_ISL_813815, EPI_ISL_814286, EPI_ISL_814293, EPI_ISL_814552, EPI_ISL_814557, EPI_ISL_838353 to 838378, EPI_ISL_838380 to 838390, EPI_ISL_838392 to 838396, EPI_ISL_838398 to 838412, EPI_ISL_838414 to 838435, EPI_ISL_838437 to 838441, EPI_ISL_838443 to 838466, EPI_ISL_838468 to 838473, EPI_ISL_838475 to 838503, EPI_ISL_838505 to 838514, EPI_ISL_838516 to 838576, EPI_ISL_838578 to 838581, EPI_ISL_838583, EPI_ISL_838585, EPI_ISL_838589 to 838591, EPI_ISL_838594 to 838596, EPI_ISL_838601, EPI_ISL_838604, EPI_ISL_838606 to 838609, EPI_ISL_838611, EPI_ISL_838615 to 838616, EPI_ISL_838621, EPI_ISL_838623 to 838624, EPI_ISL_838629 to 838631, EPI_ISL_838634 to 838637, EPI_ISL_838639, EPI_ISL_838642 to 838644, EPI_ISL_838646 to 838651, EPI_ISL_838654, EPI_ISL_838656 to 838659, EPI_ISL_838661, EPI_ISL_838663 to 838664, EPI_ISL_838666, EPI_ISL_838672, EPI_ISL_865164 to 865169, EPI_ISL_865171, EPI_ISL_865174, EPI_ISL_865176 to 865177, EPI_ISL_865207 to 865259, EPI_ISL_865261 to 865269, EPI_ISL_865278, EPI_ISL_865354 to 865363, EPI_ISL_865371 to 865372, EPI_ISL_865374 to 865382, EPI_ISL_865386, EPI_ISL_865393 to 865407, EPI_ISL_865410 to 865411, EPI_ISL_865415 to 865429, EPI_ISL_865431 to 865432, EPI_ISL_865434 to 865448, EPI_ISL_865451 to 865453, EPI_ISL_865455, EPI_ISL_865458, EPI_ISL_865460 to 865461, EPI_ISL_865464 to 865465

| | | | |
|---|---|---|---|
| see above | Liverpool Clinical Laboratories | COVID-19 Genomics UK (COG-UK) Consortium | A Alrezaihi; Alessandro Gerada; Alistair Darby; Angela Cowell; Anita Lucaci; Anu Chawla; Cassie Olateju; Catherine Hartley; Charlotte Nelson; Ecaterina Vamos; Elaine O'Toole; Elaine O'ÄôToole; Eleanor G Bentley; Ghada T Shawli; Isabel García-Dorival; Isabel Garca-Dorival; James Johnson; James P Stewart; Jenifer Manson; Joanne Watts; Jones Benjamin; Jordan J Clark; Julian Hiscox; L Luu; Lucille Rainbow; M Almsaud; Margaret Hughes; Mark Whitehead; Matthew Gemmell; Miren Iturriza-Gomara; Muhannad Alruwaili; N.P Randle; Neil Swainston; PKF Gilmore; Parul Sharma; Rebekah Penrice-Randal; Rebekah Penrice-Randal¬†; Richard Eccles; Richard Gregory; Sam Haldenby; Steve Paterson; Stuart D Armstrong; Trevor Ian Robinson; Ximeng Han |
| EPI_ISL_434658 | Lundens VC | The Public Health Agency of Sweden | Anna Risberg; Anna-Malin Linde; Karin Tegmark-Wisell; Maria Lind Karlberg; Marita Dagner; Mia Brytting; Olov Svartstrom; Oskar Karlsson Lindsjo; Theresa Enkirch |

EPI_ISL_568877 to 568933, EPI_ISL_568935 to 568936, EPI_ISL_568939 to 568944, EPI_ISL_568947 to 568984, EPI_ISL_568986, EPI_ISL_568990 to 569037, EPI_ISL_569040 to 569045, EPI_ISL_569047 to 569082, EPI_ISL_569084 to 569093, EPI_ISL_569095 to 569134, EPI_ISL_569136 to 569149, EPI_ISL_569151 to 569167, EPI_ISL_569169 to 569242, EPI_ISL_569244 to 569286, EPI_ISL_569288 to 569291, EPI_ISL_569293 to 569305, EPI_ISL_569307 to 569319, EPI_ISL_569321, EPI_ISL_569323 to 569348, EPI_ISL_569350 to 569352, EPI_ISL_569354 to 569375, EPI_ISL_569377 to 569384, EPI_ISL_569386 to 569390, EPI_ISL_569392 to 569408, EPI_ISL_569410 to 569451, EPI_ISL_569453, EPI_ISL_569457 to 569470, EPI_ISL_569472 to 569479, EPI_ISL_569481, EPI_ISL_569483 to 569486, EPI_ISL_569488 to 569516, EPI_ISL_569518 to 569533, EPI_ISL_569535 to 569538, EPI_ISL_569541 to 569546, EPI_ISL_569548 to 569580, EPI_ISL_569582 to 569584, EPI_ISL_569586 to 569608, EPI_ISL_644375 to 644376, EPI_ISL_644378 to 644384, EPI_ISL_644387 to 644388, EPI_ISL_644390 to 644396, EPI_ISL_644398 to 644424, EPI_ISL_644426 to 644438, EPI_ISL_644440 to 644454, EPI_ISL_644456 to 644465, EPI_ISL_644467 to 644497, EPI_ISL_644499 to 644508, EPI_ISL_644510 to 644518, EPI_ISL_644520 to 644521, EPI_ISL_644523 to 644535, EPI_ISL_644537 to 644548, EPI_ISL_644550 to 644561, EPI_ISL_644563 to 644564, EPI_ISL_804467 to 804476, EPI_ISL_804478 to 804560, EPI_ISL_804563, EPI_ISL_804565 to 804585, EPI_ISL_804587 to 804591, EPI_ISL_860093 to 860094

| | | | |
|---|---|---|---|
| see above | MEPHI, Aix Marseille University | MEPHI, Aix Marseille University | Anthony LEVASSEUR; Anthony Levasseur |
| EPI_ISL_413571 | MHC Brabant Zuidoost | Erasmus Medical Center | Anne van der Linden; Annemiek van der Eijk; Aura Timen; Bas Oude Munnink; Claudia Schapendonk; Corien Swaan; Corine GeurtsvanKessel; David Nieuwenhuijse; Irina Chestakova; Jeroen van Kampen; Jolanda Voermans; Madelief Mollers; Manon Haverkate; Marion Koopmans; Mark Pronk; Mart Stein; Pascal Lexmond; Reina Sikkema; Richard Molenkamp; Sandra Kengne Kamga Mobou; on behalf of the Dutch national COVID-19 response team. |
| EPI_ISL_413568 | MHC Drente | Erasmus Medical Center | Anne van der Linden; Annemiek van der Eijk; Aura Timen; Bas Oude Munnink; Claudia Schapendonk; Corien Swaan; Corine GeurtsvanKessel; David Nieuwenhuijse; Irina Chestakova; Jeroen van Kampen; Jolanda Voermans; Madelief Mollers; Manon Haverkate; Marion Koopmans; Mark Pronk; Mart Stein; Pascal Lexmond; Reina Sikkema; Richard Molenkamp; Sandra Kengne Kamga Mobou; on behalf of the Dutch national COVID-19 response team. |
| EPI_ISL_413591 | MHC Flevoland | Erasmus Medical Center | Anne van der Linden; Annemiek van der Eijk; Aura Timen; Bas Oude Munnink; Claudia Schapendonk; Corien Swaan; Corine GeurtsvanKessel; David Nieuwenhuijse; Irina Chestakova; Jeroen van Kampen; Jolanda Voermans; Madelief Mollers; Manon Haverkate; Marion Koopmans; Mark Pronk; Mart Stein; Pascal Lexmond; Reina Sikkema; Richard Molenkamp; Sandra Kengne Kamga Mobou; on behalf of the Dutch national COVID-19 response team. |
| EPI_ISL_413566, EPI_ISL_413577 | MHC Gooi & Vechtstreek | Erasmus Medical Center | Anne van der Linden; Annemiek van der Eijk; Aura Timen; Bas Oude Munnink; Claudia Schapendonk; Corien Swaan; Corine GeurtsvanKessel; David Nieuwenhuijse; Irina Chestakova; Jeroen van Kampen; Jolanda Voermans; Madelief Mollers; Manon Haverkate; Marion Koopmans; Mark Pronk; Mart |

| | | | |
|---|---|---|---|
| | | | Stein; Pascal Lexmond; Reina Sikkema; Richard Molenkamp; Sandra Kengne Kamga Mobou; on behalf of the Dutch national COVID-19 response team. |
| EPI_ISL_413579 | MHC Haaglanden | Erasmus Medical Center | Anne van der Linden; Annemiek van der Eijk; Aura Timen; Bas Oude Munnink; Claudia Schapendonk; Corien Swaan; Corine GeurtsvanKessel; David Nieuwenhuijse; Irina Chestakova; Jeroen van Kampen; Jolanda Voermans; Madelief Mollers; Manon Haverkate; Marion Koopmans; Mark Pronk; Mart Stein; Pascal Lexmond; Reina Sikkema; Richard Molenkamp; Sandra Kengne Kamga Mobou; on behalf of the Dutch national COVID-19 response team. |
| EPI_ISL_413580 | MHC Hart voor Brabant | Erasmus Medical Center | Anne van der Linden; Annemiek van der Eijk; Aura Timen; Bas Oude Munnink; Claudia Schapendonk; Corien Swaan; Corine GeurtsvanKessel; David Nieuwenhuijse; Irina Chestakova; Jeroen van Kampen; Jolanda Voermans; Madelief Mollers; Manon Haverkate; Marion Koopmans; Mark Pronk; Mart Stein; Pascal Lexmond; Reina Sikkema; Richard Molenkamp; Sandra Kengne Kamga Mobou; on behalf of the Dutch national COVID-19 response team. |
| EPI_ISL_413572 | MHC Kennemerland | Erasmus Medical Center | Anne van der Linden; Annemiek van der Eijk; Aura Timen; Bas Oude Munnink; Claudia Schapendonk; Corien Swaan; Corine GeurtsvanKessel; David Nieuwenhuijse; Irina Chestakova; Jeroen van Kampen; Jolanda Voermans; Madelief Mollers; Manon Haverkate; Marion Koopmans; Mark Pronk; Mart Stein; Pascal Lexmond; Reina Sikkema; Richard Molenkamp; Sandra Kengne Kamga Mobou; on behalf of the Dutch national COVID-19 response team. |
| EPI_ISL_413583 | MHC Rotterdam-Rijnmond | Erasmus Medical Center | Anne van der Linden; Annemiek van der Eijk; Aura Timen; Bas Oude Munnink; Claudia Schapendonk; Corien Swaan; Corine GeurtsvanKessel; David Nieuwenhuijse; Irina Chestakova; Jeroen van Kampen; Jolanda Voermans; Madelief Mollers; Manon Haverkate; Marion Koopmans; Mark Pronk; Mart Stein; Pascal Lexmond; Reina Sikkema; Richard Molenkamp; Sandra Kengne Kamga Mobou; on behalf of the Dutch national COVID-19 response team. |
| EPI_ISL_413588 to 413590 | MHC Utrecht | Erasmus Medical Center | Anne van der Linden; Annemiek van der Eijk; Aura Timen; Bas Oude Munnink; Claudia Schapendonk; Corien Swaan; Corine GeurtsvanKessel; David Nieuwenhuijse; Irina Chestakova; Jeroen van Kampen; Jolanda Voermans; Madelief Mollers; Manon Haverkate; Marion Koopmans; Mark Pronk; Mart Stein; Pascal Lexmond; Reina Sikkema; Richard Molenkamp; Sandra Kengne Kamga Mobou; on behalf of the Dutch national COVID-19 response team. |
| EPI_ISL_413564, EPI_ISL_413574 | MHC West-Brabant | Erasmus Medical Center | Anne van der Linden; Annemiek van der Eijk; Aura Timen; Bas Oude Munnink; Claudia Schapendonk; Corien Swaan; Corine GeurtsvanKessel; David Nieuwenhuijse; Irina Chestakova; Jeroen van Kampen; Jolanda Voermans; Madelief Mollers; Manon Haverkate; Marion Koopmans; Mark Pronk; Mart Stein; Pascal Lexmond; Reina Sikkema; Richard Molenkamp; Sandra Kengne Kamga Mobou; on behalf of the Dutch national COVID-19 response team. |
| EPI_ISL_710592 to 710593 | MLF Care AB | The Public Health Agency of Sweden | Department of Microbiology; The Public Health Agency of Sweden |
| EPI_ISL_631277 to 631303, EPI_ISL_632311 | MVZ DIAMEDIS Diagnostische Medizin Sennestadt GmbH | Bielefeld University | Alexander Sczyrba; Christiane Scherer; David Brandt; Jörn Kalinowski; Levin-Joe Klages; Marina Simunovic; Markus Haak; Svenja Vinke; Tobias Busche |
| EPI_ISL_831403, EPI_ISL_831406, EPI_ISL_831408 | MVZ Düsseldorf Zentrum GbR | Robert Koch Institute, Influenza and respiratory viruses FG17 & Bioinformatics MF1, Berlin, Germany | Aleksandar Radonic; Dr. Finzer; Marianne Wedde; Oliver Drechsel; Ralf Duerrwald; Rene Kmiecinski; Stefan Kroeger; Stefan Fuchs; Thorsten Wolff |
| EPI_ISL_860318, EPI_ISL_860322 to 860328, EPI_ISL_860330 to 860343, EPI_ISL_860345 to 860346, EPI_ISL_860348 to 860364, EPI_ISL_860367, EPI_ISL_860370 to 860372, EPI_ISL_860375, EPI_ISL_860378 to 860391, EPI_ISL_860394 to 860395, EPI_ISL_860397, EPI_ISL_860400 to 860409, EPI_ISL_860411 to 860413, EPI_ISL_860415, EPI_ISL_860417 to 860418, EPI_ISL_860420 to 860423, EPI_ISL_860427, EPI_ISL_860429 to 860430, EPI_ISL_860432 to 860434, EPI_ISL_860436 to 860441, EPI_ISL_860443 to 860447, EPI_ISL_860450 to 860459, EPI_ISL_860461 to 860472, EPI_ISL_860474, EPI_ISL_860476 to 860478, EPI_ISL_860481 to 860483, EPI_ISL_860485 to 860486, EPI_ISL_860488 to 860489, EPI_ISL_860492 to 860493, EPI_ISL_860495, EPI_ISL_860497 to 860500, EPI_ISL_860502, EPI_ISL_860504 to 860512, EPI_ISL_860514 to 860519, EPI_ISL_860522, EPI_ISL_860525 to 860526, EPI_ISL_860529 to 860534, EPI_ISL_860536 to 860537, EPI_ISL_860539, EPI_ISL_860542 to 860544 | | | |
| see above | MVZ Labor Krone GbR | Center of Medical Microbiology, Virology, and Hospital Hygiene, University of Duesseldorf | Alexander Dilthey; Andreas Walker; André Heimbach; Bärbel Lippke; Carsten Tiemann; Dennis Deschka; Janine Silvery; Julia Fazaal; Jörg Timm; Kerstin Ludwig; Klaus Pfeffer; Malte Kohns Vasconcelos; Per Hoffmann; Tobias Wienemann; Torsten Houwaart |
| EPI_ISL_640219 to 640264, EPI_ISL_640266 to 640269, EPI_ISL_640271 to 640272 | MVZ Laborärzte Singen | MVZ Laborärzte Singen | Folker Wenzel; Frithjof Blessing; Jonas Schmidt; Sandro Berghaus |
| EPI_ISL_428348 | Maison de Santé du Val d'Ormois | National Reference Center for Viruses of Respiratory Infections, Institut Pasteur, Paris | Angela Brisebarre; Etienne Simon-Lorière; Flora Donati; Marion Barbet; Maud Vanpeene; Mélanie Albert; Méline Bizard; Sylvie Behillil; Sylvie van der Werf; Vincent Enouf |
| EPI_ISL_420898 to 420909, EPI_ISL_420911 to 420912 | Max von Pettenkofer Institute, Virology, National Reference Center for Retroviruses, LMU Munich | Laboratory for Functional Genome Analysis, Dept. Genomics, Gene Center of the LMU Munich | Alexander Graf; Ashok Varadharajan; Helmut Blum; Max Muenchhoff; Oliver Keppler; Stefan Krebs |
| EPI_ISL_437204 to 437225, EPI_ISL_437227 to 437297, EPI_ISL_437359, EPI_ISL_451937 to 451947, EPI_ISL_452103 to 452104, EPI_ISL_466874 to 466925, EPI_ISL_548949 to 548950, EPI_ISL_548952 to 548961, EPI_ISL_852564, EPI_ISL_852566, EPI_ISL_852568 to 852569, EPI_ISL_852571 to 852603 | | | |
| see above | Max von Pettenkofer Institute, Virology, National Reference Center for Retroviruses, LMU München | Laboratory for Functional Genome Analysis, Dept. Genomics, Gene Center of the LMU Munich | Alexander Graf; Helmut Blum; Max Muenchhoff; Oliver Keppler; Stefan Krebs |
| EPI_ISL_476832 | Medical Biology Department, Kocaeli University | Medical Genetics Department, Kocaeli University | Akhan S; Akpinar G; Canturk NZ.; Cine N; Eren-Keskin S; Ilgazli A; Karadenizli A; Kasap M; Savli H; Sayan M; Sunnetci-Akkoyunlu D |
| EPI_ISL_447838 to 447839, EPI_ISL_481216, EPI_ISL_493382 to 493383, EPI_ISL_549039 to 549041, EPI_ISL_549059, EPI_ISL_549070, EPI_ISL_549081, EPI_ISL_549085 to 549086, EPI_ISL_549164, EPI_ISL_549166, EPI_ISL_590879 to 590880, EPI_ISL_590893, EPI_ISL_590929, EPI_ISL_590932 to 590933, EPI_ISL_590945, EPI_ISL_635088, EPI_ISL_635092, EPI_ISL_635099, EPI_ISL_635101, EPI_ISL_635134, EPI_ISL_635159, EPI_ISL_635169, EPI_ISL_635192 to 635193, EPI_ISL_668400, EPI_ISL_708041, EPI_ISL_708115 to 708117, EPI_ISL_708136 to 708139, EPI_ISL_708196, EPI_ISL_708199, EPI_ISL_759977 to 759980, EPI_ISL_759983, EPI_ISL_775309 to 775311, EPI_ISL_775348, EPI_ISL_775394 to 775397, EPI_ISL_775442, EPI_ISL_796651, EPI_ISL_860114 to 860116, EPI_ISL_860216 to 860217, EPI_ISL_860224 to 860225, EPI_ISL_860237 | | | |
| see above | Medical Microbiology Unit, Department for Laboratory Medicine, Drammen Hospital, Vestre Viken Health Trust, | Norwegian Institute of Public Health, Department of Virology | Atiya R Ali; Hilde Elshaug; Hilde Synnøve Vollan; Hilde Vollan; Kamilla Heddeland Instefjord; Karoline Bragstad; Kathrine Stene-Johansen; Marie Paulsen Madsen; Olav Hungnes; Rasmus Riis Kopperud |
| EPI_ISL_482745 to 482746 | Medical Microbiology, Leiden University Medical Center | Medical Microbiology, Leiden University Medical Center | Dalebout; E.J.; J.C.; J.C. and Sidorov, I.; J.J. and Sidorov, I.; N.S.; Ogando; Snijder; T.J.; Zevenhoven; de Vries |
| EPI_ISL_832820 | Medico Competente P.O. L'Aquila | Istituto Zooprofilattico Sperimentale dell'Abruzzo e Molise "G.Caporale" | Ancora M; Calistri P; Cammà C; Curini V; Di Domenico M; Di Pasquale A; Lorusso A; Mangone I; Marcacci M; Puglia I; Rinaldi A; Savini G. |
| EPI_ISL_581452 to 581454, EPI_ISL_581456 to 581461, EPI_ISL_581463 to 581466, EPI_ISL_581471, EPI_ISL_581473, EPI_ISL_581476, EPI_ISL_581478 to 581485 | | | |
| see above | Medizinische Klinik Innere Medizin I, Universitätsklinikum Tübingen | NGS Competence Center Tübingen, Institut für Medizinische Mikrobiologie und Hygiene, Universitätsklinikum Tübingen | Angel Angelov |
| EPI_ISL_763078 to 763085, EPI_ISL_977162, EPI_ISL_977164, EPI_ISL_977167 to 977179, EPI_ISL_977207 to 977210, EPI_ISL_977212 | Microbiologia e Virologia | Istituto Zooprofilattico Sperimentale delle Venezie | Adelaide Milani; Alessia Schivo; Alice Fusaro; Ambra Pastori; Annalisa Salviato; Antonia Ricci; Bianca Zecchin; Calogero Terregino; Erika Giorgia Quaranta; Isabella Monne |
| EPI_ISL_528788 to 528797, EPI_ISL_528799 to 528808, EPI_ISL_541753, EPI_ISL_541756, EPI_ISL_541767, EPI_ISL_541773 to 541774 | Microbiology Department, Barking Havering and Redbridge University Hospitals NHS trust | Wellcome Sanger Institute for the COVID-19 Genomics UK (COG-UK) consortium | Amy Ash; Cherian Koshy and Alex Alderton; Cordelia Langford; David K. Jackson; Dominic Kwiatkowski; Ewan Harrison; Fatima Ali; Ian Johnston; John Sillitoe on behalf of the Wellcome Sanger Institute COVID-19 Surveillance Team; John Sillitoe on behalf of the Wellcome Sanger Institute COVID-19 Surveillance Team (http://www.sanger.ac.uk/covid-team); Roberto Amato; Sonia Goncalves |
| EPI_ISL_482069, EPI_ISL_482071 to 482074, EPI_ISL_482076, EPI_ISL_482078 to 482081, EPI_ISL_482083 to 482086, EPI_ISL_482088 to 482090, EPI_ISL_482092 to 482095, EPI_ISL_482099 to 482100, EPI_ISL_482103 to 482104, EPI_ISL_482106 to 482114, EPI_ISL_482116 to 482119, EPI_ISL_488837 to 488839, EPI_ISL_488841 to 488844, EPI_ISL_488846, EPI_ISL_488848, EPI_ISL_488850 to 488852, EPI_ISL_488857 to 488860, EPI_ISL_488862, EPI_ISL_488865 to 488871, EPI_ISL_488873 | | | |
| see above | Microbiology Department, Hereford County Hospital | Wellcome Sanger Institute for the COVID-19 Genomics UK (COG-UK) consortium | Alison Johnson; Cordelia Langford; David K. Jackson; Dominic Kwiatkowski; Ewan Harrison; Fenella Halstead; Ian Johnston; Jane Thomas; John Sillitoe on behalf of the Wellcome Sanger Institute COVID-19 Surveillance Team (http://www.sanger.ac.uk/covid-team); Roberto Amato; Samantha Lamb and Alex Alderton; Sonia Goncalves; Venkat Sivaprakasam; Wendy Hogsden |
| EPI_ISL_833183, EPI_ISL_848199 | Microbiology Department. Complexo Hospitalario Universitario de Vigo | Microbiology Department. Complexo Hospitalario Universitario de Vigo | Complexo Hospitalario Universitario de Vigo (CHUVI). EPICOVIGAL.; Complexo Hospitalario Universitario de Vigo. EPICOVIGAL.; Microbiology Department |
| EPI_ISL_833184 | Microbiology Department. Hospital Clinico Universitario de Santiago | Microbiology Department. Complexo Hospitalario Universitario de Vigo | Microbiology Departments from Complexo Hospitalario Universitario de Vigo and Hospital Clinico Universitario de Santiago. EPICOVIGAL. |
| EPI_ISL_468331 to 468337, EPI_ISL_468340 to 468344 | Microbiology Service, University Hospital of A Coruna-Biomedical Research Institute | Genomes & Disease, Center for Research in Molecular Medicine and Chronic Diseases, University of Santiago de Compostela | Ana Pequeno-Valtierra; Cristina Gomez; Daniel Garcia-Souto; G Bou; Javier Temes; Jorge Arca; Jorge Rodriguez-Castro; Jose M. C. Tubio; Juan A. Vallejo; Kelly Conde; M Poza; Martin Santamarina; Soraya Rumbo |
| EPI_ISL_738147, EPI_ISL_738194, EPI_ISL_738243, EPI_ISL_825155 to 825156, EPI_ISL_911524 | Microbiology and Virology Unit, Florence Careggi University Hospital | Microbiology and Virology Unit, Florence Careggi University Hospital | Alberto Antonelli; Emanuele Gori; Fabio Morecchiato; Gian Maria Rossolini; Ilaria Baccani; Marco Coppi; Noemi Aiezza; Simona Pollini; Vincenzo Di Pilato |
| EPI_ISL_613459 to 613462 | Microbiology, Koc University | Microbiology, Koc University | Can, F.; Dogan, O.; Kuskucu; M.A.; Nurtop, E.; Ozer, B. |

EPI_ISL_547433 to 547442, EPI_ISL_547444, EPI_ISL_613567, EPI_ISL_613570, EPI_ISL_613636 to 613638, EPI_ISL_613640 to 613649, EPI_ISL_613651 to 613668, EPI_ISL_613670 to 613681, EPI_ISL_613683 to 613693, EPI_ISL_613695 to 613705, EPI_ISL_614983 to 614986, EPI_ISL_614988, EPI_ISL_614990 to 614994, EPI_ISL_614997 to 614999, EPI_ISL_615002 to 615003, EPI_ISL_615009 to 615021, EPI_ISL_615023 to 615025, EPI_ISL_615027 to 615033, EPI_ISL_615035, EPI_ISL_615037 to 615038, EPI_ISL_615040 to 615044, EPI_ISL_615046 to 615049, EPI_ISL_615051 to 615052, EPI_ISL_615054 to 615056, EPI_ISL_615058 to 615061, EPI_ISL_632797 to 632801, EPI_ISL_632804 to 632813, EPI_ISL_632815 to 632817, EPI_ISL_632820 to 632821, EPI_ISL_632846

| | | | |
|---|---|---|---|
| see above | Microbiology, Department of Pathology, St. Bernard's Hospital, Gibraltar Health Authority | Respiratory Virus Unit, Microbiology Services Colindale, Public Health England | Charlotte Gillborn-Jones (Gibraltar); Dr Nicholas Cortes (Gibraltar); PHE Covid Sequencing Team |

EPI_ISL_709948, EPI_ISL_709950 to 709951, EPI_ISL_709954 to 709955, EPI_ISL_709957, EPI_ISL_709959 to 709961, EPI_ISL_709963 to 709970, EPI_ISL_709972 to 710025, EPI_ISL_710027 to 710033

| | | | |
|---|---|---|---|
| see above | Microbiology, Department of Pathology, St. Bernard's Hospital, Gibraltar Health Authority | Respiratory Virus Unit, National Infection Service, Public Health England | Charlotte Gillborn-Jones (Gibraltar); Dr Nicholas Cortes (Gibraltar); PHE Covid Sequencing Team |
| EPI_ISL_513585, EPI_ISL_513587 to 513588 | Microbiology, Pathology, Kettering General Hospital | Wellcome Sanger Institute for the COVID-19 Genomics UK (COG-UK) consortium | Anita Kenyon; Cordelia Langford; David K. Jackson; Dominic Kwiatkowski; Ewan Harrison; Ian Johnston; John Sillitoe on behalf of the Wellcome Sanger Institute COVID-19 Surveillance Team (http://www.sanger.ac.uk/covid-team); Roberto Amato; Sahar El-dirdiri; Sonia Goncalves; Thomas Davis and Alex Alderton |
| EPI_ISL_475717 to 475721 | Microbiology, University Hospital Donostia | Microbiology, University Hospital Donostia | Cilla, G.; J.M.; Marimon; Montes, M.; Pineiro, L. |
| EPI_ISL_486646 to 486660, EPI_ISL_486662 to 486665 | Microbiology, Virology and Biemergency Laboratory-ASST FBF Sacco | Microbiology, Virology and Biemergency Laboratory-ASST FBF Sacco | Comandatore F; Mancon A; Micheli V; Rimoldi SG; Romeri F |
| EPI_ISL_794624 | Middlemore Hospital | Institute of Environmental Science and Research (ESR) | Anja Werno; Antje van der Linden; Arlo Upton; Chris Mansell; David Hammer; Dragana Drinkovic; Erasmus Smit; Gary McAuliffe; Hana Sofia Andersson; Hermes Perez; James Ussher; Jill Sherwood; Jing Wang; Joep de Ligt; Josh Freeman; Julia Howard; Juliet Elvy; Lauren Jelly; Mary DeAlmeida; Matt Blakiston; Matt Storey; Matthew Rogers; Max Bloomfield; Michael Addidle; Michelle Balm; Muhammad Faisal; Nikki Freed; Olin Silander; Sally Roberts; Sarah Jefferies; Sharmini Muttaiyah; Susan Morpeth; Susan Taylor; Timothy Blackmore; Vani Sathyendran; Veronica Playle; Virginia Hope; Xiaoyun Ren |
| EPI_ISL_661291 to 661292 | Mikrobiologen | The Public Health Agency of Sweden | Department of Microbiology; The Public Health Agency of Sweden |
| EPI_ISL_678273 to 678289 | Mikrobiologie, RARI | Mikrobiologie, RARI | A.V.; Badanin; D.V.; E.A.; Fedorov; Guseva; Krasnov; Kutyrev; N.A.; N.P.; Naryshkina; Portenko; S.A.; Sharapova; Shcherbakova; Sosedova; V.V.; Y.M. |
| EPI_ISL_718132, EPI_ISL_718142, EPI_ISL_718165 | Ministry of Health Hospitals | Institute of Health and Community Medicine | Chan Chia Jui; Chua Hock Hin; David Perera; Ooi Mong How; Tonnii Sia Loong Loong; Wong Jyn Shan; Wong Kiing Aik |

EPI_ISL_417413, EPI_ISL_428712 to 428713, EPI_ISL_428715, EPI_ISL_428717, EPI_ISL_428719 to 428722, EPI_ISL_429861, EPI_ISL_429863 to 429865, EPI_ISL_429867 to 429868, EPI_ISL_429872 to 429873, EPI_ISL_437304 to 437316, EPI_ISL_437318, EPI_ISL_437320 to 437322, EPI_ISL_437330, EPI_ISL_437333 to 437335, EPI_ISL_811136 to 811139, EPI_ISL_811143, EPI_ISL_812761 to 812771, EPI_ISL_812774 to 812777, EPI_ISL_812779 to 812781, EPI_ISL_812873 to 812878, EPI_ISL_812880 to 812921, EPI_ISL_814062 to 814077, EPI_ISL_814079 to 814091

| | | | |
|---|---|---|---|
| see above | Ministry of Health Turkey | Ministry of Health Turkey | Aye Baak Alta; Fatma Bayrakdar; Gülay Korukluolu; Selçuk Klç; Süleyman Yalcin; Süleyman Yalçn; Tülin Demir; Yasemin Cogun |
| EPI_ISL_462995 to 463000 | Molecular Genetics | Molecular Genetics | Coto, E.; Gomez, J. |
| EPI_ISL_451306 to 451309, EPI_ISL_460079 to 460095 | Molecular Virology Unit, Fondazione IRCCS Policlinico San Matteo , Pavia | Laboratory of Virology, INMI Lazzaro Spallanzani IRCCS | Antonino Di Caro; Antonio Piralla; Barbara Bartolini; Cesare E.M. Gruber; Fausto Baldanti; Maria R. Capobianchi; Martina Rueca |

EPI_ISL_460096, EPI_ISL_460604 to 460605, EPI_ISL_462149 to 462150, EPI_ISL_467774 to 467775, EPI_ISL_470539, EPI_ISL_479620 to 479624, EPI_ISL_486815 to 486829, EPI_ISL_534337 to 534349, EPI_ISL_539299, EPI_ISL_610227 to 610244, EPI_ISL_653922 to 653932, EPI_ISL_660435 to 660438, EPI_ISL_717965 to 717974, EPI_ISL_732659 to 732676, EPI_ISL_745223 to 745259, EPI_ISL_754196 to 754214

| | | | |
|---|---|---|---|
| see above | Molecular diagnostic laboratory of Federal Budget Institution of Science "Central Research Institute of Epidemiology" of The Federal Service on Customers' Rights Protection and Human Well-being Surveillance | Group of Genomics and Postgenomic Technologies of Central Research Institute of Epidemilology | Akimkin VG; Bukharina A.Y.; Bukharina AY; Bulanenko VP; Dudorova A.V.; Dudorova AV; Kaptelova VV; Korneenko EV; Saenko SS; Samoilov AE; Shipulina OY; Sizova TV; Speranskaya AS; Tivanova EV; Valdokhina AV |
| EPI_ISL_426883 to 426897 | Motol University Hospital | Institute of Applied Biotechnologies a.s. | Adam Novotný; Jan Geryk; Kateina Kvapilová; Martin Kašný; Milan Macek; Pavel Devínek; Petr Brož; Petr Klempt; Petr Kvapil |
| EPI_ISL_730622, EPI_ISL_730626 to 730629, EPI_ISL_730631 to 730633, EPI_ISL_730637 to 730639 | Männedorf Hospital | Institute of Medical Virology, University of Zurich | Alexandra Trkola; Gabriela Ziltener; Jürg Böni; Maryam Zaheri; Michael Huber; Stefan Schmutz; Verena Kufner |
| EPI_ISL_490205 to 490208 | München Klinik Schwabing | MGZ Medical Genetics Center | Dieter A. Wolf; Elke Holinski-Feder |
| EPI_ISL_510810 to 510816 | NA | The Public Health Agency of Sweden | Anna Risberg; Anna-Malin Linde; Karin Tegmark-Wisell; Maria Lind Karlberg; Mattias Haukland; Mia Brytting; Olov Svartstrom; Oskar Karlsson Lindsjo; Petra Edquist; Reza Advani; Sandra Broddesson |
| EPI_ISL_825044 to 825047, EPI_ISL_825050 to 825053, EPI_ISL_825059 to 825060, EPI_ISL_825063 | NHL Municipal Medical College, Ahmedbad | Gujarat Biotechnology Research Centre | Afzal Ansari; Apurvasinh Puvar; Atit Shah; Bhavin Prajapati; Bimal Chauhan; Chaitanya Joshi; Dinesh Kumar; Janvi Raval; Jayshri Pethani; Labdhi Pandya; Madhvi Joshi; Monila Patel; NM Shaikh; Nikha Trivedi; Nitin Savaliya; Ramesh Pandit; Tanmay Mehta; Zarna Patel; Zuber Saiyed |
| EPI_ISL_632242 to 632249 | NHSGGC West of Scotland Specialist Virology Centre | MRC-University of Glasgow Centre for Virus Research | Alasdair MacLean; Alex Alderton; Ana da Silva Filipe; Antonia Ho; Cordelia Langford; Daniel Mair; David K. Jackson; David L Robertson; Dominic Kwiatkowski; Emma Thomson; Ewan Harrison; Ian Johnston; James Shepherd; Jenna Nichols; John Sillitoe on behalf of the Wellcome Sanger Institute COVID-19 Surveillance Team (http://www.sanger.ac.uk/covid-team); Joseph Hughes; Kathy Li; Kathy Smollett; Kyriaki Nomikou; Lily Tong; Natasha Jesudason; Natasha Johnson; Rajiv Shah; Richard Orton; Roberto Amato; Rory Gunson; Sonia Goncalves; Sreenu Vattipally; Stephen Carmichael |
| EPI_ISL_459108, EPI_ISL_489581, EPI_ISL_534395, EPI_ISL_534634 | NHSGGC West of Scotland Specialist Virology Centre / MRC-University of Glasgow Centre for Virus Research | Wellcome Sanger Institute for the COVID-19 Genomics UK (COG-UK) Consortium | Alasdair MacLean; Alice Broos; Ana da Silva Filipe; Antonia Ho; Cordelia Langford; Daniel Mair; David K. Jackson; David L Robertson; Dominic Kwiatkowski; Elihu Aranday-Cortes; Emma Thomson and Alex Alderton; Ewan Harrison; Ian Johnston; James Shepherd; Jenna Nichols; John Sillitoe on behalf of the Wellcome Sanger Institute COVID-19 Surveillance Team; Joseph Hughes; Kathy Li; Kathy Smollett; Kirstyn Brunker; Kyriaki Nomikou; Lily Tong; Marc Niebel; Natasha Jesudason; Natasha Johnson; Patawee Asamaphan; Rajiv Shah; Richard Orton; Roberto Amato; Rory Gunson; Sarah McDonald; Sonia Goncalves; Sreenu Vattipally; Stephen Carmichael; Yasmin Parr |

EPI_ISL_459095 to 459107, EPI_ISL_459109 to 459122, EPI_ISL_459124 to 459127, EPI_ISL_459129 to 459140, EPI_ISL_459142 to 459145, EPI_ISL_459147 to 459165, EPI_ISL_459505 to 459510, EPI_ISL_459512 to 459566, EPI_ISL_459568 to 459580, EPI_ISL_459582 to 459619, EPI_ISL_459621 to 459660, EPI_ISL_459662 to 459686, EPI_ISL_459688 to 459721, EPI_ISL_459723 to 459724, EPI_ISL_469930 to 469938, EPI_ISL_469940 to 470012, EPI_ISL_489380, EPI_ISL_489391 to 489393, EPI_ISL_489396, EPI_ISL_489401, EPI_ISL_489407 to 489408, EPI_ISL_489413, EPI_ISL_489418, EPI_ISL_489429, EPI_ISL_489442, EPI_ISL_489449, EPI_ISL_489455, EPI_ISL_489470, EPI_ISL_489480, EPI_ISL_489489, EPI_ISL_489523, EPI_ISL_489530, EPI_ISL_489536, EPI_ISL_489543, EPI_ISL_489547, EPI_ISL_489567, EPI_ISL_489578 to 489580, EPI_ISL_489582 to 489643, EPI_ISL_489645 to 489650, EPI_ISL_489654 to 489658, EPI_ISL_489660 to 489661, EPI_ISL_489663 to 489670, EPI_ISL_489672 to 489706, EPI_ISL_501633 to 501635, EPI_ISL_524494, EPI_ISL_524496, EPI_ISL_524499 to 524500, EPI_ISL_524504, EPI_ISL_524507, EPI_ISL_524510 to 524514, EPI_ISL_524516 to 524520, EPI_ISL_524523 to 524524, EPI_ISL_524526 to 524527, EPI_ISL_524532 to 524536, EPI_ISL_524539 to 524540, EPI_ISL_524542, EPI_ISL_524545, EPI_ISL_524547, EPI_ISL_524549 to 524550, EPI_ISL_524552, EPI_ISL_524554 to 524557, EPI_ISL_524559, EPI_ISL_524561 to 524563, EPI_ISL_524565 to 524567, EPI_ISL_524569, EPI_ISL_524571 to 524572, EPI_ISL_530361, EPI_ISL_530366, EPI_ISL_530370, EPI_ISL_530377, EPI_ISL_530386, EPI_ISL_530388, EPI_ISL_530391, EPI_ISL_530398, EPI_ISL_530410, EPI_ISL_530429, EPI_ISL_530435, EPI_ISL_530440, EPI_ISL_530468, EPI_ISL_530480, EPI_ISL_530488 to 530489, EPI_ISL_530523, EPI_ISL_530528, EPI_ISL_530533, EPI_ISL_530535, EPI_ISL_530542, EPI_ISL_530553, EPI_ISL_530562, EPI_ISL_530596, EPI_ISL_530642, EPI_ISL_530659, EPI_ISL_530673, EPI_ISL_530682, EPI_ISL_530689, EPI_ISL_530708, EPI_ISL_530722, EPI_ISL_530740, EPI_ISL_530743, EPI_ISL_530746, EPI_ISL_530754, EPI_ISL_530764, EPI_ISL_530766 to 530768, EPI_ISL_530826, EPI_ISL_530836, EPI_ISL_530845, EPI_ISL_530862, EPI_ISL_530871, EPI_ISL_530873, EPI_ISL_530904, EPI_ISL_530906, EPI_ISL_530923, EPI_ISL_530929, EPI_ISL_530953, EPI_ISL_530959, EPI_ISL_530961 to 530962, EPI_ISL_530973, EPI_ISL_530976, EPI_ISL_530979, EPI_ISL_530983, EPI_ISL_530985 to 530986, EPI_ISL_531002, EPI_ISL_531004, EPI_ISL_531006, EPI_ISL_531017, EPI_ISL_531020, EPI_ISL_531032 to 531033, EPI_ISL_531038, EPI_ISL_531040, EPI_ISL_531045, EPI_ISL_531048 to 531049, EPI_ISL_531052, EPI_ISL_531054, EPI_ISL_531056 to 531057, EPI_ISL_531065, EPI_ISL_531079, EPI_ISL_531083, EPI_ISL_531087, EPI_ISL_531091, EPI_ISL_531096, EPI_ISL_531099 to 531100, EPI_ISL_531103 to 531104, EPI_ISL_531114, EPI_ISL_531116, EPI_ISL_531118, EPI_ISL_531123, EPI_ISL_531126, EPI_ISL_531130 to 531131, EPI_ISL_531142, EPI_ISL_531147, EPI_ISL_531154 to 531157, EPI_ISL_531159, EPI_ISL_531166, EPI_ISL_531174 to 531175, EPI_ISL_531188 to 531189, EPI_ISL_531198, EPI_ISL_531207, EPI_ISL_531212, EPI_ISL_531214, EPI_ISL_531223, EPI_ISL_531249, EPI_ISL_531266, EPI_ISL_531321, EPI_ISL_531330, EPI_ISL_531365, EPI_ISL_531374, EPI_ISL_531382, EPI_ISL_531412, EPI_ISL_531416, EPI_ISL_531420, EPI_ISL_531422, EPI_ISL_531432, EPI_ISL_531442, EPI_ISL_531464, EPI_ISL_531470, EPI_ISL_531473, EPI_ISL_531476 to 531477, EPI_ISL_531480, EPI_ISL_531483 to 531485, EPI_ISL_531492, EPI_ISL_531495, EPI_ISL_531500, EPI_ISL_531504, EPI_ISL_531524, EPI_ISL_531546, EPI_ISL_531548, EPI_ISL_531557, EPI_ISL_531563 to 531565, EPI_ISL_531569 to 531570, EPI_ISL_531575, EPI_ISL_531577, EPI_ISL_531583, EPI_ISL_531590, EPI_ISL_531595, EPI_ISL_531598 to 531599, EPI_ISL_531607, EPI_ISL_531609, EPI_ISL_531617 to 531618, EPI_ISL_531625, EPI_ISL_531631, EPI_ISL_531634, EPI_ISL_531638 to 531639, EPI_ISL_531649, EPI_ISL_531660, EPI_ISL_531668, EPI_ISL_531672, EPI_ISL_531677 to 531678, EPI_ISL_531689 to 531691, EPI_ISL_531702 to 531703, EPI_ISL_531714, EPI_ISL_531727, EPI_ISL_531730, EPI_ISL_531732, EPI_ISL_531743, EPI_ISL_531753 to 531755, EPI_ISL_531768, EPI_ISL_531779, EPI_ISL_531788, EPI_ISL_531792, EPI_ISL_531801 to 531802, EPI_ISL_531805, EPI_ISL_531814 to 531815, EPI_ISL_531821, EPI_ISL_531824, EPI_ISL_531832, EPI_ISL_531842, EPI_ISL_531851, EPI_ISL_531856 to 531858, EPI_ISL_531862 to 531863, EPI_ISL_531865, EPI_ISL_531873, EPI_ISL_531877, EPI_ISL_531880 to 531881, EPI_ISL_531884, EPI_ISL_531887, EPI_ISL_531889 to 531890, EPI_ISL_531892, EPI_ISL_531894, EPI_ISL_531905, EPI_ISL_531912 to 531913, EPI_ISL_531918 to 531919, EPI_ISL_531925, EPI_ISL_531929, EPI_ISL_531932 to 531933, EPI_ISL_531935 to 531936, EPI_ISL_531943, EPI_ISL_531945, EPI_ISL_531951, EPI_ISL_531954, EPI_ISL_531978 to 531979, EPI_ISL_531982, EPI_ISL_531984, EPI_ISL_531987, EPI_ISL_531990, EPI_ISL_531995, EPI_ISL_532003 to 532005, EPI_ISL_532007, EPI_ISL_532018, EPI_ISL_532024, EPI_ISL_532032, EPI_ISL_532038, EPI_ISL_532049, EPI_ISL_532053, EPI_ISL_532055, EPI_ISL_532057, EPI_ISL_532062, EPI_ISL_532070 to 532071, EPI_ISL_532076 to 532078, EPI_ISL_532080 to 532081, EPI_ISL_532085, EPI_ISL_532093, EPI_ISL_532098, EPI_ISL_532102, EPI_ISL_532105, EPI_ISL_532118, EPI_ISL_532123, EPI_ISL_532125, EPI_ISL_532129 to 532131, EPI_ISL_532168, EPI_ISL_532178, EPI_ISL_532186, EPI_ISL_532192, EPI_ISL_532195 to 532197, EPI_ISL_532200, EPI_ISL_532202, EPI_ISL_532210, EPI_ISL_532214, EPI_ISL_532244, EPI_ISL_532258, EPI_ISL_532263, EPI_ISL_532271 to 532272, EPI_ISL_532275 to 532276, EPI_ISL_532278, EPI_ISL_532283, EPI_ISL_532288, EPI_ISL_532293, EPI_ISL_532301, EPI_ISL_532311, EPI_ISL_532316 to 532317, EPI_ISL_532321, EPI_ISL_532327 to 532328, EPI_ISL_532334 to 532335, EPI_ISL_532338, EPI_ISL_532340, EPI_ISL_532347, EPI_ISL_532349, EPI_ISL_532351, EPI_ISL_532353, EPI_ISL_532355 to 532356, EPI_ISL_532363 to 532364, EPI_ISL_532379, EPI_ISL_532384, EPI_ISL_532388, EPI_ISL_532390, EPI_ISL_532394, EPI_ISL_532399, EPI_ISL_532402, EPI_ISL_532404, EPI_ISL_532414, EPI_ISL_532430 to 532432, EPI_ISL_532438, EPI_ISL_532440, EPI_ISL_532444, EPI_ISL_532446, EPI_ISL_532452, EPI_ISL_532454, EPI_ISL_532456, EPI_ISL_532463, EPI_ISL_532469, EPI_ISL_532472 to 532473, EPI_ISL_532480, EPI_ISL_532484, EPI_ISL_532490, EPI_ISL_532492 to 532493, EPI_ISL_532506, EPI_ISL_532510, EPI_ISL_532512, EPI_ISL_532514 to 532515, EPI_ISL_532518, EPI_ISL_532523,

| | | | |
|---|---|---|---|
| EPI_ISL_532531, EPI_ISL_532538, EPI_ISL_532540, EPI_ISL_532563 to 532564, EPI_ISL_532572, EPI_ISL_532582, EPI_ISL_532587, EPI_ISL_532594, EPI_ISL_532598, EPI_ISL_532620 to 532621, EPI_ISL_532625 to 532626, EPI_ISL_532633, EPI_ISL_532639, EPI_ISL_532648, EPI_ISL_532670, EPI_ISL_532697, EPI_ISL_532715, EPI_ISL_532733, EPI_ISL_532738, EPI_ISL_532744, EPI_ISL_532746, EPI_ISL_532750, EPI_ISL_532753, EPI_ISL_532755, EPI_ISL_532765, EPI_ISL_532767, EPI_ISL_532784, EPI_ISL_532786, EPI_ISL_532791, EPI_ISL_532811, EPI_ISL_532838, EPI_ISL_532878, EPI_ISL_532886, EPI_ISL_532894, EPI_ISL_532896, EPI_ISL_532915, EPI_ISL_532933, EPI_ISL_532988, EPI_ISL_533016, EPI_ISL_533027, EPI_ISL_533072, EPI_ISL_533228, EPI_ISL_533230 to 533232, EPI_ISL_533236 to 533237, EPI_ISL_533241, EPI_ISL_533252, EPI_ISL_533256, EPI_ISL_533262, EPI_ISL_533270 to 533271, EPI_ISL_533283, EPI_ISL_533289, EPI_ISL_533298, EPI_ISL_533311, EPI_ISL_533343, EPI_ISL_533346, EPI_ISL_533350 to 533351, EPI_ISL_533358, EPI_ISL_533365, EPI_ISL_533376, EPI_ISL_533394, EPI_ISL_533406, EPI_ISL_533419, EPI_ISL_533421, EPI_ISL_534365 to 534378, EPI_ISL_534380, EPI_ISL_534382 to 534394, EPI_ISL_534396 to 534402, EPI_ISL_534404 to 534416, EPI_ISL_534418 to 534431, EPI_ISL_534433 to 534513, EPI_ISL_534515 to 534633, EPI_ISL_534635 to 534688, EPI_ISL_534690 | | | |
| see above | NHSGGC West of Scotland Specialist Virology Centre / MRC-University of Glasgow Centre for Virus Research | Wellcome Sanger Institute for the COVID-19 Genomics UK (COG-UK) consortium | Alasdair MacLean; Alice Broos; Ana da Silva Filipe; Antonia Ho; Cordelia Langford; Daniel Mair; David K. Jackson; David L Robertson; Dominic Kwiatkowski; Elihu Aranday-Cortes; Emma Thomson and Alex Alderton; Ewan Harrison; Ian Johnston; James Shepherd; Jenna Nichols; John Sillitoe; John Sillitoe on behalf of the Wellcome Sanger Institute COVID-19 Surveillance Team (http://www.sanger.ac.uk/covid-team); Joseph Hughes; Kathy Li; Kathy Smollett; Kirstyn Brunker; Kyriaki Nomikou; Lily Tong; Marc Niebel; Natasha Jesudason; Natasha Johnson; Patawee Asamaphan; Rajiv Shah; Richard Orton; Roberto Amato; Rory Gunson; Sarah McDonald; Sonia Goncalves; Sreenu Vattipally; Stephen Carmichael; Yasmin Parr |
| EPI_ISL_416742 to 416743 | NRL for Influenza, Centrum Epidemiology and Microbiology of National Institute of Public Health, Czech Republic | Charite Universitaetsmedizin Berlin, Institute of Virology | Akexander Nagy; Barbara Muehlemann; Christian Drosten; Dusan Trnka; Helena Jirincova; Jaromira Vecerova; Jorn Beheim-Schwarzbach; Julia Schneider; Ludmila Novakova; Talitha Veith; Terry Jones; Victor M Corman |
| EPI_ISL_458712, EPI_ISL_492929, EPI_ISL_492931 | NU-OMICS DNA Sequencing research facility, Northumbria University | Wellcome Sanger Institute for the COVID-19 Genomics UK (COG-UK) Consortium | Andrew Nelson; Brendan Payne; Chris Duncan; Clive Graham; Cordelia Langford; Darren Smith and Alex Alderton; David K. Jackson; Debra Padgett; Dominic Kwiatkowski; Edward Barton; Emma Swindells; Ewan Harrison; Garren Scott; Gary Black; Gary Eltringham; Greg Young; Ian Johnston; Jane Greenaway; Jennifer Collins; John Allan; John Sillitoe on behalf of the Wellcome Sanger Institute COVID-19 Surveillance Team; Joshua Loh; Lynn Dover; Matthew Bashton; Paul Baker; Roberto Amato; Sarah Essex; Sheia Waugh; Shirelle Burton-Fanning; Sonia Goncalves; Steve Ligget; Wen Yew; Yusri Taha |
| EPI_ISL_458578 to 458581, EPI_ISL_458583 to 458598, EPI_ISL_458600 to 458601, EPI_ISL_458603 to 458639, EPI_ISL_458641 to 458711, EPI_ISL_458713 to 458718, EPI_ISL_469302 to 469303, EPI_ISL_469305, EPI_ISL_469307 to 469313, EPI_ISL_469315 to 469316, EPI_ISL_469318 to 469320, EPI_ISL_469322 to 469335, EPI_ISL_469337 to 469344, EPI_ISL_469805, EPI_ISL_469915, EPI_ISL_488003 to 488051, EPI_ISL_488053 to 488059, EPI_ISL_488061 to 488082, EPI_ISL_488084 to 488184, EPI_ISL_488186, EPI_ISL_488456 to 488461, EPI_ISL_488463 to 488532, EPI_ISL_488534 to 488547, EPI_ISL_488549 to 488574, EPI_ISL_488576 to 488747, EPI_ISL_488749 to 488816, EPI_ISL_488818 to 488836, EPI_ISL_489064 to 489111, EPI_ISL_489113 to 489119, EPI_ISL_489121 to 489124, EPI_ISL_489126 to 489155, EPI_ISL_492207 to 492210, EPI_ISL_492450, EPI_ISL_492452 to 492456, EPI_ISL_492458 to 492466, EPI_ISL_492469, EPI_ISL_492471 to 492494, EPI_ISL_492916 to 492928, EPI_ISL_492930, EPI_ISL_492932 to 492933, EPI_ISL_492935 to 492939, EPI_ISL_492941 to 492957, EPI_ISL_492959 to 492960, EPI_ISL_492962 to 492966, EPI_ISL_492968 to 492977 | | | |
| see above | NU-OMICS DNA Sequencing research facility, Northumbria University | Wellcome Sanger Institute for the COVID-19 Genomics UK (COG-UK) consortium | Andrew Nelson; Brendan Payne; Chris Duncan; Clive Graham; Cordelia Langford; Darren Smith and Alex Alderton; David K. Jackson; Debra Padgett; Dominic Kwiatkowski; Edward Barton; Emma Swindells; Ewan Harrison; Garren Scott; Gary Black; Gary Eltringham; Greg Young; Ian Johnston; Jane Greenaway; Jennifer Collins; John Allan; John Sillitoe on behalf of the Wellcome Sanger Institute COVID-19 Surveillance Team (http://www.sanger.ac.uk/covid-team); Joshua Loh; Lynn Dover; Matthew Bashton; Paul Baker; Roberto Amato; Sarah Essex; Sheia Waugh; Shirelle Burton-Fanning; Sonia Goncalves; Steve Ligget; Wen Yew; Yusri Taha |
| EPI_ISL_632313 | NU-sjukvården | Clinical Microbiology, Sahlgrenska University Hospital, | Hedvig Engström Jakobsson; Johan Ringlander; Josefin Olausson; Magnus Lindh |
| EPI_ISL_632312 | NU-sjukvården | Clinical microbiology, Sahlgrenska University Hospital | Hedvig Engström Jakobsson; Johan Ringlander; Josefin Olausson; Magnus Lindh |
| EPI_ISL_434660, EPI_ISL_434674, EPI_ISL_445229, EPI_ISL_445236, EPI_ISL_450817, EPI_ISL_450823, EPI_ISL_450825, EPI_ISL_452229 to 452230, EPI_ISL_452235, EPI_ISL_452239, EPI_ISL_475565 | | | |
| see above | Narhalsan Backa vardcentral | The Public Health Agency of Sweden | Anna Risberg; Anna-Malin Linde; Karin Tegmark-Wisell; Maria Lind Karlberg; Mats Olsson; Mattias Haukland; Mia Brytting; Olov Svartstrom; Oskar Karlsson Lindsjo; Reza Advani; Sandra Broddesson; Theresa Enkirch |
| EPI_ISL_539885, EPI_ISL_582840, EPI_ISL_710589 to 710591, EPI_ISL_732655 to 732656 | Narhalsan Fjallbacka VC | The Public Health Agency of Sweden | Anna Risberg; Anna-Malin Linde; Department of Microbiology; Karin Tegmark-Wisell; Maria Lind Karlberg; Mattias Haukland; Mia Brytting; Olov Svartstrom; Oskar Karlsson Lindsjo; Petra Edquist; Reza Advani; Sandra Broddesson; The Public Health Agency of Sweden; Theresa Enkirch |
| EPI_ISL_455107 | Narhalsan Mellerud | The Public Health Agency of Sweden | Anna Risberg; Anna-Malin Linde; Karin Tegmark-Wisell; Maria Lind Karlberg; Maria Nykvist; Mia Brytting; Olov Svartstrom; Oskar Karlsson Lindsjo; Theresa Enkirch |
| EPI_ISL_434670, EPI_ISL_445237, EPI_ISL_450815, EPI_ISL_450820 | Narhalsan Molnlycke, Barn och ungdomsmedicin | The Public Health Agency of Sweden | Anna Risberg; Anna-Malin Linde; Karin Tegmark-Wisell; Maria Lind Karlberg; Mats Reimer; Mia Brytting; Olov Svartstrom; Oskar Karlsson Lindsjo; Theresa Enkirch |
| EPI_ISL_475537 | Narhalsan Oden VC | The Public Health Agency of Sweden | Anna Risberg; Anna-Malin Linde; Karin Tegmark-Wisell; Maria Lind Karlberg; Mattias Haukland; Mia Brytting; Olov Svartstrom; Oskar Karlsson Lindsjo; Reza Advani; Sandra Broddesson |
| EPI_ISL_445224, EPI_ISL_450830, EPI_ISL_469069, EPI_ISL_539882 | Narhalsan Olskroken VC | The Public Health Agency of Sweden | Anna Risberg; Anna-Malin Linde; Karin Tegmark-Wisell; Mahin Ghoroghi; Maria Lind Karlberg; Mattias Haukland; Mia Brytting; Olov Svartstrom; Oskar Karlsson Lindsjo; Petra Edquist; Reza Advani; Sandra Broddesson; Shamam Muradrasoli; Theresa Enkirch |
| EPI_ISL_434671, EPI_ISL_450829, EPI_ISL_452238, EPI_ISL_469058, EPI_ISL_469060 to 469061, EPI_ISL_475524, EPI_ISL_475539 | | | |
| see above | Narhalsan Sjobo vardcentral | The Public Health Agency of Sweden | Anna Risberg; Anna-Malin Linde; Karin Tegmark-Wisell; Lovisa Hjerten; Maria Lind Karlberg; Mattias Haukland; Mia Brytting; Olov Svartstrom; Oskar Karlsson Lindsjo; Petra Edquist; Reza Advani; Sandra Broddesson; Shamam Muradrasoli; Theresa Enkirch |
| EPI_ISL_523960, EPI_ISL_523962, EPI_ISL_523964, EPI_ISL_523966, EPI_ISL_523968 | National Agency for Public Health, Republic of Moldova | Charite Universitatsmedizin Berlin, Institute of Virology | Ala Halacu; Barbara Muhlemann; Barbara Mühlemann; Christian Drosten; Elizabeta Jancheska; Golubinka Bosevska; Joern Beheim-Schwarzbach; Jorn Beheim-Schwarzbach; Julia Schneider; Maja Kuzmanovska; Mariana Apostol; Talitha Veith; Terry Jones; Victor M Corman |
| EPI_ISL_464112 | National Health Laboratory Service (NHLS), Tygerberg | Division of Medical Virology, Stellenbosch University and National Health Laboratory Service (NHLS) | Bronwyn Kleinhans; Eduan Wilkindon; Gert van Zyl; Houriiyah Tegally; Kayla Delaney; Susan Engelbrecht; Tulio de Oliveira; Wolfgang Preiser |
| EPI_ISL_416028, EPI_ISL_416036 | National Influenza Center - Instituto Adolfo Lutz | Instituto Adolfo Lutz, Interdiciplinary Procedures Center, Strategic Laboratory | Adriana Bugno; Adriano Abbud; Carlos Henrique Camargo; Claudia Regina Gonçalves; Claudio Tavares Sacchi; Daniela Bernardes Borges da Silva; Erica Valessa Ramos Gomes; Fabiana Cristina Pereira dos Santos; Maria do Carmo Sampaio Tavares Timenetsky; Simone Guadagnucci Morillo; Terezinha Maria de Paiva |
| EPI_ISL_420543, EPI_ISL_420545, EPI_ISL_420547, EPI_ISL_420549, EPI_ISL_420553 | National Influenza Center, Indian Council of Medical Research - National Institute of Virology | Indian Council of Medical Research-National Institute of Virology, Microbial Containment Complex | Amita Jain; Anita M. Shete; Atanu Basu; Bharati Malhotra; Dimpal A. Nyayanit; Gajanan Sapkal; Lalit Dar; M Choudhary; Pragya D. Yadav. Savita Patil; Pranita Gawande; Prasad Sarkale; Priya Abraham; Sarah Cherian; Varsha Potdar |
| EPI_ISL_467778 to 467781 | National Influenza Centre Romania | Charite Universitatsmedizin Berlin, Institute of Virology | Barbara Muehlemann; Christian Drosten; Jorn Beheim-Schwarzbach; Julia Schneider; L. Ustea; M. Lazar; N. Paraschiv; Talitha Veith; Terry Jones; Victor M Corman |
| EPI_ISL_476835, EPI_ISL_476837 to 476838, EPI_ISL_476841, EPI_ISL_476843, EPI_ISL_476845, EPI_ISL_476847, EPI_ISL_476851 | | | |
| see above | National Influenza Centre for Northern Greece | National Influenza Centre for Northern Greece | Maria Christoforidi |
| EPI_ISL_476836 | National Influenza Centre for Nothern Greece | National Influenza Centre for Nothern Greece | Maria Christoforidi |
| EPI_ISL_710542 to 710546, EPI_ISL_718262 to 718265, EPI_ISL_721624 to 721628, EPI_ISL_728279 to 728287, EPI_ISL_735510, EPI_ISL_746826 to 746835, EPI_ISL_765567 to 765573, EPI_ISL_803897 to 803898, EPI_ISL_803900 | | | |
| see above | National Institute for Infectious Diseases, INMI, "L. Spallanzani" IRCCS | National Institute for Infectious Diseases, INMI, "L. Spallanzani" IRCCS | A Di Caro; A. Di Caro; B Bartolini; B. Bartolini; C.E.M Gruber; E Giombini; E. Giombini; F Messina; F. Carletti; F. Messina; G Parisi; M Rueca; M. Rueca; M.R Capobianchi; MR Capobianchi; O Butera |
| EPI_ISL_862832 | National Institute of Infectious Diseases-Prof. Dr. Matei Bals Molecular Diagnostics Laboratory | National Institute of Infectious Diseases-Prof. Dr. Matei Bals Molecular Diagnostics Laboratory | Andreea Tudor; Corina Casangiu; Dan Otelea; Leontina Banica; Marius Surleac; Petre Milu; Simona Paraschiv |
| EPI_ISL_539776, EPI_ISL_539778 to 539782 | National Institute of Public Health (Czech Republic) | State Veterinary Institute Prague | A; D; H; Jirincova; L; Nagy; Novakova; Trnka; Vecerova, J. |
| EPI_ISL_846587, EPI_ISL_856910 to 856911 | National Institute of Public Health - National Institute of Hygiene | National Institute of Public Health - National Institute of Hygiene | Gawor Jan; Wokowicz Tomasz; Zacharczuk Katarzyna |
| EPI_ISL_864580 | National Institute of Public Health - National Institute of Hygiene | National Institute of Public Health - National Institute of Hygiene, DNA Sequencing and Synthesis Facility (oligo.pl) - Institute of Biochemistry and Biophysics PAS | Gawor Jan; Wokowicz Tomasz; Zacharczuk Katarzyna |

| | | | |
|---|---|---|---|
| EPI_ISL_449799, EPI_ISL_846582, EPI_ISL_852616 | National Laboratory for Health, Environment and Food | National Laboratory for Health, Environment and Food | Aleksander Mahnic; Blazun Vosner H.; Duh D.; Hedzet S.; Janezic S.; Mahnic A.; Maja Rupnik; Rupnik M.; Sandra Janezic; Zavrsnik J. |
| EPI_ISL_512597 to 512612, EPI_ISL_512614 to 512637, EPI_ISL_512639 to 512644, EPI_ISL_512810 | National Laboratory for Influenza/Virology reference laboratory, Public Health Center of the Ministry of Health of Ukraine | Respiratory Virus Unit, Microbiology Services Colindale, Public Health England | Dr. Iryna Demchyshyna; PHE Covid Sequencing Team |
| EPI_ISL_483637, EPI_ISL_671414 to 671416, EPI_ISL_671418 | National Laboratory of Virology, Szentágothai Research Centre | National Laboratory of Virology, Szentágothai Research Centre | Balázs Somogyi; Brigitta; Endre Gábor Tóth; Ferenc Jakab; Gábor Kemenesi |
| EPI_ISL_416741, EPI_ISL_450496 to 450497 | National Public Health Surveillance Laboratory, Vilnius, Lithuania | Charite Universitaetsmedizin Berlin, Institute of Virology | Ana Steponkiene; Barbara Muehlemann; Christian Drosten; Jorn Beheim-Schwarzbach; Jorn Beheim-Schwarzbach; Julia Schneider; Talitha Veith; Terry Jones; Victor M Corman |
| EPI_ISL_480224, EPI_ISL_480297 to 480310 | National Reference Laboratory "Influenza and acute respiratory diseases" | NRL-HIV | Ivailo Alexiev; Ivan Ivanov; Ivva Philipova |
| EPI_ISL_507008 | National Veterinary Institute | National Veterinary Institute | Siamak Zohari |
| EPI_ISL_666688 to 666710 | National Virus Reference Lab (NVRL) | Irish Coronavirus Sequencing Consortium-Teagasc Grange | Aljandro Abner Garcia Leon; Calum Walsh; Fiona Crispie; Matthew McCabe; Michael Carr; Paul Cotter |
| EPI_ISL_605058 to 605076, EPI_ISL_605078 to 605080 | National Virus Reference Laboratory | Irish Coronavirus Sequencing Consortium - Helixworks | Conor Crosbie; Nimesh Pinnamaneni; Sachin Chalapati |
| EPI_ISL_848084 to 848098, EPI_ISL_848100 to 848171 | National Virus Reference Laboratory | Irish Coronavirus Sequencing Consortium - National University of Ireland Galway | Grainne Mc Andrew; Kate Reddington; Simone Coughlan |
| EPI_ISL_671337 to 671381, EPI_ISL_671383 to 671403, EPI_ISL_681885 to 681928, EPI_ISL_682057, EPI_ISL_763350, EPI_ISL_767717 to 767825, EPI_ISL_767827 to 767838, EPI_ISL_768744 to 768750, EPI_ISL_791280 to 791316, EPI_ISL_791318 to 791326, EPI_ISL_869146 to 869165, EPI_ISL_871815 | | | |
| see above | National Virus Reference Laboratory | Irish Coronavirus Sequencing Consortium - Teagasc Moorepark | Alejandro Abner Garcia Leon; Calm Walsh; Calum Walsh; Fiona Crispie; Genuity Ireland; John Kenny; Matthew McCabe; Paddy Mallon; Paul Cotter |
| EPI_ISL_708508 to 708520, EPI_ISL_708522, EPI_ISL_708524 to 708527 | National Virus Reference Laboratory | Irish Coronavirus Sequencing Consortium - Teagasc Oakpark | Ewen Mullins; Matthew McCabe; Michele Della Bartola |
| EPI_ISL_500573 to 500595, EPI_ISL_501252 to 501274, EPI_ISL_512087 to 512088, EPI_ISL_512090 to 512094, EPI_ISL_512096 to 512101, EPI_ISL_512103 to 512106, EPI_ISL_525352 to 525418, EPI_ISL_528440 to 528473, EPI_ISL_528475 to 528484, EPI_ISL_578202 to 578215, EPI_ISL_578217 to 578230, EPI_ISL_578232 to 578238, EPI_ISL_578241 to 578334, EPI_ISL_596889 to 596925, EPI_ISL_596927, EPI_ISL_639825 to 639903, EPI_ISL_639905 to 639908, EPI_ISL_644610, EPI_ISL_671864 to 671940, EPI_ISL_732374 to 732377, EPI_ISL_732379 to 732413, EPI_ISL_732415 to 732435, EPI_ISL_732438 to 732443, EPI_ISL_732445 to 732452, EPI_ISL_732457 to 732523, EPI_ISL_735384 to 735390, EPI_ISL_752512 to 752533, EPI_ISL_752535 to 752566, EPI_ISL_752568 to 752572, EPI_ISL_752574 to 752597, EPI_ISL_789063 to 789073, EPI_ISL_789076, EPI_ISL_789079, EPI_ISL_789083 to 789091, EPI_ISL_789093, EPI_ISL_789095, EPI_ISL_789098, EPI_ISL_837341 to 837346, EPI_ISL_837348, EPI_ISL_837350 to 837386, EPI_ISL_837388 to 837400, EPI_ISL_837402 to 837432 | | | |
| see above | National Virus Reference Laboratory | National Virus Reference Laboratory | Aditi Chaturvedi; Cillian F De Gascun; Dana Alalwan; Daniel Hare; Gabriel Gonzalez; Jonathan Dean; Michael Carr; Suzie Coughlan |
| EPI_ISL_516934, EPI_ISL_516936, EPI_ISL_516938 | Nicolae Testemitanu State University of Medicine and Pharmacy | International Centre for Genetic Engineering and Biotechnology (ICGEB) and ARGO Open Lab Platform for Genome Sequencing | Dal Monego S; Licastro D; Marcello A; Rajasekharan S; Ulinici M |
| EPI_ISL_420151 | Nordland Hospital - Bodo, Laboratory Department, Molecular Biology Unit | Norwegian Institute of Public Health, Department of Virology | Hilde Elshaug; Kamilla Heddeland Instefjord; Karoline Bragstad; Kathrine Stene-Johansen; Olav Hungnes |
| EPI_ISL_635142, EPI_ISL_635145, EPI_ISL_635182, EPI_ISL_668398 to 668399, EPI_ISL_668419, EPI_ISL_668422 to 668423, EPI_ISL_668429 to 668432, EPI_ISL_668434, EPI_ISL_708097 to 708105, EPI_ISL_775277, EPI_ISL_775281 to 775283, EPI_ISL_775286 to 775292, EPI_ISL_775295 to 775298, EPI_ISL_775301 to 775302, EPI_ISL_775312 to 775313, EPI_ISL_775319 to 775321, EPI_ISL_775330, EPI_ISL_775334, EPI_ISL_775372 to 775383, EPI_ISL_775398 to 775405, EPI_ISL_775438 to 775439, EPI_ISL_775480, EPI_ISL_775485 to 775487, EPI_ISL_796650, EPI_ISL_796667 to 796669, EPI_ISL_796672, EPI_ISL_796678, EPI_ISL_796689 to 796694 | | | |
| see above | Nordland Hospital - Bodo, Laboratory Department, Molecular Biology Unit | Norwegian Institute of Public Health, Department of Virology | Atiya R Ali; Hilde Elshaug; Hilde Vollan; Kamilla Heddeland Instefjord; Karoline Bragstad; Kathrine Stene-Johansen; Marie Paulsen Madsen; Olav Hungnes; Rasmus Riis Kopperud |
| EPI_ISL_534239 to 534243 | Norra Alvsborgs lanssjukhus | The Public Health Agency of Sweden | Anna Risberg; Anna-Malin Linde; Karin Tegmark-Wisell; Maria Lind Karlberg; Mattias Haukland; Mia Brytting; Olov Svartstrom; Oskar Karlsson Lindsjo; Petra Edquist; Reza Advani; Sandra Broddesson |
| EPI_ISL_482138 | North West London Pathology, Imperial College Healthcare NHS Trust | Wellcome Sanger Institute for the COVID-19 Genomics UK (COG-UK) Consortium | Aileen Rowan; Alison Holmes; Anjna Badhan; Carolina Herrera and Alex Alderton; Cordelia Langford; David K. Jackson; David Muir; Dominic Kwiatkowski; Ewan Harrison; Frankie Bolt; Graham Taylor; Ian Johnston; James Price; John Sillitoe on behalf of the Wellcome Sanger Institute COVID-19 Surveillance Team; Ling Li; Paul Randell; Roberto Amato; Sonia Goncalves |
| EPI_ISL_482124, EPI_ISL_482131, EPI_ISL_516831 to 516844, EPI_ISL_516846 to 516864, EPI_ISL_516866 to 516870, EPI_ISL_516872 to 516883, EPI_ISL_524619 to 524623, EPI_ISL_524625 to 524626, EPI_ISL_524628 to 524631, EPI_ISL_524633 to 524638, EPI_ISL_524640, EPI_ISL_524642 to 524643, EPI_ISL_524645 to 524649, EPI_ISL_524651 to 524656, EPI_ISL_524658 to 524677, EPI_ISL_524679 to 524699, EPI_ISL_524701 to 524712 | | | |
| see above | North West London Pathology, Imperial College Healthcare NHS Trust | Wellcome Sanger Institute for the COVID-19 Genomics UK (COG-UK) consortium | Aileen Rowan; Alison Holmes; Anjna Badhan; Carolina Herrera and Alex Alderton; Cordelia Langford; David K. Jackson; David Muir; Dominic Kwiatkowski; Ewan Harrison; Frankie Bolt; Graham Taylor; Ian Johnston; James Price; John Sillitoe on behalf of the Wellcome Sanger Institute COVID-19 Surveillance Team (http://www.sanger.ac.uk/covid-team); Ling Li; Paul Randell; Roberto Amato; Sonia Goncalves |
| EPI_ISL_453486 to 453511, EPI_ISL_453513 to 453531, EPI_ISL_453533 to 453542, EPI_ISL_453544, EPI_ISL_457303 to 457319, EPI_ISL_457321 to 457325, EPI_ISL_461791 to 461804, EPI_ISL_472155 to 472160, EPI_ISL_472162 to 472174, EPI_ISL_472176 to 472187, EPI_ISL_472189 to 472237, EPI_ISL_472240 to 472290, EPI_ISL_478473 to 478498, EPI_ISL_478500 to 478511, EPI_ISL_478513, EPI_ISL_478516 to 478518, EPI_ISL_478520 to 478532, EPI_ISL_478534 to 478614, EPI_ISL_478616 to 478622, EPI_ISL_478624 to 478629, EPI_ISL_478631 to 478638, EPI_ISL_478641 to 478642, EPI_ISL_478644 to 478652, EPI_ISL_478654 to 478668, EPI_ISL_484264 to 484327, EPI_ISL_490440 to 490454, EPI_ISL_490457 to 490476, EPI_ISL_490478 to 490491, EPI_ISL_490493 to 490495, EPI_ISL_493452 to 493500, EPI_ISL_493502 to 493519, EPI_ISL_493521 to 493540, EPI_ISL_499472, EPI_ISL_499474, EPI_ISL_499478, EPI_ISL_499485, EPI_ISL_499493 to 499494, EPI_ISL_499502 to 499504, EPI_ISL_499506 to 499508, EPI_ISL_499510, EPI_ISL_499515, EPI_ISL_499546 to 499553, EPI_ISL_499555 to 499556, EPI_ISL_499558 to 499561, EPI_ISL_499563, EPI_ISL_499568, EPI_ISL_499572, EPI_ISL_499587, EPI_ISL_499595 to 499598, EPI_ISL_499602, EPI_ISL_499606, EPI_ISL_499609 to 499610, EPI_ISL_499626, EPI_ISL_499777 to 499807, EPI_ISL_507110 to 507129, EPI_ISL_512348 to 512349, EPI_ISL_512352, EPI_ISL_512354 to 512358, EPI_ISL_512361 to 512365, EPI_ISL_572409 to 572410, EPI_ISL_572424, EPI_ISL_572438 to 572439, EPI_ISL_572450, EPI_ISL_572452, EPI_ISL_572464, EPI_ISL_572480, EPI_ISL_572488, EPI_ISL_572513, EPI_ISL_572515, EPI_ISL_572517, EPI_ISL_572525, EPI_ISL_572530, EPI_ISL_572532, EPI_ISL_572568, EPI_ISL_572570 to 572572, EPI_ISL_572579, EPI_ISL_572581, EPI_ISL_572594 to 572597, EPI_ISL_572648, EPI_ISL_572663, EPI_ISL_572700 to 572704, EPI_ISL_572712 to 572713, EPI_ISL_572744, EPI_ISL_572747, EPI_ISL_572751, EPI_ISL_572753, EPI_ISL_572783, EPI_ISL_572795, EPI_ISL_572798, EPI_ISL_572802, EPI_ISL_572845, EPI_ISL_572909 to 572911, EPI_ISL_572942, EPI_ISL_572950, EPI_ISL_573266 to 573323, EPI_ISL_573325 to 573330, EPI_ISL_573332 to 573333, EPI_ISL_573335 to 573358, EPI_ISL_573360 to 573363, EPI_ISL_573365 to 573413, EPI_ISL_573760 to 573761, EPI_ISL_573799, EPI_ISL_573871 to 573872, EPI_ISL_576986 to 577000, EPI_ISL_577002 to 577007, EPI_ISL_577009 to 577039, EPI_ISL_584681 to 584702, EPI_ISL_584704 to 584715, EPI_ISL_584847 to 584882, EPI_ISL_585504 to 585532, EPI_ISL_594987 to 594991, EPI_ISL_594993 to 595005, EPI_ISL_595007 to 595028, EPI_ISL_595030 to 595045, EPI_ISL_611524, EPI_ISL_611572, EPI_ISL_611574, EPI_ISL_611576 to 611577, EPI_ISL_611591, EPI_ISL_611643 to 611649, EPI_ISL_611657, EPI_ISL_611668, EPI_ISL_611746 to 611747, EPI_ISL_611752, EPI_ISL_611756 to 611757, EPI_ISL_611759 to 611760, EPI_ISL_611766 to 611770, EPI_ISL_611839, EPI_ISL_611846, EPI_ISL_611864, EPI_ISL_611868, EPI_ISL_611917, EPI_ISL_611930, EPI_ISL_611941 to 611953, EPI_ISL_611980, EPI_ISL_612008, EPI_ISL_612026 to 612027, EPI_ISL_612030, EPI_ISL_612054, EPI_ISL_612085, EPI_ISL_612090, EPI_ISL_612097, EPI_ISL_612100, EPI_ISL_612104, EPI_ISL_612134, EPI_ISL_612523 to 612545, EPI_ISL_612549 to 612570, EPI_ISL_637280, EPI_ISL_637283, EPI_ISL_637311, EPI_ISL_637322, EPI_ISL_637352, EPI_ISL_637373 to 637377, EPI_ISL_637455, EPI_ISL_637520 to 637523, EPI_ISL_637581, EPI_ISL_637605, EPI_ISL_637784 to 637797, EPI_ISL_637883, EPI_ISL_637902, EPI_ISL_637943, EPI_ISL_637979, EPI_ISL_637994, EPI_ISL_638009 to 638010, EPI_ISL_638555 to 638562, EPI_ISL_638564, EPI_ISL_638566 to 638580, EPI_ISL_650125, EPI_ISL_650142, EPI_ISL_650147 to 650148, EPI_ISL_650156, EPI_ISL_650241, EPI_ISL_650246 to 650247, EPI_ISL_650279, EPI_ISL_650282, EPI_ISL_650348, EPI_ISL_650351, EPI_ISL_650355, EPI_ISL_650359, EPI_ISL_650393 to 650394, EPI_ISL_650428, EPI_ISL_650572, EPI_ISL_650586 to 650588, EPI_ISL_650624 to 650625, EPI_ISL_650640, EPI_ISL_650684, EPI_ISL_650703, EPI_ISL_650785, EPI_ISL_650950, EPI_ISL_650967 to 650968, EPI_ISL_651013, EPI_ISL_651060 to 651063, EPI_ISL_651117, EPI_ISL_651126, EPI_ISL_651314, EPI_ISL_651345, EPI_ISL_651354, EPI_ISL_651356, EPI_ISL_651369, EPI_ISL_651388, EPI_ISL_652111 to 652116, EPI_ISL_652337 to 652344, EPI_ISL_664107, EPI_ISL_664121, EPI_ISL_664190, EPI_ISL_664192, EPI_ISL_664238, EPI_ISL_664240 to 664247, EPI_ISL_664360, EPI_ISL_664373, EPI_ISL_664385 to 664396, EPI_ISL_664503, EPI_ISL_664541, EPI_ISL_664544, EPI_ISL_664560 to 664564, EPI_ISL_664639, EPI_ISL_664696, EPI_ISL_664856, EPI_ISL_665261 to 665262, EPI_ISL_665264, EPI_ISL_665271 to 665272, EPI_ISL_665275, EPI_ISL_665278, EPI_ISL_665280, EPI_ISL_665300, EPI_ISL_665303, EPI_ISL_665305, EPI_ISL_665307 to 665308, EPI_ISL_665310 to 665311, EPI_ISL_665313 to 665314, EPI_ISL_665317 to 665319, EPI_ISL_665322 to 665323, EPI_ISL_665329, EPI_ISL_665349 to 665350, EPI_ISL_665368, EPI_ISL_665379, EPI_ISL_665387 to 665390, EPI_ISL_665392 to 665405, EPI_ISL_665407 to 665410, EPI_ISL_665412, EPI_ISL_665432, EPI_ISL_665489 to 665503, EPI_ISL_665506, EPI_ISL_665509, EPI_ISL_665540, EPI_ISL_665600, EPI_ISL_665617 to 665618, EPI_ISL_665620 to 665621, EPI_ISL_665626 to 665631, EPI_ISL_665651, EPI_ISL_665667, EPI_ISL_665673, EPI_ISL_665676, EPI_ISL_665679 to 665681, EPI_ISL_665687, EPI_ISL_665701, EPI_ISL_665706, EPI_ISL_665717 to 665718, EPI_ISL_665735 to 665736, EPI_ISL_665739, EPI_ISL_665742, EPI_ISL_665744, EPI_ISL_665784, EPI_ISL_665787, EPI_ISL_665868 to 665875, EPI_ISL_665964 to 665970, EPI_ISL_665983 to 666012, EPI_ISL_666014 to 666021, EPI_ISL_666084 to 666106, EPI_ISL_666591 to 666592, EPI_ISL_702495, EPI_ISL_702505, EPI_ISL_702520, EPI_ISL_702525, EPI_ISL_702533, EPI_ISL_702539, EPI_ISL_702547, EPI_ISL_702747, EPI_ISL_702784, EPI_ISL_702803, EPI_ISL_702809, EPI_ISL_702812, EPI_ISL_702815, EPI_ISL_702820, EPI_ISL_702823, EPI_ISL_702827, EPI_ISL_702835 to 702836, EPI_ISL_702855, EPI_ISL_702959, EPI_ISL_702982, EPI_ISL_702990, EPI_ISL_703002, EPI_ISL_703005, EPI_ISL_703007, EPI_ISL_703010, EPI_ISL_703013, EPI_ISL_703034, EPI_ISL_703064, EPI_ISL_703096, EPI_ISL_703105, EPI_ISL_703377, EPI_ISL_703390, EPI_ISL_703484, EPI_ISL_703487, EPI_ISL_703533, EPI_ISL_703535, EPI_ISL_703537, EPI_ISL_703540, EPI_ISL_703542, EPI_ISL_703545, EPI_ISL_703599, EPI_ISL_703701, EPI_ISL_703704, EPI_ISL_703727 to 703728, EPI_ISL_704120, EPI_ISL_704166, EPI_ISL_704198, EPI_ISL_704262, EPI_ISL_704387, EPI_ISL_704422, EPI_ISL_704425, EPI_ISL_704564, EPI_ISL_704567, EPI_ISL_704570, EPI_ISL_704572 to 704579, EPI_ISL_704976, EPI_ISL_705010, EPI_ISL_705067, EPI_ISL_705102, EPI_ISL_705105, EPI_ISL_705137, EPI_ISL_705145, EPI_ISL_705148, EPI_ISL_705201, EPI_ISL_705207, EPI_ISL_705212, EPI_ISL_705214, EPI_ISL_705219, EPI_ISL_705224 to 705226, EPI_ISL_705266, EPI_ISL_705273, EPI_ISL_705279 to 705281, EPI_ISL_705293, EPI_ISL_705295, EPI_ISL_705302, EPI_ISL_705341, EPI_ISL_705343, EPI_ISL_705353, EPI_ISL_705355 to 705357, EPI_ISL_705361, EPI_ISL_705364, EPI_ISL_705367, EPI_ISL_705381, EPI_ISL_705457 to 705458, EPI_ISL_705486, EPI_ISL_705494 to 705496, EPI_ISL_706276 to 706282, EPI_ISL_706284 to 706286, EPI_ISL_706288 to 706314, EPI_ISL_706316 to 706339, EPI_ISL_706341 to 706354, EPI_ISL_723558 to 723622, EPI_ISL_723624 to 723630, EPI_ISL_724063 to 724104, EPI_ISL_724106 to 724123, EPI_ISL_724837, EPI_ISL_724841 to 724842, EPI_ISL_724845, EPI_ISL_724848 to 724976, EPI_ISL_724978 to 724983, EPI_ISL_724985 to 724994, EPI_ISL_724996 to 725028, EPI_ISL_741425 to 741452, EPI_ISL_819448 to 819457, EPI_ISL_819460 to 819461, EPI_ISL_819568 to 819570, EPI_ISL_819572 to 819576, EPI_ISL_839484 to 839485, EPI_ISL_839488, EPI_ISL_839490 to 839491, EPI_ISL_839493 to 839494, EPI_ISL_839497, EPI_ISL_839500, EPI_ISL_839507 to 839510, EPI_ISL_839516 to 839518, EPI_ISL_839520 to 839523, EPI_ISL_839525 to 839548, EPI_ISL_839550 to 839556, EPI_ISL_839558 to 839566, EPI_ISL_839568 to 839572, EPI_ISL_839574, EPI_ISL_839576 to 839581, EPI_ISL_839583, EPI_ISL_839585 to 839592, EPI_ISL_839600, EPI_ISL_839603, EPI_ISL_839607 to 839634, EPI_ISL_839636 to 839642, EPI_ISL_839644 to 839662, EPI_ISL_839664 to 839666, EPI_ISL_839669 to 839671, EPI_ISL_839673 to 839675, EPI_ISL_839677 to 839681, EPI_ISL_839683 to 839684, EPI_ISL_839686 to 839698, EPI_ISL_839702, EPI_ISL_839705 to 839708, EPI_ISL_839710 to 839712, EPI_ISL_839714 to 839719, EPI_ISL_839721 to 839726, EPI_ISL_839728 to 839732, EPI_ISL_839735 to 839745, EPI_ISL_839747 to 839777, EPI_ISL_839779 to 839816, EPI_ISL_839818 to 839822, | | | |

| | | | |
|---|---|---|---|
| EPI_ISL_866400, EPI_ISL_866402 to 866418, EPI_ISL_866420 to 866424, EPI_ISL_866427, EPI_ISL_866430, EPI_ISL_866433 to 866434, EPI_ISL_866436, EPI_ISL_866438, EPI_ISL_866440, EPI_ISL_866443 to 866446, EPI_ISL_866449 to 866454, EPI_ISL_866456, EPI_ISL_866463 to 866483, EPI_ISL_866485 to 866487, EPI_ISL_866489 to 866499, EPI_ISL_866501 to 866504, EPI_ISL_866506 to 866508, EPI_ISL_866510 to 866512, EPI_ISL_866515 to 866519, EPI_ISL_866522, EPI_ISL_866524 to 866525 | | | |
| see above | Northumbria University / South Tees Hospitals NHS Foundation Trust / North Cumbria Integrated Care NHS Foundation Trust / North Tees and Hartlepool NHS Foundation Trust / Newcastle Hospitals NHS Foundation Trust | COVID-19 Genomics UK (COG-UK) Consortium | Andrew Nelson; Brendan Payne; Clive Graham; Darren L Smith; Debra Padgett; Edward Barton; Emma Swindells; Garren Scott; Gary Black; Gary Eltringham; Giles S Holt; Greg R Young; Jane Greenaway; Jennifer Collins; John Allan; Joshua Loh; Lynn Dover; Matthew Bashton; Mohammad A Tariq; Paul Baker; Sarah Essex; Steve Liggett; Wen C Yew; Yusri Taha |
| EPI_ISL_453543 | Northumbria University / South Tees Hospitals NHS Foundation Trust / North Cumbria Integrated Care NHS Foundation Trust / North Tees and Hartlepool NHS Foundation Trust / Newcastle Hospitals NHS Foundation Trust | Northumbria University | Andrew Nelson; Brendan Payne; Clive Graham; Darren L Smith; Debra Padgett; Edward Barton; Emma Swindells; Garren Scott; Gary Black; Gary Eltringham; Giles S Holt; Greg R Young; Jane Greenaway; Jennifer Collins; John Allan; Joshua Loh; Lynn Dover; Matthew Bashton; Mohammad A Tariq; Paul Baker; Sarah Essex; Steve Liggett; Wen C Yew; Yusri Taha |
| EPI_ISL_635066 to 635067, EPI_ISL_635074 to 635079, EPI_ISL_635178, EPI_ISL_635187 to 635188, EPI_ISL_668410 to 668411, EPI_ISL_668435 to 668437, EPI_ISL_708028 to 708029, EPI_ISL_708040, EPI_ISL_708049 to 708051, EPI_ISL_708140 to 708141, EPI_ISL_708169 to 708170, EPI_ISL_708180 to 708181, EPI_ISL_759981, EPI_ISL_759989, EPI_ISL_775356, EPI_ISL_775414, EPI_ISL_775443 to 775444, EPI_ISL_775459 to 775462, EPI_ISL_775509 to 775515, EPI_ISL_796654 to 796657, EPI_ISL_796659 to 796662, EPI_ISL_796675 to 796676, EPI_ISL_796684 to 796685, EPI_ISL_796710, EPI_ISL_813976, EPI_ISL_842633, EPI_ISL_860209, EPI_ISL_860233 to 860234 | | | |
| see above | Norwegian Institute of Public Health, Department of Virology | Norwegian Institute of Public Health, Department of Virology | Atiya R Ali; Hilde Elshaug; Hilde Vollan; Kamilla Heddeland Instefjord; Karoline Bragstad; Kathrine Stene-Johansen; Marie Paulsen Madsen; Olav Hungnes; Rasmus Riis Kopperud |
| EPI_ISL_833047 to 833048, EPI_ISL_833077 to 833078 | OSPEDALE CIVILE ATRI | Istituto Zooprofilattico Sperimentale dell'Abruzzo e Molise "G. Caporale" | Ancora M; Calistri P; Cammà C; Curini V; Delli Compagni E; Di Domenico M; Di Pasquale A; Lorusso A; Mangone I; Marcacci M; Puglia I; Rinaldi A; Savini G |
| EPI_ISL_833079 to 833080 | OSPEDALE CIVILE TERAMO | Istituto Zooprofilattico Sperimentale dell'Abruzzo e Molise "G. Caporale" | Ancora M; Calistri P; Cammà C; Curini V; Delli Compagni E; Di Domenico M; Di Pasquale A; Lorusso A; Mangone I; Marcacci M; Puglia I; Rinaldi A; Savini G |
| EPI_ISL_833081 | OSPEDALE S.S. ANNUNZIATA CHIETI | Istituto Zooprofilattico Sperimentale dell'Abruzzo e Molise "G. Caporale" | Ancora M; Calistri P; Cammà C; Curini V; Delli Compagni E; Di Domenico M; Di Pasquale A; Lorusso A; Mangone I; Marcacci M; Puglia I; Rinaldi A; Savini G |
| EPI_ISL_833230 | OSPEDALE S.S. ANNUNZIATA CHIETI - CLINICA MEDICA (MEDICINA GENERALE 1) | Istitituto Zooprofilattico Sperimentale dell'Abruzzo e Molise "G.Caporale" | Ancora M; Calistri P; Cammà C; Curini V; Di Domenico M; Di Pasquale A; Lorusso A; Mangone I; Marcacci M; Puglia I; Rinaldi A; Savini G. |
| EPI_ISL_833082 | OSPEDALE SAN SALVATORE | Istituto Zooprofilattico Sperimentale dell'Abruzzo e Molise "G. Caporale" | Ancora M; Calistri P; Cammà C; Curini V; Delli Compagni E; Di Domenico M; Di Pasquale A; Lorusso A; Mangone I; Marcacci M; Puglia I; Rinaldi A; Savini G |
| EPI_ISL_833042 | OSPEDALE SAN SALVATORE L'AQUILA UOC PNEUMOLOGIAE UTSIR | Istituto Zooprofilattico Sperimentale dell'Abruzzo e Molise "G. Caporale" | Ancora M; Calistri P; Cammà C; Curini V; Delli Compagni E; Di Domenico M; Di Pasquale A; Lorusso A; Mangone I; Marcacci M; Puglia I; Rinaldi A; Savini G |
| EPI_ISL_455111 | Olof Norrby | The Public Health Agency of Sweden | Anna Risberg; Anna-Malin Linde; Bla Kustens halsocentral; Karin Tegmark-Wisell; Maria Lind Karlberg; Mia Brytting; Olov Svartstrom; Oskar Karlsson Lindsjo; Theresa Enkirch |
| EPI_ISL_569735 to 569857, EPI_ISL_629031 to 629034, EPI_ISL_639912 to 639947, EPI_ISL_733482 to 733493 | Omsk Research Institute of Natural Focal Infections | WHO National Influenza Centre Russian Federation | Aleksei Vasilenko; Andrey Komissarov; Anna Ivanova; Artem Fadeev; Daria Danilenko; Daria Nashatyreva; Dmitry Bazhenov; Dmitry Lioznov; Ekaterina Gradoboeva; Ekaterina Savkina; Elena Nabieva; Elena Poleshchuk; Georgii Bazykin; Ksenia Safina; Kseniya Komissarova; Valery Yakimenko |
| EPI_ISL_434661 to 434662, EPI_ISL_434664, EPI_ISL_434673, EPI_ISL_475528, EPI_ISL_475533 to 475535, EPI_ISL_475538, EPI_ISL_539883 to 539884, EPI_ISL_710587 to 710588 | | | |
| see above | Omtanken Grimmered | The Public Health Agency of Sweden | Anna Risberg; Anna-Malin Linde; Bernd Sengpiel; Department of Microbiology; Karin Tegmark-Wisell; Maria Lind Karlberg; Mattias Haukland; Mia Brytting; Olov Svartstrom; Oskar Karlsson Lindsjo; Reza Advani; Sandra Broddesson; The Public Health Agency of Sweden; Theresa Enkirch |
| EPI_ISL_648207 to 648208 | Orebro | The Public Health Agency of Sweden | Anna Risberg; Anna-Malin Linde; Karin Tegmark-Wisell; Maria Lind Karlberg; Mattias Haukland; Mia Brytting; Olov Svartstrom; Oskar Karlsson Lindsjo; Petra Edquist; Reza Advani; Sandra Broddesson |
| EPI_ISL_455840 to 455841, EPI_ISL_475128 to 475135, EPI_ISL_475139, EPI_ISL_510817, EPI_ISL_560972, EPI_ISL_582785 to 582789, EPI_ISL_582791 to 582793, EPI_ISL_582841, EPI_ISL_615079, EPI_ISL_615111 to 615114, EPI_ISL_615120, EPI_ISL_648204 to 648206, EPI_ISL_654943, EPI_ISL_660378 to 660383, EPI_ISL_660422 to 660426, EPI_ISL_661282, EPI_ISL_676493, EPI_ISL_676513, EPI_ISL_831960 to 831968, EPI_ISL_831970 to 831972, EPI_ISL_831976 | | | |
| see above | Orebro klinisk mikrobiologi | The Public Health Agency of Sweden | Anna Risberg; Anna-Malin Linde; Department of Microbiology; Karin Tegmark-Wisell; Maria Lind Karlberg; Mattias Haukland; Mia Brytting; Olov Svartstrom; Oskar Karlsson Lindsjo; Petra Edquist; Reza Advani; Sandra Broddesson; Shamam Muradrasoli; The Public Health Agency of Sweden |
| EPI_ISL_475511 | Orestadsklinikens VC | The Public Health Agency of Sweden | Anna Risberg; Anna-Malin Linde; Karin Tegmark-Wisell; Maria Lind Karlberg; Mattias Haukland; Mia Brytting; Olov Svartstrom; Oskar Karlsson Lindsjo; Reza Advani; Sandra Broddesson |
| EPI_ISL_840370 to 840372, EPI_ISL_840374 to 840378, EPI_ISL_840380 to 840385, EPI_ISL_840387 to 840503, EPI_ISL_840505 to 840507, EPI_ISL_840509 to 840510, EPI_ISL_840514, EPI_ISL_840516 to 840551, EPI_ISL_840553, EPI_ISL_840556 to 840569, EPI_ISL_840571 to 840578, EPI_ISL_840581 to 840583, EPI_ISL_840585, EPI_ISL_840587 to 840589, EPI_ISL_840591 to 840595, EPI_ISL_840599 to 840600, EPI_ISL_840604, EPI_ISL_840606, EPI_ISL_840609 to 840612, EPI_ISL_840615, EPI_ISL_840617 to 840636, EPI_ISL_840638 to 840641, EPI_ISL_840643 to 840649, EPI_ISL_840651, EPI_ISL_840653, EPI_ISL_840655 to 840658, EPI_ISL_840662, EPI_ISL_840667 to 840672, EPI_ISL_840695 to 840703, EPI_ISL_840706 to 840710, EPI_ISL_840712 to 840714, EPI_ISL_840716, EPI_ISL_840719 to 840720, EPI_ISL_840723, EPI_ISL_840725 to 840726, EPI_ISL_840728 to 840730, EPI_ISL_840733 to 840734, EPI_ISL_840737 to 840740, EPI_ISL_840742 to 840746, EPI_ISL_840749 to 840752, EPI_ISL_840754, EPI_ISL_840756 to 840767, EPI_ISL_840769, EPI_ISL_840774 to 840779, EPI_ISL_840781 to 840783, EPI_ISL_840785, EPI_ISL_840787, EPI_ISL_840790, EPI_ISL_840794 to 840796, EPI_ISL_840798, EPI_ISL_840801 to 840806, EPI_ISL_840808, EPI_ISL_840810, EPI_ISL_841321, EPI_ISL_841355 to 841357, EPI_ISL_841359 to 841361, EPI_ISL_841503 to 841505, EPI_ISL_841507, EPI_ISL_841521 to 841522, EPI_ISL_841529, EPI_ISL_841531, EPI_ISL_841559 to 841566, EPI_ISL_841568 to 841575, EPI_ISL_841577, EPI_ISL_841579 to 841581, EPI_ISL_841584, EPI_ISL_841587, EPI_ISL_867198 to 867201, EPI_ISL_867216 to 867218, EPI_ISL_867271 to 867297, EPI_ISL_867427, EPI_ISL_867437 to 867438, EPI_ISL_867440 to 867442, EPI_ISL_867499 to 867501 | | | |
| see above | Originating lab: Wales Specialist Virology Centre Sequencing lab: Pathogen Genomics Unit | Public Health Wales Microbiology Cardiff Wales Specialist Virology Centre | Alec Birchley; Alexander Adams; Amy Gaskin; Angela Marchbank; Bree Gatica-Wilcox; Catherine Moore; Jason Coombes; Joanne Watkins; Joel Southgate; Johnathan Evans; Laura Gifford; Lauren Gilbert; Lee Graham; Malorie Perry; Matthew Bull; Nicole Pacchiarini; Sally Corden; Sara Kumziene-Summerhayes; Sara Rey; Sarah Taylor; Simon Cottrell; Sophie Jones; Tom Connor |
| EPI_ISL_455100, EPI_ISL_475519, EPI_ISL_710585 to 710586 | Orsa VC | The Public Health Agency of Sweden | Anna Risberg; Anna-Karin Lundqvist; Anna-Malin Linde; Department of Microbiology; Karin Tegmark-Wisell; Maria Lind Karlberg; Mattias Haukland; Mia Brytting; Olov Svartstrom; Oskar Karlsson Lindsjo; Reza Advani; Sandra Broddesson; The Public Health Agency of Sweden; Theresa Enkirch |
| EPI_ISL_417483 | Oslo University Hospital, Department of Medical Microbiology | Norwegian Institute of Public Health | Hilde Elshaug; Kamilla Heddeland Instefjord; Karoline Bragstad; Kathrine Stene-Johansen; Olav Hungnes |
| EPI_ISL_417488 to 417490, EPI_ISL_420135, EPI_ISL_420150, EPI_ISL_471175, EPI_ISL_471177, EPI_ISL_481215, EPI_ISL_481217 to 481218, EPI_ISL_493351 to 493355, EPI_ISL_493385 to 493389, EPI_ISL_500800, EPI_ISL_549029 to 549035, EPI_ISL_590878, EPI_ISL_590900, EPI_ISL_590908 to 590910, EPI_ISL_590913 to 590914, EPI_ISL_590916, EPI_ISL_590918, EPI_ISL_590923, EPI_ISL_590927 to 590928, EPI_ISL_590930, EPI_ISL_590934 to 590935, EPI_ISL_590978 to 590980, EPI_ISL_591011, EPI_ISL_591020, EPI_ISL_635085, EPI_ISL_635093 to 635095, EPI_ISL_635119, EPI_ISL_635129, EPI_ISL_635155, EPI_ISL_635172, EPI_ISL_635197, EPI_ISL_668401 to 668402, EPI_ISL_708033 to 708034, EPI_ISL_708052 to 708054, EPI_ISL_708096, EPI_ISL_708118 to 708121, EPI_ISL_708171 to 708175, EPI_ISL_759982, EPI_ISL_775278, EPI_ISL_775284, EPI_ISL_775293, EPI_ISL_775307 to 775308, EPI_ISL_775316 to 775318, EPI_ISL_775391 to 775392, EPI_ISL_775410 to 775413, EPI_ISL_775415 to 775426, EPI_ISL_775457, EPI_ISL_860220 to 860221, EPI_ISL_860235 | | | |
| see above | Oslo University Hospital, Department of Medical Microbiology | Norwegian Institute of Public Health, Department of Virology | Atiya R Ali; Hilde Elshaug; Hilde Synnøve Vollan; Hilde Vollan; Kamilla Heddeland Instefjord; Karoline Bragstad; Kathrine Stene-Johansen; Marie Paulsen Madsen; Olav Hungnes; Rasmus Riis Kopperud |
| EPI_ISL_788943 to 788965, EPI_ISL_788968 to 788978, EPI_ISL_872003 to 872036 | Ospedale "Di Venere" | Beaconlab (Bioinformatics, Evolution and Comparative Genomics lab), Dept of Biosciences, University on Milan | Chiara M; Iacobellis M; Manzari C; Parisi A; Pesole G; Piluscio R; d'Avenia M |
| EPI_ISL_528922, EPI_ISL_528924, EPI_ISL_528926 to 528928, EPI_ISL_529018, EPI_ISL_529026 | Ospedale "Giuseppe Mazzini"-Teramo | Istitituto Zooprofilattico Sperimentale dell'Abruzzo e Molise "G.Caporale" | Ancora M; Cammà C; Curini V; Di Domenico M; Di Pasquale A; Lorusso A; Mangone I; Marcacci M; Puglia I; Rinaldi A; Savini G. |
| EPI_ISL_529014 to 529015, EPI_ISL_529022 | Ospedale "Ss. Annunziata" | Istitituto Zooprofilattico Sperimentale dell'Abruzzo e Molise "G.Caporale" | Ancora M; Cammà C; Curini V; Di Domenico M; Di Pasquale A; Lorusso A; Mangone I; Marcacci M; Puglia I; Rinaldi A; Savini G. |
| EPI_ISL_794753 | Ospedale Cardinale G. Panico | Istituto Zooprofilattico Sperimentale della Puglia e della Basilicata | Bianco A.; Capozzi L.; Cipolletta D.; Del Sambro L.; Galante D.; Manzulli V; Pace L.; Parisi A.; Rondinone V. |
| EPI_ISL_832818 | Ospedale Civile Atri | Istitituto Zooprofilattico Sperimentale dell'Abruzzo e Molise "G.Caporale" | Ancora M; Calistri P; Cammà C; Curini V; Di Domenico M; Di Pasquale A; Lorusso A; Mangone I; Marcacci M; Puglia I; Rinaldi A; Savini G. |
| EPI_ISL_833255 to 833257, EPI_ISL_833331 | Ospedale Civile Atri | Istituto Zooprofilattico Sperimentale dell'Abruzzo e Molise "G. | Ancora M; Calistri P; Cammà C; Curini V; Di Domenico M; Di Pasquale A; Lorusso A; Mangone I; Marcacci M; Puglia I; Rinaldi A; Savini G |

| | | | |
|---|---|---|---|
| | | Caporale" | |
| EPI_ISL_833251 | Ospedale Civile Atri-Medicina Interna | Istituto Zooprofilattico Sperimentale dell'Abruzzo e Molise "G. Caporale" | Ancora M; Calistri P; Cammà C; Curini V; Di Domenico M; Di Pasquale A; Lorusso A; Mangone I; Marcacci M; Puglia I; Rinaldi A; Savini G |
| EPI_ISL_420564 | Ospedale Civile Castel Di Sangro | Istituto Zooprofilattico Sperimentale dell'Abruzzo e Molise "G.Caporale" | Ancora M; Cammà C; Curini V; Di Domenico M; Di Pasquale A; Lorusso A; Mangone I; Marcacci M; Puglia I; Rinaldi A; Savini G |
| EPI_ISL_806784 to 806785, EPI_ISL_833276 | Ospedale Civile G.Mazzini-Teramo | Istituto Zooprofilattico Sperimentale dell'Abruzzo e Molise "G. Caporale" | Ancora M; Calistri P; Cammà C; Curini V; Di Domenico M; Di Pasquale A; Lorusso A; Mangone I; Marcacci M; Puglia I; Rinaldi A; Savini G |
| EPI_ISL_806789, EPI_ISL_833297 | Ospedale Civile Giulianova | Istituto Zooprofilattico Sperimentale dell'Abruzzo e Molise "G. Caporale" | Ancora M; Calistri P; Cammà C; Curini V; Di Domenico M; Di Pasquale A; Lorusso A; Mangone I; Marcacci M; Puglia I; Rinaldi A; Savini G |
| EPI_ISL_833252 | Ospedale Civile Giulianova-Pronto Soccorso | Istituto Zooprofilattico Sperimentale dell'Abruzzo e Molise "G. Caporale" | Ancora M; Calistri P; Cammà C; Curini V; Di Domenico M; Di Pasquale A; Lorusso A; Mangone I; Marcacci M; Puglia I; Rinaldi A; Savini G |
| EPI_ISL_418260 to 418261, EPI_ISL_420563, EPI_ISL_429228, EPI_ISL_429231 to 429235 | Ospedale Civile Giuseppe Mazzini | Istituto Zooprofilattico Sperimentale dell'Abruzzo e Molise "G. Caporale" | Ancora M; Camma C; Cammà C; Curini V; Di Domenico M; Di Pasquale A; Lorusso A; Mangone I; Marcacci M; Puglia I; Rinaldi A; Savini G; Savini G. |
| EPI_ISL_420565, EPI_ISL_420568 to 420569, EPI_ISL_420583, EPI_ISL_420592, EPI_ISL_435145, EPI_ISL_436723 | Ospedale Civile Giuseppe Mazzini | Istituto Zooprofilattico Sperimentale dell'Abruzzo e Molise "G.Caporale" | Ancora M; Cammà C; Curini V; Di Domenico M; Di Pasquale A; Lorusso A; Mangone I; Marcacci M; Puglia I; Rinaldi A; Savini G |
| EPI_ISL_418257 | Ospedale Civile Giuseppe Mazzini, Teramo | Istituto Zooprofilattico Sperimentale dell'Abruzzo e Molise "G.Caporale" | Ancora M; Cammà C; Curini V; Di Domenico M; Di Pasquale A; Lorusso A; Mangone I; Marcacci M; Puglia I; Rinaldi A; Savini G. |
| EPI_ISL_528990 | Ospedale Civile Maria SS. dello Splendore | Istitituto Zooprofilattico Sperimentale dell'Abruzzo e Molise "G.Caporale" | Ancora M; Cammà C; Curini V; Di Domenico M; Di Pasquale A; Lorusso A; Mangone I; Marcacci M; Puglia I; Rinaldi A; Savini G. |
| EPI_ISL_429236 | Ospedale Civile S. Liberatore di Atri | Istituto Zooprofilattico Sperimentale dell'Abruzzo e Molise "G. Caporale" | Ancora M; Camma C; Curini V; Di Domenico M; Di Pasquale A; Lorusso A; Mangone I; Marcacci M; Puglia I; Rinaldi A; Savini G |
| EPI_ISL_436719 to 436722, EPI_ISL_436724, EPI_ISL_436731 to 436732 | Ospedale Civile S. Liberatore di Atri | Istituto Zooprofilattico Sperimentale dell'Abruzzo e Molise "G.Caporale" | Ancora M; Cammà C; Curini V; Di Domenico M; Di Pasquale A; Lorusso A; Mangone I; Marcacci M; Puglia I; Rinaldi A; Savini G |
| EPI_ISL_528919, EPI_ISL_528923, EPI_ISL_528929, EPI_ISL_528993, EPI_ISL_529007, EPI_ISL_529009, EPI_ISL_529020 to 529021 | | | |
| see above | Ospedale Civile S. Liberatore-Atri | Istitituto Zooprofilattico Sperimentale dell'Abruzzo e Molise "G.Caporale" | Ancora M; Cammà C; Curini V; Di Domenico M; Di Pasquale A; Lorusso A; Mangone I; Marcacci M; Puglia I; Rinaldi A; Savini G. |
| EPI_ISL_745192 to 745193 | Ospedale Di Venere | Istituto Zooprofilattico Sperimentale della Puglia e della Basilicata | Bianco A.; Capozzi L.; Chiara M.; De Sabato L.; Del Sambro L.; Iacobellis M; Iacobellis M.; Parisi A.; Pesole G. |
| EPI_ISL_747465 to 747470, EPI_ISL_747484 to 747485 | Ospedale Mater Salutis | Istituto Zooprofilattico Sperimentale delle Venezie | Adelaide Milani; Alessia Schivo; Alice Fusaro; Ambra Pastori; Annalisa Salviato; Antonia Ricci; Bianca Zecchin; Calogero Terregino; Erika Giorgia Quaranta; Isabella Monne |
| EPI_ISL_429229 | Ospedale Regionale San Salvatore | Istituto Zooprofilattico Sperimentale dell'Abruzzo e Molise "G. Caporale" | Ancora M; Camma C; Curini V; Di Domenico M; Di Pasquale A; Lorusso A; Mangone I; Marcacci M; Puglia I; Rinaldi A; Savini G |
| EPI_ISL_420566 to 420567, EPI_ISL_436718 | Ospedale Regionale San Salvatore | Istituto Zooprofilattico Sperimentale dell'Abruzzo e Molise "G.Caporale" | Ancora M; Cammà C; Curini V; Di Domenico M; Di Pasquale A; Lorusso A; Mangone I; Marcacci M; Puglia I; Rinaldi A; Savini G |
| EPI_ISL_528925 | Ospedale Regionale San Salvatore-L'Aquila | Istitituto Zooprofilattico Sperimentale dell'Abruzzo e Molise "G.Caporale" | Ancora M; Cammà C; Curini V; Di Domenico M; Di Pasquale A; Lorusso A; Mangone I; Marcacci M; Puglia I; Rinaldi A; Savini G. |
| EPI_ISL_806773 to 806774 | Ospedale S.Salvatore-Medicina Interna L'Aquila | Istituto Zooprofilattico Sperimentale dell'Abruzzo e Molise "G. Caporale" | Ancora M; Calistri P; Cammà C; Curini V; Di Domenico M; Di Pasquale A; Lorusso A; Mangone I; Marcacci M; Puglia I; Rinaldi A; Savini G |
| EPI_ISL_435148, EPI_ISL_435150 to 435151 | Ospedale SS Annunziata | Istituto Zooprofilattico Sperimentale dell'Abruzzo e Molise "G.Caporale" | Ancora M; Cammà C; Curini V; Di Domenico M; Di Pasquale A; Lorusso A; Mangone I; Marcacci M; Puglia I; Rinaldi A; Savini G |
| EPI_ISL_833265 to 833269, EPI_ISL_833289, EPI_ISL_833325, EPI_ISL_833327 | Ospedale SS Annunziata Chieti | Istituto Zooprofilattico Sperimentale dell'Abruzzo e Molise "G. Caporale" | Ancora M; Calistri P; Cammà C; Curini V; Di Domenico M; Di Pasquale A; Lorusso A; Mangone I; Marcacci M; Puglia I; Rinaldi A; Savini G |
| EPI_ISL_528991 to 528992, EPI_ISL_529016 | Ospedale SS Annunziata-Sulmona | Istitituto Zooprofilattico Sperimentale dell'Abruzzo e Molise "G.Caporale" | Ancora M; Cammà C; Curini V; Di Domenico M; Di Pasquale A; Lorusso A; Mangone I; Marcacci M; Puglia I; Rinaldi A; Savini G. |
| EPI_ISL_747459 to 747464, EPI_ISL_747482 to 747483 | Ospedale San Bonifacio | Istituto Zooprofilattico Sperimentale delle Venezie | Adelaide Milani; Alessia Schivo; Alice Fusaro; Ambra Pastori; Annalisa Salviato; Antonia Ricci; Bianca Zecchin; Calogero Terregino; Erika Giorgia Quaranta; Isabella Monne |
| EPI_ISL_794750, EPI_ISL_876742, EPI_ISL_918408 to 918409 | Ospedale Santa Caterina Novella | Istituto Zooprofilattico Sperimentale della Puglia e della Basilicata | Bianco A.; Capozzi L.; Cipolletta D.; Del Sambro L.; Galante D.; Manzulli V; Pace L.; Parisi A.; Rondinone V.; Simone D. |
| EPI_ISL_794747, EPI_ISL_794751 | Ospedale Vito Fazzi | Istituto Zooprofilattico Sperimentale della Puglia e della Basilicata | Bianco A.; Capozzi L.; Cipolletta D.; Del Sambro L.; Galante D.; Manzulli V; Pace L.; Parisi A.; Rondinone V. |
| EPI_ISL_418256 | Ospedale "San Liberatore" di Atri | Istituto Zooprofilattico Sperimentale dell'Abruzzo e Molise "G. Caporale" | Ancora M; Cammà C; Curini V; Di Domenico M; Di Pasquale A; Lorusso A; Mangone I; Marcacci M; Puglia I; Rinaldi A; Savini G. |
| EPI_ISL_475124, EPI_ISL_476145 to 476147, EPI_ISL_548248 | Ostersund klinisk mikrobiologi | The Public Health Agency of Sweden | Anna Risberg; Anna-Malin Linde; Karin Tegmark-Wisell; Maria Lind Karlberg; Mattias Haukland; Mia Brytting; Olov Svartstrom; Oskar Karlsson Lindsjo; Petra Edquist; Reza Advani; Sandra Broddesson; Shamam Muradrasoli |
| EPI_ISL_449786, EPI_ISL_471174, EPI_ISL_481211 to 481213, EPI_ISL_549042 to 549047, EPI_ISL_549092 to 549111, EPI_ISL_549113 to 549118, EPI_ISL_549126 to 549131, EPI_ISL_549133 to 549142, EPI_ISL_549144 to 549153, EPI_ISL_549155 to 549163, EPI_ISL_590894 to 590897, EPI_ISL_590921 to 590922, EPI_ISL_590931, EPI_ISL_590948, EPI_ISL_590986 to 591006, EPI_ISL_591009, EPI_ISL_591015 to 591017, EPI_ISL_635107 to 635109, EPI_ISL_635116, EPI_ISL_635149 to 635152, EPI_ISL_635164, EPI_ISL_635174 to 635175, EPI_ISL_635185, EPI_ISL_668433, EPI_ISL_668438 to 668440, EPI_ISL_708032, EPI_ISL_708110 to 708114, EPI_ISL_775315, EPI_ISL_775322, EPI_ISL_775337 to 775343, EPI_ISL_775345, EPI_ISL_796674, EPI_ISL_796695 to 796697, EPI_ISL_860206 to 860207, EPI_ISL_860218, EPI_ISL_860229 to 860230, EPI_ISL_860293 | | | |
| see above | Ostfold Hospital Trust - Kalnes, Centre for Laboratory Medicine, Section for gene technology and infection serology | Norwegian Institute of Public Health, Department of Virology | Atiya R Ali; Hilde Elshaug; Hilde Synnøve Vollan; Hilde Vollan; Kamilla Heddeland Instefjord; Karoline Bragstad; Kathrine Stene-Johansen; Marie Paulsen Madsen; Olav Hungnes; Rasmus Riis Kopperud |
| EPI_ISL_420148 | Ostfold Hospital Trust -Kalnes Centre for Laboratory Medicine Section for gene technology and infection serology | Norwegian Institute of Public Health, Department of Virology | Hilde Elshaug; Kamilla Heddeland Instefjord; Karoline Bragstad; Kathrine Stene-Johansen; Olav Hungnes |
| EPI_ISL_448449 to 448547, EPI_ISL_448549, EPI_ISL_448551, EPI_ISL_448553, EPI_ISL_448555, EPI_ISL_448557, EPI_ISL_448559, EPI_ISL_448561, EPI_ISL_448564 to 448565, EPI_ISL_448774 to 448808, EPI_ISL_448810 to 448822, EPI_ISL_448824 to 448825, EPI_ISL_478729, EPI_ISL_478731 to 478732, EPI_ISL_478734 to 478736, EPI_ISL_478738 to 478744, EPI_ISL_478746 to 478749, EPI_ISL_478751 to 478754, EPI_ISL_478756 to 478759, EPI_ISL_478761, EPI_ISL_478763 to 478764, EPI_ISL_478766 to 478777, EPI_ISL_478779 to 478780, EPI_ISL_478782 to 478787, EPI_ISL_478789 to 478792, EPI_ISL_478794 to 478795, EPI_ISL_478797 to 478799, EPI_ISL_478801 to 478803, EPI_ISL_478806 to 478816, EPI_ISL_478819 to 478820, EPI_ISL_478822 to 478824, EPI_ISL_478826 to 478830, EPI_ISL_478832, EPI_ISL_478834 to 478845, EPI_ISL_478847 to 478848, EPI_ISL_478850 to 478852, EPI_ISL_478854 to 478861, EPI_ISL_478863 to 478882, EPI_ISL_478884, EPI_ISL_478886 to 478889, EPI_ISL_478891, EPI_ISL_478893 to 478895, EPI_ISL_478897 to 478898, EPI_ISL_478900 to 478901, EPI_ISL_478903 to 478909, EPI_ISL_478911 to 478912, EPI_ISL_478914 to 478915, EPI_ISL_478917 to 478920, EPI_ISL_478922 to 478924, EPI_ISL_478926 to 478933, EPI_ISL_478935 to 478940, EPI_ISL_478942 to 478943, EPI_ISL_478945 to 478946, EPI_ISL_478948 to 478954, EPI_ISL_478956 to 478957, EPI_ISL_478959 to 478960, EPI_ISL_478962 to 478964, EPI_ISL_478967 to 478969, EPI_ISL_478971 to 478972, EPI_ISL_478974 to 478977, EPI_ISL_478979 to 478980, EPI_ISL_478982 to 478984, EPI_ISL_478986 to 478987, EPI_ISL_478989, EPI_ISL_478991 to 478992, EPI_ISL_478994 to 478995, EPI_ISL_478997 to 478999, EPI_ISL_479001, EPI_ISL_479003 to 479004, EPI_ISL_479006, EPI_ISL_479008, EPI_ISL_479010, EPI_ISL_479012, EPI_ISL_479014, EPI_ISL_479017, EPI_ISL_479020, EPI_ISL_479022, EPI_ISL_479026 to 479031, EPI_ISL_479033 to 479038, EPI_ISL_479041 to 479054, EPI_ISL_479056 to 479062, EPI_ISL_479064, EPI_ISL_479066 to 479069, EPI_ISL_479071 to 479076, EPI_ISL_479078 to 479080, EPI_ISL_479082 to 479087, EPI_ISL_479089 to 479091, EPI_ISL_479093, EPI_ISL_479095 to 479097, EPI_ISL_479099 to 479104, EPI_ISL_479108 to 479114, EPI_ISL_479116 to 479154, EPI_ISL_479156 to 479167, EPI_ISL_479169 to 479174, EPI_ISL_534763 to 534922, EPI_ISL_534924 to 534935, EPI_ISL_534937 to 534950, EPI_ISL_534952 to 534956, EPI_ISL_534958 to 535023, EPI_ISL_549329 to 549333, EPI_ISL_549335, EPI_ISL_549337 to 549340, EPI_ISL_549342, EPI_ISL_549345 to 549348, EPI_ISL_549396 to 549459, EPI_ISL_549523 to 549546, EPI_ISL_559753 to 559897, EPI_ISL_559984 to 560053, EPI_ISL_560069 to 560082, EPI_ISL_560305 to 560306, EPI_ISL_572714, EPI_ISL_572723 to 572724, EPI_ISL_572726 to 572728, EPI_ISL_572846, EPI_ISL_572849 to 572850, EPI_ISL_572857 to 572861, EPI_ISL_572869 to 572870, EPI_ISL_572873, EPI_ISL_572891 to 572897, EPI_ISL_572902 to 572903, EPI_ISL_572917 to 572926, EPI_ISL_572933 to 572937, EPI_ISL_572947, EPI_ISL_572952 to 572953, | | | |

| | | | |
|---|---|---|---|
| EPI_ISL_572958, EPI_ISL_572960, EPI_ISL_572962 to 573084, EPI_ISL_573154 to 573235, EPI_ISL_573566 to 573651, EPI_ISL_573759, EPI_ISL_573811 to 573855, EPI_ISL_573866 to 573869, EPI_ISL_574257, EPI_ISL_576569 to 576748, EPI_ISL_576750 to 576849, EPI_ISL_576851 to 576876, EPI_ISL_576920 to 576960, EPI_ISL_577068 to 577095, EPI_ISL_577383 to 577384, EPI_ISL_577401 to 577416, EPI_ISL_584680, EPI_ISL_594467 to 594483, EPI_ISL_594617 to 594644, EPI_ISL_594889 to 594922, EPI_ISL_595326 to 595328, EPI_ISL_595617 to 595798, EPI_ISL_627079, EPI_ISL_627085, EPI_ISL_627250 to 627251, EPI_ISL_627255 to 627263, EPI_ISL_627281 to 627290, EPI_ISL_627292, EPI_ISL_627294, EPI_ISL_627297, EPI_ISL_627299, EPI_ISL_627302 to 627303, EPI_ISL_627305, EPI_ISL_628334 to 628368, EPI_ISL_628414 to 628420, EPI_ISL_628422 to 628700, EPI_ISL_637855, EPI_ISL_637858, EPI_ISL_637861, EPI_ISL_637867, EPI_ISL_637871, EPI_ISL_638037, EPI_ISL_638073 to 638078, EPI_ISL_638086 to 638089, EPI_ISL_638144 to 638145, EPI_ISL_638164, EPI_ISL_638170 to 638171, EPI_ISL_638173, EPI_ISL_638175 to 638176, EPI_ISL_638207, EPI_ISL_638459 to 638531, EPI_ISL_638736 to 638876, EPI_ISL_638894, EPI_ISL_638944 to 639001, EPI_ISL_639627 to 639630, EPI_ISL_651086 to 651088, EPI_ISL_651103 to 651106, EPI_ISL_651417, EPI_ISL_651421, EPI_ISL_651431 to 651433, EPI_ISL_651470 to 651483, EPI_ISL_651485, EPI_ISL_651505 to 651511, EPI_ISL_651513, EPI_ISL_651517, EPI_ISL_651525, EPI_ISL_651529, EPI_ISL_651541, EPI_ISL_651544 to 651545, EPI_ISL_651548 to 651549, EPI_ISL_651587, EPI_ISL_652117 to 652150, EPI_ISL_652711 to 653026, EPI_ISL_679629 to 679851, EPI_ISL_680579, EPI_ISL_704594, EPI_ISL_704676, EPI_ISL_704679, EPI_ISL_704682, EPI_ISL_704685, EPI_ISL_704687, EPI_ISL_704690, EPI_ISL_704692, EPI_ISL_704695, EPI_ISL_704698, EPI_ISL_704701, EPI_ISL_704703, EPI_ISL_704706, EPI_ISL_704812, EPI_ISL_704815, EPI_ISL_704817, EPI_ISL_704836, EPI_ISL_704839, EPI_ISL_704845, EPI_ISL_704861, EPI_ISL_704864, EPI_ISL_704866, EPI_ISL_704869, EPI_ISL_704872, EPI_ISL_704874, EPI_ISL_704877, EPI_ISL_704879, EPI_ISL_704882, EPI_ISL_704885, EPI_ISL_705414, EPI_ISL_705417, EPI_ISL_705426 to 705430, EPI_ISL_705443 to 705444, EPI_ISL_705450 to 705456, EPI_ISL_705464 to 705470, EPI_ISL_705473 to 705485, EPI_ISL_705489, EPI_ISL_705493, EPI_ISL_705497 to 705500, EPI_ISL_705526, EPI_ISL_705972 to 706193, EPI_ISL_706195 to 706275, EPI_ISL_706891 to 706974, EPI_ISL_723631 to 723672, EPI_ISL_723674 to 723850, EPI_ISL_723949 to 724062, EPI_ISL_724778 to 724835, EPI_ISL_727807 to 727860, EPI_ISL_740897 to 740952, EPI_ISL_741018 to 741185, EPI_ISL_741349 to 741356, EPI_ISL_741362 to 741420, EPI_ISL_741760 to 741771, EPI_ISL_741773 to 741935, EPI_ISL_742247 to 742259, EPI_ISL_742261, EPI_ISL_763838, EPI_ISL_763841, EPI_ISL_763844 to 763846, EPI_ISL_763859 to 763863, EPI_ISL_763928 to 763974, EPI_ISL_764066 to 764068, EPI_ISL_764077 to 764079, EPI_ISL_764085 to 764093, EPI_ISL_764117, EPI_ISL_764243, EPI_ISL_764250, EPI_ISL_764264 to 764265, EPI_ISL_764271, EPI_ISL_764525 to 764572, EPI_ISL_812970, EPI_ISL_813356, EPI_ISL_814466 to 814467, EPI_ISL_814572, EPI_ISL_814594, EPI_ISL_814604 to 814605, EPI_ISL_814688 to 814694, EPI_ISL_819578, EPI_ISL_819585 to 819593, EPI_ISL_819605 to 819608, EPI_ISL_819612 to 819646, EPI_ISL_819653 to 819670, EPI_ISL_819676 to 819685, EPI_ISL_819689 to 819723, EPI_ISL_819732 to 819756, EPI_ISL_819759 to 819762, EPI_ISL_819764 to 819794, EPI_ISL_819796 to 819798, EPI_ISL_819803 to 819820, EPI_ISL_819822 to 819827, EPI_ISL_819835 to 819850, EPI_ISL_819852 to 819863, EPI_ISL_819866, EPI_ISL_819868 to 819872, EPI_ISL_839360, EPI_ISL_839362 to 839365, EPI_ISL_839367 to 839384, EPI_ISL_839386 to 839411, EPI_ISL_839413 to 839436, EPI_ISL_839438 to 839443, EPI_ISL_839445 to 839483, EPI_ISL_840186 to 840199, EPI_ISL_840201 to 840202, EPI_ISL_840204 to 840207, EPI_ISL_840209 to 840217, EPI_ISL_840220 to 840224, EPI_ISL_840226, EPI_ISL_840228 to 840246, EPI_ISL_840249 to 840250, EPI_ISL_840252 to 840254, EPI_ISL_840256 to 840257, EPI_ISL_840259 to 840272, EPI_ISL_840274, EPI_ISL_840276, EPI_ISL_840278 to 840284, EPI_ISL_840286, EPI_ISL_840288 to 840292, EPI_ISL_840294 to 840336, EPI_ISL_840338 to 840345, EPI_ISL_840348 to 840351, EPI_ISL_840353, EPI_ISL_840357 to 840363, EPI_ISL_840365 to 840366, EPI_ISL_842107, EPI_ISL_842111 to 842113, EPI_ISL_842116 to 842120, EPI_ISL_842122 to 842136, EPI_ISL_842138, EPI_ISL_842140, EPI_ISL_842143 to 842145, EPI_ISL_842147 to 842152, EPI_ISL_842154 to 842160, EPI_ISL_842162 to 842163, EPI_ISL_842165 to 842171, EPI_ISL_842175, EPI_ISL_842178 to 842181, EPI_ISL_842184, EPI_ISL_842186 to 842188, EPI_ISL_842190 to 842192, EPI_ISL_842194 to 842200, EPI_ISL_867035, EPI_ISL_867037 to 867041, EPI_ISL_867044 to 867069, EPI_ISL_867071 to 867075, EPI_ISL_867077 to 867082, EPI_ISL_867084, EPI_ISL_867087 to 867090, EPI_ISL_867093 to 867098, EPI_ISL_867130, EPI_ISL_867155, EPI_ISL_867159, EPI_ISL_867161 to 867162, EPI_ISL_867168 to 867175, EPI_ISL_867177 to 867179, EPI_ISL_867181 to 867185 | | | |
| see above | Oxford Viromics, NDM, University of Oxford; Oxford University Hospitals; Basingstoke and North Hampshire Hospital | COVID-19 Genomics UK (COG-UK) Consortium | Alex Mobbs; Amy Trebes; Anita Justice; Catrin Moore; Christophe Fraser; David Bonsall; David Buck; Emma Wise; George Macintyre; Jessica Lynch; John Todd; Mariateresa de Cesare; Matilde Mori; Monique Andersson; Nathan Moore; Nick Cortes; Robert Shaw; Stephen Kidd; Tanya Golubchik; Timothy Peto |
| EPI_ISL_458491, EPI_ISL_458786, EPI_ISL_458800, EPI_ISL_458872, EPI_ISL_459039, EPI_ISL_482028, EPI_ISL_487593, EPI_ISL_488409, EPI_ISL_488411, EPI_ISL_492514, EPI_ISL_492594 | | | |
| see above | PHE South West Regional Laboratory, National Infection Service | Wellcome Sanger Institute for the COVID-19 Genomics UK (COG-UK) Consortium | Barry Vipond; Cordelia Langford; David K. Jackson; Dominic Kwiatkowski; Dr Peter Muir; Ewan Harrison; Hannah Pymont; Ian Johnston; John Sillitoe on behalf of the Wellcome Sanger Institute COVID-19 Surveillance Team; Rich Hopes; Roberto Amato; Sonia Goncalves; Stephanie Hutchings; and Alex Alderton |
| EPI_ISL_439953, EPI_ISL_439957, EPI_ISL_439959 to 439960, EPI_ISL_439962, EPI_ISL_439964 to 439967, EPI_ISL_439969, EPI_ISL_439971 to 439974, EPI_ISL_439976, EPI_ISL_439978 to 439979, EPI_ISL_439981 to 439986, EPI_ISL_439988, EPI_ISL_439992, EPI_ISL_439998, EPI_ISL_440000 to 440002, EPI_ISL_440005 to 440006, EPI_ISL_440008, EPI_ISL_440013, EPI_ISL_440016, EPI_ISL_440020 to 440022, EPI_ISL_440026 to 440029, EPI_ISL_440031 to 440033, EPI_ISL_440035 to 440039, EPI_ISL_440042 to 440043, EPI_ISL_440050 to 440052, EPI_ISL_440054, EPI_ISL_440056, EPI_ISL_440058, EPI_ISL_440063, EPI_ISL_440065, EPI_ISL_440068, EPI_ISL_440070 to 440071, EPI_ISL_440081, EPI_ISL_440084, EPI_ISL_440087, EPI_ISL_440091 to 440095, EPI_ISL_440098, EPI_ISL_440100 to 440103, EPI_ISL_440105 to 440106, EPI_ISL_440113 to 440116, EPI_ISL_440118 to 440119, EPI_ISL_440123 to 440124, EPI_ISL_440126 to 440127, EPI_ISL_440130 to 440131, EPI_ISL_440134 to 440137, EPI_ISL_440139 to 440142, EPI_ISL_440145, EPI_ISL_440148 to 440149, EPI_ISL_440151, EPI_ISL_440153 to 440154, EPI_ISL_440156 to 440158, EPI_ISL_440160, EPI_ISL_440162, EPI_ISL_440165, EPI_ISL_440167 to 440168, EPI_ISL_440172, EPI_ISL_440174 to 440175, EPI_ISL_440178, EPI_ISL_440187, EPI_ISL_440189 to 440192, EPI_ISL_440194, EPI_ISL_440196, EPI_ISL_440198 to 440199, EPI_ISL_440201, EPI_ISL_440204 to 440206, EPI_ISL_440210, EPI_ISL_440212, EPI_ISL_440214 to 440216, EPI_ISL_440218 to 440222, EPI_ISL_440224 to 440225, EPI_ISL_440227 to 440229, EPI_ISL_440232 to 440233, EPI_ISL_440235 to 440236, EPI_ISL_440238, EPI_ISL_440241 to 440243, EPI_ISL_440245, EPI_ISL_440248 to 440253, EPI_ISL_440255 to 440256, EPI_ISL_440262, EPI_ISL_440266, EPI_ISL_440269, EPI_ISL_440272 to 440277, EPI_ISL_440286, EPI_ISL_440291, EPI_ISL_440294, EPI_ISL_440298 to 440299, EPI_ISL_440623 to 440730, EPI_ISL_440732 to 440749, EPI_ISL_440751 to 440804, EPI_ISL_440806 to 440809, EPI_ISL_442624 to 442651, EPI_ISL_442653 to 442672, EPI_ISL_442674 to 442722, EPI_ISL_442724 to 442810, EPI_ISL_443686 to 443742, EPI_ISL_443744 to 443772, EPI_ISL_443774 to 443823, EPI_ISL_443825 to 443837, EPI_ISL_443839 to 443843, EPI_ISL_443845 to 443849, EPI_ISL_443851 to 443902, EPI_ISL_443904 to 443993, EPI_ISL_443995, EPI_ISL_443997 to 444021, EPI_ISL_458299 to 458322, EPI_ISL_458324 to 458339, EPI_ISL_458341 to 458345, EPI_ISL_458347 to 458363, EPI_ISL_458365 to 458370, EPI_ISL_458372 to 458392, EPI_ISL_458394 to 458423, EPI_ISL_458425 to 458431, EPI_ISL_458433 to 458435, EPI_ISL_458437 to 458454, EPI_ISL_458456 to 458469, EPI_ISL_458471 to 458490, EPI_ISL_458492 to 458495, EPI_ISL_458497 to 458510, EPI_ISL_458513 to 458515, EPI_ISL_458719, EPI_ISL_458721 to 458729, EPI_ISL_458731 to 458740, EPI_ISL_458742 to 458743, EPI_ISL_458745 to 458785, EPI_ISL_458787 to 458799, EPI_ISL_458801 to 458804, EPI_ISL_458806 to 458871, EPI_ISL_458873 to 458887, EPI_ISL_458889 to 458892, EPI_ISL_458894 to 458896, EPI_ISL_458898 to 458904, EPI_ISL_458906 to 458917, EPI_ISL_458919 to 458928, EPI_ISL_458930 to 458998, EPI_ISL_459001 to 459038, EPI_ISL_459040 to 459046, EPI_ISL_459048 to 459053, EPI_ISL_459055 to 459066, EPI_ISL_459068 to 459094, EPI_ISL_459725 to 459727, EPI_ISL_467693 to 467695, EPI_ISL_467697 to 467700, EPI_ISL_467702 to 467719, EPI_ISL_469345 to 469364, EPI_ISL_469366 to 469377, EPI_ISL_469380 to 469387, EPI_ISL_469389 to 469440, EPI_ISL_469442 to 469450, EPI_ISL_469452 to 469466, EPI_ISL_469468 to 469469, EPI_ISL_469471 to 469500, EPI_ISL_469502 to 469527, EPI_ISL_469529, EPI_ISL_469531 to 469533, EPI_ISL_469535 to 469544, EPI_ISL_469546 to 469562, EPI_ISL_469564 to 469566, EPI_ISL_469568 to 469584, EPI_ISL_469586 to 469627, EPI_ISL_469629 to 469630, EPI_ISL_469632, EPI_ISL_469634 to 469639, EPI_ISL_469641 to 469644, EPI_ISL_469646 to 469676, EPI_ISL_469679 to 469687, EPI_ISL_469689 to 469728, EPI_ISL_469730 to 469739, EPI_ISL_469741 to 469763, EPI_ISL_469765 to 469775, EPI_ISL_469777 to 469779, EPI_ISL_469781 to 469783, EPI_ISL_469785 to 469804, EPI_ISL_469806 to 469843, EPI_ISL_469914, EPI_ISL_469916 to 469917, EPI_ISL_470356 to 470357, EPI_ISL_470529, EPI_ISL_481763 to 481764, EPI_ISL_481766 to 481807, EPI_ISL_481809 to 481821, EPI_ISL_481823 to 481844, EPI_ISL_481846 to 481879, EPI_ISL_481881 to 481896, EPI_ISL_481898 to 481922, EPI_ISL_481924 to 481992, EPI_ISL_481994 to 481996, EPI_ISL_481998 to 482027, EPI_ISL_482029 to 482031, EPI_ISL_487524, EPI_ISL_487526 to 487528, EPI_ISL_487530 to 487533, EPI_ISL_487535 to 487536, EPI_ISL_487538 to 487539, EPI_ISL_487541 to 487545, EPI_ISL_487547 to 487549, EPI_ISL_487551 to 487555, EPI_ISL_487560 to 487561, EPI_ISL_487563, EPI_ISL_487565 to 487567, EPI_ISL_487570 to 487571, EPI_ISL_487573 to 487577, EPI_ISL_487579 to 487580, EPI_ISL_487583, EPI_ISL_487586 to 487588, EPI_ISL_487590 to 487592, EPI_ISL_487594, EPI_ISL_487596, EPI_ISL_487598 to 487599, EPI_ISL_487601 to 487605, EPI_ISL_487608, EPI_ISL_487610 to 487613, EPI_ISL_487617 to 487620, EPI_ISL_487622 to 487624, EPI_ISL_487629, EPI_ISL_487634, EPI_ISL_488187 to 488195, EPI_ISL_488197 to 488203, EPI_ISL_488205 to 488208, EPI_ISL_488210 to 488216, EPI_ISL_488218 to 488228, EPI_ISL_488230 to 488242, EPI_ISL_488244 to 488287, EPI_ISL_488289 to 488341, EPI_ISL_488343 to 488357, EPI_ISL_488359 to 488386, EPI_ISL_488388 to 488397, EPI_ISL_488399 to 488408, EPI_ISL_488410, EPI_ISL_488412 to 488455, EPI_ISL_488877 to 488878, EPI_ISL_489120, EPI_ISL_492185 to 492188, EPI_ISL_492195, EPI_ISL_492197, EPI_ISL_492203, EPI_ISL_492211 to 492260, EPI_ISL_492262 to 492302, EPI_ISL_492304 to 492315, EPI_ISL_492317 to 492323, EPI_ISL_492325 to 492332, EPI_ISL_492334 to 492339, EPI_ISL_492341, EPI_ISL_492343 to 492356, EPI_ISL_492358 to 492449, EPI_ISL_492505 to 492513, EPI_ISL_492515 to 492516, EPI_ISL_492518 to 492523, EPI_ISL_492526 to 492587, EPI_ISL_492589 to 492593, EPI_ISL_492595, EPI_ISL_492597 to 492629, EPI_ISL_492631 to 492636, EPI_ISL_492638 to 492703, EPI_ISL_492705 to 492714, EPI_ISL_492716 to 492717, EPI_ISL_492720 to 492724, EPI_ISL_492726 to 492745, EPI_ISL_492764 to 492785, EPI_ISL_492787 to 492804, EPI_ISL_492806 to 492819, EPI_ISL_492821 to 492824, EPI_ISL_492826 to 492835, EPI_ISL_495097 to 495114, EPI_ISL_495116 to 495121, EPI_ISL_501555 to 501556, EPI_ISL_501558 to 501561, EPI_ISL_501563 to 501568, EPI_ISL_501570, EPI_ISL_501572 to 501584, EPI_ISL_501586 to 501592, EPI_ISL_501594 to 501599, EPI_ISL_501601, EPI_ISL_501603 to 501612, EPI_ISL_511955, EPI_ISL_511957, EPI_ISL_511960 to 511962, EPI_ISL_511964 to 511969, EPI_ISL_511971 to 511981, EPI_ISL_513584, EPI_ISL_524486 to 524492, EPI_ISL_539896 | | | |
| see above | PHE South West Regional Laboratory, National Infection Service | Wellcome Sanger Institute for the COVID-19 Genomics UK (COG-UK) consortium | Alex Alderton; Barry Vipond; Cordelia Langford; David K. Jackson; Dominic Kwiatkowski; Dr Peter Muir; Ewan Harrison; Hannah Pymont; Ian Johnston; John Sillitoe on behalf of the Wellcome Sanger Institute COVID-19 Surveillance Team; John Sillitoe on behalf of the Wellcome Sanger Institute COVID-19 Surveillance Team (http://www.sanger.ac.uk/covid-team); Rich Hopes; Roberto Amato; Sonia Goncalves; Stephanie Hutchings; and Alex Alderton |
| EPI_ISL_678257 | PHI Institute of Nephrology - Struga | Research Center for Genetic Engineering and Biotechnology "Georgi D. Efremov" , Macedonian Academy of Sciences and Arts | RCGEB - MASA |
| EPI_ISL_833288, EPI_ISL_833290 | PRESIDIO TERRITORIALE CASOLI CH | Istituto Zooprofilattico Sperimentale dell'Abruzzo e Molise "G. Caporale" | Ancora M; Calistri P; Cammà C; Curini V; Di Domenico M; Di Pasquale A; Lorusso A; Mangone I; Marcacci M; Puglia I; Rinaldi A; Savini G |
| EPI_ISL_707936 to 707939, EPI_ISL_708185 to 708186, EPI_ISL_708188 to 708190, EPI_ISL_708192 to 708195 | Pamukkale University Hospital | Pamukkale University Department of Medical Genetics | Onur TOKGUN et al. |
| EPI_ISL_421502 to 421503 | Parc des Dames | National Reference Center for Viruses of Respiratory Infections, Institut Pasteur, Paris | Angela Brisebarre; Etienne Simon-Lorière; Flora Donati; Marion Barbet; Maud Vanpeene; Mélanie Albert; Méline Bizard; Sylvie Behillil; Sylvie van der Werf; Vincent Enouf |
| EPI_ISL_591348 to 591351, EPI_ISL_591354 to 591361, EPI_ISL_591364 to 591365, EPI_ISL_591376 to 591377, EPI_ISL_591460, EPI_ISL_591535, EPI_ISL_667567, EPI_ISL_667569, EPI_ISL_667581, EPI_ISL_667594 to 667597, EPI_ISL_667601, EPI_ISL_667615, EPI_ISL_667617, EPI_ISL_667620, EPI_ISL_667635, EPI_ISL_667638, EPI_ISL_667658, EPI_ISL_667667, EPI_ISL_667669, EPI_ISL_667671, EPI_ISL_667684, EPI_ISL_667690, EPI_ISL_667692, EPI_ISL_667716, EPI_ISL_667721, EPI_ISL_667728 to 667729, EPI_ISL_667732, EPI_ISL_667734, EPI_ISL_667736, EPI_ISL_667739, EPI_ISL_721569, EPI_ISL_721573 to 721574, EPI_ISL_721577, EPI_ISL_721602, EPI_ISL_721604, EPI_ISL_721610 to 721611, EPI_ISL_721614, EPI_ISL_721618, EPI_ISL_721623, EPI_ISL_735439 to 735443, EPI_ISL_736891 to 736892, EPI_ISL_736907 to 736909, EPI_ISL_736917, EPI_ISL_736922, EPI_ISL_768634 to 768641, EPI_ISL_768643 to 768658, EPI_ISL_768660 to 768667, EPI_ISL_768694 to 768695, EPI_ISL_768700 to 768701, EPI_ISL_768703, EPI_ISL_768706 to 768708, EPI_ISL_768722, EPI_ISL_779209, EPI_ISL_779220 to 779232, EPI_ISL_779253, EPI_ISL_779670, EPI_ISL_779672, EPI_ISL_779698, EPI_ISL_779701, EPI_ISL_803944 | | | |
| see above | Pathogen Genomics Center, National Institute of Infectious Diseases | Pathogen Genomics Center, National Institute of Infectious Diseases | Kentaro Itokawa; Makoto Kuroda; Masanori Hashino; Rina Tanaka; Tsuyoshi Sekizuka |
| EPI_ISL_470897 to 470899, EPI_ISL_872628 to 872640, EPI_ISL_872643 | Pathogenic Microorganisms Variability Laboratory | Pathogenic Microorganisms Variability Laboratory | Alexander Gintsburg; Alexey Shchetinin; Andrei Siniavin; Andrey Botikov; Andrey Pochtovyy; Anna Ignatieva; Denis Logunov; Elena Burtseva; Elena Shidlovskaya; Elizaveta Divisenko; Evgeniya Mukasheva; Evgeny Usachev; Kirill Krasnoslobotsev; Ludmila Kolobukhina; Maria Nikiforova; Nadezhda Kuznetsova; Olesya Venchakova; Olga Burgasova; Svetlana Smetanina; Svetlana Trushakova; Valeria Bacalin; Vladimir Gushchin |
| EPI_ISL_566077 to 566083, EPI_ISL_596318 to 596352, EPI_ISL_733156 to 733182, EPI_ISL_733185 to 733222, EPI_ISL_770670 | Pathogenic Microorganisms Variability Laboratory | WHO National Influenza Centre Russian Federation | Alexander Gintsburg; Alexey Shchetinin; Andrey Komissarov; Andrey Pochtovyy; Anna Ignatieva; Anna Ivanova; Artem Fadeev; Artem Tkachuk; Daria Danilenko; Dmitry Bazhenov; Dmitry Lioznov; Ekaterina Milashenko; Elena Burtseva; Elena Nabieva; Elena Shidlovskaya; Elena Vokalova; Elizaveta Divisenko; Evgeniya Mukasheva; Evgeny Usachev; Evgeny Usachev; Georgii Bazykin; Kirill Krasnoslobotsev; Ksenia Safina; Kseniya Komissarova; Ludmila Kolobukhina; Maria Nikiforova; Maxim Rubalsky; Nadezhda Kuznetsova; Oleg Rubalsky; Olga Burgasova; Sergey Alkhovsky; Svetlana Smetanina; Svetlana Trushakova; Tatyana Vishnevskaya; Valeria Bacalin; Vladimir Gushchin |
| EPI_ISL_583619 to 583629 | Pathologie-Labor Dr. Obrist- Dr. Brunhuber | Bergthaler laboratory, CeMM Research Center for Molecular Medicine of the Austrian Academy of Sciences | Adi Steinrigl; Alexander Lercher; Alexandra Popa; Andreas Bergthaler; Benedikt Agerer; Christian Paar; Christoph Bock; Daniela Schmid; Dorothee von Laer; Elisabeth Puchhammer-Stoeckl; Franz Allerberger; Gernot Walder; Gregor Hörmann; Guenter Weiss; Gunther Vogl; Henrique Colaco; Jakob-Wendelin Genger; Jan Laine; Judith Aberle; Kinga Rigler-Hohenwarter; Lukas Endler; Manfred Nairz; Mark Smyth; Martin Senekowitsch; Michael Schuster; Peter Hufnagl; Peter Obrist; Rainer Gattringer; Sabine Sussitz-Rack; Stephan Aberle; Thomas Penz; Wegene Borena |
| EPI_ISL_853754, EPI_ISL_853769 to 853774, EPI_ISL_853798 to 853799, EPI_ISL_853805, EPI_ISL_853874 to 853876, EPI_ISL_853917 to 853923, EPI_ISL_854217, EPI_ISL_854255 to 854257, EPI_ISL_854259 to 854260, EPI_ISL_854300, EPI_ISL_854302 to 854303 | | | |

| | | | |
|---|---|---|---|
| see above | Pharmgenetix GmbH | Bergthaler laboratory, CeMM Research Center for Molecular Medicine of the Austrian Academy of Sciences | Alexander Lercher; Alexandra Popa; Andreas Bergthaler; Anna Schedl; Benedikt Agerer; Christoph Bock; Jakob-Wendelin Genger; Jan Laine; Lukas Endler; Martin Senekowitsch; Michael Schuster; Thomas Penz |
| EPI_ISL_514750 | Pirogov Russian National Research Medical University | Pirogov Russian National Research Medical University, Research and Development | Blagodatskikh; K.A. |
| EPI_ISL_443308 | Plaisance | National Reference Center for Viruses of Respiratory Infections, Institut Pasteur, Paris | Angela Brisebarre; Etienne Simon-Lorière; Flora Donati; Marion Barbet; Maud Vanpeene; Mélanie Albert; Méline Bizard; Sylvie Behillil; Sylvie van der Werf; Vincent Enouf |
| EPI_ISL_792016 to 792023 | Plateforme COVID IDF | National Reference Center for Viruses of Respiratory Infections, Institut Pasteur, Paris | Angela Brisebarre; Camille Capel; Etienne Simon-Lorière; Jacques Fourgeaud; Marion Barbet; Maud Vanpeene; Méline Bizard; Sylvie Behillil; Sylvie van der Werf; Vincent Enouf |
| EPI_ISL_856973, EPI_ISL_857040, EPI_ISL_857052, EPI_ISL_857055 | Platform BIS UZA/UAntwerpen, University Hospital Antwerp, Edegem, Belgium | UAntwerp, Laboratory of Medical Microbiology, Campus Drie Eiken S6.26, Universiteitsplein 1, 2610, Wilrijk, Antwerp, Belgium | Basil Britto Xavier; Christine Lammens; Herman Goossens; Jasmine Coppens; Veerle Matheeussen |
| EPI_ISL_833149 | Platform BIS UZA/UAntwerpen, University Hospital Antwerp, Edegem, Belgium | UAntwerp, Laboratory of Medical Microbiology, Campus Drie Eiken S6.26, Universiteitsplein 1, 2610, Wilrijk, Belgium | Basil Britto Xavier; Christine Lammens; Herman Goossens; Jasmine Coppens; Sahaya Glingston Rajakani; Veerle Matheeussen |
| EPI_ISL_522547 to 522548 | Platforme CYROI | UMR PIMIT Université de La Réunion | Camille Lebarbenchon; David Wilkinson; Patrick Mavingui |
| EPI_ISL_418255 | Presidio Ospedaliero "S. Spirito" - PESCARA | Istituto Zooprofilattico Sperimentale dell'Abruzzo e Molise "G. Caporale" | Cammà C; Di Pasquale A; Lorusso A; Mangone I; Marcacci M; Monaco F; Puglia I; Rinaldi A; Savini G |
| EPI_ISL_529013 | Presidio Ospedaliero "S.Filippo e Nicola"-Avezzano | Istitituto Zooprofilattico Sperimentale dell'Abruzzo e Molise "G.Caporale" | Ancora M; Cammà C; Curini V; Di Domenico M; Di Pasquale A; Lorusso A; Mangone I; Marcacci M; Puglia I; Rinaldi A; Savini G. |
| EPI_ISL_528920 to 528921 | Presidio Ospedaliero "Santo Spirito"-Pescara | Istitituto Zooprofilattico Sperimentale dell'Abruzzo e Molise "G.Caporale" | Ancora M; Cammà C; Curini V; Di Domenico M; Di Pasquale A; Lorusso A; Mangone I; Marcacci M; Puglia I; Rinaldi A; Savini G. |
| EPI_ISL_806730 | Presidio Ospedaliero S.Liberatore Atri | Istituto Zooprofilattico Sperimentale dell'Abruzzo e Molise "G. Caporale" | Ancora M; Calistri P; Cammà C; Curini V; Di Domenico M; Di Pasquale A; Lorusso A; Mangone I; Marcacci M; Puglia I; Rinaldi A; Savini G |
| EPI_ISL_429226 to 429227 | Presidio Ospedaliero Santo Spirito | Istituto Zooprofilattico Sperimentale dell'Abruzzo e Molise "G. Caporale" | Ancora M; Camma C; Curini V; Di Domenico M; Di Pasquale A; Lorusso A; Mangone I; Marcacci M; Puglia I; Rinaldi A; Savini G |
| EPI_ISL_794756 to 794758, EPI_ISL_876743 | Presidio di Brindisi Di Summa - Perrino | Istituto Zooprofilattico Sperimentale della Puglia e della Basilicata | Bianco A.; Capozzi L.; Cipolletta D.; Del Sambro L.; Galante D.; Manzulli V; Pace L.; Parisi A.; Rondinone V. |
| EPI_ISL_418258 to 418259 | Presidio ospedaliero "Santo Spirito" | Istituto Zooprofilattico Sperimentale dell'Abruzzo e Molise "G. Caporale" | Ancora M; Cammà C; Curini V; Di Domenico M; Di Pasquale A; Lorusso A; Mangone I; Marcacci M; Puglia I; Rinaldi A; Savini G. |
| EPI_ISL_770533 to 770549 | Private lab | Lithuanian University of Health Sciences, Molecular cardiology lab. | Arnoldas Pautienius; Astra Vitkauskiene; Dovydas Gecys; Ingrida Olendrait; Kamile Tamusauskaite; Laura Pareckaite; Lukas Zemaitis; Vaiva Lesauskaite |
| EPI_ISL_477204, EPI_ISL_514432 | Prof. Massimo Zollo CEINGE TASK-FORCE COVID19 - Regione Campania | Prof. Massimo Zollo CEINGE TASK-FORCE COVID19 - Regione Campania | ; 2; 2* 1 CEINGE Biotecnologie Avanzate; 8* and Massimo Zollo1; Angelo Boccia; Angelo Boccia2; Azienda Sanitaria Ospedali dei Colli; Claudia Tiberio; Claudia Tiberio4; Dae young Kong; Dae young Kong8; Fatemeh asadzadeh; Fatemeh asadzadeh1; Giorgia Borriello; Giorgia Borriello3; Giovanna Fusco; Giovanna Fusco3; Giovanni Paolella; Giovanni Paolella1; Giuseppe Castaldo; Giuseppe Castaldo1; Hong Yeoul Kim 7; Hong Yeoul Kim and Massimo Zollo; Indust; Italia 2 Dipartimento di Medicina Molecolare e Biotecnologie Mediche DMMBM University of Naples Federico II; Italia 3 Istituto Zooprofilattico Sperimentale del Mezzogiorno; Italia 4 -U.O.C. di Patologia Clinica Ospedale D. Cotugno; Italia 6 Department of Microbiology; Italy. 5 Università La Sapienza di Roma; Jae Ho Jung; Jae Ho Jung6; Jae Myun Lee; Jae Myun Lee5; Korea 8 Haim bio co.; Korea 7 Department of Surgery; Kyong Seop Yun; Kyong Seop Yun7; Laura Marrone; Laura Marrone1; Lorenzo Chiariotti; Lorenzo Chiariotti1; Ltd; Luigi Atripaldi; Luigi Atripaldi4; Marika Comegna; Marika Comegna1; Martina Bianchi; Martina Bianchi4; Maurizio Viscardi; Maurizio Viscardi3; Naples; Rino Cerino; Rino Cerino3; Roberto Siciliano; Roberto Siciliano1; Seoul; Sergio Brandi; Sergio Brandi3; Stefano Pascarella; Stefano Pascarella4; Veronica Ferrucci; Veronica Ferrucci1; Yonsei University College of Medicine |
| EPI_ISL_582134 | Protzer Lab | Protzer Lab | Alexander Karollus; Andrea Theumer; Dieter Hoffmann; Eva Schulte; Julien Gagneur; Max von Kleist; Oliver Drechsel; Ulrike Protzer |
| EPI_ISL_572329 | Public Health Authority of the Slovak Republic, Bratislava | Faculty of Natural Sciences, Comenius University in Bratislava | Boris Klempa; Broa Brejová; Dominika Friová; Edita Staroová; Elena Tichá; Jozef Nosek; Juraj Kopáek; Kristína Boršová; Martina Liková; Martina Neboháová; Monika Sláviková; Sabina Fumaová Havlíková; Tomáš Vina; Viktória Hodorová; Viktória abanová; ubomíra Lukáiková |
| EPI_ISL_516988 to 516989 | Public Health Authority of the Slovak Republic, Department of Medical Microbiology | Charite Universitatsmedizin Berlin, Institute of Virology | Barbara Muehlemann; Christian Drosten; Julia Schneider; Jörn Beheim-Schwarzbach; Mgr. Edita Staronova; Talitha Veith; Terry Jones; Victor M Corman |
| EPI_ISL_513310 | Public Health, United States Air Force School of Aerospace Medicine | Public Health, United States Air Force School of Aerospace Medicine | A.C.; A.K.; A.W.; B.C.; C.R.; Chapleau; Connors; E.A.; Fries; J.R.; Javorina; Lambert; Macias; Meyer; Purves; R.R. and Starr; S.M. |
| EPI_ISL_513311 | Public Health, United States Air Force School of Aerospace Medicine | Public Health, United States Air Force School of Aerospace Medicine | A.C.; A.K.; A.W.; B.C.; C.R.; Chapleau; Connors; E.A.; Fries; J.R.; Javorina; Lambert; Macias; Meyer; Purves; R.R. and Starr; S.M. |

EPI_ISL_448223 to 448227, EPI_ISL_448229 to 448235, EPI_ISL_448237 to 448239, EPI_ISL_448242 to 448246, EPI_ISL_448248 to 448249, EPI_ISL_448251, EPI_ISL_448254 to 448256, EPI_ISL_448258 to 448259, EPI_ISL_448262, EPI_ISL_448265 to 448270, EPI_ISL_448272 to 448274, EPI_ISL_448276 to 448279, EPI_ISL_448281 to 448291, EPI_ISL_448293 to 448301, EPI_ISL_448304 to 448305, EPI_ISL_448307, EPI_ISL_448309, EPI_ISL_448311 to 448312, EPI_ISL_448314 to 448317, EPI_ISL_448319 to 448355, EPI_ISL_448357 to 448385, EPI_ISL_448387 to 448388, EPI_ISL_448979 to 448981, EPI_ISL_448983 to 448986, EPI_ISL_448988 to 448992, EPI_ISL_448994 to 449016, EPI_ISL_449018 to 449098, EPI_ISL_449100 to 449102, EPI_ISL_449104 to 449105, EPI_ISL_449107 to 449109, EPI_ISL_449111 to 449113, EPI_ISL_449116 to 449125, EPI_ISL_449127 to 449133, EPI_ISL_449135 to 449136, EPI_ISL_449139 to 449141, EPI_ISL_449143, EPI_ISL_449145 to 449146, EPI_ISL_449148 to 449175, EPI_ISL_453545 to 453550, EPI_ISL_453552 to 453575, EPI_ISL_453577 to 453580, EPI_ISL_453582 to 453589, EPI_ISL_453591 to 453601, EPI_ISL_453603 to 453613, EPI_ISL_457326 to 457339, EPI_ISL_457341 to 457387, EPI_ISL_457389 to 457391, EPI_ISL_457393, EPI_ISL_457395 to 457449, EPI_ISL_457451 to 457473, EPI_ISL_457475 to 457489, EPI_ISL_457491 to 457495, EPI_ISL_457497 to 457518, EPI_ISL_457520 to 457521, EPI_ISL_457523 to 457525, EPI_ISL_457527 to 457535, EPI_ISL_457537 to 457556, EPI_ISL_457558 to 457572, EPI_ISL_461805 to 461844, EPI_ISL_461849 to 461885, EPI_ISL_461887 to 461888, EPI_ISL_461890 to 461892, EPI_ISL_472293 to 472294, EPI_ISL_472296 to 472298, EPI_ISL_472300 to 472328, EPI_ISL_472330, EPI_ISL_472332 to 472333, EPI_ISL_472335 to 472338, EPI_ISL_472342 to 472343, EPI_ISL_472349, EPI_ISL_472352, EPI_ISL_472354, EPI_ISL_472356, EPI_ISL_472358 to 472361, EPI_ISL_472363 to 472368, EPI_ISL_472375, EPI_ISL_472380, EPI_ISL_478719 to 478720, EPI_ISL_480329 to 480330, EPI_ISL_484328, EPI_ISL_484330 to 484332, EPI_ISL_484335, EPI_ISL_484337 to 484338, EPI_ISL_490496, EPI_ISL_490517 to 490520, EPI_ISL_490522 to 490523, EPI_ISL_490525, EPI_ISL_490528 to 490529, EPI_ISL_490532 to 490533, EPI_ISL_490535, EPI_ISL_490539 to 490542, EPI_ISL_490544 to 490545, EPI_ISL_490547 to 490550, EPI_ISL_490553 to 490557, EPI_ISL_490559, EPI_ISL_493541 to 493546, EPI_ISL_499490, EPI_ISL_499495, EPI_ISL_499562, EPI_ISL_499588, EPI_ISL_499591 to 499592, EPI_ISL_499599, EPI_ISL_499601, EPI_ISL_499603, EPI_ISL_499623, EPI_ISL_499631, EPI_ISL_507130, EPI_ISL_512366, EPI_ISL_512369 to 512371, EPI_ISL_512373 to 512375, EPI_ISL_512377 to 512378, EPI_ISL_529267, EPI_ISL_529281 to 529282, EPI_ISL_529378 to 529381, EPI_ISL_529389, EPI_ISL_529437, EPI_ISL_529451 to 529452, EPI_ISL_529489, EPI_ISL_529523, EPI_ISL_529557 to 529589, EPI_ISL_540691 to 540698, EPI_ISL_549344, EPI_ISL_549350 to 549351, EPI_ISL_549355, EPI_ISL_549367, EPI_ISL_549382, EPI_ISL_549479 to 549481, EPI_ISL_549488, EPI_ISL_572400 to 572401, EPI_ISL_572403, EPI_ISL_572406, EPI_ISL_572423, EPI_ISL_572430, EPI_ISL_572437, EPI_ISL_572440 to 572441, EPI_ISL_572447, EPI_ISL_572449, EPI_ISL_572453 to 572454, EPI_ISL_572456, EPI_ISL_572459, EPI_ISL_572462 to 572463, EPI_ISL_572485 to 572486, EPI_ISL_572492 to 572493, EPI_ISL_572495, EPI_ISL_572497 to 572499, EPI_ISL_572503 to 572504, EPI_ISL_572520, EPI_ISL_572533, EPI_ISL_572546, EPI_ISL_572548 to 572551, EPI_ISL_572553, EPI_ISL_572556, EPI_ISL_572563, EPI_ISL_572569, EPI_ISL_572591, EPI_ISL_572600, EPI_ISL_572602, EPI_ISL_572607, EPI_ISL_572615, EPI_ISL_572646, EPI_ISL_572649, EPI_ISL_572651, EPI_ISL_572653, EPI_ISL_572655 to 572656, EPI_ISL_572658, EPI_ISL_572660, EPI_ISL_572665 to 572666, EPI_ISL_572670 to 572671, EPI_ISL_572673, EPI_ISL_572711, EPI_ISL_572717, EPI_ISL_572719, EPI_ISL_572732, EPI_ISL_572735, EPI_ISL_572742, EPI_ISL_572745, EPI_ISL_572749, EPI_ISL_572755, EPI_ISL_572759, EPI_ISL_572768, EPI_ISL_572775, EPI_ISL_572780, EPI_ISL_572785 to 572787, EPI_ISL_572789, EPI_ISL_572791, EPI_ISL_572793, EPI_ISL_572797, EPI_ISL_572804, EPI_ISL_572806, EPI_ISL_572811 to 572812, EPI_ISL_572819, EPI_ISL_572823, EPI_ISL_572825 to 572826, EPI_ISL_572835, EPI_ISL_572842, EPI_ISL_572844, EPI_ISL_572898 to 572901, EPI_ISL_572940 to 572941, EPI_ISL_572954, EPI_ISL_573085 to 573125, EPI_ISL_573127 to 573131, EPI_ISL_573414 to 573417, EPI_ISL_577096 to 577105, EPI_ISL_577107 to 577109, EPI_ISL_584165 to 584166, EPI_ISL_584716 to 584725, EPI_ISL_584727 to 584740, EPI_ISL_584742 to 584748, EPI_ISL_584906 to 584912, EPI_ISL_584914 to 584916, EPI_ISL_584918 to 584922, EPI_ISL_584924 to 584940, EPI_ISL_584942 to 585008, EPI_ISL_585260, EPI_ISL_585583 to 585619, EPI_ISL_586240 to 586242, EPI_ISL_595115 to 595148, EPI_ISL_595150 to 595152, EPI_ISL_595154 to 595164, EPI_ISL_595167 to 595208, EPI_ISL_595221, EPI_ISL_595225 to 595226, EPI_ISL_595229, EPI_ISL_595837 to 595841, EPI_ISL_595843 to 595860, EPI_ISL_595862 to 595873, EPI_ISL_595875 to 595880, EPI_ISL_595882 to 595932, EPI_ISL_595934 to 595938, EPI_ISL_595940 to 595944, EPI_ISL_595946 to 595947, EPI_ISL_595949 to 595951, EPI_ISL_595953 to 595966, EPI_ISL_595969 to 595999, EPI_ISL_596001 to 596006, EPI_ISL_596008 to 596051, EPI_ISL_596053 to 596097, EPI_ISL_596100 to 596113, EPI_ISL_596115 to 596147, EPI_ISL_596149 to 596157, EPI_ISL_596159 to 596172, EPI_ISL_596174 to 596183, EPI_ISL_596185 to 596190, EPI_ISL_626656 to 626657, EPI_ISL_626660, EPI_ISL_626663, EPI_ISL_626667, EPI_ISL_626681, EPI_ISL_626693, EPI_ISL_626698, EPI_ISL_626700 to 626702, EPI_ISL_626704 to 626705, EPI_ISL_626737, EPI_ISL_626741, EPI_ISL_626744 to 626745, EPI_ISL_626753, EPI_ISL_626756, EPI_ISL_626759, EPI_ISL_626765, EPI_ISL_626779, EPI_ISL_626790, EPI_ISL_626795, EPI_ISL_626808, EPI_ISL_626814, EPI_ISL_626819, EPI_ISL_626860, EPI_ISL_626864, EPI_ISL_626868, EPI_ISL_626893 to 626894, EPI_ISL_626914, EPI_ISL_626954 to 626956, EPI_ISL_626964, EPI_ISL_627009, EPI_ISL_627024, EPI_ISL_627080 to 627081, EPI_ISL_627121, EPI_ISL_627148, EPI_ISL_627170, EPI_ISL_627180, EPI_ISL_627193, EPI_ISL_627198, EPI_ISL_627213 to 627214, EPI_ISL_627300, EPI_ISL_627570 to 627583, EPI_ISL_627586 to 627658, EPI_ISL_637284, EPI_ISL_637286, EPI_ISL_637301, EPI_ISL_637442, EPI_ISL_637536, EPI_ISL_637550, EPI_ISL_637557, EPI_ISL_637588 to 637589, EPI_ISL_637602, EPI_ISL_637857, EPI_ISL_637864 to 637866, EPI_ISL_637878, EPI_ISL_638033, EPI_ISL_638581 to 638583, EPI_ISL_650113, EPI_ISL_650116 to 650118, EPI_ISL_650120, EPI_ISL_650128, EPI_ISL_650145, EPI_ISL_650170, EPI_ISL_650174, EPI_ISL_650185, EPI_ISL_650188, EPI_ISL_650190, EPI_ISL_650193, EPI_ISL_650195, EPI_ISL_650197, EPI_ISL_650211, EPI_ISL_650214, EPI_ISL_650224, EPI_ISL_650227, EPI_ISL_650229 to 650231, EPI_ISL_650285, EPI_ISL_650287 to 650288, EPI_ISL_650294, EPI_ISL_650309, EPI_ISL_650317, EPI_ISL_650319, EPI_ISL_650322 to 650323, EPI_ISL_650325, EPI_ISL_650327, EPI_ISL_650330, EPI_ISL_650332, EPI_ISL_650335, EPI_ISL_650343, EPI_ISL_650345, EPI_ISL_650371, EPI_ISL_650385, EPI_ISL_650387, EPI_ISL_650402, EPI_ISL_650404, EPI_ISL_650409, EPI_ISL_650413, EPI_ISL_650417, EPI_ISL_650421, EPI_ISL_650429, EPI_ISL_650446, EPI_ISL_650454, EPI_ISL_650457 to 650462, EPI_ISL_650464 to 650470, EPI_ISL_650472, EPI_ISL_650474 to 650476, EPI_ISL_650480, EPI_ISL_650485, EPI_ISL_650487, EPI_ISL_650489, EPI_ISL_650493, EPI_ISL_650502, EPI_ISL_650512, EPI_ISL_650516, EPI_ISL_650522, EPI_ISL_650528, EPI_ISL_650530, EPI_ISL_650534, EPI_ISL_650541, EPI_ISL_650545 to 650546, EPI_ISL_650548, EPI_ISL_650552, EPI_ISL_650555, EPI_ISL_650557, EPI_ISL_650559, EPI_ISL_650673, EPI_ISL_650677, EPI_ISL_650681, EPI_ISL_650683, EPI_ISL_650685 to 650687, EPI_ISL_650696, EPI_ISL_650716, EPI_ISL_650719 to 650722, EPI_ISL_650737, EPI_ISL_650756, EPI_ISL_650758, EPI_ISL_650761, EPI_ISL_650765 to 650766, EPI_ISL_650817 to 650818, EPI_ISL_650828 to 650829, EPI_ISL_650878 to 650880, EPI_ISL_650883, EPI_ISL_650885, EPI_ISL_650890, EPI_ISL_650894 to 650895, EPI_ISL_650900, EPI_ISL_650903, EPI_ISL_650912 to 650913, EPI_ISL_650915, EPI_ISL_650917 to 650918, EPI_ISL_650931, EPI_ISL_650938, EPI_ISL_650940, EPI_ISL_650943 to 650944, EPI_ISL_650953 to 650955, EPI_ISL_650958,

| | | | |
|---|---|---|---|
| EPI_ISL_650974 to 650977, EPI_ISL_651048, EPI_ISL_651079, EPI_ISL_651082, EPI_ISL_651089 to 651094, EPI_ISL_651100 to 651102, EPI_ISL_651113, EPI_ISL_651116, EPI_ISL_651118, EPI_ISL_651124, EPI_ISL_651133, EPI_ISL_651139, EPI_ISL_651147, EPI_ISL_651150 to 651151, EPI_ISL_651156 to 651157, EPI_ISL_651159, EPI_ISL_651173, EPI_ISL_651182, EPI_ISL_651185, EPI_ISL_651194, EPI_ISL_651198, EPI_ISL_651203, EPI_ISL_651209 to 651210, EPI_ISL_651216 to 651218, EPI_ISL_651220, EPI_ISL_651226, EPI_ISL_651237, EPI_ISL_651244, EPI_ISL_651246 to 651247, EPI_ISL_651254, EPI_ISL_651257 to 651258, EPI_ISL_651261 to 651262, EPI_ISL_651268, EPI_ISL_651317, EPI_ISL_651323 to 651324, EPI_ISL_651326, EPI_ISL_651332 to 651333, EPI_ISL_651335, EPI_ISL_651338, EPI_ISL_651340, EPI_ISL_651349, EPI_ISL_651351 to 651352, EPI_ISL_651371, EPI_ISL_651392, EPI_ISL_651397, EPI_ISL_651399, EPI_ISL_651409, EPI_ISL_651411 to 651412, EPI_ISL_651418 to 651419, EPI_ISL_651434, EPI_ISL_651438, EPI_ISL_651484, EPI_ISL_651512, EPI_ISL_651515 to 651516, EPI_ISL_651526, EPI_ISL_651588, EPI_ISL_652151 to 652153, EPI_ISL_652155 to 652156, EPI_ISL_652216 to 652226, EPI_ISL_652228 to 652236, EPI_ISL_652238 to 652244, EPI_ISL_652246 to 652250, EPI_ISL_652252 to 652285, EPI_ISL_652348 to 652363, EPI_ISL_653027 to 653049, EPI_ISL_653051 to 653056, EPI_ISL_653058, EPI_ISL_664110, EPI_ISL_664135, EPI_ISL_664154, EPI_ISL_664158 to 664159, EPI_ISL_664169, EPI_ISL_664186, EPI_ISL_664222, EPI_ISL_664224, EPI_ISL_664227, EPI_ISL_664235, EPI_ISL_664248, EPI_ISL_664290, EPI_ISL_664334, EPI_ISL_664344, EPI_ISL_664347, EPI_ISL_664351, EPI_ISL_664357 to 664358, EPI_ISL_664366, EPI_ISL_664369, EPI_ISL_664441 to 664442, EPI_ISL_664450, EPI_ISL_664452, EPI_ISL_664460 to 664464, EPI_ISL_664467, EPI_ISL_664476 to 664477, EPI_ISL_664482 to 664485, EPI_ISL_664488, EPI_ISL_664495, EPI_ISL_664502, EPI_ISL_664504, EPI_ISL_664506, EPI_ISL_664508, EPI_ISL_664515, EPI_ISL_664520, EPI_ISL_664522, EPI_ISL_664565 to 664566, EPI_ISL_664628, EPI_ISL_664638, EPI_ISL_664641 to 664642, EPI_ISL_664645, EPI_ISL_664657, EPI_ISL_664688, EPI_ISL_664704, EPI_ISL_664706, EPI_ISL_664714, EPI_ISL_664735, EPI_ISL_664741, EPI_ISL_664743, EPI_ISL_664747, EPI_ISL_664756 to 664758, EPI_ISL_664760, EPI_ISL_664764, EPI_ISL_664838, EPI_ISL_664857 to 664864, EPI_ISL_664866 to 664873, EPI_ISL_664875 to 664940, EPI_ISL_665263, EPI_ISL_665265, EPI_ISL_665268 to 665269, EPI_ISL_665293, EPI_ISL_665295 to 665296, EPI_ISL_665298, EPI_ISL_665306, EPI_ISL_665316, EPI_ISL_665336 to 665337, EPI_ISL_665342 to 665343, EPI_ISL_665346, EPI_ISL_665351, EPI_ISL_665354 to 665358, EPI_ISL_665360, EPI_ISL_665362, EPI_ISL_665364, EPI_ISL_665369, EPI_ISL_665371 to 665372, EPI_ISL_665375, EPI_ISL_665377, EPI_ISL_665380 to 665381, EPI_ISL_665406, EPI_ISL_665411, EPI_ISL_665423, EPI_ISL_665427, EPI_ISL_665429, EPI_ISL_665431, EPI_ISL_665437 to 665438, EPI_ISL_665440, EPI_ISL_665444 to 665450, EPI_ISL_665453 to 665454, EPI_ISL_665456 to 665458, EPI_ISL_665461, EPI_ISL_665463, EPI_ISL_665465, EPI_ISL_665475, EPI_ISL_665481, EPI_ISL_665508, EPI_ISL_665518 to 665520, EPI_ISL_665533 to 665534, EPI_ISL_665542, EPI_ISL_665560, EPI_ISL_665567 to 665568, EPI_ISL_665574, EPI_ISL_665577, EPI_ISL_665581, EPI_ISL_665583, EPI_ISL_665587, EPI_ISL_665590 to 665591, EPI_ISL_665594, EPI_ISL_665596, EPI_ISL_665599, EPI_ISL_665603, EPI_ISL_665638 to 665639, EPI_ISL_665668, EPI_ISL_665692 to 665693, EPI_ISL_665702, EPI_ISL_665712, EPI_ISL_665730 to 665732, EPI_ISL_665746 to 665747, EPI_ISL_665749, EPI_ISL_665788, EPI_ISL_665850, EPI_ISL_665865 to 665867, EPI_ISL_665876 to 665897, EPI_ISL_665957 to 665963, EPI_ISL_678916 to 678919, EPI_ISL_702478, EPI_ISL_702749, EPI_ISL_702840, EPI_ISL_702873, EPI_ISL_702882, EPI_ISL_702902, EPI_ISL_702925, EPI_ISL_702941, EPI_ISL_702944, EPI_ISL_703099, EPI_ISL_703113, EPI_ISL_703187, EPI_ISL_703200, EPI_ISL_703211, EPI_ISL_703264, EPI_ISL_703295, EPI_ISL_703424, EPI_ISL_703590, EPI_ISL_703616, EPI_ISL_703714, EPI_ISL_703733, EPI_ISL_703736, EPI_ISL_703764, EPI_ISL_703769, EPI_ISL_703800, EPI_ISL_703802, EPI_ISL_703805, EPI_ISL_703905, EPI_ISL_703932, EPI_ISL_703945, EPI_ISL_703947, EPI_ISL_703963, EPI_ISL_703979, EPI_ISL_703982, EPI_ISL_703998, EPI_ISL_704054, EPI_ISL_704073, EPI_ISL_704100, EPI_ISL_704123, EPI_ISL_704139, EPI_ISL_704142, EPI_ISL_704145, EPI_ISL_704148, EPI_ISL_704150, EPI_ISL_704158, EPI_ISL_704174, EPI_ISL_704176, EPI_ISL_704181, EPI_ISL_704217, EPI_ISL_704233, EPI_ISL_704236, EPI_ISL_704238, EPI_ISL_704241, EPI_ISL_704580, EPI_ISL_704598 to 704599, EPI_ISL_704609, EPI_ISL_704619 to 704635, EPI_ISL_704709, EPI_ISL_704711, EPI_ISL_704714, EPI_ISL_704716, EPI_ISL_704719, EPI_ISL_704722, EPI_ISL_704725, EPI_ISL_704728, EPI_ISL_704731, EPI_ISL_704734, EPI_ISL_704736, EPI_ISL_704739, EPI_ISL_704741, EPI_ISL_704744, EPI_ISL_704746, EPI_ISL_704749, EPI_ISL_704752, EPI_ISL_704754, EPI_ISL_704757, EPI_ISL_704759, EPI_ISL_704762, EPI_ISL_704765, EPI_ISL_704768, EPI_ISL_704770, EPI_ISL_704772, EPI_ISL_704775, EPI_ISL_704777, EPI_ISL_704786, EPI_ISL_704788, EPI_ISL_704791, EPI_ISL_704794, EPI_ISL_704797, EPI_ISL_704802, EPI_ISL_704820, EPI_ISL_704823, EPI_ISL_704826, EPI_ISL_704831, EPI_ISL_704842, EPI_ISL_704847, EPI_ISL_704850, EPI_ISL_704853, EPI_ISL_704855, EPI_ISL_704887, EPI_ISL_704890, EPI_ISL_704892, EPI_ISL_704908, EPI_ISL_704911, EPI_ISL_704914, EPI_ISL_704916, EPI_ISL_704921, EPI_ISL_704923, EPI_ISL_704926, EPI_ISL_704929, EPI_ISL_705115, EPI_ISL_705211, EPI_ISL_705228, EPI_ISL_705256, EPI_ISL_705262, EPI_ISL_705375, EPI_ISL_705432, EPI_ISL_705491, EPI_ISL_705567 to 705588, EPI_ISL_705590 to 705610, EPI_ISL_705612, EPI_ISL_706357, EPI_ISL_706359 to 706365, EPI_ISL_706367 to 706382, EPI_ISL_706384 to 706389, EPI_ISL_706391, EPI_ISL_706394, EPI_ISL_706396 to 706405, EPI_ISL_706407 to 706411, EPI_ISL_706413 to 706419, EPI_ISL_723673, EPI_ISL_724124 to 724125, EPI_ISL_724127 to 724133, EPI_ISL_724135 to 724188, EPI_ISL_724190 to 724195, EPI_ISL_724197 to 724209, EPI_ISL_725029 to 725060, EPI_ISL_725062 to 725070, EPI_ISL_725072 to 725074, EPI_ISL_725076 to 725079, EPI_ISL_725081 to 725088, EPI_ISL_725090 to 725097, EPI_ISL_725099, EPI_ISL_725101 to 725103, EPI_ISL_725105 to 725117, EPI_ISL_725119 to 725120, EPI_ISL_725122 to 725123, EPI_ISL_725125 to 725135, EPI_ISL_725137 to 725154, EPI_ISL_725156 to 725157, EPI_ISL_725160 to 725162, EPI_ISL_725164, EPI_ISL_725166 to 725173, EPI_ISL_725175 to 725190, EPI_ISL_725192 to 725197, EPI_ISL_725203, EPI_ISL_725205 to 725208, EPI_ISL_725219, EPI_ISL_725221 to 725222, EPI_ISL_725224 to 725234, EPI_ISL_725236 to 725253, EPI_ISL_725255 to 725271, EPI_ISL_725273 to 725276, EPI_ISL_725278 to 725279, EPI_ISL_725281 to 725283, EPI_ISL_725285 to 725287, EPI_ISL_725289 to 725295, EPI_ISL_725298, EPI_ISL_725300, EPI_ISL_725302 to 725315, EPI_ISL_725317 to 725332, EPI_ISL_725335 to 725358, EPI_ISL_725360, EPI_ISL_725363 to 725366, EPI_ISL_725368 to 725371, EPI_ISL_725373, EPI_ISL_725375 to 725405, EPI_ISL_725407 to 725410, EPI_ISL_725412 to 725417, EPI_ISL_725420 to 725423, EPI_ISL_725426 to 725429, EPI_ISL_725431 to 725461, EPI_ISL_725463, EPI_ISL_725465 to 725467, EPI_ISL_725469 to 725478, EPI_ISL_725480 to 725484, EPI_ISL_725487 to 725489, EPI_ISL_725491, EPI_ISL_725493 to 725495, EPI_ISL_725497 to 725499, EPI_ISL_725501 to 725502, EPI_ISL_725505 to 725511, EPI_ISL_725513 to 725516, EPI_ISL_725518, EPI_ISL_741453 to 741480, EPI_ISL_741482 to 741501, EPI_ISL_741503 to 741508, EPI_ISL_741510 to 741521, EPI_ISL_741523 to 741524, EPI_ISL_741526 to 741531, EPI_ISL_741533 to 741597, EPI_ISL_741599 to 741608, EPI_ISL_763372, EPI_ISL_763406, EPI_ISL_763456, EPI_ISL_763466, EPI_ISL_763478, EPI_ISL_763483, EPI_ISL_763490, EPI_ISL_763502, EPI_ISL_763537, EPI_ISL_763549, EPI_ISL_763560, EPI_ISL_763587, EPI_ISL_763655, EPI_ISL_763662, EPI_ISL_763665, EPI_ISL_763681, EPI_ISL_763683 to 763684, EPI_ISL_763689, EPI_ISL_763727, EPI_ISL_763737, EPI_ISL_763749, EPI_ISL_763769, EPI_ISL_763777, EPI_ISL_763780 to 763781, EPI_ISL_763797, EPI_ISL_763851 to 763853, EPI_ISL_763908 to 763924, EPI_ISL_764113 to 764116, EPI_ISL_764126, EPI_ISL_764130, EPI_ISL_764194, EPI_ISL_764205, EPI_ISL_764207, EPI_ISL_764237, EPI_ISL_764445 to 764452, EPI_ISL_764454 to 764461, EPI_ISL_764463 to 764484, EPI_ISL_764486 to 764492, EPI_ISL_819885 to 819887, EPI_ISL_819890, EPI_ISL_819894 to 819918, EPI_ISL_819928, EPI_ISL_819931 to 819935, EPI_ISL_819945, EPI_ISL_819953, EPI_ISL_819959 to 819960, EPI_ISL_819963, EPI_ISL_819970 to 819977, EPI_ISL_819990, EPI_ISL_819996, EPI_ISL_820001 to 820005, EPI_ISL_820011, EPI_ISL_820015, EPI_ISL_820021 to 820023, EPI_ISL_820025 to 820030, EPI_ISL_820041 to 820044, EPI_ISL_820056, EPI_ISL_820062, EPI_ISL_820067 to 820080, EPI_ISL_820082, EPI_ISL_820088, EPI_ISL_820090 to 820093, EPI_ISL_820095 to 820098, EPI_ISL_820102 to 820108, EPI_ISL_820110, EPI_ISL_820112, EPI_ISL_820119, EPI_ISL_820124, EPI_ISL_820126, EPI_ISL_820131, EPI_ISL_820134, EPI_ISL_820136, EPI_ISL_820141, EPI_ISL_820144, EPI_ISL_820146, EPI_ISL_820149, EPI_ISL_820151, EPI_ISL_820154, EPI_ISL_820156, EPI_ISL_820159, EPI_ISL_820161, EPI_ISL_820164, EPI_ISL_820167, EPI_ISL_820169, EPI_ISL_820172, EPI_ISL_820175, EPI_ISL_820178, EPI_ISL_820180, EPI_ISL_820183, EPI_ISL_820186, EPI_ISL_820188, EPI_ISL_820191, EPI_ISL_820193, EPI_ISL_820196, EPI_ISL_820199, EPI_ISL_820202, EPI_ISL_820221, EPI_ISL_820223, EPI_ISL_820231, EPI_ISL_820234, EPI_ISL_820238, EPI_ISL_820241, EPI_ISL_820244, EPI_ISL_820253, EPI_ISL_820261, EPI_ISL_820284, EPI_ISL_820287, EPI_ISL_820289, EPI_ISL_820292, EPI_ISL_820294, EPI_ISL_820300, EPI_ISL_820302, EPI_ISL_820304, EPI_ISL_820306, EPI_ISL_820311, EPI_ISL_820323, EPI_ISL_820353, EPI_ISL_820355, EPI_ISL_820358, EPI_ISL_820360, EPI_ISL_820363, EPI_ISL_820365, EPI_ISL_820368, EPI_ISL_820371, EPI_ISL_820378, EPI_ISL_820388, EPI_ISL_820391, EPI_ISL_820399, EPI_ISL_820404, EPI_ISL_820407, EPI_ISL_820410, EPI_ISL_820412, EPI_ISL_820415, EPI_ISL_820426, EPI_ISL_820436, EPI_ISL_820439, EPI_ISL_820441, EPI_ISL_820444, EPI_ISL_820446, EPI_ISL_820449, EPI_ISL_820451, EPI_ISL_820454, EPI_ISL_820456, EPI_ISL_820459, EPI_ISL_820462, EPI_ISL_820464, EPI_ISL_820467, EPI_ISL_820470, EPI_ISL_820472, EPI_ISL_820475, EPI_ISL_820478, EPI_ISL_820480, EPI_ISL_820483, EPI_ISL_820485, EPI_ISL_820488, EPI_ISL_820490, EPI_ISL_820493, EPI_ISL_820496, EPI_ISL_820498, EPI_ISL_820501, EPI_ISL_820504, EPI_ISL_820506, EPI_ISL_820509, EPI_ISL_820511, EPI_ISL_820514, EPI_ISL_820516, EPI_ISL_820519, EPI_ISL_820521, EPI_ISL_820524, EPI_ISL_820526, EPI_ISL_820528, EPI_ISL_820531, EPI_ISL_820536, EPI_ISL_820539, EPI_ISL_820541, EPI_ISL_820543, EPI_ISL_820546, EPI_ISL_820549, EPI_ISL_820552, EPI_ISL_820554, EPI_ISL_820557, EPI_ISL_842100 to 842106, EPI_ISL_842109, EPI_ISL_842114 to 842115, EPI_ISL_866577 to 866586, EPI_ISL_866802, EPI_ISL_866804 to 866805, EPI_ISL_866807 to 866830 | | | |
| see above | Quadram Institute Bioscience | COVID-19 Genomics UK (COG-UK) Consortium | Alexander J Trotter; Alison E. Mather; Alp Aydin; Ana P. Tedim; Anastasia Kolyva; Andrew Bell; Andrew J. Page; Claire Stuart; Dave J. Baker; Gemma L. Kay; John Wain; Justin O'Grady; Leonardo de Oliveira Martins; Lizzie Meadows; Maria Diaz; Mark Webber; Muhammed Yasir; Nabil-Fareed Alikhan; Ngozi Elumogo; Nicholas M. Thomson; Rachael Stanley; Rachel Gilroy; Reenesh Prakash; Samir Dervisevic; Samuel Bloomfield; Steven Rudder; Thanh Le-Viet |
| EPI_ISL_425462 to 425485, EPI_ISL_425487 to 425495, EPI_ISL_425497 to 425530, EPI_ISL_425532 to 425537, EPI_ISL_425539, EPI_ISL_425541 to 425542, EPI_ISL_425544, EPI_ISL_425546 to 425551, EPI_ISL_425553 to 425555, EPI_ISL_425557 to 425558, EPI_ISL_425560 to 425568, EPI_ISL_425570 to 425571, EPI_ISL_425573 to 425577, EPI_ISL_425579, EPI_ISL_425581 to 425590, EPI_ISL_425592 to 425593, EPI_ISL_425596 to 425597, EPI_ISL_425599 to 425600, EPI_ISL_425602 to 425609, EPI_ISL_425611, EPI_ISL_425613 to 425623, EPI_ISL_425626 to 425633, EPI_ISL_425635 to 425646, EPI_ISL_432668 to 432680, EPI_ISL_432682 to 432683, EPI_ISL_432685 to 432694, EPI_ISL_432696 to 432700, EPI_ISL_432702 to 432709, EPI_ISL_432901 to 432908, EPI_ISL_432910 to 432912, EPI_ISL_432914 to 432917, EPI_ISL_432919 to 432921, EPI_ISL_432923, EPI_ISL_432925 to 432931, EPI_ISL_432933 to 432936, EPI_ISL_432938 to 432940, EPI_ISL_432942, EPI_ISL_432944 to 432951, EPI_ISL_432953 to 432963, EPI_ISL_432965 to 432968, EPI_ISL_432970 to 432975, EPI_ISL_432977 to 432979, EPI_ISL_432981 to 432990, EPI_ISL_432992 to 433021, EPI_ISL_433024 to 433030, EPI_ISL_433032 to 433033, EPI_ISL_433035 to 433039, EPI_ISL_433041 to 433049, EPI_ISL_433051 to 433066, EPI_ISL_441437 to 441444, EPI_ISL_441446 to 441466, EPI_ISL_441468 to 441482, EPI_ISL_441484 to 441486, EPI_ISL_441488 to 441490, EPI_ISL_441492 to 441498, EPI_ISL_441500 to 441503, EPI_ISL_441505 to 441511, EPI_ISL_441515, EPI_ISL_441517 to 441520, EPI_ISL_441524 to 441532, EPI_ISL_441534 to 441546, EPI_ISL_441848 to 441857, EPI_ISL_441861 to 441897, EPI_ISL_448389 to 448403, EPI_ISL_448405 to 448422, EPI_ISL_448424 to 448425, EPI_ISL_448427 to 448428, EPI_ISL_448430 to 448432, EPI_ISL_448434 to 448448, EPI_ISL_453614 to 453616, EPI_ISL_453618 to 453635, EPI_ISL_453637 to 453642, EPI_ISL_453644 to 453653, EPI_ISL_453655 to 453664, EPI_ISL_457573 to 457580, EPI_ISL_461894 to 461898, EPI_ISL_461900 to 461903, EPI_ISL_461905 to 461969, EPI_ISL_472384 to 472412, EPI_ISL_472414 to 472428, EPI_ISL_472430 to 472431, EPI_ISL_478721 to 478728, EPI_ISL_484340 to 484353, EPI_ISL_484374 to 484375, EPI_ISL_484390 to 484391, EPI_ISL_490560 to 490562, EPI_ISL_493547 to 493548, EPI_ISL_493601 to 493602, EPI_ISL_493606 to 493609, EPI_ISL_499266 to 499269, EPI_ISL_499808 to 499809, EPI_ISL_512379 to 512383, EPI_ISL_514452 to 514454, EPI_ISL_526337 to 526358, EPI_ISL_526360 to 526364, EPI_ISL_526366, EPI_ISL_526395 to 526430, EPI_ISL_529264, EPI_ISL_529266, EPI_ISL_529279, EPI_ISL_529288, EPI_ISL_529308 to 529311, EPI_ISL_529383, EPI_ISL_529385, EPI_ISL_529387, EPI_ISL_529410 to 529412, EPI_ISL_529414, EPI_ISL_529423, EPI_ISL_529436, EPI_ISL_529450, EPI_ISL_529454 to 529456, EPI_ISL_529463, EPI_ISL_529467, EPI_ISL_529479, EPI_ISL_529483, EPI_ISL_529486, EPI_ISL_529504, EPI_ISL_529519, EPI_ISL_529528 to 529556, EPI_ISL_529667, EPI_ISL_534756 to 534757, EPI_ISL_535183, EPI_ISL_540640 to 540647, EPI_ISL_540649 to 540690, EPI_ISL_540699 to 540702, EPI_ISL_540704 to 540711, EPI_ISL_540713 to 540716, EPI_ISL_540718, EPI_ISL_540894 to 540896, EPI_ISL_549334, EPI_ISL_549357, EPI_ISL_549376, EPI_ISL_549491 to 549516, EPI_ISL_572444, EPI_ISL_572460, EPI_ISL_572475 to 572476, EPI_ISL_572509, EPI_ISL_572582, EPI_ISL_572598, EPI_ISL_572603, EPI_ISL_572706, EPI_ISL_572799, EPI_ISL_572872, EPI_ISL_573418 to 573424, EPI_ISL_573426 to 573427, EPI_ISL_573429 to 573436, EPI_ISL_573450 to 573477, EPI_ISL_577110 to 577166, EPI_ISL_577168 to 577192, EPI_ISL_584381 to 584439, EPI_ISL_584750 to 584765, EPI_ISL_584767 to 584778, EPI_ISL_584780 to 584799, EPI_ISL_584801 to 584805, EPI_ISL_584808, EPI_ISL_585627 to 585629, EPI_ISL_594923 to 594924, EPI_ISL_594926 to 594948, EPI_ISL_594951 to 594986, EPI_ISL_595046 to 595099, EPI_ISL_595232 to 595293, EPI_ISL_595295 to 595300, EPI_ISL_595302 to 595317, EPI_ISL_595319 to 595325, EPI_ISL_611546, EPI_ISL_611582, EPI_ISL_611604, EPI_ISL_611659, EPI_ISL_611662, EPI_ISL_611670, EPI_ISL_611676 to 611677, EPI_ISL_611680, EPI_ISL_611753, EPI_ISL_611791, EPI_ISL_611810, EPI_ISL_611812 to 611813, EPI_ISL_611831, EPI_ISL_611833, EPI_ISL_611840, EPI_ISL_611919, EPI_ISL_611954 to 611956, EPI_ISL_611963, EPI_ISL_612032, EPI_ISL_612077, EPI_ISL_612107, EPI_ISL_612158 to 612159, EPI_ISL_612181 to 612183, EPI_ISL_612192, EPI_ISL_612197, EPI_ISL_612571 to 612606, EPI_ISL_613307 to 613355, EPI_ISL_626670, EPI_ISL_626688, EPI_ISL_626695, EPI_ISL_626740, EPI_ISL_626750, EPI_ISL_626811, EPI_ISL_626816, EPI_ISL_626834, EPI_ISL_627053, EPI_ISL_627068, EPI_ISL_627147, EPI_ISL_627215 to 627216, EPI_ISL_627304, EPI_ISL_627309 to 627310, EPI_ISL_627666, EPI_ISL_627668, EPI_ISL_627670 to 627729, EPI_ISL_637298, EPI_ISL_637300, EPI_ISL_637342 to 637344, EPI_ISL_637419, EPI_ISL_637427, EPI_ISL_637555, EPI_ISL_637558, EPI_ISL_637582, EPI_ISL_637660 to 637661, EPI_ISL_637798 to 637800, EPI_ISL_638584 to 638596, EPI_ISL_638598 to 638613, EPI_ISL_650136, EPI_ISL_650150, EPI_ISL_650154, EPI_ISL_650157, EPI_ISL_650178, EPI_ISL_650186 to 650187, EPI_ISL_650238, EPI_ISL_650266, EPI_ISL_650276, EPI_ISL_650291, EPI_ISL_650295, EPI_ISL_650314, EPI_ISL_650347, EPI_ISL_650386, EPI_ISL_650430, EPI_ISL_650443, EPI_ISL_650449 to 650450, EPI_ISL_650471, EPI_ISL_650556, EPI_ISL_650558, EPI_ISL_650599, EPI_ISL_650641, EPI_ISL_650701, EPI_ISL_650738, EPI_ISL_650767, EPI_ISL_650786, EPI_ISL_650803, EPI_ISL_650811, EPI_ISL_650820 to 650821, EPI_ISL_650830, EPI_ISL_650835, EPI_ISL_650846, EPI_ISL_650937, EPI_ISL_650979, EPI_ISL_650987, EPI_ISL_651065, EPI_ISL_651071 to 651073, EPI_ISL_651096, EPI_ISL_651107, EPI_ISL_651166, EPI_ISL_651189, EPI_ISL_651232, EPI_ISL_651242, EPI_ISL_651249, EPI_ISL_651274, EPI_ISL_651286, EPI_ISL_651293, EPI_ISL_651308 to 651309, EPI_ISL_651313, EPI_ISL_651386, EPI_ISL_651389, EPI_ISL_651398, EPI_ISL_651401, EPI_ISL_651466, EPI_ISL_651504, EPI_ISL_651528, EPI_ISL_651584 to 651585, EPI_ISL_652184 to 652215, EPI_ISL_652364 to 652387, EPI_ISL_652389 to 652396, EPI_ISL_652398 to 652413, EPI_ISL_652627 to 652674, EPI_ISL_665290 to 665291, EPI_ISL_665345, EPI_ISL_665365 to 665367, EPI_ISL_665415, EPI_ISL_665420 to 665421, EPI_ISL_665424, EPI_ISL_665426, EPI_ISL_665430, EPI_ISL_665441, EPI_ISL_665443, EPI_ISL_665452, EPI_ISL_665510, EPI_ISL_665512 to 665513, EPI_ISL_665528, EPI_ISL_665543 to 665545, EPI_ISL_665550 to 665551, EPI_ISL_665561 to 665562, EPI_ISL_665601, EPI_ISL_665610, EPI_ISL_665612, EPI_ISL_665622 to 665623, EPI_ISL_665650, EPI_ISL_665699, EPI_ISL_665724, EPI_ISL_665740, EPI_ISL_665761, EPI_ISL_665789, EPI_ISL_665898 to 665901, EPI_ISL_665903 to 665945, EPI_ISL_665971 to 665982, EPI_ISL_678920 to 678967, EPI_ISL_678969 to 679007, EPI_ISL_679249 to 679308, EPI_ISL_679310 to 679314, EPI_ISL_679924 to 679930, EPI_ISL_679932 to 679945, EPI_ISL_702841, EPI_ISL_702846, EPI_ISL_702879, EPI_ISL_702889, EPI_ISL_703124, EPI_ISL_703720, EPI_ISL_703775 to 703776, EPI_ISL_703789, EPI_ISL_703792, EPI_ISL_703794, EPI_ISL_703853, EPI_ISL_704315, EPI_ISL_704581 to 704583, EPI_ISL_704780, EPI_ISL_705094, EPI_ISL_705213, EPI_ISL_705223, EPI_ISL_705380, EPI_ISL_705487, EPI_ISL_705613 to 705614, EPI_ISL_706420 to 706437, EPI_ISL_725519 to 725539, EPI_ISL_725605 to 725617, EPI_ISL_725619 to 725669, EPI_ISL_741609 to 741653, EPI_ISL_741655 to 741657, EPI_ISL_763389, EPI_ISL_763444, EPI_ISL_763449, EPI_ISL_763547, EPI_ISL_763580, EPI_ISL_763650, EPI_ISL_763663, EPI_ISL_763673, EPI_ISL_763675, EPI_ISL_763678, EPI_ISL_763726, EPI_ISL_763819, EPI_ISL_763854 to 763858, EPI_ISL_763925 to 763927, EPI_ISL_764189 to 764190, EPI_ISL_764230, EPI_ISL_764233, EPI_ISL_764493 to 764524, EPI_ISL_819458 to 819459, EPI_ISL_819464, EPI_ISL_819558, EPI_ISL_819577, EPI_ISL_819579 to 819584, EPI_ISL_819594 to 819604, EPI_ISL_819609 to 819611, EPI_ISL_819647 to 819652, EPI_ISL_819671 to 819674, EPI_ISL_819686 to 819688, EPI_ISL_819724 to 819731, EPI_ISL_819757 to 819758, EPI_ISL_819763, EPI_ISL_819795, EPI_ISL_819799 to 819802, EPI_ISL_819821, EPI_ISL_819828 to 819834, EPI_ISL_819851, EPI_ISL_819864 to 819865, EPI_ISL_819867, EPI_ISL_819942, EPI_ISL_820006, EPI_ISL_820297, EPI_ISL_820394, EPI_ISL_820402, EPI_ISL_820418, EPI_ISL_820423, EPI_ISL_820428, EPI_ISL_820431, EPI_ISL_820433, EPI_ISL_838346, EPI_ISL_838348, EPI_ISL_839981 to 839994, EPI_ISL_866889 to 866891, EPI_ISL_866893 to 866905, EPI_ISL_866990 to 866991 | | | |
| see above | Queens Medical Centre, Clinical Microbiology Department / DeepSeq Nottingham | COVID-19 Genomics UK (COG-UK) Consortium | Christopher Moore; Fei Sang; Gemma Clark; Hannah Howson-Wells; Joeseph Chappell; Johnny Debebe; Jonathan Ball; Joseph Chappell; Manjinder Khakh; Matthew Carlisle; Matthew Loose; Michelle M Lister; Nadine Holmes; Nichola Duckworth; Patrick McClure; Sarah Walsh; Theocharis Tsoleridis; Tim Sloan; Vicki M Fleming; Victoria Wright; Wendy Smith |
| EPI_ISL_833228 to 833229 | R.P. GUARDIAGRELE Ospedale di Comunità | Istituto Zooprofilattico Sperimentale dell'Abruzzo e Molise "G.Caporale" | Ancora M; Calistri P; Cammà C; Curini V; Di Domenico M; Di Pasquale A; Lorusso A; Mangone I; Marcacci M; Puglia I; Rinaldi A; Savini G. |
| EPI_ISL_413569 to 413570, EPI_ISL_413575 to 413576, EPI_ISL_413581 | RIVM | Erasmus Medical Center | Anne van der Linden; Annemiek van der Eijk; Aura Timen; Bas Oude Munnink; Claudia Schapendonk; Corien Swaan; Corine GeurtsvanKessel; David Nieuwenhuijse; Irina Chestakova; Jeroen van Kampen; Jolanda Voermans; Madelief Mollers; Manon Haverkate; Marion Koopmans; Mark Pronk; Mart Stein; Pascal Lexmond; Reina Sikkema; Richard Molenkamp; Sandra Kengne Kamga Mobou; on behalf of the Dutch national COVID-19 response team. |
| EPI_ISL_833083 to 833084 | RPS | Istituto Zooprofilattico Sperimentale dell'Abruzzo e Molise "G. Caporale" | Ancora M; Calistri P; Cammà C; Curini V; Delli Compagni E; Di Domenico M; Di Pasquale A; Lorusso A; Mangone I; Marcacci M; Puglia I; Rinaldi A; Savini G |
| EPI_ISL_806745 | RSA Giulianova | Istituto Zooprofilattico Sperimentale dell'Abruzzo e Molise "G. Caporale" | Ancora M; Calistri P; Cammà C; Curini V; Di Domenico M; Di Pasquale A; Lorusso A; Mangone I; Marcacci M; Puglia I; Rinaldi A; Savini G |
| EPI_ISL_529017 | RSA San Raffaele Sulmona | Istituto Zooprofilattico Sperimentale dell'Abruzzo e Molise | Ancora M; Cammà C; Curini V; Di Domenico M; Di Pasquale A; Lorusso A; Mangone I; Marcacci M; Puglia I; Rinaldi A; Savini G. |

| | | | |
|---|---|---|---|
| | | "G.Caporale" | |
| EPI_ISL_529019 | RSA/RP Villa San Giovanni - Gruppo Edos | Istitituto Zooprofilattico Sperimentale dell'Abruzzo e Molise "G.Caporale" | Ancora M; Cammà C; Curini V; Di Domenico M; Di Pasquale A; Lorusso A; Mangone I; Marcacci M; Puglia I; Rinaldi A; Savini G. |
| EPI_ISL_436725 | RSA/RP Villa San Giovanni - Gruppo Edos | Istituto Zooprofilattico Sperimentale dell'Abruzzo e Molise "G.Caporale" | Ancora M; Cammà C; Curini V; Di Domenico M; Di Pasquale A; Lorusso A; Mangone I; Marcacci M; Puglia I; Rinaldi A; Savini G |
| EPI_ISL_450511, EPI_ISL_450513, EPI_ISL_450515, EPI_ISL_450517 | Rafik Hariri University Hospital | Rafik Hariri University Hospital | Rita Feghali |
| EPI_ISL_414949 | Regional Virus Laboratory, Belfast | Public Health Wales Microbiology Cardiff | Catherine Moore; Conall McCaughey; Joanne Watkins; Sally Corden; Tanya Curran; Tom Connor |
| EPI_ISL_441355 to 441366, EPI_ISL_441368 to 441376, EPI_ISL_441378 to 441412, EPI_ISL_441414 to 441436, EPI_ISL_448918 to 448936, EPI_ISL_448938 to 448951, EPI_ISL_448954 to 448974, EPI_ISL_448976 to 448977, EPI_ISL_453465 to 453485, EPI_ISL_461773 to 461790, EPI_ISL_472148, EPI_ISL_585092 to 585097, EPI_ISL_585099 to 585116, EPI_ISL_585118 to 585193, EPI_ISL_585195 to 585229, EPI_ISL_585231 to 585247, EPI_ISL_585249 to 585259, EPI_ISL_611531, EPI_ISL_611807 to 611808, EPI_ISL_612546 to 612548, EPI_ISL_626828, EPI_ISL_626859, EPI_ISL_626953, EPI_ISL_627000, EPI_ISL_627008, EPI_ISL_627038, EPI_ISL_627046, EPI_ISL_627070, EPI_ISL_627199, EPI_ISL_627518 to 627556, EPI_ISL_627558 to 627569, EPI_ISL_650329, EPI_ISL_650334, EPI_ISL_650378, EPI_ISL_650415, EPI_ISL_650452, EPI_ISL_650513, EPI_ISL_650524, EPI_ISL_650535, EPI_ISL_650543, EPI_ISL_650598, EPI_ISL_650810, EPI_ISL_650888, EPI_ISL_650898, EPI_ISL_650901, EPI_ISL_650923, EPI_ISL_650930, EPI_ISL_651097, EPI_ISL_651128, EPI_ISL_651215, EPI_ISL_651318, EPI_ISL_651329, EPI_ISL_651387, EPI_ISL_651404, EPI_ISL_651501, EPI_ISL_652304 to 652317, EPI_ISL_652319 to 652336, EPI_ISL_680198 to 680226, EPI_ISL_680228 to 680235, EPI_ISL_680239 to 680241, EPI_ISL_680243 to 680252, EPI_ISL_680254, EPI_ISL_680256 to 680258, EPI_ISL_680260 to 680262, EPI_ISL_680264 to 680282, EPI_ISL_680284 to 680286, EPI_ISL_680289 to 680291, EPI_ISL_680293, EPI_ISL_680295 to 680297, EPI_ISL_680299 to 680317, EPI_ISL_680319 to 680322, EPI_ISL_680324 to 680346, EPI_ISL_680348 to 680349, EPI_ISL_680351 to 680409, EPI_ISL_680415 to 680417, EPI_ISL_680419 to 680421, EPI_ISL_680423 to 680439, EPI_ISL_680441, EPI_ISL_742118 to 742160, EPI_ISL_742162 to 742174, EPI_ISL_763367, EPI_ISL_763373, EPI_ISL_763388, EPI_ISL_763400 to 763401, EPI_ISL_763405, EPI_ISL_763408, EPI_ISL_763440, EPI_ISL_763446, EPI_ISL_763463, EPI_ISL_763475, EPI_ISL_763482, EPI_ISL_763524, EPI_ISL_763573, EPI_ISL_763583, EPI_ISL_763596, EPI_ISL_763642, EPI_ISL_763646, EPI_ISL_763719, EPI_ISL_763723, EPI_ISL_763729, EPI_ISL_764127, EPI_ISL_764143, EPI_ISL_764148, EPI_ISL_764154, EPI_ISL_764173, EPI_ISL_764187, EPI_ISL_764206, EPI_ISL_764261 to 764263, EPI_ISL_764387 to 764425, EPI_ISL_764427 to 764444, EPI_ISL_843151 to 843161, EPI_ISL_843163 to 843166, EPI_ISL_843168 to 843169, EPI_ISL_866349 to 866352, EPI_ISL_866364 to 866365, EPI_ISL_866372 | | | |
| see above | Regional Virus Laboratory, Belfast Health and Social Care Trust | COVID-19 Genomics UK (COG-UK) Consortium | Alison Watt; Ciara Cox; Conall McCaughey; David Simpson; Derek Fairley; James McKenna; Mairead Connor; Susan Feeney; Tanya Curran; Zoltan Molnar |
| EPI_ISL_459358, EPI_ISL_459372, EPI_ISL_489213, EPI_ISL_489358 | Regional Virus Laboratory, Belfast Health and Social Care Trust | Wellcome Sanger Institute for the COVID-19 Genomics UK (COG-UK) Consortium | Alison Watt; Ciara Cox; Conall McCaughey; Cordelia Langford; David K. Jackson; David Simpson; Derek Fairley; Dominic Kwiatkowski; Ewan Harrison; Ian Johnston; James McKenna; John Sillitoe on behalf of the Wellcome Sanger Institute COVID-19 Surveillance Team; Mairead Connor; Roberto Amato; Sonia Goncalves; Susan Feeney; Tanya Curran; Zoltan Molnar; and Alex Alderton |
| EPI_ISL_441659, EPI_ISL_441662 to 441663, EPI_ISL_441670, EPI_ISL_441675 to 441676, EPI_ISL_441690 to 441691, EPI_ISL_441693 to 441694, EPI_ISL_441696 to 441699, EPI_ISL_441708, EPI_ISL_441712, EPI_ISL_441714, EPI_ISL_441716 to 441717, EPI_ISL_441719, EPI_ISL_441730, EPI_ISL_441733, EPI_ISL_441737, EPI_ISL_441747, EPI_ISL_441751, EPI_ISL_441753, EPI_ISL_441761, EPI_ISL_441765 to 441766, EPI_ISL_441771, EPI_ISL_441775, EPI_ISL_441781, EPI_ISL_459324 to 459338, EPI_ISL_459340 to 459354, EPI_ISL_459357, EPI_ISL_459359 to 459369, EPI_ISL_459371, EPI_ISL_459373, EPI_ISL_459375 to 459383, EPI_ISL_459385 to 459398, EPI_ISL_459400 to 459409, EPI_ISL_469844 to 469857, EPI_ISL_469859 to 469890, EPI_ISL_470013 to 470065, EPI_ISL_470067 to 470089, EPI_ISL_470534, EPI_ISL_482033 to 482036, EPI_ISL_482038 to 482050, EPI_ISL_482052 to 482056, EPI_ISL_482070, EPI_ISL_482075, EPI_ISL_482077, EPI_ISL_482082, EPI_ISL_482091, EPI_ISL_482096, EPI_ISL_482101 to 482102, EPI_ISL_482105, EPI_ISL_482115, EPI_ISL_482132 to 482137, EPI_ISL_482139 to 482159, EPI_ISL_488874 to 488876, EPI_ISL_489156 to 489168, EPI_ISL_489170 to 489191, EPI_ISL_489193 to 489212, EPI_ISL_489214 to 489357, EPI_ISL_489359 to 489379, EPI_ISL_500954 to 500972, EPI_ISL_500974 to 501012, EPI_ISL_501014 to 501071 | | | |
| see above | Regional Virus Laboratory, Belfast Health and Social Care Trust | Wellcome Sanger Institute for the COVID-19 Genomics UK (COG-UK) consortium | Alex Alderton; Alison Watt; Ciara Cox; Conall McCaughey; Cordelia Langford; David K. Jackson; David Simpson; Derek Fairley; Dominic Kwiatkowski; Ewan Harrison; Ian Johnston; James McKenna; John Sillitoe on behalf of the Wellcome Sanger Institute COVID-19 Surveillance Team (http://www.sanger.ac.uk/covid-team); Mairead Connor; Roberto Amato; Sonia Goncalves; Susan Feeney; Tanya Curran; Zoltan Molnar; and Alex Alderton |
| EPI_ISL_451934 | Research Unit, University Hospital for Infectious Diseases "Dr. Fran Mihaljevi" | Cicin Sain lab, Helmholtz Centre for Infection Research | Ivan-Christian Kurolt; Kathrin Eschke; Zeeshan Chaudhry; Željka Maak Šafranko |
| EPI_ISL_637114 to 637119, EPI_ISL_637121 to 637122, EPI_ISL_637124 to 637149, EPI_ISL_637151 to 637184, EPI_ISL_639821 to 639822, EPI_ISL_644162, EPI_ISL_645133 to 645136, EPI_ISL_649108, EPI_ISL_649110, EPI_ISL_649786 to 649790, EPI_ISL_649793 to 649797, EPI_ISL_649799 to 649803, EPI_ISL_649806 to 649807, EPI_ISL_649809 to 649825, EPI_ISL_649827 to 649892, EPI_ISL_649936 to 649937, EPI_ISL_653933, EPI_ISL_653935 to 653949, EPI_ISL_653951 to 654015, EPI_ISL_660546 to 660550, EPI_ISL_660601, EPI_ISL_660604, EPI_ISL_660743 to 660745, EPI_ISL_660747, EPI_ISL_660751 to 660753, EPI_ISL_660755 to 660767, EPI_ISL_660769 to 660794, EPI_ISL_661175, EPI_ISL_664028, EPI_ISL_664031, EPI_ISL_664035, EPI_ISL_664042 to 664043, EPI_ISL_664055, EPI_ISL_664061 to 664062, EPI_ISL_664065, EPI_ISL_664067, EPI_ISL_664070, EPI_ISL_664080 to 664084, EPI_ISL_666726, EPI_ISL_666728 to 666730, EPI_ISL_666732 to 666736, EPI_ISL_666739 to 666741, EPI_ISL_666745, EPI_ISL_666748 to 666770, EPI_ISL_668445, EPI_ISL_676511, EPI_ISL_678644 to 678649, EPI_ISL_678651 to 678679, EPI_ISL_678681 to 678686, EPI_ISL_678688 to 678704, EPI_ISL_678706 to 678717, EPI_ISL_678719 to 678720, EPI_ISL_678722 to 678724, EPI_ISL_678738, EPI_ISL_678745, EPI_ISL_678749, EPI_ISL_678752, EPI_ISL_678756, EPI_ISL_678768, EPI_ISL_678775, EPI_ISL_678780, EPI_ISL_678787 to 678788, EPI_ISL_678800, EPI_ISL_678804, EPI_ISL_678813, EPI_ISL_678828, EPI_ISL_683452 to 683455, EPI_ISL_683457 to 683463, EPI_ISL_683465 to 683475, EPI_ISL_683478 to 683512, EPI_ISL_683514 to 683530, EPI_ISL_683532 to 683533, EPI_ISL_683536 to 683537, EPI_ISL_683539 to 683549, EPI_ISL_683551 to 683562, EPI_ISL_683564 to 683591, EPI_ISL_686108, EPI_ISL_686536 to 686545, EPI_ISL_686547 to 686548, EPI_ISL_686550 to 686586, EPI_ISL_686588 to 686596, EPI_ISL_686598 to 686611, EPI_ISL_693393 to 693418, EPI_ISL_693420 to 693435, EPI_ISL_693437 to 693460, EPI_ISL_693462 to 693470, EPI_ISL_693484 to 693487 | | | |
| see above | Respiratory Virus Unit, Microbiology Services Colindale, Public Health England | COVID-19 Genomics UK (COG-UK) Consortium | PHE Covid Sequencing Team |
| EPI_ISL_414005 to 414013, EPI_ISL_414040 to 414044, EPI_ISL_414522 to 414526, EPI_ISL_415129 to 415136, EPI_ISL_415138 to 415150, EPI_ISL_417213 to 417235, EPI_ISL_417237, EPI_ISL_417239 to 417244, EPI_ISL_417246 to 417256, EPI_ISL_417258, EPI_ISL_417260 to 417276, EPI_ISL_417278 to 417315, EPI_ISL_418667 to 418740, EPI_ISL_418742 to 418770, EPI_ISL_420457 to 420460, EPI_ISL_420462 to 420530, EPI_ISL_420631 to 420638, EPI_ISL_420640, EPI_ISL_420642 to 420647, EPI_ISL_420649 to 420656, EPI_ISL_420658 to 420705, EPI_ISL_420707 to 420726, EPI_ISL_420728 to 420742, EPI_ISL_420744, EPI_ISL_420746 to 420776, EPI_ISL_420910, EPI_ISL_421768 to 421803, EPI_ISL_421805 to 421841, EPI_ISL_421843, EPI_ISL_421845 to 421857, EPI_ISL_421859, EPI_ISL_421861 to 421870, EPI_ISL_421873 to 421877, EPI_ISL_421879 to 421887, EPI_ISL_421889 to 421896, EPI_ISL_421899 to 421916, EPI_ISL_421918 to 421939, EPI_ISL_421941 to 421949, EPI_ISL_421951, EPI_ISL_421953 to 421954, EPI_ISL_421956 to 421984, EPI_ISL_421986 to 422012, EPI_ISL_423044 to 423052, EPI_ISL_423054 to 423127, EPI_ISL_423129 to 423145, EPI_ISL_423147 to 423151, EPI_ISL_423153 to 423154, EPI_ISL_423156 to 423170, EPI_ISL_423172 to 423186, EPI_ISL_423188 to 423191, EPI_ISL_423193 to 423201, EPI_ISL_423204 to 423238, EPI_ISL_423240 to 423246, EPI_ISL_423248 to 423256, EPI_ISL_423258 to 423287, EPI_ISL_423289 to 423309, EPI_ISL_423311 to 423359, EPI_ISL_423361 to 423374, EPI_ISL_423379 to 423381, EPI_ISL_423384 to 423433, EPI_ISL_423435 to 423451, EPI_ISL_423453, EPI_ISL_423455 to 423461, EPI_ISL_423465 to 423477, EPI_ISL_423479 to 423518, EPI_ISL_423520 to 423524, EPI_ISL_423526 to 423530, EPI_ISL_423532 to 423536, EPI_ISL_423544 to 423545, EPI_ISL_423549 to 423550, EPI_ISL_423552 to 423557, EPI_ISL_423559 to 423563, EPI_ISL_423565 to 423568, EPI_ISL_423570 to 423576, EPI_ISL_423579, EPI_ISL_423581 to 423632, EPI_ISL_423635 to 423645, EPI_ISL_423648 to 423662, EPI_ISL_423664 to 423667, EPI_ISL_423669 to 423675, EPI_ISL_423677 to 423704, EPI_ISL_423706, EPI_ISL_423709 to 423713, EPI_ISL_423715 to 423725, EPI_ISL_423727 to 423735, EPI_ISL_423738 to 423741, EPI_ISL_423745 to 423746, EPI_ISL_423748 to 423756, EPI_ISL_423758 to 423759, EPI_ISL_423761 to 423764, EPI_ISL_423766 to 423780, EPI_ISL_423782 to 423787, EPI_ISL_423790 to 423791, EPI_ISL_423795 to 423799, EPI_ISL_423801 to 423805, EPI_ISL_423807 to 423813, EPI_ISL_423817 to 423818, EPI_ISL_423820, EPI_ISL_423822 to 423826, EPI_ISL_423828 to 423830, EPI_ISL_423832, EPI_ISL_423834 to 423842, EPI_ISL_423844 to 423861, EPI_ISL_423863 to 423866, EPI_ISL_423868 to 423874, EPI_ISL_423876 to 423878, EPI_ISL_423880 to 423890, EPI_ISL_423892 to 423895, EPI_ISL_423897 to 423921, EPI_ISL_423923 to 423944, EPI_ISL_423946 to 423958, EPI_ISL_423960 to 423970, EPI_ISL_423972 to 423976, EPI_ISL_423978 to 423997, EPI_ISL_423999 to 424009, EPI_ISL_424011 to 424017, EPI_ISL_424019 to 424020, EPI_ISL_424022 to 424055, EPI_ISL_424057 to 424084, EPI_ISL_424086 to 424095, EPI_ISL_424098 to 424101, EPI_ISL_424103 to 424121, EPI_ISL_424124 to 424136, EPI_ISL_424139 to 424154, EPI_ISL_424156 to 424165, EPI_ISL_425024, EPI_ISL_452249 to 452253, EPI_ISL_452255 to 452258, EPI_ISL_459958 to 459961, EPI_ISL_464170 to 464215, EPI_ISL_464217 to 464234, EPI_ISL_464236 to 464254, EPI_ISL_464256, EPI_ISL_464258 to 464263, EPI_ISL_464265 to 464286, EPI_ISL_464288 to 464298, EPI_ISL_464300 to 464327, EPI_ISL_464329 to 464334, EPI_ISL_464336 to 464340, EPI_ISL_464342 to 464357, EPI_ISL_464359 to 464389, EPI_ISL_464391, EPI_ISL_464393 to 464402, EPI_ISL_464404 to 464407, EPI_ISL_464409 to 464414, EPI_ISL_464416 to 464419, EPI_ISL_464422 to 464423, EPI_ISL_464426 to 464443, EPI_ISL_464445 to 464452, EPI_ISL_464454 to 464460, EPI_ISL_464462, EPI_ISL_464465 to 464468, EPI_ISL_464470, EPI_ISL_464472 to 464473, EPI_ISL_464475 to 464478, EPI_ISL_464480 to 464494, EPI_ISL_464496 to 464508, EPI_ISL_464511 to 464512, EPI_ISL_464514 to 464520, EPI_ISL_464522 to 464536, EPI_ISL_464538 to 464545, EPI_ISL_464548 to 464558, EPI_ISL_464560 to 464607, EPI_ISL_464609, EPI_ISL_464611, EPI_ISL_464613 to 464624, EPI_ISL_464626 to 464627, EPI_ISL_464629 to 464675, EPI_ISL_464677 to 464713, EPI_ISL_464715 to 464716, EPI_ISL_464718 to 464727, EPI_ISL_464729 to 464735, EPI_ISL_464737 to 464749, EPI_ISL_464751 to 464752, EPI_ISL_464754 to 464768, EPI_ISL_464771 to 464786, EPI_ISL_464788 to 464801, EPI_ISL_464803 to 464810, EPI_ISL_464812 to 464827, EPI_ISL_464829 to 464831, EPI_ISL_464833 to 464843, EPI_ISL_464845 to 464847, EPI_ISL_464849 to 464871, EPI_ISL_464873 to 464884, EPI_ISL_464886 to 464899, EPI_ISL_464903 to 464914, EPI_ISL_464916 to 464920, EPI_ISL_464922 to 464930, EPI_ISL_464932, EPI_ISL_464934 to 464941, EPI_ISL_464943 to 464965, EPI_ISL_464967 to 464978, EPI_ISL_464980 to 464987, EPI_ISL_464989 to 465003, EPI_ISL_465005 to 465008, EPI_ISL_465010 to 465024, EPI_ISL_465026 to 465036, EPI_ISL_465038 to 465046, EPI_ISL_465048 to 465057, EPI_ISL_465059 to 465065, EPI_ISL_465068, EPI_ISL_465070 to 465072, EPI_ISL_465075 to 465086, EPI_ISL_465088 to 465098, EPI_ISL_465100 to 465101, EPI_ISL_465103 to 465117, EPI_ISL_465119 to 465121, EPI_ISL_465123 to 465135, EPI_ISL_465137 to 465147, EPI_ISL_465150 to 465151, EPI_ISL_465154 to 465162, EPI_ISL_465165, EPI_ISL_465167 to 465170, EPI_ISL_465173 to 465174, EPI_ISL_465177 to 465178, EPI_ISL_465180 to 465184, EPI_ISL_465186 to 465188, EPI_ISL_465190, EPI_ISL_465192 to 465193, EPI_ISL_465195 to 465199, EPI_ISL_465202 to 465206, EPI_ISL_465208, EPI_ISL_465210 to 465211, EPI_ISL_465213 to 465215, EPI_ISL_465217 to 465222, EPI_ISL_465225 to 465226, EPI_ISL_465228 to 465230, EPI_ISL_465235, EPI_ISL_465237 to 465246, EPI_ISL_465248 to 465255, EPI_ISL_465257 to 465266, EPI_ISL_465270 to 465278, EPI_ISL_465280 to 465291, EPI_ISL_465294, EPI_ISL_465297 to 465304, EPI_ISL_465306 to 465315, EPI_ISL_465317 to 465325, EPI_ISL_465327 to 465328, EPI_ISL_465332, EPI_ISL_465334 to 465335, EPI_ISL_465337 to 465339, EPI_ISL_465341 to 465344, EPI_ISL_465347 to 465353, EPI_ISL_465357 to 465359, EPI_ISL_465361 to 465377, EPI_ISL_465381 to 465393, EPI_ISL_465395 to 465400, EPI_ISL_465402, EPI_ISL_465404 to 465407, EPI_ISL_465409 to 465423, EPI_ISL_465425 to 465431, EPI_ISL_465433 to 465450, EPI_ISL_465452 to 465459, EPI_ISL_465462 to 465464, EPI_ISL_465466, EPI_ISL_465468 to 465474, EPI_ISL_465476 to 465488, EPI_ISL_465490 to 465501, EPI_ISL_465503 to 465506, EPI_ISL_465508, EPI_ISL_465510 to 465517, EPI_ISL_465519, EPI_ISL_465521 to 465528, EPI_ISL_465530 to 465538, EPI_ISL_465540 to 465546, EPI_ISL_465548 to 465550, EPI_ISL_465552 to 465572, EPI_ISL_465574 to 465581, EPI_ISL_465583 to 465584, EPI_ISL_465586 to 465595, EPI_ISL_465597 to 465611, EPI_ISL_465613 to 465631, EPI_ISL_465636 to 465646, EPI_ISL_465648 to 465655, EPI_ISL_465657 to 465670, EPI_ISL_465672 to 465678, EPI_ISL_465704 to 465715, EPI_ISL_465718 to 465722, EPI_ISL_465724 to 465726, EPI_ISL_465728, EPI_ISL_465730 to 465732, EPI_ISL_465734 to 465744, EPI_ISL_465746 to 465761, EPI_ISL_465763 to 465769, EPI_ISL_465771 to 465775, EPI_ISL_465777, EPI_ISL_465780 to 465783, EPI_ISL_465785 to 465798, EPI_ISL_465800 to 465809, EPI_ISL_465811 to 465814, EPI_ISL_465816 to 465826, EPI_ISL_465828 to 465838, EPI_ISL_465840 to 465844, EPI_ISL_465846 to 465847, EPI_ISL_465849 to 465854, EPI_ISL_465856 to 465861, EPI_ISL_465863 to 465888, EPI_ISL_465890 to 465893, EPI_ISL_465895 to 465896, EPI_ISL_465898 to 465904, EPI_ISL_465906 to 465908, EPI_ISL_465910 to 465915, EPI_ISL_465917, EPI_ISL_465919 to 465923, EPI_ISL_465928 to 465929, EPI_ISL_465933 to 465937, EPI_ISL_465939 to 465942, EPI_ISL_465944 to 465947, EPI_ISL_465949 to 465957, EPI_ISL_465959 to 465964, EPI_ISL_465966 to 465969, EPI_ISL_465971 to 465974, EPI_ISL_465976 to 465983, EPI_ISL_465985 to 465995, EPI_ISL_465997 to 466012, EPI_ISL_466014 to 466021, EPI_ISL_466024 to 466031, EPI_ISL_466033 to 466034, EPI_ISL_466036 to 466038, EPI_ISL_466040 to 466104, EPI_ISL_466106 to 466108, EPI_ISL_466110 to 466122, EPI_ISL_466124 to 466182, EPI_ISL_466185 to 466201, EPI_ISL_466203 to 466209, EPI_ISL_466211 to 466226, EPI_ISL_466228 to 466235, EPI_ISL_466237 to 466267, EPI_ISL_466269 to 466283, EPI_ISL_466285 to 466294, EPI_ISL_466296, EPI_ISL_466298 to 466300, EPI_ISL_466302, EPI_ISL_466305, EPI_ISL_466307 to 466312, EPI_ISL_466314 to 466322, EPI_ISL_466324 to 466331, EPI_ISL_466333 to 466334, EPI_ISL_466336 to 466337, EPI_ISL_466339 to 466342, EPI_ISL_466344 to 466348, EPI_ISL_466350 to 466352, EPI_ISL_466355 to 466370, EPI_ISL_466372, EPI_ISL_466374, EPI_ISL_466376 to 466385, EPI_ISL_466388 to 466389, EPI_ISL_466391 to 466395, EPI_ISL_466397 to 466405, EPI_ISL_466408 to 466410, EPI_ISL_466412 to 466415, EPI_ISL_466417, EPI_ISL_466420, EPI_ISL_466422 to 466432, EPI_ISL_466434, EPI_ISL_466436 to 466437, EPI_ISL_466441, EPI_ISL_466444 to 466445, EPI_ISL_466447, EPI_ISL_466450 to 466453, EPI_ISL_466456 to 466457, EPI_ISL_466461 to 466476, EPI_ISL_466478 to 466481, EPI_ISL_466485 to 466488, EPI_ISL_466491 to 466495, EPI_ISL_466497 to 466499, EPI_ISL_466501 to 466502, EPI_ISL_466505 to 466508, EPI_ISL_466511 to 466514, EPI_ISL_466516 to 466517, EPI_ISL_466519 to 466525, EPI_ISL_466527 to 466548, EPI_ISL_466550 to 466565, EPI_ISL_466567 to 466574, EPI_ISL_466577, EPI_ISL_466579 to 466580, EPI_ISL_466583 to 466585, EPI_ISL_466588 to 466603, EPI_ISL_466605 to 466613, EPI_ISL_466615 to 466625, EPI_ISL_471510, EPI_ISL_471512 to 471518, EPI_ISL_471520 to 471524, EPI_ISL_471527, EPI_ISL_485604 to 485608, EPI_ISL_485610 to 485611, EPI_ISL_490224 to 490228, EPI_ISL_490230 to 490248, EPI_ISL_490250 to 490254, EPI_ISL_491710, EPI_ISL_491712 to 491722, EPI_ISL_491724 to 491735, EPI_ISL_493002 to 493003, EPI_ISL_500707, EPI_ISL_500709 to 500712, EPI_ISL_500715, EPI_ISL_500952 to 500953, EPI_ISL_514343 to 514353, EPI_ISL_523951, EPI_ISL_526537 to 526539, EPI_ISL_526545 to 526546, EPI_ISL_526549 to 526550, EPI_ISL_526755 to 526757, EPI_ISL_526759 to 526760, EPI_ISL_526762 to 526773, EPI_ISL_527329, EPI_ISL_527331, EPI_ISL_527333 to 527334, EPI_ISL_527336 to 527340, EPI_ISL_527343 to 527349, EPI_ISL_527351 to 527358, EPI_ISL_528388 to 528389, EPI_ISL_528392 to 528393, EPI_ISL_528395 to 528398, EPI_ISL_528400 to 528401, EPI_ISL_528403, EPI_ISL_528405 to 528409, EPI_ISL_528411, EPI_ISL_528413, EPI_ISL_528415 to 528418, EPI_ISL_528431 to 528435, EPI_ISL_528438, EPI_ISL_528930, EPI_ISL_528932, EPI_ISL_528950, EPI_ISL_529027, EPI_ISL_534717 to 534719, EPI_ISL_534722 to 534729, EPI_ISL_539880, EPI_ISL_566045, EPI_ISL_566047 to 566067, EPI_ISL_566069 to 566074, EPI_ISL_566076, EPI_ISL_574697 to 574704, EPI_ISL_583968 to 583981, EPI_ISL_583983, EPI_ISL_583985 to 583993, EPI_ISL_584053 to 584062, EPI_ISL_584064 to 584066, EPI_ISL_584068, EPI_ISL_593804 to 593805, EPI_ISL_593810, EPI_ISL_593824 to 593825, EPI_ISL_593829, EPI_ISL_593834, EPI_ISL_593837, EPI_ISL_593843, EPI_ISL_593846, EPI_ISL_593848, EPI_ISL_593850, EPI_ISL_593998, EPI_ISL_603188 to 603215, EPI_ISL_609827 to 609829, EPI_ISL_609831, EPI_ISL_609833, EPI_ISL_609835 to 609868, EPI_ISL_609870 to 609871, | | | |

| | | | |
|---|---|---|---|
| EPI_ISL_609875 to 609876, EPI_ISL_609879 to 609880, EPI_ISL_609882 to 609896, EPI_ISL_609898 to 609909, EPI_ISL_613565, EPI_ISL_613571 to 613582, EPI_ISL_613584 to 613586, EPI_ISL_613588 to 613629, EPI_ISL_613631 to 613635, EPI_ISL_622833, EPI_ISL_622835 to 622842, EPI_ISL_622844 to 622857, EPI_ISL_622859 to 622867, EPI_ISL_622869 to 622871, EPI_ISL_622873 to 622878, EPI_ISL_622880 to 622881, EPI_ISL_622883 to 622893, EPI_ISL_632796, EPI_ISL_636688 to 636697, EPI_ISL_636699 to 636703, EPI_ISL_636705 to 636715, EPI_ISL_636719 to 636723, EPI_ISL_636725 to 636727, EPI_ISL_636729 to 636730, EPI_ISL_636732 to 636736, EPI_ISL_636974, EPI_ISL_637097 | | | |
| see above | Respiratory Virus Unit, Microbiology Services Colindale, Public Health England | Respiratory Virus Unit, Microbiology Services Colindale, Public Health England | Angie Lackenby; Joanna Ellis; Jonathan Hubb; Kirstin Edwards; Leena Bhaw; Maria Zambon; Monica Galiano; Omolola Akinbami; PHE Covid Sequencing Team; Richard Myers; Shahjahan Miah; Steven Platt; Tiina Talts; Tina Talts |
| EPI_ISL_700624, EPI_ISL_700637, EPI_ISL_700651, EPI_ISL_700658, EPI_ISL_700665, EPI_ISL_700686 to 700692, EPI_ISL_700694 to 700698, EPI_ISL_707703 to 707705, EPI_ISL_709958, EPI_ISL_722967 to 723020, EPI_ISL_723022 to 723044, EPI_ISL_728343, EPI_ISL_733581, EPI_ISL_733583 to 733584, EPI_ISL_733586, EPI_ISL_733588 to 733593, EPI_ISL_747490 to 747519, EPI_ISL_754242 to 754265, EPI_ISL_754267 to 754288, EPI_ISL_754291 to 754299, EPI_ISL_754306, EPI_ISL_754309 to 754342, EPI_ISL_754344 to 754357, EPI_ISL_754359 to 754382, EPI_ISL_754384 to 754387, EPI_ISL_756136 to 756163, EPI_ISL_762996, EPI_ISL_766059, EPI_ISL_766062 to 766065, EPI_ISL_766069, EPI_ISL_766072 to 766075, EPI_ISL_766077 to 766090, EPI_ISL_766092 to 766093, EPI_ISL_766096 to 766098, EPI_ISL_766100 to 766101, EPI_ISL_766104 to 766108, EPI_ISL_766110 to 766111, EPI_ISL_766113 to 766131, EPI_ISL_766133 to 766196, EPI_ISL_766198 to 766218, EPI_ISL_766220 to 766221, EPI_ISL_766223 to 766232, EPI_ISL_766235 to 766239, EPI_ISL_766243, EPI_ISL_766245 to 766254, EPI_ISL_766257 to 766261, EPI_ISL_766263 to 766264, EPI_ISL_766266 to 766275, EPI_ISL_766277, EPI_ISL_766279 to 766280, EPI_ISL_766282 to 766283, EPI_ISL_766285 to 766317, EPI_ISL_766395 to 766424, EPI_ISL_766426 to 766428, EPI_ISL_766430 to 766451, EPI_ISL_766453 to 766457, EPI_ISL_766459, EPI_ISL_766461, EPI_ISL_766469 to 766471, EPI_ISL_766474 to 766493, EPI_ISL_766495 to 766508, EPI_ISL_766510 to 766522, EPI_ISL_766553 to 766555, EPI_ISL_766558 to 766567, EPI_ISL_766842, EPI_ISL_766853, EPI_ISL_766859, EPI_ISL_768800 to 768814, EPI_ISL_769865 to 769866, EPI_ISL_769868 to 769869, EPI_ISL_769874 to 769876, EPI_ISL_769878, EPI_ISL_770441, EPI_ISL_770462 to 770467, EPI_ISL_770469, EPI_ISL_791183 to 791191, EPI_ISL_791193, EPI_ISL_791204 to 791208, EPI_ISL_791210 to 791221, EPI_ISL_804214 to 804215, EPI_ISL_804218 to 804223, EPI_ISL_804226 to 804248, EPI_ISL_804250 to 804258, EPI_ISL_804260 to 804275, EPI_ISL_804277 to 804329, EPI_ISL_804331 to 804358, EPI_ISL_804360 to 804368, EPI_ISL_804812, EPI_ISL_811040 to 811056, EPI_ISL_811058 to 811062, EPI_ISL_811065 to 811071, EPI_ISL_811073 to 811102, EPI_ISL_811104 to 811105, EPI_ISL_811115, EPI_ISL_811117 to 811120, EPI_ISL_811122, EPI_ISL_825492 to 825514, EPI_ISL_825516 to 825549, EPI_ISL_825553 to 825557, EPI_ISL_825559 to 825572, EPI_ISL_825576 to 825597, EPI_ISL_825599 to 825613, EPI_ISL_825615, EPI_ISL_826277 to 826281, EPI_ISL_836979 to 836980, EPI_ISL_836982 to 837005, EPI_ISL_837007 to 837010, EPI_ISL_837012 to 837031, EPI_ISL_837033 to 837048, EPI_ISL_837050 to 837051, EPI_ISL_837061 to 837062, EPI_ISL_837068, EPI_ISL_837070 to 837072, EPI_ISL_837074 to 837076, EPI_ISL_837078 to 837086, EPI_ISL_837088 to 837093, EPI_ISL_837095 to 837134, EPI_ISL_837136 to 837171, EPI_ISL_837184 to 837194, EPI_ISL_837198 to 837229, EPI_ISL_837231 to 837238, EPI_ISL_846588 to 846590, EPI_ISL_846593, EPI_ISL_855576, EPI_ISL_855583, EPI_ISL_855585 to 855587, EPI_ISL_860635 to 860650, EPI_ISL_860653 to 860658, EPI_ISL_860660 to 860661, EPI_ISL_860664 to 860665, EPI_ISL_860685, EPI_ISL_860687, EPI_ISL_860689 to 860690, EPI_ISL_860782, EPI_ISL_862190, EPI_ISL_862251 | | | |
| see above | Respiratory Virus Unit, National Infection Service, Public Health England | COVID-19 Genomics UK (COG-UK) Consortium | PHE Covid Sequencing Team |
| EPI_ISL_639740 | Respiratory virus Laboratory, Chinese Academy of Medical Science | Respiratory virus Laboratory, Chinese Academy of Medical Science | Chen; Li, J.; Y. and Liu, L.; Zhen, H. |
| EPI_ISL_803894 | Robert Koch Institut | Robert Koch Institut | Alexander Dalpke; Andreas Nitsche; Annika Brinkmann; Dirk Lindemann; Dominique Seifert; Janine Michel; Kristina Hochauf-Stange; Lars Schaade; Leo Büttner; Livia Schrick; Steven Uddin |
| EPI_ISL_803890 | Robert Koch Institute | Robert Koch Institute | Andreas Nitsche; Annika Brinkmann; Dominique Seifert; Janine Michel; Lars Schaade; Livia Schrick; Merle Corty; Steven Uddin |
| EPI_ISL_481253, EPI_ISL_481256, EPI_ISL_481262, EPI_ISL_481264 | Robert Koch Institute, National Reference center for Influenza, Berlin, Germany | Robert Koch Institute, Bioinformatics MF1, Berlin, Germany | Andrea Thuermer; Marianne Wedde; Max v. Kleist; Oliver Drechsel; Ralf Duerrwald; Rene Kmiecinski; Stephan Fuchs; Thorsten Wolff |
| EPI_ISL_483139 to 483141, EPI_ISL_483143 to 483155, EPI_ISL_483157 | Robert Koch Institute, ZBS1 Highly Pathogenic Viruses, Berlin, Germany | Robert Koch Institute, Bioinformatics MF1, Berlin, Germany | Andrea Thuermer; Andreas Nitsche; Janine Michel; Max v. Kleist; Oliver Drechsel; Rene Kmiecinski; Stephan Fuchs |
| EPI_ISL_825159 to 825164, EPI_ISL_825166 to 825169, EPI_ISL_825171 to 825172 | Robert Koch Institute, ZBS1 Highly Pathogenic Viruses, Berlin, Germany | Robert Koch Institute, ZBS1 Highly Pathogenic Viruses & Bioinformatics MF1, Berlin, Germany | Alexander Dalpke; Andreas Nitsche; Annika Brinkmann; Dirk Lindemann; Dominique Seifert; Janine Michel; Kristina Hochauf-Stange; Lars Schaade; Leo Büttner; Livia Schrick; Steven Uddin |
| EPI_ISL_492862 | Royal Free Hospital / Health Services Laboratories | Wellcome Sanger Institute for the COVID-19 Genomics UK (COG-UK) Consortium | Cate Goodlad; Cordelia Langford; David K. Jackson; Dianne Irish; Dominic Kwiatkowski; Ewan Harrison; Ian Johnston; Jenny Cross; John Sillitoe on behalf of the Wellcome Sanger Institute COVID-19 Surveillance Team; Judith Heaney and Alex Alderton; Roberto Amato; Sonia Goncalves; Tabitha Mahungu; Tanzina Haque |
| EPI_ISL_492837, EPI_ISL_492840, EPI_ISL_492843 to 492847, EPI_ISL_492850 to 492852, EPI_ISL_492856, EPI_ISL_492863 to 492865, EPI_ISL_492868 to 492869, EPI_ISL_492872, EPI_ISL_492874, EPI_ISL_492876 to 492877, EPI_ISL_492879 to 492880, EPI_ISL_492882, EPI_ISL_492885, EPI_ISL_492887 to 492888, EPI_ISL_492890, EPI_ISL_492894, EPI_ISL_492902, EPI_ISL_492905 to 492906, EPI_ISL_492911, EPI_ISL_492914 | | | |
| see above | Royal Free Hospital / Health Services Laboratories | Wellcome Sanger Institute for the COVID-19 Genomics UK (COG-UK) consortium | Cate Goodlad; Cordelia Langford; David K. Jackson; Dianne Irish; Dominic Kwiatkowski; Ewan Harrison; Ian Johnston; Jenny Cross; John Sillitoe on behalf of the Wellcome Sanger Institute COVID-19 Surveillance Team (http://www.sanger.ac.uk/covid-team); Judith Heaney and Alex Alderton; Roberto Amato; Sonia Goncalves; Tabitha Mahungu; Tanzina Haque |
| EPI_ISL_421275, EPI_ISL_454732, EPI_ISL_470896 | Russian State Collection of Viruses | Pathogenic Microorganisms Variability Laboratory | Alexander Gintsburg; Alexey Mazus; Alexey Shchetinin; Andrey Botikov; Daria Grousova; Denis Logunov; Denis Protsenko; Ekaterina Aksenova; Elena Shidlovskaya; Inna Dolzhikova; Maria Nikiforova; Marina Kunda; Nadezhda Kuznetsova; Natalia Ryzhova; Olga Voronina; Vladimir Gushchin |
| EPI_ISL_420051 | Résidence Eleusis | National Reference Center for Viruses of Respiratory Infections, Institut Pasteur, Paris | Angela Brisebarre; Etienne Simon-Lorière; Flora Donati; Marion Barbet; Maud Vanpeene; Mélanie Albert; Méline Bizard; Sylvie Behillil; Sylvie van der Werf; Vincent Enouf |
| EPI_ISL_443304 | Résidence Esterel | National Reference Center for Viruses of Respiratory Infections, Institut Pasteur, Paris | Angela Brisebarre; Etienne Simon-Lorière; Flora Donati; Marion Barbet; Maud Vanpeene; Mélanie Albert; Méline Bizard; Sylvie Behillil; Sylvie van der Werf; Vincent Enouf |
| EPI_ISL_443303 | Résidence Les Marines | National Reference Center for Viruses of Respiratory Infections, Institut Pasteur, Paris | Angela Brisebarre; Etienne Simon-Lorière; Flora Donati; Marion Barbet; Maud Vanpeene; Mélanie Albert; Méline Bizard; Sylvie Behillil; Sylvie van der Werf; Vincent Enouf |
| EPI_ISL_443258 to 443259 | Résidence Ornano | National Reference Center for Viruses of Respiratory Infections, Institut Pasteur, Paris | Angela Brisebarre; Etienne Simon-Lorière; Flora Donati; Marion Barbet; Maud Vanpeene; Mélanie Albert; Méline Bizard; Sylvie Behillil; Sylvie van der Werf; Vincent Enouf |
| EPI_ISL_420046 to 420047 | Résidence Villa Caroline | National Reference Center for Viruses of Respiratory Infections, Institut Pasteur, Paris | Angela Brisebarre; Etienne Simon-Lorière; Flora Donati; Marion Barbet; Maud Vanpeene; Mélanie Albert; Méline Bizard; Sylvie Behillil; Sylvie van der Werf; Vincent Enouf |
| EPI_ISL_420054 | Résidence de maintenon | National Reference Center for Viruses of Respiratory Infections, Institut Pasteur, Paris | Angela Brisebarre; Etienne Simon-Lorière; Flora Donati; Marion Barbet; Maud Vanpeene; Mélanie Albert; Méline Bizard; Sylvie Behillil; Sylvie van der Werf; Vincent Enouf |
| EPI_ISL_420052 | Résidence les Marines | National Reference Center for Viruses of Respiratory Infections, Institut Pasteur, Paris | Angela Brisebarre; Etienne Simon-Lorière; Flora Donati; Marion Barbet; Maud Vanpeene; Mélanie Albert; Méline Bizard; Sylvie Behillil; Sylvie van der Werf; Vincent Enouf |
| EPI_ISL_882635 | SC (UCO) Igiene e Sanità Pubblica (funzione integrata con SC Microbiologia e Virologia) e Laboratory of Molecular Virology of the International Centre for Genetic Engineering and Biotechnology (ICGEB) | ARGO Laboratorio Genomica ed EpigenomicA | D'Agaro P; Dal Monego S; Degasperi M; Licastro D; Marcello A |
| EPI_ISL_804038, EPI_ISL_804040 to 804042, EPI_ISL_804044, EPI_ISL_804048, EPI_ISL_804050 to 804054, EPI_ISL_833435 to 833455, EPI_ISL_882636 | | | |
| see above | SC (UCO) Igiene e Sanità Pubblica (funzione integrata con SC Microbiologia e Virologia) e Laboratory of Molecular Virology of the International Centre for Genetic Engineering and Biotechnology (ICGEB) | ARGO Laboratorio Genomica ed Epigenomica | D'Agaro P; Dal Monego S; Degasperi M; Licastro D; Marcello A |
| EPI_ISL_435149, EPI_ISL_435153 to 435155, EPI_ISL_436726 to 436729 | SERVIZIO DI IGIENE E SANITÀ PUBBLICA ASL Teramo | Istituto Zooprofilattico Sperimentale dell'Abruzzo e Molise "G.Caporale" | Ancora M; Cammà C; Curini V; Di Domenico M; Di Pasquale A; Lorusso A; Mangone I; Marcacci M; Puglia I; Rinaldi A; Savini G |
| EPI_ISL_806804 to 806805 | SIESP CHIETI - DISTRETTO SANITARIO CHIETI | Istituto Zooprofilattico Sperimentale dell'Abruzzo e Molise "G. Caporale" | Ancora M; Calistri P; Cammà C; Curini V; Di Domenico M; Di Pasquale A; Lorusso A; Mangone I; Marcacci M; Puglia I; Rinaldi A; Savini G |
| EPI_ISL_832821, EPI_ISL_833232 | SIESP CHIETI - DRIVE IN CHIETI | Istitituto Zooprofilattico Sperimentale dell'Abruzzo e Molise "G.Caporale" | Ancora M; Calistri P; Cammà C; Curini V; Di Domenico M; Di Pasquale A; Lorusso A; Mangone I; Marcacci M; Puglia I; Rinaldi A; Savini G. |
| EPI_ISL_806733, EPI_ISL_833044, EPI_ISL_833090 to 833096, EPI_ISL_833317 to 833318, EPI_ISL_833323 | SIESP CHIETI - DRIVE IN CHIETI | Istituto Zooprofilattico Sperimentale dell'Abruzzo e Molise "G. Caporale" | Ancora M; Calistri P; Cammà C; Curini V; Delli Compagni E; Di Domenico M; Di Pasquale A; Lorusso A; Mangone I; Marcacci M; Puglia I; Rinaldi A; Savini G |
| EPI_ISL_806800 to 806803, EPI_ISL_833271, EPI_ISL_833316 | SIESP CHIETI - DRIVE IN GISSI | Istituto Zooprofilattico Sperimentale dell'Abruzzo e Molise "G. Caporale" | Ancora M; Calistri P; Cammà C; Curini V; Di Domenico M; Di Pasquale A; Lorusso A; Mangone I; Marcacci M; Puglia I; Rinaldi A; Savini G |

| | | | |
|---|---|---|---|
| EPI_ISL_832815 to 832816, EPI_ISL_832822, EPI_ISL_833231, EPI_ISL_833242 | SIESP CHIETI - DRIVE IN LANCIANO | Istitituto Zooprofilattico Sperimentale dell'Abruzzo e Molise "G.Caporale" | Ancora M; Calistri P; Cammà C; Curini V; Di Domenico M; Di Pasquale A; Lorusso A; Mangone I; Marcacci M; Puglia I; Rinaldi A; Savini G. |
| EPI_ISL_738044, EPI_ISL_806734, EPI_ISL_806775 to 806776, EPI_ISL_806799, EPI_ISL_833049, EPI_ISL_833097 to 833098, EPI_ISL_833292 to 833294, EPI_ISL_833298, EPI_ISL_833300, EPI_ISL_833313 to 833315 | | | |
| see above | SIESP CHIETI - DRIVE IN LANCIANO | Istituto Zooprofilattico Sperimentale dell'Abruzzo e Molise "G. Caporale" | Ancora M; Calistri P; Cammà C; Curini V; Delli Compagni E; Di Domenico M; Di Pasquale A; Lorusso A; Mangone I; Marcacci M; Puglia I; Rinaldi A; Savini G |
| EPI_ISL_738046 to 738048, EPI_ISL_806762, EPI_ISL_806791 to 806798, EPI_ISL_833045, EPI_ISL_833099 to 833100, EPI_ISL_833250, EPI_ISL_833270, EPI_ISL_833272 to 833275, EPI_ISL_833284 to 833287, EPI_ISL_833306 to 833310, EPI_ISL_833328 to 833330 | | | |
| see above | SIESP CHIETI - DRIVE IN ORTONA | Istituto Zooprofilattico Sperimentale dell'Abruzzo e Molise "G. Caporale" | Ancora M; Calistri P; Cammà C; Curini V; Delli Compagni E; Di Domenico M; Di Pasquale A; Lorusso A; Mangone I; Marcacci M; Puglia I; Rinaldi A; Savini G |
| EPI_ISL_833278 to 833280, EPI_ISL_833311 to 833312 | SIESP CHIETI- DRIVE IN CHIETI | Istituto Zooprofilattico Sperimentale dell'Abruzzo e Molise "G. Caporale" | Ancora M; Calistri P; Cammà C; Curini V; Di Domenico M; Di Pasquale A; Lorusso A; Mangone I; Marcacci M; Puglia I; Rinaldi A; Savini G |
| EPI_ISL_833233 to 833238, EPI_ISL_833241, EPI_ISL_833243 to 833247, EPI_ISL_849634, EPI_ISL_849637 to 849646 | SIESP DIPARTIMENTO DI PREVENZIONE CHIETI | Istitituto Zooprofilattico Sperimentale dell'Abruzzo e Molise "G.Caporale" | Ancora M; Calistri P; Cammà C; Curini V; Di Domenico M; Di Pasquale A; Lorusso A; Mangone I; Marcacci M; Puglia I; Rinaldi A; Savini G. |
| EPI_ISL_738045, EPI_ISL_806731 to 806732, EPI_ISL_806806, EPI_ISL_833043, EPI_ISL_833101 to 833102, EPI_ISL_833262 to 833264, EPI_ISL_833299, EPI_ISL_833321, EPI_ISL_833324, EPI_ISL_833326 | | | |
| see above | SIESP DIPARTIMENTO DI PREVENZIONE CHIETI | Istituto Zooprofilattico Sperimentale dell'Abruzzo e Molise "G. Caporale" | Ancora M; Calistri P; Cammà C; Curini V; Delli Compagni E; Di Domenico M; Di Pasquale A; Lorusso A; Mangone I; Marcacci M; Puglia I; Rinaldi A; Savini G |
| EPI_ISL_806790, EPI_ISL_833291 | SIESP DIPARTIMENTO DI PREVENZIONE DELL'AQUILA | Istituto Zooprofilattico Sperimentale dell'Abruzzo e Molise "G. Caporale" | Ancora M; Calistri P; Cammà C; Curini V; Di Domenico M; Di Pasquale A; Lorusso A; Mangone I; Marcacci M; Puglia I; Rinaldi A; Savini G |
| EPI_ISL_832819 | SIESP DIPARTIMENTO DI PREVENZIONE SULMONA | Istitituto Zooprofilattico Sperimentale dell'Abruzzo e Molise "G.Caporale" | Ancora M; Calistri P; Cammà C; Curini V; Di Domenico M; Di Pasquale A; Lorusso A; Mangone I; Marcacci M; Puglia I; Rinaldi A; Savini G. |
| EPI_ISL_806749 to 806757, EPI_ISL_833319 to 833320, EPI_ISL_833322 | SIESP DIPARTIMENTO DI PREVENZIONE SULMONA | Istituto Zooprofilattico Sperimentale dell'Abruzzo e Molise "G. Caporale" | Ancora M; Calistri P; Cammà C; Curini V; Di Domenico M; Di Pasquale A; Lorusso A; Mangone I; Marcacci M; Puglia I; Rinaldi A; Savini G |
| EPI_ISL_832817, EPI_ISL_833239 to 833240 | SIESP DIPARTIMENTO DI PREVENZIONE TERAMO | Istitituto Zooprofilattico Sperimentale dell'Abruzzo e Molise "G.Caporale" | Ancora M; Calistri P; Cammà C; Curini V; Di Domenico M; Di Pasquale A; Lorusso A; Mangone I; Marcacci M; Puglia I; Rinaldi A; Savini G. |
| EPI_ISL_806735 to 806744, EPI_ISL_806746 to 806748, EPI_ISL_806766 to 806772, EPI_ISL_806777 to 806780, EPI_ISL_806786 to 806788, EPI_ISL_833258 to 833259, EPI_ISL_833281 to 833282, EPI_ISL_833295 to 833296, EPI_ISL_833302 to 833305 | | | |
| see above | SIESP DIPARTIMENTO DI PREVENZIONE TERAMO | Istituto Zooprofilattico Sperimentale dell'Abruzzo e Molise "G. Caporale" | Ancora M; Calistri P; Cammà C; Curini V; Di Domenico M; Di Pasquale A; Lorusso A; Mangone I; Marcacci M; Puglia I; Rinaldi A; Savini G |
| EPI_ISL_833103 to 833118 | SIESP DIPARTIMENTO DI PREVENZIONE TERAMO C.DA CASALENA | Istituto Zooprofilattico Sperimentale dell'Abruzzo e Molise "G. Caporale" | Ancora M; Calistri P; Cammà C; Curini V; Delli Compagni E; Di Domenico M; Di Pasquale A; Lorusso A; Mangone I; Marcacci M; Puglia I; Rinaldi A; Savini G |
| EPI_ISL_833119 | SIESP L'AQUILA | Istituto Zooprofilattico Sperimentale dell'Abruzzo e Molise "G. Caporale" | Ancora M; Calistri P; Cammà C; Curini V; Delli Compagni E; Di Domenico M; Di Pasquale A; Lorusso A; Mangone I; Marcacci M; Puglia I; Rinaldi A; Savini G |
| EPI_ISL_833120 to 833122 | SIESP SULMONA | Istituto Zooprofilattico Sperimentale dell'Abruzzo e Molise "G. Caporale" | Ancora M; Calistri P; Cammà C; Curini V; Delli Compagni E; Di Domenico M; Di Pasquale A; Lorusso A; Mangone I; Marcacci M; Puglia I; Rinaldi A; Savini G |
| EPI_ISL_849647 to 849649 | SIESP- DRIVE IN CHIETI | Istitituto Zooprofilattico Sperimentale dell'Abruzzo e Molise "G.Caporale" | Ancora M; Cammà C; Curini V; Di Domenico M; Di Pasquale A; Lorusso A; Mangone I; Marcacci M; Puglia I; Rinaldi A; Savini G. |
| EPI_ISL_794746, EPI_ISL_794748 | SISP ASL BA AREA SUD | Istituto Zooprofilattico Sperimentale della Puglia e della Basilicata | Bianco A.; Capozzi L.; Cipolletta D.; Del Sambro L.; Galante D.; Manzulli V; Pace L.; Parisi A.; Rondinone V. |
| EPI_ISL_733478 to 733481 | SOGAZ PROFMEDICINA LLC | WHO National Influenza Centre Russian Federation | Andrey Komissarov; Anna Ivanova; Artem Fadeev; Daria Danilenko; Dmitry Bazhenov; Dmitry Lioznov; Elena Nabieva; Georgii Bazykin; Ksenia Safina; Kseniya Komissarova |
| EPI_ISL_457719 to 457720, EPI_ISL_457722 to 457723, EPI_ISL_457725, EPI_ISL_457727, EPI_ISL_457729, EPI_ISL_457731, EPI_ISL_457734 to 457735, EPI_ISL_457737 to 457740, EPI_ISL_516627 | | | |
| see above | SYNLAB Eesti OU | Charite Universitatsmedizin Berlin, Institute of Virology | Barbara Muhlemann; Christian Drosten; Jorn Beheim-Schwarzbach; Julia Schneider; Paul Naaber; Talitha Veith; Terry Jones; Victor M Corman |
| EPI_ISL_420540 | SYNLAB Eesti OÜ | Charite Universitatsmedizin Berlin, Institute of Virology | Barbara Mühlemann; Christian Drosten; Jorn Beheim-Schwarzbach; Julia Schneider; Paul Naaber; Talitha Veith; Terry Jones; Victor M Corman |
| EPI_ISL_542278 to 542399 | San Matteo Hospital Pavia | Dep. Of Oncology and Hemato-Oncology University of Milan | Antonio Piralla; Carlo Federico Perno; Chiara Vismara; Claudia Alteri; Elisa Matarazzo; Fausto Baldanti; Federica Giardina; Federica Novazzi; Luna Colagrossi; Maria Antonello; Massimo Puoti; Monica Tallarita; Oscar Massimiliano Epis; Roberto Fumagalli; Silvia Renica; Stefano Gaiarsa; Valentino Costabile; Valeria Cento |
| EPI_ISL_434654, EPI_ISL_445222, EPI_ISL_445226, EPI_ISL_450808, EPI_ISL_450810, EPI_ISL_452231 to 452232 | Saroledens Familjelakare | The Public Health Agency of Sweden | Anna Risberg; Anna-Malin Linde; Karin Tegmark-Wisell; Katarina Jarbur; Maria Lind Karlberg; Mia Brytting; Olov Svartstrom; Oskar Karlsson Lindsjo; Theresa Enkirch |
| EPI_ISL_455110 | Scania Halsocenter, B288 | The Public Health Agency of Sweden | Anna Risberg; Anna-Malin Linde; Christina Lergin; Karin Tegmark-Wisell; Maria Lind Karlberg; Mia Brytting; Olov Svartstrom; Oskar Karlsson Lindsjo; Theresa Enkirch |
| EPI_ISL_661179 to 661189, EPI_ISL_661191 to 661200, EPI_ISL_676574 to 676603 | Scientific Veterinary Institute Novi Sad | Veterinary Specialized Institute "Kraljevo", Serbia | Afonso, C.; Banovic Djeri, B.; Jankovic, M.; Jovanovic, T.; Knezevic, A.; Petrovic, T.; Sekler, M.; Tesovic, B.; Vidanovic, D.; Volkening, J. |
| EPI_ISL_832128 to 832129 | Sentinelles IDF | National Reference Center for Viruses of Respiratory Infections, Institut Pasteur, Paris | Angela Brisebarre; Camille Capel; Claire Watermez; Etienne Simon-Lorière; Marion Barbet; Maud Vanpeene; Méline Bizard; Patricia Lefebure; Sylvie Behillil; Sylvie van der Werf; Vincent Enouf |
| EPI_ISL_832167 to 832168 | Sentinelles Province | National Reference Center for Viruses of Respiratory Infections, Institut Pasteur, Paris | Angela Brisebarre; Camille Capel; Denis Calvet; Etienne Simon-Lorière; Jean-Michel Monnier; Marion Barbet; Maud Vanpeene; Méline Bizard; Sylvie Behillil; Sylvie van der Werf; Vincent Enouf |
| EPI_ISL_420038, EPI_ISL_420045, EPI_ISL_420055, EPI_ISL_421514 | Sentinelles network | National Reference Center for Viruses of Respiratory Infections, Institut Pasteur, Paris | Angela Brisebarre; Etienne Simon-Lorière; Flora Donati; Marion Barbet; Maud Vanpeene; Mélanie Albert; Méline Bizard; Sylvie Behillil; Sylvie van der Werf; Vincent Enouf |
| EPI_ISL_593934 | Sentinelles, Chanteloup-En-Brie | National Reference Center for Viruses of Respiratory Infections, Institut Pasteur, Paris | Etienne Simon-Lorière; Fabiana Gambaro; Maud Vanpeene; Sylvie Behillil; Sylvie van der Werf; Vincent Enouf |
| EPI_ISL_593936 | Sentinelles, Fondettes | National Reference Center for Viruses of Respiratory Infections, Institut Pasteur, Paris | Etienne Simon-Lorière; Fabiana Gambaro; Maud Vanpeene; Sylvie Behillil; Sylvie van der Werf; Vincent Enouf |
| EPI_ISL_593902 | Sentinelles, Limay | National Reference Center for Viruses of Respiratory Infections, Institut Pasteur, Paris | Etienne Simon-Lorière; Fabiana Gambaro; Maud Vanpeene; Sylvie Behillil; Sylvie van der Werf; Vincent Enouf |
| EPI_ISL_593935 | Sentinelles, Plessis-Trevise | National Reference Center for Viruses of Respiratory Infections, Institut Pasteur, Paris | Etienne Simon-Lorière; Fabiana Gambaro; Maud Vanpeene; Sylvie Behillil; Sylvie van der Werf; Vincent Enouf |
| EPI_ISL_420048, EPI_ISL_420058 to 420060, EPI_ISL_420062, EPI_ISL_420064, EPI_ISL_421501, EPI_ISL_421504 to 421506, EPI_ISL_421512, EPI_ISL_428347, EPI_ISL_428349 | | | |
| see above | Service de Biologie Médicale - BP 125 | National Reference Center for Viruses of Respiratory Infections, Institut Pasteur, Paris | Angela Brisebarre; Christine Lambert; Etienne Simon-Lorière; Flora Donati; Marion Barbet; Maud Vanpeene; Mélanie Albert; Méline Bizard; Sylvie Behillil; Sylvie van der Werf; Vincent Enouf |

| | | | |
|---|---|---|---|
| EPI_ISL_420042, EPI_ISL_421513 | Service de Biologie clinique | National Reference Center for Viruses of Respiratory Infections, Institut Pasteur, Paris | Angela Brisebarre; Christine Lambert; Etienne Simon-Lorière; Flora Donati; Marion Barbet; Maud Vanpeene; Mélanie Albert; Méline Bizard; Sylvie Behillil; Sylvie van der Werf; Vincent Enouf |
| EPI_ISL_469283 to 469284 | Service de Virologie Hôpital Saint-Louis | Laboratory Cell Biology of Viral Infection-INSERM unit 944 | Ali Amara; Constance Delaugerre; Laurent Meertens; Lucie Bonnet-Madin; Maud SALMONA; Séverine Mercier-Delarue |
| EPI_ISL_469282 | Service de Virologie Hôpital Saint-Louis | Laboratory of Cell Biology of viral infection, Unit INSERM-U944 | Ali Amara; Constance Delaugerre; Laurent Meertens; Lucie Bonnet-Madin |
| EPI_ISL_418232 to 418233 | Service des Urgences | National Reference Center for Viruses of Respiratory Infections, Institut Pasteur, Paris | Angela Brisebarre; Boubkeur; Etienne Simon-Lorière; Flora Donati; Marion Barbet; Maud Vanpeene; Mélanie Albert; Méline Bizard; Sylvie Behillil; Sylvie van der Werf; Vincent Enouf |
| EPI_ISL_414598 | Servicio Microbiologia, Hospital Clinico Universitario, Valencia | Sequencing and Bioinformatics Service and Molecular Epidemiology Research Group. FISABIO-Public Health. | David Navarro; Fernando Gonzalez-Candelas; Giuseppe D'Auria; Griselda De Marco; Maria Alma Bracho; Neris Garcia-Gonzalez |
| EPI_ISL_414496 | Servicio Microbiologia. Hospital Clinico Universitario. Valencia. | Sequencing and Bioinformatics Service. Molecular Epidemiology Laboratory. FISABIO-Public Health | David Navarro; Fernando Gonzalez-Candelas; Giuseppe D'Auria; Griselda De Marco; María Alma Bracho; Neris Garcia-Gonzalez |
| EPI_ISL_414495 | Servicio Microbiología. Hospital Clínico Universitario. Valencia. | Sequencing and Bioinformatics Service. Molecular Epidemiology Laboratory. FISABIO-Public Health | David Navarro; Fernando Gonzalez-Candelas; Giuseppe D'Auria; Griselda De Marco; Maria Alma Bracho; Neris Garcia-Gonzalez |
| EPI_ISL_849155, EPI_ISL_856756 to 856759 | Servicio Virosis Respiratorias-Departamento Virología-INEI | Instituto Nacional Enfermedades Infecciosas C.G.Malbran | Avaro M.; Baumeister E.; Benedetti E.; Campos J.; Cisterna D.; Dattero ME; Lorenzo F.; Molina V.; Perandones C.; Poklepovich T.; Pontoriero A.; Russo M.; Tuduri E. |
| EPI_ISL_416485, EPI_ISL_417206 | Servicio de Microbiologia. Consorcio Hospital General Universitario de Valencia | Sequencing and Bioinformatics Service and Molecular Epidemiology Research Group. FISABIO-Public Health | Concepcion Gimeno; Fernando Gonzalez-Candelas; Giuseppe D'Auria; Griselda De Marco; Maria Alma Bracho; Maria Dolores Ocete; Neris Garcia-Gonzalez |
| EPI_ISL_537997 to 538002 | Servicio de Microbiologia. Hospital Arnau de Vilanova | SeqCOVID-SPAIN consortium/IBV(CSIC) | Fernando Gonzalez-Candelas and SeqCOVID-SPAIN consortium; Giuseppe D'Auria; Griselda De Marco; Inma Galán Vendrell; Ivan Ansari; Lidia Ruiz Roldan; Llúcia Martínez-Priego; Loreto Ferrús Abad; Maria Alma Bracho; Mariana Reyes-Prieto; Neris Garcia-Gonzalez; Paula Ruiz-Hueso; Sandra Carbo; Vicente Soriano Chirona; Victoria Dominguez |
| EPI_ISL_539221 to 539229, EPI_ISL_539231, EPI_ISL_539235 to 539239, EPI_ISL_539243 to 539244, EPI_ISL_539288 | Servicio de Microbiologia. Hospital Clinico Universitario de Valencia | SeqCOVID-SPAIN consortium/IBV(CSIC) | Beatriz Beamud; Concepcion Gimeno; Fernando Gonzalez-Candelas and SeqCOVID-SPAIN consortium; Giuseppe D'Auria; Griselda De Marco; Inma Galán Vendrell; Lidia Ruiz Roldan; Llúcia Martínez-Priego; Loreto Ferrús Abad; Maria Alma Bracho; Maria Dolores Ocete; Mariana Reyes-Prieto; Marta Pla Diaz; Neris Garcia-Gonzalez; Paula Ruiz-Hueso; Vicente Soriano Chirona |
| EPI_ISL_420125 to 420132, EPI_ISL_421515 to 421520, EPI_ISL_425199 to 425211, EPI_ISL_436278 to 436280, EPI_ISL_436282 to 436300, EPI_ISL_436302 to 436321, EPI_ISL_436323 to 436354, EPI_ISL_436356 to 436360, EPI_ISL_436362 to 436371, EPI_ISL_436396 to 436411, EPI_ISL_447470 to 447479, EPI_ISL_447481 to 447491, EPI_ISL_447493 to 447498, EPI_ISL_447500 to 447531 | | | |
| see above | Servicio de Microbiologia. Hospital Clinico Universitario de Valencia | Sequencing and Bioinformatics Service and Molecular Epidemiology Research Group. FISABIO-Public Health | Beatriz Beamud; Concepcion Gimeno; David Navarro; Eliseo Albert; Fernando Gonzalez-Candelas; Giuseppe 'Auria; Giuseppe D'Auria; Griselda De Marco; Inma Galán Vendrell; Ivan Ansari; Lidia Ruiz Roldan; Llúcia Martínez-Priego; Loreto Ferrús Abad; Maria Alma Bracho; Maria Dolores Ocete; Mariana Reyes-Prieto; Marta Pla Diaz; Neris Garcia-Gonzalez; Paula Ruiz-Hueso; Sandra Carbo; Vicente Soriano Chirona |
| EPI_ISL_455721 to 455730, EPI_ISL_455732 to 455734, EPI_ISL_455737 to 455739, EPI_ISL_455741 to 455744 | Servicio de Microbiologia. Hospital Clinico Universitario de Valencia | Sequencing and Bioinformatics Service and Molecular Epidemiology Research Group. FISABIO-Public Health, and SeqCOVID-Spain Consortium | David Navarro; Eliseo Albert; Fernando Gonzalez-Candelas; Giuseppe 'Auria; Griselda De Marco; Inma Galán Vendrell; Ivan Ansari; Lidia Ruiz Roldan; Llúcia Martínez-Priego; Loreto Ferrús Abad; Maria Alma Bracho; Mariana Reyes-Prieto; Neris Garcia-Gonzalez; Paula Ruiz-Hueso; Sandra Carbo; Vicente Soriano Chirona |
| EPI_ISL_436235 to 436242, EPI_ISL_436244 to 436270, EPI_ISL_436372 to 436381 | Servicio de Microbiologia. Hospital Universitario Doctor Peset | Sequencing and Bioinformatics Service and Molecular Epidemiology Research Group. FISABIO-Public Health | Beatriz Beamud; Fernando Gonzalez-Candelas; Giuseppe D'Auria; Griselda De Marco; Inma Galán Vendrell; Ivan Ansari; Jose Miguel Nogueira Coito; Juan Alberola Enguídanos; Juan Jose Camarena Miñana; Lidia Ruiz Roldan; Llúcia Martínez-Priego; Loreto Ferrús Abad; Maria Alma Bracho; Mariana Reyes-Prieto; Marta Pla Diaz; Neris Garcia-Gonzalez; Paula Ruiz-Hueso; Rosa González Pellicer; Sandra Carbo; Vicente Soriano Chirona |
| EPI_ISL_455735 to 455736 | Servicio de Microbiologia. Hospital_Arnau_de_Vilanova | Sequencing and Bioinformatics Service and Molecular Epidemiology Research Group. FISABIO-Public Health, and SeqCOVID-Spain Consortium | Fernando Gonzalez-Candelas; Giuseppe 'Auria; Griselda De Marco; Inma Galán Vendrell; Ivan Ansari; Lidia Ruiz Roldan; Llúcia Martínez-Priego; Loreto Ferrús Abad; Maria Alma Bracho; Mariana Reyes-Prieto; Neris Garcia-Gonzalez; Paula Ruiz-Hueso; Sandra Carbo; Vicente Soriano Chirona; Victoria Dominguez |
| EPI_ISL_779730, EPI_ISL_846654, EPI_ISL_871908 to 871910, EPI_ISL_871939 to 871941, EPI_ISL_871954, EPI_ISL_871956, EPI_ISL_871960 to 871964 | | | |
| see above | Servicio de Microbiología Clínica (Complejo Hospitalario de Navarra, Pamplona), Instituto de Investigación Sanitaria de Navarra (IdiSNA) | SeqCOVID-SPAIN consortium/IBV(CSIC) | Ana Miqueleiz and SeqCOVID-SPAIN consortium; Ana Navascués; Carmen Ezpeleta Baquedano |
| EPI_ISL_538055 to 538061, EPI_ISL_538119 to 538123, EPI_ISL_538125 to 538129, EPI_ISL_538131 to 538132, EPI_ISL_538134 to 538135, EPI_ISL_538137 to 538143, EPI_ISL_538145 to 538158, EPI_ISL_538160 to 538162, EPI_ISL_538164 to 538211, EPI_ISL_538213 to 538230, EPI_ISL_538568 to 538578, EPI_ISL_538580 to 538581, EPI_ISL_538583 to 538587, EPI_ISL_538589 to 538593, EPI_ISL_538596 to 538603, EPI_ISL_538605 to 538607 | | | |
| see above | Servicio de Microbiología y Parasitología clínica. UCEIMP. Hospital Universitario Virgen del Rocío/IBIS/CSIC/US | SeqCOVID-SPAIN consortium/IBV(CSIC) | Guillermo Martín Gutiérrez; Javier Aznar Martín and SeqCOVID-SPAIN consortium; Lidia Gálvez Benitez; Verónica González Galán; Ángel Rodríguez Villodres |
| EPI_ISL_452544 to 452562, EPI_ISL_452564 to 452590, EPI_ISL_452592 to 452597, EPI_ISL_452599 to 452606, EPI_ISL_452608 to 452616 | Servicio de Microbiología y Parasitología clínica. UCEIMP. Hospital Universitario Virgen del Rocío/IBIS/CSIC/US. | SeqCOVID-SPAIN consortium/IBV(CSIC) | Guillermo Martí-n Gutiérrez; Javier Aznar Martí-n and SeqCOVID-SPAIN consortium; Lidia Gálvez Benitez; Verónica González Galán; Ángel Rodrí-guez Villodres |
| EPI_ISL_468765 to 468795, EPI_ISL_468797 to 468847, EPI_ISL_468849 to 468859, EPI_ISL_500287 to 500288, EPI_ISL_500290 to 500291, EPI_ISL_500336 to 500338, EPI_ISL_500341, EPI_ISL_500343 to 500347, EPI_ISL_500349 to 500351, EPI_ISL_500354 to 500356, EPI_ISL_500360, EPI_ISL_500362, EPI_ISL_500365 to 500366, EPI_ISL_510333 to 510334, EPI_ISL_510336 to 510379, EPI_ISL_510381 to 510390, EPI_ISL_537729 to 537733, EPI_ISL_537735 to 537784, EPI_ISL_537786 to 537787, EPI_ISL_654402 to 654404, EPI_ISL_654406 to 654413, EPI_ISL_654415 to 654434, EPI_ISL_654437 to 654467, EPI_ISL_722210, EPI_ISL_722212 to 722215, EPI_ISL_722217, EPI_ISL_722219, EPI_ISL_722221, EPI_ISL_722223 to 722228, EPI_ISL_722230 to 722233, EPI_ISL_722237, EPI_ISL_722242 to 722243, EPI_ISL_722245 to 722258, EPI_ISL_722260 to 722262, EPI_ISL_722264 to 722271, EPI_ISL_779732, EPI_ISL_779734 to 779735, EPI_ISL_779737, EPI_ISL_779740, EPI_ISL_779753, EPI_ISL_779774 to 779779 | | | |
| see above | Servicio de Microbiología, Hospital Miguel Servet, Zaragoza | SeqCOVID-SPAIN consortium/IBV(CSIC) | Alexander Tristancho Baró; Ana Milagro; Antonio Rezusta López; Nieves Martínez Cameo and SeqCOVID-SPAIN consortium; Yolanda Gracia Grataloup |
| EPI_ISL_541008 to 541009, EPI_ISL_541012 to 541013, EPI_ISL_541015 to 541036 | Servicio de Microbiología, Hospital Miguel Servet, Zaragoza | SeqCOVID-SPAIN consortium/Institute of Biomedicine of Valencia, IBV-CSIC | Alexander Tristancho Baró; Ana Milagro; Antonio Rezusta López; Nieves Martínez Cameo and SeqCOVID-SPAIN consortium; Yolanda Gracia Grataloup |
| EPI_ISL_654616, EPI_ISL_654619 to 654625, EPI_ISL_654627 to 654653, EPI_ISL_654655 to 654684, EPI_ISL_722241 | Servicio de Microbiología, Hospital Universitario Central de Asturias | SeqCOVID-SPAIN consortium/IBV(CSIC) | Cristián Castelló Abietar; Jose A. Boga; Marta Elena Álvarez-Argüelles; Santiago Melón and SeqCOVID-SPAIN consortium; Susana Rojo-Alba |
| EPI_ISL_582695 to 582714, EPI_ISL_582716 to 582750, EPI_ISL_582752 to 582754, EPI_ISL_582756 to 582766, EPI_ISL_654618 | Servicio de Microbiología, Hospital Universitario Cruces, Bilbao | SeqCOVID-SPAIN consortium/IBV(CSIC) | Maitane Aranzamendi Zaldumbide and SeqCOVID-SPAIN consortium |
| EPI_ISL_467055 to 467058, EPI_ISL_468952 to 468957, EPI_ISL_468959 to 468962, EPI_ISL_468966 to 468968, EPI_ISL_468970 to 468995, EPI_ISL_468997, EPI_ISL_468999 to 469001, EPI_ISL_469003, EPI_ISL_469005 to 469015, EPI_ISL_541949 to 541953, EPI_ISL_541956 to 541957, EPI_ISL_541959 to 541969, EPI_ISL_691612 to 691616, EPI_ISL_691618 to 691622, EPI_ISL_691624 to 691633, EPI_ISL_691635 to 691637, EPI_ISL_691641 to 691643, EPI_ISL_691645 to 691660, EPI_ISL_691663, EPI_ISL_691665, EPI_ISL_691667 to 691676, EPI_ISL_691678 to 691680, EPI_ISL_779850 to 779873, EPI_ISL_779875, EPI_ISL_779877, EPI_ISL_779879 to 779887, EPI_ISL_779889 to 779891, EPI_ISL_779893, EPI_ISL_779895 to 779911, EPI_ISL_779913 to 779921, EPI_ISL_819128 to 819129, EPI_ISL_819131 to 819133, EPI_ISL_819135 to 819138, EPI_ISL_819140, EPI_ISL_819142 to 819144, EPI_ISL_819149 to 819150, EPI_ISL_819156 to 819157, EPI_ISL_819163, EPI_ISL_819167, EPI_ISL_819169 to 819176, EPI_ISL_819178, EPI_ISL_819180 to 819190, EPI_ISL_819192 to 819202 | | | |
| see above | Servicio de Microbiología, Hospital Universitario Son Espases | SeqCOVID-SPAIN consortium/IBV(CSIC) | Antonio Oliver and SeqCOVID-SPAIN consortium; Carla López-Causapé; Jordi Reina; Jordi Reina y Antonio Oliver and SeqCOVID-SPAIN consortium |
| EPI_ISL_510510 to 510513, EPI_ISL_510515 to 510523, EPI_ISL_654511, EPI_ISL_654513 to 654514, EPI_ISL_654516 to 654518, EPI_ISL_654520 to 654522, EPI_ISL_654524 to 654538, EPI_ISL_660265 to 660301, EPI_ISL_660303 to 660322, EPI_ISL_671834 to 671851, EPI_ISL_671853 to 671858, EPI_ISL_671860 to 671862, EPI_ISL_683598 to 683610, EPI_ISL_683612 to 683620, EPI_ISL_683622 to 683623, EPI_ISL_683625 to 683640, EPI_ISL_683643 to 683656, EPI_ISL_779731, EPI_ISL_855416, EPI_ISL_855424, EPI_ISL_855426 to 855446, EPI_ISL_855448 to 855485, EPI_ISL_855487 to 855488, EPI_ISL_855490 to 855493, EPI_ISL_871891, EPI_ISL_871914 to 871915, EPI_ISL_871917 to 871919, EPI_ISL_871922 to 871934, EPI_ISL_871937 | | | |
| see above | Servicio de Microbiología, Laboratori Clínic Metropolitana Nord. Hospital Universitari Germans Trias i Pujol. Institut d'Investigació en Ciències de la Salut Germans Trias i Pujol | SeqCOVID-SPAIN consortium/IBV(CSIC) | Adrián Antuori; Anabel Fernández; Anna Not; Antoni E. Bordoy; Cristina Casañ and SeqCOVID-SPAIN consortium; Elisa Martró; Nona Romaní; Nona Romaní and SeqCOVID-SPAIN consortium; Verónica Saludes |

| | | | |
|---|---|---|---|
| | (IGTP) | | |
| EPI_ISL_537608, EPI_ISL_539234, EPI_ISL_539240 to 539242, EPI_ISL_539292 to 539295, EPI_ISL_539297 to 539298, EPI_ISL_846652 | Servicio de Microbiología. Consorcio Hospital General Universitario de Valencia | SeqCOVID-SPAIN consortium/IBV(CSIC) | Beatriz Beamud; Begoña Fuster Escrivá; Concepción Gimeno Cardona and SeqCOVID-SPAIN consortium; David Navalpotro; David Navarro; Fernando Gonzalez-Candelas and SeqCOVID-SPAIN consortium; Giuseppe D'Auria; Griselda De Marco; Inma Galán Vendrell; Ivan Ansari; Lidia Ruiz Roldan; Llúcia Martínez-Priego; Loreto Ferrús Abad; Maria Alma Bracho; Mariana Reyes-Prieto; Marta Pla Diaz; María Dolores Ocete; Neris Garcia-Gonzalez; Paula Ruiz-Hueso; Rafael Medina González; Sandra Carbo; Vicente Soriano Chirona |
| EPI_ISL_416483 to 416484, EPI_ISL_416486 to 416487, EPI_ISL_417205, EPI_ISL_419675 to 419685, EPI_ISL_420112 to 420124, EPI_ISL_425178 to 425198, EPI_ISL_425212 to 425223, EPI_ISL_436195 to 436234, EPI_ISL_436271 to 436277, EPI_ISL_436361, EPI_ISL_436382 to 436395 | | | |
| see above | Servicio de Microbiología. Consorcio Hospital General Universitario de Valencia | Sequencing and Bioinformatics Service and Molecular Epidemiology Research Group. FISABIO-Public Health | ; Beatriz Beamud; Concepcion Gimeno; David Navarro; Fernando Gonzalez-Candelas; Giuseppe D'Auria; Griselda De Marco; Inma Galán Vendrell; Ivan Ansari; Lidia Ruiz Roldan; Llúcia Martínez-Priego; Llúcia Martínez-PriegoGriselda De Marco; Loreto Ferrús Abad; Maria Alma Bracho; Maria Dolores Ocete; Mariana Reyes-Prieto; Marta Pla Diaz; Neris Garcia-Gonzalez; Paula Ruiz-Hueso; Sandra Carbo; Vicente Soriano Chirona |
| EPI_ISL_452365 to 452394, EPI_ISL_452396 to 452401, EPI_ISL_452403 to 452451, EPI_ISL_509602 to 509609, EPI_ISL_509611, EPI_ISL_509619 to 509622, EPI_ISL_509624 to 509631, EPI_ISL_510044 to 510048, EPI_ISL_510050 to 510053, EPI_ISL_510055, EPI_ISL_510057 to 510059, EPI_ISL_510061 to 510062, EPI_ISL_565910 to 565916, EPI_ISL_565918 | | | |
| see above | Servicio de Microbiología. HRU de Málaga. Servicio Andaluz de Salud | SeqCOVID-SPAIN consortium/IBV(CSIC) | Begoña Palop Borrás and SeqCOVID-SPAIN consortium; Inmaculada de Toro Peinado; Inmaculada de Toro Peinado. MªConcepción Mediavilla Gradolph. Begoña Palop Borrás and SeqCOVID-SPAIN consortium; Ma Concepción Mediavilla Gradolph; Maria Concepción Mediavilla Gradolph |
| EPI_ISL_654484 to 654497, EPI_ISL_728358 | Servicio de Microbiología. Hospital Clínico Universitario de Valencia | SeqCOVID-SPAIN consortium/IBV(CSIC) | David Navarro Ortega; Eliseo Albert Vicent; Ignacio Torres and SeqCOVID-SPAIN consortium |
| EPI_ISL_537946 to 537969, EPI_ISL_537971 to 537995, EPI_ISL_538613 to 538645, EPI_ISL_538647 to 538671, EPI_ISL_539284 to 539287, EPI_ISL_583918 to 583940, EPI_ISL_583942 to 583952, EPI_ISL_654468 to 654473, EPI_ISL_654475 to 654478, EPI_ISL_654480 to 654482, EPI_ISL_657450, EPI_ISL_657452, EPI_ISL_657455, EPI_ISL_657458, EPI_ISL_657460, EPI_ISL_657463, EPI_ISL_657465 to 657467, EPI_ISL_722211, EPI_ISL_722216, EPI_ISL_722235 to 722236, EPI_ISL_722238 to 722239, EPI_ISL_796064 to 796065, EPI_ISL_796067 to 796078, EPI_ISL_796080 to 796098, EPI_ISL_796100, EPI_ISL_796102 to 796114, EPI_ISL_796116 to 796126, EPI_ISL_796135 to 796138, EPI_ISL_796140 to 796157 | | | |
| see above | Servicio de Microbiología. Hospital General Universitario de Castellón | SeqCOVID-SPAIN consortium/IBV(CSIC) | María Dolores Tirado Balaguer and SeqCOVID-SPAIN consortium; María Dolores Tirado and SeqCOVID-SPAIN consortium; Rosario Moreno; Rosario Moreno Muñoz |
| EPI_ISL_480952 to 480958, EPI_ISL_480960, EPI_ISL_480962 to 480963, EPI_ISL_480965 to 480974, EPI_ISL_480976 to 480980, EPI_ISL_480982 to 480988, EPI_ISL_480990 to 480994, EPI_ISL_480996 to 481002, EPI_ISL_481004, EPI_ISL_481007 to 481013, EPI_ISL_481015 to 481016, EPI_ISL_481018 to 481024, EPI_ISL_481026 to 481028, EPI_ISL_481030 to 481033, EPI_ISL_481036 to 481037, EPI_ISL_481039 to 481040, EPI_ISL_500292 to 500296, EPI_ISL_500298 to 500334, EPI_ISL_509601, EPI_ISL_509610, EPI_ISL_509612 to 509613, EPI_ISL_509633 to 509654, EPI_ISL_510391 to 510423, EPI_ISL_510467 to 510476, EPI_ISL_510478 to 510509, EPI_ISL_538062 to 538081, EPI_ISL_565920 to 565921, EPI_ISL_565923, EPI_ISL_582035 to 582036, EPI_ISL_582041 to 582042, EPI_ISL_582045, EPI_ISL_582048 to 582050, EPI_ISL_582052, EPI_ISL_582055, EPI_ISL_582057, EPI_ISL_582060, EPI_ISL_582062, EPI_ISL_582064, EPI_ISL_582066, EPI_ISL_582072 to 582074, EPI_ISL_582076 to 582085, EPI_ISL_582088, EPI_ISL_582090, EPI_ISL_654539 to 654544, EPI_ISL_654547 to 654558, EPI_ISL_654560 to 654568, EPI_ISL_654570 to 654574, EPI_ISL_654579, EPI_ISL_654581 to 654583, EPI_ISL_654585 to 654595, EPI_ISL_654597 to 654603, EPI_ISL_654605 to 654614 | | | |
| see above | Servicio de Microbiología. Hospital Universitario Donostia. OSI Donostialdea. Área de Enfermedades Infecciosas, Grupo de Infección Respiratoria y Resistencia Antimicrobiana. Instituto de Investigación Sanitaria Biodonostia | SeqCOVID-SPAIN consortium/IBV(CSIC) | Ane Sorrarain; Gustavo Cilla; Gustavo Cilla Eguiluz; Jose Maria Marimón and SeqCOVID-SPAIN consortium; Luis Piñeiro; Luis Piñeiro Vázquez; Milagrosa Montes; Milagrosa Montes Ros |
| EPI_ISL_541084, EPI_ISL_541088 to 541098, EPI_ISL_541100 to 541110, EPI_ISL_541112 to 541130, EPI_ISL_541132 to 541135, EPI_ISL_541137 | Servicio de Microbiología. Hospital Universitario Donostia. OSI Donostialdea. Área de Enfermedades Infecciosas, Grupo de Infección Respiratoria y Resistencia Antimicrobiana. Instituto de Investigación Sanitaria Biodonostia | SeqCOVID-SPAIN consortium/Institute of Biomedicine of Valencia, IBV-CSIC | Gustavo Cilla; Jose Maria Marimón and SeqCOVID-SPAIN consortium; Luis Piñeiro; Milagrosa Montes |
| EPI_ISL_452617, EPI_ISL_452619 to 452621, EPI_ISL_452623 to 452653, EPI_ISL_452655 to 452685, EPI_ISL_452687 to 452691, EPI_ISL_468860 to 468883, EPI_ISL_468885 to 468893, EPI_ISL_468895 to 468913, EPI_ISL_468915 to 468951 | | | |
| see above | Servicio de Microbiología. Hospital Universitario Donostia. OSI Donostialdea. Área de Enfermedades Infecciosas, Grupo de Infección Respiratoria y Resistencia Antimicrobiana. Instituto de Investigación Sanitaria Biodonostia. | SeqCOVID-SPAIN consortium/IBV(CSIC) | Gustavo Cilla; Jose Maria Marimón and SeqCOVID-SPAIN consortium; Luis Piñeiro; Milagrosa Montes |
| EPI_ISL_529010 to 529012, EPI_ISL_849654, EPI_ISL_849656 to 849661 | Servizio Igiene Epidemiologia e Sanità Pubblica (SIESP)-L'Aquila | Istituto Zooprofilattico Sperimentale dell'Abruzzo e Molise "G.Caporale" | Ancora M; Cammà C; Curini V; Di Domenico M; Di Pasquale A; Lorusso A; Mangone I; Marcacci M; Puglia I; Rinaldi A; Savini G. |
| EPI_ISL_833085 to 833086 | Servizio di Igiene Epidemiologia e Sanità Pubblica (SIESP) CHIETI - DRIVE IN GISSI | Istituto Zooprofilattico Sperimentale dell'Abruzzo e Molise "G. Caporale" | Ancora M; Calistri P; Cammà C; Curini V; Delli Compagni E; Di Domenico M; Di Pasquale A; Lorusso A; Mangone I; Marcacci M; Puglia I; Rinaldi A; Savini G |
| EPI_ISL_833087 | Servizio di Igiene Epidemiologia e Sanità Pubblica (SIESP) L'AQUILA | Istituto Zooprofilattico Sperimentale dell'Abruzzo e Molise "G. Caporale" | Ancora M; Calistri P; Cammà C; Curini V; Delli Compagni E; Di Domenico M; Di Pasquale A; Lorusso A; Mangone I; Marcacci M; Puglia I; Rinaldi A; Savini G |
| EPI_ISL_833088 to 833089 | Servizio di Igiene Epidemiologia e Sanità Pubblica (SIESP) SULMONA | Istituto Zooprofilattico Sperimentale dell'Abruzzo e Molise "G. Caporale" | Ancora M; Calistri P; Cammà C; Curini V; Delli Compagni E; Di Domenico M; Di Pasquale A; Lorusso A; Mangone I; Marcacci M; Puglia I; Rinaldi A; Savini G |
| EPI_ISL_435152 | Servizio di Igiene, Epidemiologia e Sanità Pubblica (SIESP) Avezzano | Istituto Zooprofilattico Sperimentale dell'Abruzzo e Molise "G.Caporale" | Ancora M; Cammà C; Curini V; Di Domenico M; Di Pasquale A; Lorusso A; Mangone I; Marcacci M; Puglia I; Rinaldi A; Savini G |
| EPI_ISL_529006, EPI_ISL_529008, EPI_ISL_849653, EPI_ISL_849655 | Servizio di igiene e sanità pubblica (SIESP)-Teramo | Istituto Zooprofilattico Sperimentale dell'Abruzzo e Molise "G.Caporale" | Ancora M; Cammà C; Curini V; Di Domenico M; Di Pasquale A; Lorusso A; Mangone I; Marcacci M; Puglia I; Rinaldi A; Savini G. |
| EPI_ISL_528994 to 529005, EPI_ISL_529023 to 529025, EPI_ISL_849650 to 849652, EPI_ISL_849662 to 849663 | Servizio di igiene epidemiologia e sanità pubblica (SIESP)-Chieti | Istituto Zooprofilattico Sperimentale dell'Abruzzo e Molise "G.Caporale" | Ancora M; Cammà C; Curini V; Di Domenico M; Di Pasquale A; Lorusso A; Mangone I; Marcacci M; Puglia I; Rinaldi A; Savini G. |
| EPI_ISL_436730 | Servizio di igiene epidemiologia e sanità pubblica (Siesp) Chieti | Istituto Zooprofilattico Sperimentale dell'Abruzzo e Molise "G.Caporale" | Ancora M; Cammà C; Curini V; Di Domenico M; Di Pasquale A; Lorusso A; Mangone I; Marcacci M; Puglia I; Rinaldi A; Savini G |
| EPI_ISL_710547, EPI_ISL_710554, EPI_ISL_710563 | Sestre Milosrdnice University Hospital Center | Ruer Boškovic Institute; Forensic Science Centre Ivan Vueti; University of Zagreb Faculty of Science | Ana Livun; Antonela Blažekovi; Boris Maek; Danilo Licastro; Dunja Glavaš; Fran Boroveki; Gordana Maravi Vlahoviek; Ivan Šamija; Ivana elap; Josipa Skelin; Kristian Vlahoviek; Kristina Gotovac Jerei; Lidija Cvetko-Krajinovi; Lucija Markulin; Maja Kuzman; Marina Korolija; Mario Štefanovi; Mirjana Domazet-Lošo; Paula Štancl; Robert Beluži; Rosa Karli; Sanja Tadinac; Tomislav Domazet-Lošo; Vjekoslav Tomai; Vladimir Krajinovi; Željka Maak Šafranko |
| EPI_ISL_534225 to 534229, EPI_ISL_560980 | Skanes universitetssjukhus Lund | The Public Health Agency of Sweden | Anna Risberg; Anna-Malin Linde; Karin Tegmark-Wisell; Maria Lind Karlberg; Mattias Haukland; Mia Brytting; Olov Svartstrom; Oskar Karlsson Lindsjo; Petra Edquist; Reza Advani; Sandra Broddesson |
| EPI_ISL_454494 to 454496, EPI_ISL_455842 to 455845, EPI_ISL_475085, EPI_ISL_475092, EPI_ISL_475099, EPI_ISL_475105 to 475109, EPI_ISL_475111 to 475113, EPI_ISL_475140, EPI_ISL_475549 to 475550, EPI_ISL_475554 to 475555, EPI_ISL_476143 to 476144, EPI_ISL_510845 to 510851, EPI_ISL_548245 to 548246, EPI_ISL_615115 to 615116, EPI_ISL_648198 to 648199, EPI_ISL_654955 to 654956, EPI_ISL_710584 | | | |
| see above | Skovde/Unilabs | The Public Health Agency of Sweden | Anna Risberg; Anna-Malin Linde; Department of Microbiology; Karin Tegmark-Wisell; Maria Lind Karlberg; Mattias Haukland; Mia Brytting; Olov Svartstrom; Oskar Karlsson Lindsjo; Petra Edquist; Reza Advani; Sandra Broddesson; Shamam Muradrasoli; Shaman Muradrasoli; The Public Health Agency of Sweden |
| EPI_ISL_455108 | Smedby HC | The Public Health Agency of Sweden | Anna Risberg; Anna-Malin Linde; Karin Tegmark-Wisell; Maria Lind Karlberg; Mia Brytting; Olov Svartstrom; Oskar Karlsson Lindsjo; Susanne Brunby; Theresa Enkirch |
| EPI_ISL_408431 | Sorbonne Université, Inserm et Assistance Publique-Hôpitaux de Paris (Pitié Salpétrière) | National Reference Center for Viruses of Respiratory Infections, Institut Pasteur, Paris | Angela Brisebarre; Anne-Geneviève Marcelin; David Boutolleau; Elise Klément; Eric Caumes.; Flora Donati; Marion Barbet; Maud Vanpeene; Mélanie Albert; Méline Bizard; Sonia Burrel; Sylvie Behillil; Sylvie van der Werf; Valérie Pourcher; Vincent Calvez; Vincent Enouf |
| EPI_ISL_860547 to 860548, EPI_ISL_860550 | St.Olavs hospital/NTNU | IKOM, NTNU | Aleksandr Ianevski; Denis Kainov; Svein Arne Nordbo |
| EPI_ISL_450346 to 450352 | St.Olavs hospital/NTNU | Institute of Genomics Core Facility, University of Tartu | Aleksandr Ianevski; Denis Kainov; Janne-Fossum Malmring; Svein Arne Nordbø; Tuuli Reisberg |
| EPI_ISL_596666 to 596670, EPI_ISL_596672 to | St.Vincent's University Hospital | St.Vincent's University Hospital | Aisling Purcell; Gabriel Gonzalez; Guerrino Macori; Kirsten Schaffer; Lynda Fenelon; Mary Lucey; Niamh Mullane; Suzie Coughlan; Séamus Fanning; Una |

| | | | |
|---|---|---|---|
| 596676 | | | Sutton-Fitzpatrick |
| EPI_ISL_412912 | State Health Office Baden-Württemberg | Charite Universitatsmedizin Berlin, Institute of Virology | Barbara Muhlemann; Christian Drosten; Julia Schneider; Jörn Beheim-Schwarzbach; Rainer Oehme; Silke Fischer; Talitha Veith; Terry Jones; Victor M Corman |
| EPI_ISL_428860 to 428924 | State Research Center of Virology and Biotechnology VECTOR, Department of Collection of Microorganisms | State Research Center of Virology and Biotechnology VECTOR, Department of Collection of Microorganisms | Alexander N. Shvalov; Anastasiya A. Nazarenko; Anastasiya M. Smirnova; Elena V. Gavrilova; Oleg V. Pyankov; Rinat A. Maksyutov; Sergey A. Bodnev; Tatyana V. Tregubchak |
| EPI_ISL_451655 to 451664 | State Sanitary Inspectorate | Laboratory of Recombinant Vaccines | Boguslaw Szewczyk; Jaroslaw Pinkas; Krystyna Bienkowska-Szewczyk; Lukasz Rabalski |
| EPI_ISL_420149 | Stavanger University Hospital, Department of Medical Microbiology | Norwegian Institute of Public Health, Department of Virology | Hilde Elshaug; Kamilla Heddeland Instefjord; Karoline Bragstad; Kathrine Stene-Johansen; Olav Hungnes |
| EPI_ISL_648152 to 648155 | Stockholm | The Public Health Agency of Sweden | Anna Risberg; Anna-Malin Linde; Karin Tegmark-Wisell; Maria Lind Karlberg; Mattias Haukland; Mia Brytting; Olov Svartstrom; Oskar Karlsson Lindsjo; Petra Edquist; Reza Advani; Sandra Broddesson |
| EPI_ISL_582799, EPI_ISL_582839 | Stockholm_KUL Solna | The Public Health Agency of Sweden | Anna Risberg; Anna-Malin Linde; Karin Tegmark-Wisell; Maria Lind Karlberg; Mattias Haukland; Mia Brytting; Olov Svartstrom; Oskar Karlsson Lindsjo; Petra Edquist; Reza Advani; Sandra Broddesson |
| EPI_ISL_661273 | Stockholm_StGoran | The Public Health Agency of Sweden | Department of Microbiology; The Public Health Agency of Sweden |
| EPI_ISL_491036 to 491040, EPI_ISL_491042 to 491049, EPI_ISL_491052 to 491053, EPI_ISL_491058, EPI_ISL_491060 to 491061, EPI_ISL_491063 to 491070, EPI_ISL_491072, EPI_ISL_491075 to 491076, EPI_ISL_491078 to 491082, EPI_ISL_491084 to 491090, EPI_ISL_678397 to 678398 | | | |
| see above | Suceava County Emergency Hospital | "Stefan cel Mare" University Metagenomics Lab | Antoniadis Panagiotis et al.; Lobiuc Andrei; Lobiuc Andrei et al.; Puscaselu Roxana |
| EPI_ISL_486834 | Suceava County Energency Hospital "Sf. Ioan cel Nou" | SMU Metagenomics lab | Antoniadis Panagiotis; Lobiuc Andrei |
| EPI_ISL_648157 to 648161, EPI_ISL_648192 | Sundsvall | The Public Health Agency of Sweden | Anna Risberg; Anna-Malin Linde; Karin Tegmark-Wisell; Maria Lind Karlberg; Mattias Haukland; Mia Brytting; Olov Svartstrom; Oskar Karlsson Lindsjo; Petra Edquist; Reza Advani; Sandra Broddesson |
| EPI_ISL_534244, EPI_ISL_560986 | Sundsvalls sjukhus | The Public Health Agency of Sweden | Anna Risberg; Anna-Malin Linde; Karin Tegmark-Wisell; Maria Lind Karlberg; Mattias Haukland; Mia Brytting; Olov Svartstrom; Oskar Karlsson Lindsjo; Petra Edquist; Reza Advani; Sandra Broddesson |
| EPI_ISL_434663, EPI_ISL_434672, EPI_ISL_434675 to 434676, EPI_ISL_445225, EPI_ISL_455106, EPI_ISL_469066, EPI_ISL_469070, EPI_ISL_475543, EPI_ISL_475564, EPI_ISL_476136 | | | |
| see above | Surbrunns VC | The Public Health Agency of Sweden | Anna Risberg; Anna-Malin Linde; Erik Embring; Karin Tegmark-Wisell; Maria Lind Karlberg; Mattias Haukland; Mia Brytting; Olov Svartstrom; Oskar Karlsson Lindsjo; Petra Edquist; Reza Advani; Sandra Broddesson; Shamam Muradrasoli; Theresa Enkirch |
| EPI_ISL_434645, EPI_ISL_434649 | Svardsjo VC | The Public Health Agency of Sweden | Anna Risberg; Anna-Malin Linde; Karin Tegmark-Wisell; Maria Lind Karlberg; Mia Brytting; Olov Svartstrom; Oskar Karlsson Lindsjo; Theresa Enkirch; Tommy Janers |
| EPI_ISL_857489 to 857541, EPI_ISL_860730 to 860760, EPI_ISL_860762 to 860774, EPI_ISL_860776 to 860781 | Swiss National Reference Centre for Influenza | Swiss National Reference Centre for Influenza | Adrian Egli; Alfredo Mari; Ana Rita Goncalves; Ana Rita Gonçalves; Ana Rita Gonçalves Cabecinhas; Christoph Grünig; Emmanouil Dermitzakis; Florian Laubscher; Hans Hirsch; Helena MB Seth-Smith; Henri Pegeot; Ioannis Xenarios; Julia Bielicki; Karoline Leuzinger; Keith Harshman; Laurent Kaiser; Lorenzo Cerutti; Madlen Stange; Manuel Battegay; Melyssa Elies; Samuel Cordey; Tim Roloff |
| EPI_ISL_775583, EPI_ISL_775587, EPI_ISL_831986 to 831988 | Synlab Medilab, Mikrobiologi | The Public Health Agency of Sweden | Department of Microbiology; The Public Health Agency of Sweden |
| EPI_ISL_455719 | T.C. Salk Bakanl Adyaman l Salk Müdürlüü Adyaman Eitim Ve Aratrma Hastanesi | VETAL Animal Health Products Company, BSL3+ Production Laboratuary /Turkey | Abidin Ercan Yonucu; Fatma Nilay Tutak; Fethiye Sevimli; Haluk Uluca; Mehmet Turgut; Muhittin Önderci; Murat Dönen; O. Ugur Sezerman |
| EPI_ISL_766724, EPI_ISL_775593, EPI_ISL_831993 to 831995 | TATAA Biocenter | The Public Health Agency of Sweden | Department of Microbiology; The Public Health Agency of Sweden |
| EPI_ISL_482057 to 482067, EPI_ISL_482120 to 482123, EPI_ISL_482125 to 482130 | The Department of Microbiology, Torbay and South Devon NHS Foundation Trust | Wellcome Sanger Institute for the COVID-19 Genomics UK (COG-UK) consortium | Amy Hurd; Anthony Mogridge; Cheryl Bailiss; Cordelia Langford; David K. Jackson; Dominic Kwiatkowski; Ewan Harrison; Gary Booth; Helen Brown; Ian Johnston; Jack Howe; John Sillitoe on behalf of the Wellcome Sanger Institute COVID-19 Surveillance Team (http://www.sanger.ac.uk/covid-team); Mel Brown; Michelle Harrison and Alex Alderton; Roberto Amato; Sonia Goncalves; Sophie Lloyd |
| EPI_ISL_491092, EPI_ISL_530347 to 530350, EPI_ISL_539493 to 539494, EPI_ISL_539777, EPI_ISL_541082, EPI_ISL_541138, EPI_ISL_541332 to 541334, EPI_ISL_541336 to 541337, EPI_ISL_545580, EPI_ISL_545954, EPI_ISL_545956 to 545957, EPI_ISL_546436, EPI_ISL_546935 to 546936, EPI_ISL_547966 to 547968, EPI_ISL_560408 to 560412, EPI_ISL_568577, EPI_ISL_577623 to 577634, EPI_ISL_577636 to 577638, EPI_ISL_584073 to 584082, EPI_ISL_626570 to 626625, EPI_ISL_660553 to 660587, EPI_ISL_660589 to 660599, EPI_ISL_693660 to 693687, EPI_ISL_736998 to 737033, EPI_ISL_792684 to 792707, EPI_ISL_850663 to 850670, EPI_ISL_850674 to 850676, EPI_ISL_850687 | | | |
| see above | The National Institute of Public Health | State Veterinary Institute Prague | A; D; H; J; Jirincova; L; M; Nagy; Novakova; Trinklova; Trnka; Vecerova |
| EPI_ISL_471528, EPI_ISL_471530, EPI_ISL_471540, EPI_ISL_471544, EPI_ISL_471547, EPI_ISL_471550, EPI_ISL_471553, EPI_ISL_471555, EPI_ISL_476067 | | | |
| see above | The National Institute of Public Health | State Veterinary Institute Prague and The National Institute of Public Health | A; D; H; J; Jirincova; L; Nagy; Novakova; Trnka; Vecerova |
| EPI_ISL_541335 | The National Institute of Public Health | Sídlištní 136/24 165 03, Prague Czech Republic | A; D; H; J; Jirincova; L; Nagy; Novakova; Trnka; Vecerova |
| EPI_ISL_489708 to 489709, EPI_ISL_490112, EPI_ISL_491093 to 491095, EPI_ISL_491117 to 491119, EPI_ISL_513511 to 513512 | The National Institute of Public Health | The National Institute of Public Health and State Veterinary Institute Prague | A; D; H; J; Jirincova; L; Nagy; Novakova; Trnka; Vecerova |
| EPI_ISL_414477 | The National Institute of Public Health Center for Epidemiology and Microbiology | State Veterinary Institute Prague | Alexander Nagy; Dusan Trnka; Helena Jirincova; Jaromira Vecerova; Klara Labska; Ludmila Novakova; Oldrich Bartos; Olga Storkanova |
| EPI_ISL_437519 | The National Institute of Public Health Center for Epidemiology and Microbiology | The National Institute of Public Health Center for Epidemiology and Microbiology | Alexander Nagy; Dusan Trnka; Helena Jirincova; Jaromira Vecerova; Ludmila Novakova |
| EPI_ISL_426379 | The National Laboratory of Health, Environment and Food, Maribor, Slovenia | The National Laboratory of Health, Environment and Food, Maribor, Slovenia | Blazun Vosner H.; Duh D.; Hedzet S.; Janezic S.; Mahnic A.; Rupnik M.; Zavrsnik J. |
| EPI_ISL_417550 to 417551, EPI_ISL_417553, EPI_ISL_417555 to 417577, EPI_ISL_417579 to 417586, EPI_ISL_417588 to 417589, EPI_ISL_417591 to 417595, EPI_ISL_417597 to 417599, EPI_ISL_417602 to 417604, EPI_ISL_417607, EPI_ISL_417609, EPI_ISL_417612, EPI_ISL_417614, EPI_ISL_417619, EPI_ISL_417621 to 417622, EPI_ISL_417624, EPI_ISL_417626 to 417630, EPI_ISL_417632, EPI_ISL_417634 to 417637, EPI_ISL_417639 to 417640, EPI_ISL_417642 to 417647, EPI_ISL_417649 to 417654, EPI_ISL_417673 to 417675, EPI_ISL_417677 to 417685, EPI_ISL_417687 to 417688, EPI_ISL_417690 to 417716, EPI_ISL_417718 to 417719, EPI_ISL_417721 to 417723, EPI_ISL_417725 to 417755, EPI_ISL_417757 to 417761, EPI_ISL_417763 to 417777, EPI_ISL_417779 to 417788, EPI_ISL_417790, EPI_ISL_417792 to 417793, EPI_ISL_417795 to 417815, EPI_ISL_417817, EPI_ISL_417819 to 417820, EPI_ISL_417822 to 417846, EPI_ISL_417848 to 417857, EPI_ISL_417859 to 417868, EPI_ISL_417870, EPI_ISL_417872 to 417876, EPI_ISL_424367 to 424371, EPI_ISL_424375 to 424376, EPI_ISL_424379 to 424385, EPI_ISL_424388 to 424393, EPI_ISL_424396, EPI_ISL_424398, EPI_ISL_424400 to 424406, EPI_ISL_424410 to 424415, EPI_ISL_424418 to 424419, EPI_ISL_424422 to 424427, EPI_ISL_424429, EPI_ISL_424434 to 424443, EPI_ISL_424445 to 424456, EPI_ISL_424459 to 424462, EPI_ISL_424464 to 424478, EPI_ISL_424480, EPI_ISL_424482 to 424486, EPI_ISL_424489, EPI_ISL_424491 to 424494, EPI_ISL_424496, EPI_ISL_424498 to 424503, EPI_ISL_424506 to 424511, EPI_ISL_424514 to 424529, EPI_ISL_424531 to 424532, EPI_ISL_424552, EPI_ISL_424554 to 424559, EPI_ISL_424563 to 424564, EPI_ISL_424571 to 424578, EPI_ISL_424580 to 424586, EPI_ISL_424588 to 424606, EPI_ISL_424610 to 424617, EPI_ISL_424619, EPI_ISL_424621 to 424624, EPI_ISL_826677 to 826678, EPI_ISL_826681, EPI_ISL_826683, EPI_ISL_826693, EPI_ISL_826715, EPI_ISL_826792, EPI_ISL_826801, EPI_ISL_826813, EPI_ISL_826815, EPI_ISL_826817, EPI_ISL_826819, EPI_ISL_826821, EPI_ISL_826824, EPI_ISL_826828, EPI_ISL_826854 to 826856, EPI_ISL_826858, EPI_ISL_826862, EPI_ISL_826884 to 826885, EPI_ISL_826893, EPI_ISL_826897, EPI_ISL_826916, EPI_ISL_826937, EPI_ISL_826955, EPI_ISL_826970, EPI_ISL_826972 to 826974, EPI_ISL_826980 to 826981, EPI_ISL_826990, EPI_ISL_826994, EPI_ISL_826999, EPI_ISL_827009, EPI_ISL_827012, EPI_ISL_827014, EPI_ISL_827016 to 827017, EPI_ISL_827027, EPI_ISL_827032 to 827033, EPI_ISL_827035 to 827037, EPI_ISL_827040, EPI_ISL_827045, EPI_ISL_827048, EPI_ISL_827063 to 827064, EPI_ISL_827066 to 827067, EPI_ISL_827069 to 827071, EPI_ISL_827074 to 827077, EPI_ISL_827079, EPI_ISL_827081 to 827087, EPI_ISL_827090, EPI_ISL_827110, EPI_ISL_827121 to 827124, EPI_ISL_827126, EPI_ISL_827142, EPI_ISL_827163 to 827164, EPI_ISL_827195, EPI_ISL_827198 to 827199, EPI_ISL_827202 to 827203, EPI_ISL_827205 to 827208, EPI_ISL_827210, EPI_ISL_827212, EPI_ISL_827229, EPI_ISL_827238, EPI_ISL_827242, EPI_ISL_827252 to 827254, EPI_ISL_827256 to 827258, EPI_ISL_827266, EPI_ISL_827270 to 827271, EPI_ISL_827288 to 827291, EPI_ISL_827293, EPI_ISL_827306, EPI_ISL_827309 to 827310, EPI_ISL_827330 to 827331, EPI_ISL_827333 to 827335, EPI_ISL_827337, EPI_ISL_827354, EPI_ISL_827359 to 827361, EPI_ISL_827363, EPI_ISL_827367 to 827368, EPI_ISL_827372, EPI_ISL_827374, EPI_ISL_827383, EPI_ISL_827395, EPI_ISL_827413, EPI_ISL_827419, EPI_ISL_827435, EPI_ISL_827437, EPI_ISL_827461, EPI_ISL_827484 to 827485, EPI_ISL_827492 to 827494, EPI_ISL_827496, EPI_ISL_827513 to 827514, EPI_ISL_827529, EPI_ISL_827531 to 827537, EPI_ISL_827539 to 827544, EPI_ISL_827548 to 827551, EPI_ISL_827553, EPI_ISL_827575 to 827577, EPI_ISL_827582 to 827584, EPI_ISL_827586 to 827590, EPI_ISL_827609, EPI_ISL_827644, EPI_ISL_827660, EPI_ISL_827663, EPI_ISL_827687, EPI_ISL_827692 to 827693, EPI_ISL_827695 to 827697, EPI_ISL_827713 to 827716, EPI_ISL_827718, EPI_ISL_827722 to 827725, EPI_ISL_827737, EPI_ISL_827746, EPI_ISL_827754, EPI_ISL_827781 to 827783, EPI_ISL_827800 to 827805, EPI_ISL_827821 to 827822, EPI_ISL_827825, EPI_ISL_827831, EPI_ISL_827859 to 827860, EPI_ISL_827863, EPI_ISL_827865 to 827870, EPI_ISL_827876 to 827881, EPI_ISL_827895, EPI_ISL_827902 to 827905, EPI_ISL_827907 to 827911, EPI_ISL_827913, EPI_ISL_827935, EPI_ISL_827939 to 827940, EPI_ISL_827942, EPI_ISL_828008 to 828014, EPI_ISL_828016, EPI_ISL_828020 to 828023, EPI_ISL_828025, EPI_ISL_828027, EPI_ISL_828029 to 828033, EPI_ISL_828035 to 828038, EPI_ISL_828053, EPI_ISL_828064, EPI_ISL_828070, EPI_ISL_828087, EPI_ISL_828089, EPI_ISL_828099, EPI_ISL_828137, EPI_ISL_828142 to 828144, EPI_ISL_828147, EPI_ISL_828157 to 828160, EPI_ISL_828162, EPI_ISL_828174 to 828175, EPI_ISL_828177, EPI_ISL_828188, EPI_ISL_828190 to 828191, EPI_ISL_828205, EPI_ISL_828209, EPI_ISL_828211 to 828212, EPI_ISL_828214 to 828215, EPI_ISL_828224 to 828227, EPI_ISL_828229, EPI_ISL_828241 to 828244, EPI_ISL_828250 to 828251, EPI_ISL_828265 to 828266, EPI_ISL_828270, EPI_ISL_828283, EPI_ISL_828285 to 828287, EPI_ISL_828320, EPI_ISL_828322 to 828327, EPI_ISL_828330 to 828333, EPI_ISL_828336, EPI_ISL_828352, EPI_ISL_828360, EPI_ISL_828363 to 828364, EPI_ISL_828383, EPI_ISL_828397, EPI_ISL_828399 to 828402, EPI_ISL_828405 to 828406, EPI_ISL_828408, EPI_ISL_828413, EPI_ISL_828464 to 828467, EPI_ISL_828469, EPI_ISL_828483 to 828487, EPI_ISL_828493 to 828495, EPI_ISL_828503 to 828507, EPI_ISL_828518, EPI_ISL_828522, EPI_ISL_828530, EPI_ISL_828537, EPI_ISL_828539 to 828541, EPI_ISL_828543, EPI_ISL_828554, EPI_ISL_828557, EPI_ISL_828589, EPI_ISL_828591, EPI_ISL_828593 to 828596, EPI_ISL_828630 to 828631, EPI_ISL_828634 to | | | |

| | | | |
|---|---|---|---|
| 828635, EPI_ISL_828641 to 828644, EPI_ISL_828646, EPI_ISL_828653, EPI_ISL_828655, EPI_ISL_828657 to 828660, EPI_ISL_828662 to 828663, EPI_ISL_828677, EPI_ISL_828679, EPI_ISL_828730, EPI_ISL_828741, EPI_ISL_828753 to 828755, EPI_ISL_828757 to 828759, EPI_ISL_828761 to 828762, EPI_ISL_828774 to 828776, EPI_ISL_828779 to 828780, EPI_ISL_828790, EPI_ISL_828802, EPI_ISL_828804, EPI_ISL_828811, EPI_ISL_828826, EPI_ISL_828828, EPI_ISL_828843, EPI_ISL_828862 to 828863, EPI_ISL_828869, EPI_ISL_828876, EPI_ISL_828889 to 828890, EPI_ISL_828892 to 828898, EPI_ISL_828924, EPI_ISL_828931, EPI_ISL_828934, EPI_ISL_828940, EPI_ISL_828946, EPI_ISL_828948, EPI_ISL_828970 to 828971, EPI_ISL_828976 to 828977, EPI_ISL_828992, EPI_ISL_828995 to 828997, EPI_ISL_828999 to 829000, EPI_ISL_829002 to 829007, EPI_ISL_829009 to 829011, EPI_ISL_829049, EPI_ISL_829053, EPI_ISL_829055, EPI_ISL_829059 to 829060, EPI_ISL_829103, EPI_ISL_829105, EPI_ISL_829116, EPI_ISL_829132 to 829133, EPI_ISL_829142, EPI_ISL_829144, EPI_ISL_829153 to 829156, EPI_ISL_829165 to 829166, EPI_ISL_829188 to 829190, EPI_ISL_829192, EPI_ISL_829194, EPI_ISL_829196 to 829197, EPI_ISL_829199, EPI_ISL_829220, EPI_ISL_829223 to 829224, EPI_ISL_829239, EPI_ISL_829275, EPI_ISL_829286, EPI_ISL_829290 to 829293, EPI_ISL_829314 to 829315, EPI_ISL_829317 to 829318, EPI_ISL_829361, EPI_ISL_829364, EPI_ISL_829366, EPI_ISL_829369 to 829372, EPI_ISL_829376 to 829379, EPI_ISL_829381, EPI_ISL_829414, EPI_ISL_829426, EPI_ISL_829449, EPI_ISL_829464, EPI_ISL_829470 to 829471, EPI_ISL_829476 to 829477, EPI_ISL_829512, EPI_ISL_829518, EPI_ISL_829524 to 829526, EPI_ISL_829529, EPI_ISL_829543 to 829544, EPI_ISL_829581, EPI_ISL_829590, EPI_ISL_829601, EPI_ISL_829607 to 829608, EPI_ISL_829617, EPI_ISL_829622 to 829625, EPI_ISL_829675, EPI_ISL_829679, EPI_ISL_829697, EPI_ISL_829707 to 829709, EPI_ISL_829721, EPI_ISL_829725, EPI_ISL_829734 to 829735, EPI_ISL_829759, EPI_ISL_829798 to 829802, EPI_ISL_829849 to 829851, EPI_ISL_829859 to 829860, EPI_ISL_829864, EPI_ISL_829869, EPI_ISL_829881 to 829882, EPI_ISL_829885, EPI_ISL_829887 to 829889, EPI_ISL_829914 to 829916, EPI_ISL_829930 to 829931, EPI_ISL_829934, EPI_ISL_829937 to 829940, EPI_ISL_829948, EPI_ISL_829953, EPI_ISL_829955, EPI_ISL_829957, EPI_ISL_829959, EPI_ISL_829970, EPI_ISL_829989, EPI_ISL_830005, EPI_ISL_830007, EPI_ISL_830019, EPI_ISL_830033 to 830034, EPI_ISL_830037 to 830038, EPI_ISL_830069, EPI_ISL_830072, EPI_ISL_830085 to 830086, EPI_ISL_830093 to 830098, EPI_ISL_830100, EPI_ISL_830102 to 830105, EPI_ISL_830119 to 830120, EPI_ISL_830127 to 830129, EPI_ISL_830131 to 830132, EPI_ISL_830140 to 830146, EPI_ISL_830152, EPI_ISL_830168, EPI_ISL_830177, EPI_ISL_830184 to 830185, EPI_ISL_830191, EPI_ISL_830193, EPI_ISL_830209, EPI_ISL_830213, EPI_ISL_830235, EPI_ISL_830237, EPI_ISL_830240, EPI_ISL_830242, EPI_ISL_830245, EPI_ISL_830264, EPI_ISL_830266, EPI_ISL_830268, EPI_ISL_830305, EPI_ISL_830307, EPI_ISL_830309, EPI_ISL_830311, EPI_ISL_830314, EPI_ISL_830316, EPI_ISL_830320, EPI_ISL_830327 to 830328, EPI_ISL_830354, EPI_ISL_830359, EPI_ISL_830361 to 830364, EPI_ISL_830366 to 830368, EPI_ISL_830377, EPI_ISL_830379 to 830382, EPI_ISL_830387, EPI_ISL_830397, EPI_ISL_830407, EPI_ISL_830410, EPI_ISL_830415, EPI_ISL_830424 to 830425, EPI_ISL_830428 to 830429, EPI_ISL_830445, EPI_ISL_830449, EPI_ISL_830468, EPI_ISL_830471 to 830480, EPI_ISL_830487, EPI_ISL_830492, EPI_ISL_830501 to 830502, EPI_ISL_830519, EPI_ISL_830529 to 830530, EPI_ISL_830535, EPI_ISL_830537, EPI_ISL_830551 to 830555, EPI_ISL_830557 to 830559, EPI_ISL_830568 | | | |
| see above | The National University Hospital of Iceland | deCODE genetics | Agnar Helgason; Alma Moller; Arna B Agustsdottir; Arnaldur Gylfason; Asgeir Sigurdsson; Aslaug Jonasdottir; Berglind Eiriksdottir; Bjarni Thorbjornsson; Brynjar O Jensson; Daniel F Gudbjartsson; Droplaug N Magnusdottir; Elisabet E Gardarsdottir; Emil A Thorarensen; Gardar Sveinbjornsson; Gisli Masson; Gudmundur Georgsson; Gudmundur L Norddahl; Gudrun Sigmundsdottir; Hakon Jonsson; Hannes Eggertsson; Hilma Holm; Ingileif Jonsdottir; Jona Saemundsdottir; Kamilla S Josefsdottir; Kari Stefansson; Karl G Kristinsson; Kjartan R Gudmundsson; Kristin E Sveinsdottir; Kristin E Sveinsdottir; Louise le Roux; Maney Sveinsdottir; Olafia S Gretarsdottir; Olafia S Gretarsdottir; Olafur T Magnusson; Pall Melsted; Patrick Sulem; Run Fridriksdottir; Solvi Rognvaldsson; Thora R Gunnarsdottir; Thordur Kristjansson; Thorolfur Gudnason; Unnur Thorsteinsdottir |
| EPI_ISL_509436 to 509443 | The Princess Alexandra Hospital | Wellcome Sanger Institute for the COVID-19 Genomics UK (COG-UK) consortium | Claudia McCrea and Alex Alderton; Cordelia Langford; David K. Jackson; Dominic Kwiatkowski; Ewan Harrison; Ian Johnston; Jessica Scott; John Sillitoe on behalf of the Wellcome Sanger Institute COVID-19 Surveillance Team (http://www.sanger.ac.uk/covid-team); Louise Lopez; Lynn Monaghan; Nick Levene; Roberto Amato; Sonia Goncalves |
| EPI_ISL_429161 to 429163, EPI_ISL_430862, EPI_ISL_430864, EPI_ISL_648129, EPI_ISL_648132, EPI_ISL_648134 to 648139, EPI_ISL_648143, EPI_ISL_648145 to 648147, EPI_ISL_648149 to 648150, EPI_ISL_648156, EPI_ISL_648165, EPI_ISL_648170 to 648172, EPI_ISL_648174 to 648191, EPI_ISL_648194 to 648197, EPI_ISL_654498 to 654503, EPI_ISL_654506 to 654508, EPI_ISL_710581 to 710583, EPI_ISL_738133, EPI_ISL_766594 to 766607, EPI_ISL_766612, EPI_ISL_766700 to 766706 | | | |
| see above | The Public Health Agency of Sweden | The Public Health Agency of Sweden | Anna Risberg; Anna-Malin Linde; Department of Microbiology; Karin Tegmark-Wisell; Maria Lind Karlberg; Mattias Haukland; Mia Brytting; Olov Svartstrom; Oskar Karlsson Lindsjo; Petra Edquist; Reza Advani; Sandra Broddesson; Shaman Muradrasoli; Sofia Stamouli; The Public Health Agency of Sweden |
| EPI_ISL_451948 | The Republican Research and Practical Center for Epidemiology and Microbiology | Charite Universitatsmedizin Berlin, Institute of Virology | Barbara Muhlemann; Christian Drosten; Jorn Beheim-Schwarzbach; Julia Schneider; Natallia Shmialiova; Natallia Sivets; Talitha Veith; Terry Jones; Victor M Corman |
| EPI_ISL_419692 to 419693 | The Republican Research and Practical Center for Epidemiology and Microbiology | Charite Universitatsmedizin Berlin, Institute of Virology | Barbara Mühlemann; Christian Drosten; Jorn Beheim-Schwarzbach; Julia Schneider; Natallia Shmialiova; Natallia Sivets; Talitha Veith; Terry Jones; Victor M Corman |
| EPI_ISL_754229 to 754235 | The Republican Research and Practical Center for Epidemiology and Microbiology (RRPCEM) | WHO National Influenza Centre Russian Federation | Anatoly Krasko; Andrey Komissarov; Anna Ivanova; Artem Fadeev; Daria Danilenko; Dmitry Bazhenov; Dmitry Lioznov; Elena Gasich; Elena Nabieva; Georgii Bazykin; Kirill Bulda; Ksenia Safina; Kseniya Komissarova |
| EPI_ISL_779711 | The University Hospital Brno | Institute of Applied Biotechnologies a.s. | Martin Kašný; Ondej Brzo; Petr Klempt; Petr Kvapil |
| EPI_ISL_766585 | Triemli Hospital | Institute of Medical Virology, University of Zurich | Alexandra Trkola; Annette Audigé; Cyril Shah; Jon Huder; Jürg Böni; Kevin Steiner; Maria Grünberg; Maryam Zaheri; Michael Huber; Riccarda Capaul; Stefan Schmutz; Verena Kufner |
| EPI_ISL_722208 | Triemli Hospital Zurich | Institute of Medical Virology, University of Zurich | Alexandra Trkola; Daniel Röösli; Gabriela Ziltener; Gerhard Eich; Jürg Böni; Maryam Zaheri; Michael Huber; Stefan Schmutz; Verena Kufner |
| EPI_ISL_434653, EPI_ISL_434656, EPI_ISL_475518 | Trollbackens VC | The Public Health Agency of Sweden | Amelie Holmqvist; Anna Risberg; Anna-Malin Linde; Karin Tegmark-Wisell; Maria Lind Karlberg; Mattias Haukland; Mia Brytting; Olov Svartstrom; Oskar Karlsson Lindsjo; Reza Advani; Sandra Broddesson; Theresa Enkirch |
| EPI_ISL_417432 | UCD National Virus Reference Laboratory | Public Health Wales Microbiology Cardiff | ; Catherine Dempsey; Cillian F De Gascun; Michael Prentice; Olive Murphy |
| EPI_ISL_414487, EPI_ISL_414584 to 414587, EPI_ISL_418516, EPI_ISL_418548, EPI_ISL_418580 to 418584, EPI_ISL_437684 to 437688 | UCD National Virus Reference Laboratory | UCD National Virus Reference Laboratory | Alison Murphy; Brendan Crowley; Brendan Loftus; Cillian F De Gascun; Gabriel Gonzalez; Jeff Connell; Jonathan Dean; Ken Wolfe; Kevin Byrne; Michael Carr; Michael J. Carr; Suzie Coughlan |
| EPI_ISL_877765 | UCO Igiene e Sanità Pubblica, ASUGI, Trieste, Italy; Laboratorio di Virologia Molecolare, ICGEB, Trieste, Italy | Laboratorio di Genomica ed Epigenomica sistema Argo, Area SciencePark, Trieste, Italy; | Alessandro Marcello; Danilo Licastro; Pierlanfranco D'Agaro |
| EPI_ISL_584440 to 584441, EPI_ISL_584443 to 584483, EPI_ISL_584485 to 584501, EPI_ISL_584503 to 584508, EPI_ISL_584510 to 584557, EPI_ISL_584560 to 584564, EPI_ISL_584566 to 584590 | | | |
| see above | UHCW / University of Warwick | COVID-19 Genomics UK (COG-UK) Consortium | Chrystala Constantinidou; Dimitris Grammatopoulos; Grace Taylor-Joyce; Hannah Elizabeth Bridgewater; Jeff Cheng; Laura Baxter; Lucy Frost; Meera Unnikrishnan; Paul Brown; Richard Stark; Sarojini Pandey; Sascha Ott; Tauqeer Alam |
| EPI_ISL_460635 | UHCW Pathology / University of Warwick | University of Warwick, for the COVID-19 Genomics (COG) UK Consortium | Chrystala Constantinidou; Dimitris Grammatopoulos; Grace Taylor-Joyce; Hannah Elizabeth Bridgewater; Jeff Cheng; Laura Baxter; Lucy Frost; Meera Unnikrishnan; Paul Brown; Richard Stark; Sarojini Pandey; Sascha Ott; Tauqeer Alam |
| EPI_ISL_977156, EPI_ISL_977161, EPI_ISL_977242 to 977245, EPI_ISL_977247 to 977250 | ULSS 03 Venezia | Istituto Zooprofilattico Sperimentale delle Venezie | Adelaide Milani; Alessia Schivo; Alice Fusaro; Ambra Pastori; Annalisa Salviato; Antonia Ricci; Bianca Zecchin; Calogero Terregino; Erika Giorgia Quaranta; Isabella Monne |
| EPI_ISL_875551, EPI_ISL_977213 to 977230 | ULSS 1 Dolomiti | Istituto Zooprofilattico Sperimentale delle Venezie | Adelaide Milani; Alessia Schivo; Alice Fusaro; Ambra Pastori; Annalisa Salviato; Antonia Ricci; Bianca Zecchin; Calogero Terregino; Erika Giorgia Quaranta; Isabella Monne |
| EPI_ISL_766580 to 766581, EPI_ISL_875552 to 875556, EPI_ISL_977154, EPI_ISL_977163, EPI_ISL_977188 to 977193, EPI_ISL_977195 to 977199 | ULSS 2 Treviso | Istituto Zooprofilattico Sperimentale delle Venezie | Adelaide Milani; Alessia Schivo; Alice Fusaro; Ambra Pastori; Annalisa Salviato; Antonia Ricci; Bianca Zecchin; Calogero Terregino; Erika Giorgia Quaranta; Isabella Monne |
| EPI_ISL_747477 to 747481, EPI_ISL_747486 to 747489, EPI_ISL_977159, EPI_ISL_977231 to 977239 | ULSS 5 Polesana | Istituto Zooprofilattico Sperimentale delle Venezie | Adelaide Milani; Alessia Schivo; Alice Fusaro; Ambra Pastori; Annalisa Salviato; Antonia Ricci; Bianca Zecchin; Calogero Terregino; Erika Giorgia Quaranta; Isabella Monne |
| EPI_ISL_977165 to 977166, EPI_ISL_977180 to 977187 | ULSS 6 Euganea | Istituto Zooprofilattico Sperimentale delle Venezie | Adelaide Milani; Alessia Schivo; Alice Fusaro; Ambra Pastori; Annalisa Salviato; Antonia Ricci; Bianca Zecchin; Calogero Terregino; Erika Giorgia Quaranta; Isabella Monne |
| EPI_ISL_766571 to 766573, EPI_ISL_977203 to 977206 | ULSS 7 Pedemontana - Distretto 1 | Istituto Zooprofilattico Sperimentale delle Venezie | Adelaide Milani; Alessia Schivo; Alice Fusaro; Ambra Pastori; Annalisa Salviato; Antonia Ricci; Bianca Zecchin; Calogero Terregino; Erika Giorgia Quaranta; Isabella Monne |
| EPI_ISL_794826 to 794828, EPI_ISL_977200 to 977202 | ULSS 7 Pedemontana - Distretto 2 | Istituto Zooprofilattico Sperimentale delle Venezie | Adelaide Milani; Alessia Schivo; Alice Fusaro; Ambra Pastori; Annalisa Salviato; Antonia Ricci; Bianca Zecchin; Calogero Terregino; Erika Giorgia Quaranta; Isabella Monne |
| EPI_ISL_766574 to 766579, EPI_ISL_977194 | ULSS 8 Berica | Istituto Zooprofilattico Sperimentale delle Venezie | Adelaide Milani; Alessia Schivo; Alice Fusaro; Ambra Pastori; Annalisa Salviato; Antonia Ricci; Bianca Zecchin; Calogero Terregino; Erika Giorgia Quaranta; Isabella Monne |
| EPI_ISL_636467 to 636470, EPI_ISL_636474 to 636475 | ULSS6 Distretto Padova Terme Colli | Istituto Zooprofilattico Sperimentale delle Venezie | Adelaide Milani; Alessia Schivo; Alice Fusaro; Ambra Pastori; Annalisa Salviato; Antonia Ricci; Bianca Zecchin; Calogero Terregino; Erika Giorgia Quaranta; Isabella Monne |
| EPI_ISL_636462 to 636463, EPI_ISL_636465 to | ULSS6 Euganea | Istituto Zooprofilattico Sperimentale delle Venezie | Adelaide Milani; Alessia Schivo; Alice Fusaro; Ambra Pastori; Annalisa Salviato; Antonia Ricci; Bianca Zecchin; Calogero Terregino; Erika Giorgia |

| | | | |
|---|---|---|---|
| 636466 | | | Quaranta; Isabella Monne |
| EPI_ISL_636464, EPI_ISL_636471 to 636473 | ULSS6 Piove di Sacco | Istituto Zooprofilattico Sperimentale delle Venezie | Adelaide Milani; Alessia Schivo; Alice Fusaro; Ambra Pastori; Annalisa Salviato; Antonia Ricci; Bianca Zecchin; Calogero Terregino; Erika Giorgia Quaranta; Isabella Monne |
| EPI_ISL_422437 to 422438, EPI_ISL_452181 to 452189, EPI_ISL_522855, EPI_ISL_522860 to 522868, EPI_ISL_747471 to 747476 | ULSS9 Distretto di Bussolengo | Istituto Zooprofilattico Sperimentale delle Venezie | Adelaide Milani; Alessia Schivo; Alice Fusaro; Ambra Pastori; Annalisa Salviato; Antonia Ricci; Bianca Zecchin; Calogero Terregino; Erika Giorgia Quaranta; Isabella Monne |
| EPI_ISL_452190 to 452191, EPI_ISL_522856, EPI_ISL_522858 | ULSS9 Distretto di San Bonifacio | Istituto Zooprofilattico Sperimentale delle Venezie | Adelaide Milani; Alessia Schivo; Alice Fusaro; Ambra Pastori; Annalisa Salviato; Antonia Ricci; Bianca Zecchin; Calogero Terregino; Erika Giorgia Quaranta; Isabella Monne |
| EPI_ISL_522857, EPI_ISL_522859, EPI_ISL_636488 | ULSS9 Scaligera | Istituto Zooprofilattico Sperimentale delle Venezie | Adelaide Milani; Alessia Schivo; Alice Fusaro; Ambra Pastori; Annalisa Salviato; Antonia Ricci; Bianca Zecchin; Calogero Terregino; Erika Giorgia Quaranta; Isabella Monne |
| EPI_ISL_560308 to 560317, EPI_ISL_732990 to 733004, EPI_ISL_733223 to 733240, EPI_ISL_733405 to 733416, EPI_ISL_770638, EPI_ISL_770642, EPI_ISL_770647 to 770652, EPI_ISL_770654 to 770657, EPI_ISL_770659, EPI_ISL_770662 to 770665, EPI_ISL_770667 to 770668, EPI_ISL_770671 to 770672, EPI_ISL_869242 to 869246, EPI_ISL_869248 to 869249, EPI_ISL_869251 to 869261, EPI_ISL_872569 | | | |
| see above | UMMC-Health | WHO National Influenza Centre Russian Federation | Andrey Komissarov; Anna Ivanova; Artem Fadeev; Daria Danilenko; Dmitry Bazhenov; Dmitry Lioznov; Elena Nabieva; Georgii Bazykin; Ksenia Safina; Kseniya Komissarova; Mikhail Bakaev; Tatiana Platonova |
| EPI_ISL_510530 | UMR 190 - Faculte de medecine, UMR 'Emergence des Pathologies Virales' (EPV: Aix-Marseille University - IRD 190 - Inserm 1207 - EHE | UMR 190 - Faculte de medecine, UMR 'Emergence des Pathologies Virales' (EPV: Aix-Marseille University - IRD 190 - Inserm 1207 - EHE | De Laval, F.; Durand; F.-X.; G.A.; Geulen, M.; Gilles, M.; Le Flem; Leparc Goffart, I.; Peduzzi, F.; Piorkowski, G.; Pommier De Santi, V. |
| EPI_ISL_582123, EPI_ISL_583953 to 583965 | UOC Microbiologia e Virologia, Azienda Ospedaliera Universitaria Senese, Siena, Italy | Dipartimento di Biotecnologie Mediche | Claudia Gandolfo; David Pinzauti; Francesco Santoro; Gabriele Anichini; Gianni Pozzi; Maria Grazia Cusi |
| EPI_ISL_833124 to 833128 | USCA Sulmona | Istituto Zooprofilattico Sperimentale dell'Abruzzo e Molise "G. Caporale" | Ancora M; Calistri P; Cammà C; Curini V; Delli Compagni E; Di Domenico M; Di Pasquale A; Lorusso A; Mangone I; Marcacci M; Puglia I; Rinaldi A; Savini G |
| EPI_ISL_833129 | USCA Tagliacozzo | Istituto Zooprofilattico Sperimentale dell'Abruzzo e Molise "G. Caporale" | Ancora M; Calistri P; Cammà C; Curini V; Delli Compagni E; Di Domenico M; Di Pasquale A; Lorusso A; Mangone I; Marcacci M; Puglia I; Rinaldi A; Savini G |
| EPI_ISL_833253 | USCA-Avezzano | Istituto Zooprofilattico Sperimentale dell'Abruzzo e Molise "G. Caporale" | Ancora M; Calistri P; Cammà C; Curini V; Di Domenico M; Di Pasquale A; Lorusso A; Mangone I; Marcacci M; Puglia I; Rinaldi A; Savini G |
| EPI_ISL_833254 | USCA-Pescina | Istituto Zooprofilattico Sperimentale dell'Abruzzo e Molise "G. Caporale" | Ancora M; Calistri P; Cammà C; Curini V; Di Domenico M; Di Pasquale A; Lorusso A; Mangone I; Marcacci M; Puglia I; Rinaldi A; Savini G |
| EPI_ISL_833277 | USCA-Sulmona | Istituto Zooprofilattico Sperimentale dell'Abruzzo e Molise "G. Caporale" | Ancora M; Calistri P; Cammà C; Curini V; Di Domenico M; Di Pasquale A; Lorusso A; Mangone I; Marcacci M; Puglia I; Rinaldi A; Savini G |
| EPI_ISL_734482 to 734726, EPI_ISL_734728 to 734755, EPI_ISL_734757 to 734928, EPI_ISL_734930 to 734932, EPI_ISL_734934 to 735079, EPI_ISL_735081 to 735093, EPI_ISL_735095 to 735101, EPI_ISL_735103 to 735119, EPI_ISL_735121 to 735249, EPI_ISL_738195 to 738207, EPI_ISL_738209 to 738242, EPI_ISL_738360 to 738416, EPI_ISL_738418 to 738499, EPI_ISL_745222, EPI_ISL_791332 to 791333, EPI_ISL_794321 to 794323 | | | |
| see above | UZ Leuven, National Reference Laboratory for Coronaviruses, Laboratory Medicine, Leuven, Belgium | KU Leuven, Rega Institute, Clinical and Epidemiological Virology | Bert Vanmechelen; Joan Marti-Carerras; Piet Maes; Tony Wawina-Bokalanga |
| EPI_ISL_833433, EPI_ISL_833456 | UZA/UAntwerpen (BIS lab), University Hospital Antwerp, Edegem, Belgium | Laboratory of Medical Microbiology, University of Antwerp, Campus Drie Eiken, S6.26, Universiteitsplein 1, 2610, Wilrijk, Antwerp, Belgium | Basil Britto Xavier; Christine Lammens; Herman Goossens; Jasmine Coppens; Sahaya Glingston Rajakani; Veerle Matheeussen |
| EPI_ISL_434646, EPI_ISL_434667, EPI_ISL_445228, EPI_ISL_445240, EPI_ISL_450824, EPI_ISL_455104, EPI_ISL_455109, EPI_ISL_469063, EPI_ISL_469072, EPI_ISL_475521, EPI_ISL_476138 | | | |
| see above | Ulltuna Vardcentral | The Public Health Agency of Sweden | Anna Risberg; Anna-Malin Linde; Heidi Lindback; Karin Tegmark-Wisell; Maria Lind Karlberg; Mattias Haukland; Mia Brytting; Olov Svartstrom; Oskar Karlsson Lindsjo; Petra Edquist; Reza Advani; Sandra Broddesson; Shamam Muradrasoli; Theresa Enkirch |
| EPI_ISL_475122, EPI_ISL_475126 to 475127, EPI_ISL_475143, EPI_ISL_615092 to 615096 | Umea klinisk mikrobiologi | The Public Health Agency of Sweden | Anna Risberg; Anna-Malin Linde; Karin Tegmark-Wisell; Maria Lind Karlberg; Mattias Haukland; Mia Brytting; Olov Svartstrom; Oskar Karlsson Lindsjo; Petra Edquist; Reza Advani; Sandra Broddesson; Shamam Muradrasoli |
| EPI_ISL_661294 to 661298 | Unilabs | The Public Health Agency of Sweden | Department of Microbiology; The Public Health Agency of Sweden |
| EPI_ISL_775585 to 775586, EPI_ISL_789054 | Unilabs AB | The Public Health Agency of Sweden | Department of Microbiology; The Public Health Agency of Sweden |
| EPI_ISL_510827 to 510830, EPI_ISL_582794 to 582796 | Unilabs Eskilstuna | The Public Health Agency of Sweden | Anna Risberg; Anna-Malin Linde; Karin Tegmark-Wisell; Maria Lind Karlberg; Mattias Haukland; Mia Brytting; Olov Svartstrom; Oskar Karlsson Lindsjo; Petra Edquist; Reza Advani; Sandra Broddesson |
| EPI_ISL_420143, EPI_ISL_471172, EPI_ISL_549050, EPI_ISL_549123, EPI_ISL_549125, EPI_ISL_549172, EPI_ISL_549175, EPI_ISL_590882, EPI_ISL_590911, EPI_ISL_590917, EPI_ISL_591010, EPI_ISL_591013, EPI_ISL_635104, EPI_ISL_635132, EPI_ISL_635153 to 635154, EPI_ISL_635158, EPI_ISL_635162, EPI_ISL_635191, EPI_ISL_668409, EPI_ISL_708046 to 708048, EPI_ISL_708060 to 708064, EPI_ISL_708147 to 708150, EPI_ISL_738314, EPI_ISL_775305, EPI_ISL_775349 to 775352, EPI_ISL_775358 to 775361, EPI_ISL_775384 to 775385, EPI_ISL_775406, EPI_ISL_775463, EPI_ISL_775468 to 775471, EPI_ISL_775476 to 775478, EPI_ISL_775495, EPI_ISL_775516 to 775517, EPI_ISL_796673, EPI_ISL_796679 to 796680, EPI_ISL_796686, EPI_ISL_796700 to 796702, EPI_ISL_796705, EPI_ISL_860270 | | | |
| see above | Unilabs Laboratory Medicine | Norwegian Institute of Public Health, Department of Virology | Atiya R Ali; Hilde Elshaug; Hilde Synnøve Vollan; Hilde Vollan; Kamilla Heddeland Instefjord; Karoline Bragstad; Kathrine Stene-Johansen; Marie Paulsen Madsen; Olav Hungnes; Rasmus Riis Kopperud |
| EPI_ISL_429126 to 429127, EPI_ISL_450835 | Unilabs Skovde | The Public Health Agency of Sweden | Anna Risberg; Anna-Malin Linde; Helena Enroth; Karin Tegmark-Wisell; Maria Lind Karlberg; Mia Brytting; Olov Svartstrom; Oskar Karlsson Lindsjo; Shaman Muradrasoli; Theresa Enkirch; Tobias Kollberg |
| EPI_ISL_832000, EPI_ISL_832008 | Unilabs, Mikrobiologiska laboratoriet | The Public Health Agency of Sweden | Department of Microbiology; The Public Health Agency of Sweden |
| EPI_ISL_762999 to 763016, EPI_ISL_763018 to 763019, EPI_ISL_763021 to 763025, EPI_ISL_763027 to 763028, EPI_ISL_763030 to 763033, EPI_ISL_763036 to 763044, EPI_ISL_763046 to 763048, EPI_ISL_763050 to 763061, EPI_ISL_765893 to 765895, EPI_ISL_860298 to 860303, EPI_ISL_860316 | | | |
| see above | Unit 17: Influenza & Other Respiratory Viruses, German National Influenza Center | Project group Epidemiology of Highly Pathogenic Microorganisms, Robert Koch-Institute | Andreas Sachse; Ariane Düx; Djin-Ye Oh; Essia Belarbi; Fabian Leendertz; Grit Schubert; Marianne Wedde; Ralf Dürrwald; Sébastien Calvignac-Spencer; Thorsten Wolff |
| EPI_ISL_411951 | Unit for Laboratory Development and Technology Transfer, Public Health Agency of Sweden | Unit for Laboratory Development and Technology Transfer, Public Health Agency of Sweden | Appelberg, S.; Bengner, M.; Brave, A.; Lind Karlberg, M.; Lindsjo, O.; Monteil, V.; Muradrasoli; Palmerus, M.; S. and Tegmark-Wisell, K. |
| EPI_ISL_882882 to 882883, EPI_ISL_882896 | Unit of Infectious Diseases, Azienda Socio Sanitaria Territoriale di Cremona | Laboratory of Infectious Diseases, Department of Biomedical and Clinical Sciences L. Sacco, University of Milan | Alessia Lai; Alice Ferraresi; Annalisa Bergna; Antonio Coluccello; Carla Della Ventura; Chiara Fornabaio; Claudia Balotta; Fabio Sagradi; Gianguglielmo Zehender on behalf of SARS-CoV-2 ITALIAN RESEARCH ENTERPRISE-(SCIRE) Collaborative Group; Massimo Galli; Matteo Giorgi Pierfranceschi; Sophie Testa |
| EPI_ISL_510527 | Unite des virus emergents, UMR190 | Unite des virus emergents, UMR190 | Baronti, C.; Charrel; Coutard, B.; Piorkowski, G.; R. and de Lamballerie, X. |
| EPI_ISL_644703 to 644714, EPI_ISL_645212 to 645214, EPI_ISL_649188, EPI_ISL_660732 to 660741 | Unité des Virus Émergents | CNR Virus des Infections Respiratoires - France SUD | Antoine Nougairède; Antonin Bal; Bruno Lina; Gregory Destras; Gwendolyne Burfin; Hadrien Règue; Laetitia Ninove; Laurence Josset; Léa Luciani; Martine Valette; Quentin Semanas |
| EPI_ISL_537288 to 537297, EPI_ISL_537299 to 537302, EPI_ISL_537304 to 537328, EPI_ISL_537330 to 537379, EPI_ISL_537609 to 537627, EPI_ISL_537629, EPI_ISL_537631 to 537635, EPI_ISL_537637 to 537662, EPI_ISL_537664 to 537669, EPI_ISL_537671 to 537680, EPI_ISL_537682, EPI_ISL_537684, EPI_ISL_537686 to 537689 | | | |
| see above | Universidad de León | SeqCOVID-SPAIN consortium/IBV(CSIC) | Ana Carvajal; Antonio J. Molina and SeqCOVID-SPAIN consortium; Héctor Argüello; Juan M. Fregeneda; Tania Fernández-Villa; Vicente Martín |
| EPI_ISL_437933 to 437958, EPI_ISL_437960 to 437966, EPI_ISL_437968 to 437973, EPI_ISL_475764, EPI_ISL_475766 to 475768, EPI_ISL_475929 to 475936, EPI_ISL_583557 to 583559, EPI_ISL_583561 to 583562, EPI_ISL_583617 to 583618 | | | |
| see above | Universitaetsklinik für Innere Medizin II Innsbruck | Bergthaler laboratory, CeMM Research Center for Molecular | Adi Steinrigl; Alexander Lercher; Alexandra Popa; Andreas Bergthaler; Benedikt Agerer; Christian Paar; Christoph Bock; Daniela Schmid; Dorothee von |

| | | | |
|---|---|---|---|
| | | Medicine of the Austrian Academy of Sciences | Laer; Elisabeth Puchhammer-Stoeckl; Franz Allerberger; Gernot Walder; Gregor Hörmann; Guenter Weiss; Gunther Vogl; Henrique Colaco; Jakob-Wendelin Genger; Jan Laine; Judith Aberle; Kinga Rigler-Hohenwarter; Lukas Endler; Manfred Nairz; Mark Smyth; Martin Senekowitsch; Michael Schuster; Peter Hufnagl; Peter Obrist; Rainer Gattringer; Sabine Sussitz-Rack; Stephan Aberle; Thomas Penz; Wegene Borena |
| EPI_ISL_534224, EPI_ISL_534246 to 534247, EPI_ISL_560977 to 560979 | Universitetssjukhuset i Linkoping | The Public Health Agency of Sweden | Anna Risberg; Anna-Malin Linde; Karin Tegmark-Wisell; Maria Lind Karlberg; Mattias Haukland; Mia Brytting; Olov Svartstrom; Oskar Karlsson Lindsjo; Petra Edquist; Reza Advani; Sandra Broddesson |
| EPI_ISL_462990 | University Clinical Centre of the Republic of Srpska | University of Sarajevo, Veterinary Faculty | Aida, K.; Alma; Maja, T.; Mirsada, H.; Teufik, G.; Toni, E.; Š.A.; Šejla, G. |
| EPI_ISL_462753 | University Clinical Hospital of Mostar | University of Sarajevo Veterinary Faculty | Eterovic, T.; Goletic, S.; Goletic, T.; Hukic, M.; Ostojic, M.; Seho-Alic, A.; Softic, A. |
| EPI_ISL_507040 to 507109, EPI_ISL_664125, EPI_ISL_664147, EPI_ISL_664171, EPI_ISL_664187 to 664189, EPI_ISL_664196 to 664197, EPI_ISL_664225, EPI_ISL_664228, EPI_ISL_664233, EPI_ISL_664236, EPI_ISL_664281, EPI_ISL_664335, EPI_ISL_664340, EPI_ISL_664342, EPI_ISL_664352 to 664353, EPI_ISL_664356, EPI_ISL_664359, EPI_ISL_664361, EPI_ISL_664367, EPI_ISL_664372, EPI_ISL_664427, EPI_ISL_664438, EPI_ISL_664447, EPI_ISL_664449, EPI_ISL_664471 to 664472, EPI_ISL_664479, EPI_ISL_664490, EPI_ISL_664493 to 664494, EPI_ISL_664497 to 664500, EPI_ISL_664505, EPI_ISL_664507, EPI_ISL_664510, EPI_ISL_664512, EPI_ISL_664517, EPI_ISL_664521, EPI_ISL_664525, EPI_ISL_664540, EPI_ISL_664605, EPI_ISL_664618, EPI_ISL_664651, EPI_ISL_664669, EPI_ISL_664672 to 664673, EPI_ISL_664694, EPI_ISL_664701 to 664703, EPI_ISL_664707, EPI_ISL_664711 to 664712, EPI_ISL_664723 to 664724, EPI_ISL_664728, EPI_ISL_664734, EPI_ISL_664742, EPI_ISL_664744 to 664745, EPI_ISL_664753 to 664755, EPI_ISL_664839, EPI_ISL_665143 to 665253, EPI_ISL_665287, EPI_ISL_665297, EPI_ISL_665299, EPI_ISL_665320, EPI_ISL_665334, EPI_ISL_665352, EPI_ISL_665374, EPI_ISL_665376, EPI_ISL_665382, EPI_ISL_665466, EPI_ISL_665517, EPI_ISL_665525, EPI_ISL_665530, EPI_ISL_665559, EPI_ISL_665572, EPI_ISL_665580, EPI_ISL_665597, EPI_ISL_665611, EPI_ISL_665632, EPI_ISL_665634 to 665637, EPI_ISL_665643 to 665645, EPI_ISL_665648 to 665649, EPI_ISL_665757 to 665759, EPI_ISL_665785 to 665786, EPI_ISL_665801, EPI_ISL_665805 to 665848, EPI_ISL_679406 to 679446, EPI_ISL_702507, EPI_ISL_702602, EPI_ISL_702669, EPI_ISL_702862, EPI_ISL_702961, EPI_ISL_703222, EPI_ISL_703340, EPI_ISL_703369, EPI_ISL_703886, EPI_ISL_703966, EPI_ISL_704041, EPI_ISL_704052, EPI_ISL_704102, EPI_ISL_704105, EPI_ISL_704607 to 704608, EPI_ISL_704666, EPI_ISL_704669, EPI_ISL_704671, EPI_ISL_704674, EPI_ISL_705247, EPI_ISL_705378, EPI_ISL_705472, EPI_ISL_705942 to 705960, EPI_ISL_842653 to 842656, EPI_ISL_842658 to 842672, EPI_ISL_842674 to 842680, EPI_ISL_842682 to 842687, EPI_ISL_842689 to 842691, EPI_ISL_842693 to 842694, EPI_ISL_842696 to 842703, EPI_ISL_842705 to 842708, EPI_ISL_842710 to 842715, EPI_ISL_842717 to 842721, EPI_ISL_842723, EPI_ISL_842725 to 842726, EPI_ISL_842728 to 842744, EPI_ISL_842746, EPI_ISL_842749 to 842759, EPI_ISL_842762 to 842766, EPI_ISL_842769 to 842774, EPI_ISL_842776, EPI_ISL_842778 to 842779, EPI_ISL_842781 to 842784, EPI_ISL_842786 to 842789, EPI_ISL_842791 to 842792, EPI_ISL_866048 to 866074, EPI_ISL_866106 to 866107, EPI_ISL_866115 to 866116, EPI_ISL_866118 to 866119, EPI_ISL_866121 to 866124, EPI_ISL_866127, EPI_ISL_866129 to 866133, EPI_ISL_866135 to 866141, EPI_ISL_866145, EPI_ISL_866148 to 866149, EPI_ISL_866151, EPI_ISL_866162, EPI_ISL_866164, EPI_ISL_866168, EPI_ISL_866179, EPI_ISL_866183 | | | |
| see above | University College London Hospital | COVID-19 Genomics UK (COG-UK) Consortium | Catherine Houlihan; Dan Frampton; Judith Heaney; Matthew Byott; Moira Spyer and Eleni Nastouli; Stuart Kirk |
| EPI_ISL_440951 to 440976, EPI_ISL_440978 to 440991, EPI_ISL_440993 to 441043, EPI_ISL_441045 to 441051, EPI_ISL_441350, EPI_ISL_441352 to 441354, EPI_ISL_444079 to 444081, EPI_ISL_444083 to 444106, EPI_ISL_444108 to 444135, EPI_ISL_444137 to 444144, EPI_ISL_444146 to 444147, EPI_ISL_444149 to 444153, EPI_ISL_444155 to 444157, EPI_ISL_444159 to 444220, EPI_ISL_444222 to 444265, EPI_ISL_444267 to 444272, EPI_ISL_449626 to 449630, EPI_ISL_449632 to 449716, EPI_ISL_449718 to 449729, EPI_ISL_453460 to 453464, EPI_ISL_457270 to 457302, EPI_ISL_461763 to 461772, EPI_ISL_478404 to 478406, EPI_ISL_478408 to 478464, EPI_ISL_478466 to 478468, EPI_ISL_478470, EPI_ISL_478472, EPI_ISL_572684 to 572685, EPI_ISL_572709, EPI_ISL_572725, EPI_ISL_572770, EPI_ISL_572800, EPI_ISL_572807 to 572809, EPI_ISL_572828 to 572829, EPI_ISL_572837 to 572838, EPI_ISL_572957, EPI_ISL_573478 to 573500, EPI_ISL_573502 to 573525, EPI_ISL_573652 to 573670, EPI_ISL_573672 to 573682, EPI_ISL_573684 to 573685, EPI_ISL_573764, EPI_ISL_573800 to 573805, EPI_ISL_573856 to 573865, EPI_ISL_574258, EPI_ISL_576984 to 576985, EPI_ISL_584670 to 584679, EPI_ISL_584811 to 584813, EPI_ISL_584815 to 584846, EPI_ISL_585502 to 585503, EPI_ISL_594861 to 594888, EPI_ISL_611523, EPI_ISL_611527, EPI_ISL_611558 to 611559, EPI_ISL_611595 to 611597, EPI_ISL_611622 to 611623, EPI_ISL_611665, EPI_ISL_611822 to 611823, EPI_ISL_611996, EPI_ISL_612000, EPI_ISL_612014, EPI_ISL_612078 to 612079, EPI_ISL_612103, EPI_ISL_612105, EPI_ISL_612131, EPI_ISL_612198, EPI_ISL_612450 to 612457, EPI_ISL_627082, EPI_ISL_627104, EPI_ISL_627108, EPI_ISL_627110, EPI_ISL_627175, EPI_ISL_627211, EPI_ISL_627450 to 627481, EPI_ISL_627491, EPI_ISL_637912, EPI_ISL_637937, EPI_ISL_637995, EPI_ISL_637999, EPI_ISL_638003, EPI_ISL_638022, EPI_ISL_650123, EPI_ISL_651132, EPI_ISL_651179, EPI_ISL_665266, EPI_ISL_665270, EPI_ISL_665335, EPI_ISL_665339, EPI_ISL_665413, EPI_ISL_665428, EPI_ISL_665661, EPI_ISL_665674, EPI_ISL_665720, EPI_ISL_665733, EPI_ISL_665741, EPI_ISL_679447 to 679478, EPI_ISL_679482 to 679484, EPI_ISL_679486 to 679497, EPI_ISL_679499 to 679503, EPI_ISL_679505 to 679518, EPI_ISL_679520 to 679548, EPI_ISL_679550 to 679554, EPI_ISL_679556 to 679562, EPI_ISL_679564, EPI_ISL_679566 to 679570, EPI_ISL_679572 to 679575, EPI_ISL_679581, EPI_ISL_679584 to 679585, EPI_ISL_679587 to 679590, EPI_ISL_679592 to 679595, EPI_ISL_679597 to 679598, EPI_ISL_679600 to 679602, EPI_ISL_679604 to 679607, EPI_ISL_679610 to 679611, EPI_ISL_679613, EPI_ISL_679615 to 679628, EPI_ISL_702544, EPI_ISL_702610, EPI_ISL_702690, EPI_ISL_702731, EPI_ISL_702824, EPI_ISL_702831, EPI_ISL_702839, EPI_ISL_702930, EPI_ISL_702999, EPI_ISL_703069, EPI_ISL_703107, EPI_ISL_703179, EPI_ISL_703319, EPI_ISL_703332, EPI_ISL_703335, EPI_ISL_703383, EPI_ISL_703385, EPI_ISL_703706, EPI_ISL_703739 to 703740, EPI_ISL_703745, EPI_ISL_703767, EPI_ISL_703865, EPI_ISL_703879, EPI_ISL_704025, EPI_ISL_704036, EPI_ISL_704046, EPI_ISL_704084, EPI_ISL_704086, EPI_ISL_704092, EPI_ISL_704163, EPI_ISL_704179, EPI_ISL_704193, EPI_ISL_704312, EPI_ISL_704372, EPI_ISL_704955, EPI_ISL_704963, EPI_ISL_704965, EPI_ISL_705018, EPI_ISL_705035, EPI_ISL_705072, EPI_ISL_705078, EPI_ISL_705132, EPI_ISL_705143, EPI_ISL_705215 to 705216, EPI_ISL_705243, EPI_ISL_705268, EPI_ISL_705278, EPI_ISL_705283, EPI_ISL_705285, EPI_ISL_705297, EPI_ISL_705331, EPI_ISL_705337 to 705338, EPI_ISL_705342, EPI_ISL_705368, EPI_ISL_705397, EPI_ISL_705425, EPI_ISL_705961 to 705971, EPI_ISL_724541 to 724560, EPI_ISL_724562 to 724569, EPI_ISL_724572 to 724575, EPI_ISL_724577 to 724580, EPI_ISL_724582 to 724592, EPI_ISL_724594, EPI_ISL_724596, EPI_ISL_724598 to 724605, EPI_ISL_724607, EPI_ISL_724613, EPI_ISL_724615, EPI_ISL_724617 to 724619, EPI_ISL_724621, EPI_ISL_724624 to 724631, EPI_ISL_724635 to 724642, EPI_ISL_724644 to 724649, EPI_ISL_724652 to 724657, EPI_ISL_724659 to 724660, EPI_ISL_724662 to 724664, EPI_ISL_724668 to 724672, EPI_ISL_724675 to 724677, EPI_ISL_724679 to 724683, EPI_ISL_724685, EPI_ISL_724687 to 724691, EPI_ISL_724693 to 724696, EPI_ISL_724699 to 724702, EPI_ISL_724704 to 724706, EPI_ISL_724708, EPI_ISL_724710 to 724717, EPI_ISL_724719 to 724725, EPI_ISL_724727 to 724733, EPI_ISL_724735 to 724736, EPI_ISL_724738, EPI_ISL_724740 to 724743, EPI_ISL_724745 to 724748, EPI_ISL_724750, EPI_ISL_724752 to 724755, EPI_ISL_724757 to 724759, EPI_ISL_724761 to 724764, EPI_ISL_724767, EPI_ISL_724769, EPI_ISL_724771 to 724772, EPI_ISL_724774 to 724777, EPI_ISL_741186 to 741193, EPI_ISL_741195 to 741214, EPI_ISL_741216 to 741287, EPI_ISL_741289, EPI_ISL_741291 to 741323, EPI_ISL_741325 to 741332, EPI_ISL_741334 to 741346, EPI_ISL_763381, EPI_ISL_763393, EPI_ISL_763397 to 763398, EPI_ISL_763404, EPI_ISL_763442, EPI_ISL_763455, EPI_ISL_763468, EPI_ISL_763485, EPI_ISL_763491, EPI_ISL_763500, EPI_ISL_763510, EPI_ISL_763520, EPI_ISL_763540, EPI_ISL_763544 to 763545, EPI_ISL_763551 to 763552, EPI_ISL_763567, EPI_ISL_763578, EPI_ISL_763582, EPI_ISL_763628, EPI_ISL_763640, EPI_ISL_763643 to 763644, EPI_ISL_763682, EPI_ISL_763687, EPI_ISL_763700, EPI_ISL_763720, EPI_ISL_763728, EPI_ISL_763730, EPI_ISL_763745, EPI_ISL_763756, EPI_ISL_763767, EPI_ISL_763772, EPI_ISL_763784 to 763785, EPI_ISL_763809 to 763811, EPI_ISL_763813, EPI_ISL_763826, EPI_ISL_764037 to 764060, EPI_ISL_764131, EPI_ISL_764146, EPI_ISL_764160, EPI_ISL_764175, EPI_ISL_764188, EPI_ISL_764228, EPI_ISL_764238, EPI_ISL_764241, EPI_ISL_764384 to 764386, EPI_ISL_813832 to 813868, EPI_ISL_813870 to 813913, EPI_ISL_813915 to 813918, EPI_ISL_813920 to 813923, EPI_ISL_813925, EPI_ISL_813927 to 813970, EPI_ISL_819462 to 819463, EPI_ISL_819467 to 819475, EPI_ISL_819477, EPI_ISL_819483 to 819484, EPI_ISL_819486, EPI_ISL_819490 to 819492, EPI_ISL_819494 to 819498, EPI_ISL_819500 to 819502, EPI_ISL_819508 to 819519, EPI_ISL_819526 to 819528, EPI_ISL_819530, EPI_ISL_819533 to 819536, EPI_ISL_819539 to 819550, EPI_ISL_819552 to 819553, EPI_ISL_819555 to 819557, EPI_ISL_819559 to 819563, EPI_ISL_819565 to 819567, EPI_ISL_819873 to 819876, EPI_ISL_819878 to 819884, EPI_ISL_819888 to 819889, EPI_ISL_819892 to 819893, EPI_ISL_819919 to 819927, EPI_ISL_819929 to 819930, EPI_ISL_819936 to 819941, EPI_ISL_819943, EPI_ISL_819946 to 819952, EPI_ISL_819954 to 819955, EPI_ISL_819957 to 819958, EPI_ISL_819961 to 819962, EPI_ISL_819964 to 819969, EPI_ISL_819978 to 819989, EPI_ISL_819991 to 819995, EPI_ISL_819997 to 820000, EPI_ISL_820007 to 820010, EPI_ISL_820012 to 820014, EPI_ISL_820016 to 820020, EPI_ISL_820024, EPI_ISL_820031 to 820040, EPI_ISL_820045 to 820054, EPI_ISL_820057 to 820061, EPI_ISL_820063 to 820066, EPI_ISL_820081, EPI_ISL_820083 to 820087, EPI_ISL_820089, EPI_ISL_820094, EPI_ISL_820099 to 820101, EPI_ISL_820109, EPI_ISL_820111, EPI_ISL_820113 to 820118, EPI_ISL_820120 to 820123, EPI_ISL_820125, EPI_ISL_820127 to 820128, EPI_ISL_820138, EPI_ISL_820204, EPI_ISL_820206, EPI_ISL_820209, EPI_ISL_820211, EPI_ISL_820213, EPI_ISL_820216, EPI_ISL_820218, EPI_ISL_820226, EPI_ISL_820229, EPI_ISL_820236, EPI_ISL_820246, EPI_ISL_820249, EPI_ISL_820251, EPI_ISL_820256, EPI_ISL_820264, EPI_ISL_820266, EPI_ISL_820268, EPI_ISL_820271, EPI_ISL_820273, EPI_ISL_820276, EPI_ISL_820282, EPI_ISL_820314, EPI_ISL_820316, EPI_ISL_820318, EPI_ISL_820321, EPI_ISL_820326, EPI_ISL_820328, EPI_ISL_820331, EPI_ISL_820333, EPI_ISL_820335, EPI_ISL_820337, EPI_ISL_820340, EPI_ISL_820343, EPI_ISL_820345, EPI_ISL_820347, EPI_ISL_820350, EPI_ISL_820376, EPI_ISL_820381, EPI_ISL_820383, EPI_ISL_820385, EPI_ISL_820396, EPI_ISL_820420, EPI_ISL_838673 to 838709, EPI_ISL_838711 to 838716, EPI_ISL_838718 to 838760, EPI_ISL_838762 to 838764, EPI_ISL_838766 to 838777, EPI_ISL_838780 to 838782, EPI_ISL_838784 to 838786, EPI_ISL_838789 to 838797, EPI_ISL_838799 to 838806, EPI_ISL_838808 to 838817, EPI_ISL_838819 to 838827, EPI_ISL_839041 to 839043, EPI_ISL_839063 to 839069, EPI_ISL_839080 to 839084, EPI_ISL_839101 to 839114, EPI_ISL_839116, EPI_ISL_839215, EPI_ISL_839218, EPI_ISL_839220, EPI_ISL_839222 to 839223, EPI_ISL_839226 to 839227, EPI_ISL_839305 to 839308, EPI_ISL_839310, EPI_ISL_839313 to 839314, EPI_ISL_839316, EPI_ISL_839318, EPI_ISL_839320 to 839323, EPI_ISL_839325 to 839328, EPI_ISL_839330 to 839332, EPI_ISL_839334 to 839336, EPI_ISL_839338 to 839340, EPI_ISL_839342, EPI_ISL_839344 to 839357, EPI_ISL_865682 to 865704, EPI_ISL_865706 to 865759, EPI_ISL_865761 to 865772, EPI_ISL_865775 to 865794, EPI_ISL_865796 to 865812, EPI_ISL_865814 to 865821, EPI_ISL_865823 to 865840, EPI_ISL_865843, EPI_ISL_865845 to 865846, EPI_ISL_865892 to 865894, EPI_ISL_865896, EPI_ISL_865898 to 865899, EPI_ISL_865903, EPI_ISL_866334 to 866345 | | | |
| see above | University College London, Great Ormond Street Hospital for Children NHS Foundation Trust, Imperial College Healthcare NHS Trust | COVID-19 Genomics UK (COG-UK) Consortium | Alison Holmes; Charlotte Williams; Helena Tutill; Jacqueline Findlay; James Price; Judith Breuer; Julianne Brown; Kathryn Harris; Leysa Forrest; Mark Kristiansen; Paola Niola; Paola Resende Silva; Patricia Dyal; Paul Randell; Rachel Williams; Sam Weeks; Samuel Weeks; Sergi Castellano; Sunando Roy; Tony Brooks; Yasmin Panchbhaya |
| EPI_ISL_492495, EPI_ISL_492497, EPI_ISL_492748, EPI_ISL_492750 | University College London, Great Ormond Street Hospital for Children NHS Foundation Trust, Imperial College Healthcare NHS Trust | Wellcome Sanger Institute for the COVID-19 Genomics UK (COG-UK) Consortium | Alison Holmes; Charlotte Williams; Cordelia Langford; David K. Jackson; Dominic Kwiatkowski; Ewan Harrison; Helena Tutill; Ian Johnston; Jacqueline Findlay; James Price; John Sillitoe on behalf of the Wellcome Sanger Institute COVID-19 Surveillance Team; Judith Breuer and Alex Alderton; Julianne Brown; Kathryn Harris; Leysa Forrest; Mark Kristiansen; Paola Niola; Paola Resende Silva; Patricia Dyal; Paul Randell; Rachel Williams; Roberto Amato; Sam Weeks; Sergi Castellano; Sonia Goncalves; Sunando Roy; Tony Brooks; Yasmin Panchbhaya |
| EPI_ISL_482160 to 482168, EPI_ISL_482170 to 482218, EPI_ISL_487525, EPI_ISL_487529, EPI_ISL_487534, EPI_ISL_487537, EPI_ISL_487540, EPI_ISL_487546, EPI_ISL_487550, EPI_ISL_487556 to 487559, EPI_ISL_487562, EPI_ISL_487564, EPI_ISL_487568, EPI_ISL_487572, EPI_ISL_487578, EPI_ISL_487581 to 487582, EPI_ISL_487584 to 487585, EPI_ISL_487589, EPI_ISL_487595, EPI_ISL_487597, EPI_ISL_487600, EPI_ISL_487606 to 487607, EPI_ISL_487614 to 487616, EPI_ISL_487621, EPI_ISL_487625 to 487628, EPI_ISL_487630 to 487633, EPI_ISL_487635 to 487637, EPI_ISL_487639 to 487641, EPI_ISL_487643, EPI_ISL_487646, EPI_ISL_492496, EPI_ISL_492498 to 492504, EPI_ISL_492747, EPI_ISL_492751, EPI_ISL_492753 to 492757, EPI_ISL_492760, EPI_ISL_492762 | | | |
| see above | University College London, Great Ormond Street Hospital for Children NHS Foundation Trust, Imperial College Healthcare NHS Trust | Wellcome Sanger Institute for the COVID-19 Genomics UK (COG-UK) consortium | Alison Holmes; Charlotte Williams; Cordelia Langford; David K. Jackson; Dominic Kwiatkowski; Ewan Harrison; Helena Tutill; Ian Johnston; Jacqueline Findlay; James Price; John Sillitoe on behalf of the Wellcome Sanger Institute COVID-19 Surveillance Team (http://www.sanger.ac.uk/covid-team); Judith Breuer and Alex Alderton; Julianne Brown; Kathryn Harris; Leysa Forrest; Mark Kristiansen; Paola Niola; Paola Resende Silva; Patricia Dyal; Paul Randell; Rachel Williams; Roberto Amato; Sam Weeks; Sergi Castellano; Sonia Goncalves; Sunando Roy; Tony Brooks; Yasmin Panchbhaya |
| EPI_ISL_418275, EPI_ISL_418277, EPI_ISL_418279 to 418280, EPI_ISL_418282 to 418284, EPI_ISL_418433 to 418435, EPI_ISL_418438 to 418440, EPI_ISL_527916 to 527917, EPI_ISL_527919 to 527941, EPI_ISL_527943 to 527961, EPI_ISL_527963 to 527964, EPI_ISL_527966 to 527970, EPI_ISL_527972 to 527975, EPI_ISL_527977 to 528018, EPI_ISL_528020 to 528025, EPI_ISL_528027 to 528034, EPI_ISL_528036 to 528037, EPI_ISL_528039 to 528077, EPI_ISL_528079 to 528097, EPI_ISL_528099 to 528143, EPI_ISL_528145 to 528152, EPI_ISL_528154 to 528160, EPI_ISL_528162 to 528176, EPI_ISL_528178 to 528188, EPI_ISL_528190 to 528191, EPI_ISL_528193 to 528209, EPI_ISL_528211 to 528217, EPI_ISL_528221 to 528225, EPI_ISL_528227, EPI_ISL_528229, EPI_ISL_528231 to 528233, EPI_ISL_528235 to 528237, EPI_ISL_528239 to 528240, EPI_ISL_528242 to 528247, EPI_ISL_528249, EPI_ISL_528251 to 528252, EPI_ISL_528254 to 528259, EPI_ISL_528262 to 528265, EPI_ISL_528267, EPI_ISL_528269 to 528274, EPI_ISL_528276 to 528277, EPI_ISL_528279 to 528281, EPI_ISL_528283 to 528288, EPI_ISL_528292 to 528294, EPI_ISL_528296 to 528297, EPI_ISL_528299 to 528304, EPI_ISL_528308 to 528311, EPI_ISL_528313, EPI_ISL_528315 to 528321, EPI_ISL_528323 to 528341, EPI_ISL_528343 to 528345, EPI_ISL_528347 to 528378, EPI_ISL_528381, EPI_ISL_530122, EPI_ISL_581668 to 581671, EPI_ISL_581673 to 581679, EPI_ISL_581681 to 581684, EPI_ISL_581686, EPI_ISL_581689 to 581690, EPI_ISL_581693 to 581703, EPI_ISL_581705 to 581708, EPI_ISL_581710 to 581712, EPI_ISL_581714 to 581718, EPI_ISL_581721, EPI_ISL_581723, EPI_ISL_581725 to 581739, EPI_ISL_581741 to 581754, EPI_ISL_581756 to 581758, EPI_ISL_581760 to 581770, EPI_ISL_581772 to 581773, EPI_ISL_581775 to 581810, EPI_ISL_581812 to 581816, EPI_ISL_581818 to 581830, EPI_ISL_581832 to 581843, EPI_ISL_581846 to 581857, EPI_ISL_581859, EPI_ISL_581861, EPI_ISL_581863 to 581881, EPI_ISL_581883 to 581886, EPI_ISL_581888 to 581889, EPI_ISL_581891 to 581909, EPI_ISL_581911, EPI_ISL_581913, EPI_ISL_581915 to 581973, EPI_ISL_581975 to 581987, EPI_ISL_581989 to 582001, EPI_ISL_830727 to 830729, EPI_ISL_830731 to 830737, EPI_ISL_830739 to 830747, EPI_ISL_830749 to 830757, EPI_ISL_830759 to 830765, EPI_ISL_830767 to 830776, EPI_ISL_830778 to 830783, EPI_ISL_830785 to 830798, EPI_ISL_830800 to 830803, EPI_ISL_830805, EPI_ISL_830807 to 830809, EPI_ISL_830811 to 830816, EPI_ISL_830818 to 830828, EPI_ISL_830830 to 830840, EPI_ISL_830842 to 830845, EPI_ISL_830847 to 830870, EPI_ISL_830872 to 830873, EPI_ISL_830875 to 830886, EPI_ISL_830888, EPI_ISL_830890 to 830893, EPI_ISL_830896 to 830917, EPI_ISL_830919 to 830932, EPI_ISL_830934 to 830943, EPI_ISL_830945 to 830993, EPI_ISL_830995 to 831001, EPI_ISL_831003 to 831014, EPI_ISL_831016 to 831019, EPI_ISL_842646 to 842651 | | | |
| see above | University Hospital Basel, Clinical Virology | University Hospital Basel, Clinical Bacteriology | Adrian Egli; Alexander Gensch; Alfredo Mari; Christian Nickel; Egli, A.; Hans Hirsch; Hans Pargger; Helena MB Seth-Smith; Hirsch, H.; Julia Bielicki; Karoline Leuzinger; Kirstine K. Soegaard; Leuzinger, K.; Madlen Stange; Manuel Battegay; Mari, A.; Martin Siegemund; Michael Osthoff; Michael Schweitzer; Myrta Brunner; Rita Schneider-Sliwa; Roland Bingisser; Roloff, T.; Sarah Tschudin-Sutter; Seth-Smith, H.; Simon Fuchs; Stefano Bassetti; Tim Roloff |
| EPI_ISL_418271 | University Hospital Basel, Clinical Virology | University Hospital Basel, Labormedizin | Egli, A.; Hirsch, H.; Leuzinger, K.; Mari, A.; Roloff, T.; Seth-Smith, H. |
| EPI_ISL_631377 to 631386, EPI_ISL_631388 | University Hospital Cologne | Center of Medical Microbiology, Virology, and Hospital Hygiene, University of Duesseldorf | Alexander Dilthey; Andreas Walker; Ashley-Jane Duplessis; Daniel Strelow; Elena Knops; Eva Heger; Florian Klein; Gibran Rubio Quintanares; Jessica Nicolai; Jörg Timm; Klaus Pfeffer; Malte Kohns Vasconcelos; Maximilian Damagnez; Rolf Kaiser; Saleta Sierra-Aragón; Tobias Wienemann; Torsten Houwaart |

| | | | |
|---|---|---|---|
| EPI_ISL_710548 to 710553, EPI_ISL_710555 to 710562, EPI_ISL_710564 to 710574 | University Hospital Dubrava | Ruer Boškovic Institute; Forensic Science Centre Ivan Vueti; University of Zagreb Faculty of Science | Ana Livun; Antonela Blažekovi; Boris Maek; Danilo Licastro; Dunja Glavaš; Fran Boroveki; Fuad osovi; Gordana Maravi Vlahoviek; Ivan Šamija; Ivana elap; Jasna Kašman; Josipa Skelin; Katarina Marija Tupek; Kristian Vlahoviek; Kristina Gotovac Jerei; Lidija Cvetko-Krajinovi; Lucija Basi; Lucija Markulin; Maja Kuzman; Marina Korolija; Mario Štefanovi; Mirjana Domazet-Lošo; Paula Štancl; Petra Vrabec; Robert Beluži; Rosa Karli; Sanja Tadinac; Senica Pejša; Tomislav Domazet-Lošo; Valentina umlijan-Combaj; Vjekoslav Tomai; Vladimir Krajinovi; Željka Maak Šafranko |
| EPI_ISL_681705 to 681742, EPI_ISL_681744 to 681757, EPI_ISL_681759 to 681775, EPI_ISL_763351 | University Hospital Limerick | Irish Coronavirus Sequencing Consortium - Teagasc Moorepark | Amy Fitzpatrick; Calum Walsh; Carolyn Meaney; Elaine Lawton; Fiona Crispie; John Kenny; Patrick Stapleton; Paul Cotter |
| EPI_ISL_483668 to 483671, EPI_ISL_483674, EPI_ISL_483676 to 483678, EPI_ISL_483680 to 483682, EPI_ISL_483684 | University Hospital Zurich | Department of Biosystems Science and Engineering, ETH Zürich | Christian Beisel; Elodie Burcklen; Ina Nissen; Ivan Topolsky; Julia Martinez-Gomez; Mitch Levesque; Natascha Santacroce; Niko Beerenwinkel; Pedro Ferreira; Phil Cheng; Philipp Bosshard; Philipp Jablonski; Sarah Nadeau; Susana Posada-Céspedes; Tanja Stadler; Tobias Schär |
| EPI_ISL_722202 to 722207 | University Hospital Zurich | Institute of Medical Virology, University of Zurich | Alexandra Trkola; Aline Wolfensberger; Gabriela Ziltener; Jürg Böni; Maryam Zaheri; Michael Huber; Stefan Schmutz; Thomas Scheier; Verena Kufner |
| EPI_ISL_524482, EPI_ISL_524782, EPI_ISL_730634, EPI_ISL_730636, EPI_ISL_733496 to 733497, EPI_ISL_751193 | University Hospital Zürich | Institute of Medical Virology, University of Zurich | Alexandra Trkola; Andrea Zbinden; Fiona Steiner; Gabriela Ziltener; Jon Huder; Jürg Böni; Kevin Steiner; Maryam Zaheri; Michael Huber; Patrick Redli; Riccarda Capaul; Stefan Schmutz; Thomas Scheier; Verena Kufner |
| EPI_ISL_454581, EPI_ISL_454583, EPI_ISL_454588, EPI_ISL_454592 | University Hospital for Infectious Diseases "Dr. Fran Mihaljevi", Research Unit | University of Zagreb, Centre for research and knowledge transfer in biotechnology | Anamarija Slovic; Ivan-Christian Kurolt; Jelena Ivancic Jelecki |
| EPI_ISL_417485, EPI_ISL_420138, EPI_ISL_420152 to 420153, EPI_ISL_420310, EPI_ISL_590924 to 590926, EPI_ISL_590938 to 590944, EPI_ISL_590946 to 590947, EPI_ISL_590949 to 590950, EPI_ISL_635068 to 635073, EPI_ISL_635113 to 635115, EPI_ISL_635117 to 635118, EPI_ISL_635137, EPI_ISL_635141, EPI_ISL_635147 to 635148, EPI_ISL_635156 to 635157, EPI_ISL_635160, EPI_ISL_635163, EPI_ISL_635179 to 635181, EPI_ISL_635186, EPI_ISL_635190, EPI_ISL_635198, EPI_ISL_668424 to 668426, EPI_ISL_708026 to 708027, EPI_ISL_708065 to 708072, EPI_ISL_708079 to 708080, EPI_ISL_708082 to 708095, EPI_ISL_708197 to 708198, EPI_ISL_759986 to 759987, EPI_ISL_775445 to 775448, EPI_ISL_775452 to 775456, EPI_ISL_775458, EPI_ISL_796670, EPI_ISL_796704, EPI_ISL_796707 to 796709, EPI_ISL_813978 | | | |
| see above | University Hospital of Northern Norway, Department for Microbiology and Infectious Disease Control | Norwegian Institute of Public Health, Department of Virology | Atiya R Ali; Hilde Elshaug; Hilde Vollan; Kamilla Heddeland Instefjord; Karoline Bragstad; Kathrine Stene-Johansen; Marie Paulsen Madsen; Olav Hungnes; Rasmus Riis Kopperud |
| EPI_ISL_478289 to 478371, EPI_ISL_484221 to 484253, EPI_ISL_484376 to 484386, EPI_ISL_484388 to 484389, EPI_ISL_499810, EPI_ISL_499828 to 499829, EPI_ISL_499858, EPI_ISL_499861, EPI_ISL_499869, EPI_ISL_499894 to 499895, EPI_ISL_499897, EPI_ISL_499910, EPI_ISL_499912 to 499913, EPI_ISL_499933, EPI_ISL_499950, EPI_ISL_499974 to 499983 | | | |
| see above | University Hospitals Of Leicester NHS Trust and DeepSeq Nottingham | COVID-19 Genomics UK (COG-UK) Consortium | Christopher Holmes; Christopher Moore; Fei Sang; Joeseph Chappell; Johnny Debebe; Jonathan Ball; Julian Tang; Karlie Fallon; Matthew Carlisle; Matthew Loose; Nadine Holmes; Patrick McClure; Paul Bird; Thomas Helmer; Victoria Wright |
| EPI_ISL_415454 to 415459, EPI_ISL_415698 to 415708, EPI_ISL_429196 to 429197, EPI_ISL_429199 to 429200, EPI_ISL_429203 to 429204, EPI_ISL_429206 to 429215, EPI_ISL_429217 to 429223 | | | |
| see above | University Hospitals of Geneva Laboratory of Virology | University Hospitals of Geneva Laboratory of Virology | Laubscher F. |
| EPI_ISL_860247 | University Hospitals of Geneva, Laboratory of Virology | HUG, Laboratory of Virology and Universitätsspital Basel | Adrian Egli; Alfredo Mari; Ana Rita Goncalves; Hans Hirsch; Helena MB Seth-Smith; Julia Bielicki; Karoline Leuzinger; Laurent Kaiser; Madlen Stange; Manuel Battegay; Samuel Cordey; Tim Roloff |
| EPI_ISL_847843, EPI_ISL_847845 to 847846, EPI_ISL_847848 to 847851, EPI_ISL_847854, EPI_ISL_847873 to 847919, EPI_ISL_847921 to 847928, EPI_ISL_847937 to 847939, EPI_ISL_847954 to 847977 | | | |
| see above | University Hospitals of Geneva, Laboratory of Virology | HUG, Laboratory of Virology and the Health2030 Genome Center | Ana Rita Goncalves; Emmanouil Dermitzakis; Henri Pegeot; Ioannis Xenarios; Keith Harshman; Laurent Kaiser; Lorenzo Cerutti; Melyssa Elies; Samuel Cordey |
| EPI_ISL_708306 to 708308, EPI_ISL_708315, EPI_ISL_708379 to 708381, EPI_ISL_738085 | University Hospitals of Geneva, Laboratory of Virology | University Hospitals of Geneva, Laboratory of Virology | Cordey Samuel; Cordey Samuel and Laubscher Florian; Laubscher Florian |
| EPI_ISL_775607, EPI_ISL_775609 to 775619, EPI_ISL_775621, EPI_ISL_775623 to 775639, EPI_ISL_775642 to 775643, EPI_ISL_775645 to 775758, EPI_ISL_775760 to 775763, EPI_ISL_775766 to 775768, EPI_ISL_775771 to 775774, EPI_ISL_775776 to 775778, EPI_ISL_775780 to 775782, EPI_ISL_775786, EPI_ISL_775790 to 775791, EPI_ISL_775793 to 775799, EPI_ISL_775801 to 775810, EPI_ISL_775812 to 775813, EPI_ISL_775815, EPI_ISL_775817, EPI_ISL_775819, EPI_ISL_775821 to 775829, EPI_ISL_775831, EPI_ISL_775833 to 775835, EPI_ISL_775839, EPI_ISL_775843, EPI_ISL_775845, EPI_ISL_775848 to 775851, EPI_ISL_775853 to 775855, EPI_ISL_775857, EPI_ISL_775860 to 775862, EPI_ISL_775864 to 775868, EPI_ISL_775870 to 775874, EPI_ISL_775879, EPI_ISL_775883, EPI_ISL_775885 to 775891, EPI_ISL_775898 to 775914, EPI_ISL_775916 to 775921, EPI_ISL_775925 to 776566 | | | |
| see above | University Medical Center Hamburg Eppendorf | Heinrich Pette Institute, Leibniz Institute for Experimental Virology | Adam Grundhoff; Alexis Robitaille; Johannes Knobloch; Martin Aepfelbacher; Nicole Fischer; Thomas Günther |
| EPI_ISL_735504 to 735506, EPI_ISL_738144, EPI_ISL_767013, EPI_ISL_767043, EPI_ISL_826284, EPI_ISL_826458, EPI_ISL_826520, EPI_ISL_940753 | | | |
| see above | University of Bari Biomedical Sciences and Human Oncology | University of Bari Biomedical Sciences and Human Oncology | Accogli M.; Accogli Marisa; Anna Sallustio; Chironna M; Chironna M.; Chironna Maria; Daniela Loconsole; Loconsole D.; Loconsole Daniela; Maria Chironna; Marisa Accogli; Sallustio A.; Sallustio Anna |
| EPI_ISL_730653 | University of Bari, Valenzano, Italy | Istituto Zooprofilattico Sperimentale dell'Abruzzo e del Molise "G. Caporale". | A. Lorusso; C. Buonavoglia; C. Cammà; C. Desario; D. Buonavoglia, V.; E. Lorusso; G. Elia; Martella; N. Decaro; V. Curini |
| EPI_ISL_444279 to 444312, EPI_ISL_447922 to 447944, EPI_ISL_456667, EPI_ISL_456669 to 456676, EPI_ISL_473306 to 473346, EPI_ISL_473348 to 473392, EPI_ISL_473394 to 473450, EPI_ISL_477727 to 477782, EPI_ISL_483880 to 483912, EPI_ISL_484218 to 484220, EPI_ISL_493441 to 493451, EPI_ISL_526287 to 526336, EPI_ISL_529218 to 529223, EPI_ISL_529234 to 529235, EPI_ISL_529237, EPI_ISL_529239, EPI_ISL_529251 to 529252, EPI_ISL_529255 to 529256, EPI_ISL_529262, EPI_ISL_529275, EPI_ISL_529277 to 529278, EPI_ISL_529289, EPI_ISL_529291, EPI_ISL_529298, EPI_ISL_529312 to 529314, EPI_ISL_529320 to 529340, EPI_ISL_529357 to 529372, EPI_ISL_529384, EPI_ISL_529386, EPI_ISL_529390 to 529406, EPI_ISL_529408, EPI_ISL_529420 to 529421, EPI_ISL_529424 to 529430, EPI_ISL_529434 to 529435, EPI_ISL_529438 to 529444, EPI_ISL_529448, EPI_ISL_529453, EPI_ISL_529461 to 529462, EPI_ISL_529470 to 529475, EPI_ISL_529480, EPI_ISL_529482, EPI_ISL_529485, EPI_ISL_529493 to 529497, EPI_ISL_529499, EPI_ISL_529508, EPI_ISL_529512, EPI_ISL_529514 to 529515, EPI_ISL_529517 to 529518, EPI_ISL_529522, EPI_ISL_529590 to 529655, EPI_ISL_529661 to 529666, EPI_ISL_576877 to 576896, EPI_ISL_594484 to 594616, EPI_ISL_611550, EPI_ISL_611554, EPI_ISL_611556, EPI_ISL_611608, EPI_ISL_611612, EPI_ISL_611671, EPI_ISL_611692, EPI_ISL_611696, EPI_ISL_611700, EPI_ISL_611705, EPI_ISL_611776, EPI_ISL_611814, EPI_ISL_611825, EPI_ISL_611847, EPI_ISL_611855 to 611856, EPI_ISL_611858 to 611860, EPI_ISL_612011, EPI_ISL_612015, EPI_ISL_612047, EPI_ISL_612073, EPI_ISL_612207 to 612260, EPI_ISL_650119, EPI_ISL_650153, EPI_ISL_650155, EPI_ISL_650158, EPI_ISL_650171 to 650172, EPI_ISL_650176, EPI_ISL_650212, EPI_ISL_650216, EPI_ISL_650245, EPI_ISL_650250 to 650254, EPI_ISL_650271, EPI_ISL_650350, EPI_ISL_650354, EPI_ISL_650373, EPI_ISL_650418, EPI_ISL_650432, EPI_ISL_650436 to 650437, EPI_ISL_650463, EPI_ISL_650582, EPI_ISL_650613, EPI_ISL_650648, EPI_ISL_650668, EPI_ISL_650717, EPI_ISL_650760, EPI_ISL_650769 to 650773, EPI_ISL_650790 to 650792, EPI_ISL_650822 to 650825, EPI_ISL_650948, EPI_ISL_650995 to 651000, EPI_ISL_651127, EPI_ISL_651137, EPI_ISL_651140, EPI_ISL_651144, EPI_ISL_651174, EPI_ISL_651190, EPI_ISL_651193, EPI_ISL_651195, EPI_ISL_651266, EPI_ISL_651287, EPI_ISL_651304 to 651305, EPI_ISL_651342, EPI_ISL_651353, EPI_ISL_651378 to 651379, EPI_ISL_651381, EPI_ISL_651390, EPI_ISL_651393, EPI_ISL_651413, EPI_ISL_651422, EPI_ISL_651519, EPI_ISL_651590 to 651624, EPI_ISL_651627 to 651630, EPI_ISL_651632 to 651633, EPI_ISL_651635 to 651652, EPI_ISL_679008 to 679050, EPI_ISL_679054 to 679121, EPI_ISL_679123 to 679133, EPI_ISL_812971 to 813007, EPI_ISL_813009 to 813045, EPI_ISL_813047, EPI_ISL_813049 to 813091, EPI_ISL_813093 to 813101, EPI_ISL_813104, EPI_ISL_813106 to 813112, EPI_ISL_813114 to 813118, EPI_ISL_813120 to 813127, EPI_ISL_813129 to 813153 | | | |
| see above | University of Birmingham | COVID-19 Genomics UK (COG-UK) Consortium | Alex Richter; Andrew Bosworth; Andrew Bosworth. Queen Elizabeth Hospital: Anna Casey; Andrew D Beggs; Andrew D Beggs // PHE Heartlands Lab: Husam Osman; Andrew D Beggs PHE Heartlands Lab: Husam Osman; Celina M Whalley; Charlotte Poxon; Claire McMurray; Husam Osman; Institute of Microbiology; Joanne Stockton; Josh Quick; Kasun Wanigasooriya; Loman Lab: Claire McMurray; Mike Kidd; Nicholas Loman; Nicholas Loman // UHB Lab: Celina M Whalley; Nicholas Loman. University of Birmingham Testing Laboratory: Celina M Whalley; Oliver Pickles; Radoslaw Poplawski; Samuel Nicholls; University of Birmingham Testing Laboratory: Celina M Whalley; University of Birmingham: Claire McMurray; Will Rowe |
| EPI_ISL_476068 to 476077, EPI_ISL_477008 to 477013, EPI_ISL_671419 to 671495, EPI_ISL_678387 to 678396 | University of Debrecen, Department of Medical Microbiology | National Laboratory of Virology, Szentágothai Research Centre | Balázs Somogyi; Brigitta Zana; Endre Gábor Tóth; Eszter Csoma; Ferenc Jakab; Gábor Kemenesi |
| EPI_ISL_457028 to 457029, EPI_ISL_457031 to 457038, EPI_ISL_457040 to 457119, EPI_ISL_457121 to 457136, EPI_ISL_457138 to 457147, EPI_ISL_457149 to 457151, EPI_ISL_457153 to 457157, EPI_ISL_457159 to 457207, EPI_ISL_457209 to 457252, EPI_ISL_457254, EPI_ISL_457256 to 457258, EPI_ISL_471911 to 471922, EPI_ISL_471924 to 471927, EPI_ISL_471929 to 471951, EPI_ISL_471953 to 471957, EPI_ISL_471959 to 471961, EPI_ISL_471963 to 471972, EPI_ISL_471974 to 471992, EPI_ISL_478276 to 478279, EPI_ISL_478281 to 478284, EPI_ISL_478288, EPI_ISL_512336 to 512340, EPI_ISL_540579, EPI_ISL_540582 to 540583, EPI_ISL_577220 to 577231, EPI_ISL_577233 to 577245, EPI_ISL_577247 to 577251, EPI_ISL_577253 to 577256, EPI_ISL_577258 to 577264, EPI_ISL_577266 to 577267, EPI_ISL_577269 to 577277, EPI_ISL_577279 to 577289, EPI_ISL_577291 to 577298, EPI_ISL_577346, EPI_ISL_577359 to 577373, EPI_ISL_577375 to 577380, EPI_ISL_577544, EPI_ISL_584883 to 584905, EPI_ISL_585533 to 585559, EPI_ISL_585561 to 585582, EPI_ISL_611599, EPI_ISL_611641, EPI_ISL_611836 to 611838, EPI_ISL_611843 to 611844, EPI_ISL_611939 to 611940, EPI_ISL_611976, EPI_ISL_611987, EPI_ISL_612057, EPI_ISL_612118, EPI_ISL_612130, EPI_ISL_612375 to 612393, EPI_ISL_612395 to 612398, EPI_ISL_612400, EPI_ISL_626662, EPI_ISL_626666, EPI_ISL_626668 to 626669, EPI_ISL_626671 to 626673, EPI_ISL_626676, EPI_ISL_626682 to 626683, EPI_ISL_626685 to 626686, EPI_ISL_626713, EPI_ISL_626715 to 626717, EPI_ISL_626719 to 626731, EPI_ISL_626772 to 626777, EPI_ISL_626785, EPI_ISL_626793 to 626794, EPI_ISL_626797 to 626806, EPI_ISL_626810, EPI_ISL_626813, EPI_ISL_626882 to 626891, EPI_ISL_626910, EPI_ISL_626925, EPI_ISL_626937, EPI_ISL_626939 to 626946, EPI_ISL_627031, EPI_ISL_627052, EPI_ISL_627058 to 627062, EPI_ISL_627064 to 627065, EPI_ISL_627067, EPI_ISL_627102, EPI_ISL_627111 to 627112, EPI_ISL_627118, EPI_ISL_627143, EPI_ISL_627145, EPI_ISL_627149, EPI_ISL_627159, EPI_ISL_627163, EPI_ISL_627166, EPI_ISL_627182, EPI_ISL_627186, EPI_ISL_627190, EPI_ISL_627194, EPI_ISL_627196, EPI_ISL_627210, EPI_ISL_627291, EPI_ISL_627311 to 627312, EPI_ISL_627314 to 627321, EPI_ISL_627323 to 627328, EPI_ISL_627424 to 627441, EPI_ISL_627444, EPI_ISL_650141, EPI_ISL_650151, EPI_ISL_650163, EPI_ISL_650215, EPI_ISL_650219, EPI_ISL_650222, EPI_ISL_650225 to 650226, EPI_ISL_650258 to 650261, EPI_ISL_650296, EPI_ISL_650352, EPI_ISL_650357 to 650358, EPI_ISL_650360 to 650361, EPI_ISL_650377, EPI_ISL_650399, EPI_ISL_650414, EPI_ISL_650444 to 650445, EPI_ISL_650565, EPI_ISL_650597, EPI_ISL_650606 to 650607, EPI_ISL_650634 to 650639, EPI_ISL_650642 to 650645, EPI_ISL_650667, EPI_ISL_650702, EPI_ISL_650783 to 650784, EPI_ISL_650788, EPI_ISL_651053 to 651059, EPI_ISL_651074 to 651075, EPI_ISL_651083 to 651085, EPI_ISL_651130, EPI_ISL_651177, EPI_ISL_651183, EPI_ISL_651231, EPI_ISL_651238, EPI_ISL_651241, EPI_ISL_651289 to 651290, EPI_ISL_651300, EPI_ISL_651372 to 651373, EPI_ISL_651384, EPI_ISL_651439, EPI_ISL_651467 to 651468, EPI_ISL_651527, EPI_ISL_651547, EPI_ISL_652289 to 652303, EPI_ISL_652675 to 652710, EPI_ISL_664114 to 664117, EPI_ISL_664123 to 664124, EPI_ISL_664127 to 664128, EPI_ISL_664136, EPI_ISL_664144 to 664145, EPI_ISL_664153, EPI_ISL_664162, EPI_ISL_664170, EPI_ISL_664173, EPI_ISL_664183, EPI_ISL_664193 to 664194, EPI_ISL_664202, EPI_ISL_664205 to 664212, EPI_ISL_664214 to 664216, EPI_ISL_664218, EPI_ISL_664239, EPI_ISL_664250 to 664276, EPI_ISL_664279, EPI_ISL_664287, EPI_ISL_664292 to 664297, EPI_ISL_664318 to 664319, EPI_ISL_664327 to 664328, EPI_ISL_664330 to 664331, EPI_ISL_664398 to 664420, EPI_ISL_664423, EPI_ISL_664429, EPI_ISL_664439, EPI_ISL_664458 to 664459, EPI_ISL_664465, EPI_ISL_664473 to 664474, EPI_ISL_664491, EPI_ISL_664509, EPI_ISL_664545, EPI_ISL_664592, EPI_ISL_664594 to 664597, | | | |

| | | | |
|---|---|---|---|
| EPI_ISL_664603, EPI_ISL_664608 to 664609, EPI_ISL_664611, EPI_ISL_664626, EPI_ISL_664629, EPI_ISL_664644, EPI_ISL_664683, EPI_ISL_664687, EPI_ISL_664691, EPI_ISL_664709, EPI_ISL_664715 to 664716, EPI_ISL_664718, EPI_ISL_664720, EPI_ISL_664722, EPI_ISL_665046, EPI_ISL_665048 to 665050, EPI_ISL_665052 to 665054, EPI_ISL_665056 to 665062, EPI_ISL_665064 to 665112, EPI_ISL_665273, EPI_ISL_665277, EPI_ISL_665279, EPI_ISL_665282, EPI_ISL_665285, EPI_ISL_665324 to 665326, EPI_ISL_665332 to 665333, EPI_ISL_665417, EPI_ISL_665419, EPI_ISL_665422, EPI_ISL_665435, EPI_ISL_665470, EPI_ISL_665514 to 665515, EPI_ISL_665547 to 665549, EPI_ISL_665570 to 665571, EPI_ISL_665663, EPI_ISL_665666, EPI_ISL_665675, EPI_ISL_665686, EPI_ISL_665694, EPI_ISL_665700, EPI_ISL_665719, EPI_ISL_665721, EPI_ISL_665734, EPI_ISL_665738, EPI_ISL_665760, EPI_ISL_665851 to 665863, EPI_ISL_665956, EPI_ISL_702562, EPI_ISL_702790, EPI_ISL_702844, EPI_ISL_702994, EPI_ISL_705099, EPI_ISL_705134, EPI_ISL_705237, EPI_ISL_705251, EPI_ISL_705254, EPI_ISL_705373, EPI_ISL_705383, EPI_ISL_724262 to 724268, EPI_ISL_724276 to 724294, EPI_ISL_724296 to 724297, EPI_ISL_724299 to 724323, EPI_ISL_741359 to 741361, EPI_ISL_742117, EPI_ISL_763376, EPI_ISL_763382 to 763385, EPI_ISL_763387, EPI_ISL_763390 to 763391, EPI_ISL_763412, EPI_ISL_763414, EPI_ISL_763431 to 763437, EPI_ISL_763452 to 763453, EPI_ISL_763457, EPI_ISL_763459 to 763461, EPI_ISL_763496, EPI_ISL_763527 to 763536, EPI_ISL_763543, EPI_ISL_763553 to 763556, EPI_ISL_763558 to 763559, EPI_ISL_763561 to 763564, EPI_ISL_763629 to 763631, EPI_ISL_763669 to 763670, EPI_ISL_763696 to 763699, EPI_ISL_763701, EPI_ISL_763703 to 763717, EPI_ISL_763763, EPI_ISL_763818, EPI_ISL_763907, EPI_ISL_764084, EPI_ISL_764129, EPI_ISL_764135, EPI_ISL_764137, EPI_ISL_764158, EPI_ISL_764163, EPI_ISL_764165 to 764166, EPI_ISL_764170, EPI_ISL_764178 to 764179, EPI_ISL_764181 to 764185, EPI_ISL_764196 to 764197, EPI_ISL_764211, EPI_ISL_764214 to 764215, EPI_ISL_764217 to 764220, EPI_ISL_764223 to 764225, EPI_ISL_764240, EPI_ISL_764373 to 764382, EPI_ISL_813357 to 813486, EPI_ISL_813490 to 813498, EPI_ISL_813500 to 813505, EPI_ISL_813507 to 813517, EPI_ISL_813519 to 813524, EPI_ISL_813527 to 813533, EPI_ISL_813536 to 813589, EPI_ISL_813591 to 813598, EPI_ISL_813600 to 813654, EPI_ISL_838330, EPI_ISL_838332 to 838333 | | | |
| see above | University of Exeter | COVID-19 Genomics UK (COG-UK) Consortium | Aaron Jeffries; Audrey Farbos; Ben Temperton; Jane Masoli; Joanna Warwick-Dugdale; Michelle Michelsen; Robyn Manley; Stephen Michell |
| EPI_ISL_540442 to 540468 | University of Liège COVID-19 testing center | GIGA Medical Genomics | Emmanuel André; Fabrice Bureau; Keith Durkin; Laurent Gillet; Marc Van Ranst; Maria Artesi; Vincent Bours; Wouter Coppieters |
| EPI_ISL_476078, EPI_ISL_477615 to 477625, EPI_ISL_677727 to 677820, EPI_ISL_678290 to 678291 | University of Szeged, Institute of Clinical Microbiology | National Laboratory of Virology, Szentágothai Research Centre | Balázs Somogyi; Brigitta; Brigitta Zana; Endre Gábor Tóth; Ferenc Jakab; Gabriella Terhes; Gábor Kemenesi; Terhes Gabriella |
| EPI_ISL_434643, EPI_ISL_434655, EPI_ISL_434665, EPI_ISL_445227, EPI_ISL_445230 to 445231, EPI_ISL_445239, EPI_ISL_450812, EPI_ISL_450826 to 450828, EPI_ISL_475514, EPI_ISL_475516 to 475517, EPI_ISL_475526 to 475527 | | | |
| see above | Uppsala Narakut Aleris | The Public Health Agency of Sweden | Anna Risberg; Anna-Malin Linde; Annika Nilsson; Karin Tegmark-Wisell; Maria Lind Karlberg; Mattias Haukland; Mia Brytting; Olov Svartstrom; Oskar Karlsson Lindsjo; Reza Advani; Sandra Broddesson; Theresa Enkirch |
| EPI_ISL_469076, EPI_ISL_475118 to 475119, EPI_ISL_475151, EPI_ISL_582776 to 582784, EPI_ISL_615076 to 615078, EPI_ISL_623076 to 623079, EPI_ISL_648130 to 648131, EPI_ISL_654953, EPI_ISL_676521 to 676530, EPI_ISL_710579 to 710580, EPI_ISL_831975 | | | |
| see above | Uppsala klinisk mikrobiologi | The Public Health Agency of Sweden | Anna Risberg; Anna-Malin Linde; Department of Microbiology; Karin Tegmark-Wisell; Maria Lind Karlberg; Mattias Haukland; Mia Brytting; Olov Svartstrom; Oskar Karlsson Lindsjo; Petra Edquist; Reza Advani; Sandra Broddesson; Shamam Muradrasoli; The Public Health Agency of Sweden |
| EPI_ISL_833123 | Usca Pizzoli | Istituto Zooprofilattico Sperimentale dell'Abruzzo e Molise "G. Caporale" | Ancora M; Calistri P; Cammà C; Curini V; Delli Compagni E; Di Domenico M; Di Pasquale A; Lorusso A; Mangone I; Marcacci M; Puglia I; Rinaldi A; Savini G |
| EPI_ISL_452236, EPI_ISL_710576 | VC Sorgenfrimottagningen | The Public Health Agency of Sweden | Anna Risberg; Anna-Malin Linde; Department of Microbiology; Karin Tegmark-Wisell; Lisa Esbjornsson Klemendz; Maria Lind Karlberg; Mia Brytting; Olov Svartstrom; Oskar Karlsson Lindsjo; The Public Health Agency of Sweden; Theresa Enkirch |
| EPI_ISL_424366 | Vaccine Research, Development and Application Center, Erciyes University | Gen Era Diagnostics Inc. | Alper Iseri; Aykut Ozdarendeli; Can Holyavkin; Gunsu Aydin; Hazel Yetiskin; Muhammet Ali Uygut; Shaikh Terkis Islam Pavel; Zehra B Dursun; Ilhami Celik |
| EPI_ISL_445233, EPI_ISL_475520, EPI_ISL_475566, EPI_ISL_710577 to 710578, EPI_ISL_766642 to 766644 | Vardcentralen Brinken | The Public Health Agency of Sweden | Agnes Wigh; Anna Risberg; Anna-Malin Linde; Department of Microbiology; Karin Tegmark-Wisell; Maria Lind Karlberg; Mattias Haukland; Mia Brytting; Olov Svartstrom; Oskar Karlsson Lindsjo; Reza Advani; Sandra Broddesson; The Public Health Agency of Sweden; Theresa Enkirch |
| EPI_ISL_549051, EPI_ISL_549087, EPI_ISL_549173 to 549174, EPI_ISL_590885 to 590888, EPI_ISL_590919 to 590920, EPI_ISL_590982 to 590983, EPI_ISL_635103, EPI_ISL_635105 to 635106, EPI_ISL_635112, EPI_ISL_635139 to 635140, EPI_ISL_635168, EPI_ISL_635171, EPI_ISL_635195, EPI_ISL_668390, EPI_ISL_668406, EPI_ISL_708154 to 708168, EPI_ISL_775331 to 775332, EPI_ISL_775393, EPI_ISL_775449 to 775451, EPI_ISL_775472 to 775474, EPI_ISL_775482 to 775483, EPI_ISL_775498 to 775504, EPI_ISL_860231 | | | |
| see above | Vestfold Hospital, Toensberg Department of Microbiology | Norwegian Institute of Public Health, Department of Virology | Atiya R Ali; Hilde Elshaug; Hilde Synnøve Vollan; Kamilla Heddeland Instefjord; Karoline Bragstad; Kathrine Stene-Johansen; Marie Paulsen Madsen; Olav Hungnes; Rasmus Riis Kopperud |
| EPI_ISL_420137 | Vestfold Hospital, Tonsberg Department of Microbiology | Norwegian Institute of Public Health, Department of Virology | Hilde Elshaug; Kamilla Heddeland Instefjord; Karoline Bragstad; Kathrine Stene-Johansen; Olav Hungnes |
| EPI_ISL_437435 to 437436 | Veterinary Specialized Institue Kraljevo | Veterinary Specialized Institue Kraljevo | Claudio L Afonso; Dejan Vidanovic. Bojana Tesovic; Jeremy Volkening; Kazimir Matovic; Marko Dmitric; Milanko Sekler; Nikola Vaskovic; Tamas Petrovic; Zoran Debeljak |
| EPI_ISL_582524 to 582529, EPI_ISL_582531 to 582534, EPI_ISL_644568 to 644583 | Veterinary Specialized Institute "Kraljevo", Serbia | Veterinary Specialized Institute "Kraljevo", Serbia | Afonso, C.; Banovic Djeri, B.; Jankovic, M.; Jovanovic, T.; Knezevic, A.; Petrovic, T.; Sekler, M.; Tesovic, B.; Vidanovic, D.; Volkening, J. |
| EPI_ISL_678486 to 678493 | Veterinary Specialized Institute "Sabac", Serbia | Veterinary Specialized Institute "Kraljevo", Serbia | Afonso, C.; Banovic Djeri, B.; Jankovic, M.; Jovanovic, T.; Knezevic, A.; Mrkovacki, S.; Petrovic, T.; Sekler, M.; Tesovic, B.; Vidanovic, D.; Volkening, J. |
| EPI_ISL_454795, EPI_ISL_455480 | Veterinary Specialized Institute Kraljevo | Veterinary Specialized Institute Kraljevo | Banovic Djeri, B.; C.L.; Debeljak, Z.; Dmitric, M.; J. and Alfonso; Matovic, K.; Petrovic, T.; Sekler, M.; Tesovic, B.; Vaskovic, N.; Vidanovic, D.; Volkening |
| EPI_ISL_833572 | Veterinary Specialized Instute "Nis" | Scientific Veterinary Institute "Novi Sad" | Afonso, C.; Banovic Djeri, B.; Jankovic, M.; Jovanovic, T.; Knezevic, A.; Manic, M.; Petrovic, M.; Petrovic, T.; Sekler, M.; Tesovic, B.; Vidanovic, D.; Volkening, J. |
| EPI_ISL_833511 to 833521 | Veterinary Specialized Instute "Nis" | Veterinary Specialized Institute "Kraljevo", Serbia | Afonso, C.; Banovic Djeri, B.; Jankovic, M.; Jovanovic, T.; Knezevic, A.; Manic, M.; Petrovic, M.; Petrovic, T.; Sekler, M.; Tesovic, B.; Vidanovic, D.; Volkening, J. |
| EPI_ISL_833576 to 833577 | Veterinary Specialized Instute "Nis" | Veterinary Specialized Institute "Nis", Serbia | Afonso, C.; Banovic Djeri, B.; Jankovic, M.; Jovanovic, T.; Knezevic, A.; Manic, M.; Petrovic, M.; Petrovic, T.; Sekler, M.; Tesovic, B.; Vidanovic, D.; Volkening, J. |
| EPI_ISL_833573 to 833575 | Veterinary Specialized Instute "Nis" | Veterinary Specialized Institute "Sabac", Serbia | Afonso, C.; Banovic Djeri, B.; Jankovic, M.; Jovanovic, T.; Knezevic, A.; Mrkovacki, S.; Petrovic, T.; Sekler, M.; Tesovic, B.; Vidanovic, D.; Volkening, J.; Vujinovic, S., . |
| EPI_ISL_434647 to 434648, EPI_ISL_445223, EPI_ISL_450807, EPI_ISL_452233 | Victoria Vard och Halsa | The Public Health Agency of Sweden | Anna Risberg; Anna-Malin Linde; Karin Tegmark-Wisell; Maria Lind Karlberg; Mia Brytting; Olov Svartstrom; Oskar Karlsson Lindsjo; Sarah Henriksson; Theresa Enkirch |
| EPI_ISL_435146 to 435147 | Villa Serena del Dr. Leonardo Petruzzi | Istituto Zooprofilattico Sperimentale dell'Abruzzo e Molise "G.Caporale" | Ancora M; Cammà C; Curini V; Di Domenico M; Di Pasquale A; Lorusso A; Mangone I; Marcacci M; Puglia I; Rinaldi A; Savini G |
| EPI_ISL_802852 to 802863 | Vilnius University Hospital Santaros Klinikos, Vilnius University | Institute of Biotechnology, Life Sciences Center, Vilnius University | Albertas Timinskas; Alma Gedvilaite; Aurelija Zvirbliene; Daniel Naumovas; Emilija Vasiliunaite; Laimonas Griskevicius; Milda Norkiene |
| EPI_ISL_560391 to 560405 | Vilnius University Hospital Santaros Klinikos, Vilnius University | Institute of Biotechnology, Life Sciences Center, Vilnius University and Thermo Fisher Scientific | Albertas Timinskas; Alma Gedvilaite; Aurelija Zvirbliene; Daniel Naumovas; Justinas Slikas; Laimonas Griskevicius; Ligita Jancioriene; Mindaugas Paulauskas |
| EPI_ISL_451668 to 451670, EPI_ISL_451672 to 451677, EPI_ISL_451681 to 451685, EPI_ISL_451687 to 451691, EPI_ISL_451693 to 451714, EPI_ISL_451716 to 451717, EPI_ISL_451720 to 451723, EPI_ISL_451725 to 451730, EPI_ISL_451732, EPI_ISL_451734, EPI_ISL_451736, EPI_ISL_451738 to 451741, EPI_ISL_451744 to 451749, EPI_ISL_451751 to 451753, EPI_ISL_451755 to 451758, EPI_ISL_451760 to 451761, EPI_ISL_451763 to 451764, EPI_ISL_451766 to 451767, EPI_ISL_451769 to 451772, EPI_ISL_451777 to 451778, EPI_ISL_451780 to 451782, EPI_ISL_451784 to 451785, EPI_ISL_451787 to 451789, EPI_ISL_451791, EPI_ISL_451795 to 451807, EPI_ISL_451809, EPI_ISL_451812 to 451813, EPI_ISL_451816 to 451818, EPI_ISL_451821 to 451822, EPI_ISL_451827 to 451829, EPI_ISL_451831 to 451835, EPI_ISL_451839, EPI_ISL_451842 to 451845, EPI_ISL_451848 to 451854, EPI_ISL_451856 to 451857, EPI_ISL_451859 to 451866, EPI_ISL_451868 to 451880, EPI_ISL_451883 to 451886, EPI_ISL_451888 to 451894, EPI_ISL_451898, EPI_ISL_451902 to 451903, EPI_ISL_451906, EPI_ISL_451908 to 451911, EPI_ISL_451914 to 451916, EPI_ISL_466928 to 466929, EPI_ISL_466931, EPI_ISL_466933 to 466938, EPI_ISL_466941 to 466953, EPI_ISL_466955 to 466956, EPI_ISL_466958 to 466962, EPI_ISL_466964, EPI_ISL_466966 to 466972, EPI_ISL_466975 to 466985, EPI_ISL_466987 to 466992, EPI_ISL_466994 to 466997, EPI_ISL_466999 to 467004, EPI_ISL_467006 to 467007, EPI_ISL_467009 to 467010, EPI_ISL_467014, EPI_ISL_467016, EPI_ISL_468203 to 468204, EPI_ISL_468207, EPI_ISL_468209 to 468211, EPI_ISL_468213, EPI_ISL_468215, EPI_ISL_468220 to 468222, EPI_ISL_468224 to 468229, EPI_ISL_468231, EPI_ISL_468233 to 468235, EPI_ISL_468237 to 468238, EPI_ISL_468240 to 468245, EPI_ISL_468247, EPI_ISL_468250, EPI_ISL_468254, EPI_ISL_468257 to 468258, EPI_ISL_468260, EPI_ISL_468264 to 468268, EPI_ISL_468270 to 468271, EPI_ISL_468275 to 468276, EPI_ISL_468278 to 468280, EPI_ISL_468293, EPI_ISL_468299, EPI_ISL_468303, EPI_ISL_476082 to 476083, EPI_ISL_476085 to 476089, EPI_ISL_476095 to 476100, EPI_ISL_476102 to 476107, EPI_ISL_476110, EPI_ISL_476112 to 476113, EPI_ISL_476115, EPI_ISL_476117 to 476118, EPI_ISL_476121 to 476122, EPI_ISL_476124, EPI_ISL_476126 to 476127, EPI_ISL_476129 to 476130, EPI_ISL_476133 to 476134, EPI_ISL_483648 to 483649, EPI_ISL_483651 to 483652, EPI_ISL_483654, EPI_ISL_483656 to 483657, EPI_ISL_483660 to 483662, EPI_ISL_483664 to 483666, EPI_ISL_486448, EPI_ISL_486450, EPI_ISL_486452, EPI_ISL_486454 to 486455, EPI_ISL_486458, EPI_ISL_486460, EPI_ISL_486463, EPI_ISL_486466 to 486467, EPI_ISL_486470 to 486471, EPI_ISL_486473, EPI_ISL_486475 to 486477, EPI_ISL_486479, EPI_ISL_486481, EPI_ISL_486484, EPI_ISL_486488 to 486489, EPI_ISL_486491, EPI_ISL_486493 to 486497, EPI_ISL_486499, EPI_ISL_486501, EPI_ISL_486503 to 486505, EPI_ISL_486507, EPI_ISL_486509, EPI_ISL_486511 to 486514, EPI_ISL_486518 to 486519, EPI_ISL_486521, EPI_ISL_486523 to 486524, EPI_ISL_486526 to 486527, EPI_ISL_486530, EPI_ISL_486532 to 486533, EPI_ISL_486535, EPI_ISL_486538, EPI_ISL_489961 to 489963, EPI_ISL_489965 to 489967, EPI_ISL_489969, EPI_ISL_489971 to 489973, EPI_ISL_489975 to 489986, EPI_ISL_498621 to 498623, EPI_ISL_498627, EPI_ISL_500877 to 500882, EPI_ISL_500884 to 500887, EPI_ISL_500889 to 500890, EPI_ISL_500892 to 500899, EPI_ISL_500903 to 500910, EPI_ISL_500913 to 500914, EPI_ISL_500916 to 500917, EPI_ISL_500919, EPI_ISL_500921, EPI_ISL_500923 to 500924, EPI_ISL_500927, EPI_ISL_500929, EPI_ISL_500932 to 500933, EPI_ISL_500935 to 500940, EPI_ISL_500942 to 500945, EPI_ISL_510691, EPI_ISL_510693 to 510695, EPI_ISL_510698 to 510713, EPI_ISL_510715, EPI_ISL_510717 to 510722, EPI_ISL_510724, EPI_ISL_510728 to 510733, EPI_ISL_510735, EPI_ISL_510737 to 510740, EPI_ISL_510742 to 510743, EPI_ISL_510746, EPI_ISL_510748, EPI_ISL_510750 to 510751, EPI_ISL_510753 to 510754, EPI_ISL_510757 to 510763, EPI_ISL_510766 to 510767, EPI_ISL_510769 to 510775, EPI_ISL_510777 to 510781, EPI_ISL_510783 to 510785, EPI_ISL_510789, EPI_ISL_510791, EPI_ISL_510794 to 510795, EPI_ISL_510800 to 510803, EPI_ISL_510805 to 510809, EPI_ISL_511985 to 511989, EPI_ISL_511991 to 511992, EPI_ISL_511994 to 511995, EPI_ISL_511997 to 511998, EPI_ISL_512000 to 512001, EPI_ISL_512004 to 512006, EPI_ISL_512008, EPI_ISL_512010 to 512013, EPI_ISL_512016 to 512028, EPI_ISL_512030 to 512032, EPI_ISL_512034 to 512035, EPI_ISL_512037 to 512040, EPI_ISL_512043, EPI_ISL_512045 to 512046, EPI_ISL_512048 to 512054, EPI_ISL_516554 to 516556, EPI_ISL_516558 to 516565, EPI_ISL_516567, EPI_ISL_516570, EPI_ISL_516572 to 516576, EPI_ISL_516578 to 516590, EPI_ISL_516593 to 516597, EPI_ISL_516599 to 516600, EPI_ISL_516602, EPI_ISL_516605 to 516606, EPI_ISL_523856 to 523858, EPI_ISL_523860 to 523869, EPI_ISL_523871, EPI_ISL_523873 to 523877, EPI_ISL_523879, EPI_ISL_523883, EPI_ISL_523885 to 523887, EPI_ISL_523889 to 523892, EPI_ISL_523898 to 523900, EPI_ISL_523903, EPI_ISL_523905 to 523907, EPI_ISL_523909 to 523913, EPI_ISL_523915 to 523922, EPI_ISL_523924 to 523926, EPI_ISL_535585, EPI_ISL_535587 to 535589, EPI_ISL_535591, EPI_ISL_535593, EPI_ISL_535595, EPI_ISL_535597 to 535600, EPI_ISL_535602 to 535606, EPI_ISL_535608 to 535609, EPI_ISL_535611 to 535614, EPI_ISL_535617, EPI_ISL_535620 to 535621, EPI_ISL_535623, EPI_ISL_535625 to 535628, EPI_ISL_535630 to | | | |

535636, EPI_ISL_535638 to 535641, EPI_ISL_535643 to 535644, EPI_ISL_535646 to 535649, EPI_ISL_539341 to 539343, EPI_ISL_539346 to 539349, EPI_ISL_539356 to 539361, EPI_ISL_539364 to 539366, EPI_ISL_539369 to 539377, EPI_ISL_539379, EPI_ISL_539382 to 539383, EPI_ISL_539386 to 539387, EPI_ISL_539389, EPI_ISL_539393, EPI_ISL_539395 to 539399, EPI_ISL_539401 to 539403, EPI_ISL_539405 to 539406, EPI_ISL_539410 to 539412, EPI_ISL_539414, EPI_ISL_539419, EPI_ISL_539421, EPI_ISL_539423 to 539430, EPI_ISL_539435 to 539436, EPI_ISL_539439, EPI_ISL_539441 to 539442, EPI_ISL_539444 to 539448, EPI_ISL_539450 to 539462, EPI_ISL_539464 to 539465, EPI_ISL_539468, EPI_ISL_539470, EPI_ISL_539472 to 539474, EPI_ISL_539477, EPI_ISL_539479, EPI_ISL_541401 to 541405, EPI_ISL_541407 to 541417, EPI_ISL_541419 to 541423, EPI_ISL_541426 to 541433, EPI_ISL_541435 to 541437, EPI_ISL_541441 to 541443, EPI_ISL_541447 to 541454, EPI_ISL_541457 to 541466, EPI_ISL_541468, EPI_ISL_541472, EPI_ISL_541476 to 541480, EPI_ISL_541484 to 541485, EPI_ISL_541487 to 541490, EPI_ISL_541492, EPI_ISL_541496 to 541499, EPI_ISL_541502 to 541503, EPI_ISL_541506, EPI_ISL_541509 to 541511, EPI_ISL_541513, EPI_ISL_541517 to 541525, EPI_ISL_541527, EPI_ISL_541531, EPI_ISL_541533, EPI_ISL_541535 to 541536, EPI_ISL_541538, EPI_ISL_560416 to 560419, EPI_ISL_560421, EPI_ISL_560423 to 560426, EPI_ISL_560428 to 560429, EPI_ISL_560431 to 560440, EPI_ISL_560442, EPI_ISL_560445 to 560449, EPI_ISL_560451 to 560457, EPI_ISL_560459 to 560460, EPI_ISL_560462 to 560466, EPI_ISL_560468 to 560472, EPI_ISL_560474, EPI_ISL_560476 to 560477, EPI_ISL_560479 to 560482, EPI_ISL_560486 to 560487, EPI_ISL_560491, EPI_ISL_560493 to 560496, EPI_ISL_560500 to 560503, EPI_ISL_560505 to 560506, EPI_ISL_560508 to 560512, EPI_ISL_560515 to 560517, EPI_ISL_560519 to 560520, EPI_ISL_560523, EPI_ISL_560525, EPI_ISL_560528 to 560529, EPI_ISL_560531, EPI_ISL_560533 to 560535, EPI_ISL_560537 to 560539, EPI_ISL_560542 to 560546, EPI_ISL_560548 to 560553, EPI_ISL_574876 to 574883, EPI_ISL_574885 to 574886, EPI_ISL_574888 to 574892, EPI_ISL_574894 to 574897, EPI_ISL_574899, EPI_ISL_574901, EPI_ISL_574903 to 574906, EPI_ISL_574909 to 574910, EPI_ISL_574912 to 574922, EPI_ISL_574924 to 574925, EPI_ISL_574927 to 574936, EPI_ISL_574938 to 574956, EPI_ISL_574958 to 574962, EPI_ISL_574964 to 574976, EPI_ISL_574978 to 574981, EPI_ISL_574983 to 574984, EPI_ISL_574986 to 574992, EPI_ISL_574994, EPI_ISL_574996 to 575002, EPI_ISL_575004 to 575013, EPI_ISL_575015 to 575018, EPI_ISL_603267 to 603268, EPI_ISL_603270, EPI_ISL_603272, EPI_ISL_603274, EPI_ISL_603276 to 603277, EPI_ISL_603281, EPI_ISL_603288 to 603289, EPI_ISL_603293 to 603295, EPI_ISL_603298 to 603301, EPI_ISL_603305 to 603306, EPI_ISL_603308 to 603309, EPI_ISL_603311 to 603313, EPI_ISL_603315, EPI_ISL_603319 to 603325, EPI_ISL_603327 to 603328, EPI_ISL_603330 to 603332, EPI_ISL_603335, EPI_ISL_603338, EPI_ISL_603342, EPI_ISL_603345 to 603346, EPI_ISL_603350 to 603355, EPI_ISL_603357 to 603358, EPI_ISL_603360 to 603364, EPI_ISL_603366 to 603367, EPI_ISL_603369 to 603372, EPI_ISL_603375 to 603377, EPI_ISL_603379 to 603380, EPI_ISL_603383, EPI_ISL_603386, EPI_ISL_603388, EPI_ISL_603391 to 603392, EPI_ISL_603394, EPI_ISL_603396 to 603398, EPI_ISL_603400, EPI_ISL_603402 to 603404, EPI_ISL_603406 to 603409, EPI_ISL_603411 to 603413, EPI_ISL_603415, EPI_ISL_603417, EPI_ISL_603420 to 603421, EPI_ISL_603423 to 603424, EPI_ISL_603429, EPI_ISL_603431, EPI_ISL_603437 to 603441, EPI_ISL_603443 to 603444, EPI_ISL_603446 to 603447, EPI_ISL_603451, EPI_ISL_603455 to 603464, EPI_ISL_603466, EPI_ISL_603468 to 603469, EPI_ISL_603472 to 603473, EPI_ISL_603476, EPI_ISL_603478, EPI_ISL_603481 to 603486, EPI_ISL_603490, EPI_ISL_603493 to 603495, EPI_ISL_603499 to 603505, EPI_ISL_603507 to 603509, EPI_ISL_603511 to 603513, EPI_ISL_603515 to 603517, EPI_ISL_603520 to 603526, EPI_ISL_603529, EPI_ISL_603531, EPI_ISL_603536 to 603538, EPI_ISL_603541, EPI_ISL_603543 to 603544, EPI_ISL_603547 to 603549, EPI_ISL_603551 to 603554, EPI_ISL_603556 to 603557, EPI_ISL_603559 to 603561, EPI_ISL_603563, EPI_ISL_603565, EPI_ISL_603567 to 603568, EPI_ISL_603570 to 603571, EPI_ISL_603573, EPI_ISL_603577 to 603579, EPI_ISL_603583, EPI_ISL_603586, EPI_ISL_603590, EPI_ISL_603599 to 603602, EPI_ISL_603605 to 603606, EPI_ISL_603609 to 603610, EPI_ISL_603614, EPI_ISL_603616 to 603618, EPI_ISL_603620 to 603621, EPI_ISL_603623 to 603626, EPI_ISL_603630, EPI_ISL_603632, EPI_ISL_603634, EPI_ISL_603637, EPI_ISL_603639, EPI_ISL_603642 to 603644, EPI_ISL_603652 to 603654, EPI_ISL_603657, EPI_ISL_603659 to 603660, EPI_ISL_603663, EPI_ISL_603665 to 603667, EPI_ISL_603670, EPI_ISL_603672 to 603674, EPI_ISL_603676, EPI_ISL_603678, EPI_ISL_603681 to 603683, EPI_ISL_603685 to 603687, EPI_ISL_603689 to 603690, EPI_ISL_603693 to 603695, EPI_ISL_603698, EPI_ISL_603703 to 603705, EPI_ISL_603707 to 603708, EPI_ISL_603711, EPI_ISL_603713, EPI_ISL_603715 to 603716, EPI_ISL_603718, EPI_ISL_603720, EPI_ISL_603723, EPI_ISL_603725, EPI_ISL_603727 to 603729, EPI_ISL_603731 to 603734, EPI_ISL_603739, EPI_ISL_603742 to 603743, EPI_ISL_603745, EPI_ISL_603748 to 603750, EPI_ISL_603752, EPI_ISL_614893 to 614904, EPI_ISL_614906, EPI_ISL_614908 to 614912, EPI_ISL_614914 to 614917, EPI_ISL_614921 to 614928, EPI_ISL_614930 to 614931, EPI_ISL_614933 to 614937, EPI_ISL_614940 to 614943, EPI_ISL_614945 to 614946, EPI_ISL_614948 to 614953, EPI_ISL_614955 to 614956, EPI_ISL_614958, EPI_ISL_614960 to 614961, EPI_ISL_614963, EPI_ISL_614965, EPI_ISL_614967 to 614969, EPI_ISL_614971 to 614974, EPI_ISL_614976 to 614979, EPI_ISL_693775 to 693783, EPI_ISL_693785 to 693789, EPI_ISL_693792 to 693793, EPI_ISL_693795 to 693796, EPI_ISL_693798 to 693799, EPI_ISL_693803 to 693805, EPI_ISL_693807 to 693828, EPI_ISL_693830, EPI_ISL_693832 to 693844, EPI_ISL_693846, EPI_ISL_693849, EPI_ISL_693851 to 693854, EPI_ISL_693856 to 693859, EPI_ISL_693861 to 693862, EPI_ISL_693864 to 693866, EPI_ISL_693868 to 693870, EPI_ISL_693872 to 693873, EPI_ISL_693876 to 693880, EPI_ISL_693882 to 693884, EPI_ISL_693886 to 693896, EPI_ISL_693898, EPI_ISL_693900 to 693901, EPI_ISL_693905 to 693917, EPI_ISL_693919 to 693928, EPI_ISL_693930 to 693931, EPI_ISL_693934, EPI_ISL_693936 to 693946, EPI_ISL_693952 to 693961, EPI_ISL_693963 to 693968, EPI_ISL_693971, EPI_ISL_693973 to 693975, EPI_ISL_693978, EPI_ISL_693980 to 693982, EPI_ISL_693984 to 693985, EPI_ISL_693987 to 693994, EPI_ISL_693997, EPI_ISL_694000 to 694001, EPI_ISL_694004 to 694005, EPI_ISL_694007 to 694008, EPI_ISL_721670, EPI_ISL_721672, EPI_ISL_721674 to 721675, EPI_ISL_721677, EPI_ISL_721679 to 721681, EPI_ISL_721684, EPI_ISL_721688, EPI_ISL_721691 to 721701, EPI_ISL_721704 to 721706, EPI_ISL_721709 to 721715, EPI_ISL_721718 to 721720, EPI_ISL_721724 to 721726, EPI_ISL_721728 to 721729, EPI_ISL_721731 to 721733, EPI_ISL_721735, EPI_ISL_721737 to 721738, EPI_ISL_721740 to 721742, EPI_ISL_721744, EPI_ISL_721746 to 721754, EPI_ISL_721756 to 721759, EPI_ISL_721763, EPI_ISL_721765 to 721768, EPI_ISL_721770 to 721774, EPI_ISL_721776 to 721777, EPI_ISL_721781, EPI_ISL_721786 to 721795, EPI_ISL_721797 to 721799, EPI_ISL_721801 to 721802, EPI_ISL_721804 to 721815, EPI_ISL_721817, EPI_ISL_721819 to 721822, EPI_ISL_721824 to 721831, EPI_ISL_721833 to 721834, EPI_ISL_721836, EPI_ISL_721839, EPI_ISL_721842, EPI_ISL_721844 to 721851, EPI_ISL_721853, EPI_ISL_721855 to 721858, EPI_ISL_721860 to 721862, EPI_ISL_721864 to 721865, EPI_ISL_721867 to 721874, EPI_ISL_721876 to 721889, EPI_ISL_721891 to 721895, EPI_ISL_721897, EPI_ISL_721899 to 721904, EPI_ISL_721907 to 721908, EPI_ISL_721910 to 721912, EPI_ISL_721915, EPI_ISL_721918, EPI_ISL_721920 to 721922, EPI_ISL_721924, EPI_ISL_721926 to 721928, EPI_ISL_721931 to 721933, EPI_ISL_721937, EPI_ISL_721939 to 721940, EPI_ISL_721942 to 721943, EPI_ISL_721948 to 721950, EPI_ISL_721952 to 721954, EPI_ISL_721956 to 721960, EPI_ISL_721962 to 721967, EPI_ISL_721971 to 721976, EPI_ISL_721978 to 721979, EPI_ISL_728765 to 728771, EPI_ISL_728773 to 728775, EPI_ISL_728777, EPI_ISL_728779 to 728781, EPI_ISL_728783 to 728784, EPI_ISL_728786 to 728795, EPI_ISL_728797, EPI_ISL_728799, EPI_ISL_728801 to 728802, EPI_ISL_728804, EPI_ISL_728806 to 728807, EPI_ISL_728810, EPI_ISL_728812 to 728815, EPI_ISL_728817 to 728818, EPI_ISL_728820 to 728823, EPI_ISL_728825 to 728826, EPI_ISL_728830, EPI_ISL_728832 to 728842, EPI_ISL_728844 to 728856, EPI_ISL_728858 to 728870, EPI_ISL_728872 to 728875, EPI_ISL_728877 to 728883, EPI_ISL_728885 to 728889, EPI_ISL_728891 to 728892, EPI_ISL_728894, EPI_ISL_728896 to 728897, EPI_ISL_728899 to 728906, EPI_ISL_728909 to 728910, EPI_ISL_728912 to 728916, EPI_ISL_728918 to 728920, EPI_ISL_728922 to 728927, EPI_ISL_728929 to 728932, EPI_ISL_728939 to 728948, EPI_ISL_728950, EPI_ISL_728953 to 728954, EPI_ISL_728956 to 728958, EPI_ISL_728960, EPI_ISL_728974, EPI_ISL_728976, EPI_ISL_728985 to 728986, EPI_ISL_728991, EPI_ISL_728995 to 729000, EPI_ISL_729002 to 729003, EPI_ISL_729005 to 729006, EPI_ISL_729009 to 729011, EPI_ISL_729015, EPI_ISL_729017, EPI_ISL_729020 to 729024, EPI_ISL_729026 to 729027, EPI_ISL_729029, EPI_ISL_729032 to 729033, EPI_ISL_729035 to 729039, EPI_ISL_729041 to 729054, EPI_ISL_729056 to 729059, EPI_ISL_729062, EPI_ISL_729065 to 729069, EPI_ISL_729071 to 729072, EPI_ISL_729074, EPI_ISL_729076 to 729081, EPI_ISL_729083, EPI_ISL_729086 to 729087, EPI_ISL_729089 to 729095, EPI_ISL_729097 to 729098, EPI_ISL_729100 to 729102, EPI_ISL_729104 to 729110, EPI_ISL_729112, EPI_ISL_729114, EPI_ISL_729117 to 729118, EPI_ISL_729122 to 729140, EPI_ISL_729143 to 729160, EPI_ISL_729162 to 729165, EPI_ISL_729167 to 729168, EPI_ISL_729171, EPI_ISL_729173 to 729176, EPI_ISL_729178 to 729180, EPI_ISL_729182, EPI_ISL_729184, EPI_ISL_729186 to 729187, EPI_ISL_729189, EPI_ISL_729191, EPI_ISL_729194, EPI_ISL_729196 to 729204, EPI_ISL_729207 to 729209, EPI_ISL_729211 to 729212, EPI_ISL_729214, EPI_ISL_729219 to 729221, EPI_ISL_729224 to 729226, EPI_ISL_729229 to 729240, EPI_ISL_729242 to 729243, EPI_ISL_729247 to 729250, EPI_ISL_729252 to 729253, EPI_ISL_729255, EPI_ISL_729257 to 729258, EPI_ISL_737396 to 737404, EPI_ISL_737406 to 737410, EPI_ISL_737412 to 737420, EPI_ISL_737422 to 737424, EPI_ISL_737426, EPI_ISL_737429 to 737430, EPI_ISL_737432, EPI_ISL_737434, EPI_ISL_737436 to 737439, EPI_ISL_737442, EPI_ISL_737444 to 737448, EPI_ISL_737450, EPI_ISL_737452 to 737453, EPI_ISL_737455 to 737456, EPI_ISL_737458 to 737462, EPI_ISL_737464 to 737476, EPI_ISL_737478 to 737482, EPI_ISL_737484 to 737489, EPI_ISL_737491 to 737499, EPI_ISL_737504, EPI_ISL_737506 to 737510, EPI_ISL_737512, EPI_ISL_737514, EPI_ISL_737516 to 737521, EPI_ISL_737523, EPI_ISL_737525 to 737526, EPI_ISL_737528 to 737534, EPI_ISL_737536 to 737537, EPI_ISL_737539, EPI_ISL_737541 to 737546, EPI_ISL_737549 to 737554, EPI_ISL_737556 to 737557, EPI_ISL_737560 to 737564, EPI_ISL_737566 to 737570, EPI_ISL_737572, EPI_ISL_737574 to 737575, EPI_ISL_737577, EPI_ISL_737579, EPI_ISL_737581, EPI_ISL_737583 to 737584, EPI_ISL_737587, EPI_ISL_737589, EPI_ISL_737591 to 737592, EPI_ISL_737594 to 737602, EPI_ISL_737604 to 737608, EPI_ISL_737610 to 737611, EPI_ISL_737614 to 737616, EPI_ISL_737619 to 737620, EPI_ISL_737625 to 737626, EPI_ISL_737629 to 737636, EPI_ISL_737639 to 737643, EPI_ISL_737647, EPI_ISL_737649, EPI_ISL_737651 to 737655, EPI_ISL_737657 to 737658, EPI_ISL_737660, EPI_ISL_737663 to 737665, EPI_ISL_737667 to 737668, EPI_ISL_737670 to 737672, EPI_ISL_737674 to 737677, EPI_ISL_737679 to 737687, EPI_ISL_737689 to 737692, EPI_ISL_737694 to 737695, EPI_ISL_737697 to 737700, EPI_ISL_737702 to 737706, EPI_ISL_737710 to 737717, EPI_ISL_737719, EPI_ISL_737721 to 737724, EPI_ISL_737729 to 737731, EPI_ISL_737733 to 737737, EPI_ISL_737739 to 737740, EPI_ISL_737742 to 737743, EPI_ISL_737746 to 737748, EPI_ISL_737751 to 737753, EPI_ISL_737755 to 737761, EPI_ISL_737763 to 737764, EPI_ISL_737766, EPI_ISL_737768 to 737769, EPI_ISL_737771, EPI_ISL_737773 to 737775, EPI_ISL_737777 to 737786, EPI_ISL_737789 to 737790, EPI_ISL_737792 to 737794, EPI_ISL_737796 to 737798, EPI_ISL_737800 to 737801, EPI_ISL_737803 to 737806, EPI_ISL_737808, EPI_ISL_737810, EPI_ISL_737812 to 737821, EPI_ISL_737823, EPI_ISL_737825 to 737826, EPI_ISL_737828, EPI_ISL_737830, EPI_ISL_737832 to 737835, EPI_ISL_737837, EPI_ISL_737839 to 737840, EPI_ISL_737844, EPI_ISL_737846 to 737847, EPI_ISL_737849 to 737850, EPI_ISL_737852 to 737853, EPI_ISL_737855 to 737856, EPI_ISL_737860 to 737868, EPI_ISL_737870 to 737871, EPI_ISL_737873, EPI_ISL_737875 to 737876, EPI_ISL_737878, EPI_ISL_737880, EPI_ISL_737883 to 737885, EPI_ISL_737887 to 737888, EPI_ISL_737890, EPI_ISL_737892 to 737893, EPI_ISL_737897 to 737898, EPI_ISL_737902 to 737904, EPI_ISL_737906 to 737908, EPI_ISL_737910 to 737912, EPI_ISL_737915 to 737916, EPI_ISL_737918 to 737919, EPI_ISL_737922 to 737928, EPI_ISL_767941, EPI_ISL_767943, EPI_ISL_767948, EPI_ISL_767955, EPI_ISL_767961, EPI_ISL_767963, EPI_ISL_767965 to 767967, EPI_ISL_767969, EPI_ISL_767971 to 767972, EPI_ISL_767977, EPI_ISL_767981, EPI_ISL_767989, EPI_ISL_767991 to 767993, EPI_ISL_767995, EPI_ISL_767999, EPI_ISL_768003 to 768004, EPI_ISL_768006 to 768007, EPI_ISL_768012, EPI_ISL_768018 to 768022, EPI_ISL_768024, EPI_ISL_768030, EPI_ISL_768033, EPI_ISL_768043, EPI_ISL_768046, EPI_ISL_768049, EPI_ISL_768051 to 768053, EPI_ISL_768055 to 768057, EPI_ISL_768062 to 768063, EPI_ISL_768067, EPI_ISL_768069, EPI_ISL_768078 to 768079, EPI_ISL_768081, EPI_ISL_768084, EPI_ISL_768088, EPI_ISL_768091 to 768092, EPI_ISL_768096, EPI_ISL_768105 to 768106, EPI_ISL_768112 to 768113, EPI_ISL_768115 to 768117, EPI_ISL_768121 to 768122, EPI_ISL_768125, EPI_ISL_768127, EPI_ISL_768131, EPI_ISL_768135, EPI_ISL_768137, EPI_ISL_768140, EPI_ISL_768143 to 768145, EPI_ISL_768147, EPI_ISL_768157, EPI_ISL_768166 to 768167, EPI_ISL_768169 to 768171, EPI_ISL_768173 to 768174, EPI_ISL_768178 to 768180, EPI_ISL_768182 to 768188, EPI_ISL_768192 to 768194, EPI_ISL_768198, EPI_ISL_768200 to 768201, EPI_ISL_768203 to 768205, EPI_ISL_768207 to 768208, EPI_ISL_768210, EPI_ISL_768212 to 768214, EPI_ISL_768216 to 768220, EPI_ISL_768224 to 768226, EPI_ISL_768233, EPI_ISL_768238, EPI_ISL_768256, EPI_ISL_768258, EPI_ISL_768260, EPI_ISL_768265, EPI_ISL_768277, EPI_ISL_768280, EPI_ISL_768288, EPI_ISL_768299 to 768302, EPI_ISL_768310 to 768311, EPI_ISL_768314, EPI_ISL_768318, EPI_ISL_796163, EPI_ISL_796166, EPI_ISL_796173, EPI_ISL_796176, EPI_ISL_796178, EPI_ISL_796182, EPI_ISL_796186, EPI_ISL_796195, EPI_ISL_796197, EPI_ISL_796200 to 796201, EPI_ISL_796204 to 796205, EPI_ISL_796211, EPI_ISL_796214, EPI_ISL_796217, EPI_ISL_796219, EPI_ISL_796222 to 796229, EPI_ISL_796231, EPI_ISL_796233 to 796234, EPI_ISL_796238, EPI_ISL_796240, EPI_ISL_796301 to 796306, EPI_ISL_796308 to 796309, EPI_ISL_796312 to 796316, EPI_ISL_796319 to 796321, EPI_ISL_796323 to 796325, EPI_ISL_796327, EPI_ISL_796330 to 796332, EPI_ISL_796335 to 796337, EPI_ISL_796340 to 796341, EPI_ISL_796345 to 796347, EPI_ISL_796352, EPI_ISL_796354, EPI_ISL_796356 to 796357, EPI_ISL_796360 to 796364, EPI_ISL_796373 to 796375, EPI_ISL_796377, EPI_ISL_796380 to 796384, EPI_ISL_796386 to 796388, EPI_ISL_796390, EPI_ISL_796392 to 796393, EPI_ISL_796395 to 796398, EPI_ISL_796400 to 796401, EPI_ISL_796403, EPI_ISL_796405 to 796406, EPI_ISL_796409 to 796412, EPI_ISL_796416, EPI_ISL_796419 to 796420, EPI_ISL_796423 to 796424, EPI_ISL_796428, EPI_ISL_796431 to 796435, EPI_ISL_796437, EPI_ISL_796440, EPI_ISL_796443, EPI_ISL_796446, EPI_ISL_796448, EPI_ISL_796451, EPI_ISL_796454, EPI_ISL_796456, EPI_ISL_796458, EPI_ISL_796462, EPI_ISL_796464, EPI_ISL_796468, EPI_ISL_796471, EPI_ISL_796473, EPI_ISL_796475 to 796479, EPI_ISL_796482 to 796483, EPI_ISL_796485 to 796488, EPI_ISL_796491 to 796501, EPI_ISL_796503, EPI_ISL_796507 to 796508, EPI_ISL_796511 to 796513, EPI_ISL_796516, EPI_ISL_796520 to 796521, EPI_ISL_796523 to 796524, EPI_ISL_796526, EPI_ISL_796528, EPI_ISL_796530, EPI_ISL_796532, EPI_ISL_796534, EPI_ISL_796541 to 796551, EPI_ISL_796553 to 796555, EPI_ISL_796557 to 796559, EPI_ISL_796562, EPI_ISL_796564 to 796567, EPI_ISL_796569, EPI_ISL_796572, EPI_ISL_796574 to 796575, EPI_ISL_796577 to 796579, EPI_ISL_796582, EPI_ISL_796584, EPI_ISL_796586, EPI_ISL_796590 to 796593, EPI_ISL_796596 to 796598, EPI_ISL_796600 to 796601, EPI_ISL_796604 to 796605, EPI_ISL_796607, EPI_ISL_796611, EPI_ISL_796613, EPI_ISL_796615 to 796617, EPI_ISL_796624, EPI_ISL_796626, EPI_ISL_796629 to 796634, EPI_ISL_796636, EPI_ISL_796638, EPI_ISL_796641 to 796646, EPI_ISL_811132

| | | | |
|---|---|---|---|
| see above | Viollier AG | Department of Biosystems Science and Engineering, ETH Zürich | Andrea Patrignani; Andreia Cabral de Gouvea; Catharine Aquino; Chaoran Chen; Christian Beisel; Christiane Beckmann; Christoph Noppen; David Dreifuss; Deborah Penet; Doris Popovic; Elodie Burcklen; Emmanouil Dermitzakis; Griffin White; Henri Pegeot; Ina Nissen; Ioannis Xenarios; Ivan Topolsky; Jay Tracy; Katharina Jahn; Keith Harshman; Lara Fuhrmann; Laura Neff; Lennart Opitz; Lorenzo Cerutti; Maria Domenica Moccia; Maurice Redondo; Natascha Santacroce; Niko Beerenwinkel; Noemie Santamaria de Souza; Olivier Kobel; Pedro Ferreira; Philipp Jablonski; Ralph Schlapbach; Sarah Nadeau; Simon Grüter; Sophie Seidel; Susana Posada-Céspedes; Tanja Stadler; Timothy Sykes; Tobias Schär |
| EPI_ISL_416488 | ViroGenetics - BSL3 Laboratory of Virology; Human Genome Variation Research Group & Genomics Centre MCB; Bioinformatics Research Group Department of Virology | ViroGenetics - BSL3 Laboratory of Virology; Human Genome Variation Research Group & Genomics Centre MCB; Bioinformatics Research Group Department of Virology | Adrianna Klajmon; Agata Jarosz; Agnieszka Koakowska-Kulesza; Aleksandra Milewska; Ewelina Hallman-Szeliska; Ewelina Popiech; Kamila Marszaek; Katarzyna Pancer; Katarzyna Zacharczuk; Krzysztof Pyr; Magdalena Rzeczkowska; Natalia Wolaniuk; Pawe P abaj; Tomasz Wokowicz; Wojciech Branicki |
| EPI_ISL_428925 to 428932 | ViroGenetics - BSL3 Laboratory of Virology; Human Genome Variation Research Group & Genomics Centre MCB; Bioinformatics Research Group; Wojewódzka Stacja Sanitarno-Epidemiologiczna w Krakowie | ViroGenetics - BSL3 Laboratory of Virology; Human Genome Variation Research Group & Genomics Centre MCB; Bioinformatics Research Group | Adrianna Klajmon; Agnieszka Starowicz; Aleksandra Pisarek; Aleksandra Synowiec; Danuta Piniewska-Róg; Ewelina Popiech; Inga Drebot; Jarosaw Foremny; Kamila Marszaek; Katarzyna Dudek; Katarzyna Gua; Katarzyna Kopera; Krzysztof Pyr; Magda Pachota; Marek Sanak; Micha Kowalski; Pawe P abaj; Tomasz Gromowski; Wojciech Branicki |
| EPI_ISL_416744, EPI_ISL_418183, EPI_ISL_435403 to 435431 | Virological Research Group, Szentágothai Research Centre | Bioinformatics Research Group, Szentágothai Research Centre | Attila Gyenesei; Endre Gábor Tóth; Ferenc Jakab; Gábor Kemenesi; Péter Urbán; Róbert Herczeg |
| EPI_ISL_416426 | Virological Research Group, Szentágothai Research Centre, University of Pécs | Bioinformatics Research Group, Szentágothai Research Centre, University of Pécs | Attila Gyenesei; Endre Gábor Tóth; Ferenc Jakab; Gábor Kemenesi; Péter Urbán; Róbert Herczeg |
| EPI_ISL_610249 | Virologisches Institut, Universitätsklinikum Erlangen | Virologisches Institut, Universitätsklinikum Erlangen | Armin Ensser; Klaus Korn; Klaus Überla |
| EPI_ISL_710487 to 710492 | Virology (microbiology), Hospital Universitario Central de Asturias | Virology (microbiology), Hospital Universitario Central de Asturias | Abreu, F.; Alvarez-Argueelles; Boga; Castello, C.; Costales, I.; Coto, E.; Gomez-deOna, J.; J.A.; M.E.; Martin-Rodriguez, G.; Melon, S.; Perez-Martinez, Z.; Rojo, S.; Sandoval, M. |
| EPI_ISL_415629, EPI_ISL_415632 to 415640 | Virology Department, Royal Infirmary of Edinburgh, NHS Lothian | Virology Department, Royal Infirmary of Edinburgh, NHS Lothian | Dewar R; McHugh M; O'Toole Á; Rambaut A; Templeton K; Williams TC |
| EPI_ISL_487767, EPI_ISL_489043, EPI_ISL_491572, EPI_ISL_491608, EPI_ISL_491611, EPI_ISL_491616, EPI_ISL_491655 | | | |
| see above | Virology Department, Royal Infirmary of Edinburgh, NHS Lothian / School of Biological Sciences, University of Edinburgh | Wellcome Sanger Institute for the COVID-19 Genomics UK (COG-UK) Consortium | Colquhoun R; Cordelia Langford; David K. Jackson; Dewar R; Dominic Kwiatkowski; Ewan Harrison; Hill V; Ian Johnston; Jackson B; John Sillitoe on behalf of the Wellcome Sanger Institute COVID-19 Surveillance Team; McCrone JT; McHugh M; O'Toole Á; Rambaut A; Roberto Amato; Rooke S; Scher E; Sonia Goncalves; Templeton K and Alex Alderton; Yu X |

| | | | |
|---|---|---|---|
| EPI_ISL_487647 to 487692, EPI_ISL_487694 to 487750, EPI_ISL_487752 to 487765, EPI_ISL_487768 to 487793, EPI_ISL_487795 to 487807, EPI_ISL_487809 to 487818, EPI_ISL_487820 to 487855, EPI_ISL_487857 to 487895, EPI_ISL_487897 to 487905, EPI_ISL_487907 to 487954, EPI_ISL_487956 to 487967, EPI_ISL_487969 to 488002, EPI_ISL_488879 to 488929, EPI_ISL_488931 to 489010, EPI_ISL_489012 to 489042, EPI_ISL_489044 to 489063, EPI_ISL_491494 to 491522, EPI_ISL_491524 to 491527, EPI_ISL_491529 to 491536, EPI_ISL_491538 to 491539, EPI_ISL_491541 to 491542, EPI_ISL_491544 to 491547, EPI_ISL_491549 to 491571, EPI_ISL_491573 to 491599, EPI_ISL_491601 to 491607, EPI_ISL_491609 to 491610, EPI_ISL_491612 to 491615, EPI_ISL_491617 to 491622, EPI_ISL_491624 to 491645, EPI_ISL_491647 to 491654, EPI_ISL_491656 to 491668, EPI_ISL_491670 to 491674, EPI_ISL_491676 to 491683, EPI_ISL_491685 to 491700, EPI_ISL_501632, EPI_ISL_532659, EPI_ISL_532683, EPI_ISL_532689, EPI_ISL_532705, EPI_ISL_532978, EPI_ISL_532983, EPI_ISL_532990, EPI_ISL_532992, EPI_ISL_532994 | | | |
| see above | Virology Department, Royal Infirmary of Edinburgh, NHS Lothian / School of Biological Sciences, University of Edinburgh | Wellcome Sanger Institute for the COVID-19 Genomics UK (COG-UK) consortium | Colquhoun R; Cordelia Langford; David K. Jackson; Dewar R; Dominic Kwiatkowski; Ewan Harrison; Hill V; Ian Johnston; Jackson B; John Sillitoe; John Sillitoe on behalf of the Wellcome Sanger Institute COVID-19 Surveillance Team (http://www.sanger.ac.uk/covid-team); McCrone JT; McHugh M; O'Toole Á; Rambaut A; Roberto Amato; Rooke S; Scher E; Sonia Goncalves; Templeton K and Alex Alderton; Yu X |
| EPI_ISL_425819 to 425822, EPI_ISL_425824 to 425829, EPI_ISL_425831 to 425832, EPI_ISL_425834 to 425836, EPI_ISL_425838 to 425908, EPI_ISL_425910 to 425911, EPI_ISL_425913 to 426024, EPI_ISL_432868 to 432899, EPI_ISL_433068 to 433198, EPI_ISL_433200 to 433267, EPI_ISL_433374 to 433403, EPI_ISL_433405 to 433421, EPI_ISL_433423 to 433426, EPI_ISL_433428 to 433437, EPI_ISL_433439 to 433465, EPI_ISL_439144 to 439178, EPI_ISL_439180 to 439294, EPI_ISL_439296 to 439299, EPI_ISL_439301 to 439367, EPI_ISL_439665 to 439671, EPI_ISL_449259 to 449329, EPI_ISL_453099 to 453194, EPI_ISL_456890 to 457023, EPI_ISL_461706 to 461762, EPI_ISL_473783 to 473876, EPI_ISL_473879 to 473894, EPI_ISL_473896 to 473955, EPI_ISL_478163 to 478273, EPI_ISL_484680 to 484684, EPI_ISL_493898 to 493974, EPI_ISL_507180 to 507201, EPI_ISL_514516 to 514562, EPI_ISL_517583 to 517585, EPI_ISL_517587 to 517589, EPI_ISL_526459 to 526517, EPI_ISL_526519 to 526536, EPI_ISL_529285, EPI_ISL_529317 to 529318, EPI_ISL_529478, EPI_ISL_529689, EPI_ISL_529691 to 529696, EPI_ISL_540871 to 540893, EPI_ISL_559971 to 559982, EPI_ISL_572411, EPI_ISL_572413 to 572414, EPI_ISL_572416, EPI_ISL_572421, EPI_ISL_572425, EPI_ISL_572427, EPI_ISL_572442, EPI_ISL_572483, EPI_ISL_572490, EPI_ISL_572514, EPI_ISL_572535 to 572539, EPI_ISL_572542 to 572544, EPI_ISL_572590, EPI_ISL_572593, EPI_ISL_572599, EPI_ISL_572608, EPI_ISL_572610, EPI_ISL_572617, EPI_ISL_572689, EPI_ISL_572692, EPI_ISL_572707, EPI_ISL_572756, EPI_ISL_572760, EPI_ISL_572762, EPI_ISL_572774, EPI_ISL_572776 to 572777, EPI_ISL_572801, EPI_ISL_572803, EPI_ISL_572821, EPI_ISL_572834, EPI_ISL_572839, EPI_ISL_572871, EPI_ISL_572874, EPI_ISL_572955, EPI_ISL_573247 to 573252, EPI_ISL_573794 to 573798, EPI_ISL_577347 to 577358, EPI_ISL_585426 to 585501, EPI_ISL_594816 to 594848, EPI_ISL_611544, EPI_ISL_611549, EPI_ISL_611685, EPI_ISL_611707, EPI_ISL_611842, EPI_ISL_611850, EPI_ISL_611894, EPI_ISL_611924, EPI_ISL_611989, EPI_ISL_612037, EPI_ISL_612074, EPI_ISL_612127 to 612129, EPI_ISL_612187 to 612189, EPI_ISL_612314 to 612374, EPI_ISL_650135, EPI_ISL_650138, EPI_ISL_650152, EPI_ISL_650159, EPI_ISL_650165, EPI_ISL_650167, EPI_ISL_650173, EPI_ISL_650177, EPI_ISL_650179 to 650180, EPI_ISL_650182, EPI_ISL_650191, EPI_ISL_650220, EPI_ISL_650255, EPI_ISL_650263 to 650264, EPI_ISL_650267, EPI_ISL_650277, EPI_ISL_650280, EPI_ISL_650286, EPI_ISL_650300 to 650308, EPI_ISL_650311, EPI_ISL_650426 to 650427, EPI_ISL_650433, EPI_ISL_650435, EPI_ISL_650439, EPI_ISL_650456, EPI_ISL_650478, EPI_ISL_650495, EPI_ISL_650499 to 650501, EPI_ISL_650531, EPI_ISL_650618, EPI_ISL_650690, EPI_ISL_650757, EPI_ISL_650775 to 650781, EPI_ISL_650793 to 650800, EPI_ISL_650812 to 650815, EPI_ISL_650819, EPI_ISL_650826, EPI_ISL_650832 to 650834, EPI_ISL_651012, EPI_ISL_651120, EPI_ISL_651141, EPI_ISL_651158, EPI_ISL_651169, EPI_ISL_651171, EPI_ISL_651188, EPI_ISL_651221, EPI_ISL_651228 to 651230, EPI_ISL_651294, EPI_ISL_651299, EPI_ISL_651301, EPI_ISL_651316, EPI_ISL_651320, EPI_ISL_651322, EPI_ISL_651343, EPI_ISL_651350, EPI_ISL_651366 to 651367, EPI_ISL_651385, EPI_ISL_651394 to 651395, EPI_ISL_651425 to 651428, EPI_ISL_651520 to 651524, EPI_ISL_651958 to 652066, EPI_ISL_665340, EPI_ISL_665552 to 665555, EPI_ISL_665563, EPI_ISL_665646, EPI_ISL_665725, EPI_ISL_665762, EPI_ISL_665790 to 665791, EPI_ISL_666060 to 666083, EPI_ISL_680461 to 680502, EPI_ISL_680504 to 680555, EPI_ISL_702528, EPI_ISL_702594, EPI_ISL_702620, EPI_ISL_702679, EPI_ISL_702937, EPI_ISL_703144, EPI_ISL_703198, EPI_ISL_703297, EPI_ISL_703414, EPI_ISL_703622, EPI_ISL_703779, EPI_ISL_703782, EPI_ISL_703784, EPI_ISL_703787, EPI_ISL_703810, EPI_ISL_703813, EPI_ISL_703863, EPI_ISL_703950, EPI_ISL_704187, EPI_ISL_704559, EPI_ISL_705118, EPI_ISL_705391, EPI_ISL_705416, EPI_ISL_705492, EPI_ISL_705734 to 705768, EPI_ISL_705770 to 705771, EPI_ISL_705773 to 705804, EPI_ISL_724222 to 724252, EPI_ISL_742175 to 742246, EPI_ISL_763447, EPI_ISL_763511, EPI_ISL_763548, EPI_ISL_763647, EPI_ISL_763649, EPI_ISL_763694 to 763695, EPI_ISL_763732, EPI_ISL_763817, EPI_ISL_763892 to 763906, EPI_ISL_764061, EPI_ISL_764069 to 764075, EPI_ISL_764083, EPI_ISL_764147, EPI_ISL_764195, EPI_ISL_764198, EPI_ISL_764221, EPI_ISL_764278 to 764279, EPI_ISL_764356 to 764372, EPI_ISL_814333, EPI_ISL_814365, EPI_ISL_814460 to 814465, EPI_ISL_814533 to 814535, EPI_ISL_814546, EPI_ISL_814569 to 814571, EPI_ISL_814665 to 814687, EPI_ISL_838214 to 838232, EPI_ISL_838234 to 838235, EPI_ISL_838238 to 838239, EPI_ISL_838241 to 838242, EPI_ISL_838244, EPI_ISL_838247 to 838248, EPI_ISL_838250 to 838251, EPI_ISL_838254 to 838257, EPI_ISL_838259 to 838263, EPI_ISL_838265 to 838271, EPI_ISL_838273, EPI_ISL_838275, EPI_ISL_838277 to 838279, EPI_ISL_838281 to 838289, EPI_ISL_838313 to 838314, EPI_ISL_865069 to 865072, EPI_ISL_865079 to 865088, EPI_ISL_865090 to 865091, EPI_ISL_865142, EPI_ISL_865146 to 865148, EPI_ISL_865153 to 865162 | | | |
| see above | Virology Department, Royal Infirmary of Edinburgh, NHS Lothian / School of Biological Sciences, University of Edinburgh / Institute of Genetics and Molecular Medicine, University of Edinburgh | COVID-19 Genomics UK (COG-UK) Consortium | Balcaza C; Colquhoun R; Dewar R; Gallagher M; Hill V; Jackson B; McCrone JT; McHugh M; O'Toole A; O'Toole Á; O'ÄôToole Å; Rambaut A; Rooke S; Scher E; Templeton K; Williams TC; Yu X |
| EPI_ISL_414500 to 414501 | Virology Department, Sheffield Teaching Hospitals NHS Foundation Trust | Department of Infection, Immunity and Cardiovascular Disease, The Florey Institute, The Medical School, University of Sheffield | Cariad Evans; Matthew Parker; Matthew Wyles; Mehmet Yavuz; Mohammad Raza; Thushan de Silva |
| EPI_ISL_416730 to 416740, EPI_ISL_418286 to 418321, EPI_ISL_420154 to 420181, EPI_ISL_420183 to 420194, EPI_ISL_420196 to 420233, EPI_ISL_420235 to 420244, EPI_ISL_420246 to 420267, EPI_ISL_420269, EPI_ISL_420271 to 420292 | | | |
| see above | Virology Department, Sheffield Teaching Hospitals NHS Foundation Trust | Department of Infection, Immunity and Cardiovascular Disease, The Florey Institute, The Medical School, University of Sheffield | Adri Angyal; Alex Keeley; Benjamin Lindsey; Cariad Evans; Danielle Groves; Dave Partridge; Luke Green; Matthew Parker; Matthew Wyles; Mehmet Yavuz; Mohammad Raza; Paul Parsons; Rachel Tucker; Rebecca Brown; Thushan de Silva |
| EPI_ISL_537254, EPI_ISL_537285 | Virology Department, Sheffield Teaching Hospitals NHS Foundation Trust / Department of Infection, Immunity and Cardiovascular Disease, The Medical School, University of Sheffield | Wellcome Sanger Institute for the COVID-19 Genomics UK (COG-UK) Consortium | Adri Angyal; Alex Keeley; Benjamin Lindsey; Cariad Evans and Alex Alderton; Cordelia Langford; Danielle Groves; Dave Partridge; David K. Jackson; Dominic Kwiatkowski; Ewan Harrison; Ian Johnston; John Sillitoe on behalf of the Wellcome Sanger Institute COVID-19 Surveillance Team; Luke Green; Matthew Parker; Matthew Wyles; Mehmet Yavuz; Mohammad Raza; Paul Parsons; Rachel Tucker; Rebecca Brown; Roberto Amato; Sonia Goncalves; Thushan de Silva |
| EPI_ISL_469918 to 469920, EPI_ISL_469922 to 469929, EPI_ISL_537206 to 537209, EPI_ISL_537211 to 537212, EPI_ISL_537214 to 537218, EPI_ISL_537220 to 537221, EPI_ISL_537223 to 537227, EPI_ISL_537229 to 537232, EPI_ISL_537234, EPI_ISL_537236 to 537253, EPI_ISL_537255 to 537267, EPI_ISL_537269 to 537270, EPI_ISL_537272 to 537274, EPI_ISL_537277, EPI_ISL_537280, EPI_ISL_537282 to 537284 | | | |
| see above | Virology Department, Sheffield Teaching Hospitals NHS Foundation Trust / Department of Infection, Immunity and Cardiovascular Disease, The Medical School, University of Sheffield | Wellcome Sanger Institute for the COVID-19 Genomics UK (COG-UK) consortium | Adri Angyal; Alex Keeley; Benjamin Lindsey; Cariad Evans and Alex Alderton; Cordelia Langford; Danielle Groves; Dave Partridge; David K. Jackson; Dominic Kwiatkowski; Ewan Harrison; Ian Johnston; John Sillitoe on behalf of the Wellcome Sanger Institute COVID-19 Surveillance Team; John Sillitoe on behalf of the Wellcome Sanger Institute COVID-19 Surveillance Team (http://www.sanger.ac.uk/covid-team); Luke Green; Matthew Parker; Matthew Wyles; Mehmet Yavuz; Mohammad Raza; Paul Parsons; Rachel Tucker; Rebecca Brown; Roberto Amato; Sonia Goncalves; Thushan de Silva |
| EPI_ISL_432451 to 432480, EPI_ISL_432482 to 432593, EPI_ISL_432595 to 432613, EPI_ISL_432615 to 432626, EPI_ISL_432628 to 432667, EPI_ISL_432710 to 432747, EPI_ISL_432749 to 432753, EPI_ISL_432755 to 432867 | | | |
| see above | Virology Department, Sheffield Teaching Hospitals NHS Foundation Trust / Virology Department, Sheffield Teaching Hospitals NHS Foundation Trust | COVID-19 Genomics UK (COG-UK) Consortium | Adri Angyal; Alex Keeley; Benjamin Lindsey; Cariad Evans; Danielle Groves; Dave Partridge; Luke Green; Matthew Parker; Matthew Wyles; Mehmet Yavuz; Mohammad Raza; Paul Parsons; Rachel Tucker; Rebecca Brown; Thushan de Silva |
| EPI_ISL_441898 to 441913, EPI_ISL_441915 to 441959, EPI_ISL_441961 to 442018, EPI_ISL_442020 to 442043, EPI_ISL_442344 to 442418, EPI_ISL_442420 to 442445, EPI_ISL_442447 to 442511, EPI_ISL_442513 to 442522, EPI_ISL_448826 to 448843, EPI_ISL_448845 to 448849, EPI_ISL_448851 to 448853, EPI_ISL_448855 to 448863, EPI_ISL_448865, EPI_ISL_448867 to 448893, EPI_ISL_448895, EPI_ISL_448897 to 448901, EPI_ISL_448903, EPI_ISL_448905, EPI_ISL_448907 to 448917, EPI_ISL_453696 to 453778, EPI_ISL_457581 to 457609, EPI_ISL_457611 to 457633, EPI_ISL_457635 to 457686, EPI_ISL_461999 to 462013, EPI_ISL_462015 to 462025, EPI_ISL_462027 to 462041, EPI_ISL_462043 to 462054, EPI_ISL_462057 to 462062, EPI_ISL_462064 to 462084, EPI_ISL_475342 to 475345, EPI_ISL_475347 to 475349, EPI_ISL_475351 to 475354, EPI_ISL_475356 to 475363, EPI_ISL_475365 to 475387, EPI_ISL_475389 to 475409, EPI_ISL_475411 to 475461, EPI_ISL_475463 to 475510, EPI_ISL_479195 to 479283, EPI_ISL_484433 to 484439, EPI_ISL_484441 to 484490, EPI_ISL_484492 to 484518, EPI_ISL_490581 to 490619, EPI_ISL_490621 to 490645, EPI_ISL_493666 to 493697, EPI_ISL_493699 to 493740, EPI_ISL_499330 to 499345, EPI_ISL_499347 to 499351, EPI_ISL_507160 to 507166, EPI_ISL_507168 to 507169, EPI_ISL_507172 to 507174, EPI_ISL_507176 to 507179, EPI_ISL_517543 to 517557, EPI_ISL_517559 to 517579, EPI_ISL_526437 to 526450, EPI_ISL_529301, EPI_ISL_529353 to 529356, EPI_ISL_529382, EPI_ISL_529407, EPI_ISL_529433, EPI_ISL_529446, EPI_ISL_529449, EPI_ISL_529500 to 529503, EPI_ISL_529521, EPI_ISL_529675 to 529683, EPI_ISL_535043 to 535058, EPI_ISL_535060 to 535101, EPI_ISL_540719 to 540774, EPI_ISL_559898 to 559925, EPI_ISL_559927 to 559946, EPI_ISL_560054 to 560068, EPI_ISL_572417 to 572418, EPI_ISL_572420, EPI_ISL_572422, EPI_ISL_572428, EPI_ISL_572432 to 572433, EPI_ISL_572457 to 572458, EPI_ISL_572467, EPI_ISL_572481 to 572482, EPI_ISL_572516, EPI_ISL_572522, EPI_ISL_572529, EPI_ISL_572567, EPI_ISL_572587, EPI_ISL_572589, EPI_ISL_572609, EPI_ISL_572613, EPI_ISL_572618 to 572619, EPI_ISL_572678, EPI_ISL_572722, EPI_ISL_572743, EPI_ISL_572752, EPI_ISL_572757, EPI_ISL_572767, EPI_ISL_572796, EPI_ISL_572820, EPI_ISL_572841, EPI_ISL_572848, EPI_ISL_572912 to 572916, EPI_ISL_572927 to 572930, EPI_ISL_572945 to 572946, EPI_ISL_572948 to 572949, EPI_ISL_572956, EPI_ISL_572959, EPI_ISL_572961, EPI_ISL_573527 to 573548, EPI_ISL_573550 to 573565, EPI_ISL_573686 to 573693, EPI_ISL_573695 to 573727, EPI_ISL_573729 to 573758, EPI_ISL_573762 to 573763, EPI_ISL_573806 to 573810, EPI_ISL_584167, EPI_ISL_584188 to 584192, EPI_ISL_584196 to 584199, EPI_ISL_584207 to 584211, EPI_ISL_584217 to 584230, EPI_ISL_584232 to 584282, EPI_ISL_584324 to 584377, EPI_ISL_584379 to 584380, EPI_ISL_585009 to 585015, EPI_ISL_585017 to 585037, EPI_ISL_585039 to 585091, EPI_ISL_585620 to 585626, EPI_ISL_595802 to 595821, EPI_ISL_595823 to 595836, EPI_ISL_596191 to 596227, EPI_ISL_611520, EPI_ISL_611525, EPI_ISL_611528, EPI_ISL_611536, EPI_ISL_611541, EPI_ISL_611545, EPI_ISL_611583 to 611584, EPI_ISL_611587, EPI_ISL_611594, EPI_ISL_611598, EPI_ISL_611602, EPI_ISL_611606, EPI_ISL_611650 to 611651, EPI_ISL_611655 to 611656, EPI_ISL_611661, EPI_ISL_611669, EPI_ISL_611678 to 611679, EPI_ISL_611681 to 611682, EPI_ISL_611690, EPI_ISL_611698 to 611699, EPI_ISL_611733, EPI_ISL_611744 to 611745, EPI_ISL_611774 to 611775, EPI_ISL_611777 to 611783, EPI_ISL_611787 to 611789, EPI_ISL_611793 to 611795, EPI_ISL_611797 to 611806, EPI_ISL_611816 to 611817, EPI_ISL_611829 to 611830, EPI_ISL_611861 to 611862, EPI_ISL_611869, EPI_ISL_611920, EPI_ISL_611936, EPI_ISL_611979, EPI_ISL_611988, EPI_ISL_611997, EPI_ISL_612005, EPI_ISL_612020, EPI_ISL_612022 to 612023, EPI_ISL_612033, EPI_ISL_612050, EPI_ISL_612055, EPI_ISL_612058, EPI_ISL_612062, EPI_ISL_612067, EPI_ISL_612084, EPI_ISL_612102, EPI_ISL_612196, EPI_ISL_613356 to 613409, EPI_ISL_626664, EPI_ISL_626679 to 626680, EPI_ISL_626710, EPI_ISL_626734 to 626735, EPI_ISL_626739, EPI_ISL_626743, EPI_ISL_626747, EPI_ISL_626752, EPI_ISL_626754, EPI_ISL_626757, EPI_ISL_626771, EPI_ISL_626781, EPI_ISL_626786, EPI_ISL_626791, EPI_ISL_626809, EPI_ISL_626817, EPI_ISL_626821, EPI_ISL_626823 to 626824, EPI_ISL_626827, EPI_ISL_626830, EPI_ISL_626852, EPI_ISL_626896, EPI_ISL_626898 to 626903, EPI_ISL_626906 to 626909, EPI_ISL_626911 to 626913, EPI_ISL_626915 to 626916, EPI_ISL_626918, EPI_ISL_626922 to 626924, EPI_ISL_626929, EPI_ISL_626932, EPI_ISL_626938, EPI_ISL_626947 to 626951, EPI_ISL_627019, EPI_ISL_627103, EPI_ISL_627130, EPI_ISL_627164, EPI_ISL_627184, EPI_ISL_627191, EPI_ISL_627252 to 627254, EPI_ISL_627264, EPI_ISL_627280, EPI_ISL_627298, EPI_ISL_627313, EPI_ISL_627492 to 627517, EPI_ISL_628369 to 628413, EPI_ISL_628421, EPI_ISL_637273, EPI_ISL_637285, EPI_ISL_637290 to 637291, EPI_ISL_637297, EPI_ISL_637320, EPI_ISL_637326, EPI_ISL_637358, EPI_ISL_637361, EPI_ISL_637409, EPI_ISL_637436, EPI_ISL_637531, EPI_ISL_637533, EPI_ISL_637535, EPI_ISL_637538 to 637541, EPI_ISL_637553, EPI_ISL_637559, EPI_ISL_637561, EPI_ISL_637580, EPI_ISL_637584 to 637585, EPI_ISL_637594, EPI_ISL_637621, EPI_ISL_637862 to 637863, EPI_ISL_637872, EPI_ISL_637876, EPI_ISL_637879, EPI_ISL_637881, EPI_ISL_637887, EPI_ISL_637901, EPI_ISL_637905, EPI_ISL_637944, EPI_ISL_637946, EPI_ISL_637973, EPI_ISL_637985, EPI_ISL_638017, EPI_ISL_638024, EPI_ISL_638090, EPI_ISL_638168, EPI_ISL_638532 to 638554, EPI_ISL_638877 to 638893, EPI_ISL_639008 to 639010, EPI_ISL_650121, EPI_ISL_650124, EPI_ISL_650131, EPI_ISL_650134, EPI_ISL_650208 to 650210, EPI_ISL_650237, EPI_ISL_650244, EPI_ISL_650283, EPI_ISL_650349, EPI_ISL_650362 to 650363, EPI_ISL_650367 to 650370, EPI_ISL_650372, EPI_ISL_650374, EPI_ISL_650384, EPI_ISL_650395, EPI_ISL_650406 to 650408, EPI_ISL_650422 to 650423, EPI_ISL_650425, EPI_ISL_650488, EPI_ISL_650554, EPI_ISL_650646, EPI_ISL_650650 to 650653, EPI_ISL_650659, EPI_ISL_650662 to 650663, EPI_ISL_650670, EPI_ISL_650674 to 650676, EPI_ISL_650678 to 650680, EPI_ISL_650689, EPI_ISL_650691, EPI_ISL_650705 to 650706, EPI_ISL_650713, EPI_ISL_650715, EPI_ISL_650718, EPI_ISL_650807 to 650808, EPI_ISL_651076 to 651078, EPI_ISL_651109, EPI_ISL_651121, EPI_ISL_651152, EPI_ISL_651184, EPI_ISL_651224, EPI_ISL_651233, EPI_ISL_651243, EPI_ISL_651291, EPI_ISL_651341, EPI_ISL_651374, EPI_ISL_651414, EPI_ISL_651437, EPI_ISL_651514, EPI_ISL_651518, EPI_ISL_651530, EPI_ISL_651715 to 651732, EPI_ISL_652180 to 652183, EPI_ISL_653059 to 653097, EPI_ISL_664283, EPI_ISL_664298, EPI_ISL_664370, EPI_ISL_664425 to 664426, EPI_ISL_664431, EPI_ISL_664437, EPI_ISL_664600, EPI_ISL_664602, EPI_ISL_664604, EPI_ISL_664630, EPI_ISL_664689, EPI_ISL_664713, EPI_ISL_664752, EPI_ISL_665023 to 665045, EPI_ISL_665864, EPI_ISL_679852 to 679856, EPI_ISL_679858 to 679923, EPI_ISL_679986 to 680015, EPI_ISL_680101 to 680110, EPI_ISL_680112 to 680135, EPI_ISL_680137 to 680150, EPI_ISL_680152 to 680197, EPI_ISL_680556 to 680561, EPI_ISL_702476, EPI_ISL_702487, EPI_ISL_702518, EPI_ISL_702531, EPI_ISL_702550, EPI_ISL_702564, EPI_ISL_702579, EPI_ISL_702607, EPI_ISL_702633, EPI_ISL_702682, EPI_ISL_702705, EPI_ISL_702761, EPI_ISL_702776, EPI_ISL_702798, EPI_ISL_702829 to 702830, EPI_ISL_702833, EPI_ISL_702842 to 702843, EPI_ISL_702852, EPI_ISL_702880 to 702881, EPI_ISL_702964, EPI_ISL_703018, EPI_ISL_703020, EPI_ISL_703026, EPI_ISL_703029, EPI_ISL_703038, EPI_ISL_703040, EPI_ISL_703048, EPI_ISL_703056, EPI_ISL_703133, EPI_ISL_703400, EPI_ISL_703449, EPI_ISL_703569, EPI_ISL_703575, EPI_ISL_703579, EPI_ISL_703582, EPI_ISL_703585, EPI_ISL_703587, EPI_ISL_703593, EPI_ISL_703601, EPI_ISL_703604, EPI_ISL_703606, EPI_ISL_703636, EPI_ISL_703639, EPI_ISL_703644, EPI_ISL_703647, EPI_ISL_703649, EPI_ISL_703655, EPI_ISL_703658, EPI_ISL_703660, EPI_ISL_703671, EPI_ISL_703674, EPI_ISL_703676, EPI_ISL_703684, EPI_ISL_703765 to 703766, EPI_ISL_703894, EPI_ISL_704190, EPI_ISL_704600 to 704605, EPI_ISL_704636 to 704639, EPI_ISL_704828, EPI_ISL_704960, EPI_ISL_704978, EPI_ISL_705026, EPI_ISL_705088, EPI_ISL_705096, EPI_ISL_705129, EPI_ISL_705153, EPI_ISL_705163, EPI_ISL_705195, EPI_ISL_705204, EPI_ISL_705210, EPI_ISL_705220, EPI_ISL_705230, EPI_ISL_705233, EPI_ISL_705270, EPI_ISL_705272, EPI_ISL_705276, EPI_ISL_705288, EPI_ISL_705317, EPI_ISL_705319, EPI_ISL_705335, EPI_ISL_705351, EPI_ISL_705371, EPI_ISL_705374, EPI_ISL_705405, EPI_ISL_705421, EPI_ISL_705615 to 705628, EPI_ISL_706975 to 707006, EPI_ISL_727861 to 727886, EPI_ISL_727888 to 727890, EPI_ISL_727892 to 727893, EPI_ISL_727895, EPI_ISL_727897 to 727900, EPI_ISL_727902 to 727905, EPI_ISL_727907 to 727922, EPI_ISL_727924 to 727951, EPI_ISL_727953 to 727954, EPI_ISL_727956 to 727976, EPI_ISL_727978, EPI_ISL_727980 to 727997, EPI_ISL_741936 to 741941, EPI_ISL_741943 to 741950, EPI_ISL_741952 to 741998, EPI_ISL_742001 to 742024, EPI_ISL_742026 to 742116, EPI_ISL_763365, EPI_ISL_763368 to 763369, EPI_ISL_763415, EPI_ISL_763419, EPI_ISL_763424, EPI_ISL_763427 to 763429, EPI_ISL_763439, EPI_ISL_763499, EPI_ISL_763501, EPI_ISL_763504, | | | |

| | | | |
|---|---|---|---|
| EPI_ISL_763506 to 763507, EPI_ISL_763514, EPI_ISL_763588, EPI_ISL_763632, EPI_ISL_763634 to 763636, EPI_ISL_763639, EPI_ISL_763645, EPI_ISL_763651 to 763654, EPI_ISL_763677, EPI_ISL_763842, EPI_ISL_763866 to 763871, EPI_ISL_764032 to 764033, EPI_ISL_764076, EPI_ISL_764099 to 764100, EPI_ISL_764103 to 764110, EPI_ISL_764112, EPI_ISL_764144 to 764145, EPI_ISL_764153, EPI_ISL_764167, EPI_ISL_764210, EPI_ISL_764213, EPI_ISL_764216, EPI_ISL_764226 to 764227, EPI_ISL_764231, EPI_ISL_764269, EPI_ISL_765149 to 765158, EPI_ISL_765160 to 765166, EPI_ISL_765168 to 765187, EPI_ISL_765189 to 765208, EPI_ISL_816226 to 816228, EPI_ISL_816230 to 816234, EPI_ISL_816236, EPI_ISL_816238 to 816242, EPI_ISL_816246 to 816250, EPI_ISL_816252, EPI_ISL_816254 to 816257, EPI_ISL_816259, EPI_ISL_816261 to 816263, EPI_ISL_816266, EPI_ISL_816269, EPI_ISL_816271 to 816277, EPI_ISL_816279 to 816280, EPI_ISL_816282 to 816283, EPI_ISL_816285, EPI_ISL_816288 to 816292, EPI_ISL_816295 to 816298, EPI_ISL_816300, EPI_ISL_816302, EPI_ISL_816304 to 816306, EPI_ISL_816308, EPI_ISL_816310 to 816311, EPI_ISL_816313 to 816314, EPI_ISL_816317 to 816320, EPI_ISL_816323 to 816324, EPI_ISL_816327, EPI_ISL_816330 to 816332, EPI_ISL_816335, EPI_ISL_816337, EPI_ISL_816339 to 816340, EPI_ISL_816342 to 816343, EPI_ISL_816346 to 816354, EPI_ISL_816356 to 816357, EPI_ISL_816359 to 816362, EPI_ISL_816364 to 816365, EPI_ISL_816368, EPI_ISL_816370, EPI_ISL_816372, EPI_ISL_816374 to 816375, EPI_ISL_816379 to 816381, EPI_ISL_816383, EPI_ISL_816385 to 816386, EPI_ISL_816388, EPI_ISL_816390, EPI_ISL_816392 to 816393, EPI_ISL_816396 to 816398, EPI_ISL_816400 to 816401, EPI_ISL_816403 to 816407, EPI_ISL_816409 to 816410, EPI_ISL_816414 to 816415, EPI_ISL_816418 to 816419, EPI_ISL_816421 to 816423, EPI_ISL_816425 to 816431, EPI_ISL_816434 to 816435, EPI_ISL_816437 to 816438, EPI_ISL_816440 to 816449, EPI_ISL_816453 to 816454, EPI_ISL_816456 to 816457, EPI_ISL_816459, EPI_ISL_816461 to 816462, EPI_ISL_816464 to 816469, EPI_ISL_816472 to 816475, EPI_ISL_816477 to 816478, EPI_ISL_816480 to 816485, EPI_ISL_816488 to 816492, EPI_ISL_816494 to 816497, EPI_ISL_816499 to 816501, EPI_ISL_816503 to 816505, EPI_ISL_816507 to 816508, EPI_ISL_816510 to 816516, EPI_ISL_816521 to 816522, EPI_ISL_816525, EPI_ISL_816528 to 816534, EPI_ISL_816536 to 816537, EPI_ISL_816539 to 816540, EPI_ISL_816542, EPI_ISL_816544 to 816545, EPI_ISL_816547 to 816549, EPI_ISL_816551, EPI_ISL_816554 to 816558, EPI_ISL_816560 to 816563, EPI_ISL_816566 to 816571, EPI_ISL_816574, EPI_ISL_816576 to 816582, EPI_ISL_816584 to 816587, EPI_ISL_816589, EPI_ISL_816592 to 816595, EPI_ISL_816598 to 816599, EPI_ISL_816601 to 816602, EPI_ISL_816604 to 816607, EPI_ISL_816610 to 816613, EPI_ISL_816615 to 816616, EPI_ISL_816619 to 816621, EPI_ISL_816625, EPI_ISL_816627 to 816631, EPI_ISL_816633 to 816634, EPI_ISL_816637 to 816638, EPI_ISL_816640 to 816643, EPI_ISL_816645 to 816647, EPI_ISL_816651, EPI_ISL_842204, EPI_ISL_842209, EPI_ISL_842217 to 842218, EPI_ISL_842228, EPI_ISL_842230, EPI_ISL_842233, EPI_ISL_842248, EPI_ISL_842254 to 842256, EPI_ISL_842263, EPI_ISL_842265 to 842266, EPI_ISL_842270, EPI_ISL_842272 to 842273, EPI_ISL_842281 to 842282, EPI_ISL_842291 to 842293, EPI_ISL_842297, EPI_ISL_842306, EPI_ISL_842312 to 842313, EPI_ISL_842316 to 842319, EPI_ISL_842325, EPI_ISL_842335, EPI_ISL_842338, EPI_ISL_842341, EPI_ISL_842347 to 842354, EPI_ISL_868372, EPI_ISL_868414, EPI_ISL_868419 to 868420, EPI_ISL_868423, EPI_ISL_868436, EPI_ISL_868438, EPI_ISL_868475, EPI_ISL_868483, EPI_ISL_868485, EPI_ISL_868512, EPI_ISL_868559, EPI_ISL_868570, EPI_ISL_868576 to 868577, EPI_ISL_868614, EPI_ISL_868628 to 868629, EPI_ISL_868635, EPI_ISL_868645, EPI_ISL_868649, EPI_ISL_868669, EPI_ISL_868673 | | | |
| see above | Virology Department, Sheffield Teaching Hospitals NHS Foundation Trust/Department of Infection, Immunity and Cardiovascular Disease, The Medical School, University of Sheffield | COVID-19 Genomics UK (COG-UK) Consortium | Adri Angyal; Alex Keeley; Benjamin Lindsey; Cariad Evans; Danielle Groves; Dave Partridge; Katie Johnson; Laura Carrilero; Luke Green; Matthew Parker; Matthew Wyles; Mehmet Yavuz; Mohammad Raza; Nikki Smith; Paul Parsons; Rachel Tucker; Rebecca Brown; Thushan de Silva |
| EPI_ISL_778868 | Virology Lab, Ospedali Riuniti di Ancona | Dipartimento di Scienze Biomediche e Cliniche, L.Sacco, Università di Milano | Alessia Lai; Annalisa Bergna; Claudia Balotta; Gianni Zehender; Laura Di Sante; Massimo Galli; Patrizia Bagnarelli; Roberta Longo; Sara Caucci; Sofia Maria Luigia Tiano; Stefano Menzo |
| EPI_ISL_778869 | Virology Lab, Ospedali Riuniti, Ancona | Dipartimento di Scenze Biomediche e Cliniche, L. Sacco, Università di Milano | Alessia Lai; Annalisa Bergna; Claudia Balotta; Gianni Zehender; Laura Di Sante; Massimo Galli; Patrizia Bagnarelli; Roberta Longo; Sara Caucci; Sofia Maria Luigia Tiano; Stefano Menzo |
| EPI_ISL_448844, EPI_ISL_448850, EPI_ISL_448864, EPI_ISL_448866, EPI_ISL_448894, EPI_ISL_448896, EPI_ISL_448902, EPI_ISL_448906 | | | |
| see above | Virology Laboratory, Castle Hill Hospital, Hull University Teaching Hospitals NHS Trust/Department of Infection, Immunity and Cardiovascular Disease, The Medical School, University of Sheffield | COVID-19 Genomics UK (COG-UK) Consortium | Adri Angyal; Alex Keeley; Benjamin Lindsey; Cariad Evans; Danielle Groves; Dave Partridge; Katie Johnson; Laura Carrilero; Luke Green; Matthew Parker; Matthew Wyles; Mehmet Yavuz; Mohammad Raza; Nikki Smith; Paul Parsons; Rachel Tucker; Rebecca Brown; Thushan de Silva |
| EPI_ISL_417491 | Virology Laboratory, Department of Biomedical Sciences and Public Health, University Politecnica delle Marche | Virology and Legal Medicine Laboratories, Department of Biomedical Sciences and Public Health, University Politecnica delle Marche | Alessandrini, F.; Bagnarelli, P.; Caucci, S.; Di Sante, L.; Menzo, S.; Onofri, V.; Tagliabracci, A.; Turchi, C. |
| EPI_ISL_856869 to 856870, EPI_ISL_856872 to 856879, EPI_ISL_856882 to 856883, EPI_ISL_856885 | Virology Laboratory, Scientific Department, Army Medical Center | Virology Laboratory, Scientific Department, Army Medical Center | Anella Monte; Anna Anselmo; Antonella Fortunato; Filippo Molinari; Florigio Lista; Francesco Giordani; Giancarlo Petralito; Giovanni Faggioni; Nino D'Amore; Riccardo De Sanctis; Silvia Fillo; Vanessa Vera Fain |
| EPI_ISL_508862 to 508863 | Virology Unit, Institut Pasteur de Madagascar | Virology Unit, Institut Pasteur de Madagascar | Cara Brook; Christian Ranaivoson; Cristina M. Tato; Helisoa Razafimanjato; Jean-Michel Heraud; Joseph L. DeRisi; Michelle Tan; Norosoa Razanajatovo; Soa Fy Andriamandimby; Tsiry Randriambolamanantsoa; Vida Ahyong; Vololoniaina Raharinosy |
| EPI_ISL_803896 | Virology Unit, Pisa University Hospital and Retrovirus Center, University of Pisa | National Institute for Infectious Diseases, INMI, "L. Spallanzani" IRCCS | A Di Caro; A Rosellini; B Bartolini; C.E.M Gruber; E Giombini; F Messina; M Pistello; M Rueca; MR Capobianchi; O Butera; P Mazzetti |
| EPI_ISL_779254 | Virology, CHU Pitie Salpetriere Charles Foix | Virology, CHU Pitie Salpetriere Charles Foix | A.-G.; Boutolleau, D.; Bruel, T.; Burrel, S.; Calvez; Dorgham, K.; Gorochov, G.; Gothland, A.; Jary, A.; Leducq, V.; Malet, I.; Marot, S.; Planas, D.; Schwartz, O.; Sterlin, D.; V. and Marcelin; Zafilaza, K. |
| EPI_ISL_862814 | Virology, Ecole Nationale Veterinaire de Toulouse | Virology, Ecole Nationale Veterinaire de Toulouse | Bessiere, P.; Cadiergues; Croville, G.; Fusade-Boyer, M.; J.-L.; Lebre, L.; M.-C. and Guerin; Walch, M. |
| EPI_ISL_610245 | Virology, National Institute for Biological Standards and Control | Virology, National Institute for Biological Standards and Control | Bentley, E.; E.T.; Fritzsche; M. and Mattiuzzo, G.; Mate, R.; Mee; Routley, S. |
| EPI_ISL_707943, EPI_ISL_707947 to 707951, EPI_ISL_707953 to 707968, EPI_ISL_707970 to 708003, EPI_ISL_708010 to 708015 | Virology, Universitätsklinikum des Saarlandes | Epigenetics, Saarland University | Jörn Walter; Kathrin Kattler; Markus Vogelgesang; Sascha Tierling; Sigrun Smola; Stefan Lohse |
| EPI_ISL_568579 | Virus Molecular Laboratory of the Microbiology and Virology Department | INMI Lazzaro Spallanzani IRCCS | Antonino Di Caro; Barbara Bartolini; Cesare E.M. Gruber; Concetta Castilletti; Edoardo Vecchio Nepita; Elena Percivalle; Fausto Baldanti; Francesca Colavita; Francesco Messina; Irene Cassaniti; Maria R. Capobianchi; Martina Rueca; Silvia Meschi |
| EPI_ISL_722201 | Vitalis Mostar | Alea Genetic Center | Konjhodzic R.; Pecar D.; Salihefendic L. |
| EPI_ISL_415710, EPI_ISL_420080 to 420081, EPI_ISL_427307 to 427339, EPI_ISL_427815, EPI_ISL_430067 to 430112, EPI_ISL_450241 to 450293, EPI_ISL_507205, EPI_ISL_507216 to 507227, EPI_ISL_507229 to 507291, EPI_ISL_507293 to 507296, EPI_ISL_507300, EPI_ISL_523995 to 524052, EPI_ISL_596228 to 596249, EPI_ISL_596264 to 596267, EPI_ISL_596353 to 596356, EPI_ISL_602413 to 602416, EPI_ISL_732971 to 732989, EPI_ISL_733005 to 733022, EPI_ISL_733154 to 733155, EPI_ISL_733183 to 733184, EPI_ISL_733241 to 733300, EPI_ISL_733392, EPI_ISL_733394 to 733395, EPI_ISL_733402 to 733404, EPI_ISL_733459 to 733477, EPI_ISL_733494 to 733495, EPI_ISL_770633 to 770637, EPI_ISL_770639 to 770641, EPI_ISL_770643 to 770646, EPI_ISL_770653, EPI_ISL_770658, EPI_ISL_770660 to 770661, EPI_ISL_770666, EPI_ISL_770669, EPI_ISL_770673 to 770687, EPI_ISL_872898 to 872927, EPI_ISL_872929 to 872930, EPI_ISL_872932 to 872951, EPI_ISL_872953 to 872964, EPI_ISL_872966 to 872971 | | | |
| see above | WHO National Influenza Centre Russian Federation | WHO National Influenza Centre Russian Federation | Andrey Komissarov; Anna Ivanova; Artem Fadeev; Daria Danilenko; Dmitry Bazhenov; Dmitry Lioznov; Elena Nabieva; Georgii Bazykin; Ksenia Komissarova; Ksenia Safina; Kseniya Komissarova; Mariia Pisareva; Mariia Sergeeva; Mariia Timofeeva; Mikhail Bakaev; Tamila Musaeva; Veronica Eder |
| EPI_ISL_860797 to 860798, EPI_ISL_860800 to 860818, EPI_ISL_861456 to 861458 | WHO/Minsk | Charité Universitätsmedizin Berlin, Institut für Virologie | Barbara Mühlemann; Christian Drosten; Julia Schneider; Julia Tesch; Jörn Beheim-Schwarzbach; Shmialiova Natallia; Sivets Natallia; Talitha Veith; Terry Jones; Tobias Bleicker; Victor M Corman |
| EPI_ISL_413555 to 413556, EPI_ISL_414486, EPI_ISL_414488, EPI_ISL_415041, EPI_ISL_415435, EPI_ISL_415453, EPI_ISL_415536 to 415537, EPI_ISL_415655 to 415657, EPI_ISL_415659, EPI_ISL_415916, EPI_ISL_415918 to 415920, EPI_ISL_415977 to 415978, EPI_ISL_415991, EPI_ISL_416024, EPI_ISL_416026, EPI_ISL_418083, EPI_ISL_418085 to 418087, EPI_ISL_418090, EPI_ISL_418093 to 418094, EPI_ISL_418096, EPI_ISL_418100 to 418104, EPI_ISL_418107, EPI_ISL_418110, EPI_ISL_418112, EPI_ISL_418114 to 418115, EPI_ISL_418117, EPI_ISL_418120 to 418140, EPI_ISL_418142 to 418147, EPI_ISL_418149 to 418151, EPI_ISL_418153 to 418163, EPI_ISL_418165, EPI_ISL_419399 to 419400, EPI_ISL_419402 to 419413, EPI_ISL_419415 to 419418, EPI_ISL_419420, EPI_ISL_419422, EPI_ISL_419424 to 419428, EPI_ISL_419430 to 419434, EPI_ISL_419436 to 419437, EPI_ISL_419439 to 419451, EPI_ISL_419453 to 419455, EPI_ISL_419457 to 419466, EPI_ISL_419468 to 419472, EPI_ISL_419474 to 419493, EPI_ISL_419495 to 419499, EPI_ISL_419502 to 419504, EPI_ISL_419506 to 419511, EPI_ISL_420913, EPI_ISL_420918 to 420920, EPI_ISL_420922, EPI_ISL_420925 to 420933, EPI_ISL_420935 to 420941, EPI_ISL_420943 to 420945, EPI_ISL_420947 to 420951, EPI_ISL_420953, EPI_ISL_420955 to 420956, EPI_ISL_420958 to 420974, EPI_ISL_420976 to 420978, EPI_ISL_420980 to 421011, EPI_ISL_422016 to 422086, EPI_ISL_422088 to 422100, EPI_ISL_422102, EPI_ISL_422104 to 422112, EPI_ISL_422114 to 422139, EPI_ISL_422141 to 422151, EPI_ISL_422153, EPI_ISL_422155 to 422161, EPI_ISL_422163 to 422170, EPI_ISL_422172 to 422175, EPI_ISL_422179 to 422185, EPI_ISL_422188 to 422196, EPI_ISL_422199 to 422210, EPI_ISL_422212 to 422214, EPI_ISL_422216 to 422244, EPI_ISL_422246 to 422261, EPI_ISL_422263 to 422269, EPI_ISL_422271 to 422273, EPI_ISL_422275 to 422287, EPI_ISL_422289 to 422292, EPI_ISL_422294 to 422296, EPI_ISL_422298, EPI_ISL_422301 to 422307, EPI_ISL_422309 to 422335, EPI_ISL_422337 to 422339, EPI_ISL_422341 to 422350, EPI_ISL_422352 to 422355, EPI_ISL_422357 to 422358, EPI_ISL_422360 to 422363, EPI_ISL_422365 to 422366, EPI_ISL_422368 to 422370, EPI_ISL_422376 to 422377, EPI_ISL_431901 to 431915, EPI_ISL_431917 to 431950, EPI_ISL_431952 to 431957, EPI_ISL_431959 to 431968, EPI_ISL_431970 to 431977, EPI_ISL_431981 to 431992, EPI_ISL_431994 to 432032, EPI_ISL_432034 to 432043, EPI_ISL_432045 to 432057, EPI_ISL_432059 to 432060, EPI_ISL_432062, EPI_ISL_432065 to 432067, EPI_ISL_432069, EPI_ISL_432072 to 432086, EPI_ISL_432088 to 432092, EPI_ISL_432094 to 432101, EPI_ISL_432103, EPI_ISL_432105, EPI_ISL_432107 to 432115, EPI_ISL_432117 to 432118, EPI_ISL_432120, EPI_ISL_432122 to 432124, EPI_ISL_432126 to 432127, EPI_ISL_432130 to 432132, EPI_ISL_432134 to 432135, EPI_ISL_432137 to 432138, EPI_ISL_432140, EPI_ISL_432142, EPI_ISL_432144 to 432147, EPI_ISL_432151 to 432154, EPI_ISL_432158 to 432159, EPI_ISL_432164, EPI_ISL_432169 to 432170, EPI_ISL_432175 to 432183, EPI_ISL_432185 to 432235, EPI_ISL_432237 to 432281, EPI_ISL_432283 to 432319, EPI_ISL_432321 to 432374, EPI_ISL_432376 to 432403, EPI_ISL_432405 to 432450, EPI_ISL_445382 to 445385, EPI_ISL_445387 to 445400, EPI_ISL_445402 to 445451, EPI_ISL_445453 to 445492, EPI_ISL_445494, EPI_ISL_445496 to 445515, EPI_ISL_445519 to 445523, EPI_ISL_445525 to 445527, EPI_ISL_445529 to 445531, EPI_ISL_445534 to 445542, EPI_ISL_445544 to 445546, EPI_ISL_445548 to 445549, EPI_ISL_445551 to 445555, EPI_ISL_445560 to 445561, EPI_ISL_445563 to 445567, EPI_ISL_445571 to 445574, EPI_ISL_445576, EPI_ISL_445578, EPI_ISL_445582 to 445585, EPI_ISL_445588 to 445602, EPI_ISL_445604 to 445608, EPI_ISL_445610, EPI_ISL_445612, EPI_ISL_445614 to 445619, EPI_ISL_445621 to 445657, EPI_ISL_445659 to 445675, EPI_ISL_445677 to 445682, EPI_ISL_445684 to 445703, EPI_ISL_445705, EPI_ISL_445707, EPI_ISL_445709 to 445712, EPI_ISL_445714 to 445728, EPI_ISL_445731 to 445738, EPI_ISL_445740 to 445741, EPI_ISL_445743 to 445744, EPI_ISL_445746 to 445749, EPI_ISL_445751, EPI_ISL_445753 to 445757, EPI_ISL_445759 to 445764, EPI_ISL_445766 to 445768, EPI_ISL_445770 to 445772, EPI_ISL_445774 to 445776, EPI_ISL_445778 to 445779, EPI_ISL_445781 to 445782, EPI_ISL_445784 to 445800, EPI_ISL_445802 to 445803, EPI_ISL_445805 to 445813, EPI_ISL_445815 to 445824, EPI_ISL_445826 to 445834, EPI_ISL_445836 to 445843, EPI_ISL_445845 to 445857, EPI_ISL_445859 to 445879, EPI_ISL_445881 to 445882, EPI_ISL_445884, EPI_ISL_445886 to 445890, EPI_ISL_445892 to 445898, EPI_ISL_445901 to 445902, EPI_ISL_445904 to 445917, EPI_ISL_445919 to 445928, EPI_ISL_445931 to 445934, EPI_ISL_445937 to 445939, EPI_ISL_445941 to 445946, EPI_ISL_445948 to 445956, EPI_ISL_445958 to 445966, EPI_ISL_445968 to 445980, EPI_ISL_445982 to 445988, EPI_ISL_445990, EPI_ISL_445992 to 446002, EPI_ISL_446004 to 446008, EPI_ISL_446011 to 446018, EPI_ISL_446024 to 446035, EPI_ISL_446037 to 446056, EPI_ISL_446058, EPI_ISL_446060 to 446072, EPI_ISL_446074 to 446077, EPI_ISL_446079 to 446083, EPI_ISL_446085 to 446087, EPI_ISL_446089 to 446090, EPI_ISL_446093 to 446094, EPI_ISL_446098 to 446109, EPI_ISL_446111 to 446179, EPI_ISL_446181 to 446200, EPI_ISL_446202 to 446260, EPI_ISL_446263, EPI_ISL_446265 to 446273, EPI_ISL_446276, EPI_ISL_446278, EPI_ISL_446280 to 446297, EPI_ISL_446299 to 446357, EPI_ISL_446359 to 446373, EPI_ISL_446375 to 446396, EPI_ISL_446402 to 446405, EPI_ISL_446407 to 446408, EPI_ISL_446410 to 446430, EPI_ISL_446432 to 446449, EPI_ISL_446452 to 446455, EPI_ISL_446457 to 446461, EPI_ISL_446463 to 446468, EPI_ISL_446470 to 446494, EPI_ISL_446496 to 446498, EPI_ISL_446501 to 446512, EPI_ISL_446514 to 446525, EPI_ISL_446527 to 446560, EPI_ISL_446562 to 446566, EPI_ISL_446568 to 446574, EPI_ISL_446576 to 446584, EPI_ISL_446586 to 446615, EPI_ISL_446617 to 446651, EPI_ISL_446653 to 446654, EPI_ISL_446656 to 446658, EPI_ISL_446660, EPI_ISL_446662 to 446699, EPI_ISL_446701 to 446707, EPI_ISL_446709, EPI_ISL_446711 to 446737, EPI_ISL_446739, EPI_ISL_446741 to 446742, EPI_ISL_446744 to 446746, EPI_ISL_446748 to 446749, EPI_ISL_446751 to 446825, EPI_ISL_446827 to 446828, EPI_ISL_446830 to 446891, EPI_ISL_446893 to 446897, EPI_ISL_446900 to 446906, EPI_ISL_446908 to 446909, EPI_ISL_446911 to 446912, EPI_ISL_446914 to 446924, EPI_ISL_446926 to 446934, EPI_ISL_446936 to 446943, EPI_ISL_446946 to 446950, EPI_ISL_446952 to 446964, EPI_ISL_446966 to 446968, EPI_ISL_446974, EPI_ISL_446976, EPI_ISL_446978 to 446979, EPI_ISL_446981 to 446984, EPI_ISL_446986 to 446989, EPI_ISL_446991 to 446992, EPI_ISL_446994 to 446995 | | | |

| | | | |
|---|---|---|---|
| see above | Wales Specialist Virology Centre | Public Health Wales Microbiology Cardiff | Alec Birchley; Alexander Adams; Amy Gaskin; Bree Gatica-Wilcox; Catherine Moore; Cen Sabu; Jason Coombes; Joanne Watkins; Joanne watkins; Johnathan Evans; Laura Gifford; Lauren Gilbert; Lee Graham; Malorie Perry; Matt Bull; Matthew Bull; Nicole Pacchiarini; Sally Corden; Sara Kumziene-Summerhayes; Sara Rey; Sarah Taylor; Simon Cottrell; Simon Cottrell Sara Rey; Sophie Jones; Tim Jones; Tom Connor |

EPI_ISL_472432 to 472433, EPI_ISL_472435 to 472446, EPI_ISL_472448 to 472466, EPI_ISL_472468 to 472470, EPI_ISL_472472 to 472486, EPI_ISL_472488 to 472489, EPI_ISL_472491 to 472501, EPI_ISL_472503 to 472506, EPI_ISL_472508 to 472524, EPI_ISL_472526 to 472576, EPI_ISL_472578 to 472587, EPI_ISL_472589 to 472598, EPI_ISL_472601 to 472603, EPI_ISL_472605 to 472619, EPI_ISL_472621 to 472631, EPI_ISL_472633 to 472662, EPI_ISL_472664 to 472678, EPI_ISL_472680 to 472689, EPI_ISL_472691 to 472697, EPI_ISL_472699 to 472720, EPI_ISL_472722 to 472737, EPI_ISL_472740 to 472744, EPI_ISL_472746 to 472750, EPI_ISL_472753 to 472755, EPI_ISL_472757 to 472758, EPI_ISL_472760 to 472773, EPI_ISL_472775 to 472787, EPI_ISL_472789 to 472795, EPI_ISL_472798 to 472801, EPI_ISL_472803 to 472806, EPI_ISL_472808 to 472821, EPI_ISL_472823 to 472828, EPI_ISL_472830, EPI_ISL_472833 to 472839, EPI_ISL_472843 to 472863, EPI_ISL_472865 to 472880, EPI_ISL_472882 to 472899, EPI_ISL_472901 to 472914, EPI_ISL_472916 to 472920, EPI_ISL_472922 to 472927, EPI_ISL_472929 to 472944, EPI_ISL_472946 to 472954, EPI_ISL_472956 to 472977, EPI_ISL_472979, EPI_ISL_472981 to 472988, EPI_ISL_472990 to 473012, EPI_ISL_473015 to 473016, EPI_ISL_473018 to 473033, EPI_ISL_473035 to 473048, EPI_ISL_473050 to 473076, EPI_ISL_473078 to 473079, EPI_ISL_473081 to 473099, EPI_ISL_473101 to 473105, EPI_ISL_473108 to 473112, EPI_ISL_473115 to 473146, EPI_ISL_473148 to 473149, EPI_ISL_473151, EPI_ISL_473153 to 473158, EPI_ISL_473160 to 473191, EPI_ISL_473193 to 473207, EPI_ISL_473209 to 473224, EPI_ISL_473226 to 473230, EPI_ISL_473232 to 473234, EPI_ISL_473236 to 473239, EPI_ISL_473241 to 473251, EPI_ISL_473253 to 473256, EPI_ISL_473258 to 473267, EPI_ISL_473269 to 473270, EPI_ISL_473272 to 473280, EPI_ISL_473282 to 473283, EPI_ISL_473285 to 473287, EPI_ISL_473289, EPI_ISL_473291 to 473298, EPI_ISL_473300 to 473305, EPI_ISL_473956 to 473970, EPI_ISL_473972, EPI_ISL_473974 to 473996, EPI_ISL_473998 to 474031, EPI_ISL_474033 to 474055, EPI_ISL_474057 to 474067, EPI_ISL_474069 to 474075, EPI_ISL_474077 to 474086, EPI_ISL_474088 to 474089, EPI_ISL_474091 to 474092, EPI_ISL_474094 to 474096, EPI_ISL_474098 to 474102, EPI_ISL_474104 to 474137, EPI_ISL_474139 to 474143, EPI_ISL_474146 to 474153, EPI_ISL_474155 to 474175, EPI_ISL_474177 to 474221, EPI_ISL_474223 to 474227, EPI_ISL_474230 to 474233, EPI_ISL_474235 to 474242, EPI_ISL_474244 to 474246, EPI_ISL_474248 to 474250, EPI_ISL_474253 to 474256, EPI_ISL_474258, EPI_ISL_474260 to 474268, EPI_ISL_474270 to 474277, EPI_ISL_474280 to 474299, EPI_ISL_474301 to 474310, EPI_ISL_474312 to 474319, EPI_ISL_474321 to 474326, EPI_ISL_474328 to 474381, EPI_ISL_474383 to 474402, EPI_ISL_474404 to 474408, EPI_ISL_474419, EPI_ISL_474431 to 474433, EPI_ISL_474435 to 474440, EPI_ISL_474442 to 474446, EPI_ISL_474448 to 474455, EPI_ISL_474457 to 474465, EPI_ISL_474467 to 474475, EPI_ISL_474477 to 474487, EPI_ISL_474489 to 474500, EPI_ISL_474502 to 474509, EPI_ISL_474511 to 474535, EPI_ISL_474537 to 474545, EPI_ISL_474547 to 474556, EPI_ISL_474558, EPI_ISL_474560 to 474576, EPI_ISL_474578 to 474600, EPI_ISL_474603 to 474639, EPI_ISL_474641 to 474672, EPI_ISL_474674 to 474691, EPI_ISL_474693 to 474733, EPI_ISL_474735 to 474744, EPI_ISL_474747 to 474755, EPI_ISL_474757 to 474796, EPI_ISL_479284 to 479300, EPI_ISL_479302 to 479305, EPI_ISL_479307 to 479312, EPI_ISL_479314 to 479319, EPI_ISL_479321, EPI_ISL_479323 to 479334, EPI_ISL_479337 to 479339, EPI_ISL_479341 to 479344, EPI_ISL_479346 to 479352, EPI_ISL_479354 to 479357, EPI_ISL_479359 to 479365, EPI_ISL_479368, EPI_ISL_479370 to 479373, EPI_ISL_479376 to 479377, EPI_ISL_479379 to 479381, EPI_ISL_479383 to 479388, EPI_ISL_479391 to 479410, EPI_ISL_479412 to 479415, EPI_ISL_479418 to 479424, EPI_ISL_479426 to 479443, EPI_ISL_479445 to 479448, EPI_ISL_479450 to 479457, EPI_ISL_479460 to 479466, EPI_ISL_479468 to 479470, EPI_ISL_479472 to 479476, EPI_ISL_479478 to 479481, EPI_ISL_484685 to 484691, EPI_ISL_490710 to 490719, EPI_ISL_490722 to 490727, EPI_ISL_490729 to 490736, EPI_ISL_490739 to 490741, EPI_ISL_490743 to 490747, EPI_ISL_490750 to 490751, EPI_ISL_490753 to 490777, EPI_ISL_490780 to 490783, EPI_ISL_490785 to 490788, EPI_ISL_490790 to 490800, EPI_ISL_490802, EPI_ISL_490804 to 490809, EPI_ISL_490811 to 490812, EPI_ISL_490814 to 490818, EPI_ISL_490820 to 490836, EPI_ISL_490838 to 490845, EPI_ISL_490847 to 490848, EPI_ISL_490850 to 490859, EPI_ISL_490861 to 490866, EPI_ISL_493975 to 493976, EPI_ISL_493979, EPI_ISL_493982, EPI_ISL_493984 to 494066, EPI_ISL_494068 to 494094, EPI_ISL_494096 to 494120, EPI_ISL_494122 to 494156, EPI_ISL_494158 to 494159, EPI_ISL_494161 to 494182, EPI_ISL_494184 to 494185, EPI_ISL_494187 to 494192, EPI_ISL_494194 to 494197, EPI_ISL_494200 to 494203, EPI_ISL_494205 to 494208, EPI_ISL_494210 to 494237, EPI_ISL_494239 to 494284, EPI_ISL_494286 to 494288, EPI_ISL_494290 to 494292, EPI_ISL_494294 to 494315, EPI_ISL_494317 to 494356, EPI_ISL_494359 to 494371, EPI_ISL_499355 to 499359, EPI_ISL_499361 to 499363, EPI_ISL_499365 to 499390, EPI_ISL_499392 to 499406, EPI_ISL_499408 to 499459, EPI_ISL_512481 to 512546, EPI_ISL_514563, EPI_ISL_514565 to 514579, EPI_ISL_517590 to 517609, EPI_ISL_529226 to 529233, EPI_ISL_529253, EPI_ISL_529297, EPI_ISL_529418, EPI_ISL_529447, EPI_ISL_529527, EPI_ISL_529697 to 529717, EPI_ISL_535184 to 535186, EPI_ISL_535188 to 535215, EPI_ISL_535217 to 535262, EPI_ISL_540897 to 540904, EPI_ISL_540906 to 540920, EPI_ISL_560083 to 560088, EPI_ISL_560090 to 560235, EPI_ISL_560237 to 560250, EPI_ISL_560252 to 560304, EPI_ISL_572405, EPI_ISL_572407 to 572408, EPI_ISL_572434, EPI_ISL_572446, EPI_ISL_572455, EPI_ISL_572461, EPI_ISL_572465, EPI_ISL_572468 to 572469, EPI_ISL_572474, EPI_ISL_572477, EPI_ISL_572487, EPI_ISL_572494, EPI_ISL_572496, EPI_ISL_572501, EPI_ISL_572512, EPI_ISL_572531, EPI_ISL_572540, EPI_ISL_572545, EPI_ISL_572547, EPI_ISL_572555, EPI_ISL_572558, EPI_ISL_572560 to 572561, EPI_ISL_572564 to 572566, EPI_ISL_572574 to 572575, EPI_ISL_572577, EPI_ISL_572580, EPI_ISL_572586, EPI_ISL_572605 to 572606, EPI_ISL_572611, EPI_ISL_572614, EPI_ISL_572643, EPI_ISL_572645, EPI_ISL_572647, EPI_ISL_572652, EPI_ISL_572654, EPI_ISL_572657, EPI_ISL_572659, EPI_ISL_572661 to 572662, EPI_ISL_572667 to 572668, EPI_ISL_572672, EPI_ISL_572674 to 572676, EPI_ISL_572680 to 572681, EPI_ISL_572683, EPI_ISL_572690, EPI_ISL_572693 to 572694, EPI_ISL_572697 to 572698, EPI_ISL_572708, EPI_ISL_572731, EPI_ISL_572754, EPI_ISL_572766, EPI_ISL_572772, EPI_ISL_572778, EPI_ISL_572781 to 572782, EPI_ISL_572784, EPI_ISL_572790, EPI_ISL_572805, EPI_ISL_572816, EPI_ISL_572818, EPI_ISL_572830, EPI_ISL_572836, EPI_ISL_572853 to 572856, EPI_ISL_572875 to 572890, EPI_ISL_572931 to 572932, EPI_ISL_572951, EPI_ISL_573873 to 573893, EPI_ISL_573895 to 573927, EPI_ISL_573929 to 573930, EPI_ISL_573932 to 573936, EPI_ISL_573938 to 573985, EPI_ISL_573987 to 574003, EPI_ISL_574005 to 574053, EPI_ISL_574055 to 574064, EPI_ISL_574066 to 574110, EPI_ISL_574112 to 574240, EPI_ISL_574242 to 574256, EPI_ISL_577417 to 577455, EPI_ISL_577457 to 577480, EPI_ISL_577482 to 577483, EPI_ISL_577485 to 577513, EPI_ISL_577515 to 577543, EPI_ISL_585631, EPI_ISL_585633 to 585656, EPI_ISL_585658 to 585679, EPI_ISL_585681 to 585694, EPI_ISL_585696 to 585785, EPI_ISL_585787 to 585802, EPI_ISL_585804 to 585829, EPI_ISL_585831 to 585839, EPI_ISL_585841 to 585866, EPI_ISL_585868 to 585893, EPI_ISL_585895 to 585940, EPI_ISL_585942 to 585946, EPI_ISL_585948, EPI_ISL_585950 to 585952, EPI_ISL_585954 to 586011, EPI_ISL_586013 to 586030, EPI_ISL_586032 to 586045, EPI_ISL_586047 to 586051, EPI_ISL_586053, EPI_ISL_586055 to 586094, EPI_ISL_586096 to 586100, EPI_ISL_586102 to 586151, EPI_ISL_586153 to 586211, EPI_ISL_586213 to 586224, EPI_ISL_586226 to 586235, EPI_ISL_586237 to 586239, EPI_ISL_595329 to 595363, EPI_ISL_595365 to 595399, EPI_ISL_595401 to 595423, EPI_ISL_595425 to 595440, EPI_ISL_595442 to 595450, EPI_ISL_595452 to 595467, EPI_ISL_595469 to 595478, EPI_ISL_595480 to 595529, EPI_ISL_595531 to 595587, EPI_ISL_595589 to 595596, EPI_ISL_595598 to 595612, EPI_ISL_595614 to 595616, EPI_ISL_611518 to 611519, EPI_ISL_611529 to 611530, EPI_ISL_611534, EPI_ISL_611560 to 611564, EPI_ISL_611566, EPI_ISL_611573, EPI_ISL_611578, EPI_ISL_611581, EPI_ISL_611586, EPI_ISL_611588, EPI_ISL_611601, EPI_ISL_611605, EPI_ISL_611609, EPI_ISL_611624 to 611626, EPI_ISL_611631, EPI_ISL_611633, EPI_ISL_611636, EPI_ISL_611640, EPI_ISL_611652, EPI_ISL_611663, EPI_ISL_611666, EPI_ISL_611673 to 611674, EPI_ISL_611683, EPI_ISL_611686 to 611688, EPI_ISL_611691, EPI_ISL_611697, EPI_ISL_611704, EPI_ISL_611706, EPI_ISL_611711, EPI_ISL_611722, EPI_ISL_611725 to 611726, EPI_ISL_611732, EPI_ISL_611735, EPI_ISL_611738, EPI_ISL_611740, EPI_ISL_611742, EPI_ISL_611748 to 611749, EPI_ISL_611792, EPI_ISL_611809, EPI_ISL_611819, EPI_ISL_611824, EPI_ISL_611828, EPI_ISL_611832, EPI_ISL_611834 to 611835, EPI_ISL_611849, EPI_ISL_611863, EPI_ISL_611865, EPI_ISL_611886 to 611888, EPI_ISL_611896, EPI_ISL_611900 to 611907, EPI_ISL_611909, EPI_ISL_611912 to 611913, EPI_ISL_611922 to 611923, EPI_ISL_611926, EPI_ISL_611931 to 611932, EPI_ISL_611968 to 611970, EPI_ISL_611972 to 611973, EPI_ISL_611975, EPI_ISL_611982, EPI_ISL_611986, EPI_ISL_611992, EPI_ISL_611994, EPI_ISL_612002, EPI_ISL_612006, EPI_ISL_612012 to 612013, EPI_ISL_612018, EPI_ISL_612029, EPI_ISL_612038, EPI_ISL_612040, EPI_ISL_612048 to 612049, EPI_ISL_612052 to 612053, EPI_ISL_612076, EPI_ISL_612081, EPI_ISL_612083, EPI_ISL_612109, EPI_ISL_612116 to 612117, EPI_ISL_612119 to 612122, EPI_ISL_612132 to 612133, EPI_ISL_612135 to 612157, EPI_ISL_612160 to 612175, EPI_ISL_612177 to 612180, EPI_ISL_612184 to 612185, EPI_ISL_612193 to 612194, EPI_ISL_612199 to 612206, EPI_ISL_612458 to 612459, EPI_ISL_612461 to 612463, EPI_ISL_612465 to 612486, EPI_ISL_612489 to 612510, EPI_ISL_612512 to 612522, EPI_ISL_612607 to 612655, EPI_ISL_612657 to 612674, EPI_ISL_612676 to 612693, EPI_ISL_612695 to 612721, EPI_ISL_612723 to 612726, EPI_ISL_612728 to 612732, EPI_ISL_612734 to 612771, EPI_ISL_612773 to 612846, EPI_ISL_612848 to 612853, EPI_ISL_612855 to 612870, EPI_ISL_612872 to 612873, EPI_ISL_612875 to 612903, EPI_ISL_612905 to 612945, EPI_ISL_612947 to 612950, EPI_ISL_612952 to 612979, EPI_ISL_612981 to 613007, EPI_ISL_613009 to 613111, EPI_ISL_613113 to 613127, EPI_ISL_613129 to 613138, EPI_ISL_613140 to 613204, EPI_ISL_613206 to 613214, EPI_ISL_613216 to 613273, EPI_ISL_613275 to 613281, EPI_ISL_626658, EPI_ISL_626674 to 626675, EPI_ISL_626696, EPI_ISL_626703, EPI_ISL_626706 to 626708, EPI_ISL_626711, EPI_ISL_626746, EPI_ISL_626748, EPI_ISL_626762 to 626763, EPI_ISL_626768 to 626769, EPI_ISL_626780, EPI_ISL_626783 to 626784, EPI_ISL_626787, EPI_ISL_626796, EPI_ISL_626820, EPI_ISL_626836, EPI_ISL_626850, EPI_ISL_626853, EPI_ISL_626856 to 626857, EPI_ISL_626861 to 626863, EPI_ISL_626865, EPI_ISL_626867, EPI_ISL_626869 to 626872, EPI_ISL_626874 to 626877, EPI_ISL_626920 to 626921, EPI_ISL_626928, EPI_ISL_626931, EPI_ISL_626959 to 626961, EPI_ISL_626965, EPI_ISL_626993, EPI_ISL_626996, EPI_ISL_626998, EPI_ISL_627001 to 627003, EPI_ISL_627006 to 627007, EPI_ISL_627010 to 627013, EPI_ISL_627015, EPI_ISL_627018, EPI_ISL_627021 to 627023, EPI_ISL_627025 to 627029, EPI_ISL_627032 to 627035, EPI_ISL_627037, EPI_ISL_627039, EPI_ISL_627042 to 627044, EPI_ISL_627048, EPI_ISL_627050 to 627051, EPI_ISL_627054 to 627055, EPI_ISL_627069, EPI_ISL_627078, EPI_ISL_627086 to 627097, EPI_ISL_627100 to 627101, EPI_ISL_627105, EPI_ISL_627113, EPI_ISL_627115, EPI_ISL_627119 to 627120, EPI_ISL_627122, EPI_ISL_627124, EPI_ISL_627126, EPI_ISL_627129, EPI_ISL_627134, EPI_ISL_627146, EPI_ISL_627154 to 627155, EPI_ISL_627157, EPI_ISL_627161, EPI_ISL_627167 to 627169, EPI_ISL_627172, EPI_ISL_627197, EPI_ISL_627203 to 627207, EPI_ISL_627212, EPI_ISL_627218 to 627249, EPI_ISL_627265 to 627266, EPI_ISL_627268 to 627279, EPI_ISL_627293, EPI_ISL_627295 to 627296, EPI_ISL_627301, EPI_ISL_627306 to 627308, EPI_ISL_627482 to 627487, EPI_ISL_627489 to 627490, EPI_ISL_627730 to 627752, EPI_ISL_627755 to 627790, EPI_ISL_627792 to 627955, EPI_ISL_627957 to 628015, EPI_ISL_628017 to 628065, EPI_ISL_628067 to 628119, EPI_ISL_628121 to 628262, EPI_ISL_628264 to 628287, EPI_ISL_628289 to 628307, EPI_ISL_628309 to 628311, EPI_ISL_637256, EPI_ISL_637258, EPI_ISL_637288 to 637289, EPI_ISL_637302, EPI_ISL_637319, EPI_ISL_637327, EPI_ISL_637330, EPI_ISL_637332 to 637333, EPI_ISL_637346 to 637347, EPI_ISL_637349, EPI_ISL_637351, EPI_ISL_637378 to 637380, EPI_ISL_637385, EPI_ISL_637395, EPI_ISL_637400, EPI_ISL_637412, EPI_ISL_637422, EPI_ISL_637445, EPI_ISL_637447 to 637450, EPI_ISL_637452, EPI_ISL_637454, EPI_ISL_637458, EPI_ISL_637524 to 637525, EPI_ISL_637537, EPI_ISL_637542 to 637544, EPI_ISL_637546, EPI_ISL_637551, EPI_ISL_637556, EPI_ISL_637586 to 637587, EPI_ISL_637593, EPI_ISL_637596 to 637597, EPI_ISL_637599 to 637601, EPI_ISL_637603 to 637604, EPI_ISL_637606 to 637609, EPI_ISL_637611 to 637612, EPI_ISL_637614 to 637616, EPI_ISL_637618 to 637619, EPI_ISL_637622, EPI_ISL_637625, EPI_ISL_637627, EPI_ISL_637667, EPI_ISL_637675, EPI_ISL_637854, EPI_ISL_637868 to 637870, EPI_ISL_637882, EPI_ISL_637884, EPI_ISL_637886, EPI_ISL_637894, EPI_ISL_637896, EPI_ISL_637899, EPI_ISL_637907, EPI_ISL_637909, EPI_ISL_637915, EPI_ISL_637918, EPI_ISL_637924, EPI_ISL_637928, EPI_ISL_637952, EPI_ISL_637956 to 637958, EPI_ISL_637962, EPI_ISL_637964, EPI_ISL_637997 to 637998, EPI_ISL_638007, EPI_ISL_638012, EPI_ISL_638028, EPI_ISL_638034 to 638035, EPI_ISL_638041 to 638049, EPI_ISL_638051 to 638064, EPI_ISL_638084 to 638085, EPI_ISL_638091 to 638130, EPI_ISL_638147 to 638163, EPI_ISL_638172, EPI_ISL_638206, EPI_ISL_638682 to 638688, EPI_ISL_638690 to 638695, EPI_ISL_638697 to 638710, EPI_ISL_638712 to 638725, EPI_ISL_638727, EPI_ISL_638729 to 638730, EPI_ISL_638928 to 638935, EPI_ISL_639011 to 639016, EPI_ISL_639018 to 639021, EPI_ISL_639023 to 639057, EPI_ISL_639059 to 639105, EPI_ISL_639107 to 639180, EPI_ISL_639182, EPI_ISL_639184 to 639199, EPI_ISL_639201 to 639227, EPI_ISL_639229 to 639292, EPI_ISL_639294 to 639316, EPI_ISL_639318 to 639322, EPI_ISL_639324 to 639394, EPI_ISL_639396, EPI_ISL_639398 to 639421, EPI_ISL_639423, EPI_ISL_639425 to 639475, EPI_ISL_639477 to 639493, EPI_ISL_639495 to 639498, EPI_ISL_639500 to 639547, EPI_ISL_639549 to 639565, EPI_ISL_639567 to 639606, EPI_ISL_639608 to 639616, EPI_ISL_639618 to 639623, EPI_ISL_639626, EPI_ISL_650204, EPI_ISL_650206, EPI_ISL_650320, EPI_ISL_650338 to 650339, EPI_ISL_650431, EPI_ISL_650549, EPI_ISL_650551, EPI_ISL_650660, EPI_ISL_650695, EPI_ISL_650787, EPI_ISL_650884, EPI_ISL_650896, EPI_ISL_650905 to 650906, EPI_ISL_650914, EPI_ISL_650919, EPI_ISL_650929, EPI_ISL_651108, EPI_ISL_651112, EPI_ISL_651154, EPI_ISL_651191, EPI_ISL_651207, EPI_ISL_651264, EPI_ISL_651415 to 651416, EPI_ISL_651420, EPI_ISL_651440 to 651465, EPI_ISL_651502 to 651503, EPI_ISL_651543, EPI_ISL_651546, EPI_ISL_652414 to 652424, EPI_ISL_652426 to 652439, EPI_ISL_652441 to 652558, EPI_ISL_652560 to 652561, EPI_ISL_652563 to 652573, EPI_ISL_652575 to 652592, EPI_ISL_652594 to 652599, EPI_ISL_652601 to 652619, EPI_ISL_665281, EPI_ISL_665284, EPI_ISL_665288 to 665289, EPI_ISL_665302, EPI_ISL_665309, EPI_ISL_665315, EPI_ISL_665331, EPI_ISL_665361, EPI_ISL_665373, EPI_ISL_665386, EPI_ISL_665442, EPI_ISL_665455, EPI_ISL_665462, EPI_ISL_665464, EPI_ISL_665468 to 665469, EPI_ISL_665473 to 665474, EPI_ISL_665504, EPI_ISL_665511, EPI_ISL_665526 to 665527, EPI_ISL_665539, EPI_ISL_665546, EPI_ISL_665556 to 665558, EPI_ISL_665569, EPI_ISL_665573, EPI_ISL_665575 to 665576, EPI_ISL_665582, EPI_ISL_665584 to 665586, EPI_ISL_665588 to 665589, EPI_ISL_665592, EPI_ISL_665595, EPI_ISL_665598, EPI_ISL_665602, EPI_ISL_665604 to 665608, EPI_ISL_665613, EPI_ISL_665619, EPI_ISL_665642, EPI_ISL_665647, EPI_ISL_665653 to 665655, EPI_ISL_665658 to 665660, EPI_ISL_665670, EPI_ISL_665677, EPI_ISL_665691, EPI_ISL_665713, EPI_ISL_665722, EPI_ISL_665728, EPI_ISL_665743, EPI_ISL_665750 to 665756, EPI_ISL_665763 to 665783, EPI_ISL_665793 to 665800, EPI_ISL_665802 to 665803, EPI_ISL_666110 to 666127, EPI_ISL_666129, EPI_ISL_666131 to 666144, EPI_ISL_666146, EPI_ISL_666148 to 666156, EPI_ISL_666158 to 666298, EPI_ISL_666300 to 666310, EPI_ISL_666312 to 666313, EPI_ISL_666315 to 666325, EPI_ISL_666327 to 666342, EPI_ISL_666344 to 666371, EPI_ISL_666373 to 666433, EPI_ISL_666435 to 666473, EPI_ISL_666475, EPI_ISL_666477 to 666495, EPI_ISL_666497 to 666505, EPI_ISL_666507 to 666529, EPI_ISL_666531 to 666567, EPI_ISL_666569 to 666586, EPI_ISL_666588 to 666590, EPI_ISL_680580 to 680593, EPI_ISL_680595 to 680720, EPI_ISL_680722 to 680752, EPI_ISL_680754 to 680758, EPI_ISL_680760 to 680763, EPI_ISL_680765 to 680776, EPI_ISL_680778 to 680786, EPI_ISL_680788 to 680795, EPI_ISL_680797 to 680802, EPI_ISL_680804 to 680815, EPI_ISL_680817 to 680820, EPI_ISL_680822 to 680858, EPI_ISL_680860 to 680873, EPI_ISL_680875 to 680933, EPI_ISL_680935 to 681008, EPI_ISL_681010 to 681079, EPI_ISL_681081 to 681118, EPI_ISL_681120 to 681182, EPI_ISL_681184 to 681259, EPI_ISL_702692, EPI_ISL_702742, EPI_ISL_702817, EPI_ISL_702857, EPI_ISL_702859, EPI_ISL_702863 to 702864, EPI_ISL_702890, EPI_ISL_702895 to 702896, EPI_ISL_702905, EPI_ISL_703066, EPI_ISL_703074, EPI_ISL_703129, EPI_ISL_703139, EPI_ISL_703203, EPI_ISL_703227, EPI_ISL_703251, EPI_ISL_703259, EPI_ISL_703278, EPI_ISL_703299, EPI_ISL_703302, EPI_ISL_703321, EPI_ISL_703330, EPI_ISL_703343, EPI_ISL_703346, EPI_ISL_703351, EPI_ISL_703359, EPI_ISL_703392, EPI_ISL_703395, EPI_ISL_703398, EPI_ISL_703443, EPI_ISL_703609, EPI_ISL_703614, EPI_ISL_703627, EPI_ISL_703642, EPI_ISL_703721 to 703722, EPI_ISL_703746, EPI_ISL_703760 to 703761, EPI_ISL_703768, EPI_ISL_703855, EPI_ISL_703858, EPI_ISL_703860, EPI_ISL_703868, EPI_ISL_703870, EPI_ISL_703873, EPI_ISL_703899, EPI_ISL_703902, EPI_ISL_703908, EPI_ISL_703918, EPI_ISL_703940, EPI_ISL_703942, EPI_ISL_703955, EPI_ISL_703985, EPI_ISL_703987, EPI_ISL_704001, EPI_ISL_704014, EPI_ISL_704017, EPI_ISL_704030, EPI_ISL_704038, EPI_ISL_704044, EPI_ISL_704081, EPI_ISL_704095, EPI_ISL_704111 to 704112, EPI_ISL_704126, EPI_ISL_704161, EPI_ISL_704171, EPI_ISL_704317, EPI_ISL_704320, EPI_ISL_704584, EPI_ISL_704918, EPI_ISL_704931, EPI_ISL_704981, EPI_ISL_705015, EPI_ISL_705070, EPI_ISL_705166, EPI_ISL_705229, EPI_ISL_705239, EPI_ISL_705244, EPI_ISL_705265, EPI_ISL_705306, EPI_ISL_705327 to 705328, EPI_ISL_705366, EPI_ISL_705384, EPI_ISL_705390, EPI_ISL_705412 to 705413, EPI_ISL_705418 to 705420, EPI_ISL_705433 to 705439, EPI_ISL_705441 to 705442, EPI_ISL_705459 to 705463, EPI_ISL_705488, EPI_ISL_705527, EPI_ISL_706438 to 706461, EPI_ISL_706463 to 706519, EPI_ISL_706521 to 706630, EPI_ISL_706632 to 706780, EPI_ISL_706782 to 706813, EPI_ISL_706815 to 706863, EPI_ISL_725670 to 725697, EPI_ISL_725699 to 725745, EPI_ISL_725747 to 725792, EPI_ISL_725794 to 725834, EPI_ISL_725836 to 725838, EPI_ISL_725840 to 725857, EPI_ISL_725859 to 725870, EPI_ISL_725872 to 725908, EPI_ISL_725910 to 725927, EPI_ISL_725929 to 726007, EPI_ISL_726009 to 726056, EPI_ISL_726058 to 726059, EPI_ISL_726061 to 726092, EPI_ISL_726094 to 726143, EPI_ISL_726145 to 726161, EPI_ISL_726163 to 726241, EPI_ISL_726243 to 726251, EPI_ISL_726253, EPI_ISL_726255 to 726339, EPI_ISL_726341 to 726367, EPI_ISL_726369 to 726379, EPI_ISL_726381 to 726410, EPI_ISL_726412 to 726454, EPI_ISL_726456 to 726492, EPI_ISL_726494 to 726510, EPI_ISL_726512 to 726513, EPI_ISL_726516, EPI_ISL_726518 to 726520, EPI_ISL_726522 to 726551, EPI_ISL_726553 to 726575, EPI_ISL_726577 to 726667, EPI_ISL_726669 to 726686, EPI_ISL_726688 to 726700, EPI_ISL_726702 to 726721, EPI_ISL_726723 to 726826, EPI_ISL_726828 to 726862, EPI_ISL_726864 to 726941, EPI_ISL_726943 to 726948, EPI_ISL_726951 to 726993, EPI_ISL_726995 to 727033, EPI_ISL_727035 to 727101, EPI_ISL_727103 to 727121, EPI_ISL_727123 to 727194, EPI_ISL_727196 to 727223, EPI_ISL_727225 to 727294, EPI_ISL_727296 to 727399, EPI_ISL_727401 to 727408, EPI_ISL_727411 to 727434, EPI_ISL_727436 to 727442, EPI_ISL_727444 to 727448, EPI_ISL_727450 to 727492, EPI_ISL_727494 to 727502, EPI_ISL_727504 to 727547, EPI_ISL_727549 to 727631, EPI_ISL_727633 to 727665, EPI_ISL_727667 to 727696, EPI_ISL_741347, EPI_ISL_742262 to 742334, EPI_ISL_742336 to 742365, EPI_ISL_742367 to 742513, EPI_ISL_742515 to 742562, EPI_ISL_742564 to 742613, EPI_ISL_742615, EPI_ISL_742617 to 742620, EPI_ISL_742622 to 742632, EPI_ISL_742634 to 742668, EPI_ISL_742670 to 742795, EPI_ISL_742797 to 742872, EPI_ISL_742874 to 742891, EPI_ISL_742893 to 742896, EPI_ISL_742898 to 742927, EPI_ISL_742929 to 743037, EPI_ISL_743039 to 743116, EPI_ISL_743118 to 743183, EPI_ISL_743185 to 743249, EPI_ISL_743251 to 743326, EPI_ISL_743328 to 743418, EPI_ISL_743420 to 743464, EPI_ISL_743466 to 743504, EPI_ISL_743506 to 743516, EPI_ISL_743518 to 743588, EPI_ISL_743590 to 743611, EPI_ISL_743613 to 743614, EPI_ISL_743616, EPI_ISL_743618 to 743635, EPI_ISL_743637 to 743652, EPI_ISL_743654 to 743741, EPI_ISL_743743 to 743808, EPI_ISL_743810 to 743815, EPI_ISL_743817 to 743934, EPI_ISL_743936, EPI_ISL_743938 to 743942, EPI_ISL_743944 to 743945, EPI_ISL_743948, EPI_ISL_743951 to 743964, EPI_ISL_743966 to 744023, EPI_ISL_744025 to 744081, EPI_ISL_744083 to 744134, EPI_ISL_763371, EPI_ISL_763378, EPI_ISL_763392, EPI_ISL_763409 to 763411, EPI_ISL_763413, EPI_ISL_763416, EPI_ISL_763418, EPI_ISL_763420, EPI_ISL_763422 to 763423, EPI_ISL_763443, EPI_ISL_763445, EPI_ISL_763454, EPI_ISL_763458, EPI_ISL_763462, EPI_ISL_763470, EPI_ISL_763472 to 763474, EPI_ISL_763480 to 763481, EPI_ISL_763484, EPI_ISL_763487 to 763489, EPI_ISL_763492 to 763494, EPI_ISL_763497, EPI_ISL_763509, EPI_ISL_763519, EPI_ISL_763522, EPI_ISL_763525, EPI_ISL_763539, EPI_ISL_763546, EPI_ISL_763557, EPI_ISL_763565 to 763566, EPI_ISL_763568, EPI_ISL_763570 to 763572, EPI_ISL_763574 to 763577, EPI_ISL_763579, EPI_ISL_763584 to 763585, EPI_ISL_763589 to 763591, EPI_ISL_763594, EPI_ISL_763597, EPI_ISL_763599, EPI_ISL_763633, EPI_ISL_763657 to 763658, EPI_ISL_763664, EPI_ISL_763668, EPI_ISL_763671, EPI_ISL_763674, EPI_ISL_763679 to 763680, EPI_ISL_763686, EPI_ISL_763692, EPI_ISL_763718, EPI_ISL_763721 to 763722, EPI_ISL_763724 to 763725, EPI_ISL_763733, EPI_ISL_763735, EPI_ISL_763738 to 763739, EPI_ISL_763748, EPI_ISL_763750 to 763753, EPI_ISL_763758 to 763762, EPI_ISL_763764 to 763766, EPI_ISL_763770 to 763771, EPI_ISL_763773 to 763776, EPI_ISL_763778 to 763779, EPI_ISL_763782 to 763783, EPI_ISL_763786 to 763787, EPI_ISL_763814, EPI_ISL_763820 to 763822, EPI_ISL_763864, EPI_ISL_763975 to 764001, EPI_ISL_764003 to 764005, EPI_ISL_764034 to 764035, EPI_ISL_764063 to 764065, EPI_ISL_764080, EPI_ISL_764133, EPI_ISL_764138, EPI_ISL_764142, EPI_ISL_764150, EPI_ISL_764155 to 764157, EPI_ISL_764161, EPI_ISL_764168, EPI_ISL_764180, EPI_ISL_764186, EPI_ISL_764192, EPI_ISL_764199 to 764201, EPI_ISL_764222, EPI_ISL_764229, EPI_ISL_764232, EPI_ISL_764234 to 764235, EPI_ISL_764244 to 764249, EPI_ISL_764251 to 764260, EPI_ISL_764266 to 764268, EPI_ISL_764273 to 764275, EPI_ISL_764277, EPI_ISL_764282 to 764283, EPI_ISL_764573 to 764611, EPI_ISL_764613 to 764637, EPI_ISL_764639 to 764689, EPI_ISL_764691 to 764692, EPI_ISL_764694 to 764723, EPI_ISL_764725 to 764749, EPI_ISL_764751 to 764832, EPI_ISL_764834 to 764837, EPI_ISL_764839 to 764841, EPI_ISL_764843 to 764845, EPI_ISL_764847 to 764851, EPI_ISL_764853 to 764870, EPI_ISL_764872 to 764884, EPI_ISL_764886 to 764890, EPI_ISL_764892 to 764938, EPI_ISL_764940 to 764974, EPI_ISL_764976, EPI_ISL_764978 to 765003, EPI_ISL_765005 to 765017, EPI_ISL_765019 to 765023, EPI_ISL_765025 to 765081, EPI_ISL_765083 to 765097, EPI_ISL_765099 to 765102, EPI_ISL_765104, EPI_ISL_765106 to 765109, EPI_ISL_765111, EPI_ISL_814274 to 814284, EPI_ISL_814291, EPI_ISL_814294 to 814311, EPI_ISL_814313 to 814317, EPI_ISL_814319 to 814325, EPI_ISL_814327 to 814332, EPI_ISL_814334, EPI_ISL_814336 to 814338, EPI_ISL_814340 to 814343, EPI_ISL_814346 to 814347, EPI_ISL_814349 to 814350, EPI_ISL_814352 to 814354, EPI_ISL_814356, EPI_ISL_814362 to 814364, EPI_ISL_814366 to 814394, EPI_ISL_814396 to 814413, EPI_ISL_814415 to 814416, EPI_ISL_814422 to 814423, EPI_ISL_814468 to 814488, EPI_ISL_814491 to 814503, EPI_ISL_814505 to 814516, EPI_ISL_814523, EPI_ISL_814526 to 814529, EPI_ISL_814531, EPI_ISL_814536 to 814537, EPI_ISL_814540 to 814541, EPI_ISL_814545, EPI_ISL_814550

| | | | |
|---|---|---|---|
| to 814551, EPI_ISL_814553 to 814556, EPI_ISL_814558 to 814559, EPI_ISL_814561 to 814567, EPI_ISL_814573 to 814589, EPI_ISL_814595 to 814603, EPI_ISL_814606 to 814607, EPI_ISL_814695 to 814807, EPI_ISL_814809 to 814823, EPI_ISL_814825 to 814969, EPI_ISL_814971 to 815032, EPI_ISL_815034 to 815042, EPI_ISL_815044 to 815111, EPI_ISL_815113 to 815150, EPI_ISL_815152 to 815206, EPI_ISL_815209 to 815254, EPI_ISL_822345 to 822368, EPI_ISL_822370 to 822388, EPI_ISL_822390 to 822447, EPI_ISL_822449 to 822525, EPI_ISL_822527 to 822536, EPI_ISL_822538 to 822635, EPI_ISL_822637 to 822658, EPI_ISL_822660 to 822675, EPI_ISL_822677 to 822714, EPI_ISL_822716 to 822762, EPI_ISL_822764 to 822805, EPI_ISL_822838 to 822843, EPI_ISL_822845 to 822858, EPI_ISL_822860 to 822881, EPI_ISL_822883 to 822892, EPI_ISL_822894 to 822921, EPI_ISL_822923 to 822937, EPI_ISL_822939 to 822968, EPI_ISL_822970 to 822997, EPI_ISL_823000 to 823102, EPI_ISL_823104 to 823112, EPI_ISL_823114 to 823234, EPI_ISL_823236 to 823284, EPI_ISL_823286 to 823318, EPI_ISL_823320 to 823324, EPI_ISL_823326 to 823397, EPI_ISL_823399 to 823404, EPI_ISL_823406 to 823466, EPI_ISL_823468 to 823502, EPI_ISL_823504 to 823629, EPI_ISL_823631 to 823656, EPI_ISL_823658 to 823692, EPI_ISL_823694 to 823734, EPI_ISL_823736 to 823762, EPI_ISL_823764 to 823806, EPI_ISL_823808 to 823812 | | | |
| see above | Wales Specialist Virology Centre Sequencing lab: Pathogen Genomics Unit | COVID-19 Genomics UK (COG-UK) Consortium | Alec Birchley; Alexander Adams; Amy Gaskin; Angela Marchbank; Bree Gatica-Wilcox; Catherine Moore; Jason Coombes; Joanne Watkins; Joel Southgate; Johnathan Evans; Laura Gifford; Lauren Gilbert; Lee Graham; Malorie Perry; Matthew Bull; Nicole Pacchiarini; Sally Corden; Sara Kumziene-Summerhayes; Sara Rey; Sarah Taylor; Simon Cottrell; Sophie Jones; Tom Connor |
| EPI_ISL_840835 to 840837, EPI_ISL_840844, EPI_ISL_840885 to 840888, EPI_ISL_840890 to 840891, EPI_ISL_840893 to 840895, EPI_ISL_840897 to 840900, EPI_ISL_841057 to 841058, EPI_ISL_841065 to 841066, EPI_ISL_841083, EPI_ISL_841087 to 841097, EPI_ISL_841099 to 841107, EPI_ISL_841109 to 841113, EPI_ISL_841302 to 841305 | | | |
| see above | Wales Specialist Virology Centre Sequencing lab: Pathogen Genomics Unit | Public Health Wales Microbiology Cardiff Wales Specialist Virology Centre | Alec Birchley; Alexander Adams; Amy Gaskin; Angela Marchbank; Bree Gatica-Wilcox; Catherine Moore; Jason Coombes; Joanne Watkins; Joel Southgate; Johnathan Evans; Laura Gifford; Lauren Gilbert; Lee Graham; Malorie Perry; Matthew Bull; Nicole Pacchiarini; Sally Corden; Sara Kumziene-Summerhayes; Sara Rey; Sarah Taylor; Simon Cottrell; Sophie Jones; Tom Connor |
| EPI_ISL_445221, EPI_ISL_445234 to 445235, EPI_ISL_469071, EPI_ISL_476137 | Wasterlakarna | The Public Health Agency of Sweden | Anna Risberg; Anna-Malin Linde; Frida Ahlfors; Karin Tegmark-Wisell; Maria Lind Karlberg; Mattias Haukland; Mia Brytting; Olov Svartstrom; Oskar Karlsson Lindsjo; Petra Edquist; Reza Advani; Sandra Broddesson; Shamam Muradrasoli; Theresa Enkirch |
| EPI_ISL_452242 | Wernstedt Medical AB | The Public Health Agency of Sweden | Anna Risberg; Anna-Malin Linde; Eva Sandberg; Karin Tegmark-Wisell; Maria Lind Karlberg; Mia Brytting; Olov Svartstrom; Oskar Karlsson Lindsjo; Theresa Enkirch |
| EPI_ISL_413221, EPI_ISL_414024 to 414027, EPI_ISL_415628, EPI_ISL_415630 to 415631, EPI_ISL_629000 | West of Scotland Specialist Virology Centre, NHSGGC | MRC-University of Glasgow Centre for Virus Research | Alasdair MacLean; Ana da Silva Filipe; Andrew Rambaut; Antonia Ho; Daniel Mair; David L Robertson; Emma Thomson; Emma Thomson.; James Shephard; James Shepherd; Jenna Nichols; Joseph Hughes; Josh Singer; Kathy Li; Kathy Smollett; Lily Tong; Natasha Jesudason; Natasha Johnson; Rajiv Shah; Richard Orton; Rory Gunson; Rory Gunson.; Sarah McDonald; Shirin Ashraf; Sreenu Vattipally; Stephen Carmichael |
| EPI_ISL_425647 to 425650, EPI_ISL_425652 to 425671, EPI_ISL_425673 to 425680, EPI_ISL_425682 to 425688, EPI_ISL_425690 to 425705, EPI_ISL_425707 to 425783, EPI_ISL_425785 to 425818, EPI_ISL_433268 to 433270, EPI_ISL_433272 to 433314, EPI_ISL_433316 to 433336, EPI_ISL_433338 to 433362, EPI_ISL_433364 to 433373, EPI_ISL_433498 to 433505, EPI_ISL_433507 to 433615, EPI_ISL_433618 to 433622, EPI_ISL_433624 to 433665, EPI_ISL_438749 to 438757, EPI_ISL_438759 to 438802, EPI_ISL_438804, EPI_ISL_438806 to 438807, EPI_ISL_438811 to 438819, EPI_ISL_438821 to 438858, EPI_ISL_438860 to 438864, EPI_ISL_438866 to 438913, EPI_ISL_438915 to 438921, EPI_ISL_438923 to 438926, EPI_ISL_438928 to 438935, EPI_ISL_438937 to 438941, EPI_ISL_438943 to 438946, EPI_ISL_438973 to 438995, EPI_ISL_438997 to 439014, EPI_ISL_439016, EPI_ISL_439018 to 439029, EPI_ISL_439031 to 439073, EPI_ISL_439075 to 439143, EPI_ISL_448117 to 448134, EPI_ISL_448136 to 448173, EPI_ISL_448175 to 448207, EPI_ISL_448209 to 448221, EPI_ISL_449176 to 449183, EPI_ISL_449185 to 449217, EPI_ISL_449219 to 449223, EPI_ISL_449225 to 449258, EPI_ISL_453006 to 453008, EPI_ISL_453010 to 453015, EPI_ISL_453017 to 453021, EPI_ISL_453024 to 453027, EPI_ISL_453029 to 453049, EPI_ISL_453051 to 453057, EPI_ISL_453059 to 453070, EPI_ISL_453072, EPI_ISL_453074 to 453091, EPI_ISL_453096 to 453098, EPI_ISL_456756 to 456815, EPI_ISL_456817 to 456889, EPI_ISL_461589 to 461609, EPI_ISL_461611 to 461639, EPI_ISL_461641 to 461705, EPI_ISL_473508 to 473536, EPI_ISL_473538 to 473563, EPI_ISL_473565 to 473590, EPI_ISL_473592 to 473605, EPI_ISL_473607 to 473628, EPI_ISL_473630 to 473643, EPI_ISL_473645 to 473663, EPI_ISL_473665 to 473666, EPI_ISL_473668 to 473748, EPI_ISL_473750 to 473782, EPI_ISL_477820 to 477892, EPI_ISL_477894 to 477916, EPI_ISL_477918 to 478052, EPI_ISL_478055 to 478099, EPI_ISL_478102 to 478123, EPI_ISL_478125 to 478162, EPI_ISL_484519 to 484535, EPI_ISL_484537 to 484549, EPI_ISL_484551 to 484552, EPI_ISL_484555 to 484574, EPI_ISL_484576 to 484601, EPI_ISL_484603 to 484607, EPI_ISL_484609 to 484612, EPI_ISL_484614 to 484627, EPI_ISL_484629 to 484679, EPI_ISL_490330, EPI_ISL_490646 to 490682, EPI_ISL_490684 to 490709, EPI_ISL_493741 to 493742, EPI_ISL_493745 to 493754, EPI_ISL_493756 to 493764, EPI_ISL_493766 to 493783, EPI_ISL_493785 to 493800, EPI_ISL_493802, EPI_ISL_493804 to 493808, EPI_ISL_493810 to 493814, EPI_ISL_493816 to 493820, EPI_ISL_493823, EPI_ISL_493831, EPI_ISL_493891 to 493892, EPI_ISL_493895 to 493897, EPI_ISL_499352 to 499354, EPI_ISL_512436 to 512480, EPI_ISL_514514 to 514515, EPI_ISL_517580 to 517582, EPI_ISL_526451 to 526453, EPI_ISL_526456 to 526458, EPI_ISL_529315 to 529316, EPI_ISL_529319, EPI_ISL_529409, EPI_ISL_529684 to 529688, EPI_ISL_535102 to 535109, EPI_ISL_535111 to 535182, EPI_ISL_540775 to 540811, EPI_ISL_540813 to 540815, EPI_ISL_540817 to 540822, EPI_ISL_572415, EPI_ISL_572470, EPI_ISL_572518, EPI_ISL_572523, EPI_ISL_572528, EPI_ISL_572534, EPI_ISL_572585, EPI_ISL_572620 to 572641, EPI_ISL_572686, EPI_ISL_572741, EPI_ISL_572746, EPI_ISL_572763, EPI_ISL_572788, EPI_ISL_572832, EPI_ISL_573765 to 573768, EPI_ISL_573770 to 573785, EPI_ISL_573787 to 573793, EPI_ISL_585261 to 585265, EPI_ISL_585267 to 585269, EPI_ISL_585271 to 585296, EPI_ISL_585422 to 585425, EPI_ISL_594645 to 594686, EPI_ISL_594688 to 594689, EPI_ISL_594691 to 594751, EPI_ISL_594753 to 594809, EPI_ISL_594811 to 594815, EPI_ISL_611533, EPI_ISL_611547 to 611548, EPI_ISL_611610 to 611611, EPI_ISL_611614, EPI_ISL_611660, EPI_ISL_611693, EPI_ISL_611708 to 611710, EPI_ISL_611712 to 611720, EPI_ISL_611818, EPI_ISL_611820, EPI_ISL_611826, EPI_ISL_611848, EPI_ISL_611852, EPI_ISL_611866 to 611867, EPI_ISL_611870 to 611884, EPI_ISL_611890, EPI_ISL_611990, EPI_ISL_612063, EPI_ISL_612113, EPI_ISL_612124 to 612125, EPI_ISL_612284 to 612313, EPI_ISL_626687, EPI_ISL_626691 to 626692, EPI_ISL_626749, EPI_ISL_626758, EPI_ISL_626812, EPI_ISL_626818, EPI_ISL_626822, EPI_ISL_626825 to 626826, EPI_ISL_626831, EPI_ISL_626835, EPI_ISL_626837 to 626849, EPI_ISL_626851, EPI_ISL_626855, EPI_ISL_626897, EPI_ISL_626904, EPI_ISL_626917, EPI_ISL_626927, EPI_ISL_626933 to 626935, EPI_ISL_626952, EPI_ISL_626957 to 626958, EPI_ISL_626962 to 626963, EPI_ISL_626966 to 626992, EPI_ISL_626995, EPI_ISL_627014, EPI_ISL_627030, EPI_ISL_627040, EPI_ISL_627045, EPI_ISL_627049, EPI_ISL_627063, EPI_ISL_627165, EPI_ISL_627173, EPI_ISL_627195, EPI_ISL_627200 to 627202, EPI_ISL_627208 to 627209, EPI_ISL_627329 to 627402, EPI_ISL_627404 to 627420, EPI_ISL_627422 to 627423, EPI_ISL_650146, EPI_ISL_650164, EPI_ISL_650175, EPI_ISL_650292 to 650293, EPI_ISL_650297 to 650298, EPI_ISL_650310, EPI_ISL_650312 to 650313, EPI_ISL_650316, EPI_ISL_650328, EPI_ISL_650375, EPI_ISL_650412, EPI_ISL_650424, EPI_ISL_650438, EPI_ISL_650451, EPI_ISL_650477, EPI_ISL_650482 to 650484, EPI_ISL_650490 to 650492, EPI_ISL_650494, EPI_ISL_650497 to 650498, EPI_ISL_650503 to 650511, EPI_ISL_650514, EPI_ISL_650517 to 650521, EPI_ISL_650523, EPI_ISL_650525 to 650526, EPI_ISL_650538, EPI_ISL_650550, EPI_ISL_650649, EPI_ISL_650709, EPI_ISL_650712, EPI_ISL_650725 to 650728, EPI_ISL_650730 to 650732, EPI_ISL_650734 to 650736, EPI_ISL_650739, EPI_ISL_650745 to 650754, EPI_ISL_650759, EPI_ISL_650762 to 650764, EPI_ISL_650789, EPI_ISL_650802, EPI_ISL_650804 to 650806, EPI_ISL_650816, EPI_ISL_650837 to 650843, EPI_ISL_650845, EPI_ISL_650848, EPI_ISL_650850 to 650876, EPI_ISL_650925, EPI_ISL_650927, EPI_ISL_650963, EPI_ISL_651011, EPI_ISL_651125, EPI_ISL_651160, EPI_ISL_651310, EPI_ISL_651315, EPI_ISL_651330, EPI_ISL_651359, EPI_ISL_651377, EPI_ISL_651380, EPI_ISL_651402, EPI_ISL_651406, EPI_ISL_651493 to 651498, EPI_ISL_651531 to 651540, EPI_ISL_651583, EPI_ISL_651733 to 651789, EPI_ISL_651811 to 651848, EPI_ISL_651850 to 651885, EPI_ISL_651887 to 651956, EPI_ISL_665283, EPI_ISL_665286, EPI_ISL_665321, EPI_ISL_665363, EPI_ISL_665385, EPI_ISL_665416, EPI_ISL_665425, EPI_ISL_665436, EPI_ISL_665439, EPI_ISL_665521 to 665524, EPI_ISL_665529, EPI_ISL_665531 to 665532, EPI_ISL_665535 to 665538, EPI_ISL_665541, EPI_ISL_665564 to 665566, EPI_ISL_665578 to 665579, EPI_ISL_665609, EPI_ISL_665633, EPI_ISL_665640 to 665641, EPI_ISL_665685, EPI_ISL_665727, EPI_ISL_665748, EPI_ISL_665792, EPI_ISL_665804, EPI_ISL_666023 to 666059, EPI_ISL_680442 to 680460, EPI_ISL_702618, EPI_ISL_702659, EPI_ISL_702771, EPI_ISL_702867, EPI_ISL_702892 to 702893, EPI_ISL_703136, EPI_ISL_703141, EPI_ISL_703155, EPI_ISL_703190, EPI_ISL_703192, EPI_ISL_703233, EPI_ISL_703240, EPI_ISL_703243, EPI_ISL_703246, EPI_ISL_703249, EPI_ISL_703269, EPI_ISL_703287, EPI_ISL_703430, EPI_ISL_703717 to 703719, EPI_ISL_703737, EPI_ISL_703743, EPI_ISL_703748 to 703758, EPI_ISL_703771, EPI_ISL_703773, EPI_ISL_703829, EPI_ISL_703831, EPI_ISL_703834, EPI_ISL_703837, EPI_ISL_703839, EPI_ISL_703842, EPI_ISL_703845, EPI_ISL_703911, EPI_ISL_703921, EPI_ISL_703924, EPI_ISL_703927, EPI_ISL_703929, EPI_ISL_703968, EPI_ISL_704089, EPI_ISL_704118, EPI_ISL_704184, EPI_ISL_704206, EPI_ISL_704228, EPI_ISL_704595 to 704597, EPI_ISL_704804, EPI_ISL_704807, EPI_ISL_704809, EPI_ISL_704952, EPI_ISL_705124, EPI_ISL_705253, EPI_ISL_705296, EPI_ISL_705300, EPI_ISL_705316, EPI_ISL_705399, EPI_ISL_705490, EPI_ISL_705636 to 705718, EPI_ISL_705720 to 705733, EPI_ISL_724210 to 724217, EPI_ISL_724219 to 724221, EPI_ISL_814312, EPI_ISL_814335, EPI_ISL_814339, EPI_ISL_814344 to 814345, EPI_ISL_814348, EPI_ISL_814355, EPI_ISL_814439 to 814459, EPI_ISL_814547, EPI_ISL_814549, EPI_ISL_814568, EPI_ISL_814590 to 814593, EPI_ISL_814608 to 814609, EPI_ISL_814620 to 814664, EPI_ISL_838055 to 838163, EPI_ISL_838165 to 838177, EPI_ISL_838179 to 838213, EPI_ISL_864960 to 864980, EPI_ISL_864982 to 864983, EPI_ISL_864992 to 864993 | | | |
| see above | West of Scotland Specialist Virology Centre, NHSGGC / MRC-University of Glasgow Centre for Virus Research | COVID-19 Genomics UK (COG-UK) Consortium | Alasdair MacLean; Alice Broos; Ana da Silva Filipe; Antonia Ho; Daniel Mair; David L Robertson; Elihu Aranday-Cortes; Emma Thomson; Guy Mollett; Igor Starinskij; James Shepherd; Jenna Nichols; Joseph Hughes; Kathy Li; Kathy Smollett; Kirstyn Brunker; Kyriaki Nomikou; Lily Tong; Marc Niebel; Matthew Holden; Natasha Jesudason; Natasha Johnson; Patawee Asamaphan; Rachel Blacow; Rajiv Shah; Richard Orton; Rory Gunson; Rory Gunson.; Sarah McDonald; Sharif Shaaban; Sreenu Vattipally; Stephen Carmichael; Yasmin Parr |
| EPI_ISL_450831, EPI_ISL_450833, EPI_ISL_455105 | Wetterhalsan | The Public Health Agency of Sweden | Anders Tengblad; Anna Risberg; Anna-Malin Linde; Karin Tegmark-Wisell; Maria Lind Karlberg; Mia Brytting; Olov Svartstrom; Oskar Karlsson Lindsjo; Theresa Enkirch |
| EPI_ISL_455332, EPI_ISL_539565 to 539566 | Xerencia de Xestión Integrada de Pontevedra e o Salnés | Instituto de Salud Carlos III | A. Monzón; F. Casas; I; I. Jiménez; I. Pérez; Iglesias-Caballero; J. López; M. Camarero; M. Cuesta; M. Garcia; M. González-Esguevillas; M. Molinero Calamita; M. Zaballos; P. Jiménez; S. Juliá; S. Pozo; S. Varona |
| EPI_ISL_766588 | Xundheitszentrum Wengen | Institute of Medical Virology, University of Zurich | Alexandra Trkola; Annette Audigé; Cyril Shah; Jon Huder; Jürg Böni; Kevin Steiner; Maria Grünberg; Maryam Zaheri; Michael Huber; Riccarda Capaul; Stefan Schmutz; Verena Kufner |
| EPI_ISL_539577, EPI_ISL_539587 to 539589, EPI_ISL_539591 to 539596, EPI_ISL_539599 to 539615, EPI_ISL_631304 to 631320, EPI_ISL_631344 to 631346, EPI_ISL_631348 to 631360, EPI_ISL_631362 to 631376, EPI_ISL_631387, EPI_ISL_666623 to 666632, EPI_ISL_666635 to 666652, EPI_ISL_718027 to 718046, EPI_ISL_718048 to 718052, EPI_ISL_718054 to 718059, EPI_ISL_718061 to 718064, EPI_ISL_718067 to 718104, EPI_ISL_733501 to 733506, EPI_ISL_733508 to 733523, EPI_ISL_770688 to 770728, EPI_ISL_770730, EPI_ISL_770732 to 770733, EPI_ISL_770735 to 770756, EPI_ISL_770758 to 770774, EPI_ISL_778844, EPI_ISL_779954, EPI_ISL_819203, EPI_ISL_861509 to 861520, EPI_ISL_861522 to 861527 | | | |
| see above | ZOTZ KLIMAS MVZ Düsseldorf-Centrum GbR ÜBAG für Labormedizin, Genetik, Zytologie, Pathologie | Center of Medical Microbiology, Virology, and Hospital Hygiene, University of Duesseldorf | Alexander Dilthey; Andreas Walker; Ashley-Jane Duplessis; Daniel Strelow; Jessica Nicolai; Jörg Timm; Katrin Hoffmann; Klaus Pfeffer; Lisanna Hülse; Malte Kohns Vasconcelos; Marek Korencak; Maximilian Damagnez; Nadine Lübke; Patrick Finzer; Rainer Zotz; Tobias Wienemann; Torsten Houwaart |
| EPI_ISL_475887 to 475891, EPI_ISL_475893 to 475897, EPI_ISL_475899 to 475906, EPI_ISL_475908 to 475909, EPI_ISL_583599 | Zentralinstitut für medizinische und chemische Labordiagnostik, Universitätskliniken Insbruck | Bergthaler laboratory, CeMM Research Center for Molecular Medicine of the Austrian Academy of Sciences | Adi Steinrigl; Alexander Lercher; Alexandra Popa; Andreas Bergthaler; Benedikt Agerer; Christian Paar; Christoph Bock; Daniela Schmid; Dorothee von Laer; Elisabeth Puchhammer-Stoeckl; Franz Allerberger; Gernot Walder; Gregor Hörmann; Guenter Weiss; Gunther Vogl; Henrique Colaco; Jakob-Wendelin Genger; Jan Laine; Judith Aberle; Kinga Rigler-Hohenwarter; Lukas Endler; Manfred Nairz; Mark Smyth; Martin Senekowitsch; Michael Schuster; Peter Hufnagl; Peter Obrist; Rainer Gattringer; Sabine Sussitz-Rack; Stephan Aberle; Thomas Penz; Wegene Borena |
| EPI_ISL_468134 to 468158 | [Romania, Bucharest] National Institute for Infectious Diseases "Prof. Dr. Matei Bal" | [Romania, Bucharest] National Institute for Infectious Diseases "Prof. Dr. Matei Bal" | Corina Casangiu; Leontina Banica; Marius Cotic; Marius Surleac; Simona Paraschiv |
| EPI_ISL_417481, EPI_ISL_417535 to 417549, EPI_ISL_417552, EPI_ISL_417590, EPI_ISL_417655 to 417664, EPI_ISL_417666 to 417670, EPI_ISL_417672, EPI_ISL_417676, EPI_ISL_424377 to 424378, EPI_ISL_424533 to 424538, EPI_ISL_424540 to 424542, EPI_ISL_424545, EPI_ISL_424548 to 424551, EPI_ISL_424560 to 424562, EPI_ISL_424567 to 424568, EPI_ISL_424570, EPI_ISL_424608 to 424609, EPI_ISL_826671 to 826676, EPI_ISL_826679 to 826680, EPI_ISL_826682, EPI_ISL_826684 to 826692, EPI_ISL_826694 to 826699, EPI_ISL_826701 to 826702, EPI_ISL_826705 to 826714, EPI_ISL_826716, EPI_ISL_826718 to 826725, EPI_ISL_826732 to 826739, EPI_ISL_826741, EPI_ISL_826743 to 826746, EPI_ISL_826748 to 826751, EPI_ISL_826753 to 826776, EPI_ISL_826778 to 826791, EPI_ISL_826793 to 826795, EPI_ISL_826797, EPI_ISL_826799, EPI_ISL_826803, EPI_ISL_826806, EPI_ISL_826808, EPI_ISL_826831, EPI_ISL_826833, EPI_ISL_826835, EPI_ISL_826837 to 826841, EPI_ISL_826843 to 826853, EPI_ISL_826857, EPI_ISL_826860, EPI_ISL_826863 to 826879, EPI_ISL_826881 to 826883, EPI_ISL_826886 to 826892, EPI_ISL_826895, EPI_ISL_826898 to 826914, EPI_ISL_826917, EPI_ISL_826921 to 826924, EPI_ISL_826929, EPI_ISL_826931 to 826936, EPI_ISL_826938 to 826948, EPI_ISL_826950 to 826954, EPI_ISL_826956 to 826968, EPI_ISL_826975 to 826979, EPI_ISL_826982 to 826989, EPI_ISL_826991 to 826993, EPI_ISL_826995 to 826996, EPI_ISL_826998, EPI_ISL_827000, EPI_ISL_827002 to 827008, EPI_ISL_827010, EPI_ISL_827013, EPI_ISL_827015, EPI_ISL_827018 to 827026, EPI_ISL_827028 to 827030, EPI_ISL_827034, EPI_ISL_827039, EPI_ISL_827046 to 827047, EPI_ISL_827049, EPI_ISL_827052 to 827062, EPI_ISL_827065, EPI_ISL_827068, EPI_ISL_827072 to 827073, EPI_ISL_827078, EPI_ISL_827080, EPI_ISL_827089, EPI_ISL_827091 to 827100, EPI_ISL_827102 to 827109, EPI_ISL_827111 to 827119, EPI_ISL_827127, EPI_ISL_827130 to 827141, EPI_ISL_827143 to 827148, EPI_ISL_827150 to 827162, EPI_ISL_827166 to 827178, EPI_ISL_827183 to 827184, EPI_ISL_827186, EPI_ISL_827189, EPI_ISL_827193 to 827194, EPI_ISL_827196, EPI_ISL_827201, EPI_ISL_827209, EPI_ISL_827211, EPI_ISL_827213 to 827228, EPI_ISL_827231 to 827234, EPI_ISL_827236, EPI_ISL_827239 to 827241, EPI_ISL_827243 to 827249, EPI_ISL_827251, EPI_ISL_827255, EPI_ISL_827259 to 827260, EPI_ISL_827262, EPI_ISL_827264 to 827265, EPI_ISL_827267 to 827268, EPI_ISL_827274 to 827278, EPI_ISL_827280 to 827285, EPI_ISL_827292, EPI_ISL_827294 to 827305, EPI_ISL_827308, EPI_ISL_827311 to 827319, EPI_ISL_827321 to 827323, EPI_ISL_827325 to 827329, EPI_ISL_827332, EPI_ISL_827336, EPI_ISL_827338 to 827342, EPI_ISL_827344, EPI_ISL_827346 to 827353, EPI_ISL_827355 to 827356, EPI_ISL_827364 to 827366, EPI_ISL_827369 to 827370, EPI_ISL_827373, EPI_ISL_827376, EPI_ISL_827378, EPI_ISL_827380 to 827381, EPI_ISL_827384, EPI_ISL_827386 to 827391, EPI_ISL_827393 to 827394, EPI_ISL_827397 to 827412, EPI_ISL_827414 to 827415, EPI_ISL_827417 to 827418, EPI_ISL_827420 to 827426, EPI_ISL_827428 to 827434, EPI_ISL_827436, EPI_ISL_827440, EPI_ISL_827442, EPI_ISL_827444, EPI_ISL_827447 to 827448, EPI_ISL_827450 to 827458, EPI_ISL_827460, EPI_ISL_827462, EPI_ISL_827464 to 827467, EPI_ISL_827470 to 827472, EPI_ISL_827474 to 827475, EPI_ISL_827478 to 827481, EPI_ISL_827483, | | | |

| | | | |
|---|---|---|---|
| EPI_ISL_827486 to 827491, EPI_ISL_827495, EPI_ISL_827497 to 827512, EPI_ISL_827515 to 827516, EPI_ISL_827518 to 827528, EPI_ISL_827530, EPI_ISL_827538, EPI_ISL_827547, EPI_ISL_827552, EPI_ISL_827555 to 827558, EPI_ISL_827560 to 827566, EPI_ISL_827568 to 827574, EPI_ISL_827578, EPI_ISL_827580 to 827581, EPI_ISL_827585, EPI_ISL_827591 to 827606, EPI_ISL_827610 to 827617, EPI_ISL_827619 to 827632, EPI_ISL_827634 to 827643, EPI_ISL_827645, EPI_ISL_827647 to 827659, EPI_ISL_827661, EPI_ISL_827664 to 827666, EPI_ISL_827668 to 827678, EPI_ISL_827680 to 827682, EPI_ISL_827686, EPI_ISL_827691, EPI_ISL_827694, EPI_ISL_827698 to 827712, EPI_ISL_827717, EPI_ISL_827719 to 827721, EPI_ISL_827726 to 827735, EPI_ISL_827738, EPI_ISL_827740 to 827745, EPI_ISL_827747 to 827753, EPI_ISL_827755 to 827756, EPI_ISL_827758 to 827779, EPI_ISL_827787 to 827788, EPI_ISL_827790 to 827799, EPI_ISL_827806 to 827807, EPI_ISL_827809 to 827815, EPI_ISL_827820, EPI_ISL_827823 to 827824, EPI_ISL_827826 to 827827, EPI_ISL_827830, EPI_ISL_827832 to 827839, EPI_ISL_827841 to 827853, EPI_ISL_827855 to 827858, EPI_ISL_827861, EPI_ISL_827873 to 827874, EPI_ISL_827883 to 827894, EPI_ISL_827906, EPI_ISL_827912, EPI_ISL_827914 to 827920, EPI_ISL_827924, EPI_ISL_827926 to 827929, EPI_ISL_827932, EPI_ISL_827934, EPI_ISL_827936 to 827937, EPI_ISL_827941, EPI_ISL_827944 to 827950, EPI_ISL_827952 to 827968, EPI_ISL_827971 to 827981, EPI_ISL_827983 to 827991, EPI_ISL_827993 to 828002, EPI_ISL_828004 to 828007, EPI_ISL_828017 to 828019, EPI_ISL_828026, EPI_ISL_828028, EPI_ISL_828034, EPI_ISL_828039 to 828049, EPI_ISL_828051 to 828052, EPI_ISL_828054 to 828063, EPI_ISL_828065, EPI_ISL_828068 to 828069, EPI_ISL_828071 to 828076, EPI_ISL_828078 to 828086, EPI_ISL_828088, EPI_ISL_828090 to 828098, EPI_ISL_828100 to 828120, EPI_ISL_828122 to 828128, EPI_ISL_828130 to 828136, EPI_ISL_828138 to 828141, EPI_ISL_828145, EPI_ISL_828148 to 828153, EPI_ISL_828155, EPI_ISL_828161, EPI_ISL_828164 to 828165, EPI_ISL_828167 to 828173, EPI_ISL_828176, EPI_ISL_828178 to 828181, EPI_ISL_828185 to 828187, EPI_ISL_828189, EPI_ISL_828193 to 828199, EPI_ISL_828201 to 828203, EPI_ISL_828206 to 828208, EPI_ISL_828210, EPI_ISL_828213, EPI_ISL_828217 to 828218, EPI_ISL_828221 to 828222, EPI_ISL_828228, EPI_ISL_828230, EPI_ISL_828232 to 828240, EPI_ISL_828245 to 828249, EPI_ISL_828252 to 828254, EPI_ISL_828257 to 828261, EPI_ISL_828263 to 828264, EPI_ISL_828269, EPI_ISL_828271 to 828272, EPI_ISL_828274 to 828282, EPI_ISL_828284, EPI_ISL_828288 to 828304, EPI_ISL_828306 to 828309, EPI_ISL_828311 to 828319, EPI_ISL_828321, EPI_ISL_828328 to 828329, EPI_ISL_828335, EPI_ISL_828337 to 828338, EPI_ISL_828340 to 828351, EPI_ISL_828353 to 828355, EPI_ISL_828357 to 828358, EPI_ISL_828361 to 828362, EPI_ISL_828365 to 828366, EPI_ISL_828368 to 828372, EPI_ISL_828374 to 828382, EPI_ISL_828384 to 828396, EPI_ISL_828403 to 828404, EPI_ISL_828407, EPI_ISL_828409 to 828412, EPI_ISL_828414 to 828417, EPI_ISL_828419 to 828434, EPI_ISL_828436 to 828449, EPI_ISL_828451, EPI_ISL_828453 to 828454, EPI_ISL_828456 to 828463, EPI_ISL_828468, EPI_ISL_828471 to 828479, EPI_ISL_828481 to 828482, EPI_ISL_828488 to 828490, EPI_ISL_828492, EPI_ISL_828496, EPI_ISL_828498 to 828500, EPI_ISL_828508 to 828513, EPI_ISL_828515 to 828516, EPI_ISL_828520 to 828521, EPI_ISL_828524 to 828529, EPI_ISL_828531, EPI_ISL_828533 to 828536, EPI_ISL_828542, EPI_ISL_828546 to 828549, EPI_ISL_828551, EPI_ISL_828553, EPI_ISL_828558 to 828561, EPI_ISL_828563 to 828575, EPI_ISL_828578 to 828584, EPI_ISL_828586 to 828587, EPI_ISL_828590, EPI_ISL_828592, EPI_ISL_828599 to 828600, EPI_ISL_828602 to 828606, EPI_ISL_828608 to 828617, EPI_ISL_828619 to 828626, EPI_ISL_828628 to 828629, EPI_ISL_828632, EPI_ISL_828636 to 828640, EPI_ISL_828645, EPI_ISL_828647 to 828652, EPI_ISL_828654, EPI_ISL_828664 to 828675, EPI_ISL_828678, EPI_ISL_828680 to 828708, EPI_ISL_828710 to 828718, EPI_ISL_828720 to 828725, EPI_ISL_828727 to 828729, EPI_ISL_828731 to 828740, EPI_ISL_828742 to 828743, EPI_ISL_828746 to 828751, EPI_ISL_828760, EPI_ISL_828763 to 828773, EPI_ISL_828777 to 828778, EPI_ISL_828781, EPI_ISL_828783 to 828784, EPI_ISL_828786 to 828789, EPI_ISL_828791 to 828801, EPI_ISL_828803, EPI_ISL_828805 to 828806, EPI_ISL_828809, EPI_ISL_828813 to 828820, EPI_ISL_828822 to 828825, EPI_ISL_828827, EPI_ISL_828830 to 828835, EPI_ISL_828837 to 828842, EPI_ISL_828844 to 828846, EPI_ISL_828848 to 828861, EPI_ISL_828864, EPI_ISL_828866 to 828868, EPI_ISL_828870 to 828872, EPI_ISL_828874 to 828875, EPI_ISL_828877 to 828880, EPI_ISL_828882 to 828888, EPI_ISL_828891, EPI_ISL_828899 to 828903, EPI_ISL_828905 to 828923, EPI_ISL_828925, EPI_ISL_828927 to 828928, EPI_ISL_828933, EPI_ISL_828935 to 828938, EPI_ISL_828941 to 828943, EPI_ISL_828945, EPI_ISL_828947, EPI_ISL_828949 to 828964, EPI_ISL_828967 to 828969, EPI_ISL_828972 to 828975, EPI_ISL_828978 to 828991, EPI_ISL_828998, EPI_ISL_829012 to 829029, EPI_ISL_829031 to 829035, EPI_ISL_829038, EPI_ISL_829040 to 829041, EPI_ISL_829043 to 829045, EPI_ISL_829047 to 829048, EPI_ISL_829051 to 829052, EPI_ISL_829054, EPI_ISL_829056, EPI_ISL_829058, EPI_ISL_829061, EPI_ISL_829063 to 829065, EPI_ISL_829067 to 829072, EPI_ISL_829075 to 829079, EPI_ISL_829082 to 829102, EPI_ISL_829104, EPI_ISL_829106 to 829109, EPI_ISL_829111 to 829115, EPI_ISL_829117 to 829118, EPI_ISL_829120 to 829131, EPI_ISL_829134 to 829141, EPI_ISL_829143, EPI_ISL_829145, EPI_ISL_829148 to 829152, EPI_ISL_829158 to 829164, EPI_ISL_829167, EPI_ISL_829169 to 829178, EPI_ISL_829181 to 829187, EPI_ISL_829191, EPI_ISL_829198, EPI_ISL_829200 to 829219, EPI_ISL_829221 to 829222, EPI_ISL_829225, EPI_ISL_829228 to 829235, EPI_ISL_829240, EPI_ISL_829242 to 829248, EPI_ISL_829252 to 829260, EPI_ISL_829262 to 829272, EPI_ISL_829274, EPI_ISL_829276 to 829278, EPI_ISL_829280 to 829285, EPI_ISL_829287, EPI_ISL_829289, EPI_ISL_829294 to 829295, EPI_ISL_829297 to 829304, EPI_ISL_829306 to 829313, EPI_ISL_829321, EPI_ISL_829324, EPI_ISL_829327 to 829334, EPI_ISL_829336 to 829359, EPI_ISL_829362 to 829363, EPI_ISL_829365, EPI_ISL_829367 to 829368, EPI_ISL_829373 to 829375, EPI_ISL_829380, EPI_ISL_829382 to 829409, EPI_ISL_829411 to 829413, EPI_ISL_829415 to 829425, EPI_ISL_829427 to 829428, EPI_ISL_829430 to 829433, EPI_ISL_829435 to 829445, EPI_ISL_829447 to 829448, EPI_ISL_829450 to 829458, EPI_ISL_829461 to 829463, EPI_ISL_829466 to 829469, EPI_ISL_829472 to 829475, EPI_ISL_829479 to 829484, EPI_ISL_829486 to 829511, EPI_ISL_829513 to 829517, EPI_ISL_829519 to 829523, EPI_ISL_829527 to 829528, EPI_ISL_829530 to 829535, EPI_ISL_829537 to 829542, EPI_ISL_829545 to 829546, EPI_ISL_829548 to 829549, EPI_ISL_829551 to 829557, EPI_ISL_829559 to 829567, EPI_ISL_829569, EPI_ISL_829571 to 829572, EPI_ISL_829574 to 829576, EPI_ISL_829578 to 829580, EPI_ISL_829582 to 829584, EPI_ISL_829586 to 829589, EPI_ISL_829591 to 829598, EPI_ISL_829600, EPI_ISL_829602 to 829605, EPI_ISL_829609 to 829616, EPI_ISL_829618 to 829621, EPI_ISL_829626 to 829628, EPI_ISL_829630 to 829641, EPI_ISL_829643 to 829653, EPI_ISL_829655, EPI_ISL_829657 to 829664, EPI_ISL_829666 to 829670, EPI_ISL_829672 to 829674, EPI_ISL_829676 to 829678, EPI_ISL_829682 to 829685, EPI_ISL_829687 to 829689, EPI_ISL_829691 to 829696, EPI_ISL_829699 to 829702, EPI_ISL_829704 to 829706, EPI_ISL_829710 to 829713, EPI_ISL_829716, EPI_ISL_829718 to 829720, EPI_ISL_829722 to 829724, EPI_ISL_829726 to 829733, EPI_ISL_829736, EPI_ISL_829739 to 829741, EPI_ISL_829744 to 829758, EPI_ISL_829760 to 829766, EPI_ISL_829768 to 829779, EPI_ISL_829781, EPI_ISL_829783 to 829792, EPI_ISL_829794, EPI_ISL_829796, EPI_ISL_829803 to 829806, EPI_ISL_829808 to 829810, EPI_ISL_829812 to 829815, EPI_ISL_829817 to 829837, EPI_ISL_829839 to 829843, EPI_ISL_829845 to 829848, EPI_ISL_829852 to 829854, EPI_ISL_829856 to 829858, EPI_ISL_829861 to 829863, EPI_ISL_829865 to 829868, EPI_ISL_829871 to 829874, EPI_ISL_829877 to 829880, EPI_ISL_829883, EPI_ISL_829886, EPI_ISL_829892 to 829896, EPI_ISL_829898 to 829901, EPI_ISL_829903 to 829906, EPI_ISL_829908, EPI_ISL_829910 to 829913, EPI_ISL_829917, EPI_ISL_829919, EPI_ISL_829921 to 829922, EPI_ISL_829924, EPI_ISL_829926 to 829929, EPI_ISL_829932, EPI_ISL_829935 to 829936, EPI_ISL_829941 to 829943, EPI_ISL_829947, EPI_ISL_829949 to 829951, EPI_ISL_829954, EPI_ISL_829960 to 829969, EPI_ISL_829971, EPI_ISL_829973 to 829976, EPI_ISL_829978 to 829980, EPI_ISL_829983 to 829984, EPI_ISL_829986 to 829987, EPI_ISL_829990 to 830004, EPI_ISL_830006, EPI_ISL_830008 to 830009, EPI_ISL_830011 to 830017, EPI_ISL_830020, EPI_ISL_830022 to 830023, EPI_ISL_830026 to 830032, EPI_ISL_830035 to 830036, EPI_ISL_830039 to 830060, EPI_ISL_830063 to 830066, EPI_ISL_830068, EPI_ISL_830070 to 830071, EPI_ISL_830073 to 830075, EPI_ISL_830077 to 830084, EPI_ISL_830087 to 830092, EPI_ISL_830106 to 830113, EPI_ISL_830115 to 830118, EPI_ISL_830121 to 830124, EPI_ISL_830126, EPI_ISL_830133 to 830139, EPI_ISL_830148 to 830150, EPI_ISL_830153 to 830166, EPI_ISL_830170 to 830176, EPI_ISL_830178 to 830183, EPI_ISL_830187 to 830189, EPI_ISL_830195 to 830197, EPI_ISL_830207, EPI_ISL_830222, EPI_ISL_830224, EPI_ISL_830226, EPI_ISL_830233, EPI_ISL_830247, EPI_ISL_830249, EPI_ISL_830251, EPI_ISL_830256, EPI_ISL_830258, EPI_ISL_830260, EPI_ISL_830262, EPI_ISL_830271, EPI_ISL_830273, EPI_ISL_830275, EPI_ISL_830277, EPI_ISL_830279, EPI_ISL_830281, EPI_ISL_830283, EPI_ISL_830285, EPI_ISL_830287, EPI_ISL_830290, EPI_ISL_830292, EPI_ISL_830295, EPI_ISL_830297, EPI_ISL_830299, EPI_ISL_830301, EPI_ISL_830303, EPI_ISL_830322, EPI_ISL_830324, EPI_ISL_830330, EPI_ISL_830332, EPI_ISL_830336, EPI_ISL_830339, EPI_ISL_830341, EPI_ISL_830343, EPI_ISL_830345, EPI_ISL_830347, EPI_ISL_830350, EPI_ISL_830352, EPI_ISL_830357 to 830358, EPI_ISL_830360, EPI_ISL_830369 to 830372, EPI_ISL_830374, EPI_ISL_830376, EPI_ISL_830383 to 830386, EPI_ISL_830388, EPI_ISL_830392, EPI_ISL_830394 to 830395, EPI_ISL_830398 to 830404, EPI_ISL_830406, EPI_ISL_830408 to 830409, EPI_ISL_830411 to 830412, EPI_ISL_830414, EPI_ISL_830416, EPI_ISL_830418 to 830423, EPI_ISL_830426 to 830427, EPI_ISL_830430 to 830433, EPI_ISL_830435 to 830437, EPI_ISL_830439 to 830444, EPI_ISL_830446 to 830448, EPI_ISL_830450 to 830454, EPI_ISL_830456 to 830458, EPI_ISL_830460 to 830467, EPI_ISL_830469 to 830470, EPI_ISL_830481, EPI_ISL_830483 to 830485, EPI_ISL_830488 to 830491, EPI_ISL_830494 to 830500, EPI_ISL_830503 to 830518, EPI_ISL_830520 to 830528, EPI_ISL_830531 to 830534, EPI_ISL_830536, EPI_ISL_830544, EPI_ISL_830547 to 830548, EPI_ISL_830550, EPI_ISL_830556, EPI_ISL_830561 to 830566, EPI_ISL_830569 to 830570 | | | |
| see above | deCODE genetics | deCODE genetics | Agnar Helgason; Alma Moller; Arna B Agustsdottir; Arnaldur Gylfason; Asgeir Sigurdsson; Aslaug Jonasdottir; Berglind Eiriksdottir; Bjarni Thorbjornsson; Brynjar O Jensson; Daniel F Gudbjartsson; Droplaug N Magnusdottir; Elisabet E Gardarsdottir; Emil A Thorarensen; Gardar Sveinbjornsson; Gisli Masson; Gudmundur Georgsson; Gudmundur L Norddahl; Gudrun Sigmundsdottir; Hakon Jonsson; Hannes Eggertsson; Hilma Holm; Ingileif Jonsdottir; Jona Saemundsdottir; Kamilla S Josefsdottir; Kari Stefansson; Karl G Kristinsson; Kjartan R Gudmundsson; Kristin E Sveinsdottir; Kristin E Sveinsdottir; Louise le Roux; Maney Sveinsdottir; Olafia S Gretarsdottir; Olafia S Gretarsdottir; Olafur T Magnusson; Pall Melsted; Patrick Sulem; Run Fridriksdottir; Solvi Rognvaldsson; Thora R Gunnarsdottir; Thordur Kristjansson; Thorolfur Gudnason; Unnur Thorsteinsdottir |
| EPI_ISL_509017 to 509024, EPI_ISL_509026, EPI_ISL_509028 to 509029, EPI_ISL_509031, EPI_ISL_509033, EPI_ISL_509035, EPI_ISL_509037 to 509038, EPI_ISL_509051, EPI_ISL_732827 to 732881, EPI_ISL_732883 to 732920, EPI_ISL_732923 to 732926 | | | |
| see above | genXone SA, Molecular Diagnostics Laboratory / NZOZ | genXone SA, Research & Development Laboratory | Grzegorz Nowicki; Jakub Grabowski; Maciej Sykulski; Micha Kaszuba; Monika Makowska-Woniak; Natalia Drwska-Matelska; ukasz Krych |
| EPI_ISL_560591 to 560597, EPI_ISL_594163 to 594165 | hopital | National Reference Center for Viruses of Respiratory Infections, Institut Pasteur, Paris | Etienne Simon-Lorière; Fabiana Gambaro; Maud Vanpeene; Sylvie Behillil; Sylvie van der Werf; Vincent Enouf |
| EPI_ISL_693765, EPI_ISL_792039 to 792041, EPI_ISL_792054 to 792055 | hospital | National Reference Center for Viruses of Respiratory Infections, Institut Pasteur, Paris | Angela Brisebarre; Camille Capel; Esther Gyde; Etienne Simon-Lorière; Gisèle Lagathu; Laurent Roudière; Marion Barbet; Maud Vanpeene; Méline Bizard; Patricia Stoessel; Sylvie Behillil; Sylvie van der Werf; Vincent Enouf |
| EPI_ISL_560568 to 560580 | hôpital | National Reference Center for Viruses of Respiratory Infections, Institut Pasteur, Paris | Etienne Simon-Lorière; Fabiana Gambaro; Maud Vanpeene; Sylvie Behillil; Sylvie van der Werf; Vincent Enouf |
| EPI_ISL_430837 | n/a | Thai National Influenza Center, Department of medical Science, Ministry of Public Health, Thailand | Chittaganpitch; Malinee; Okada; Parnmen; Phuygun; Pilailuk; Siripaporn; Sittiporn; Sunthareeya; Thanadachakul; Thanutsapa; Waicharoen; Warawan; Wongboot |
| EPI_ISL_454574 | nstitute for Public Health | Laboratory for advanced genomics | Filip Roki; Igor Jurak; Lovro Trgovec-Greif; Neven Sui; Oliver Vugrek; Tomislav Rukavina |
| EPI_ISL_450816 | orestadsklinikens VC | The Public Health Agency of Sweden | Anna Risberg; Anna-Malin Linde; Karin Tegmark-Wisell; Lisa Kjellberg / Laura Plavitu; Maria Lind Karlberg; Mia Brytting; Olov Svartstrom; Oskar Karlsson Lindsjo; Theresa Enkirch |
| EPI_ISL_447635 to 447653, EPI_ISL_447832 to 447836 | unknown | Department of Medicine | Bampali, M.; Dovrolis, N.; Froukala, E.; Gatzidou, E.; Kassela, K.; N. and Karakasiliotis, I.; Spanakis; Stavropoulou, A.; Tsakris, A.; Veletza, S. |
| EPI_ISL_413584 | unknown | Erasmus Medical Center | Anne van der Linden; Annemiek van der Eijk; Aura Timen; Bas Oude Munnink; Claudia Schapendonk; Corien Swaan; Corine GeurtsvanKessel; David Nieuwenhuijse; Irina Chestakova; Jeroen van Kampen; Jolanda Voermans; Madelief Mollers; Manon Haverkate; Marion Koopmans; Mark Pronk; Mart Stein; Pascal Lexmond; Reina Sikkema; Richard Molenkamp; Sandra Kengne Kamga Mobou; on behalf of the Dutch national COVID-19 response team. |
| EPI_ISL_447656 | unknown | Genomic platform | Canoui-Poitrine, F.; Clinical Study Group, C.; De Prost, N.; Demontant, V.; Deveaux, I.; Fourati, S.; Gricourt, G.; J.-M.; Lamoureux, C.; Lepeule, R.; Mekontso-Dessap, A.; N'debi, M.; P.-L.; Pawlotsky; Picard, O.; Rodrigue, C.; Schmitz, D.; Surgers, L.; Trawinski, E.; Woerther |
| EPI_ISL_453813 to 453837, EPI_ISL_453839 to 453856, EPI_ISL_453859 to 453870, EPI_ISL_453872 to 453879, EPI_ISL_453881, EPI_ISL_453884 to 453899, EPI_ISL_453901, EPI_ISL_453905 to 453906, EPI_ISL_453911 to 453917, EPI_ISL_453920, EPI_ISL_453922, EPI_ISL_453924 to 453927, EPI_ISL_453929, EPI_ISL_453931, EPI_ISL_453933 to 453935, EPI_ISL_453937, EPI_ISL_453939 to 453940, EPI_ISL_453942 to 453944, EPI_ISL_453946 to 453948, EPI_ISL_453951, EPI_ISL_453954, EPI_ISL_453958 to 453959, EPI_ISL_453961 to 453967, EPI_ISL_453969 to 453979, EPI_ISL_453982 to 453990, EPI_ISL_453992 to 453994, EPI_ISL_453996 to 454023, EPI_ISL_454025 to 454026, EPI_ISL_454028 to 454053, EPI_ISL_454055 to 454078, EPI_ISL_454080 to 454083, EPI_ISL_454085 to 454101, EPI_ISL_454103 to 454107, EPI_ISL_454109 to 454124, EPI_ISL_454126 to 454137, EPI_ISL_454139 to 454147, EPI_ISL_454149 to 454150, EPI_ISL_454154, EPI_ISL_454158 to 454160, EPI_ISL_454162 to 454164, EPI_ISL_454167 to 454175, EPI_ISL_454178, EPI_ISL_454181 to 454255, EPI_ISL_454257 to 454272, EPI_ISL_454274 to 454305, EPI_ISL_454307, EPI_ISL_454309 to 454315, EPI_ISL_454317 to 454352, EPI_ISL_455625 to 455627, EPI_ISL_582516 to 582519, EPI_ISL_582521 | | | |
| see above | unknown | Instituto Nacional de Saude (INSA) | Borges et al |
| EPI_ISL_582514 to 582515, EPI_ISL_582522 to 582523 | unknown | Instituto Nacional de Saude (INSA) and Instituto Gulbenkian de Ciencia (IGC) | Borges et al |
| EPI_ISL_462434 to 462438 | unknown | Laboratory Diagnostic | Afonso; Banovic Djeri, B.; C.L.; Dmitric, M.; Jankovic, M.; Knezevic, A.; Petrovic, T.; Sekler, M.; Tesovic, B.; Vidanovic, D.; Volkening, J. |
| EPI_ISL_478669 to 478670 | unknown | Microbiology, Koc University | Can, F.; Dogan, O.; Nurtop, E.; Ozer, B. |
| EPI_ISL_414626, EPI_ISL_415649, EPI_ISL_415651 to 415652 | unknown | National Reference Center for Viruses of Respiratory Infections, Institut Pasteur, Paris | Angela Brisebarre; Flora Donati Vincent Enouf; Marion Barbet; Maud Vanpeene; Méline Bizard; Mélnie Albert; Sylvie Behillil; Sylvie van der Werf |
| EPI_ISL_460618 to 460619 | unknown | Physiology | Caykara, B.; Cevher Keskin, B.; H.H.; Kara, A.; Pence; Pence, S.; Tekin, S.; Yiyit, N. |
| EPI_ISL_660415, EPI_ISL_766614, | unknown | The Public Health Agency of Sweden | Anna Risberg; Anna-Malin Linde; Department of Microbiology; Karin Tegmark-Wisell; Maria Lind Karlberg; Mattias Haukland; Mia Brytting; Olov |

| | | | |
|---|---|---|---|
| EPI_ISL_789046, EPI_ISL_831969 | | | Svartstrom; Oskar Karlsson Lindsjo; Petra Edquist; Reza Advani; Sandra Broddesson; The Public Health Agency of Sweden |
| EPI_ISL_445241 | Ã-restadsklinikens VC | The Public Health Agency of Sweden | Anna Risberg; Anna-Malin Linde; Karin Tegmark-Wisell; Lisa Kjellberg / Laura Plavitu; Maria Lind Karlberg; Mia Brytting; Olov Svartstrom; Oskar Karlsson Lindsjo; Theresa Enkirch |
| EPI_ISL_445238 | Ã-resundslakarna | The Public Health Agency of Sweden | Anna Risberg; Anna-Malin Linde; Del Akrawi; Karin Tegmark-Wisell; Maria Lind Karlberg; Mia Brytting; Olov Svartstrom; Oskar Karlsson Lindsjo; Theresa Enkirch |
